# Stability of Dynamic Radiomics Features in Cardiac MRI

**DOI:** 10.1101/2025.04.18.25326051

**Authors:** Mike D. Klaus, Fabian Laqua, Bettina Baeßler, Markus J. Ankenbrand

## Abstract

**Background:** Radiomic studies on cardiac MR mainly focus on images from distinct time points rather than considering the system’s dynamic nature. Recent studies have shown that radiomic features exhibit considerable variation across the cardiac cycle and that dynamic features can improve classification accuracy in downstream tasks. However, it is unclear whether the dynamic temporal evolution of radiomic features is sufficiently stable in the presence of noise.

**Purpose:** In this work, we evaluate the stability of radiomic feature curves of cine CMR images under noise.

**Methods:** We extracted over 800 radiomic features from all time points of cine CMR images of 35 subjects from three cohorts with various levels of artificially added noise. The stability of feature curves is evaluated based on pairwise normalized mean squared errors, and features are ranked by their stability.

**Results:** Features exhibit a varying degree of stability, but stability is consistent across subjects. Besides generally stable and unstable features, some features are stable within the same noise level but unstable otherwise.

**Conclusion:** Some radiomic feature curves remain stable under noise while showing variability over the cardiac cycle. These features are promising candidates for improving models using dynamic rather than static feature values.

## Introduction

Radiomics is the process of quantifying textural information contained within medical images^1^. The concept behind radiomics is that images generally contain information not visible to the human eye^2,3^. Those features can be used to train machine learning models for detecting, classifying, and prognosis of medical conditions^4^. Radiomics has been successfully applied to many medical imaging modalities, including CT^5^, MRI^6^, and positron emission tomography^7^.

Most papers researching the application of radiomics in cardiac magnetic resonance (CMR) only take the end-systolic (ES) or end-diastolic (ED) images of the cardiac cycle into account for feature extraction^8–10^. However, there is proof that radiomics features differ throughout the cardiac cycle^11,12^. Features calculated on 4D CMR images are more robust than features based on end-systolic or end-diastolic images^11^. Rather than combining features from all frames in a single value, it might be beneficial to consider values from all frames separately^13^.

The way a radiomic feature changes throughout the cardiac cycle could encode valuable information about certain cardiac conditions, especially if the dynamics and function of the heart play a vital role^14^. We refer to radiomic studies that consider the temporal dynamics of a system as *dynamic radiomics* and the sequence of feature values as *feature curves*.

Generally, feature stability and repeatability are a primary concern^15–17^. This limits the usage of radiomics as a prognostic and diagnostic tool since image acquisition settings and scanners differ in different hospitals^18^. Hence, further research is necessary to explore possibilities to ensure reproducible features and thus allow clinical adoption for radiomics workflows^19,20^. The stability problem might be exacerbated when adding a temporal dimension with additional inherent variation. To be useful for downstream tasks, radiomic feature curves must exhibit some stability under noise^21^. So far, it is unclear whether radiomic feature curves of the left ventricular myocardium have this stability in cine CMR.

To answer this question, we analyze the impact of Gaussian noise on the stability of feature curves for 819 radiomic features on 35 subjects with four noise levels.

We hypothesize that some radiomic feature curves are insensitive to noise. Identifying those feature curves should help train more reliable machine learning models to differentiate diseases in medical images.

Besides quantifying the stability of radiomic feature curves for this specific setting, we publish all code as an open-source Snakemake^22^ workflow, thus providing a framework for generally assessing feature curve stability for dynamic radiomics.

## Methods

### 2.1 Study design

This study elucidates how stable the progression of radiomic features in the myocardium over the cardiac cycle (*feature curves*) is at different noise levels (Fig. 1). A combination of simulated and retrospective data analysis is used. Data for this study comes from three sources: (1) simulations with MRXCAT, (2) the public ACDC dataset, and (3) data from a previous study by Baessler et al.^23^ (referred to as BAE). All cine time points of a single central slice are used for each subject. Radiomic features are calculated for every time point. We refer to the sequence of feature values over the cardiac cycle as *feature curves*. The stability of these feature curves under noise is examined.

**Figure 1.**
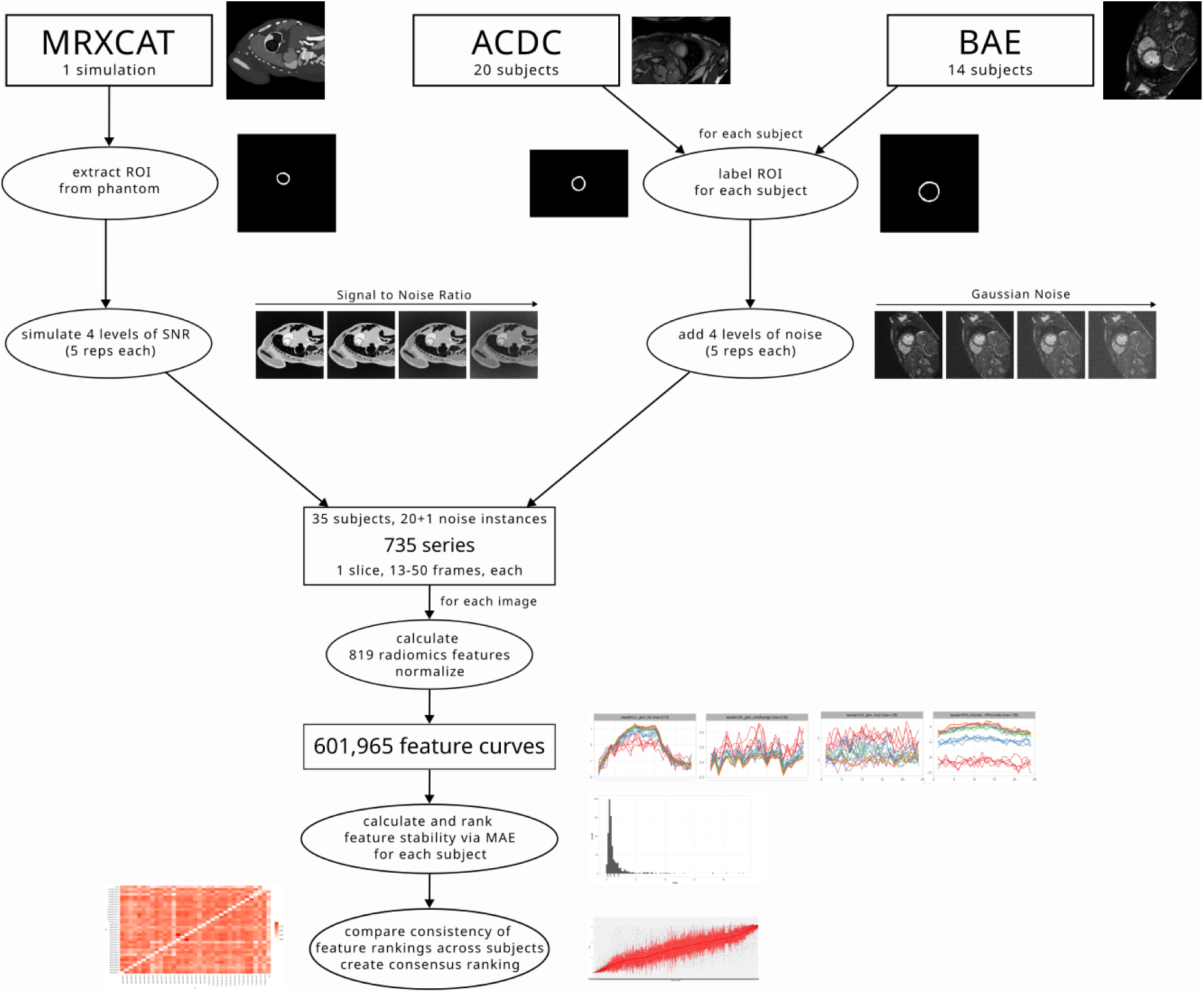
Overview of the data processing for the three data sources

### 2.2 Study populations

The total study population (n=35) consists of the MRXCAT phantom (n=1), the patients from the ACDC dataset (n=20), and the volunteers from the BAE dataset (n=14). We refer to any individual in the study population as a subject, whether a phantom, patient, or volunteer.

### 2.2.1 MRXCAT

MRXCAT^24^ is a program that creates realistic simulations of MR images over the cardiac cycle via an XCAT^25^ phantom of the whole body. It simulates different aspects of MRI acquisition by considering physiological properties and MR parameters. In contrast to actual MRI, we can precisely control the signal-to-noise ratio (SNR). We used MRXCAT to simulate images with the included breath-hold phantom and different SNRs. Five replicates for each SNR value (5, 10, 20, 30) were generated. Another simulation with SNR 50 is considered the reference. The phantom consists of a single slice of 1024 x 1024 pixels over 24 time points along the cardiac cycle. The field of view for the simulations was restricted to the central 512 x 512 pixels. The left ventricular myocardium, excluding papillary muscles, is defined as the region of interest (ROI) and precisely extracted from the phantom.

#### 2.2.2 ACDC

The ACDC dataset was published as part of the Automatic Cardiac Detection Challenge. It consists of cine MRI scans of 150 subjects from clinical exams at the University Hospital of Dijon (France)^26^. We used data from the 20 subjects of the NOR training set. These are individuals with normal cardiac anatomy and function (defined by the authors of ACDC as ejection fraction greater than 50%, wall thickness in diastole lower than 12 mm, LV diastolic volume below 90 mL/m^2^ for men and 80 mL/m^2^ for women, RV volume less than 100 mL/m^2^ and RV ejection fraction above 40%, normal visual analysis of the segmental LV and RV myocardial contraction). The left ventricular myocardium was segmented with nnU-Net^27^ for each subject, and a central slice was selected for further analysis. The number of frames varies between 14 and 35.

#### 2.2.3 BAE

The full BAE dataset consists of 30 healthy volunteers, of which 15 subjects were initially selected for this study, and one subject had to be excluded because automatic segmentation failed, resulting in 14 subjects (8 male, 6 female)^23^. All subjects were healthy and did not suffer from any abnormal cardiac conditions (inclusion criteria: (i) no significant medical history, (ii) no signs of inflammation, (iii) no symptoms indicating cardiovascular dysfunction, and (iv) normal cardiac dimensions and function confirmed by cine CMR. Volunteers with a history of inflammatory disease, including the common cold virus, in the last four weeks before the scans, were excluded). We used short-axis slice stacks with 50 phases per cardiac cycle for each subject. The examinations were conducted on a 1.5T scanner (Achieva 1.5T, Philips Medical Systems, Best, The Netherlands). The imaging parameters used for the 1.5 T short axes stacks were repetition time (TR) 28 ms, echo time (TE) 1.4 ms, flip angle (FA) 60◦, field of view (FOV) 343 × 380 mm^2^, matrix 256 × 256, slice thickness 8 mm, 50 cardiac phases. Each subject’s left ventricular myocardium was segmented with a pre-trained artificial neural network^28^, and a central slice was selected for further analysis.

### 2.3 Noise

For the MRXCAT phantom, a reference (SNR=50) and five replicates for each of the four noise classes with precisely defined SNR (30,20,10,5) were directly simulated. For the real subject data (BAE and ACDC), images with different SNRs were derived by adding artificial (Gaussian) noise with TorchIO^29^. The maximum SNR for each subject was given with the original images. Adding increasing amounts of artificial noise gradually reduces the SNR. Using TorchIO’s RandomNoise function with different values for standard deviation (std), five replicates of MR images for four selected noise levels were generated. Values for std include 0.010, 0.020, 0.030, 0.040. These were chosen by visual inspection of the generated noise levels and intended to be comparable to the noise of the different SNR classes generated in the MRXCAT simulations.

### 2.4 Radiomic feature curves

Texture analysis on all subjects was performed via AutoRadiomics^30^, a radiomics tool for automatic preprocessing, feature selection, modeling, and model evaluation, which uses pyradiomics^31^ for feature extraction. For each subject, all frames of a single central slice with left ventricular myocardium as ROI were used. The default preprocessing of AutoRadiomics for MRI was applied to the data. Features were extracted on raw and wavelet-filtered data in seven categories: shape, first-order intensity statistics, grey level co-occurrence matrix (GLCM), grey level run length matrix (GLRLM), grey level size zone matrix (GLSZM), grey level dependence matrix (GLDM), neighborhood grey tone difference matrix (NGTDM). We excluded shape features from further analysis as they only depend on the region of interest, which was the same for all noise levels. In total, 819 filter-feature combinations were extracted for every frame of each noise instance and subject. The corresponding feature values across frames are combined into feature curves, leading to 601,965 feature curves for the 35 subjects with 21 noise instances and 819 features each (Fig. 1).

### 2.5 Feature curve stability

For comparability, all feature curves for each feature and subject (i.e., across all noise instances) were normalized such that the curve with the least noise (i.e., SNR 50 for MRXCAT and no noise added for ACDC and BAE) had a mean of 0 and standard deviation of 1. Three features were excluded from further analysis (the first-order RootMeanSquared with wavelet filters HHH, HLH, and LHH) because they were constant across the cardiac cycle.

We calculated the pairwise distance of all noise instances for each feature and subject. The distance was measured as the mean absolute error (MAE) of normalized feature curves, calculated with the package similarity measures^32^. The subject-level stability for each feature under noise is given by the mean of pairwise MAE values, with lower values indicating higher stability (0 would be the theoretical maximum stability). Ordering features by these noise values results in a subject-specific feature ranking. To assess the consistency of these rankings, we calculated pairwise correlations of all subjects using the Spearman correlation coefficient. A consensus ranking across subjects was achieved by ordering features by their median rank. A further measure of stability was calculated by restricting the pairwise distances to feature curves of the same noise level. This allows us to identify features sensitive to differences in the amount of noise but not the specific noise itself.

### 2.6 Code and data availability

The whole analysis is implemented as a Snakemake^22^ workflow. All code is available via GitHub (https://github.com/BioMeDS/cmr-dynamic-radiomics-stability) and Zenodo (https://doi.org/10.5281/zenodo.15239556). Large files, including simulation data, result tables, and figures are available as dvcstore via Zenodo (https://doi.org/10.5281/zenodo.15239918). ACDC data is available through their website^26^. BAE data is not publicly available.

## Results

As expected, the radiomic features show variability over the cardiac cycle. The extent to which the resulting feature curves for the reference images (SNR 50, no additional noise) remain stable under noise varies across features and subjects. The stability of features for the MRXCAT phantom, as measured by the mean pairwise MAEs of the normalized curves (mpMAE), ranges from 0 to 24, with most features having values below 2.5 (Fig. 2A). For mpMAE values below 1, the cases with lower SNR well reproduced the pattern of the reference feature curve, while the SNR 5 cases show the highest variability (Fig. 2B, C). For values above 1, there are apparent deviations from the reference curve. Some curves show high variability around the original values (Fig. 2D). In contrast, others have an explicit dependency on the amount of noise; thus, curves from the same SNR level are consistent, but they are shifted compared to other SNR levels and the reference curve (Fig. 2E). To quantify this, we calculated a noise-level dependent score as the mpMAE between all pairs of the same SNR/noise level.

**Figure 2.**
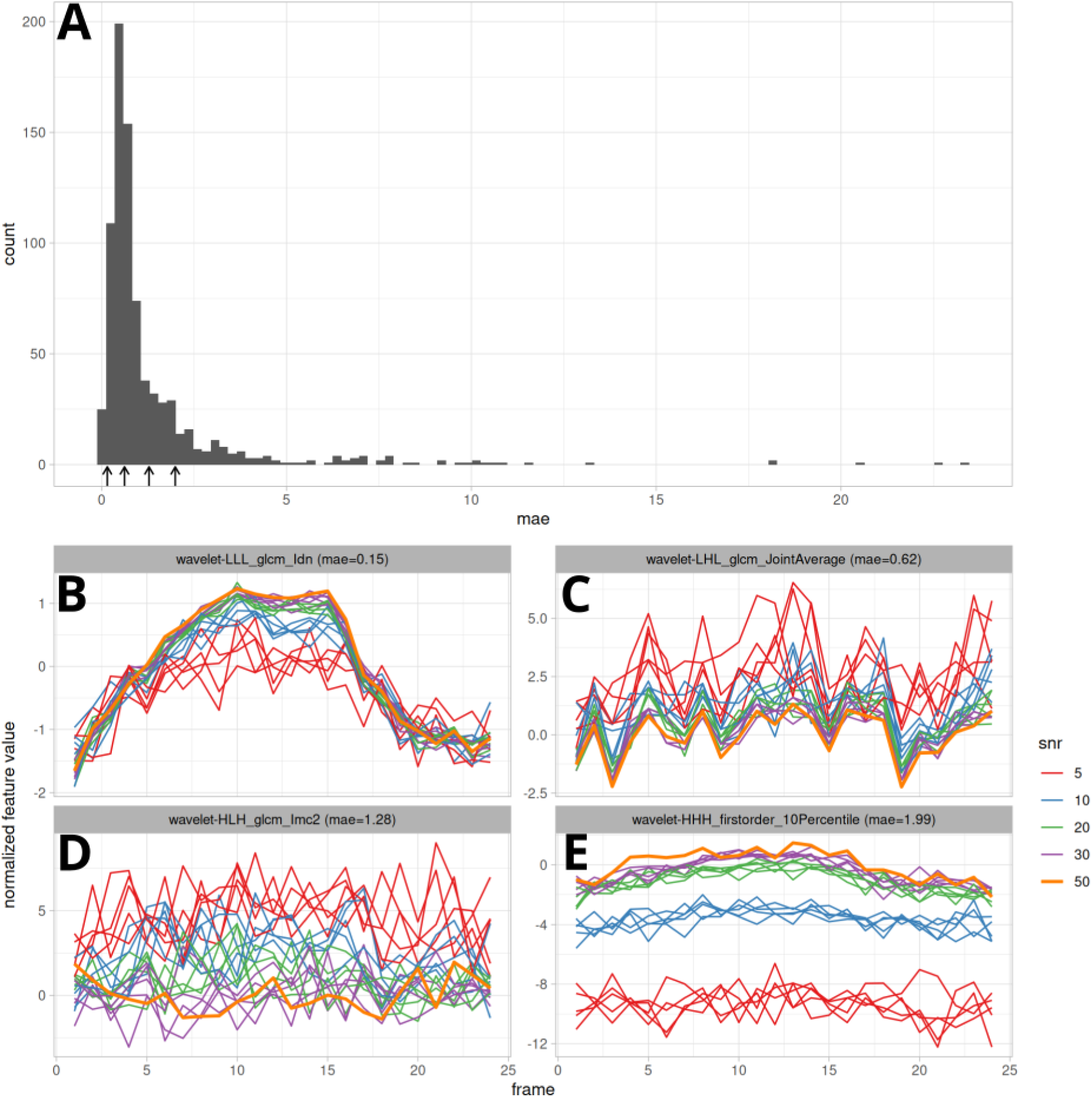
MAE score distribution for the MRXCAT simulation (A) and exemplary feature curves for a range of MAE values (B-E). Feature curves were processed and plotted via tidyverse^33^ and ggplot2^34^ in R^35^.

Comparing all feature curves for a given feature across subjects shows that feature curves and their stability are similar for subjects from all three sets (Fig. 3 and Supplementary File S1).

**Figure 3.**
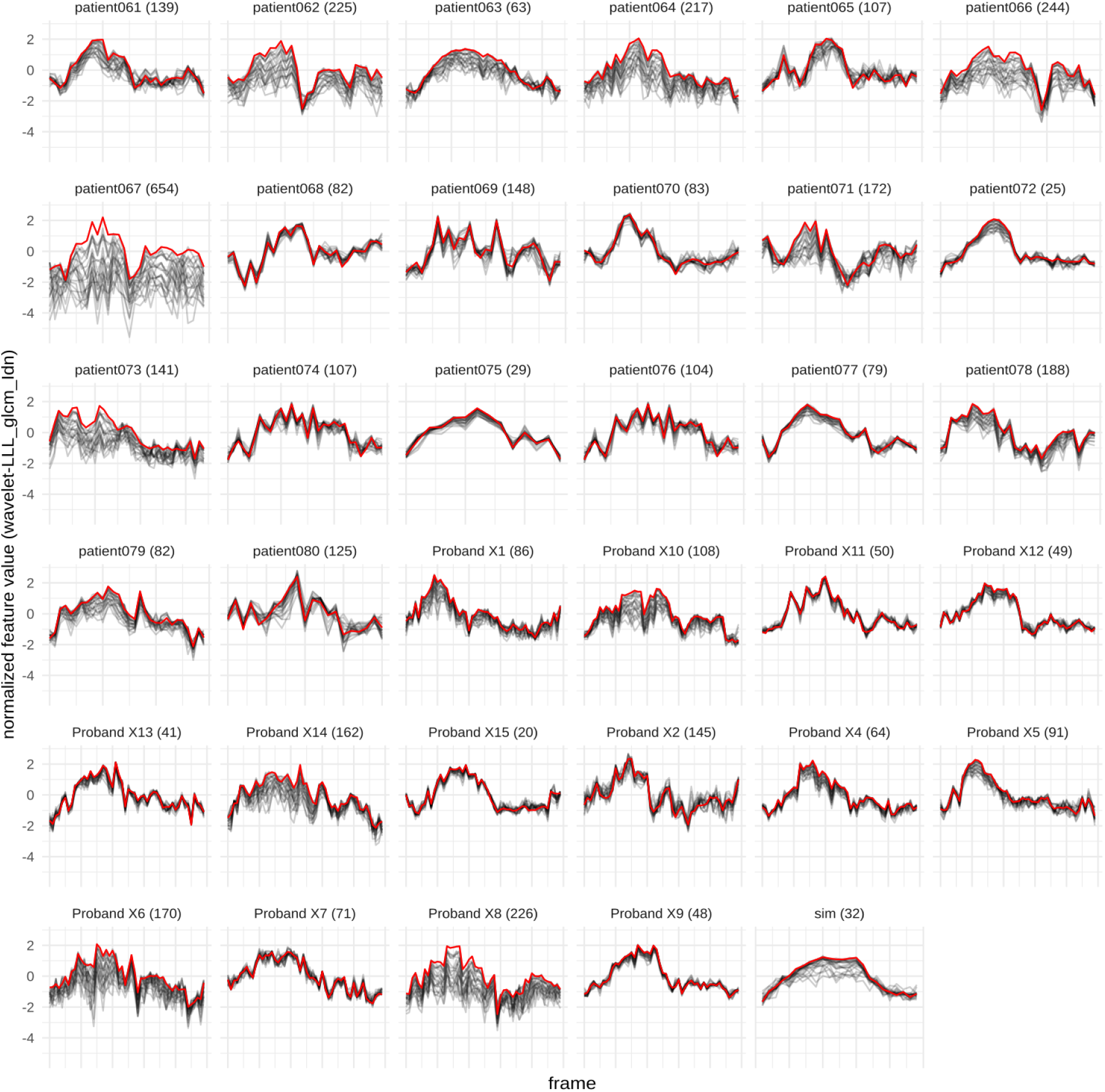
Feature curves for wavelet-LLL_glcm_ldn for all subjects. Reference curve (SNR50/no noise added) are shown in red, all noisy curves are shown in gray. The median rank for this feature is 80, the rank for each subject is printed in parentheses. Subjects starting with patient are from ACDC, subjects starting with Proband are from BAE and sim is the MRXCAT simulation. Plots for all 816 features are in supplementary file S1

The correlation coefficients range from -0.06 to 1.00, with a median Spearman correlation coefficient of 0.59 (Fig. 4). A consensus ranking of all features was created by ordering features by their median rank (Supp. Fig. 1). In this global ranking, features from all categories show varying stability; however, features from category *glszm* tend to be more stable than features from category *firstorder* (Fig. 6).

**Figure 4.**
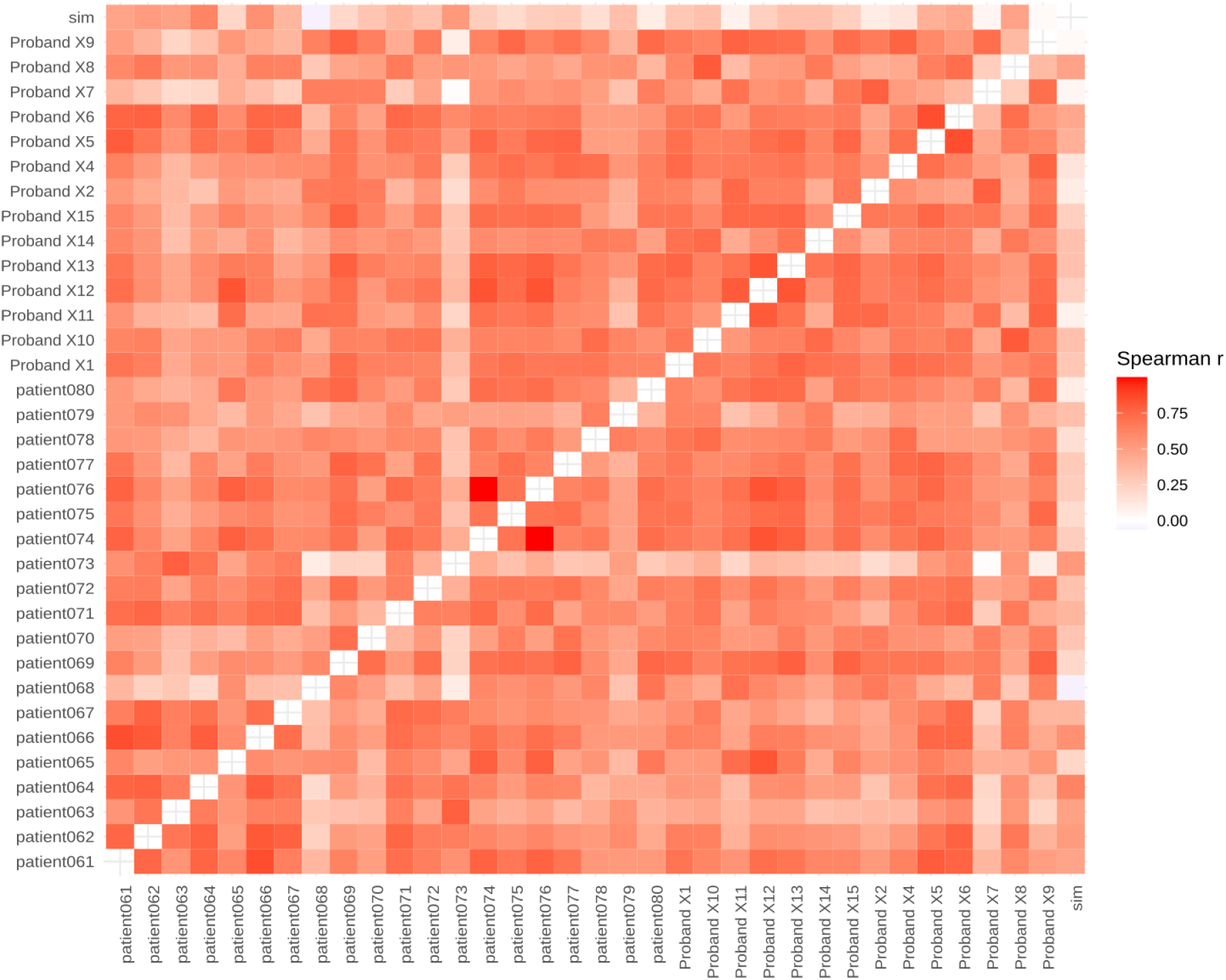
Pairwise Spearman correlation coefficient of feature stability scores (mae). White indicates no correlation, and dark red indicates a high correlation. The diagonal is empty as the correlation between a subject and itself is trivially 1.

**Figure 5.**
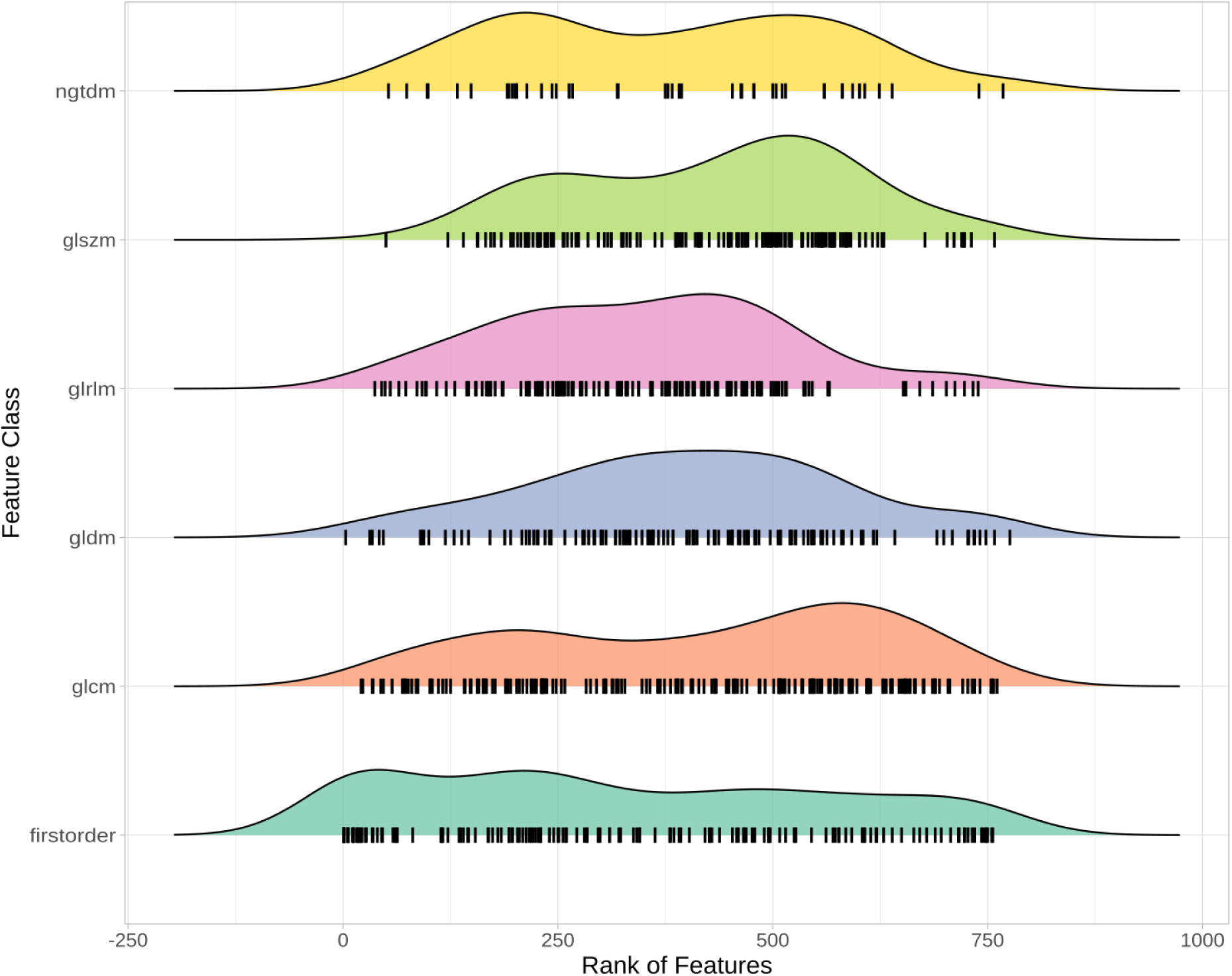
Distribution of consensus ranks across feature classes. Each feature is drawn as a vertical black bar with density plots drawn above.

In addition to the consensus rank for each feature, the median mpMAE across subjects and the rank and median score for the noise-level dependent distance are reported (Supp. Tab. 1). Comparing overall mpMAE and noise-level dependent mpMAE reveals features sensitive to the amount of noise but less so to the noise itself (Supp Fig. 2).

## Discussion

As previously demonstrated, radiomic features vary along the cardiac cycle^11^. Thus, considering the entire curve rather than limiting the analysis to distinct time points might increase the power. However, to be considered valuable inputs for predictive models, these feature curves must be robust to small perturbations, e.g., random noise^21^. This work aimed to determine if the curves of any radiomic features are sufficiently stable under noise. We used data from 35 subjects and added four levels of noise. The resulting feature curves and mean pairwise MAE scores show that many features are robust to even high noise levels (Fig. 2AB, Fig. 3).

In contrast, other features are susceptible to noise or at least the amount of noise (Fig. 2ADE). The observation that feature curves show different noise sensitivity led to the question of whether this noise sensitivity is subject-dependent. Indeed, mpMAE scores and ranks differ between subjects, in some cases considerably (e.g., Fig. 3, patient067). However, overall, the rankings are consistent, with a median Spearman correlation coefficient of 0.59 across all pairwise comparisons (Fig. 4). Correlations are similarly strong between subjects of the same and different cohorts (Fig. 4). Therefore, our consensus scoring and ranking of radiomic features by their curve stability under noise (Supp. Tab. 1) can serve as the foundation for feature selection in dynamic radiomics studies. The stability of feature curves under noise is a prerequisite for further use in machine learning models. However, it does not guarantee that these feature curves improve the performance compared to single feature values. This question can now be addressed in follow-up studies.

An interesting observation is that some features show high sensitivity to the amount of noise in the image while being relatively robust to different noise instances (Fig. 2E, Supp. Fig. 2). These features can be considered in studies with uniform noise levels but should be avoided if some elements in the data set have higher levels of noise than others.

To allow users maximum flexibility in selecting the relevant features for their dynamic radiomics studies with LV myocardium on CMR, we provide overall and noise-level dependent rankings and scores for all radiomic features (Supp. Tab. 1).

All our code and most of the data (except for BAE) are publicly available, thus allowing anyone to reproduce our findings and apply this approach to different datasets, regions of interest, or modalities. Hence, besides the concrete results for feature curve stability in LV myocardium in CMR, we provide a general framework for studying noise sensitivity in dynamic radiomics.

## Limitations

Several limitations of this study merit consideration. As with all observational studies, no causal conclusions could be drawn from the data. Hence, results from the non-experimental study design should be interpreted as hypothesis-generating. The scope of this study is limited to the stability of radiomic feature curves for a central slice of the left ventricular myocardium on cardiac magnetic resonance images of healthy subjects to Gaussian noise. In particular, only within-subject stability was considered. This study is limited to simulated noise that might not resemble noise in a realistic setting. The approach must be applied to scan-rescan data to ensure realistic noise. The results of this study do not indicate the diagnostic or prognostic value of these feature curves.

The variability of features and, thus, feature curves is expected to change in response to different acquisition methods and scanners and might require additional harmonization^36^. While the reported feature curve stability might not be transferable to data from other hospitals, the described methodology can be applied to re-evaluate the features in these settings.

## Conclusion

We show that for many radiomic features, the way they vary along the cardiac cycle is stable under noise. This is a prerequisite for these feature curves to be considered as inputs to classification or regression models. We score and rank all features by overall and noise-level dependent stability based on one phantom and 34 subjects from two data sources. This table can be used to select features in follow-up studies utilizing dynamic radiomics. Exploring the potential incremental predictive value of feature curves over single features in supervised machine-learning tasks and medical conditions will be the subject of future research.

## Data Availability

All code is available via GitHub (https://github.com/BioMeDS/cmr-dynamic-radiomics-stability) and Zenodo (https://doi.org/10.5281/zenodo.15239556). Large files, including simulation data, result tables, and figures are available as dvcstore via Zenodo (https://doi.org/10.5281/zenodo.15239918). ACDC data is available through their website. BAE data is not publicly available.

https://doi.org/10.5281/zenodo.15239556

https://doi.org/10.5281/zenodo.15239918

https://www.creatis.insa-lyon.fr/Challenge/acdc/index.html

## Supplementary Figures

**Supplementary Figure 1.**
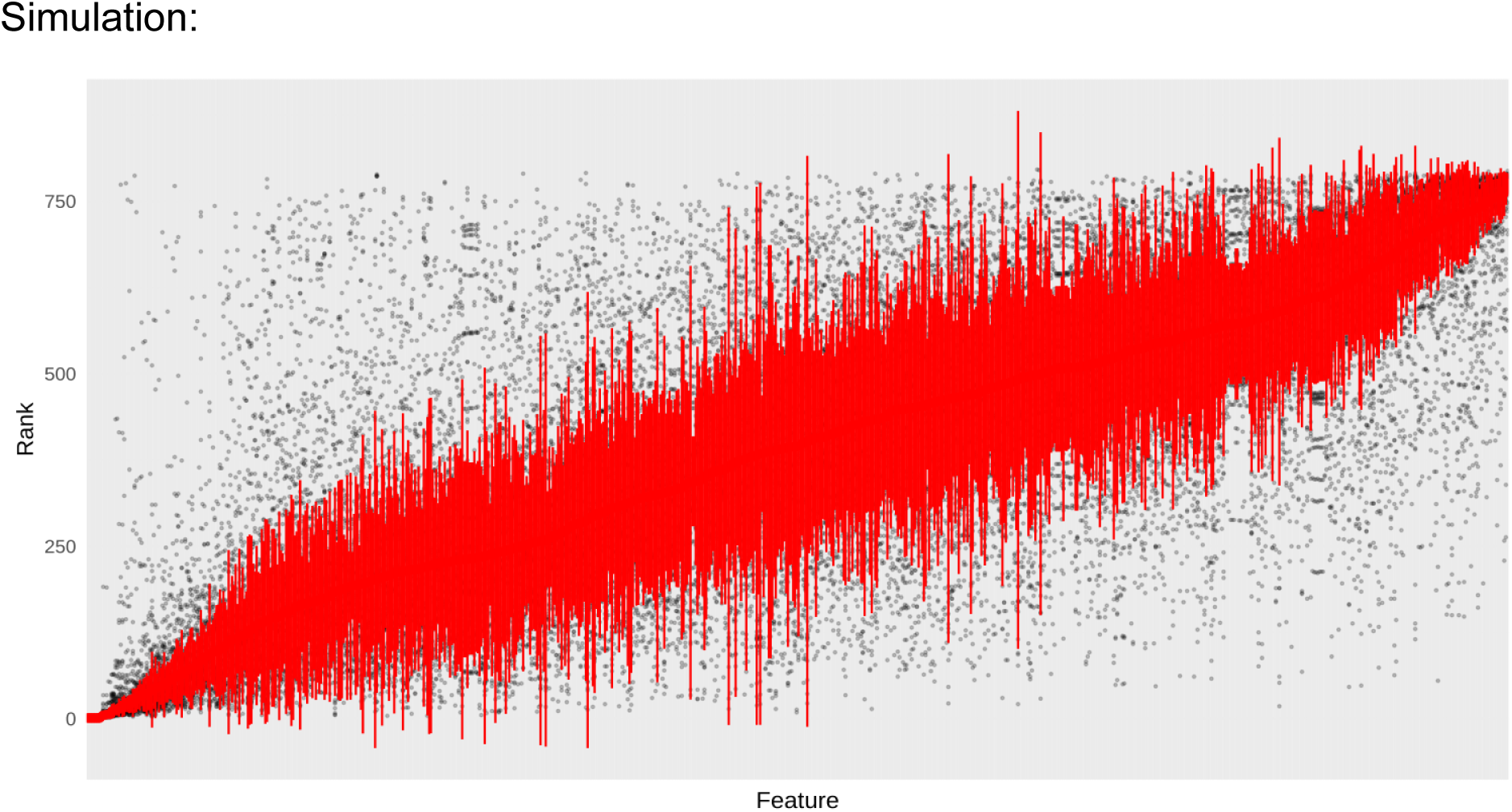
Consensus ranking of features. Features are on the x-axis, ordered by median rank (median and mad shown in red), and individual ranks for each subject are shown as gray dots.

**Supplementary Figure 2.**
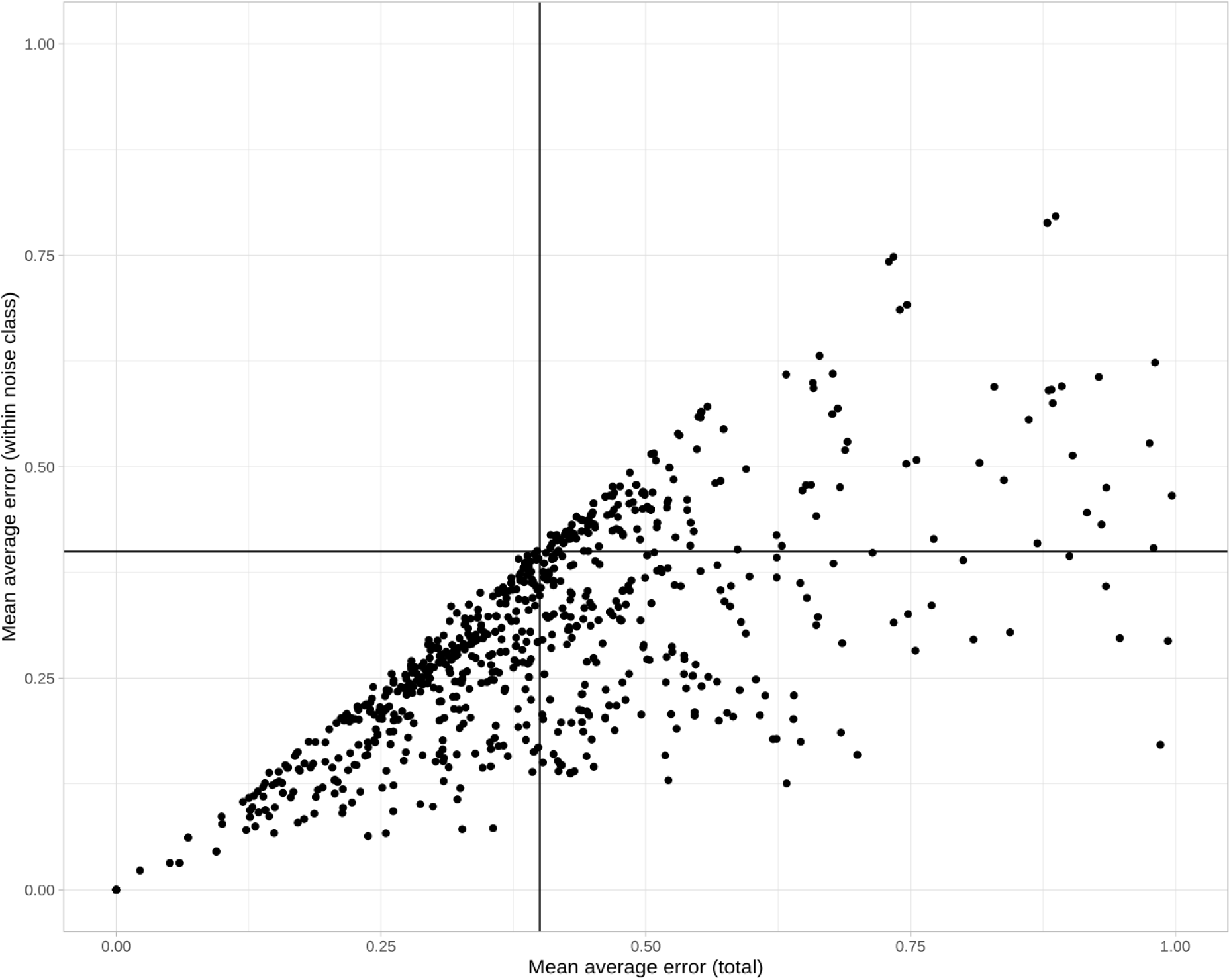
Mean mae within noise level compared to mean mae across all noise levels. Each dot represents a feature. Range of x- and y-axis restricted to 0.0-1.0 (33 features are out of range and therefore not shown)

