## Supplemental File S1 for "Stability of Dynamic Radiomics Features in Cardiac MRI"

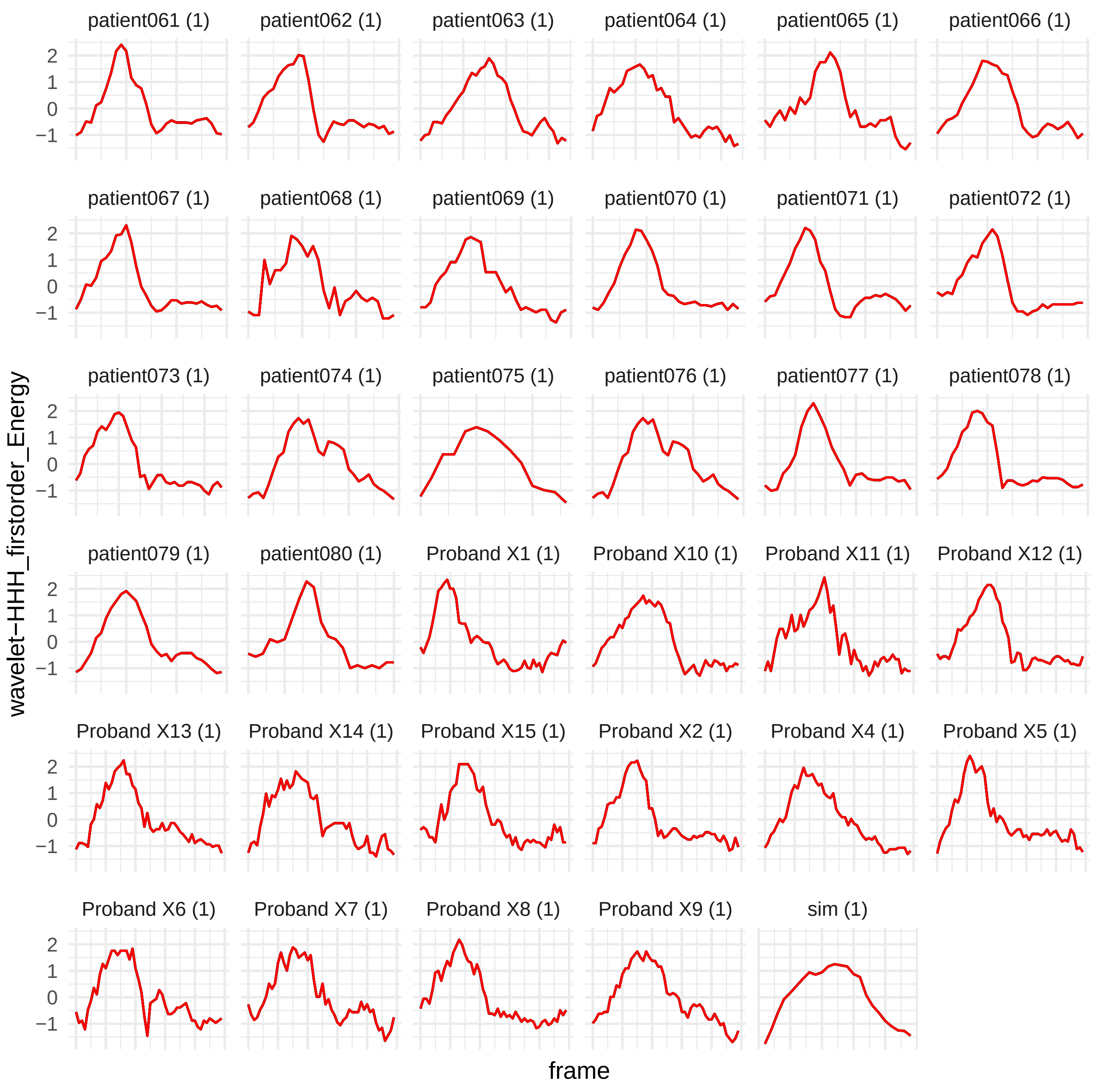

patient061 (1)
patient062 (1)
patient063 (1)
patient064 (1)
patient065 (1)
patient066 (1)
patient067 (1)
patient068 (1)
patient069 (1)
patient070 (1)
patient071 (1)
patient072 (1)
patient073 (1)
patient074 (1)
patient075 (1)
patient076 (1)
patient077 (1)
patient078 (1)
patient079 (1)
patient080 (1)
Proband X1 (1)
Proband X10 (1)
Proband X11 (1)
Proband X12 (1)
Proband X13 (1)
Proband X14 (1)
Proband X15 (1)
Proband X2 (1)
Proband X4 (1)
Proband X5 (1)
Proband X6 (1)
Proband X7 (1)
Proband X8 (1)
Proband X9 (1)
sim (1)
2
1
0
−1
wavelet−HHH_firstorder_Energy
frame

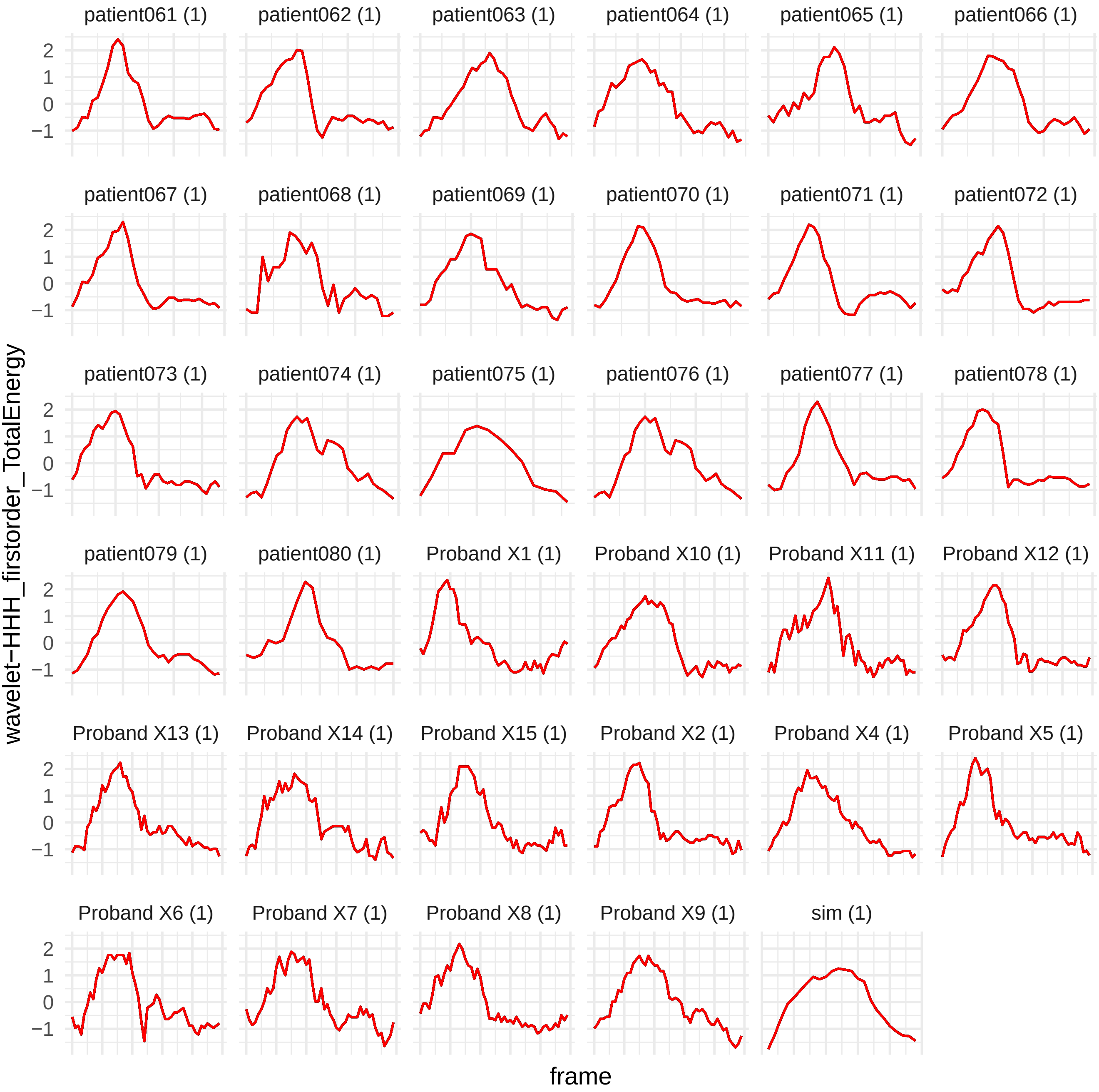
patient061 (1)
patient062 (1)
patient063 (1)
patient064 (1)
patient065 (1)
patient066 (1)
patient067 (1)
patient068 (1)
patient069 (1)
patient070 (1)
patient071 (1)
patient072 (1)
patient073 (1)
patient074 (1)
patient075 (1)
patient076 (1)
patient077 (1)
patient078 (1)
patient079 (1)
patient080 (1)
Proband X1 (1)
Proband X10 (1)
Proband X11 (1)
Proband X12 (1)
Proband X13 (1)
Proband X14 (1)
Proband X15 (1)
Proband X2 (1)
Proband X4 (1)
Proband X5 (1)
Proband X6 (1)
Proband X7 (1)
Proband X8 (1)
Proband X9 (1)
sim (1)
2
1
0
−1
wavelet−HHH_firstorder_TotalEnergy
frame

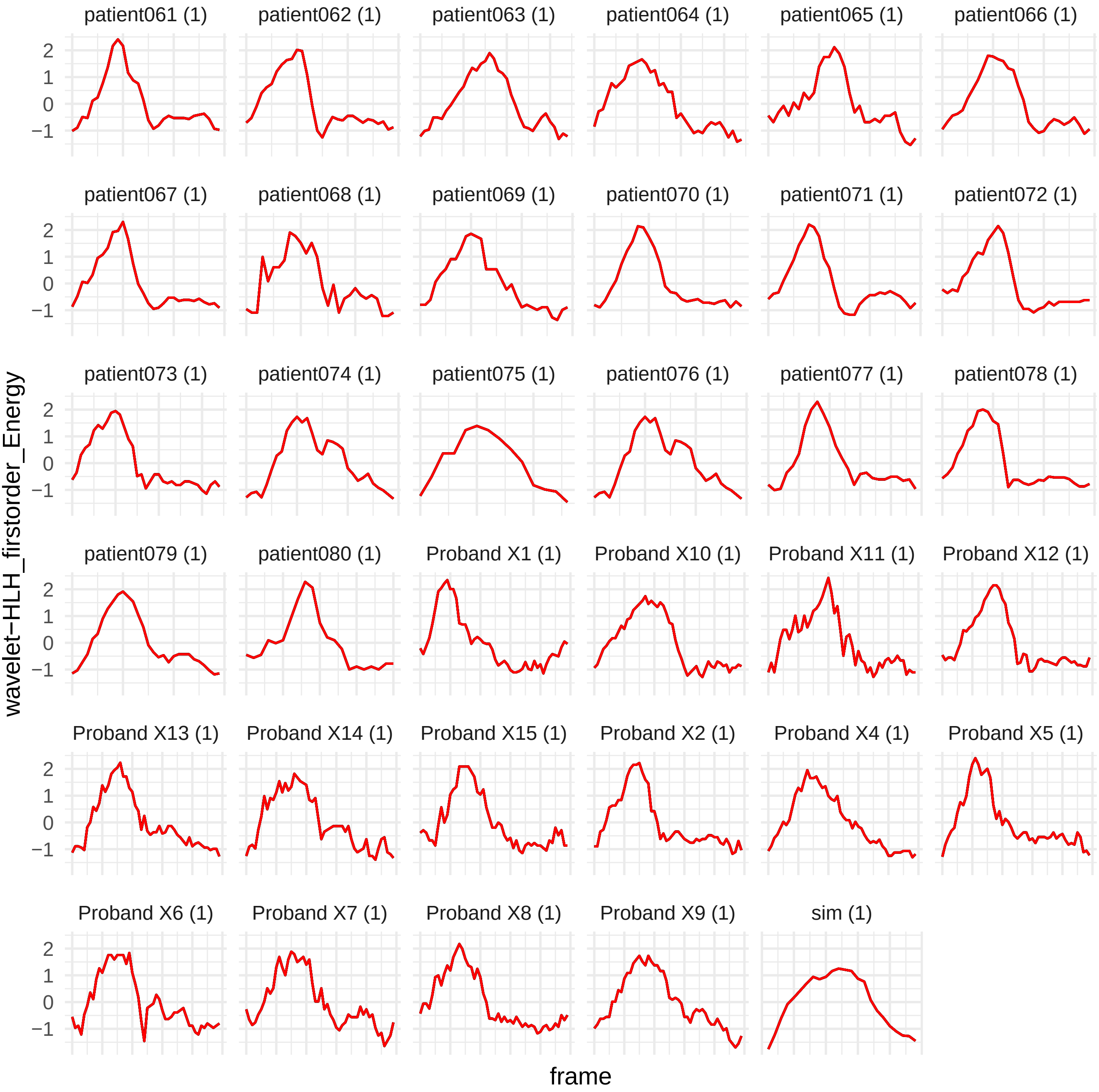

patient061 (1)
patient062 (1)
patient063 (1)
patient064 (1)
patient065 (1)
patient066 (1)
patient067 (1)
patient068 (1)
patient069 (1)
patient070 (1)
patient071 (1)
patient072 (1)
patient073 (1)
patient074 (1)
patient075 (1)
patient076 (1)
patient077 (1)
patient078 (1)
patient079 (1)
patient080 (1)
Proband X1 (1)
Proband X10 (1)
Proband X11 (1)
Proband X12 (1)
Proband X13 (1)
Proband X14 (1)
Proband X15 (1)
Proband X2 (1)
Proband X4 (1)
Proband X5 (1)
Proband X6 (1)
Proband X7 (1)
Proband X8 (1)
Proband X9 (1)
sim (1)
wavelet−HLH_firstorder_Energy
frame
2
1
0
−1

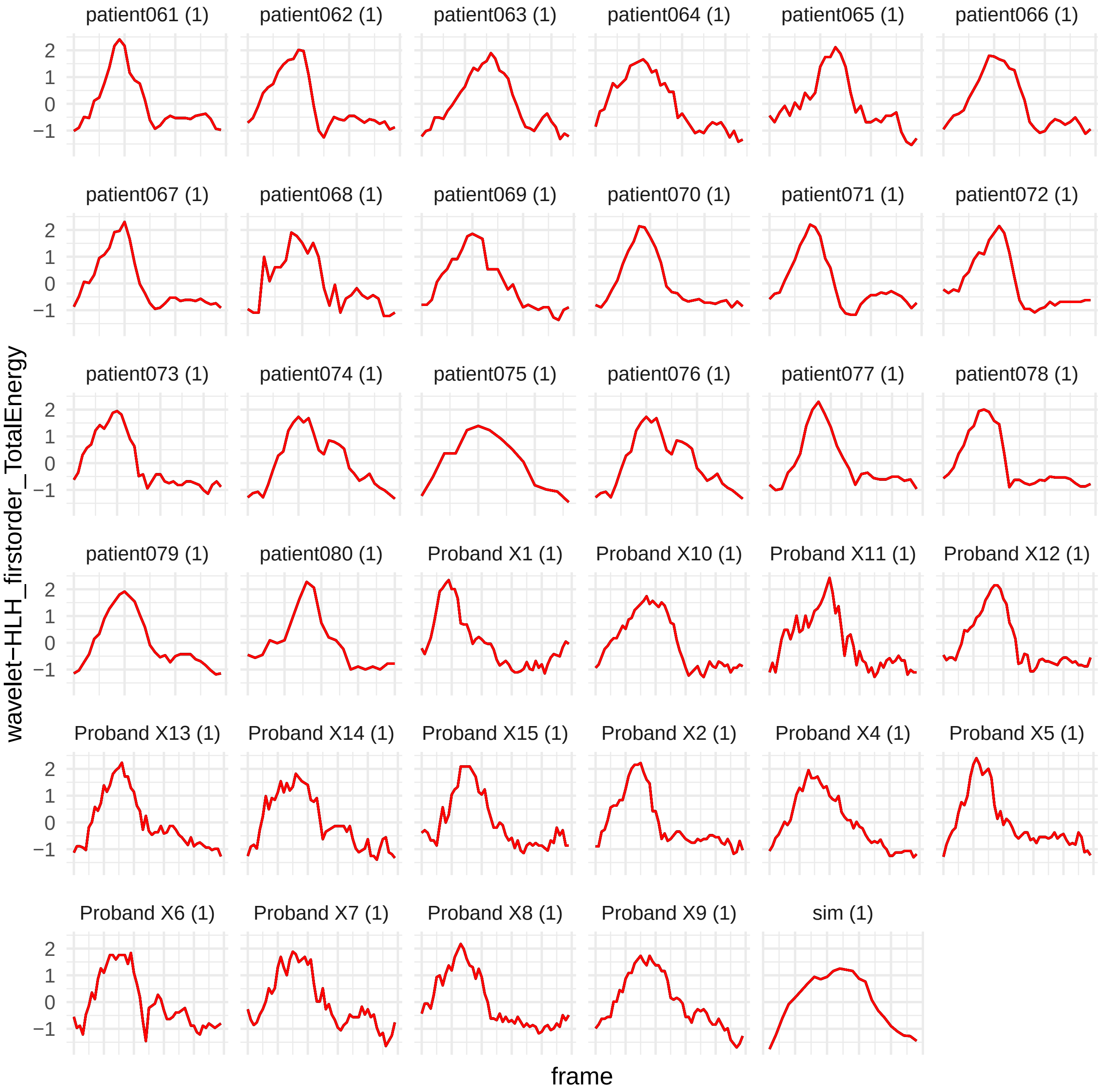

patient061 (1)
patient062 (1)
patient063 (1)
patient064 (1)
patient065 (1)
patient066 (1)
patient067 (1)
patient068 (1)
patient069 (1)
patient070 (1)
patient071 (1)
patient072 (1)
patient073 (1)
patient074 (1)
patient075 (1)
patient076 (1)
patient077 (1)
patient078 (1)
patient079 (1)
patient080 (1)
Proband X1 (1)
Proband X10 (1)
Proband X11 (1)
Proband X12 (1)
Proband X13 (1)
Proband X14 (1)
Proband X15 (1)
Proband X2 (1)
Proband X4 (1)
Proband X5 (1)
Proband X6 (1)
Proband X7 (1)
Proband X8 (1)
Proband X9 (1)
sim (1)
2
1
0
−1
wavelet−HLH_firstorder_TotalEnergy
frame

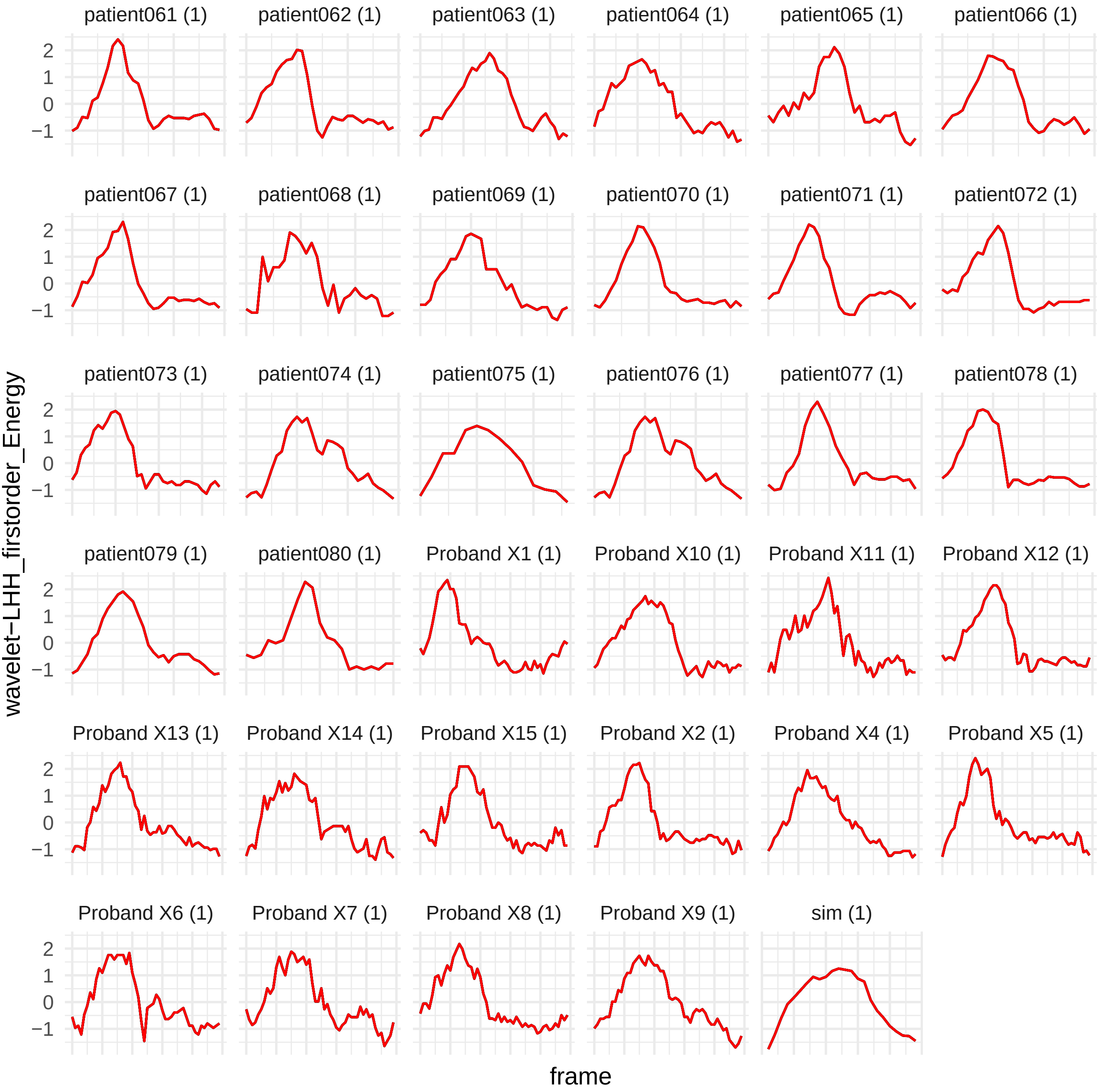

patient061 (1)
patient062 (1)
patient063 (1)
patient064 (1)
patient065 (1)
patient066 (1)
patient067 (1)
patient068 (1)
patient069 (1)
patient070 (1)
patient071 (1)
patient072 (1)
patient073 (1)
patient074 (1)
patient075 (1)
patient076 (1)
patient077 (1)
patient078 (1)
patient079 (1)
patient080 (1)
Proband X1 (1)
Proband X10 (1)
Proband X11 (1)
Proband X12 (1)
Proband X13 (1)
Proband X14 (1)
Proband X15 (1)
Proband X2 (1)
Proband X4 (1)
Proband X5 (1)
Proband X6 (1)
Proband X7 (1)
Proband X8 (1)
Proband X9 (1)
sim (1)
2
1
0
−1
wavelet−LHH_firstorder_Energy
frame

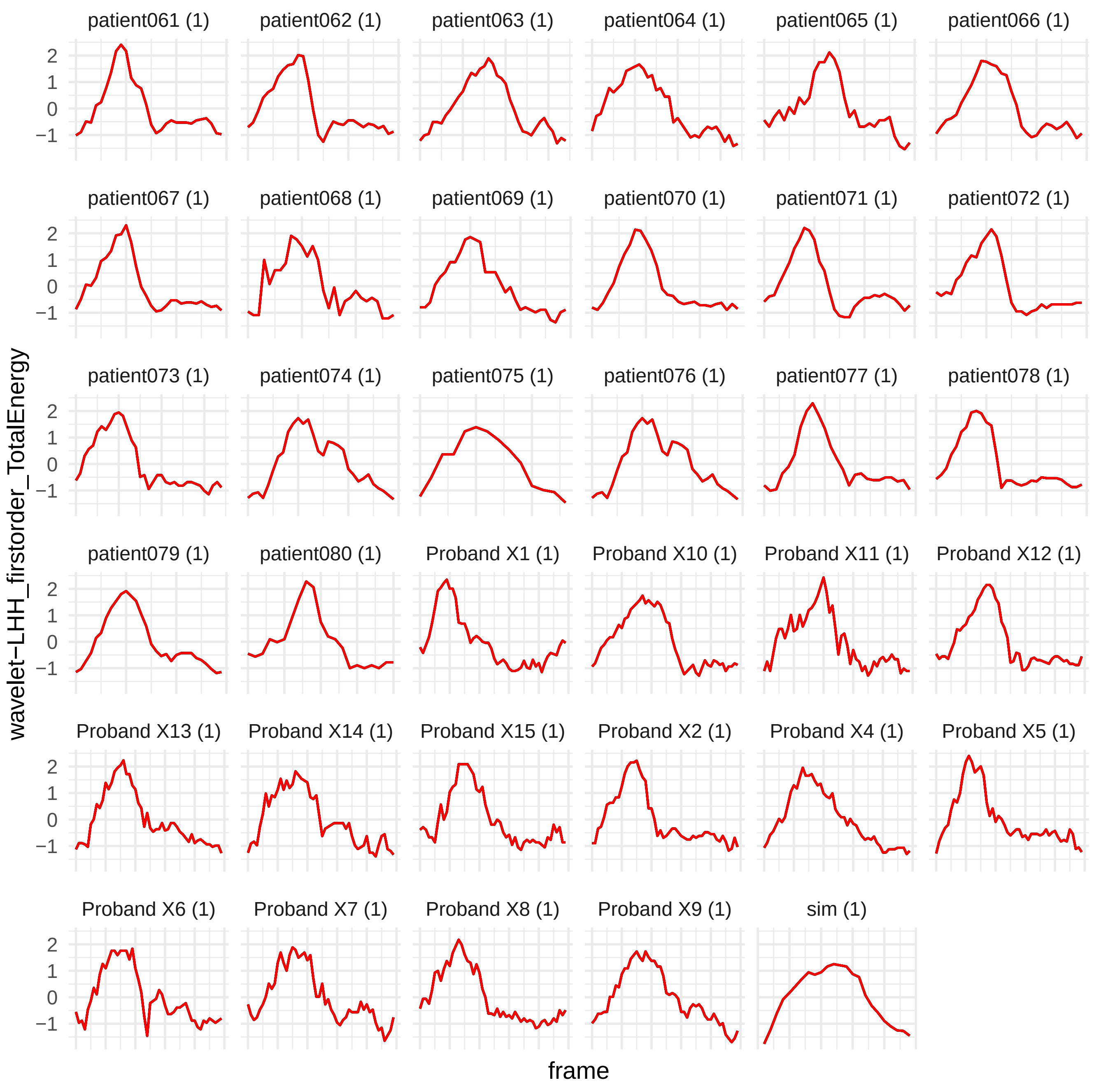
patient061 (1)
patient062 (1)
patient063 (1)
patient064 (1)
patient065 (1)
patient066 (1)
patient067 (1)
patient068 (1)
patient069 (1)
patient070 (1)
patient071 (1)
patient072 (1)
patient073 (1)
patient074 (1)
patient075 (1)
patient076 (1)
patient077 (1)
patient078 (1)
patient079 (1)
patient080 (1)
Proband X1 (1)
Proband X10 (1)
Proband X11 (1)
Proband X12 (1)
Proband X13 (1)
Proband X14 (1)
Proband X15 (1)
Proband X2 (1)
Proband X4 (1)
Proband X5 (1)
Proband X6 (1)
Proband X7 (1)
Proband X8 (1)
Proband X9 (1)
sim (1)
2
1
0
−1
wavelet−LHH_firstorder_TotalEnergy
frame

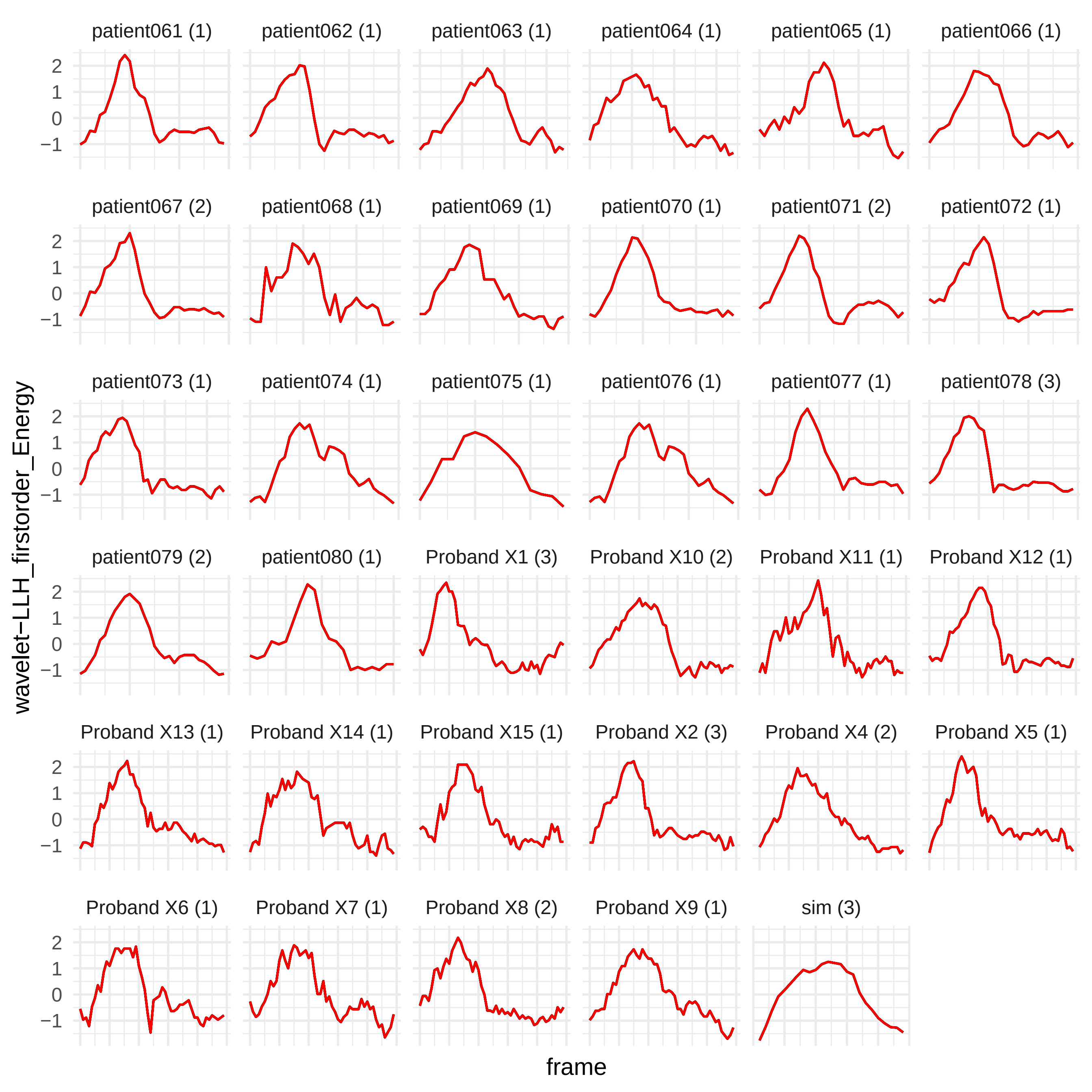

patient061 (1)
patient062 (1)
patient063 (1)
patient064 (1)
patient065 (1)
patient066 (1)
patient067 (2)
patient068 (1)
patient069 (1)
patient070 (1)
patient071 (2)
patient072 (1)
patient073 (1)
patient074 (1)
patient075 (1)
patient076 (1)
patient077 (1)
patient078 (3)
patient079 (2)
patient080 (1)
Proband X1 (3)
Proband X10 (2)
Proband X11 (1)
Proband X12 (1)
Proband X13 (1)
Proband X14 (1)
Proband X15 (1)
Proband X2 (3)
Proband X4 (2)
Proband X5 (1)
Proband X6 (1)
Proband X7 (1)
Proband X8 (2)
Proband X9 (1)
sim (3)
2
1
0
−1
wavelet−LLH_firstorder_Energy
frame

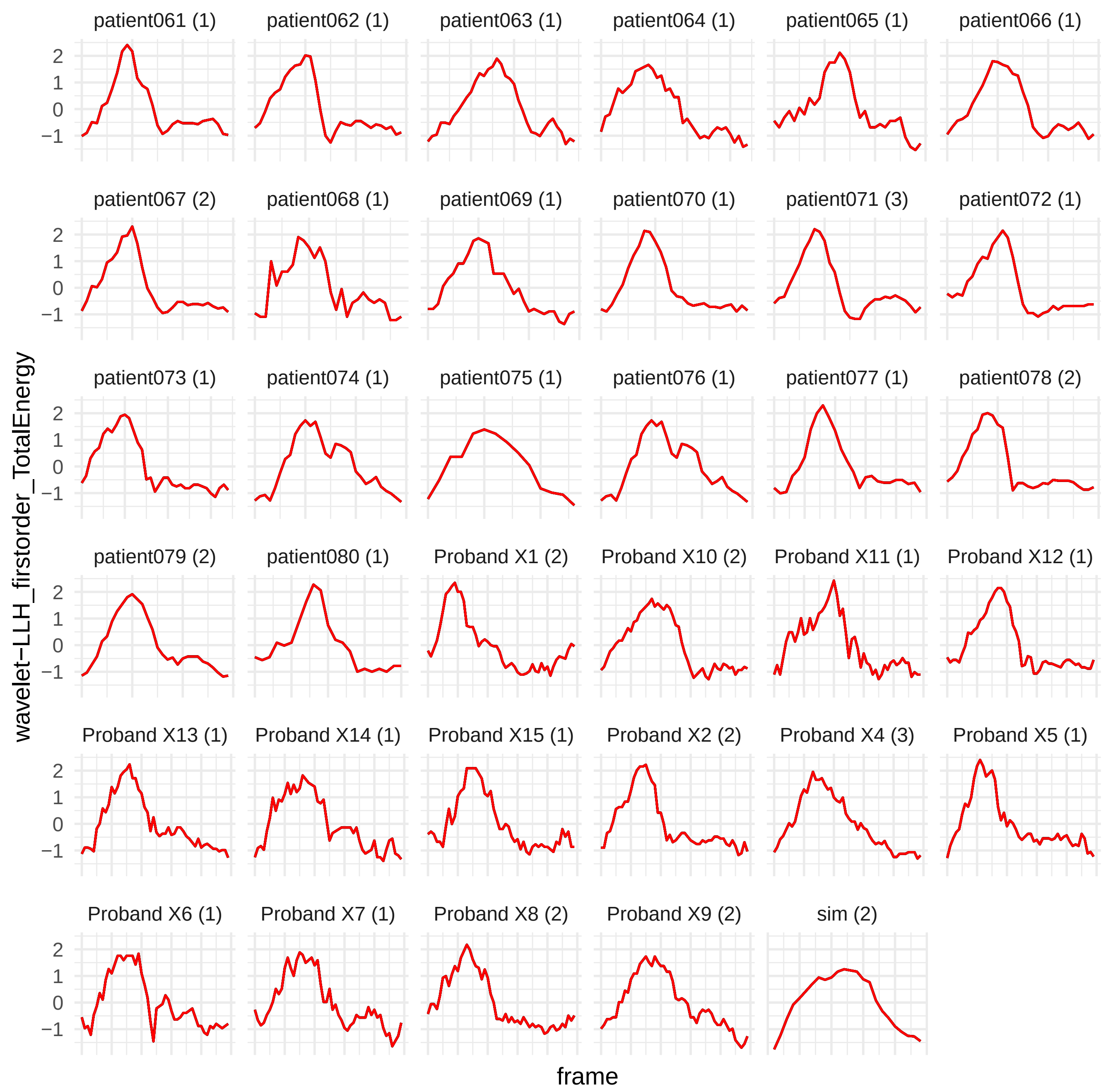
patient061 (1)
patient062 (1)
patient063 (1)
patient064 (1)
patient065 (1)
patient066 (1)
patient067 (2)
patient068 (1)
patient069 (1)
patient070 (1)
patient071 (3)
patient072 (1)
patient073 (1)
patient074 (1)
patient075 (1)
patient076 (1)
patient077 (1)
patient078 (2)
patient079 (2)
patient080 (1)
Proband X1 (2)
Proband X10 (2)
Proband X11 (1)
Proband X12 (1)
Proband X13 (1)
Proband X14 (1)
Proband X15 (1)
Proband X2 (2)
Proband X4 (3)
Proband X5 (1)
Proband X6 (1)
Proband X7 (1)
Proband X8 (2)
Proband X9 (2)
sim (2)
wavelet−LLH_firstorder_TotalEnergy
frame
2
1
0
−1

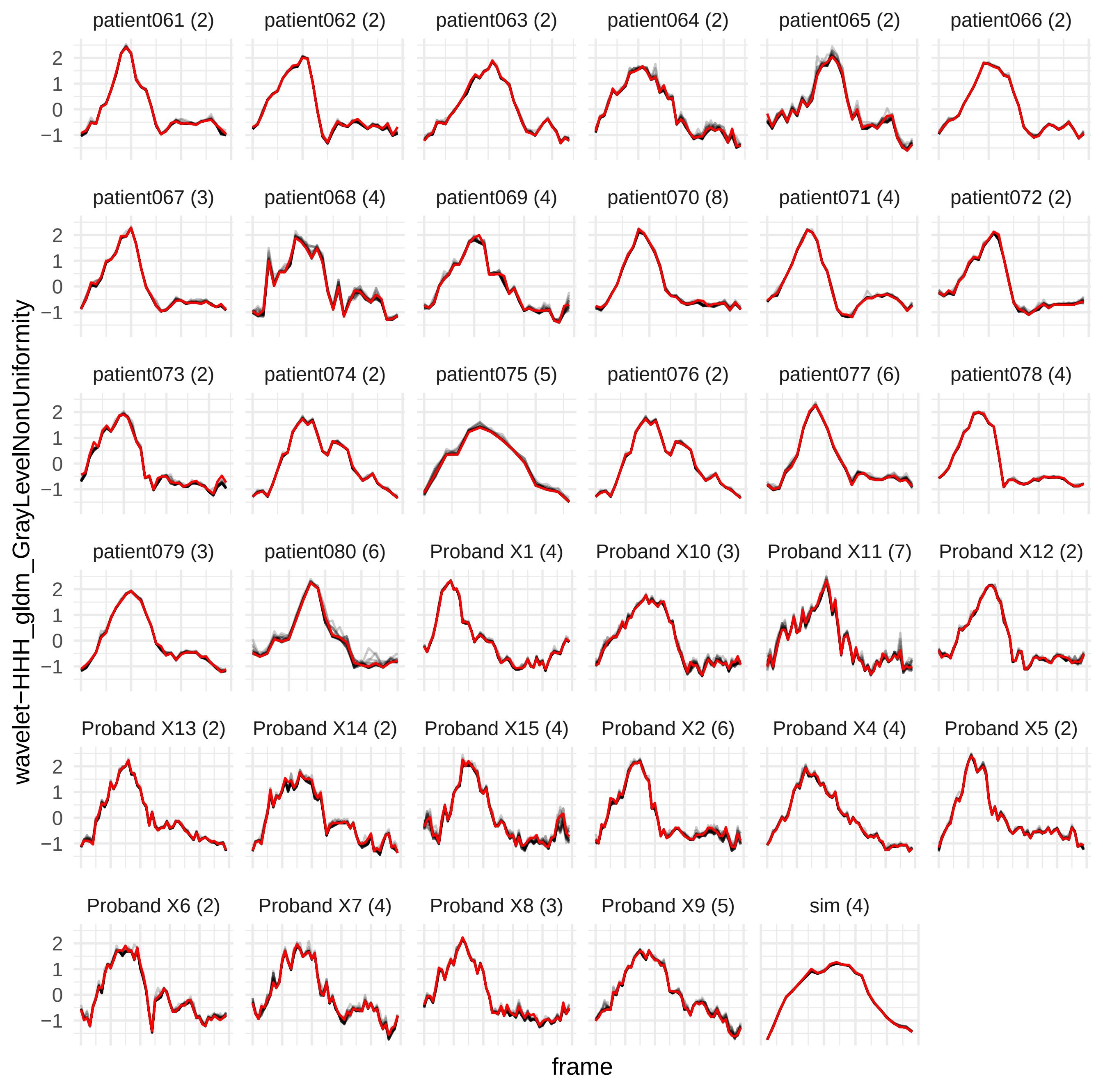
patient061 (2)
patient062 (2)
patient063 (2)
patient064 (2)
patient065 (2)
patient066 (2)
patient067 (3)
patient068 (4)
patient069 (4)
patient070 (8)
patient071 (4)
patient072 (2)
patient073 (2)
patient074 (2)
patient075 (5)
patient076 (2)
patient077 (6)
patient078 (4)
patient079 (3)
patient080 (6)
Proband X1 (4)
Proband X10 (3)
Proband X11 (7)
Proband X12 (2)
Proband X13 (2)
Proband X14 (2)
Proband X15 (4)
Proband X2 (6)
Proband X4 (4)
Proband X5 (2)
Proband X6 (2)
Proband X7 (4)
Proband X8 (3)
Proband X9 (5)
sim (4)
2
1
0
−1
wavelet−HHH_gldm_GrayLevelNonUniformity
frame

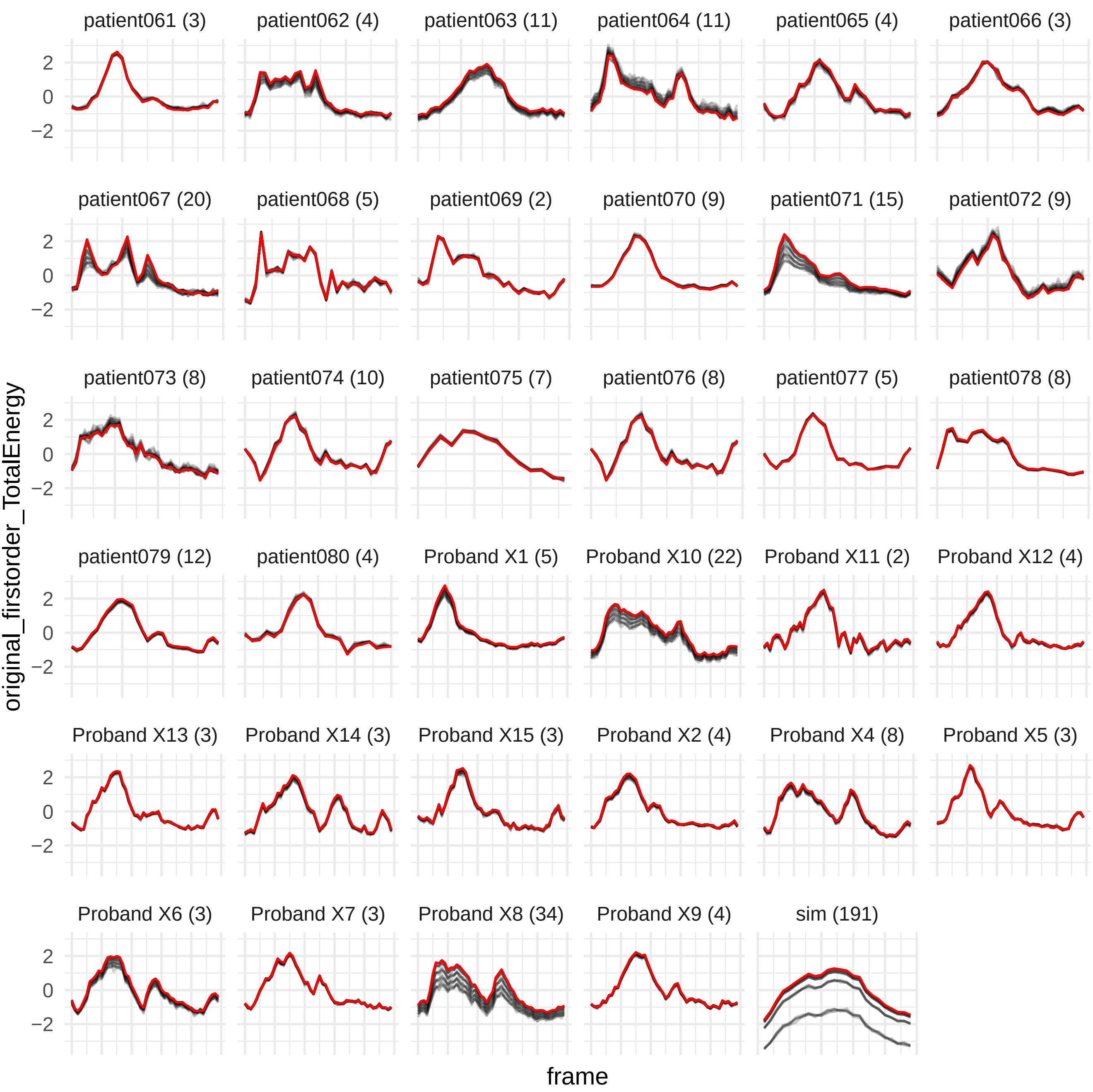

patient061 (3)
patient062 (4)
patient063 (11)
patient064 (11)
patient065 (4)
patient066 (3)
patient067 (20)
patient068 (5)
patient069 (2)
patient070 (9)
patient071 (15)
patient072 (9)
patient073 (8)
patient074 (10)
patient075 (7)
patient076 (8)
patient077 (5)
patient078 (8)
patient079 (12)
patient080 (4)
Proband X1 (5)
Proband X10 (22)
Proband X11 (2)
Proband X12 (4)
Proband X13 (3)
Proband X14 (3)
Proband X15 (3)
Proband X2 (4)
Proband X4 (8)
Proband X5 (3)
Proband X6 (3)
Proband X7 (3)
Proband X8 (34)
Proband X9 (4)
sim (191)
2
0
−2
original_firstorder_TotalEnergy
frame

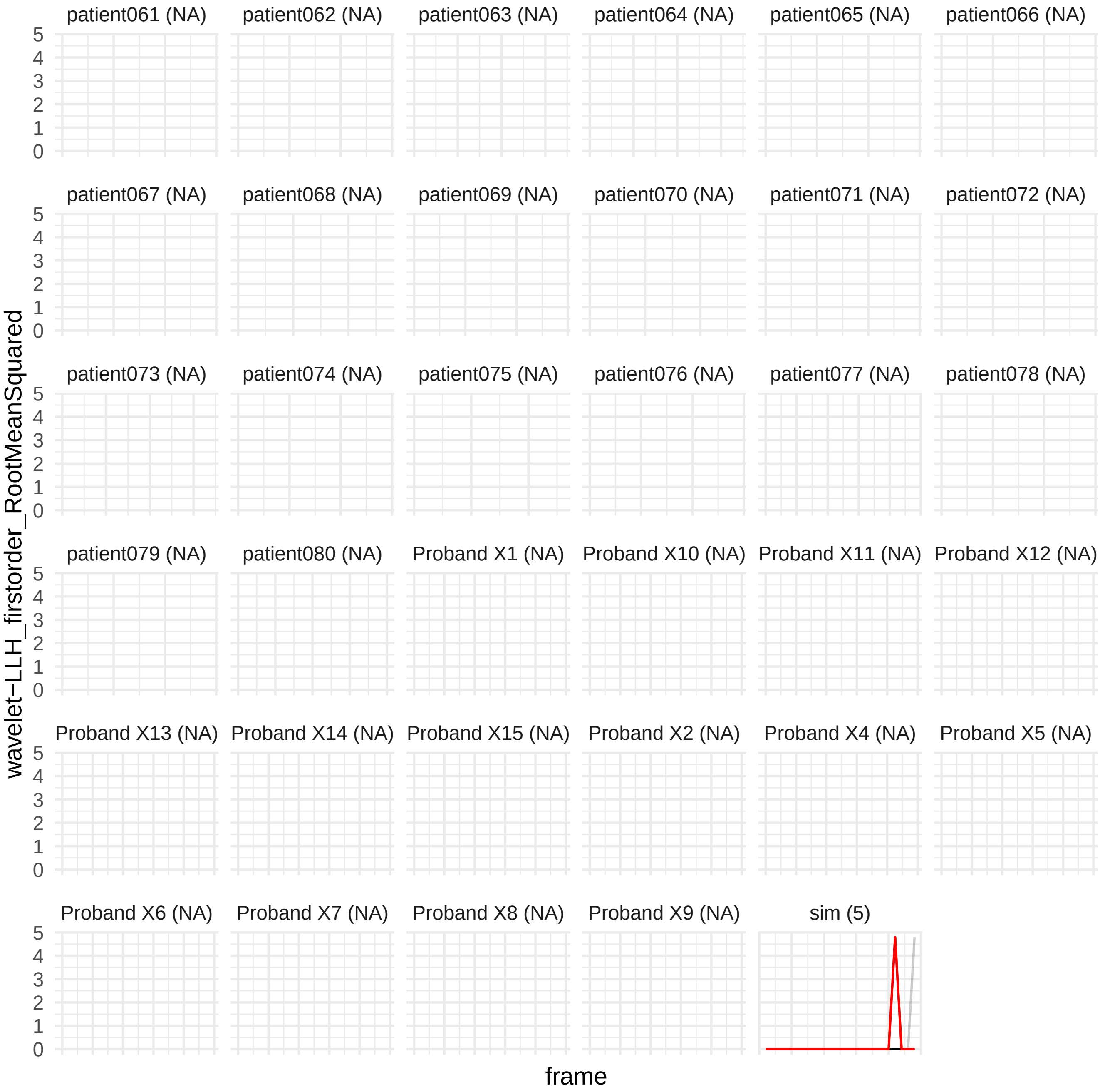

patient061 (NA)
patient062 (NA)
patient063 (NA)
patient064 (NA)
patient065 (NA)
patient066 (NA)
patient067 (NA)
patient068 (NA)
patient069 (NA)
patient070 (NA)
patient071 (NA)
patient072 (NA)
patient073 (NA)
patient074 (NA)
patient075 (NA)
patient076 (NA)
patient077 (NA)
patient078 (NA)
patient079 (NA)
patient080 (NA)
Proband X1 (NA)
Proband X10 (NA)
Proband X11 (NA)
Proband X12 (NA)
Proband X13 (NA)
Proband X14 (NA)
Proband X15 (NA)
Proband X2 (NA)
Proband X4 (NA)
Proband X5 (NA)
Proband X6 (NA)
Proband X7 (NA)
Proband X8 (NA)
Proband X9 (NA)
sim (5)
0
1
2
3
4
5
wavelet−LLH_firstorder_RootMeanSquared
frame

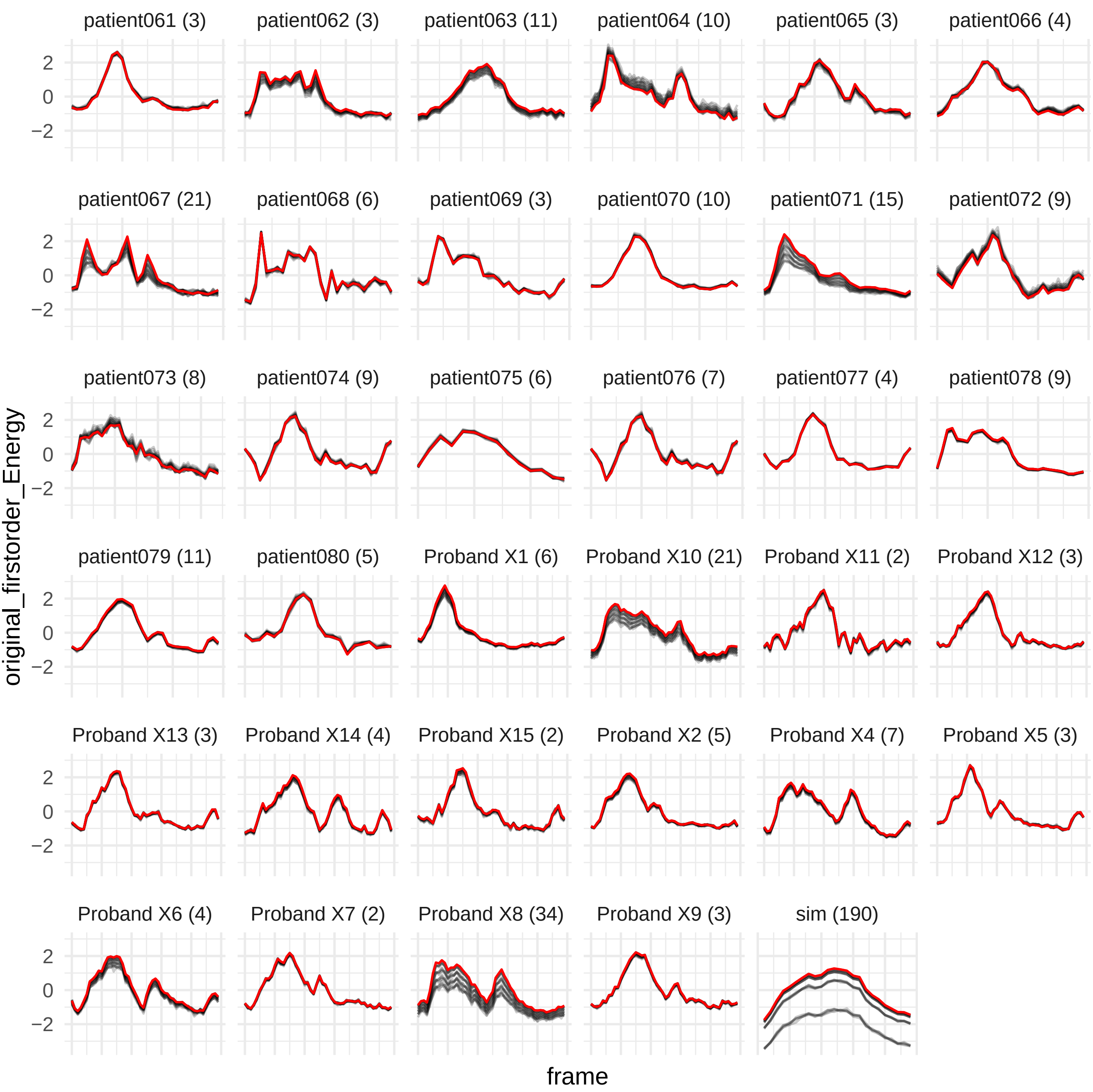

patient061 (3)
patient062 (3)
patient063 (11)
patient064 (10)
patient065 (3)
patient066 (4)
patient067 (21)
patient068 (6)
patient069 (3)
patient070 (10)
patient071 (15)
patient072 (9)
patient073 (8)
patient074 (9)
patient075 (6)
patient076 (7)
patient077 (4)
patient078 (9)
patient079 (11)
patient080 (5)
Proband X1 (6)
Proband X10 (21)
Proband X11 (2)
Proband X12 (3)
Proband X13 (3)
Proband X14 (4)
Proband X15 (2)
Proband X2 (5)
Proband X4 (7)
Proband X5 (3)
Proband X6 (4)
Proband X7 (2)
Proband X8 (34)
Proband X9 (3)
sim (190)
original_firstorder_Energy
frame
2
0
−2

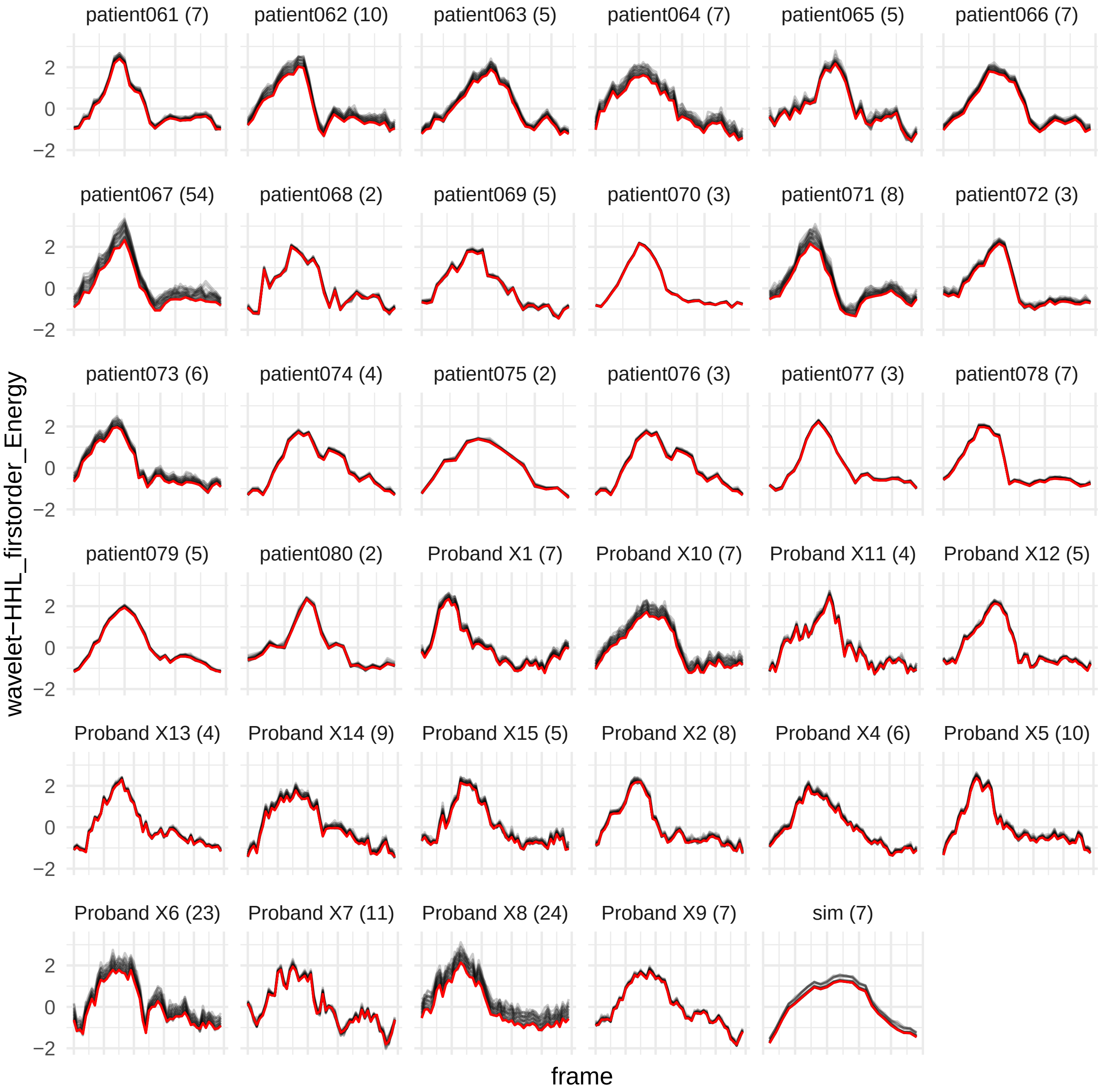
patient061 (7)
patient062 (10)
patient063 (5)
patient064 (7)
patient065 (5)
patient066 (7)
patient067 (54)
patient068 (2)
patient069 (5)
patient070 (3)
patient071 (8)
patient072 (3)
patient073 (6)
patient074 (4)
patient075 (2)
patient076 (3)
patient077 (3)
patient078 (7)
patient079 (5)
patient080 (2)
Proband X1 (7)
Proband X10 (7)
Proband X11 (4)
Proband X12 (5)
Proband X13 (4)
Proband X14 (9)
Proband X15 (5)
Proband X2 (8)
Proband X4 (6)
Proband X5 (10)
Proband X6 (23)
Proband X7 (11)
Proband X8 (24)
Proband X9 (7)
sim (7)
wavelet−HHL_firstorder_Energy
frame
2
0
−2

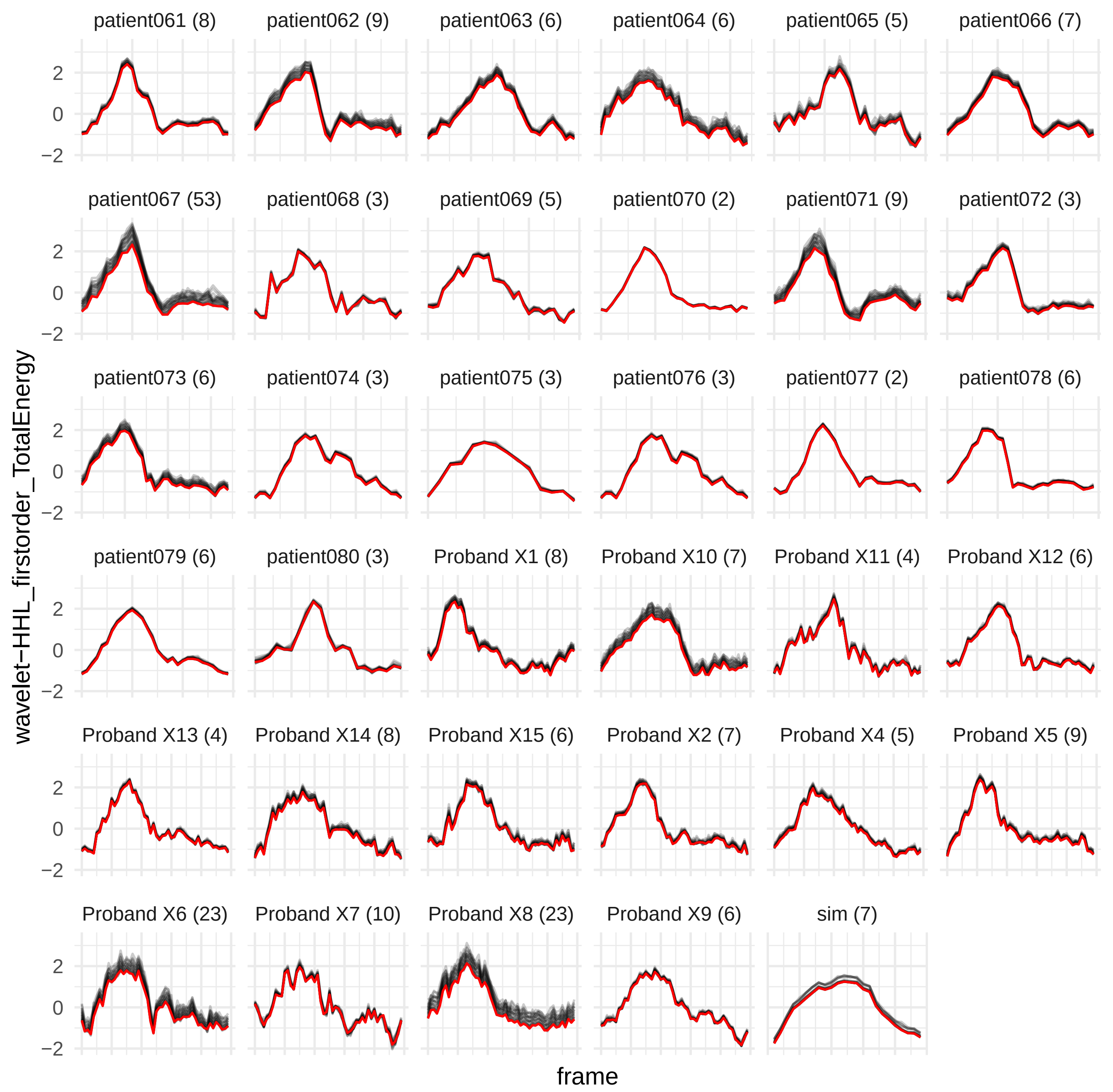

patient061 (8)
patient062 (9)
patient063 (6)
patient064 (6)
patient065 (5)
patient066 (7)
patient067 (53)
patient068 (3)
patient069 (5)
patient070 (2)
patient071 (9)
patient072 (3)
patient073 (6)
patient074 (3)
patient075 (3)
patient076 (3)
patient077 (2)
patient078 (6)
patient079 (6)
patient080 (3)
Proband X1 (8)
Proband X10 (7)
Proband X11 (4)
Proband X12 (6)
Proband X13 (4)
Proband X14 (8)
Proband X15 (6)
Proband X2 (7)
Proband X4 (5)
Proband X5 (9)
Proband X6 (23)
Proband X7 (10)
Proband X8 (23)
Proband X9 (6)
sim (7)
2
0
−2
wavelet−HHL_firstorder_TotalEnergy
frame

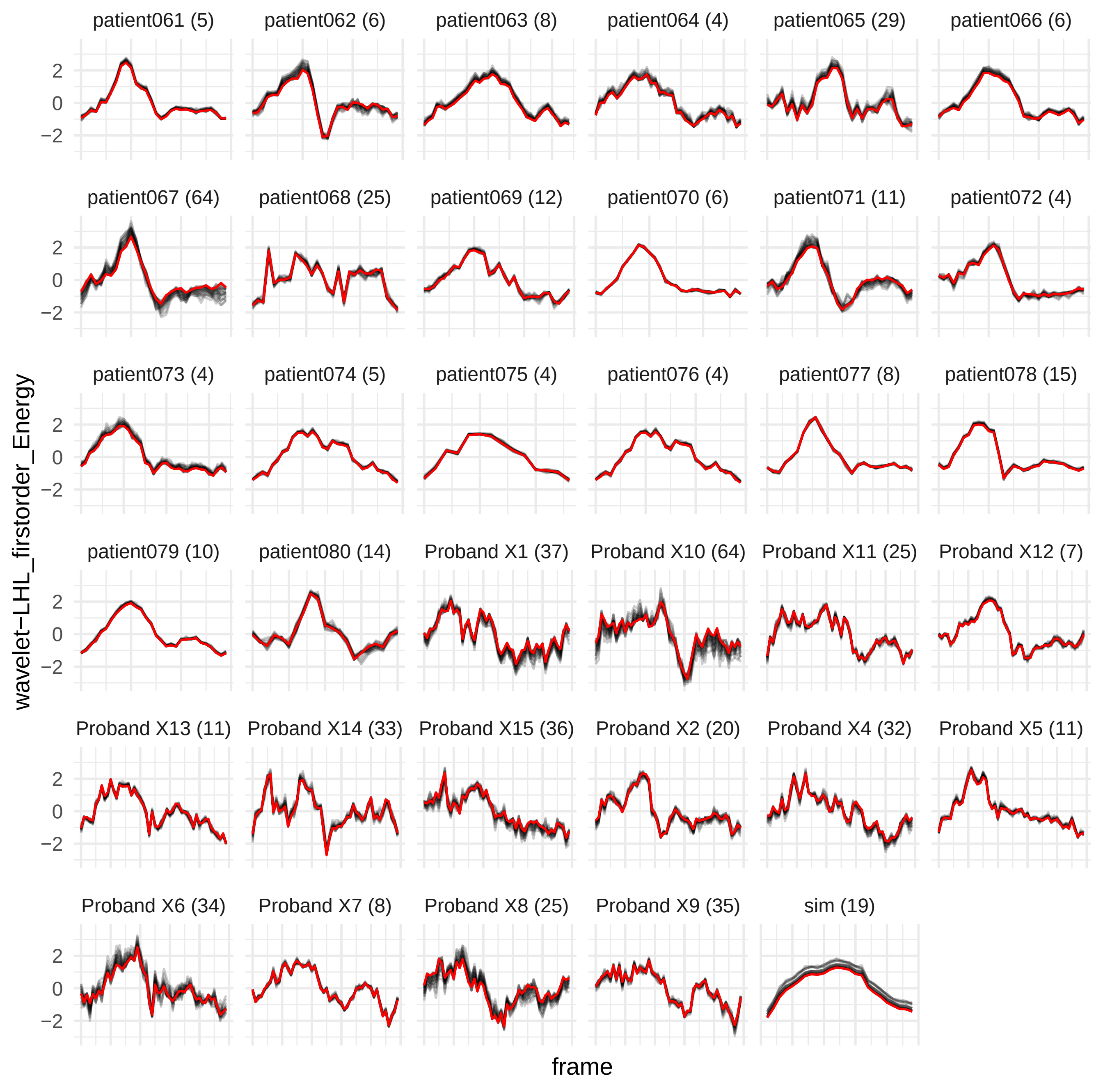
patient061 (5)
patient062 (6)
patient063 (8)
patient064 (4)
patient065 (29)
patient066 (6)
patient067 (64)
patient068 (25)
patient069 (12)
patient070 (6)
patient071 (11)
patient072 (4)
patient073 (4)
patient074 (5)
patient075 (4)
patient076 (4)
patient077 (8)
patient078 (15)
patient079 (10)
patient080 (14)
Proband X1 (37)
Proband X10 (64)
Proband X11 (25)
Proband X12 (7)
Proband X13 (11)
Proband X14 (33)
Proband X15 (36)
Proband X2 (20)
Proband X4 (32)
Proband X5 (11)
Proband X6 (34)
Proband X7 (8)
Proband X8 (25)
Proband X9 (35)
sim (19)
2
0
−2
wavelet−LHL_firstorder_Energy
frame

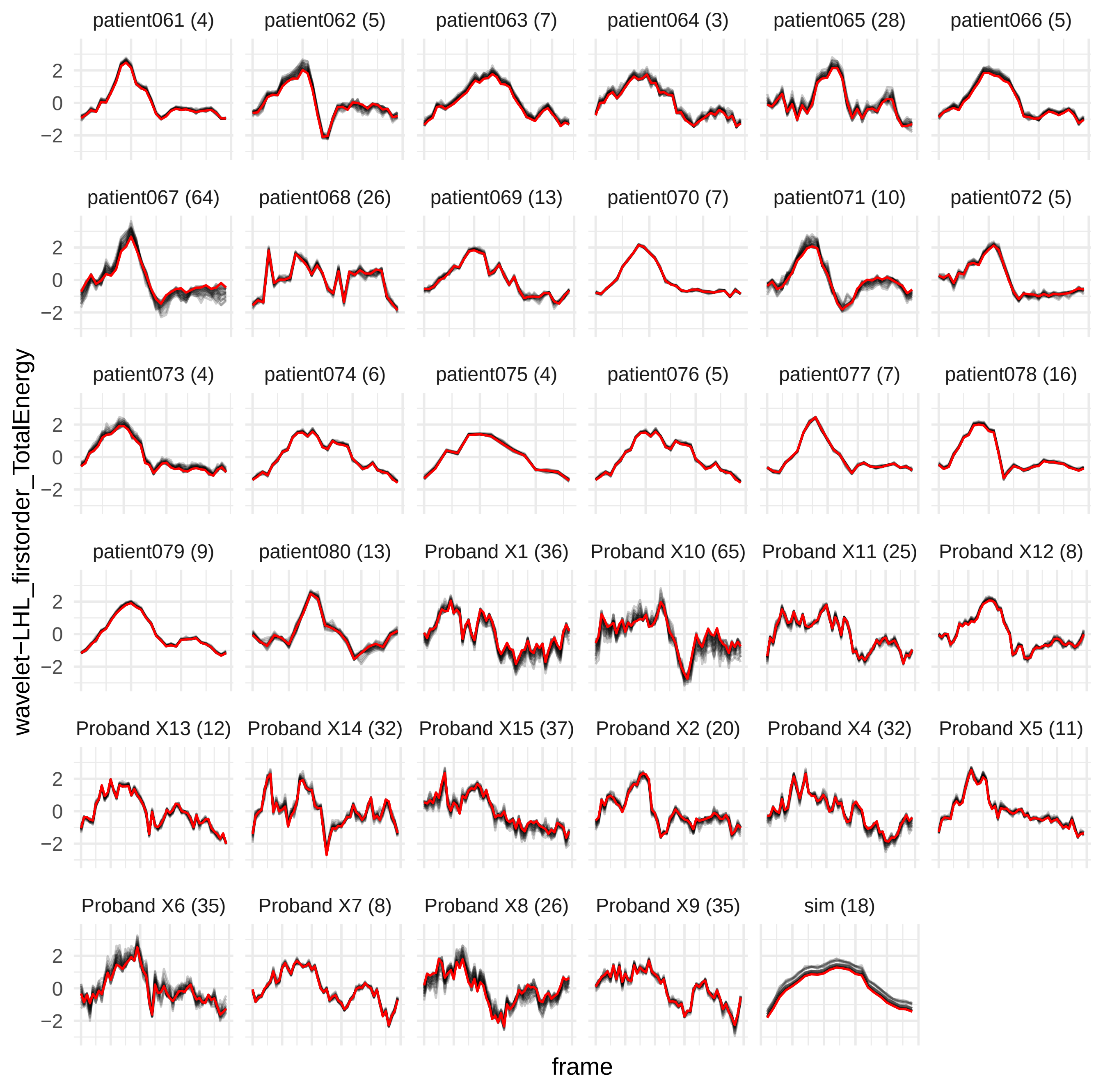

patient061 (4)
patient062 (5)
patient063 (7)
patient064 (3)
patient065 (28)
patient066 (5)
patient067 (64)
patient068 (26)
patient069 (13)
patient070 (7)
patient071 (10)
patient072 (5)
patient073 (4)
patient074 (6)
patient075 (4)
patient076 (5)
patient077 (7)
patient078 (16)
patient079 (9)
patient080 (13)
Proband X1 (36)
Proband X10 (65)
Proband X11 (25)
Proband X12 (8)
Proband X13 (12)
Proband X14 (32)
Proband X15 (37)
Proband X2 (20)
Proband X4 (32)
Proband X5 (11)
Proband X6 (35)
Proband X7 (8)
Proband X8 (26)
Proband X9 (35)
sim (18)
2
0
−2
wavelet−LHL_firstorder_TotalEnergy
frame

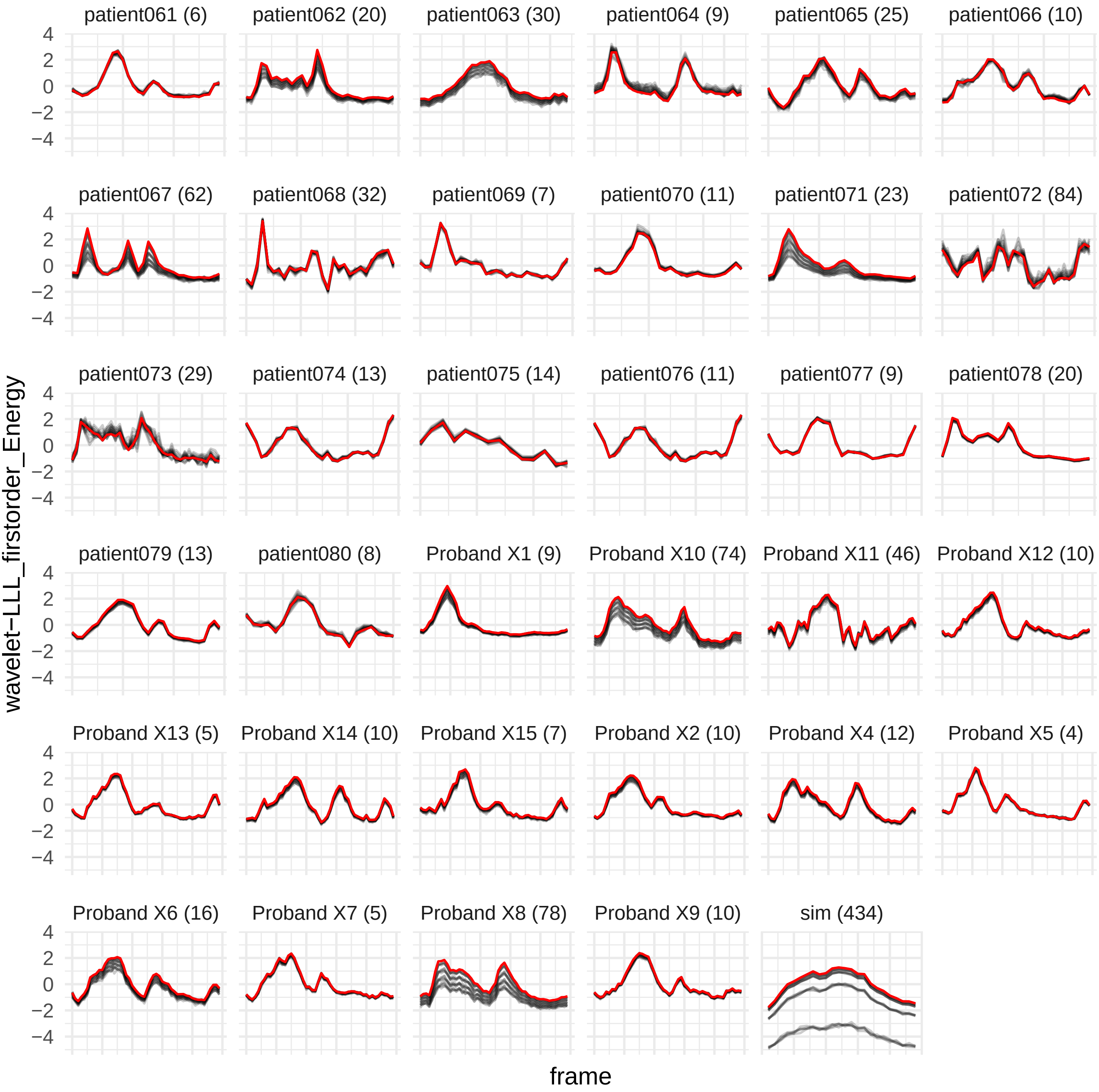
patient061 (6)
patient062 (20)
patient063 (30)
patient064 (9)
patient065 (25)
patient066 (10)
patient067 (62)
patient068 (32)
patient069 (7)
patient070 (11)
patient071 (23)
patient072 (84)
patient073 (29)
patient074 (13)
patient075 (14)
patient076 (11)
patient077 (9)
patient078 (20)
patient079 (13)
patient080 (8)
Proband X1 (9)
Proband X10 (74)
Proband X11 (46)
Proband X12 (10)
Proband X13 (5)
Proband X14 (10)
Proband X15 (7)
Proband X2 (10)
Proband X4 (12)
Proband X5 (4)
Proband X6 (16)
Proband X7 (5)
Proband X8 (78)
Proband X9 (10)
sim (434)
4
2
0
−2
−4
wavelet−LLL_firstorder_Energy
frame

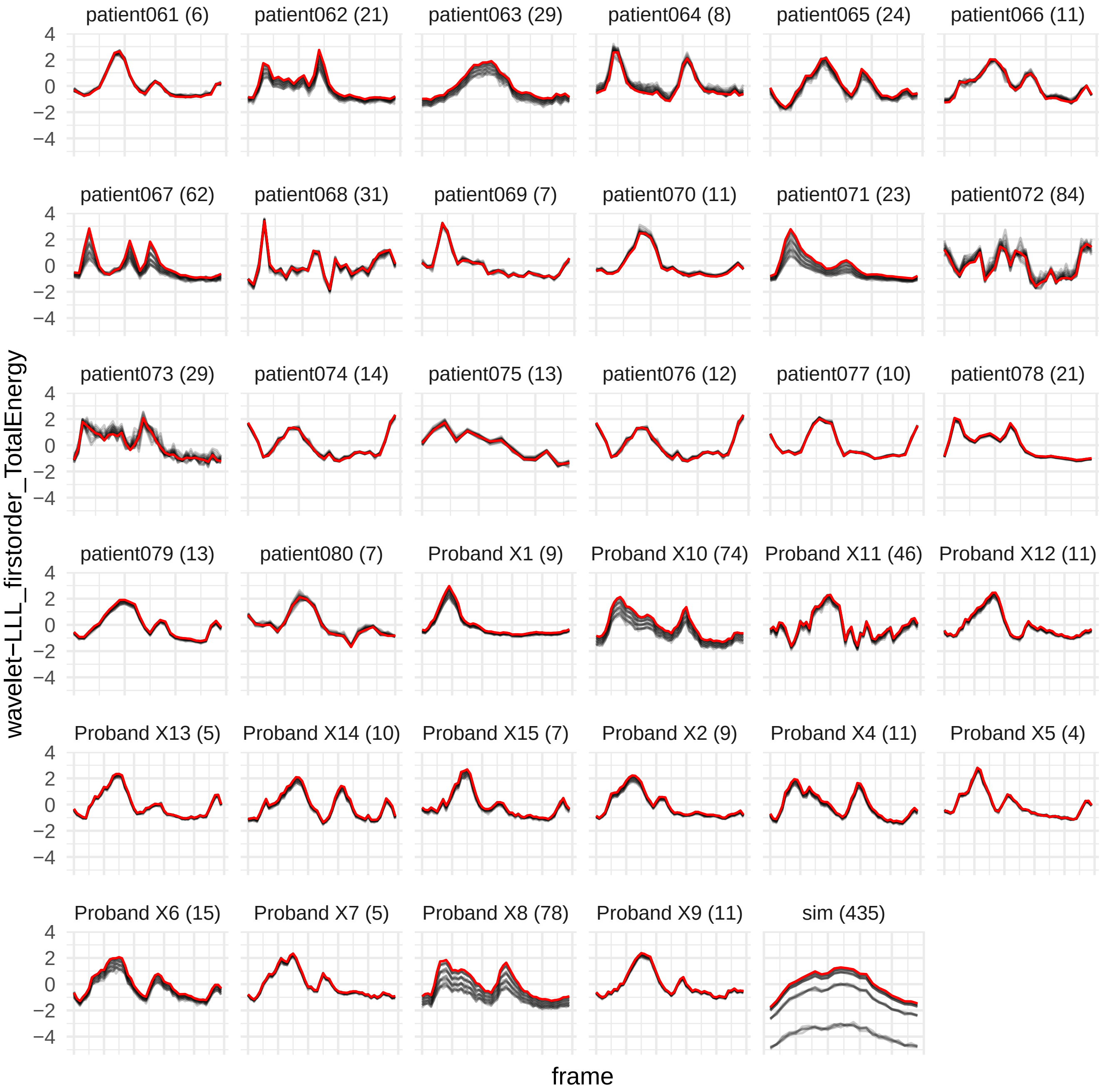

patient061 (6)
patient062 (21)
patient063 (29)
patient064 (8)
patient065 (24)
patient066 (11)
patient067 (62)
patient068 (31)
patient069 (7)
patient070 (11)
patient071 (23)
patient072 (84)
patient073 (29)
patient074 (14)
patient075 (13)
patient076 (12)
patient077 (10)
patient078 (21)
patient079 (13)
patient080 (7)
Proband X1 (9)
Proband X10 (74)
Proband X11 (46)
Proband X12 (11)
Proband X13 (5)
Proband X14 (10)
Proband X15 (7)
Proband X2 (9)
Proband X4 (11)
Proband X5 (4)
Proband X6 (15)
Proband X7 (5)
Proband X8 (78)
Proband X9 (11)
sim (435)
4
2
0
−2
−4
wavelet−LLL_firstorder_TotalEnergy
frame

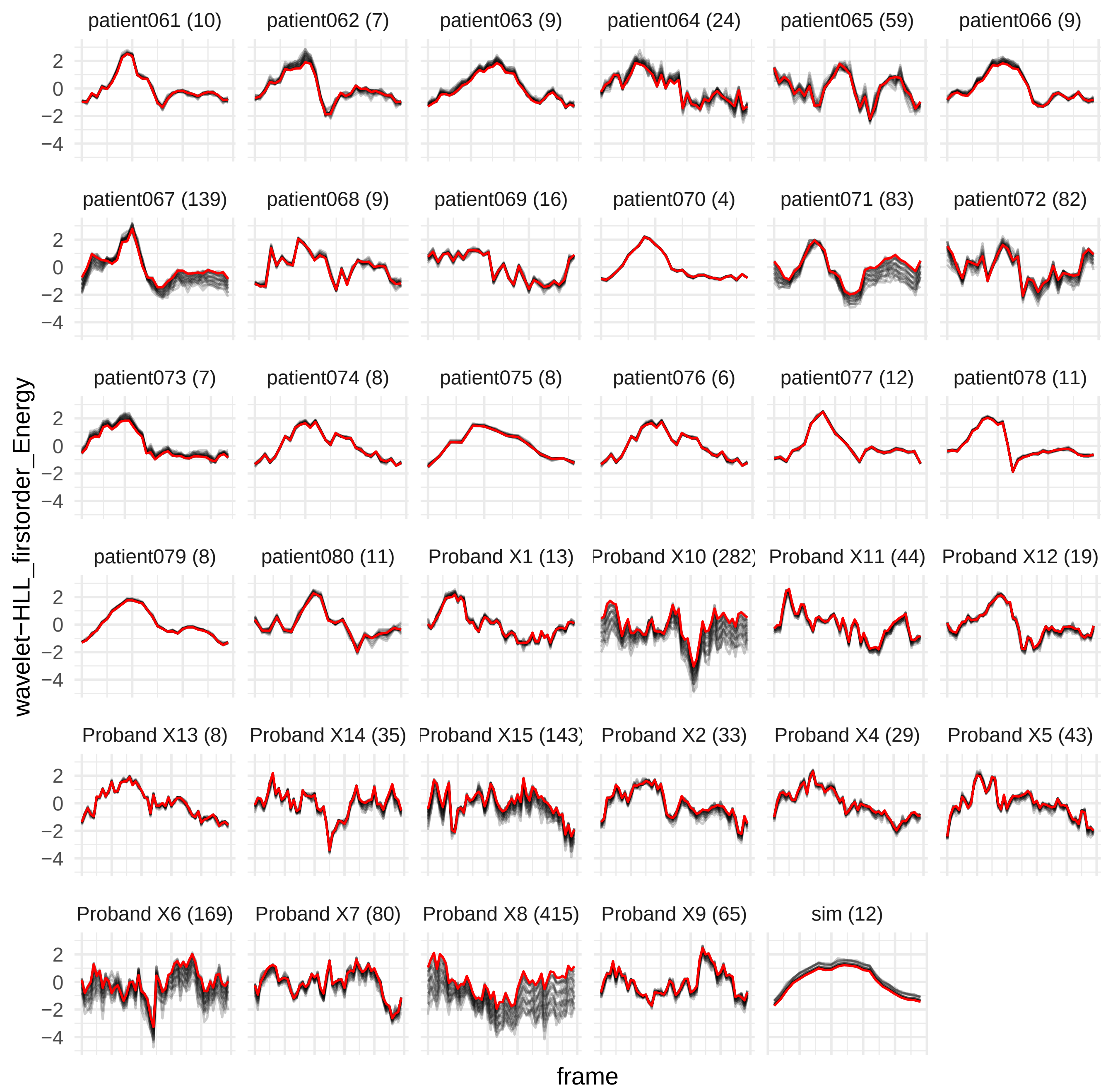
patient061 (10)
patient062 (7)
patient063 (9)
patient064 (24)
patient065 (59)
patient066 (9)
patient067 (139)
patient068 (9)
patient069 (16)
patient070 (4)
patient071 (83)
patient072 (82)
patient073 (7)
patient074 (8)
patient075 (8)
patient076 (6)
patient077 (12)
patient078 (11)
patient079 (8)
patient080 (11)
Proband X1 (13)
Proband X10 (282)
Proband X11 (44)
Proband X12 (19)
Proband X13 (8)
Proband X14 (35)
Proband X15 (143)
Proband X2 (33)
Proband X4 (29)
Proband X5 (43)
Proband X6 (169)
Proband X7 (80)
Proband X8 (415)
Proband X9 (65)
sim (12)
2
0
−2
−4
wavelet−HLL_firstorder_Energy
frame

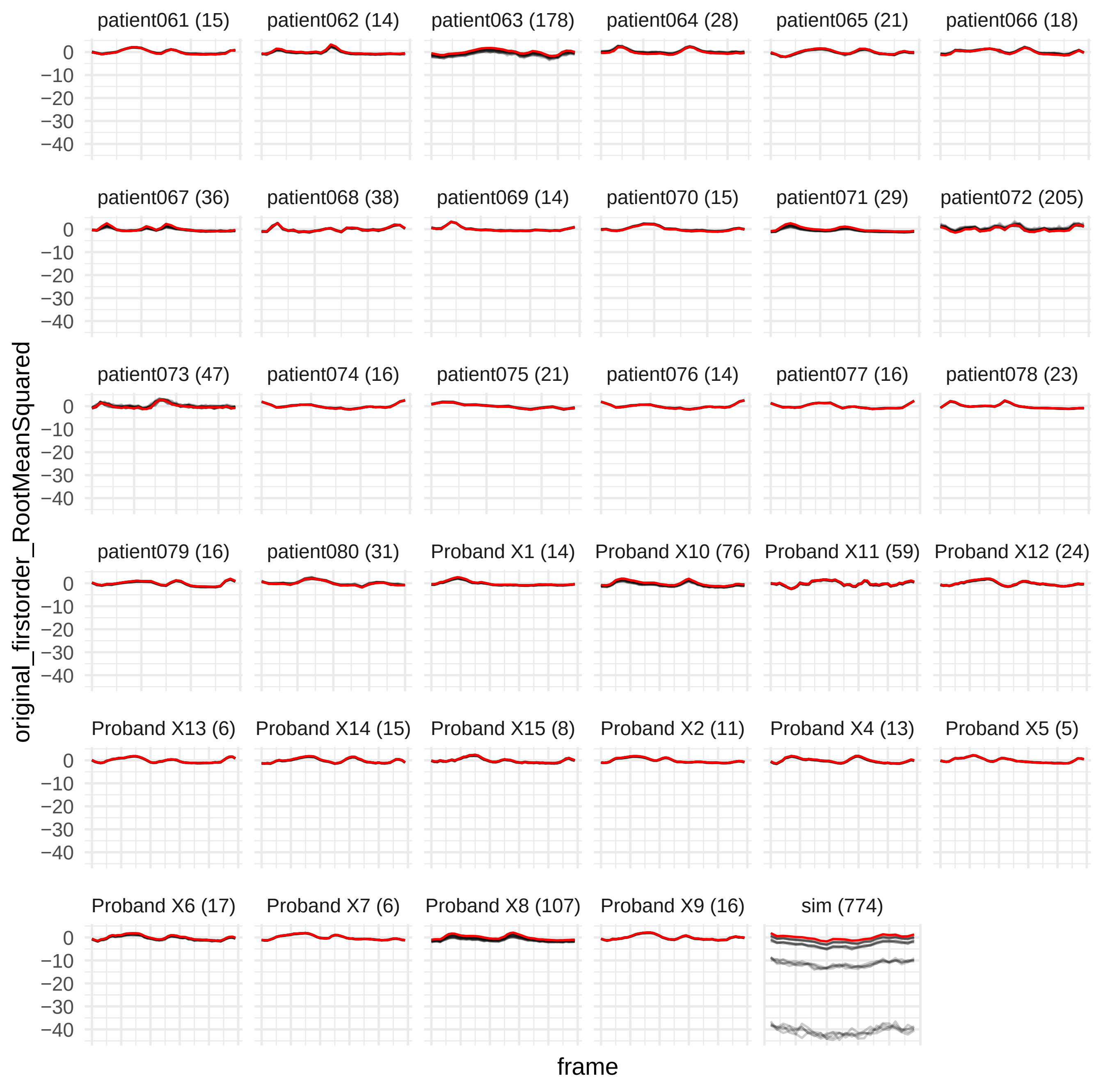
patient061 (15)
patient062 (14)
patient063 (178)
patient064 (28)
patient065 (21)
patient066 (18)
patient067 (36)
patient068 (38)
patient069 (14)
patient070 (15)
patient071 (29)
patient072 (205)
patient073 (47)
patient074 (16)
patient075 (21)
patient076 (14)
patient077 (16)
patient078 (23)
patient079 (16)
patient080 (31)
Proband X1 (14)
Proband X10 (76)
Proband X11 (59)
Proband X12 (24)
Proband X13 (6)
Proband X14 (15)
Proband X15 (8)
Proband X2 (11)
Proband X4 (13)
Proband X5 (5)
Proband X6 (17)
Proband X7 (6)
Proband X8 (107)
Proband X9 (16)
sim (774)
original_firstorder_RootMeanSquared
0
−10
−20
−30
−40
frame

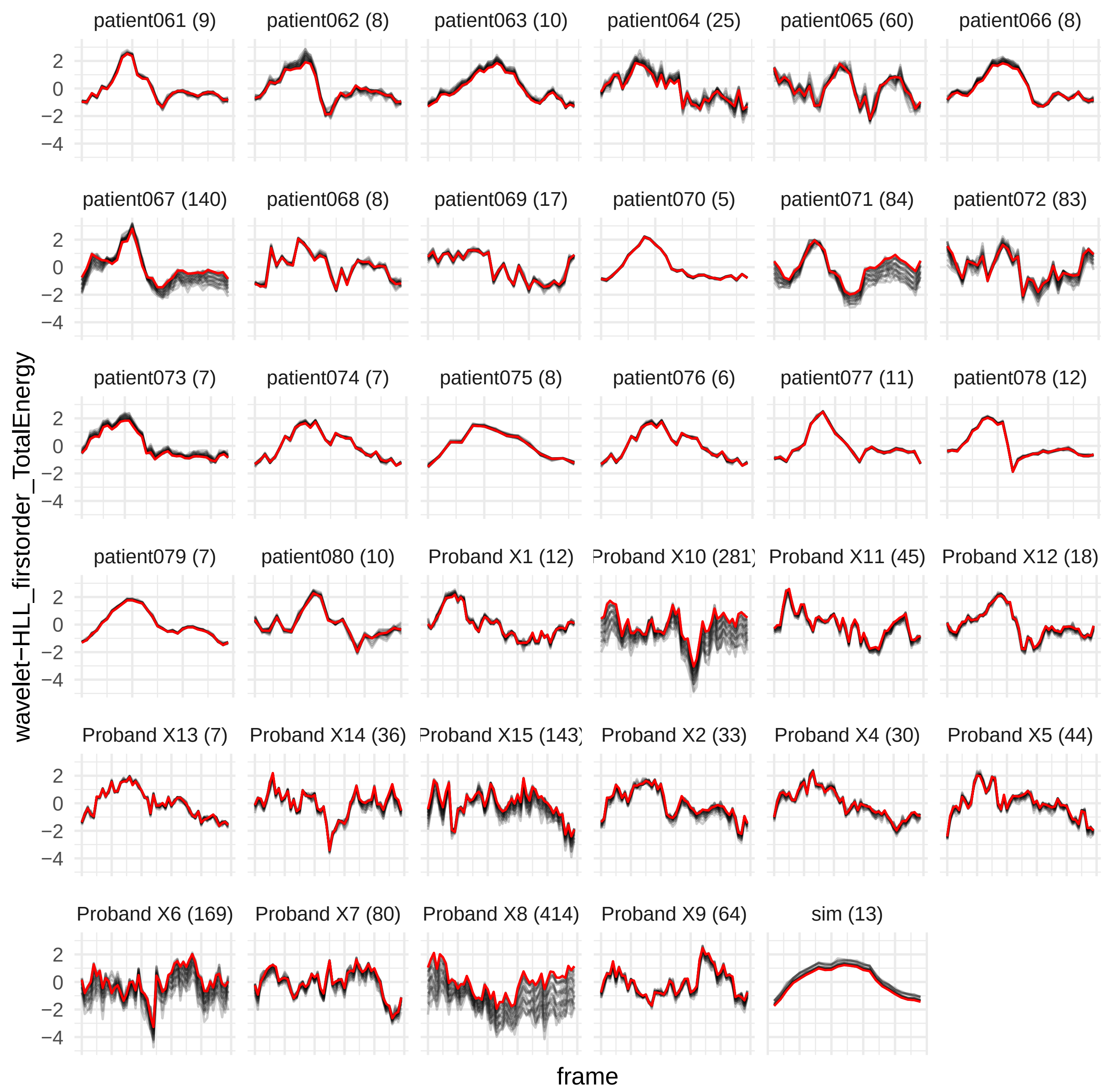

patient061 (9)
patient062 (8)
patient063 (10)
patient064 (25)
patient065 (60)
patient066 (8)
patient067 (140)
patient068 (8)
patient069 (17)
patient070 (5)
patient071 (84)
patient072 (83)
patient073 (7)
patient074 (7)
patient075 (8)
patient076 (6)
patient077 (11)
patient078 (12)
patient079 (7)
patient080 (10)
Proband X1 (12)
Proband X10 (281)
Proband X11 (45)
Proband X12 (18)
Proband X13 (7)
Proband X14 (36)
Proband X15 (143)
Proband X2 (33)
Proband X4 (30)
Proband X5 (44)
Proband X6 (169)
Proband X7 (80)
Proband X8 (414)
Proband X9 (64)
sim (13)
2
0
−2
−4
wavelet−HLL_firstorder_TotalEnergy
frame

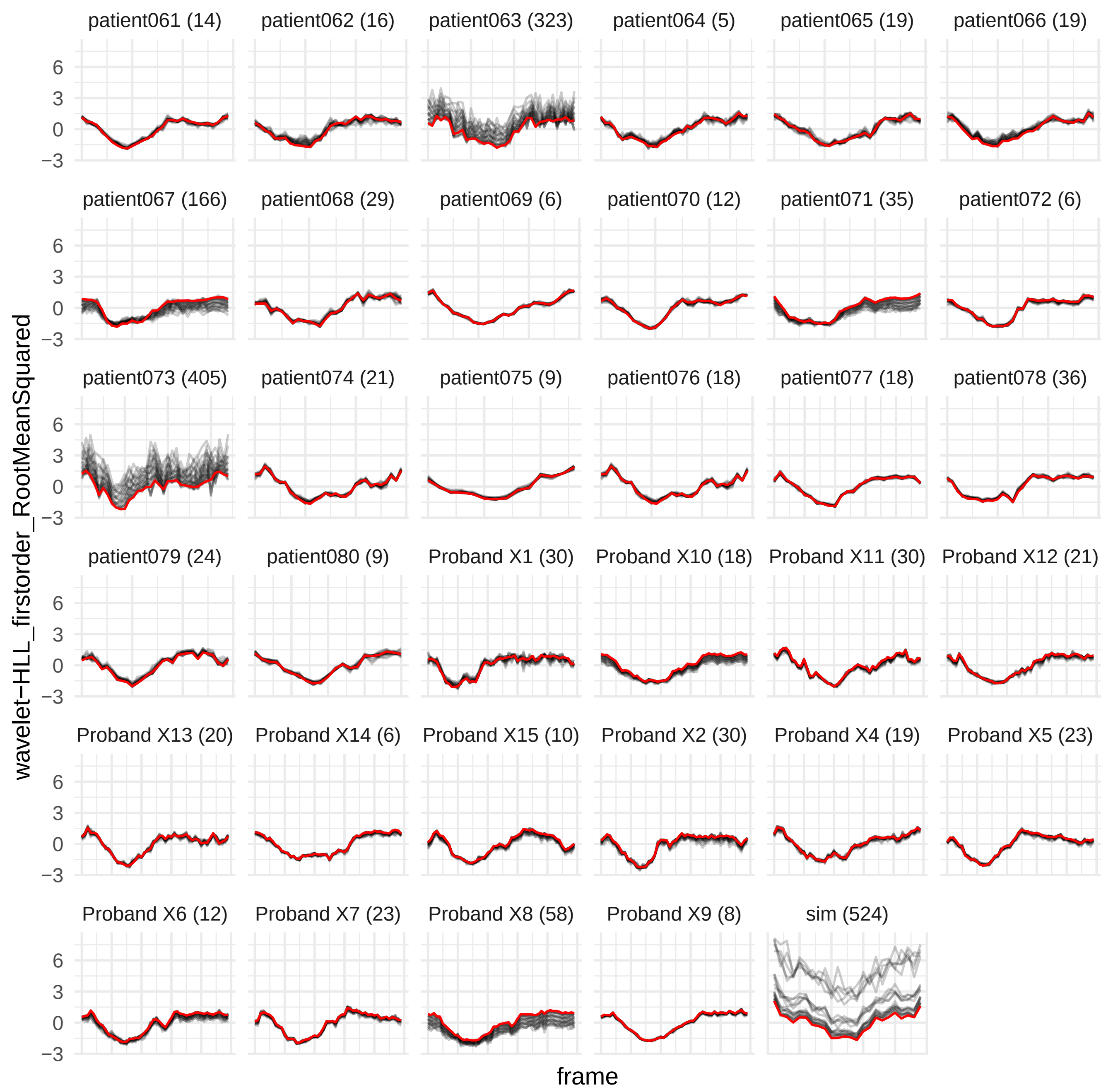
patient061 (14)
patient062 (16)
patient063 (323)
patient064 (5)
patient065 (19)
patient066 (19)
patient067 (166)
patient068 (29)
patient069 (6)
patient070 (12)
patient071 (35)
patient072 (6)
patient073 (405)
patient074 (21)
patient075 (9)
patient076 (18)
patient077 (18)
patient078 (36)
patient079 (24)
patient080 (9)
Proband X1 (30)
Proband X10 (18)
Proband X11 (30)
Proband X12 (21)
Proband X13 (20)
Proband X14 (6)
Proband X15 (10)
Proband X2 (30)
Proband X4 (19)
Proband X5 (23)
Proband X6 (12)
Proband X7 (23)
Proband X8 (58)
Proband X9 (8)
sim (524)
6
3
0
−3
wavelet−HLL_firstorder_RootMeanSquared
frame

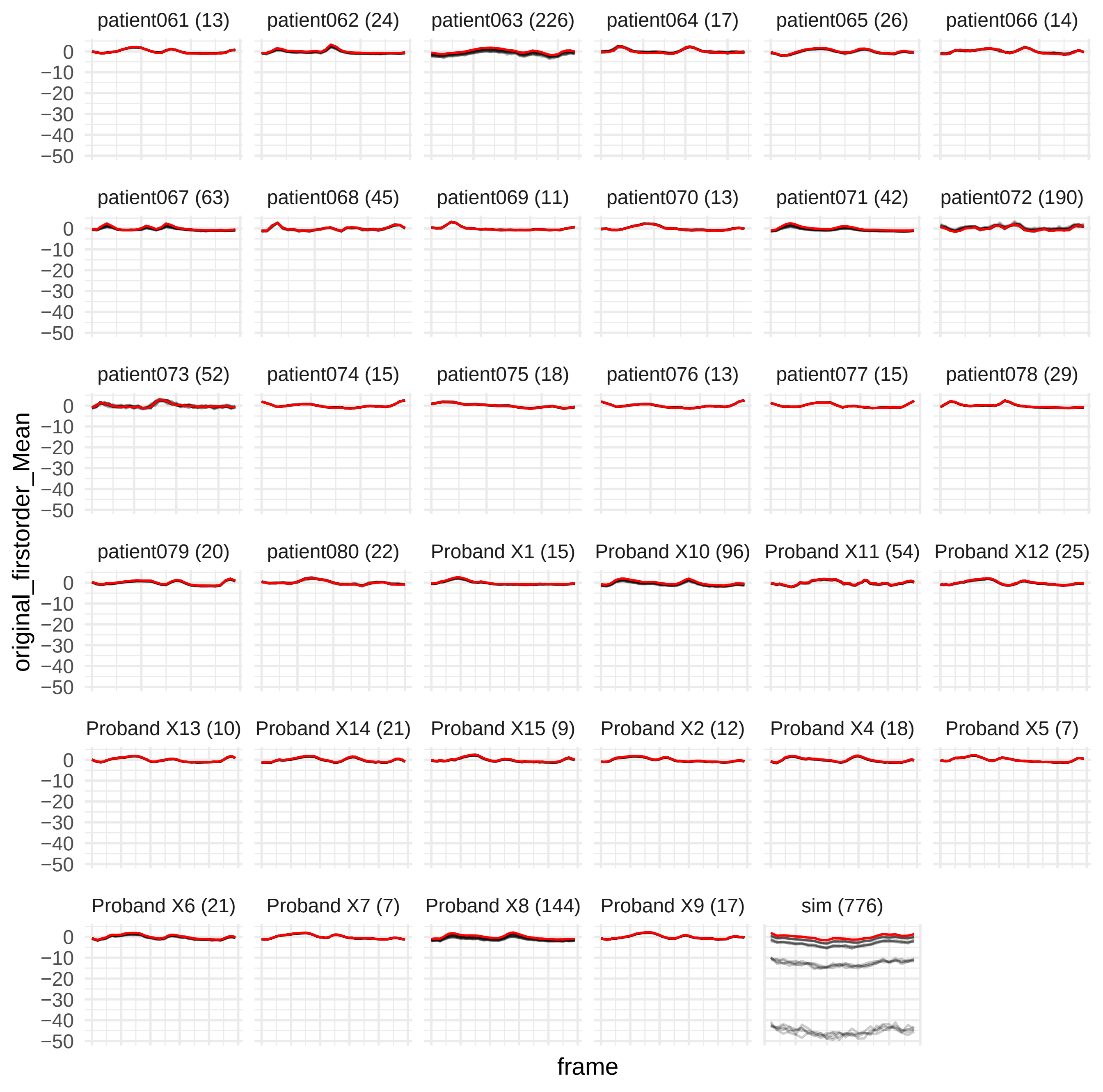
patient061 (13)
patient062 (24)
patient063 (226)
patient064 (17)
patient065 (26)
patient066 (14)
patient067 (63)
patient068 (45)
patient069 (11)
patient070 (13)
patient071 (42)
patient072 (190)
patient073 (52)
patient074 (15)
patient075 (18)
patient076 (13)
patient077 (15)
patient078 (29)
patient079 (20)
patient080 (22)
Proband X1 (15)
Proband X10 (96)
Proband X11 (54)
Proband X12 (25)
Proband X13 (10)
Proband X14 (21)
Proband X15 (9)
Proband X2 (12)
Proband X4 (18)
Proband X5 (7)
Proband X6 (21)
Proband X7 (7)
Proband X8 (144)
Proband X9 (17)
sim (776)
0
−10
−20
−30
−40
−50
original_firstorder_Mean
frame

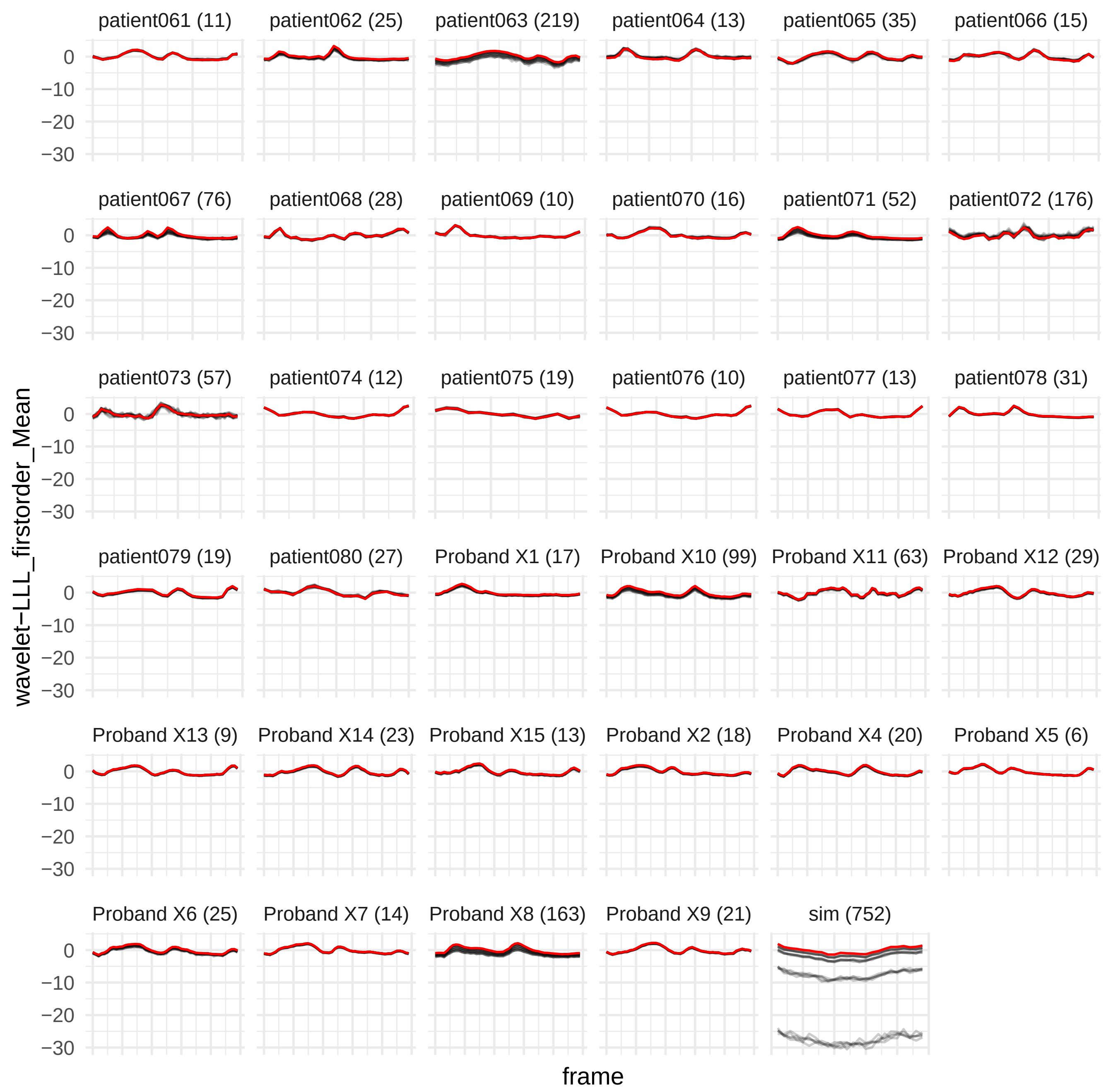

patient061 (11)
patient062 (25)
patient063 (219)
patient064 (13)
patient065 (35)
patient066 (15)
patient067 (76)
patient068 (28)
patient069 (10)
patient070 (16)
patient071 (52)
patient072 (176)
patient073 (57)
patient074 (12)
patient075 (19)
patient076 (10)
patient077 (13)
patient078 (31)
patient079 (19)
patient080 (27)
Proband X1 (17)
Proband X10 (99)
Proband X11 (63)
Proband X12 (29)
Proband X13 (9)
Proband X14 (23)
Proband X15 (13)
Proband X2 (18)
Proband X4 (20)
Proband X5 (6)
Proband X6 (25)
Proband X7 (14)
Proband X8 (163)
Proband X9 (21)
sim (752)
0
−10
−20
−30
wavelet−LLL_firstorder_Mean
frame

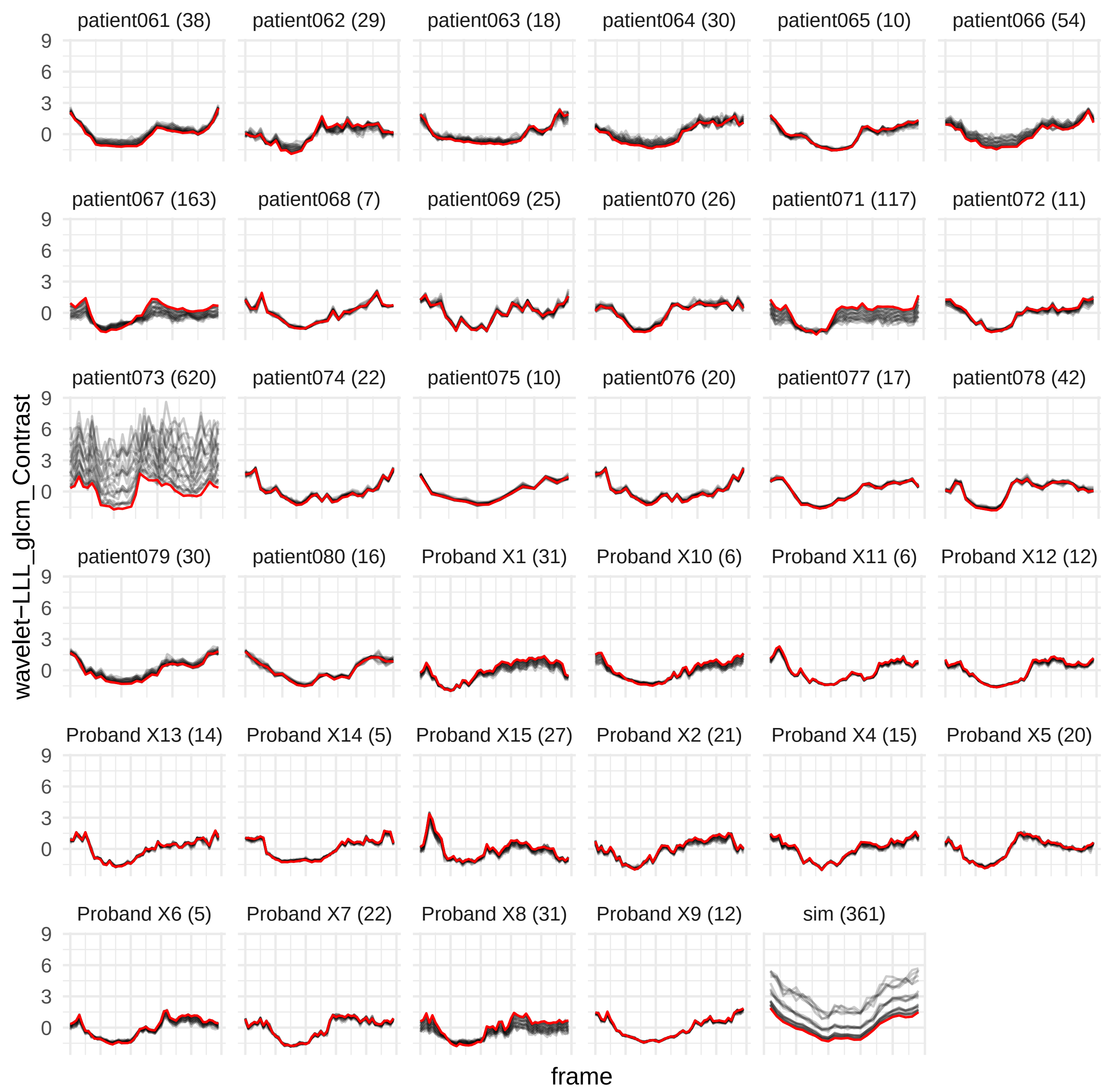

patient061 (38)
patient062 (29)
patient063 (18)
patient064 (30)
patient065 (10)
patient066 (54)
patient067 (163)
patient068 (7)
patient069 (25)
patient070 (26)
patient071 (117)
patient072 (11)
patient073 (620)
patient074 (22)
patient075 (10)
patient076 (20)
patient077 (17)
patient078 (42)
patient079 (30)
patient080 (16)
Proband X1 (31)
Proband X10 (6)
Proband X11 (6)
Proband X12 (12)
Proband X13 (14)
Proband X14 (5)
Proband X15 (27)
Proband X2 (21)
Proband X4 (15)
Proband X5 (20)
Proband X6 (5)
Proband X7 (22)
Proband X8 (31)
Proband X9 (12)
sim (361)
9
6
3
0
wavelet−LLL_glcm_Contrast
frame

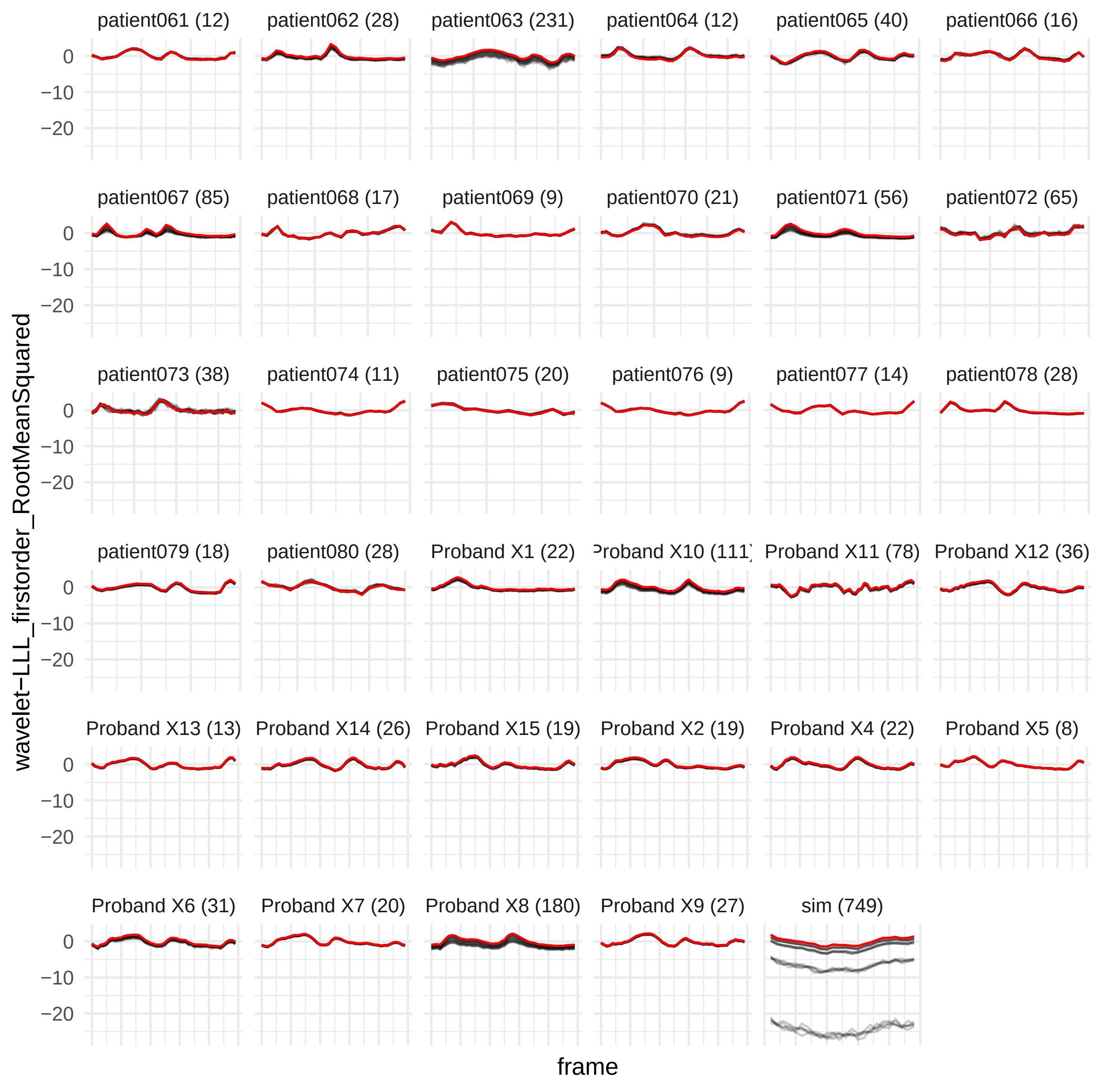

patient061 (12)
patient062 (28)
patient063 (231)
patient064 (12)
patient065 (40)
patient066 (16)
patient067 (85)
patient068 (17)
patient069 (9)
patient070 (21)
patient071 (56)
patient072 (65)
patient073 (38)
patient074 (11)
patient075 (20)
patient076 (9)
patient077 (14)
patient078 (28)
patient079 (18)
patient080 (28)
Proband X1 (22)
Proband X10 (111)
Proband X11 (78)
Proband X12 (36)
Proband X13 (13)
Proband X14 (26)
Proband X15 (19)
Proband X2 (19)
Proband X4 (22)
Proband X5 (8)
Proband X6 (31)
Proband X7 (20)
Proband X8 (180)
Proband X9 (27)
sim (749)
0
−10
−20
wavelet−LLL_firstorder_RootMeanSquared
frame

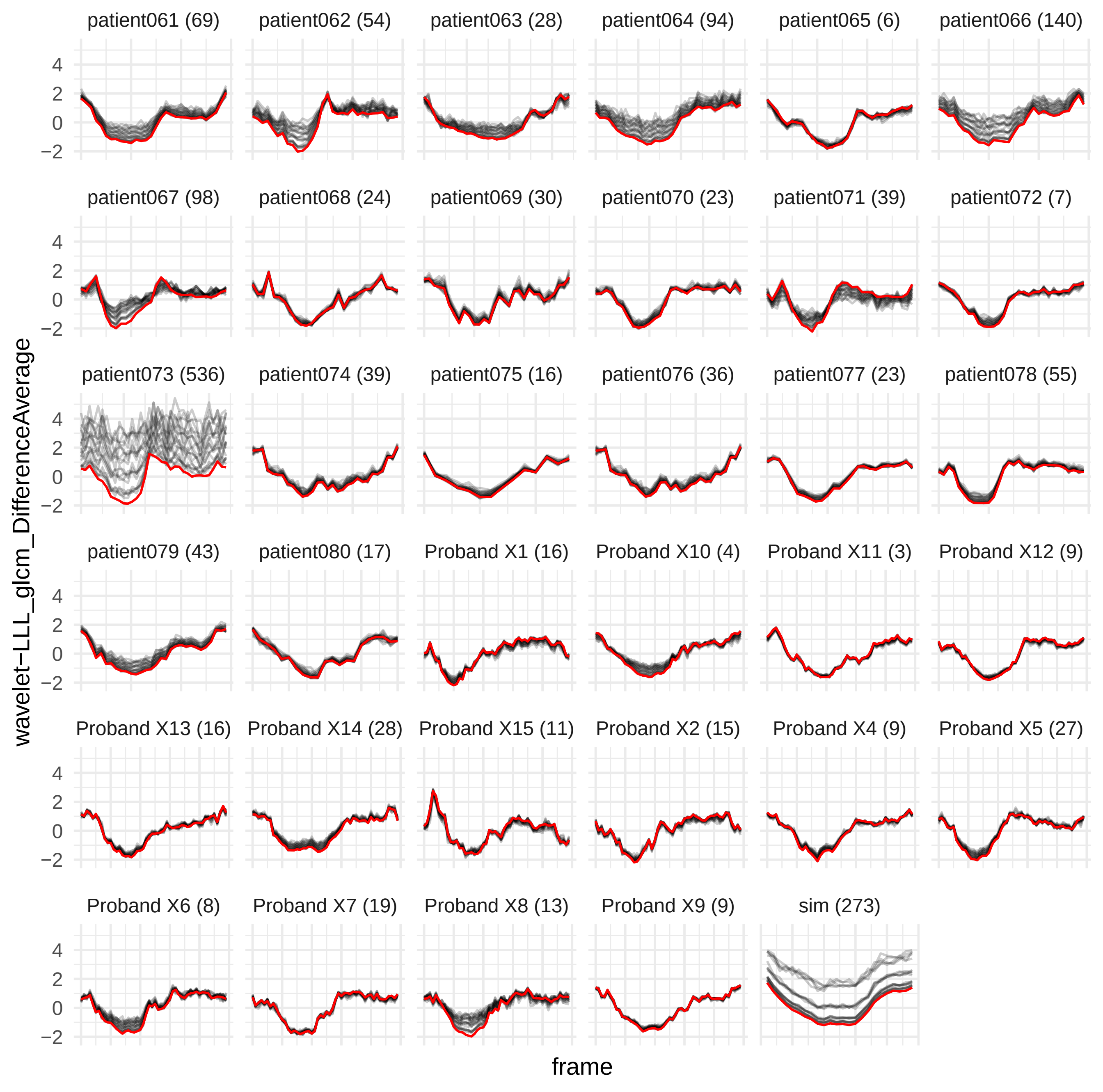
patient061 (69)
patient062 (54)
patient063 (28)
patient064 (94)
patient065 (6)
patient066 (140)
patient067 (98)
patient068 (24)
patient069 (30)
patient070 (23)
patient071 (39)
patient072 (7)
patient073 (536)
patient074 (39)
patient075 (16)
patient076 (36)
patient077 (23)
patient078 (55)
patient079 (43)
patient080 (17)
Proband X1 (16)
Proband X10 (4)
Proband X11 (3)
Proband X12 (9)
Proband X13 (16)
Proband X14 (28)
Proband X15 (11)
Proband X2 (15)
Proband X4 (9)
Proband X5 (27)
Proband X6 (8)
Proband X7 (19)
Proband X8 (13)
Proband X9 (9)
sim (273)
wavelet−LLL_glcm_DifferenceAverage
4
2
0
−2
frame

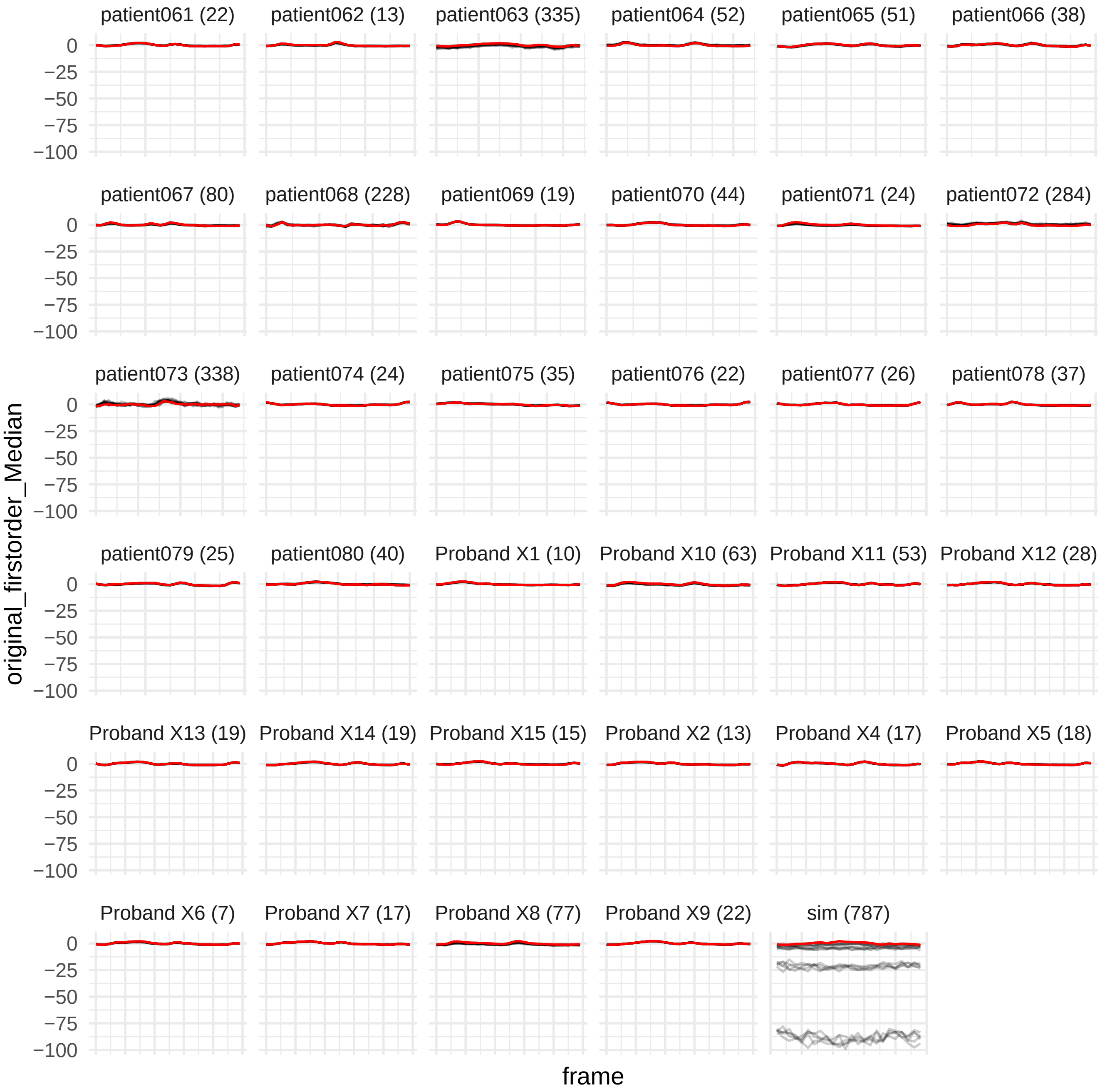

patient061 (22)
patient062 (13)
patient063 (335)
patient064 (52)
patient065 (51)
patient066 (38)
patient067 (80)
patient068 (228)
patient069 (19)
patient070 (44)
patient071 (24)
patient072 (284)
patient073 (338)
patient074 (24)
patient075 (35)
patient076 (22)
patient077 (26)
patient078 (37)
patient079 (25)
patient080 (40)
Proband X1 (10)
Proband X10 (63)
Proband X11 (53)
Proband X12 (28)
Proband X13 (19)
Proband X14 (19)
Proband X15 (15)
Proband X2 (13)
Proband X4 (17)
Proband X5 (18)
Proband X6 (7)
Proband X7 (17)
Proband X8 (77)
Proband X9 (22)
sim (787)
0
−25
−50
−75
−100
original_firstorder_Median
frame

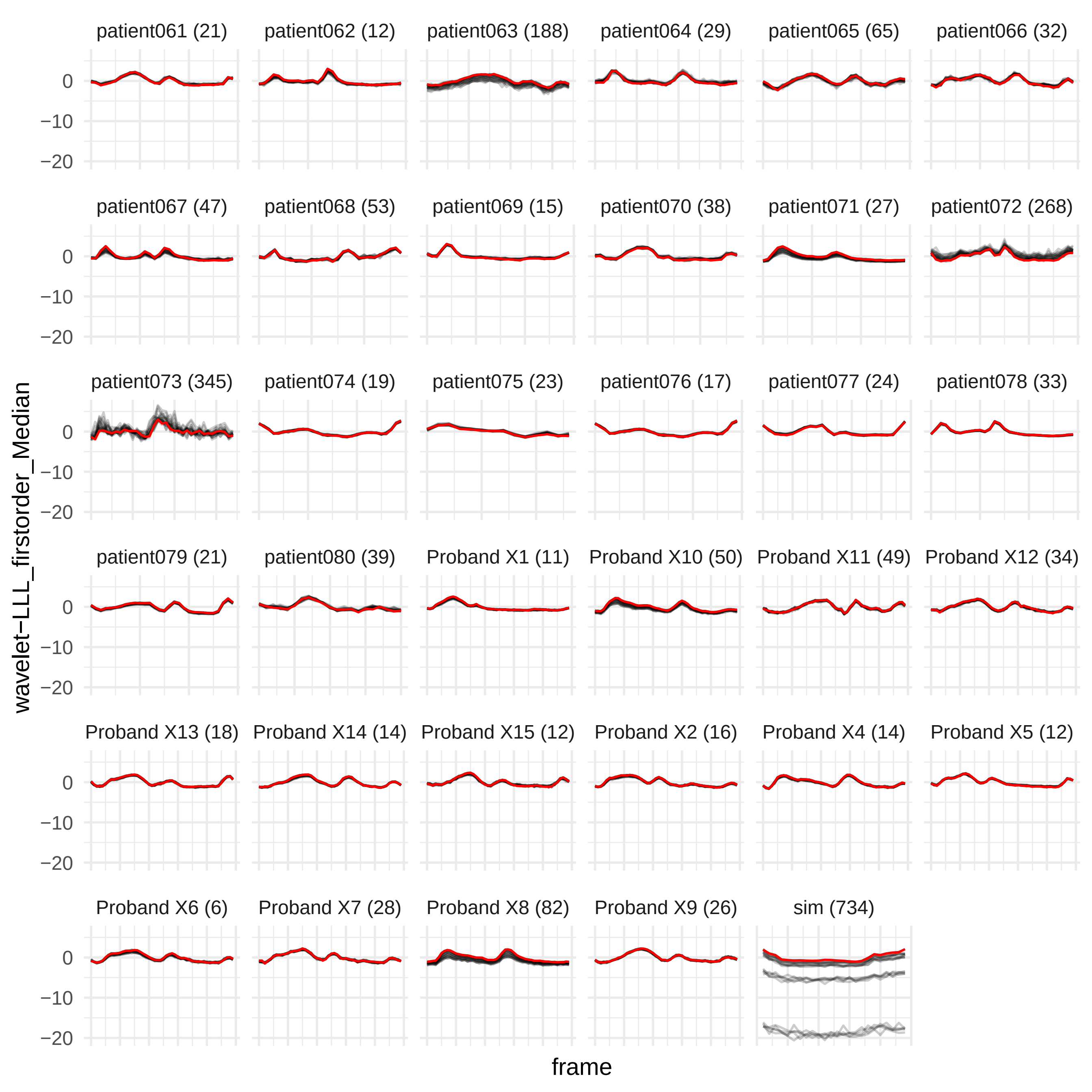

patient061 (21)
patient062 (12)
patient063 (188)
patient064 (29)
patient065 (65)
patient066 (32)
patient067 (47)
patient068 (53)
patient069 (15)
patient070 (38)
patient071 (27)
patient072 (268)
patient073 (345)
patient074 (19)
patient075 (23)
patient076 (17)
patient077 (24)
patient078 (33)
patient079 (21)
patient080 (39)
Proband X1 (11)
Proband X10 (50)
Proband X11 (49)
Proband X12 (34)
Proband X13 (18)
Proband X14 (14)
Proband X15 (12)
Proband X2 (16)
Proband X4 (14)
Proband X5 (12)
Proband X6 (6)
Proband X7 (28)
Proband X8 (82)
Proband X9 (26)
sim (734)
0
−10
−20
wavelet−LLL_firstorder_Median
frame

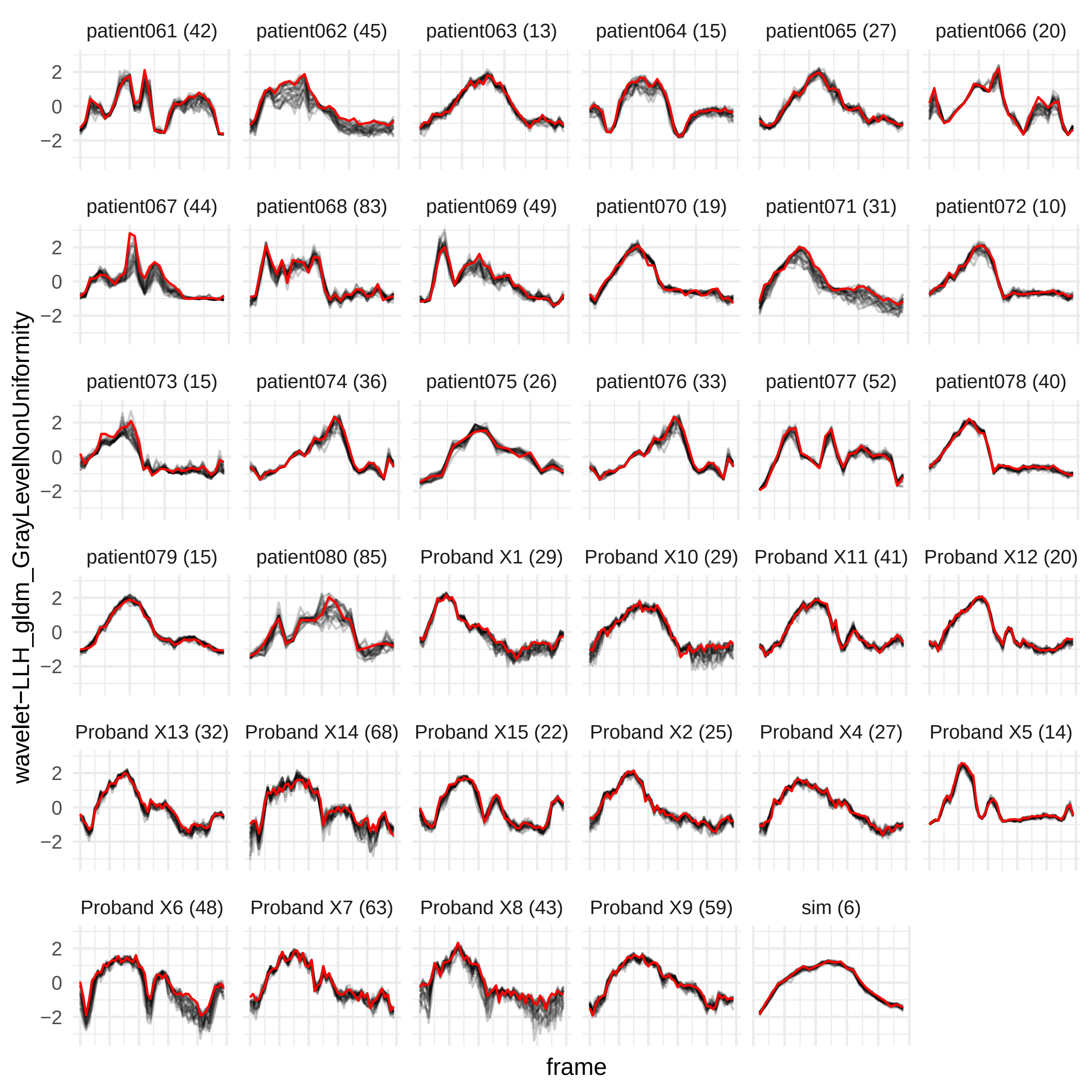

patient061 (42)
patient062 (45)
patient063 (13)
patient064 (15)
patient065 (27)
patient066 (20)
patient067 (44)
patient068 (83)
patient069 (49)
patient070 (19)
patient071 (31)
patient072 (10)
patient073 (15)
patient074 (36)
patient075 (26)
patient076 (33)
patient077 (52)
patient078 (40)
patient079 (15)
patient080 (85)
Proband X1 (29)
Proband X10 (29)
Proband X11 (41)
Proband X12 (20)
Proband X13 (32)
Proband X14 (68)
Proband X15 (22)
Proband X2 (25)
Proband X4 (27)
Proband X5 (14)
Proband X6 (48)
Proband X7 (63)
Proband X8 (43)
Proband X9 (59)
sim (6)
wavelet−LLH_gldm_GrayLevelNonUniformity
frame
2
0
−2

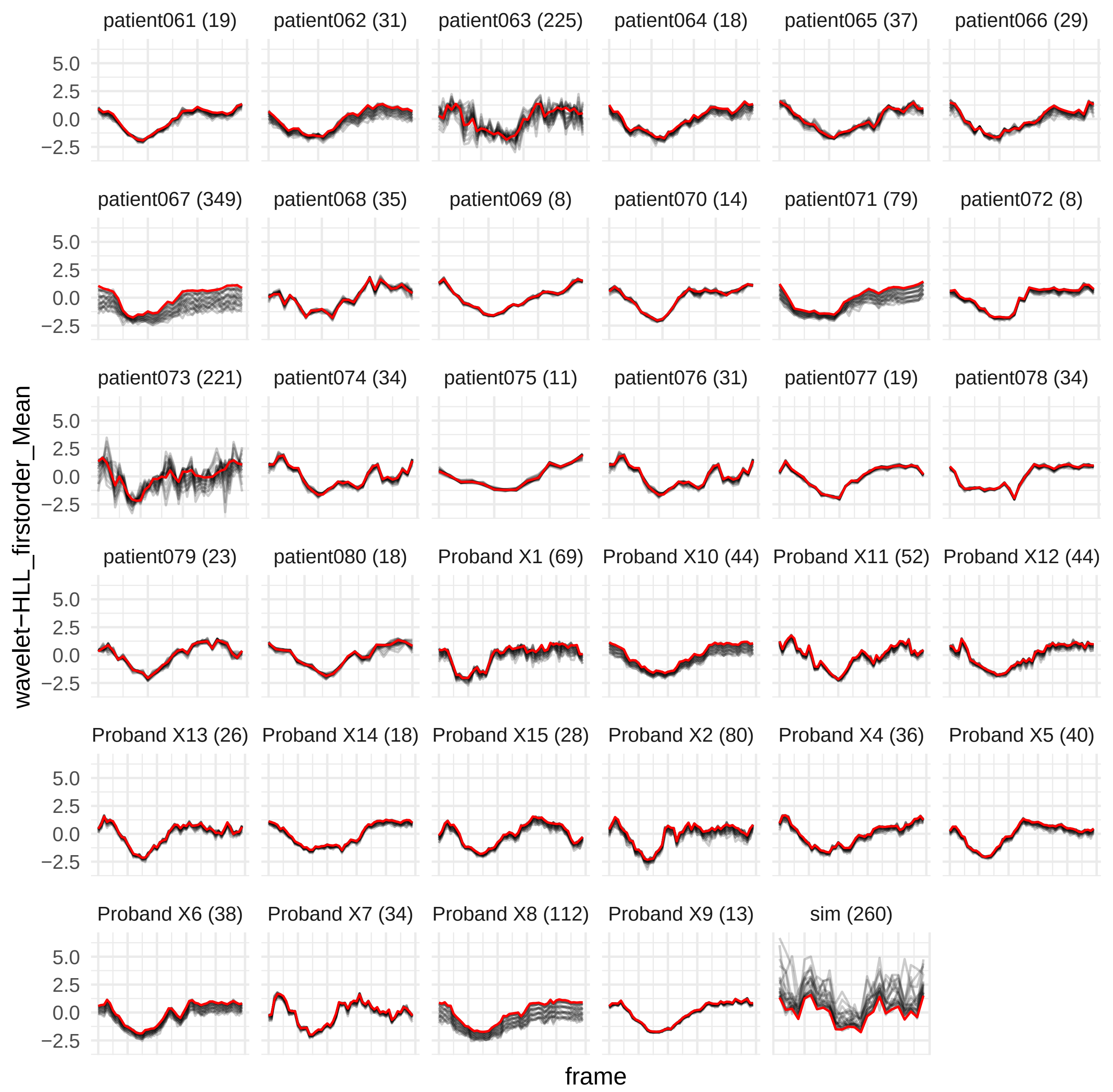

patient061 (19)
patient062 (31)
patient063 (225)
patient064 (18)
patient065 (37)
patient066 (29)
patient067 (349)
patient068 (35)
patient069 (8)
patient070 (14)
patient071 (79)
patient072 (8)
patient073 (221)
patient074 (34)
patient075 (11)
patient076 (31)
patient077 (19)
patient078 (34)
patient079 (23)
patient080 (18)
Proband X1 (69)
Proband X10 (44)
Proband X11 (52)
Proband X12 (44)
Proband X13 (26)
Proband X14 (18)
Proband X15 (28)
Proband X2 (80)
Proband X4 (36)
Proband X5 (40)
Proband X6 (38)
Proband X7 (34)
Proband X8 (112)
Proband X9 (13)
sim (260)
5.0
2.5
0.0
−2.5
wavelet−HLL_firstorder_Mean
frame

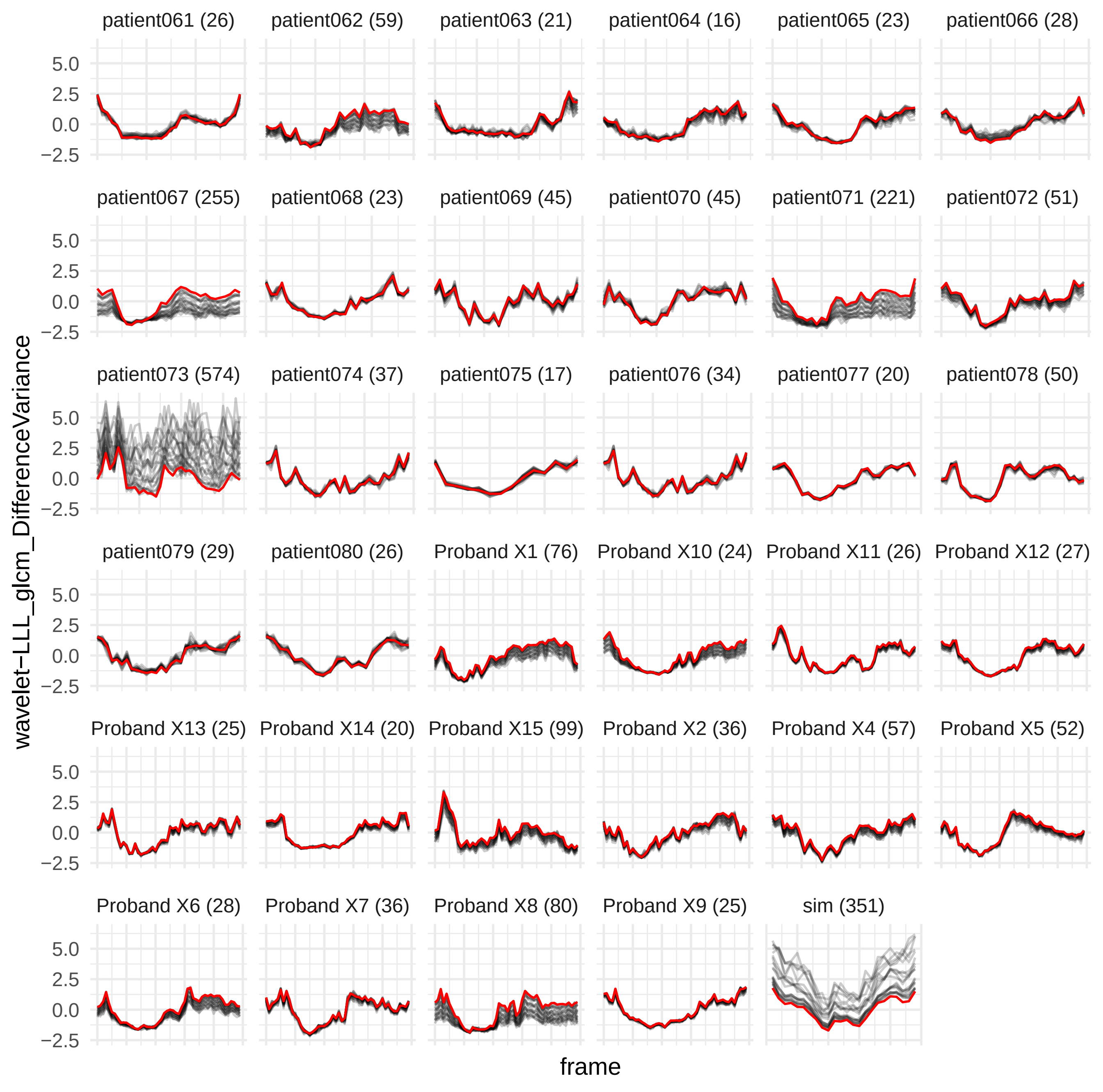
patient061 (26)
patient062 (59)
patient063 (21)
patient064 (16)
patient065 (23)
patient066 (28)
patient067 (255)
patient068 (23)
patient069 (45)
patient070 (45)
patient071 (221)
patient072 (51)
patient073 (574)
patient074 (37)
patient075 (17)
patient076 (34)
patient077 (20)
patient078 (50)
patient079 (29)
patient080 (26)
Proband X1 (76)
Proband X10 (24)
Proband X11 (26)
Proband X12 (27)
Proband X13 (25)
Proband X14 (20)
Proband X15 (99)
Proband X2 (36)
Proband X4 (57)
Proband X5 (52)
Proband X6 (28)
Proband X7 (36)
Proband X8 (80)
Proband X9 (25)
sim (351)
5.0
2.5
0.0
−2.5
wavelet−LLL_glcm_DifferenceVariance
frame

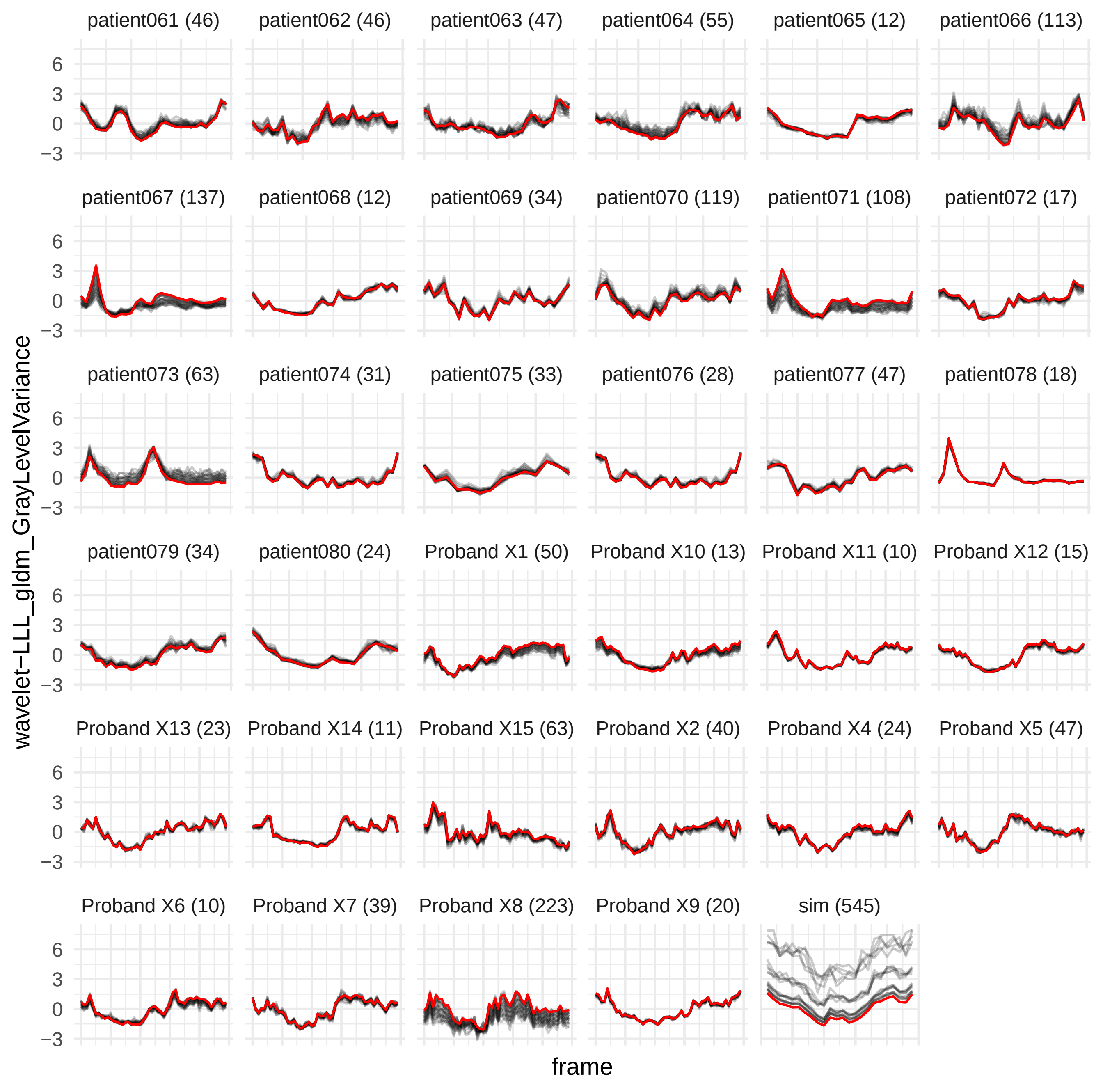
patient061 (46)
patient062 (46)
patient063 (47)
patient064 (55)
patient065 (12)
patient066 (113)
patient067 (137)
patient068 (12)
patient069 (34)
patient070 (119)
patient071 (108)
patient072 (17)
patient073 (63)
patient074 (31)
patient075 (33)
patient076 (28)
patient077 (47)
patient078 (18)
patient079 (34)
patient080 (24)
Proband X1 (50)
Proband X10 (13)
Proband X11 (10)
Proband X12 (15)
Proband X13 (23)
Proband X14 (11)
Proband X15 (63)
Proband X2 (40)
Proband X4 (24)
Proband X5 (47)
Proband X6 (10)
Proband X7 (39)
Proband X8 (223)
Proband X9 (20)
sim (545)
6
3
0
−3
wavelet−LLL_gldm_GrayLevelVariance
frame

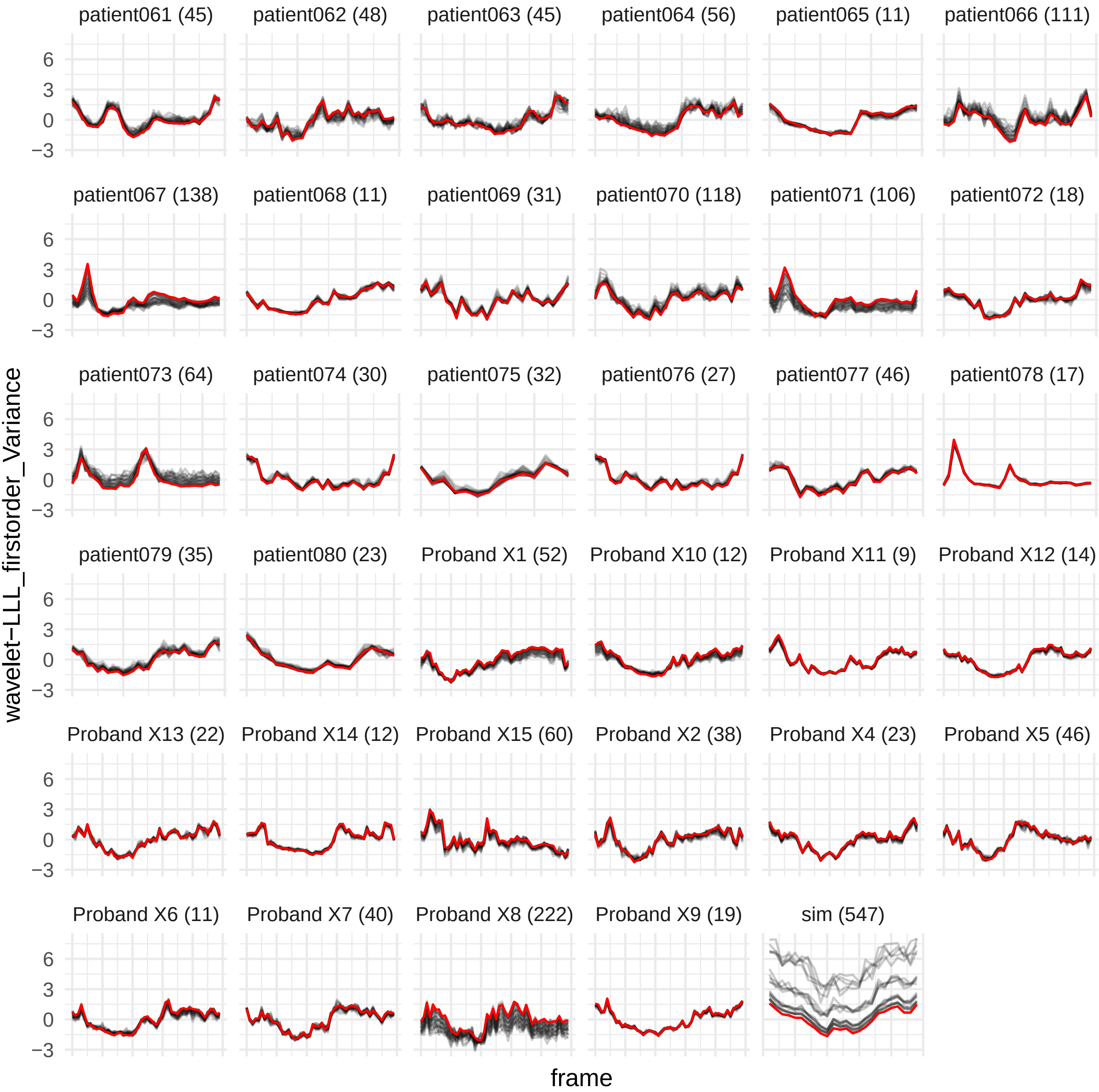

patient061 (45)
patient062 (48)
patient063 (45)
patient064 (56)
patient065 (11)
patient066 (111)
patient067 (138)
patient068 (11)
patient069 (31)
patient070 (118)
patient071 (106)
patient072 (18)
patient073 (64)
patient074 (30)
patient075 (32)
patient076 (27)
patient077 (46)
patient078 (17)
patient079 (35)
patient080 (23)
Proband X1 (52)
Proband X10 (12)
Proband X11 (9)
Proband X12 (14)
Proband X13 (22)
Proband X14 (12)
Proband X15 (60)
Proband X2 (38)
Proband X4 (23)
Proband X5 (46)
Proband X6 (11)
Proband X7 (40)
Proband X8 (222)
Proband X9 (19)
sim (547)
6
3
0
−3
wavelet−LLL_firstorder_Variance
frame

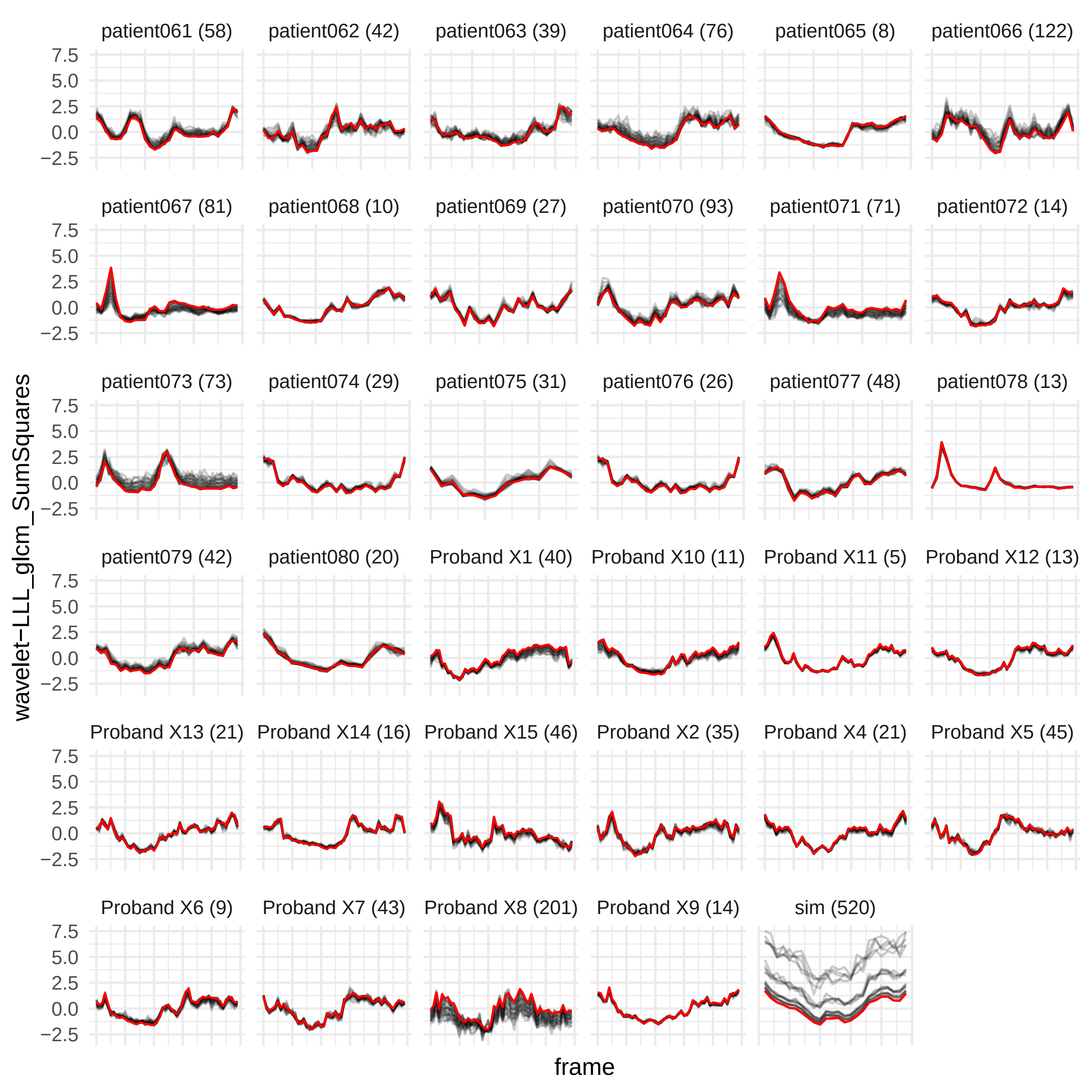

patient061 (58)
patient062 (42)
patient063 (39)
patient064 (76)
patient065 (8)
patient066 (122)
patient067 (81)
patient068 (10)
patient069 (27)
patient070 (93)
patient071 (71)
patient072 (14)
patient073 (73)
patient074 (29)
patient075 (31)
patient076 (26)
patient077 (48)
patient078 (13)
patient079 (42)
patient080 (20)
Proband X1 (40)
Proband X10 (11)
Proband X11 (5)
Proband X12 (13)
Proband X13 (21)
Proband X14 (16)
Proband X15 (46)
Proband X2 (35)
Proband X4 (21)
Proband X5 (45)
Proband X6 (9)
Proband X7 (43)
Proband X8 (201)
Proband X9 (14)
sim (520)
7.5
5.0
2.5
0.0
−2.5
wavelet−LLL_glcm_SumSquares
frame

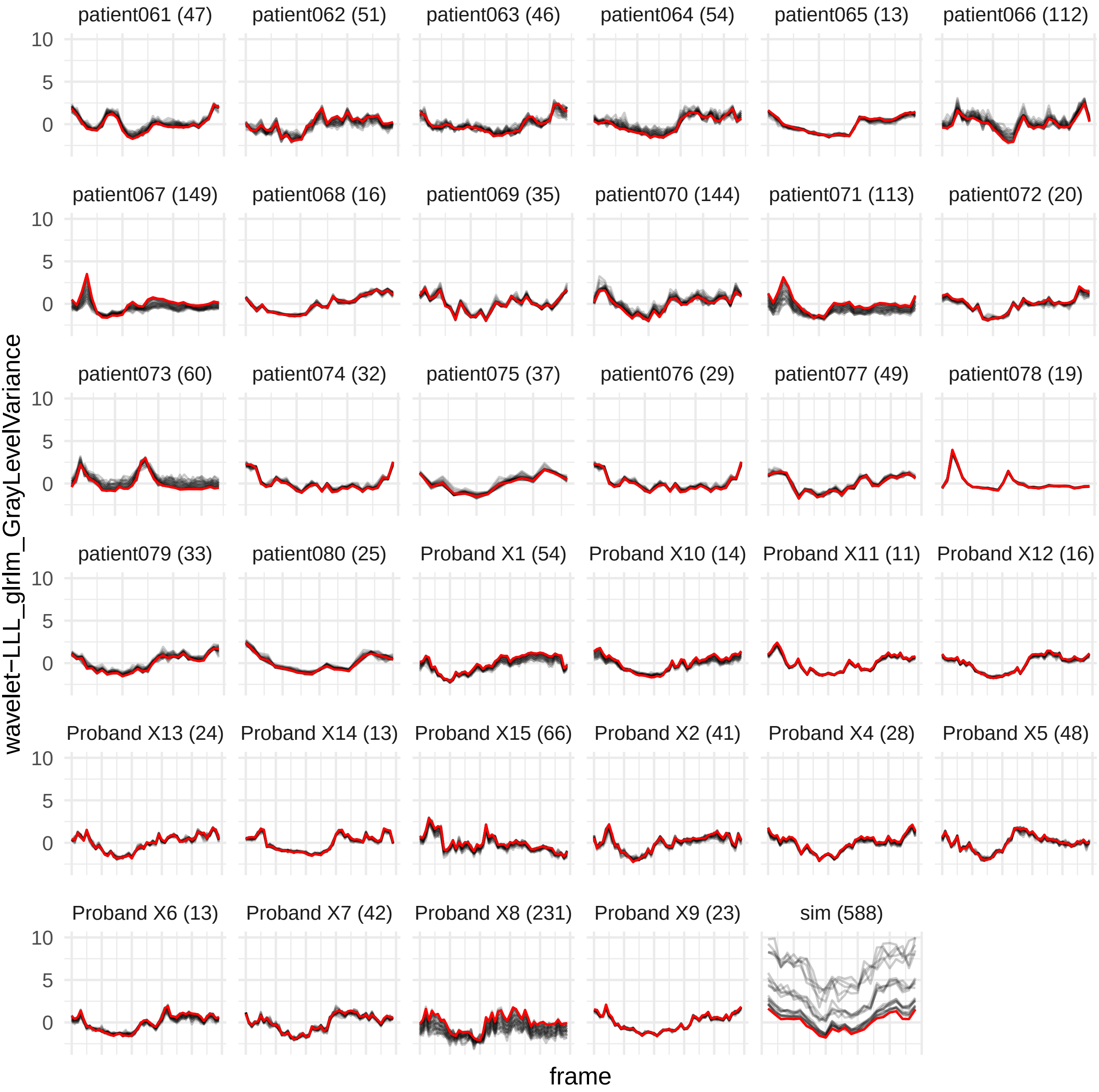

patient061 (47)
patient062 (51)
patient063 (46)
patient064 (54)
patient065 (13)
patient066 (112)
patient067 (149)
patient068 (16)
patient069 (35)
patient070 (144)
patient071 (113)
patient072 (20)
patient073 (60)
patient074 (32)
patient075 (37)
patient076 (29)
patient077 (49)
patient078 (19)
patient079 (33)
patient080 (25)
Proband X1 (54)
Proband X10 (14)
Proband X11 (11)
Proband X12 (16)
Proband X13 (24)
Proband X14 (13)
Proband X15 (66)
Proband X2 (41)
Proband X4 (28)
Proband X5 (48)
Proband X6 (13)
Proband X7 (42)
Proband X8 (231)
Proband X9 (23)
sim (588)
10
5
0
wavelet−LLL_glrlm_GrayLevelVariance
frame

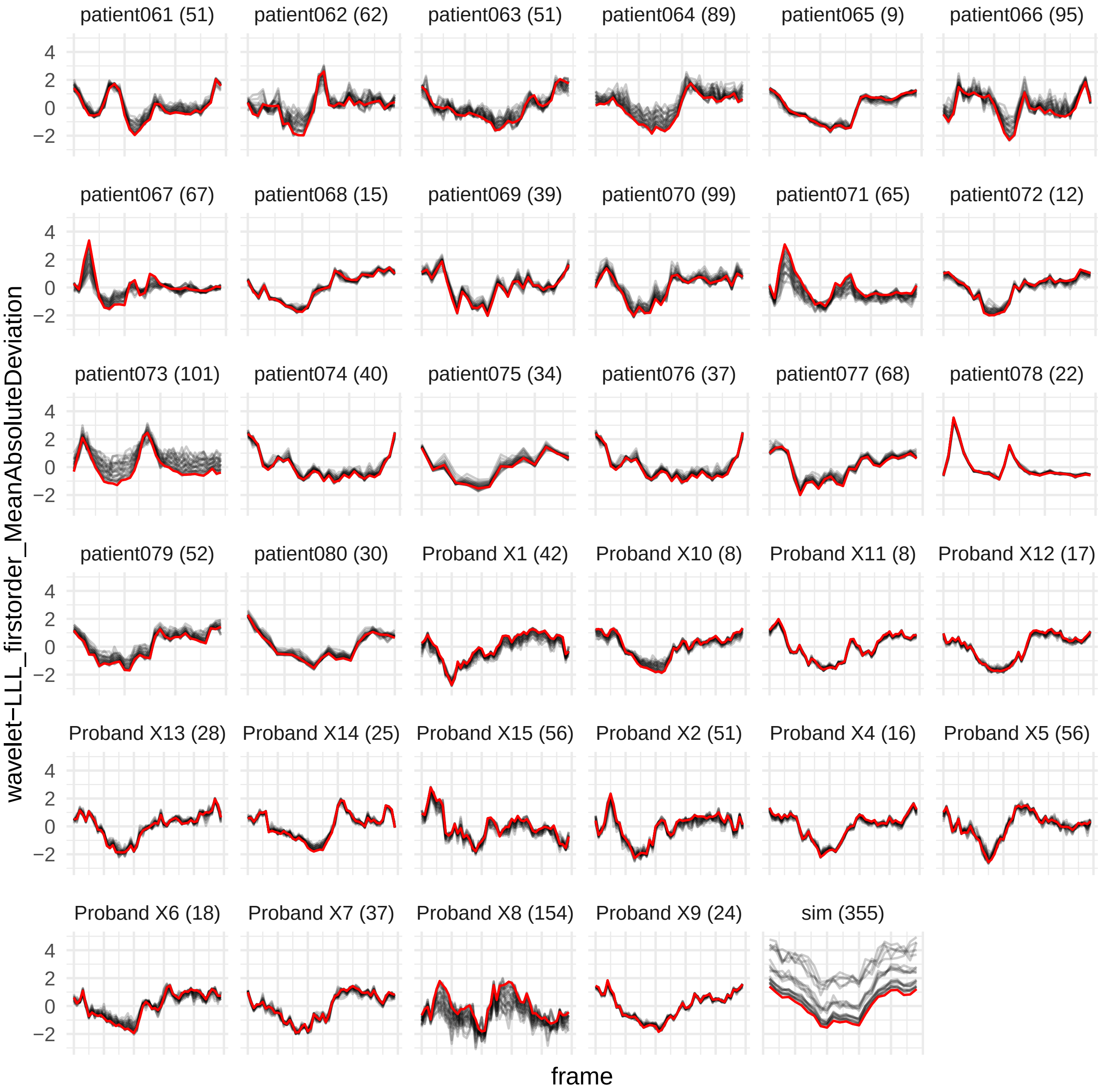

patient061 (51)
patient062 (62)
patient063 (51)
patient064 (89)
patient065 (9)
patient066 (95)
patient067 (67)
patient068 (15)
patient069 (39)
patient070 (99)
patient071 (65)
patient072 (12)
patient073 (101)
patient074 (40)
patient075 (34)
patient076 (37)
patient077 (68)
patient078 (22)
patient079 (52)
patient080 (30)
Proband X1 (42)
Proband X10 (8)
Proband X11 (8)
Proband X12 (17)
Proband X13 (28)
Proband X14 (25)
Proband X15 (56)
Proband X2 (51)
Proband X4 (16)
Proband X5 (56)
Proband X6 (18)
Proband X7 (37)
Proband X8 (154)
Proband X9 (24)
sim (355)
wavelet−LLL_firstorder_MeanAbsoluteDeviation
frame
4
2
0
−2

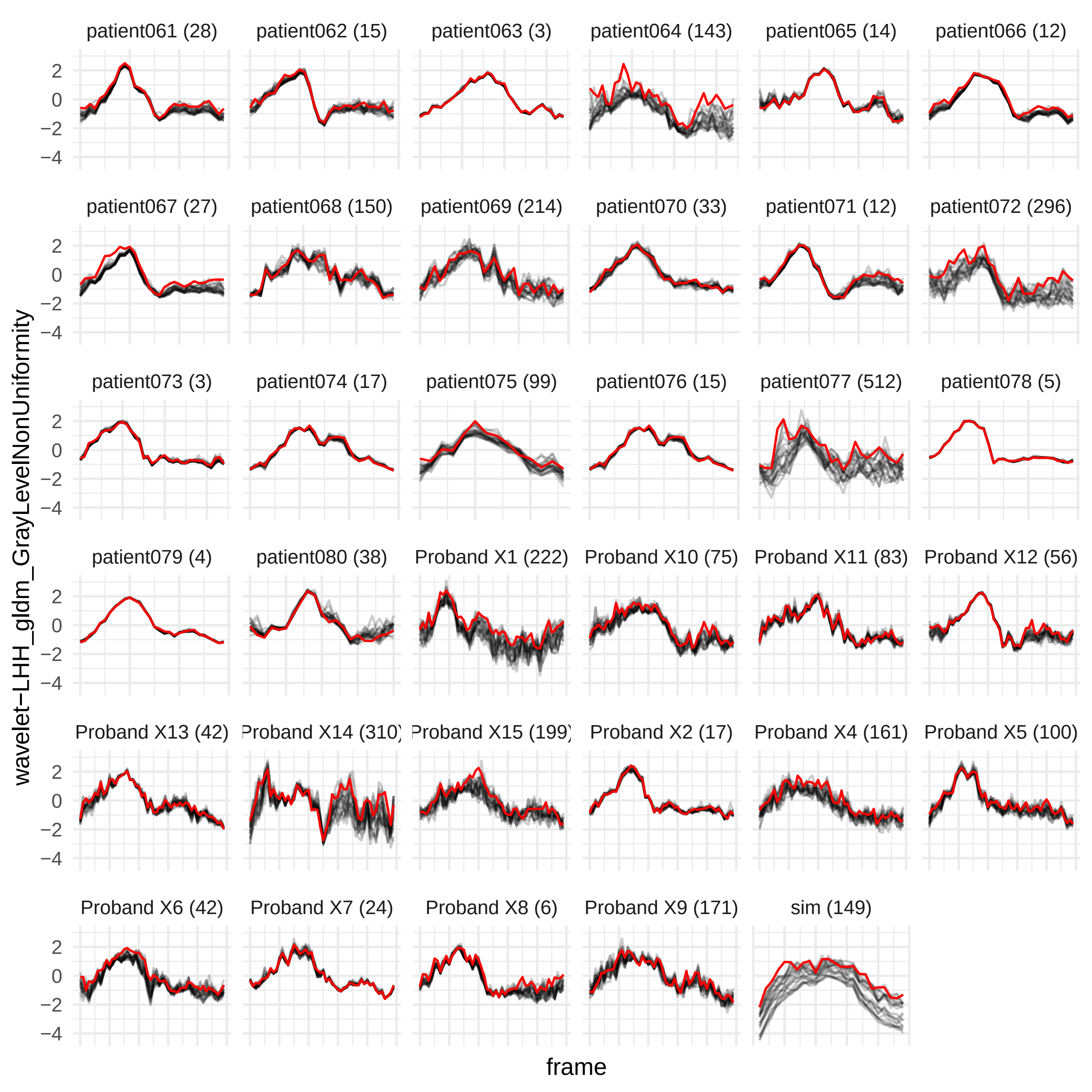

patient061 (28)
patient062 (15)
patient063 (3)
patient064 (143)
patient065 (14)
patient066 (12)
patient067 (27)
patient068 (150)
patient069 (214)
patient070 (33)
patient071 (12)
patient072 (296)
patient073 (3)
patient074 (17)
patient075 (99)
patient076 (15)
patient077 (512)
patient078 (5)
patient079 (4)
patient080 (38)
Proband X1 (222)
Proband X10 (75)
Proband X11 (83)
Proband X12 (56)
Proband X13 (42)
Proband X14 (310)
Proband X15 (199)
Proband X2 (17)
Proband X4 (161)
Proband X5 (100)
Proband X6 (42)
Proband X7 (24)
Proband X8 (6)
Proband X9 (171)
sim (149)
wavelet−LHH_gldm_GrayLevelNonUniformity
frame
2
0
−2
−4

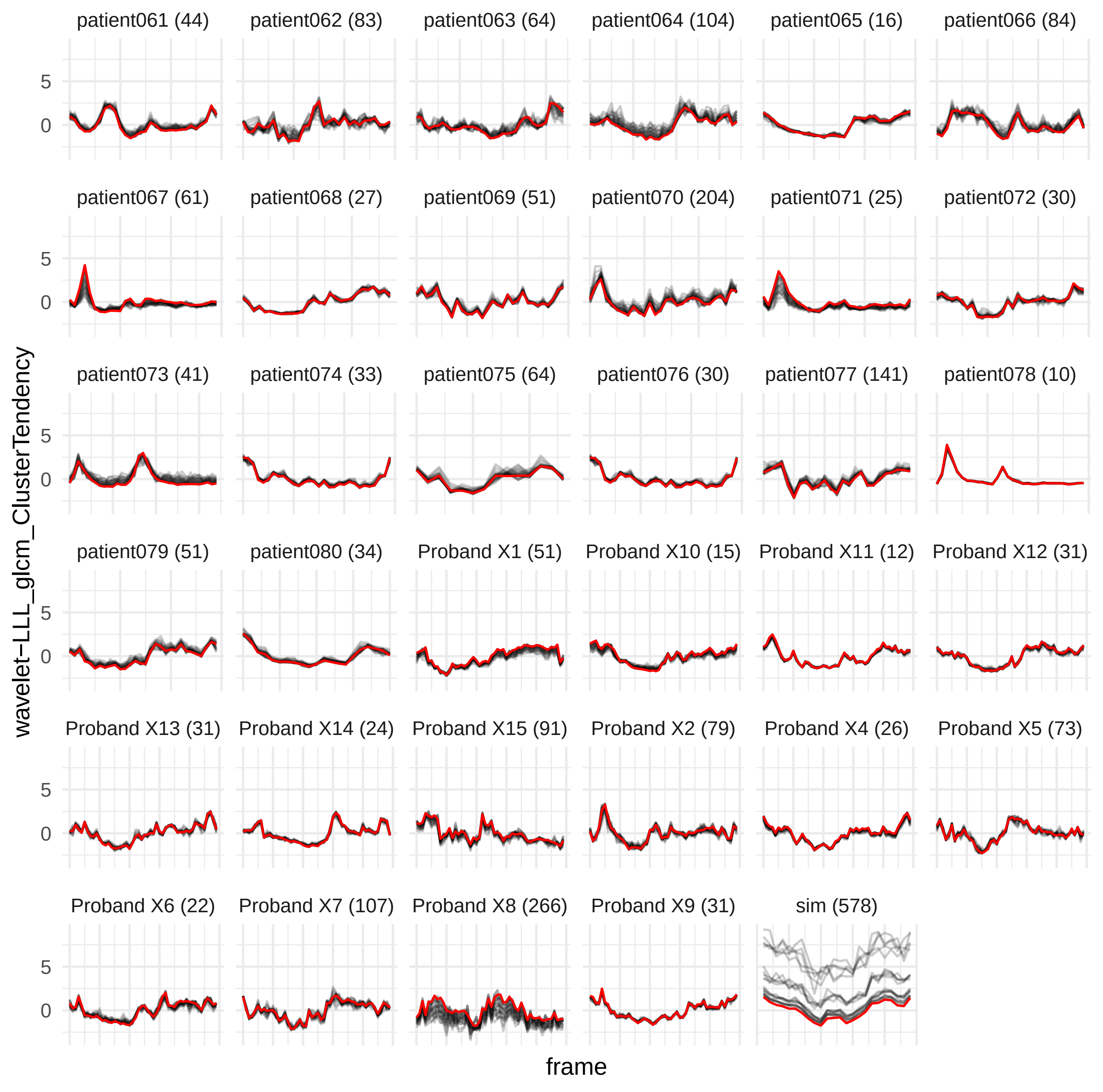
patient061 (44)
patient062 (83)
patient063 (64)
patient064 (104)
patient065 (16)
patient066 (84)
patient067 (61)
patient068 (27)
patient069 (51)
patient070 (204)
patient071 (25)
patient072 (30)
patient073 (41)
patient074 (33)
patient075 (64)
patient076 (30)
patient077 (141)
patient078 (10)
patient079 (51)
patient080 (34)
Proband X1 (51)
Proband X10 (15)
Proband X11 (12)
Proband X12 (31)
Proband X13 (31)
Proband X14 (24)
Proband X15 (91)
Proband X2 (79)
Proband X4 (26)
Proband X5 (73)
Proband X6 (22)
Proband X7 (107)
Proband X8 (266)
Proband X9 (31)
sim (578)
wavelet−LLL_glcm_ClusterTendency
5
0
frame

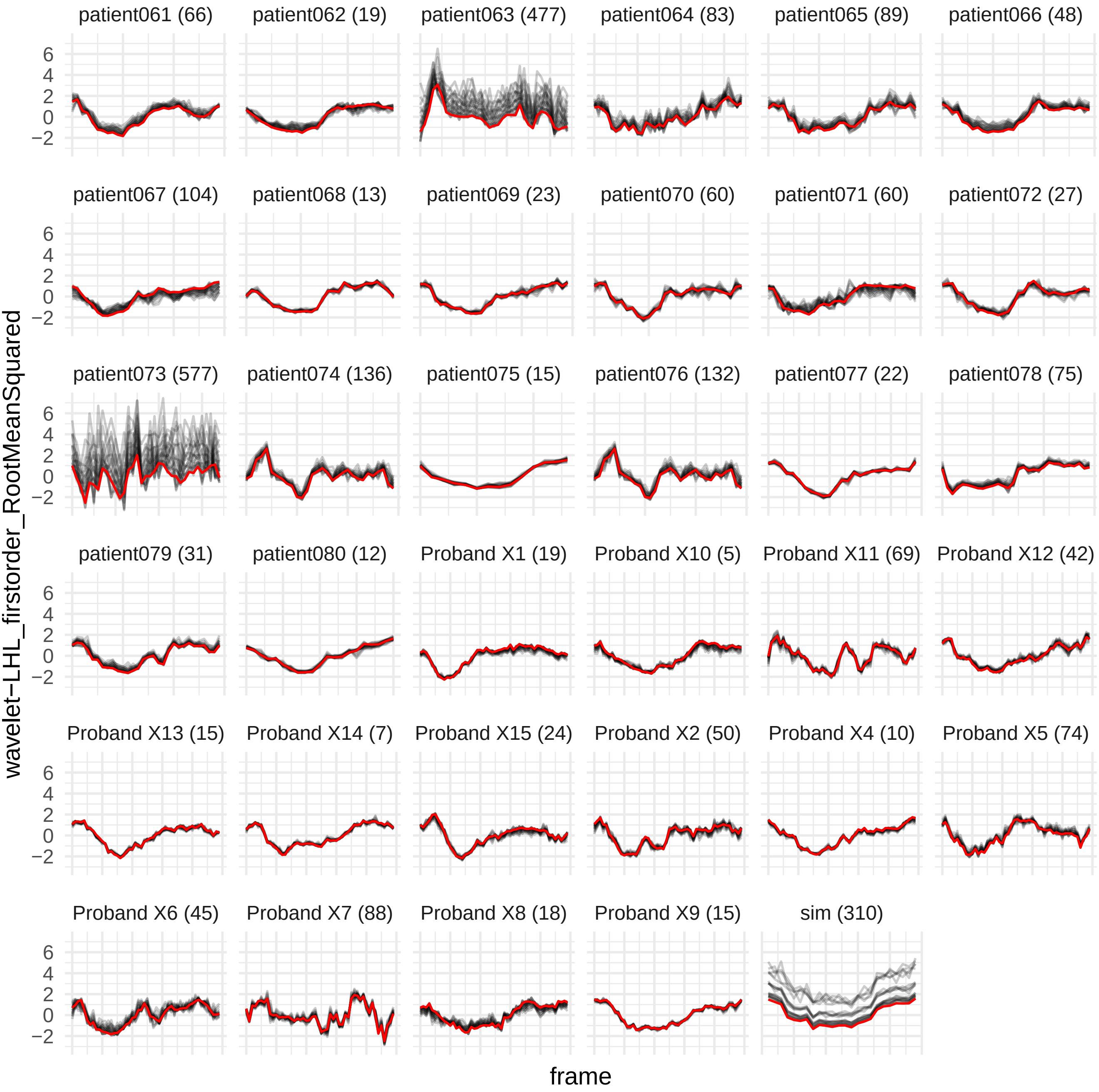

patient061 (66)
patient062 (19)
patient063 (477)
patient064 (83)
patient065 (89)
patient066 (48)
patient067 (104)
patient068 (13)
patient069 (23)
patient070 (60)
patient071 (60)
patient072 (27)
patient073 (577)
patient074 (136)
patient075 (15)
patient076 (132)
patient077 (22)
patient078 (75)
patient079 (31)
patient080 (12)
Proband X1 (19)
Proband X10 (5)
Proband X11 (69)
Proband X12 (42)
Proband X13 (15)
Proband X14 (7)
Proband X15 (24)
Proband X2 (50)
Proband X4 (10)
Proband X5 (74)
Proband X6 (45)
Proband X7 (88)
Proband X8 (18)
Proband X9 (15)
sim (310)
wavelet−LHL_firstorder_RootMeanSquared
frame
6
4
2
0
−2

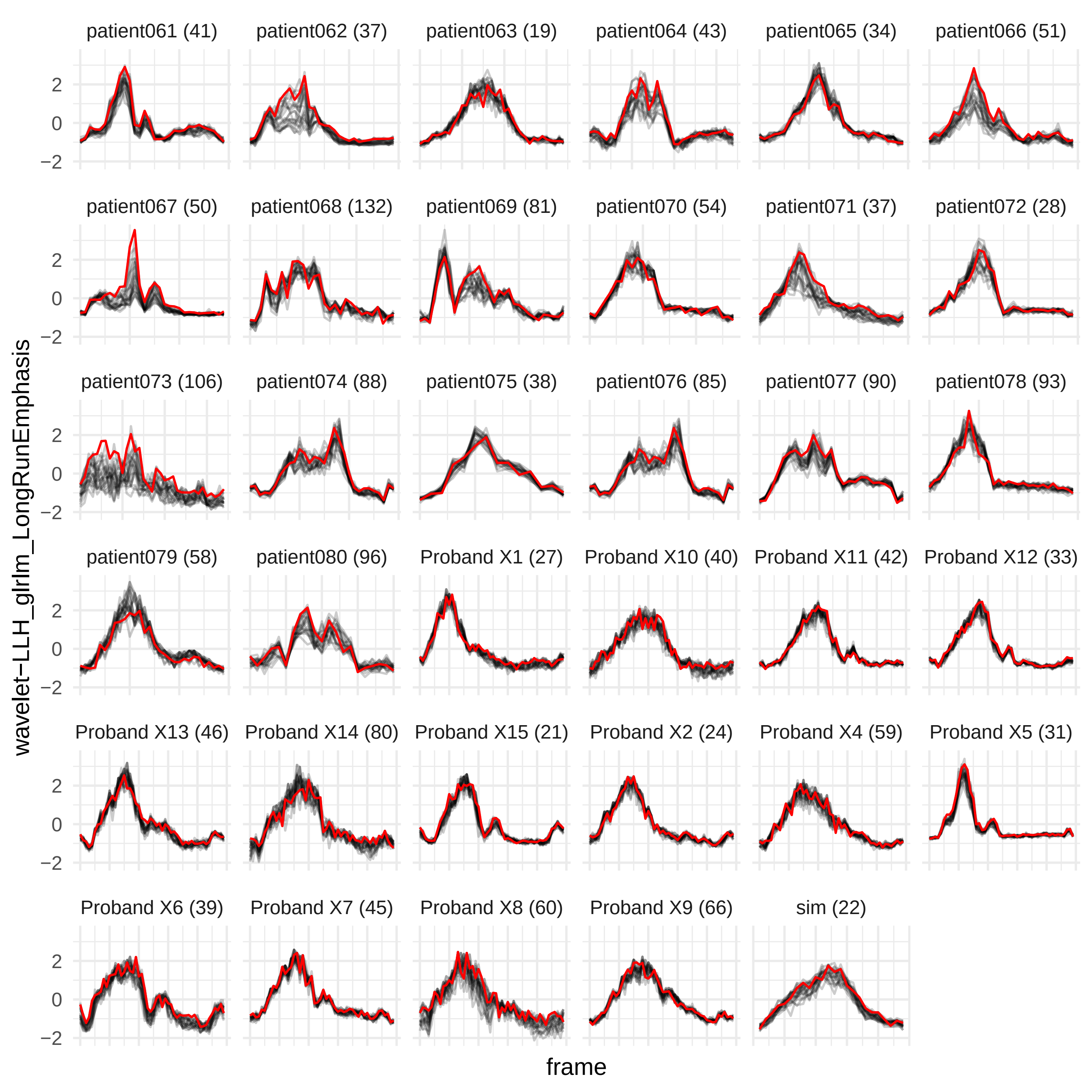

patient061 (41)
patient062 (37)
patient063 (19)
patient064 (43)
patient065 (34)
patient066 (51)
patient067 (50)
patient068 (132)
patient069 (81)
patient070 (54)
patient071 (37)
patient072 (28)
patient073 (106)
patient074 (88)
patient075 (38)
patient076 (85)
patient077 (90)
patient078 (93)
patient079 (58)
patient080 (96)
Proband X1 (27)
Proband X10 (40)
Proband X11 (42)
Proband X12 (33)
Proband X13 (46)
Proband X14 (80)
Proband X15 (21)
Proband X2 (24)
Proband X4 (59)
Proband X5 (31)
Proband X6 (39)
Proband X7 (45)
Proband X8 (60)
Proband X9 (66)
sim (22)
wavelet−LLH_glrlm_LongRunEmphasis
frame
2
0
−2

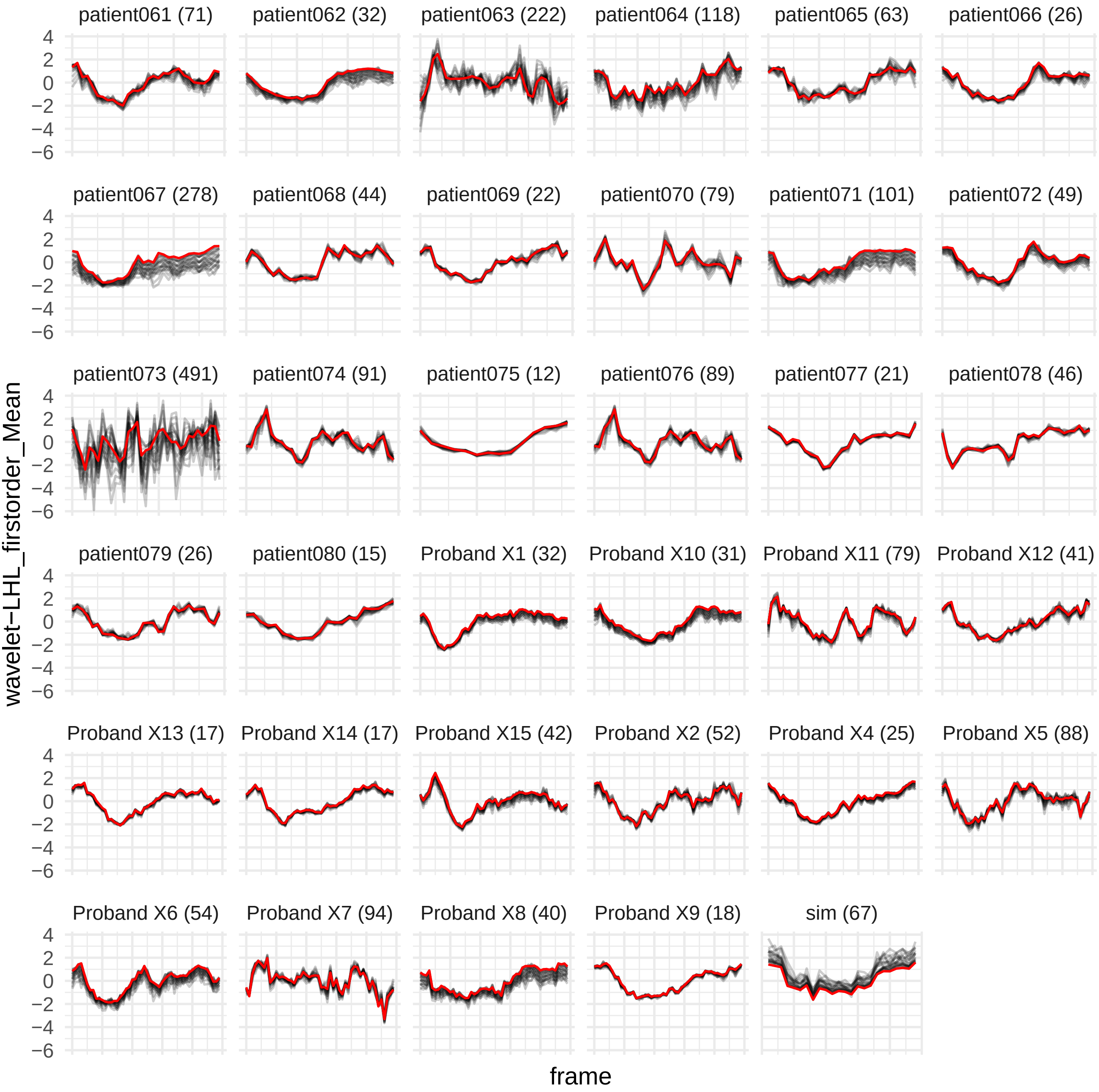

patient061 (71)
patient062 (32)
patient063 (222)
patient064 (118)
patient065 (63)
patient066 (26)
patient067 (278)
patient068 (44)
patient069 (22)
patient070 (79)
patient071 (101)
patient072 (49)
patient073 (491)
patient074 (91)
patient075 (12)
patient076 (89)
patient077 (21)
patient078 (46)
patient079 (26)
patient080 (15)
Proband X1 (32)
Proband X10 (31)
Proband X11 (79)
Proband X12 (41)
Proband X13 (17)
Proband X14 (17)
Proband X15 (42)
Proband X2 (52)
Proband X4 (25)
Proband X5 (88)
Proband X6 (54)
Proband X7 (94)
Proband X8 (40)
Proband X9 (18)
sim (67)
4
2
0
−2
−4
−6
wavelet−LHL_firstorder_Mean
frame

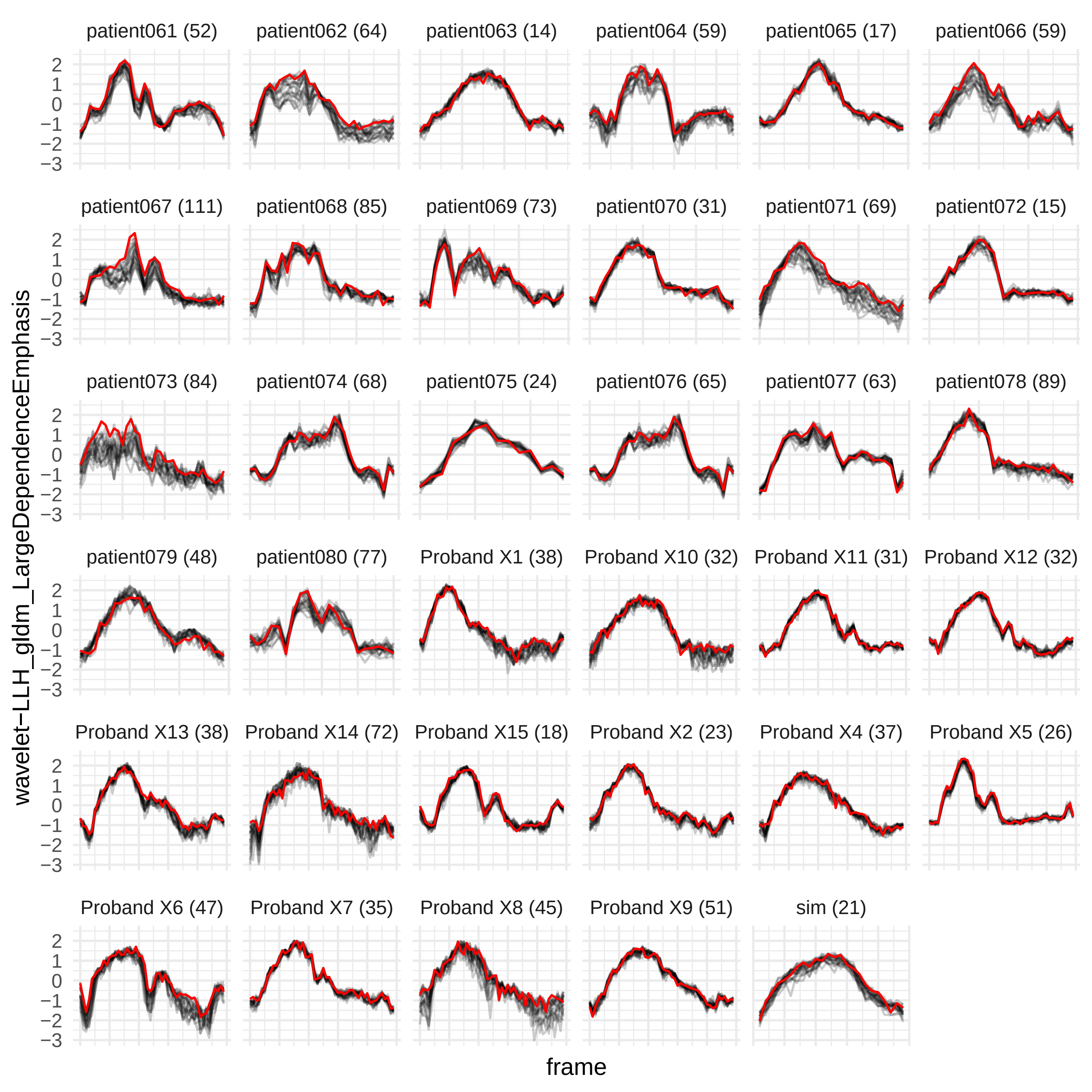

patient061 (52)
patient062 (64)
patient063 (14)
patient064 (59)
patient065 (17)
patient066 (59)
patient067 (111)
patient068 (85)
patient069 (73)
patient070 (31)
patient071 (69)
patient072 (15)
patient073 (84)
patient074 (68)
patient075 (24)
patient076 (65)
patient077 (63)
patient078 (89)
patient079 (48)
patient080 (77)
Proband X1 (38)
Proband X10 (32)
Proband X11 (31)
Proband X12 (32)
Proband X13 (38)
Proband X14 (72)
Proband X15 (18)
Proband X2 (23)
Proband X4 (37)
Proband X5 (26)
Proband X6 (47)
Proband X7 (35)
Proband X8 (45)
Proband X9 (51)
sim (21)
wavelet−LLH_gldm_LargeDependenceEmphasis
frame

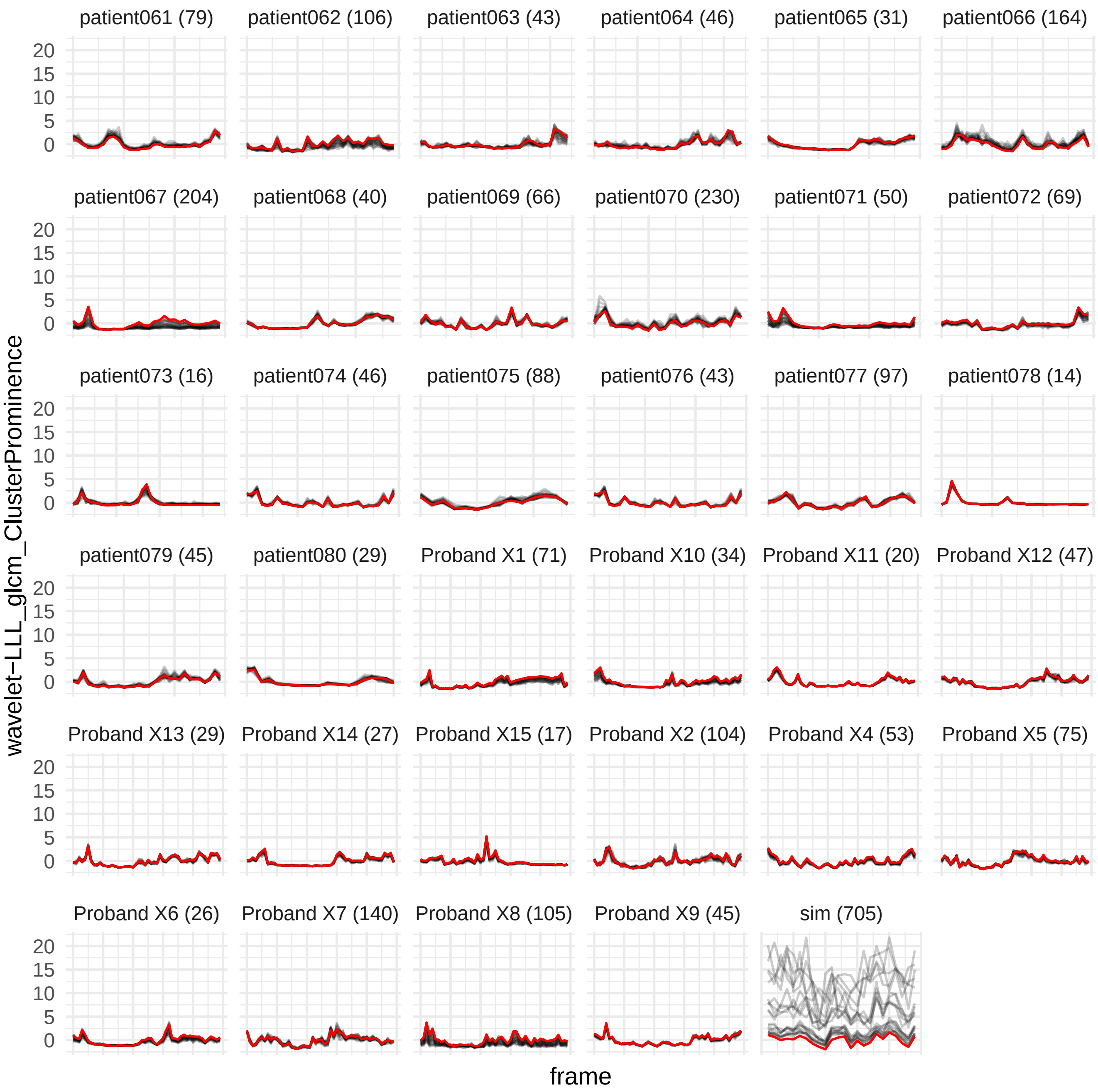
patient061 (79)
patient062 (106)
patient063 (43)
patient064 (46)
patient065 (31)
patient066 (164)
patient067 (204)
patient068 (40)
patient069 (66)
patient070 (230)
patient071 (50)
patient072 (69)
patient073 (16)
patient074 (46)
patient075 (88)
patient076 (43)
patient077 (97)
patient078 (14)
patient079 (45)
patient080 (29)
Proband X1 (71)
Proband X10 (34)
Proband X11 (20)
Proband X12 (47)
Proband X13 (29)
Proband X14 (27)
Proband X15 (17)
Proband X2 (104)
Proband X4 (53)
Proband X5 (75)
Proband X6 (26)
Proband X7 (140)
Proband X8 (105)
Proband X9 (45)
sim (705)
0
5
10
15
20
wavelet−LLL_glcm_ClusterProminence
frame

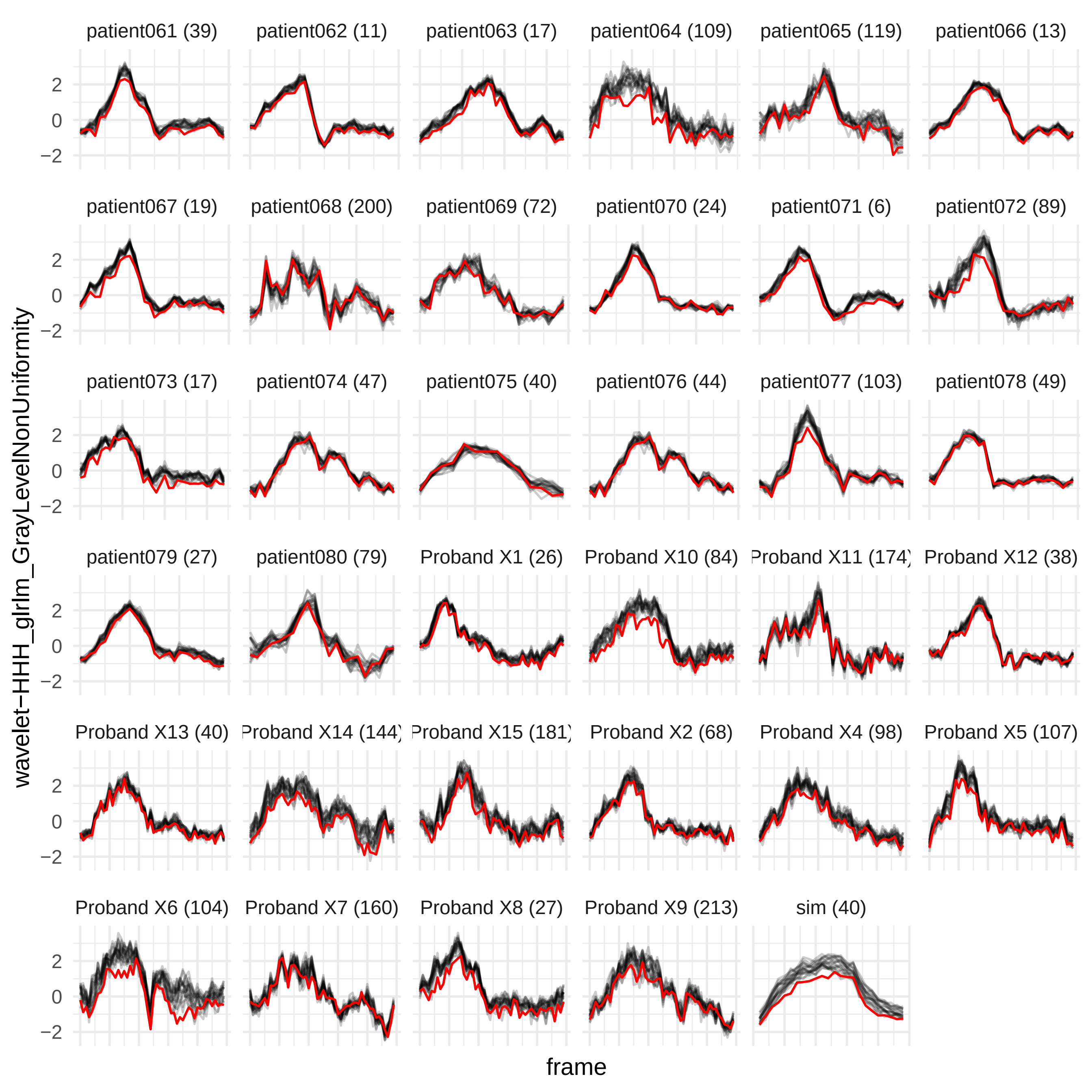

patient061 (39)
patient062 (11)
patient063 (17)
patient064 (109)
patient065 (119)
patient066 (13)
patient067 (19)
patient068 (200)
patient069 (72)
patient070 (24)
patient071 (6)
patient072 (89)
patient073 (17)
patient074 (47)
patient075 (40)
patient076 (44)
patient077 (103)
patient078 (49)
patient079 (27)
patient080 (79)
Proband X1 (26)
Proband X10 (84)
Proband X11 (174)
Proband X12 (38)
Proband X13 (40)
Proband X14 (144)
Proband X15 (181)
Proband X2 (68)
Proband X4 (98)
Proband X5 (107)
Proband X6 (104)
Proband X7 (160)
Proband X8 (27)
Proband X9 (213)
sim (40)
2
0
−2
wavelet−HHH_glrlm_GrayLevelNonUniformity
frame

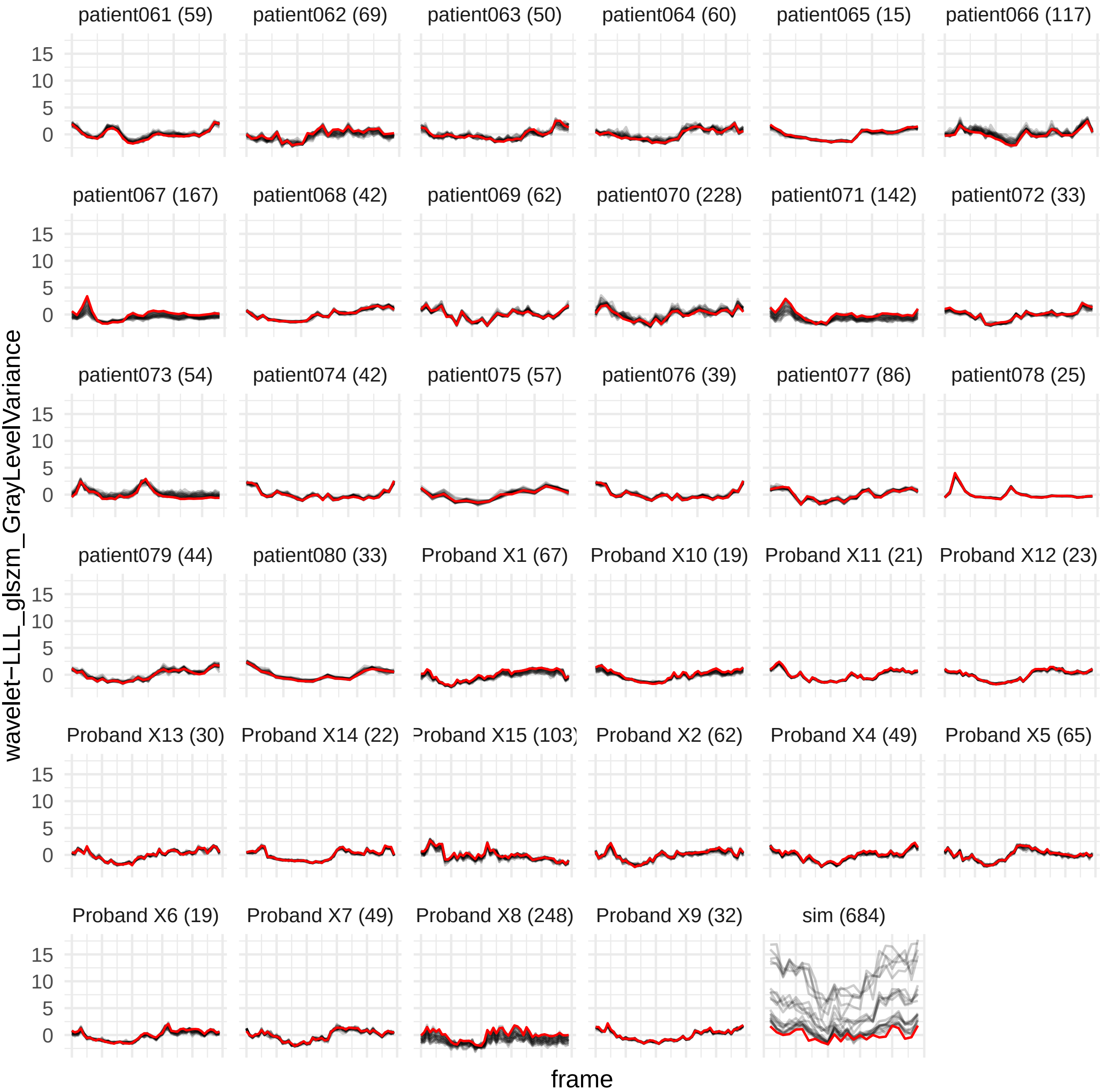
patient061 (59)
patient062 (69)
patient063 (50)
patient064 (60)
patient065 (15)
patient066 (117)
patient067 (167)
patient068 (42)
patient069 (62)
patient070 (228)
patient071 (142)
patient072 (33)
patient073 (54)
patient074 (42)
patient075 (57)
patient076 (39)
patient077 (86)
patient078 (25)
patient079 (44)
patient080 (33)
Proband X1 (67)
Proband X10 (19)
Proband X11 (21)
Proband X12 (23)
Proband X13 (30)
Proband X14 (22)
Proband X15 (103)
Proband X2 (62)
Proband X4 (49)
Proband X5 (65)
Proband X6 (19)
Proband X7 (49)
Proband X8 (248)
Proband X9 (32)
sim (684)
15
10
5
0
wavelet−LLL_glszm_GrayLevelVariance
frame

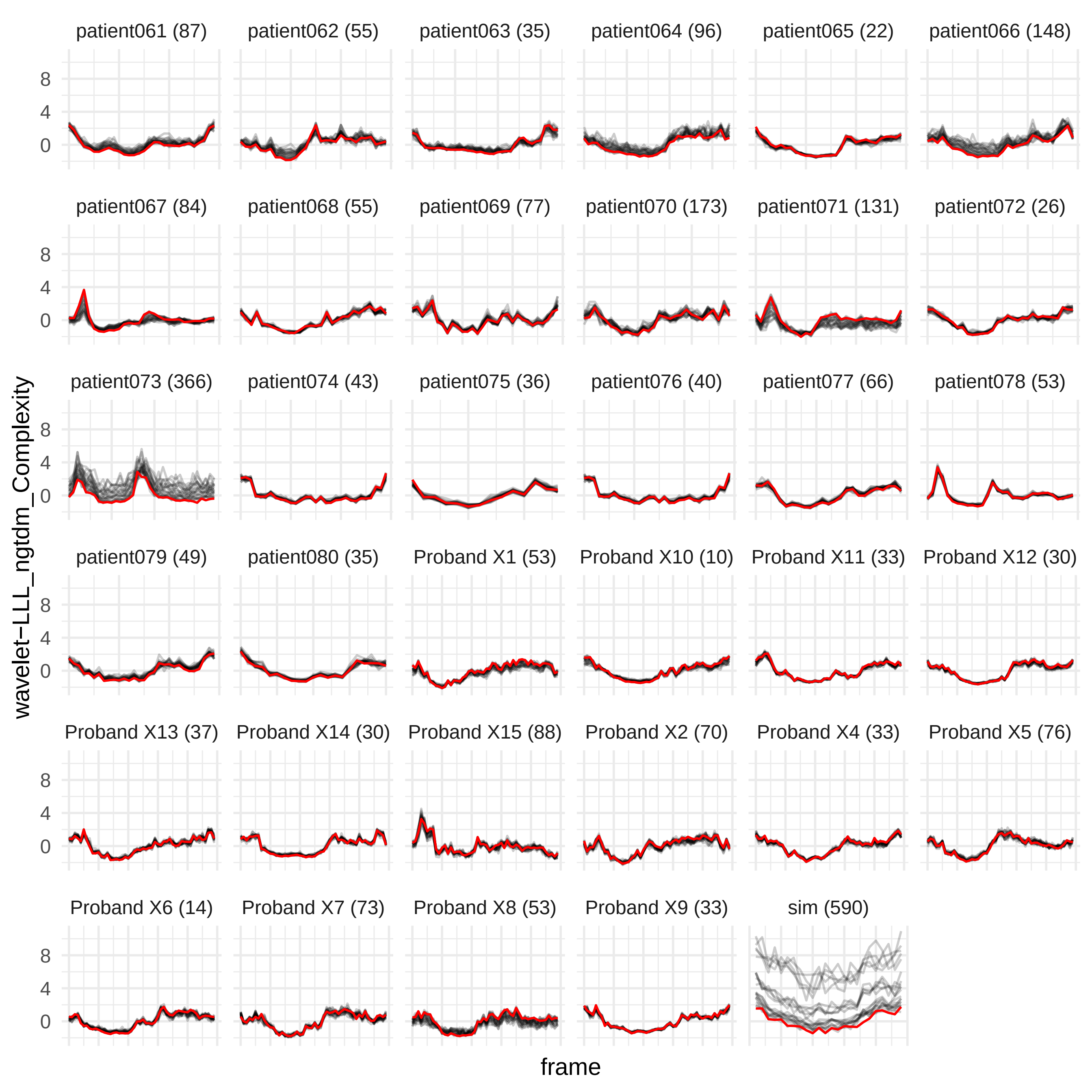

patient061 (87)
patient062 (55)
patient063 (35)
patient064 (96)
patient065 (22)
patient066 (148)
patient067 (84)
patient068 (55)
patient069 (77)
patient070 (173)
patient071 (131)
patient072 (26)
patient073 (366)
patient074 (43)
patient075 (36)
patient076 (40)
patient077 (66)
patient078 (53)
patient079 (49)
patient080 (35)
Proband X1 (53)
Proband X10 (10)
Proband X11 (33)
Proband X12 (30)
Proband X13 (37)
Proband X14 (30)
Proband X15 (88)
Proband X2 (70)
Proband X4 (33)
Proband X5 (76)
Proband X6 (14)
Proband X7 (73)
Proband X8 (53)
Proband X9 (33)
sim (590)
wavelet−LLL_ngtdm_Complexity
8
4
0
frame

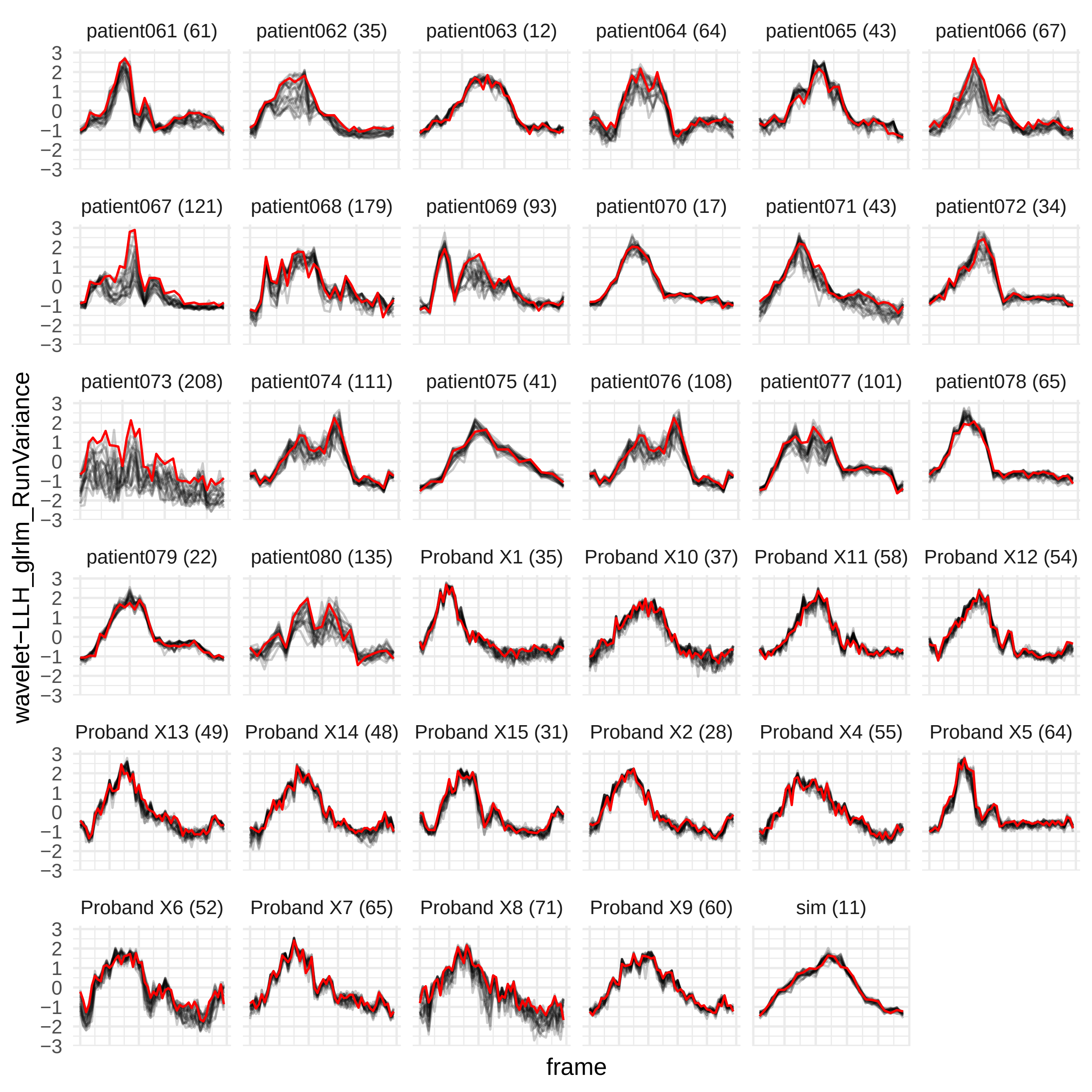

patient061 (61)
patient062 (35)
patient063 (12)
patient064 (64)
patient065 (43)
patient066 (67)
patient067 (121)
patient068 (179)
patient069 (93)
patient070 (17)
patient071 (43)
patient072 (34)
patient073 (208)
patient074 (111)
patient075 (41)
patient076 (108)
patient077 (101)
patient078 (65)
patient079 (22)
patient080 (135)
Proband X1 (35)
Proband X10 (37)
Proband X11 (58)
Proband X12 (54)
Proband X13 (49)
Proband X14 (48)
Proband X15 (31)
Proband X2 (28)
Proband X4 (55)
Proband X5 (64)
Proband X6 (52)
Proband X7 (65)
Proband X8 (71)
Proband X9 (60)
sim (11)
wavelet−LLH_glrlm_RunVariance
frame
3
2
1
0
−1
−2
−3

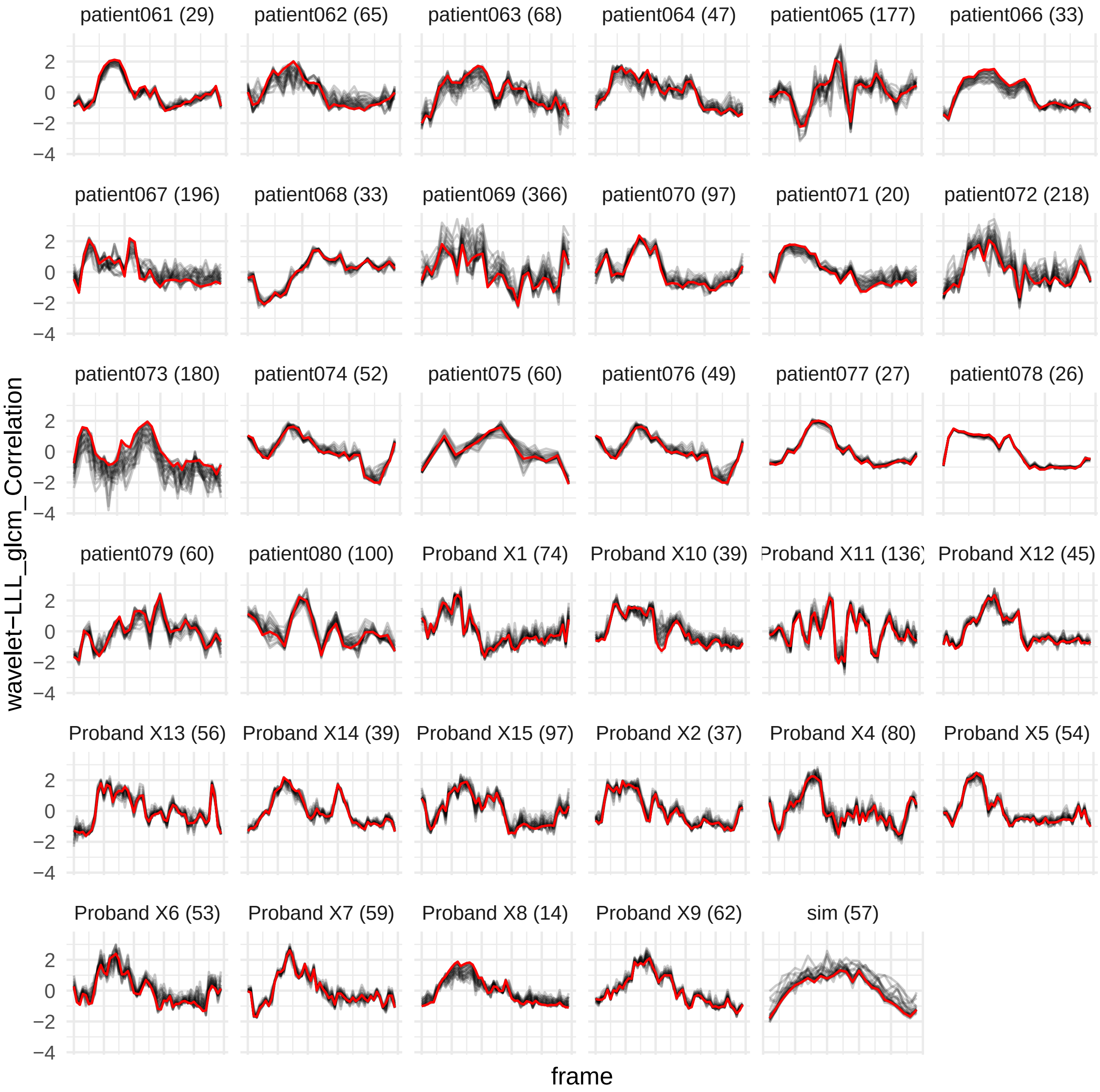
patient061 (29)
patient062 (65)
patient063 (68)
patient064 (47)
patient065 (177)
patient066 (33)
patient067 (196)
patient068 (33)
patient069 (366)
patient070 (97)
patient071 (20)
patient072 (218)
patient073 (180)
patient074 (52)
patient075 (60)
patient076 (49)
patient077 (27)
patient078 (26)
patient079 (60)
patient080 (100)
Proband X1 (74)
Proband X10 (39)
Proband X11 (136)
Proband X12 (45)
Proband X13 (56)
Proband X14 (39)
Proband X15 (97)
Proband X2 (37)
Proband X4 (80)
Proband X5 (54)
Proband X6 (53)
Proband X7 (59)
Proband X8 (14)
Proband X9 (62)
sim (57)
wavelet−LLL_glcm_Correlation
frame
2
0
−2
−4

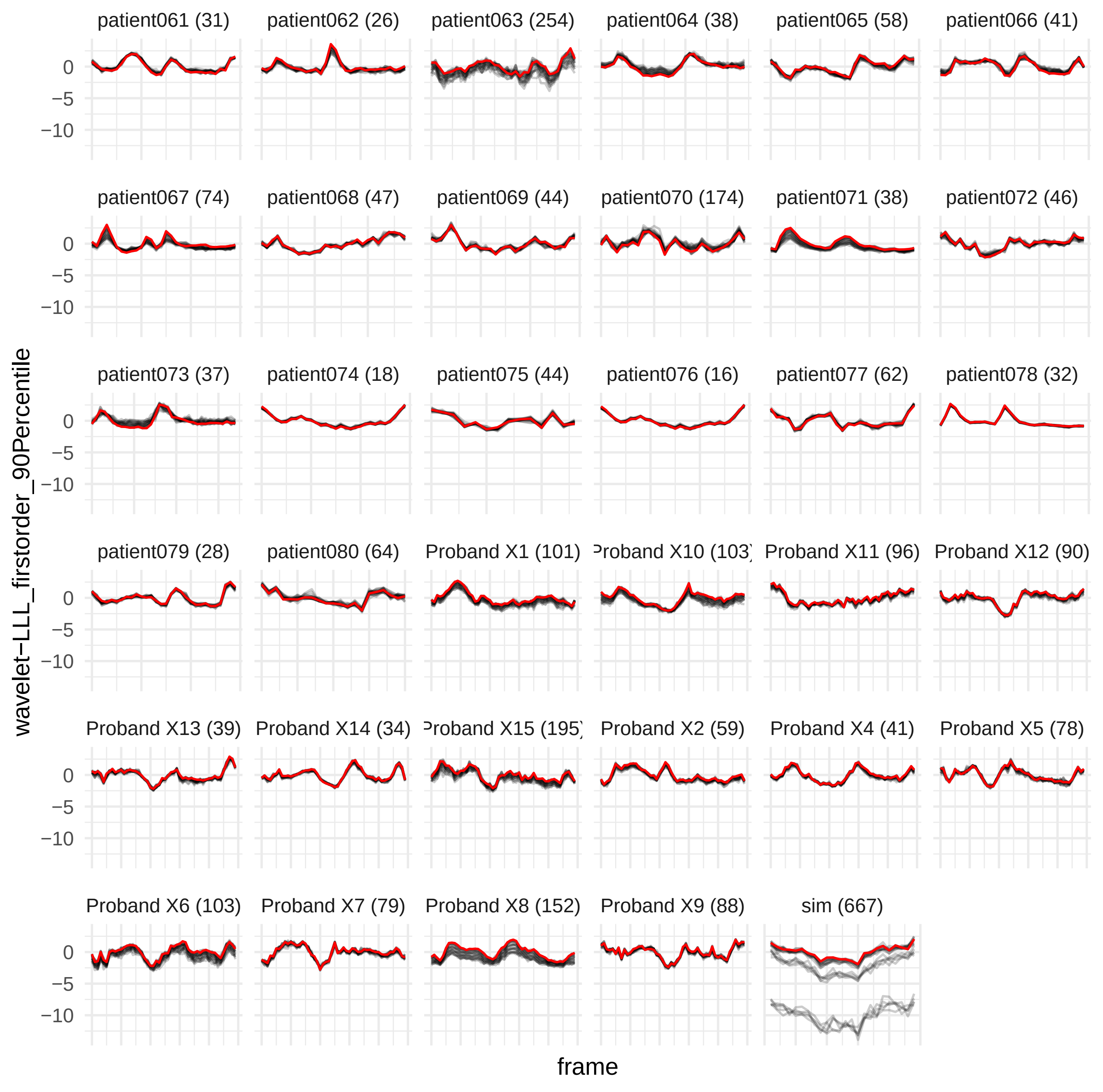

patient061 (31)
patient062 (26)
patient063 (254)
patient064 (38)
patient065 (58)
patient066 (41)
patient067 (74)
patient068 (47)
patient069 (44)
patient070 (174)
patient071 (38)
patient072 (46)
patient073 (37)
patient074 (18)
patient075 (44)
patient076 (16)
patient077 (62)
patient078 (32)
patient079 (28)
patient080 (64)
Proband X1 (101)
Proband X10 (103)
Proband X11 (96)
Proband X12 (90)
Proband X13 (39)
Proband X14 (34)
Proband X15 (195)
Proband X2 (59)
Proband X4 (41)
Proband X5 (78)
Proband X6 (103)
Proband X7 (79)
Proband X8 (152)
Proband X9 (88)
sim (667)
0
−5
−10
wavelet−LLL_firstorder_90Percentile
frame

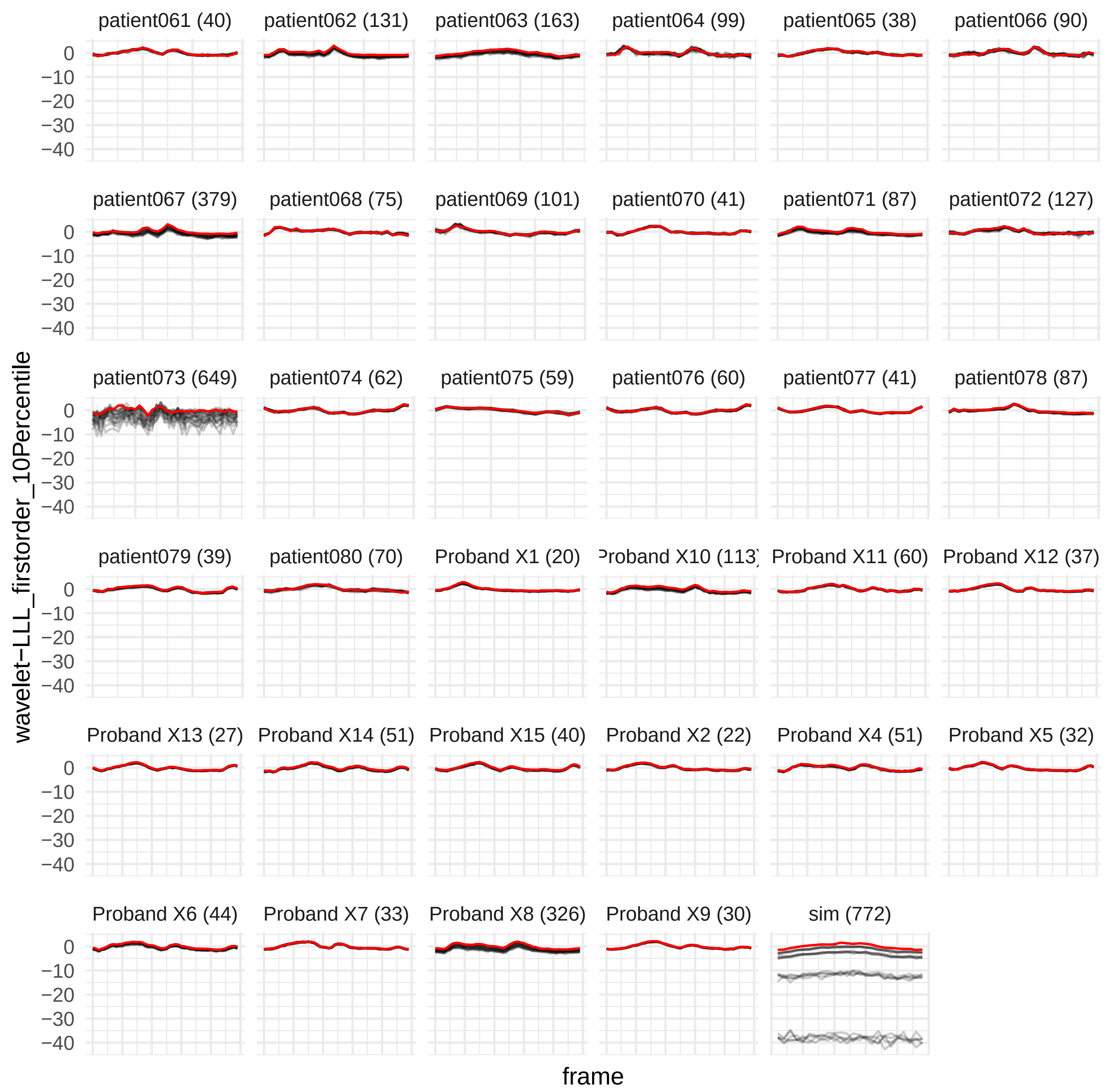

patient061 (40)
patient062 (131)
patient063 (163)
patient064 (99)
patient065 (38)
patient066 (90)
patient067 (379)
patient068 (75)
patient069 (101)
patient070 (41)
patient071 (87)
patient072 (127)
patient073 (649)
patient074 (62)
patient075 (59)
patient076 (60)
patient077 (41)
patient078 (87)
patient079 (39)
patient080 (70)
Proband X1 (20)
Proband X10 (113)
Proband X11 (60)
Proband X12 (37)
Proband X13 (27)
Proband X14 (51)
Proband X15 (40)
Proband X2 (22)
Proband X4 (51)
Proband X5 (32)
Proband X6 (44)
Proband X7 (33)
Proband X8 (326)
Proband X9 (30)
sim (772)
0
−10
−20
−30
−40
wavelet−LLL_firstorder_10Percentile
frame

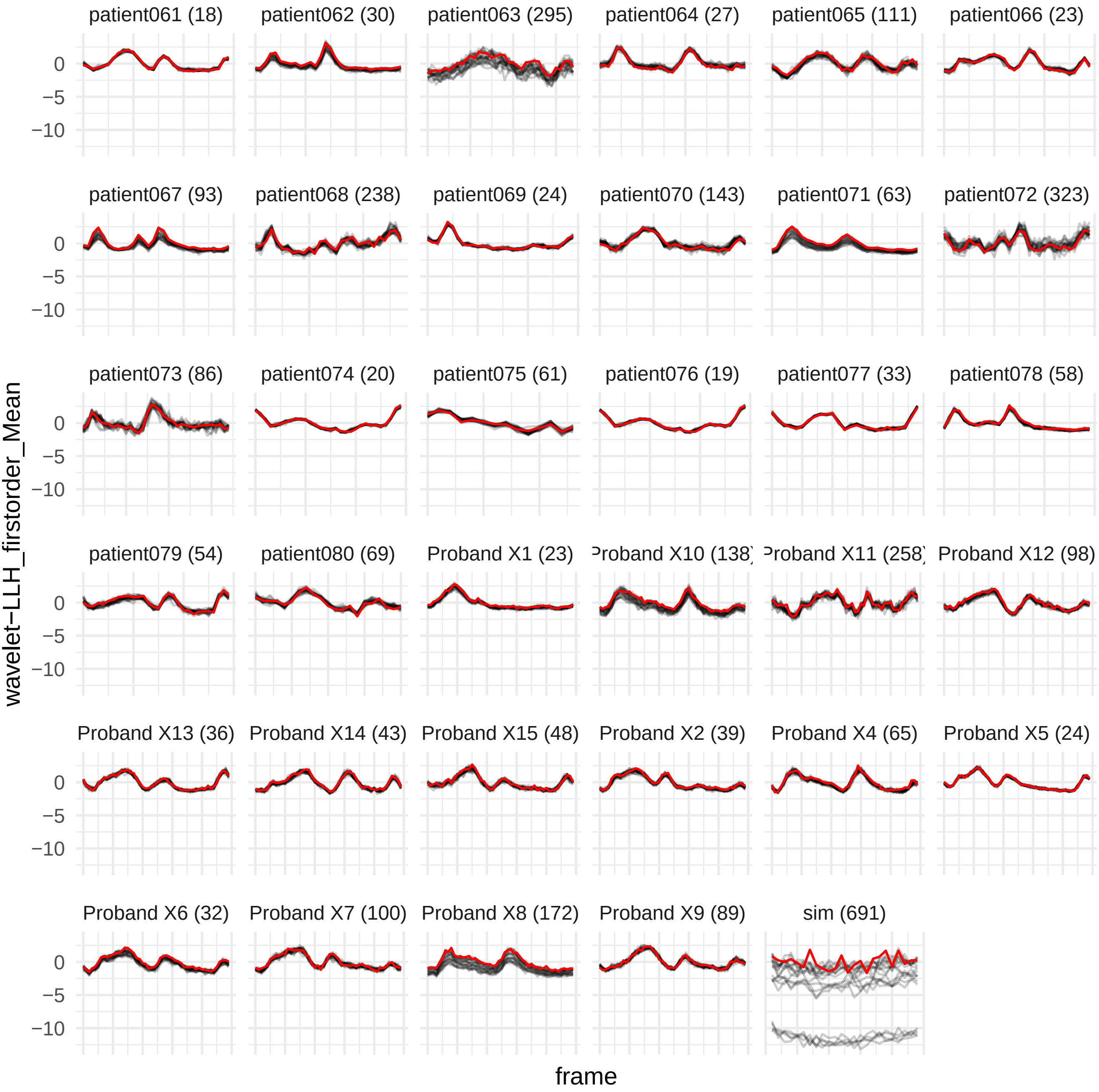
patient061 (18)
patient062 (30)
patient063 (295)
patient064 (27)
patient065 (111)
patient066 (23)
patient067 (93)
patient068 (238)
patient069 (24)
patient070 (143)
patient071 (63)
patient072 (323)
patient073 (86)
patient074 (20)
patient075 (61)
patient076 (19)
patient077 (33)
patient078 (58)
patient079 (54)
patient080 (69)
Proband X1 (23)
Proband X10 (138)
Proband X11 (258)
Proband X12 (98)
Proband X13 (36)
Proband X14 (43)
Proband X15 (48)
Proband X2 (39)
Proband X4 (65)
Proband X5 (24)
Proband X6 (32)
Proband X7 (100)
Proband X8 (172)
Proband X9 (89)
sim (691)
0
−5
−10
wavelet−LLH_firstorder_Mean
frame

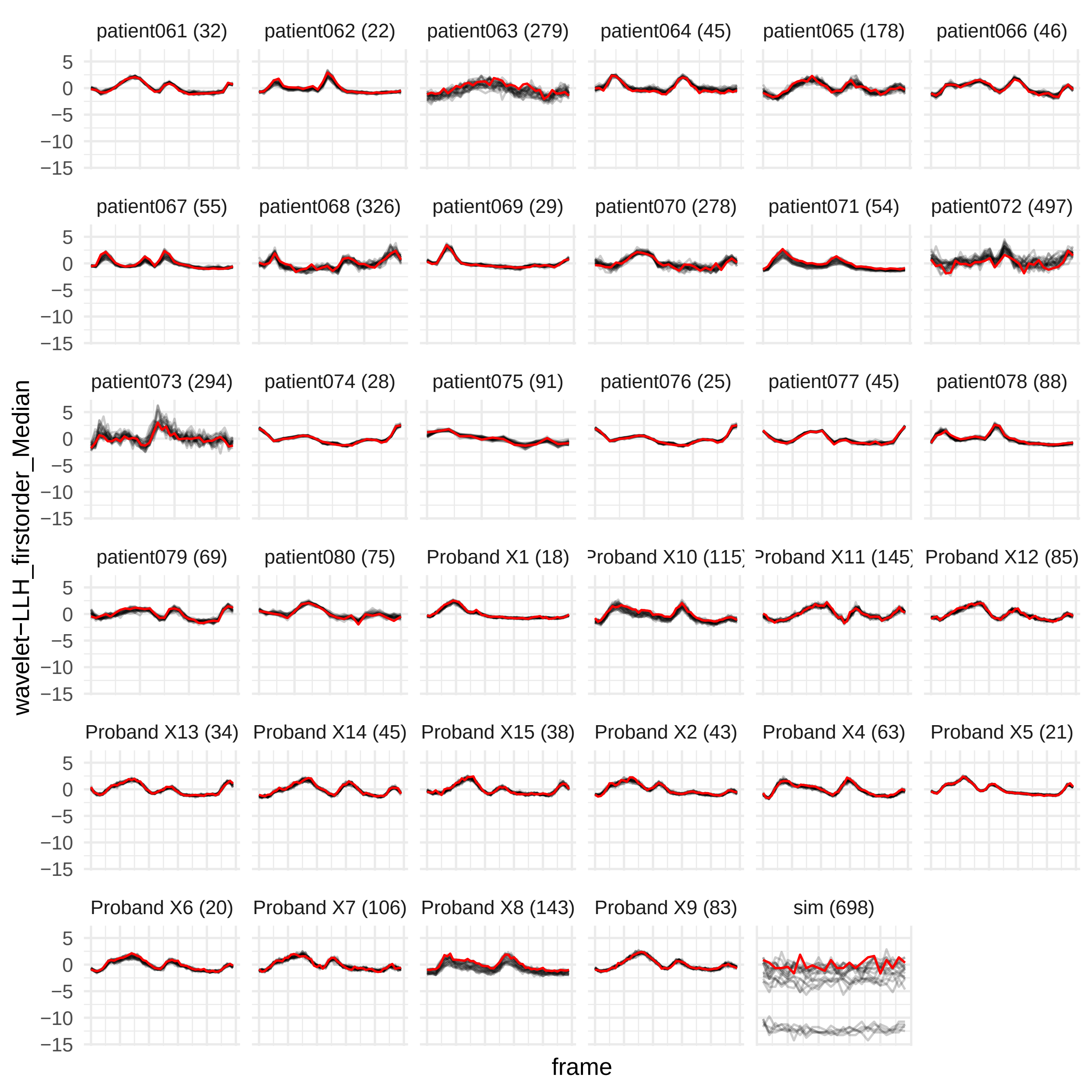
patient061 (32)
patient062 (22)
patient063 (279)
patient064 (45)
patient065 (178)
patient066 (46)
patient067 (55)
patient068 (326)
patient069 (29)
patient070 (278)
patient071 (54)
patient072 (497)
patient073 (294)
patient074 (28)
patient075 (91)
patient076 (25)
patient077 (45)
patient078 (88)
patient079 (69)
patient080 (75)
Proband X1 (18)
Proband X10 (115)
Proband X11 (145)
Proband X12 (85)
Proband X13 (34)
Proband X14 (45)
Proband X15 (38)
Proband X2 (43)
Proband X4 (63)
Proband X5 (21)
Proband X6 (20)
Proband X7 (106)
Proband X8 (143)
Proband X9 (83)
sim (698)
5
0
−5
−10
−15
wavelet−LLH_firstorder_Median
frame

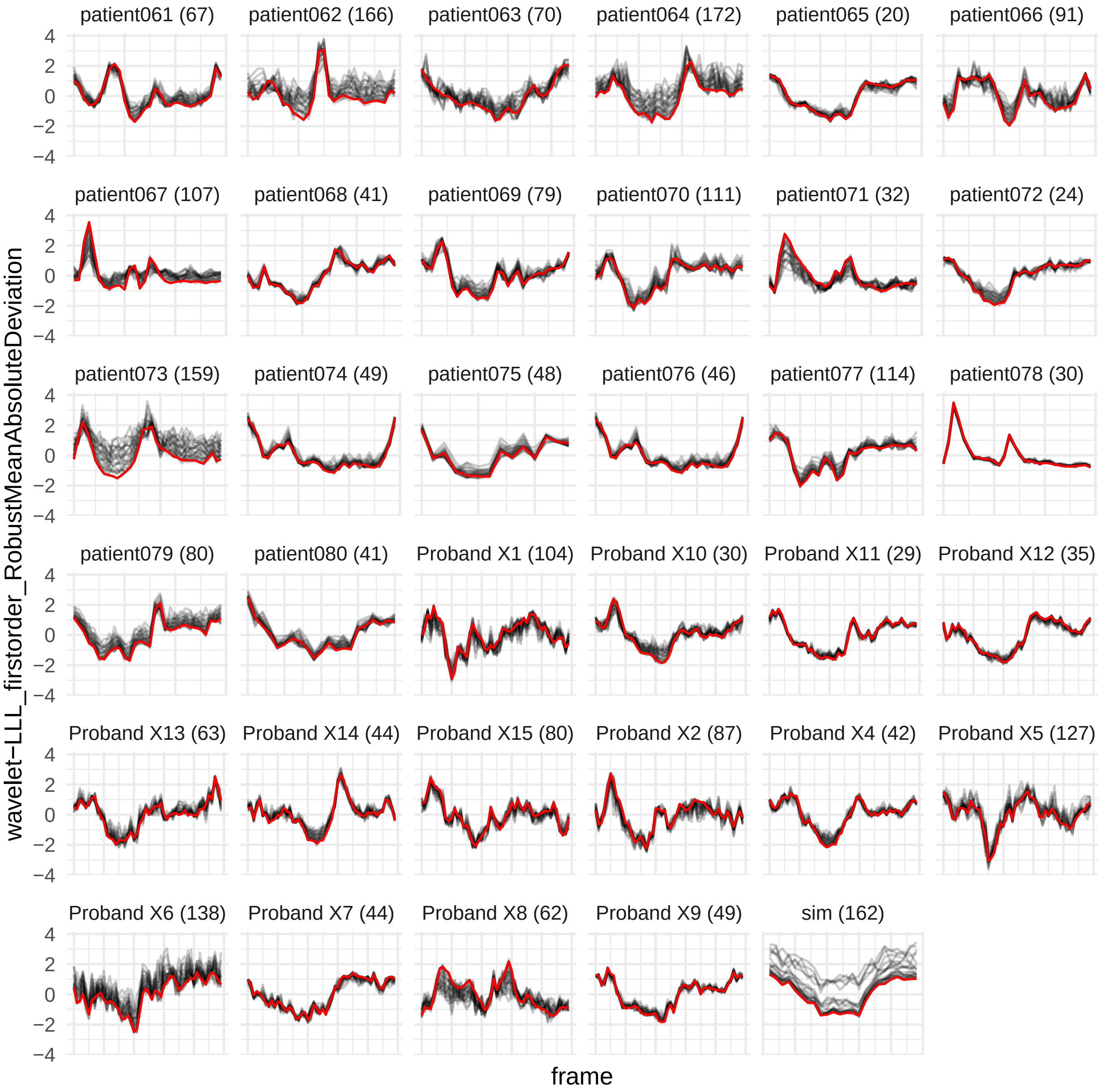
patient061 (67)
patient062 (166)
patient063 (70)
patient064 (172)
patient065 (20)
patient066 (91)
patient067 (107)
patient068 (41)
patient069 (79)
patient070 (111)
patient071 (32)
patient072 (24)
patient073 (159)
patient074 (49)
patient075 (48)
patient076 (46)
patient077 (114)
patient078 (30)
patient079 (80)
patient080 (41)
Proband X1 (104)
Proband X10 (30)
Proband X11 (29)
Proband X12 (35)
Proband X13 (63)
Proband X14 (44)
Proband X15 (80)
Proband X2 (87)
Proband X4 (42)
Proband X5 (127)
Proband X6 (138)
Proband X7 (44)
Proband X8 (62)
Proband X9 (49)
sim (162)
4
2
0
−2
−4
wavelet−LLL_firstorder_RobustMeanAbsoluteDeviation
frame

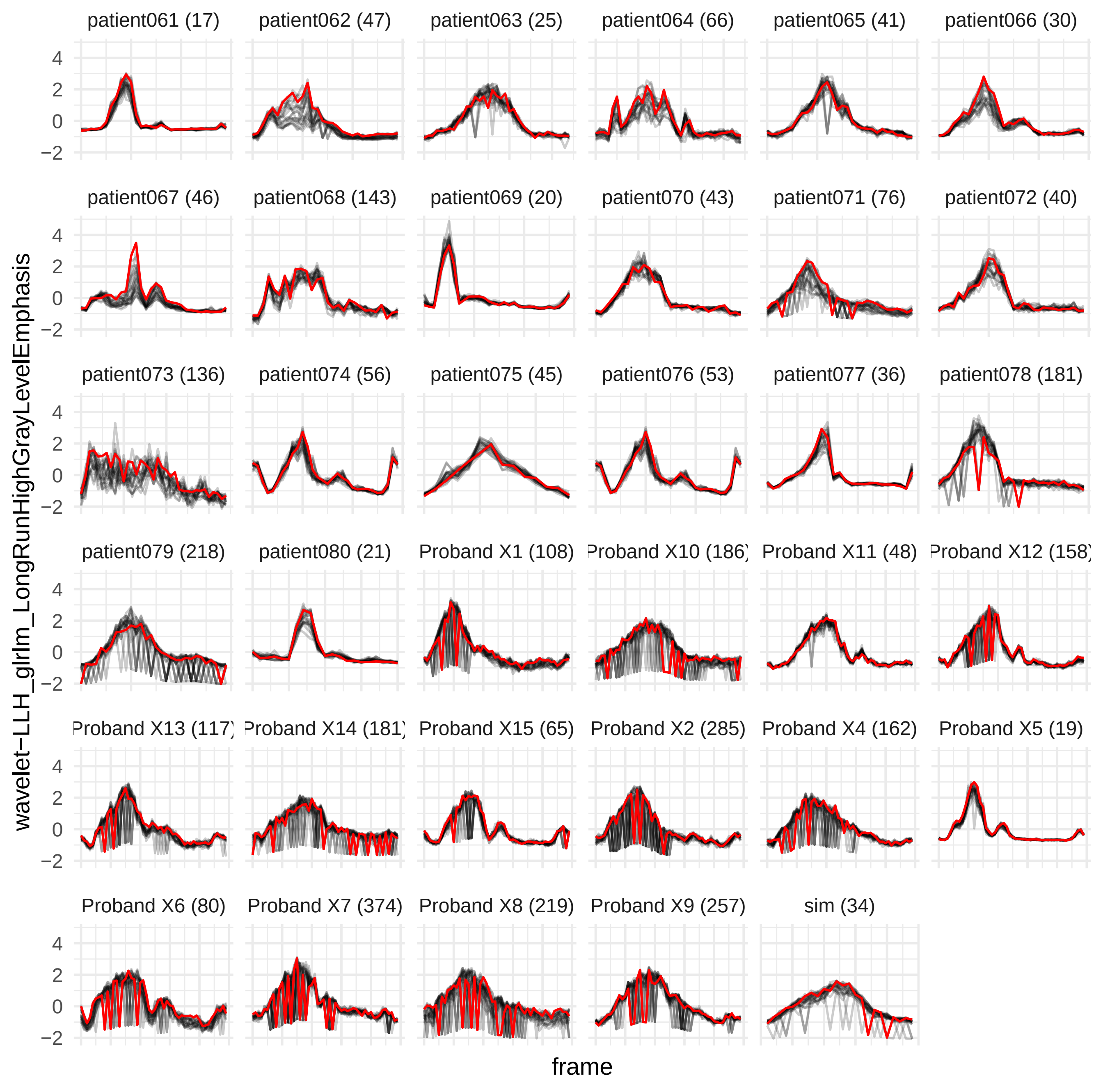
patient061 (17)
patient062 (47)
patient063 (25)
patient064 (66)
patient065 (41)
patient066 (30)
patient067 (46)
patient068 (143)
patient069 (20)
patient070 (43)
patient071 (76)
patient072 (40)
patient073 (136)
patient074 (56)
patient075 (45)
patient076 (53)
patient077 (36)
patient078 (181)
patient079 (218)
patient080 (21)
Proband X1 (108)
Proband X10 (186)
Proband X11 (48)
Proband X12 (158)
Proband X13 (117)
Proband X14 (181)
Proband X15 (65)
Proband X2 (285)
Proband X4 (162)
Proband X5 (19)
Proband X6 (80)
Proband X7 (374)
Proband X8 (219)
Proband X9 (257)
sim (34)
wavelet−LLH_glrlm_LongRunHighGrayLevelEmphasis
frame
4
2
0
−2

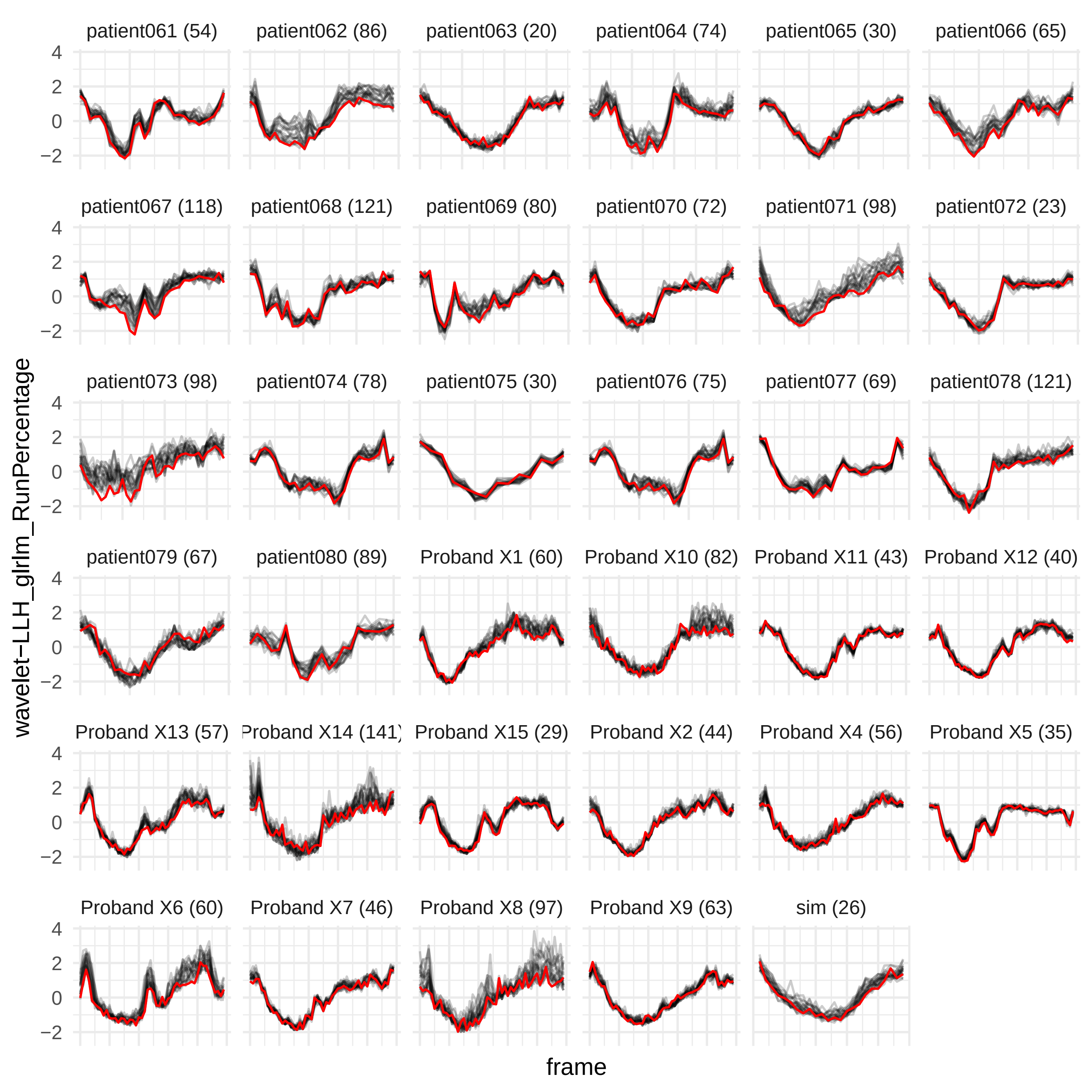
patient061 (54)
patient062 (86)
patient063 (20)
patient064 (74)
patient065 (30)
patient066 (65)
patient067 (118)
patient068 (121)
patient069 (80)
patient070 (72)
patient071 (98)
patient072 (23)
patient073 (98)
patient074 (78)
patient075 (30)
patient076 (75)
patient077 (69)
patient078 (121)
patient079 (67)
patient080 (89)
Proband X1 (60)
Proband X10 (82)
Proband X11 (43)
Proband X12 (40)
Proband X13 (57)
Proband X14 (141)
Proband X15 (29)
Proband X2 (44)
Proband X4 (56)
Proband X5 (35)
Proband X6 (60)
Proband X7 (46)
Proband X8 (97)
Proband X9 (63)
sim (26)
wavelet−LLH_glrlm_RunPercentage
frame
4
2
0
−2

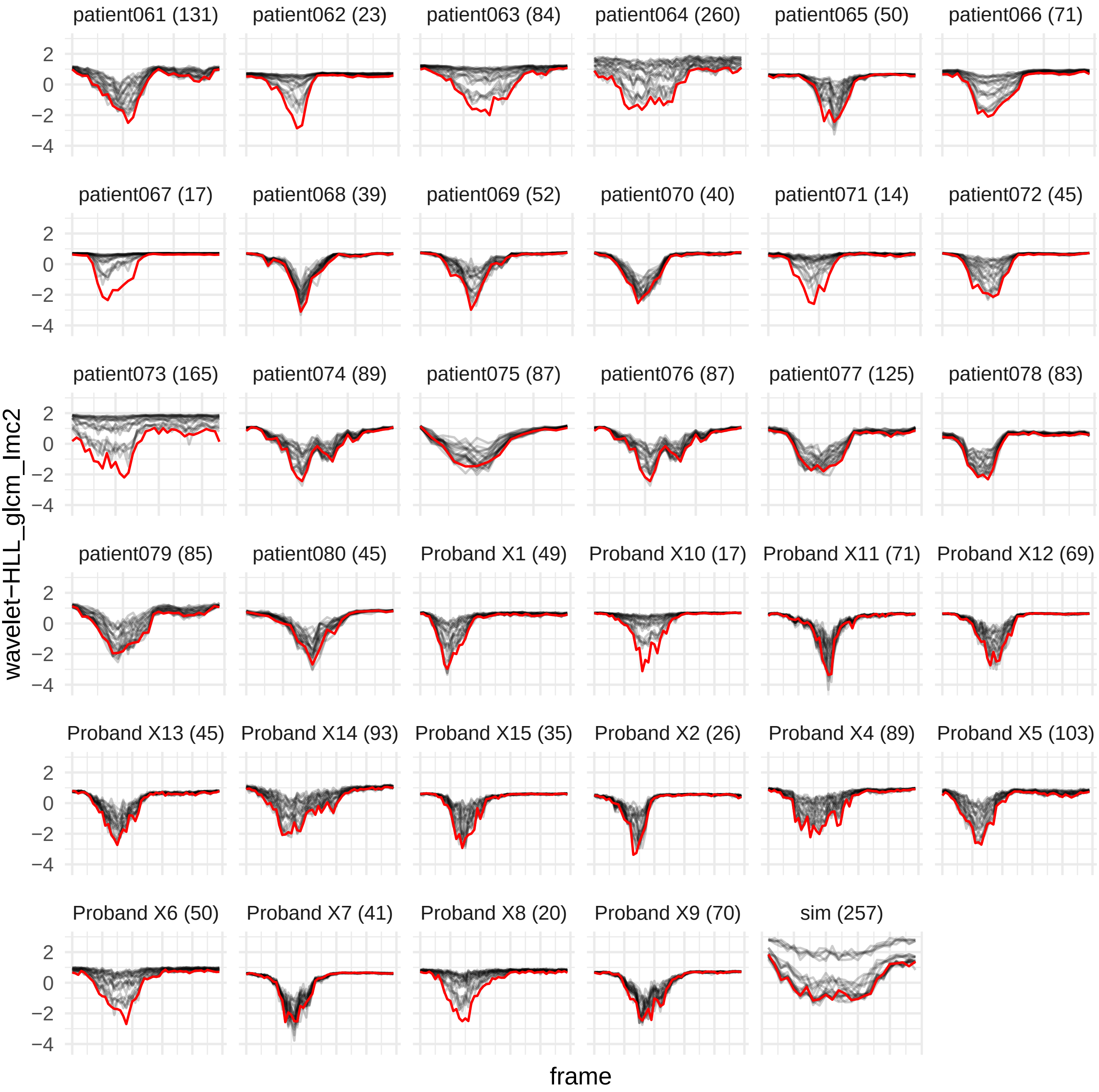

patient061 (131)
patient062 (23)
patient063 (84)
patient064 (260)
patient065 (50)
patient066 (71)
patient067 (17)
patient068 (39)
patient069 (52)
patient070 (40)
patient071 (14)
patient072 (45)
patient073 (165)
patient074 (89)
patient075 (87)
patient076 (87)
patient077 (125)
patient078 (83)
patient079 (85)
patient080 (45)
Proband X1 (49)
Proband X10 (17)
Proband X11 (71)
Proband X12 (69)
Proband X13 (45)
Proband X14 (93)
Proband X15 (35)
Proband X2 (26)
Proband X4 (89)
Proband X5 (103)
Proband X6 (50)
Proband X7 (41)
Proband X8 (20)
Proband X9 (70)
sim (257)
2
0
−2
−4
wavelet−HLL_glcm_Imc2
frame

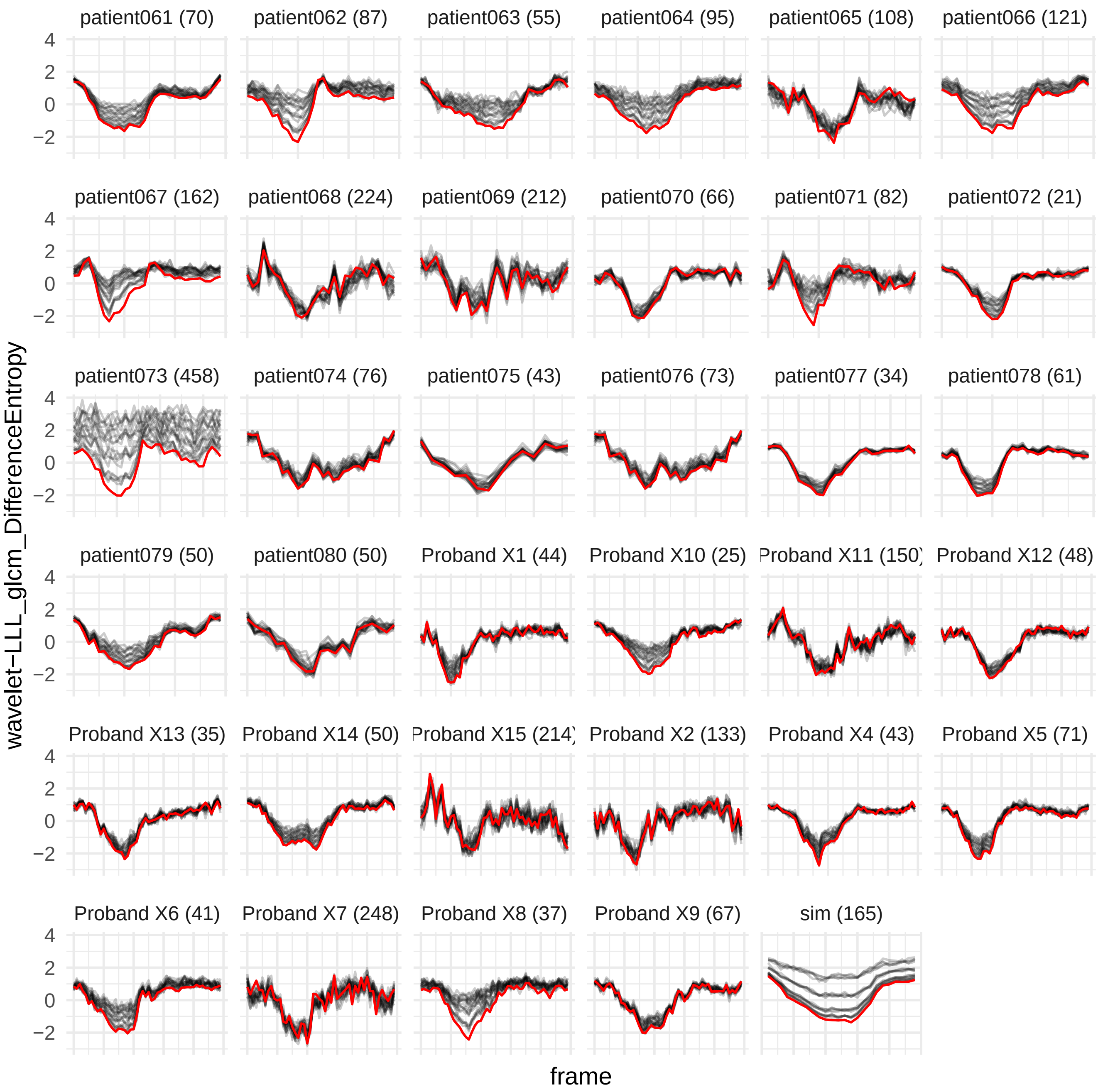
patient061 (70)
patient062 (87)
patient063 (55)
patient064 (95)
patient065 (108)
patient066 (121)
patient067 (162)
patient068 (224)
patient069 (212)
patient070 (66)
patient071 (82)
patient072 (21)
patient073 (458)
patient074 (76)
patient075 (43)
patient076 (73)
patient077 (34)
patient078 (61)
patient079 (50)
patient080 (50)
Proband X1 (44)
Proband X10 (25)
Proband X11 (150)
Proband X12 (48)
Proband X13 (35)
Proband X14 (50)
Proband X15 (214)
Proband X2 (133)
Proband X4 (43)
Proband X5 (71)
Proband X6 (41)
Proband X7 (248)
Proband X8 (37)
Proband X9 (67)
sim (165)
wavelet−LLL_glcm_DifferenceEntropy
frame
4
2
0
−2

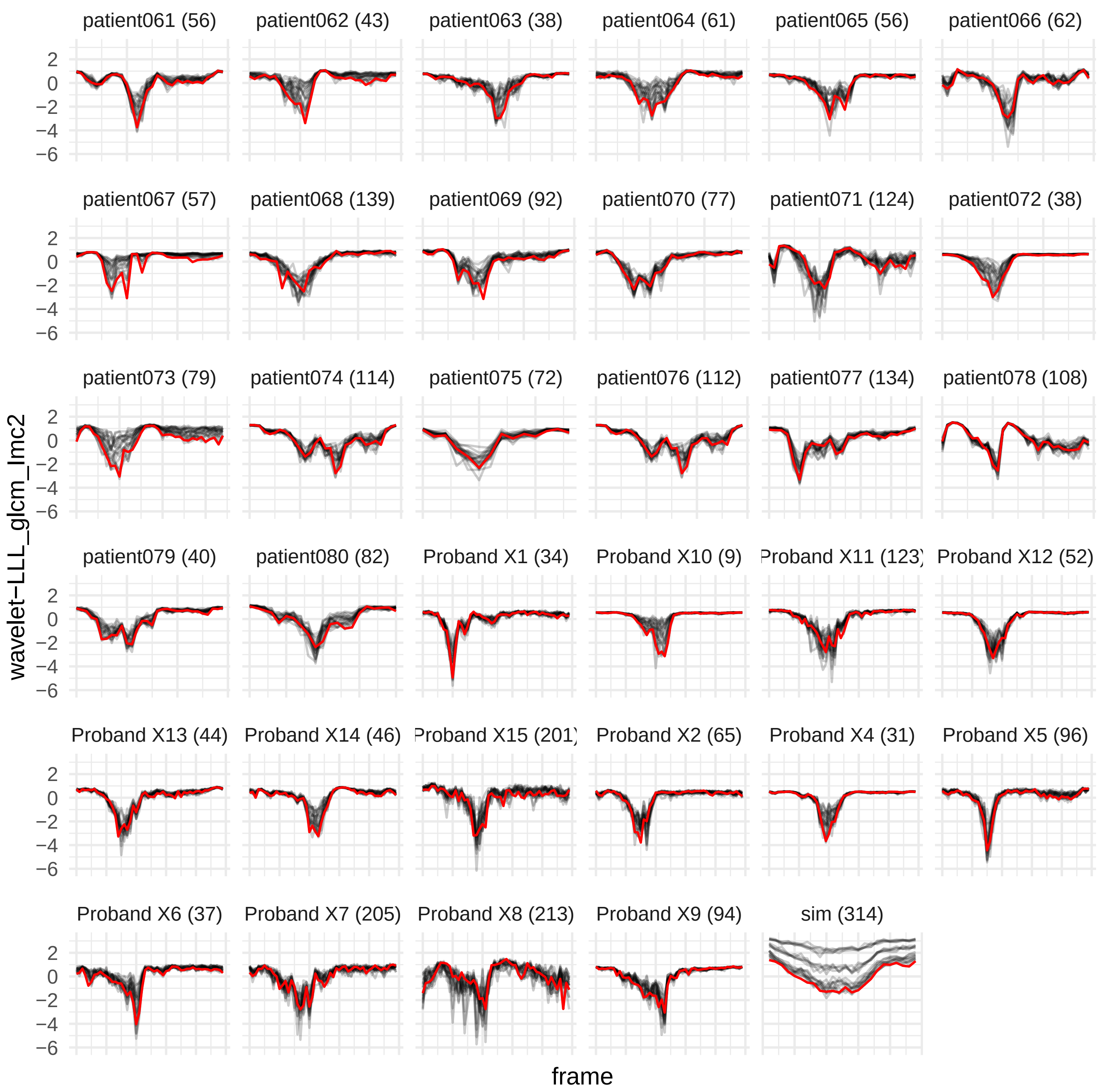

patient061 (56)
patient062 (43)
patient063 (38)
patient064 (61)
patient065 (56)
patient066 (62)
patient067 (57)
patient068 (139)
patient069 (92)
patient070 (77)
patient071 (124)
patient072 (38)
patient073 (79)
patient074 (114)
patient075 (72)
patient076 (112)
patient077 (134)
patient078 (108)
patient079 (40)
patient080 (82)
Proband X1 (34)
Proband X10 (9)
Proband X11 (123)
Proband X12 (52)
Proband X13 (44)
Proband X14 (46)
Proband X15 (201)
Proband X2 (65)
Proband X4 (31)
Proband X5 (96)
Proband X6 (37)
Proband X7 (205)
Proband X8 (213)
Proband X9 (94)
sim (314)
2
0
−2
−4
−6
wavelet−LLL_glcm_Imc2
frame

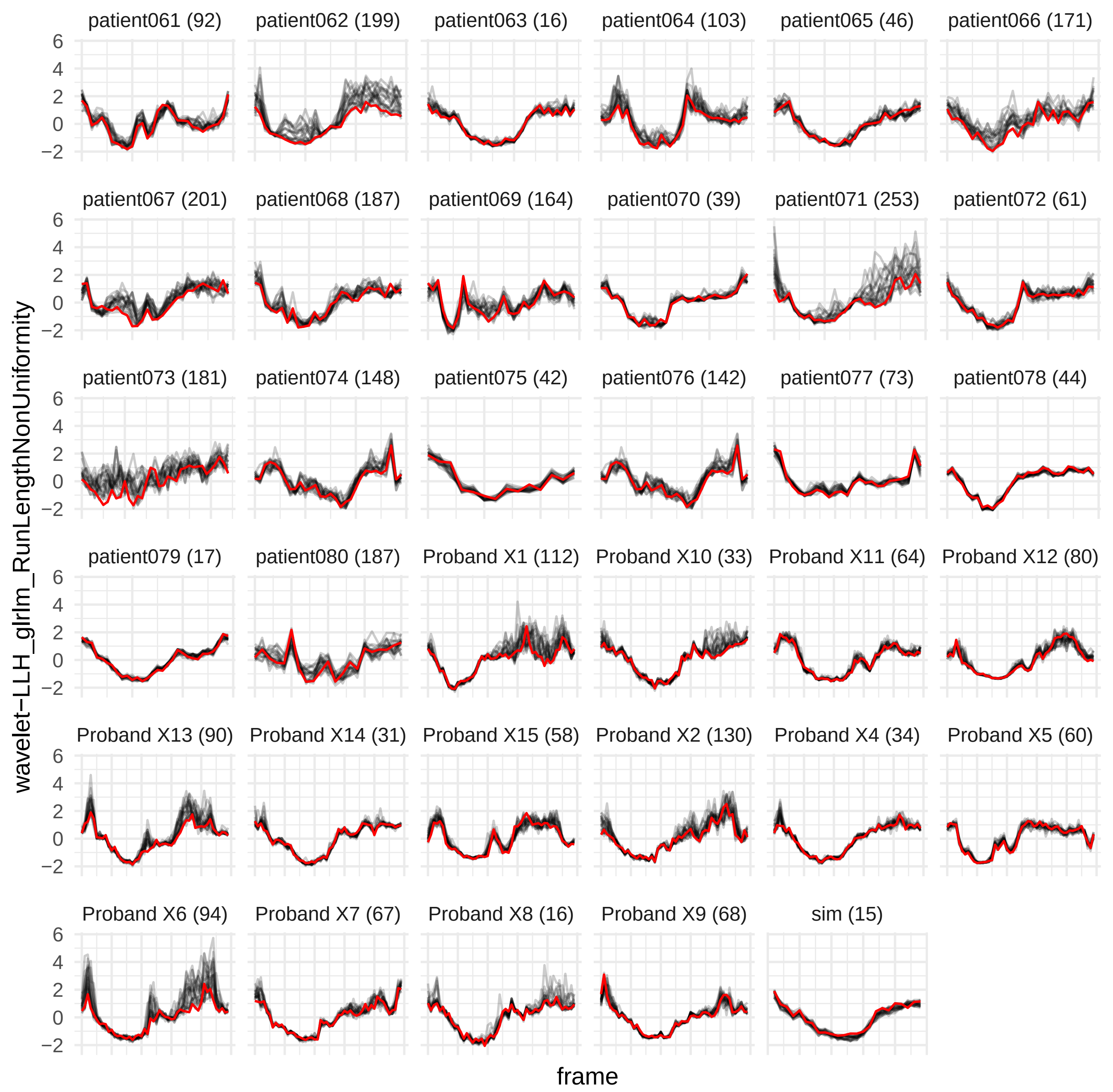

patient061 (92)
patient062 (199)
patient063 (16)
patient064 (103)
patient065 (46)
patient066 (171)
patient067 (201)
patient068 (187)
patient069 (164)
patient070 (39)
patient071 (253)
patient072 (61)
patient073 (181)
patient074 (148)
patient075 (42)
patient076 (142)
patient077 (73)
patient078 (44)
patient079 (17)
patient080 (187)
Proband X1 (112)
Proband X10 (33)
Proband X11 (64)
Proband X12 (80)
Proband X13 (90)
Proband X14 (31)
Proband X15 (58)
Proband X2 (130)
Proband X4 (34)
Proband X5 (60)
Proband X6 (94)
Proband X7 (67)
Proband X8 (16)
Proband X9 (68)
sim (15)
wavelet−LLH_glrlm_RunLengthNonUniformity
frame
6
4
2
0
−2

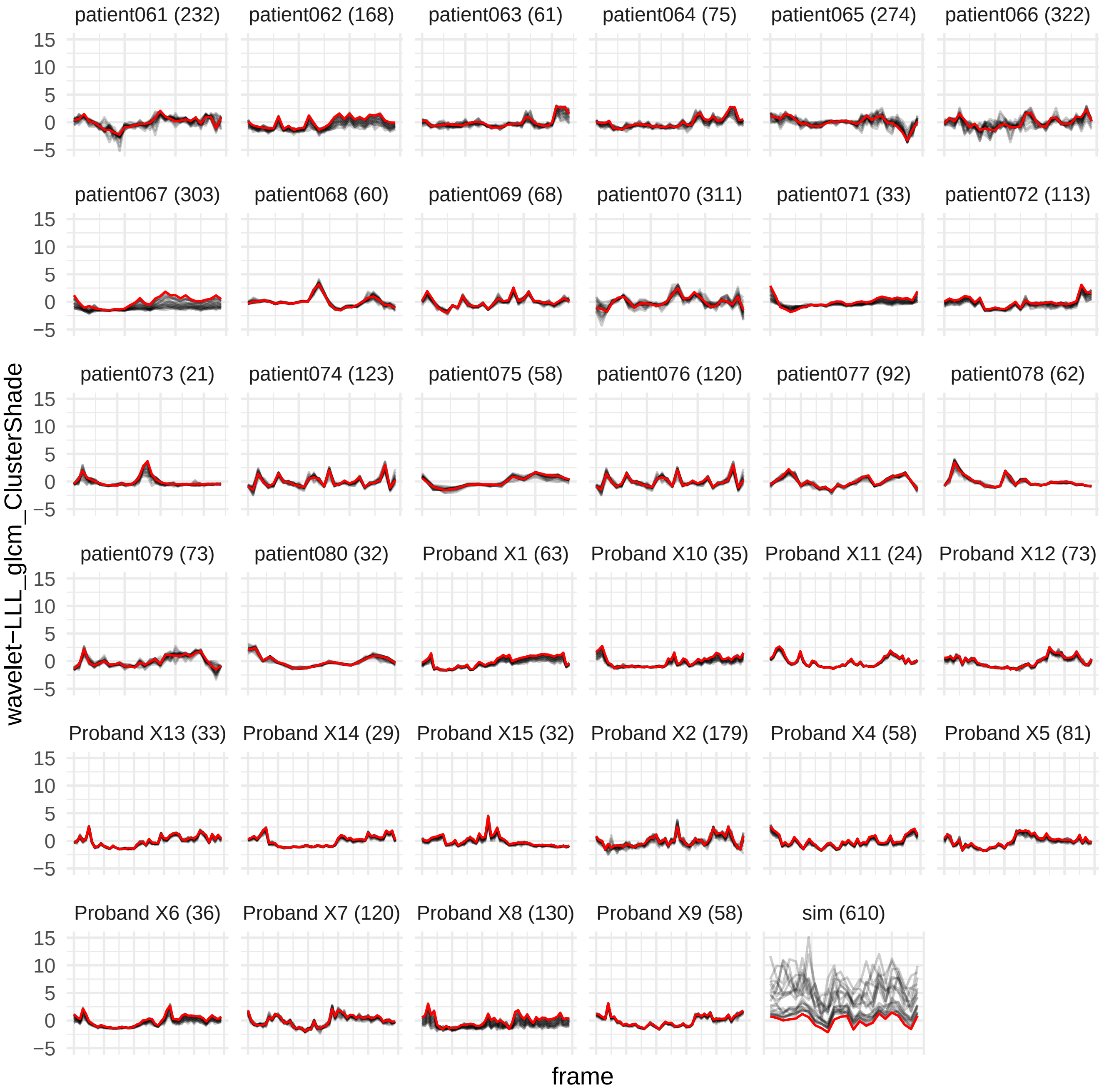

patient061 (232)
patient062 (168)
patient063 (61)
patient064 (75)
patient065 (274)
patient066 (322)
patient067 (303)
patient068 (60)
patient069 (68)
patient070 (311)
patient071 (33)
patient072 (113)
patient073 (21)
patient074 (123)
patient075 (58)
patient076 (120)
patient077 (92)
patient078 (62)
patient079 (73)
patient080 (32)
Proband X1 (63)
Proband X10 (35)
Proband X11 (24)
Proband X12 (73)
Proband X13 (33)
Proband X14 (29)
Proband X15 (32)
Proband X2 (179)
Proband X4 (58)
Proband X5 (81)
Proband X6 (36)
Proband X7 (120)
Proband X8 (130)
Proband X9 (58)
sim (610)
15
10
5
0
−5
wavelet−LLL_glcm_ClusterShade
frame

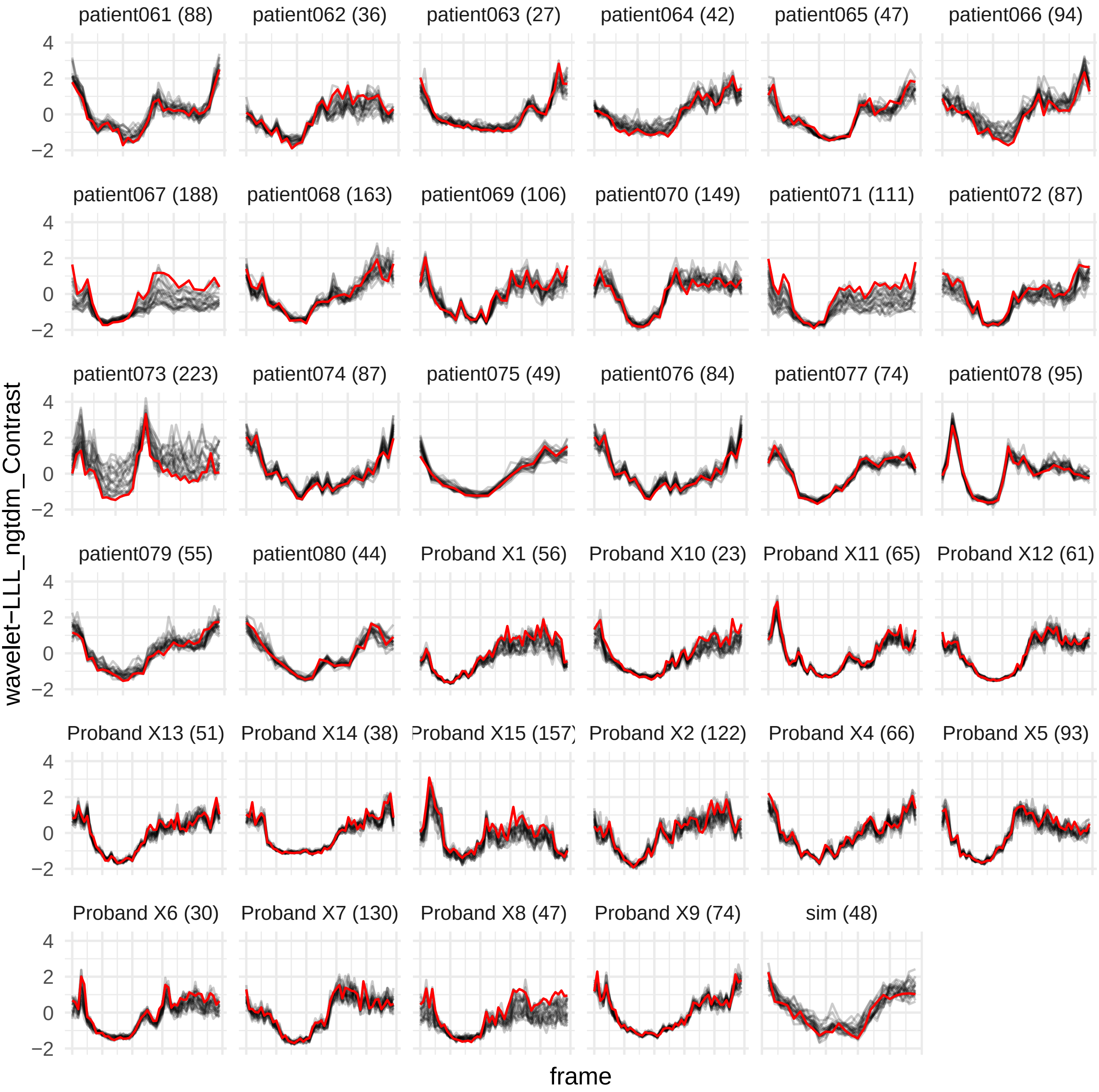

patient061 (88)
patient062 (36)
patient063 (27)
patient064 (42)
patient065 (47)
patient066 (94)
patient067 (188)
patient068 (163)
patient069 (106)
patient070 (149)
patient071 (111)
patient072 (87)
patient073 (223)
patient074 (87)
patient075 (49)
patient076 (84)
patient077 (74)
patient078 (95)
patient079 (55)
patient080 (44)
Proband X1 (56)
Proband X10 (23)
Proband X11 (65)
Proband X12 (61)
Proband X13 (51)
Proband X14 (38)
Proband X15 (157)
Proband X2 (122)
Proband X4 (66)
Proband X5 (93)
Proband X6 (30)
Proband X7 (130)
Proband X8 (47)
Proband X9 (74)
sim (48)
4
2
0
−2
wavelet−LLL_ngtdm_Contrast
frame

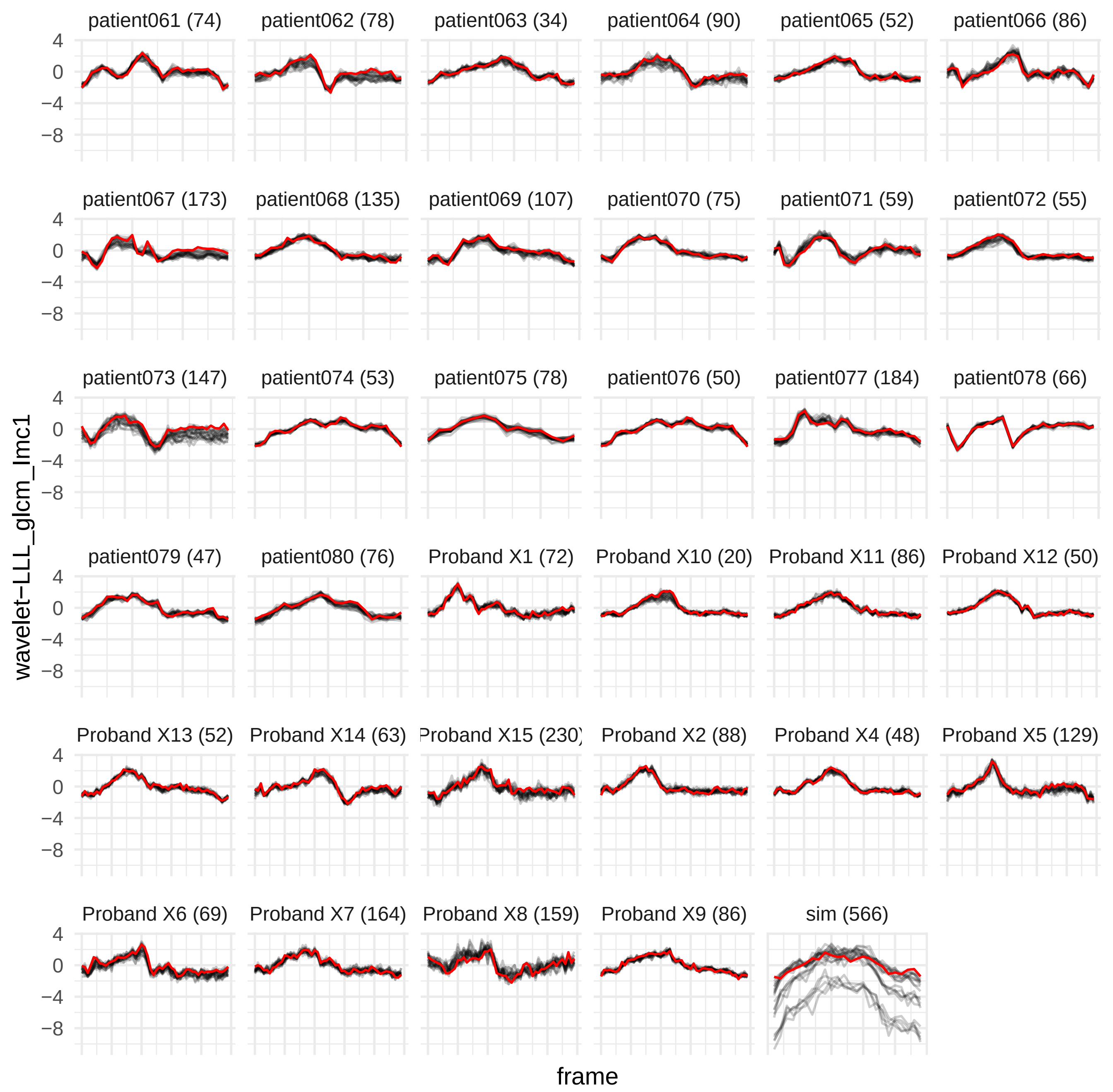
patient061 (74)
patient062 (78)
patient063 (34)
patient064 (90)
patient065 (52)
patient066 (86)
patient067 (173)
patient068 (135)
patient069 (107)
patient070 (75)
patient071 (59)
patient072 (55)
patient073 (147)
patient074 (53)
patient075 (78)
patient076 (50)
patient077 (184)
patient078 (66)
patient079 (47)
patient080 (76)
Proband X1 (72)
Proband X10 (20)
Proband X11 (86)
Proband X12 (50)
Proband X13 (52)
Proband X14 (63)
Proband X15 (230)
Proband X2 (88)
Proband X4 (48)
Proband X5 (129)
Proband X6 (69)
Proband X7 (164)
Proband X8 (159)
Proband X9 (86)
sim (566)
4
0
−4
−8
wavelet−LLL_glcm_Imc1
frame

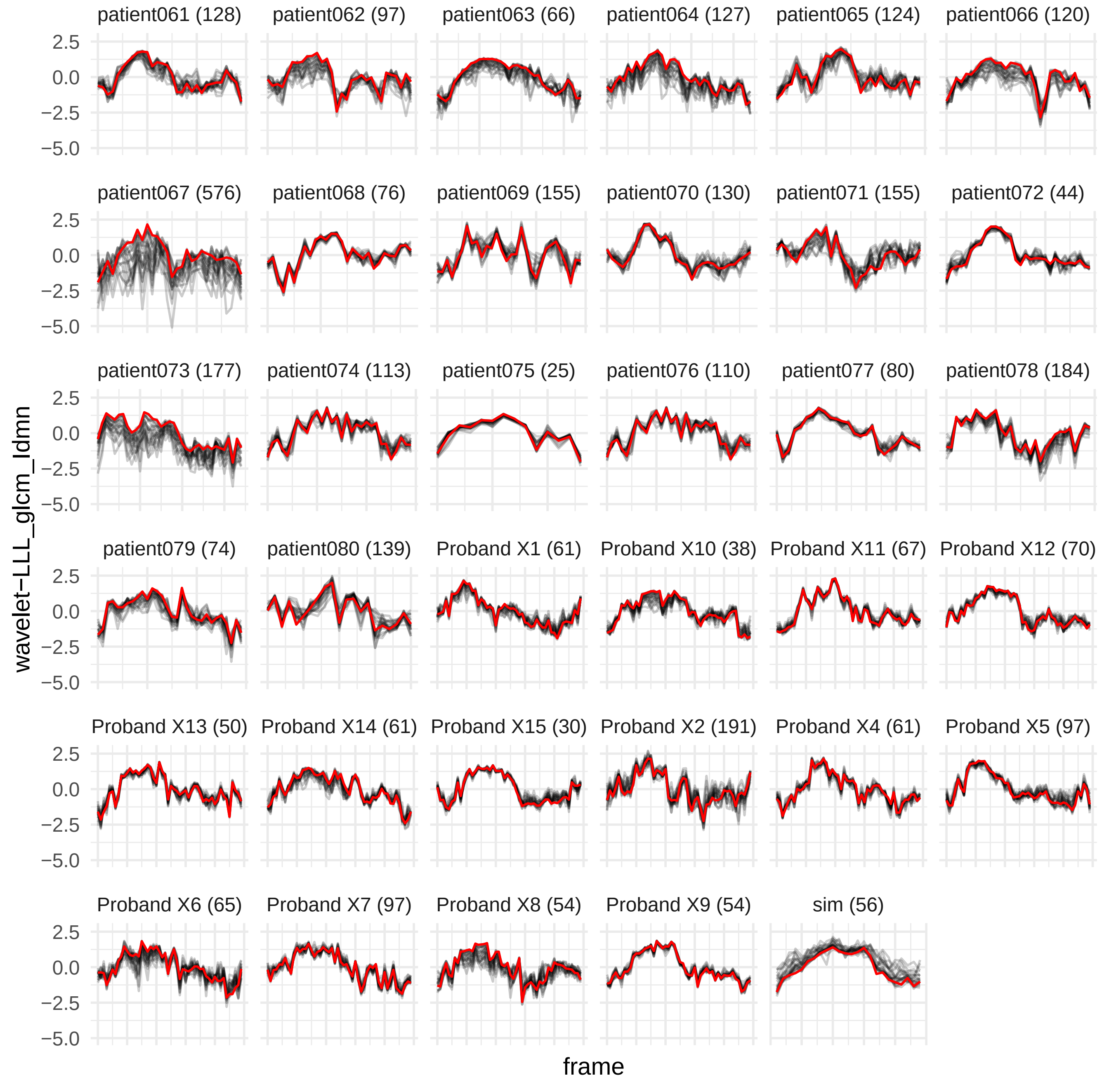

patient061 (128)
patient062 (97)
patient063 (66)
patient064 (127)
patient065 (124)
patient066 (120)
patient067 (576)
patient068 (76)
patient069 (155)
patient070 (130)
patient071 (155)
patient072 (44)
patient073 (177)
patient074 (113)
patient075 (25)
patient076 (110)
patient077 (80)
patient078 (184)
patient079 (74)
patient080 (139)
Proband X1 (61)
Proband X10 (38)
Proband X11 (67)
Proband X12 (70)
Proband X13 (50)
Proband X14 (61)
Proband X15 (30)
Proband X2 (191)
Proband X4 (61)
Proband X5 (97)
Proband X6 (65)
Proband X7 (97)
Proband X8 (54)
Proband X9 (54)
sim (56)
2.5
0.0
−2.5
−5.0
wavelet−LLL_glcm_Idmn
frame

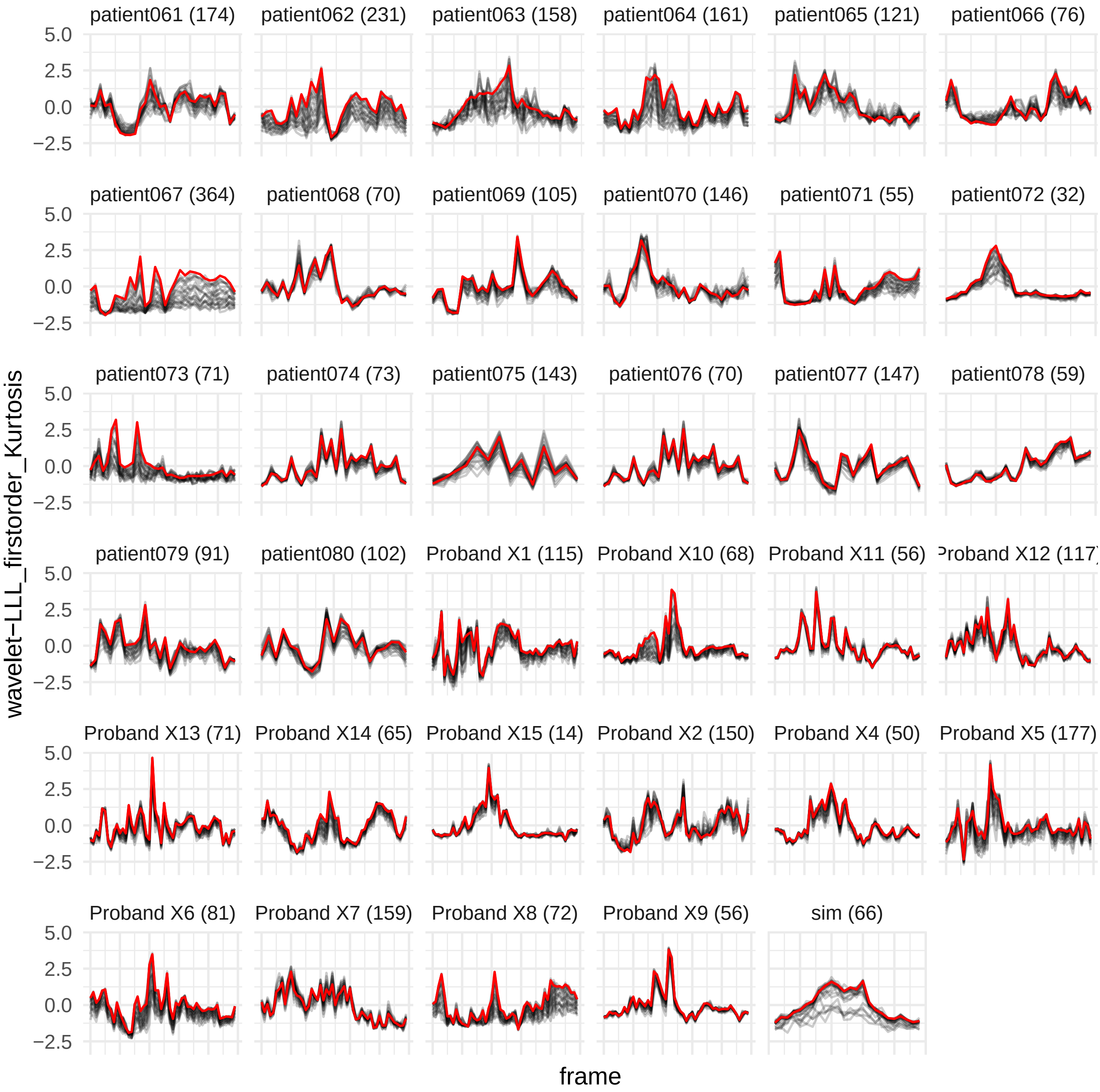

wavelet−LLL_firstorder_Kurtosis
frame
5.0
2.5
0.0
−2.5
patient061 (174)
patient062 (231)
patient063 (158)
patient064 (161)
patient065 (121)
patient066 (76)
patient067 (364)
patient068 (70)
patient069 (105)
patient070 (146)
patient071 (55)
patient072 (32)
patient073 (71)
patient074 (73)
patient075 (143)
patient076 (70)
patient077 (147)
patient078 (59)
patient079 (91)
patient080 (102)
Proband X1 (115)
Proband X10 (68)
Proband X11 (56)
Proband X13 (71)
Proband X14 (65)
Proband X15 (14)
Proband X2 (150)
Proband X4 (50)
Proband X5 (177)
Proband X6 (81)
Proband X7 (159)
Proband X8 (72)
Proband X9 (56)
sim (66)

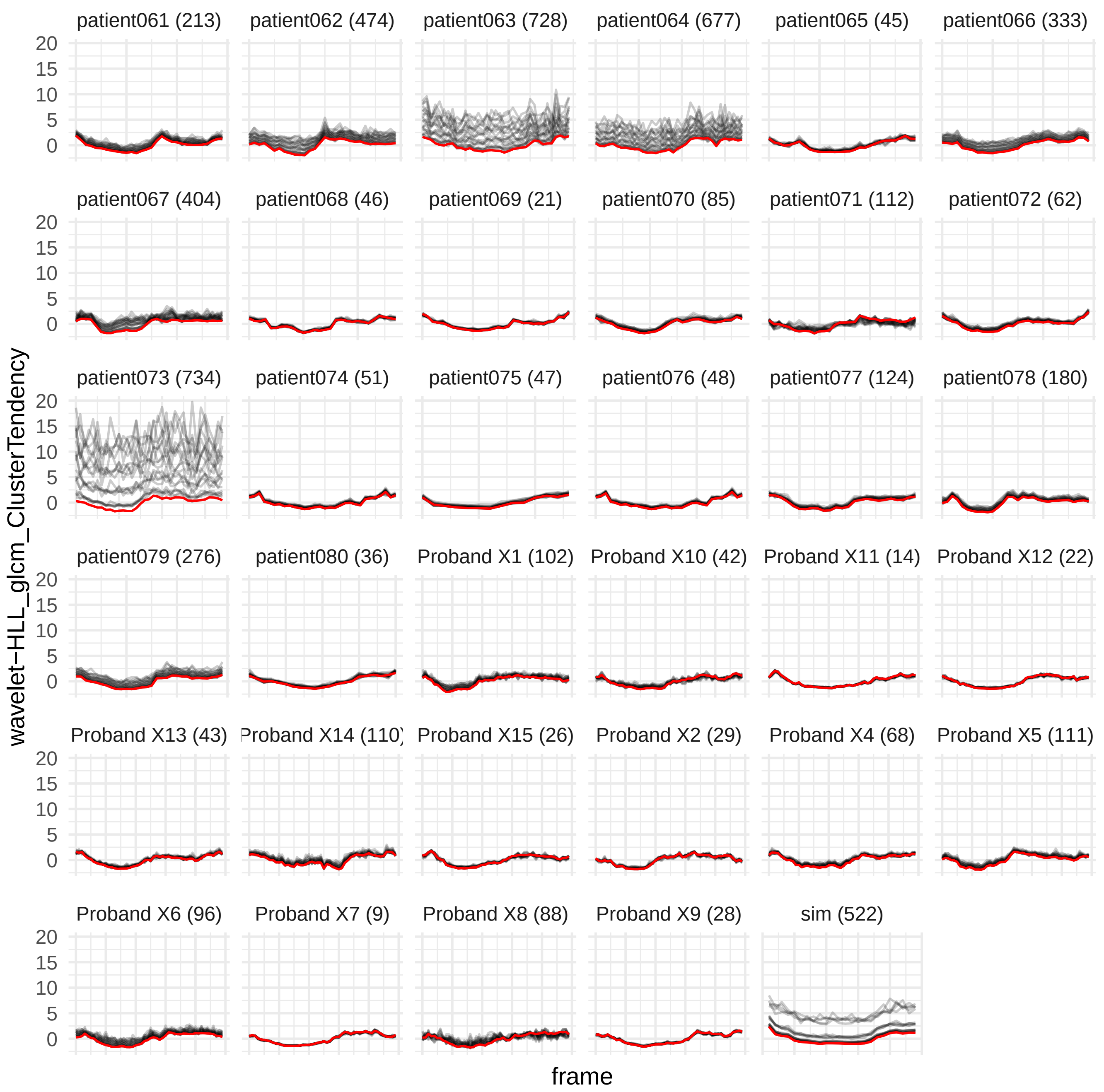
patient061 (213)
patient062 (474)
patient063 (728)
patient064 (677)
patient065 (45)
patient066 (333)
patient067 (404)
patient068 (46)
patient069 (21)
patient070 (85)
patient071 (112)
patient072 (62)
patient073 (734)
patient074 (51)
patient075 (47)
patient076 (48)
patient077 (124)
patient078 (180)
patient079 (276)
patient080 (36)
Proband X1 (102)
Proband X10 (42)
Proband X11 (14)
Proband X12 (22)
Proband X13 (43)
Proband X14 (110)
Proband X15 (26)
Proband X2 (29)
Proband X4 (68)
Proband X5 (111)
Proband X6 (96)
Proband X7 (9)
Proband X8 (88)
Proband X9 (28)
sim (522)
0
5
10
15
20
wavelet−HLL_glcm_ClusterTendency
frame

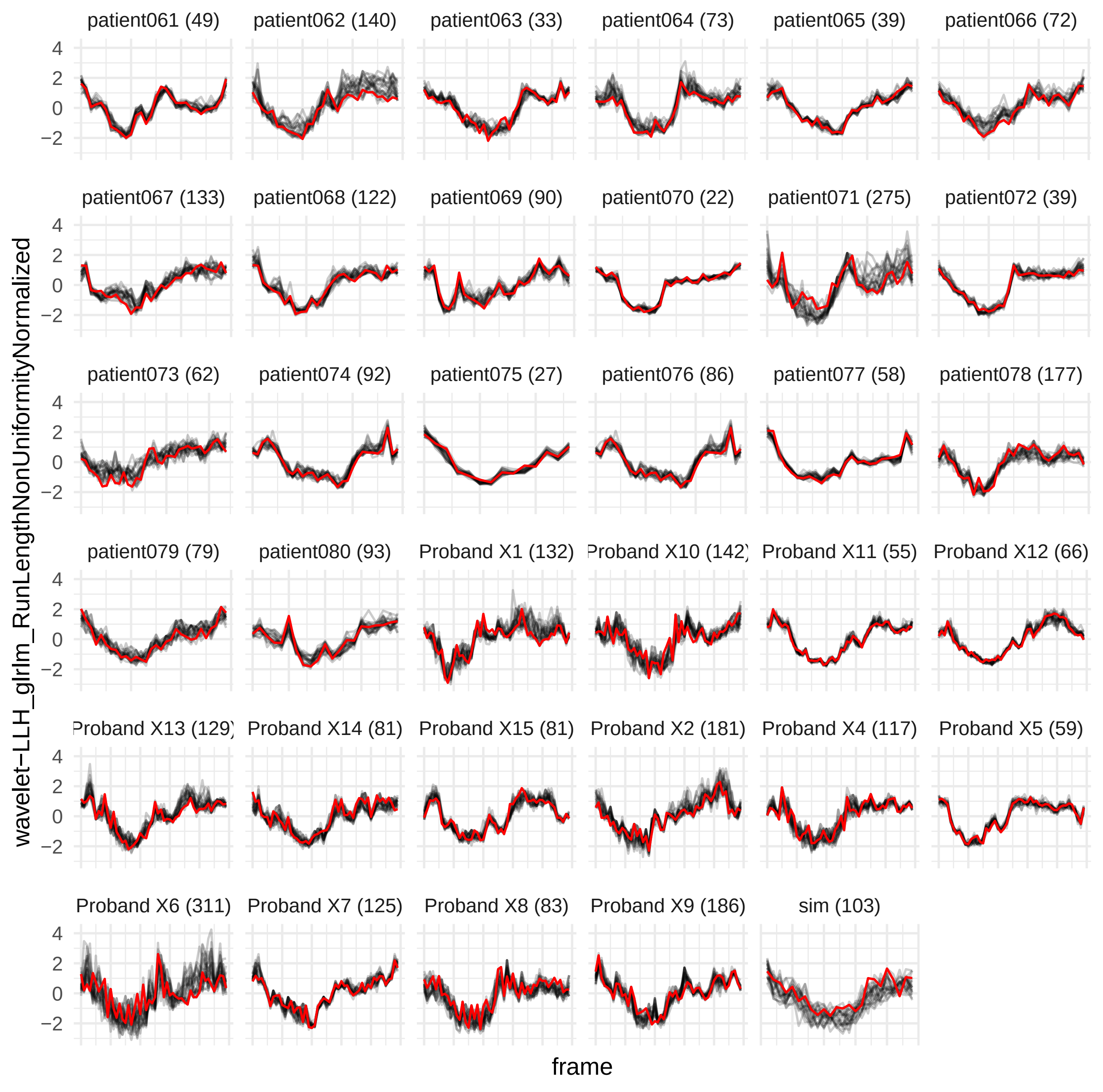
patient061 (49)
patient062 (140)
patient063 (33)
patient064 (73)
patient065 (39)
patient066 (72)
patient067 (133)
patient068 (122)
patient069 (90)
patient070 (22)
patient071 (275)
patient072 (39)
patient073 (62)
patient074 (92)
patient075 (27)
patient076 (86)
patient077 (58)
patient078 (177)
patient079 (79)
patient080 (93)
Proband X1 (132)
Proband X10 (142)
Proband X11 (55)
Proband X12 (66)
Proband X13 (129)
Proband X14 (81)
Proband X15 (81)
Proband X2 (181)
Proband X4 (117)
Proband X5 (59)
Proband X6 (311)
Proband X7 (125)
Proband X8 (83)
Proband X9 (186)
sim (103)
wavelet−LLH_glrlm_RunLengthNonUniformityNormalized
frame
4
2
0
−2

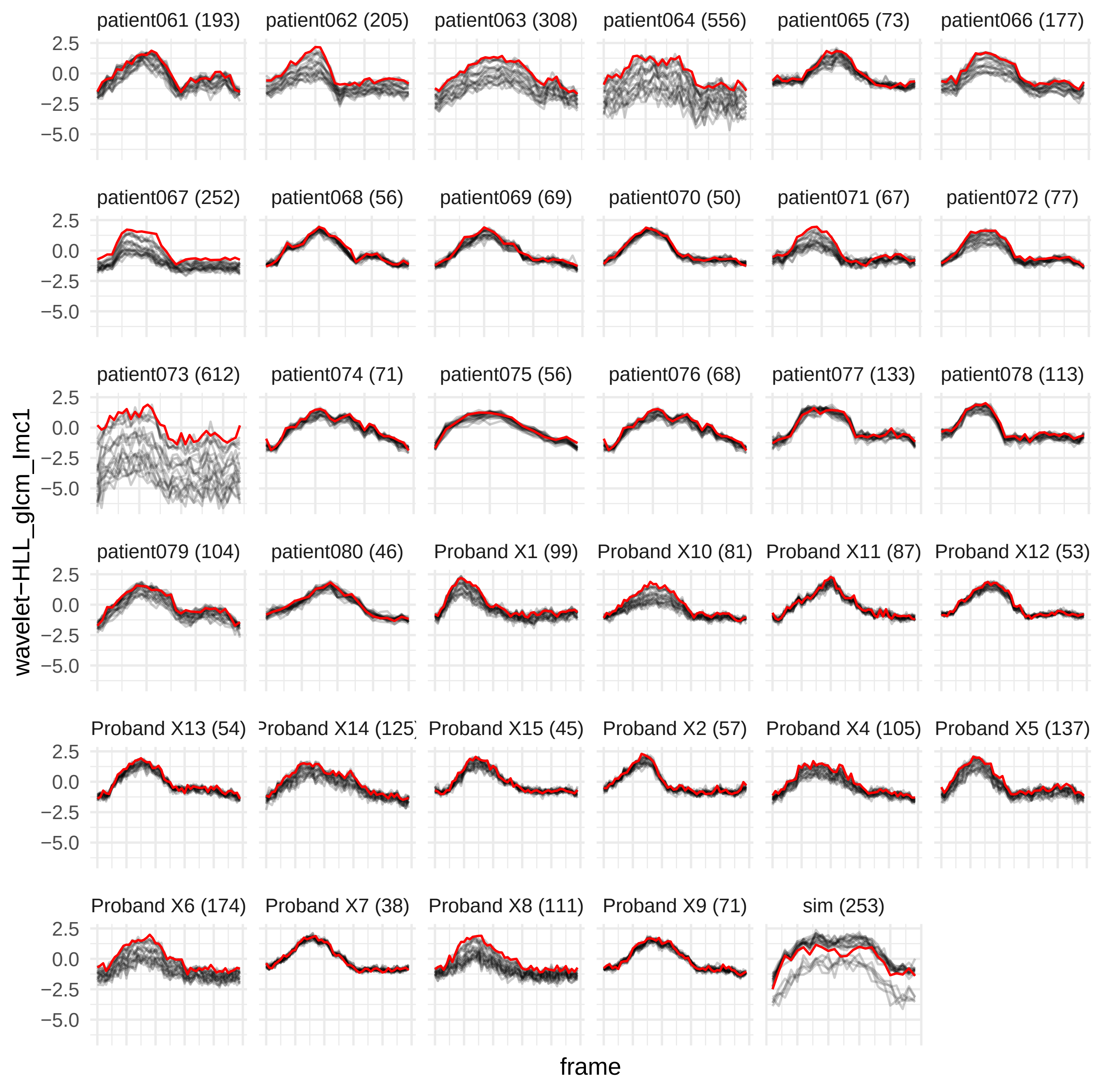
patient061 (193)
patient062 (205)
patient063 (308)
patient064 (556)
patient065 (73)
patient066 (177)
patient067 (252)
patient068 (56)
patient069 (69)
patient070 (50)
patient071 (67)
patient072 (77)
patient073 (612)
patient074 (71)
patient075 (56)
patient076 (68)
patient077 (133)
patient078 (113)
patient079 (104)
patient080 (46)
Proband X1 (99)
Proband X10 (81)
Proband X11 (87)
Proband X12 (53)
Proband X13 (54)
Proband X14 (125)
Proband X15 (45)
Proband X2 (57)
Proband X4 (105)
Proband X5 (137)
Proband X6 (174)
Proband X7 (38)
Proband X8 (111)
Proband X9 (71)
sim (253)
2.5
0.0
−2.5
−5.0
wavelet−HLL_glcm_Imc1
frame

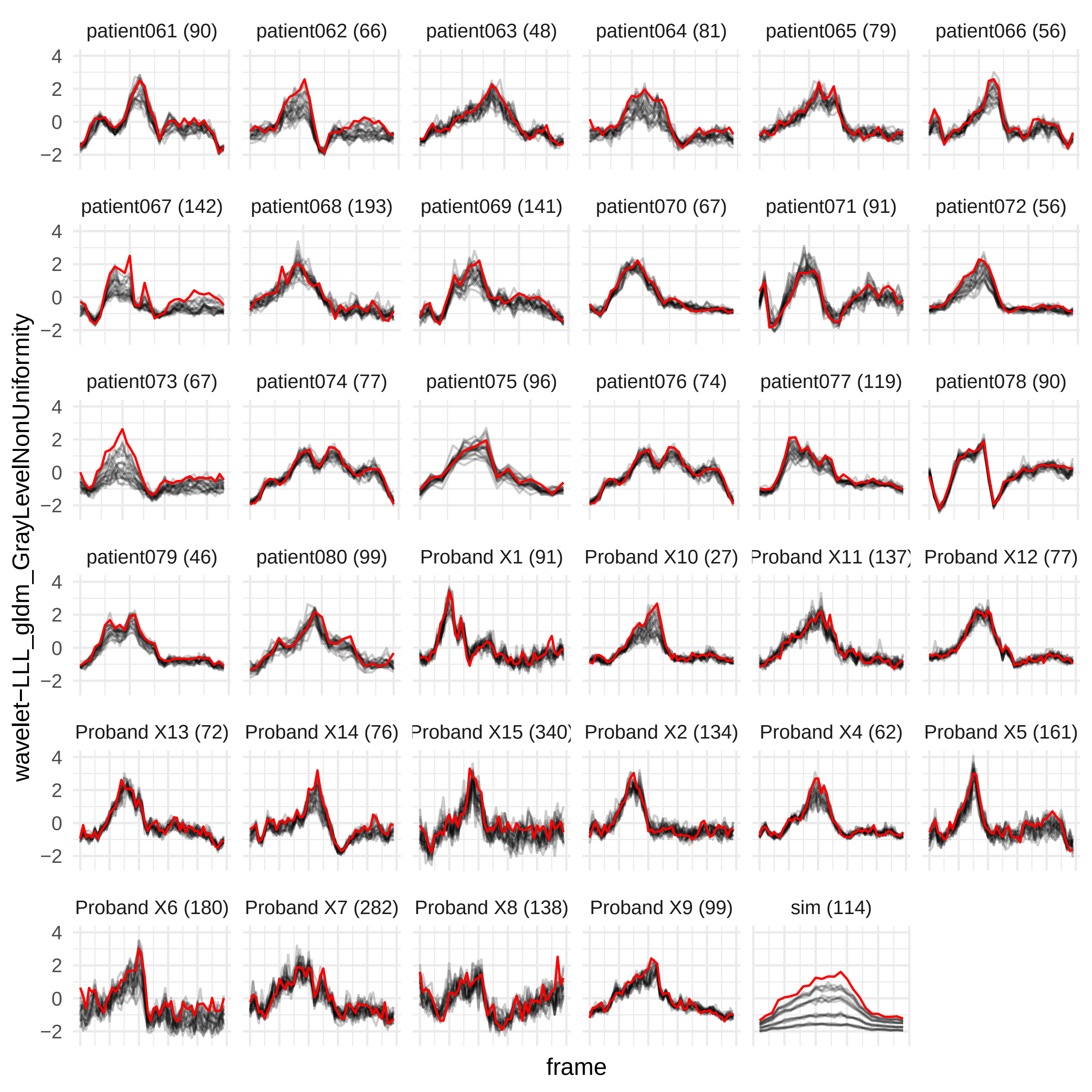

patient061 (90)
patient062 (66)
patient063 (48)
patient064 (81)
patient065 (79)
patient066 (56)
patient067 (142)
patient068 (193)
patient069 (141)
patient070 (67)
patient071 (91)
patient072 (56)
patient073 (67)
patient074 (77)
patient075 (96)
patient076 (74)
patient077 (119)
patient078 (90)
patient079 (46)
patient080 (99)
Proband X1 (91)
Proband X10 (27)
Proband X11 (137)
Proband X12 (77)
Proband X13 (72)
Proband X14 (76)
Proband X15 (340)
Proband X2 (134)
Proband X4 (62)
Proband X5 (161)
Proband X6 (180)
Proband X7 (282)
Proband X8 (138)
Proband X9 (99)
sim (114)
wavelet−LLL_gldm_GrayLevelNonUniformity
frame
4
2
0
−2

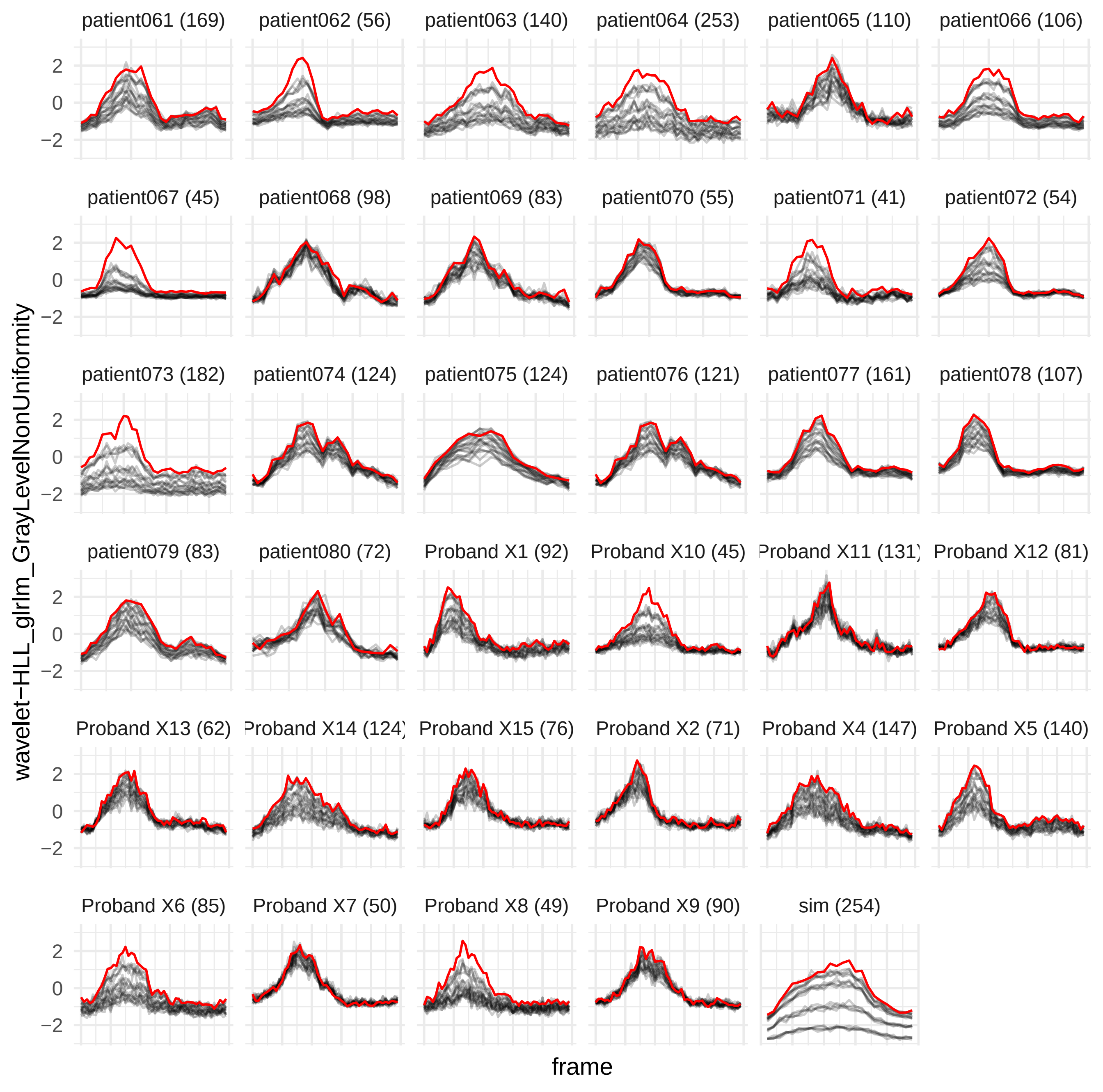
patient061 (169)
patient062 (56)
patient063 (140)
patient064 (253)
patient065 (110)
patient066 (106)
patient067 (45)
patient068 (98)
patient069 (83)
patient070 (55)
patient071 (41)
patient072 (54)
patient073 (182)
patient074 (124)
patient075 (124)
patient076 (121)
patient077 (161)
patient078 (107)
patient079 (83)
patient080 (72)
Proband X1 (92)
Proband X10 (45)
Proband X11 (131)
Proband X12 (81)
Proband X13 (62)
Proband X14 (124)
Proband X15 (76)
Proband X2 (71)
Proband X4 (147)
Proband X5 (140)
Proband X6 (85)
Proband X7 (50)
Proband X8 (49)
Proband X9 (90)
sim (254)
2
0
−2
wavelet−HLL_glrlm_GrayLevelNonUniformity
frame

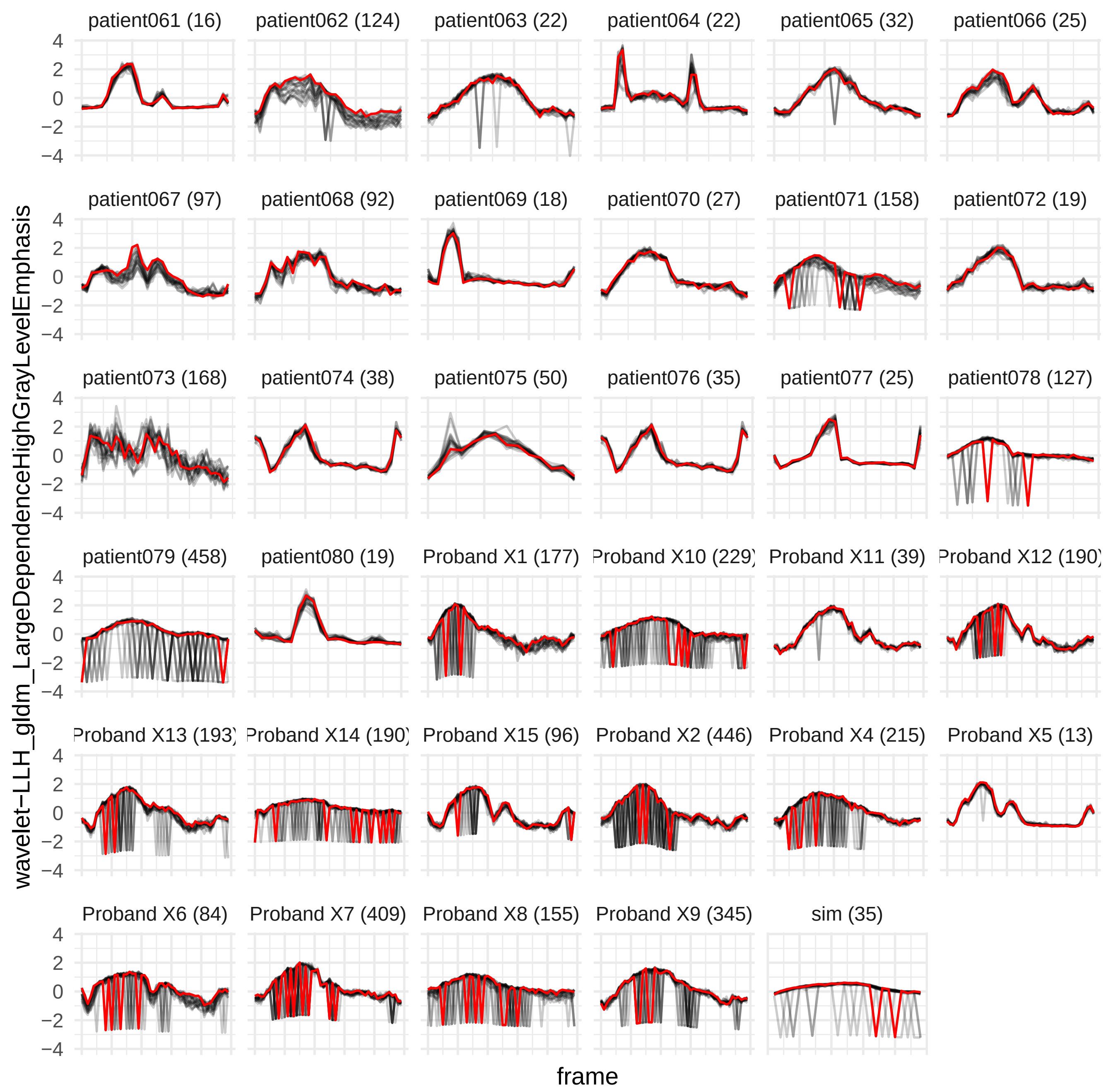

patient061 (16)
patient062 (124)
patient063 (22)
patient064 (22)
patient065 (32)
patient066 (25)
patient067 (97)
patient068 (92)
patient069 (18)
patient070 (27)
patient071 (158)
patient072 (19)
patient073 (168)
patient074 (38)
patient075 (50)
patient076 (35)
patient077 (25)
patient078 (127)
patient079 (458)
patient080 (19)
Proband X1 (177)
Proband X10 (229)
Proband X11 (39)
Proband X12 (190)
Proband X13 (193)
Proband X14 (190)
Proband X15 (96)
Proband X2 (446)
Proband X4 (215)
Proband X5 (13)
Proband X6 (84)
Proband X7 (409)
Proband X8 (155)
Proband X9 (345)
sim (35)
wavelet−LLH_gldm_LargeDependenceHighGrayLevelEmphasis
frame
4
2
0
−2
−4

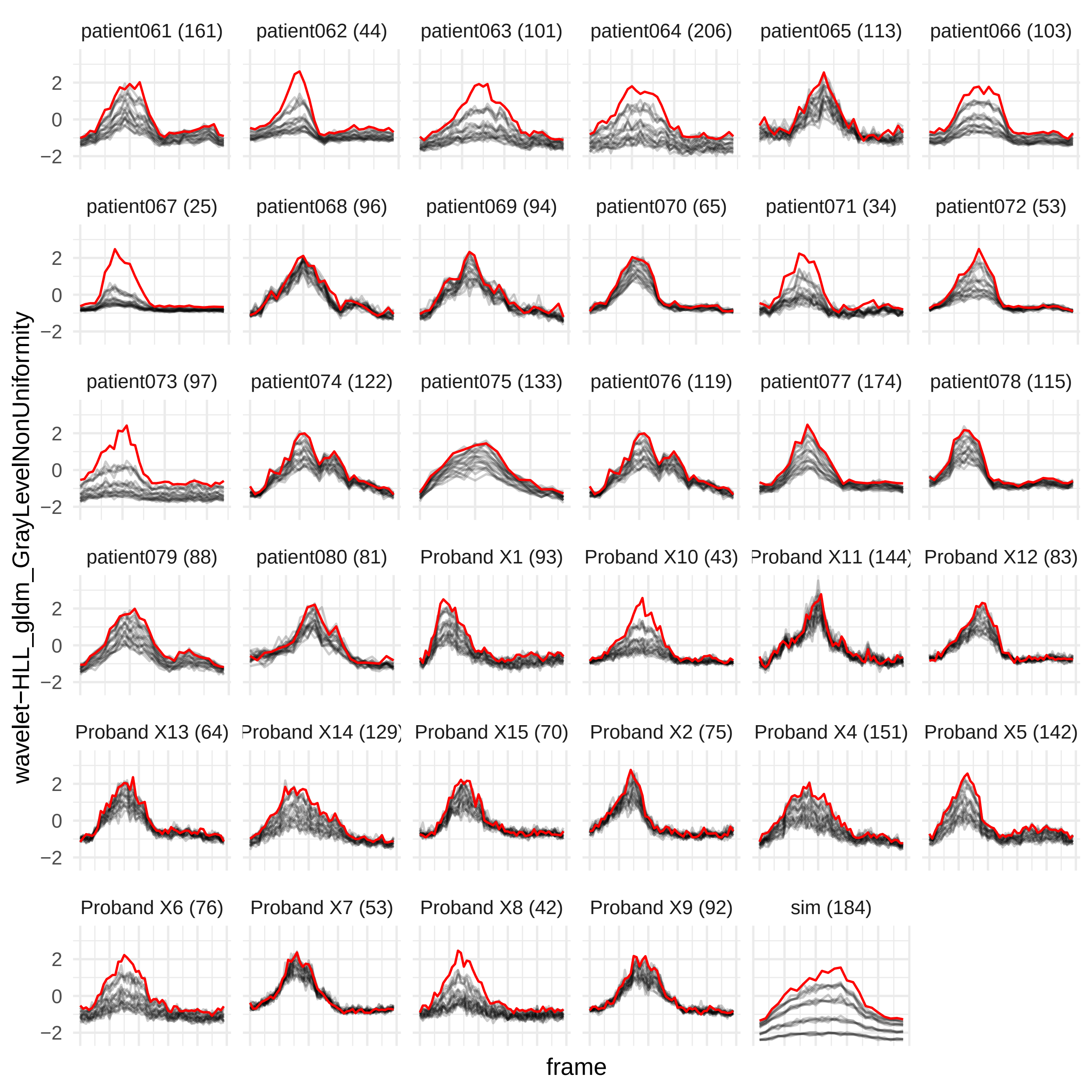
patient061 (161)
patient062 (44)
patient063 (101)
patient064 (206)
patient065 (113)
patient066 (103)
patient067 (25)
patient068 (96)
patient069 (94)
patient070 (65)
patient071 (34)
patient072 (53)
patient073 (97)
patient074 (122)
patient075 (133)
patient076 (119)
patient077 (174)
patient078 (115)
patient079 (88)
patient080 (81)
Proband X1 (93)
Proband X10 (43)
Proband X11 (144)
Proband X12 (83)
Proband X13 (64)
Proband X14 (129)
Proband X15 (70)
Proband X2 (75)
Proband X4 (151)
Proband X5 (142)
Proband X6 (76)
Proband X7 (53)
Proband X8 (42)
Proband X9 (92)
sim (184)
2
0
−2
wavelet−HLL_gldm_GrayLevelNonUniformity
frame

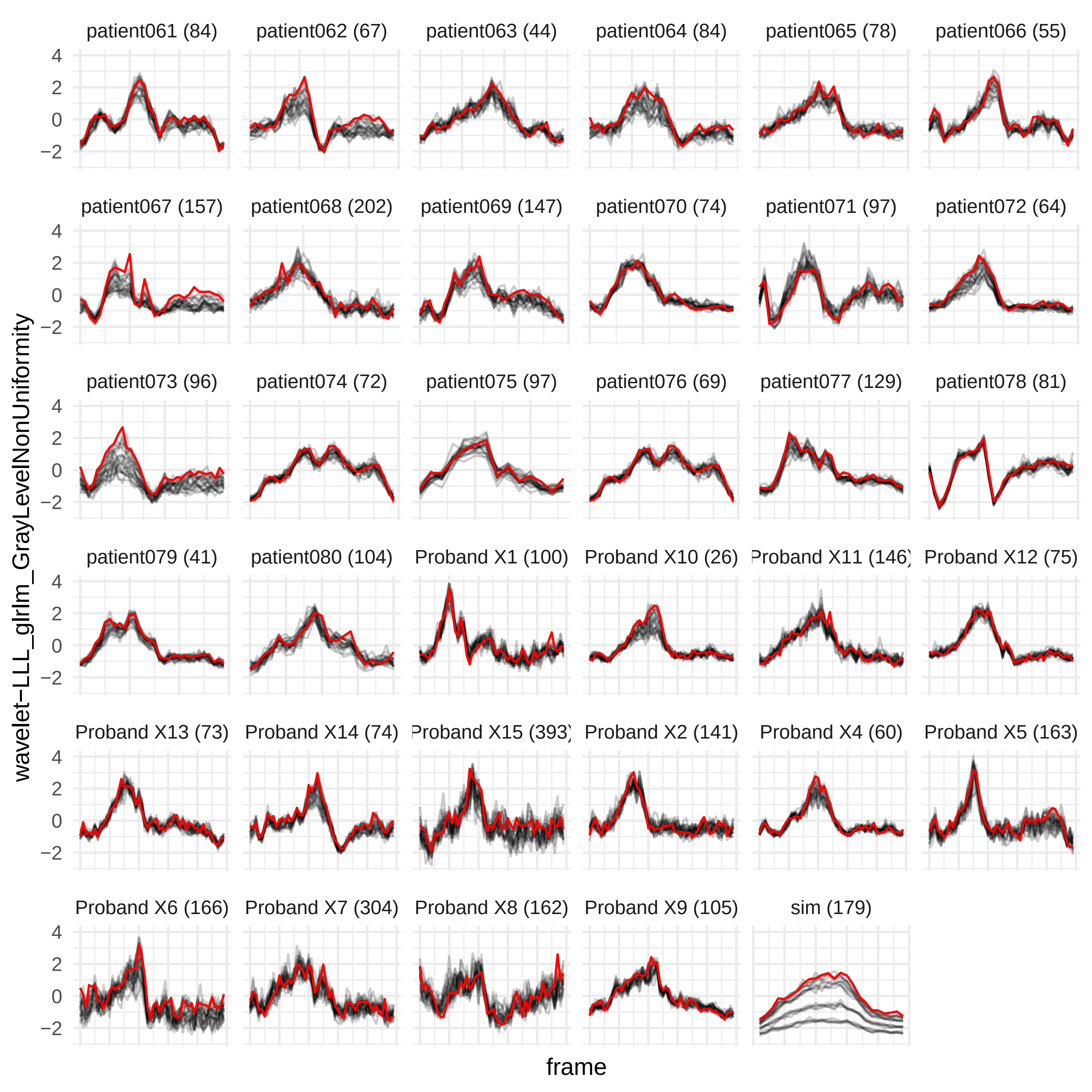

patient061 (84)
patient062 (67)
patient063 (44)
patient064 (84)
patient065 (78)
patient066 (55)
patient067 (157)
patient068 (202)
patient069 (147)
patient070 (74)
patient071 (97)
patient072 (64)
patient073 (96)
patient074 (72)
patient075 (97)
patient076 (69)
patient077 (129)
patient078 (81)
patient079 (41)
patient080 (104)
Proband X1 (100)
Proband X10 (26)
Proband X11 (146)
Proband X12 (75)
Proband X13 (73)
Proband X14 (74)
Proband X15 (393)
Proband X2 (141)
Proband X4 (60)
Proband X5 (163)
Proband X6 (166)
Proband X7 (304)
Proband X8 (162)
Proband X9 (105)
sim (179)
4
2
0
−2
wavelet−LLL_glrlm_GrayLevelNonUniformity
frame

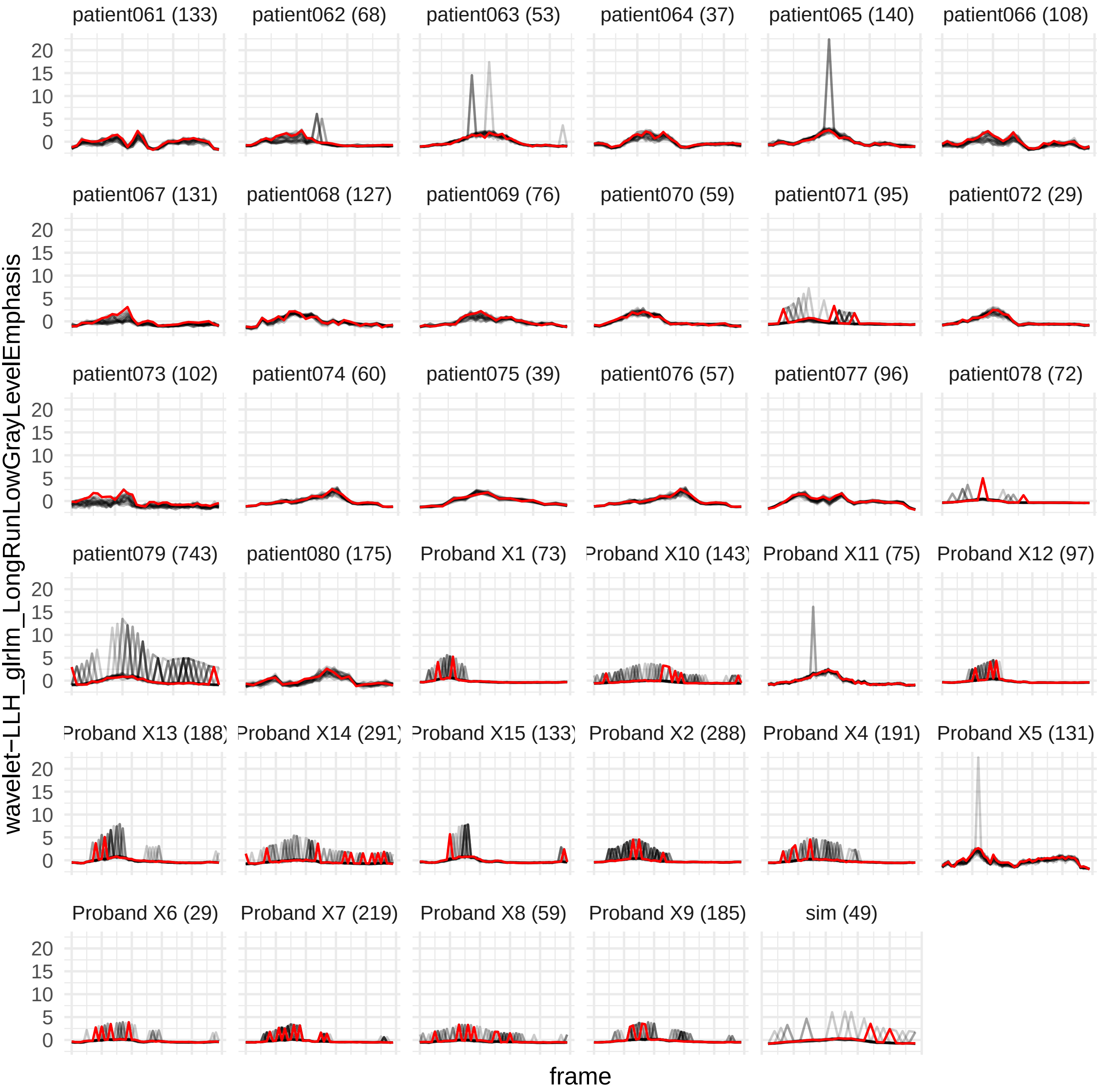

patient061 (133)
patient062 (68)
patient063 (53)
patient064 (37)
patient065 (140)
patient066 (108)
patient067 (131)
patient068 (127)
patient069 (76)
patient070 (59)
patient071 (95)
patient072 (29)
patient073 (102)
patient074 (60)
patient075 (39)
patient076 (57)
patient077 (96)
patient078 (72)
patient079 (743)
patient080 (175)
Proband X1 (73)
Proband X10 (143)
Proband X11 (75)
Proband X12 (97)
Proband X13 (188)
Proband X14 (291)
Proband X15 (133)
Proband X2 (288)
Proband X4 (191)
Proband X5 (131)
Proband X6 (29)
Proband X7 (219)
Proband X8 (59)
Proband X9 (185)
sim (49)
wavelet−LLH_glrlm_LongRunLowGrayLevelEmphasis
frame

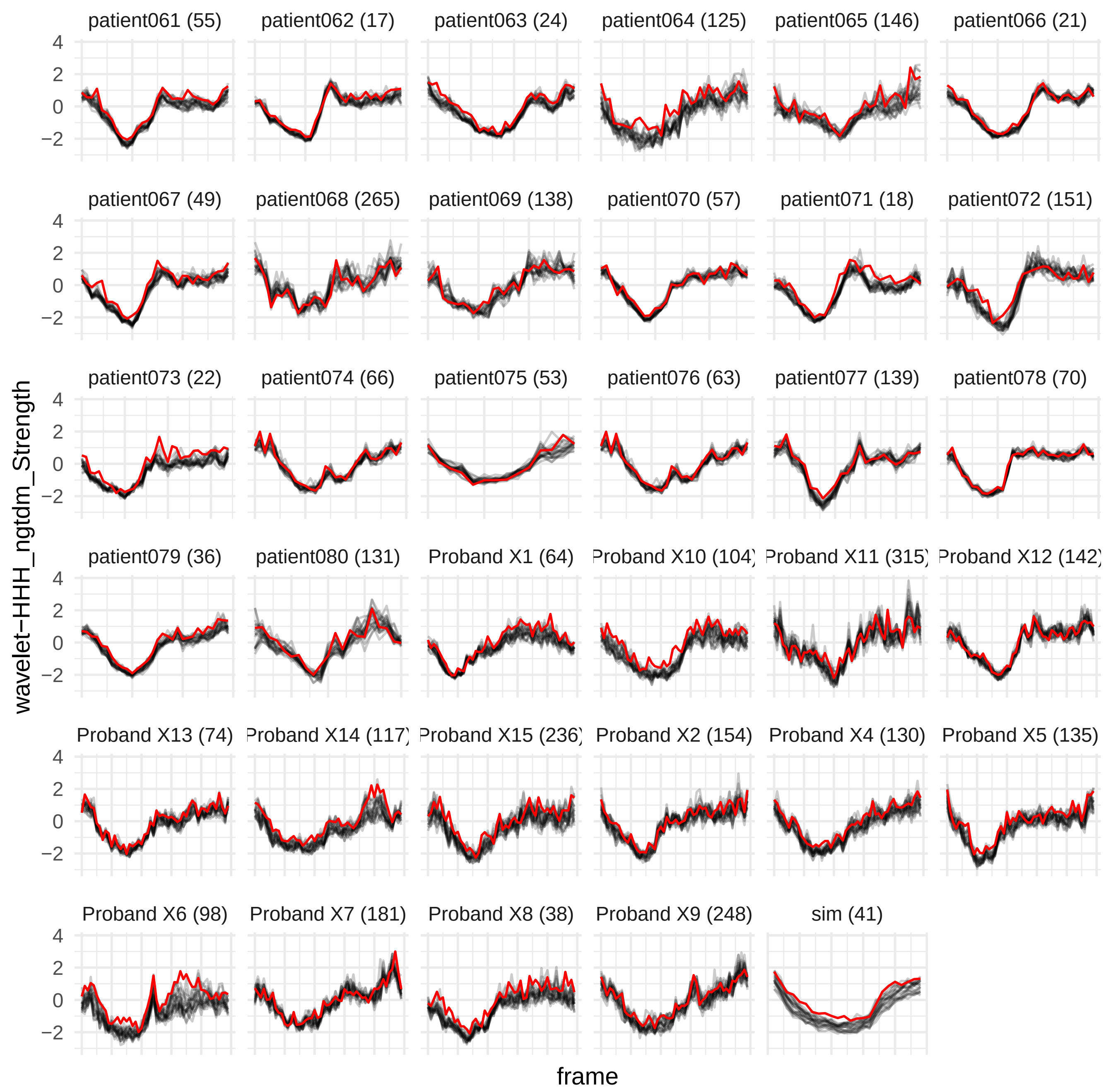

patient061 (55)
patient062 (17)
patient063 (24)
patient064 (125)
patient065 (146)
patient066 (21)
patient067 (49)
patient068 (265)
patient069 (138)
patient070 (57)
patient071 (18)
patient072 (151)
patient073 (22)
patient074 (66)
patient075 (53)
patient076 (63)
patient077 (139)
patient078 (70)
patient079 (36)
patient080 (131)
Proband X1 (64)
Proband X10 (104)
Proband X11 (315)
Proband X12 (142)
Proband X13 (74)
Proband X14 (117)
Proband X15 (236)
Proband X2 (154)
Proband X4 (130)
Proband X5 (135)
Proband X6 (98)
Proband X7 (181)
Proband X8 (38)
Proband X9 (248)
sim (41)
4
2
0
−2
wavelet−HHH_ngtdm_Strength
frame

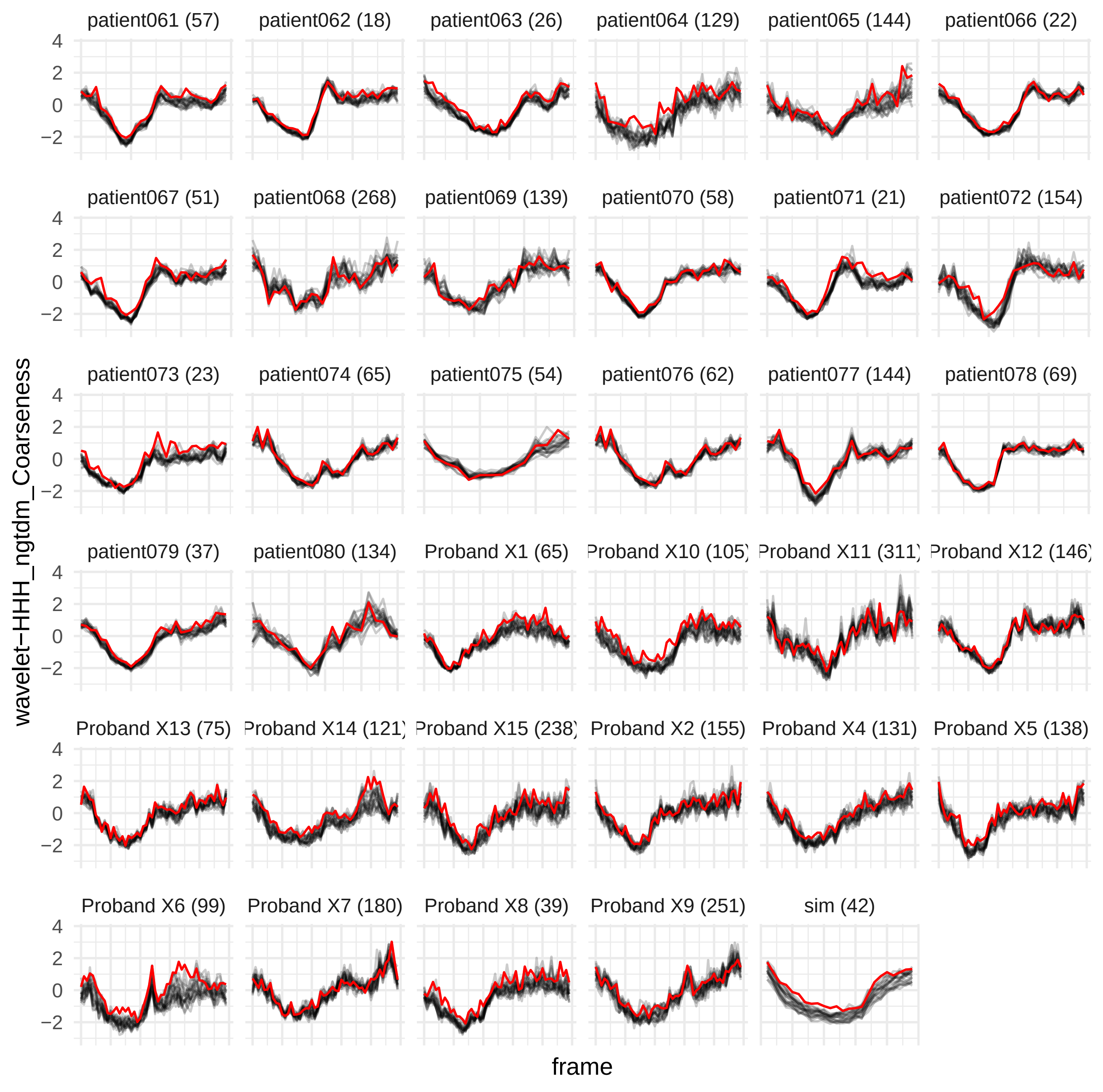
patient061 (57)
patient062 (18)
patient063 (26)
patient064 (129)
patient065 (144)
patient066 (22)
patient067 (51)
patient068 (268)
patient069 (139)
patient070 (58)
patient071 (21)
patient072 (154)
patient073 (23)
patient074 (65)
patient075 (54)
patient076 (62)
patient077 (144)
patient078 (69)
patient079 (37)
patient080 (134)
Proband X1 (65)
Proband X10 (105)
Proband X11 (311)
Proband X12 (146)
Proband X13 (75)
Proband X14 (121)
Proband X15 (238)
Proband X2 (155)
Proband X4 (131)
Proband X5 (138)
Proband X6 (99)
Proband X7 (180)
Proband X8 (39)
Proband X9 (251)
sim (42)
4
2
0
−2
wavelet−HHH_ngtdm_Coarseness
frame

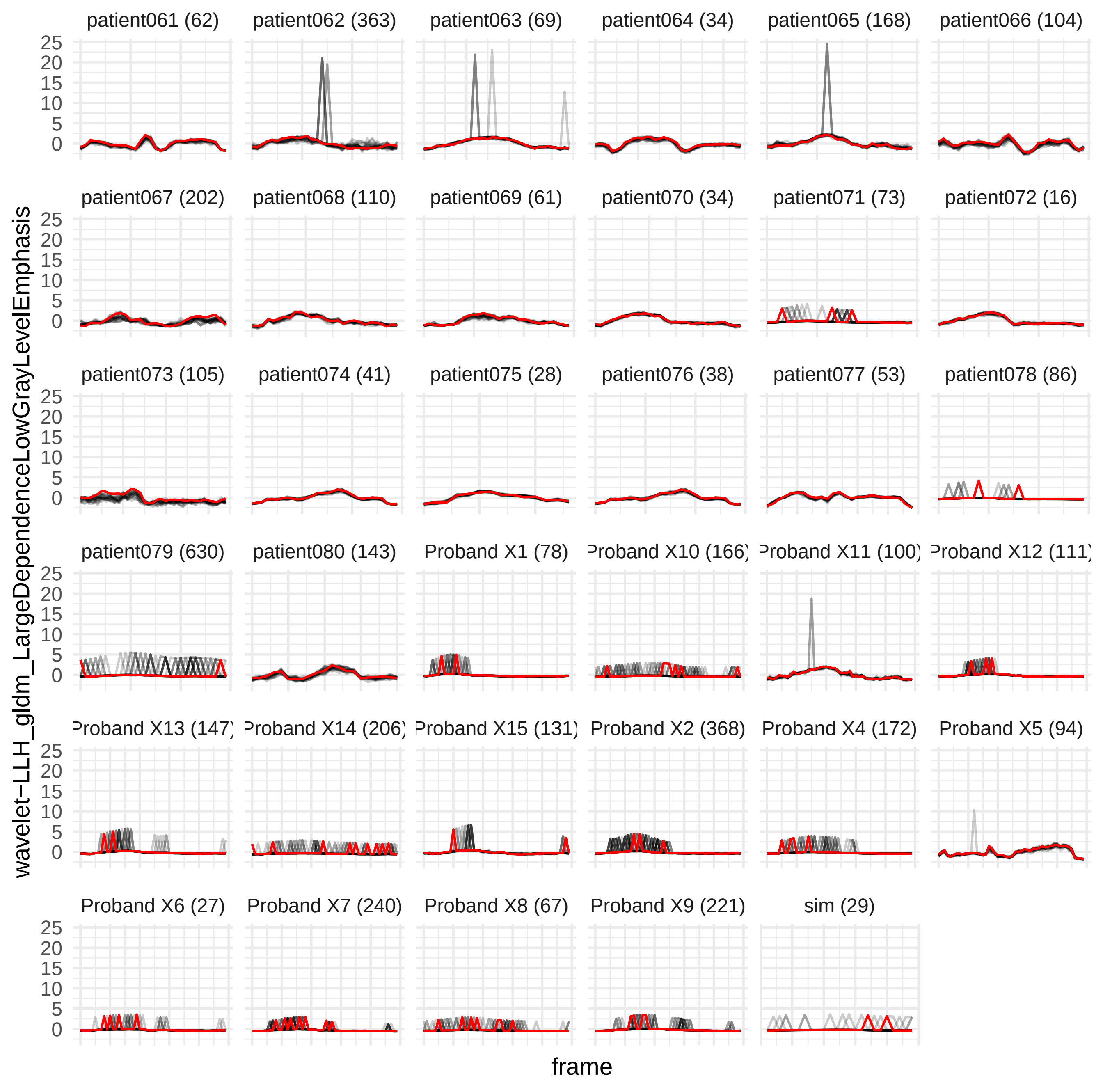
patient061 (62)
patient062 (363)
patient063 (69)
patient064 (34)
patient065 (168)
patient066 (104)
patient067 (202)
patient068 (110)
patient069 (61)
patient070 (34)
patient071 (73)
patient072 (16)
patient073 (105)
patient074 (41)
patient075 (28)
patient076 (38)
patient077 (53)
patient078 (86)
patient079 (630)
patient080 (143)
Proband X1 (78)
Proband X10 (166)
Proband X11 (100)
Proband X12 (111)
Proband X13 (147)
Proband X14 (206)
Proband X15 (131)
Proband X2 (368)
Proband X4 (172)
Proband X5 (94)
Proband X6 (27)
Proband X7 (240)
Proband X8 (67)
Proband X9 (221)
sim (29)
0
5
10
15
20
25
wavelet−LLH_gldm_LargeDependenceLowGrayLevelEmphasis
frame

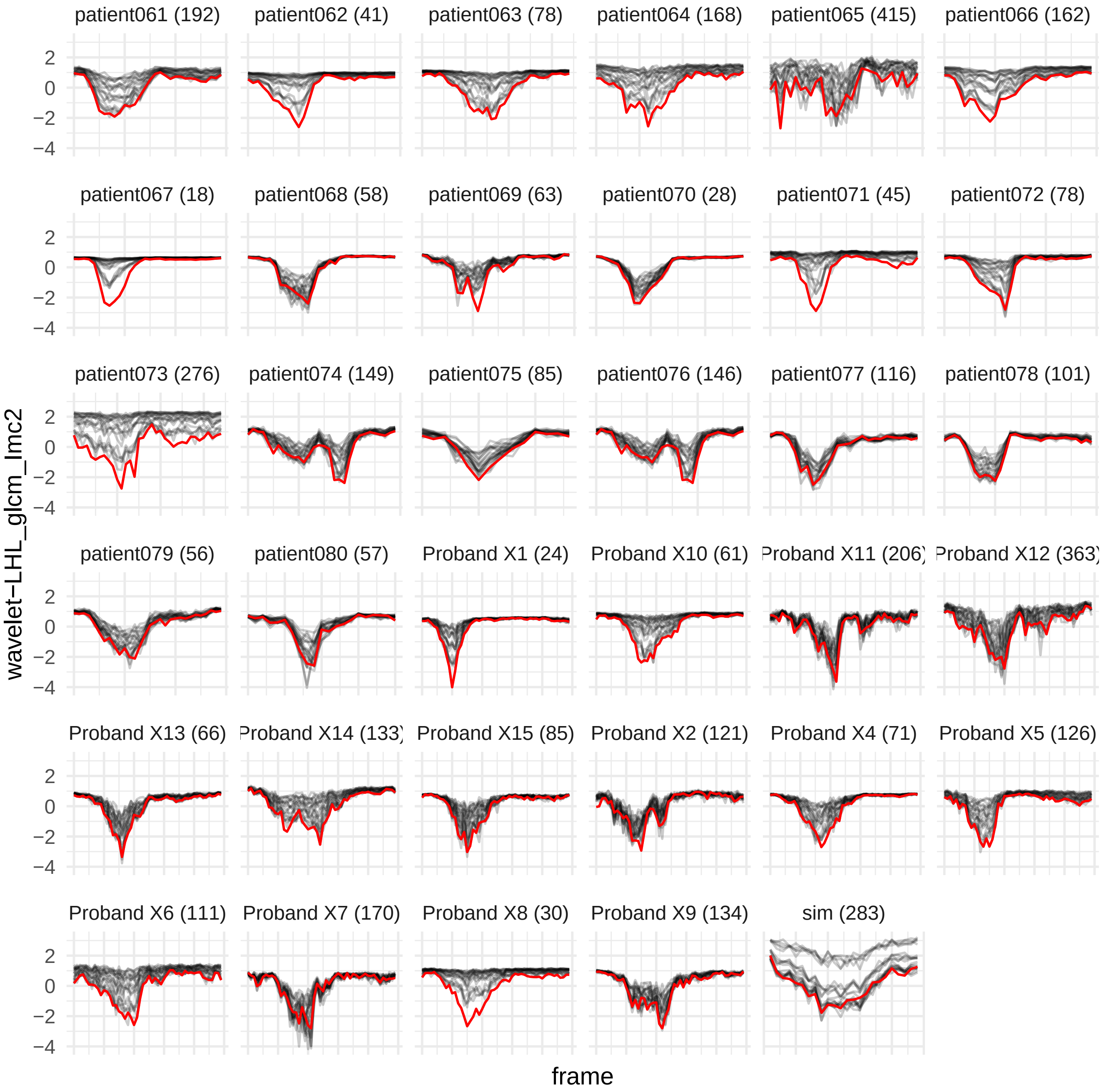
patient061 (192)
patient062 (41)
patient063 (78)
patient064 (168)
patient065 (415)
patient066 (162)
patient067 (18)
patient068 (58)
patient069 (63)
patient070 (28)
patient071 (45)
patient072 (78)
patient073 (276)
patient074 (149)
patient075 (85)
patient076 (146)
patient077 (116)
patient078 (101)
patient079 (56)
patient080 (57)
Proband X1 (24)
Proband X10 (61)
Proband X11 (206)
Proband X12 (363)
Proband X13 (66)
Proband X14 (133)
Proband X15 (85)
Proband X2 (121)
Proband X4 (71)
Proband X5 (126)
Proband X6 (111)
Proband X7 (170)
Proband X8 (30)
Proband X9 (134)
sim (283)
2
0
−2
−4
wavelet−LHL_glcm_Imc2
frame

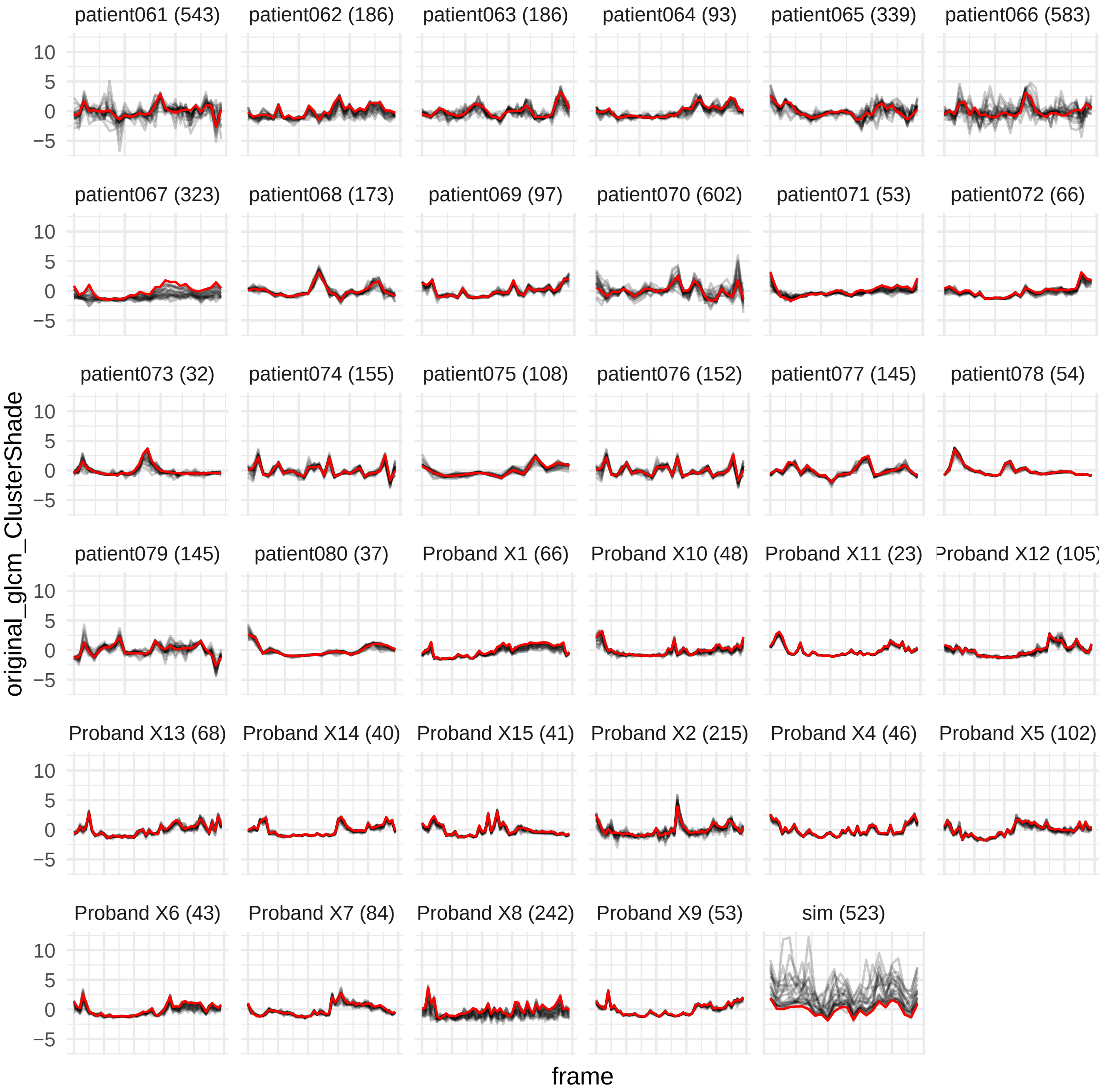

patient061 (543)
patient062 (186)
patient063 (186)
patient064 (93)
patient065 (339)
patient066 (583)
patient067 (323)
patient068 (173)
patient069 (97)
patient070 (602)
patient071 (53)
patient072 (66)
patient073 (32)
patient074 (155)
patient075 (108)
patient076 (152)
patient077 (145)
patient078 (54)
patient079 (145)
patient080 (37)
Proband X1 (66)
Proband X10 (48)
Proband X11 (23)
Proband X12 (105)
Proband X13 (68)
Proband X14 (40)
Proband X15 (41)
Proband X2 (215)
Proband X4 (46)
Proband X5 (102)
Proband X6 (43)
Proband X7 (84)
Proband X8 (242)
Proband X9 (53)
sim (523)
original_glcm_ClusterShade
frame
10
5
0
−5

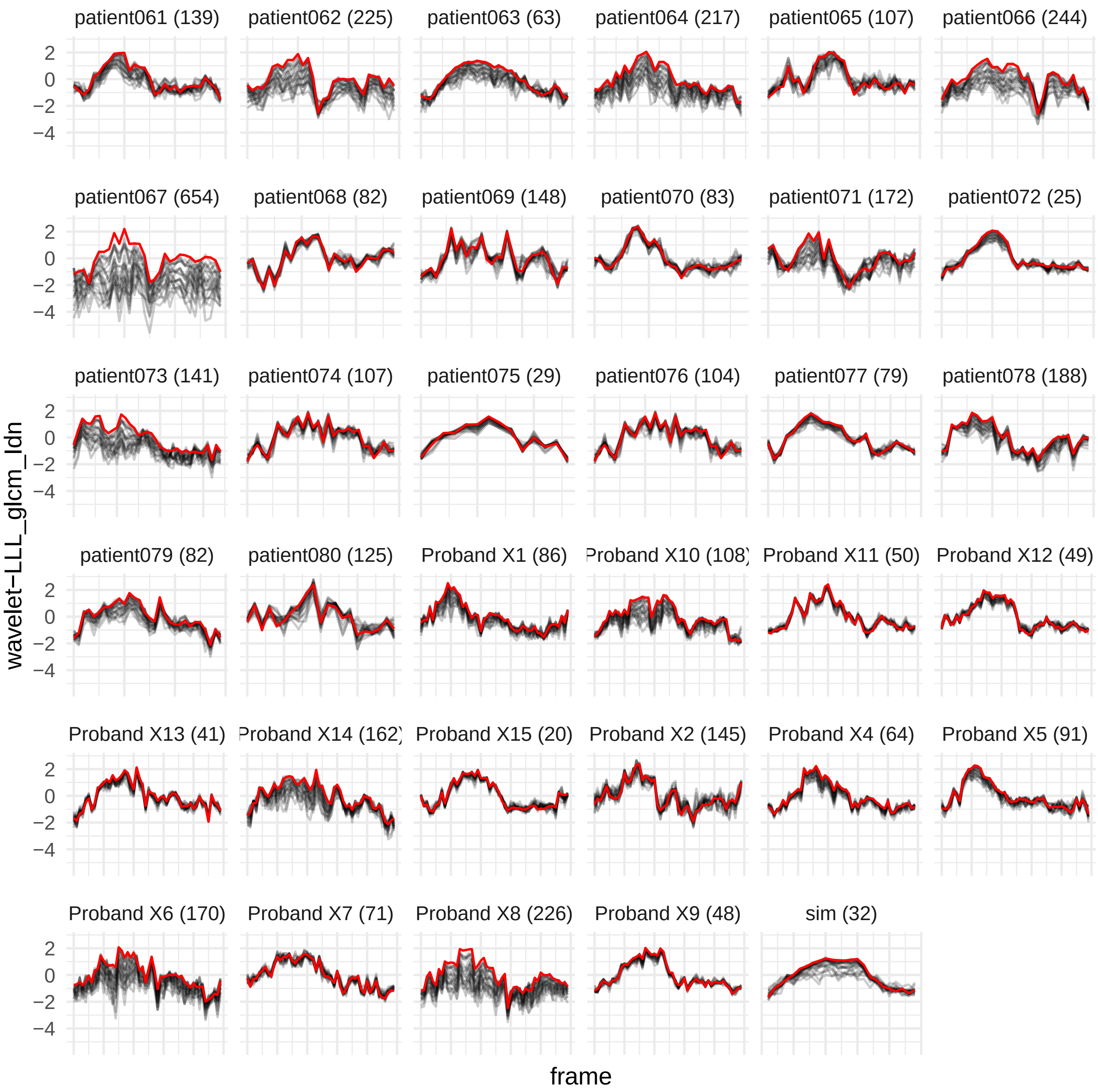

patient061 (139)
patient062 (225)
patient063 (63)
patient064 (217)
patient065 (107)
patient066 (244)
patient067 (654)
patient068 (82)
patient069 (148)
patient070 (83)
patient071 (172)
patient072 (25)
patient073 (141)
patient074 (107)
patient075 (29)
patient076 (104)
patient077 (79)
patient078 (188)
patient079 (82)
patient080 (125)
Proband X1 (86)
Proband X10 (108)
Proband X11 (50)
Proband X12 (49)
Proband X13 (41)
Proband X14 (162)
Proband X15 (20)
Proband X2 (145)
Proband X4 (64)
Proband X5 (91)
Proband X6 (170)
Proband X7 (71)
Proband X8 (226)
Proband X9 (48)
sim (32)
2
0
−2
−4
wavelet−LLL_glcm_Idn
frame

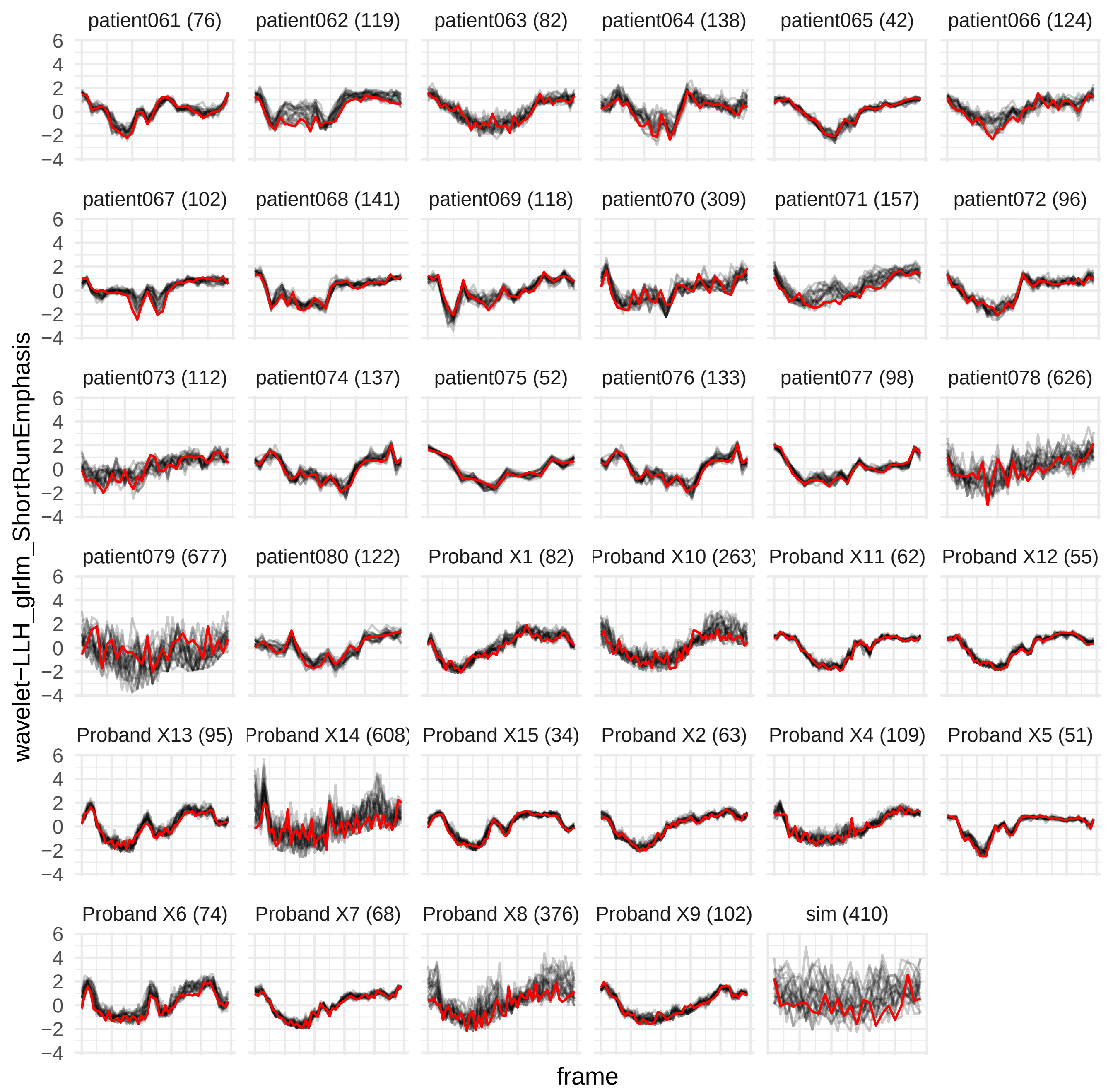

patient061 (76)
patient062 (119)
patient063 (82)
patient064 (138)
patient065 (42)
patient066 (124)
patient067 (102)
patient068 (141)
patient069 (118)
patient070 (309)
patient071 (157)
patient072 (96)
patient073 (112)
patient074 (137)
patient075 (52)
patient076 (133)
patient077 (98)
patient078 (626)
patient079 (677)
patient080 (122)
Proband X1 (82)
Proband X10 (263)
Proband X11 (62)
Proband X12 (55)
Proband X13 (95)
Proband X14 (608)
Proband X15 (34)
Proband X2 (63)
Proband X4 (109)
Proband X5 (51)
Proband X6 (74)
Proband X7 (68)
Proband X8 (376)
Proband X9 (102)
sim (410)
wavelet−LLH_glrlm_ShortRunEmphasis
frame

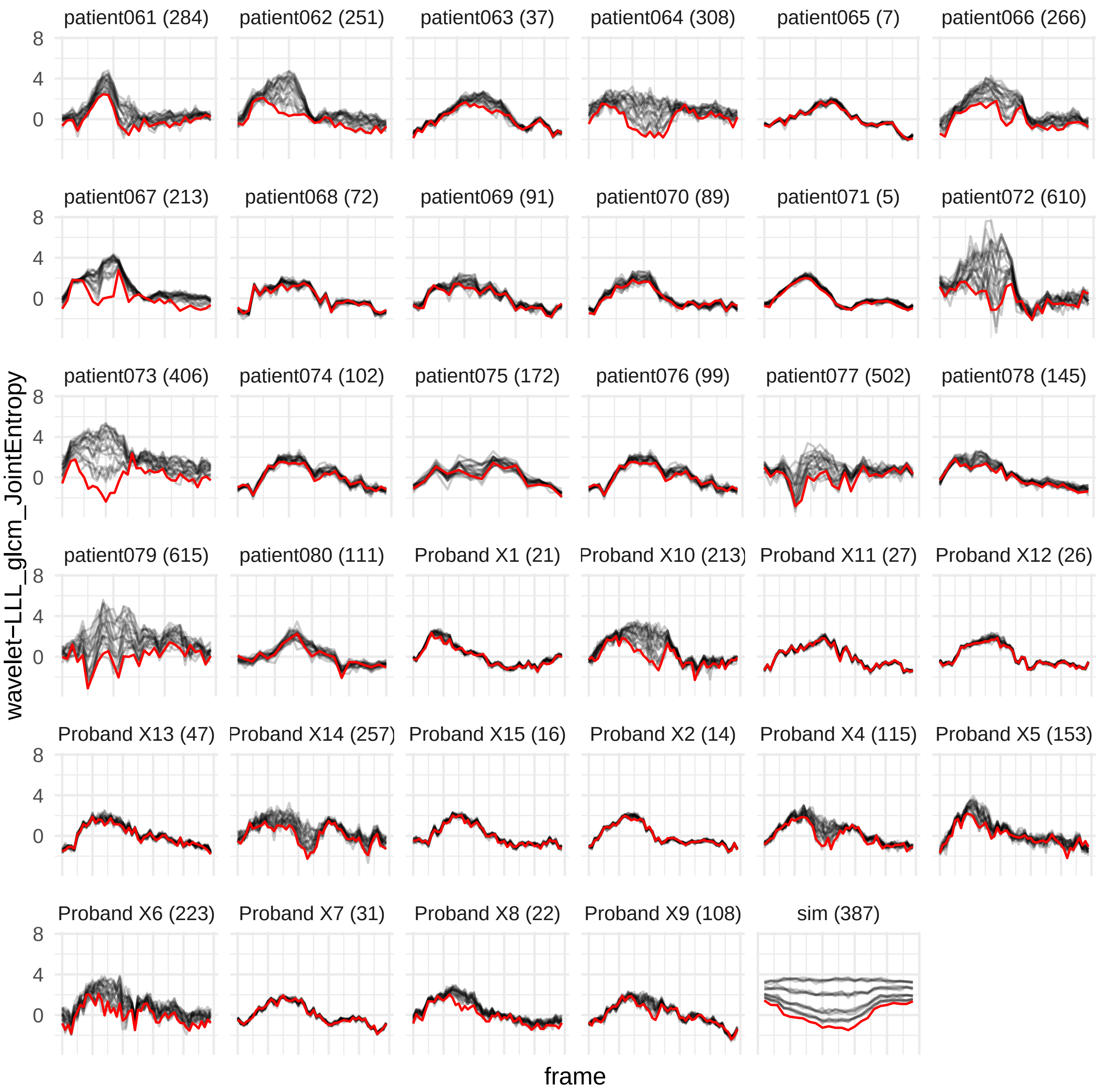
patient061 (284)
patient062 (251)
patient063 (37)
patient064 (308)
patient065 (7)
patient066 (266)
patient067 (213)
patient068 (72)
patient069 (91)
patient070 (89)
patient071 (5)
patient072 (610)
patient073 (406)
patient074 (102)
patient075 (172)
patient076 (99)
patient077 (502)
patient078 (145)
patient079 (615)
patient080 (111)
Proband X1 (21)
Proband X10 (213)
Proband X11 (27)
Proband X12 (26)
Proband X13 (47)
Proband X14 (257)
Proband X15 (16)
Proband X2 (14)
Proband X4 (115)
Proband X5 (153)
Proband X6 (223)
Proband X7 (31)
Proband X8 (22)
Proband X9 (108)
sim (387)
wavelet−LLL_glcm_JointEntropy
frame
0
4
8

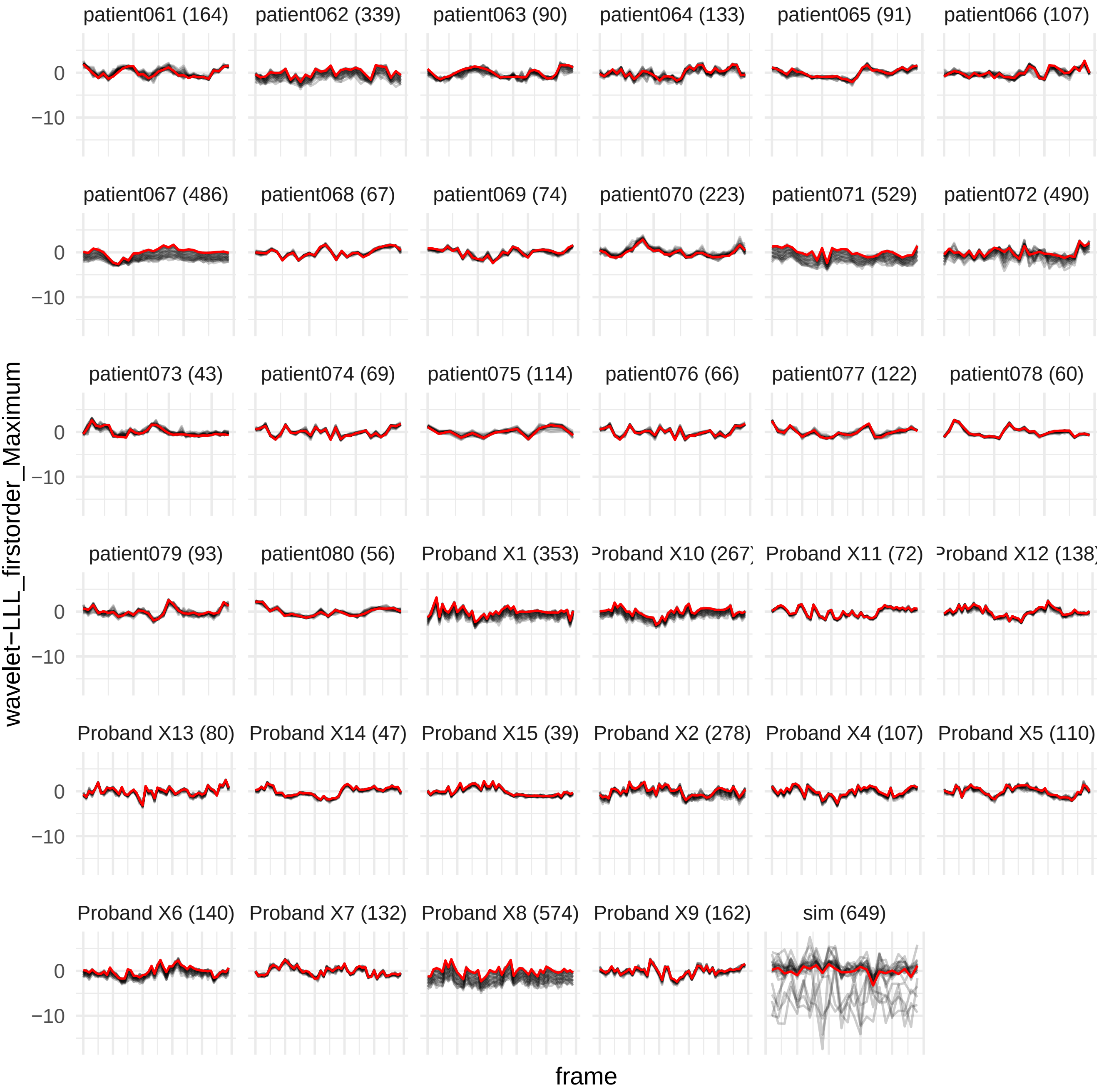
patient061 (164)
patient062 (339)
patient063 (90)
patient064 (133)
patient065 (91)
patient066 (107)
patient067 (486)
patient068 (67)
patient069 (74)
patient070 (223)
patient071 (529)
patient072 (490)
patient073 (43)
patient074 (69)
patient075 (114)
patient076 (66)
patient077 (122)
patient078 (60)
patient079 (93)
patient080 (56)
Proband X1 (353)
Proband X11 (72)
Proband X13 (80)
Proband X14 (47)
Proband X15 (39)
Proband X2 (278)
Proband X4 (107)
Proband X5 (110)
Proband X6 (140)
Proband X7 (132)
Proband X8 (574)
Proband X9 (162)
sim (649)
0
−10
wavelet−LLL_firstorder_Maximum
frame

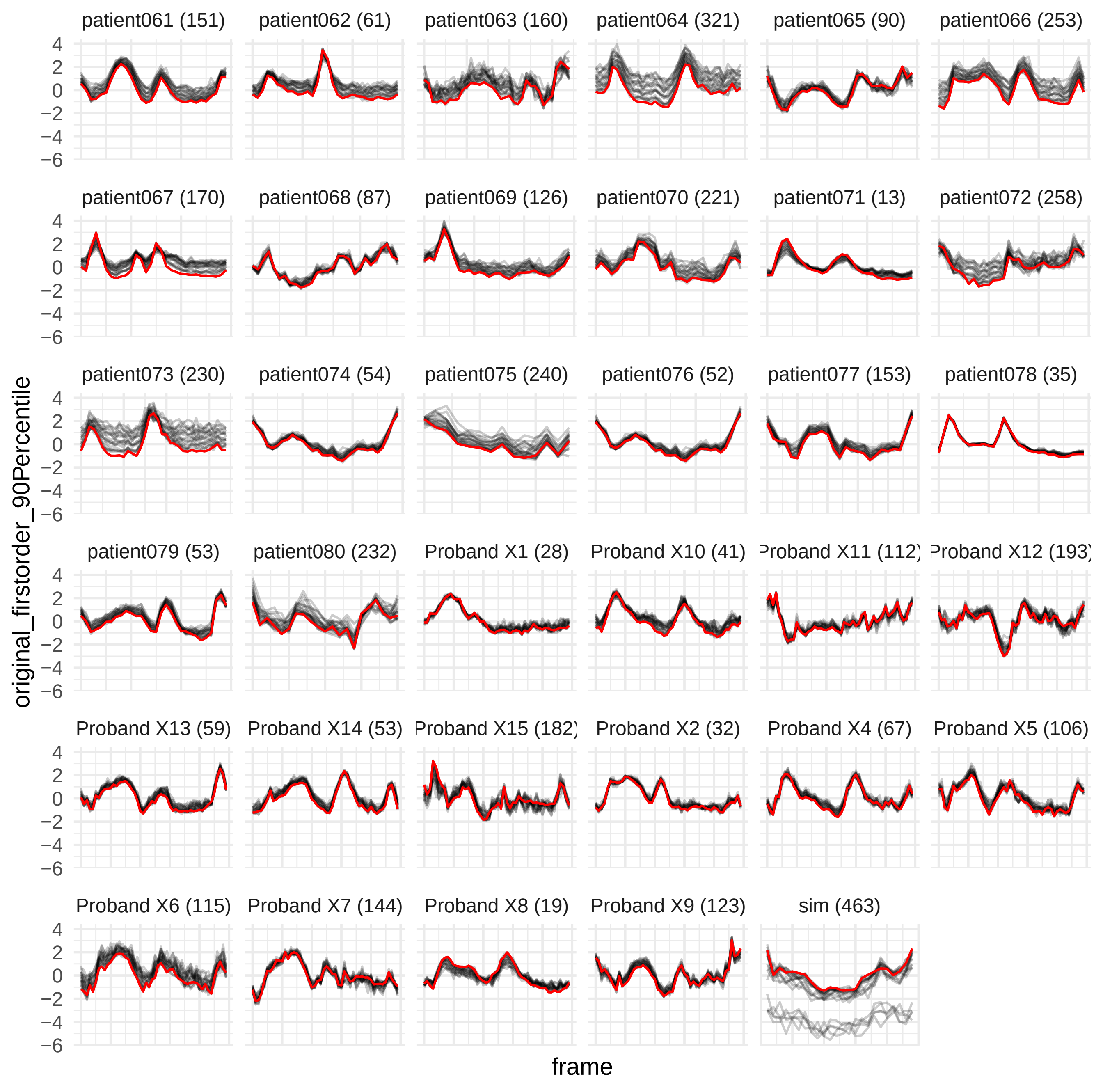

patient061 (151)
patient062 (61)
patient063 (160)
patient064 (321)
patient065 (90)
patient066 (253)
patient067 (170)
patient068 (87)
patient069 (126)
patient070 (221)
patient071 (13)
patient072 (258)
patient073 (230)
patient074 (54)
patient075 (240)
patient076 (52)
patient077 (153)
patient078 (35)
patient079 (53)
patient080 (232)
Proband X1 (28)
Proband X10 (41)
Proband X11 (112)
Proband X12 (193)
Proband X13 (59)
Proband X14 (53)
Proband X15 (182)
Proband X2 (32)
Proband X4 (67)
Proband X5 (106)
Proband X6 (115)
Proband X7 (144)
Proband X8 (19)
Proband X9 (123)
sim (463)
original_firstorder_90Percentile
frame
4
2
0
−2
−4
−6

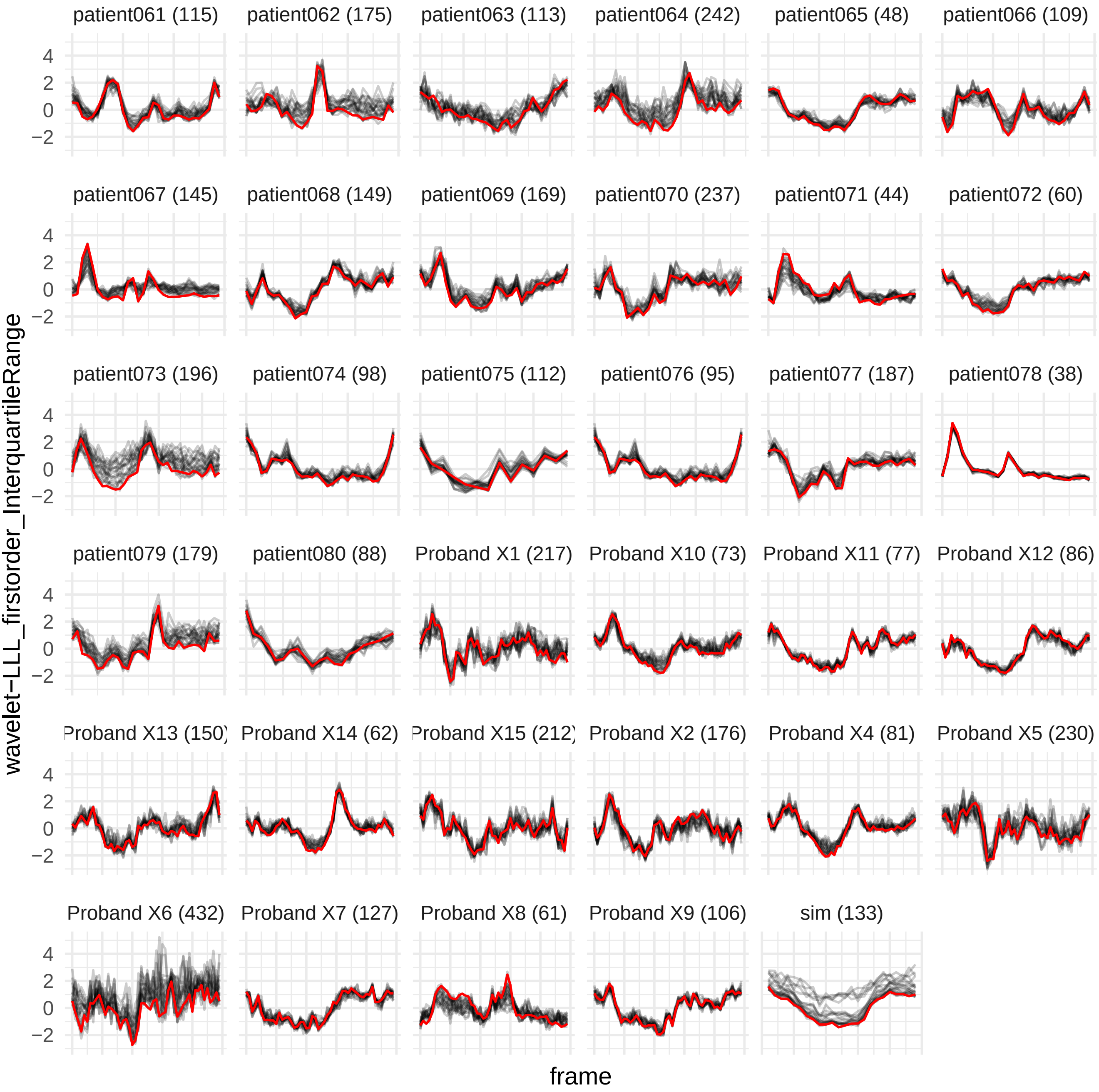
patient061 (115)
patient062 (175)
patient063 (113)
patient064 (242)
patient065 (48)
patient066 (109)
patient067 (145)
patient068 (149)
patient069 (169)
patient070 (237)
patient071 (44)
patient072 (60)
patient073 (196)
patient074 (98)
patient075 (112)
patient076 (95)
patient077 (187)
patient078 (38)
patient079 (179)
patient080 (88)
Proband X1 (217)
Proband X10 (73)
Proband X11 (77)
Proband X12 (86)
Proband X13 (150)
Proband X14 (62)
Proband X15 (212)
Proband X2 (176)
Proband X4 (81)
Proband X5 (230)
Proband X6 (432)
Proband X7 (127)
Proband X8 (61)
Proband X9 (106)
sim (133)
wavelet−LLL_firstorder_InterquartileRange
frame
4
2
0
−2

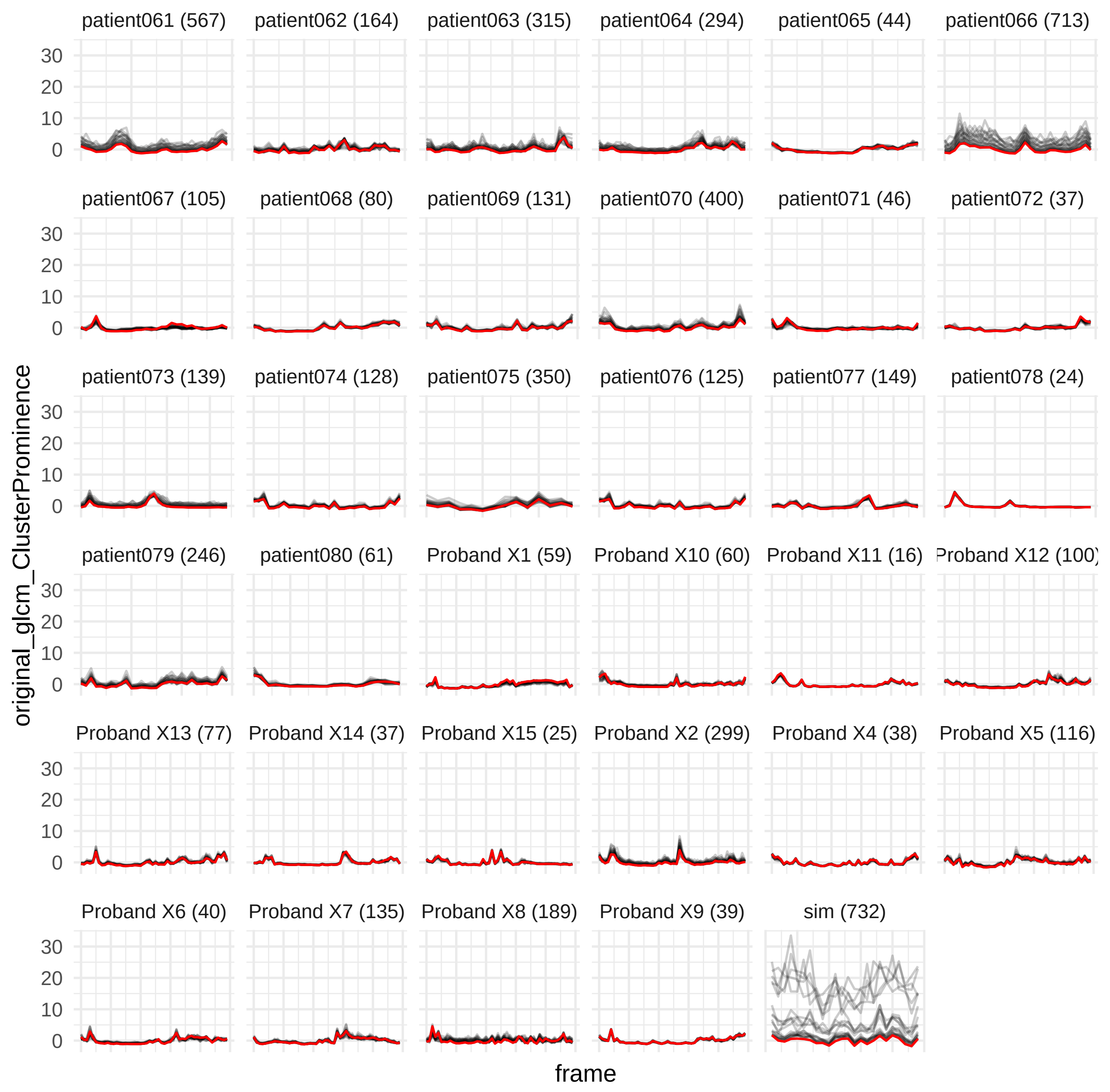

patient061 (567)
patient062 (164)
patient063 (315)
patient064 (294)
patient065 (44)
patient066 (713)
patient067 (105)
patient068 (80)
patient069 (131)
patient070 (400)
patient071 (46)
patient072 (37)
patient073 (139)
patient074 (128)
patient075 (350)
patient076 (125)
patient077 (149)
patient078 (24)
patient079 (246)
patient080 (61)
Proband X1 (59)
Proband X10 (60)
Proband X11 (16)
Proband X12 (100)
Proband X13 (77)
Proband X14 (37)
Proband X15 (25)
Proband X2 (299)
Proband X4 (38)
Proband X5 (116)
Proband X6 (40)
Proband X7 (135)
Proband X8 (189)
Proband X9 (39)
sim (732)
0
10
20
30
original_glcm_ClusterProminence
frame

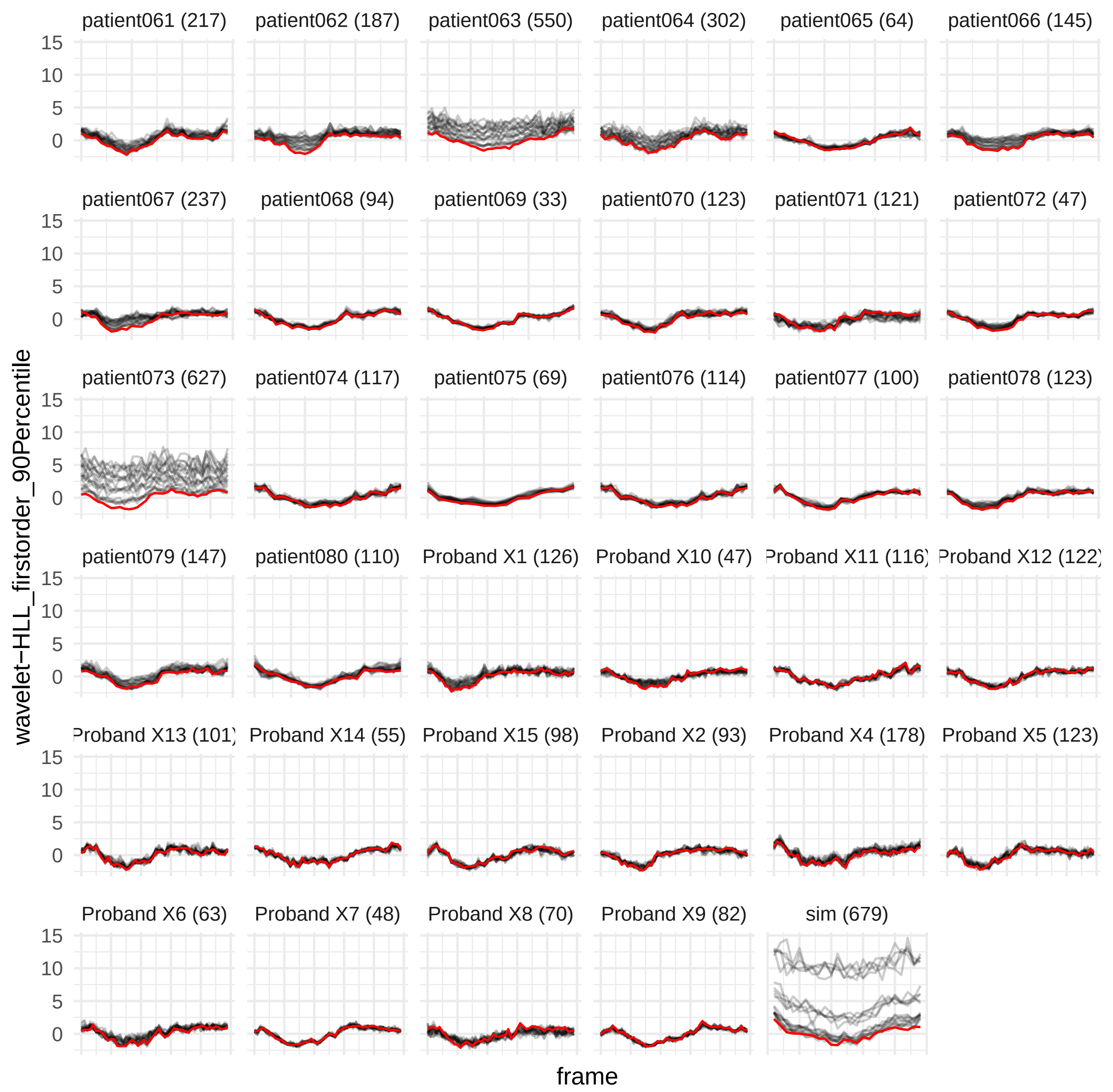

patient061 (217)
patient062 (187)
patient063 (550)
patient064 (302)
patient065 (64)
patient066 (145)
patient067 (237)
patient068 (94)
patient069 (33)
patient070 (123)
patient071 (121)
patient072 (47)
patient073 (627)
patient074 (117)
patient075 (69)
patient076 (114)
patient077 (100)
patient078 (123)
patient079 (147)
patient080 (110)
Proband X1 (126)
Proband X10 (47)
Proband X11 (116)
Proband X12 (122)
Proband X13 (101)
Proband X14 (55)
Proband X15 (98)
Proband X2 (93)
Proband X4 (178)
Proband X5 (123)
Proband X6 (63)
Proband X7 (48)
Proband X8 (70)
Proband X9 (82)
sim (679)
15
10
5
0
wavelet−HLL_firstorder_90Percentile
frame

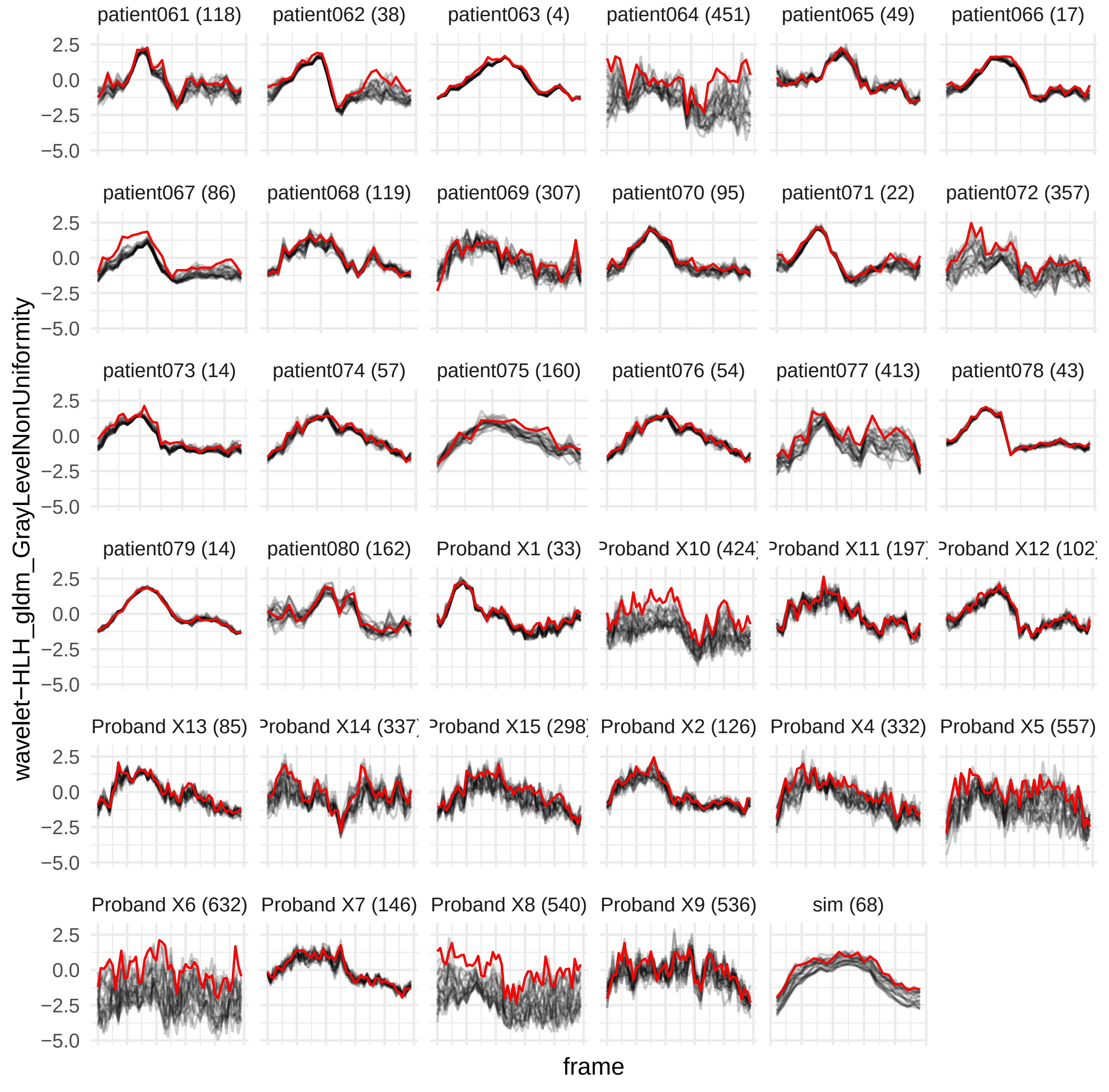
patient061 (118)
patient062 (38)
patient063 (4)
patient064 (451)
patient065 (49)
patient066 (17)
patient067 (86)
patient068 (119)
patient069 (307)
patient070 (95)
patient071 (22)
patient072 (357)
patient073 (14)
patient074 (57)
patient075 (160)
patient076 (54)
patient077 (413)
patient078 (43)
patient079 (14)
patient080 (162)
Proband X1 (33)
Proband X10 (424)
Proband X11 (197)
Proband X12 (102)
Proband X13 (85)
Proband X14 (337)
Proband X15 (298)
Proband X2 (126)
Proband X4 (332)
Proband X5 (557)
Proband X6 (632)
Proband X7 (146)
Proband X8 (540)
Proband X9 (536)
sim (68)
2.5
0.0
−2.5
−5.0
wavelet−HLH_gldm_GrayLevelNonUniformity
frame

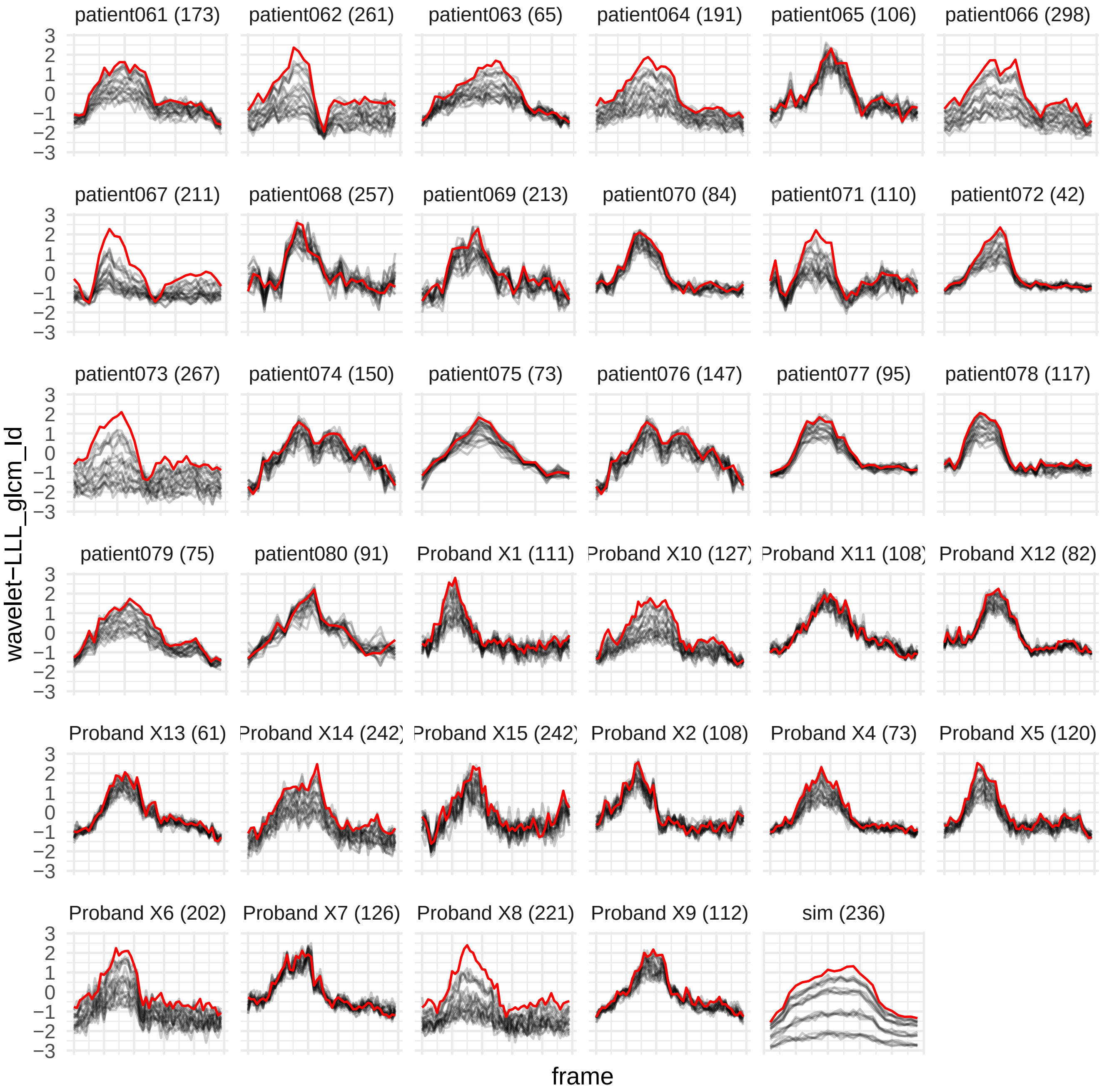

patient061 (173)
patient062 (261)
patient063 (65)
patient064 (191)
patient065 (106)
patient066 (298)
patient067 (211)
patient068 (257)
patient069 (213)
patient070 (84)
patient071 (110)
patient072 (42)
patient073 (267)
patient074 (150)
patient075 (73)
patient076 (147)
patient077 (95)
patient078 (117)
patient079 (75)
patient080 (91)
Proband X1 (111)
Proband X10 (127)
Proband X11 (108)
Proband X12 (82)
Proband X13 (61)
Proband X14 (242)
Proband X15 (242)
Proband X2 (108)
Proband X4 (73)
Proband X5 (120)
Proband X6 (202)
Proband X7 (126)
Proband X8 (221)
Proband X9 (112)
sim (236)
wavelet−LLL_glcm_Id
frame

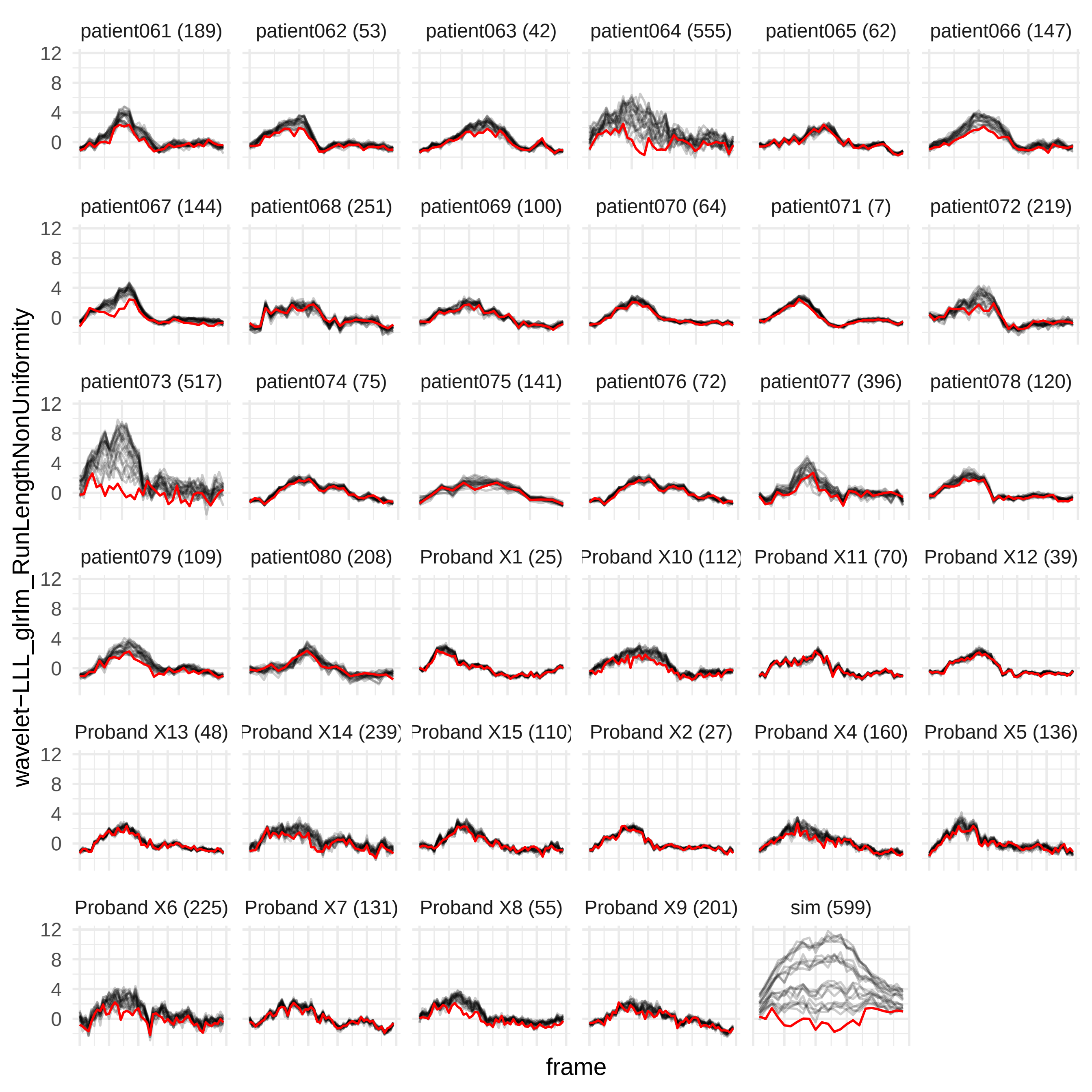

patient061 (189)
patient062 (53)
patient063 (42)
patient064 (555)
patient065 (62)
patient066 (147)
patient067 (144)
patient068 (251)
patient069 (100)
patient070 (64)
patient071 (7)
patient072 (219)
patient073 (517)
patient074 (75)
patient075 (141)
patient076 (72)
patient077 (396)
patient078 (120)
patient079 (109)
patient080 (208)
Proband X1 (25)
Proband X10 (112)
Proband X11 (70)
Proband X12 (39)
Proband X13 (48)
Proband X14 (239)
Proband X15 (110)
Proband X2 (27)
Proband X4 (160)
Proband X5 (136)
Proband X6 (225)
Proband X7 (131)
Proband X8 (55)
Proband X9 (201)
sim (599)
12
8
4
0
wavelet−LLL_glrlm_RunLengthNonUniformity
frame

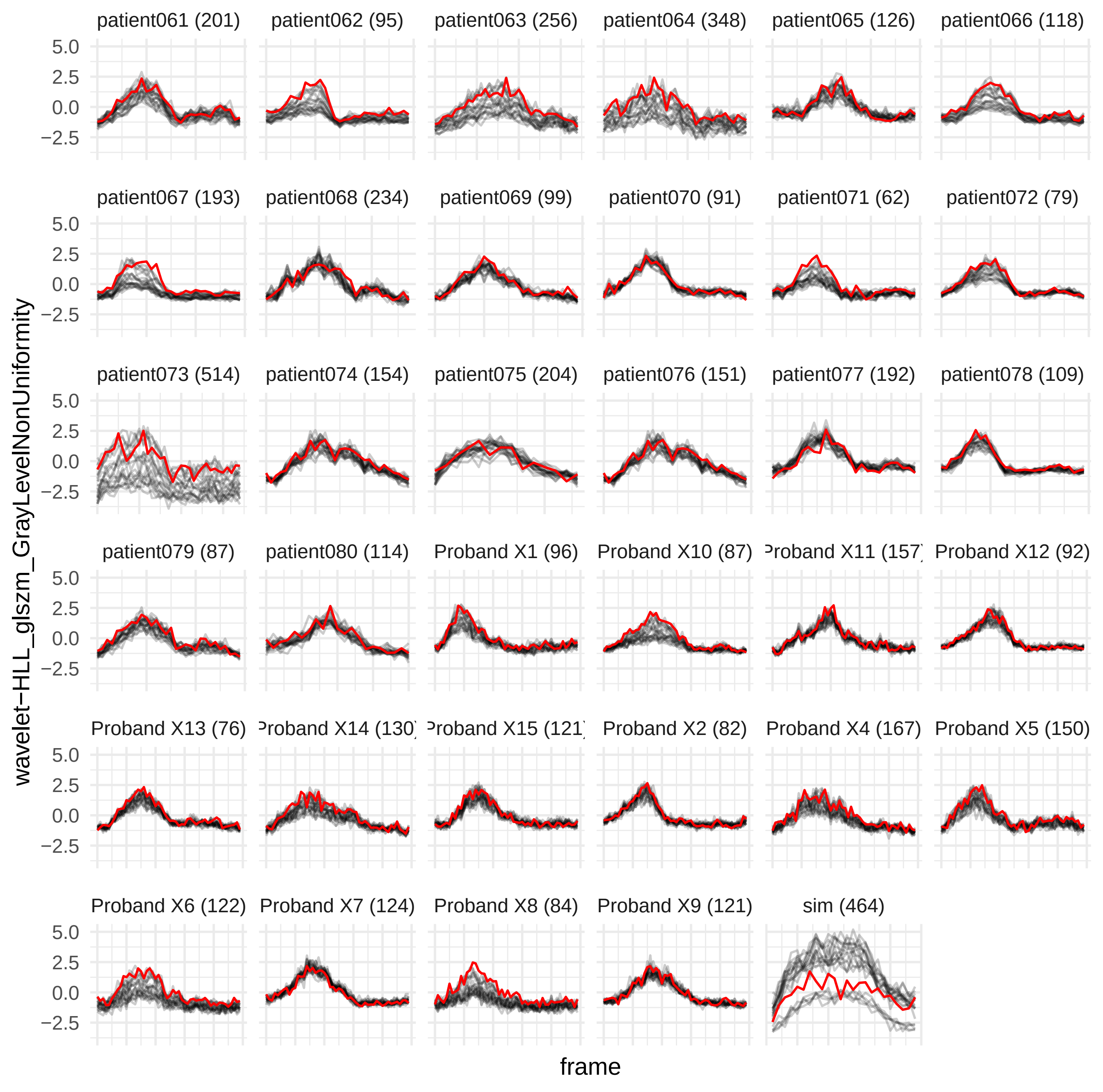

patient061 (201)
patient062 (95)
patient063 (256)
patient064 (348)
patient065 (126)
patient066 (118)
patient067 (193)
patient068 (234)
patient069 (99)
patient070 (91)
patient071 (62)
patient072 (79)
patient073 (514)
patient074 (154)
patient075 (204)
patient076 (151)
patient077 (192)
patient078 (109)
patient079 (87)
patient080 (114)
Proband X1 (96)
Proband X10 (87)
Proband X11 (157)
Proband X12 (92)
Proband X13 (76)
Proband X14 (130)
Proband X15 (121)
Proband X2 (82)
Proband X4 (167)
Proband X5 (150)
Proband X6 (122)
Proband X7 (124)
Proband X8 (84)
Proband X9 (121)
sim (464)
5.0
2.5
0.0
−2.5
wavelet−HLL_glszm_GrayLevelNonUniformity
frame

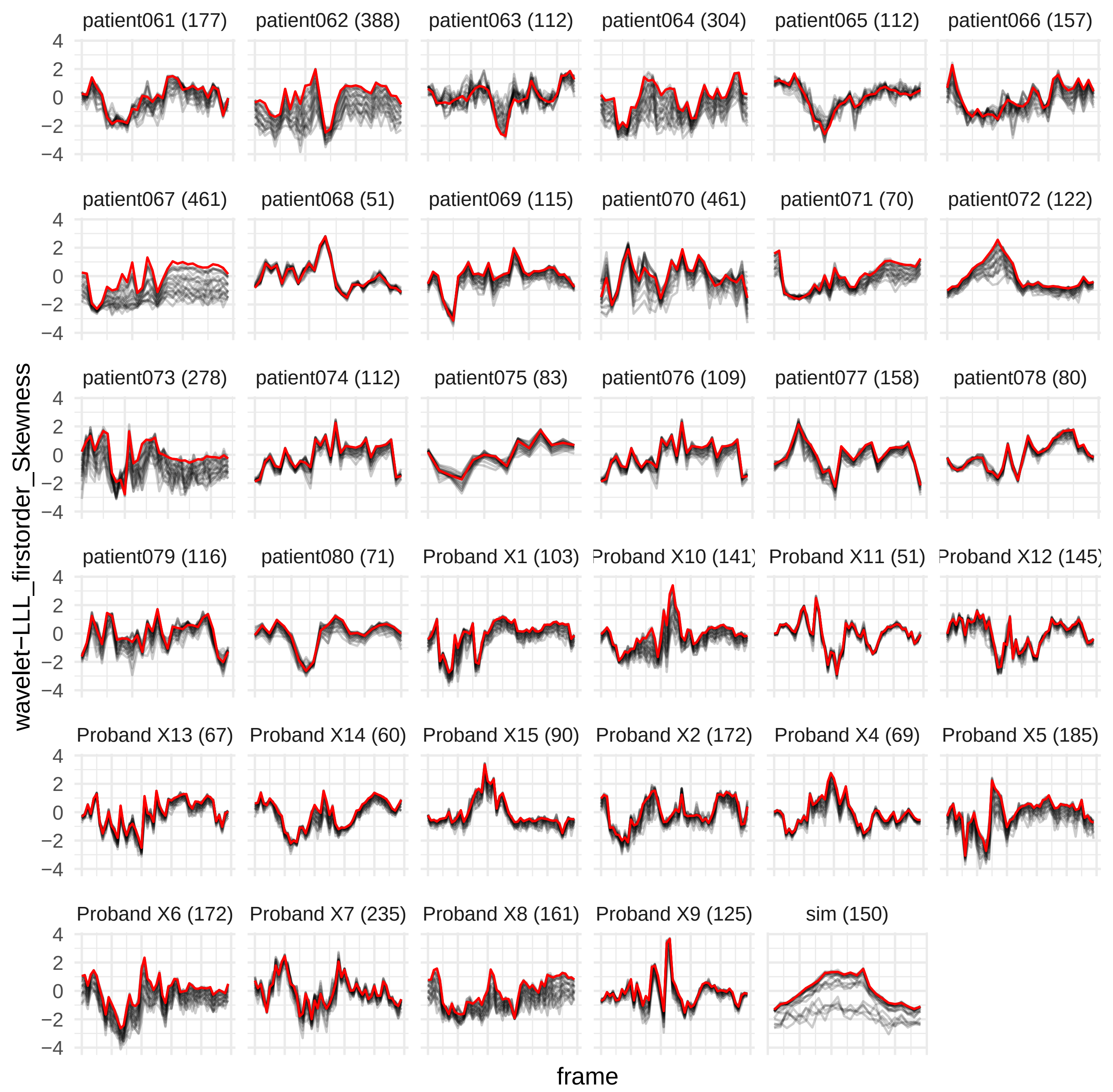

patient061 (177)
patient062 (388)
patient063 (112)
patient064 (304)
patient065 (112)
patient066 (157)
patient067 (461)
patient068 (51)
patient069 (115)
patient070 (461)
patient071 (70)
patient072 (122)
patient073 (278)
patient074 (112)
patient075 (83)
patient076 (109)
patient077 (158)
patient078 (80)
patient079 (116)
patient080 (71)
Proband X1 (103)
Proband X10 (141)
Proband X11 (51)
Proband X12 (145)
Proband X13 (67)
Proband X14 (60)
Proband X15 (90)
Proband X2 (172)
Proband X4 (69)
Proband X5 (185)
Proband X6 (172)
Proband X7 (235)
Proband X8 (161)
Proband X9 (125)
sim (150)
wavelet−LLL_firstorder_Skewness
frame

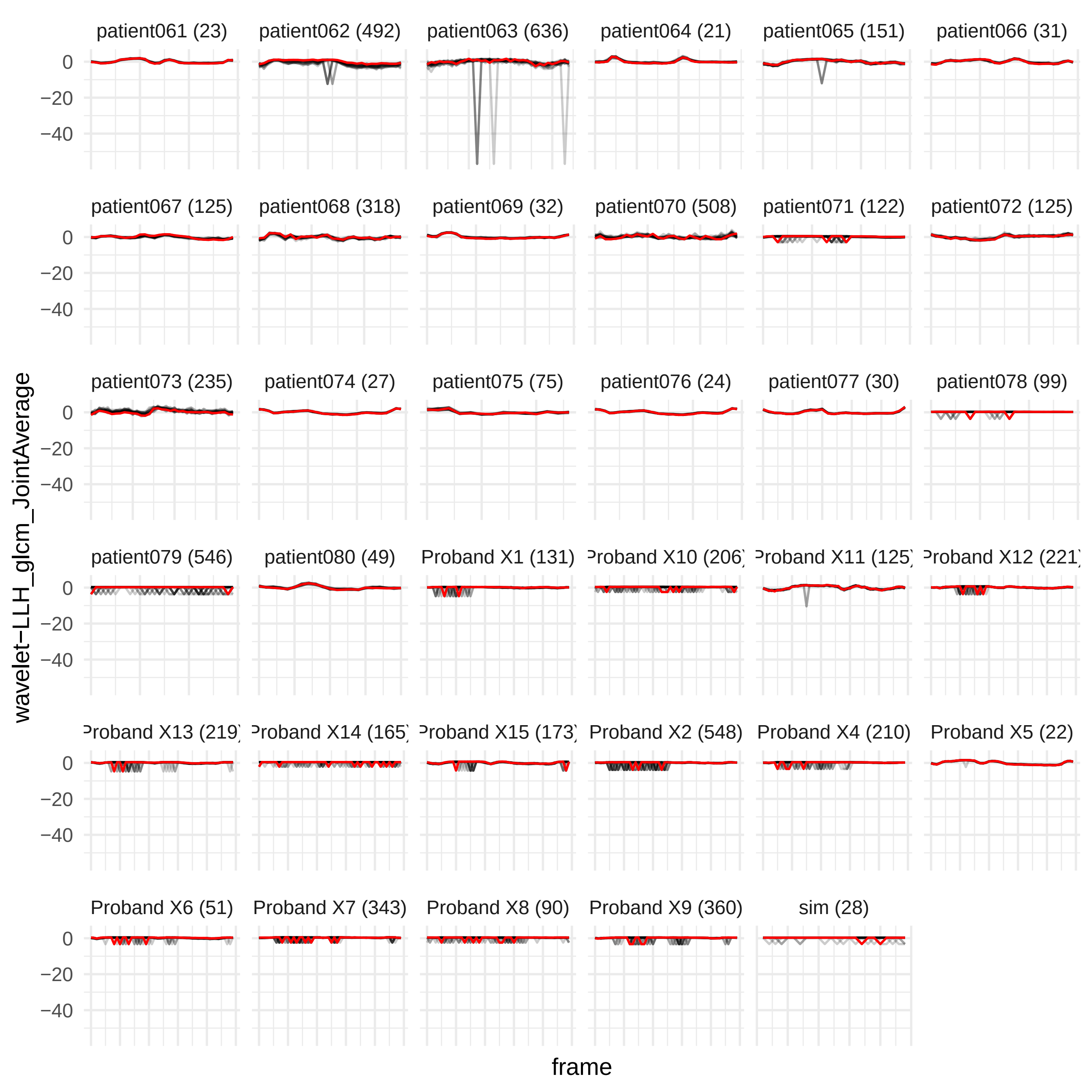
patient061 (23)
patient062 (492)
patient063 (636)
patient064 (21)
patient065 (151)
patient066 (31)
patient067 (125)
patient068 (318)
patient069 (32)
patient070 (508)
patient071 (122)
patient072 (125)
patient073 (235)
patient074 (27)
patient075 (75)
patient076 (24)
patient077 (30)
patient078 (99)
patient079 (546)
patient080 (49)
Proband X1 (131)
Proband X10 (206)
Proband X11 (125)
Proband X12 (221)
Proband X13 (219)
Proband X14 (165)
Proband X15 (173)
Proband X2 (548)
Proband X4 (210)
Proband X5 (22)
Proband X6 (51)
Proband X7 (343)
Proband X8 (90)
Proband X9 (360)
sim (28)
0
−20
−40
wavelet−LLH_glcm_JointAverage
frame

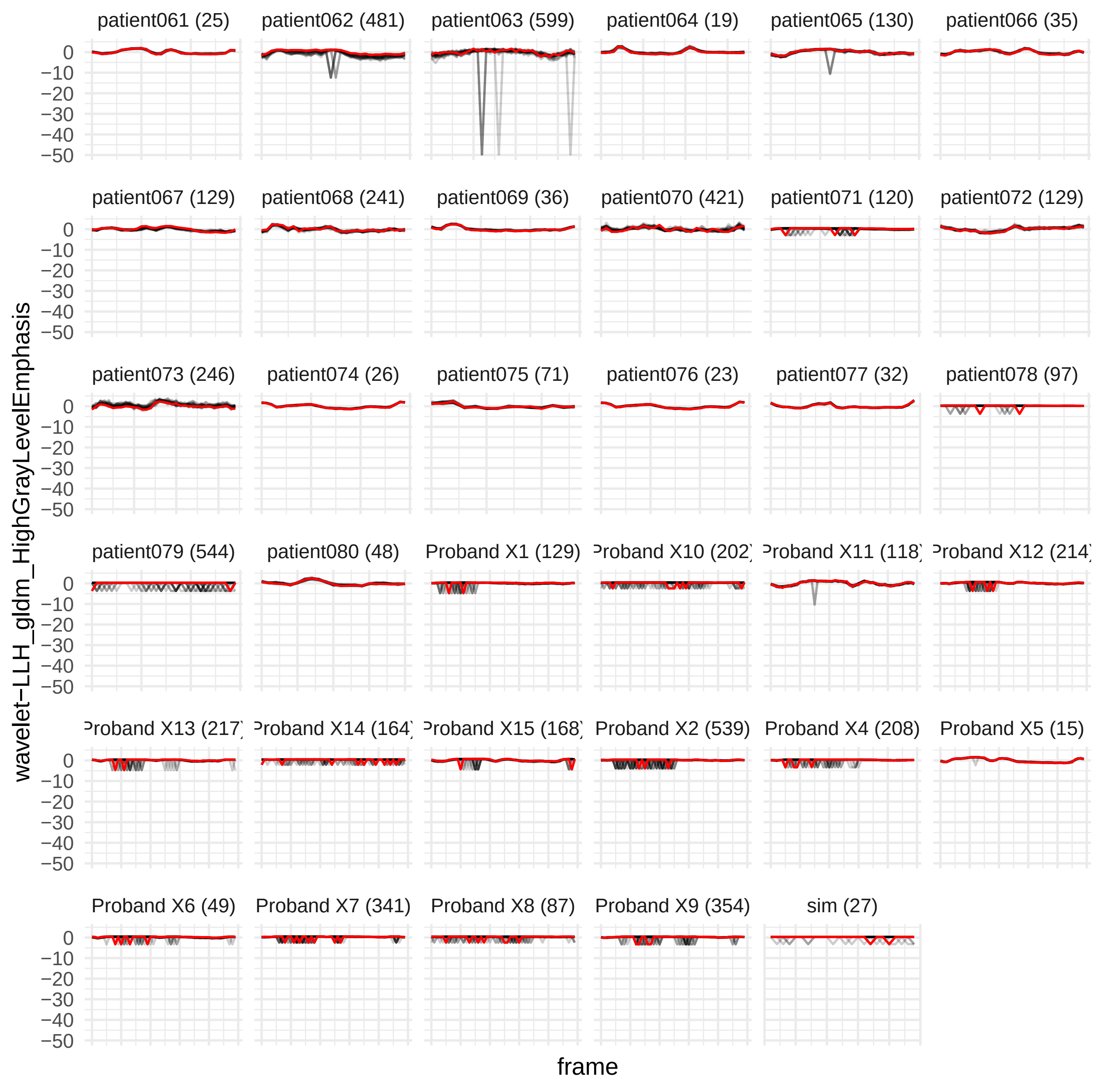

patient061 (25)
patient062 (481)
patient063 (599)
patient064 (19)
patient065 (130)
patient066 (35)
patient067 (129)
patient068 (241)
patient069 (36)
patient070 (421)
patient071 (120)
patient072 (129)
patient073 (246)
patient074 (26)
patient075 (71)
patient076 (23)
patient077 (32)
patient078 (97)
patient079 (544)
patient080 (48)
Proband X1 (129)
Proband X10 (202)
Proband X11 (118)
Proband X12 (214)
Proband X13 (217)
Proband X14 (164)
Proband X15 (168)
Proband X2 (539)
Proband X4 (208)
Proband X5 (15)
Proband X6 (49)
Proband X7 (341)
Proband X8 (87)
Proband X9 (354)
sim (27)
0
−10
−20
−30
−40
−50
wavelet−LLH_gldm_HighGrayLevelEmphasis
frame

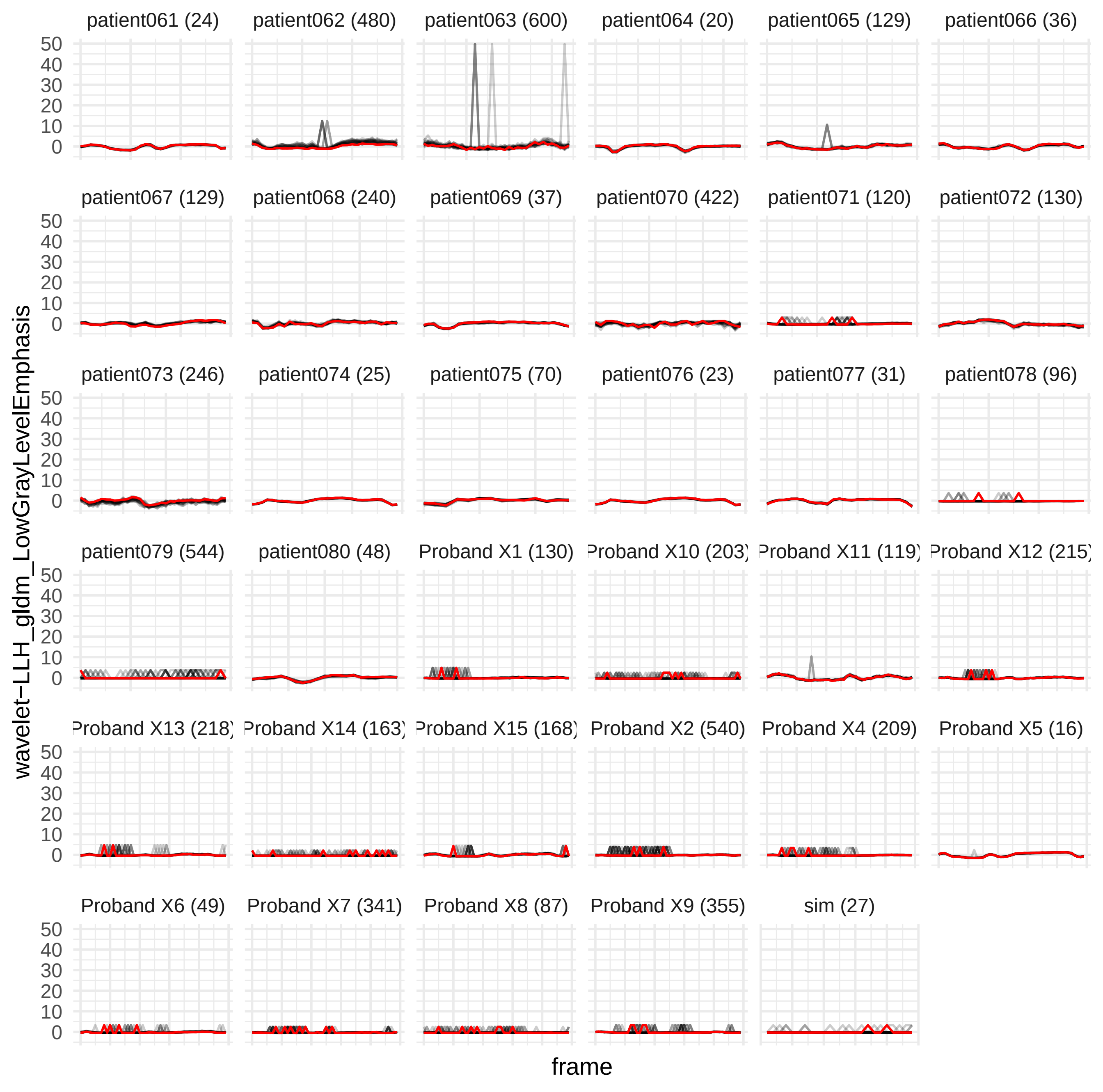

patient061 (24)
patient062 (480)
patient063 (600)
patient064 (20)
patient065 (129)
patient066 (36)
patient067 (129)
patient068 (240)
patient069 (37)
patient070 (422)
patient071 (120)
patient072 (130)
patient073 (246)
patient074 (25)
patient075 (70)
patient076 (23)
patient077 (31)
patient078 (96)
patient079 (544)
patient080 (48)
Proband X1 (130)
Proband X10 (203)
Proband X11 (119)
Proband X12 (215)
Proband X13 (218)
Proband X14 (163)
Proband X15 (168)
Proband X2 (540)
Proband X4 (209)
Proband X5 (16)
Proband X6 (49)
Proband X7 (341)
Proband X8 (87)
Proband X9 (355)
sim (27)
wavelet−LLH_gldm_LowGrayLevelEmphasis
frame

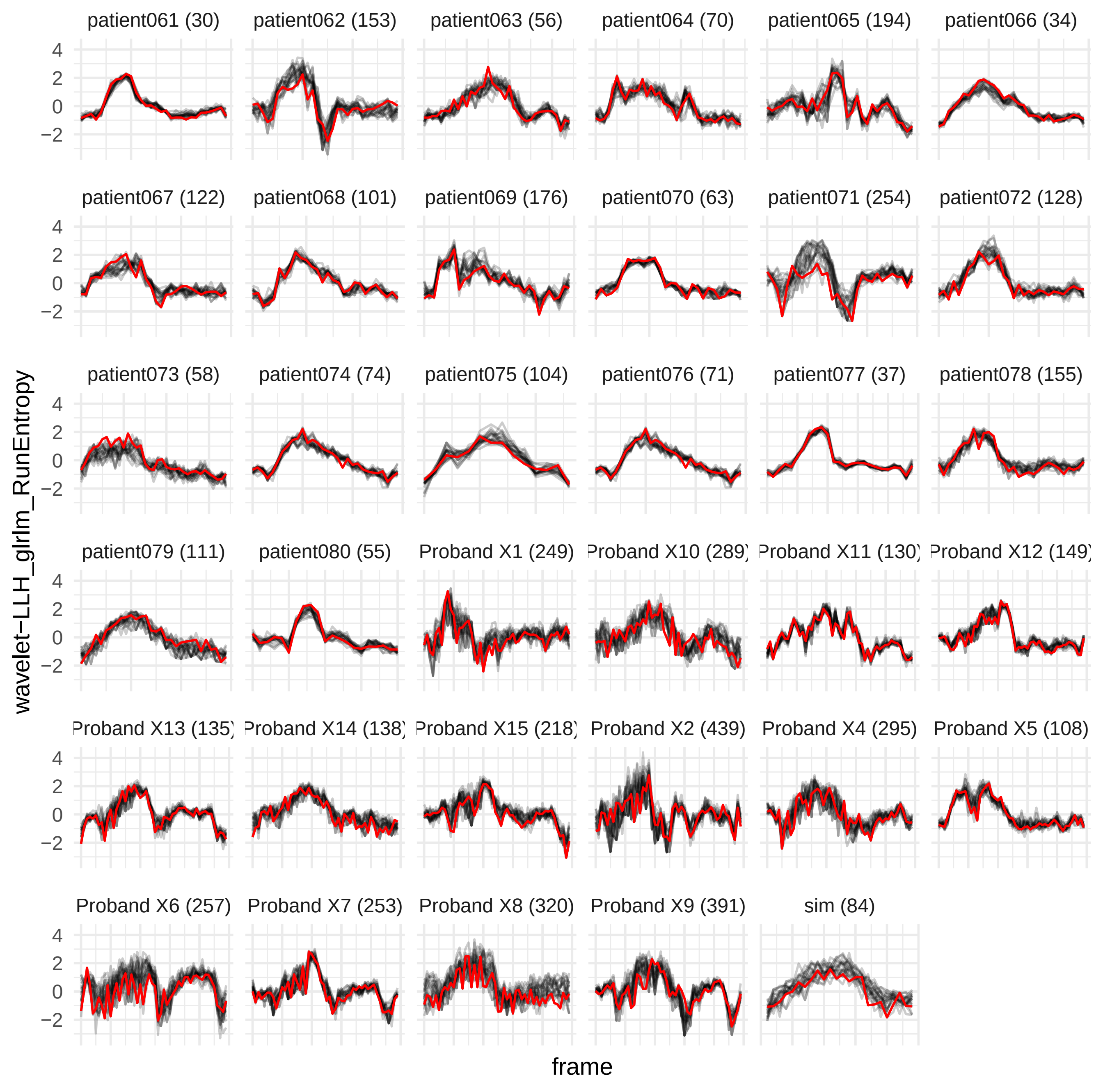

patient061 (30)
patient062 (153)
patient063 (56)
patient064 (70)
patient065 (194)
patient066 (34)
patient067 (122)
patient068 (101)
patient069 (176)
patient070 (63)
patient071 (254)
patient072 (128)
patient073 (58)
patient074 (74)
patient075 (104)
patient076 (71)
patient077 (37)
patient078 (155)
patient079 (111)
patient080 (55)
Proband X1 (249)
Proband X10 (289)
Proband X11 (130)
Proband X12 (149)
Proband X13 (135)
Proband X14 (138)
Proband X15 (218)
Proband X2 (439)
Proband X4 (295)
Proband X5 (108)
Proband X6 (257)
Proband X7 (253)
Proband X8 (320)
Proband X9 (391)
sim (84)
wavelet−LLH_glrlm_RunEntropy
frame
4
2
0
−2

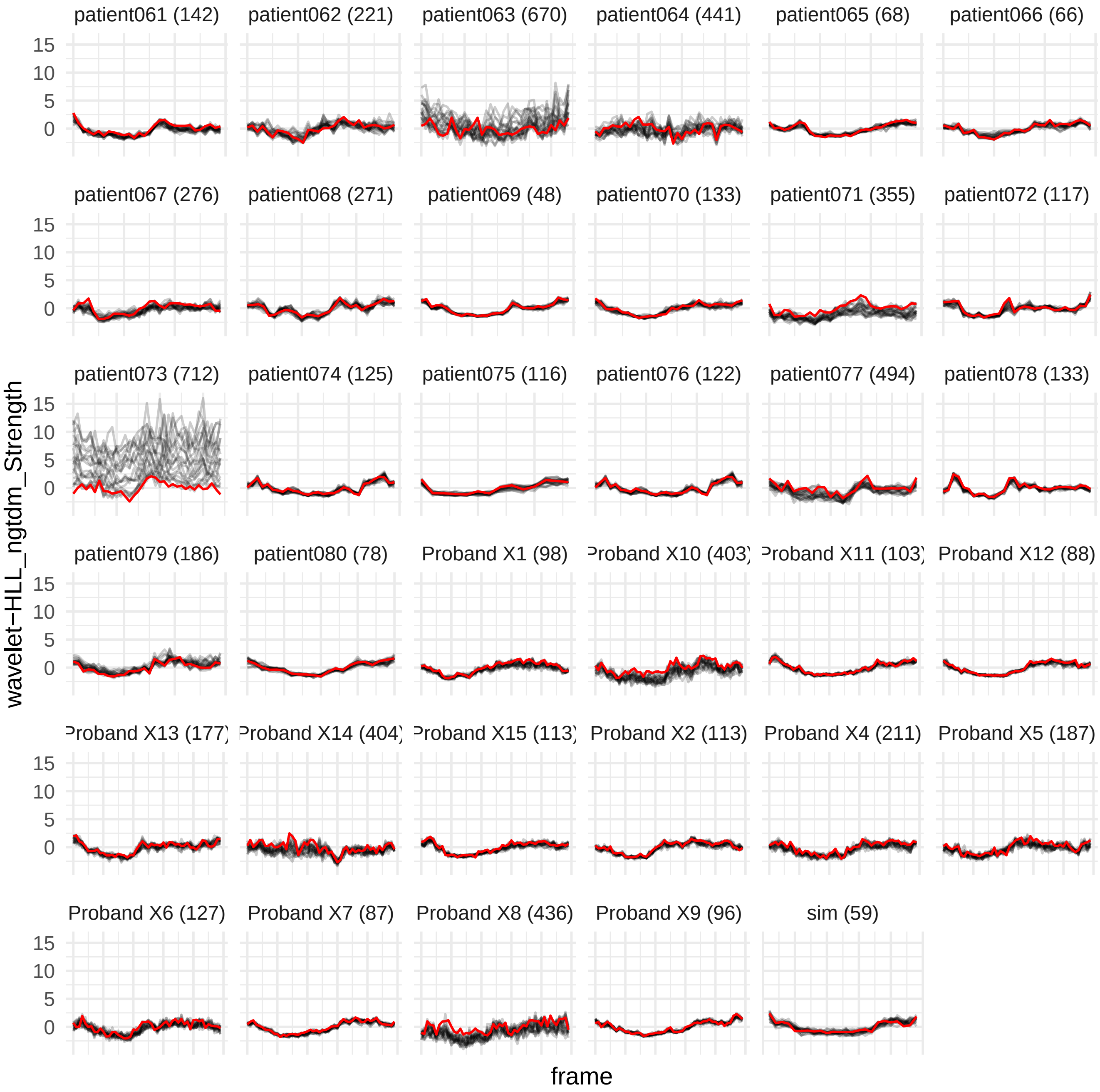

patient061 (142)
patient062 (221)
patient063 (670)
patient064 (441)
patient065 (68)
patient066 (66)
patient067 (276)
patient068 (271)
patient069 (48)
patient070 (133)
patient071 (355)
patient072 (117)
patient073 (712)
patient074 (125)
patient075 (116)
patient076 (122)
patient077 (494)
patient078 (133)
patient079 (186)
patient080 (78)
Proband X1 (98)
Proband X10 (403)
Proband X11 (103)
Proband X12 (88)
Proband X13 (177)
Proband X14 (404)
Proband X15 (113)
Proband X2 (113)
Proband X4 (211)
Proband X5 (187)
Proband X6 (127)
Proband X7 (87)
Proband X8 (436)
Proband X9 (96)
sim (59)
15
10
5
0
wavelet−HLL_ngtdm_Strength
frame

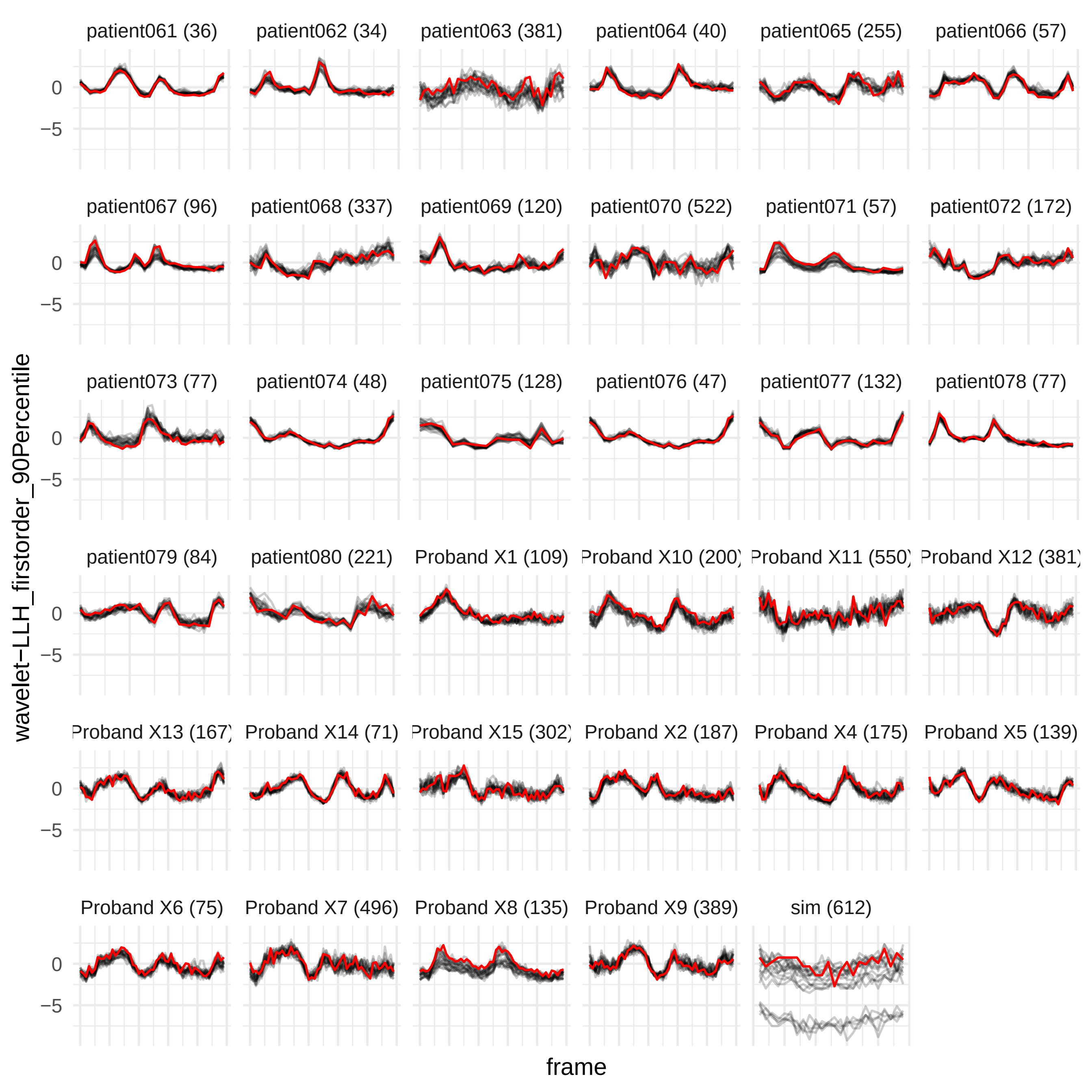

patient061 (36)
patient062 (34)
patient063 (381)
patient064 (40)
patient065 (255)
patient066 (57)
patient067 (96)
patient068 (337)
patient069 (120)
patient070 (522)
patient071 (57)
patient072 (172)
patient073 (77)
patient074 (48)
patient075 (128)
patient076 (47)
patient077 (132)
patient078 (77)
patient079 (84)
patient080 (221)
Proband X1 (109)
Proband X10 (200)
Proband X11 (550)
Proband X12 (381)
Proband X13 (167)
Proband X14 (71)
Proband X15 (302)
Proband X2 (187)
Proband X4 (175)
Proband X5 (139)
Proband X6 (75)
Proband X7 (496)
Proband X8 (135)
Proband X9 (389)
sim (612)
0
−5
wavelet−LLH_firstorder_90Percentile
frame

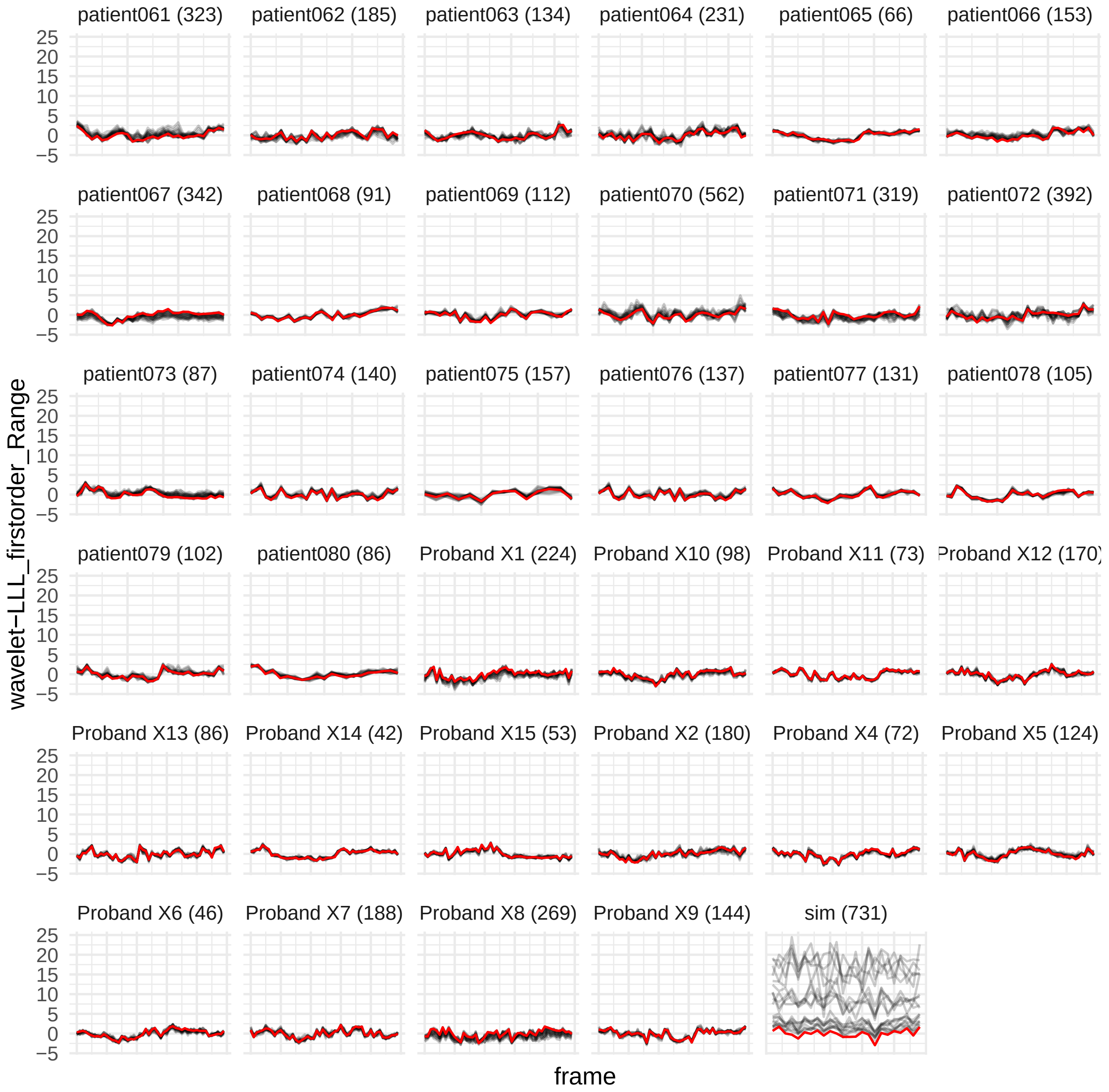

patient061 (323)
patient062 (185)
patient063 (134)
patient064 (231)
patient065 (66)
patient066 (153)
patient067 (342)
patient068 (91)
patient069 (112)
patient070 (562)
patient071 (319)
patient072 (392)
patient073 (87)
patient074 (140)
patient075 (157)
patient076 (137)
patient077 (131)
patient078 (105)
patient079 (102)
patient080 (86)
Proband X1 (224)
Proband X10 (98)
Proband X11 (73)
Proband X12 (170)
Proband X13 (86)
Proband X14 (42)
Proband X15 (53)
Proband X2 (180)
Proband X4 (72)
Proband X5 (124)
Proband X6 (46)
Proband X7 (188)
Proband X8 (269)
Proband X9 (144)
sim (731)
25
20
15
10
5
0
−5
wavelet−LLL_firstorder_Range
frame

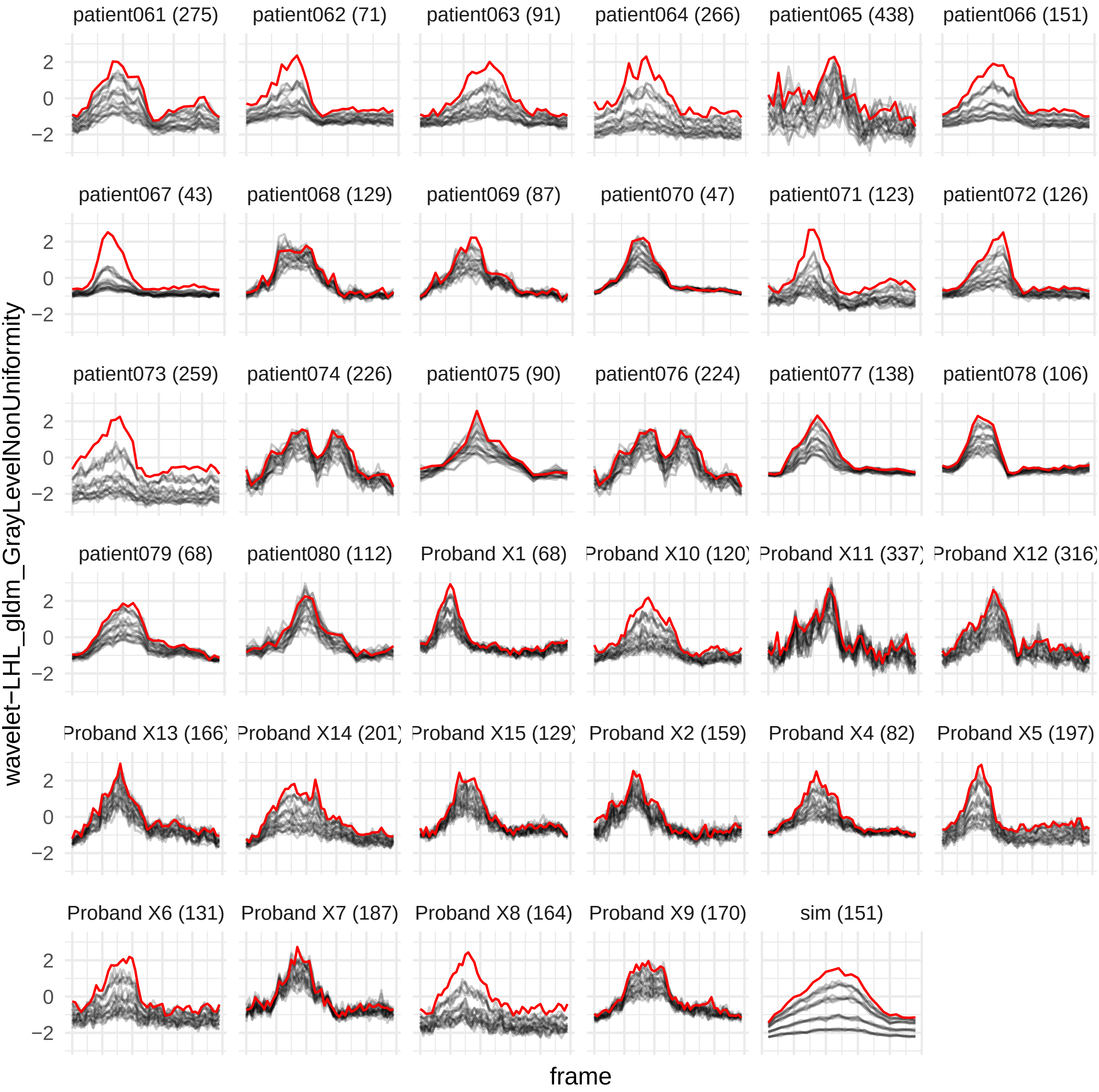
patient061 (275)
patient062 (71)
patient063 (91)
patient064 (266)
patient065 (438)
patient066 (151)
patient067 (43)
patient068 (129)
patient069 (87)
patient070 (47)
patient071 (123)
patient072 (126)
patient073 (259)
patient074 (226)
patient075 (90)
patient076 (224)
patient077 (138)
patient078 (106)
patient079 (68)
patient080 (112)
Proband X1 (68)
Proband X10 (120)
Proband X11 (337)
Proband X12 (316)
Proband X13 (166)
Proband X14 (201)
Proband X15 (129)
Proband X2 (159)
Proband X4 (82)
Proband X5 (197)
Proband X6 (131)
Proband X7 (187)
Proband X8 (164)
Proband X9 (170)
sim (151)
wavelet−LHL_gldm_GrayLevelNonUniformity
frame
2
0
−2

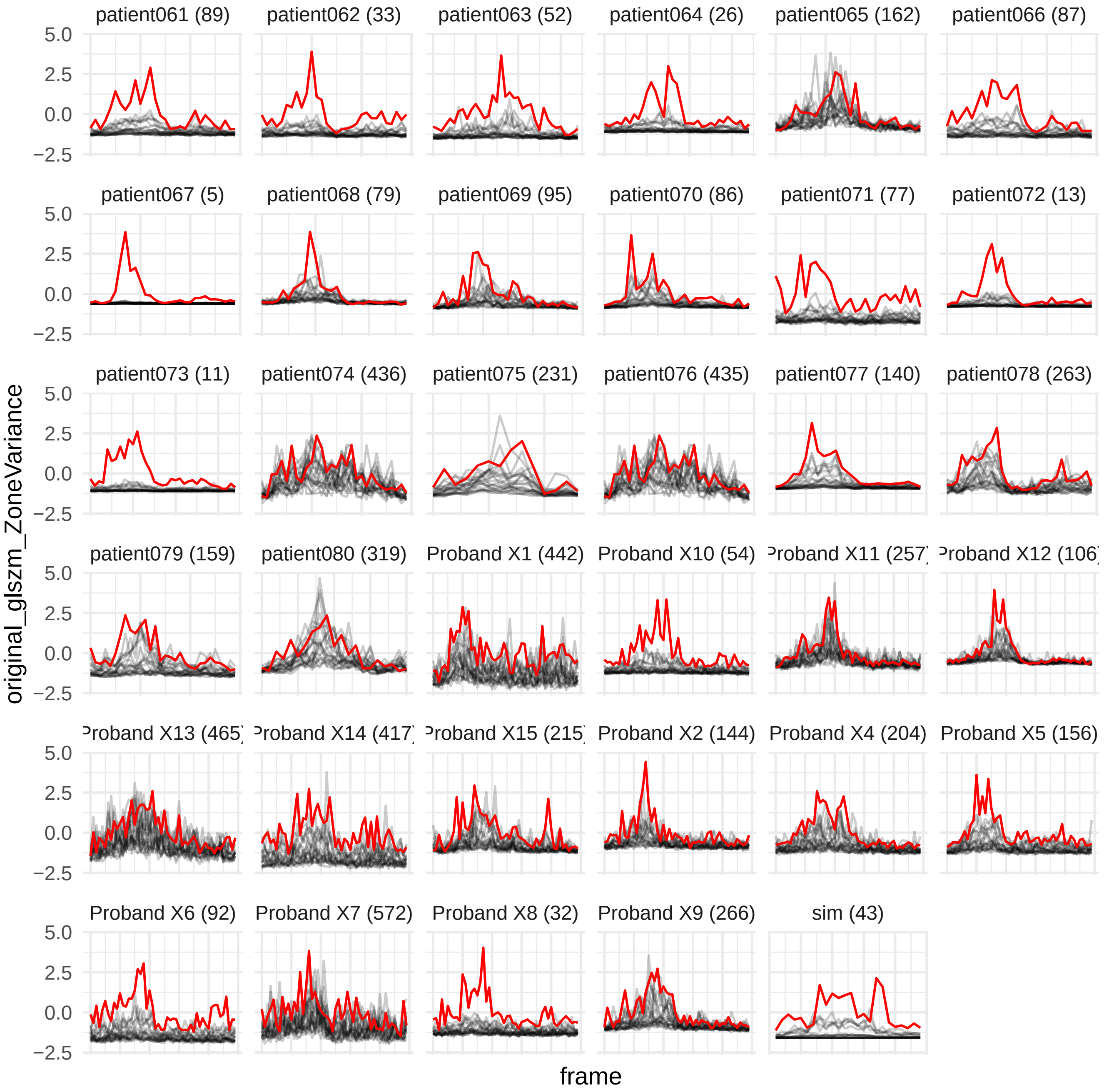
patient061 (89)
patient062 (33)
patient063 (52)
patient064 (26)
patient065 (162)
patient066 (87)
patient067 (5)
patient068 (79)
patient069 (95)
patient070 (86)
patient071 (77)
patient072 (13)
patient073 (11)
patient074 (436)
patient075 (231)
patient076 (435)
patient077 (140)
patient078 (263)
patient079 (159)
patient080 (319)
Proband X1 (442)
Proband X10 (54)
Proband X11 (257)
Proband X12 (106)
Proband X13 (465)
Proband X14 (417)
Proband X15 (215)
Proband X2 (144)
Proband X4 (204)
Proband X5 (156)
Proband X6 (92)
Proband X7 (572)
Proband X8 (32)
Proband X9 (266)
sim (43)
5.0
2.5
0.0
−2.5
original_glszm_ZoneVariance
frame

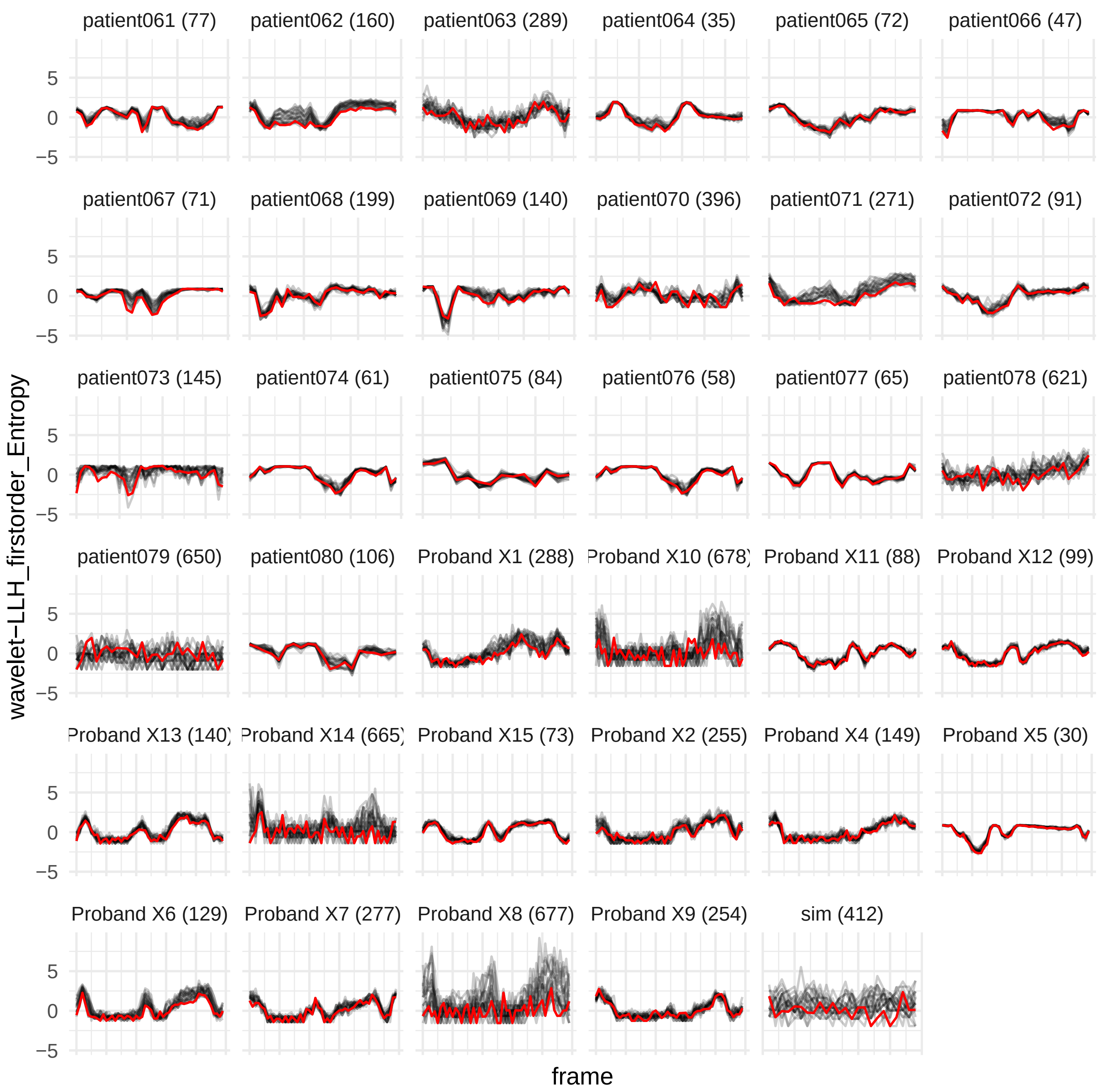

patient061 (77)
patient062 (160)
patient063 (289)
patient064 (35)
patient065 (72)
patient066 (47)
patient067 (71)
patient068 (199)
patient069 (140)
patient070 (396)
patient071 (271)
patient072 (91)
patient073 (145)
patient074 (61)
patient075 (84)
patient076 (58)
patient077 (65)
patient078 (621)
patient079 (650)
patient080 (106)
Proband X1 (288)
Proband X10 (678)
Proband X11 (88)
Proband X12 (99)
Proband X13 (140)
Proband X14 (665)
Proband X15 (73)
Proband X2 (255)
Proband X4 (149)
Proband X5 (30)
Proband X6 (129)
Proband X7 (277)
Proband X8 (677)
Proband X9 (254)
sim (412)
5
0
−5
wavelet−LLH_firstorder_Entropy
frame

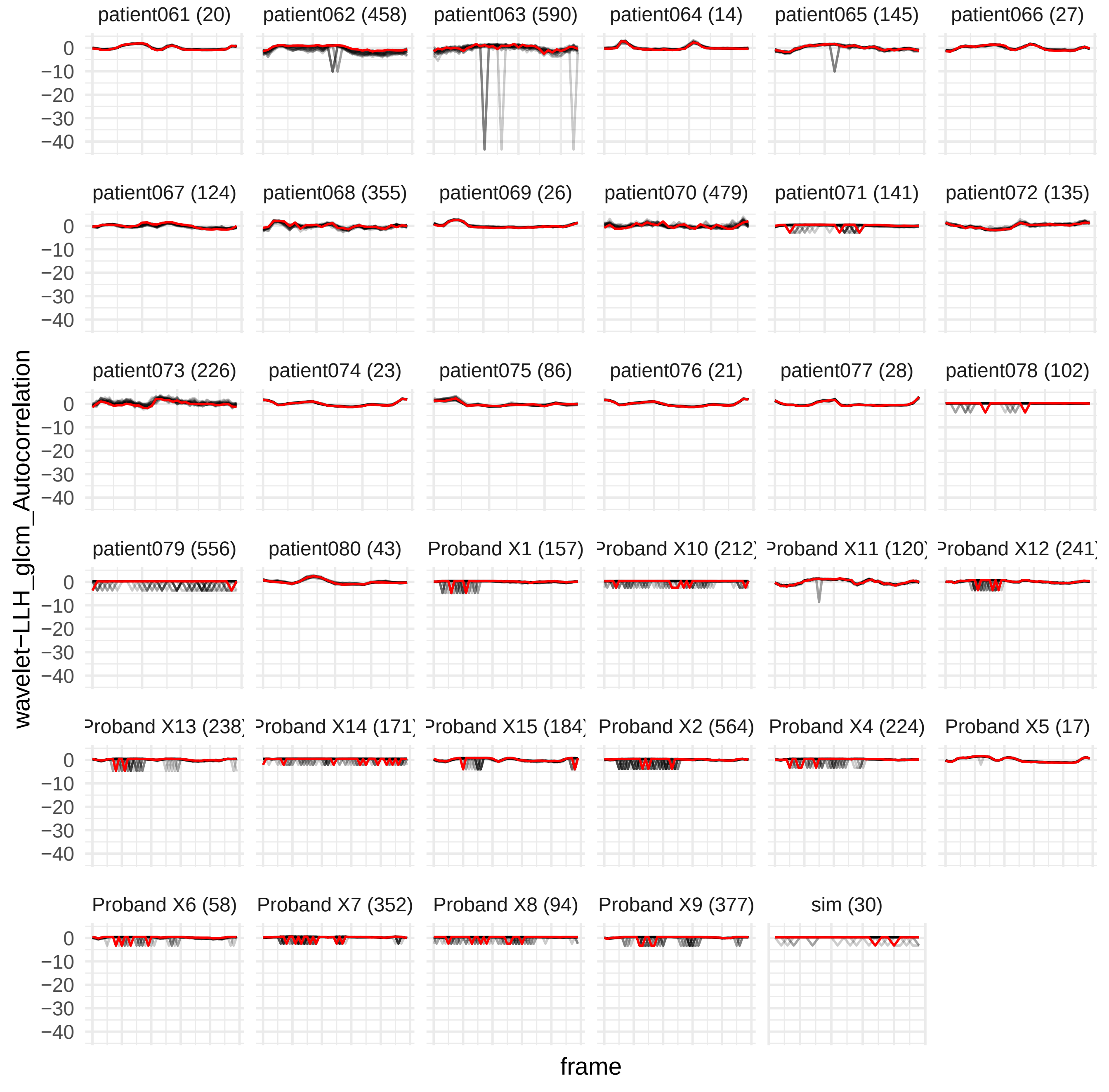

patient061 (20)
patient062 (458)
patient063 (590)
patient064 (14)
patient065 (145)
patient066 (27)
patient067 (124)
patient068 (355)
patient069 (26)
patient070 (479)
patient071 (141)
patient072 (135)
patient073 (226)
patient074 (23)
patient075 (86)
patient076 (21)
patient077 (28)
patient078 (102)
patient079 (556)
patient080 (43)
Proband X1 (157)
Proband X10 (212)
Proband X11 (120)
Proband X12 (241)
Proband X13 (238)
Proband X14 (171)
Proband X15 (184)
Proband X2 (564)
Proband X4 (224)
Proband X5 (17)
Proband X6 (58)
Proband X7 (352)
Proband X8 (94)
Proband X9 (377)
sim (30)
0
−10
−20
−30
−40
wavelet−LLH_glcm_Autocorrelation
frame

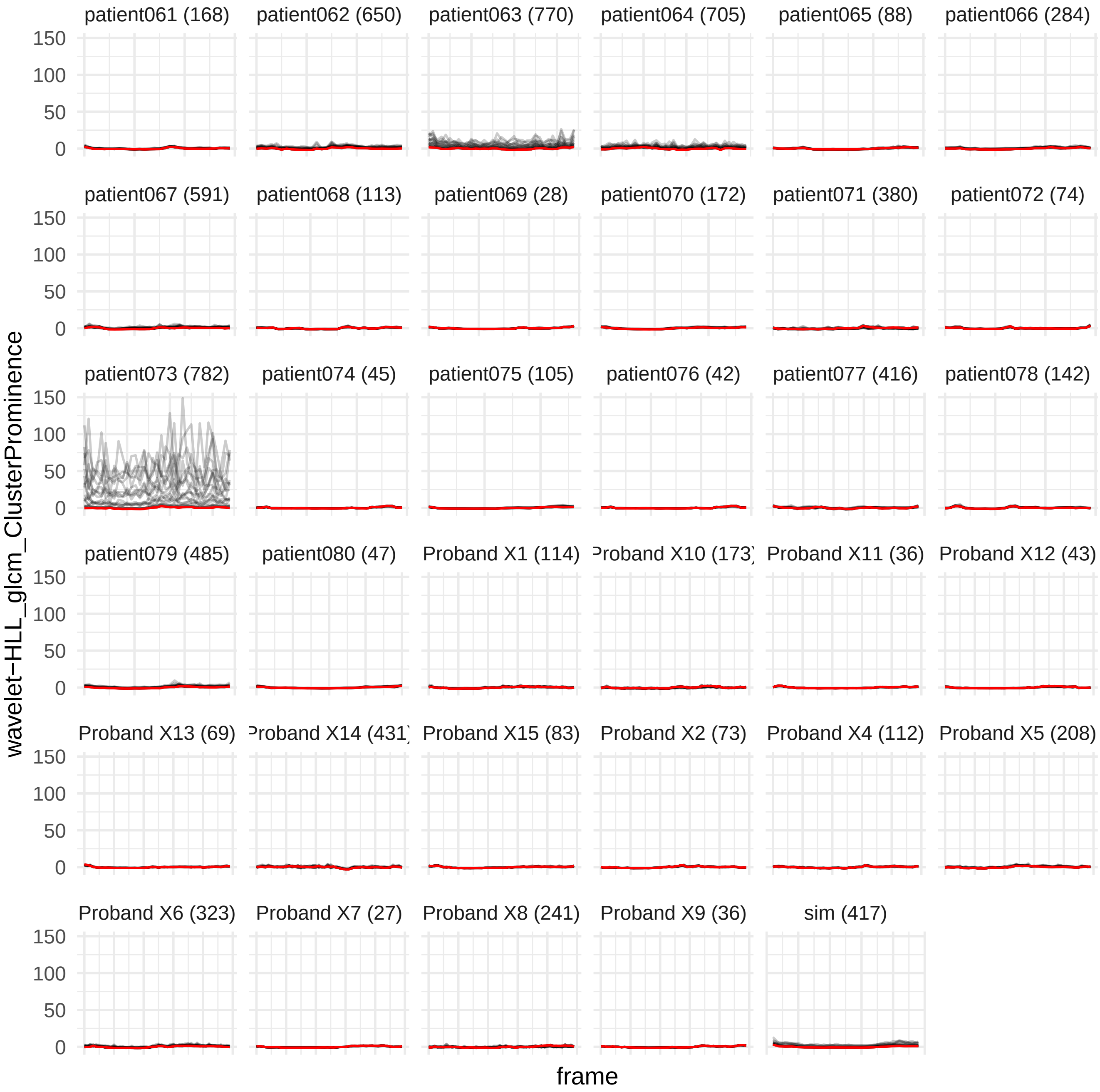

patient061 (168)
patient062 (650)
patient063 (770)
patient064 (705)
patient065 (88)
patient066 (284)
patient067 (591)
patient068 (113)
patient069 (28)
patient070 (172)
patient071 (380)
patient072 (74)
patient073 (782)
patient074 (45)
patient075 (105)
patient076 (42)
patient077 (416)
patient078 (142)
patient079 (485)
patient080 (47)
Proband X1 (114)
Proband X10 (173)
Proband X11 (36)
Proband X12 (43)
Proband X13 (69)
Proband X14 (431)
Proband X15 (83)
Proband X2 (73)
Proband X4 (112)
Proband X5 (208)
Proband X6 (323)
Proband X7 (27)
Proband X8 (241)
Proband X9 (36)
sim (417)
150
100
50
0
wavelet−HLL_glcm_ClusterProminence
frame

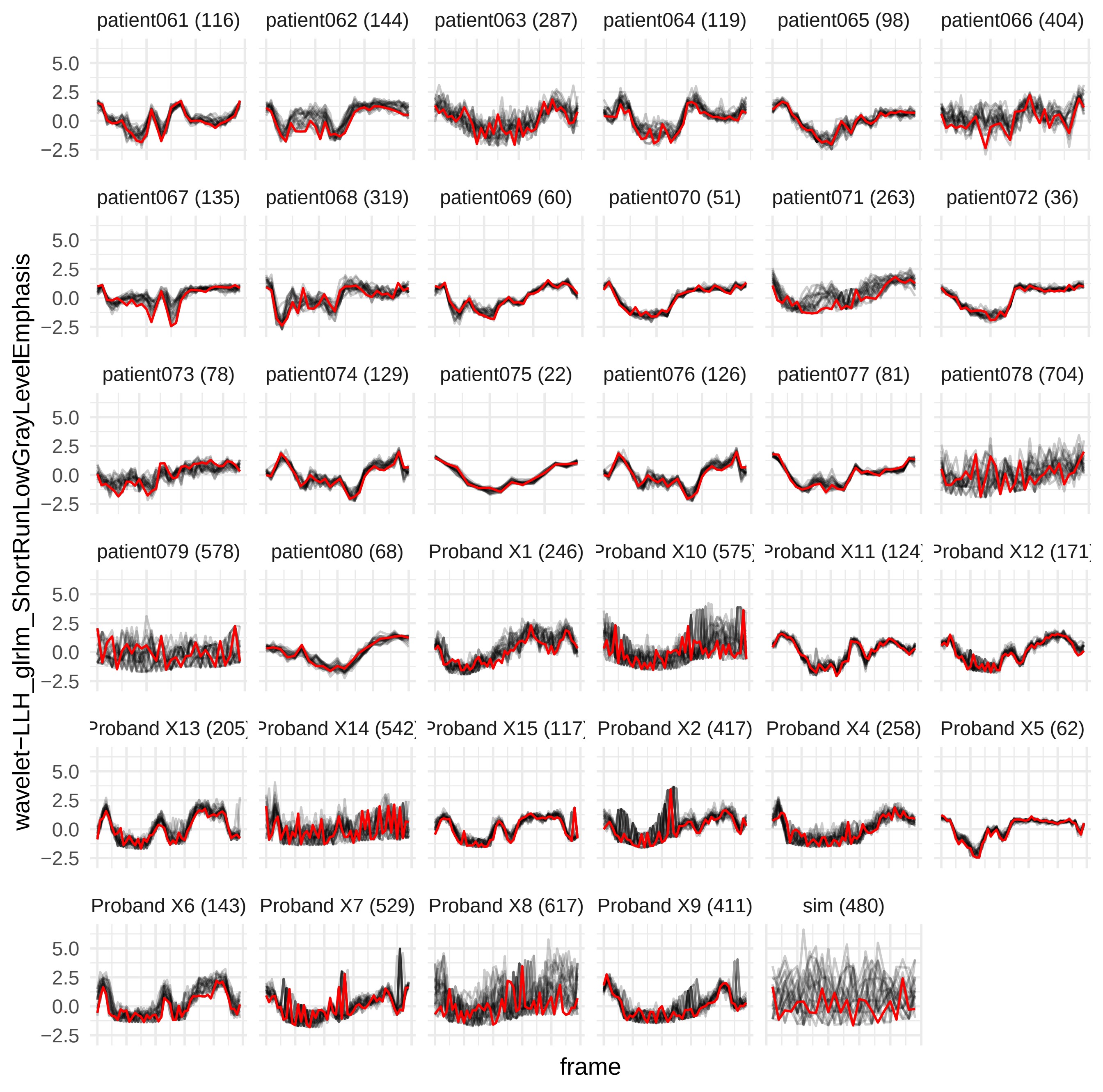

patient061 (116)
patient062 (144)
patient063 (287)
patient064 (119)
patient065 (98)
patient066 (404)
patient067 (135)
patient068 (319)
patient069 (60)
patient070 (51)
patient071 (263)
patient072 (36)
patient073 (78)
patient074 (129)
patient075 (22)
patient076 (126)
patient077 (81)
patient078 (704)
patient079 (578)
patient080 (68)
Proband X1 (246)
Proband X10 (575)
Proband X11 (124)
Proband X12 (171)
Proband X13 (205)
Proband X14 (542)
Proband X15 (117)
Proband X2 (417)
Proband X4 (258)
Proband X5 (62)
Proband X6 (143)
Proband X7 (529)
Proband X8 (617)
Proband X9 (411)
sim (480)
5.0
2.5
0.0
−2.5
wavelet−LLH_glrlm_ShortRunLowGrayLevelEmphasis
frame

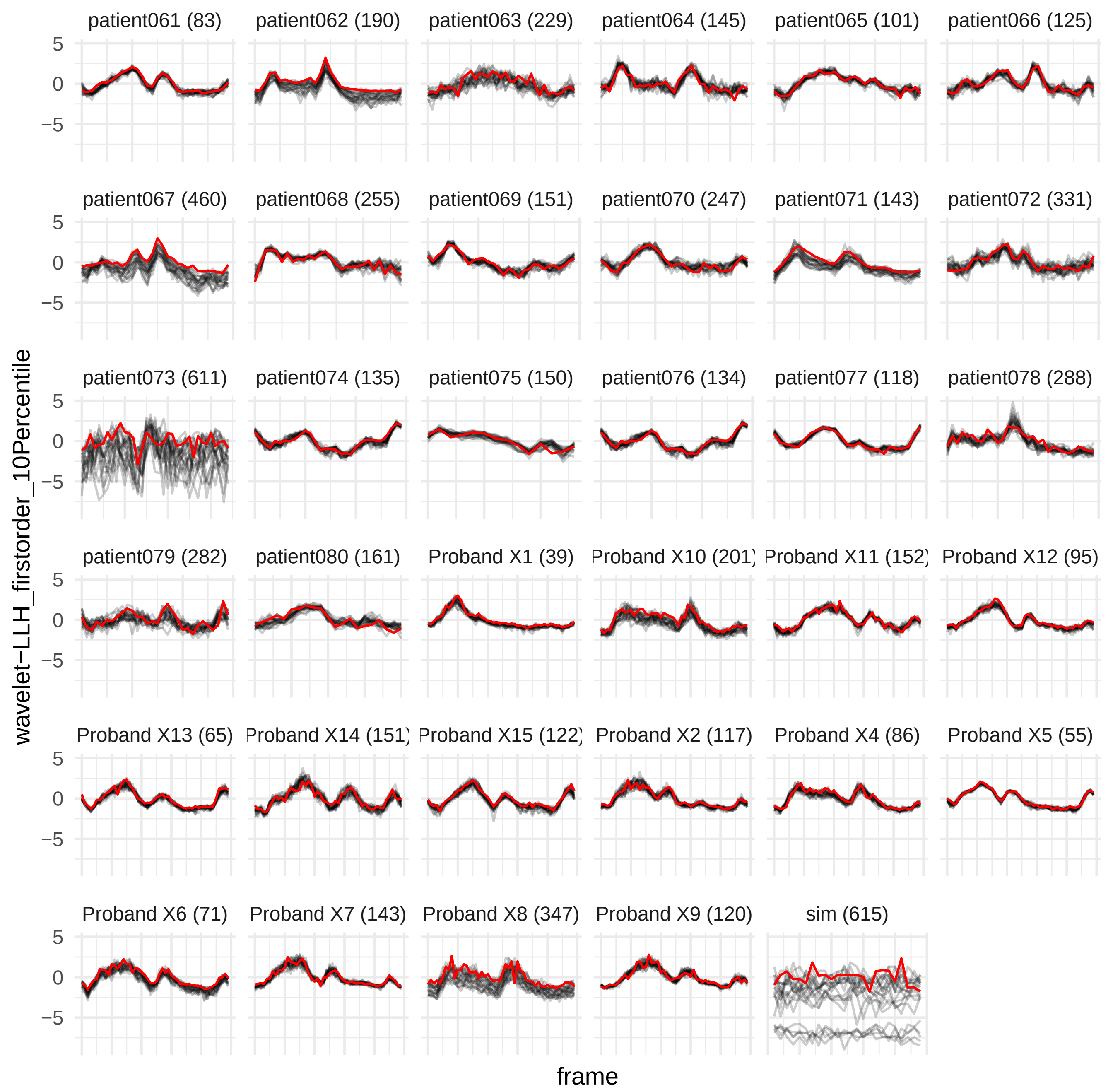
patient061 (83)
patient062 (190)
patient063 (229)
patient064 (145)
patient065 (101)
patient066 (125)
patient067 (460)
patient068 (255)
patient069 (151)
patient070 (247)
patient071 (143)
patient072 (331)
patient073 (611)
patient074 (135)
patient075 (150)
patient076 (134)
patient077 (118)
patient078 (288)
patient079 (282)
patient080 (161)
Proband X1 (39)
Proband X10 (201)
Proband X11 (152)
Proband X12 (95)
Proband X13 (65)
Proband X14 (151)
Proband X15 (122)
Proband X2 (117)
Proband X4 (86)
Proband X5 (55)
Proband X6 (71)
Proband X7 (143)
Proband X8 (347)
Proband X9 (120)
sim (615)
5
0
−5
wavelet−LLH_firstorder_10Percentile
frame

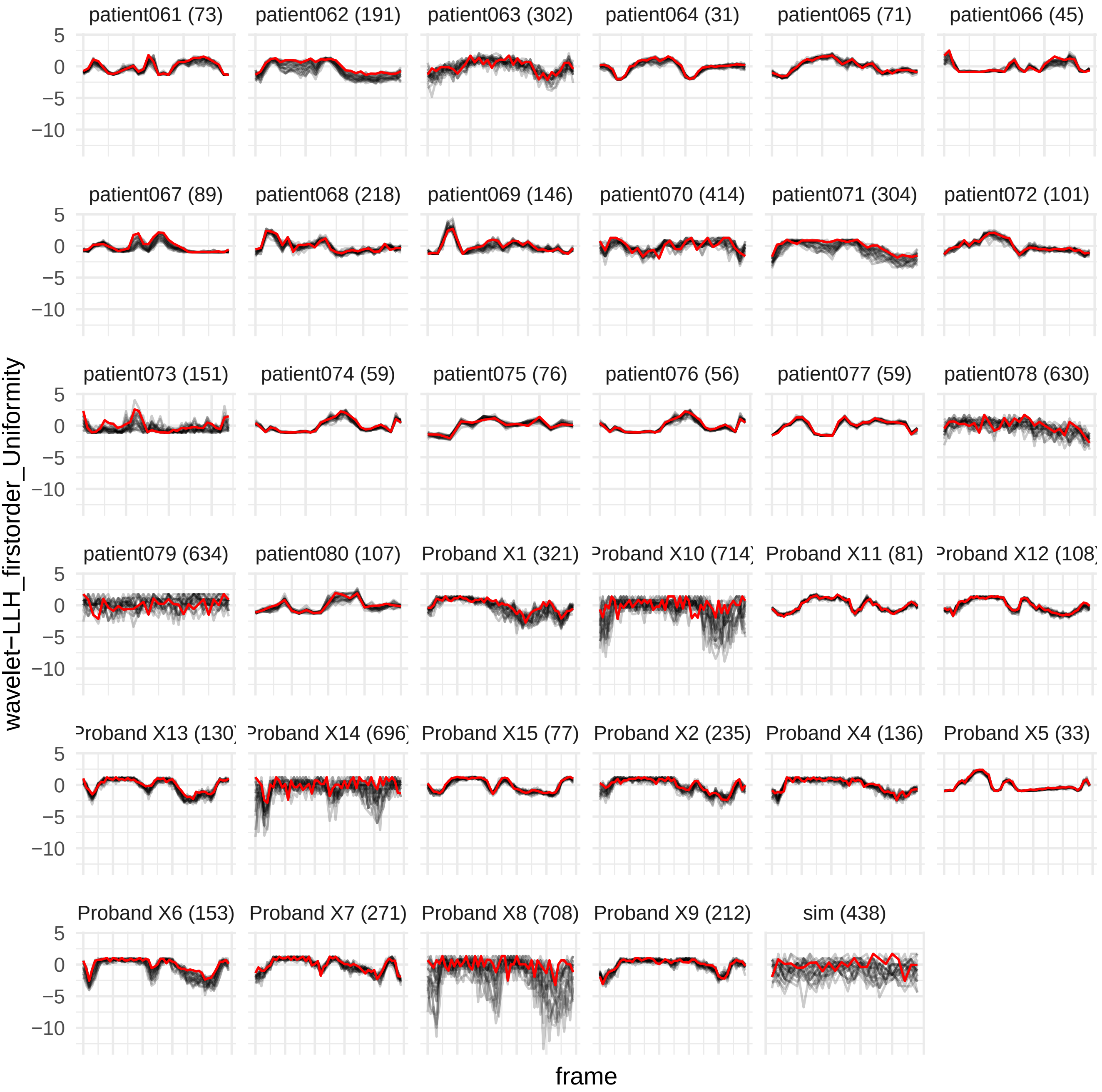

patient061 (73)
patient062 (191)
patient063 (302)
patient064 (31)
patient065 (71)
patient066 (45)
patient067 (89)
patient068 (218)
patient069 (146)
patient070 (414)
patient071 (304)
patient072 (101)
patient073 (151)
patient074 (59)
patient075 (76)
patient076 (56)
patient077 (59)
patient078 (630)
patient079 (634)
patient080 (107)
Proband X1 (321)
Proband X10 (714)
Proband X11 (81)
Proband X12 (108)
Proband X13 (130)
Proband X14 (696)
Proband X15 (77)
Proband X2 (235)
Proband X4 (136)
Proband X5 (33)
Proband X6 (153)
Proband X7 (271)
Proband X8 (708)
Proband X9 (212)
sim (438)
5
0
−5
−10
wavelet−LLH_firstorder_Uniformity
frame

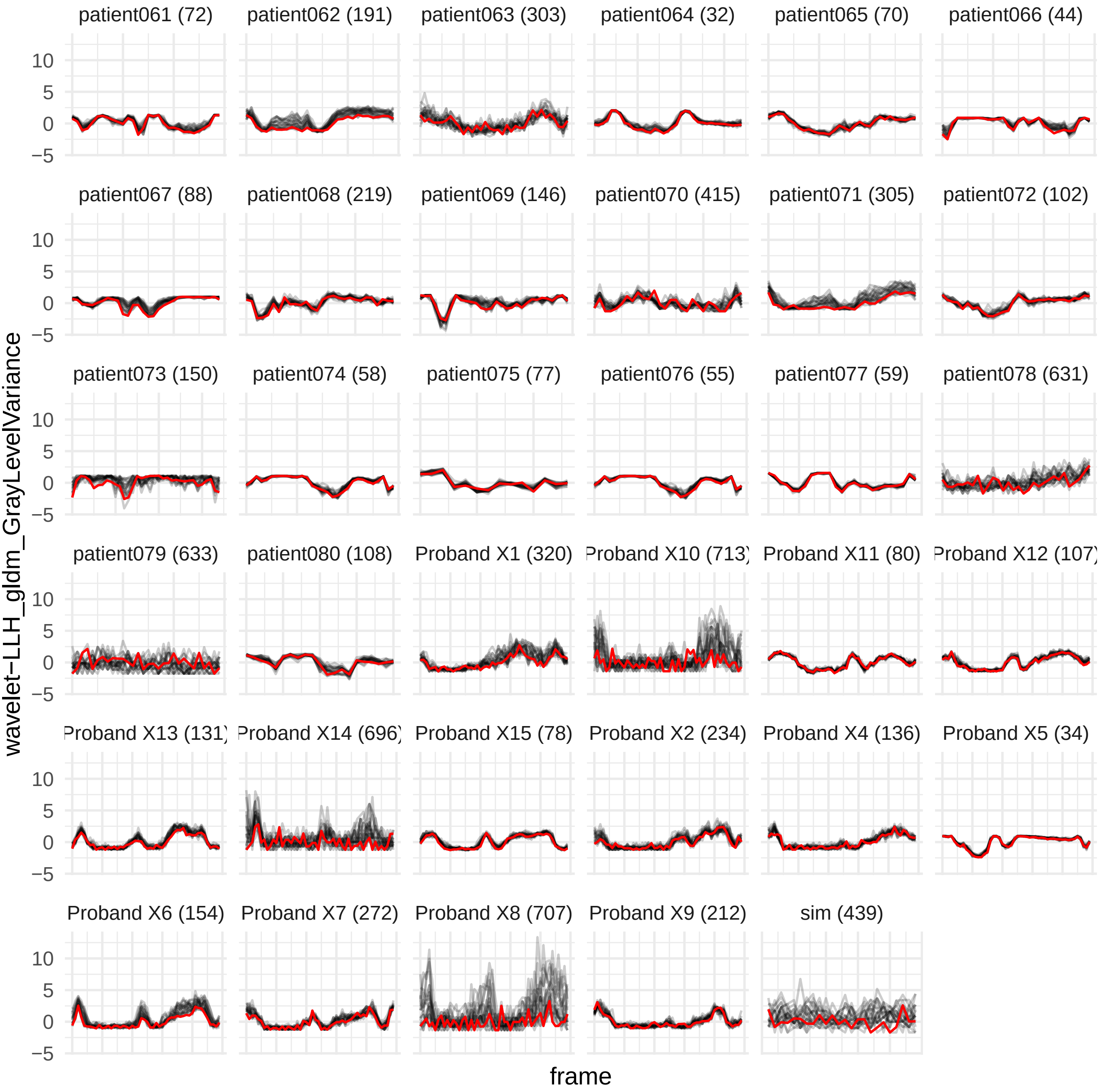
patient061 (72)
patient062 (191)
patient063 (303)
patient064 (32)
patient065 (70)
patient066 (44)
patient067 (88)
patient068 (219)
patient069 (146)
patient070 (415)
patient071 (305)
patient072 (102)
patient073 (150)
patient074 (58)
patient075 (77)
patient076 (55)
patient077 (59)
patient078 (631)
patient079 (633)
patient080 (108)
Proband X1 (320)
Proband X10 (713)
Proband X11 (80)
Proband X12 (107)
Proband X13 (131)
Proband X14 (696)
Proband X15 (78)
Proband X2 (234)
Proband X4 (136)
Proband X5 (34)
Proband X6 (154)
Proband X7 (272)
Proband X8 (707)
Proband X9 (212)
sim (439)
wavelet−LLH_gldm_GrayLevelVariance
frame
10
5
0
−5

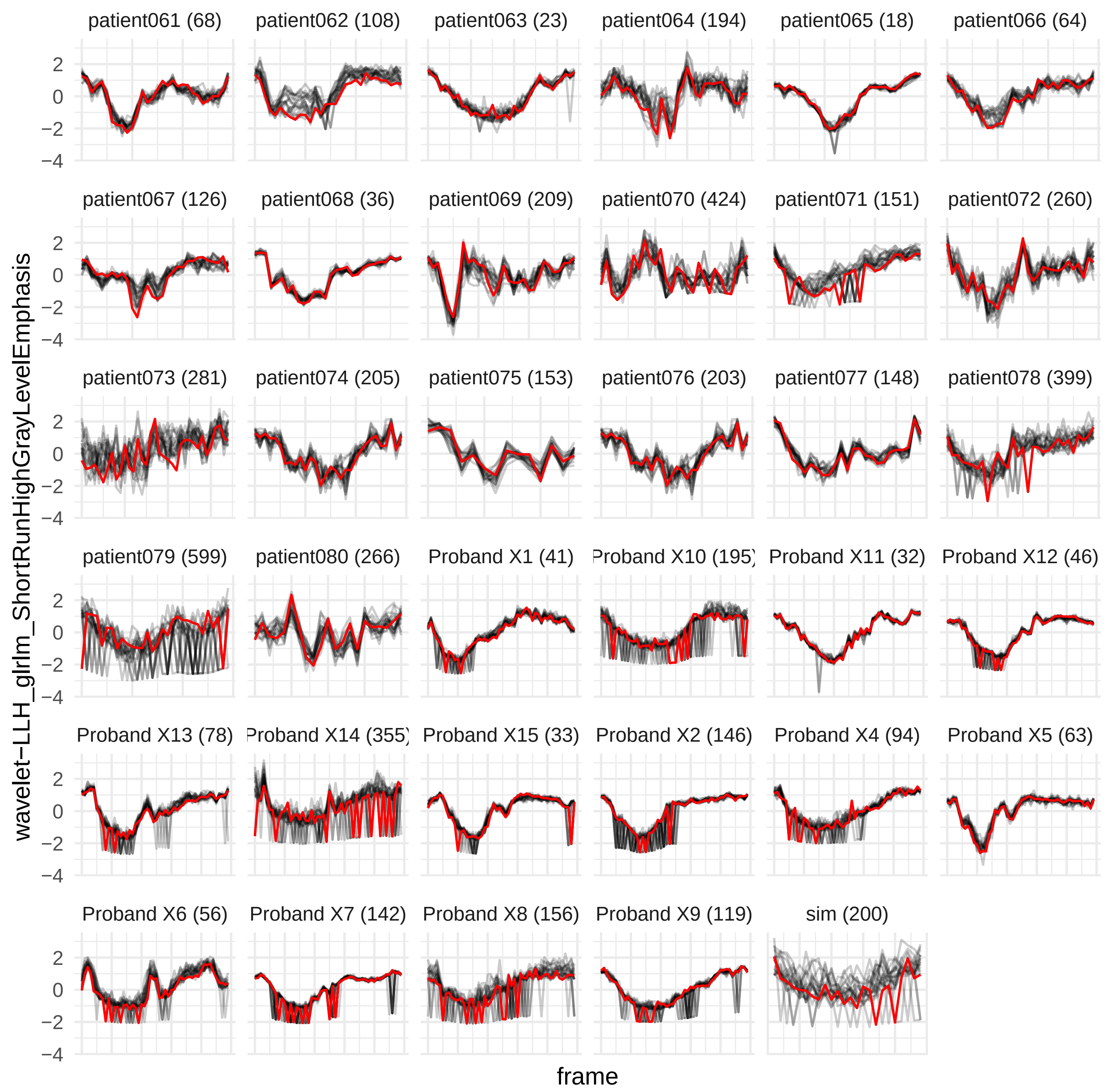

patient061 (68)
patient062 (108)
patient063 (23)
patient064 (194)
patient065 (18)
patient066 (64)
patient067 (126)
patient068 (36)
patient069 (209)
patient070 (424)
patient071 (151)
patient072 (260)
patient073 (281)
patient074 (205)
patient075 (153)
patient076 (203)
patient077 (148)
patient078 (399)
patient079 (599)
patient080 (266)
Proband X1 (41)
Proband X10 (195)
Proband X11 (32)
Proband X12 (46)
Proband X13 (78)
Proband X14 (355)
Proband X15 (33)
Proband X2 (146)
Proband X4 (94)
Proband X5 (63)
Proband X6 (56)
Proband X7 (142)
Proband X8 (156)
Proband X9 (119)
sim (200)
2
0
−2
−4
wavelet−LLH_glrlm_ShortRunHighGrayLevelEmphasis
frame

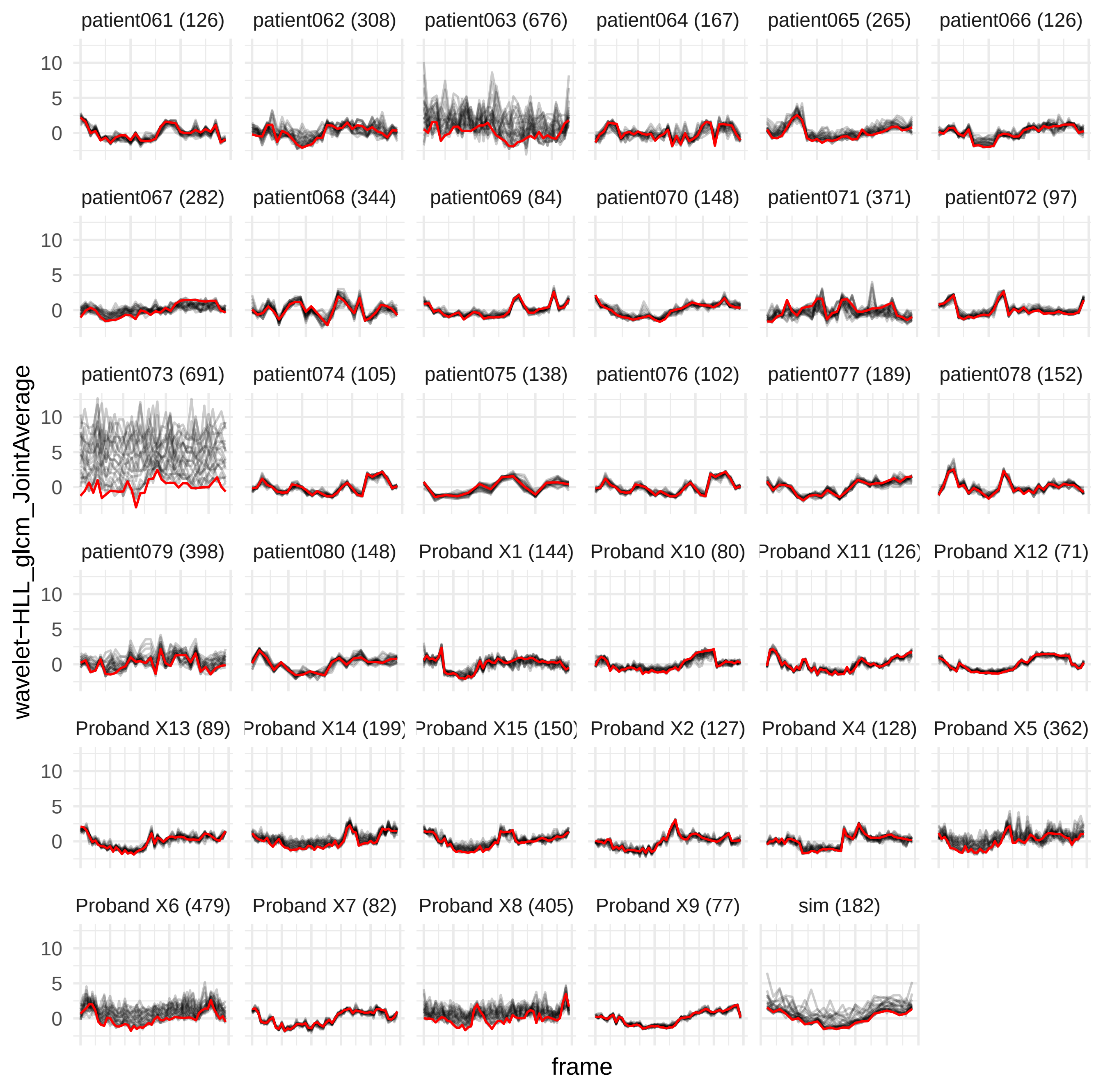

patient061 (126)
patient062 (308)
patient063 (676)
patient064 (167)
patient065 (265)
patient066 (126)
patient067 (282)
patient068 (344)
patient069 (84)
patient070 (148)
patient071 (371)
patient072 (97)
patient073 (691)
patient074 (105)
patient075 (138)
patient076 (102)
patient077 (189)
patient078 (152)
patient079 (398)
patient080 (148)
Proband X1 (144)
Proband X10 (80)
Proband X11 (126)
Proband X12 (71)
Proband X13 (89)
Proband X14 (199)
Proband X15 (150)
Proband X2 (127)
Proband X4 (128)
Proband X5 (362)
Proband X6 (479)
Proband X7 (82)
Proband X8 (405)
Proband X9 (77)
sim (182)
10
5
0
wavelet−HLL_glcm_JointAverage
frame

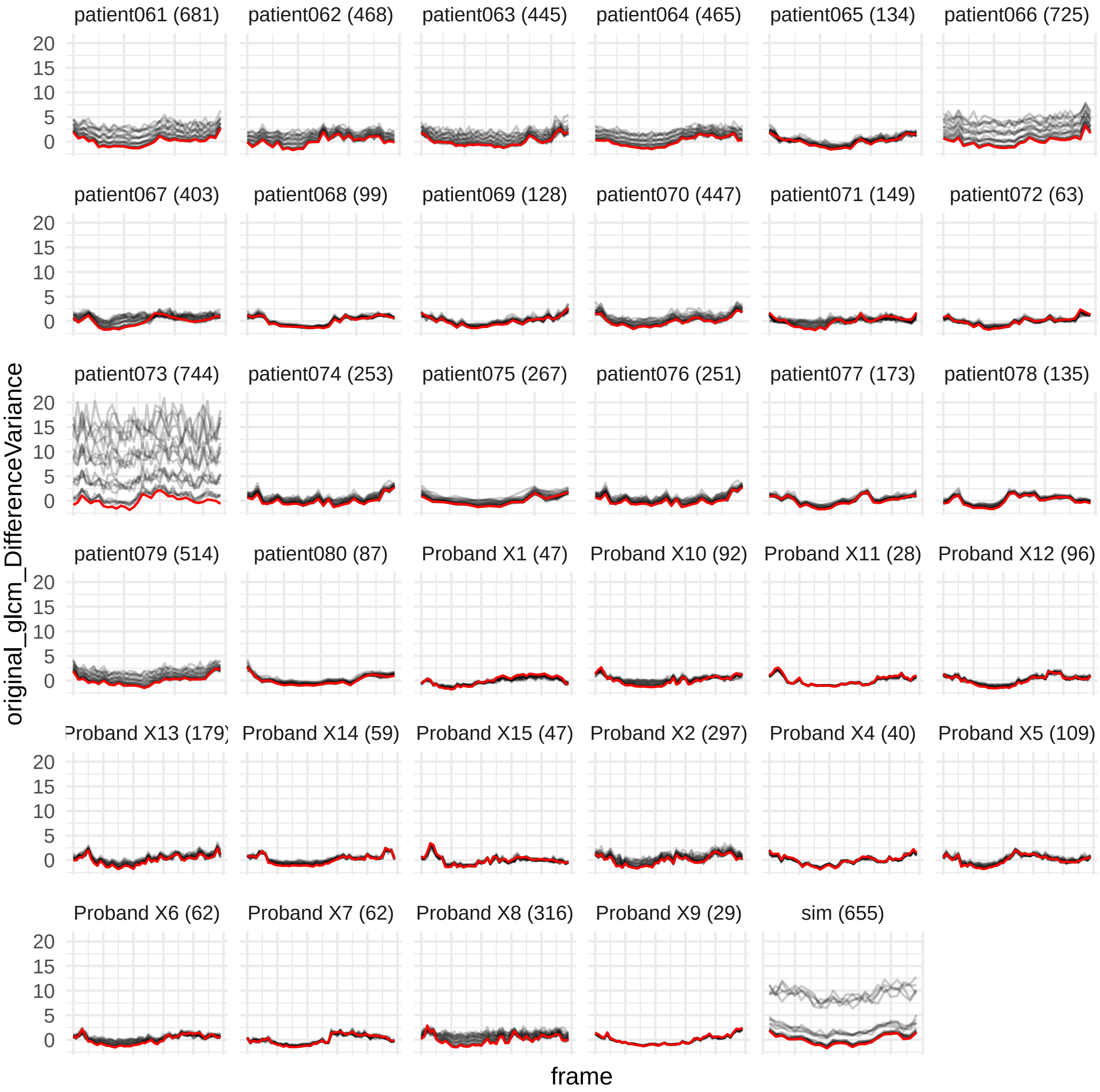

patient061 (681)
patient062 (468)
patient063 (445)
patient064 (465)
patient065 (134)
patient066 (725)
patient067 (403)
patient068 (99)
patient069 (128)
patient070 (447)
patient071 (149)
patient072 (63)
patient073 (744)
patient074 (253)
patient075 (267)
patient076 (251)
patient077 (173)
patient078 (135)
patient079 (514)
patient080 (87)
Proband X1 (47)
Proband X10 (92)
Proband X11 (28)
Proband X12 (96)
Proband X13 (179)
Proband X14 (59)
Proband X15 (47)
Proband X2 (297)
Proband X4 (40)
Proband X5 (109)
Proband X6 (62)
Proband X7 (62)
Proband X8 (316)
Proband X9 (29)
sim (655)
original_glcm_DifferenceVariance
0
5
10
15
20
frame

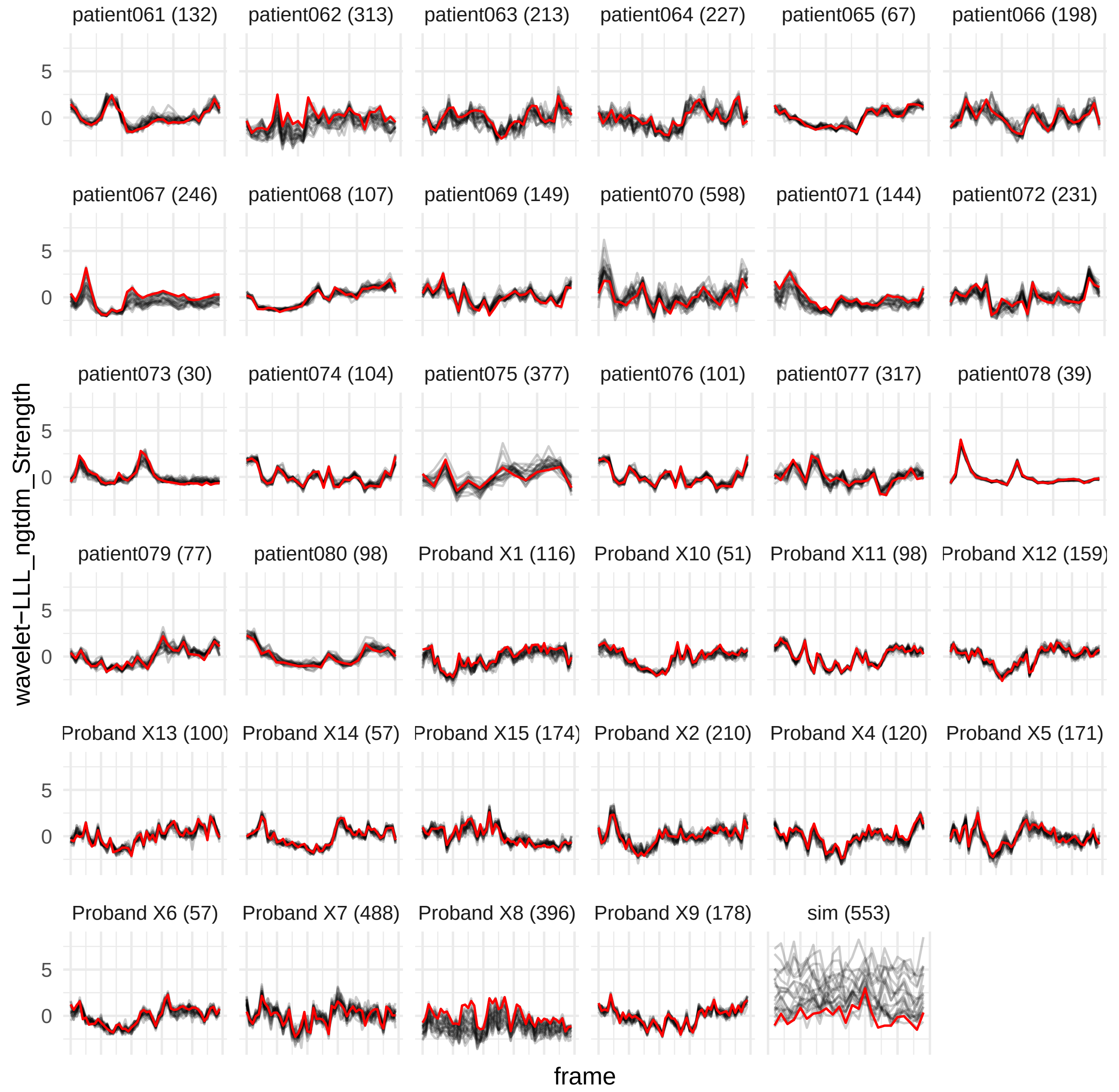

patient061 (132)
patient062 (313)
patient063 (213)
patient064 (227)
patient065 (67)
patient066 (198)
patient067 (246)
patient068 (107)
patient069 (149)
patient070 (598)
patient071 (144)
patient072 (231)
patient073 (30)
patient074 (104)
patient075 (377)
patient076 (101)
patient077 (317)
patient078 (39)
patient079 (77)
patient080 (98)
Proband X1 (116)
Proband X10 (51)
Proband X11 (98)
Proband X12 (159)
Proband X13 (100)
Proband X14 (57)
Proband X15 (174)
Proband X2 (210)
Proband X4 (120)
Proband X5 (171)
Proband X6 (57)
Proband X7 (488)
Proband X8 (396)
Proband X9 (178)
sim (553)
5
0
wavelet−LLL_ngtdm_Strength
frame

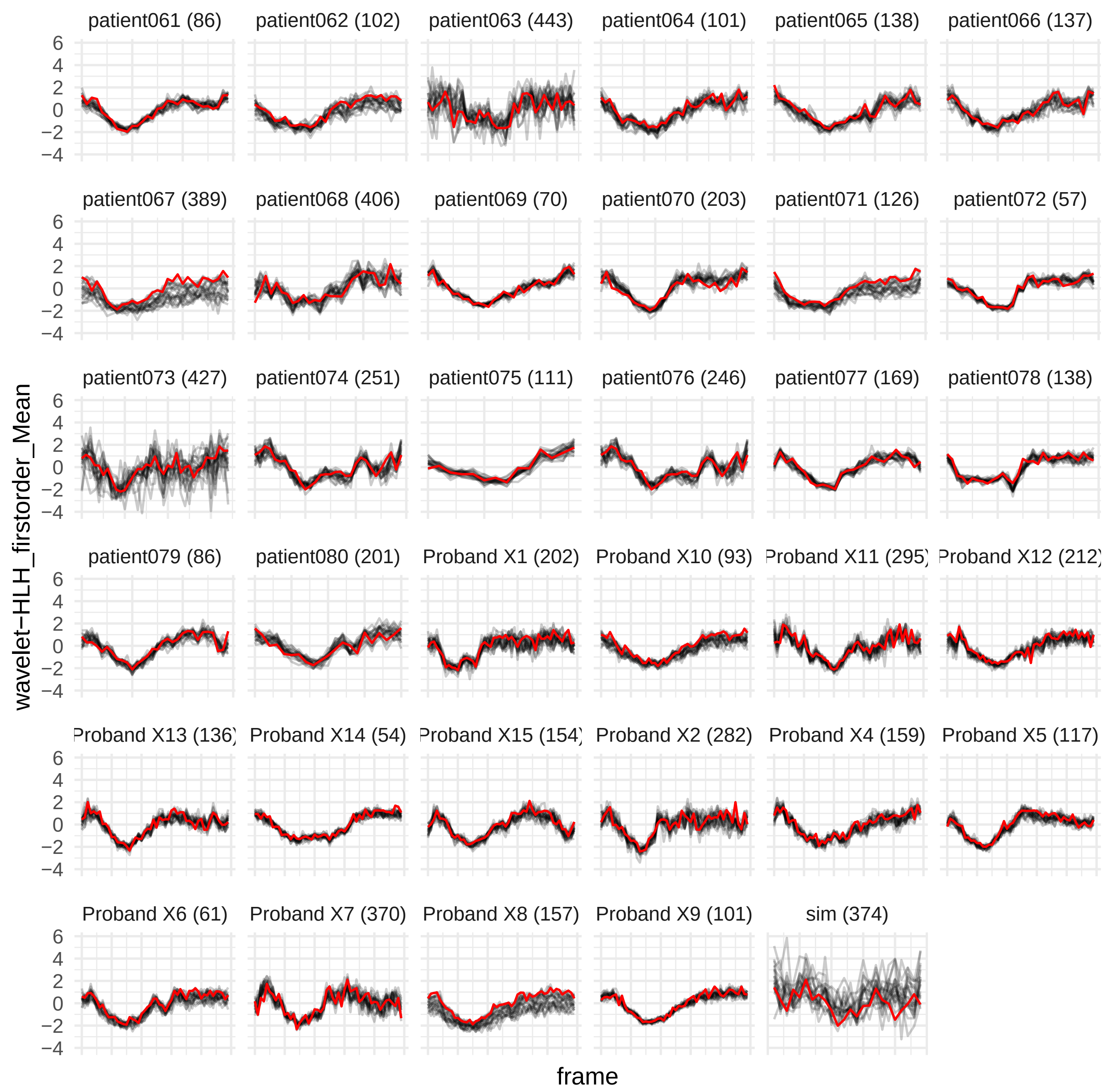
patient061 (86)
patient062 (102)
patient063 (443)
patient064 (101)
patient065 (138)
patient066 (137)
patient067 (389)
patient068 (406)
patient069 (70)
patient070 (203)
patient071 (126)
patient072 (57)
patient073 (427)
patient074 (251)
patient075 (111)
patient076 (246)
patient077 (169)
patient078 (138)
patient079 (86)
patient080 (201)
Proband X1 (202)
Proband X10 (93)
Proband X11 (295)
Proband X12 (212)
Proband X13 (136)
Proband X14 (54)
Proband X15 (154)
Proband X2 (282)
Proband X4 (159)
Proband X5 (117)
Proband X6 (61)
Proband X7 (370)
Proband X8 (157)
Proband X9 (101)
sim (374)
wavelet−HLH_firstorder_Mean
frame
6
4
2
0
−2
−4

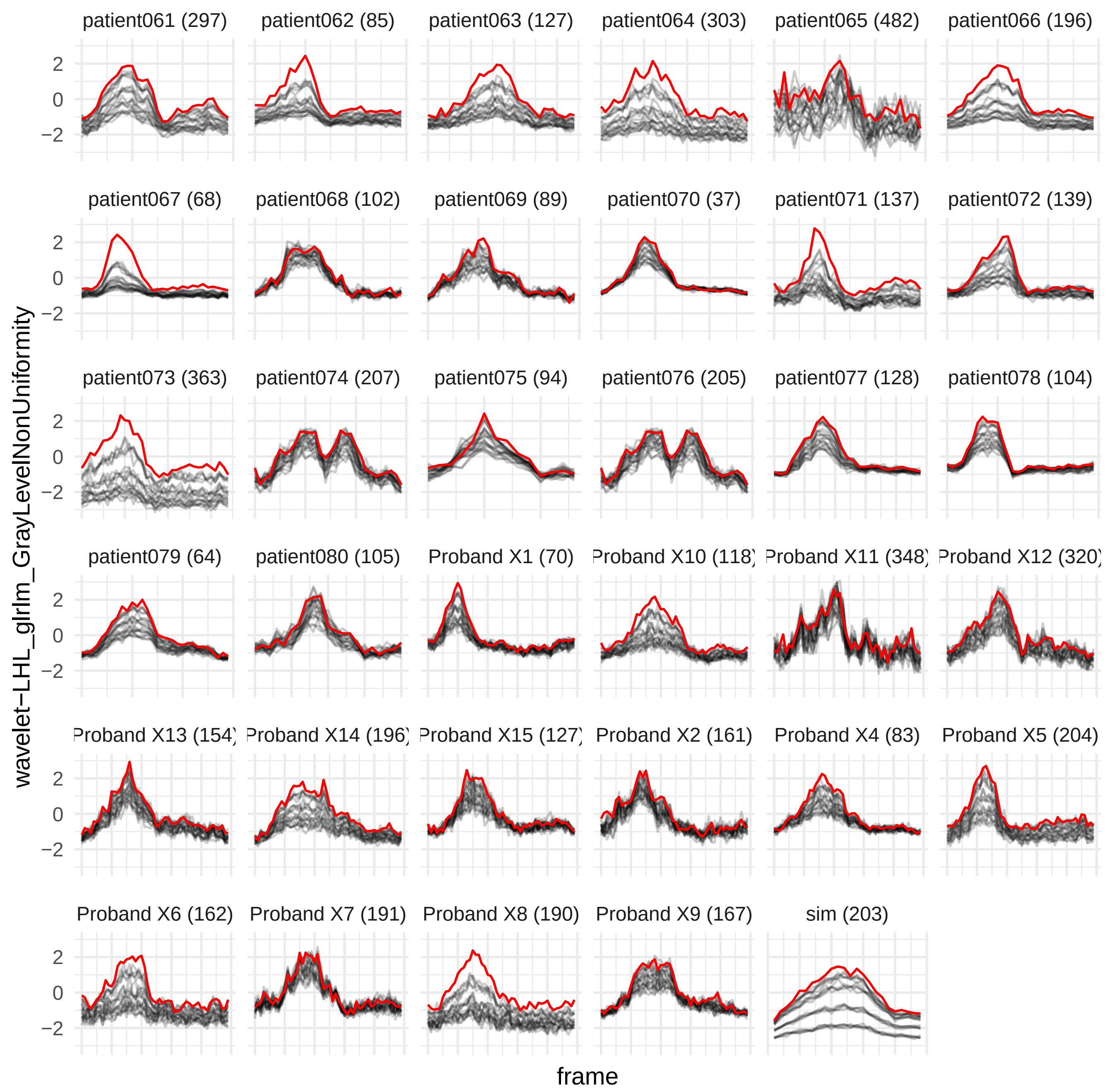

patient061 (297)
patient062 (85)
patient063 (127)
patient064 (303)
patient065 (482)
patient066 (196)
patient067 (68)
patient068 (102)
patient069 (89)
patient070 (37)
patient071 (137)
patient072 (139)
patient073 (363)
patient074 (207)
patient075 (94)
patient076 (205)
patient077 (128)
patient078 (104)
patient079 (64)
patient080 (105)
Proband X1 (70)
Proband X10 (118)
Proband X11 (348)
Proband X12 (320)
Proband X13 (154)
Proband X14 (196)
Proband X15 (127)
Proband X2 (161)
Proband X4 (83)
Proband X5 (204)
Proband X6 (162)
Proband X7 (191)
Proband X8 (190)
Proband X9 (167)
sim (203)
2
0
−2
wavelet−LHL_glrlm_GrayLevelNonUniformity
frame

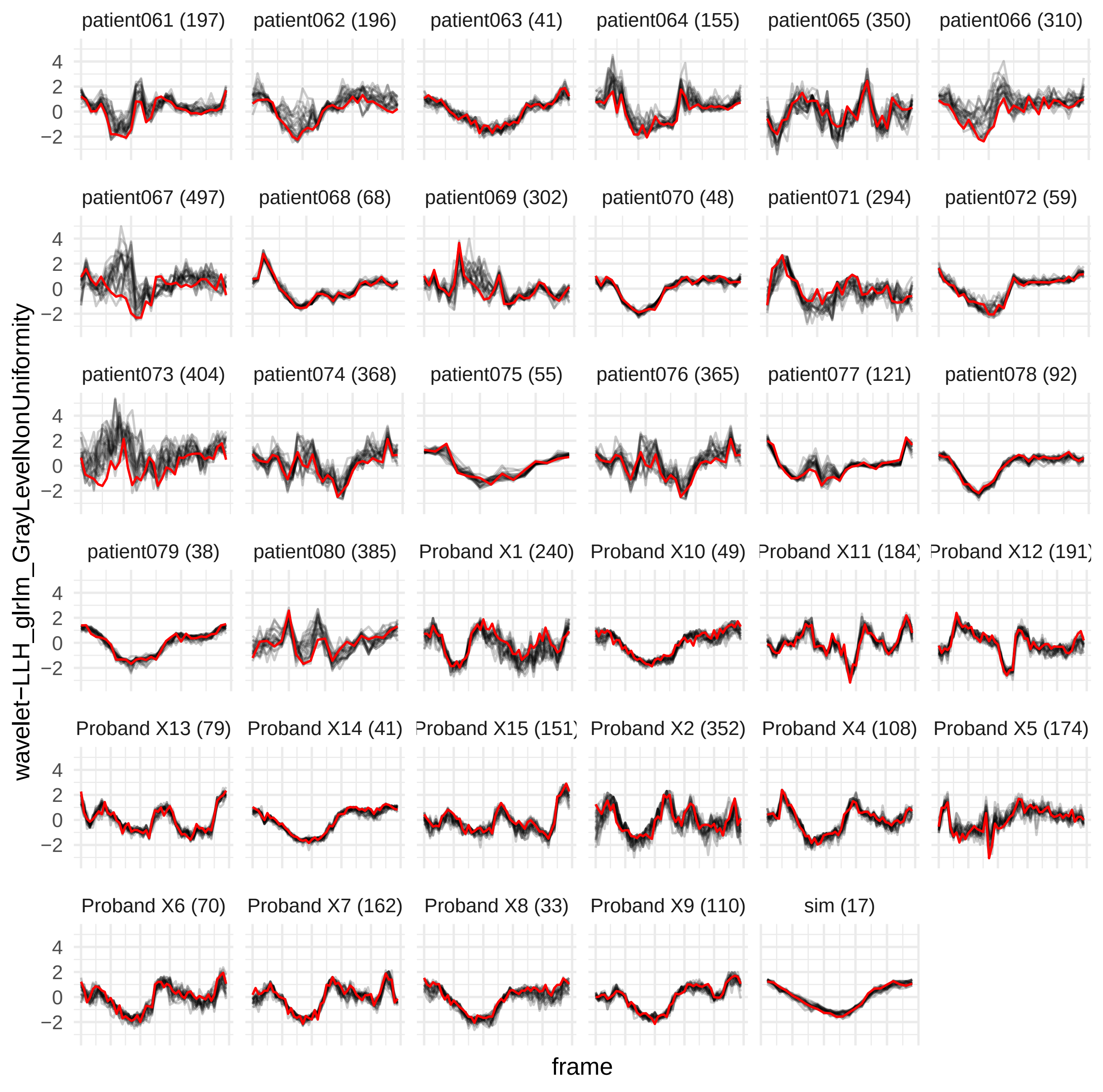
patient061 (197)
patient062 (196)
patient063 (41)
patient064 (155)
patient065 (350)
patient066 (310)
patient067 (497)
patient068 (68)
patient069 (302)
patient070 (48)
patient071 (294)
patient072 (59)
patient073 (404)
patient074 (368)
patient075 (55)
patient076 (365)
patient077 (121)
patient078 (92)
patient079 (38)
patient080 (385)
Proband X1 (240)
Proband X10 (49)
Proband X11 (184)
Proband X12 (191)
Proband X13 (79)
Proband X14 (41)
Proband X15 (151)
Proband X2 (352)
Proband X4 (108)
Proband X5 (174)
Proband X6 (70)
Proband X7 (162)
Proband X8 (33)
Proband X9 (110)
sim (17)
4
2
0
−2
wavelet−LLH_glrlm_GrayLevelNonUniformity
frame

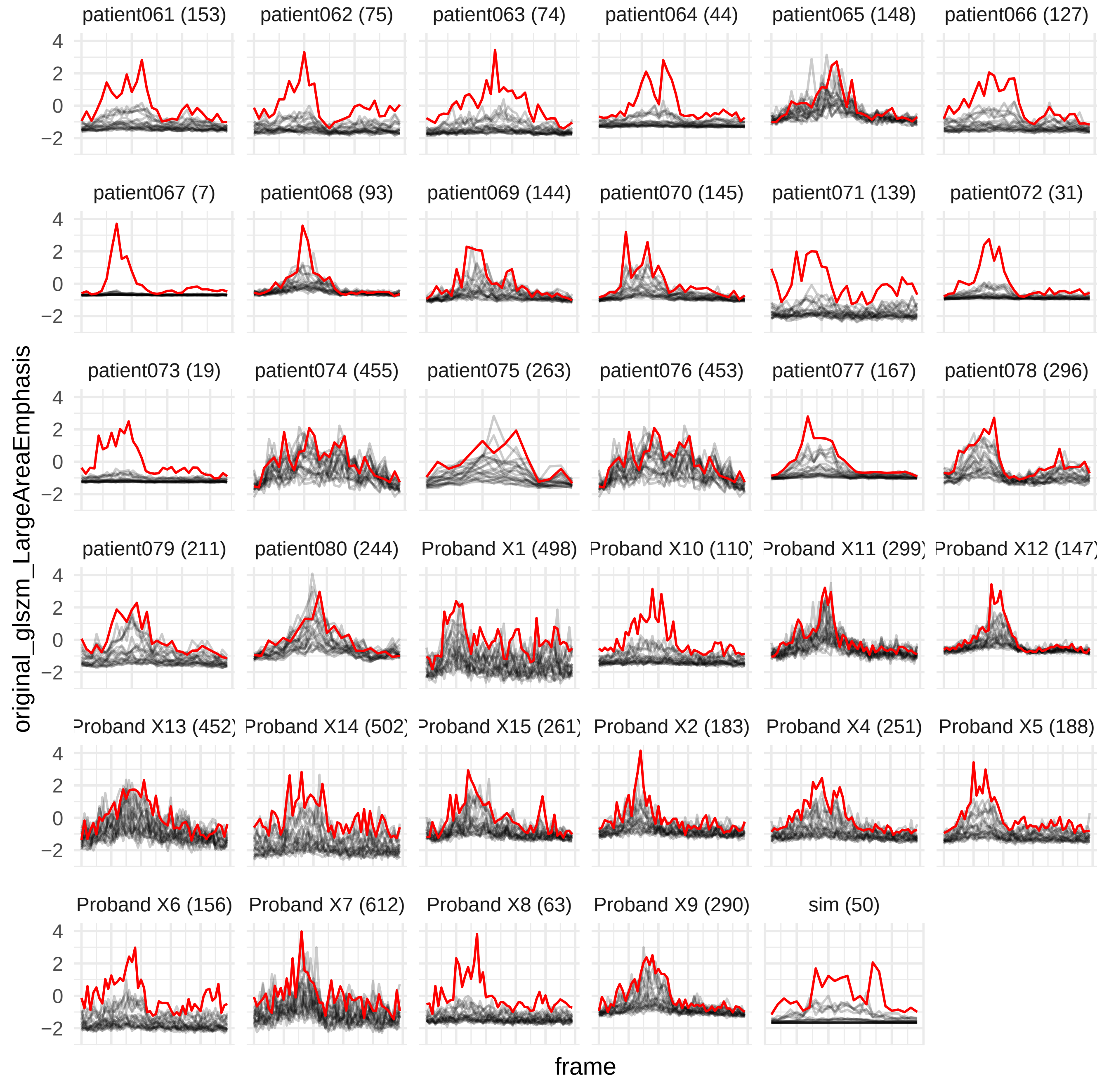

patient061 (153)
patient062 (75)
patient063 (74)
patient064 (44)
patient065 (148)
patient066 (127)
patient067 (7)
patient068 (93)
patient069 (144)
patient070 (145)
patient071 (139)
patient072 (31)
patient073 (19)
patient074 (455)
patient075 (263)
patient076 (453)
patient077 (167)
patient078 (296)
patient079 (211)
patient080 (244)
Proband X1 (498)
Proband X10 (110)
Proband X11 (299)
Proband X12 (147)
Proband X13 (452)
Proband X14 (502)
Proband X15 (261)
Proband X2 (183)
Proband X4 (251)
Proband X5 (188)
Proband X6 (156)
Proband X7 (612)
Proband X8 (63)
Proband X9 (290)
sim (50)
original_glszm_LargeAreaEmphasis
frame
4
2
0
−2

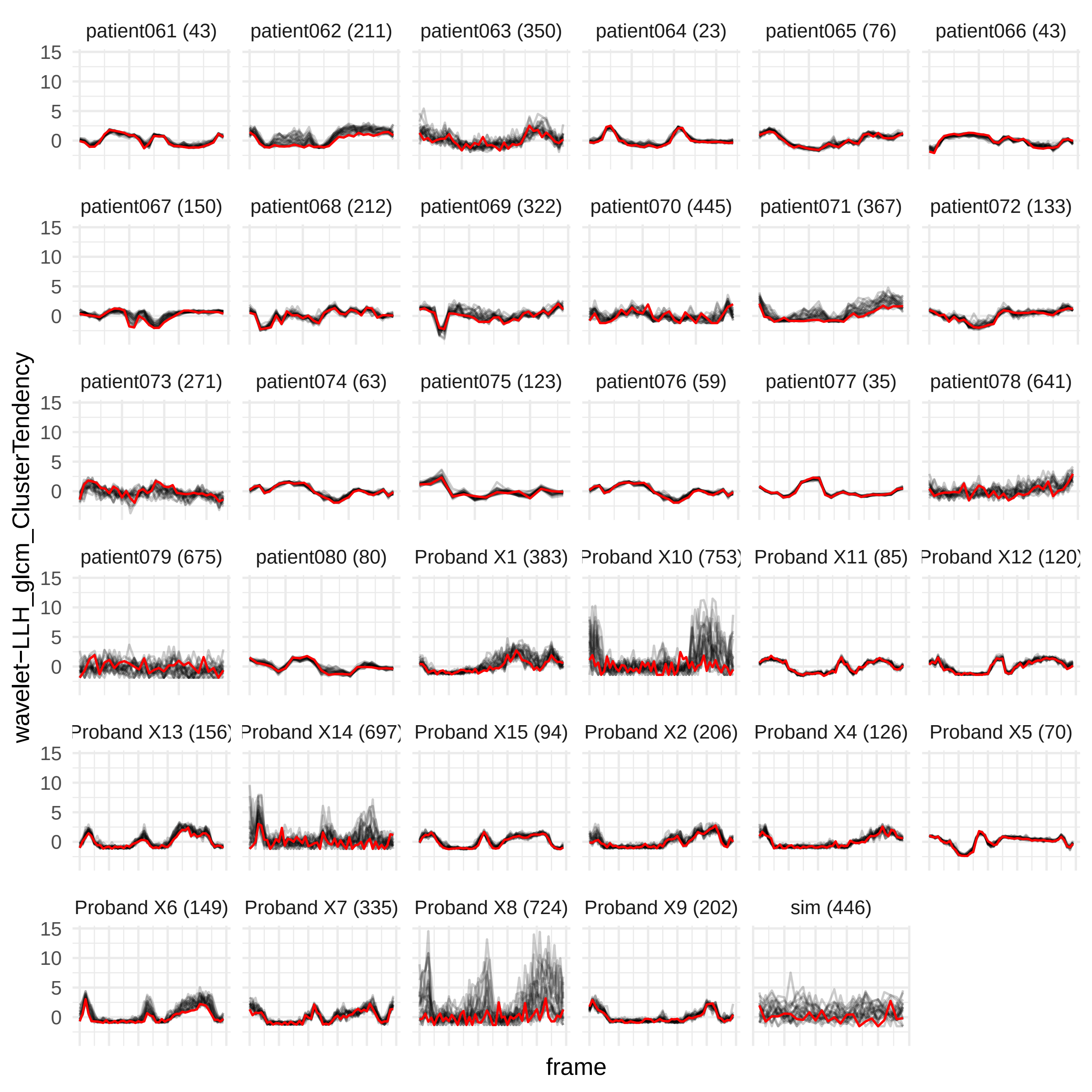
patient061 (43)
patient062 (211)
patient063 (350)
patient064 (23)
patient065 (76)
patient066 (43)
patient067 (150)
patient068 (212)
patient069 (322)
patient070 (445)
patient071 (367)
patient072 (133)
patient073 (271)
patient074 (63)
patient075 (123)
patient076 (59)
patient077 (35)
patient078 (641)
patient079 (675)
patient080 (80)
Proband X1 (383)
Proband X10 (753)
Proband X11 (85)
Proband X12 (120)
Proband X13 (156)
Proband X14 (697)
Proband X15 (94)
Proband X2 (206)
Proband X4 (126)
Proband X5 (70)
Proband X6 (149)
Proband X7 (335)
Proband X8 (724)
Proband X9 (202)
sim (446)
wavelet−LLH_glcm_ClusterTendency
frame

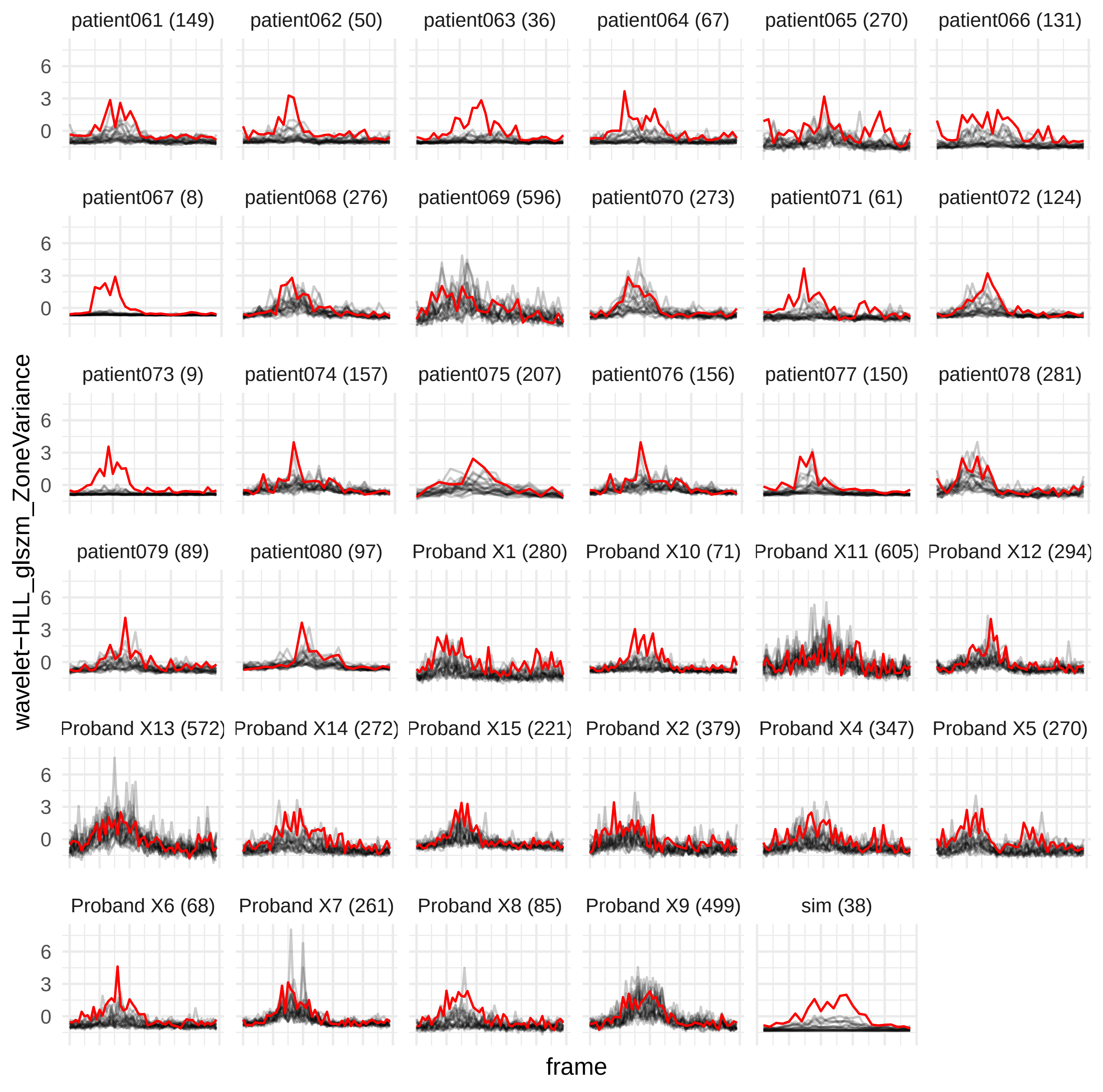
patient061 (149)
patient062 (50)
patient063 (36)
patient064 (67)
patient065 (270)
patient066 (131)
patient067 (8)
patient068 (276)
patient069 (596)
patient070 (273)
patient071 (61)
patient072 (124)
patient073 (9)
patient074 (157)
patient075 (207)
patient076 (156)
patient077 (150)
patient078 (281)
patient079 (89)
patient080 (97)
Proband X1 (280)
Proband X10 (71)
Proband X11 (605)
Proband X12 (294)
Proband X13 (572)
Proband X14 (272)
Proband X15 (221)
Proband X2 (379)
Proband X4 (347)
Proband X5 (270)
Proband X6 (68)
Proband X7 (261)
Proband X8 (85)
Proband X9 (499)
sim (38)
6
3
0
wavelet−HLL_glszm_ZoneVariance
frame

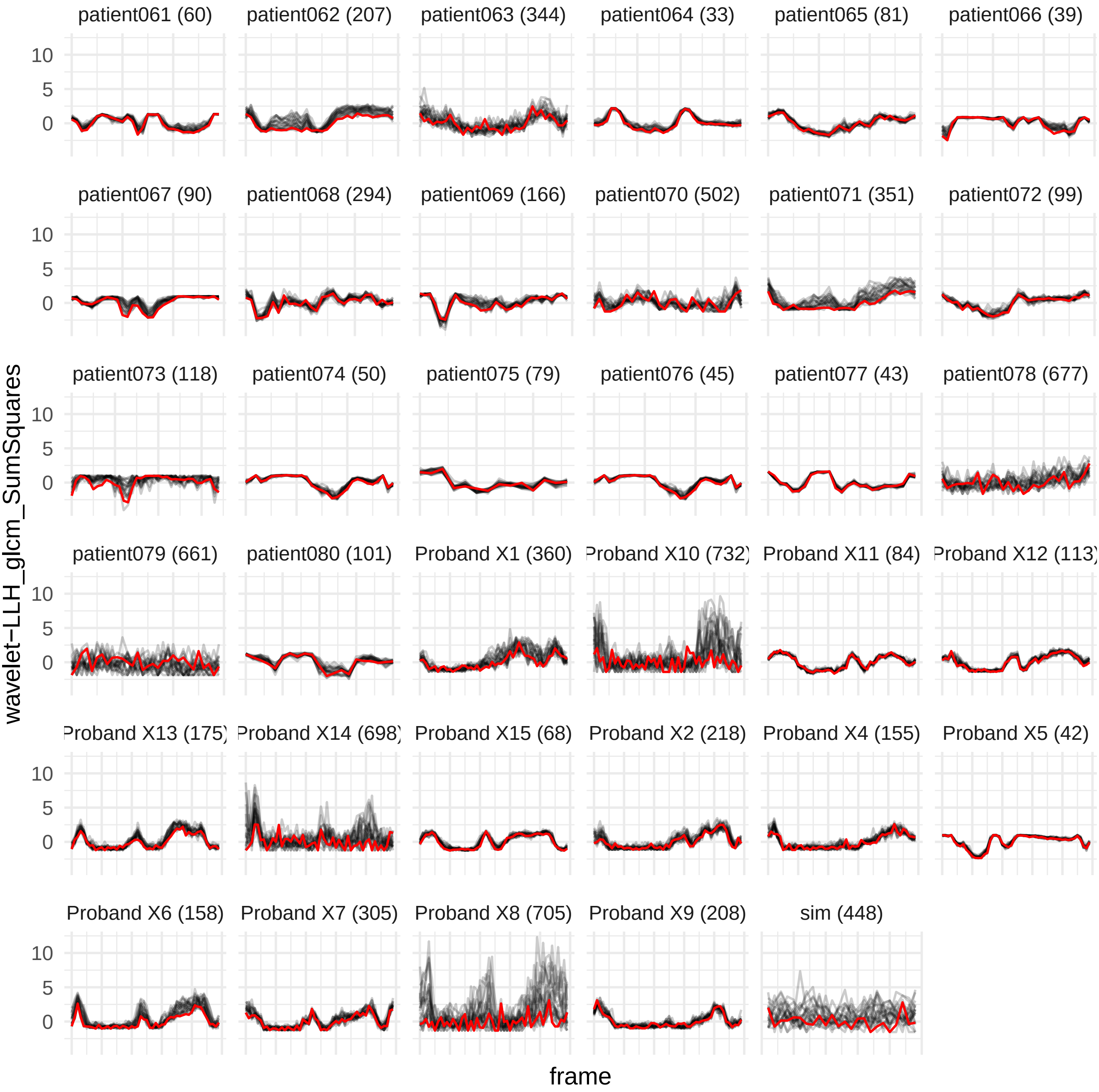

patient061 (60)
patient062 (207)
patient063 (344)
patient064 (33)
patient065 (81)
patient066 (39)
patient067 (90)
patient068 (294)
patient069 (166)
patient070 (502)
patient071 (351)
patient072 (99)
patient073 (118)
patient074 (50)
patient075 (79)
patient076 (45)
patient077 (43)
patient078 (677)
patient079 (661)
patient080 (101)
Proband X1 (360)
Proband X10 (732)
Proband X11 (84)
Proband X12 (113)
Proband X13 (175)
Proband X14 (698)
Proband X15 (68)
Proband X2 (218)
Proband X4 (155)
Proband X5 (42)
Proband X6 (158)
Proband X7 (305)
Proband X8 (705)
Proband X9 (208)
sim (448)
wavelet−LLH_glcm_SumSquares
10
5
0
frame

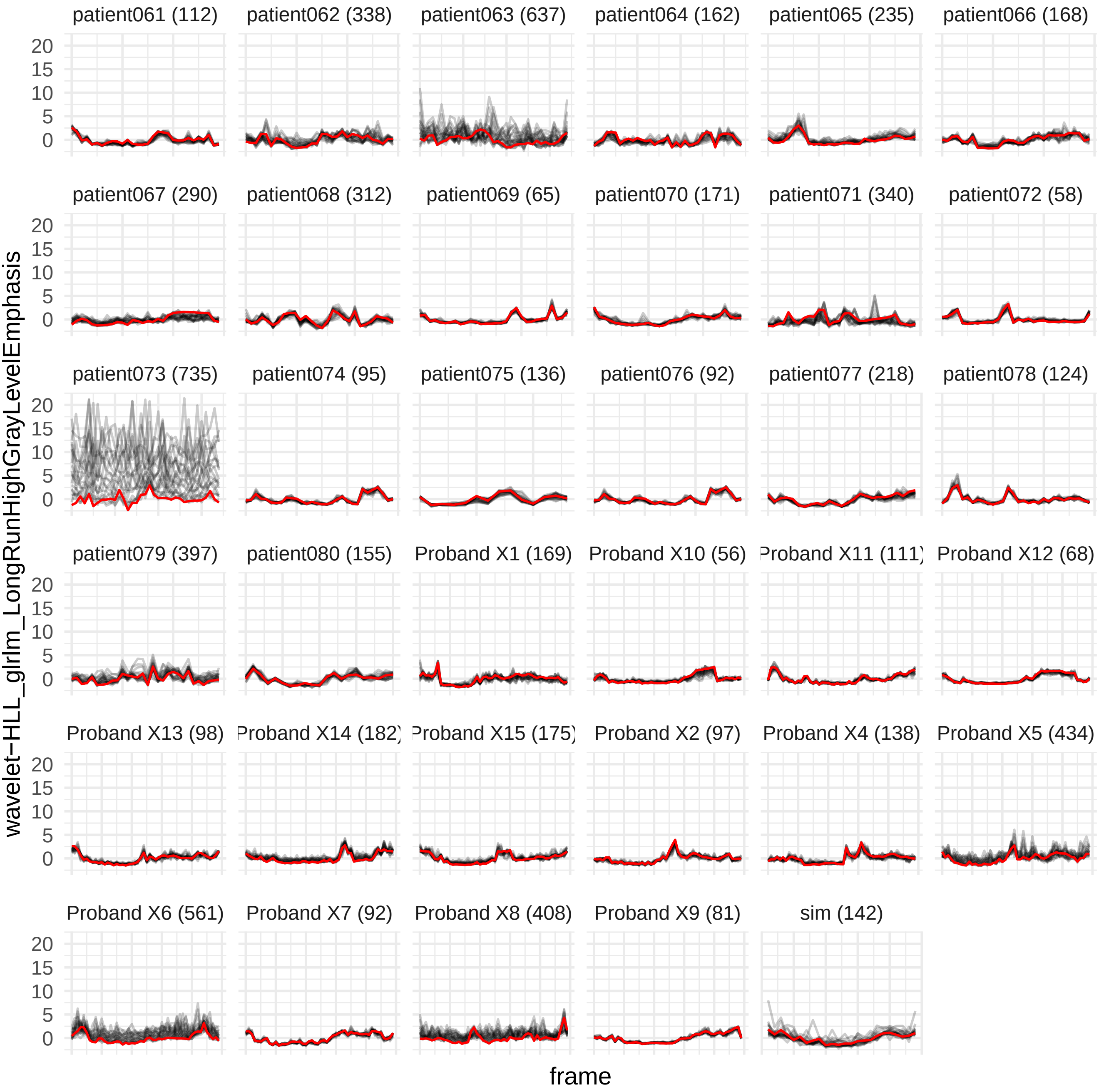
patient061 (112)
patient062 (338)
patient063 (637)
patient064 (162)
patient065 (235)
patient066 (168)
patient067 (290)
patient068 (312)
patient069 (65)
patient070 (171)
patient071 (340)
patient072 (58)
patient073 (735)
patient074 (95)
patient075 (136)
patient076 (92)
patient077 (218)
patient078 (124)
patient079 (397)
patient080 (155)
Proband X1 (169)
Proband X10 (56)
Proband X11 (111)
Proband X12 (68)
Proband X13 (98)
Proband X14 (182)
Proband X15 (175)
Proband X2 (97)
Proband X4 (138)
Proband X5 (434)
Proband X6 (561)
Proband X7 (92)
Proband X8 (408)
Proband X9 (81)
sim (142)
20
15
10
5
0
wavelet−HLL_glrlm_LongRunHighGrayLevelEmphasis
frame

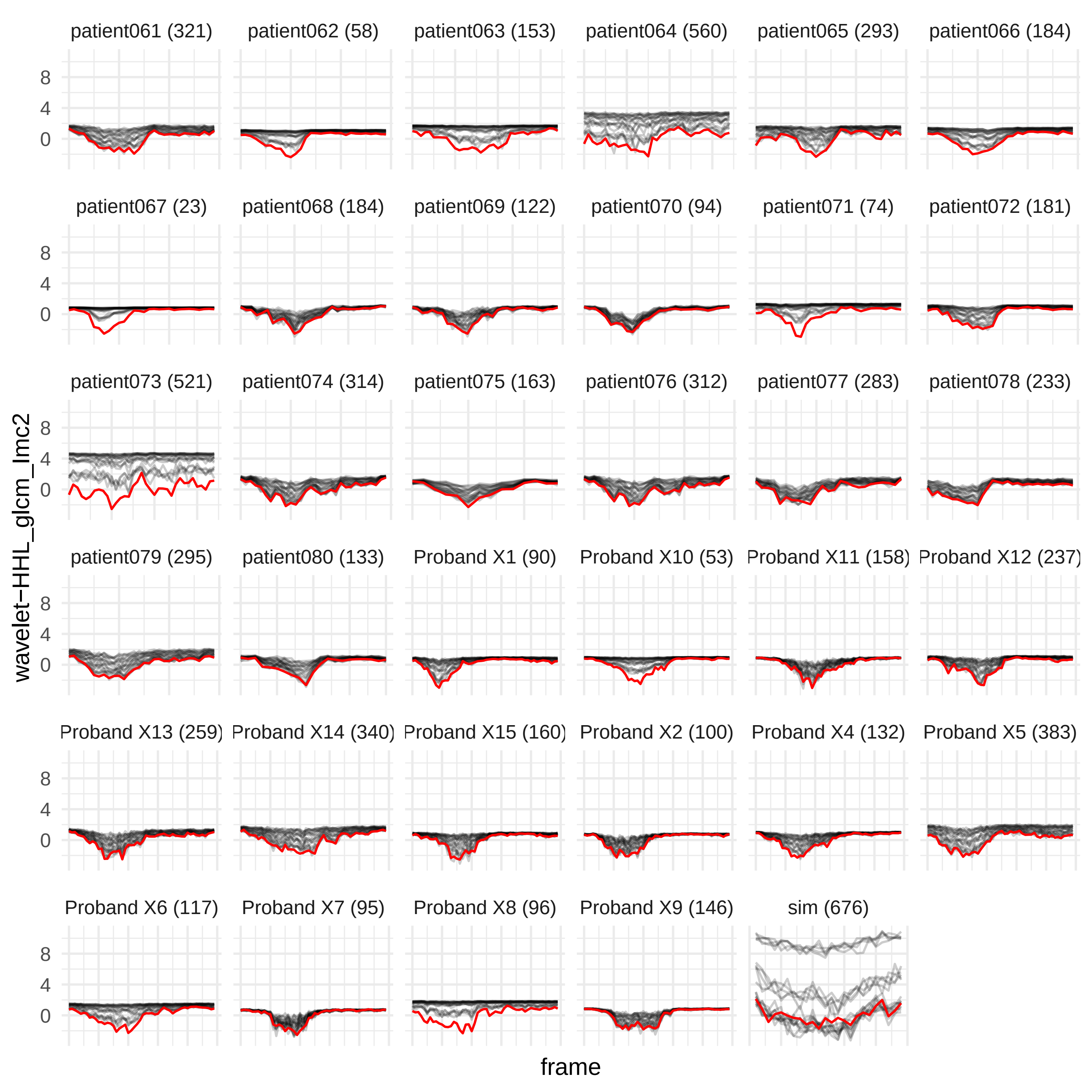

patient061 (321)
patient062 (58)
patient063 (153)
patient064 (560)
patient065 (293)
patient066 (184)
patient067 (23)
patient068 (184)
patient069 (122)
patient070 (94)
patient071 (74)
patient072 (181)
patient073 (521)
patient074 (314)
patient075 (163)
patient076 (312)
patient077 (283)
patient078 (233)
patient079 (295)
patient080 (133)
Proband X1 (90)
Proband X10 (53)
Proband X11 (158)
Proband X12 (237)
Proband X13 (259)
Proband X14 (340)
Proband X15 (160)
Proband X2 (100)
Proband X4 (132)
Proband X5 (383)
Proband X6 (117)
Proband X7 (95)
Proband X8 (96)
Proband X9 (146)
sim (676)
wavelet−HHL_glcm_Imc2
8
4
0
frame

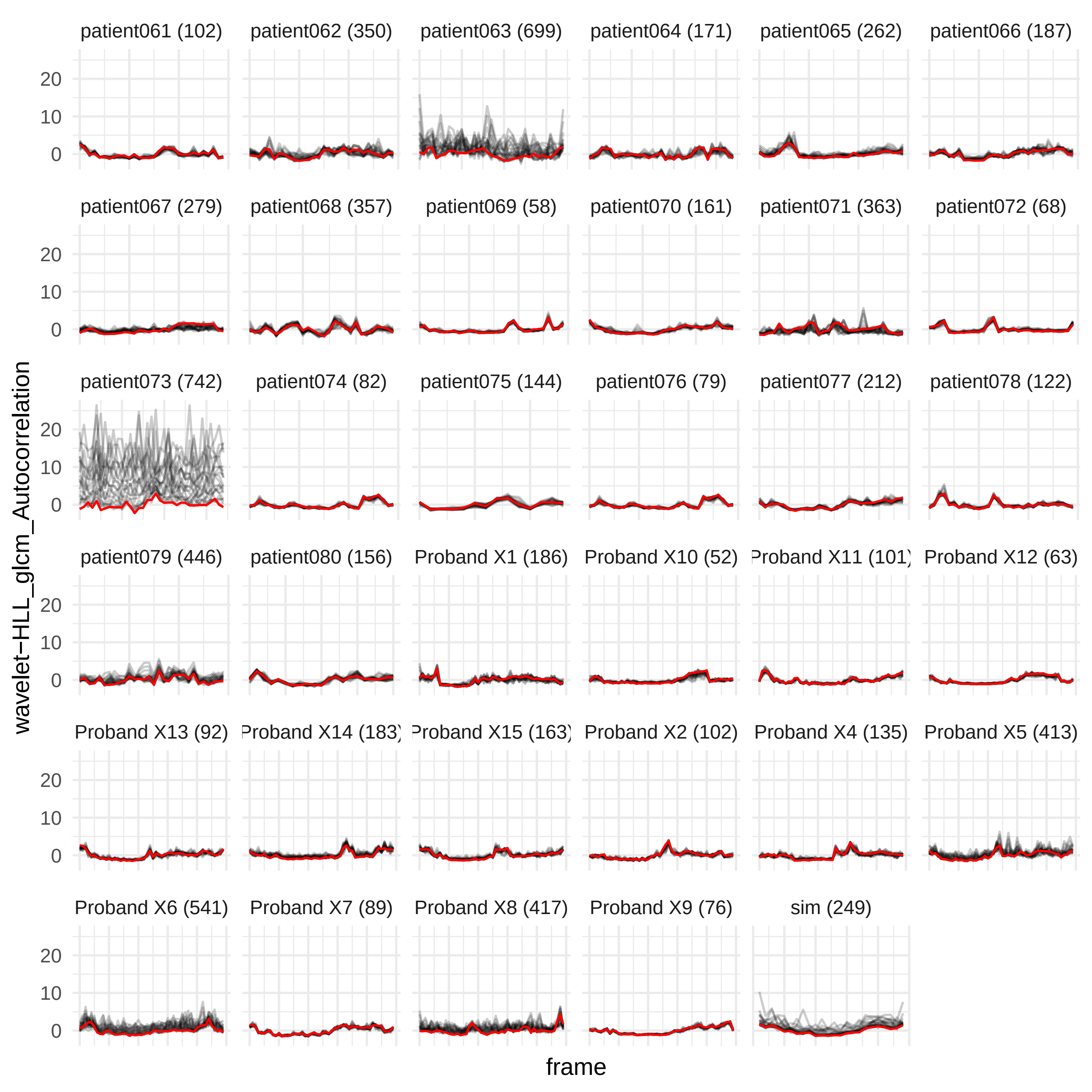
patient061 (102)
patient062 (350)
patient063 (699)
patient064 (171)
patient065 (262)
patient066 (187)
patient067 (279)
patient068 (357)
patient069 (58)
patient070 (161)
patient071 (363)
patient072 (68)
patient073 (742)
patient074 (82)
patient075 (144)
patient076 (79)
patient077 (212)
patient078 (122)
patient079 (446)
patient080 (156)
Proband X1 (186)
Proband X10 (52)
Proband X11 (101)
Proband X12 (63)
Proband X13 (92)
Proband X14 (183)
Proband X15 (163)
Proband X2 (102)
Proband X4 (135)
Proband X5 (413)
Proband X6 (541)
Proband X7 (89)
Proband X8 (417)
Proband X9 (76)
sim (249)
20
10
0
wavelet−HLL_glcm_Autocorrelation
frame

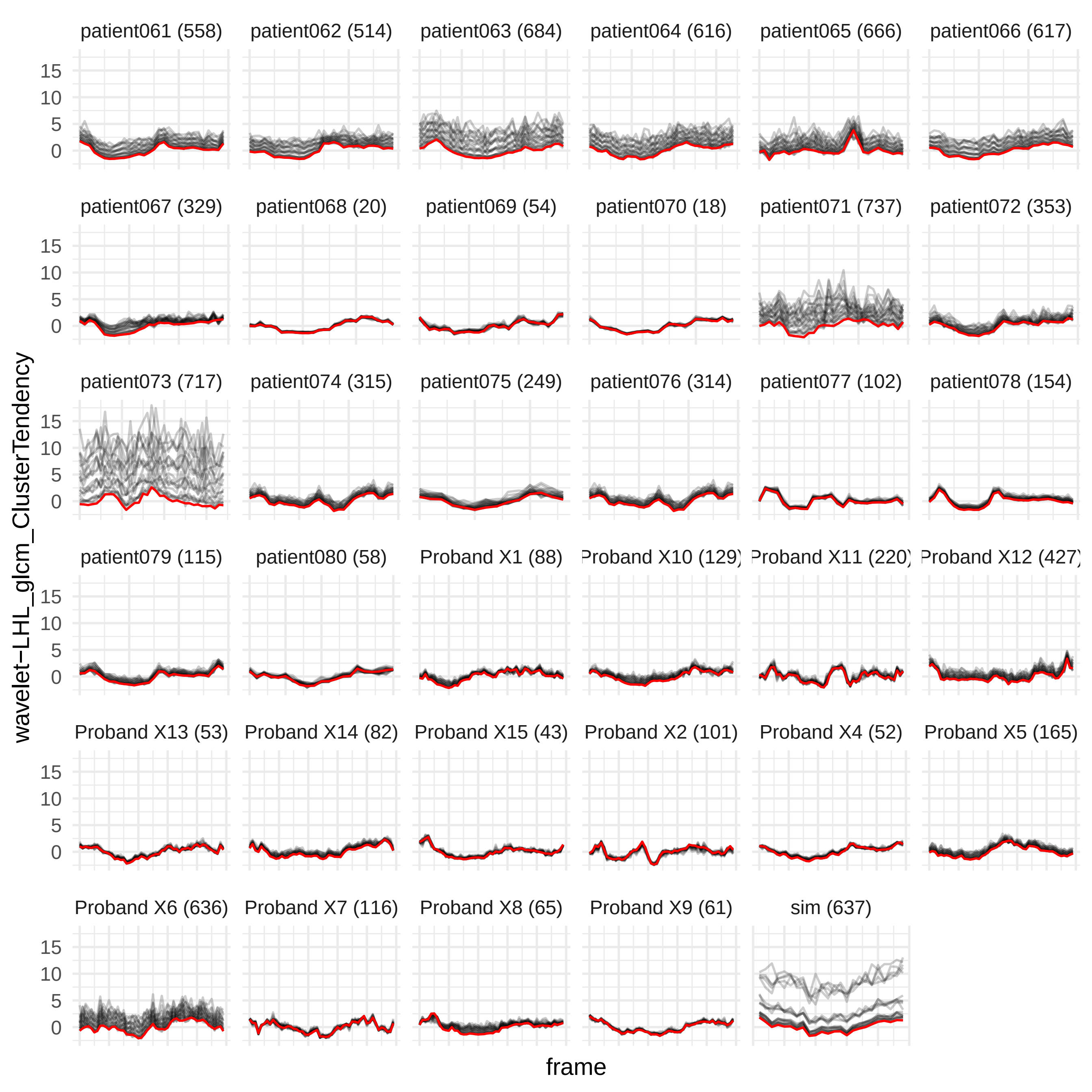
patient061 (558)
patient062 (514)
patient063 (684)
patient064 (616)
patient065 (666)
patient066 (617)
patient067 (329)
patient068 (20)
patient069 (54)
patient070 (18)
patient071 (737)
patient072 (353)
patient073 (717)
patient074 (315)
patient075 (249)
patient076 (314)
patient077 (102)
patient078 (154)
patient079 (115)
patient080 (58)
Proband X1 (88)
Proband X10 (129)
Proband X11 (220)
Proband X12 (427)
Proband X13 (53)
Proband X14 (82)
Proband X15 (43)
Proband X2 (101)
Proband X4 (52)
Proband X5 (165)
Proband X6 (636)
Proband X7 (116)
Proband X8 (65)
Proband X9 (61)
sim (637)
wavelet−LHL_glcm_ClusterTendency
frame
15
10
5
0

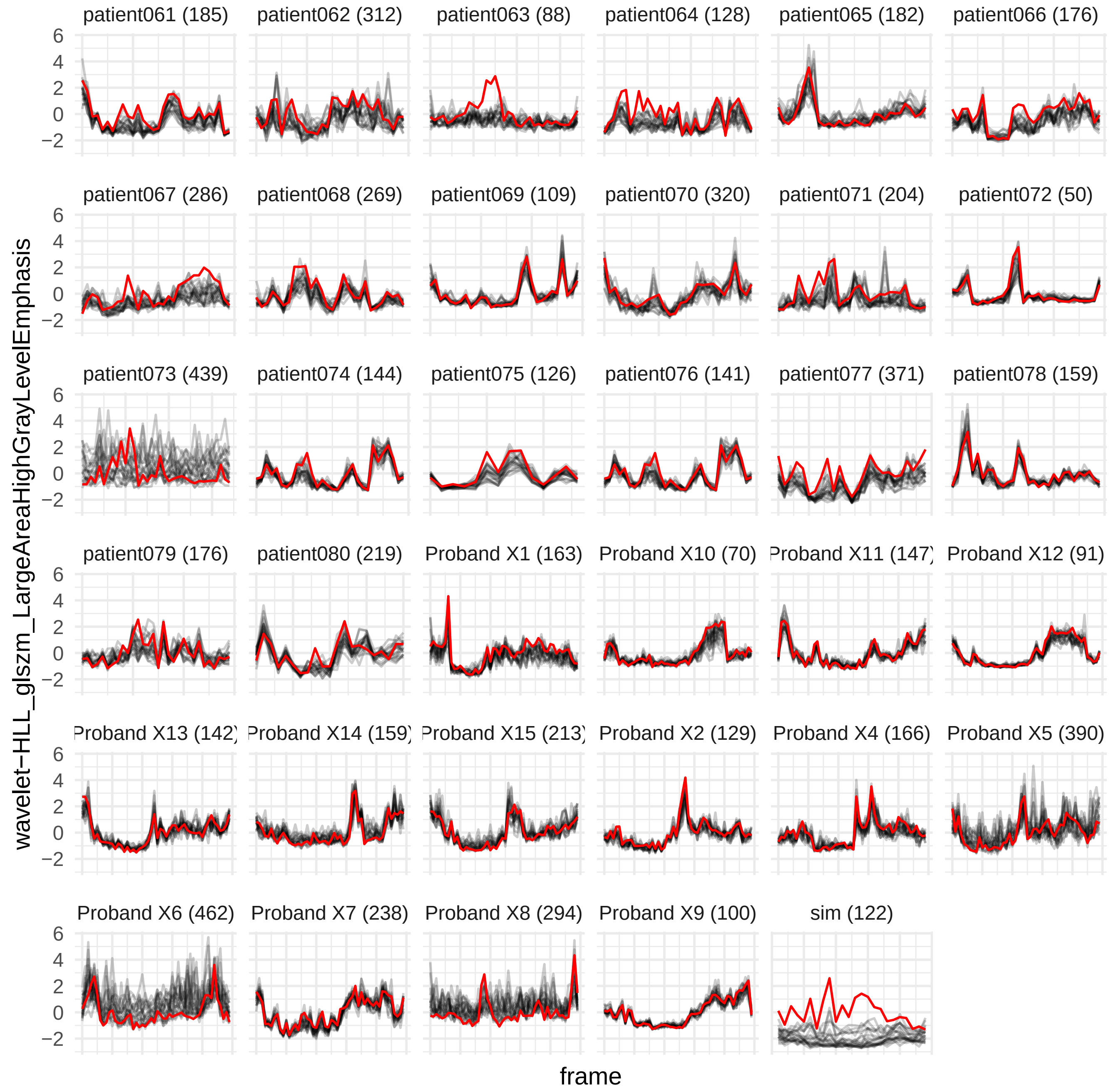
patient061 (185)
patient062 (312)
patient063 (88)
patient064 (128)
patient065 (182)
patient066 (176)
patient067 (286)
patient068 (269)
patient069 (109)
patient070 (320)
patient071 (204)
patient072 (50)
patient073 (439)
patient074 (144)
patient075 (126)
patient076 (141)
patient077 (371)
patient078 (159)
patient079 (176)
patient080 (219)
Proband X1 (163)
Proband X10 (70)
Proband X11 (147)
Proband X12 (91)
Proband X13 (142)
Proband X14 (159)
Proband X15 (213)
Proband X2 (129)
Proband X4 (166)
Proband X5 (390)
Proband X6 (462)
Proband X7 (238)
Proband X8 (294)
Proband X9 (100)
sim (122)
wavelet−HLL_glszm_LargeAreaHighGrayLevelEmphasis
frame
6
4
2
0
−2

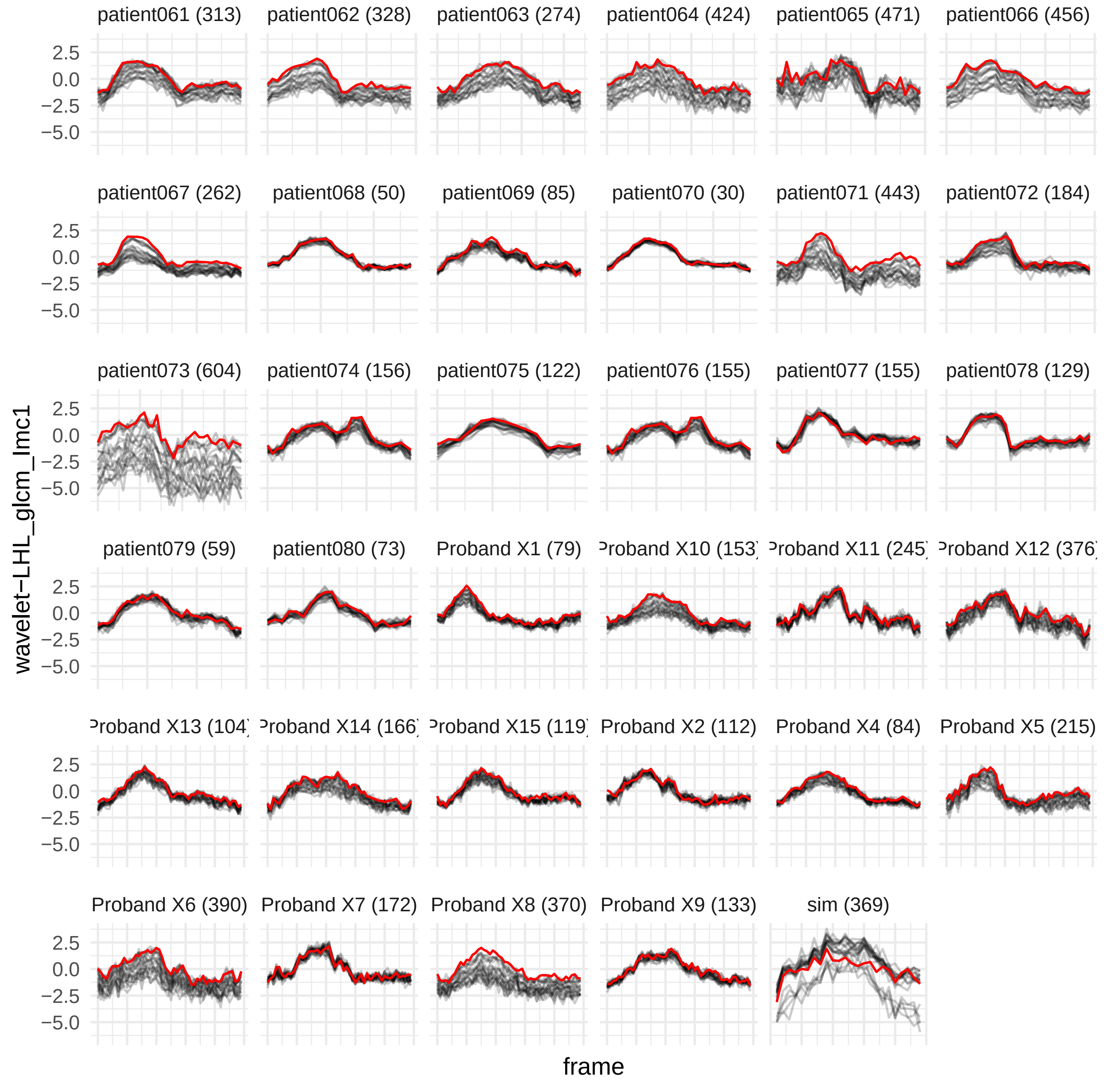

patient061 (313)
patient062 (328)
patient063 (274)
patient064 (424)
patient065 (471)
patient066 (456)
patient067 (262)
patient068 (50)
patient069 (85)
patient070 (30)
patient071 (443)
patient072 (184)
patient073 (604)
patient074 (156)
patient075 (122)
patient076 (155)
patient077 (155)
patient078 (129)
patient079 (59)
patient080 (73)
Proband X1 (79)
Proband X10 (153)
Proband X11 (245)
Proband X12 (376)
Proband X13 (104)
Proband X14 (166)
Proband X15 (119)
Proband X2 (112)
Proband X4 (84)
Proband X5 (215)
Proband X6 (390)
Proband X7 (172)
Proband X8 (370)
Proband X9 (133)
sim (369)
2.5
0.0
−2.5
−5.0
wavelet−LHL_glcm_Imc1
frame

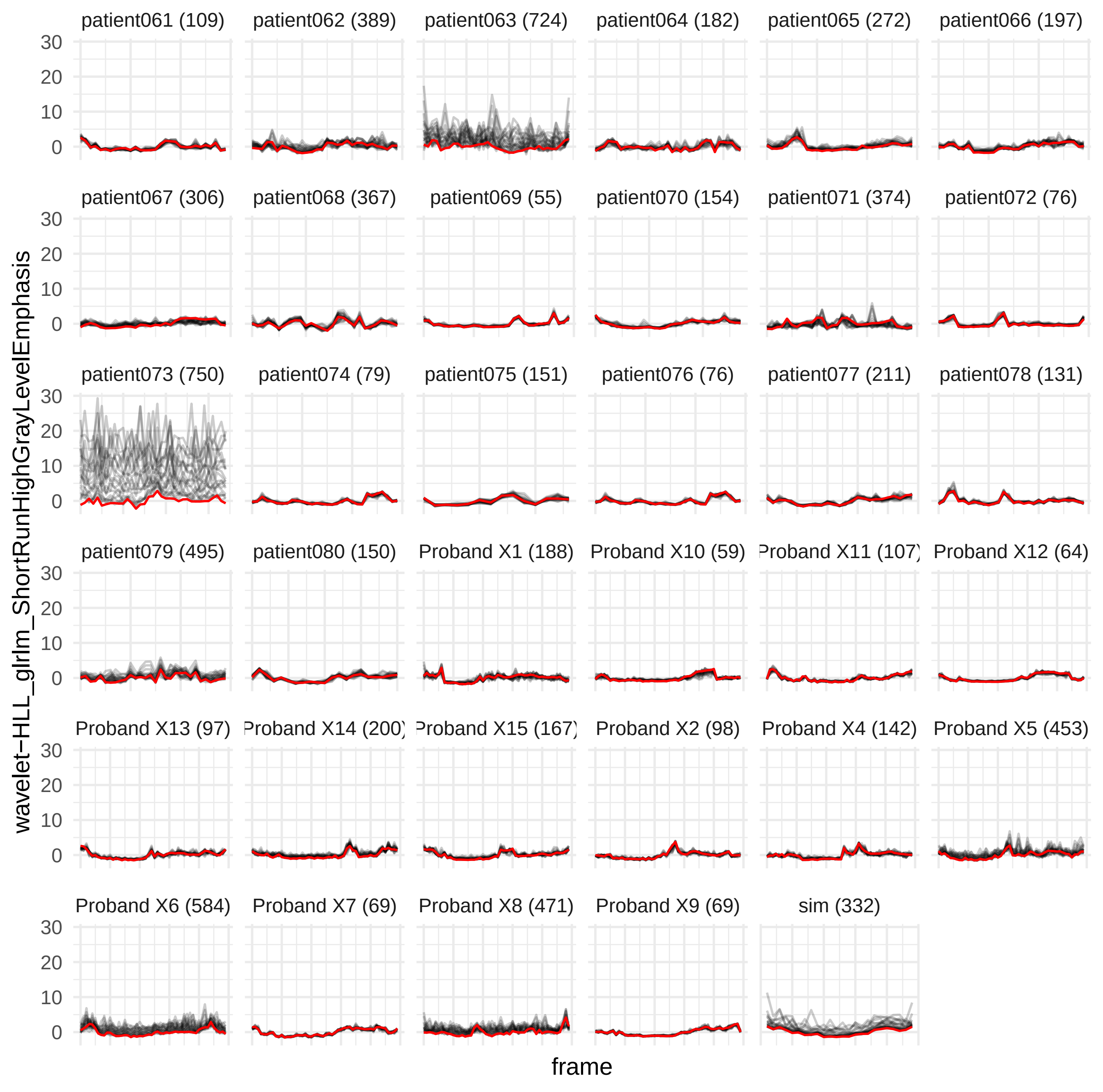
patient061 (109)
patient062 (389)
patient063 (724)
patient064 (182)
patient065 (272)
patient066 (197)
patient067 (306)
patient068 (367)
patient069 (55)
patient070 (154)
patient071 (374)
patient072 (76)
patient073 (750)
patient074 (79)
patient075 (151)
patient076 (76)
patient077 (211)
patient078 (131)
patient079 (495)
patient080 (150)
Proband X1 (188)
Proband X10 (59)
Proband X11 (107)
Proband X12 (64)
Proband X13 (97)
Proband X14 (200)
Proband X15 (167)
Proband X2 (98)
Proband X4 (142)
Proband X5 (453)
Proband X6 (584)
Proband X7 (69)
Proband X8 (471)
Proband X9 (69)
sim (332)
30
20
10
0
wavelet−HLL_glrlm_ShortRunHighGrayLevelEmphasis
frame

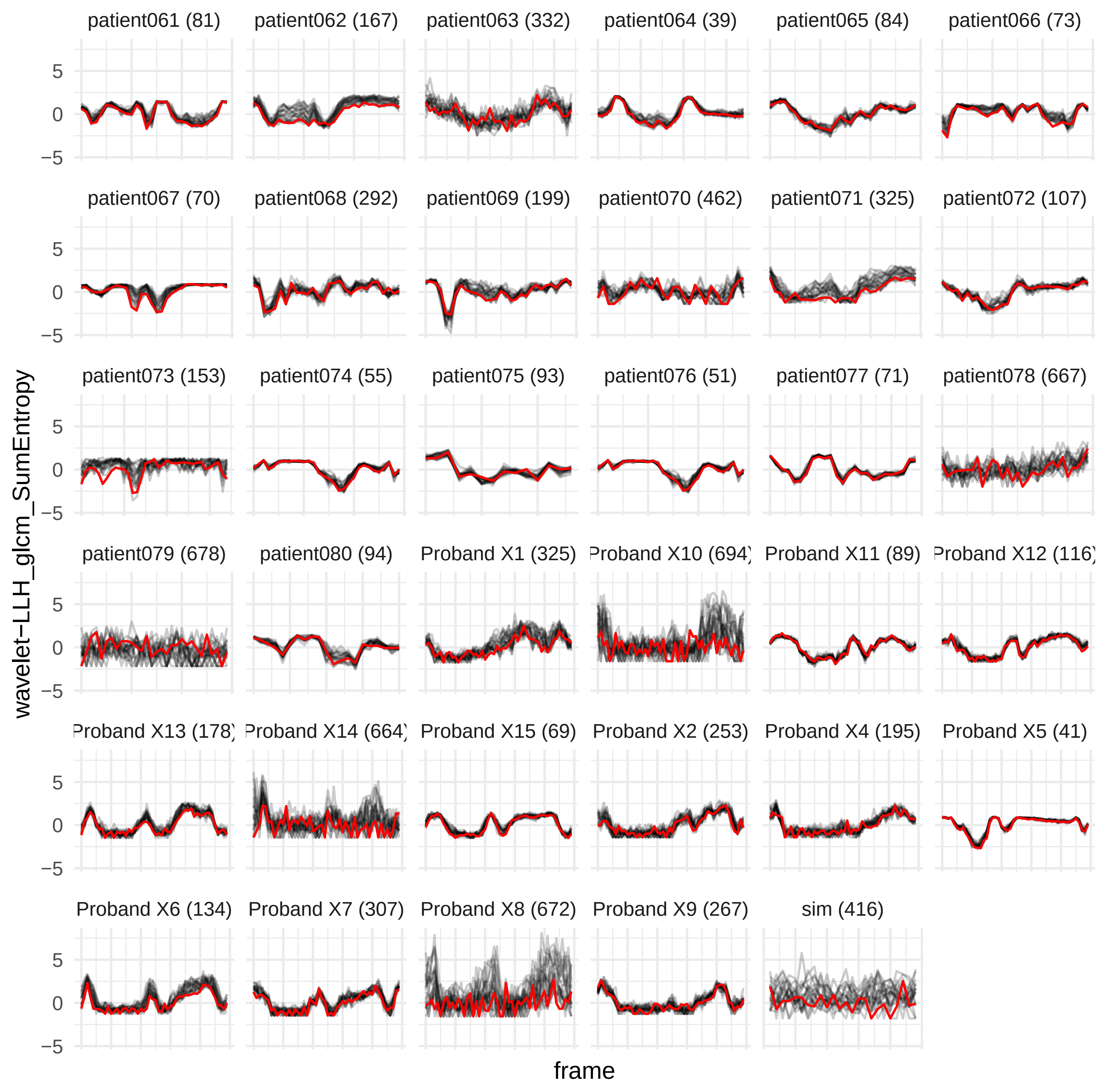

patient061 (81)
patient062 (167)
patient063 (332)
patient064 (39)
patient065 (84)
patient066 (73)
patient067 (70)
patient068 (292)
patient069 (199)
patient070 (462)
patient071 (325)
patient072 (107)
patient073 (153)
patient074 (55)
patient075 (93)
patient076 (51)
patient077 (71)
patient078 (667)
patient079 (678)
patient080 (94)
Proband X1 (325)
Proband X10 (694)
Proband X11 (89)
Proband X12 (116)
Proband X13 (178)
Proband X14 (664)
Proband X15 (69)
Proband X2 (253)
Proband X4 (195)
Proband X5 (41)
Proband X6 (134)
Proband X7 (307)
Proband X8 (672)
Proband X9 (267)
sim (416)
5
0
−5
wavelet−LLH_glcm_SumEntropy
frame

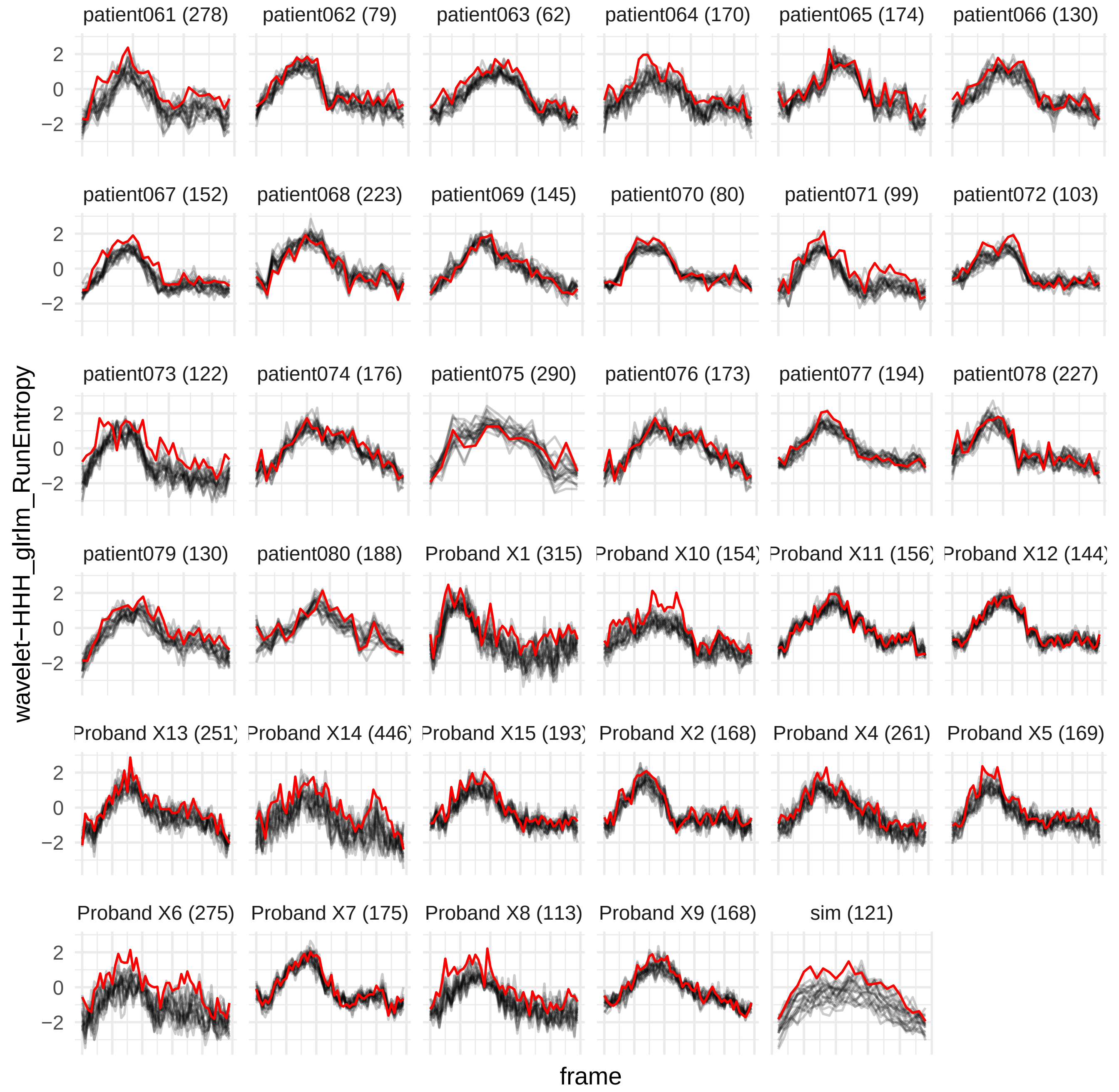

patient061 (278)
patient062 (79)
patient063 (62)
patient064 (170)
patient065 (174)
patient066 (130)
patient067 (152)
patient068 (223)
patient069 (145)
patient070 (80)
patient071 (99)
patient072 (103)
patient073 (122)
patient074 (176)
patient075 (290)
patient076 (173)
patient077 (194)
patient078 (227)
patient079 (130)
patient080 (188)
Proband X1 (315)
Proband X10 (154)
Proband X11 (156)
Proband X12 (144)
Proband X13 (251)
Proband X14 (446)
Proband X15 (193)
Proband X2 (168)
Proband X4 (261)
Proband X5 (169)
Proband X6 (275)
Proband X7 (175)
Proband X8 (113)
Proband X9 (168)
sim (121)
2
0
−2
wavelet−HHH_glrlm_RunEntropy
frame

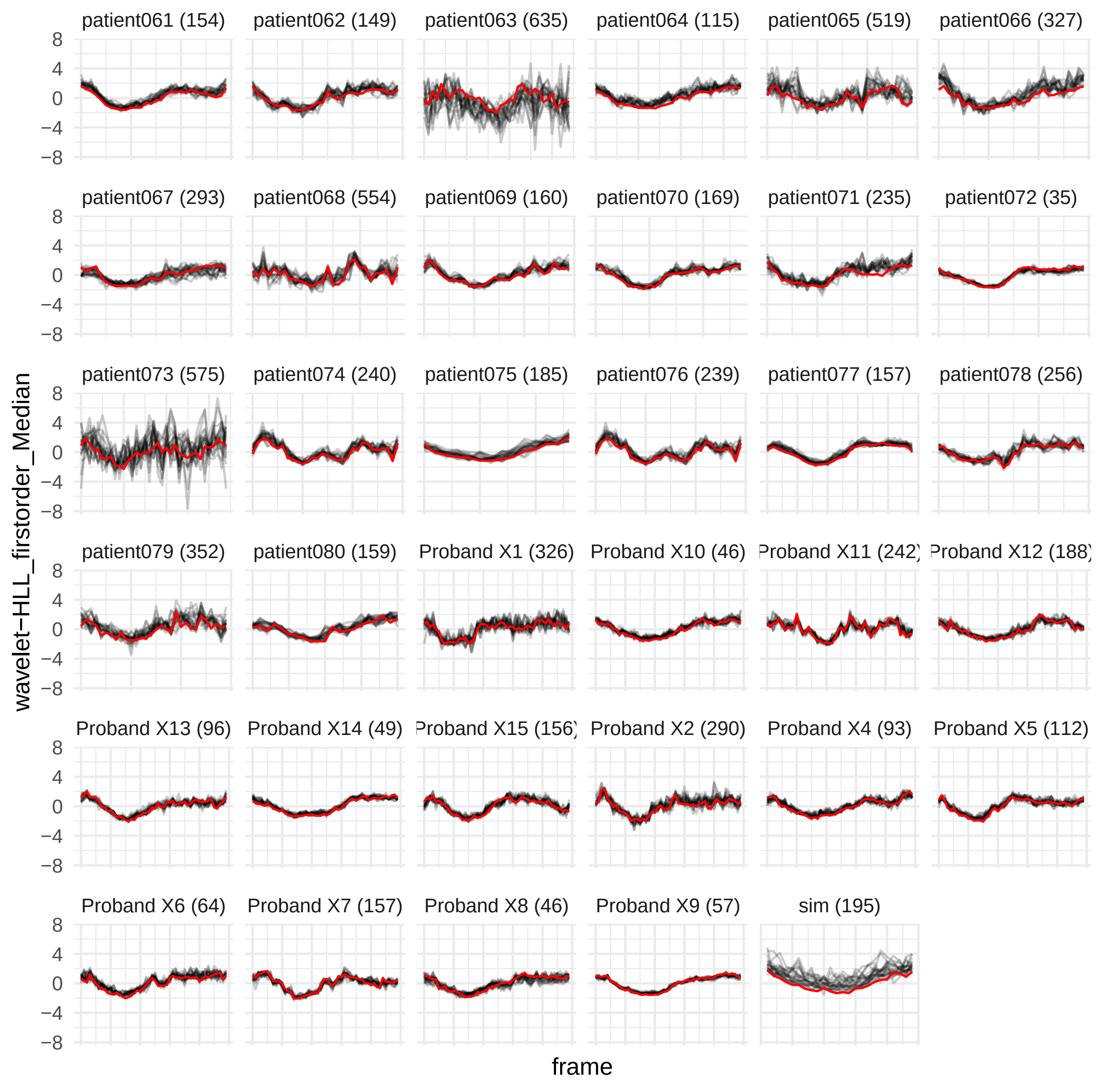
patient061 (154)
patient062 (149)
patient063 (635)
patient064 (115)
patient065 (519)
patient066 (327)
patient067 (293)
patient068 (554)
patient069 (160)
patient070 (169)
patient071 (235)
patient072 (35)
patient073 (575)
patient074 (240)
patient075 (185)
patient076 (239)
patient077 (157)
patient078 (256)
patient079 (352)
patient080 (159)
Proband X1 (326)
Proband X10 (46)
Proband X11 (242)
Proband X12 (188)
Proband X13 (96)
Proband X14 (49)
Proband X15 (156)
Proband X2 (290)
Proband X4 (93)
Proband X5 (112)
Proband X6 (64)
Proband X7 (157)
Proband X8 (46)
Proband X9 (57)
sim (195)
8
4
0
−4
−8
wavelet−HLL_firstorder_Median
frame

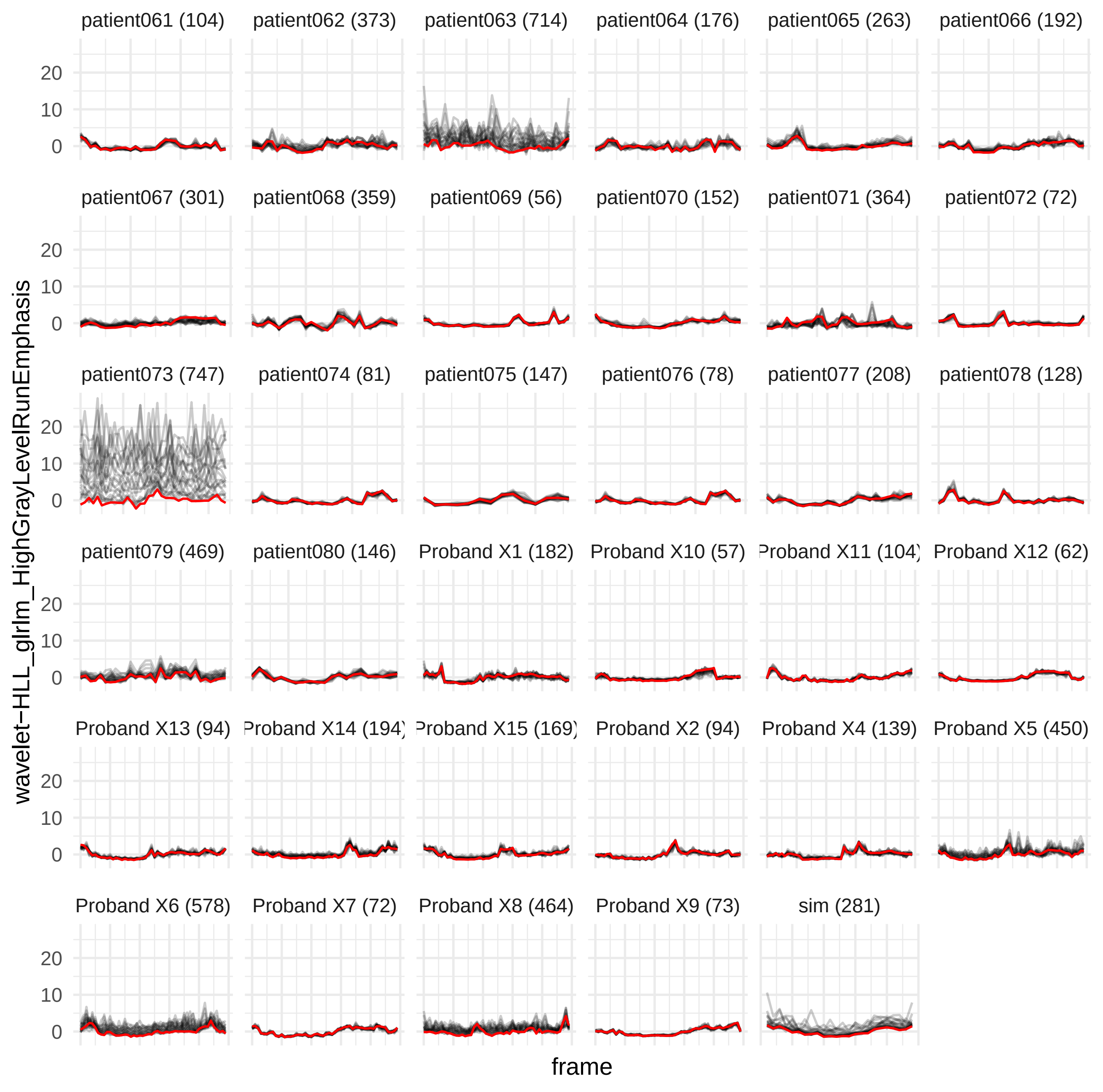
patient061 (104)
patient062 (373)
patient063 (714)
patient064 (176)
patient065 (263)
patient066 (192)
patient067 (301)
patient068 (359)
patient069 (56)
patient070 (152)
patient071 (364)
patient072 (72)
patient073 (747)
patient074 (81)
patient075 (147)
patient076 (78)
patient077 (208)
patient078 (128)
patient079 (469)
patient080 (146)
Proband X1 (182)
Proband X10 (57)
Proband X11 (104)
Proband X12 (62)
Proband X13 (94)
Proband X14 (194)
Proband X15 (169)
Proband X2 (94)
Proband X4 (139)
Proband X5 (450)
Proband X6 (578)
Proband X7 (72)
Proband X8 (464)
Proband X9 (73)
sim (281)
20
10
0
wavelet-HLL_glrlm_HighGrayLevelRunEmphasis
frame

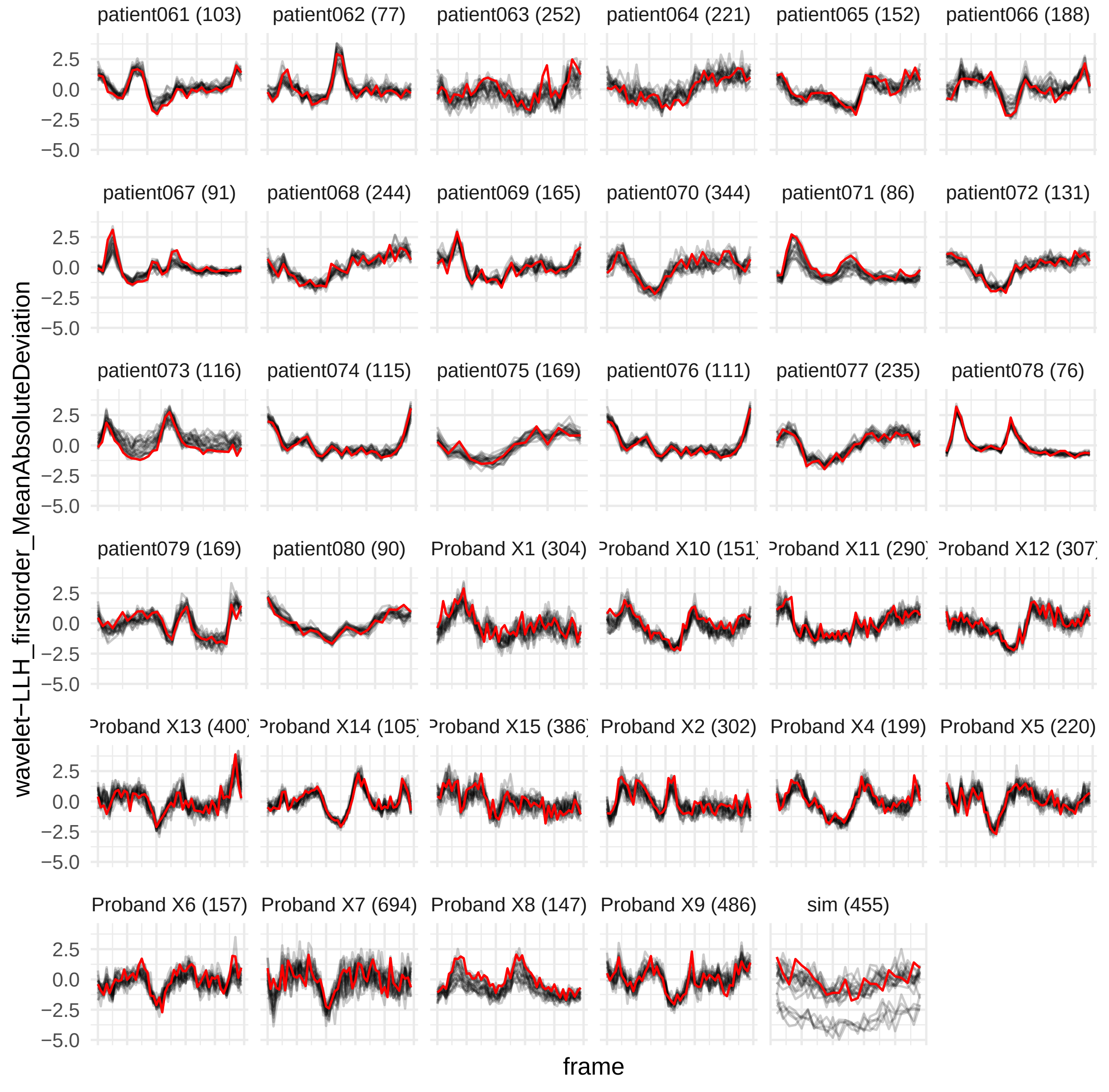

patient061 (103)
patient062 (77)
patient063 (252)
patient064 (221)
patient065 (152)
patient066 (188)
patient067 (91)
patient068 (244)
patient069 (165)
patient070 (344)
patient071 (86)
patient072 (131)
patient073 (116)
patient074 (115)
patient075 (169)
patient076 (111)
patient077 (235)
patient078 (76)
patient079 (169)
patient080 (90)
Proband X1 (304)
Proband X10 (151)
Proband X11 (290)
Proband X12 (307)
Proband X13 (400)
Proband X14 (105)
Proband X15 (386)
Proband X2 (302)
Proband X4 (199)
Proband X5 (220)
Proband X6 (157)
Proband X7 (694)
Proband X8 (147)
Proband X9 (486)
sim (455)
2.5
0.0
−2.5
−5.0
wavelet−LLH_firstorder_MeanAbsoluteDeviation
frame

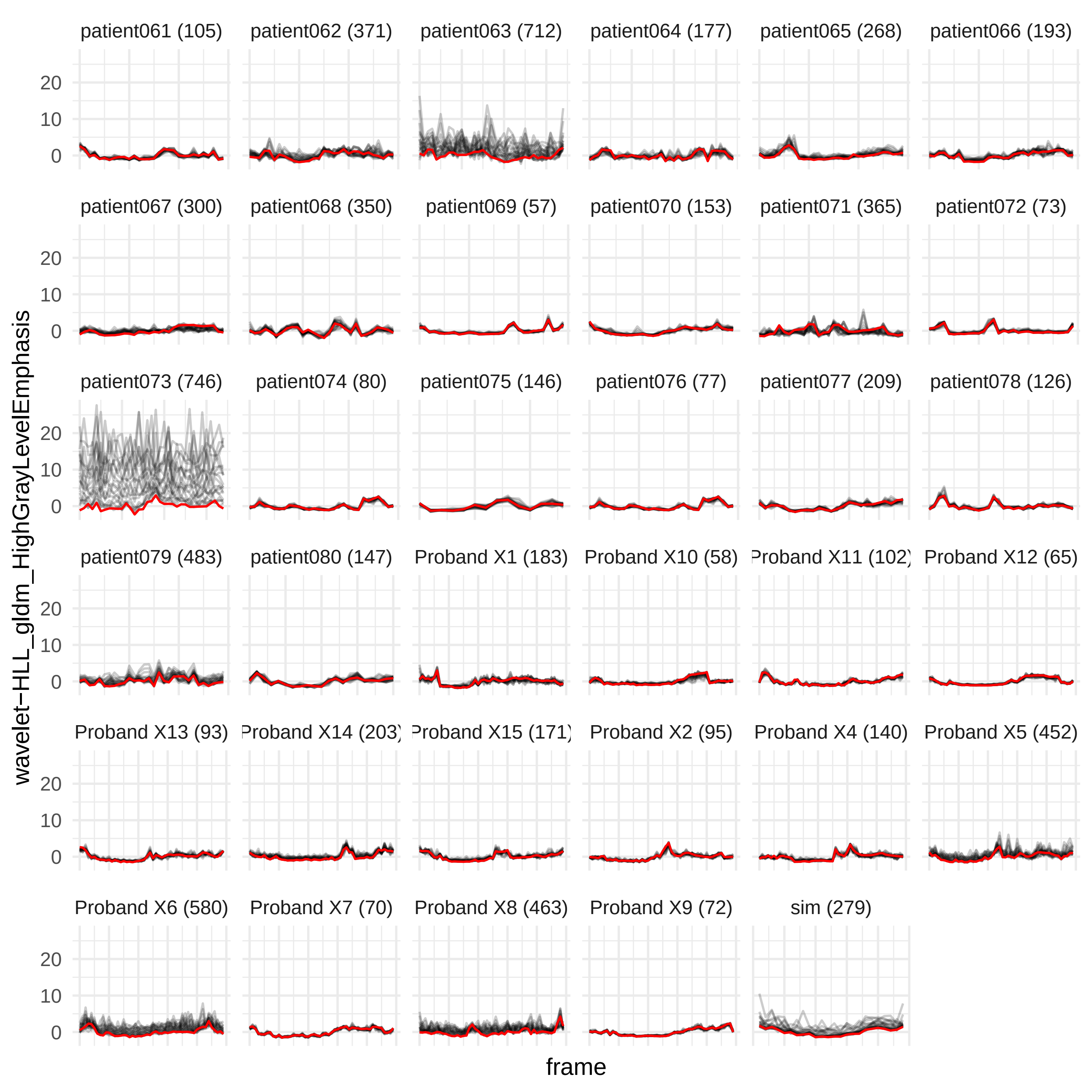
patient061 (105)
patient062 (371)
patient063 (712)
patient064 (177)
patient065 (268)
patient066 (193)
patient067 (300)
patient068 (350)
patient069 (57)
patient070 (153)
patient071 (365)
patient072 (73)
patient073 (746)
patient074 (80)
patient075 (146)
patient076 (77)
patient077 (209)
patient078 (126)
patient079 (483)
patient080 (147)
Proband X1 (183)
Proband X10 (58)
Proband X11 (102)
Proband X12 (65)
Proband X13 (93)
Proband X14 (203)
Proband X15 (171)
Proband X2 (95)
Proband X4 (140)
Proband X5 (452)
Proband X6 (580)
Proband X7 (70)
Proband X8 (463)
Proband X9 (72)
sim (279)
0
10
20
wavelet−HLL_gldm_HighGrayLevelEmphasis
frame

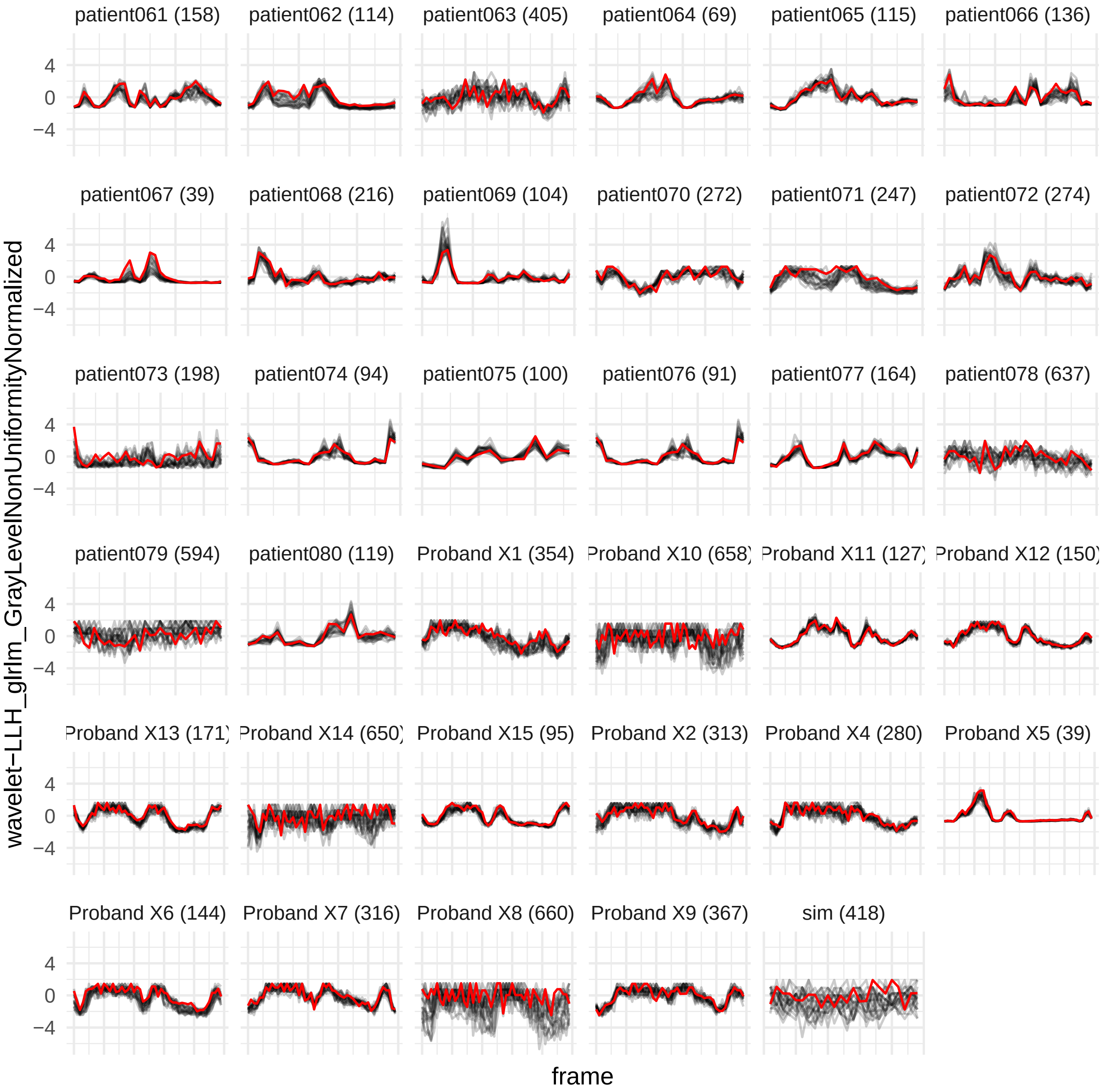

patient061 (158)
patient062 (114)
patient063 (405)
patient064 (69)
patient065 (115)
patient066 (136)
patient067 (39)
patient068 (216)
patient069 (104)
patient070 (272)
patient071 (247)
patient072 (274)
patient073 (198)
patient074 (94)
patient075 (100)
patient076 (91)
patient077 (164)
patient078 (637)
patient079 (594)
patient080 (119)
Proband X1 (354)
Proband X10 (658)
Proband X11 (127)
Proband X12 (150)
Proband X13 (171)
Proband X14 (650)
Proband X15 (95)
Proband X2 (313)
Proband X4 (280)
Proband X5 (39)
Proband X6 (144)
Proband X7 (316)
Proband X8 (660)
Proband X9 (367)
sim (418)
4
0
−4
wavelet−LLH_glrlm_GrayLevelNonUniformityNormalized
frame

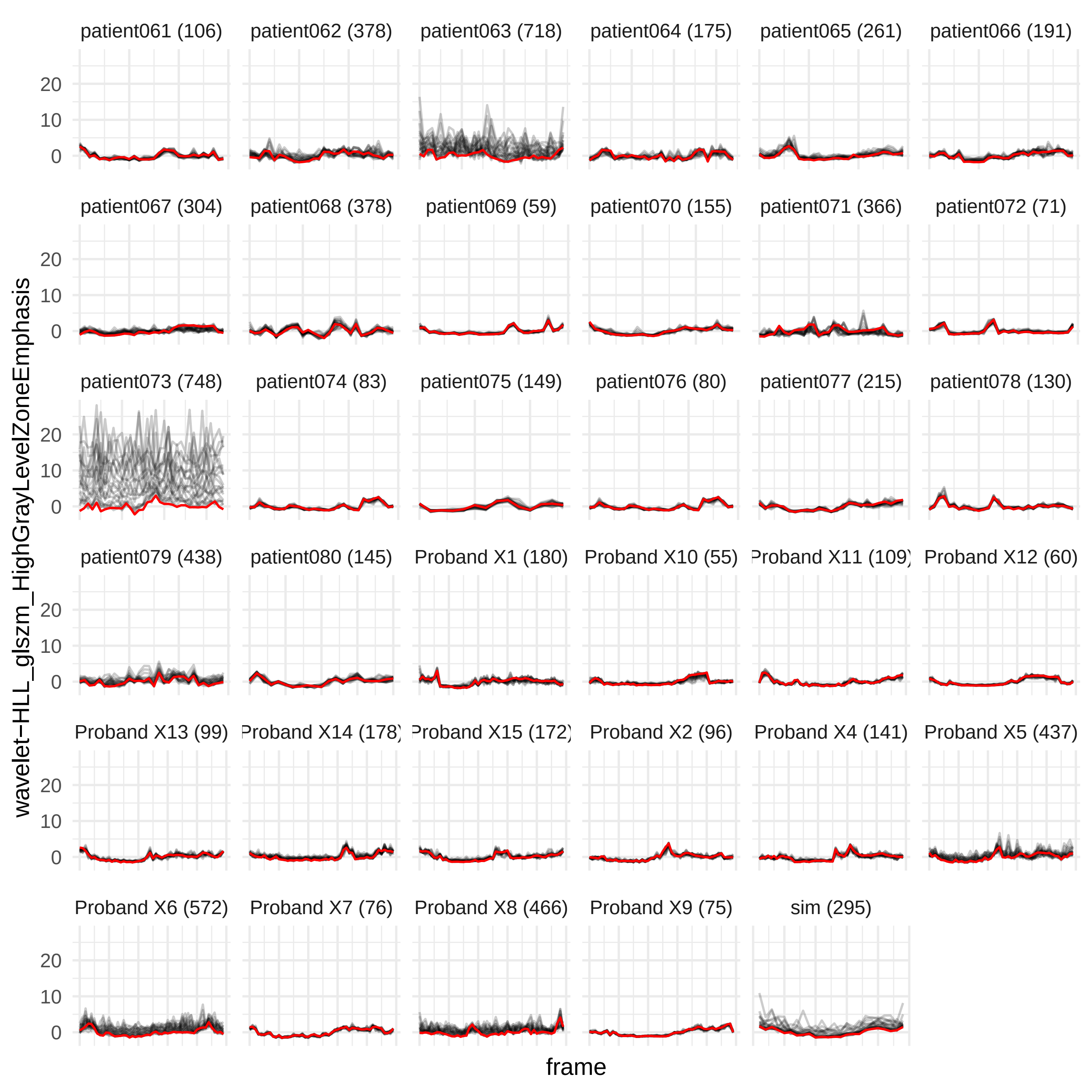

patient061 (106)
patient062 (378)
patient063 (718)
patient064 (175)
patient065 (261)
patient066 (191)
patient067 (304)
patient068 (378)
patient069 (59)
patient070 (155)
patient071 (366)
patient072 (71)
patient073 (748)
patient074 (83)
patient075 (149)
patient076 (80)
patient077 (215)
patient078 (130)
patient079 (438)
patient080 (145)
Proband X1 (180)
Proband X10 (55)
Proband X11 (109)
Proband X12 (60)
Proband X13 (99)
Proband X14 (178)
Proband X15 (172)
Proband X2 (96)
Proband X4 (141)
Proband X5 (437)
Proband X6 (572)
Proband X7 (76)
Proband X8 (466)
Proband X9 (75)
sim (295)
0
10
20
wavelet−HLL_glszm_HighGrayLevelZoneEmphasis
frame

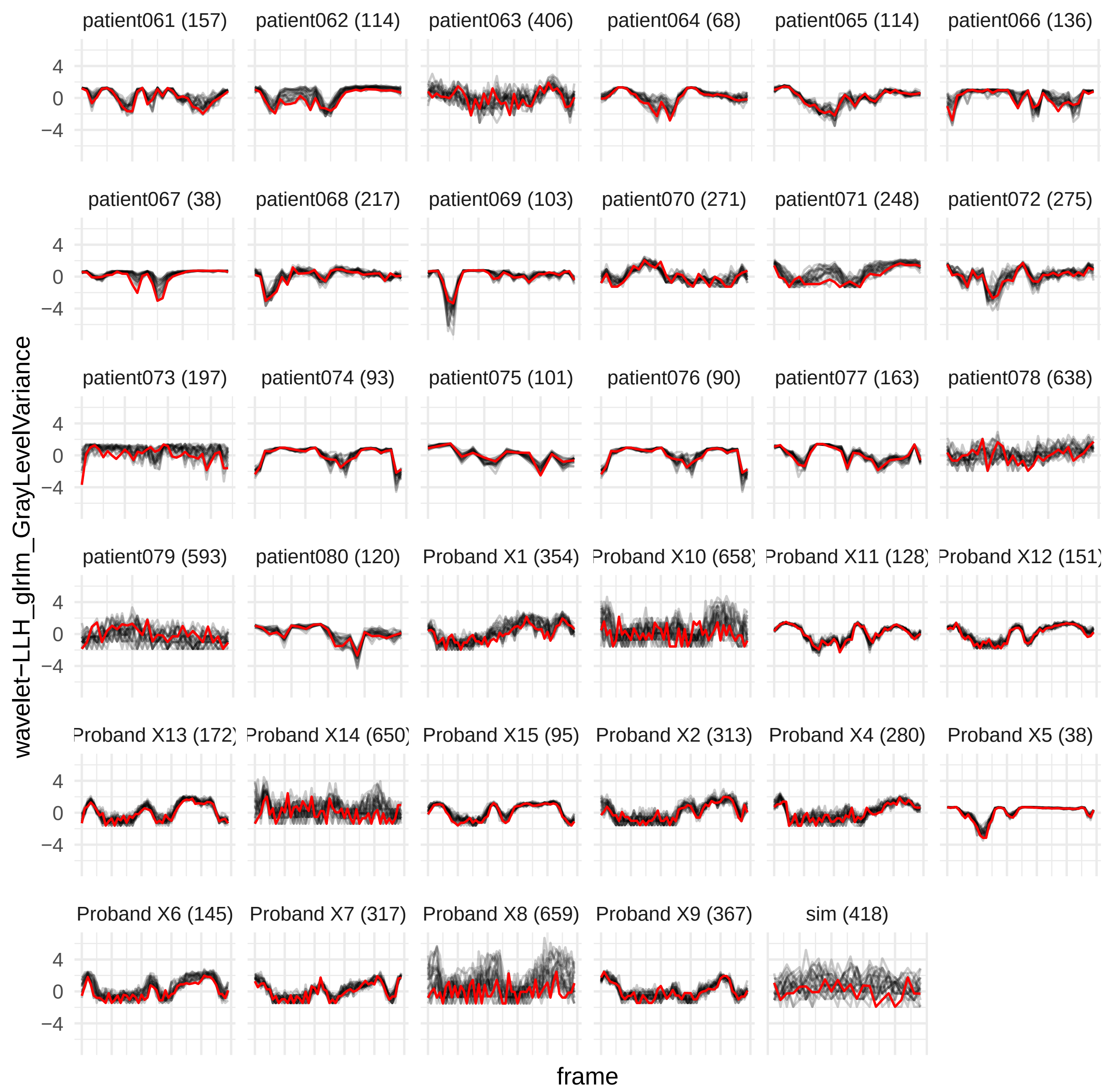

patient061 (157)
patient062 (114)
patient063 (406)
patient064 (68)
patient065 (114)
patient066 (136)
patient067 (38)
patient068 (217)
patient069 (103)
patient070 (271)
patient071 (248)
patient072 (275)
patient073 (197)
patient074 (93)
patient075 (101)
patient076 (90)
patient077 (163)
patient078 (638)
patient079 (593)
patient080 (120)
Proband X1 (354)
Proband X10 (658)
Proband X11 (128)
Proband X12 (151)
Proband X13 (172)
Proband X14 (650)
Proband X15 (95)
Proband X2 (313)
Proband X4 (280)
Proband X5 (38)
Proband X6 (145)
Proband X7 (317)
Proband X8 (659)
Proband X9 (367)
sim (418)
4
0
−4
wavelet−LLH_glrlm_GrayLevelVariance
frame

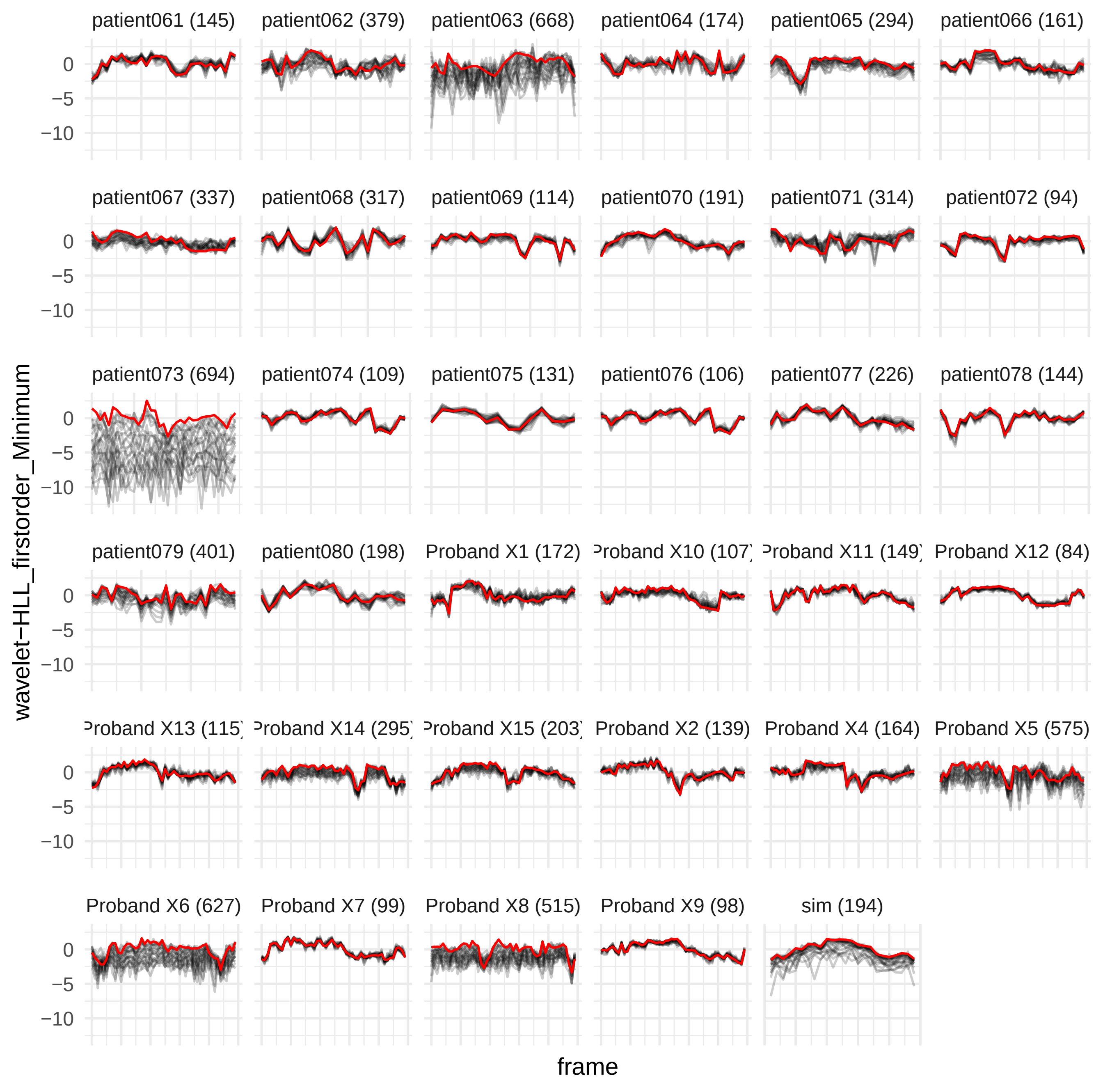

patient061 (145)
patient062 (379)
patient063 (668)
patient064 (174)
patient065 (294)
patient066 (161)
patient067 (337)
patient068 (317)
patient069 (114)
patient070 (191)
patient071 (314)
patient072 (94)
patient073 (694)
patient074 (109)
patient075 (131)
patient076 (106)
patient077 (226)
patient078 (144)
patient079 (401)
patient080 (198)
Proband X1 (172)
Proband X10 (107)
Proband X11 (149)
Proband X12 (84)
Proband X13 (115)
Proband X14 (295)
Proband X15 (203)
Proband X2 (139)
Proband X4 (164)
Proband X5 (575)
Proband X6 (627)
Proband X7 (99)
Proband X8 (515)
Proband X9 (98)
sim (194)
0
−5
−10
wavelet−HLL_firstorder_Minimum
frame

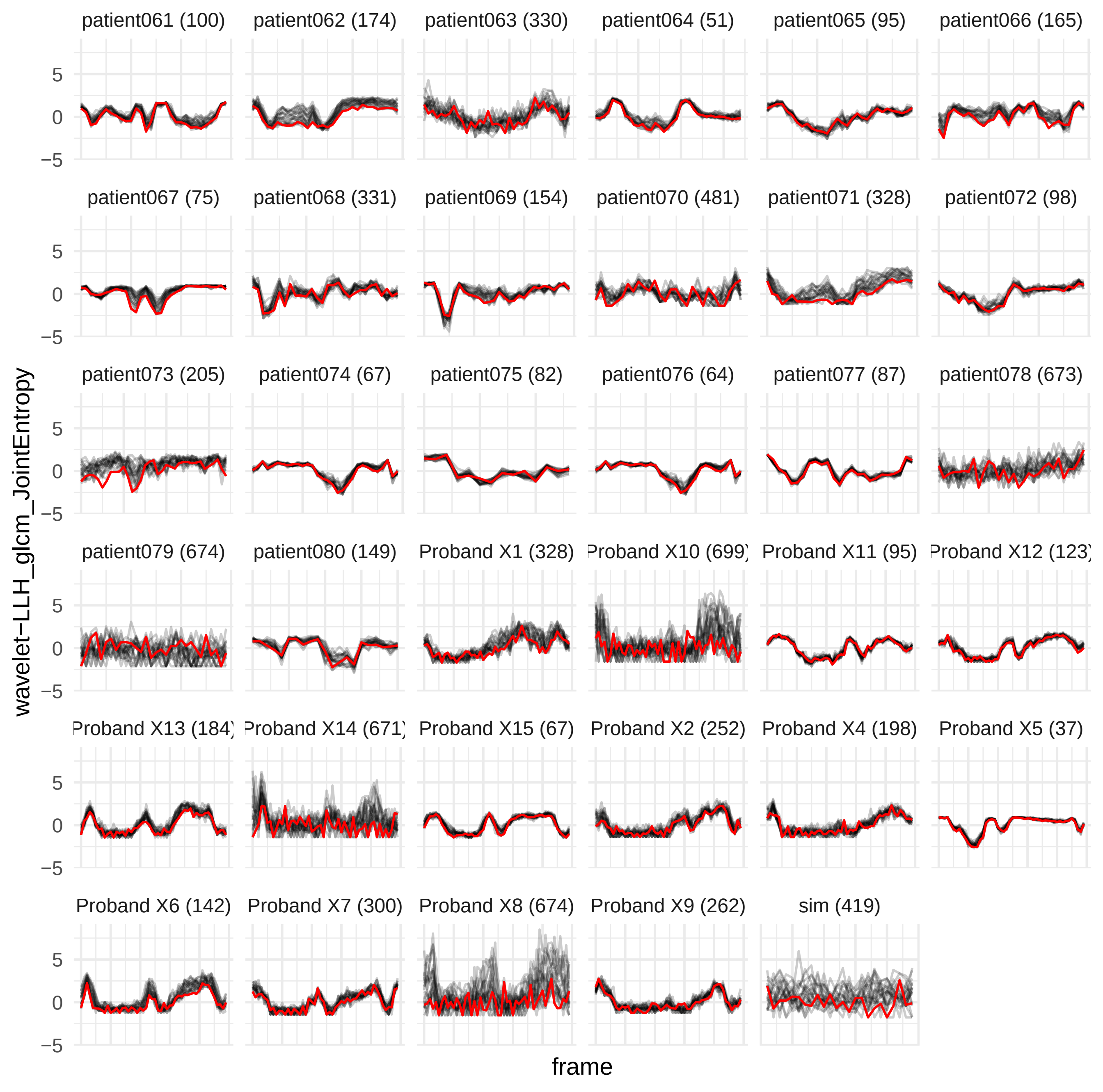
patient061 (100)
patient062 (174)
patient063 (330)
patient064 (51)
patient065 (95)
patient066 (165)
patient067 (75)
patient068 (331)
patient069 (154)
patient070 (481)
patient071 (328)
patient072 (98)
patient073 (205)
patient074 (67)
patient075 (82)
patient076 (64)
patient077 (87)
patient078 (673)
patient079 (674)
patient080 (149)
Proband X1 (328)
Proband X10 (699)
Proband X11 (95)
Proband X12 (123)
Proband X13 (184)
Proband X14 (671)
Proband X15 (67)
Proband X2 (252)
Proband X4 (198)
Proband X5 (37)
Proband X6 (142)
Proband X7 (300)
Proband X8 (674)
Proband X9 (262)
sim (419)
wavelet−LLH_glcm_JointEntropy
frame
5
0
−5

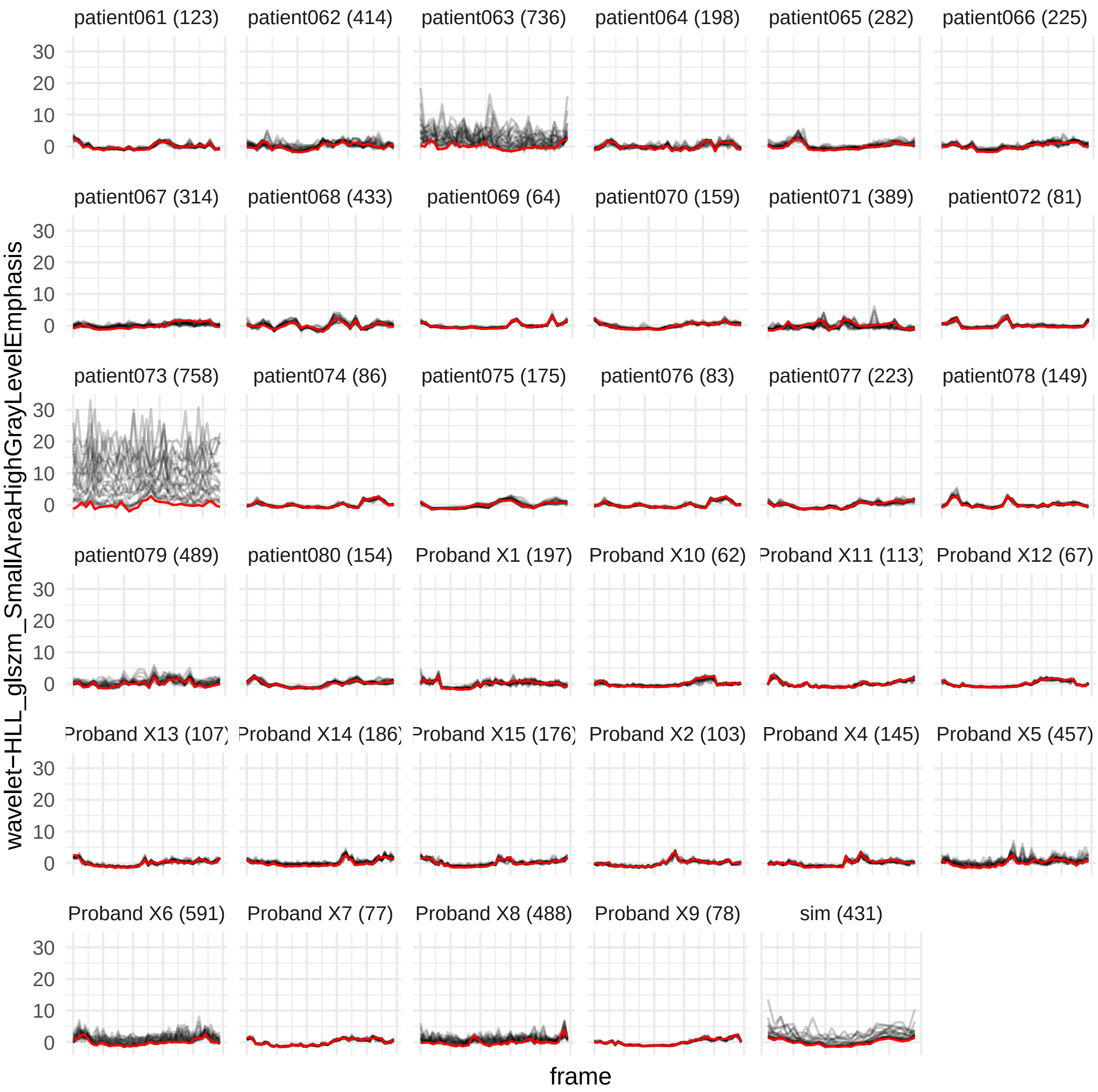
patient061 (123)
patient062 (414)
patient063 (736)
patient064 (198)
patient065 (282)
patient066 (225)
patient067 (314)
patient068 (433)
patient069 (64)
patient070 (159)
patient071 (389)
patient072 (81)
patient073 (758)
patient074 (86)
patient075 (175)
patient076 (83)
patient077 (223)
patient078 (149)
patient079 (489)
patient080 (154)
Proband X1 (197)
Proband X10 (62)
Proband X11 (113)
Proband X12 (67)
Proband X13 (107)
Proband X14 (186)
Proband X15 (176)
Proband X2 (103)
Proband X4 (145)
Proband X5 (457)
Proband X6 (591)
Proband X7 (77)
Proband X8 (488)
Proband X9 (78)
sim (431)
wavelet−HLL_glszm_SmallAreaHighGrayLevelEmphasis
frame
0
10
20
30

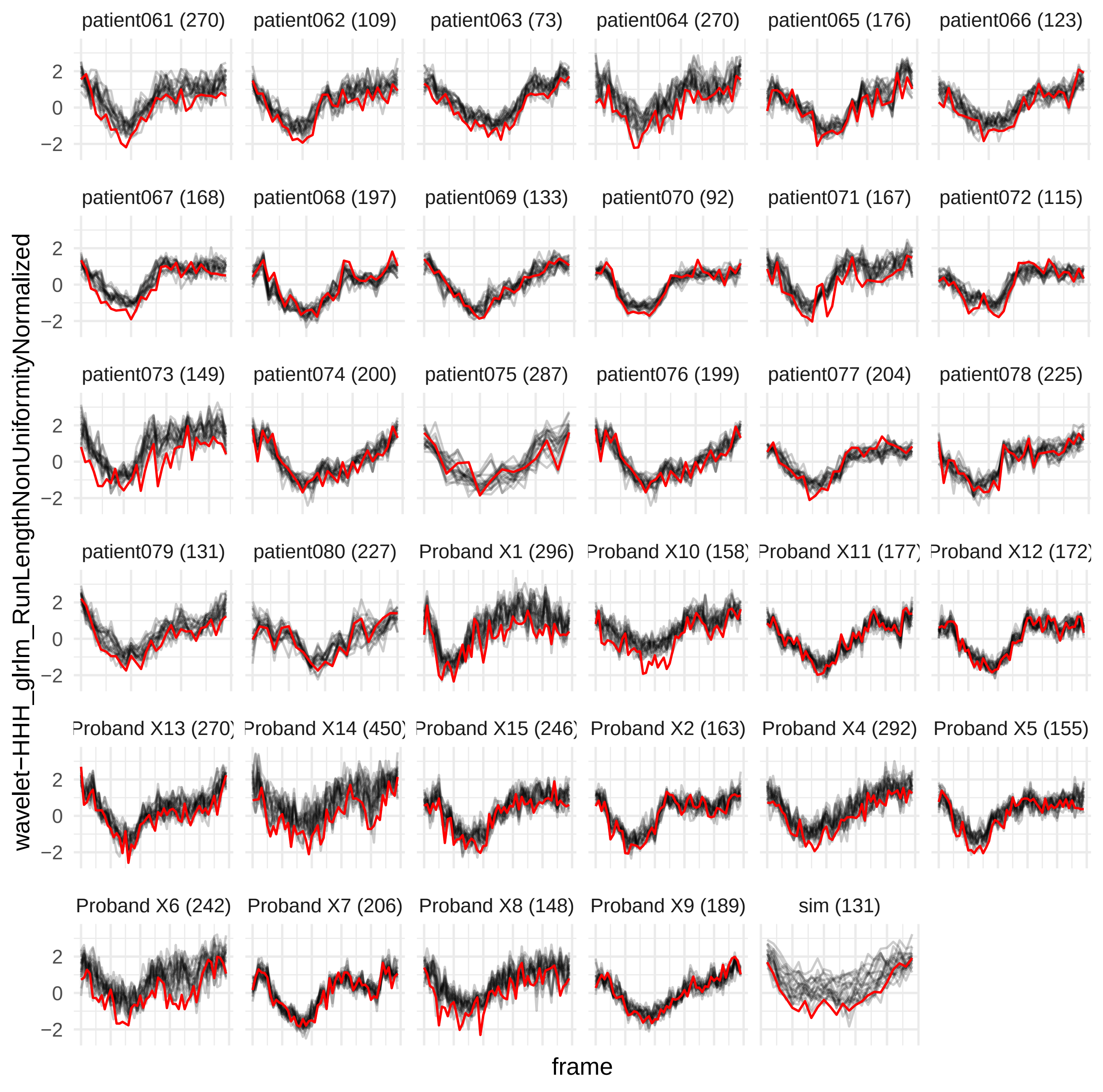

patient061 (270)
patient062 (109)
patient063 (73)
patient064 (270)
patient065 (176)
patient066 (123)
patient067 (168)
patient068 (197)
patient069 (133)
patient070 (92)
patient071 (167)
patient072 (115)
patient073 (149)
patient074 (200)
patient075 (287)
patient076 (199)
patient077 (204)
patient078 (225)
patient079 (131)
patient080 (227)
Proband X1 (296)
Proband X10 (158)
Proband X11 (177)
Proband X12 (172)
Proband X13 (270)
Proband X14 (450)
Proband X15 (246)
Proband X2 (163)
Proband X4 (292)
Proband X5 (155)
Proband X6 (242)
Proband X7 (206)
Proband X8 (148)
Proband X9 (189)
sim (131)
2
0
−2
wavelet−HHH_glrlm_RunLengthNonUniformityNormalized
frame

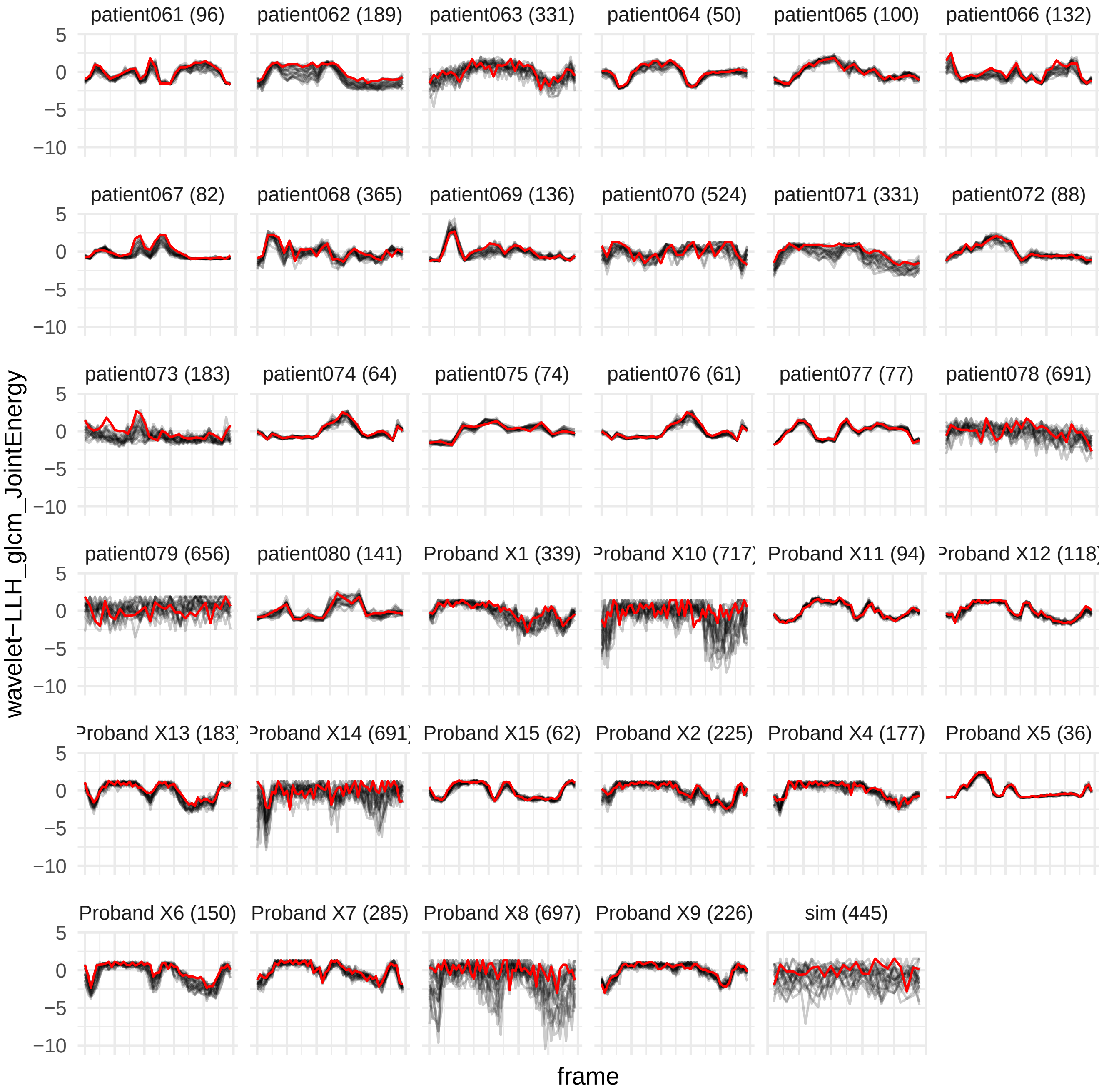

patient061 (96)
patient062 (189)
patient063 (331)
patient064 (50)
patient065 (100)
patient066 (132)
patient067 (82)
patient068 (365)
patient069 (136)
patient070 (524)
patient071 (331)
patient072 (88)
patient073 (183)
patient074 (64)
patient075 (74)
patient076 (61)
patient077 (77)
patient078 (691)
patient079 (656)
patient080 (141)
Proband X1 (339)
Proband X10 (717)
Proband X11 (94)
Proband X12 (118)
Proband X13 (183)
Proband X14 (691)
Proband X15 (62)
Proband X2 (225)
Proband X4 (177)
Proband X5 (36)
Proband X6 (150)
Proband X7 (285)
Proband X8 (697)
Proband X9 (226)
sim (445)
5
0
−5
−10
wavelet−LLH_glcm_JointEnergy
frame

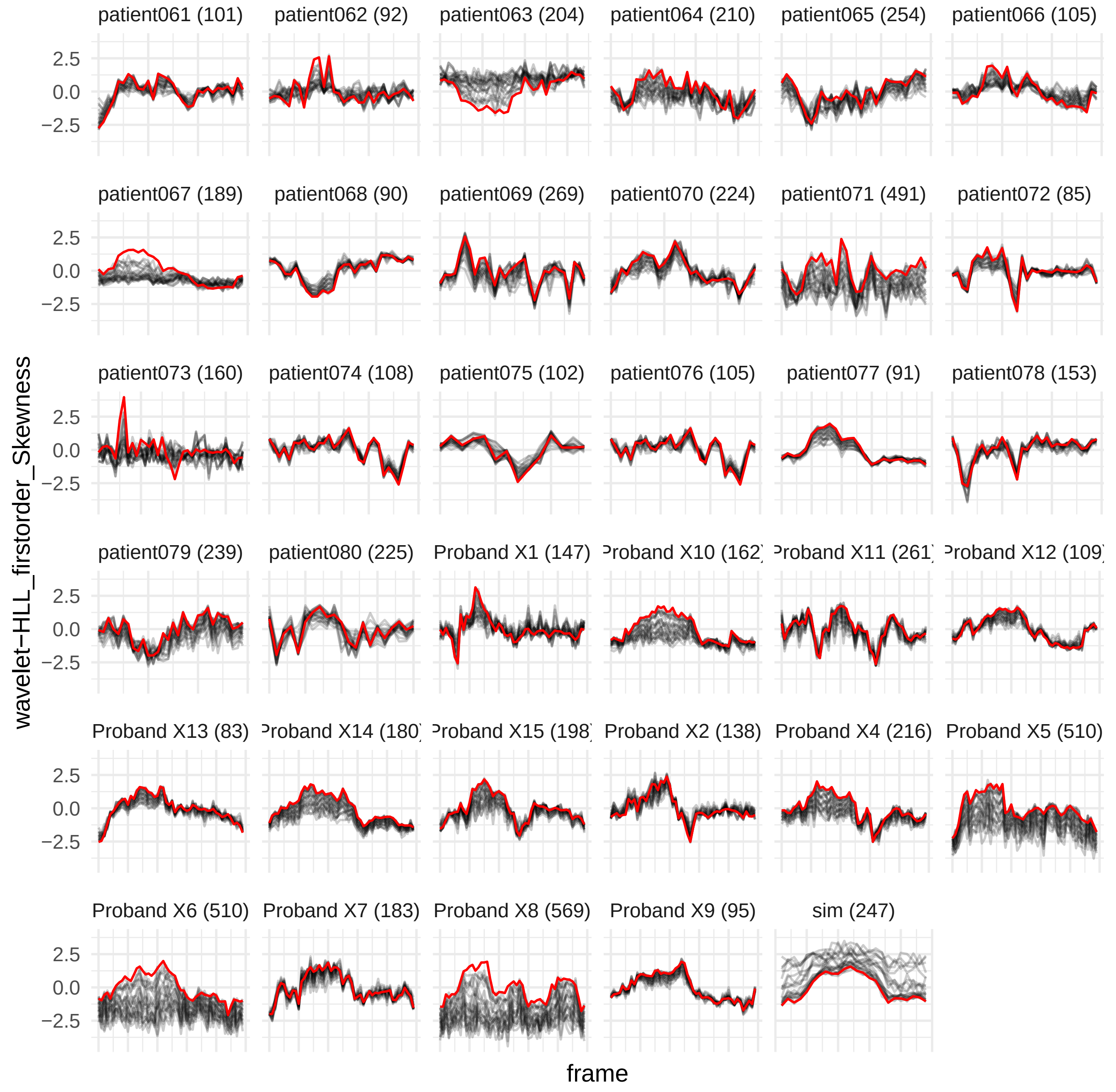

patient061 (101)
patient062 (92)
patient063 (204)
patient064 (210)
patient065 (254)
patient066 (105)
patient067 (189)
patient068 (90)
patient069 (269)
patient070 (224)
patient071 (491)
patient072 (85)
patient073 (160)
patient074 (108)
patient075 (102)
patient076 (105)
patient077 (91)
patient078 (153)
patient079 (239)
patient080 (225)
Proband X1 (147)
Proband X10 (162)
Proband X11 (261)
Proband X12 (109)
Proband X13 (83)
Proband X14 (180)
Proband X15 (198)
Proband X2 (138)
Proband X4 (216)
Proband X5 (510)
Proband X6 (510)
Proband X7 (183)
Proband X8 (569)
Proband X9 (95)
sim (247)
2.5
0.0
−2.5
wavelet−HLL_firstorder_Skewness
frame

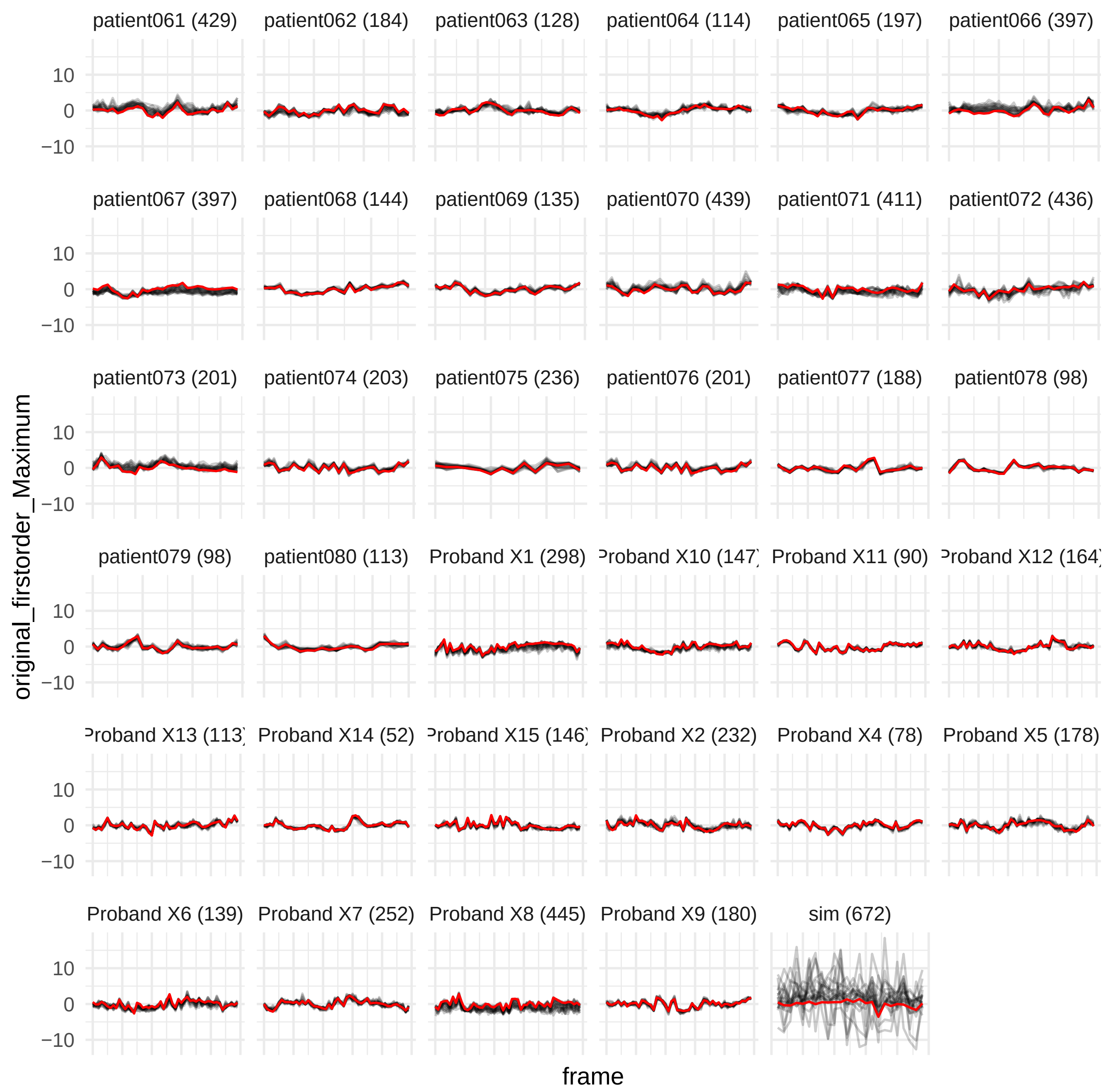

patient061 (429)
patient062 (184)
patient063 (128)
patient064 (114)
patient065 (197)
patient066 (397)
patient067 (397)
patient068 (144)
patient069 (135)
patient070 (439)
patient071 (411)
patient072 (436)
patient073 (201)
patient074 (203)
patient075 (236)
patient076 (201)
patient077 (188)
patient078 (98)
patient079 (98)
patient080 (113)
Proband X1 (298)
Proband X10 (147)
Proband X11 (90)
Proband X12 (164)
Proband X13 (113)
Proband X14 (52)
Proband X15 (146)
Proband X2 (232)
Proband X4 (78)
Proband X5 (178)
Proband X6 (139)
Proband X7 (252)
Proband X8 (445)
Proband X9 (180)
sim (672)
original_firstorder_Maximum
10
0
−10
frame

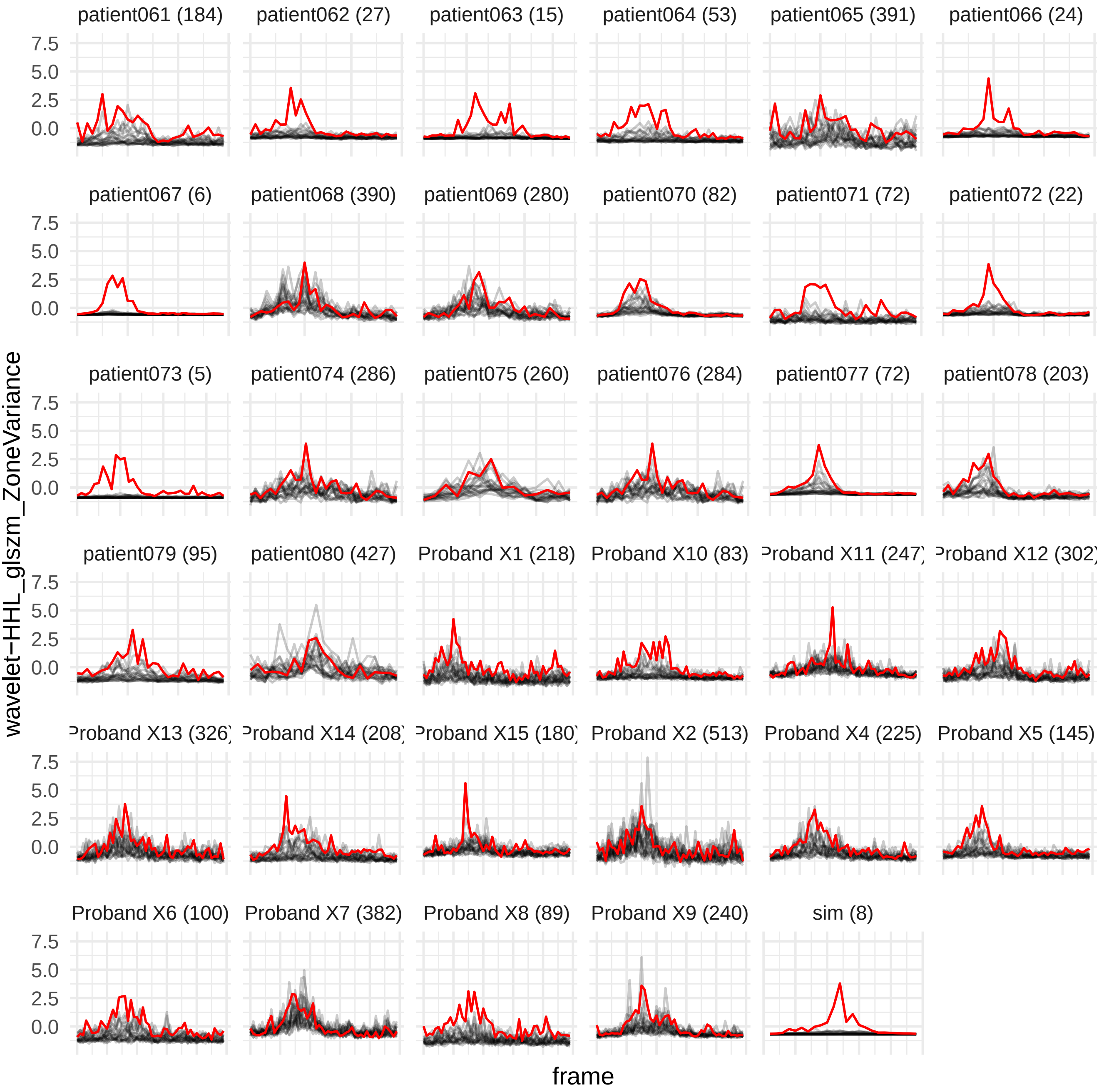

patient061 (184)
patient062 (27)
patient063 (15)
patient064 (53)
patient065 (391)
patient066 (24)
patient067 (6)
patient068 (390)
patient069 (280)
patient070 (82)
patient071 (72)
patient072 (22)
patient073 (5)
patient074 (286)
patient075 (260)
patient076 (284)
patient077 (72)
patient078 (203)
patient079 (95)
patient080 (427)
Proband X1 (218)
Proband X10 (83)
Proband X11 (247)
Proband X12 (302)
Proband X13 (326)
Proband X14 (208)
Proband X15 (180)
Proband X2 (513)
Proband X4 (225)
Proband X5 (145)
Proband X6 (100)
Proband X7 (382)
Proband X8 (89)
Proband X9 (240)
sim (8)
7.5
5.0
2.5
0.0
wavelet−HHL_glszm_ZoneVariance
frame

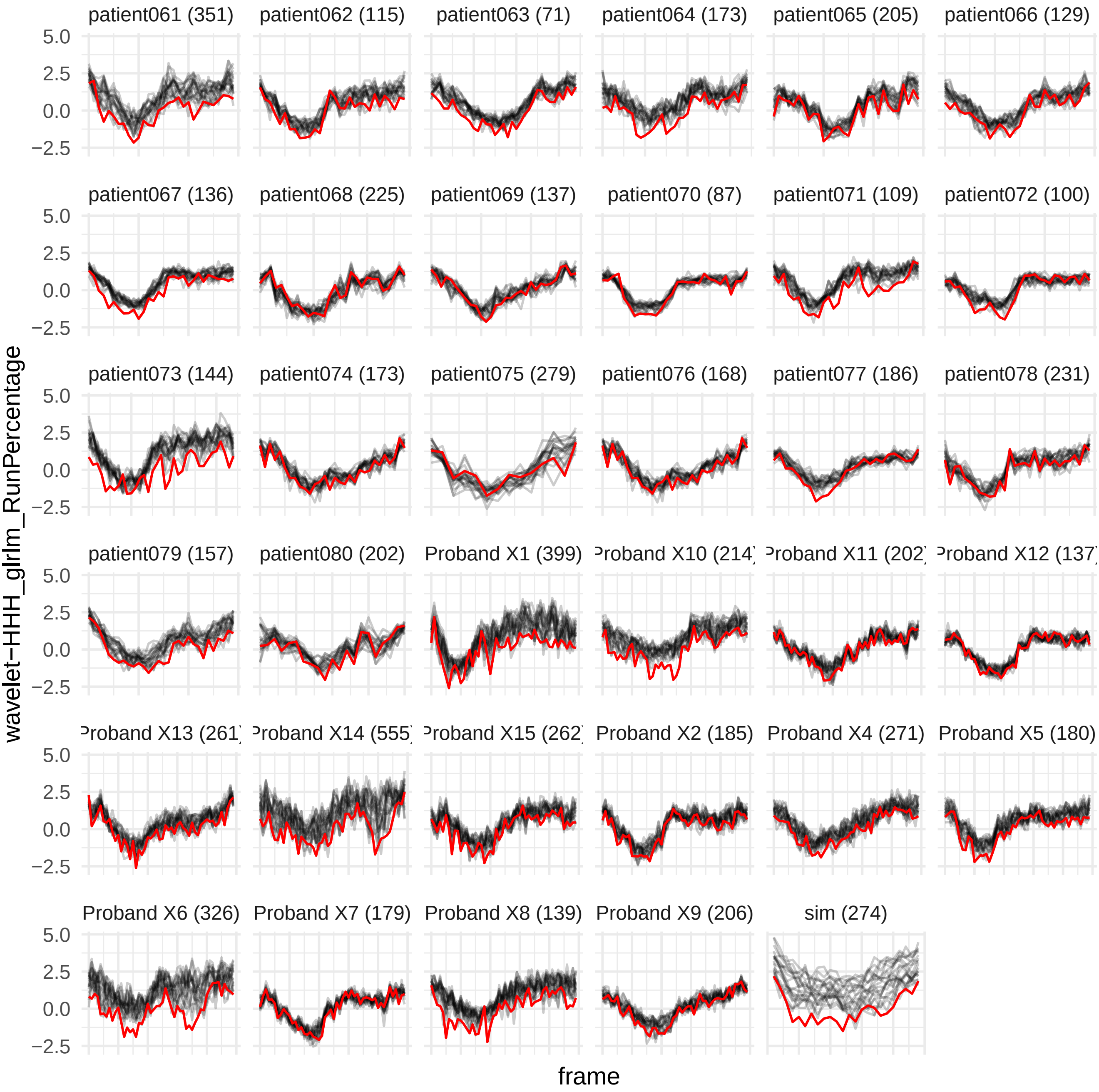

patient061 (351)
patient062 (115)
patient063 (71)
patient064 (173)
patient065 (205)
patient066 (129)
patient067 (136)
patient068 (225)
patient069 (137)
patient070 (87)
patient071 (109)
patient072 (100)
patient073 (144)
patient074 (173)
patient075 (279)
patient076 (168)
patient077 (186)
patient078 (231)
patient079 (157)
patient080 (202)
Proband X1 (399)
Proband X10 (214)
Proband X11 (202)
Proband X12 (137)
Proband X13 (261)
Proband X14 (555)
Proband X15 (262)
Proband X2 (185)
Proband X4 (271)
Proband X5 (180)
Proband X6 (326)
Proband X7 (179)
Proband X8 (139)
Proband X9 (206)
sim (274)
5.0
2.5
0.0
−2.5
wavelet−HHH_glrlm_RunPercentage
frame

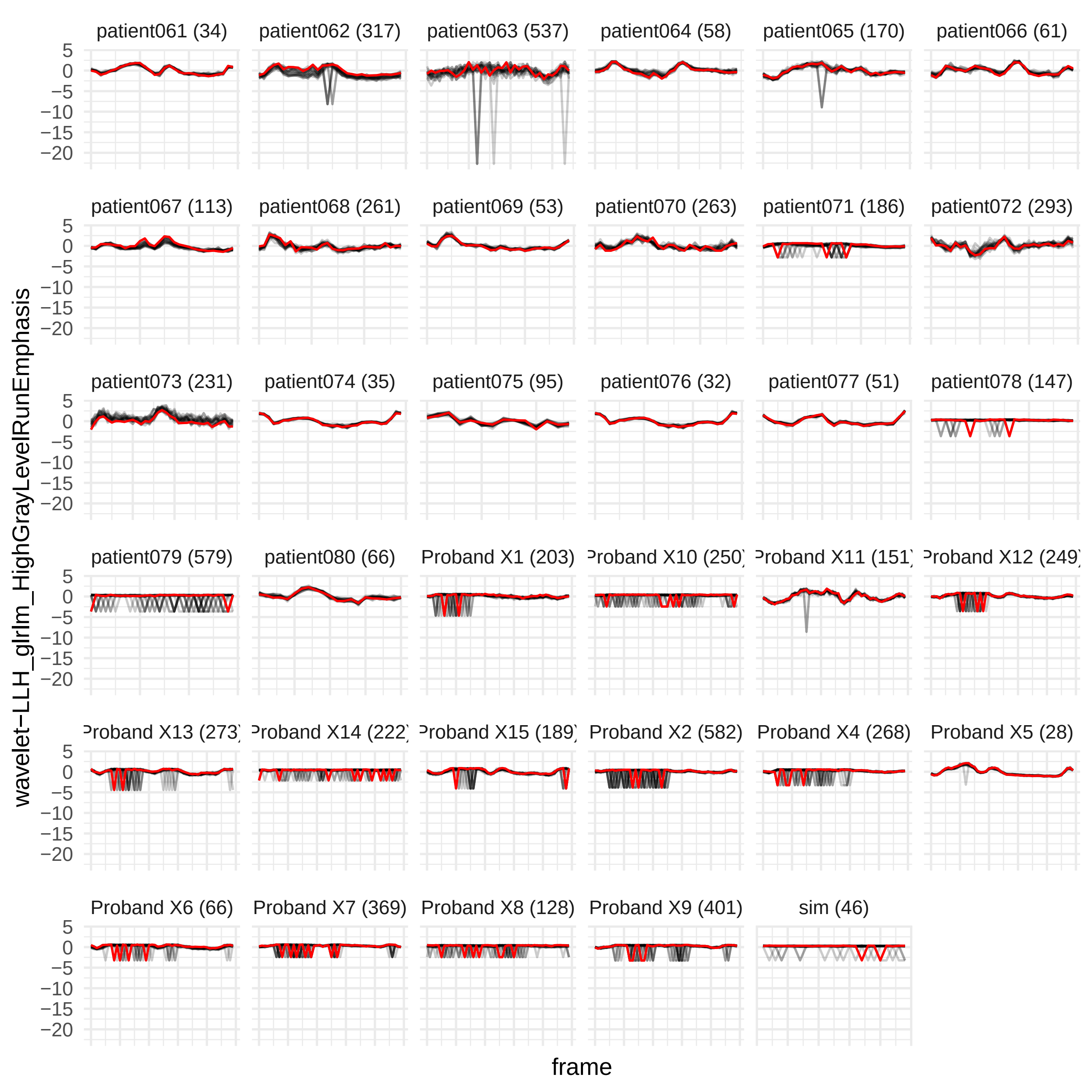
patient061 (34)
patient062 (317)
patient063 (537)
patient064 (58)
patient065 (170)
patient066 (61)
patient067 (113)
patient068 (261)
patient069 (53)
patient070 (263)
patient071 (186)
patient072 (293)
patient073 (231)
patient074 (35)
patient075 (95)
patient076 (32)
patient077 (51)
patient078 (147)
patient079 (579)
patient080 (66)
Proband X1 (203)
Proband X10 (250)
Proband X11 (151)
Proband X12 (249)
Proband X13 (273)
Proband X14 (222)
Proband X15 (189)
Proband X2 (582)
Proband X4 (268)
Proband X5 (28)
Proband X6 (66)
Proband X7 (369)
Proband X8 (128)
Proband X9 (401)
sim (46)
5
0
−5
−10
−15
−20
wavelet−LLH_glrlm_HighGrayLevelRunEmphasis
frame

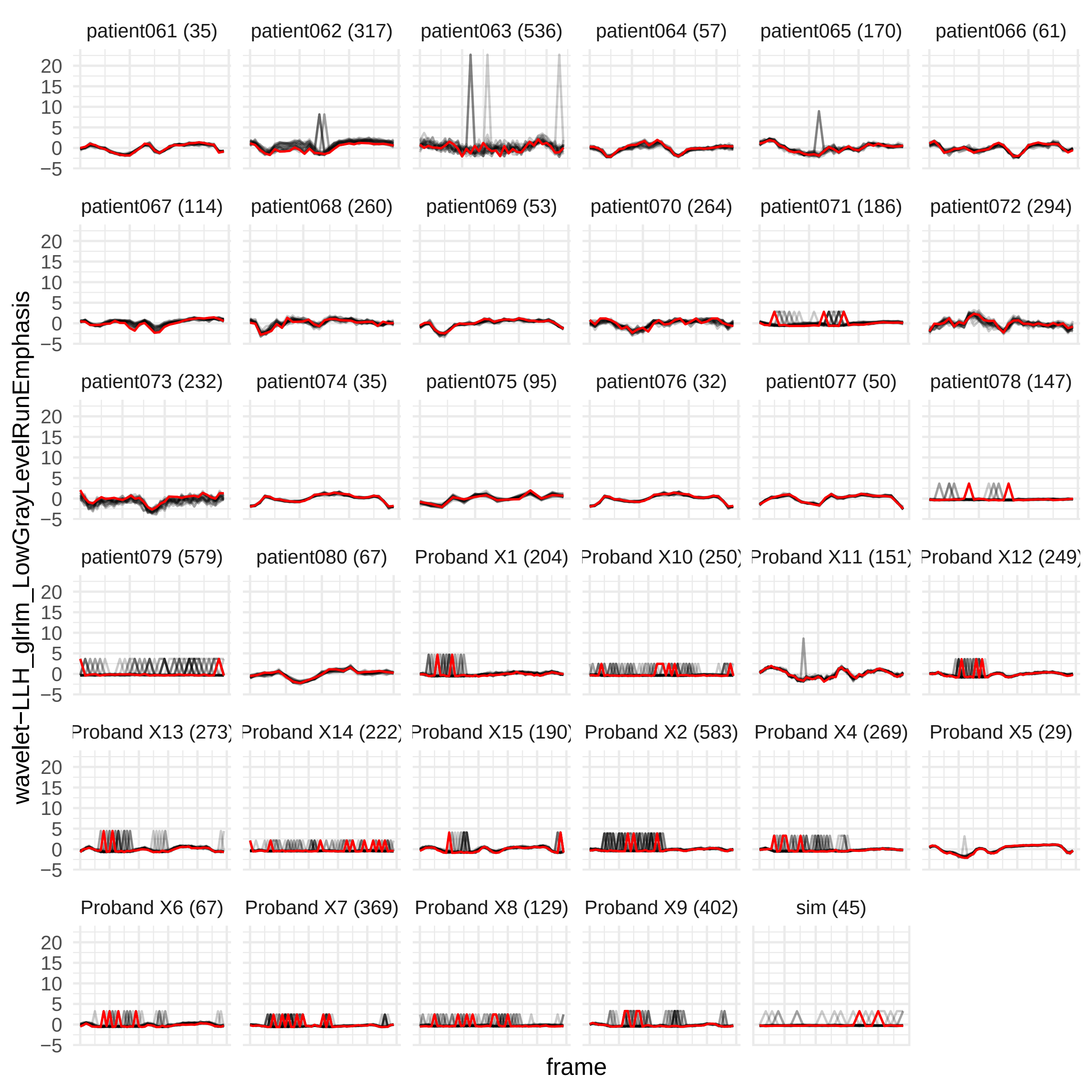
patient061 (35)
patient062 (317)
patient063 (536)
patient064 (57)
patient065 (170)
patient066 (61)
patient067 (114)
patient068 (260)
patient069 (53)
patient070 (264)
patient071 (186)
patient072 (294)
patient073 (232)
patient074 (35)
patient075 (95)
patient076 (32)
patient077 (50)
patient078 (147)
patient079 (579)
patient080 (67)
Proband X1 (204)
Proband X10 (250)
Proband X11 (151)
Proband X12 (249)
Proband X13 (273)
Proband X14 (222)
Proband X15 (190)
Proband X2 (583)
Proband X4 (269)
Proband X5 (29)
Proband X6 (67)
Proband X7 (369)
Proband X8 (129)
Proband X9 (402)
sim (45)
wavelet−LLH_glrlm_LowGrayLevelRunEmphasis
frame
20
15
10
5
0
−5

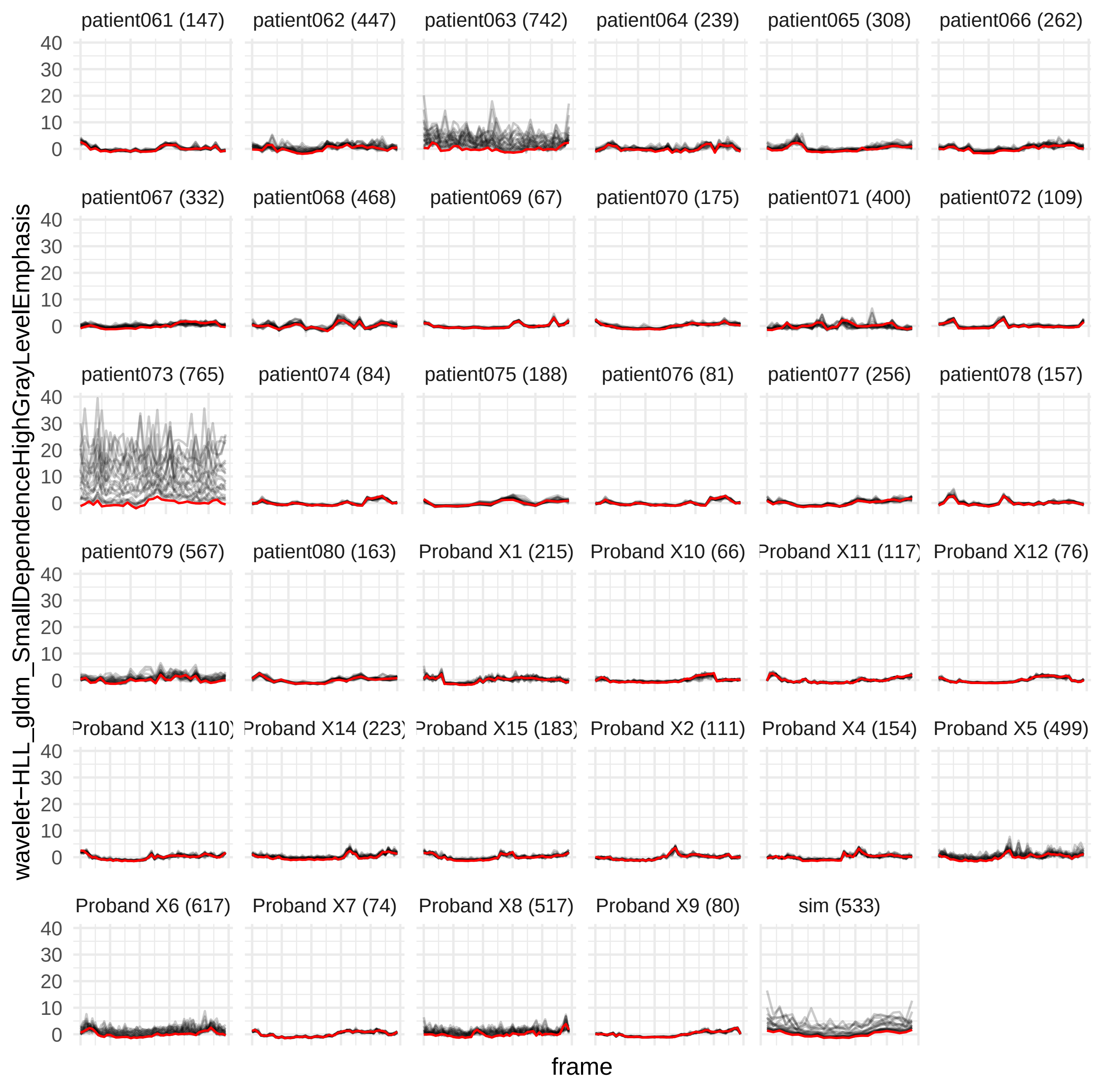

patient061 (147)
patient062 (447)
patient063 (742)
patient064 (239)
patient065 (308)
patient066 (262)
patient067 (332)
patient068 (468)
patient069 (67)
patient070 (175)
patient071 (400)
patient072 (109)
patient073 (765)
patient074 (84)
patient075 (188)
patient076 (81)
patient077 (256)
patient078 (157)
patient079 (567)
patient080 (163)
Proband X1 (215)
Proband X10 (66)
Proband X11 (117)
Proband X12 (76)
Proband X13 (110)
Proband X14 (223)
Proband X15 (183)
Proband X2 (111)
Proband X4 (154)
Proband X5 (499)
Proband X6 (617)
Proband X7 (74)
Proband X8 (517)
Proband X9 (80)
sim (533)
wavelet−HLL_gldm_SmallDependenceHighGrayLevelEmphasis
0
10
20
30
40
frame

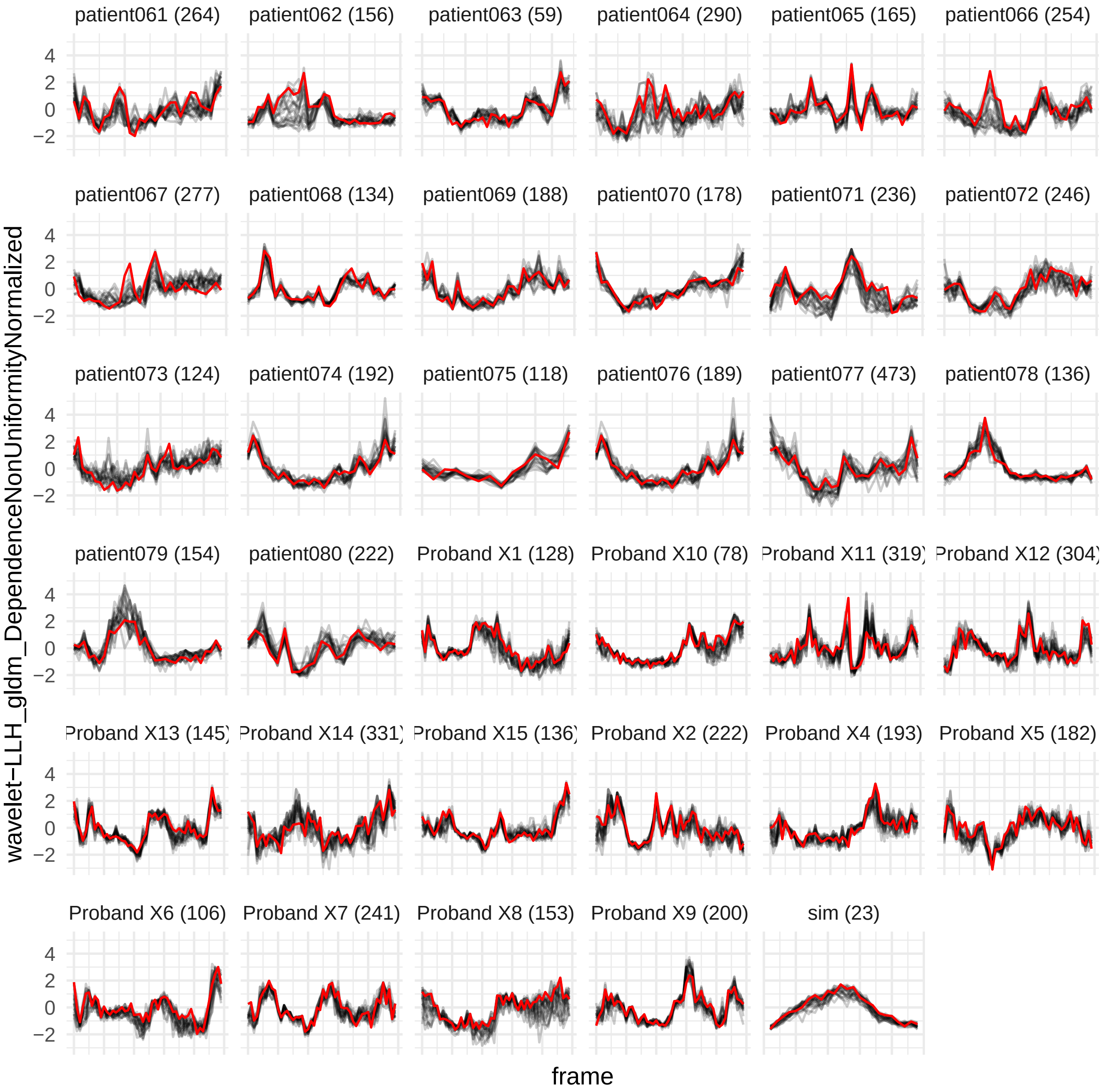

patient061 (264)
patient062 (156)
patient063 (59)
patient064 (290)
patient065 (165)
patient066 (254)
patient067 (277)
patient068 (134)
patient069 (188)
patient070 (178)
patient071 (236)
patient072 (246)
patient073 (124)
patient074 (192)
patient075 (118)
patient076 (189)
patient077 (473)
patient078 (136)
patient079 (154)
patient080 (222)
Proband X1 (128)
Proband X10 (78)
Proband X11 (319)
Proband X12 (304)
Proband X13 (145)
Proband X14 (331)
Proband X15 (136)
Proband X2 (222)
Proband X4 (193)
Proband X5 (182)
Proband X6 (106)
Proband X7 (241)
Proband X8 (153)
Proband X9 (200)
sim (23)
wavelet−LLH_gldm_DependenceNonUniformityNormalized
frame
4
2
0
−2

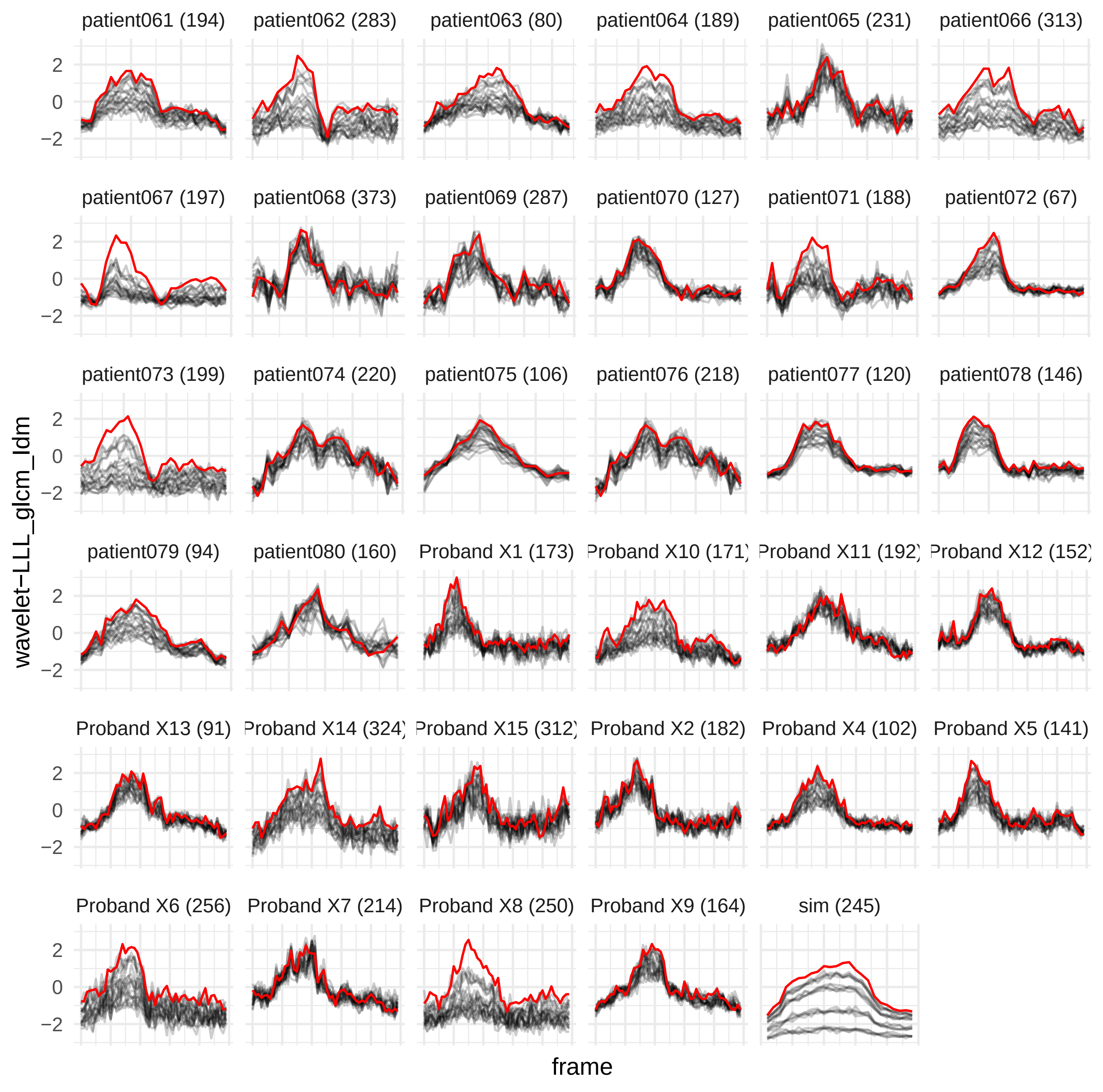
patient061 (194)
patient062 (283)
patient063 (80)
patient064 (189)
patient065 (231)
patient066 (313)
patient067 (197)
patient068 (373)
patient069 (287)
patient070 (127)
patient071 (188)
patient072 (67)
patient073 (199)
patient074 (220)
patient075 (106)
patient076 (218)
patient077 (120)
patient078 (146)
patient079 (94)
patient080 (160)
Proband X1 (173)
Proband X10 (171)
Proband X11 (192)
Proband X12 (152)
Proband X13 (91)
Proband X14 (324)
Proband X15 (312)
Proband X2 (182)
Proband X4 (102)
Proband X5 (141)
Proband X6 (256)
Proband X7 (214)
Proband X8 (250)
Proband X9 (164)
sim (245)
2
0
−2
wavelet−LLL_glcm_Idm
frame

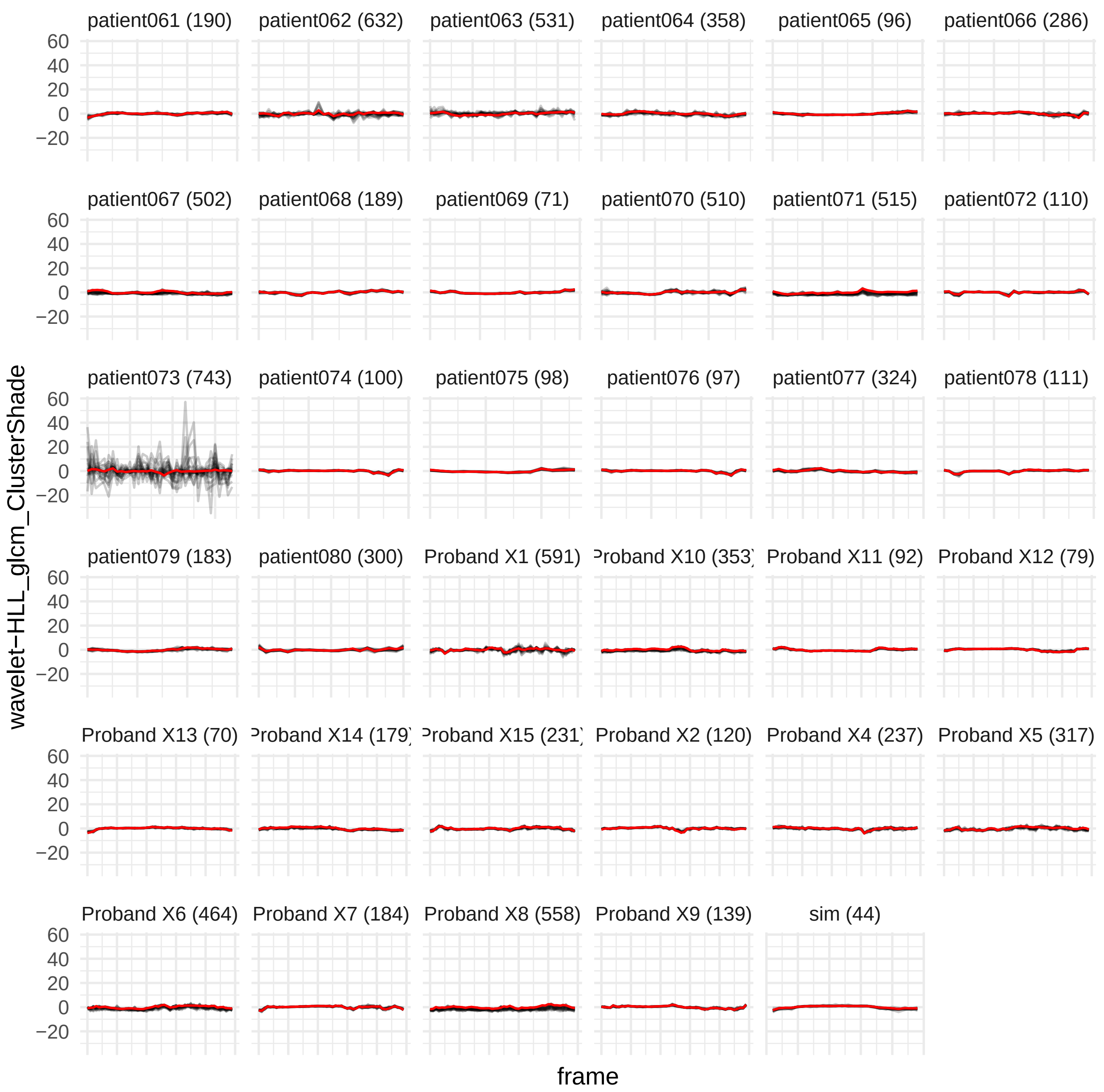

patient061 (190)
patient062 (632)
patient063 (531)
patient064 (358)
patient065 (96)
patient066 (286)
patient067 (502)
patient068 (189)
patient069 (71)
patient070 (510)
patient071 (515)
patient072 (110)
patient073 (743)
patient074 (100)
patient075 (98)
patient076 (97)
patient077 (324)
patient078 (111)
patient079 (183)
patient080 (300)
Proband X1 (591)
Proband X10 (353)
Proband X11 (92)
Proband X12 (79)
Proband X13 (70)
Proband X14 (179)
Proband X15 (231)
Proband X2 (120)
Proband X4 (237)
Proband X5 (317)
Proband X6 (464)
Proband X7 (184)
Proband X8 (558)
Proband X9 (139)
sim (44)
60
40
20
0
−20
wavelet−HLL_glcm_ClusterShade
frame

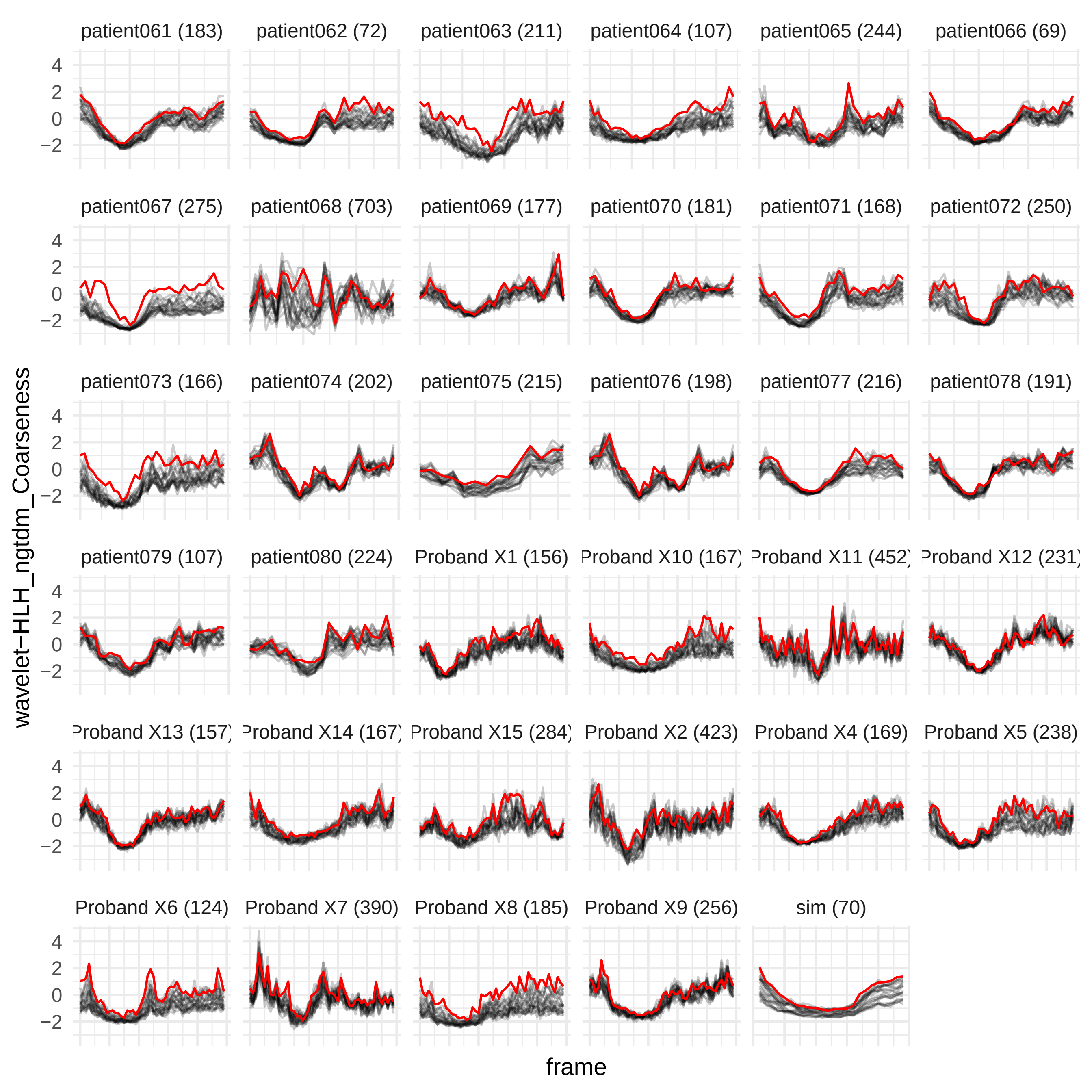

patient061 (183)
patient062 (72)
patient063 (211)
patient064 (107)
patient065 (244)
patient066 (69)
patient067 (275)
patient068 (703)
patient069 (177)
patient070 (181)
patient071 (168)
patient072 (250)
patient073 (166)
patient074 (202)
patient075 (215)
patient076 (198)
patient077 (216)
patient078 (191)
patient079 (107)
patient080 (224)
Proband X1 (156)
Proband X10 (167)
Proband X11 (452)
Proband X12 (231)
Proband X13 (157)
Proband X14 (167)
Proband X15 (284)
Proband X2 (423)
Proband X4 (169)
Proband X5 (238)
Proband X6 (124)
Proband X7 (390)
Proband X8 (185)
Proband X9 (256)
sim (70)
4
2
0
−2
wavelet−HLH_ngtdm_Coarseness
frame

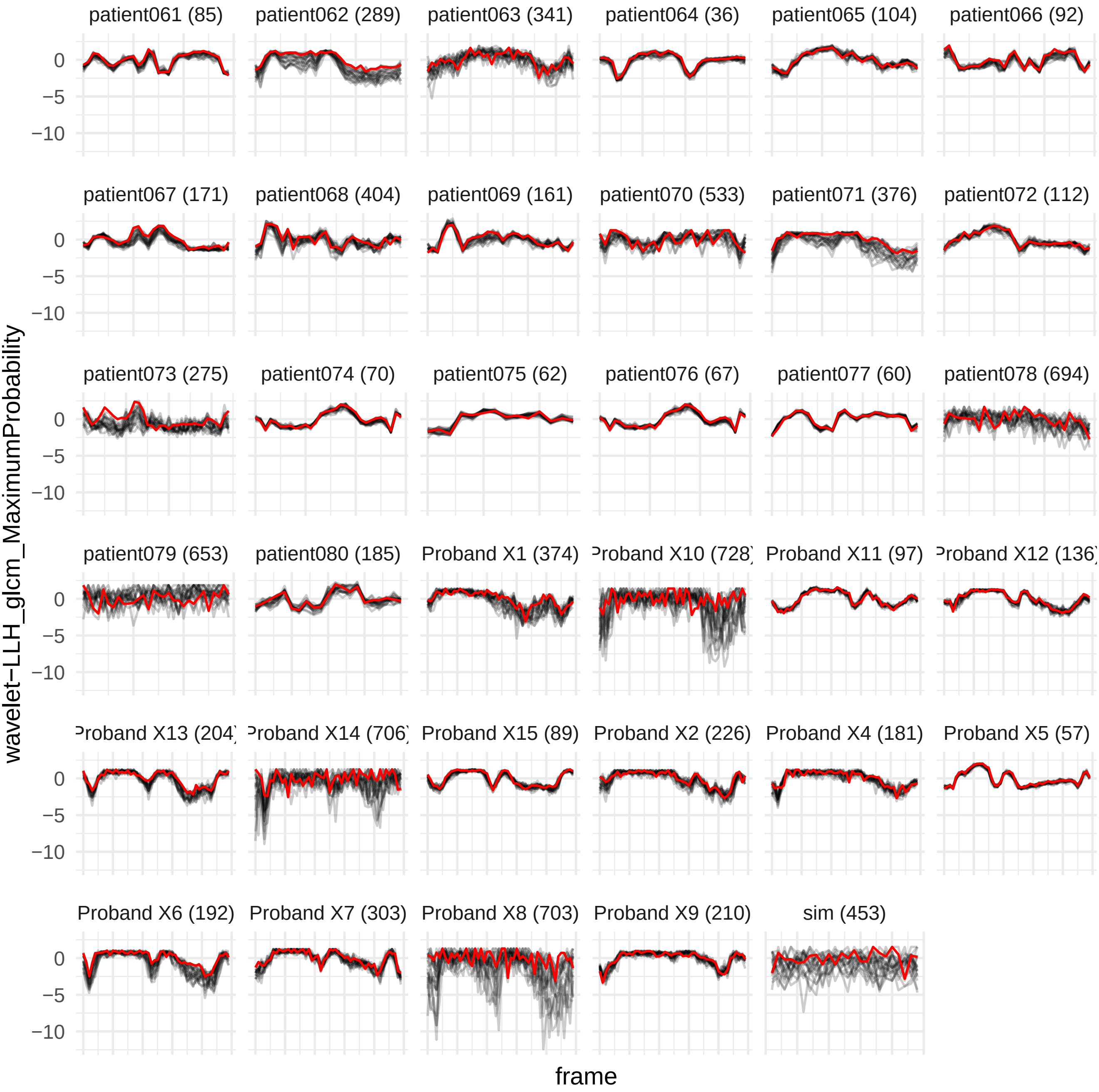
patient061 (85)
patient062 (289)
patient063 (341)
patient064 (36)
patient065 (104)
patient066 (92)
patient067 (171)
patient068 (404)
patient069 (161)
patient070 (533)
patient071 (376)
patient072 (112)
patient073 (275)
patient074 (70)
patient075 (62)
patient076 (67)
patient077 (60)
patient078 (694)
patient079 (653)
patient080 (185)
Proband X1 (374)
Proband X10 (728)
Proband X11 (97)
Proband X12 (136)
Proband X13 (204)
Proband X14 (706)
Proband X15 (89)
Proband X2 (226)
Proband X4 (181)
Proband X5 (57)
Proband X6 (192)
Proband X7 (303)
Proband X8 (703)
Proband X9 (210)
sim (453)
0
−5
−10
wavelet−LLH_glcm_MaximumProbability
frame

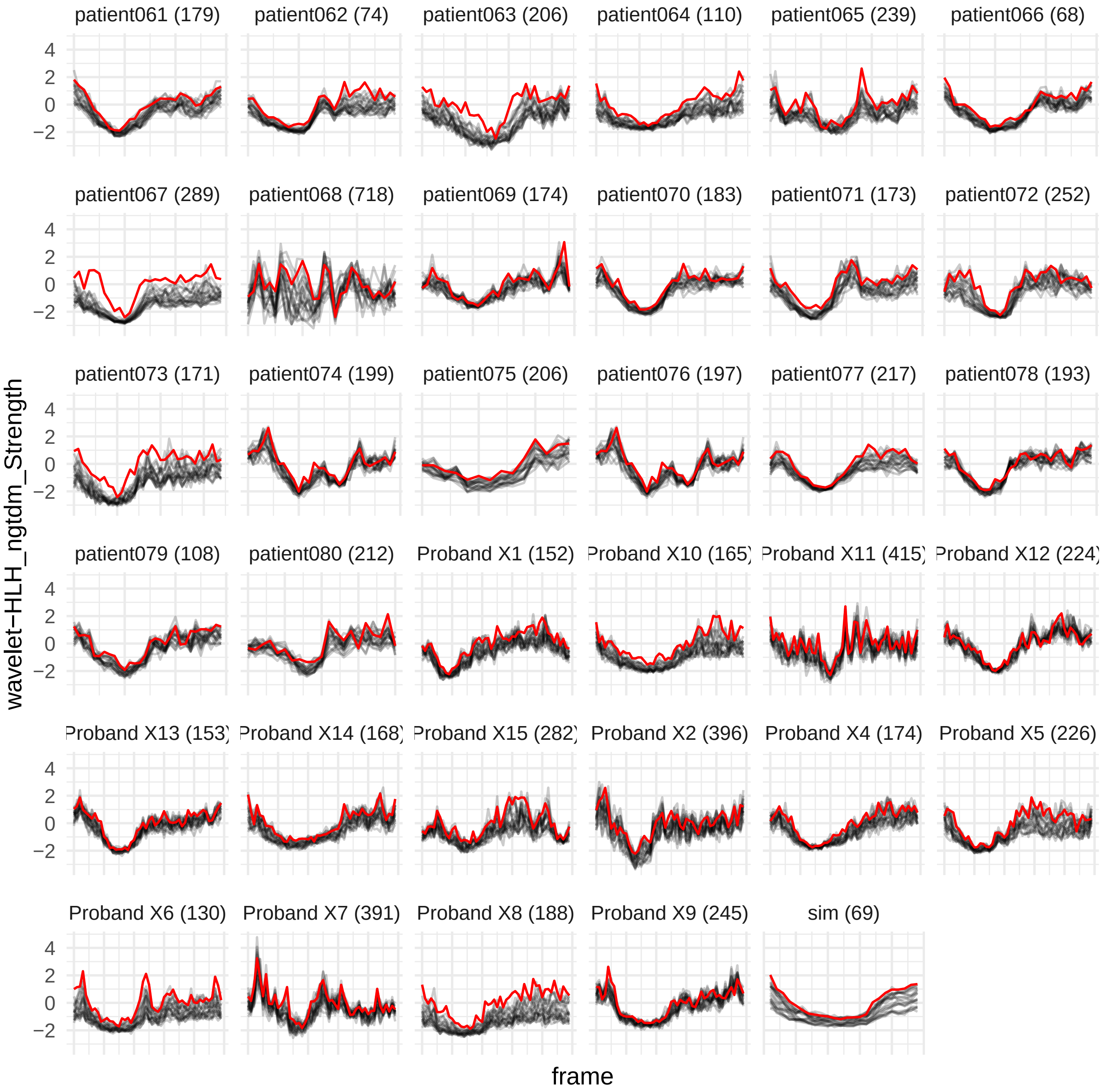
patient061 (179)
patient062 (74)
patient063 (206)
patient064 (110)
patient065 (239)
patient066 (68)
patient067 (289)
patient068 (718)
patient069 (174)
patient070 (183)
patient071 (173)
patient072 (252)
patient073 (171)
patient074 (199)
patient075 (206)
patient076 (197)
patient077 (217)
patient078 (193)
patient079 (108)
patient080 (212)
Proband X1 (152)
Proband X10 (165)
Proband X11 (415)
Proband X12 (224)
Proband X13 (153)
Proband X14 (168)
Proband X15 (282)
Proband X2 (396)
Proband X4 (174)
Proband X5 (226)
Proband X6 (130)
Proband X7 (391)
Proband X8 (188)
Proband X9 (245)
sim (69)
wavelet−HLH_ngtdm_Strength
frame

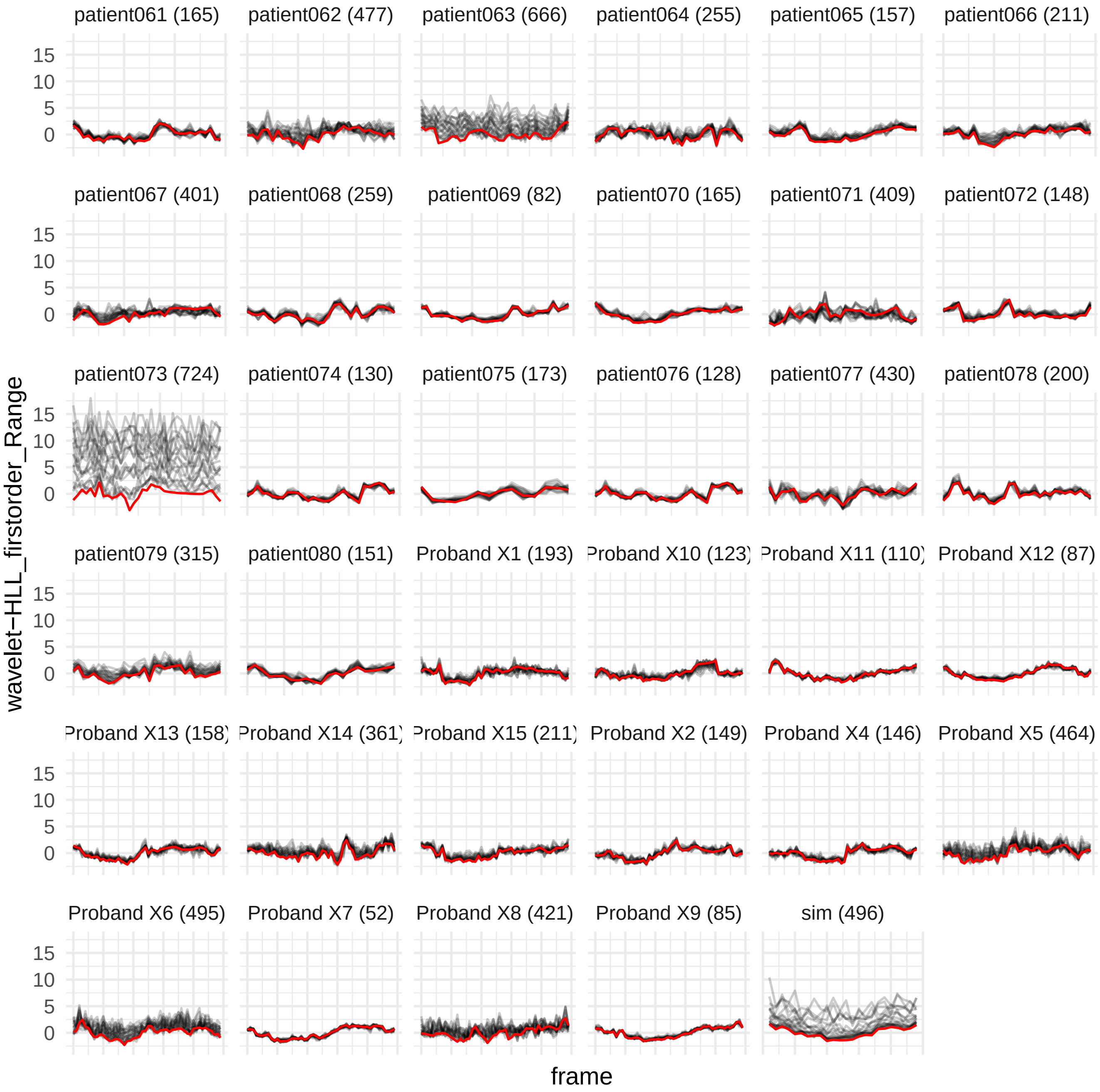
patient061 (165)
patient062 (477)
patient063 (666)
patient064 (255)
patient065 (157)
patient066 (211)
patient067 (401)
patient068 (259)
patient069 (82)
patient070 (165)
patient071 (409)
patient072 (148)
patient073 (724)
patient074 (130)
patient075 (173)
patient076 (128)
patient077 (430)
patient078 (200)
patient079 (315)
patient080 (151)
Proband X1 (193)
Proband X10 (123)
Proband X11 (110)
Proband X12 (87)
Proband X13 (158)
Proband X14 (361)
Proband X15 (211)
Proband X2 (149)
Proband X4 (146)
Proband X5 (464)
Proband X6 (495)
Proband X7 (52)
Proband X8 (421)
Proband X9 (85)
sim (496)
wavelet−HLL_firstorder_Range
15
10
5
0
frame

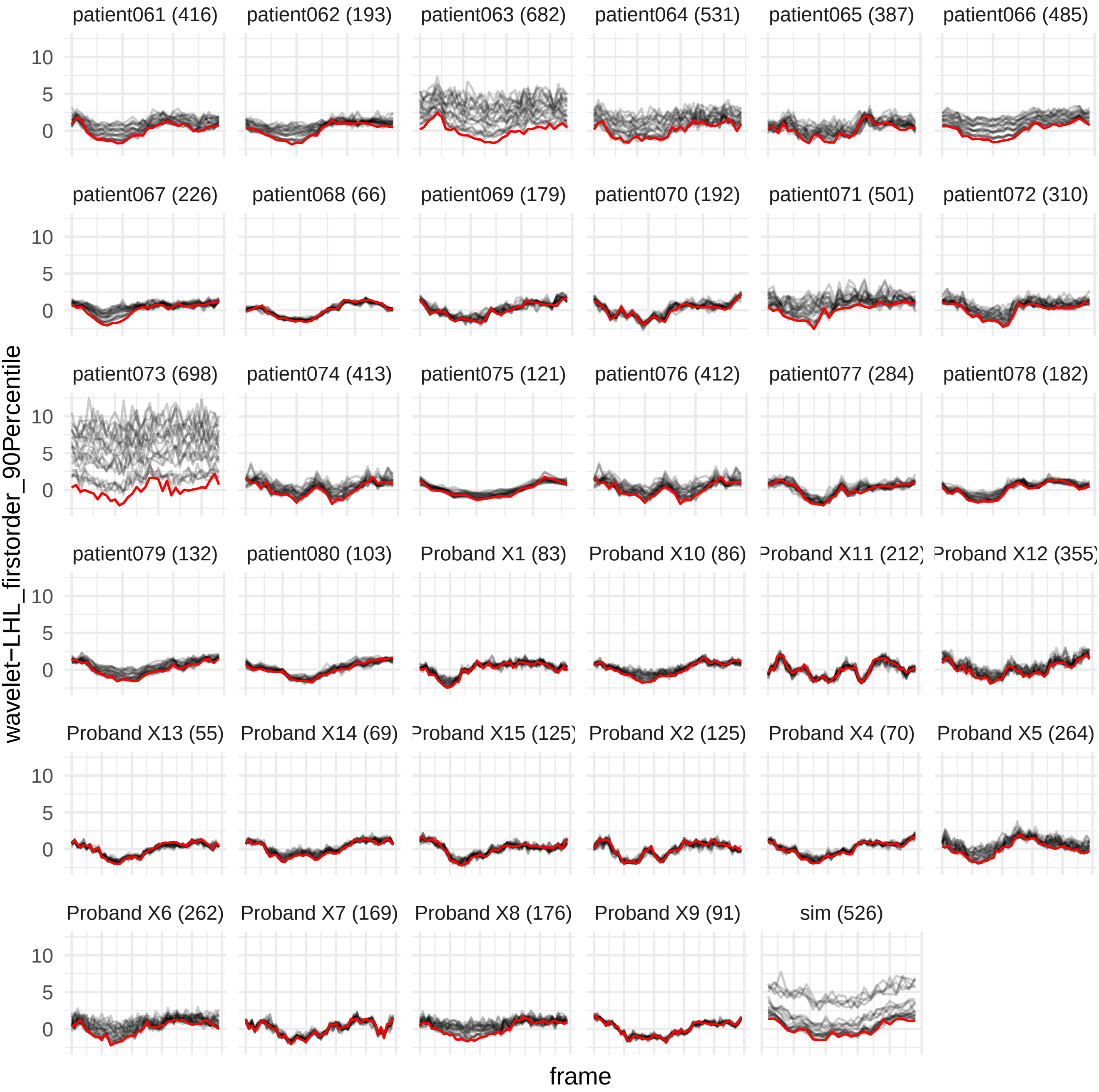
patient061 (416)
patient062 (193)
patient063 (682)
patient064 (531)
patient065 (387)
patient066 (485)
patient067 (226)
patient068 (66)
patient069 (179)
patient070 (192)
patient071 (501)
patient072 (310)
patient073 (698)
patient074 (413)
patient075 (121)
patient076 (412)
patient077 (284)
patient078 (182)
patient079 (132)
patient080 (103)
Proband X1 (83)
Proband X10 (86)
Proband X11 (212)
Proband X12 (355)
Proband X13 (55)
Proband X14 (69)
Proband X15 (125)
Proband X2 (125)
Proband X4 (70)
Proband X5 (264)
Proband X6 (262)
Proband X7 (169)
Proband X8 (176)
Proband X9 (91)
sim (526)
10
5
0
wavelet−LHL_firstorder_90Percentile
frame

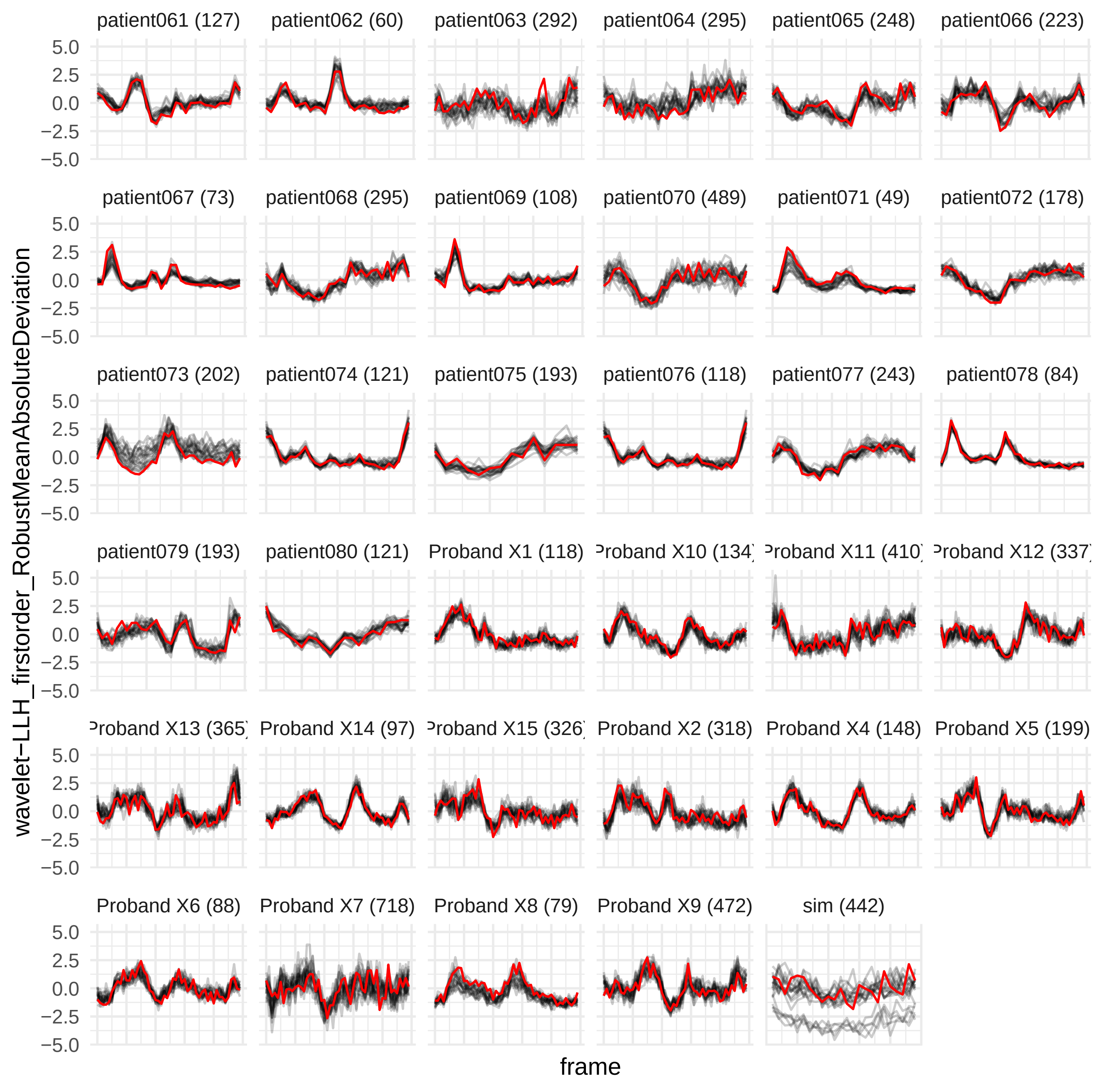

patient061 (127)
patient062 (60)
patient063 (292)
patient064 (295)
patient065 (248)
patient066 (223)
patient067 (73)
patient068 (295)
patient069 (108)
patient070 (489)
patient071 (49)
patient072 (178)
patient073 (202)
patient074 (121)
patient075 (193)
patient076 (118)
patient077 (243)
patient078 (84)
patient079 (193)
patient080 (121)
Proband X1 (118)
Proband X10 (134)
Proband X11 (410)
Proband X12 (337)
Proband X13 (365)
Proband X14 (97)
Proband X15 (326)
Proband X2 (318)
Proband X4 (148)
Proband X5 (199)
Proband X6 (88)
Proband X7 (718)
Proband X8 (79)
Proband X9 (472)
sim (442)
5.0
2.5
0.0
−2.5
−5.0
wavelet−LLH_firstorder_RobustMeanAbsoluteDeviation
frame

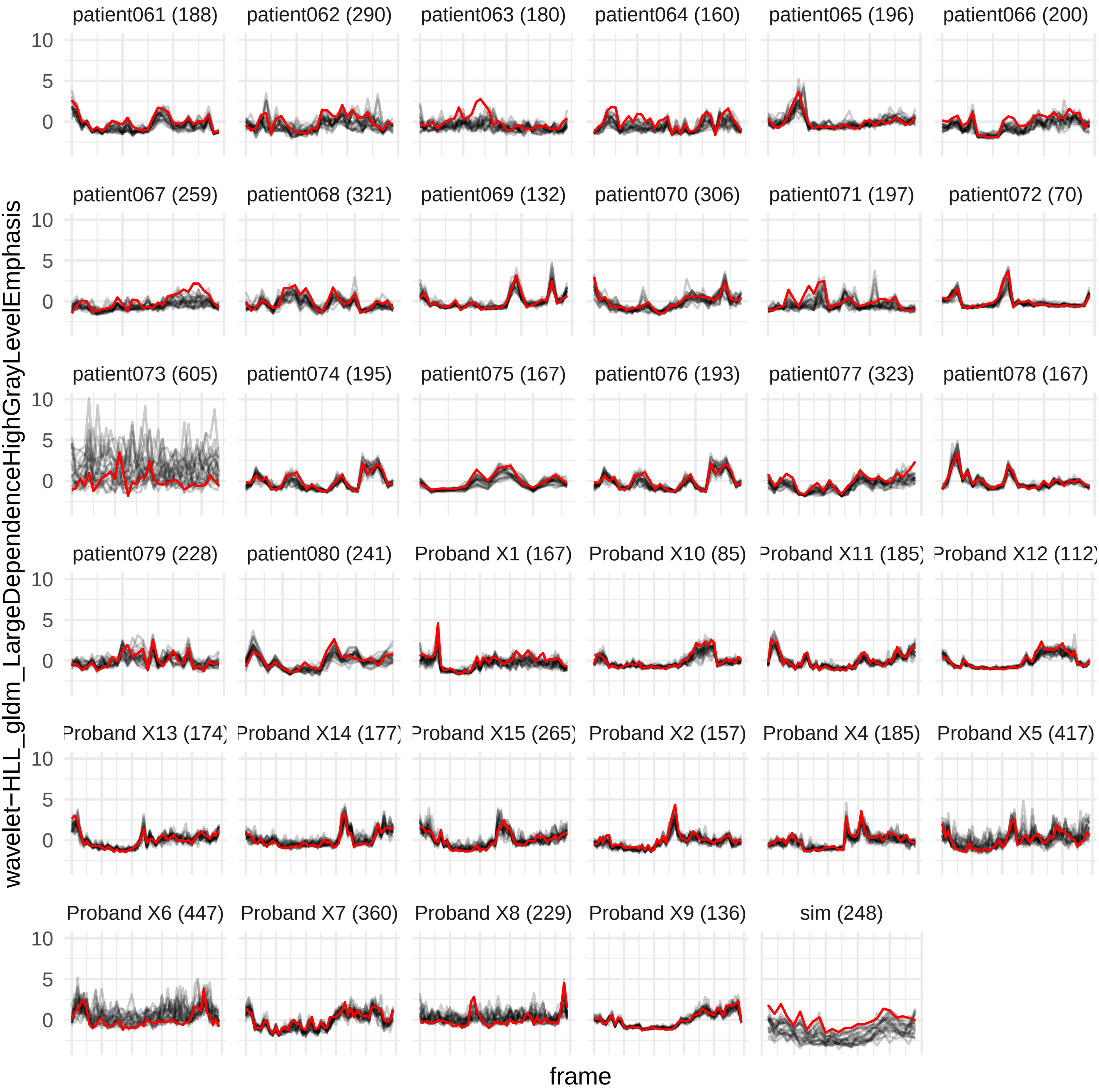
patient061 (188)
patient062 (290)
patient063 (180)
patient064 (160)
patient065 (196)
patient066 (200)
patient067 (259)
patient068 (321)
patient069 (132)
patient070 (306)
patient071 (197)
patient072 (70)
patient073 (605)
patient074 (195)
patient075 (167)
patient076 (193)
patient077 (323)
patient078 (167)
patient079 (228)
patient080 (241)
Proband X1 (167)
Proband X10 (85)
Proband X11 (185)
Proband X12 (112)
Proband X13 (174)
Proband X14 (177)
Proband X15 (265)
Proband X2 (157)
Proband X4 (185)
Proband X5 (417)
Proband X6 (447)
Proband X7 (360)
Proband X8 (229)
Proband X9 (136)
sim (248)
10
5
0
wavelet−HLL_gldm_LargeDependenceHighGrayLevelEmphasis
frame

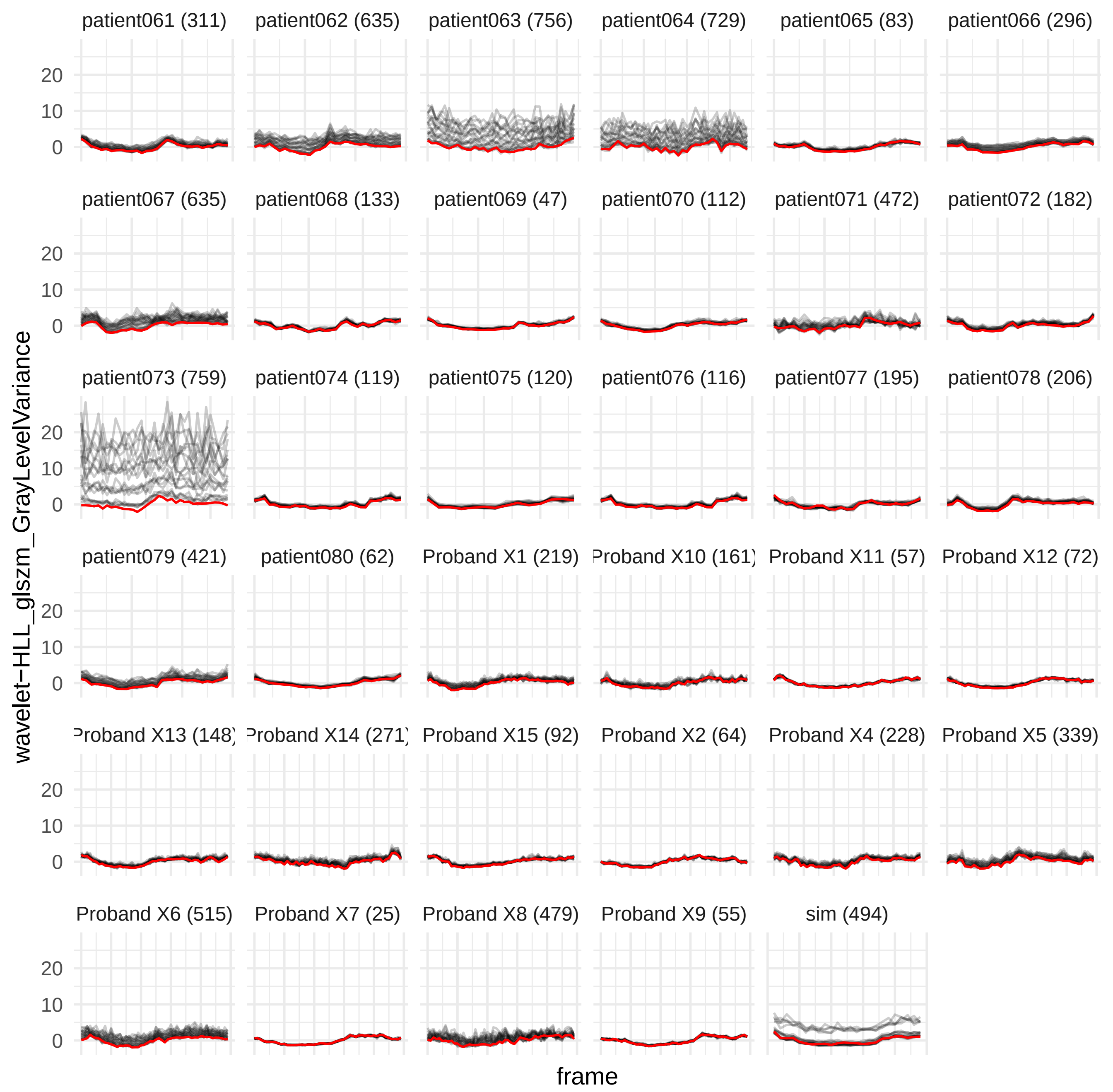
patient061 (311)
patient062 (635)
patient063 (756)
patient064 (729)
patient065 (83)
patient066 (296)
patient067 (635)
patient068 (133)
patient069 (47)
patient070 (112)
patient071 (472)
patient072 (182)
patient073 (759)
patient074 (119)
patient075 (120)
patient076 (116)
patient077 (195)
patient078 (206)
patient079 (421)
patient080 (62)
Proband X1 (219)
Proband X10 (161)
Proband X11 (57)
Proband X12 (72)
Proband X13 (148)
Proband X14 (271)
Proband X15 (92)
Proband X2 (64)
Proband X4 (228)
Proband X5 (339)
Proband X6 (515)
Proband X7 (25)
Proband X8 (479)
Proband X9 (55)
sim (494)
wavelet−HLL_glszm_GrayLevelVariance
frame
0
10
20

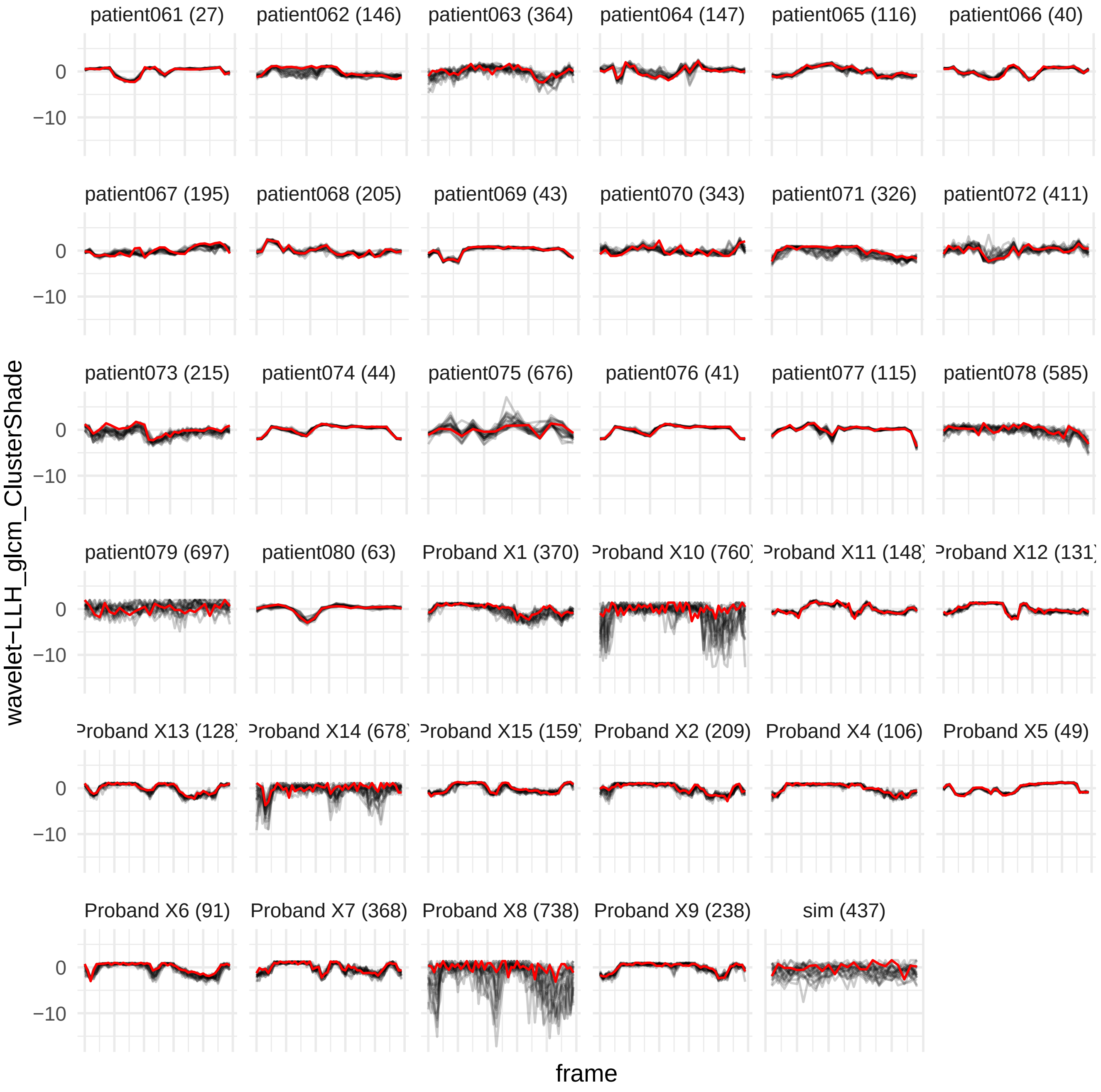
patient061 (27)
patient062 (146)
patient063 (364)
patient064 (147)
patient065 (116)
patient066 (40)
patient067 (195)
patient068 (205)
patient069 (43)
patient070 (343)
patient071 (326)
patient072 (411)
patient073 (215)
patient074 (44)
patient075 (676)
patient076 (41)
patient077 (115)
patient078 (585)
patient079 (697)
patient080 (63)
Proband X1 (370)
Proband X4 (106)
Proband X5 (49)
Proband X2 (209)
Proband X6 (91)
Proband X7 (368)
Proband X8 (738)
Proband X9 (238)
sim (437)
0
−10
wavelet−LLH_glcm_ClusterShade
frame

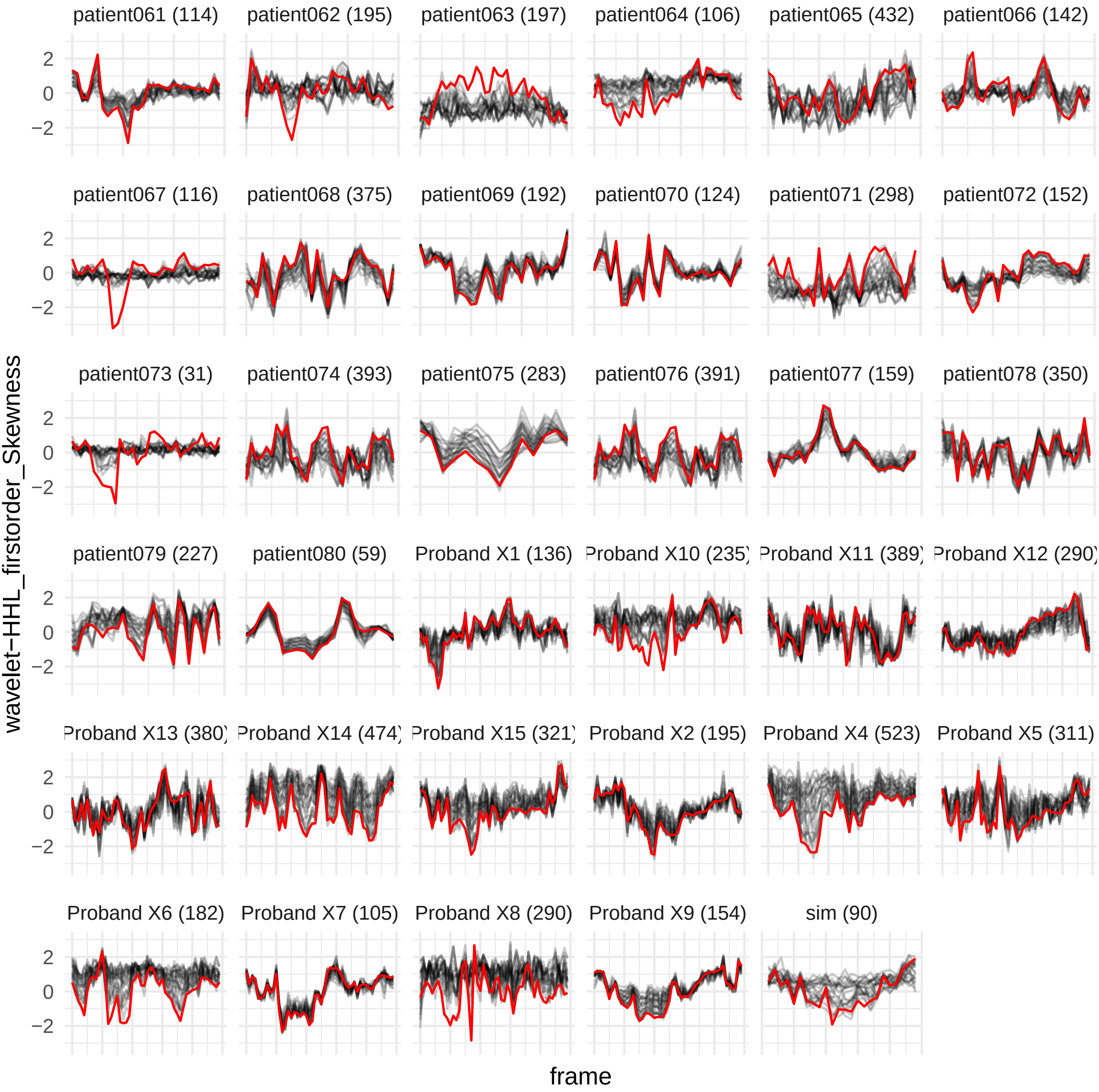
patient061 (114)
patient062 (195)
patient063 (197)
patient064 (106)
patient065 (432)
patient066 (142)
patient067 (116)
patient068 (375)
patient069 (192)
patient070 (124)
patient071 (298)
patient072 (152)
patient073 (31)
patient074 (393)
patient075 (283)
patient076 (391)
patient077 (159)
patient078 (350)
patient079 (227)
patient080 (59)
Proband X1 (136)
Proband X10 (235)
Proband X11 (389)
Proband X12 (290)
Proband X13 (380)
Proband X14 (474)
Proband X15 (321)
Proband X2 (195)
Proband X4 (523)
Proband X5 (311)
Proband X6 (182)
Proband X7 (105)
Proband X8 (290)
Proband X9 (154)
sim (90)
2
0
−2
wavelet−HHL_firstorder_Skewness
frame

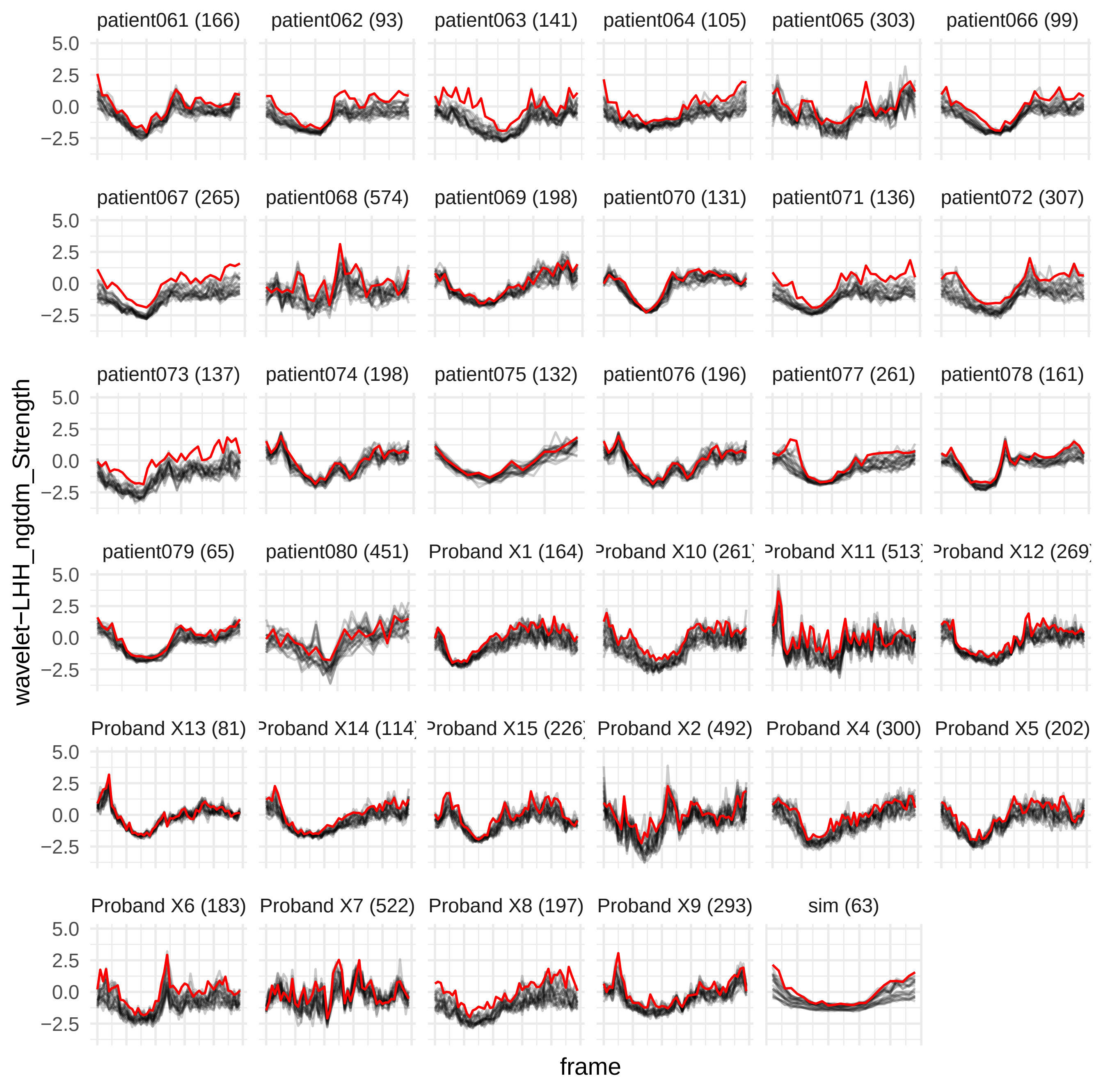

patient061 (166)
patient062 (93)
patient063 (141)
patient064 (105)
patient065 (303)
patient066 (99)
patient067 (265)
patient068 (574)
patient069 (198)
patient070 (131)
patient071 (136)
patient072 (307)
patient073 (137)
patient074 (198)
patient075 (132)
patient076 (196)
patient077 (261)
patient078 (161)
patient079 (65)
patient080 (451)
Proband X1 (164)
Proband X10 (261)
Proband X11 (513)
Proband X12 (269)
Proband X13 (81)
Proband X14 (114)
Proband X15 (226)
Proband X2 (492)
Proband X4 (300)
Proband X5 (202)
Proband X6 (183)
Proband X7 (522)
Proband X8 (197)
Proband X9 (293)
sim (63)
5.0
2.5
0.0
−2.5
wavelet−LHH_ngtdm_Strength
frame

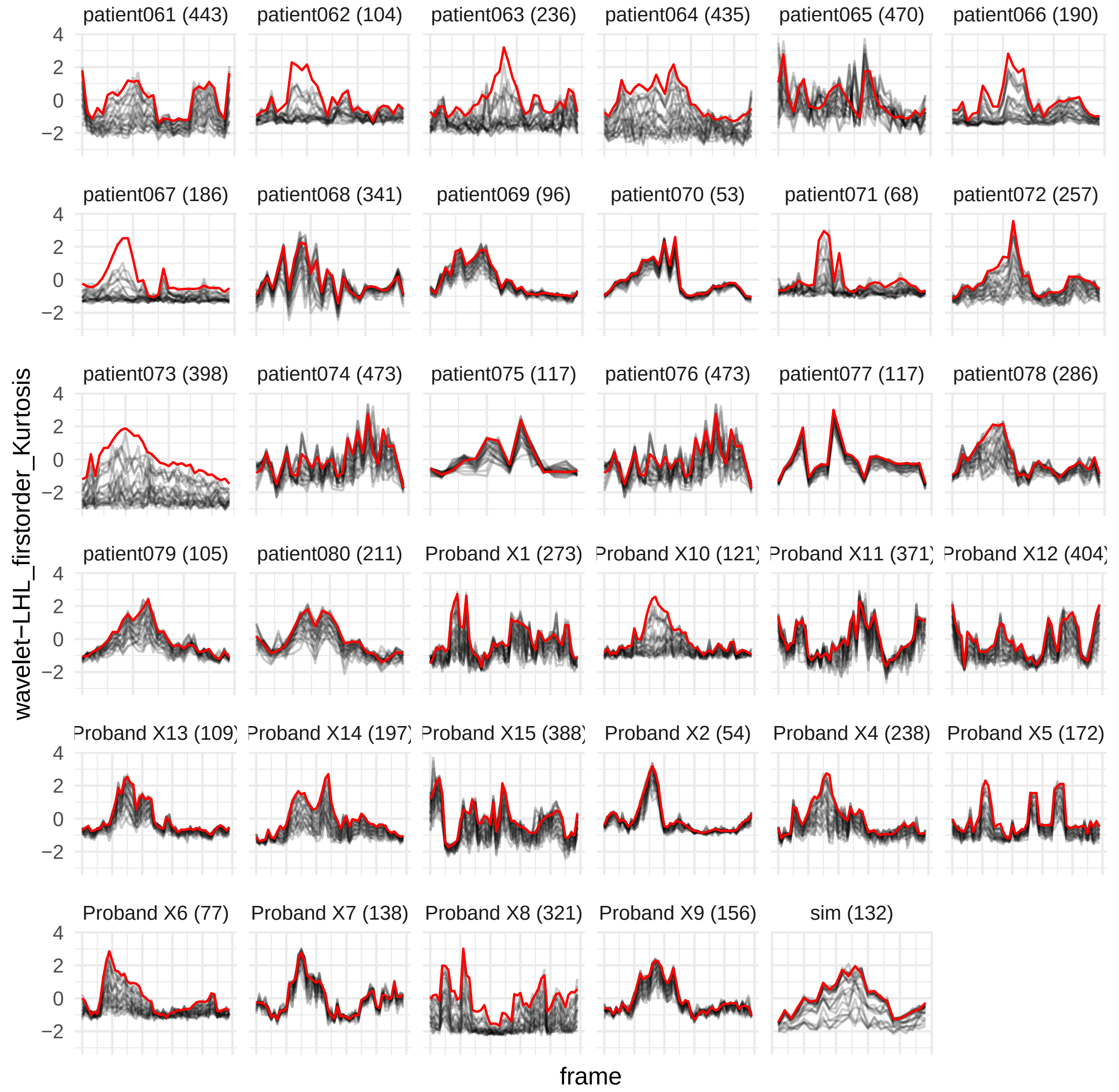
patient061 (443)
patient062 (104)
patient063 (236)
patient064 (435)
patient065 (470)
patient066 (190)
patient067 (186)
patient068 (341)
patient069 (96)
patient070 (53)
patient071 (68)
patient072 (257)
patient073 (398)
patient074 (473)
patient075 (117)
patient076 (473)
patient077 (117)
patient078 (286)
patient079 (105)
patient080 (211)
Proband X1 (273)
Proband X10 (121)
Proband X11 (371)
Proband X12 (404)
Proband X13 (109)
Proband X14 (197)
Proband X15 (388)
Proband X2 (54)
Proband X4 (238)
Proband X5 (172)
Proband X6 (77)
Proband X7 (138)
Proband X8 (321)
Proband X9 (156)
sim (132)
wavelet−LHL_firstorder_Kurtosis
frame
4
2
0
−2

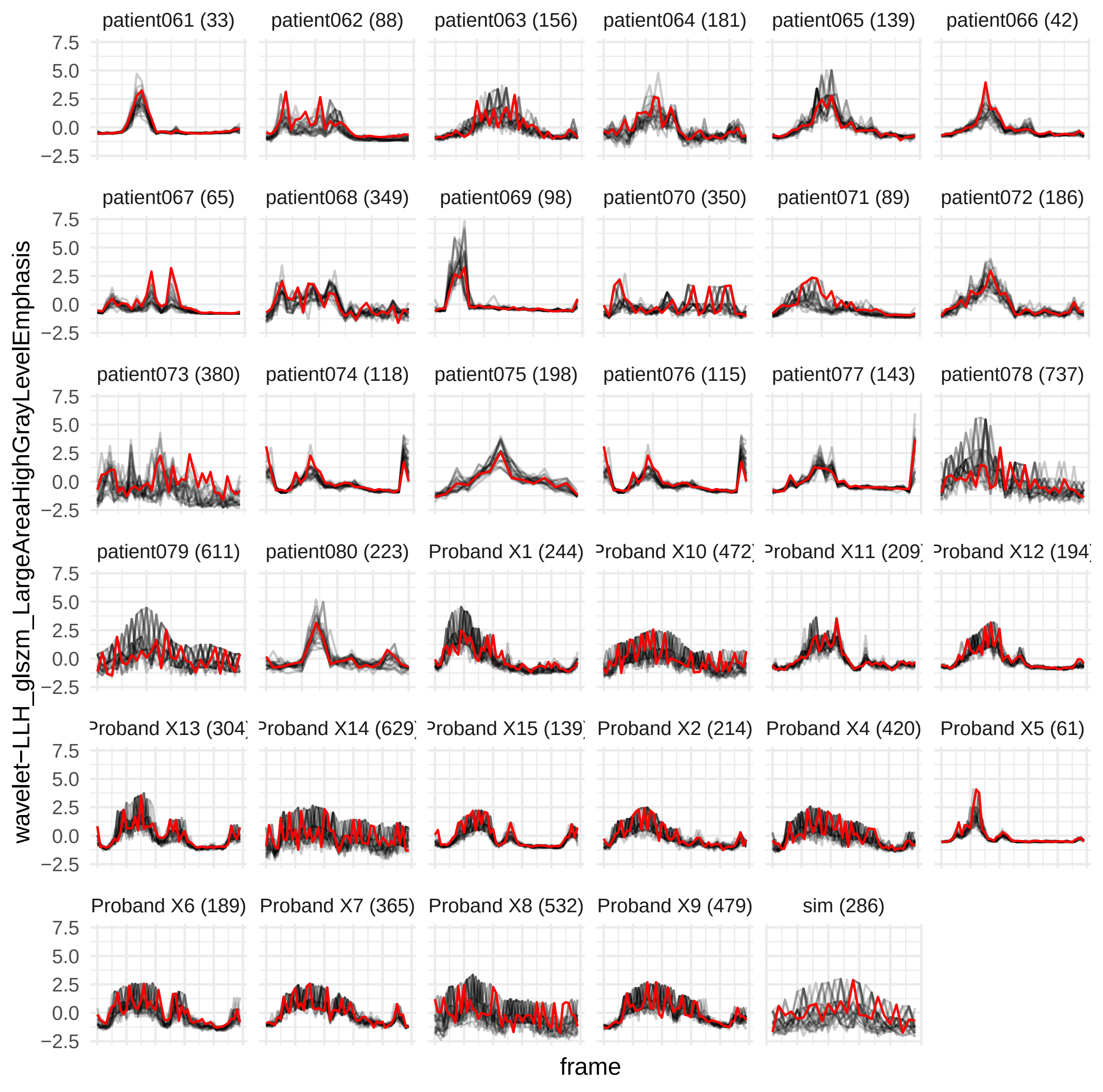

patient061 (33)
patient062 (88)
patient063 (156)
patient064 (181)
patient065 (139)
patient066 (42)
patient067 (65)
patient068 (349)
patient069 (98)
patient070 (350)
patient071 (89)
patient072 (186)
patient073 (380)
patient074 (118)
patient075 (198)
patient076 (115)
patient077 (143)
patient078 (737)
patient079 (611)
patient080 (223)
Proband X1 (244)
Proband X10 (472)
Proband X11 (209)
Proband X12 (194)
Proband X13 (304)
Proband X14 (629)
Proband X15 (139)
Proband X2 (214)
Proband X4 (420)
Proband X5 (61)
Proband X6 (189)
Proband X7 (365)
Proband X8 (532)
Proband X9 (479)
sim (286)
7.5
5.0
2.5
0.0
−2.5
wavelet−LLH_glszm_LargeAreaHighGrayLevelEmphasis
frame

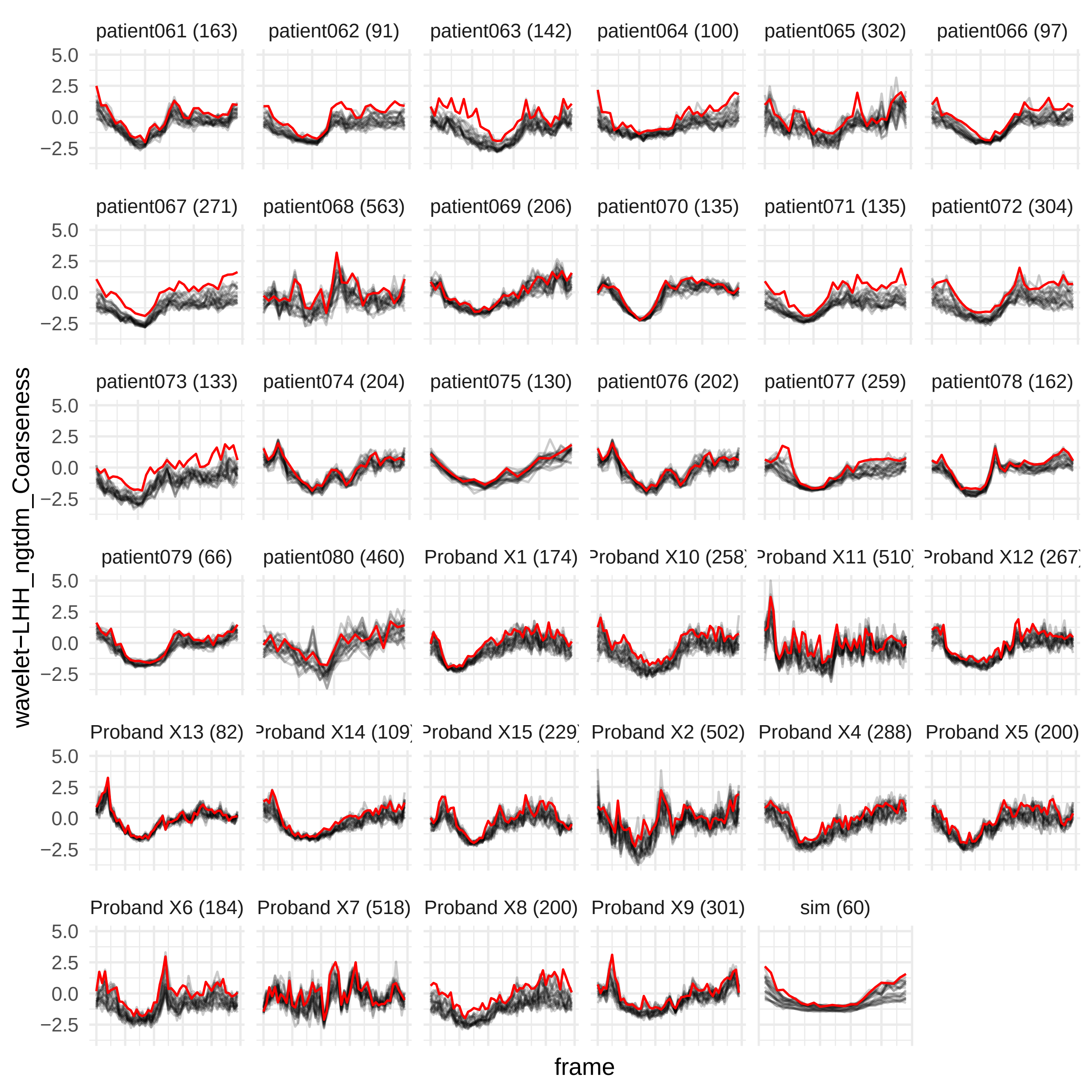

patient061 (163)
patient062 (91)
patient063 (142)
patient064 (100)
patient065 (302)
patient066 (97)
patient067 (271)
patient068 (563)
patient069 (206)
patient070 (135)
patient071 (135)
patient072 (304)
patient073 (133)
patient074 (204)
patient075 (130)
patient076 (202)
patient077 (259)
patient078 (162)
patient079 (66)
patient080 (460)
Proband X1 (174)
Proband X10 (258)
Proband X11 (510)
Proband X12 (267)
Proband X13 (82)
Proband X14 (109)
Proband X15 (229)
Proband X2 (502)
Proband X4 (288)
Proband X5 (200)
Proband X6 (184)
Proband X7 (518)
Proband X8 (200)
Proband X9 (301)
sim (60)
5.0
2.5
0.0
−2.5
wavelet−LHH_ngtdm_Coarseness
frame

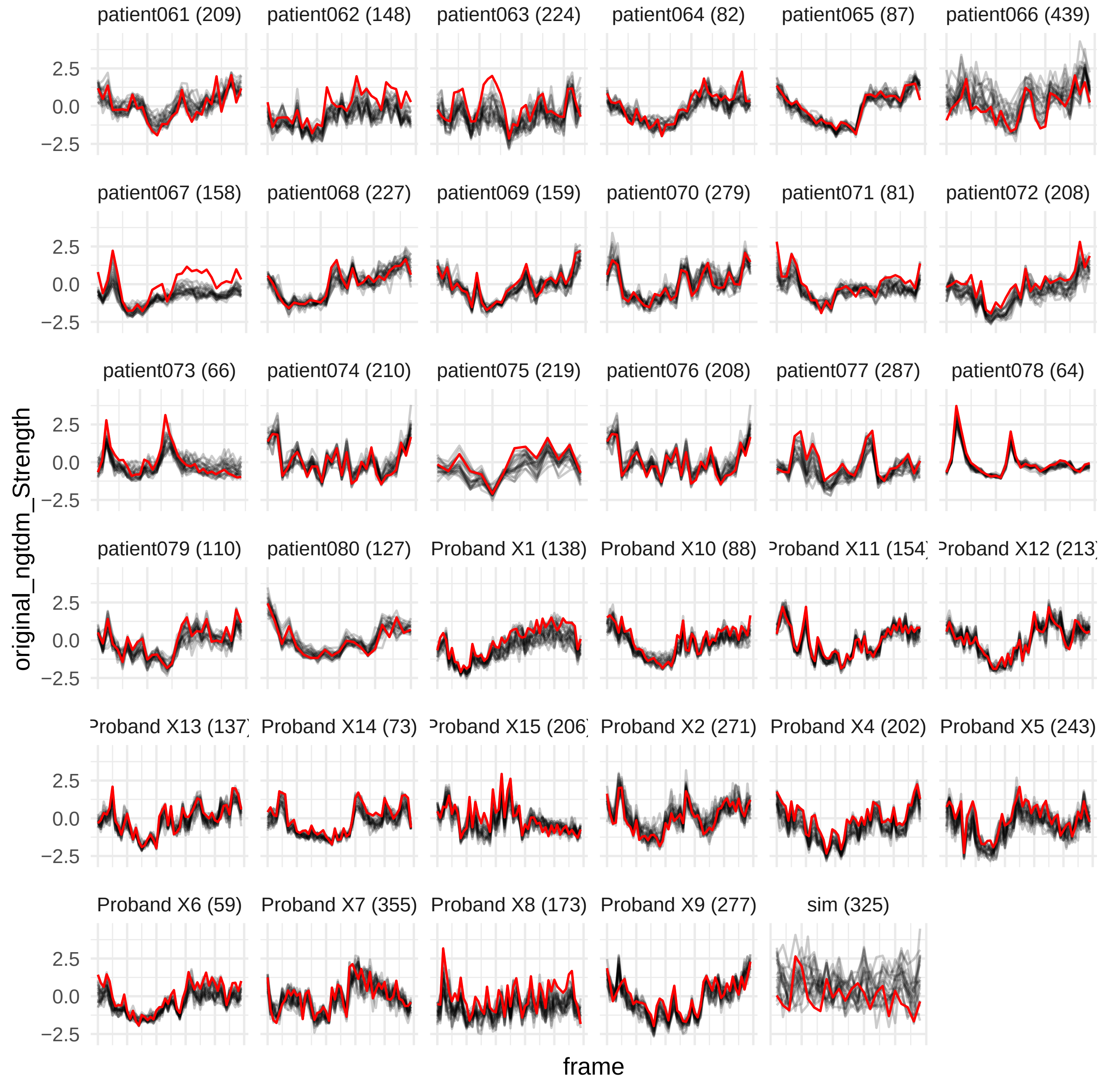
patient061 (209)
patient062 (148)
patient063 (224)
patient064 (82)
patient065 (87)
patient066 (439)
patient067 (158)
patient068 (227)
patient069 (159)
patient070 (279)
patient071 (81)
patient072 (208)
patient073 (66)
patient074 (210)
patient075 (219)
patient076 (208)
patient077 (287)
patient078 (64)
patient079 (110)
patient080 (127)
Proband X1 (138)
Proband X10 (88)
Proband X11 (154
Proband X12 (213
Proband X13 (137
Proband X14 (73)
Proband X15 (206
Proband X2 (271)
Proband X4 (202)
Proband X5 (243)
Proband X6 (59)
Proband X7 (355)
Proband X8 (173)
Proband X9 (277)
sim (325)
original_ngtdm_Strength
2.5
0.0
−2.5
frame

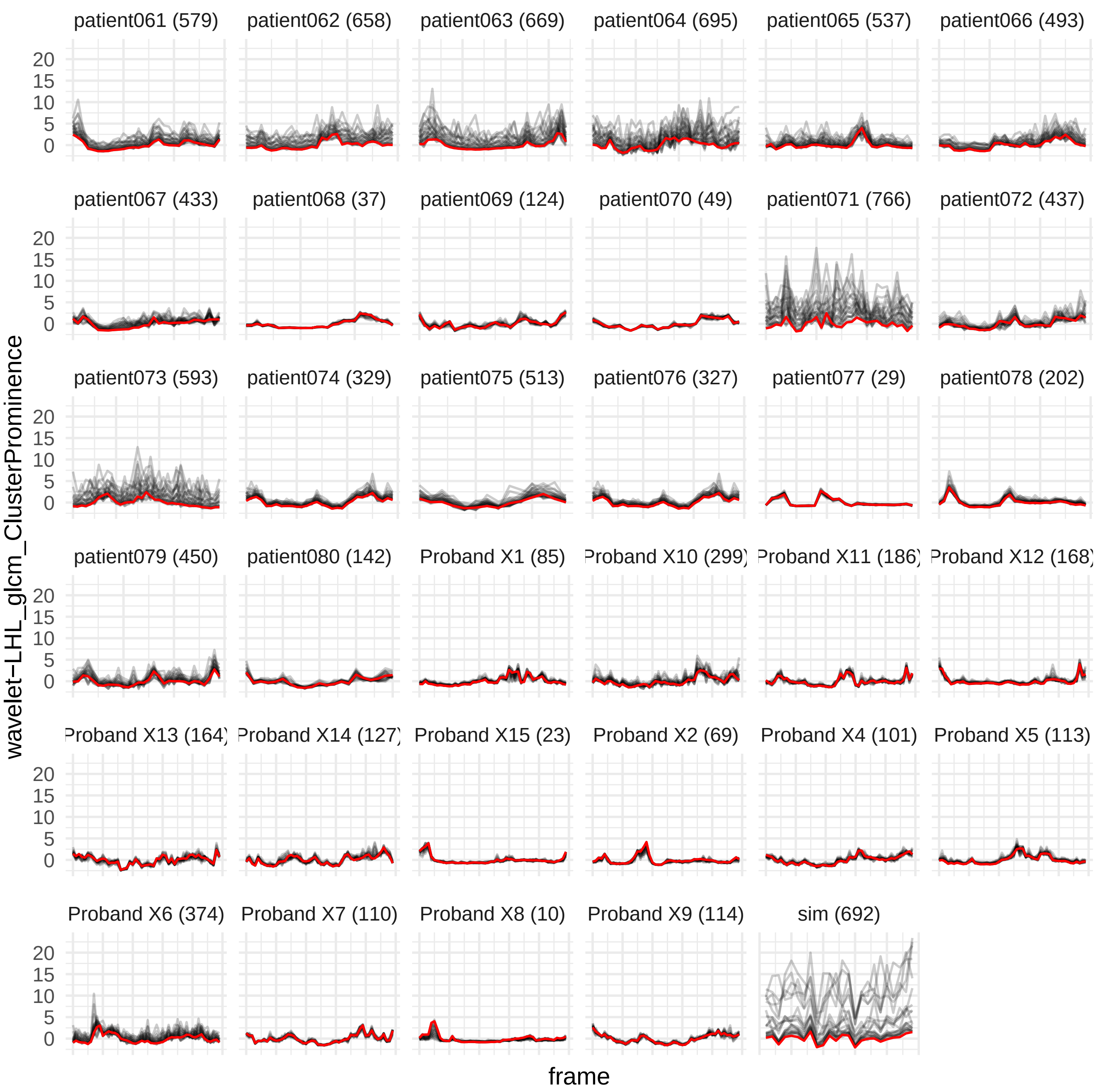

patient061 (579)
patient062 (658)
patient063 (669)
patient064 (695)
patient065 (537)
patient066 (493)
patient067 (433)
patient068 (37)
patient069 (124)
patient070 (49)
patient071 (766)
patient072 (437)
patient073 (593)
patient074 (329)
patient075 (513)
patient076 (327)
patient077 (29)
patient078 (202)
patient079 (450)
patient080 (142)
Proband X1 (85)
Proband X10 (299)
Proband X11 (186)
Proband X12 (168)
Proband X13 (164)
Proband X14 (127)
Proband X15 (23)
Proband X2 (69)
Proband X4 (101)
Proband X5 (113)
Proband X6 (374)
Proband X7 (110)
Proband X8 (10)
Proband X9 (114)
sim (692)
wavelet−LHL_glcm_ClusterProminence
0
5
10
15
20
frame

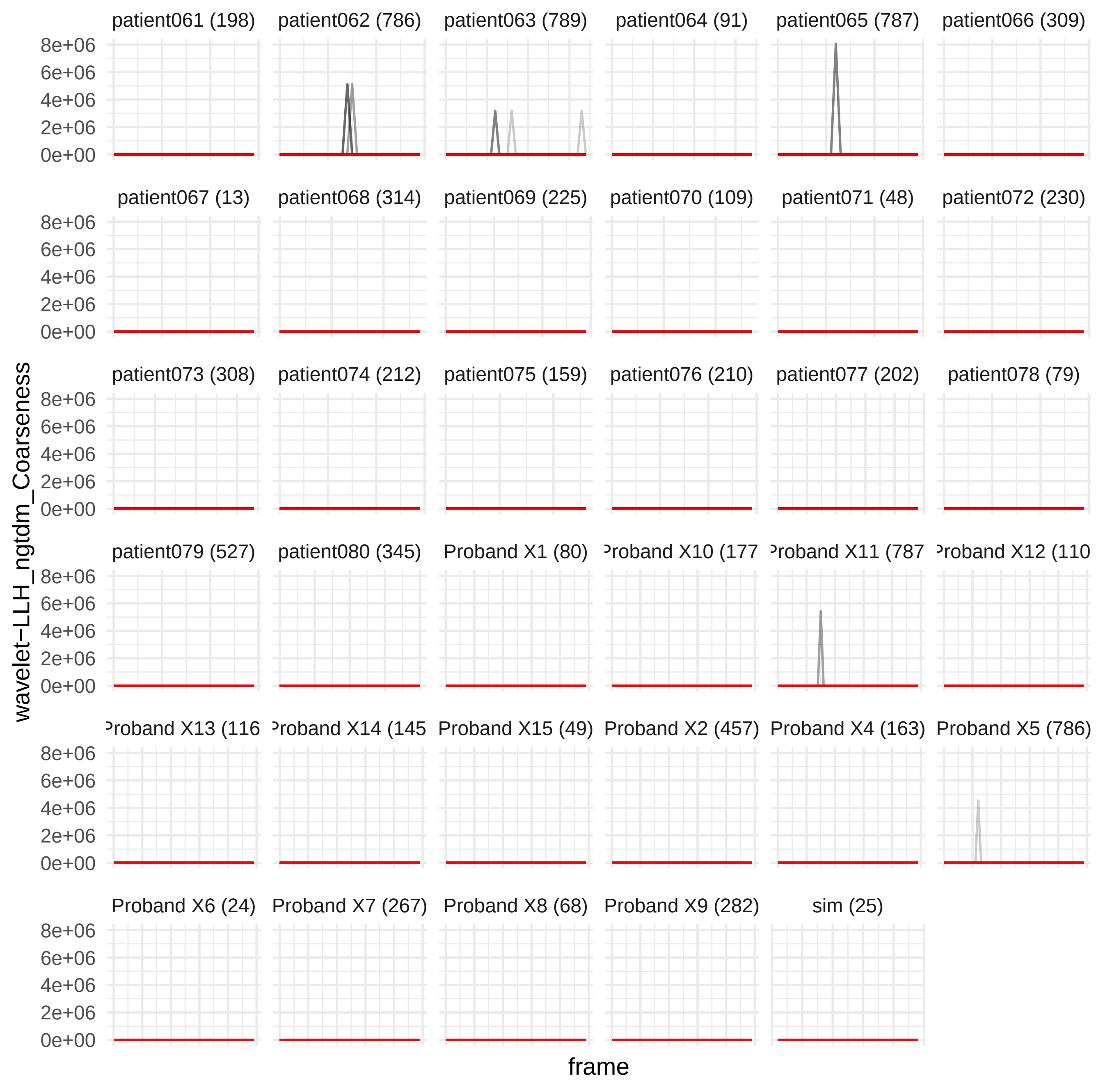

patient061 (198)
patient062 (786)
patient063 (789)
patient064 (91)
patient065 (787)
patient066 (309)
patient067 (13)
patient068 (314)
patient069 (225)
patient070 (109)
patient071 (48)
patient072 (230)
patient073 (308)
patient074 (212)
patient075 (159)
patient076 (210)
patient077 (202)
patient078 (79)
patient079 (527)
patient080 (345)
Proband X1 (80)
Proband X10 (177
Proband X11 (787
Proband X12 (110
Proband X13 (116
Proband X14 (145
Proband X15 (49)
Proband X2 (457)
Proband X4 (163)
Proband X5 (786)
Proband X6 (24)
Proband X7 (267)
Proband X8 (68)
Proband X9 (282)
sim (25)
8e+06
6e+06
4e+06
2e+06
0e+00
wavelet−LLH_ngtdm_Coarseness
frame

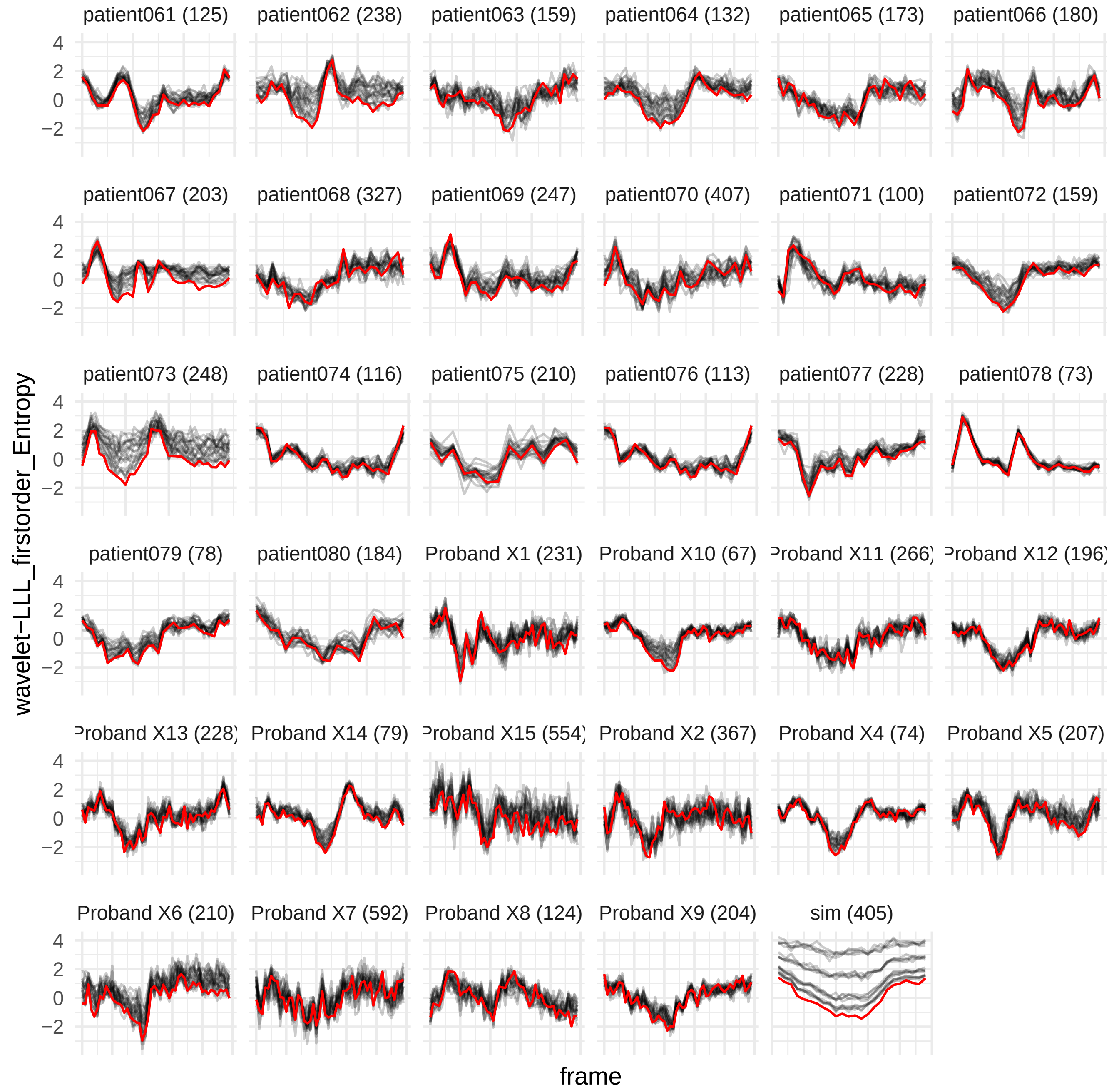
patient061 (125)
patient062 (238)
patient063 (159)
patient064 (132)
patient065 (173)
patient066 (180)
patient067 (203)
patient068 (327)
patient069 (247)
patient070 (407)
patient071 (100)
patient072 (159)
patient073 (248)
patient074 (116)
patient075 (210)
patient076 (113)
patient077 (228)
patient078 (73)
patient079 (78)
patient080 (184)
Proband X1 (231)
Proband X10 (67)
Proband X11 (266)
Proband X12 (196)
Proband X13 (228)
Proband X14 (79)
Proband X15 (554)
Proband X2 (367)
Proband X4 (74)
Proband X5 (207)
Proband X6 (210)
Proband X7 (592)
Proband X8 (124)
Proband X9 (204)
sim (405)
wavelet−LLL_firstorder_Entropy
frame

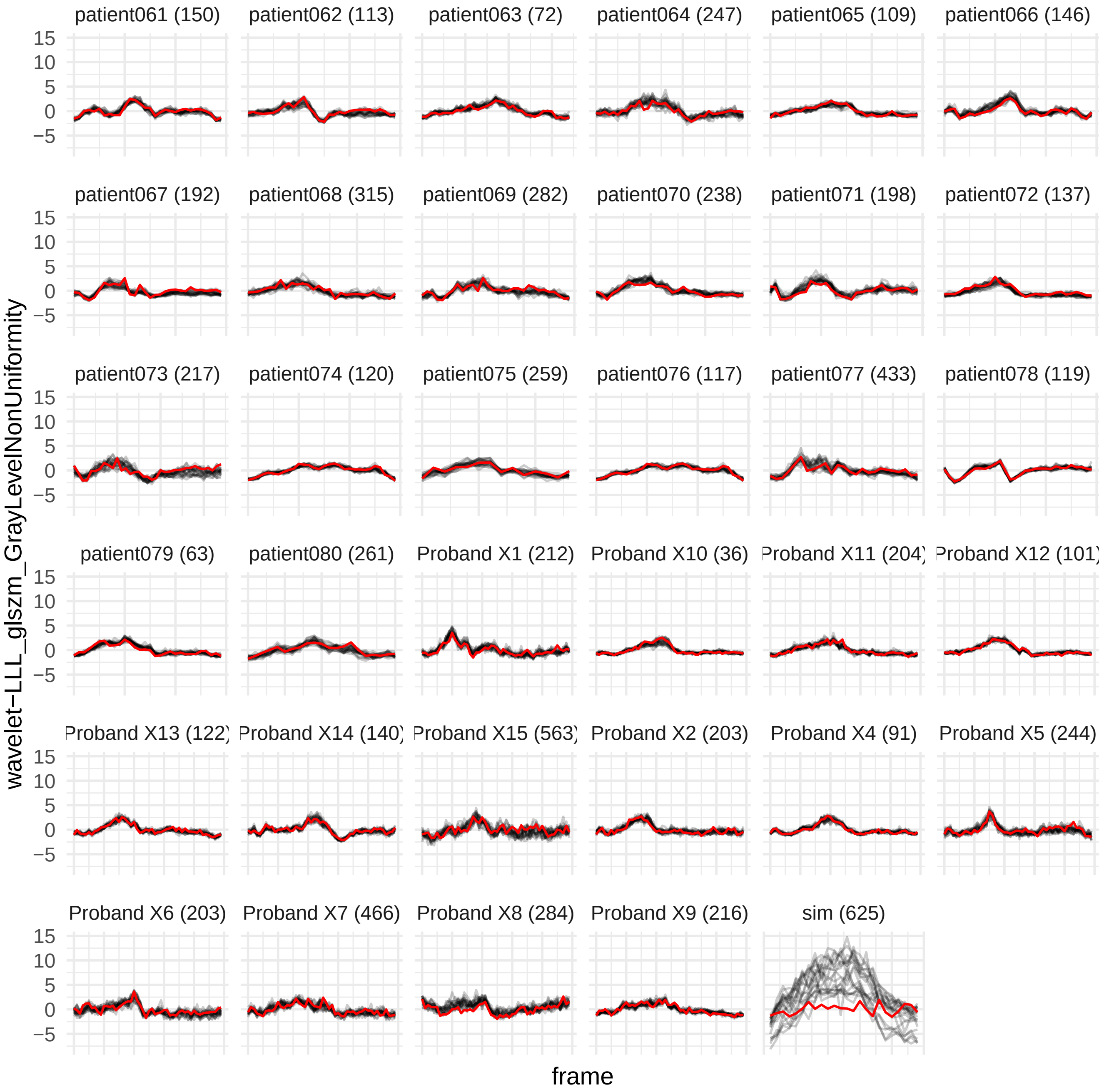

patient061 (150)
patient062 (113)
patient063 (72)
patient064 (247)
patient065 (109)
patient066 (146)
patient067 (192)
patient068 (315)
patient069 (282)
patient070 (238)
patient071 (198)
patient072 (137)
patient073 (217)
patient074 (120)
patient075 (259)
patient076 (117)
patient077 (433)
patient078 (119)
patient079 (63)
patient080 (261)
Proband X1 (212)
Proband X10 (36)
Proband X11 (204)
Proband X12 (101)
Proband X13 (122)
Proband X14 (140)
Proband X15 (563)
Proband X2 (203)
Proband X4 (91)
Proband X5 (244)
Proband X6 (203)
Proband X7 (466)
Proband X8 (284)
Proband X9 (216)
sim (625)
15
10
5
0
−5
wavelet−LLL_glszm_GrayLevelNonUniformity
frame

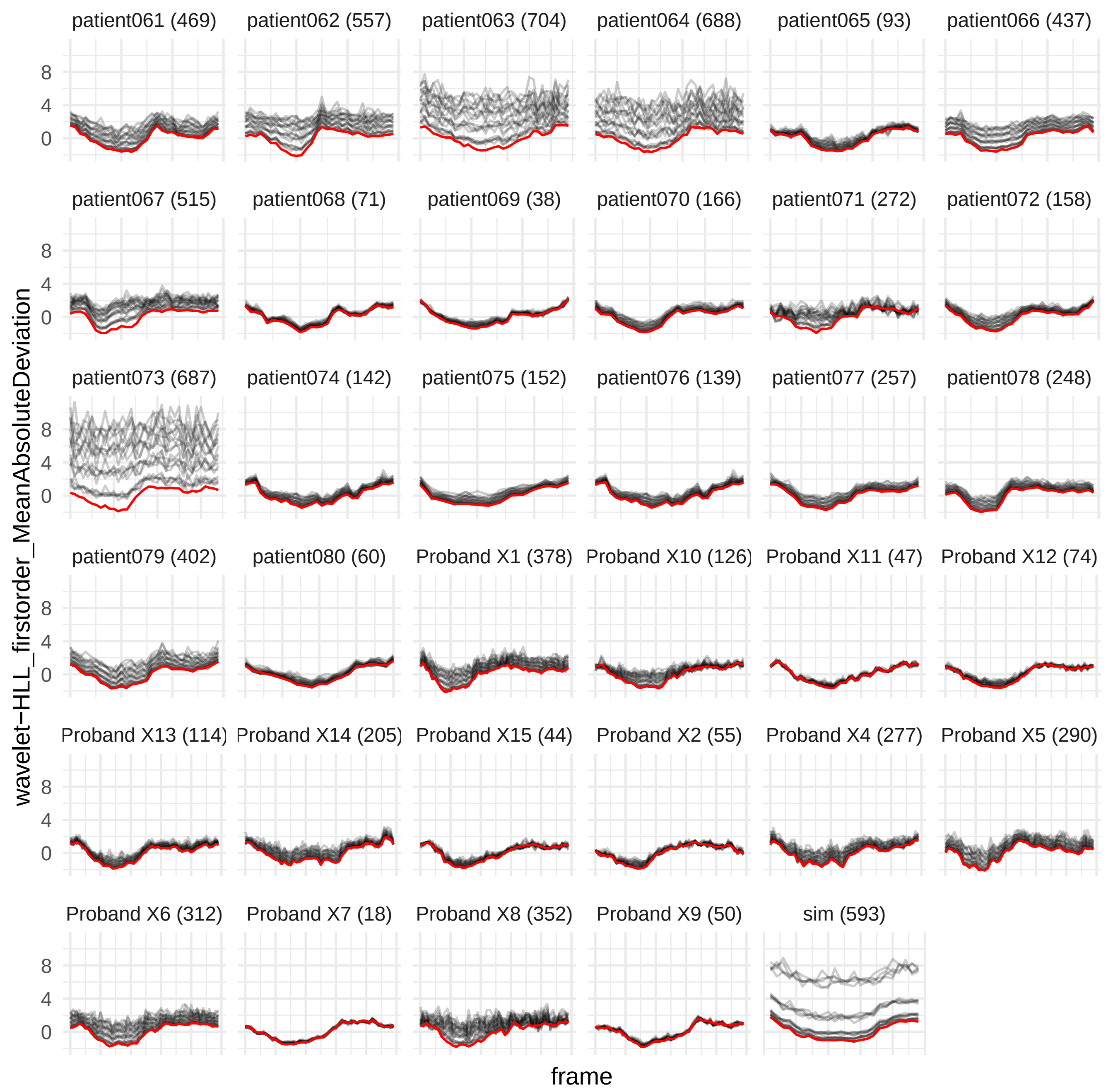

patient061 (469)
patient062 (557)
patient063 (704)
patient064 (688)
patient065 (93)
patient066 (437)
patient067 (515)
patient068 (71)
patient069 (38)
patient070 (166)
patient071 (272)
patient072 (158)
patient073 (687)
patient074 (142)
patient075 (152)
patient076 (139)
patient077 (257)
patient078 (248)
patient079 (402)
patient080 (60)
Proband X1 (378)
Proband X10 (126)
Proband X11 (47)
Proband X12 (74)
Proband X13 (114)
Proband X14 (205)
Proband X15 (44)
Proband X2 (55)
Proband X4 (277)
Proband X5 (290)
Proband X6 (312)
Proband X7 (18)
Proband X8 (352)
Proband X9 (50)
sim (593)
8
4
0
wavelet−HLL_firstorder_MeanAbsoluteDeviation
frame

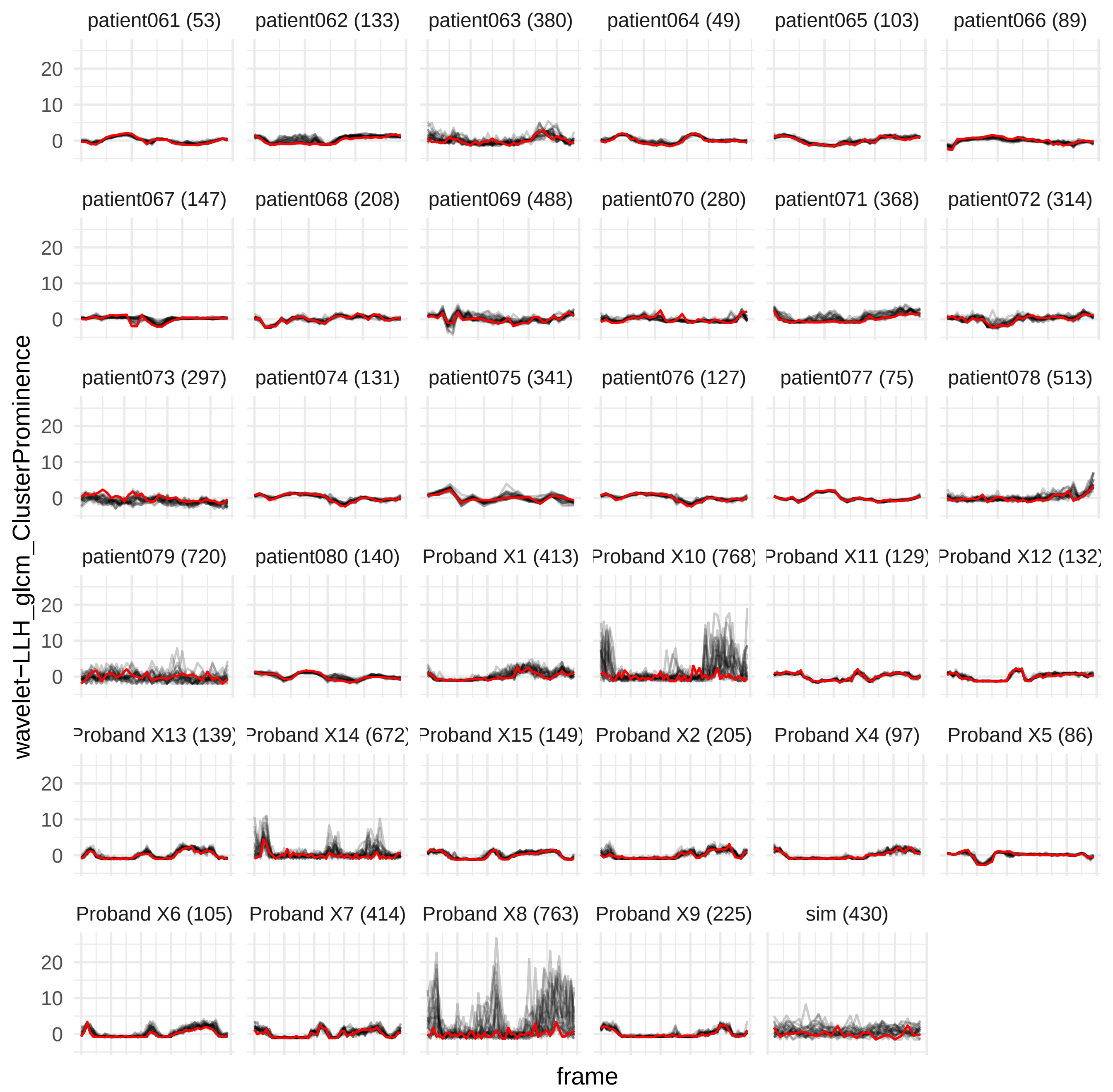

patient061 (53)
patient062 (133)
patient063 (380)
patient064 (49)
patient065 (103)
patient066 (89)
patient067 (147)
patient068 (208)
patient069 (488)
patient070 (280)
patient071 (368)
patient072 (314)
patient073 (297)
patient074 (131)
patient075 (341)
patient076 (127)
patient077 (75)
patient078 (513)
patient079 (720)
patient080 (140)
Proband X1 (413)
Proband X10 (768)
Proband X11 (129)
Proband X12 (132)
Proband X13 (139)
Proband X14 (672)
Proband X15 (149)
Proband X2 (205)
Proband X4 (97)
Proband X5 (86)
Proband X6 (105)
Proband X7 (414)
Proband X8 (763)
Proband X9 (225)
sim (430)
wavelet−LLH_glcm_ClusterProminence
frame
20
10
0

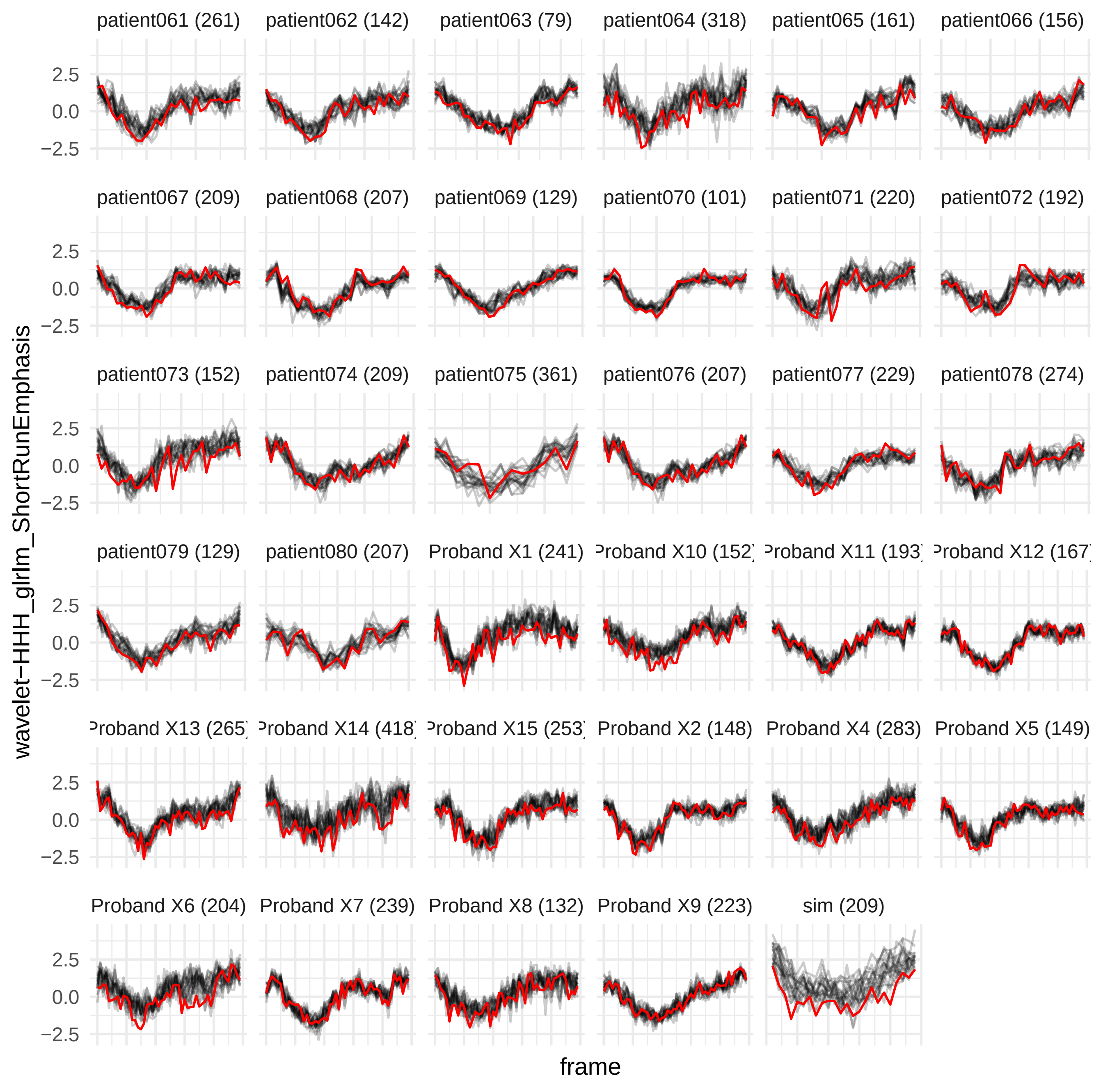
patient061 (261)
patient062 (142)
patient063 (79)
patient064 (318)
patient065 (161)
patient066 (156)
patient067 (209)
patient068 (207)
patient069 (129)
patient070 (101)
patient071 (220)
patient072 (192)
patient073 (152)
patient074 (209)
patient075 (361)
patient076 (207)
patient077 (229)
patient078 (274)
patient079 (129)
patient080 (207)
Proband X1 (241)
Proband X10 (152)
Proband X11 (193)
Proband X12 (167)
Proband X13 (265)
Proband X14 (418)
Proband X15 (253)
Proband X2 (148)
Proband X4 (283)
Proband X5 (149)
Proband X6 (204)
Proband X7 (239)
Proband X8 (132)
Proband X9 (223)
sim (209)
2.5
0.0
−2.5
wavelet−HHH_glrlm_ShortRunEmphasis
frame

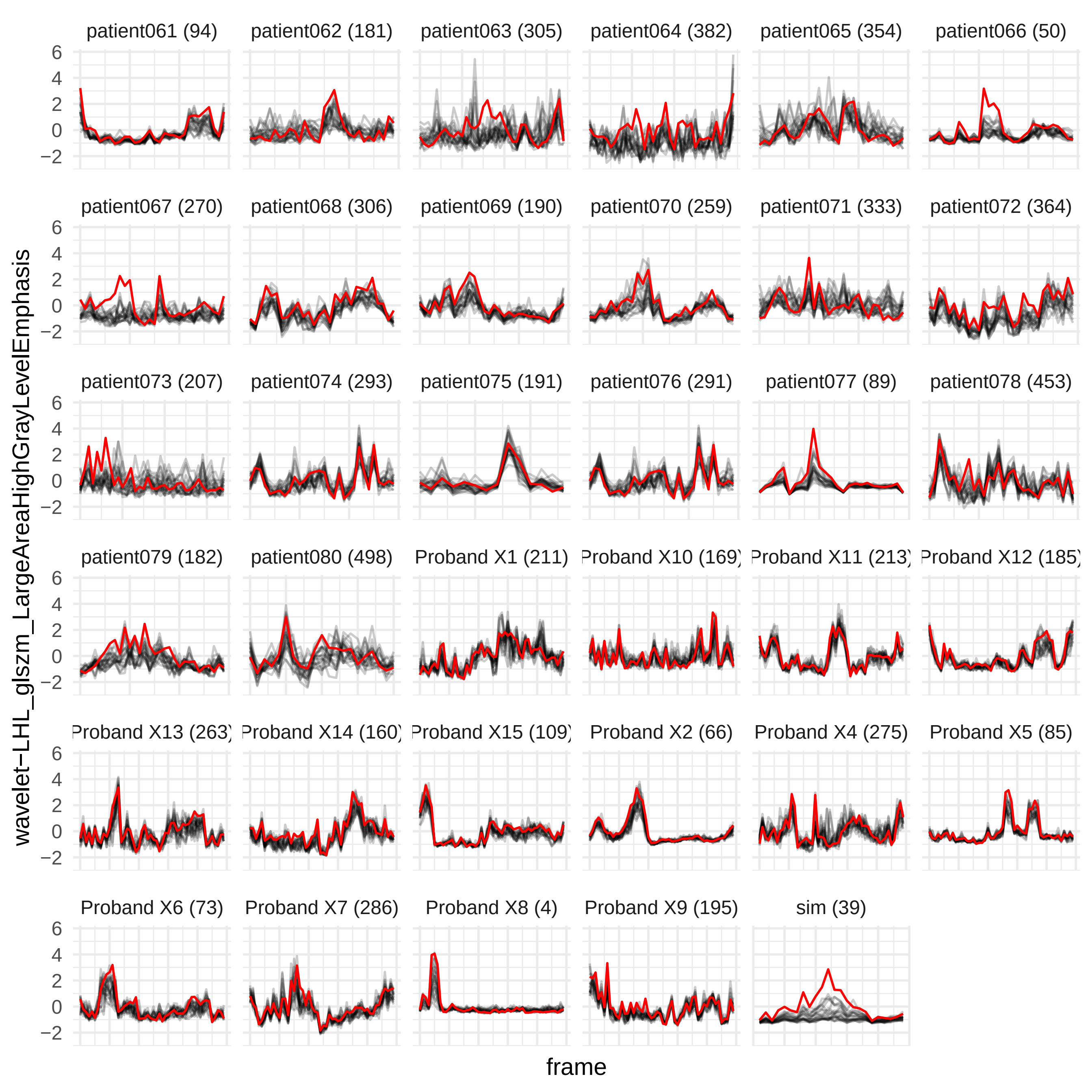

patient061 (94)
patient062 (181)
patient063 (305)
patient064 (382)
patient065 (354)
patient066 (50)
patient067 (270)
patient068 (306)
patient069 (190)
patient070 (259)
patient071 (333)
patient072 (364)
patient073 (207)
patient074 (293)
patient075 (191)
patient076 (291)
patient077 (89)
patient078 (453)
patient079 (182)
patient080 (498)
Proband X1 (211)
Proband X10 (169)
Proband X11 (213)
Proband X12 (185)
Proband X13 (263)
Proband X14 (160)
Proband X15 (109)
Proband X2 (66)
Proband X4 (275)
Proband X5 (85)
Proband X6 (73)
Proband X7 (286)
Proband X8 (4)
Proband X9 (195)
sim (39)
6
4
2
0
−2
wavelet−LHL_glszm_LargeAreaHighGrayLevelEmphasis
frame

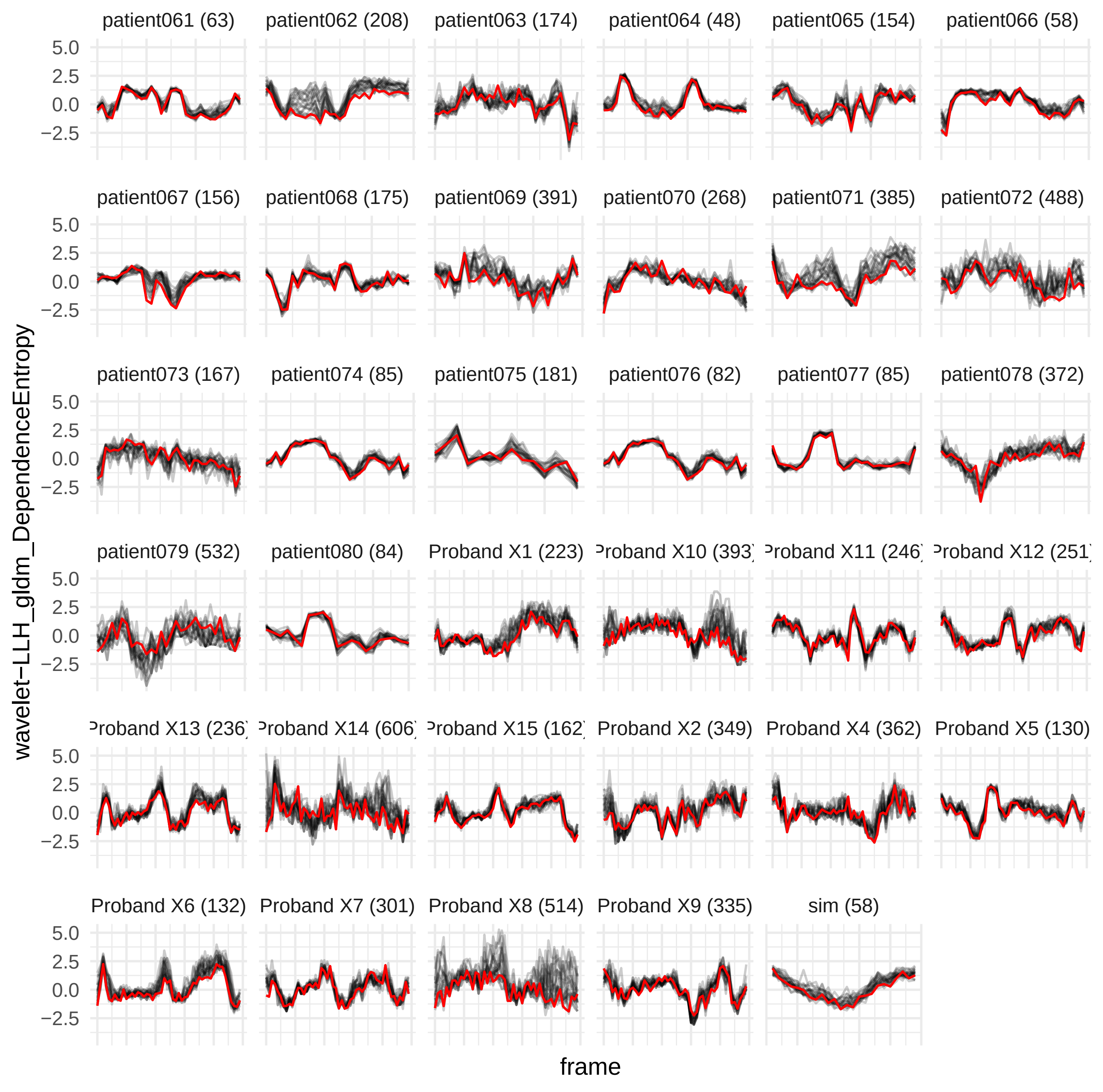

patient061 (63)
patient062 (208)
patient063 (174)
patient064 (48)
patient065 (154)
patient066 (58)
patient067 (156)
patient068 (175)
patient069 (391)
patient070 (268)
patient071 (385)
patient072 (488)
patient073 (167)
patient074 (85)
patient075 (181)
patient076 (82)
patient077 (85)
patient078 (372)
patient079 (532)
patient080 (84)
Proband X1 (223)
Proband X10 (393)
Proband X11 (246)
Proband X12 (251)
Proband X13 (236)
Proband X14 (606)
Proband X15 (162)
Proband X2 (349)
Proband X4 (362)
Proband X5 (130)
Proband X6 (132)
Proband X7 (301)
Proband X8 (514)
Proband X9 (335)
sim (58)
5.0
2.5
0.0
−2.5
wavelet−LLH_gldm_DependenceEntropy
frame

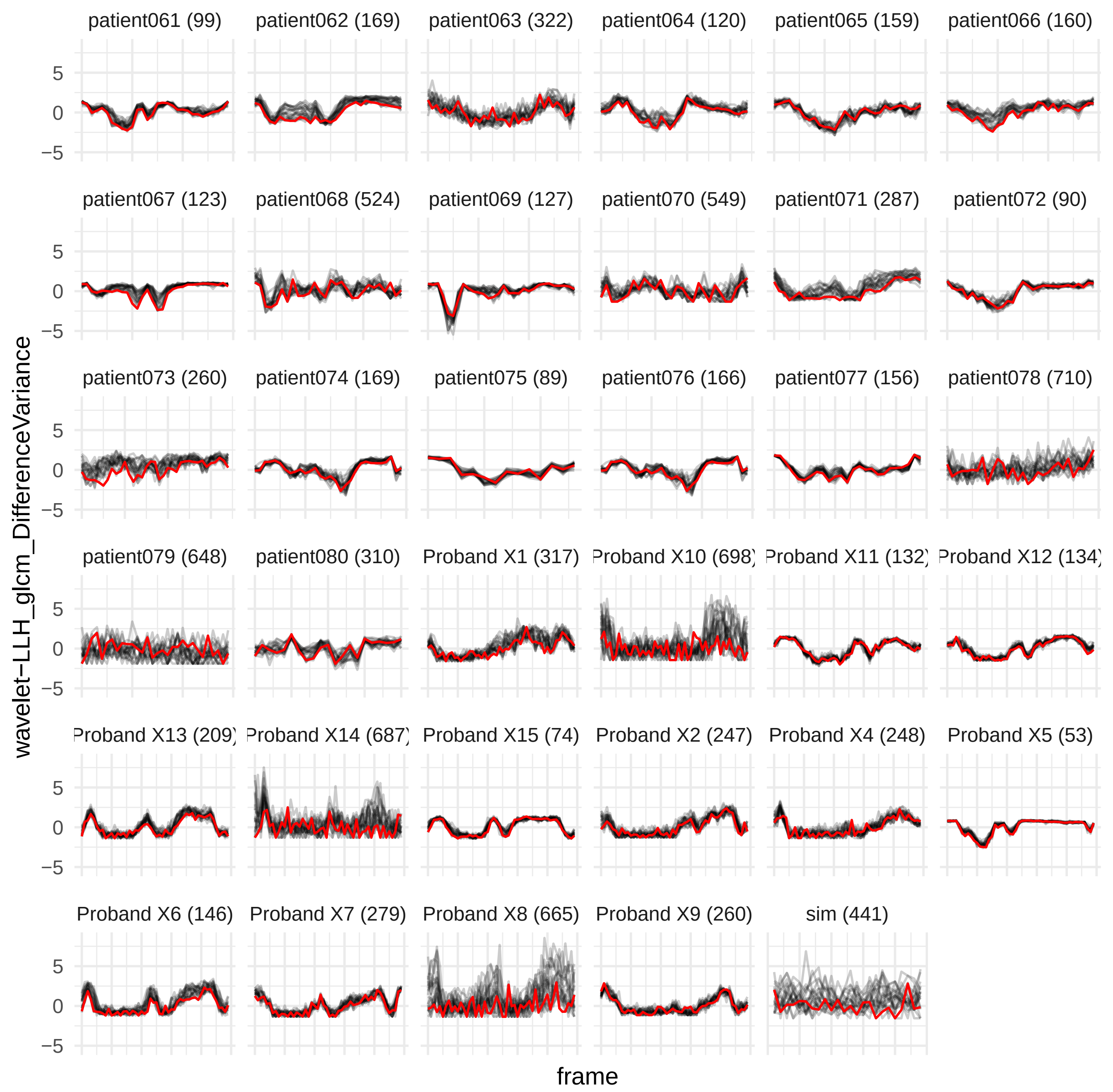
patient061 (99)
patient062 (169)
patient063 (322)
patient064 (120)
patient065 (159)
patient066 (160)
patient067 (123)
patient068 (524)
patient069 (127)
patient070 (549)
patient071 (287)
patient072 (90)
patient073 (260)
patient074 (169)
patient075 (89)
patient076 (166)
patient077 (156)
patient078 (710)
patient079 (648)
patient080 (310)
Proband X1 (317)
Proband X10 (698)
Proband X11 (132)
Proband X12 (134)
Proband X13 (209)
Proband X14 (687)
Proband X15 (74)
Proband X2 (247)
Proband X4 (248)
Proband X5 (53)
Proband X6 (146)
Proband X7 (279)
Proband X8 (665)
Proband X9 (260)
sim (441)
wavelet−LLH_glcm_DifferenceVariance
frame
5
0
−5

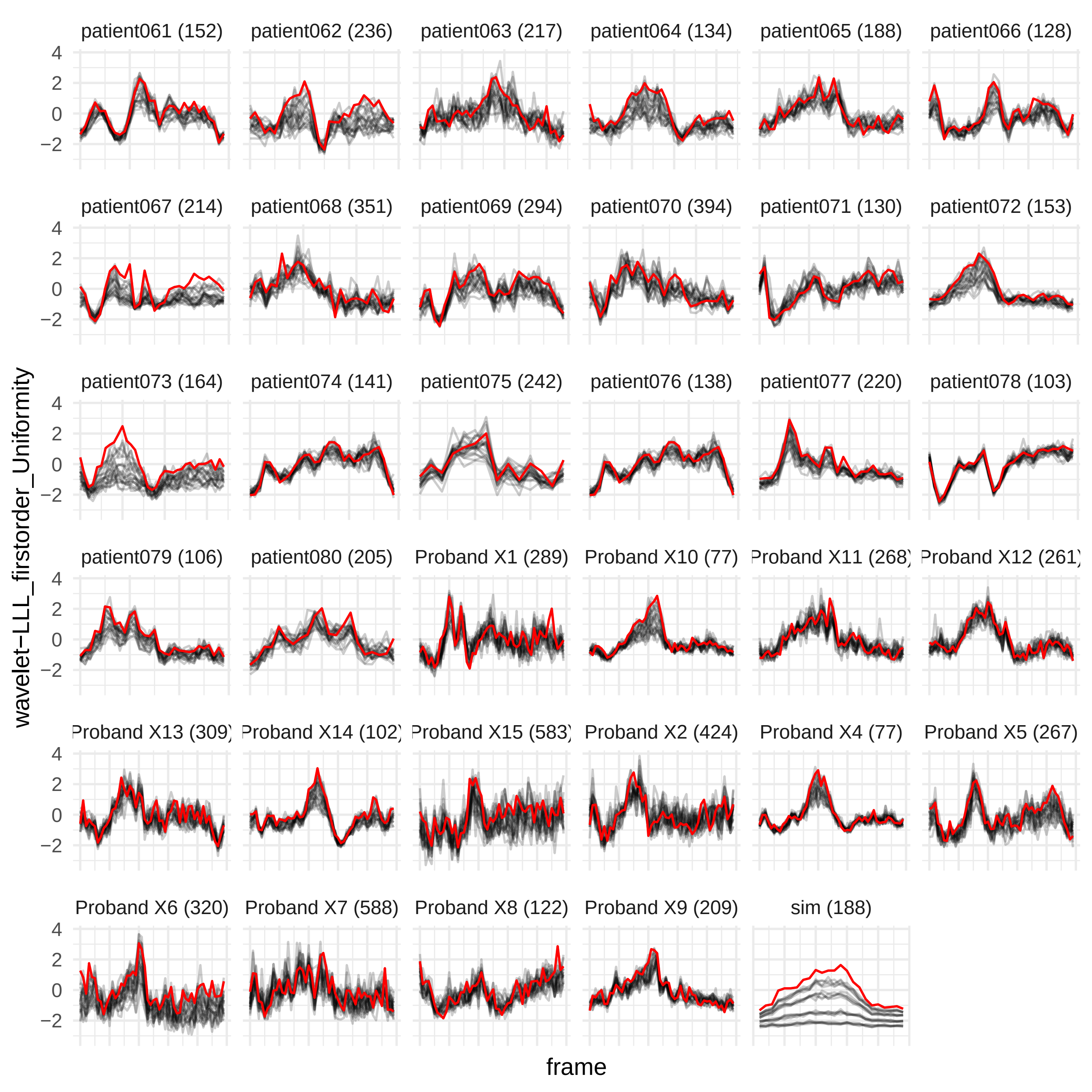
patient061 (152)
patient062 (236)
patient063 (217)
patient064 (134)
patient065 (188)
patient066 (128)
patient067 (214)
patient068 (351)
patient069 (294)
patient070 (394)
patient071 (130)
patient072 (153)
patient073 (164)
patient074 (141)
patient075 (242)
patient076 (138)
patient077 (220)
patient078 (103)
patient079 (106)
patient080 (205)
Proband X1 (289)
Proband X10 (77)
Proband X11 (268)
Proband X12 (261)
Proband X13 (309)
Proband X14 (102)
Proband X15 (583)
Proband X2 (424)
Proband X4 (77)
Proband X5 (267)
Proband X6 (320)
Proband X7 (588)
Proband X8 (122)
Proband X9 (209)
sim (188)
wavelet−LLL_firstorder_Uniformity
frame
4
2
0
−2

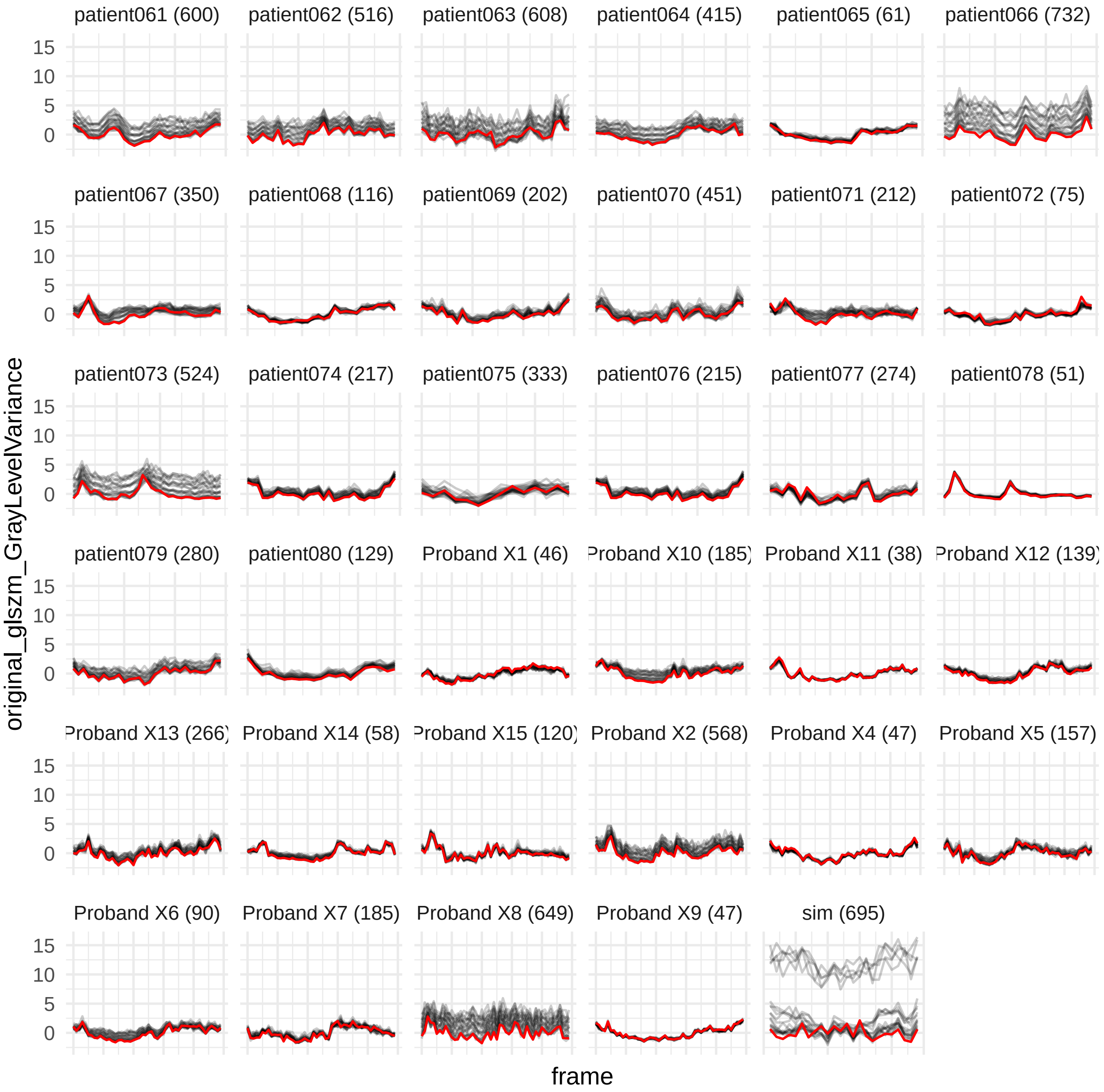

patient061 (600)
patient062 (516)
patient063 (608)
patient064 (415)
patient065 (61)
patient066 (732)
patient067 (350)
patient068 (116)
patient069 (202)
patient070 (451)
patient071 (212)
patient072 (75)
patient073 (524)
patient074 (217)
patient075 (333)
patient076 (215)
patient077 (274)
patient078 (51)
patient079 (280)
patient080 (129)
Proband X1 (46)
Proband X10 (185)
Proband X11 (38)
Proband X12 (139)
Proband X13 (266)
Proband X14 (58)
Proband X15 (120)
Proband X2 (568)
Proband X4 (47)
Proband X5 (157)
Proband X6 (90)
Proband X7 (185)
Proband X8 (649)
Proband X9 (47)
sim (695)
original_glszm_GrayLevelVariance
frame
0
5
10
15

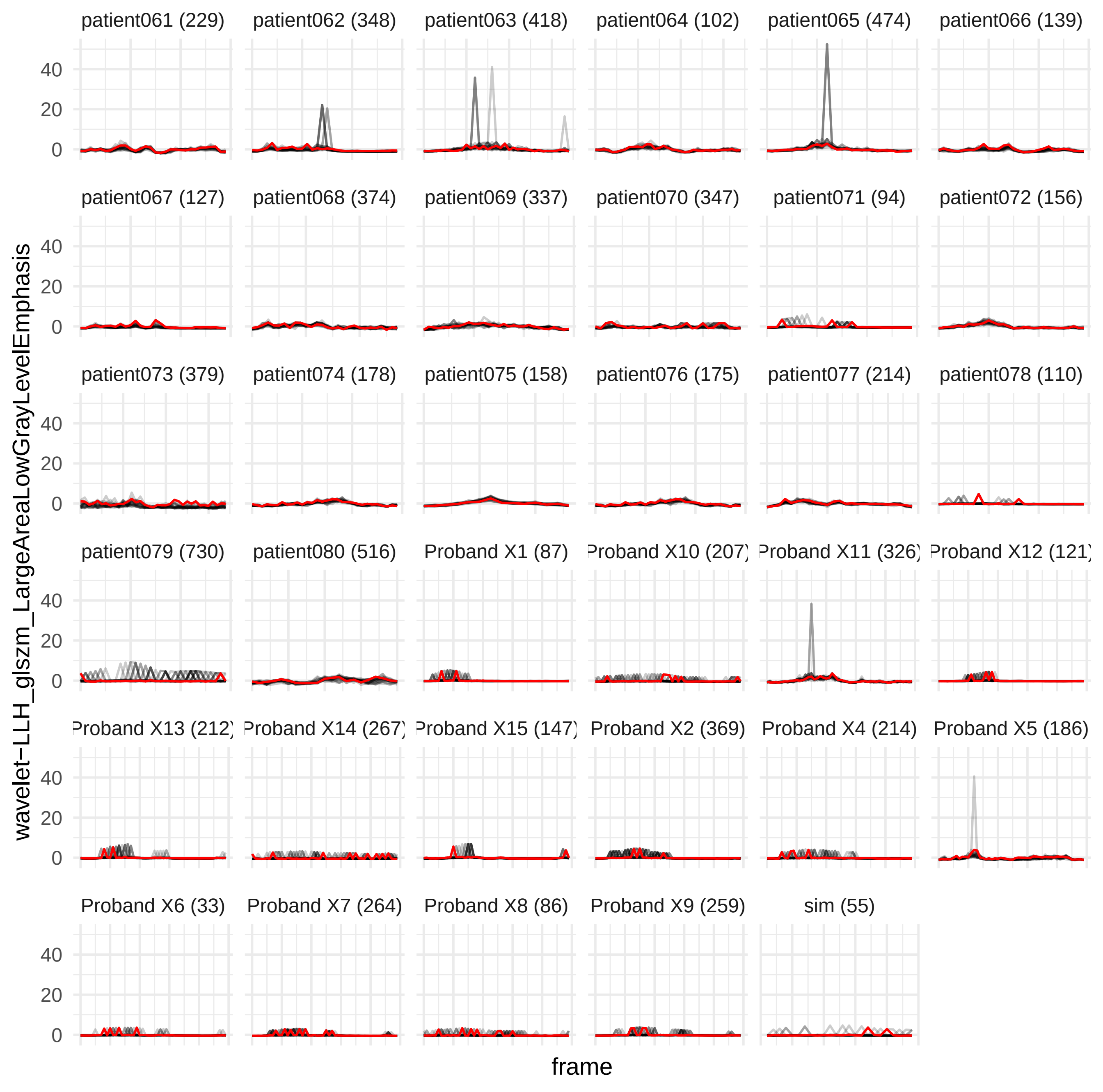

patient061 (229)
patient062 (348)
patient063 (418)
patient064 (102)
patient065 (474)
patient066 (139)
patient067 (127)
patient068 (374)
patient069 (337)
patient070 (347)
patient071 (94)
patient072 (156)
patient073 (379)
patient074 (178)
patient075 (158)
patient076 (175)
patient077 (214)
patient078 (110)
patient079 (730)
patient080 (516)
Proband X1 (87)
Proband X10 (207)
Proband X11 (326)
Proband X12 (121)
Proband X13 (212)
Proband X14 (267)
Proband X15 (147)
Proband X2 (369)
Proband X4 (214)
Proband X5 (186)
Proband X6 (33)
Proband X7 (264)
Proband X8 (86)
Proband X9 (259)
sim (55)
40
20
0
wavelet−LLH_glszm_LargeAreaLowGrayLevelEmphasis
frame

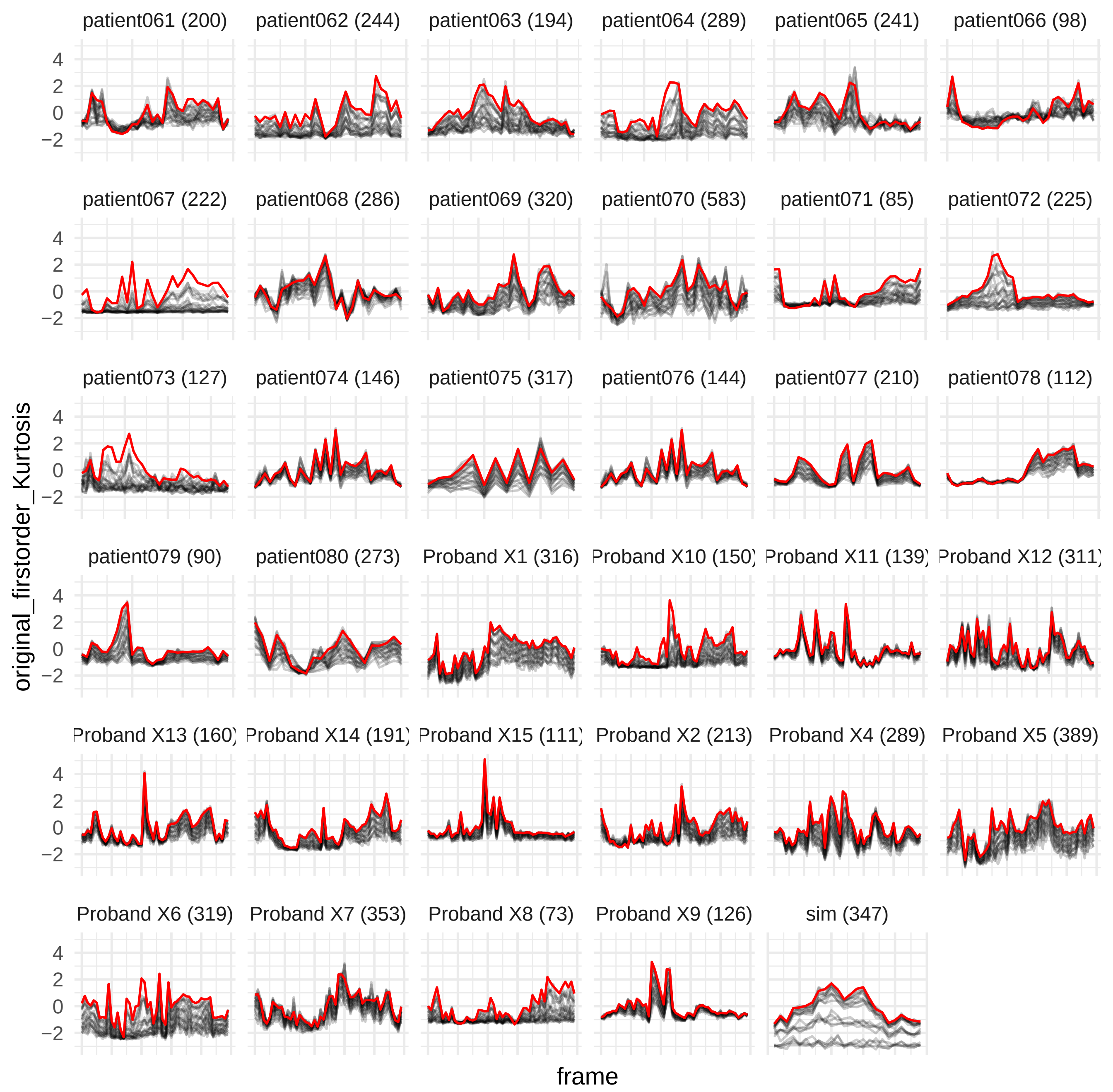

patient061 (200)
patient062 (244)
patient063 (194)
patient064 (289)
patient065 (241)
patient066 (98)
patient067 (222)
patient068 (286)
patient069 (320)
patient070 (583)
patient071 (85)
patient072 (225)
patient073 (127)
patient074 (146)
patient075 (317)
patient076 (144)
patient077 (210)
patient078 (112)
patient079 (90)
patient080 (273)
Proband X1 (316)
Proband X10 (150)
Proband X11 (139)
Proband X12 (311)
Proband X13 (160)
Proband X14 (191)
Proband X15 (111)
Proband X2 (213)
Proband X4 (289)
Proband X5 (389)
Proband X6 (319)
Proband X7 (353)
Proband X8 (73)
Proband X9 (126)
sim (347)
4
2
0
−2
original_firstorder_Kurtosis
frame

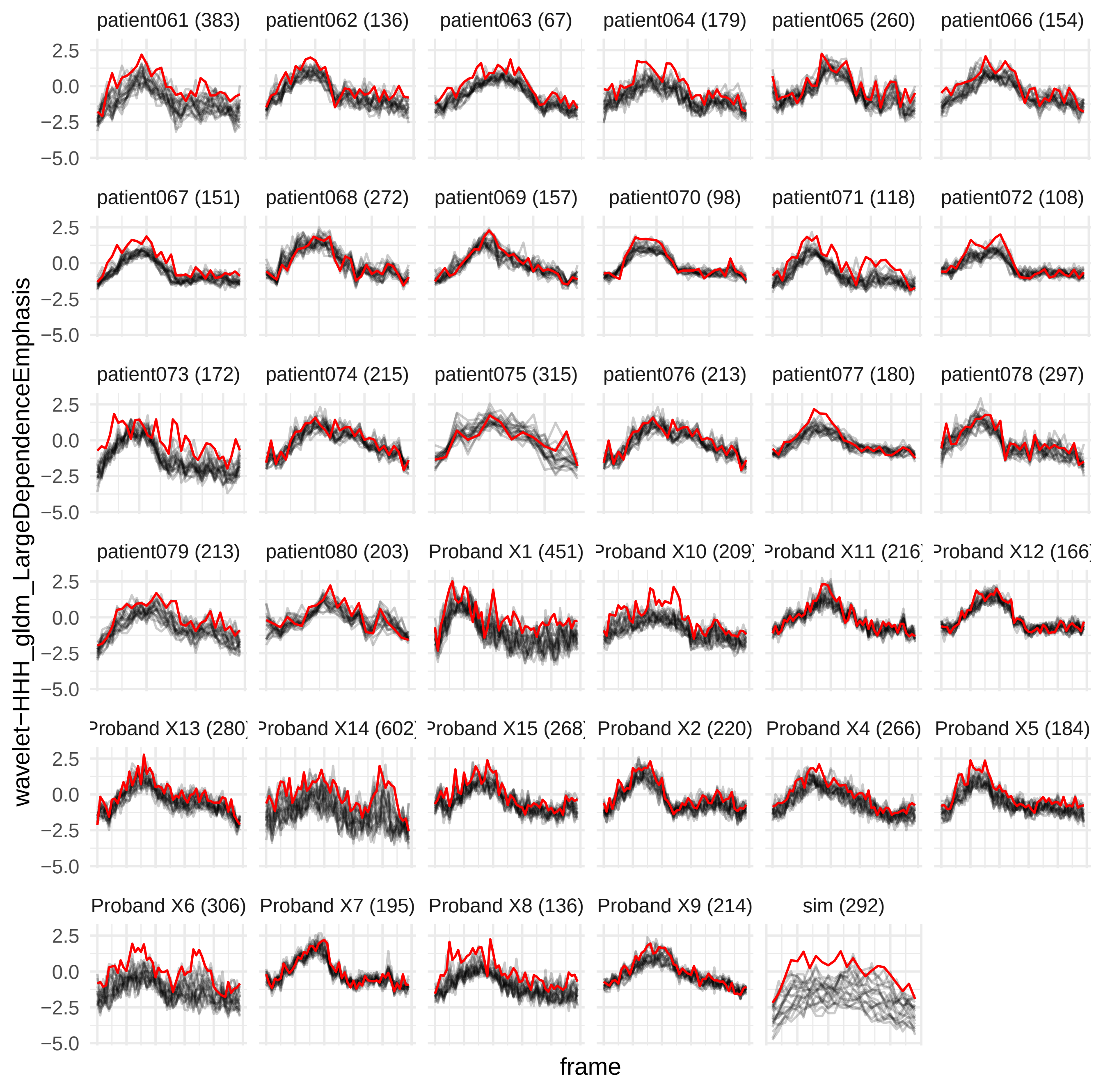

patient061 (383)
patient062 (136)
patient063 (67)
patient064 (179)
patient065 (260)
patient066 (154)
patient067 (151)
patient068 (272)
patient069 (157)
patient070 (98)
patient071 (118)
patient072 (108)
patient073 (172)
patient074 (215)
patient075 (315)
patient076 (213)
patient077 (180)
patient078 (297)
patient079 (213)
patient080 (203)
Proband X1 (451)
Proband X10 (209)
Proband X11 (216)
Proband X12 (166)
Proband X13 (280)
Proband X14 (602)
Proband X15 (268)
Proband X2 (220)
Proband X4 (266)
Proband X5 (184)
Proband X6 (306)
Proband X7 (195)
Proband X8 (136)
Proband X9 (214)
sim (292)
2.5
0.0
−2.5
−5.0
wavelet−HHH_gldm_LargeDependenceEmphasis
frame

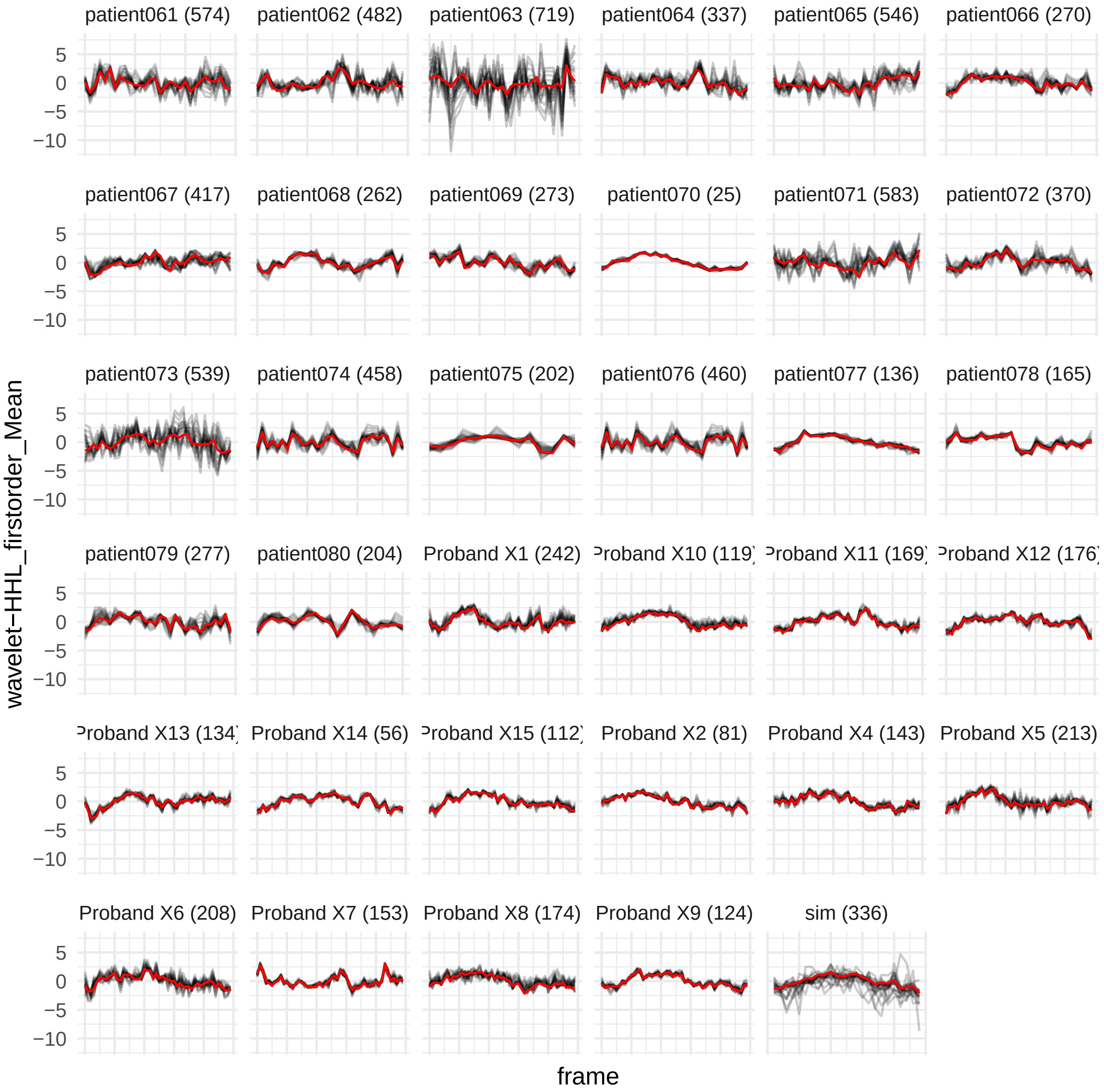
patient061 (574)
patient062 (482)
patient063 (719)
patient064 (337)
patient065 (546)
patient066 (270)
patient067 (417)
patient068 (262)
patient069 (273)
patient070 (25)
patient071 (583)
patient072 (370)
patient073 (539)
patient074 (458)
patient075 (202)
patient076 (460)
patient077 (136)
patient078 (165)
patient079 (277)
patient080 (204)
Proband X1 (242)
Proband X10 (119)
Proband X11 (169)
Proband X12 (176)
Proband X13 (134)
Proband X14 (56)
Proband X15 (112)
Proband X2 (81)
Proband X4 (143)
Proband X5 (213)
Proband X6 (208)
Proband X7 (153)
Proband X8 (174)
Proband X9 (124)
sim (336)
5
0
−5
−10
wavelet−HHL_firstorder_Mean
frame

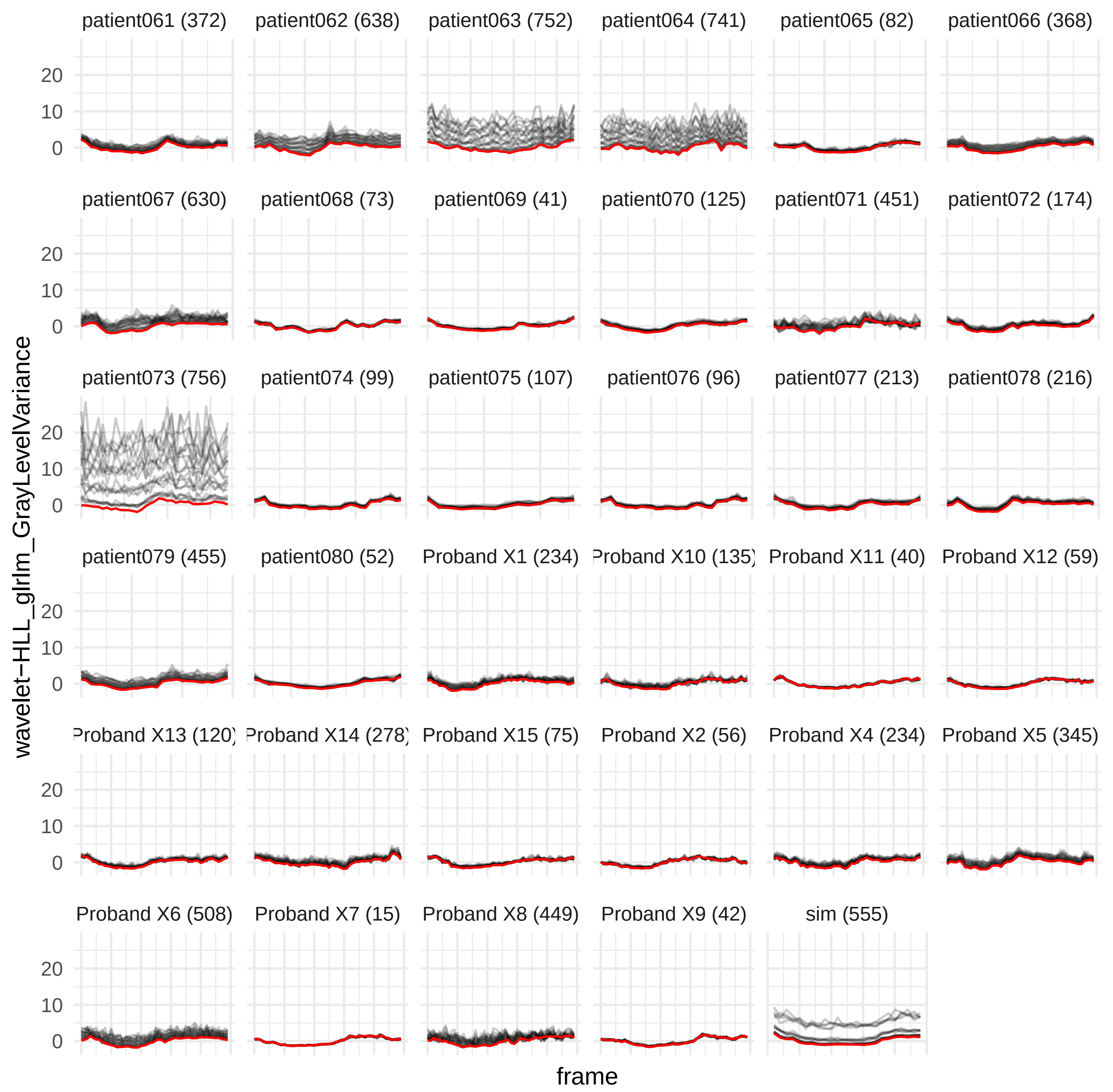
patient061 (372)
patient062 (638)
patient063 (752)
patient064 (741)
patient065 (82)
patient066 (368)
patient067 (630)
patient068 (73)
patient069 (41)
patient070 (125)
patient071 (451)
patient072 (174)
patient073 (756)
patient074 (99)
patient075 (107)
patient076 (96)
patient077 (213)
patient078 (216)
patient079 (455)
patient080 (52)
Proband X1 (234)
Proband X10 (135)
Proband X11 (40)
Proband X12 (59)
Proband X13 (120)
Proband X14 (278)
Proband X15 (75)
Proband X2 (56)
Proband X4 (234)
Proband X5 (345)
Proband X6 (508)
Proband X7 (15)
Proband X8 (449)
Proband X9 (42)
sim (555)
0
10
20
wavelet−HLL_glrlm_GrayLevelVariance
frame

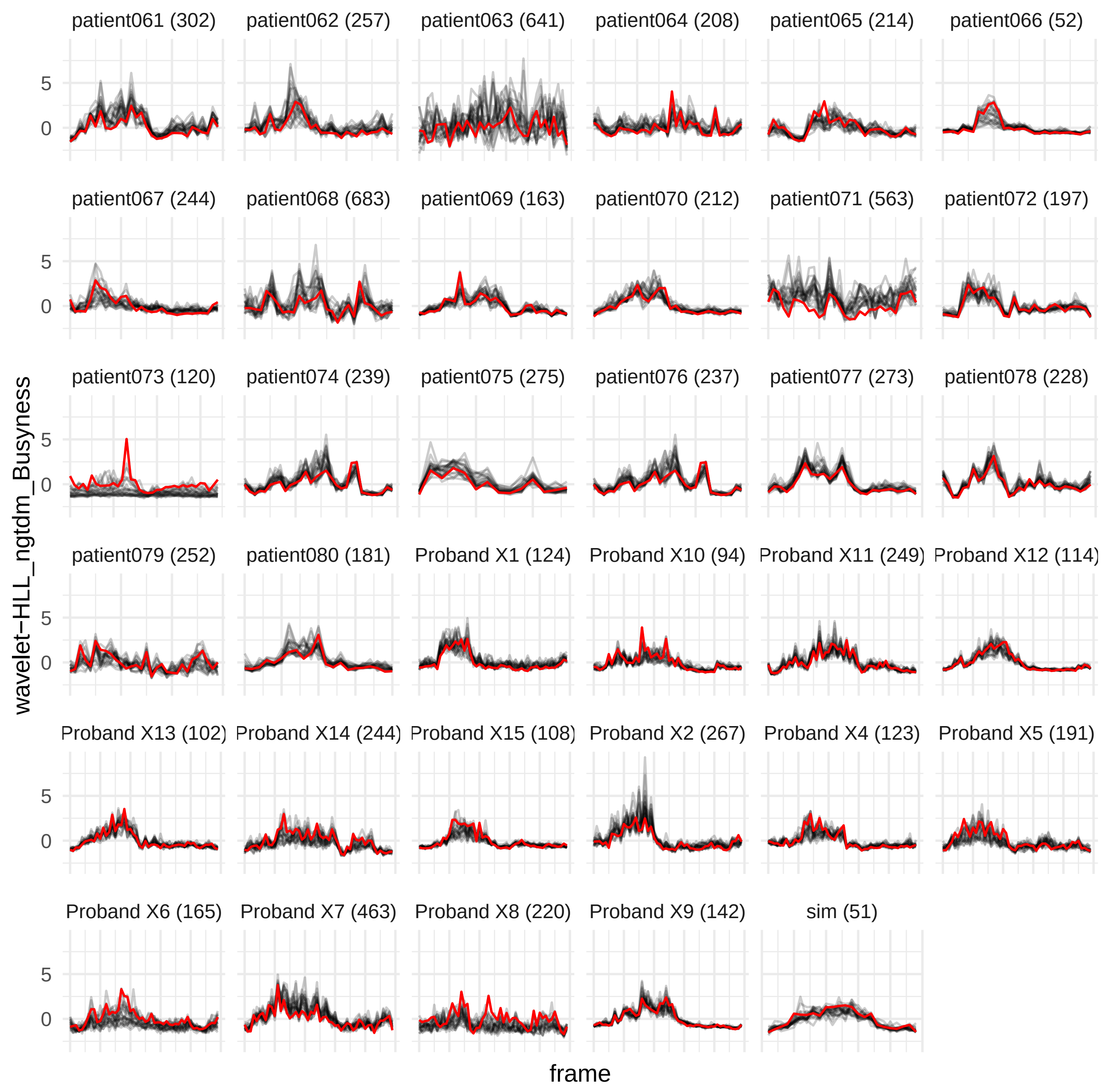

patient061 (302)
patient062 (257)
patient063 (641)
patient064 (208)
patient065 (214)
patient066 (52)
patient067 (244)
patient068 (683)
patient069 (163)
patient070 (212)
patient071 (563)
patient072 (197)
patient073 (120)
patient074 (239)
patient075 (275)
patient076 (237)
patient077 (273)
patient078 (228)
patient079 (252)
patient080 (181)
Proband X1 (124)
Proband X10 (94)
Proband X11 (249)
Proband X12 (114)
Proband X13 (102)
Proband X14 (244)
Proband X15 (108)
Proband X2 (267)
Proband X4 (123)
Proband X5 (191)
Proband X6 (165)
Proband X7 (463)
Proband X8 (220)
Proband X9 (142)
sim (51)
5
0
wavelet−HLL_ngtdm_Busyness
frame

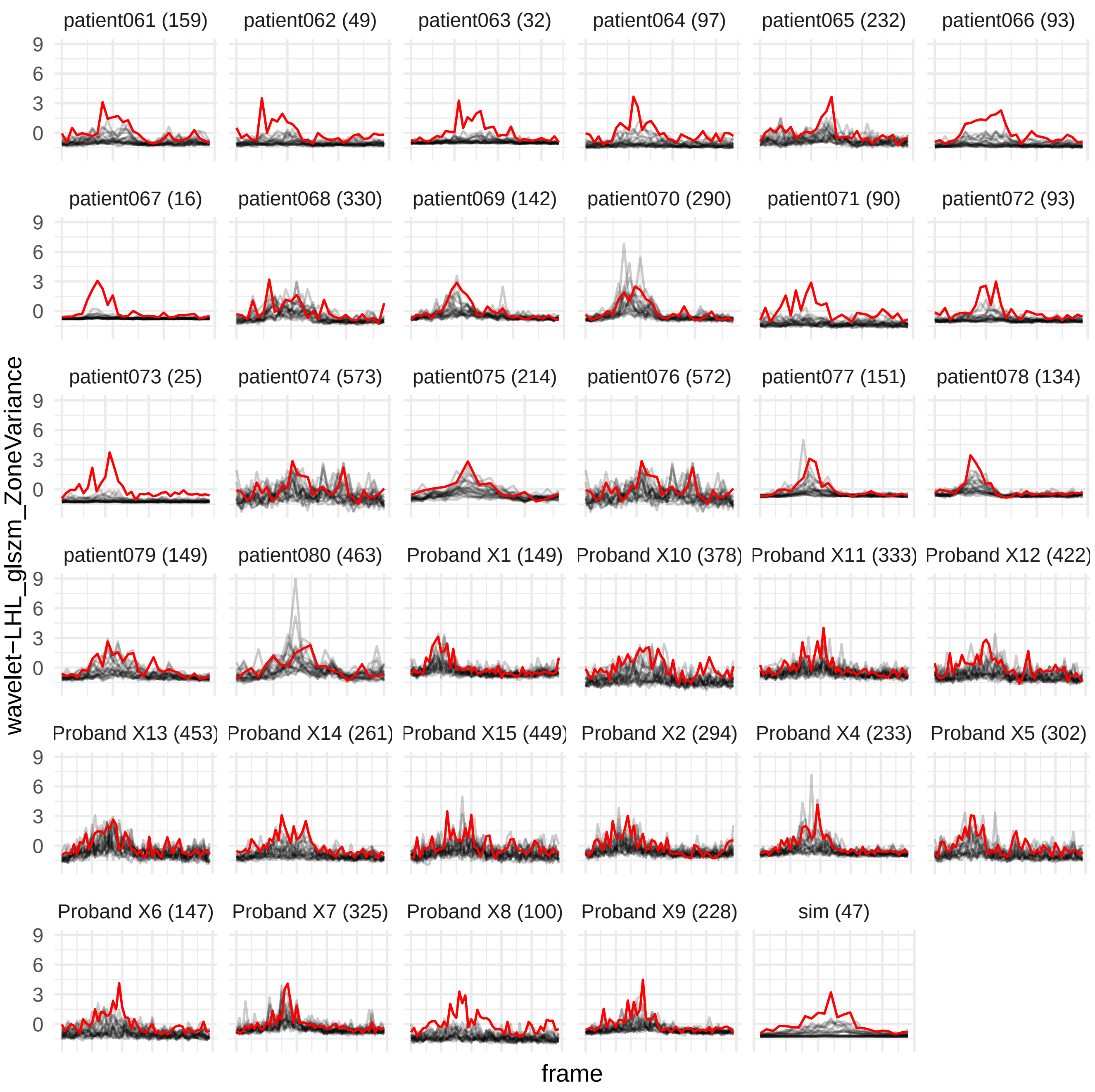

patient061 (159)
patient062 (49)
patient063 (32)
patient064 (97)
patient065 (232)
patient066 (93)
patient067 (16)
patient068 (330)
patient069 (142)
patient070 (290)
patient071 (90)
patient072 (93)
patient073 (25)
patient074 (573)
patient075 (214)
patient076 (572)
patient077 (151)
patient078 (134)
patient079 (149)
patient080 (463)
Proband X1 (149)
Proband X10 (378)
Proband X11 (333)
Proband X12 (422)
Proband X13 (453)
Proband X14 (261)
Proband X15 (449)
Proband X2 (294)
Proband X4 (233)
Proband X5 (302)
Proband X6 (147)
Proband X7 (325)
Proband X8 (100)
Proband X9 (228)
sim (47)
9
6
3
0
wavelet−LHL_glszm_ZoneVariance
frame

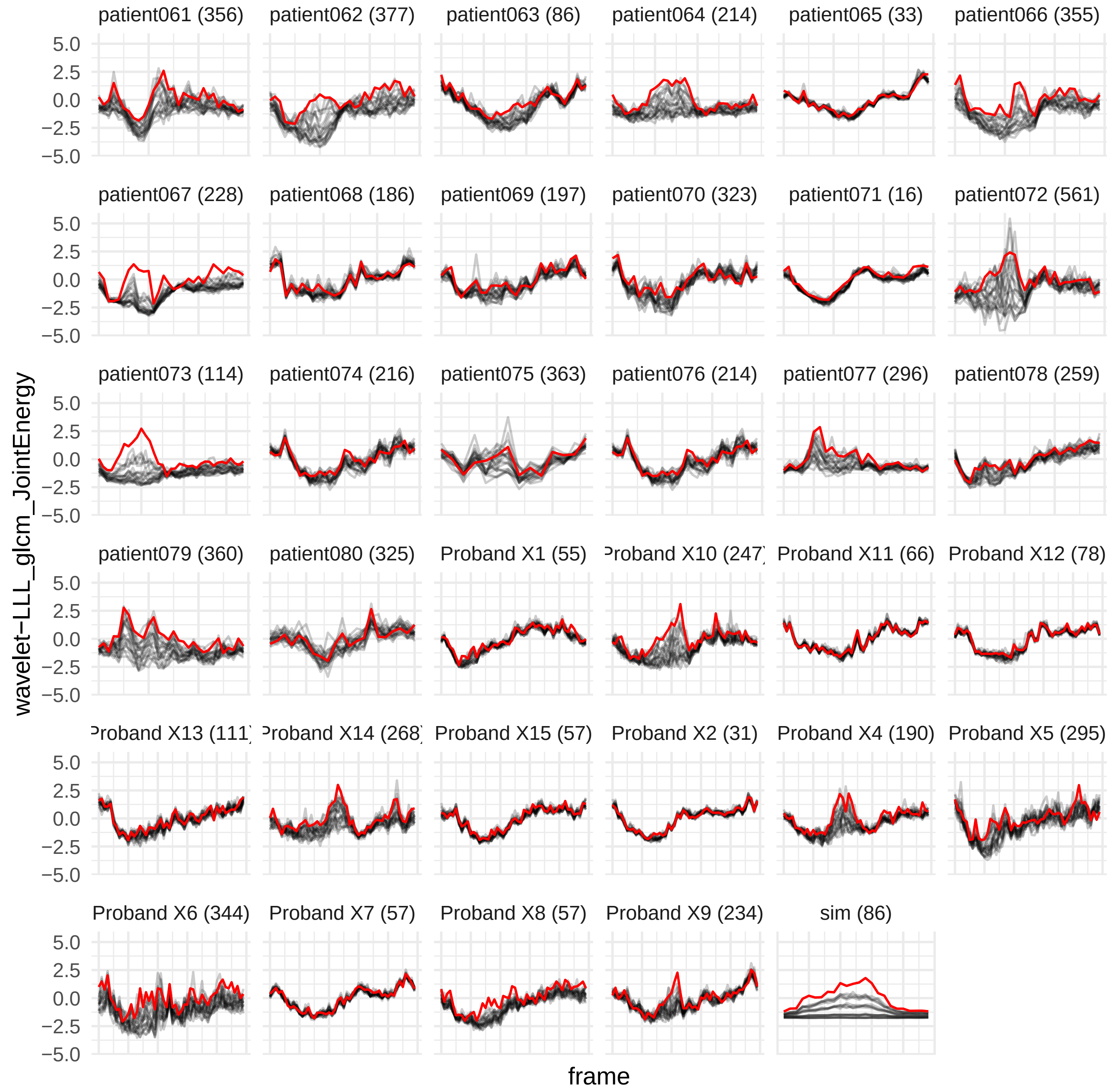

patient061 (356)
patient062 (377)
patient063 (86)
patient064 (214)
patient065 (33)
patient066 (355)
patient067 (228)
patient068 (186)
patient069 (197)
patient070 (323)
patient071 (16)
patient072 (561)
patient073 (114)
patient074 (216)
patient075 (363)
patient076 (214)
patient077 (296)
patient078 (259)
patient079 (360)
patient080 (325)
Proband X1 (55)
Proband X10 (247)
Proband X11 (66)
Proband X12 (78)
Proband X13 (111)
Proband X14 (268)
Proband X15 (57)
Proband X2 (31)
Proband X4 (190)
Proband X5 (295)
Proband X6 (344)
Proband X7 (57)
Proband X8 (57)
Proband X9 (234)
sim (86)
5.0
2.5
0.0
−2.5
−5.0
wavelet−LLL_glcm_JointEnergy
frame

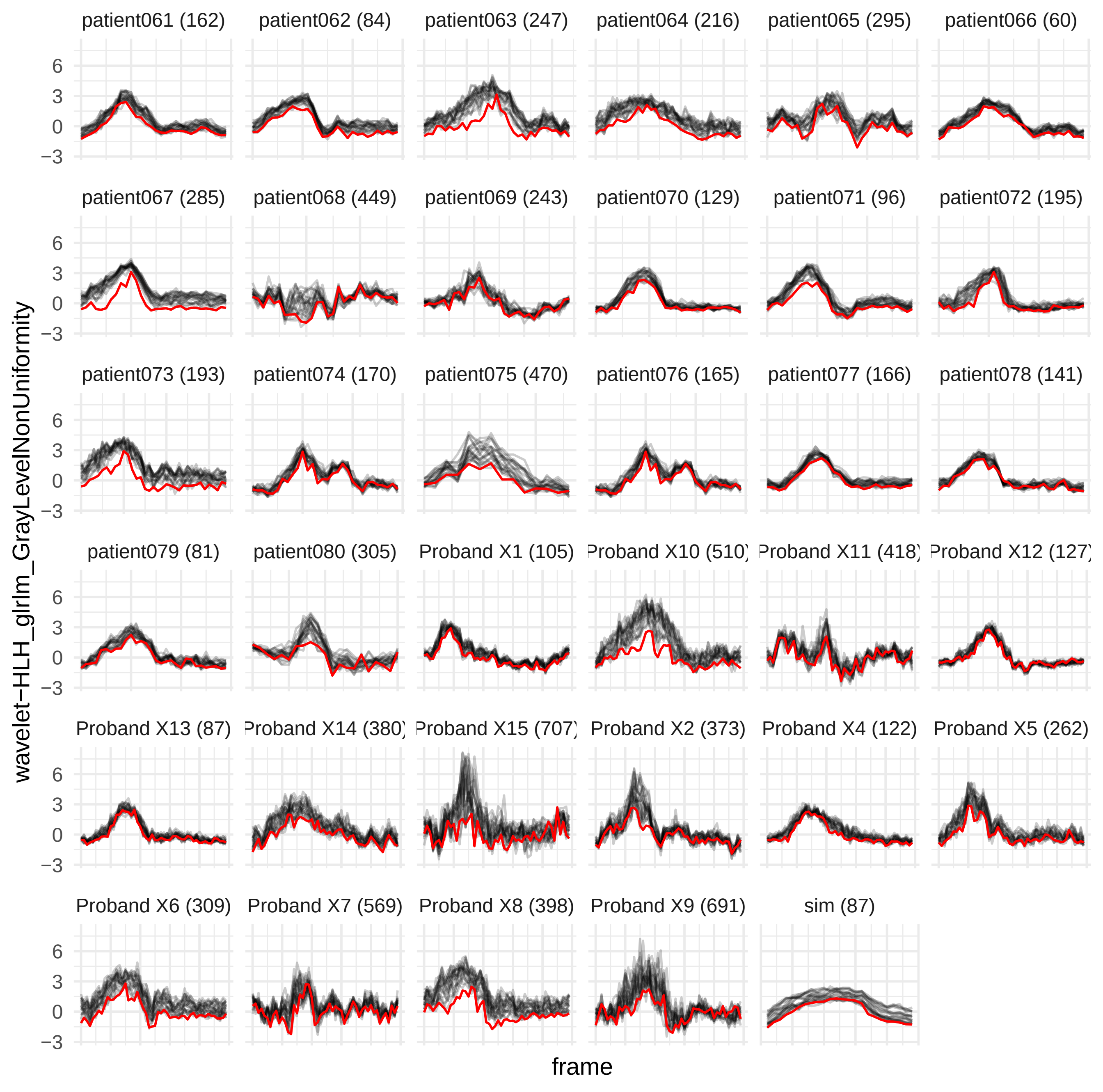

patient061 (162)
patient062 (84)
patient063 (247)
patient064 (216)
patient065 (295)
patient066 (60)
patient067 (285)
patient068 (449)
patient069 (243)
patient070 (129)
patient071 (96)
patient072 (195)
patient073 (193)
patient074 (170)
patient075 (470)
patient076 (165)
patient077 (166)
patient078 (141)
patient079 (81)
patient080 (305)
Proband X1 (105)
Proband X10 (510)
Proband X11 (418)
Proband X12 (127)
Proband X13 (87)
Proband X14 (380)
Proband X15 (707)
Proband X2 (373)
Proband X4 (122)
Proband X5 (262)
Proband X6 (309)
Proband X7 (569)
Proband X8 (398)
Proband X9 (691)
sim (87)
6
3
0
−3
wavelet−HLH_glrlm_GrayLevelNonUniformity
frame

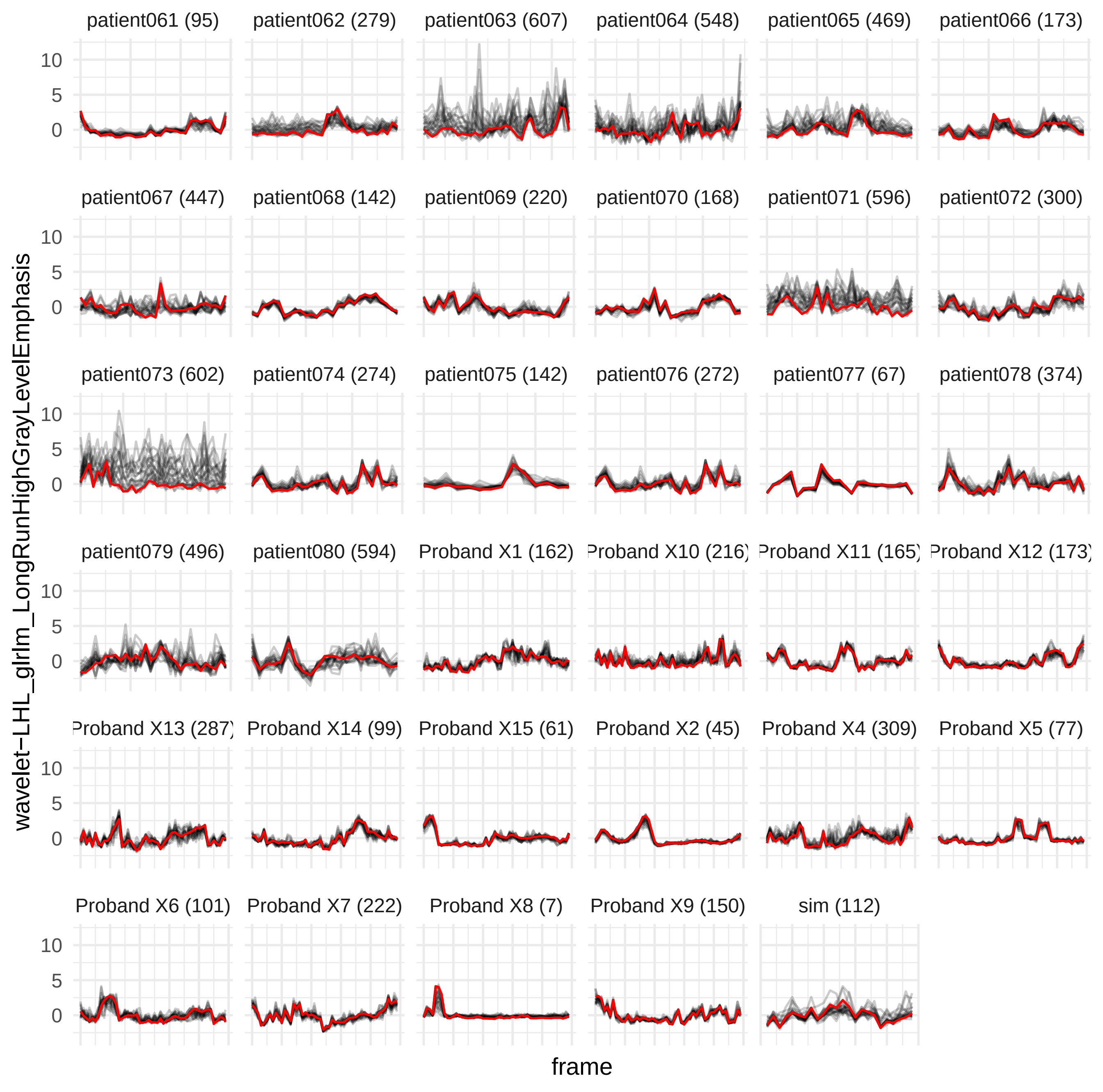
patient061 (95)
patient062 (279)
patient063 (607)
patient064 (548)
patient065 (469)
patient066 (173)
patient067 (447)
patient068 (142)
patient069 (220)
patient070 (168)
patient071 (596)
patient072 (300)
patient073 (602)
patient074 (274)
patient075 (142)
patient076 (272)
patient077 (67)
patient078 (374)
patient079 (496)
patient080 (594)
Proband X1 (162)
Proband X10 (216)
Proband X11 (165)
Proband X12 (173)
Proband X13 (287)
Proband X14 (99)
Proband X15 (61)
Proband X2 (45)
Proband X4 (309)
Proband X5 (77)
Proband X6 (101)
Proband X7 (222)
Proband X8 (7)
Proband X9 (150)
sim (112)
10
5
0
wavelet−LHL_glrlm_LongRunHighGrayLevelEmphasis
frame

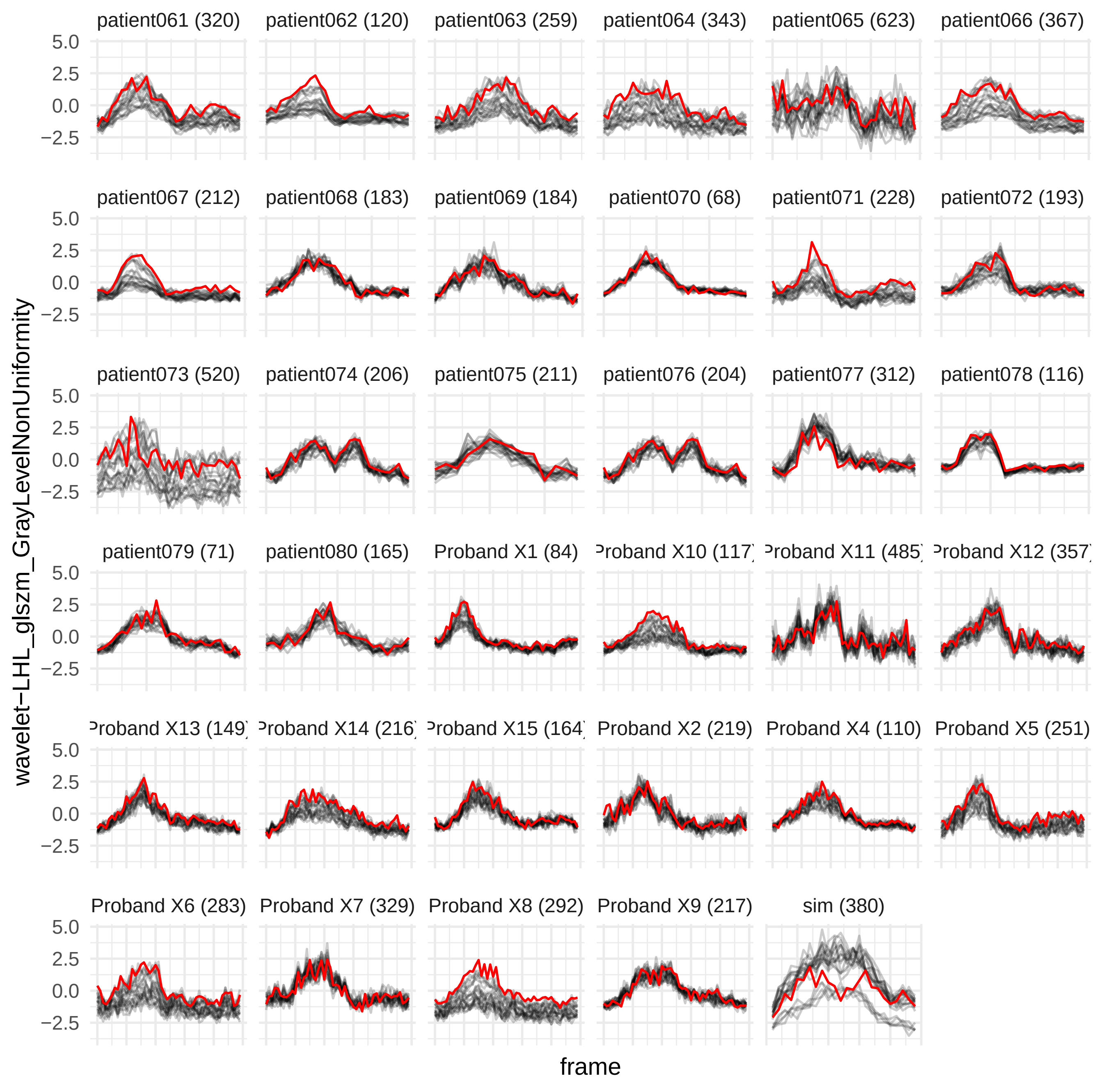
patient061 (320)
patient062 (120)
patient063 (259)
patient064 (343)
patient065 (623)
patient066 (367)
patient067 (212)
patient068 (183)
patient069 (184)
patient070 (68)
patient071 (228)
patient072 (193)
patient073 (520)
patient074 (206)
patient075 (211)
patient076 (204)
patient077 (312)
patient078 (116)
patient079 (71)
patient080 (165)
Proband X1 (84)
Proband X10 (117
Proband X11 (485
Proband X12 (357
Proband X13 (149
Proband X14 (216
Proband X15 (164
Proband X2 (219)
Proband X4 (110)
Proband X5 (251)
Proband X6 (283)
Proband X7 (329)
Proband X8 (292)
Proband X9 (217)
sim (380)
5.0
2.5
0.0
−2.5
wavelet−LHL_glszm_GrayLevelNonUniformity
frame

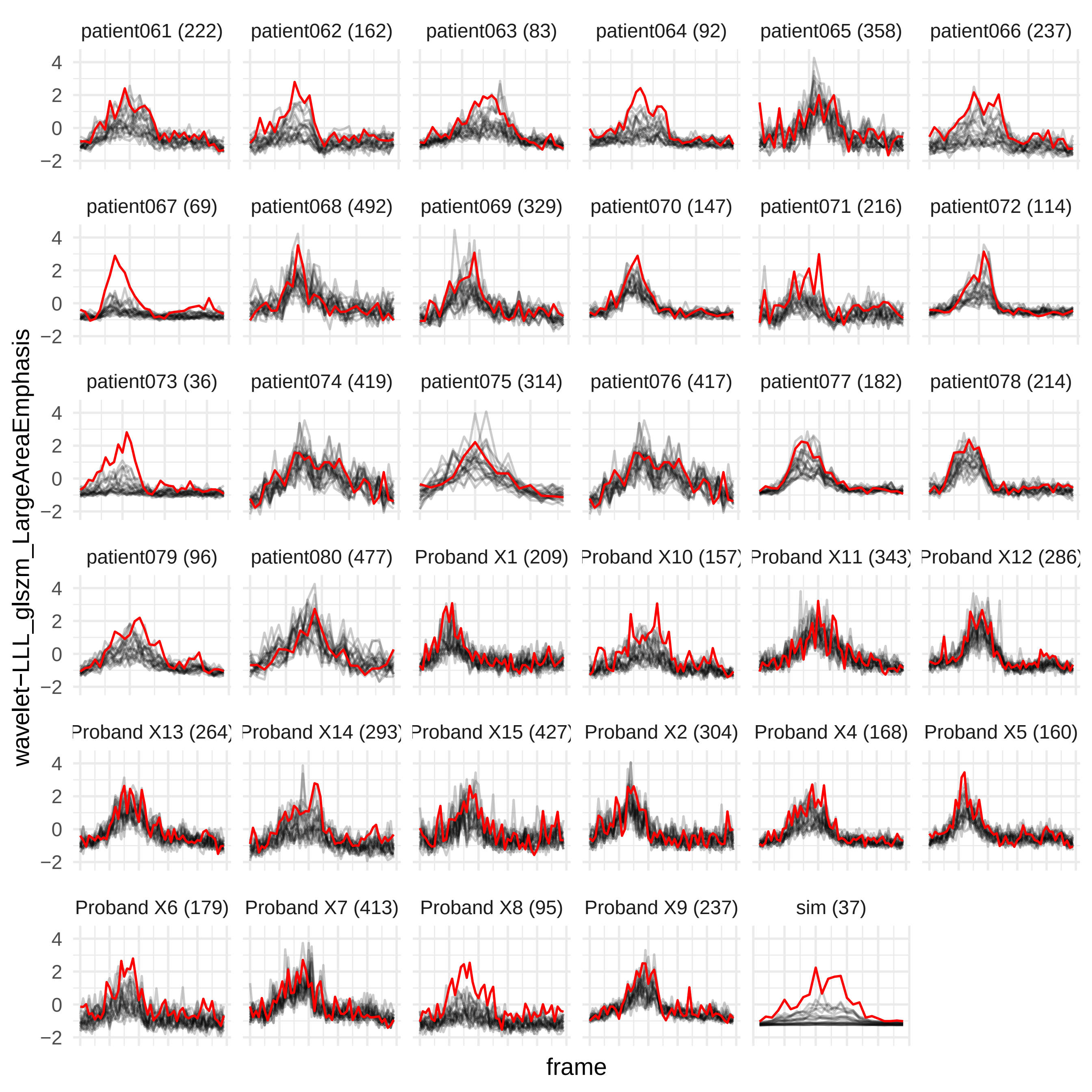

patient061 (222)
patient062 (162)
patient063 (83)
patient064 (92)
patient065 (358)
patient066 (237)
patient067 (69)
patient068 (492)
patient069 (329)
patient070 (147)
patient071 (216)
patient072 (114)
patient073 (36)
patient074 (419)
patient075 (314)
patient076 (417)
patient077 (182)
patient078 (214)
patient079 (96)
patient080 (477)
Proband X1 (209)
Proband X10 (157)
Proband X11 (343)
Proband X12 (286)
Proband X13 (264)
Proband X14 (293)
Proband X15 (427)
Proband X2 (304)
Proband X4 (168)
Proband X5 (160)
Proband X6 (179)
Proband X7 (413)
Proband X8 (95)
Proband X9 (237)
sim (37)
wavelet−LLL_glszm_LargeAreaEmphasis
frame

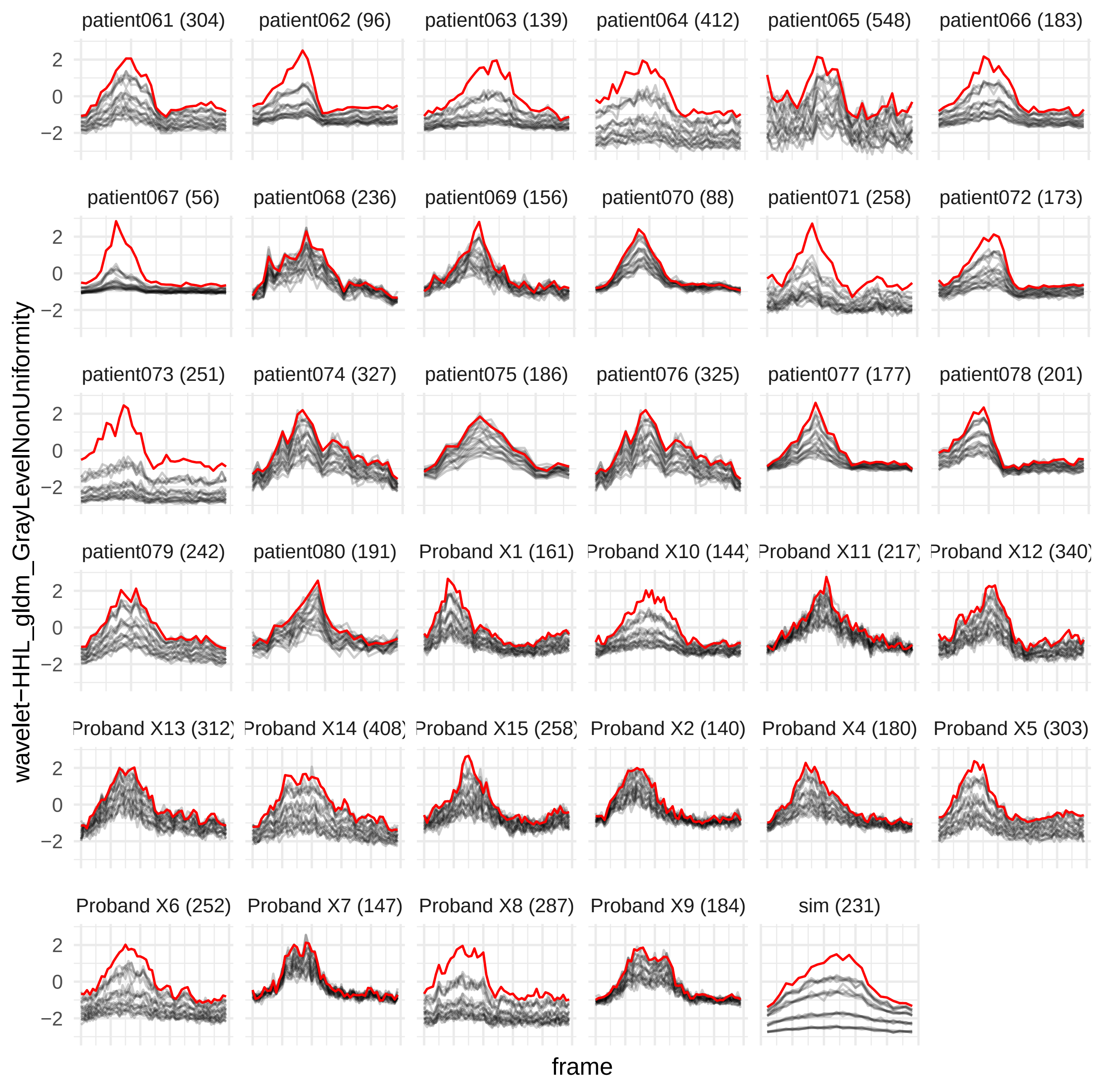

patient061 (304)
patient062 (96)
patient063 (139)
patient064 (412)
patient065 (548)
patient066 (183)
patient067 (56)
patient068 (236)
patient069 (156)
patient070 (88)
patient071 (258)
patient072 (173)
patient073 (251)
patient074 (327)
patient075 (186)
patient076 (325)
patient077 (177)
patient078 (201)
patient079 (242)
patient080 (191)
Proband X1 (161)
Proband X10 (144)
Proband X11 (217)
Proband X12 (340)
Proband X13 (312)
Proband X14 (408)
Proband X15 (258)
Proband X2 (140)
Proband X4 (180)
Proband X5 (303)
Proband X6 (252)
Proband X7 (147)
Proband X8 (287)
Proband X9 (184)
sim (231)
wavelet−HHL_gldm_GrayLevelNonUniformity
frame
2
0
−2

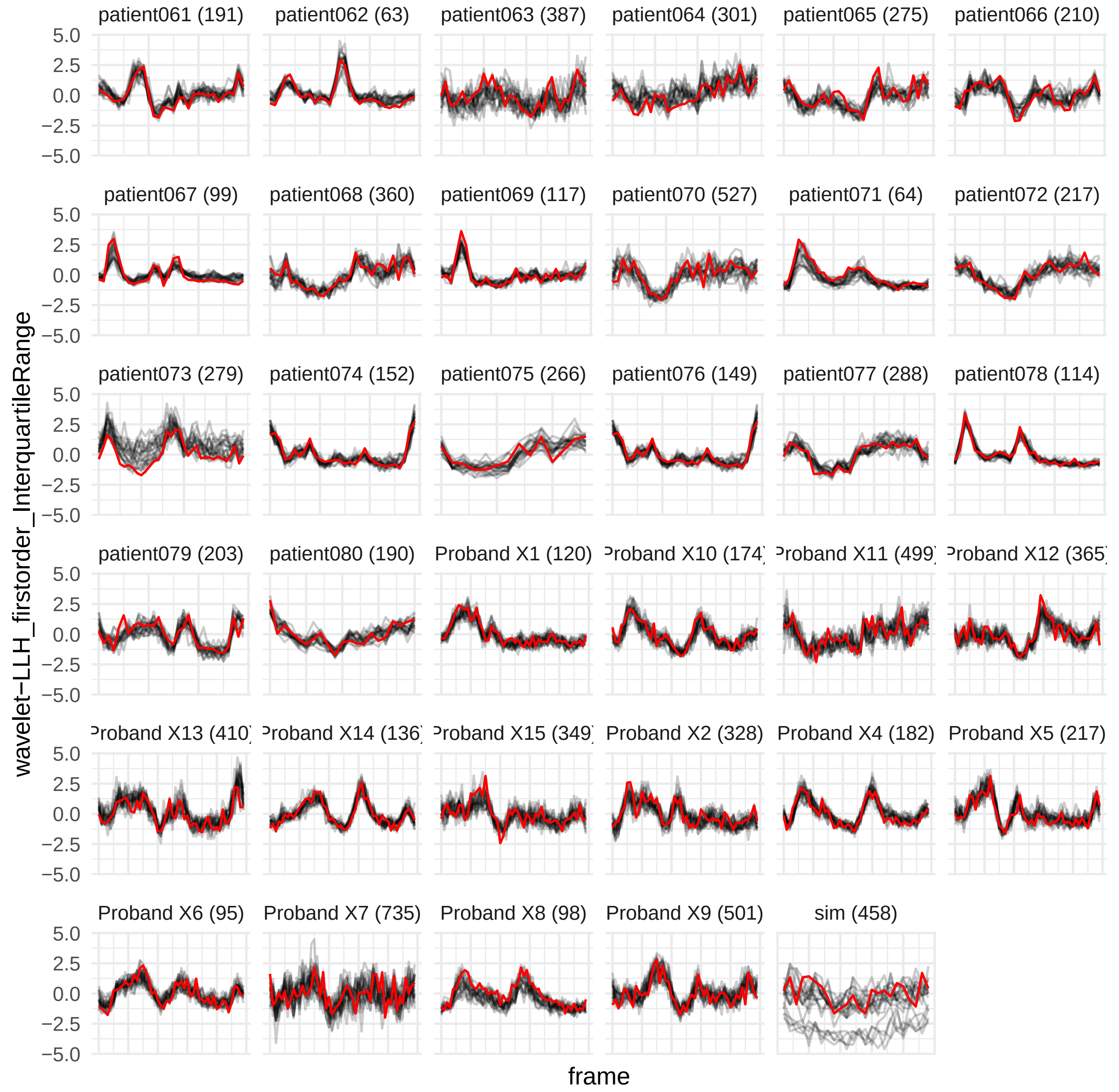

patient061 (191)
patient062 (63)
patient063 (387)
patient064 (301)
patient065 (275)
patient066 (210)
patient067 (99)
patient068 (360)
patient069 (117)
patient070 (527)
patient071 (64)
patient072 (217)
patient073 (279)
patient074 (152)
patient075 (266)
patient076 (149)
patient077 (288)
patient078 (114)
patient079 (203)
patient080 (190)
Proband X1 (120)
Proband X10 (174)
Proband X11 (499)
Proband X12 (365)
Proband X13 (410)
Proband X14 (136)
Proband X15 (349)
Proband X2 (328)
Proband X4 (182)
Proband X5 (217)
Proband X6 (95)
Proband X7 (735)
Proband X8 (98)
Proband X9 (501)
sim (458)
5.0
2.5
0.0
−2.5
−5.0
wavelet−LLH_firstorder_InterquartileRange
frame

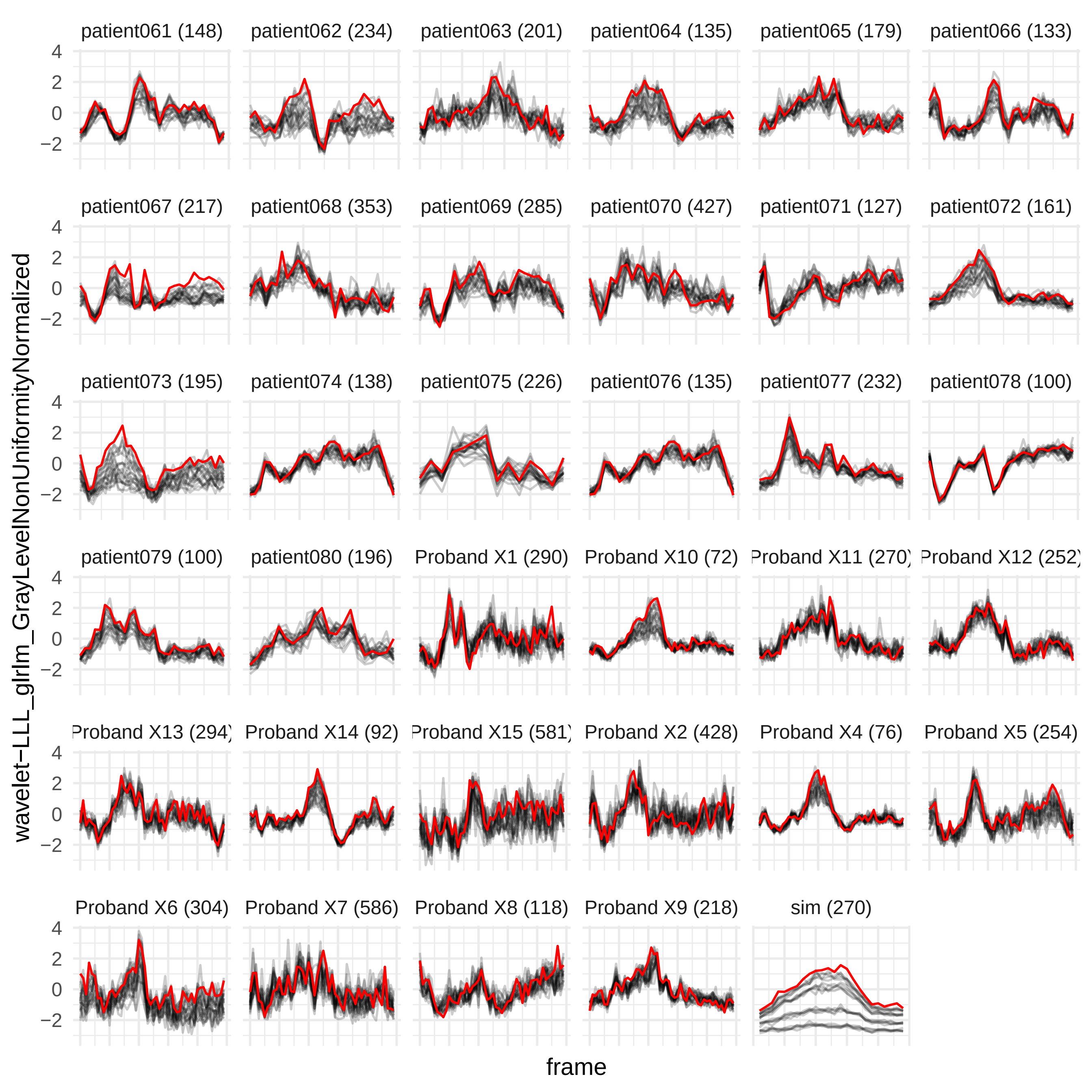

patient061 (148)
patient062 (234)
patient063 (201)
patient064 (135)
patient065 (179)
patient066 (133)
patient067 (217)
patient068 (353)
patient069 (285)
patient070 (427)
patient071 (127)
patient072 (161)
patient073 (195)
patient074 (138)
patient075 (226)
patient076 (135)
patient077 (232)
patient078 (100)
patient079 (100)
patient080 (196)
Proband X1 (290)
Proband X10 (72)
Proband X11 (270)
Proband X12 (252)
Proband X13 (294)
Proband X14 (92)
Proband X15 (581)
Proband X2 (428)
Proband X4 (76)
Proband X5 (254)
Proband X6 (304)
Proband X7 (586)
Proband X8 (118)
Proband X9 (218)
sim (270)
wavelet−LLL_glrlm_GrayLevelNonUniformityNormalized
frame

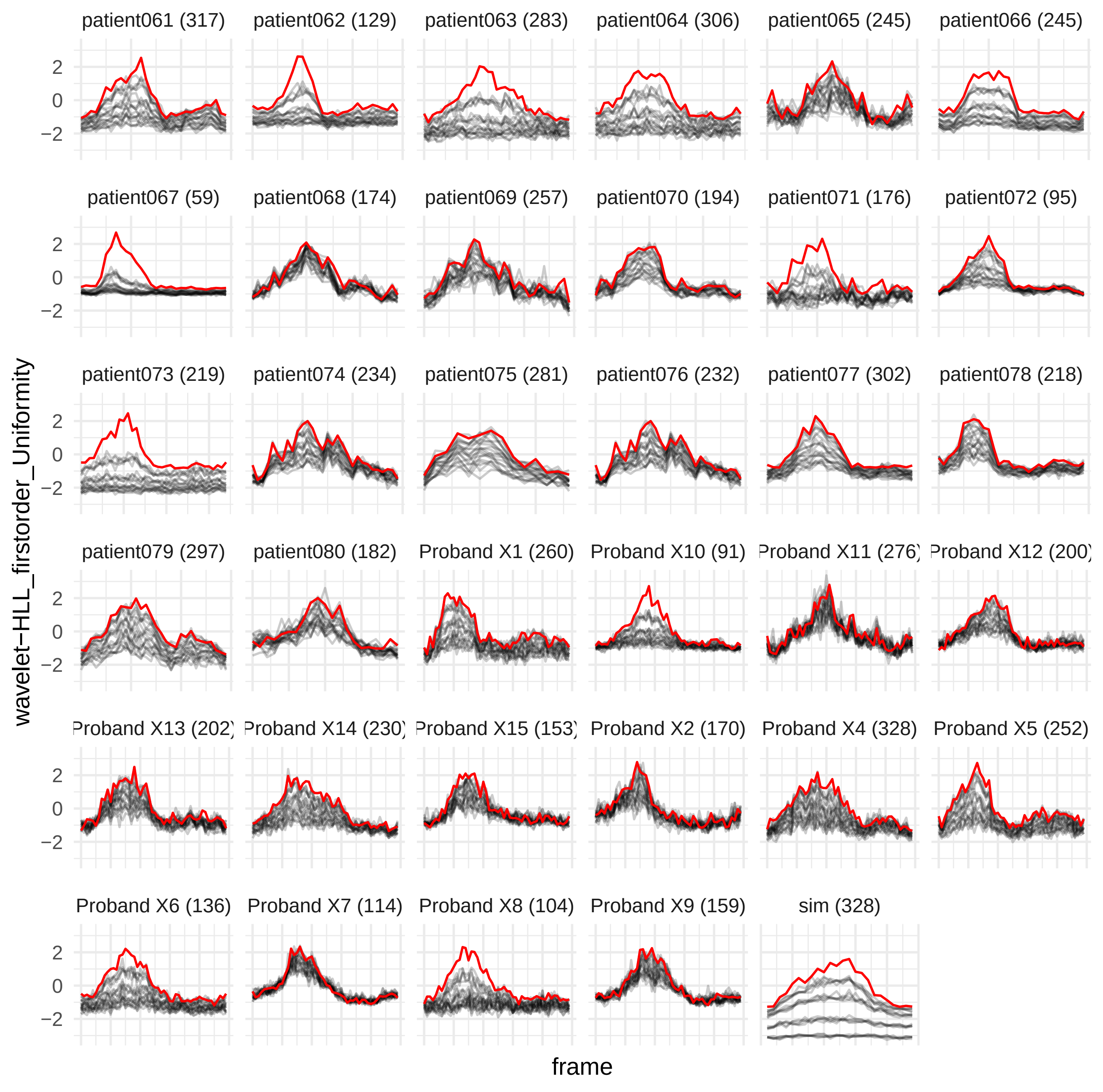

patient061 (317)
patient062 (129)
patient063 (283)
patient064 (306)
patient065 (245)
patient066 (245)
patient067 (59)
patient068 (174)
patient069 (257)
patient070 (194)
patient071 (176)
patient072 (95)
patient073 (219)
patient074 (234)
patient075 (281)
patient076 (232)
patient077 (302)
patient078 (218)
patient079 (297)
patient080 (182)
Proband X1 (260)
Proband X10 (91)
Proband X11 (276)
Proband X12 (200)
Proband X13 (202)
Proband X14 (230)
Proband X15 (153)
Proband X2 (170)
Proband X4 (328)
Proband X5 (252)
Proband X6 (136)
Proband X7 (114)
Proband X8 (104)
Proband X9 (159)
sim (328)
wavelet−HLL_firstorder_Uniformity
frame
2
0
−2

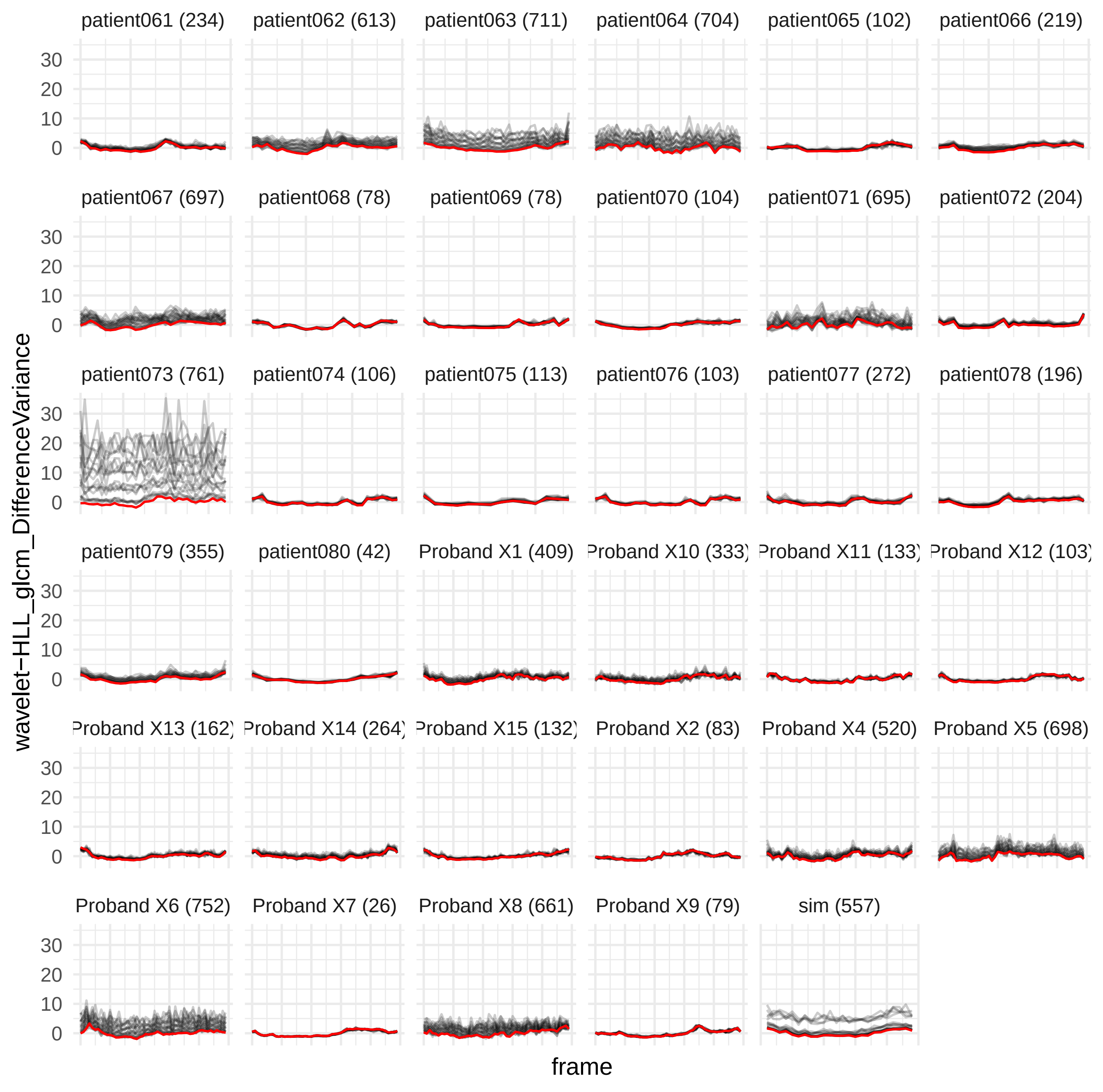
patient061 (234)
patient062 (613)
patient063 (711)
patient064 (704)
patient065 (102)
patient066 (219)
patient067 (697)
patient068 (78)
patient069 (78)
patient070 (104)
patient071 (695)
patient072 (204)
patient073 (761)
patient074 (106)
patient075 (113)
patient076 (103)
patient077 (272)
patient078 (196)
patient079 (355)
patient080 (42)
Proband X1 (409)
Proband X10 (333)
Proband X11 (133)
Proband X12 (103)
Proband X13 (162)
Proband X14 (264)
Proband X15 (132)
Proband X2 (83)
Proband X4 (520)
Proband X5 (698)
Proband X6 (752)
Proband X7 (26)
Proband X8 (661)
Proband X9 (79)
sim (557)
wavelet-HLL_glcm_DifferenceVariance
frame
0
10
20
30

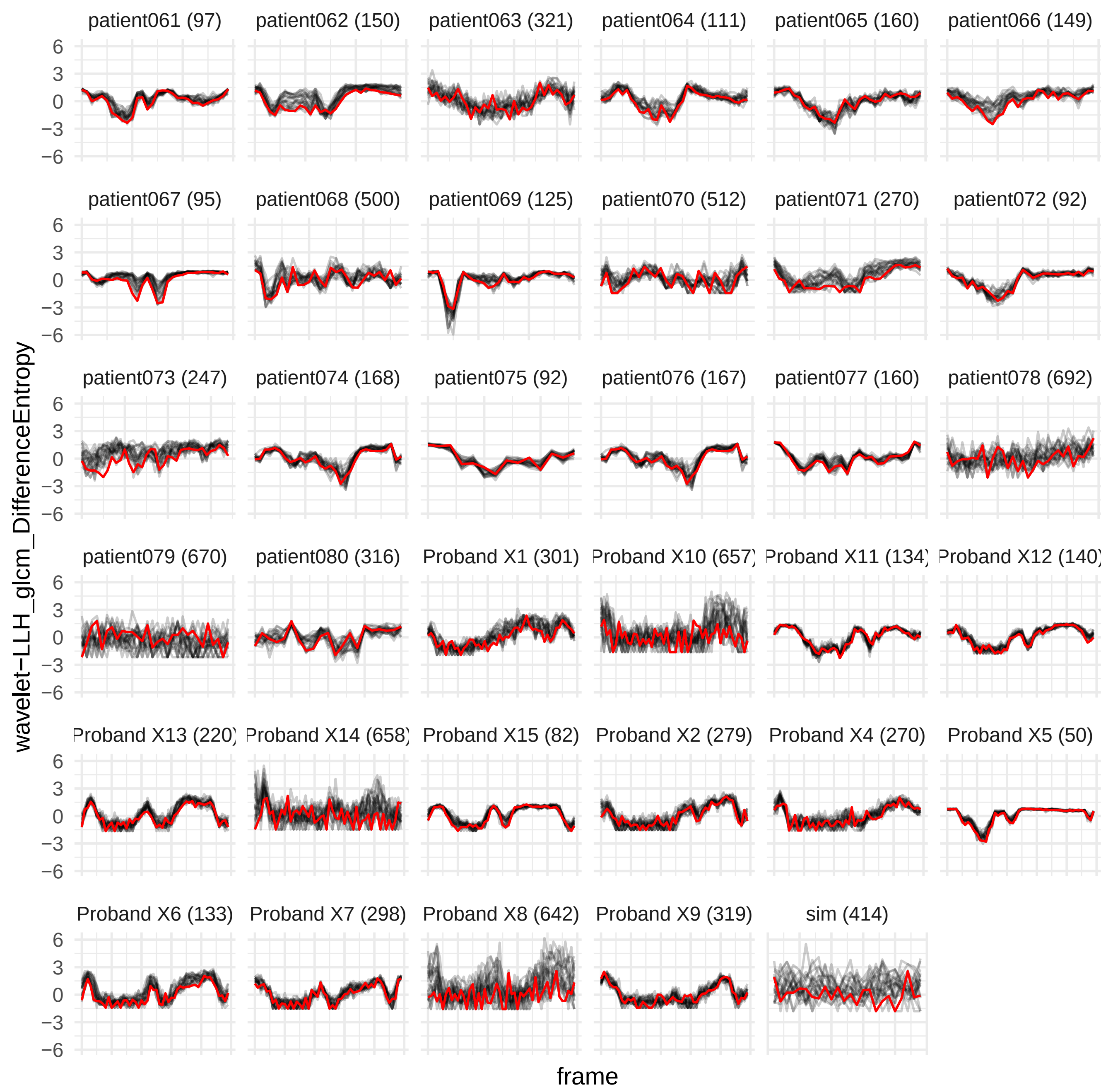

patient061 (97)
patient062 (150)
patient063 (321)
patient064 (111)
patient065 (160)
patient066 (149)
patient067 (95)
patient068 (500)
patient069 (125)
patient070 (512)
patient071 (270)
patient072 (92)
patient073 (247)
patient074 (168)
patient075 (92)
patient076 (167)
patient077 (160)
patient078 (692)
patient079 (670)
patient080 (316)
Proband X1 (301)
Proband X10 (657)
Proband X11 (134)
Proband X12 (140)
Proband X13 (220)
Proband X14 (658)
Proband X15 (82)
Proband X2 (279)
Proband X4 (270)
Proband X5 (50)
Proband X6 (133)
Proband X7 (298)
Proband X8 (642)
Proband X9 (319)
sim (414)
6
3
0
−3
−6
wavelet−LLH_glcm_DifferenceEntropy
frame

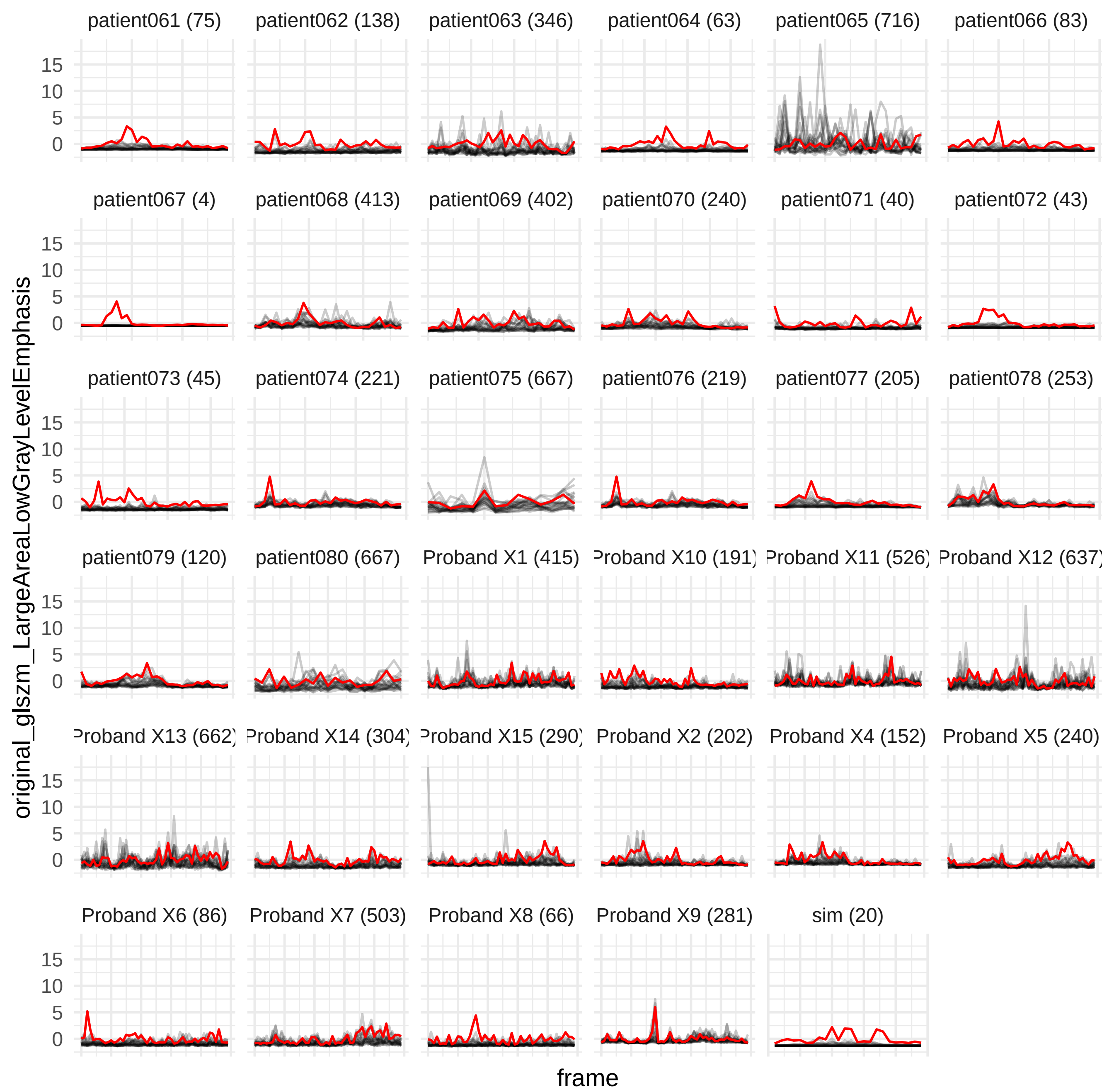

patient061 (75)
patient062 (138)
patient063 (346)
patient064 (63)
patient065 (716)
patient066 (83)
patient067 (4)
patient068 (413)
patient069 (402)
patient070 (240)
patient071 (40)
patient072 (43)
patient073 (45)
patient074 (221)
patient075 (667)
patient076 (219)
patient077 (205)
patient078 (253)
patient079 (120)
patient080 (667)
Proband X1 (415)
Proband X10 (191)
Proband X11 (526)
Proband X12 (637)
Proband X13 (662)
Proband X14 (304)
Proband X15 (290)
Proband X2 (202)
Proband X4 (152)
Proband X5 (240)
Proband X6 (86)
Proband X7 (503)
Proband X8 (66)
Proband X9 (281)
sim (20)
0
5
10
15
original_glszm_LargeAreaLowGrayLevelEmphasis
frame

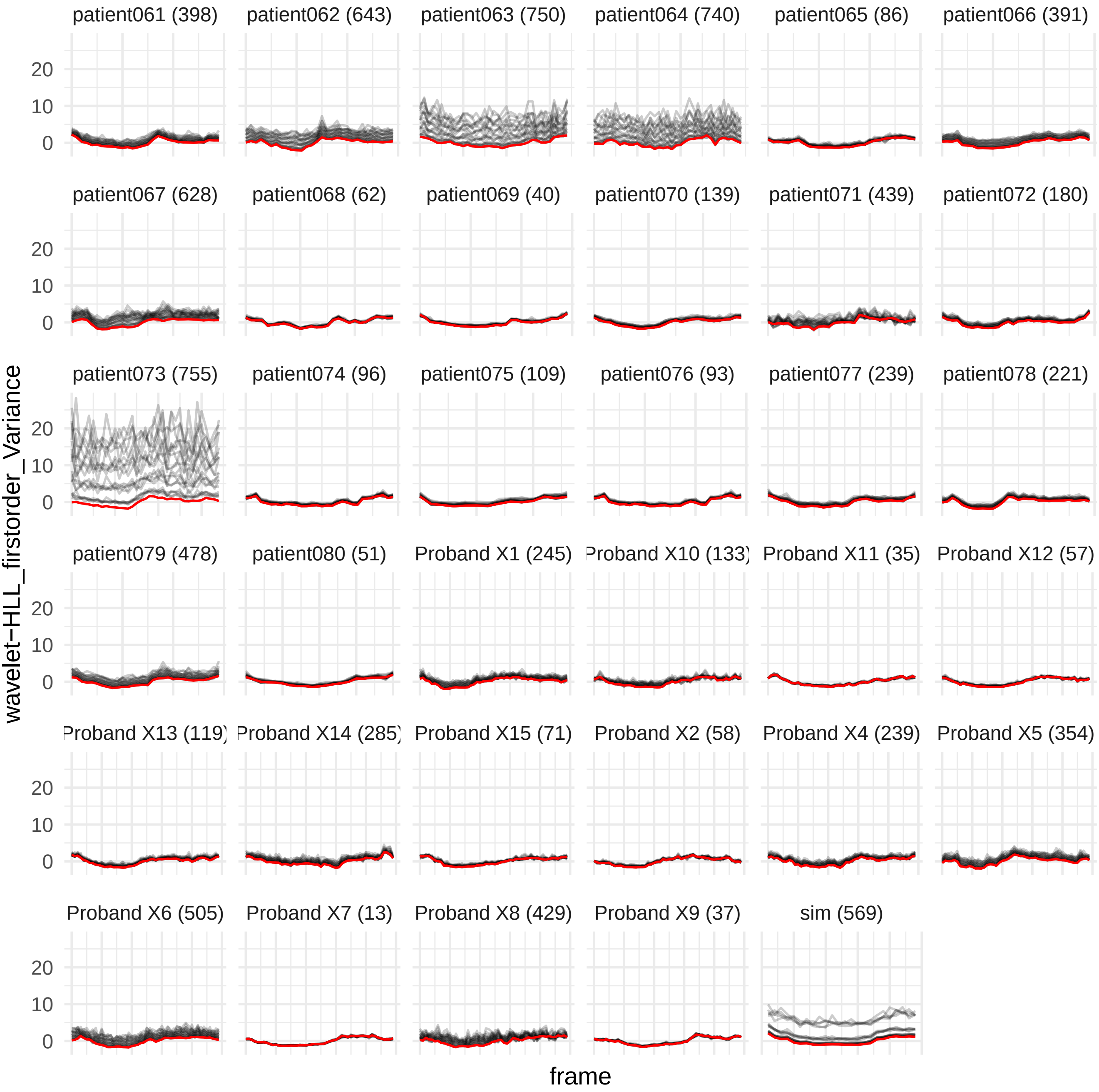

patient061 (398)
patient062 (643)
patient063 (750)
patient064 (740)
patient065 (86)
patient066 (391)
patient067 (628)
patient068 (62)
patient069 (40)
patient070 (139)
patient071 (439)
patient072 (180)
patient073 (755)
patient074 (96)
patient075 (109)
patient076 (93)
patient077 (239)
patient078 (221)
patient079 (478)
patient080 (51)
Proband X1 (245)
Proband X10 (133)
Proband X11 (35)
Proband X12 (57)
Proband X13 (119)
Proband X14 (285)
Proband X15 (71)
Proband X2 (58)
Proband X4 (239)
Proband X5 (354)
Proband X6 (505)
Proband X7 (13)
Proband X8 (429)
Proband X9 (37)
sim (569)
0
10
20
wavelet−HLL_firstorder_Variance
frame

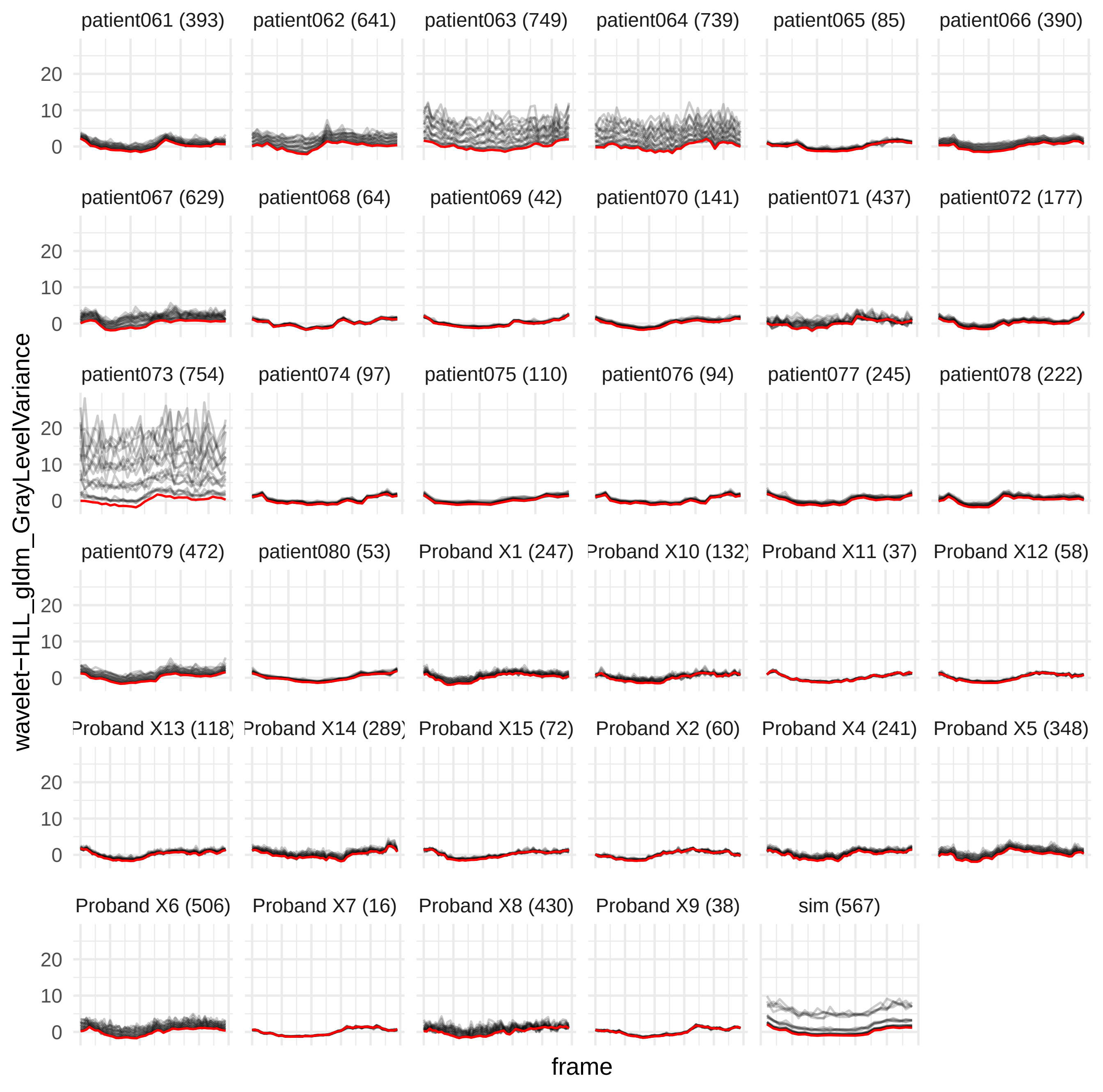
patient061 (393)
patient062 (641)
patient063 (749)
patient064 (739)
patient065 (85)
patient066 (390)
patient067 (629)
patient068 (64)
patient069 (42)
patient070 (141)
patient071 (437)
patient072 (177)
patient073 (754)
patient074 (97)
patient075 (110)
patient076 (94)
patient077 (245)
patient078 (222)
patient079 (472)
patient080 (53)
Proband X1 (247)
Proband X10 (132)
Proband X11 (37)
Proband X12 (58)
Proband X13 (118)
Proband X14 (289)
Proband X15 (72)
Proband X2 (60)
Proband X4 (241)
Proband X5 (348)
Proband X6 (506)
Proband X7 (16)
Proband X8 (430)
Proband X9 (38)
sim (567)
wavelet−HLL_gldm_GrayLevelVariance
frame
0
10
20

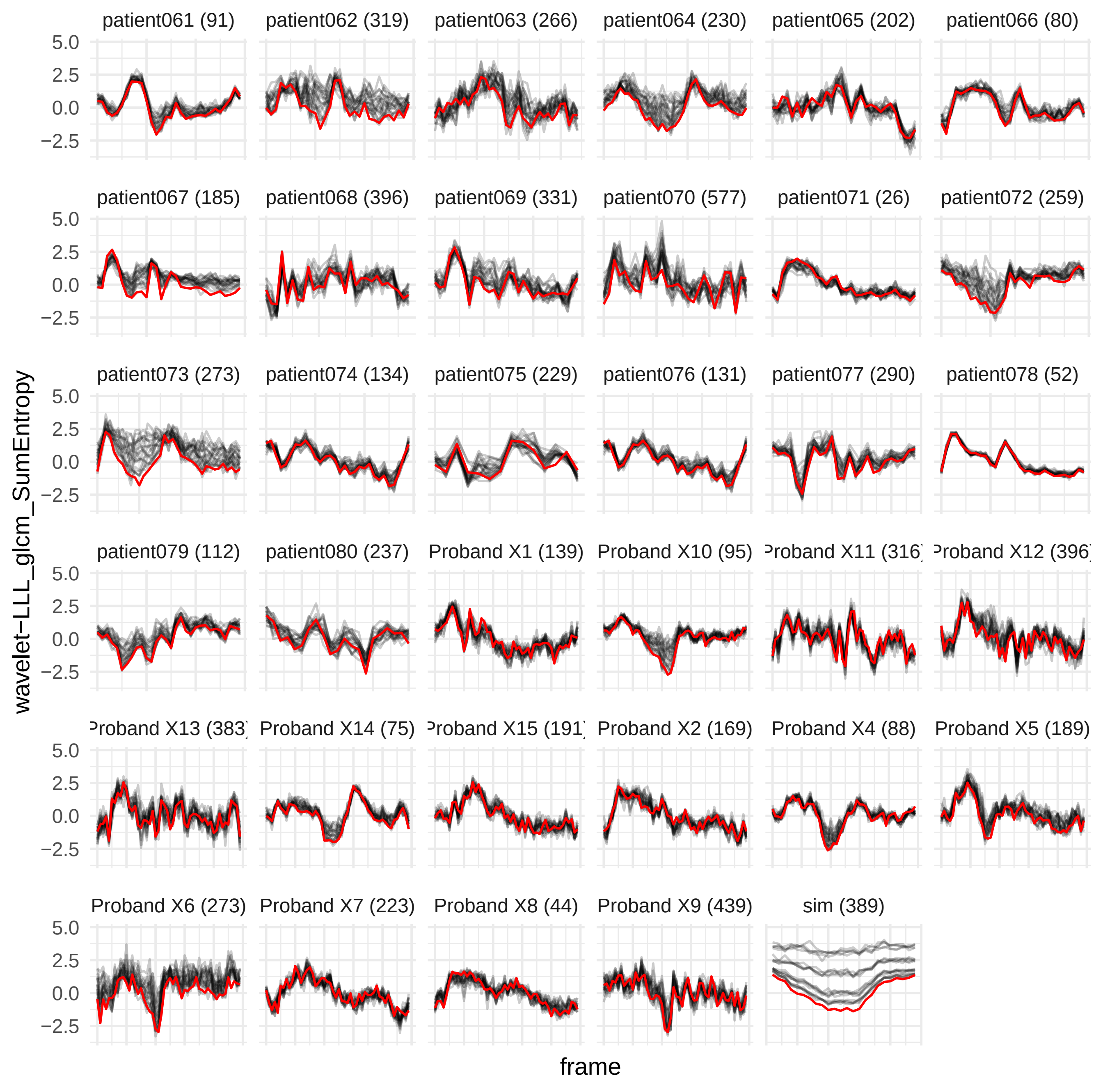

patient061 (91)
patient062 (319)
patient063 (266)
patient064 (230)
patient065 (202)
patient066 (80)
patient067 (185)
patient068 (396)
patient069 (331)
patient070 (577)
patient071 (26)
patient072 (259)
patient073 (273)
patient074 (134)
patient075 (229)
patient076 (131)
patient077 (290)
patient078 (52)
patient079 (112)
patient080 (237)
Proband X1 (139)
Proband X10 (95)
Proband X11 (316)
Proband X12 (396)
Proband X13 (383)
Proband X14 (75)
Proband X15 (191)
Proband X2 (169)
Proband X4 (88)
Proband X5 (189)
Proband X6 (273)
Proband X7 (223)
Proband X8 (44)
Proband X9 (439)
sim (389)
5.0
2.5
0.0
−2.5
wavelet−LLL_glcm_SumEntropy
frame

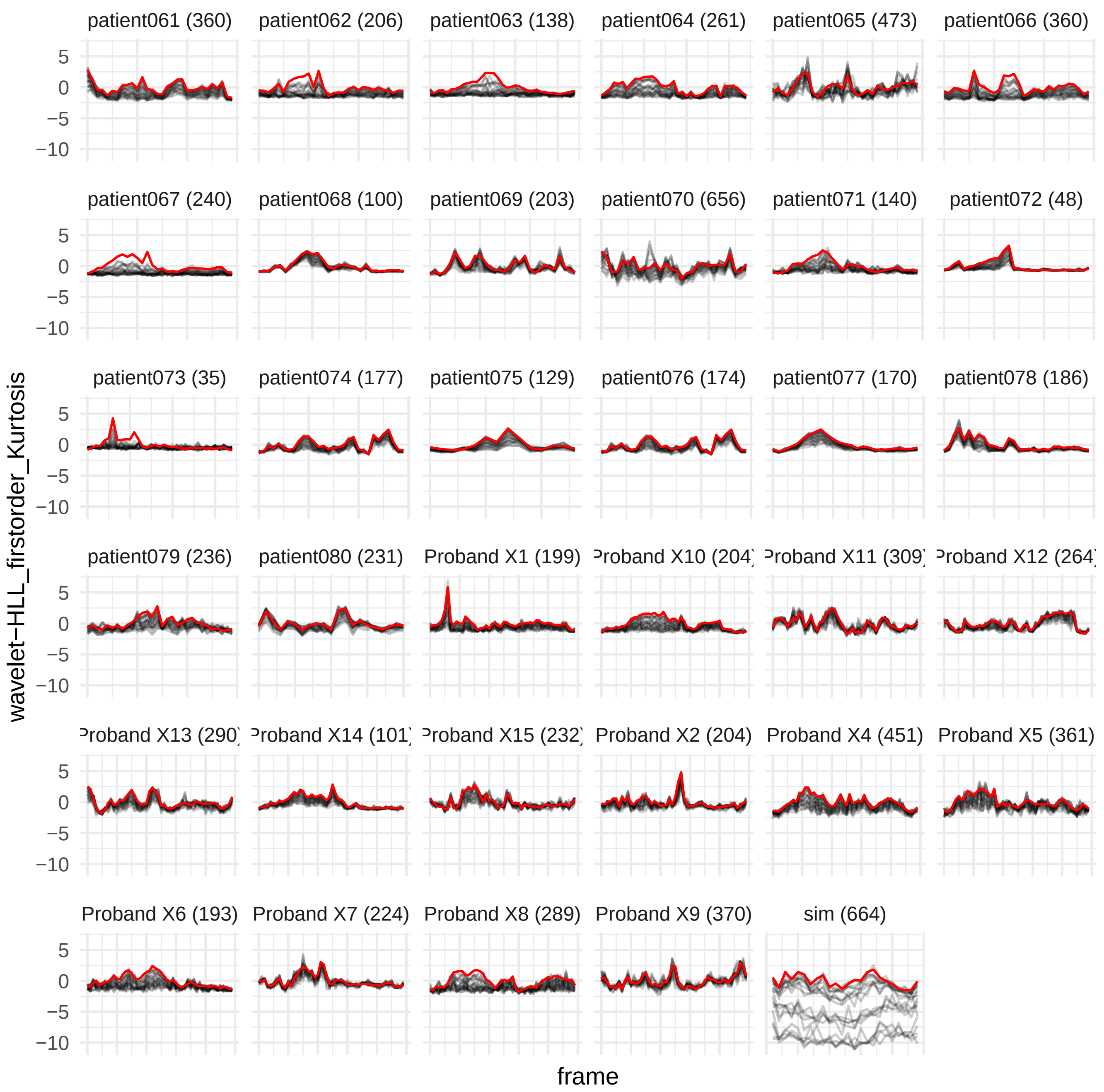

patient061 (360)
patient062 (206)
patient063 (138)
patient064 (261)
patient065 (473)
patient066 (360)
patient067 (240)
patient068 (100)
patient069 (203)
patient070 (656)
patient071 (140)
patient072 (48)
patient073 (35)
patient074 (177)
patient075 (129)
patient076 (174)
patient077 (170)
patient078 (186)
patient079 (236)
patient080 (231)
Proband X1 (199)
Proband X10 (204)
Proband X11 (309)
Proband X12 (264)
Proband X13 (290)
Proband X14 (101)
Proband X15 (232)
Proband X2 (204)
Proband X4 (451)
Proband X5 (361)
Proband X6 (193)
Proband X7 (224)
Proband X8 (289)
Proband X9 (370)
sim (664)
5
0
−5
−10
wavelet−HLL_firstorder_Kurtosis
frame

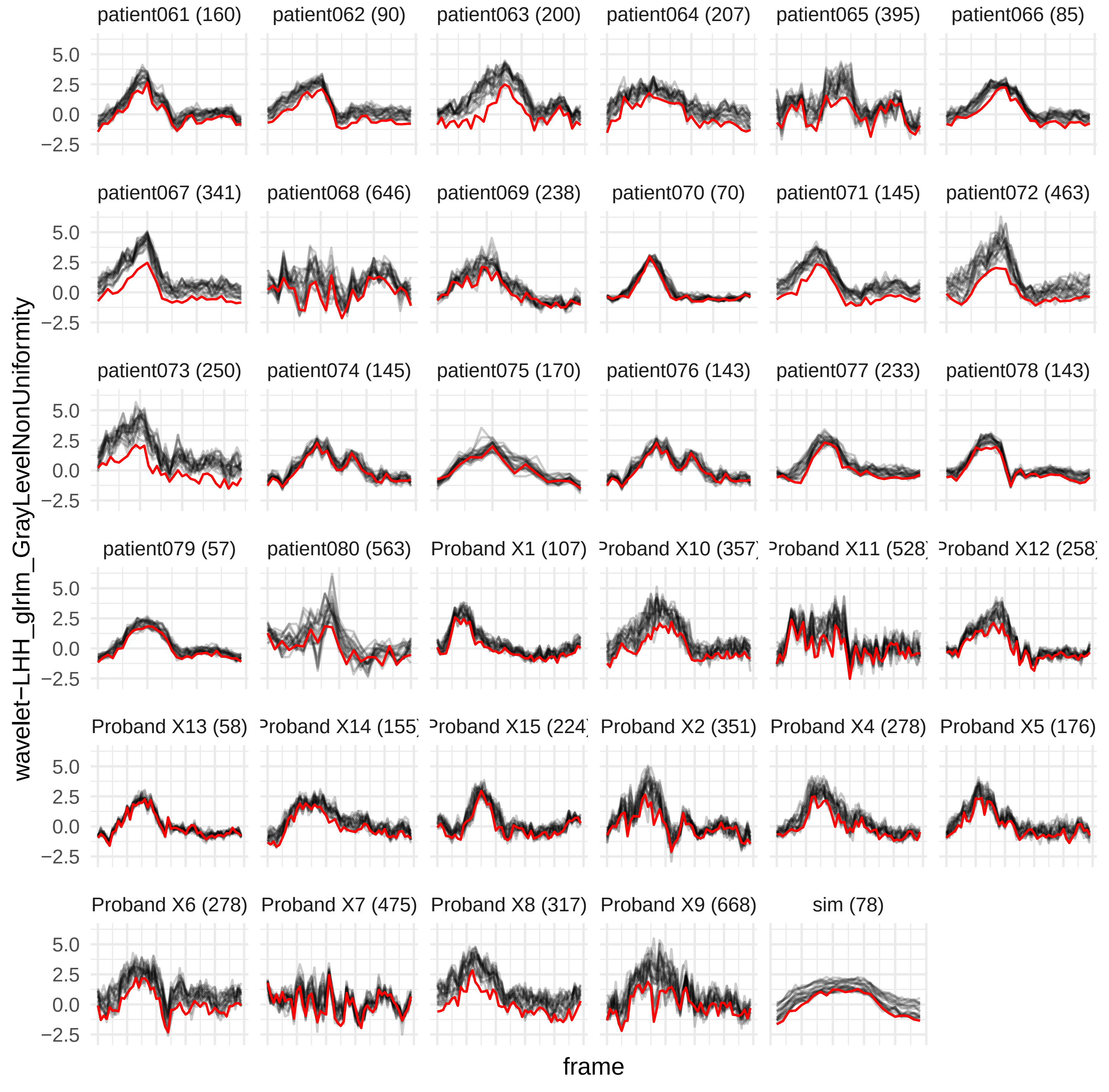

patient061 (160)
patient062 (90)
patient063 (200)
patient064 (207)
patient065 (395)
patient066 (85)
patient067 (341)
patient068 (646)
patient069 (238)
patient070 (70)
patient071 (145)
patient072 (463)
patient073 (250)
patient074 (145)
patient075 (170)
patient076 (143)
patient077 (233)
patient078 (143)
patient079 (57)
patient080 (563)
Proband X1 (107)
Proband X10 (357)
Proband X11 (528)
Proband X12 (258)
Proband X13 (58)
Proband X14 (155)
Proband X15 (224)
Proband X2 (351)
Proband X4 (278)
Proband X5 (176)
Proband X6 (278)
Proband X7 (475)
Proband X8 (317)
Proband X9 (668)
sim (78)
5.0
2.5
0.0
−2.5
wavelet−LHH_glrlm_GrayLevelNonUniformity
frame

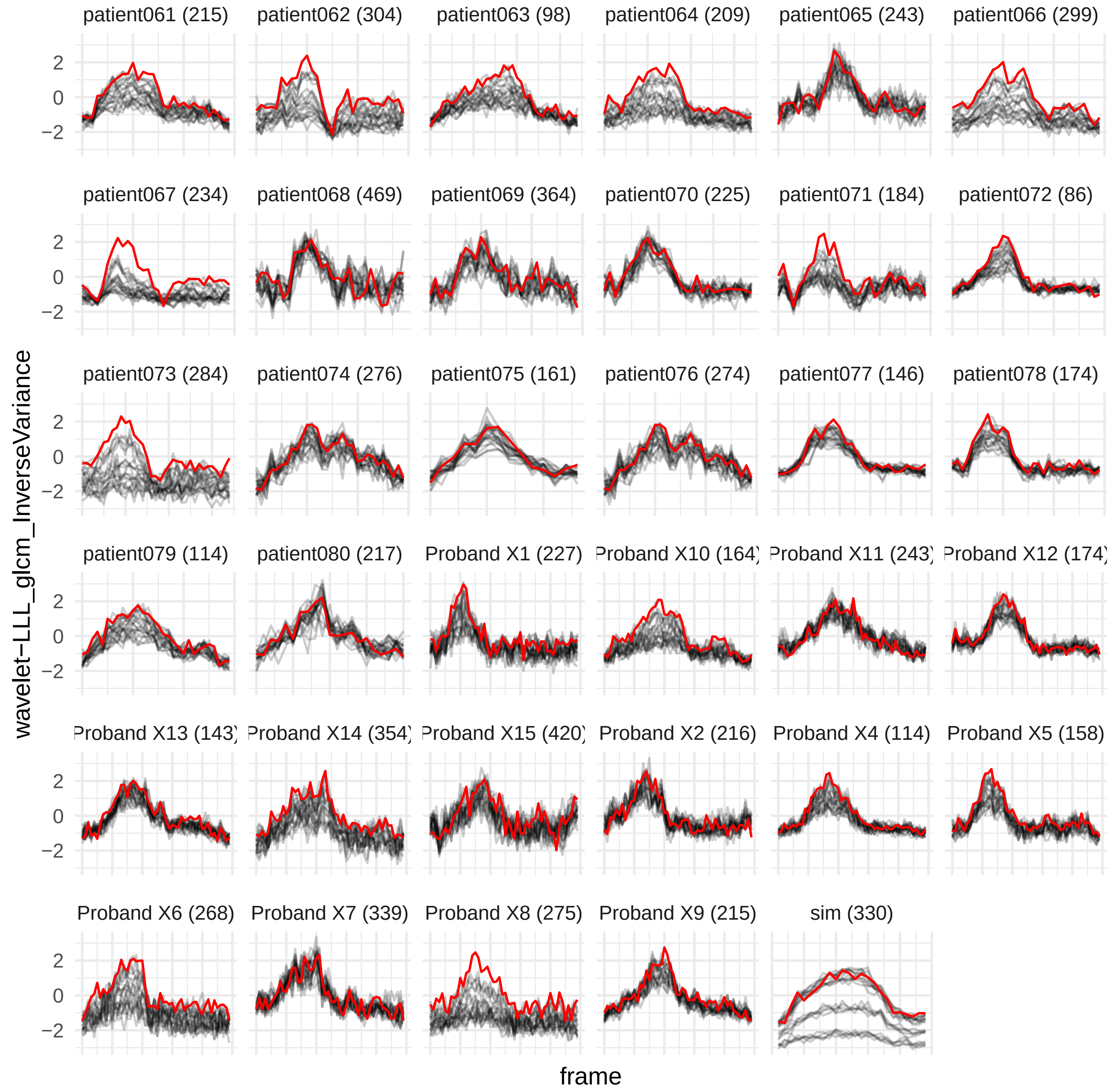

patient061 (215)
patient062 (304)
patient063 (98)
patient064 (209)
patient065 (243)
patient066 (299)
patient067 (234)
patient068 (469)
patient069 (364)
patient070 (225)
patient071 (184)
patient072 (86)
patient073 (284)
patient074 (276)
patient075 (161)
patient076 (274)
patient077 (146)
patient078 (174)
patient079 (114)
patient080 (217)
Proband X1 (227)
Proband X10 (164)
Proband X11 (243)
Proband X12 (174)
Proband X13 (143)
Proband X14 (354)
Proband X15 (420)
Proband X2 (216)
Proband X4 (114)
Proband X5 (158)
Proband X6 (268)
Proband X7 (339)
Proband X8 (275)
Proband X9 (215)
sim (330)
wavelet−LLL_glcm_InverseVariance
frame
2
0
−2

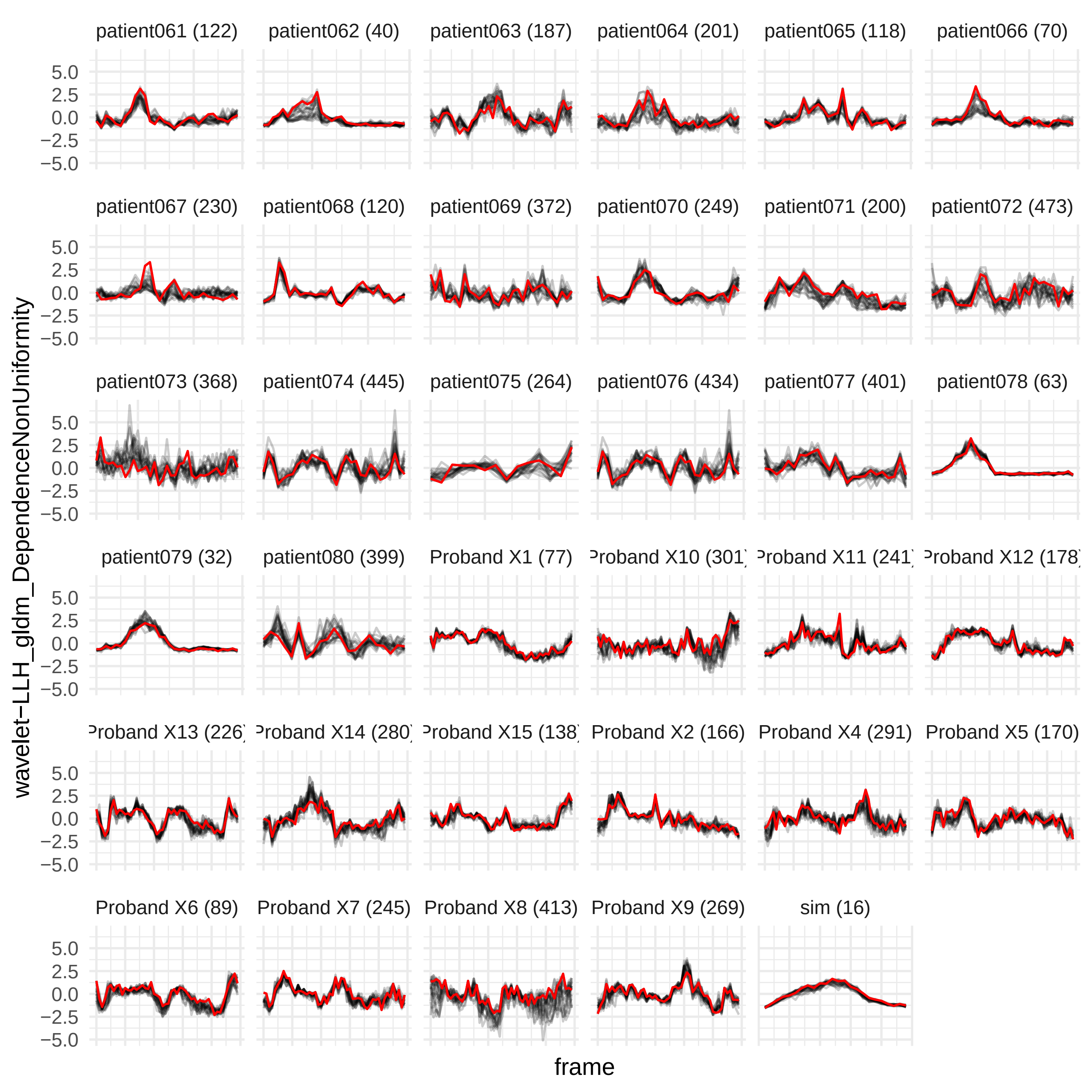

patient061 (122)
patient062 (40)
patient063 (187)
patient064 (201)
patient065 (118)
patient066 (70)
patient067 (230)
patient068 (120)
patient069 (372)
patient070 (249)
patient071 (200)
patient072 (473)
patient073 (368)
patient074 (445)
patient075 (264)
patient076 (434)
patient077 (401)
patient078 (63)
patient079 (32)
patient080 (399)
Proband X1 (77)
Proband X10 (301)
Proband X11 (241)
Proband X12 (178)
Proband X13 (226)
Proband X14 (280)
Proband X15 (138)
Proband X2 (166)
Proband X4 (291)
Proband X5 (170)
Proband X6 (89)
Proband X7 (245)
Proband X8 (413)
Proband X9 (269)
sim (16)
5.0
2.5
0.0
−2.5
−5.0
wavelet−LLH_gldm_DependenceNonUniformity
frame

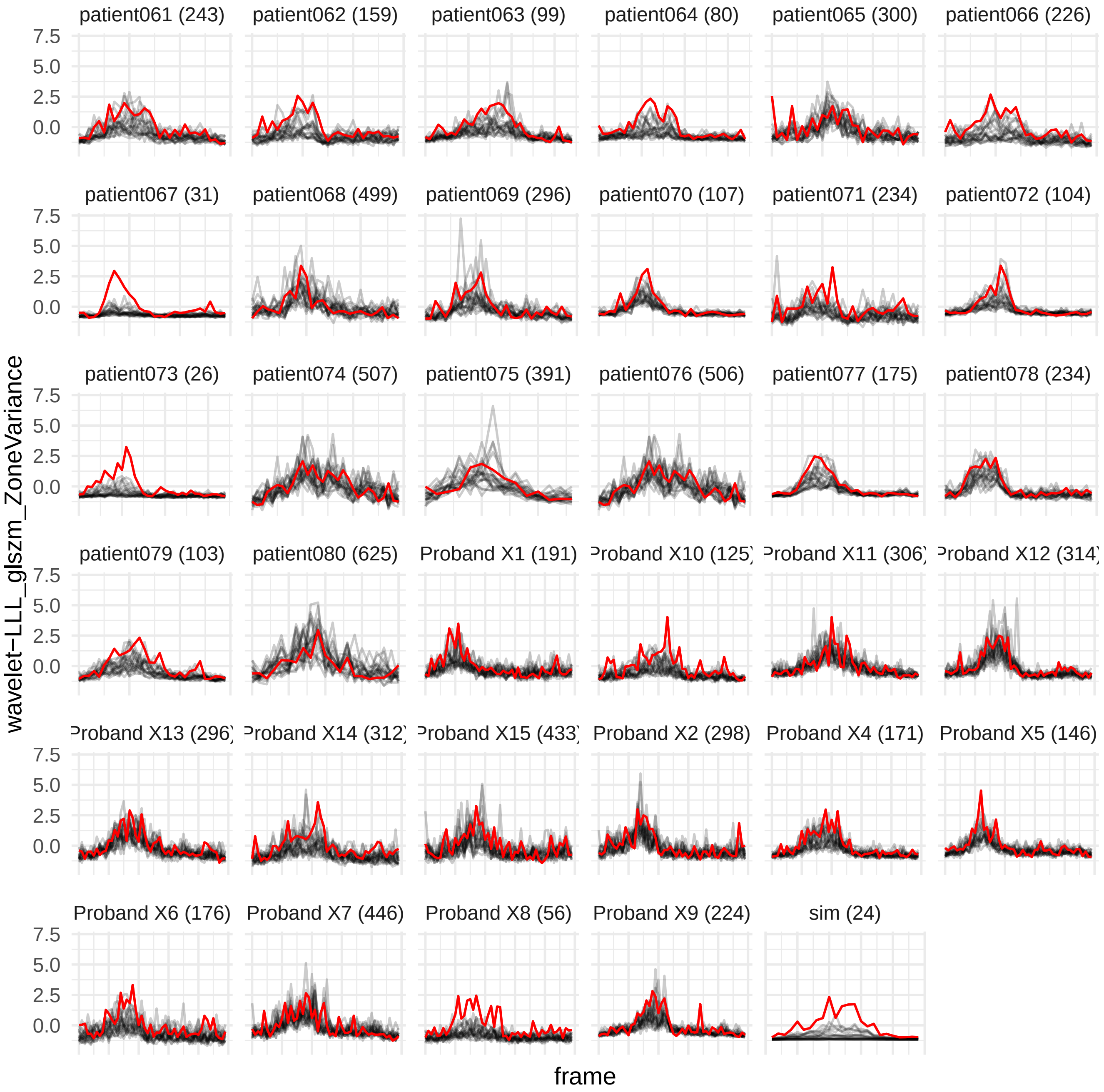

patient061 (243)
patient062 (159)
patient063 (99)
patient064 (80)
patient065 (300)
patient066 (226)
patient067 (31)
patient068 (499)
patient069 (296)
patient070 (107)
patient071 (234)
patient072 (104)
patient073 (26)
patient074 (507)
patient075 (391)
patient076 (506)
patient077 (175)
patient078 (234)
patient079 (103)
patient080 (625)
Proband X1 (191)
Proband X10 (125)
Proband X11 (306)
Proband X12 (314)
Proband X13 (296)
Proband X14 (312)
Proband X15 (433)
Proband X2 (298)
Proband X4 (171)
Proband X5 (146)
Proband X6 (176)
Proband X7 (446)
Proband X8 (56)
Proband X9 (224)
sim (24)
7.5
5.0
2.5
0.0
wavelet−LLL_glszm_ZoneVariance
frame

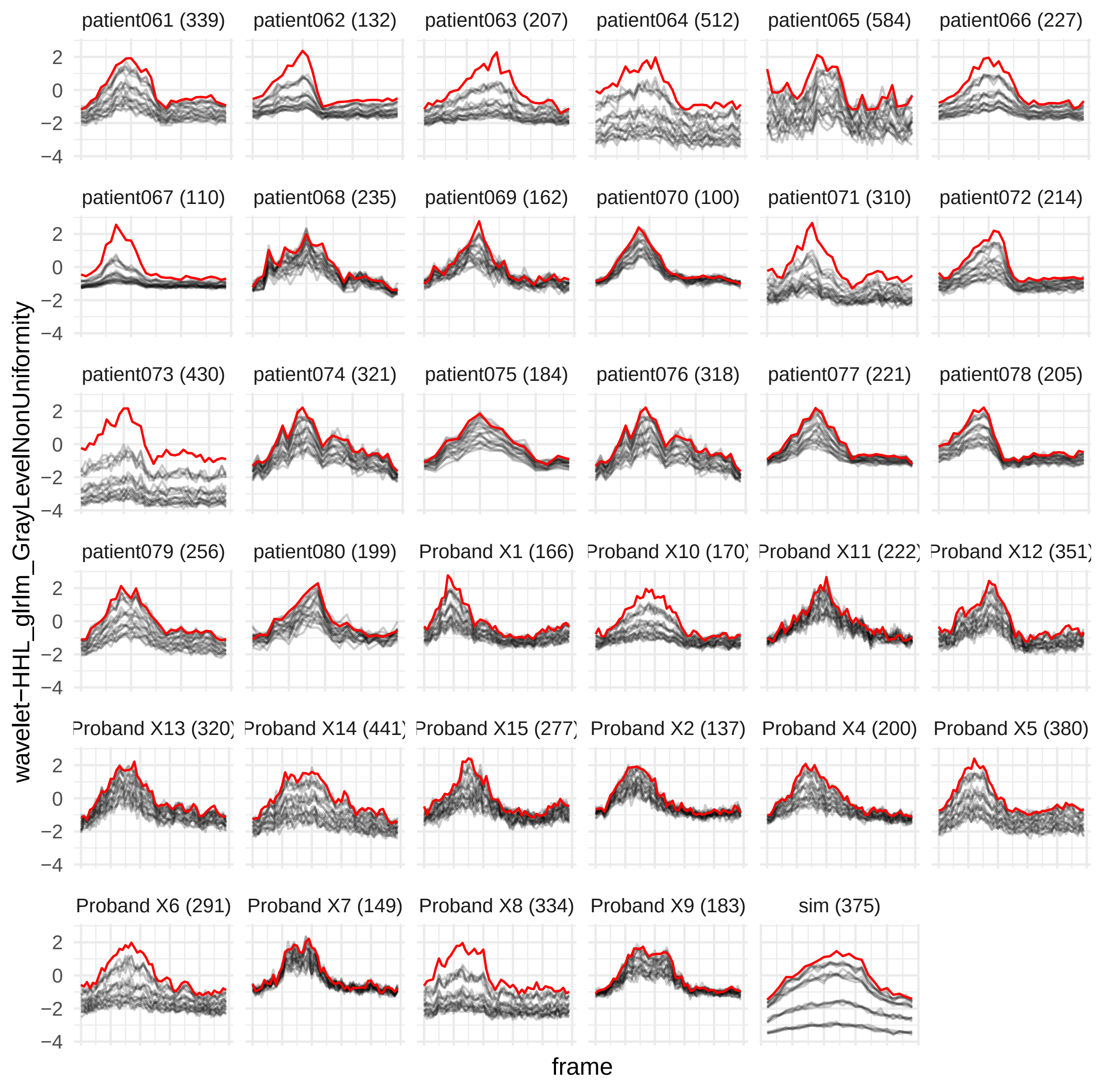

patient061 (339)
patient062 (132)
patient063 (207)
patient064 (512)
patient065 (584)
patient066 (227)
patient067 (110)
patient068 (235)
patient069 (162)
patient070 (100)
patient071 (310)
patient072 (214)
patient073 (430)
patient074 (321)
patient075 (184)
patient076 (318)
patient077 (221)
patient078 (205)
patient079 (256)
patient080 (199)
Proband X1 (166)
Proband X10 (170)
Proband X11 (222)
Proband X12 (351)
Proband X13 (320)
Proband X14 (441)
Proband X15 (277)
Proband X2 (137)
Proband X4 (200)
Proband X5 (380)
Proband X6 (291)
Proband X7 (149)
Proband X8 (334)
Proband X9 (183)
sim (375)
2
0
−2
−4
wavelet−HHL_glrlm_GrayLevelNonUniformity
frame

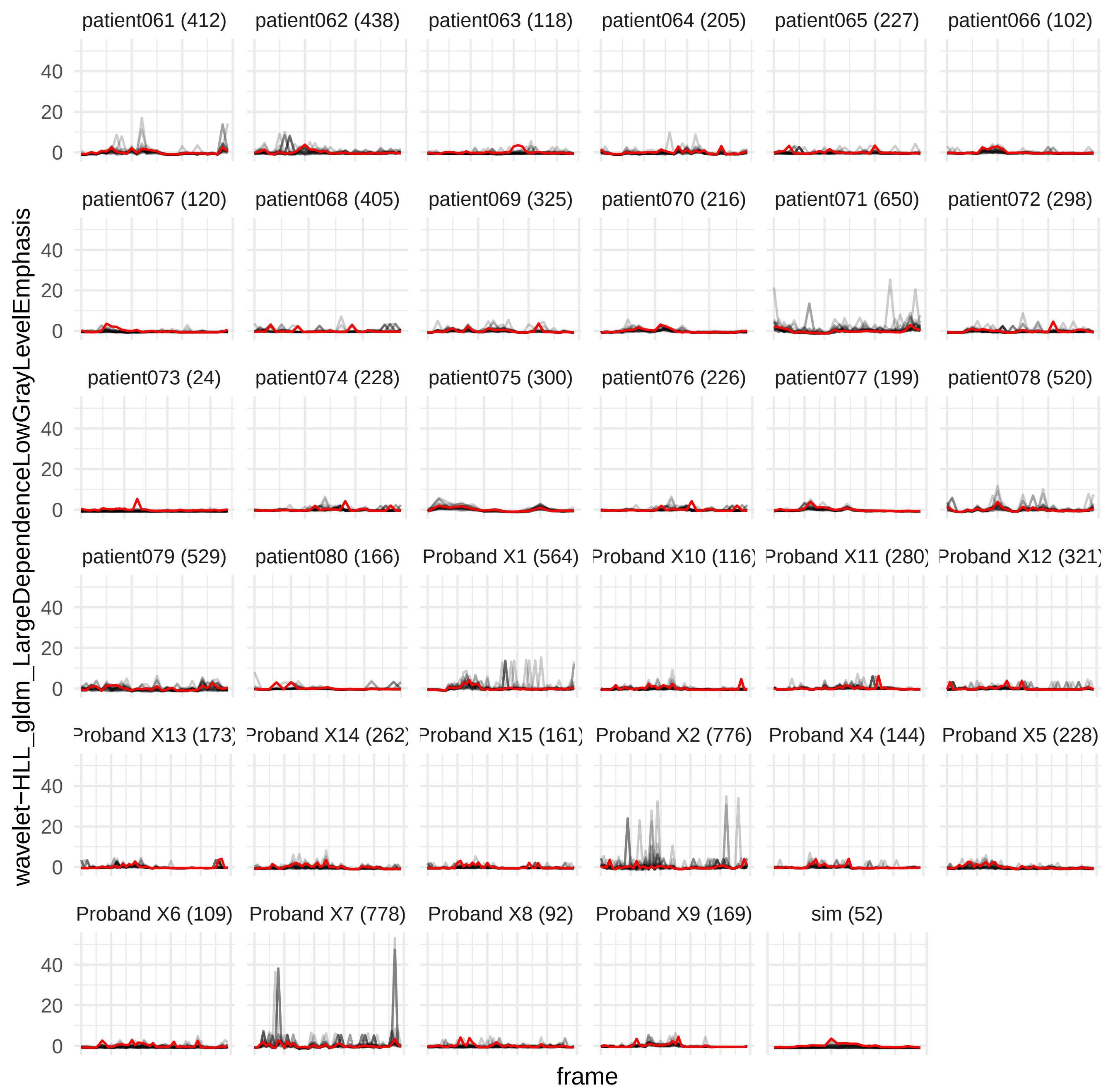
patient061 (412)
patient062 (438)
patient063 (118)
patient064 (205)
patient065 (227)
patient066 (102)
patient067 (120)
patient068 (405)
patient069 (325)
patient070 (216)
patient071 (650)
patient072 (298)
patient073 (24)
patient074 (228)
patient075 (300)
patient076 (226)
patient077 (199)
patient078 (520)
patient079 (529)
patient080 (166)
Proband X1 (564)
Proband X10 (116)
Proband X11 (280)
Proband X12 (321)
Proband X13 (173)
Proband X14 (262)
Proband X15 (161)
Proband X2 (776)
Proband X4 (144)
Proband X5 (228)
Proband X6 (109)
Proband X7 (778)
Proband X8 (92)
Proband X9 (169)
sim (52)
40
20
0
wavelet−HLL_gldm_LargeDependenceLowGrayLevelEmphasis
frame

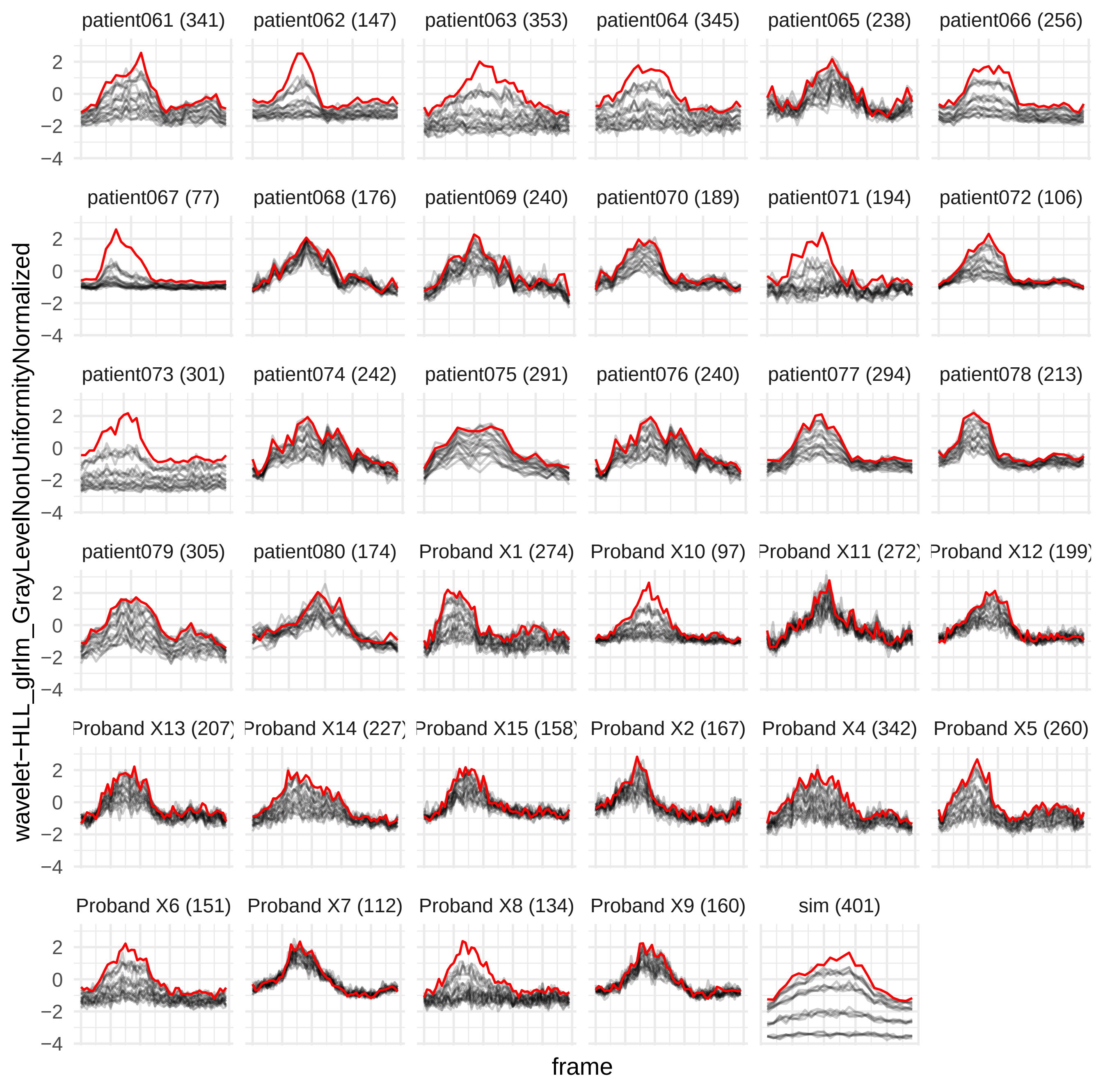

patient061 (341)
patient062 (147)
patient063 (353)
patient064 (345)
patient065 (238)
patient066 (256)
patient067 (77)
patient068 (176)
patient069 (240)
patient070 (189)
patient071 (194)
patient072 (106)
patient073 (301)
patient074 (242)
patient075 (291)
patient076 (240)
patient077 (294)
patient078 (213)
patient079 (305)
patient080 (174)
Proband X1 (274)
Proband X10 (97)
Proband X11 (272)
Proband X12 (199)
Proband X13 (207)
Proband X14 (227)
Proband X15 (158)
Proband X2 (167)
Proband X4 (342)
Proband X5 (260)
Proband X6 (151)
Proband X7 (112)
Proband X8 (134)
Proband X9 (160)
sim (401)
wavelet−HLL_glrlm_GrayLevelNonUniformityNormalized
frame
2
0
−2
−4

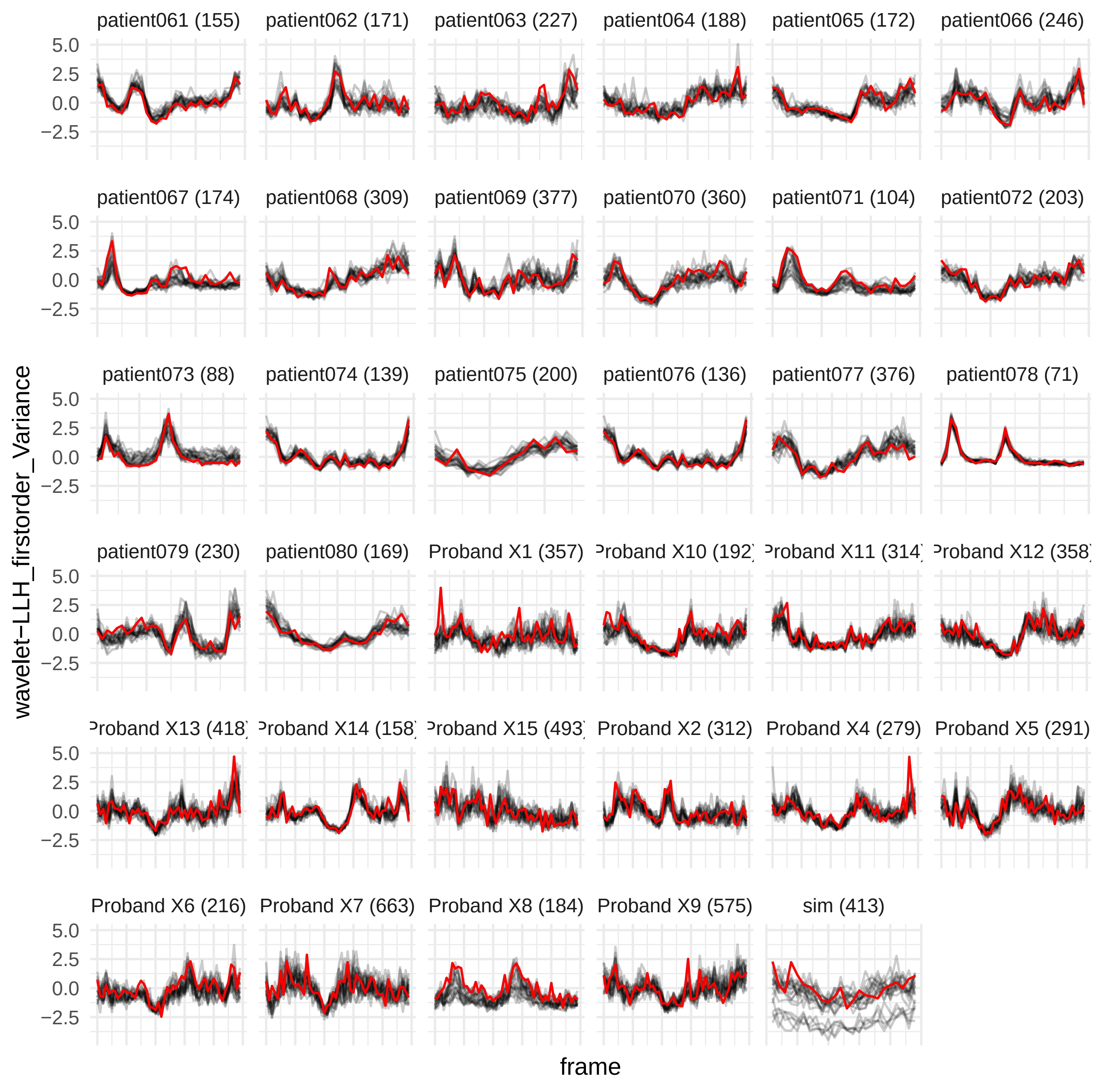
patient061 (155)
patient062 (171)
patient063 (227)
patient064 (188)
patient065 (172)
patient066 (246)
patient067 (174)
patient068 (309)
patient069 (377)
patient070 (360)
patient071 (104)
patient072 (203)
patient073 (88)
patient074 (139)
patient075 (200)
patient076 (136)
patient077 (376)
patient078 (71)
patient079 (230)
patient080 (169)
Proband X1 (357)
Proband X10 (192)
Proband X11 (314)
Proband X12 (358)
Proband X13 (418)
Proband X14 (158)
Proband X15 (493)
Proband X2 (312)
Proband X4 (279)
Proband X5 (291)
Proband X6 (216)
Proband X7 (663)
Proband X8 (184)
Proband X9 (575)
sim (413)
5.0
2.5
0.0
−2.5
wavelet−LLH_firstorder_Variance
frame

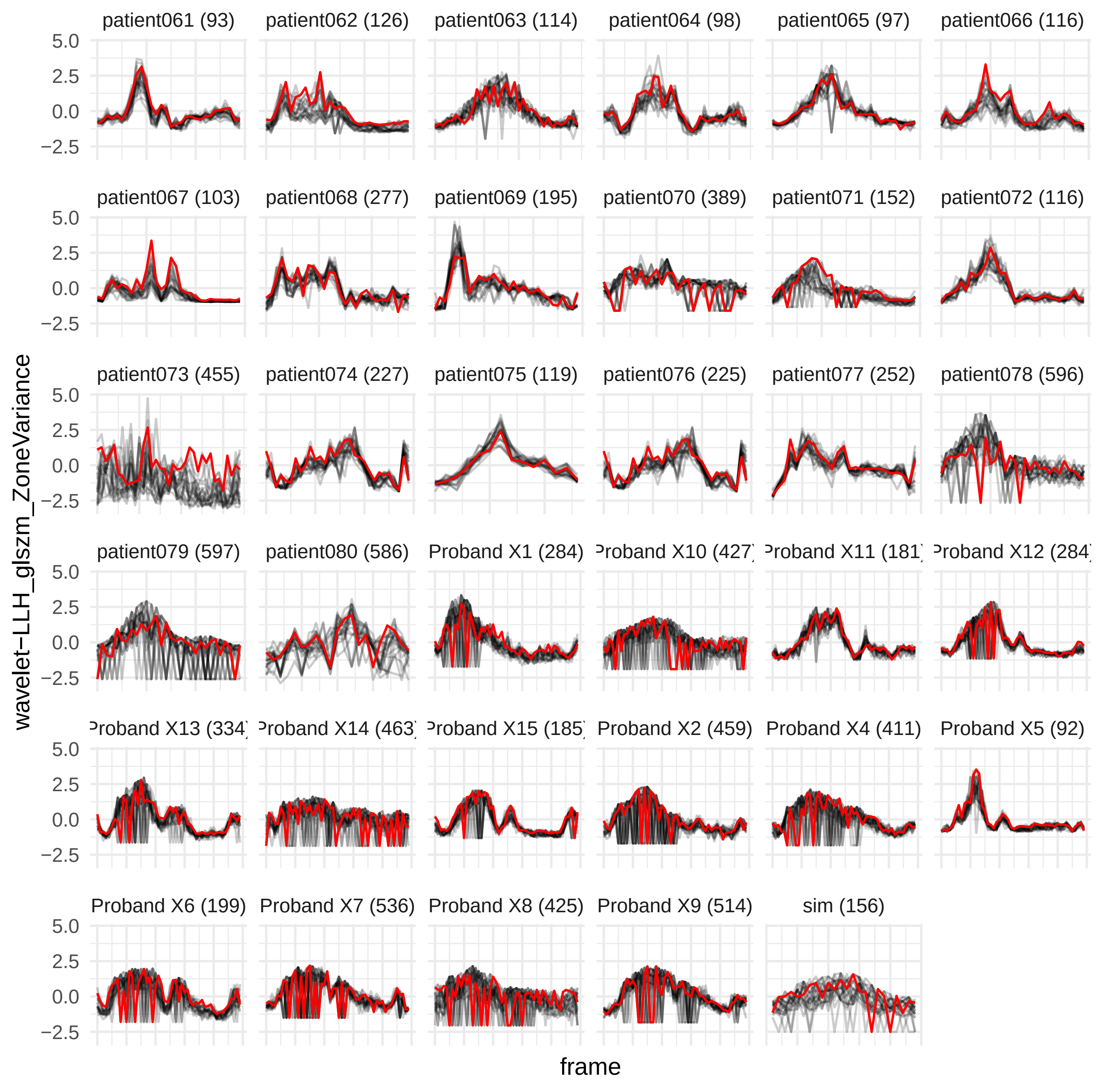

patient061 (93)
patient062 (126)
patient063 (114)
patient064 (98)
patient065 (97)
patient066 (116)
patient067 (103)
patient068 (277)
patient069 (195)
patient070 (389)
patient071 (152)
patient072 (116)
patient073 (455)
patient074 (227)
patient075 (119)
patient076 (225)
patient077 (252)
patient078 (596)
patient079 (597)
patient080 (586)
Proband X1 (284)
Proband X10 (427)
Proband X11 (181)
Proband X12 (284)
Proband X13 (334)
Proband X14 (463)
Proband X15 (185)
Proband X2 (459)
Proband X4 (411)
Proband X5 (92)
Proband X6 (199)
Proband X7 (536)
Proband X8 (425)
Proband X9 (514)
sim (156)
5.0
2.5
0.0
−2.5
wavelet−LLH_glszm_ZoneVariance
frame

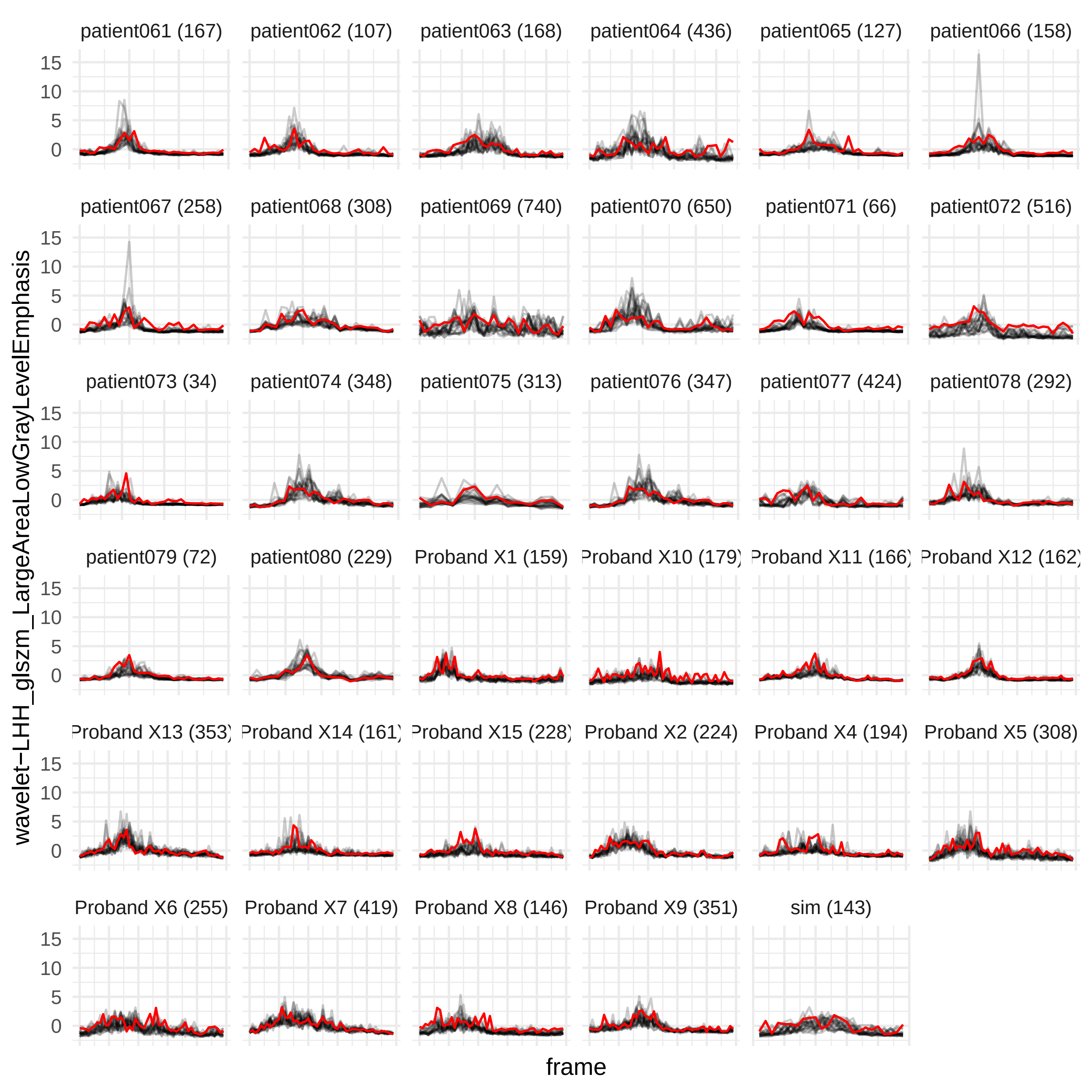
patient061 (167)
patient062 (107)
patient063 (168)
patient064 (436)
patient065 (127)
patient066 (158)
patient067 (258)
patient068 (308)
patient069 (740)
patient070 (650)
patient071 (66)
patient072 (516)
patient073 (34)
patient074 (348)
patient075 (313)
patient076 (347)
patient077 (424)
patient078 (292)
patient079 (72)
patient080 (229)
Proband X1 (159)
Proband X10 (179)
Proband X11 (166)
Proband X12 (162)
Proband X13 (353)
Proband X14 (161)
Proband X15 (228)
Proband X2 (224)
Proband X4 (194)
Proband X5 (308)
Proband X6 (255)
Proband X7 (419)
Proband X8 (146)
Proband X9 (351)
sim (143)
wavelet−LHH_glszm_LargeAreaLowGrayLevelEmphasis
frame
15
10
5
0

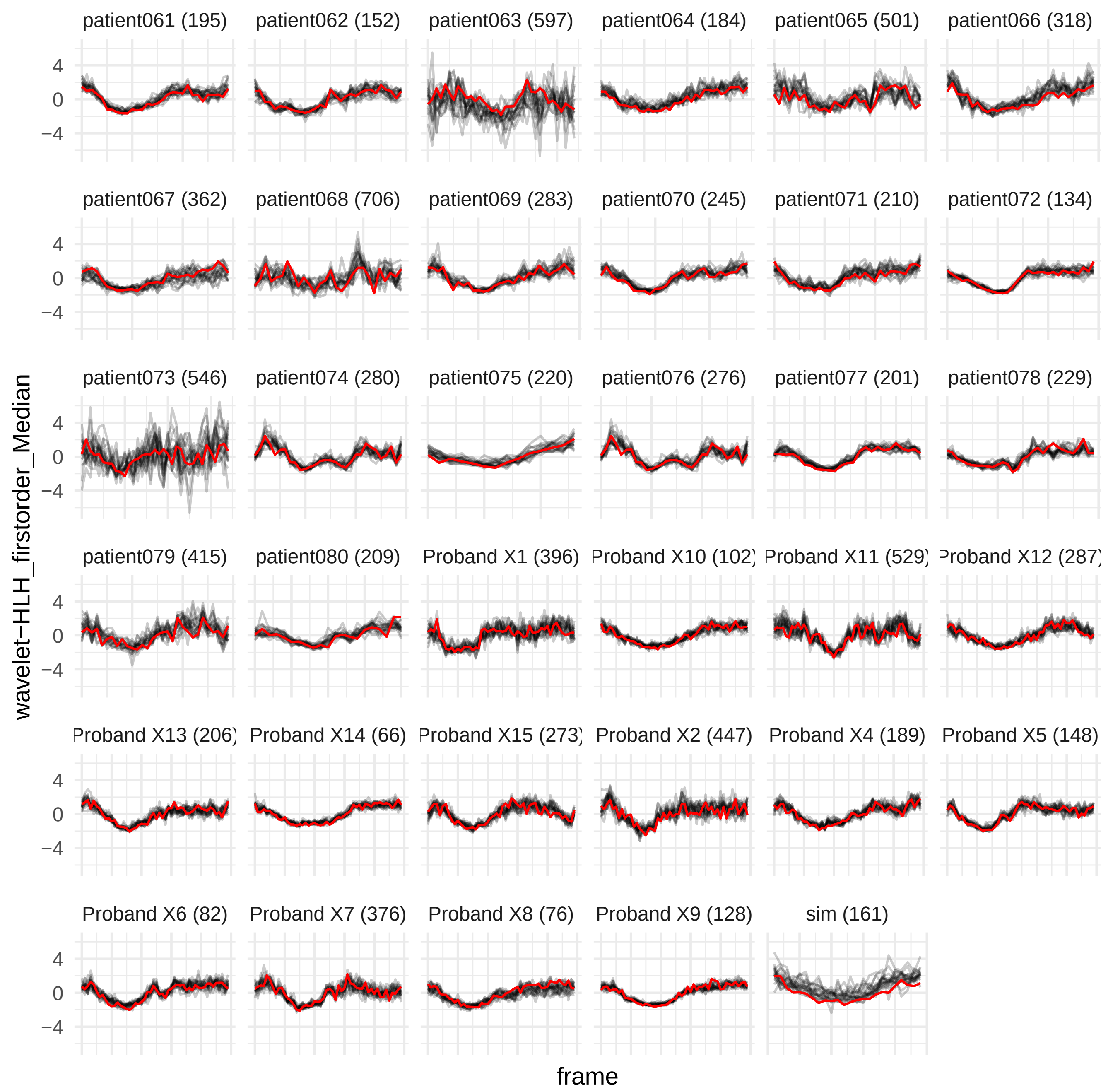
patient061 (195)
patient062 (152)
patient063 (597)
patient064 (184)
patient065 (501)
patient066 (318)
patient067 (362)
patient068 (706)
patient069 (283)
patient070 (245)
patient071 (210)
patient072 (134)
patient073 (546)
patient074 (280)
patient075 (220)
patient076 (276)
patient077 (201)
patient078 (229)
patient079 (415)
patient080 (209)
Proband X1 (396)
Proband X10 (102)
Proband X11 (529)
Proband X12 (287)
Proband X13 (206)
Proband X14 (66)
Proband X15 (273)
Proband X2 (447)
Proband X4 (189)
Proband X5 (148)
Proband X6 (82)
Proband X7 (376)
Proband X8 (76)
Proband X9 (128)
sim (161)
4
0
−4
wavelet−HLH_firstorder_Median
frame

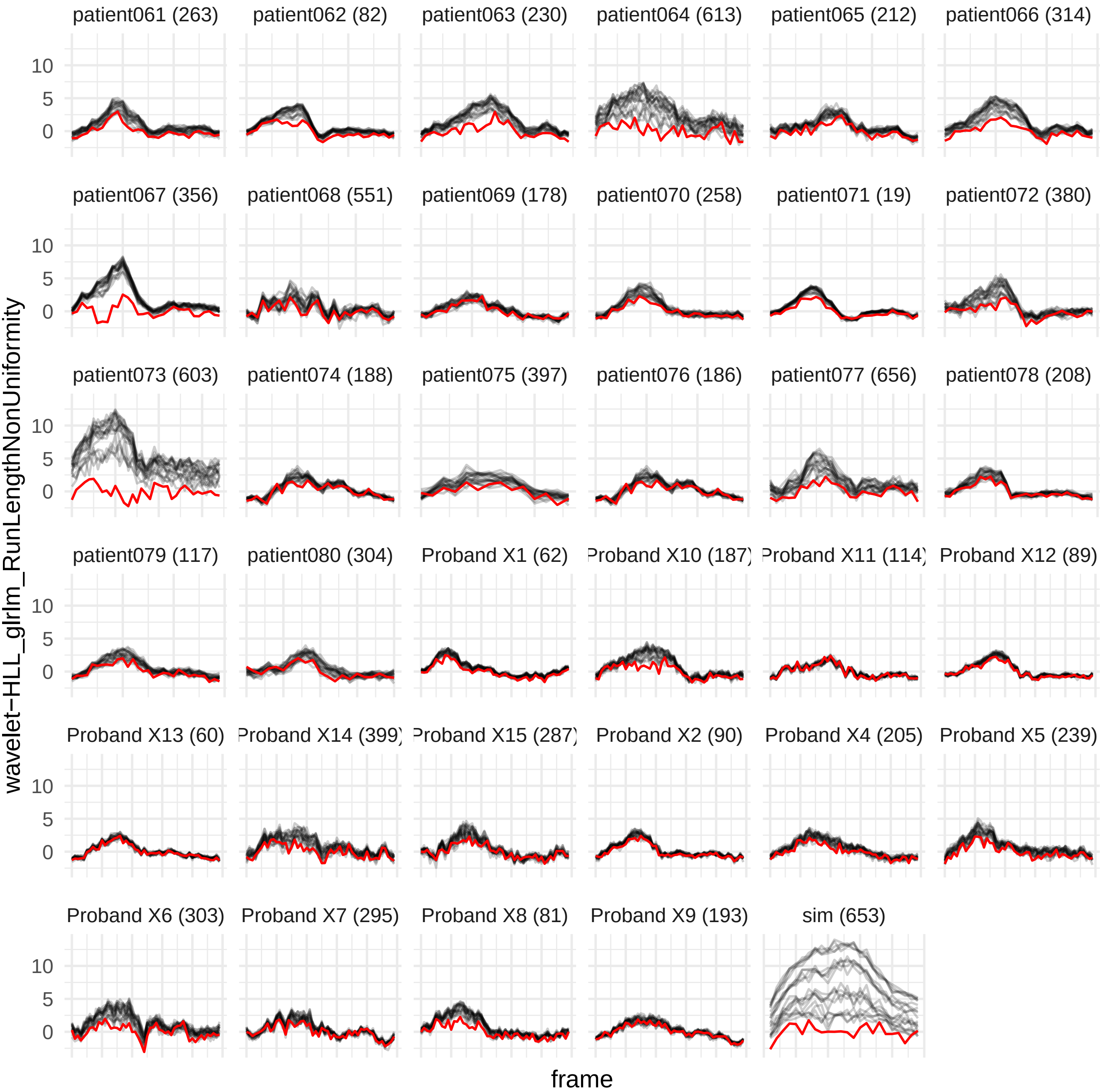
patient061 (263)
patient062 (82)
patient063 (230)
patient064 (613)
patient065 (212)
patient066 (314)
patient067 (356)
patient068 (551)
patient069 (178)
patient070 (258)
patient071 (19)
patient072 (380)
patient073 (603)
patient074 (188)
patient075 (397)
patient076 (186)
patient077 (656)
patient078 (208)
patient079 (117)
patient080 (304)
Proband X1 (62)
Proband X10 (187)
Proband X11 (114)
Proband X12 (89)
Proband X13 (60)
Proband X14 (399)
Proband X15 (287)
Proband X2 (90)
Proband X4 (205)
Proband X5 (239)
Proband X6 (303)
Proband X7 (295)
Proband X8 (81)
Proband X9 (193)
sim (653)
10
5
0
wavelet−HLL_glrlm_RunLengthNonUniformity
frame

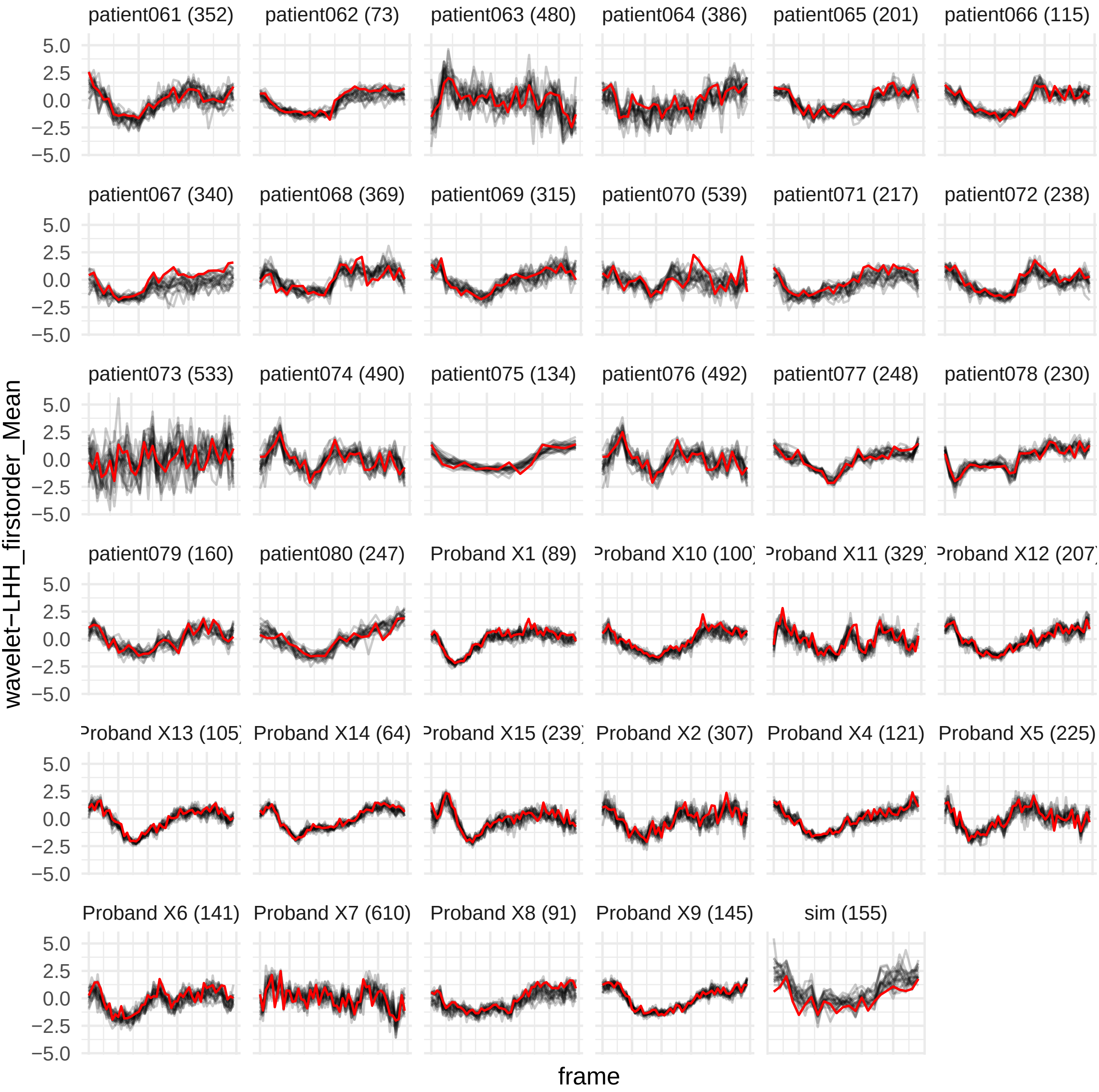

patient061 (352)
patient062 (73)
patient063 (480)
patient064 (386)
patient065 (201)
patient066 (115)
patient067 (340)
patient068 (369)
patient069 (315)
patient070 (539)
patient071 (217)
patient072 (238)
patient073 (533)
patient074 (490)
patient075 (134)
patient076 (492)
patient077 (248)
patient078 (230)
patient079 (160)
patient080 (247)
Proband X1 (89)
Proband X10 (100)
Proband X11 (329)
Proband X12 (207)
Proband X13 (105)
Proband X14 (64)
Proband X15 (239)
Proband X2 (307)
Proband X4 (121)
Proband X5 (225)
Proband X6 (141)
Proband X7 (610)
Proband X8 (91)
Proband X9 (145)
sim (155)
5.0
2.5
0.0
−2.5
−5.0
wavelet−LHH_firstorder_Mean
frame

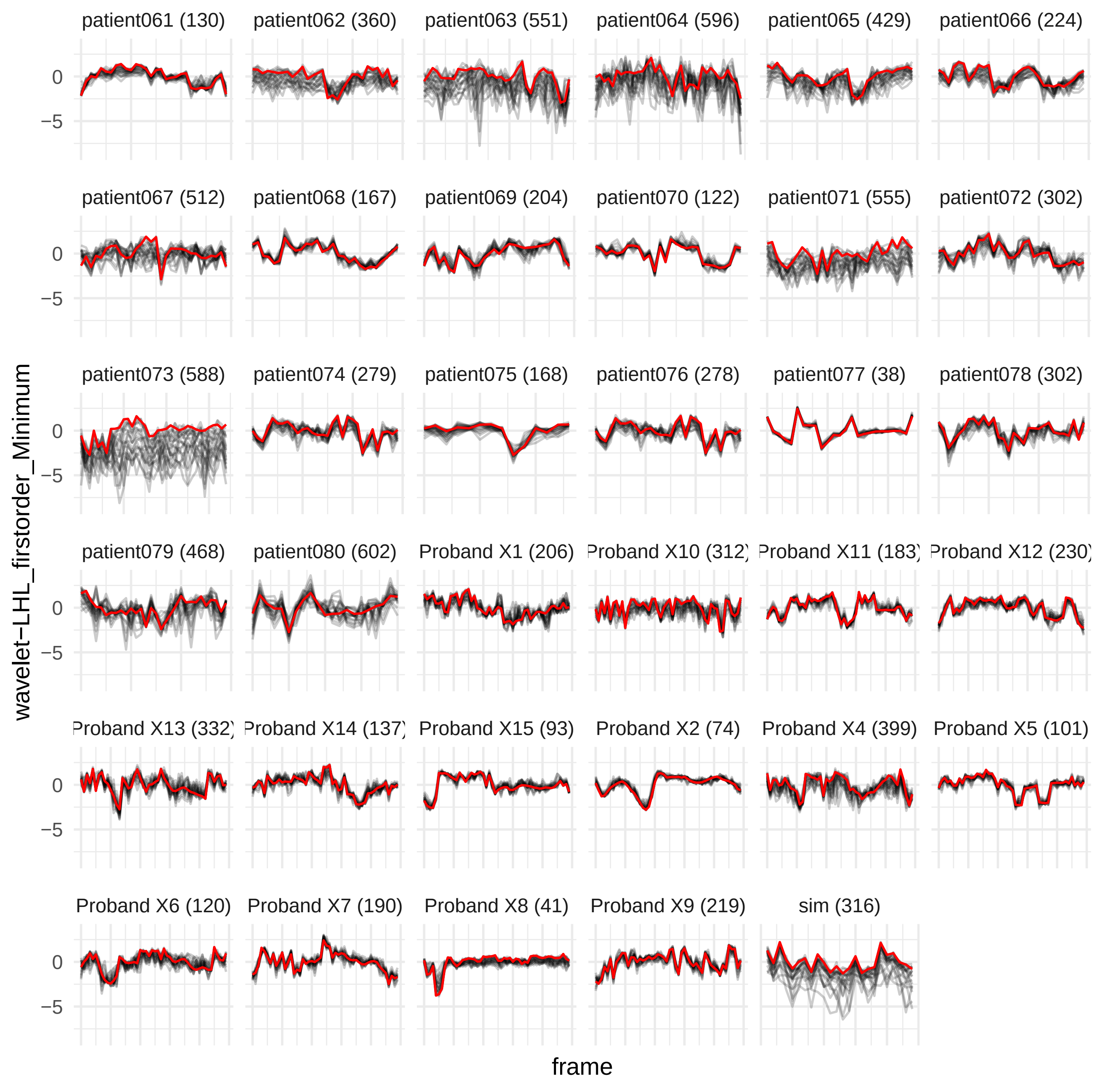

patient061 (130)
patient062 (360)
patient063 (551)
patient064 (596)
patient065 (429)
patient066 (224)
patient067 (512)
patient068 (167)
patient069 (204)
patient070 (122)
patient071 (555)
patient072 (302)
patient073 (588)
patient074 (279)
patient075 (168)
patient076 (278)
patient077 (38)
patient078 (302)
patient079 (468)
patient080 (602)
Proband X1 (206)
Proband X10 (312)
Proband X11 (183)
Proband X12 (230)
Proband X13 (332)
Proband X14 (137)
Proband X15 (93)
Proband X2 (74)
Proband X4 (399)
Proband X5 (101)
Proband X6 (120)
Proband X7 (190)
Proband X8 (41)
Proband X9 (219)
sim (316)
0
−5
wavelet−LHL_firstorder_Minimum
frame

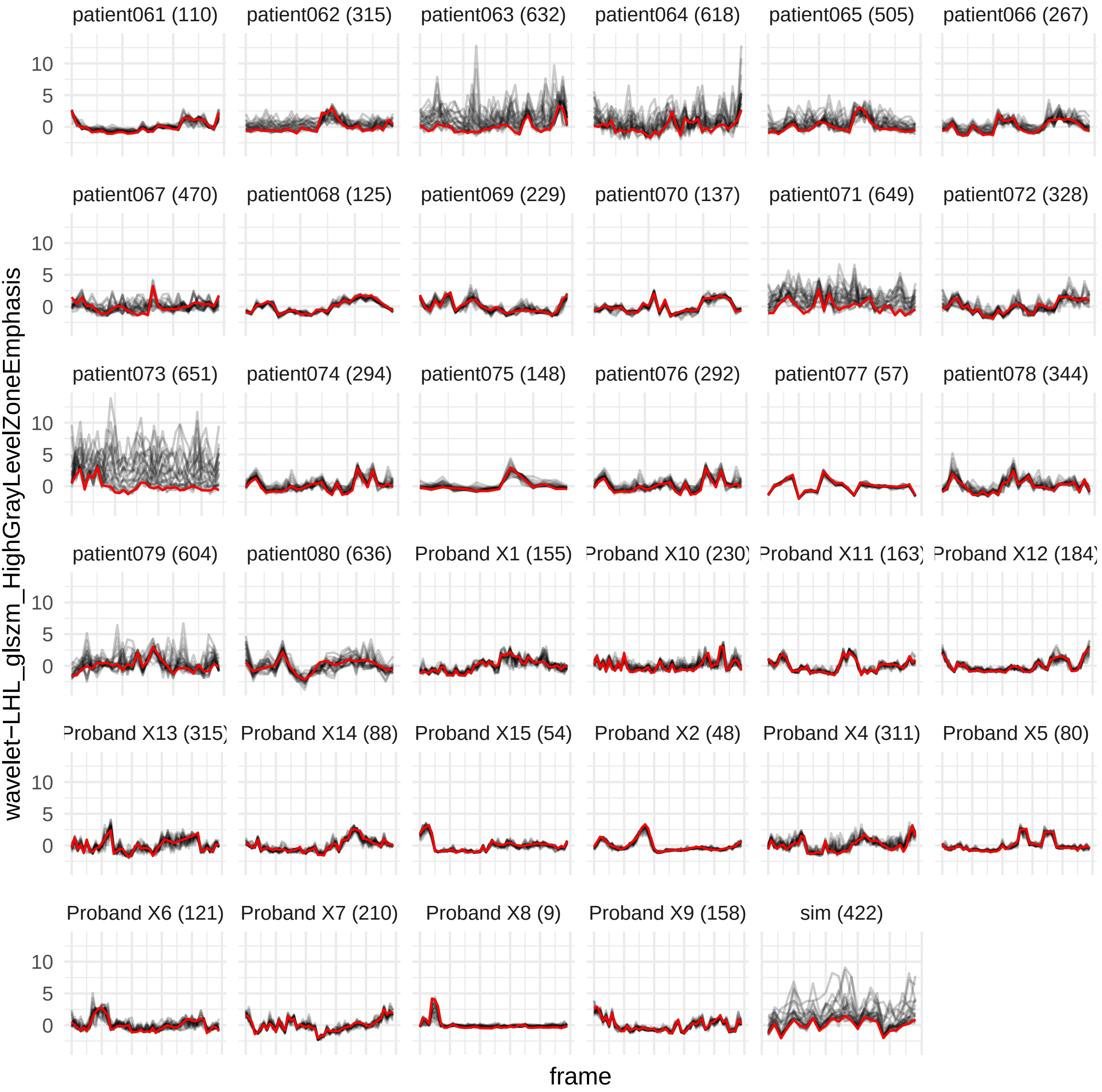

patient061 (110)
patient062 (315)
patient063 (632)
patient064 (618)
patient065 (505)
patient066 (267)
patient067 (470)
patient068 (125)
patient069 (229)
patient070 (137)
patient071 (649)
patient072 (328)
patient073 (651)
patient074 (294)
patient075 (148)
patient076 (292)
patient077 (57)
patient078 (344)
patient079 (604)
patient080 (636)
Proband X1 (155)
Proband X10 (230)
Proband X11 (163)
Proband X12 (184)
Proband X13 (315)
Proband X14 (88)
Proband X15 (54)
Proband X2 (48)
Proband X4 (311)
Proband X5 (80)
Proband X6 (121)
Proband X7 (210)
Proband X8 (9)
Proband X9 (158)
sim (422)
wavelet−LHL_glszm_HighGrayLevelZoneEmphasis
frame
10
5
0

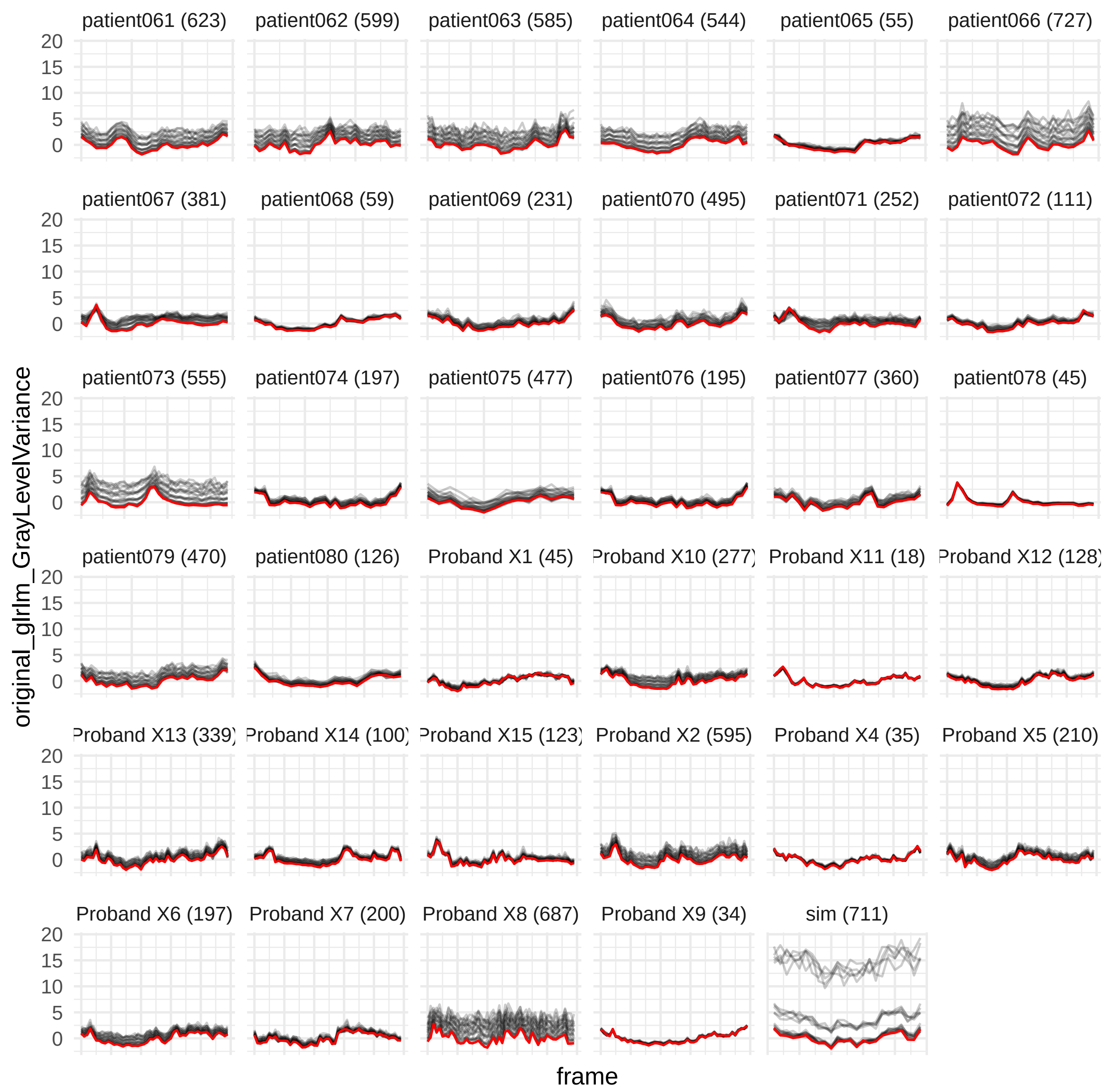
patient061 (623)
patient062 (599)
patient063 (585)
patient064 (544)
patient065 (55)
patient066 (727)
patient067 (381)
patient068 (59)
patient069 (231)
patient070 (495)
patient071 (252)
patient072 (111)
patient073 (555)
patient074 (197)
patient075 (477)
patient076 (195)
patient077 (360)
patient078 (45)
patient079 (470)
patient080 (126)
Proband X1 (45)
Proband X10 (277)
Proband X11 (18)
Proband X12 (128)
Proband X13 (339)
Proband X14 (100)
Proband X15 (123)
Proband X2 (595)
Proband X4 (35)
Proband X5 (210)
Proband X6 (197)
Proband X7 (200)
Proband X8 (687)
Proband X9 (34)
sim (711)
20
15
10
5
0
original_glrlm_GrayLevelVariance
frame

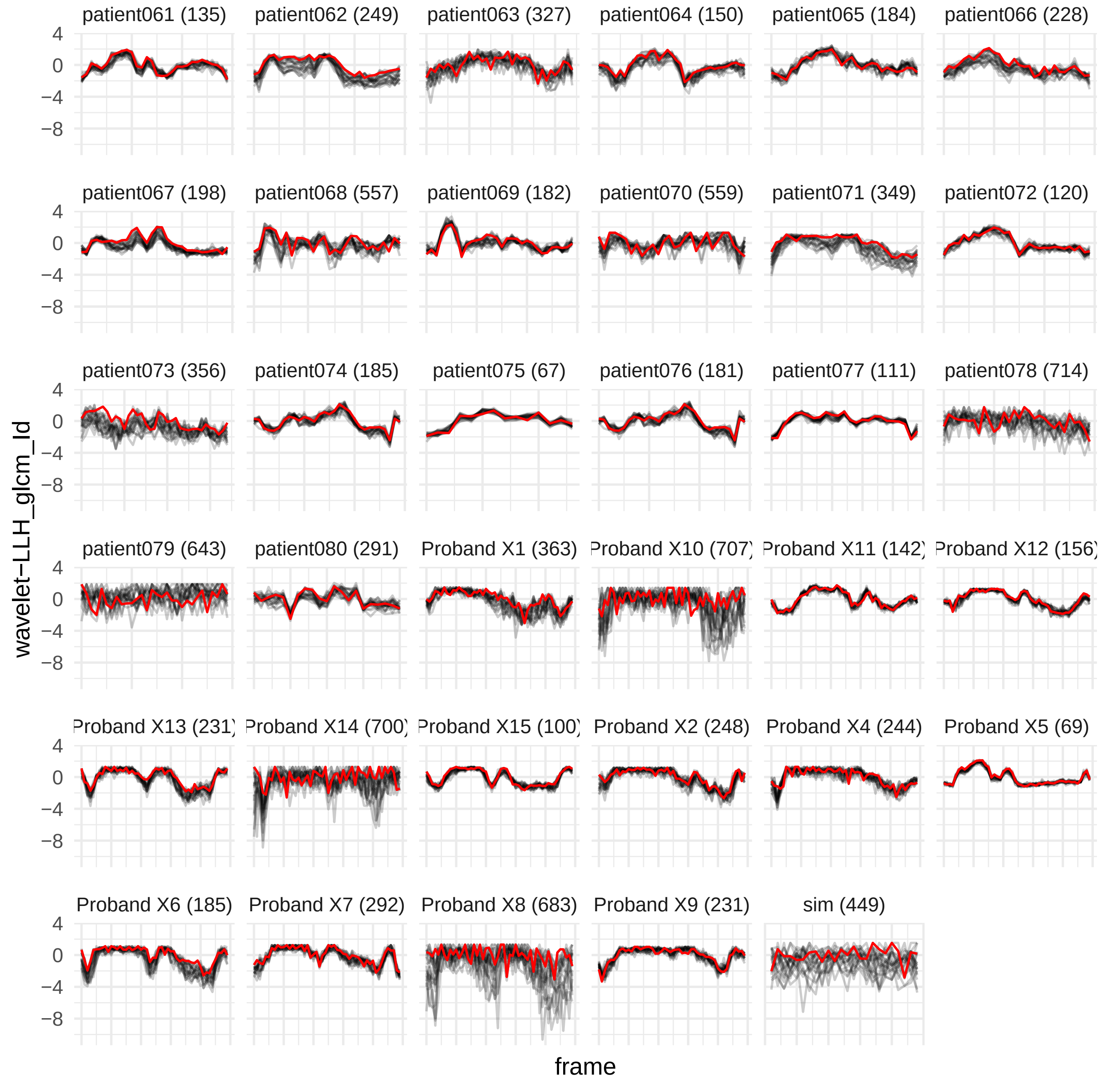

patient061 (135)
patient062 (249)
patient063 (327)
patient064 (150)
patient065 (184)
patient066 (228)
patient067 (198)
patient068 (557)
patient069 (182)
patient070 (559)
patient071 (349)
patient072 (120)
patient073 (356)
patient074 (185)
patient075 (67)
patient076 (181)
patient077 (111)
patient078 (714)
patient079 (643)
patient080 (291)
Proband X1 (363)
Proband X10 (707)
Proband X11 (142)
Proband X12 (156)
Proband X13 (231)
Proband X14 (700)
Proband X15 (100)
Proband X2 (248)
Proband X4 (244)
Proband X5 (69)
Proband X6 (185)
Proband X7 (292)
Proband X8 (683)
Proband X9 (231)
sim (449)
4
0
−4
−8
wavelet−LLH_glcm_Id
frame

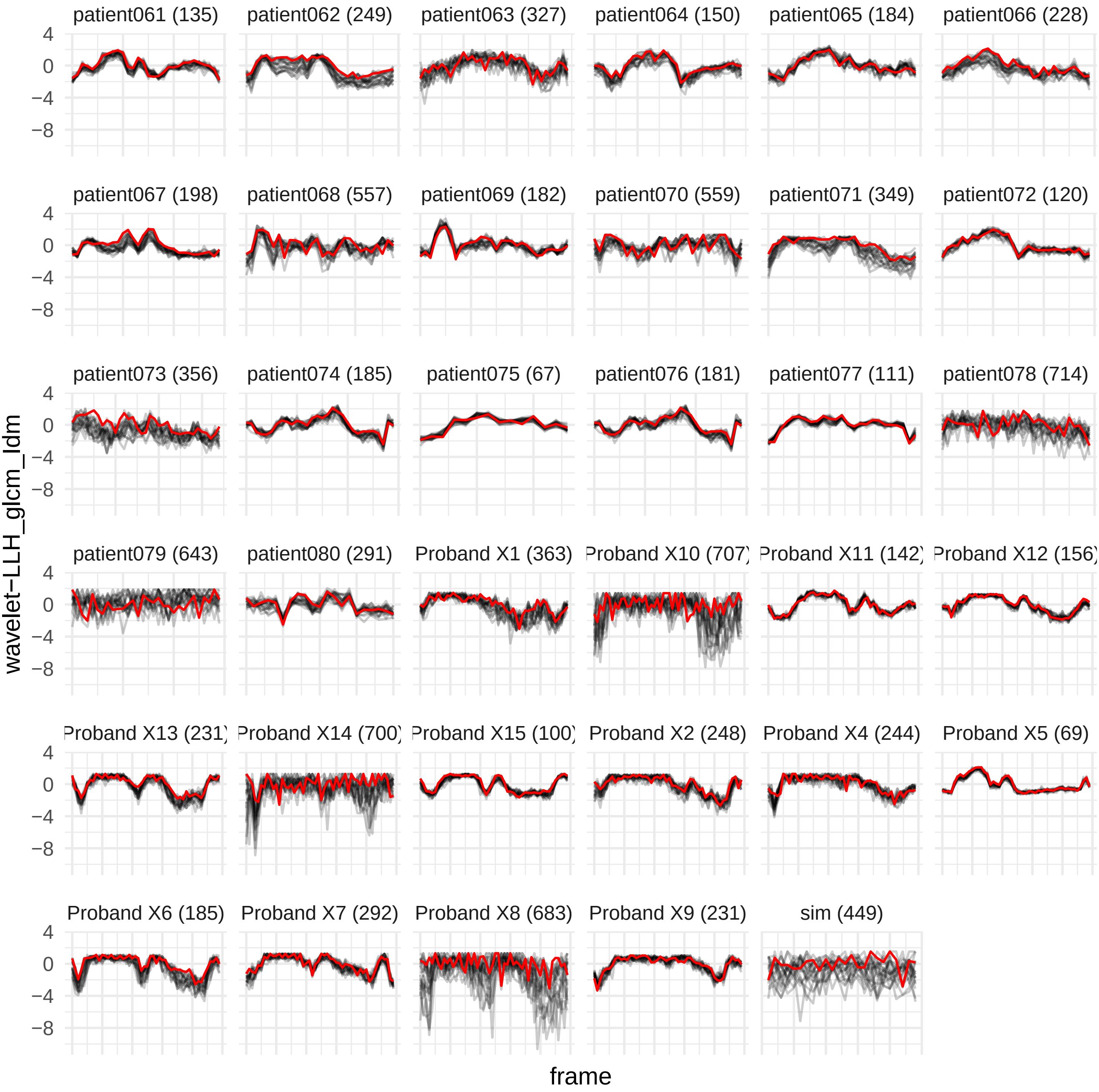

patient061 (135)
patient062 (249)
patient063 (327)
patient064 (150)
patient065 (184)
patient066 (228)
patient067 (198)
patient068 (557)
patient069 (182)
patient070 (559)
patient071 (349)
patient072 (120)
patient073 (356)
patient074 (185)
patient075 (67)
patient076 (181)
patient077 (111)
patient078 (714)
patient079 (643)
patient080 (291)
Proband X1 (363)
Proband X10 (707)
Proband X11 (142)
Proband X12 (156)
Proband X13 (231)
Proband X14 (700)
Proband X15 (100)
Proband X2 (248)
Proband X4 (244)
Proband X5 (69)
Proband X6 (185)
Proband X7 (292)
Proband X8 (683)
Proband X9 (231)
sim (449)
4
0
−4
−8
wavelet−LLH_glcm_Idm
frame

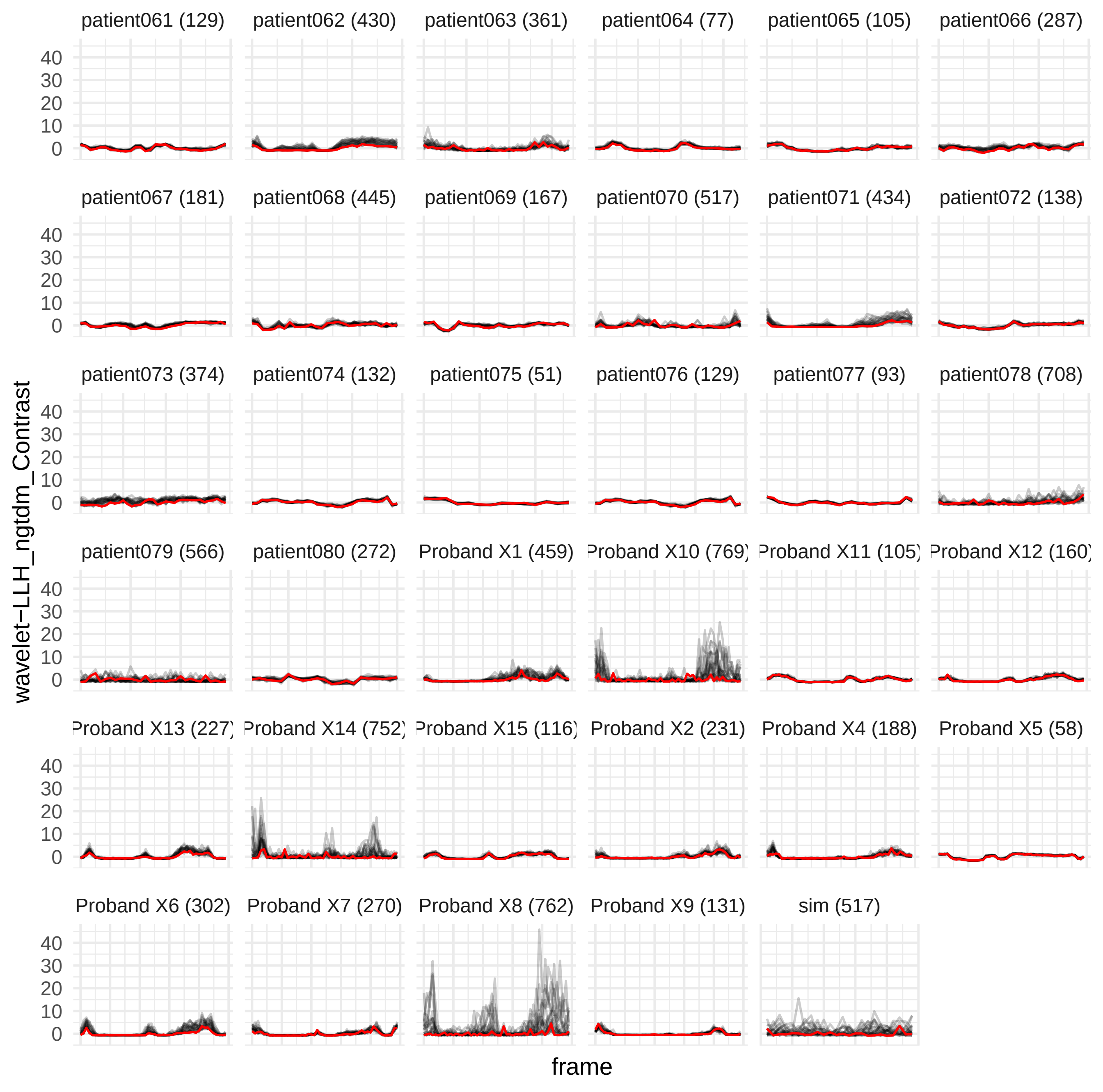

patient061 (129)
patient062 (430)
patient063 (361)
patient064 (77)
patient065 (105)
patient066 (287)
patient067 (181)
patient068 (445)
patient069 (167)
patient070 (517)
patient071 (434)
patient072 (138)
patient073 (374)
patient074 (132)
patient075 (51)
patient076 (129)
patient077 (93)
patient078 (708)
patient079 (566)
patient080 (272)
Proband X1 (459)
Proband X10 (769)
Proband X11 (105)
Proband X12 (160)
Proband X13 (227)
Proband X14 (752)
Proband X15 (116)
Proband X2 (231)
Proband X4 (188)
Proband X5 (58)
Proband X6 (302)
Proband X7 (270)
Proband X8 (762)
Proband X9 (131)
sim (517)
40
30
20
10
0
wavelet−LLH_ngtdm_Contrast
frame

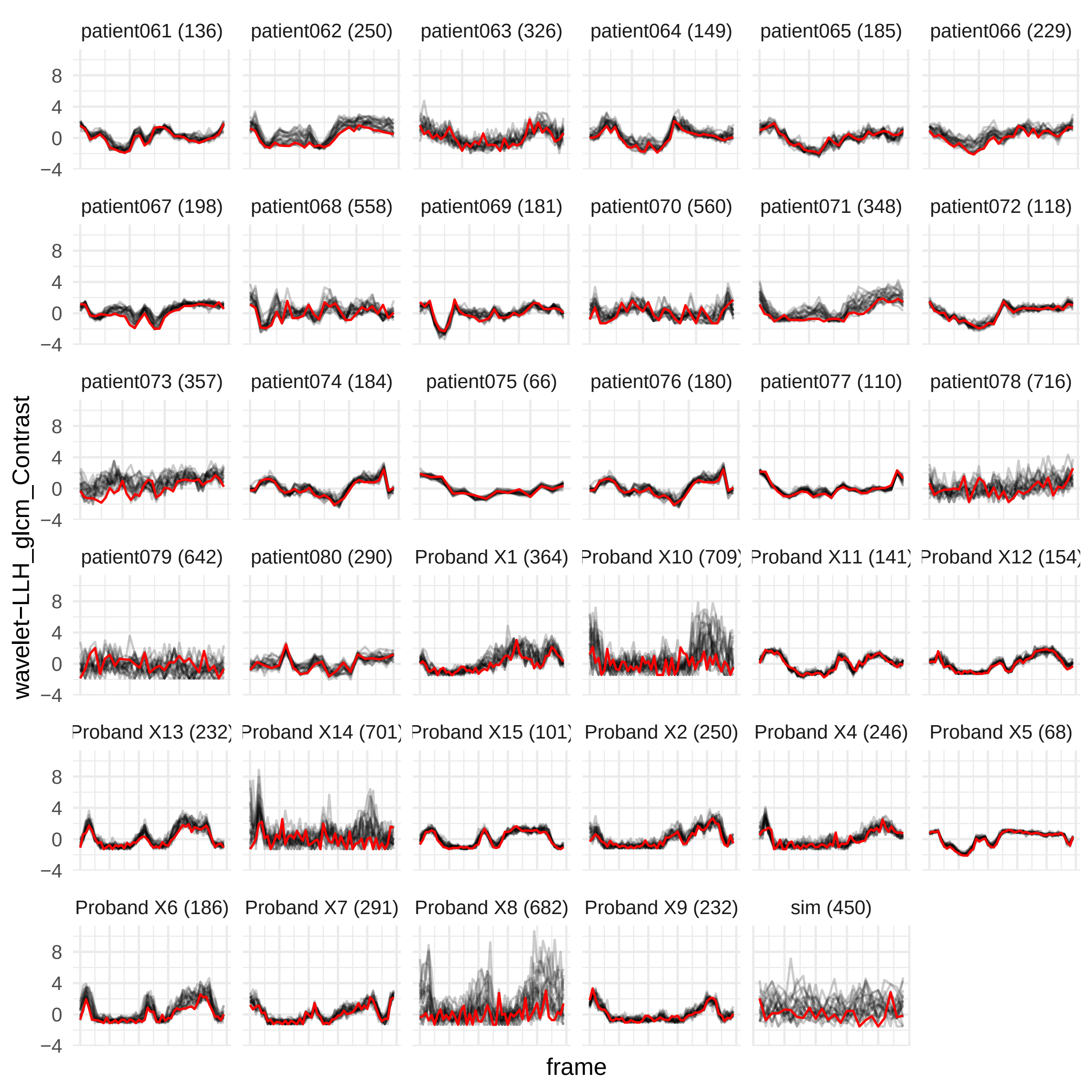
patient061 (136)
patient062 (250)
patient063 (326)
patient064 (149)
patient065 (185)
patient066 (229)
patient067 (198)
patient068 (558)
patient069 (181)
patient070 (560)
patient071 (348)
patient072 (118)
patient073 (357)
patient074 (184)
patient075 (66)
patient076 (180)
patient077 (110)
patient078 (716)
patient079 (642)
patient080 (290)
Proband X1 (364)
Proband X10 (709)
Proband X11 (141)
Proband X12 (154)
Proband X13 (232)
Proband X14 (701)
Proband X15 (101)
Proband X2 (250)
Proband X4 (246)
Proband X5 (68)
Proband X6 (186)
Proband X7 (291)
Proband X8 (682)
Proband X9 (232)
sim (450)
8
4
0
−4
wavelet−LLH_glcm_Contrast
frame

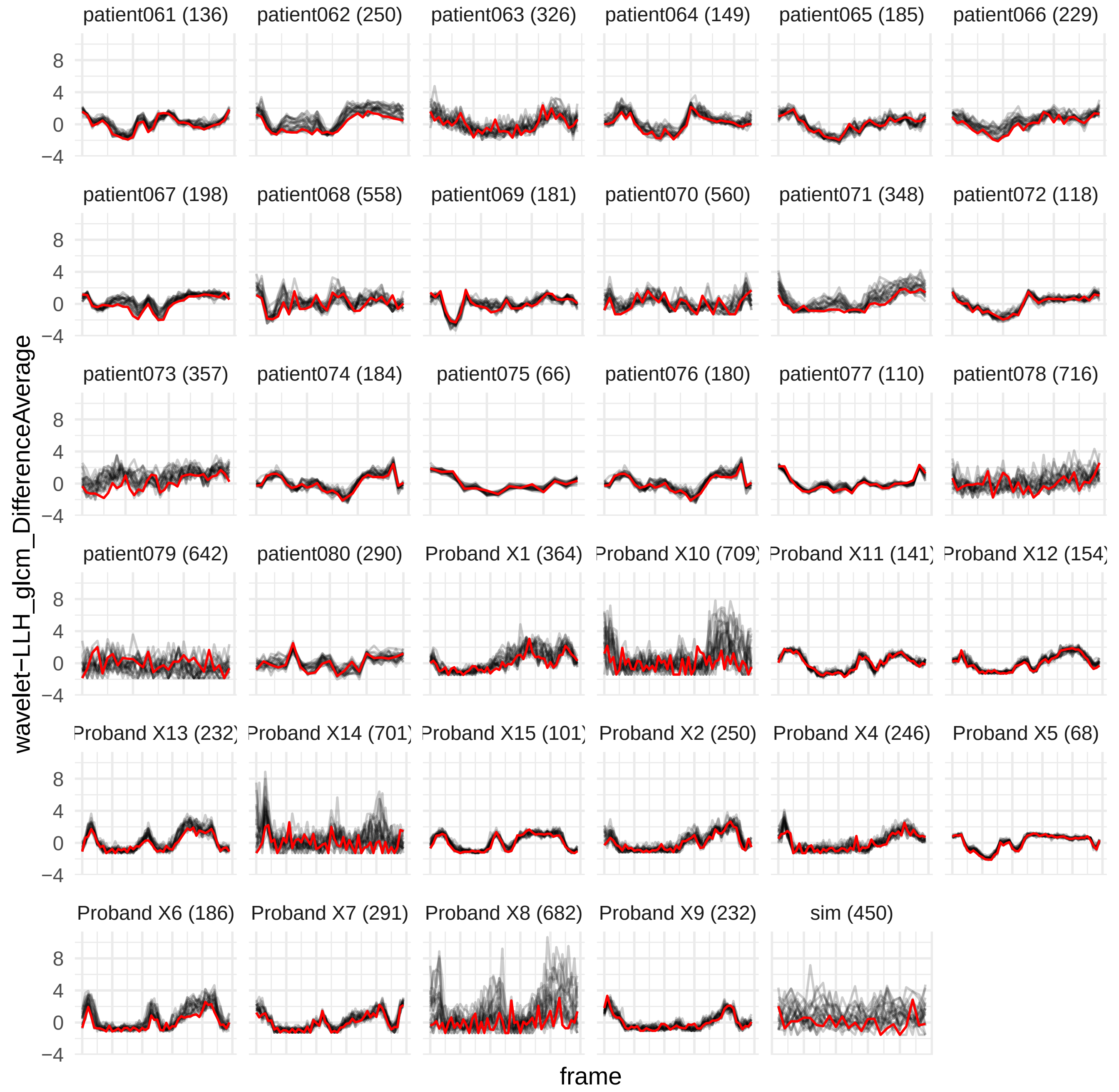

patient061 (136)
patient062 (250)
patient063 (326)
patient064 (149)
patient065 (185)
patient066 (229)
patient067 (198)
patient068 (558)
patient069 (181)
patient070 (560)
patient071 (348)
patient072 (118)
patient073 (357)
patient074 (184)
patient075 (66)
patient076 (180)
patient077 (110)
patient078 (716)
patient079 (642)
patient080 (290)
Proband X1 (364)
Proband X10 (709)
Proband X11 (141)
Proband X12 (154)
Proband X13 (232)
Proband X14 (701)
Proband X15 (101)
Proband X2 (250)
Proband X4 (246)
Proband X5 (68)
Proband X6 (186)
Proband X7 (291)
Proband X8 (682)
Proband X9 (232)
sim (450)
8
4
0
−4
wavelet−LLH_glcm_DifferenceAverage
frame

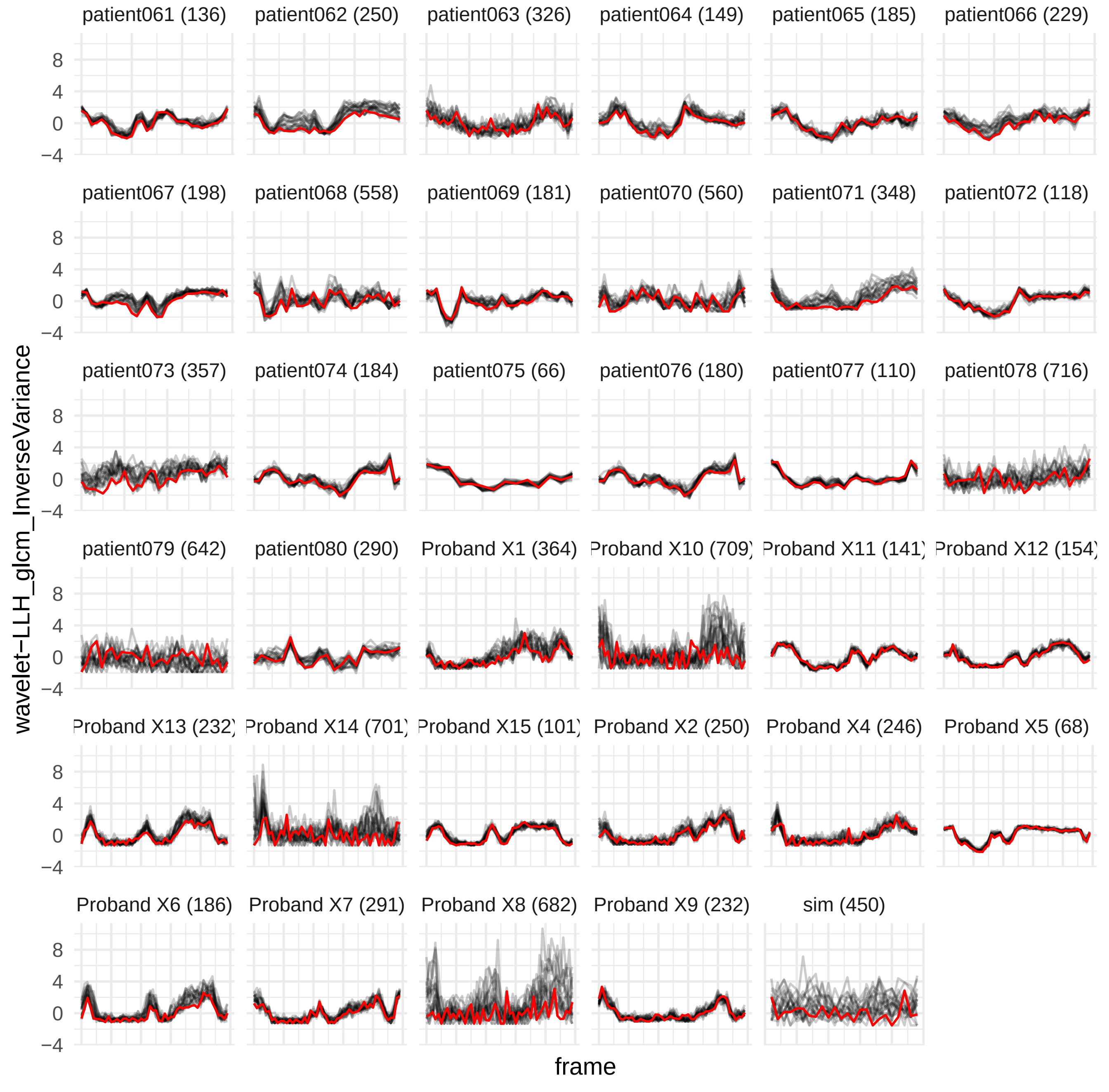
patient061 (136)
patient062 (250)
patient063 (326)
patient064 (149)
patient065 (185)
patient066 (229)
patient067 (198)
patient068 (558)
patient069 (181)
patient070 (560)
patient071 (348)
patient072 (118)
patient073 (357)
patient074 (184)
patient075 (66)
patient076 (180)
patient077 (110)
patient078 (716)
patient079 (642)
patient080 (290)
Proband X1 (364)
Proband X10 (709)
Proband X11 (141)
Proband X12 (154)
Proband X13 (232)
Proband X14 (701)
Proband X15 (101)
Proband X2 (250)
Proband X4 (246)
Proband X5 (68)
Proband X6 (186)
Proband X7 (291)
Proband X8 (682)
Proband X9 (232)
sim (450)
8
4
0
−4
wavelet−LLH_glcm_InverseVariance
frame

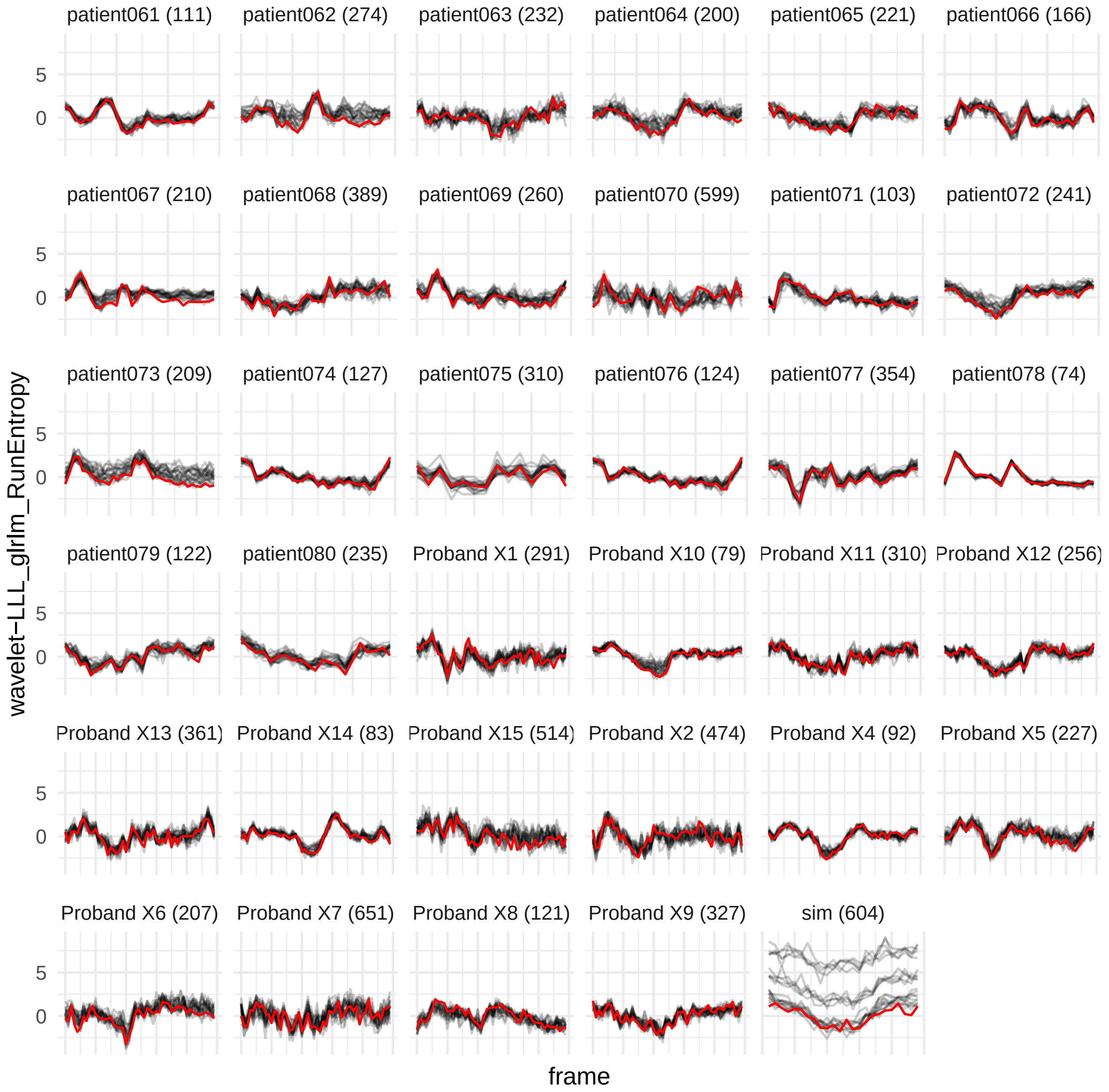
patient061 (111)
patient062 (274)
patient063 (232)
patient064 (200)
patient065 (221)
patient066 (166)
patient067 (210)
patient068 (389)
patient069 (260)
patient070 (599)
patient071 (103)
patient072 (241)
patient073 (209)
patient074 (127)
patient075 (310)
patient076 (124)
patient077 (354)
patient078 (74)
patient079 (122)
patient080 (235)
Proband X1 (291)
Proband X10 (79)
Proband X11 (310)
Proband X12 (256)
Proband X13 (361)
Proband X14 (83)
Proband X15 (514)
Proband X2 (474)
Proband X4 (92)
Proband X5 (227)
Proband X6 (207)
Proband X7 (651)
Proband X8 (121)
Proband X9 (327)
sim (604)
wavelet−LLL_glrlm_RunEntropy
frame
5
0

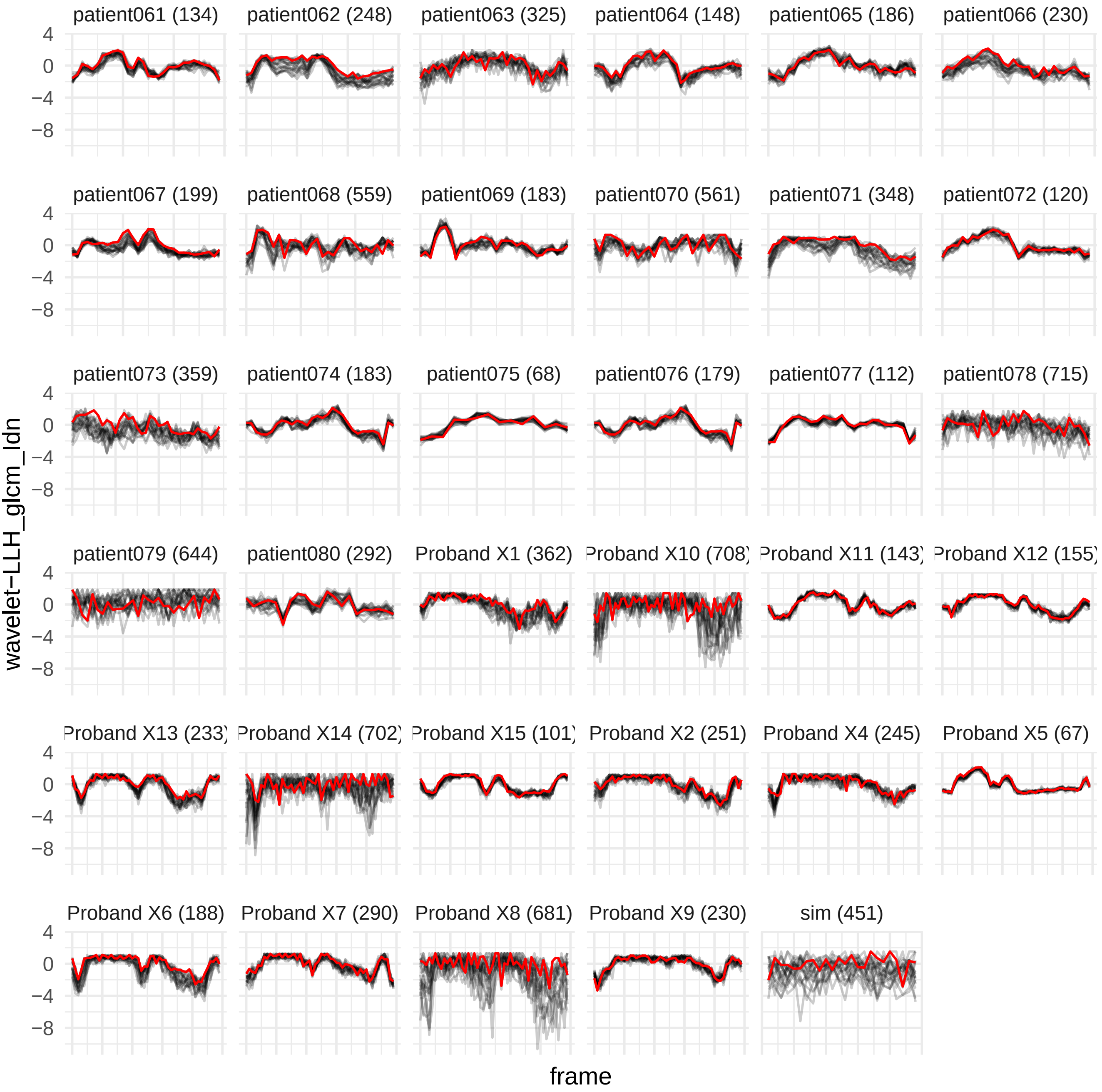
patient061 (134)
patient062 (248)
patient063 (325)
patient064 (148)
patient065 (186)
patient066 (230)
patient067 (199)
patient068 (559)
patient069 (183)
patient070 (561)
patient071 (348)
patient072 (120)
patient073 (359)
patient074 (183)
patient075 (68)
patient076 (179)
patient077 (112)
patient078 (715)
patient079 (644)
patient080 (292)
Proband X1 (362)
Proband X10 (708)
Proband X11 (143)
Proband X12 (155)
Proband X13 (233)
Proband X14 (702)
Proband X15 (101)
Proband X2 (251)
Proband X4 (245)
Proband X5 (67)
Proband X6 (188)
Proband X7 (290)
Proband X8 (681)
Proband X9 (230)
sim (451)
wavelet−LLH_glcm_Idn
frame
4
0
−4
−8

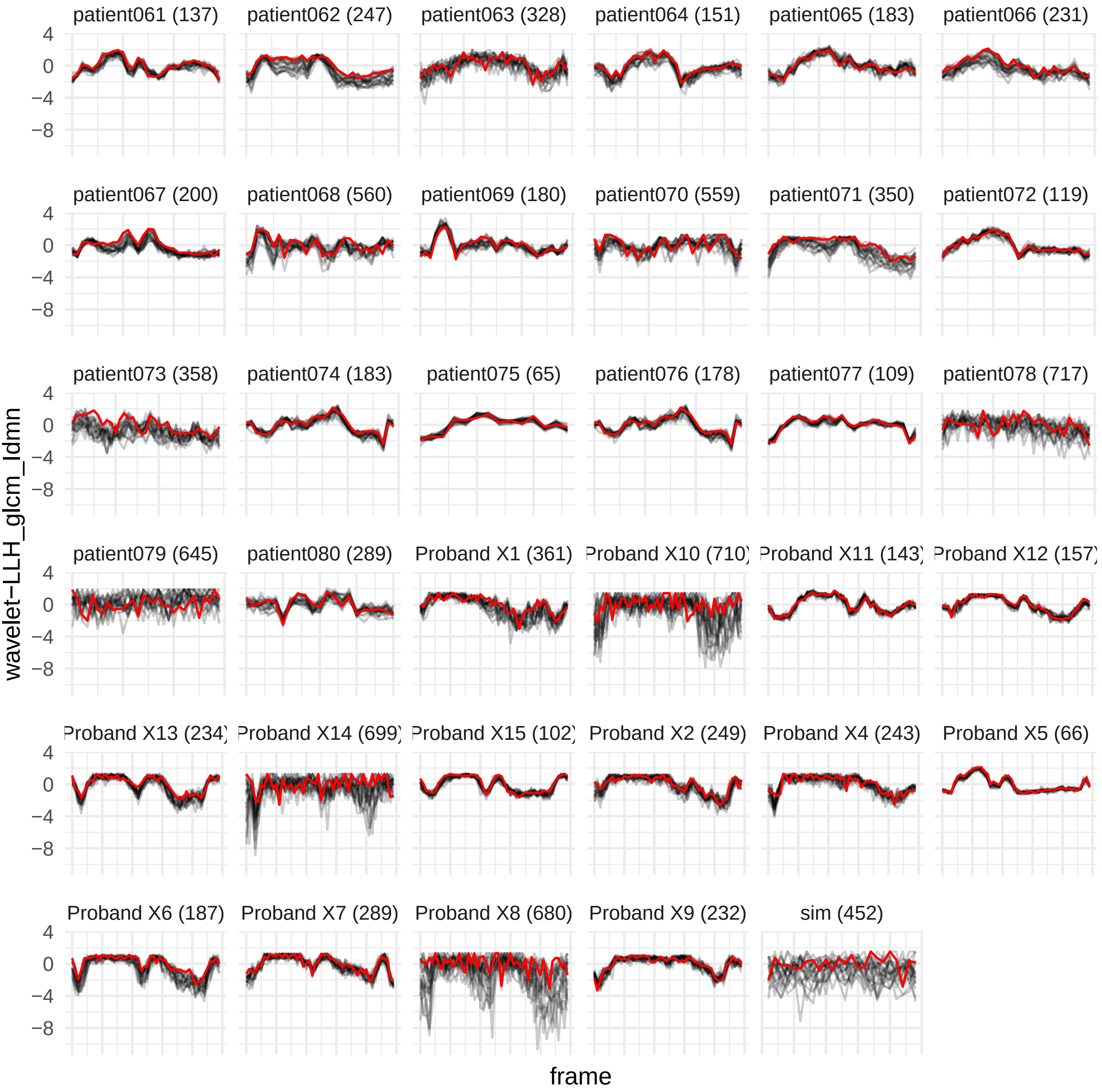
patient061 (137)
patient062 (247)
patient063 (328)
patient064 (151)
patient065 (183)
patient066 (231)
patient067 (200)
patient068 (560)
patient069 (180)
patient070 (559)
patient071 (350)
patient072 (119)
patient073 (358)
patient074 (183)
patient075 (65)
patient076 (178)
patient077 (109)
patient078 (717)
patient079 (645)
patient080 (289)
Proband X1 (361)
Proband X10 (710)
Proband X11 (143)
Proband X12 (157)
Proband X13 (234)
Proband X14 (699)
Proband X15 (102)
Proband X2 (249)
Proband X4 (243)
Proband X5 (66)
Proband X6 (187)
Proband X7 (289)
Proband X8 (680)
Proband X9 (232)
sim (452)
4
0
−4
−8
wavelet−LLH_glcm_Idmn
frame

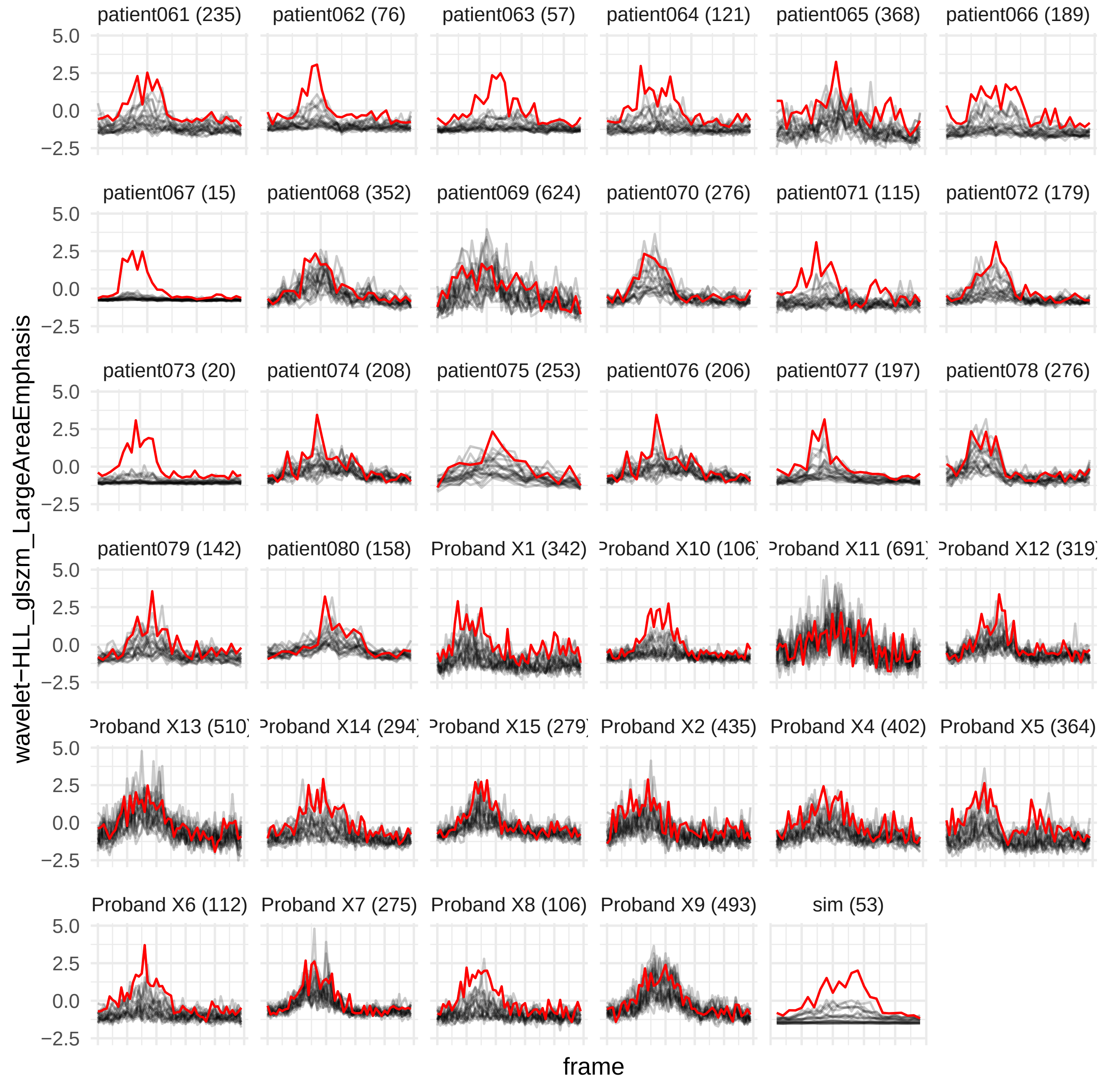

patient061 (235)
patient062 (76)
patient063 (57)
patient064 (121)
patient065 (368)
patient066 (189)
patient067 (15)
patient068 (352)
patient069 (624)
patient070 (276)
patient071 (115)
patient072 (179)
patient073 (20)
patient074 (208)
patient075 (253)
patient076 (206)
patient077 (197)
patient078 (276)
patient079 (142)
patient080 (158)
Proband X1 (342)
Proband X10 (106)
Proband X11 (691)
Proband X12 (319)
Proband X13 (510)
Proband X14 (294)
Proband X15 (279)
Proband X2 (435)
Proband X4 (402)
Proband X5 (364)
Proband X6 (112)
Proband X7 (275)
Proband X8 (106)
Proband X9 (493)
sim (53)
5.0
2.5
0.0
−2.5
wavelet−HLL_glszm_LargeAreaEmphasis
frame

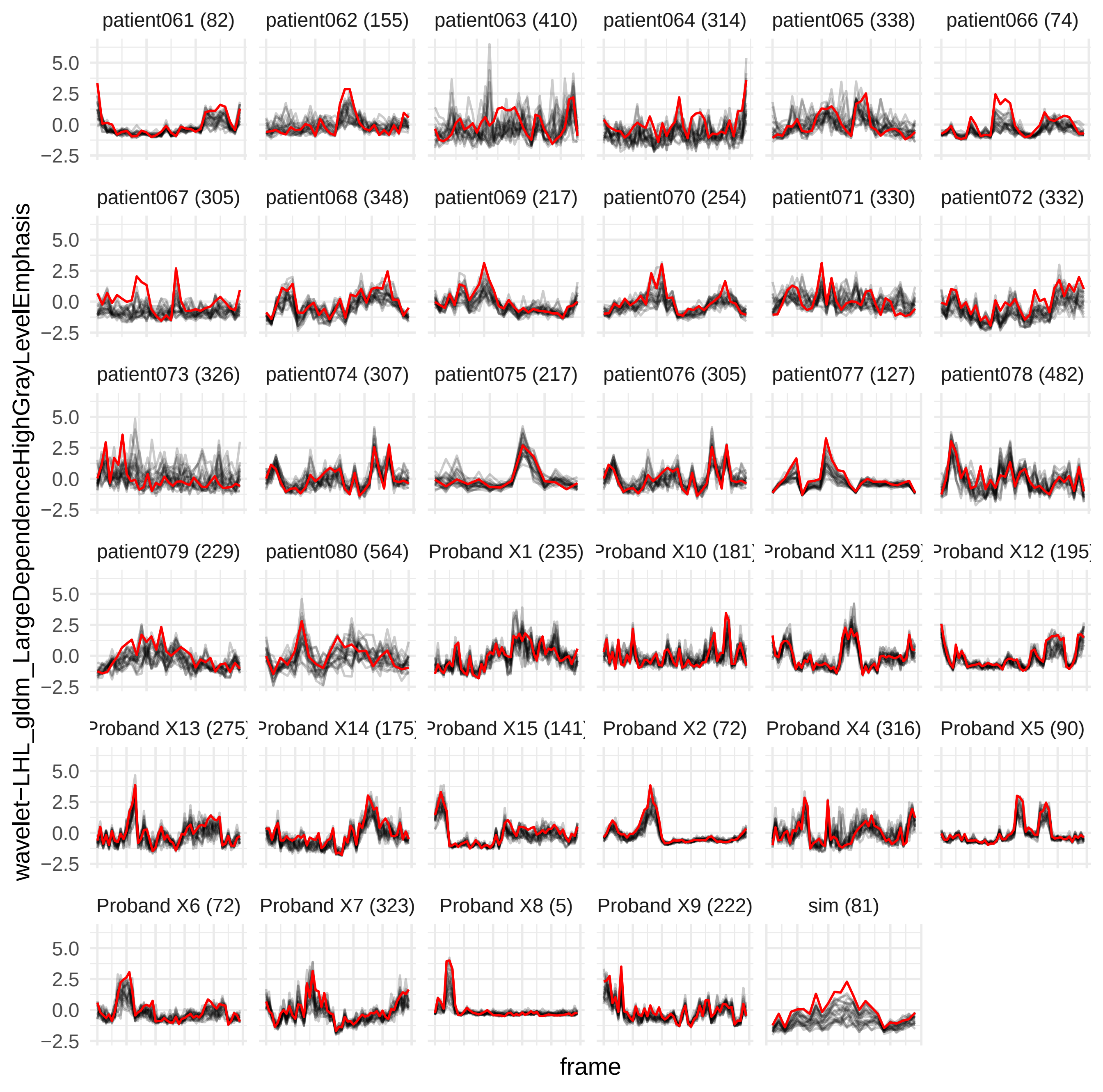
patient061 (82)
patient062 (155)
patient063 (410)
patient064 (314)
patient065 (338)
patient066 (74)
patient067 (305)
patient068 (348)
patient069 (217)
patient070 (254)
patient071 (330)
patient072 (332)
patient073 (326)
patient074 (307)
patient075 (217)
patient076 (305)
patient077 (127)
patient078 (482)
patient079 (229)
patient080 (564)
Proband X1 (235)
Proband X10 (181)
Proband X11 (259)
Proband X12 (195)
Proband X13 (275)
Proband X14 (175)
Proband X15 (141)
Proband X2 (72)
Proband X4 (316)
Proband X5 (90)
Proband X6 (72)
Proband X7 (323)
Proband X8 (5)
Proband X9 (222)
sim (81)
5.0
2.5
0.0
−2.5
wavelet−LHL_gldm_LargeDependenceHighGrayLevelEmphasis
frame

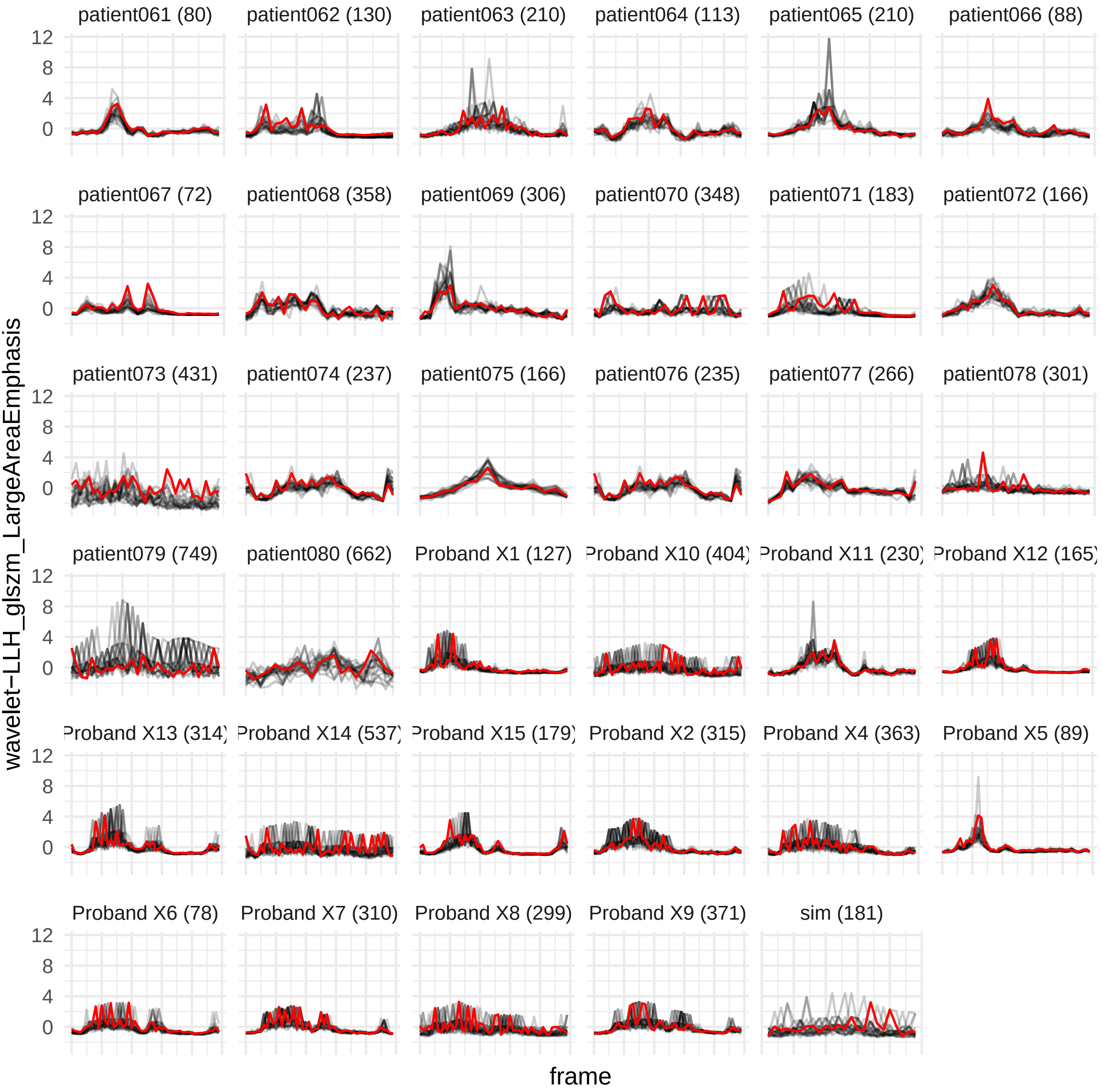
patient061 (80)
patient062 (130)
patient063 (210)
patient064 (113)
patient065 (210)
patient066 (88)
patient067 (72)
patient068 (358)
patient069 (306)
patient070 (348)
patient071 (183)
patient072 (166)
patient073 (431)
patient074 (237)
patient075 (166)
patient076 (235)
patient077 (266)
patient078 (301)
patient079 (749)
patient080 (662)
Proband X1 (127)
Proband X10 (404)
Proband X11 (230)
Proband X12 (165)
Proband X13 (314)
Proband X14 (537)
Proband X15 (179)
Proband X2 (315)
Proband X4 (363)
Proband X5 (89)
Proband X6 (78)
Proband X7 (310)
Proband X8 (299)
Proband X9 (371)
sim (181)
12
8
4
0
wavelet−LLH_glszm_LargeAreaEmphasis
frame

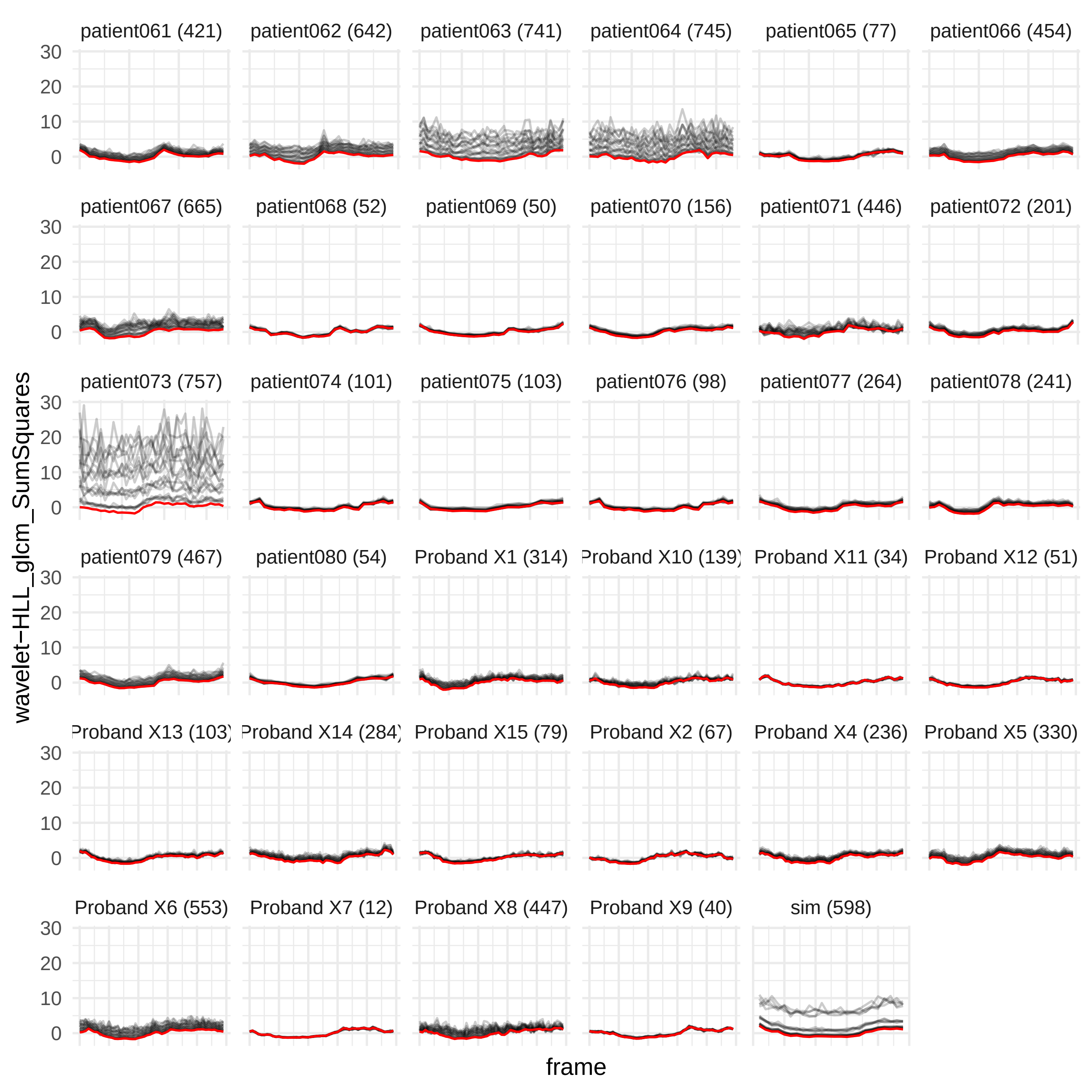

patient061 (421)
patient062 (642)
patient063 (741)
patient064 (745)
patient065 (77)
patient066 (454)
patient067 (665)
patient068 (52)
patient069 (50)
patient070 (156)
patient071 (446)
patient072 (201)
patient073 (757)
patient074 (101)
patient075 (103)
patient076 (98)
patient077 (264)
patient078 (241)
patient079 (467)
patient080 (54)
Proband X1 (314)
Proband X10 (139)
Proband X11 (34)
Proband X12 (51)
Proband X13 (103)
Proband X14 (284)
Proband X15 (79)
Proband X2 (67)
Proband X4 (236)
Proband X5 (330)
Proband X6 (553)
Proband X7 (12)
Proband X8 (447)
Proband X9 (40)
sim (598)
30
20
10
0
wavelet−HLL_glcm_SumSquares
frame

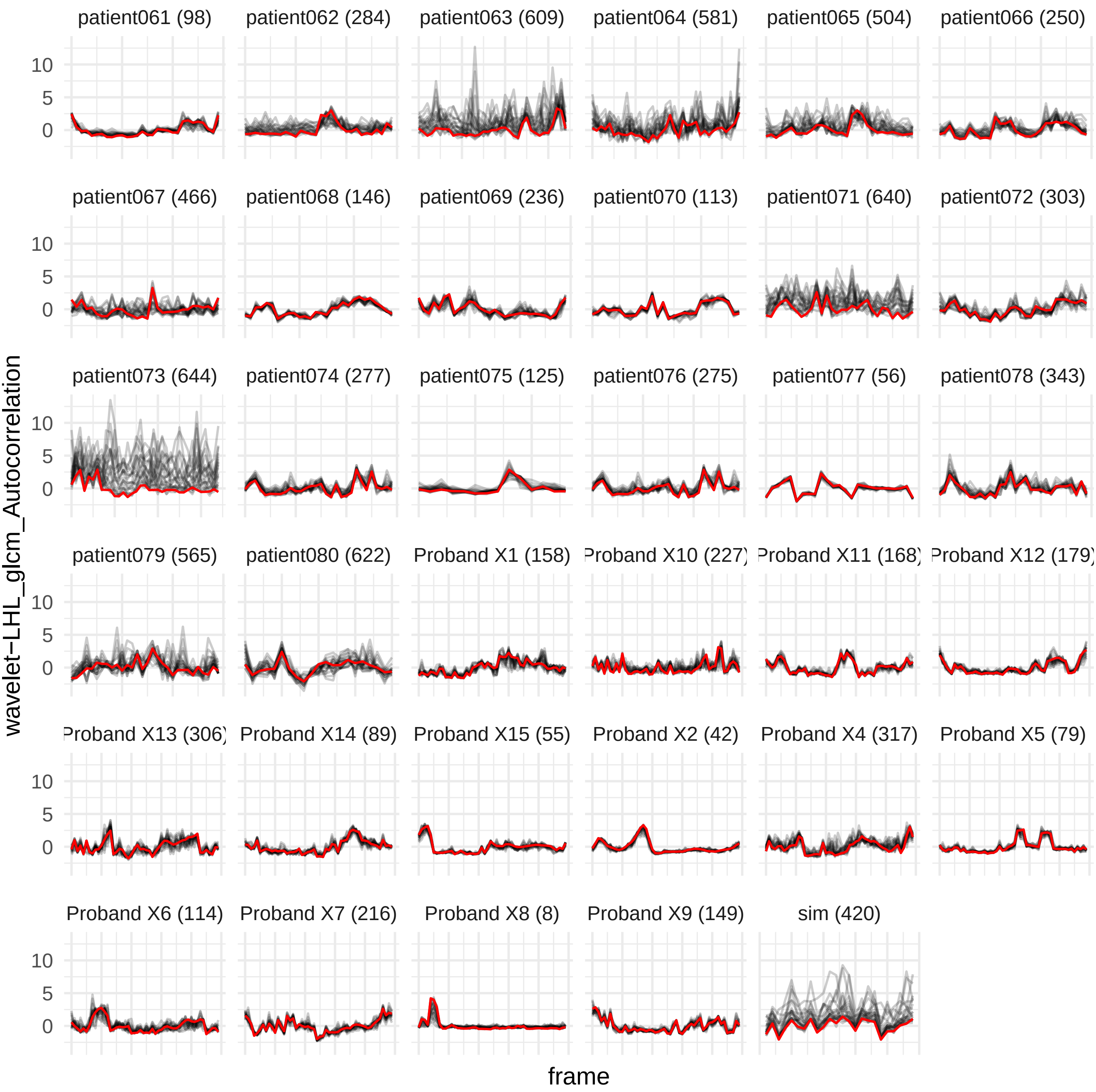
patient061 (98)
patient062 (284)
patient063 (609)
patient064 (581)
patient065 (504)
patient066 (250)
patient067 (466)
patient068 (146)
patient069 (236)
patient070 (113)
patient071 (640)
patient072 (303)
patient073 (644)
patient074 (277)
patient075 (125)
patient076 (275)
patient077 (56)
patient078 (343)
patient079 (565)
patient080 (622)
Proband X1 (158)
Proband X10 (227)
Proband X11 (168)
Proband X12 (179)
Proband X13 (306)
Proband X14 (89)
Proband X15 (55)
Proband X2 (42)
Proband X4 (317)
Proband X5 (79)
Proband X6 (114)
Proband X7 (216)
Proband X8 (8)
Proband X9 (149)
sim (420)
0
5
10
wavelet−LHL_glcm_Autocorrelation
frame

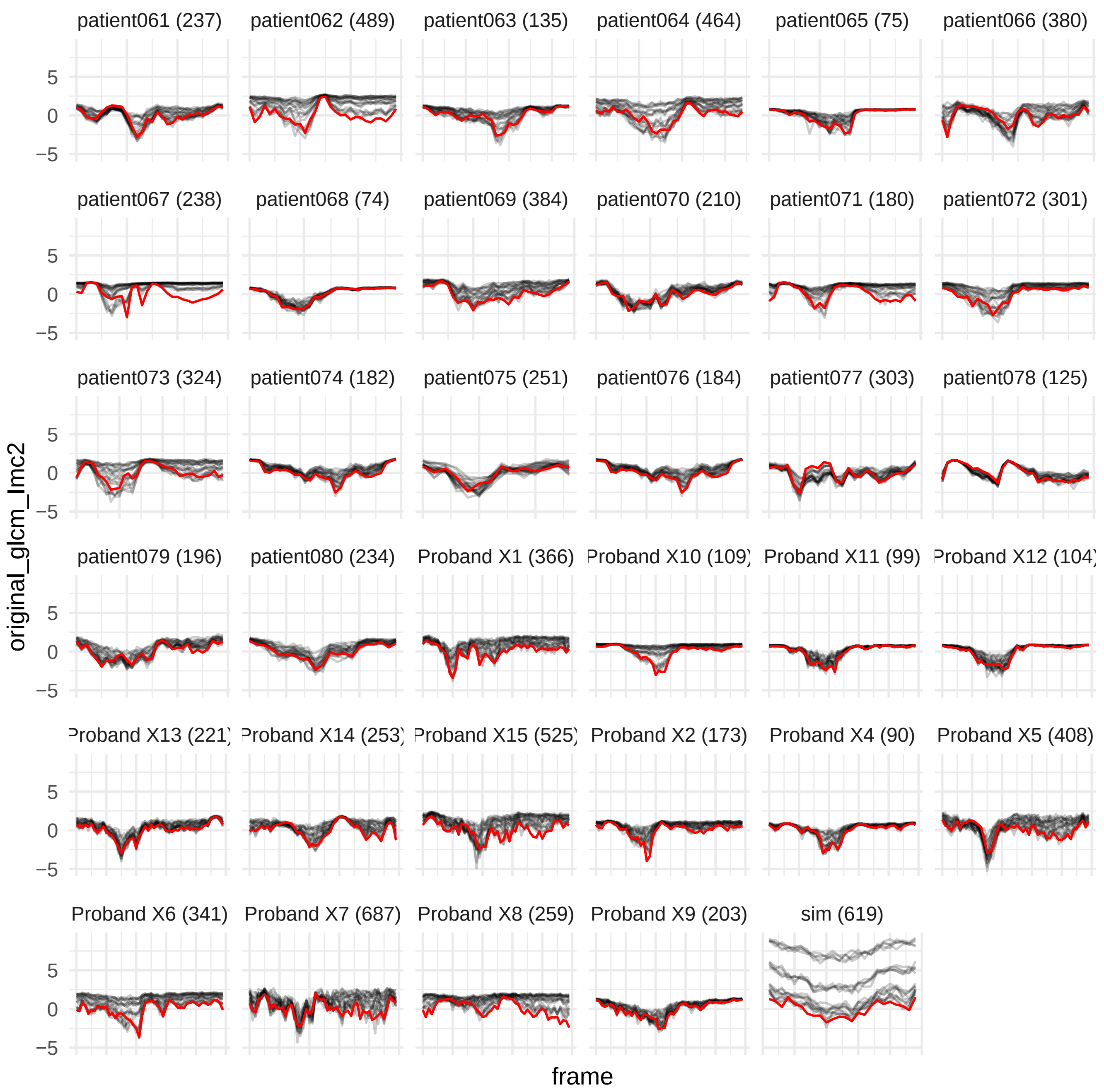

patient061 (237)
patient062 (489)
patient063 (135)
patient064 (464)
patient065 (75)
patient066 (380)
patient067 (238)
patient068 (74)
patient069 (384)
patient070 (210)
patient071 (180)
patient072 (301)
patient073 (324)
patient074 (182)
patient075 (251)
patient076 (184)
patient077 (303)
patient078 (125)
patient079 (196)
patient080 (234)
Proband X1 (366)
Proband X10 (109)
Proband X11 (99)
Proband X12 (104)
Proband X13 (221)
Proband X14 (253)
Proband X15 (525)
Proband X2 (173)
Proband X4 (90)
Proband X5 (408)
Proband X6 (341)
Proband X7 (687)
Proband X8 (259)
Proband X9 (203)
sim (619)
original_glcm_Imc2
5
0
−5
frame

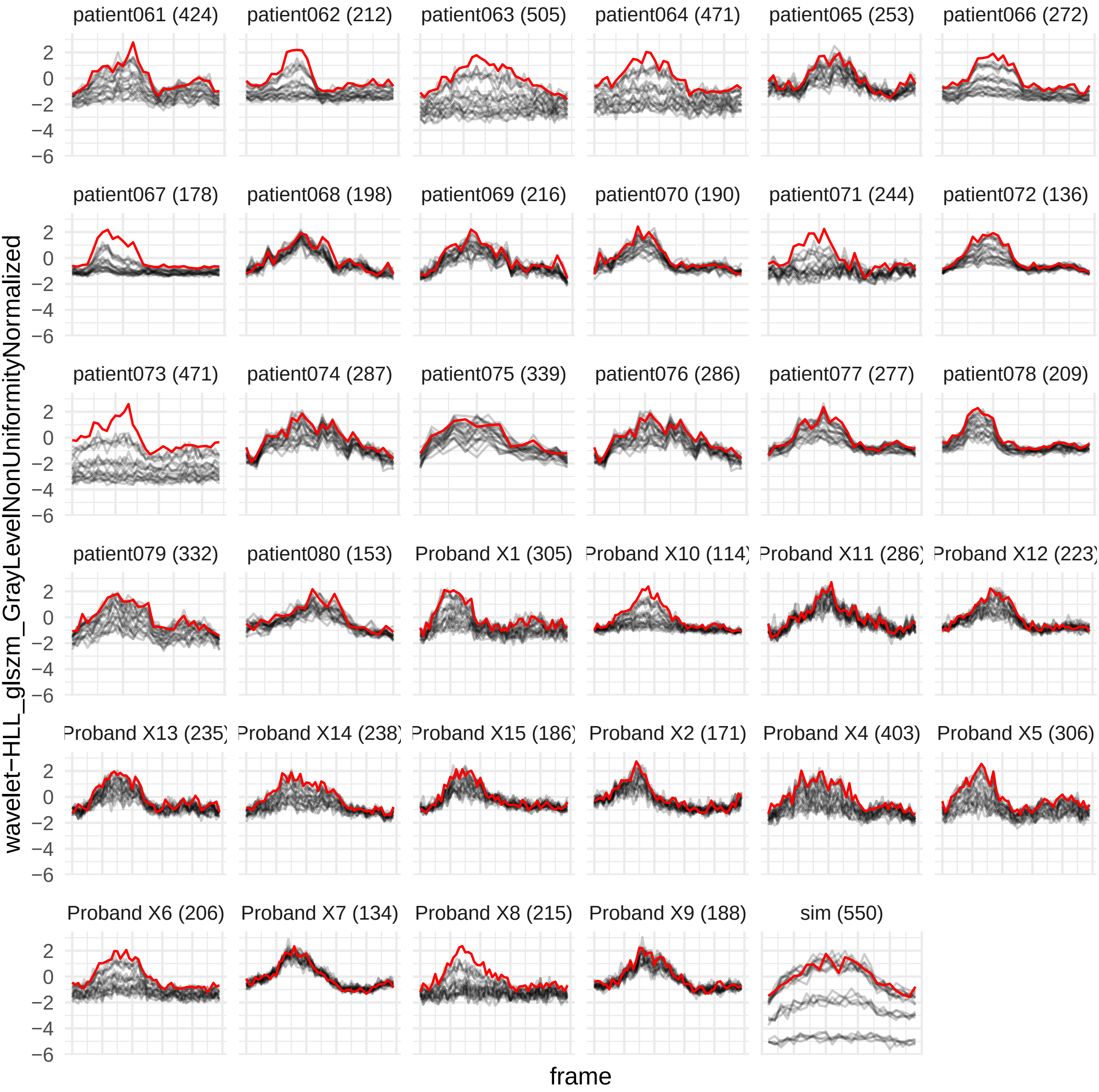
patient061 (424)
patient062 (212)
patient063 (505)
patient064 (471)
patient065 (253)
patient066 (272)
patient067 (178)
patient068 (198)
patient069 (216)
patient070 (190)
patient071 (244)
patient072 (136)
patient073 (471)
patient074 (287)
patient075 (339)
patient076 (286)
patient077 (277)
patient078 (209)
patient079 (332)
patient080 (153)
Proband X1 (305)
Proband X10 (114)
Proband X11 (286)
Proband X12 (223)
Proband X13 (235)
Proband X14 (238)
Proband X15 (186)
Proband X2 (171)
Proband X4 (403)
Proband X5 (306)
Proband X6 (206)
Proband X7 (134)
Proband X8 (215)
Proband X9 (188)
sim (550)
2
0
−2
−4
−6
wavelet−HLL_glszm_GrayLevelNonUniformityNormalized
frame

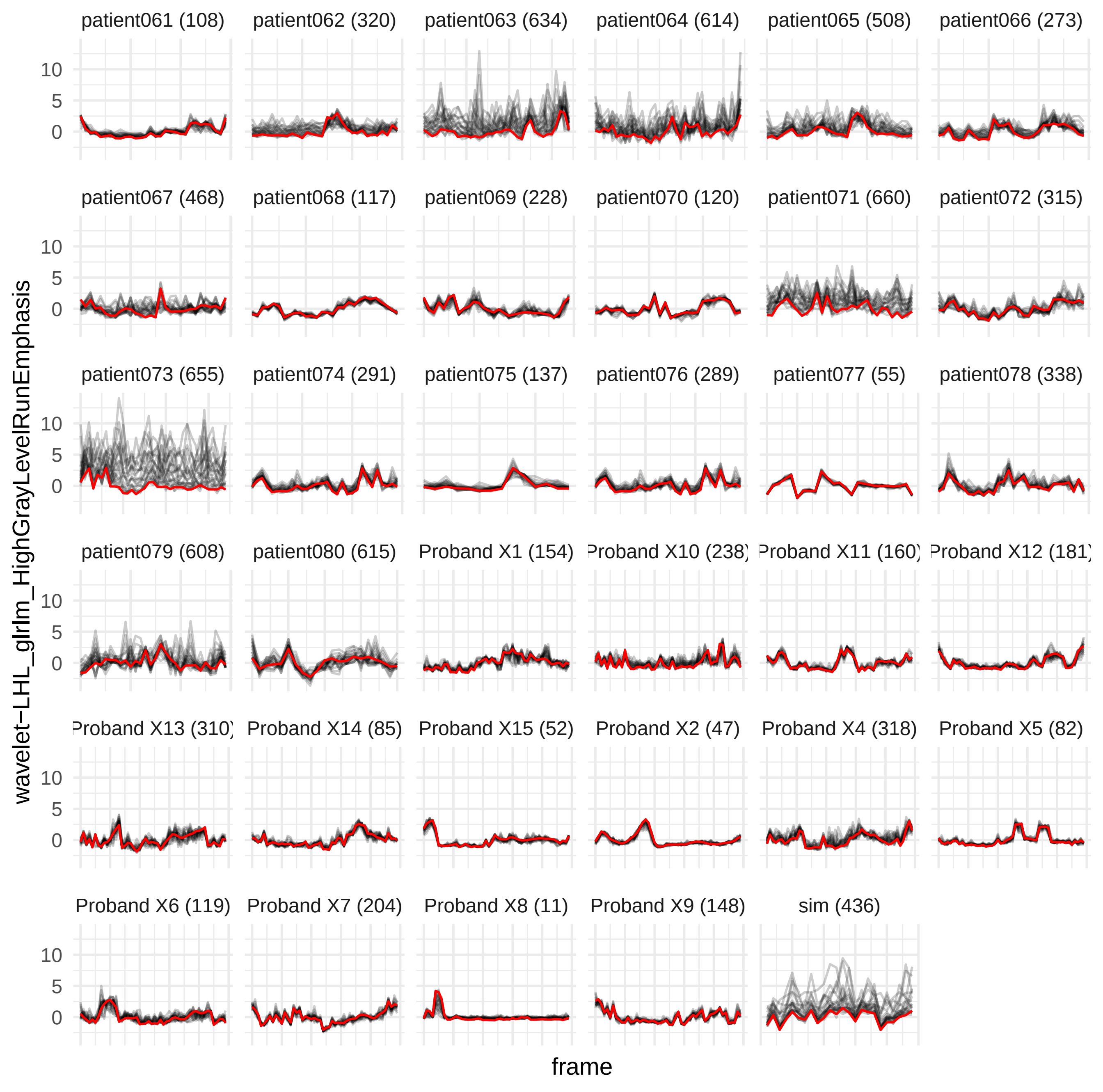

patient061 (108)
patient062 (320)
patient063 (634)
patient064 (614)
patient065 (508)
patient066 (273)
patient067 (468)
patient068 (117)
patient069 (228)
patient070 (120)
patient071 (660)
patient072 (315)
patient073 (655)
patient074 (291)
patient075 (137)
patient076 (289)
patient077 (55)
patient078 (338)
patient079 (608)
patient080 (615)
Proband X1 (154)
Proband X10 (238)
Proband X11 (160)
Proband X12 (181)
Proband X13 (310)
Proband X14 (85)
Proband X15 (52)
Proband X2 (47)
Proband X4 (318)
Proband X5 (82)
Proband X6 (119)
Proband X7 (204)
Proband X8 (11)
Proband X9 (148)
sim (436)
10
5
0
wavelet−LHL_glrlm_HighGrayLevelRunEmphasis
frame

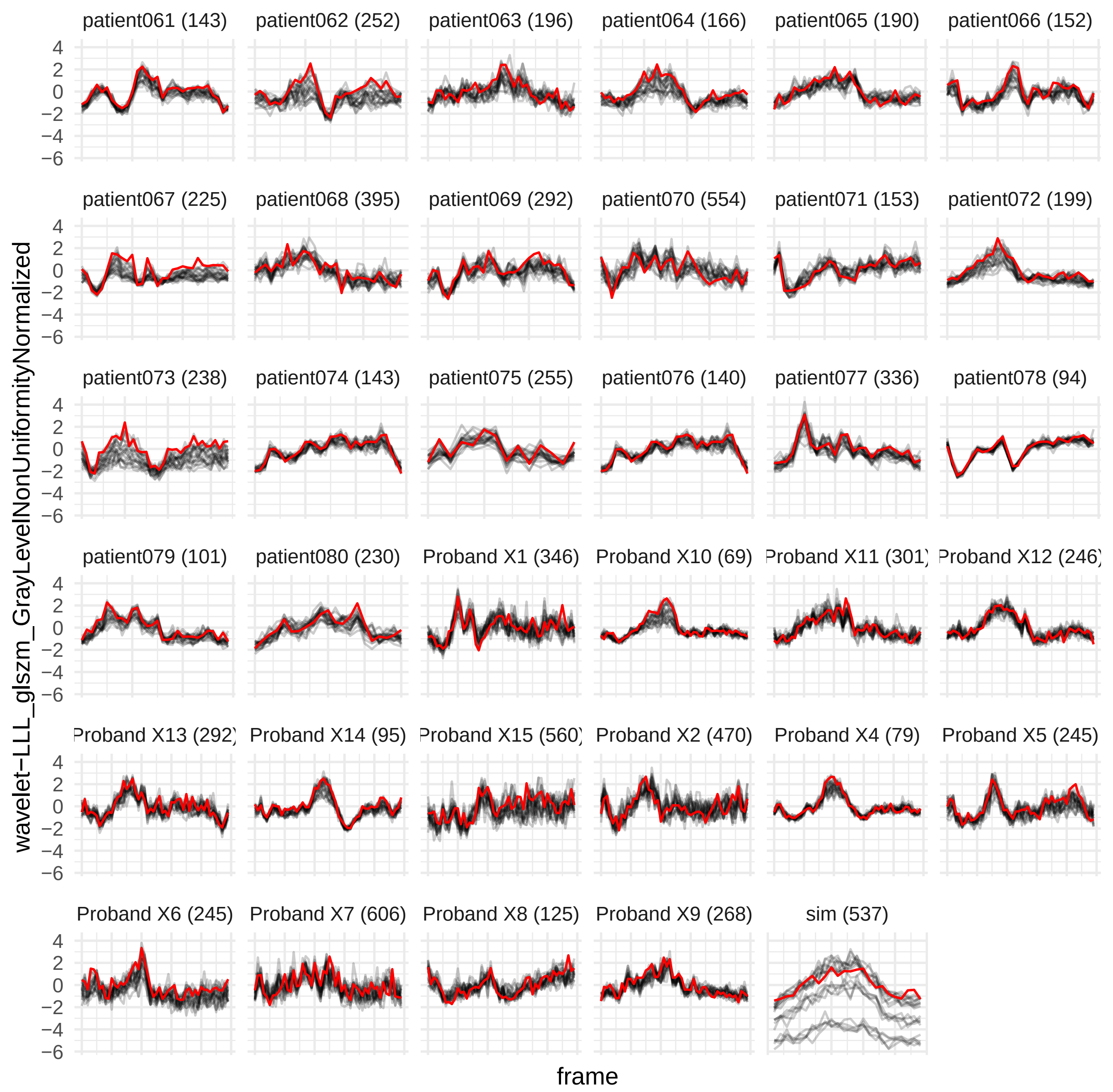

patient061 (143)
patient062 (252)
patient063 (196)
patient064 (166)
patient065 (190)
patient066 (152)
patient067 (225)
patient068 (395)
patient069 (292)
patient070 (554)
patient071 (153)
patient072 (199)
patient073 (238)
patient074 (143)
patient075 (255)
patient076 (140)
patient077 (336)
patient078 (94)
patient079 (101)
patient080 (230)
Proband X1 (346)
Proband X10 (69)
Proband X11 (301)
Proband X12 (246)
Proband X13 (292)
Proband X14 (95)
Proband X15 (560)
Proband X2 (470)
Proband X4 (79)
Proband X5 (245)
Proband X6 (245)
Proband X7 (606)
Proband X8 (125)
Proband X9 (268)
sim (537)
4
2
0
−2
−4
−6
wavelet−LLL_glszm_GrayLevelNonUniformityNormalized
frame

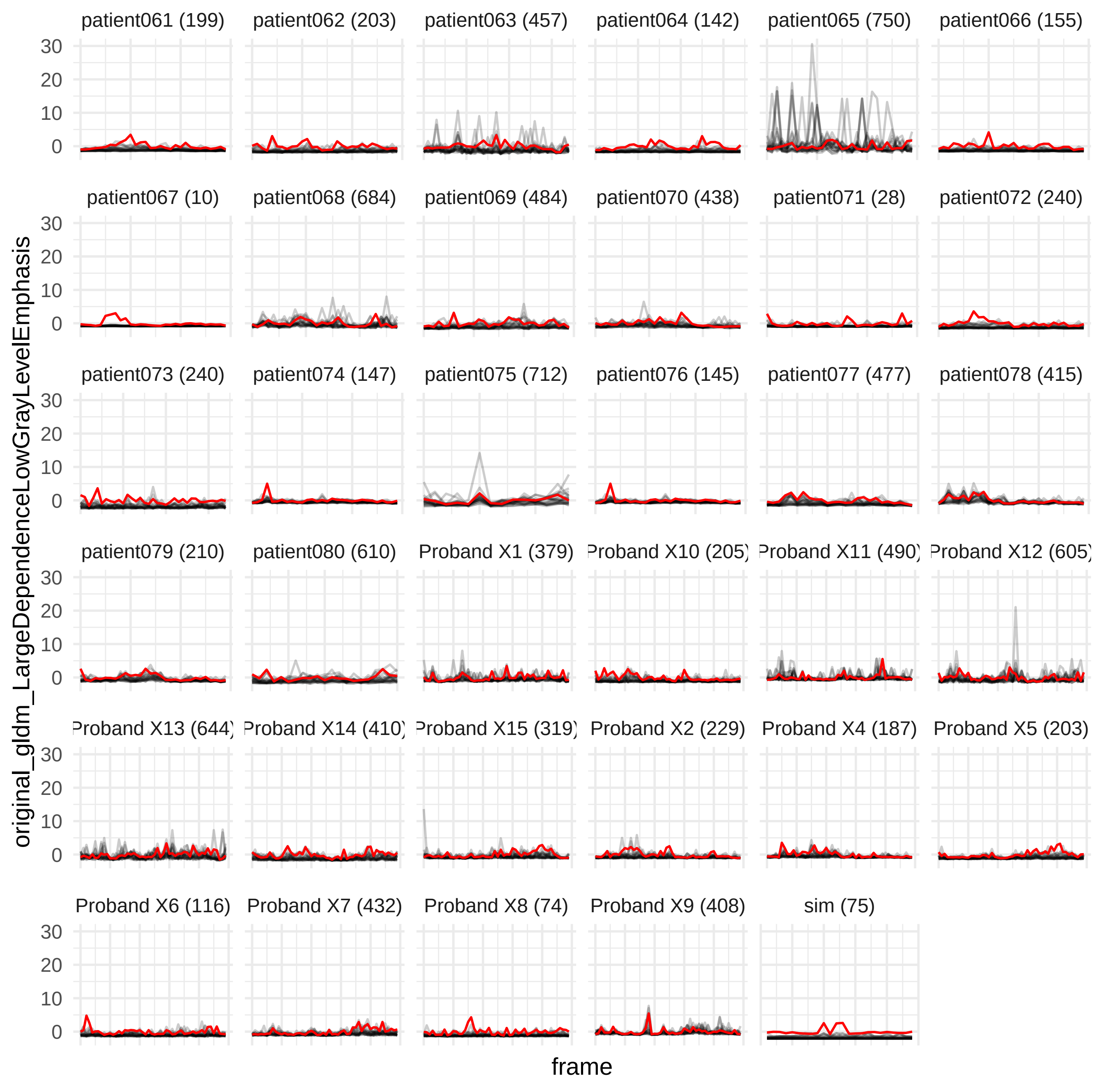
patient061 (199)
patient062 (203)
patient063 (457)
patient064 (142)
patient065 (750)
patient066 (155)
patient067 (10)
patient068 (684)
patient069 (484)
patient070 (438)
patient071 (28)
patient072 (240)
patient073 (240)
patient074 (147)
patient075 (712)
patient076 (145)
patient077 (477)
patient078 (415)
patient079 (210)
patient080 (610)
Proband X1 (379)
Proband X10 (205)
Proband X11 (490)
Proband X12 (605)
Proband X13 (644)
Proband X14 (410)
Proband X15 (319)
Proband X2 (229)
Proband X4 (187)
Proband X5 (203)
Proband X6 (116)
Proband X7 (432)
Proband X8 (74)
Proband X9 (408)
sim (75)
original_gldm_LargeDependenceLowGrayLevelEmphasis
frame
0
10
20
30

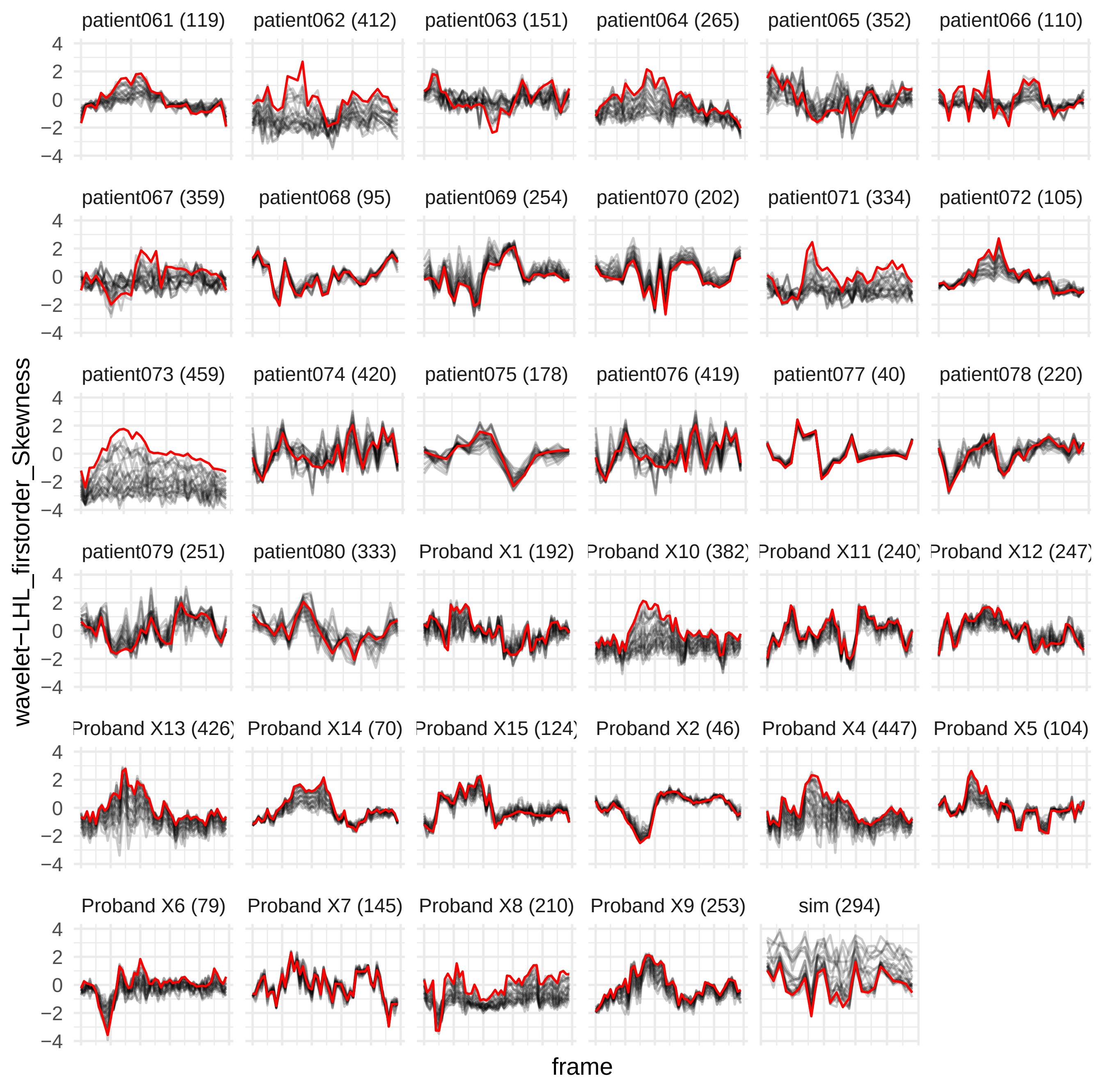

patient061 (119)
patient062 (412)
patient063 (151)
patient064 (265)
patient065 (352)
patient066 (110)
patient067 (359)
patient068 (95)
patient069 (254)
patient070 (202)
patient071 (334)
patient072 (105)
patient073 (459)
patient074 (420)
patient075 (178)
patient076 (419)
patient077 (40)
patient078 (220)
patient079 (251)
patient080 (333)
Proband X1 (192)
Proband X10 (382)
Proband X11 (240)
Proband X12 (247)
Proband X13 (426)
Proband X14 (70)
Proband X15 (124)
Proband X2 (46)
Proband X4 (447)
Proband X5 (104)
Proband X6 (79)
Proband X7 (145)
Proband X8 (210)
Proband X9 (253)
sim (294)
wavelet−LHL_firstorder_Skewness
frame

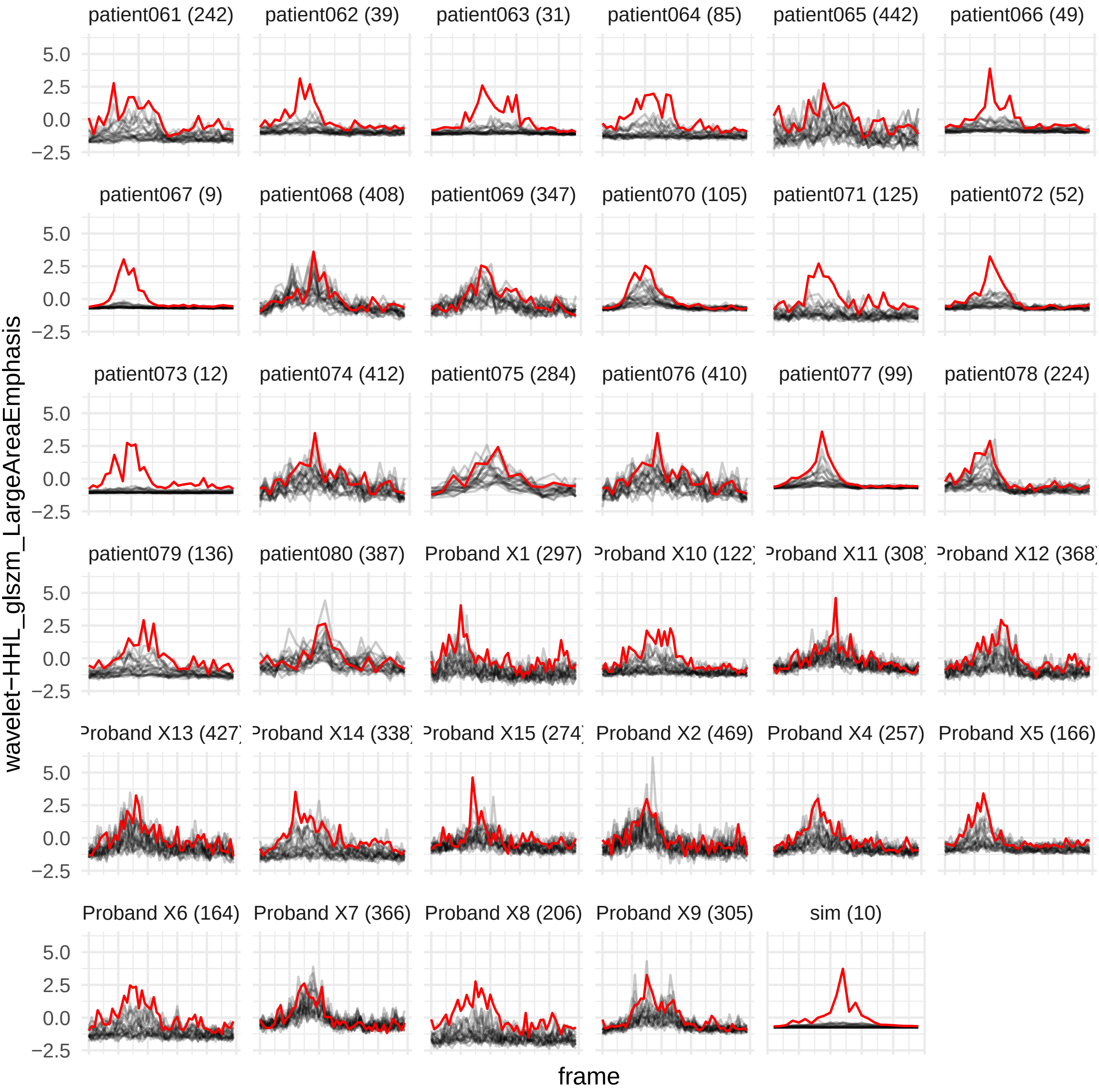
patient061 (242)
patient062 (39)
patient063 (31)
patient064 (85)
patient065 (442)
patient066 (49)
patient067 (9)
patient068 (408)
patient069 (347)
patient070 (105)
patient071 (125)
patient072 (52)
patient073 (12)
patient074 (412)
patient075 (284)
patient076 (410)
patient077 (99)
patient078 (224)
patient079 (136)
patient080 (387)
Proband X1 (297)
Proband X10 (122)
Proband X11 (308)
Proband X12 (368)
Proband X13 (427)
Proband X14 (338)
Proband X15 (274)
Proband X2 (469)
Proband X4 (257)
Proband X5 (166)
Proband X6 (164)
Proband X7 (366)
Proband X8 (206)
Proband X9 (305)
sim (10)
5.0
2.5
0.0
−2.5
wavelet−HHL_glszm_LargeAreaEmphasis
frame

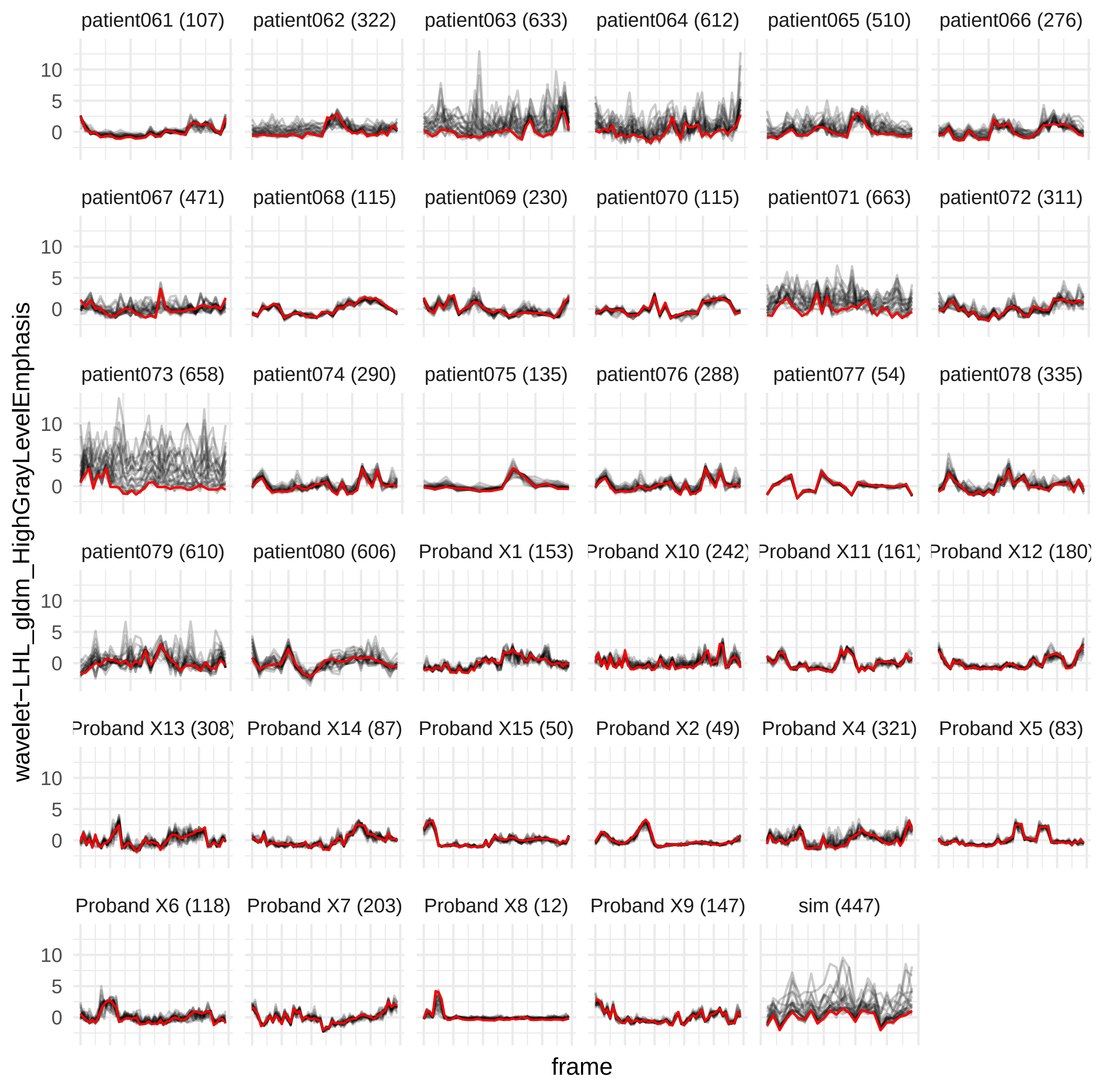

patient061 (107)
patient062 (322)
patient063 (633)
patient064 (612)
patient065 (510)
patient066 (276)
patient067 (471)
patient068 (115)
patient069 (230)
patient070 (115)
patient071 (663)
patient072 (311)
patient073 (658)
patient074 (290)
patient075 (135)
patient076 (288)
patient077 (54)
patient078 (335)
patient079 (610)
patient080 (606)
Proband X1 (153)
Proband X10 (242)
Proband X11 (161)
Proband X12 (180)
Proband X13 (308)
Proband X14 (87)
Proband X15 (50)
Proband X2 (49)
Proband X4 (321)
Proband X5 (83)
Proband X6 (118)
Proband X7 (203)
Proband X8 (12)
Proband X9 (147)
sim (447)
wavelet−LHL_gldm_HighGrayLevelEmphasis
frame
10
5
0

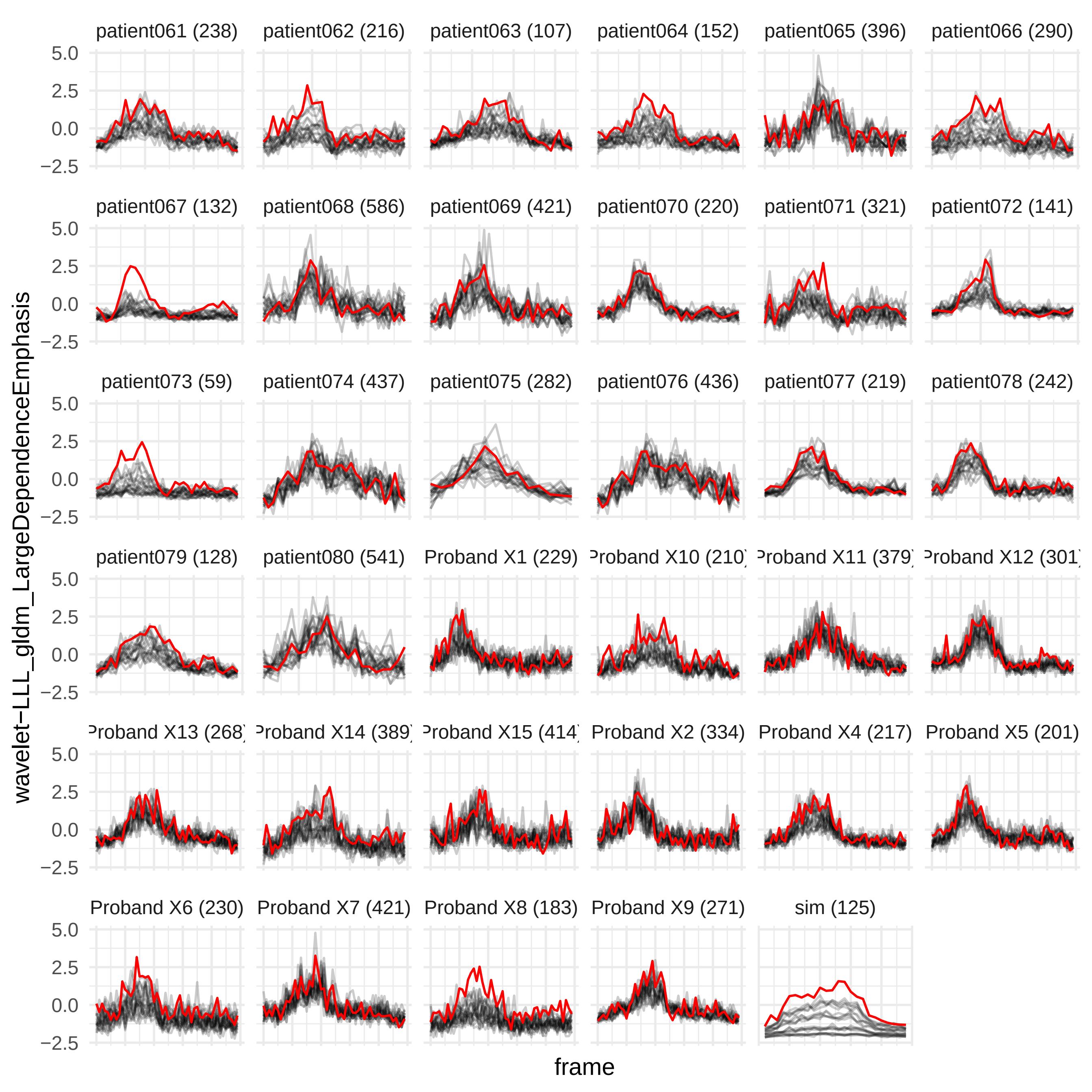

patient061 (238)
patient062 (216)
patient063 (107)
patient064 (152)
patient065 (396)
patient066 (290)
patient067 (132)
patient068 (586)
patient069 (421)
patient070 (220)
patient071 (321)
patient072 (141)
patient073 (59)
patient074 (437)
patient075 (282)
patient076 (436)
patient077 (219)
patient078 (242)
patient079 (128)
patient080 (541)
Proband X1 (229)
Proband X10 (210)
Proband X11 (379)
Proband X12 (301)
Proband X13 (268)
Proband X14 (389)
Proband X15 (414)
Proband X2 (334)
Proband X4 (217)
Proband X5 (201)
Proband X6 (230)
Proband X7 (421)
Proband X8 (183)
Proband X9 (271)
sim (125)
5.0
2.5
0.0
−2.5
wavelet−LLL_gldm_LargeDependenceEmphasis
frame

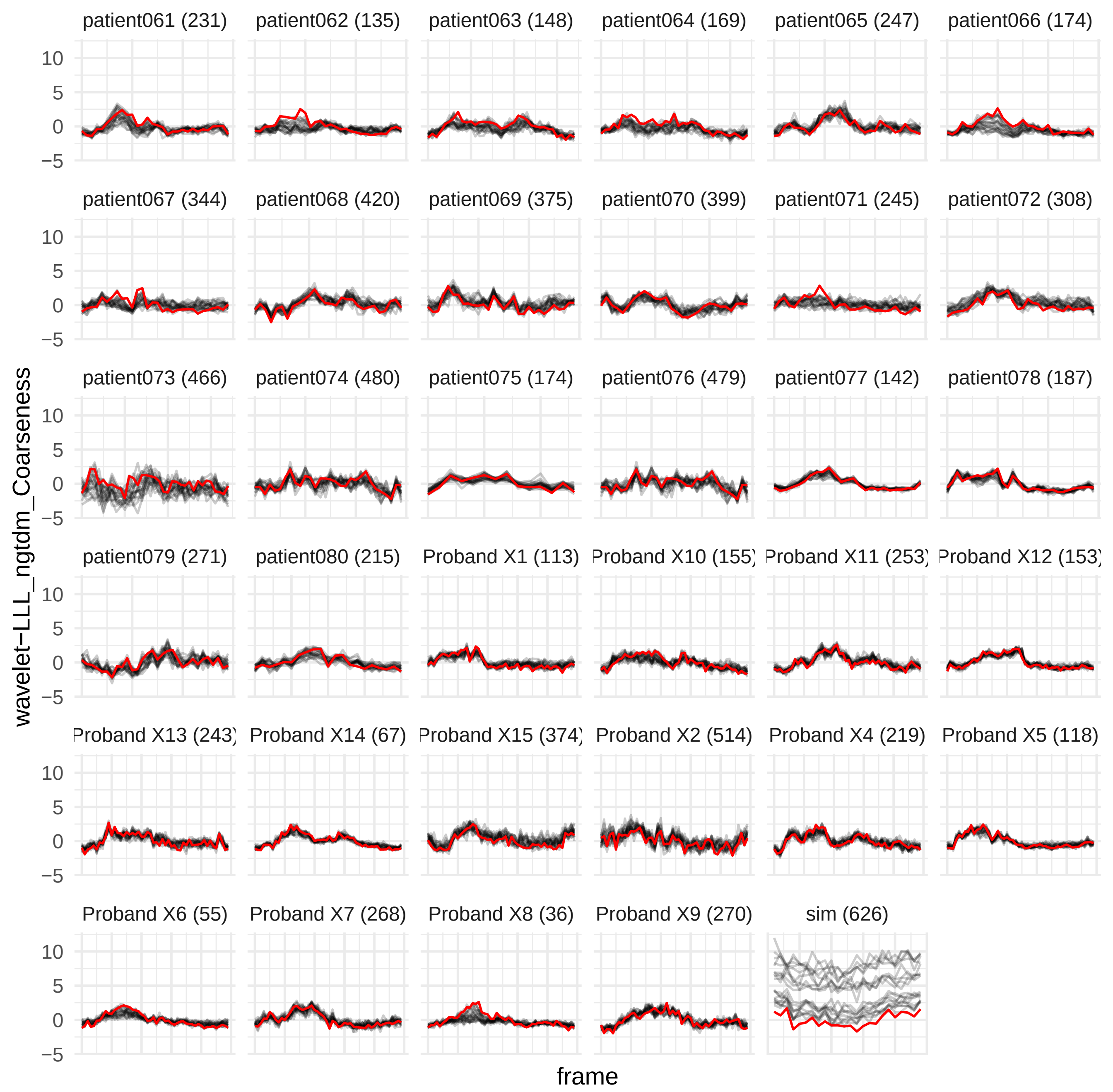
patient061 (231)
patient062 (135)
patient063 (148)
patient064 (169)
patient065 (247)
patient066 (174)
patient067 (344)
patient068 (420)
patient069 (375)
patient070 (399)
patient071 (245)
patient072 (308)
patient073 (466)
patient074 (480)
patient075 (174)
patient076 (479)
patient077 (142)
patient078 (187)
patient079 (271)
patient080 (215)
Proband X1 (113)
Proband X10 (155)
Proband X11 (253)
Proband X12 (153)
Proband X13 (243)
Proband X14 (67)
Proband X15 (374)
Proband X2 (514)
Proband X4 (219)
Proband X5 (118)
Proband X6 (55)
Proband X7 (268)
Proband X8 (36)
Proband X9 (270)
sim (626)
wavelet−LLL_ngtdm_Coarseness
frame
10
5
0
−5

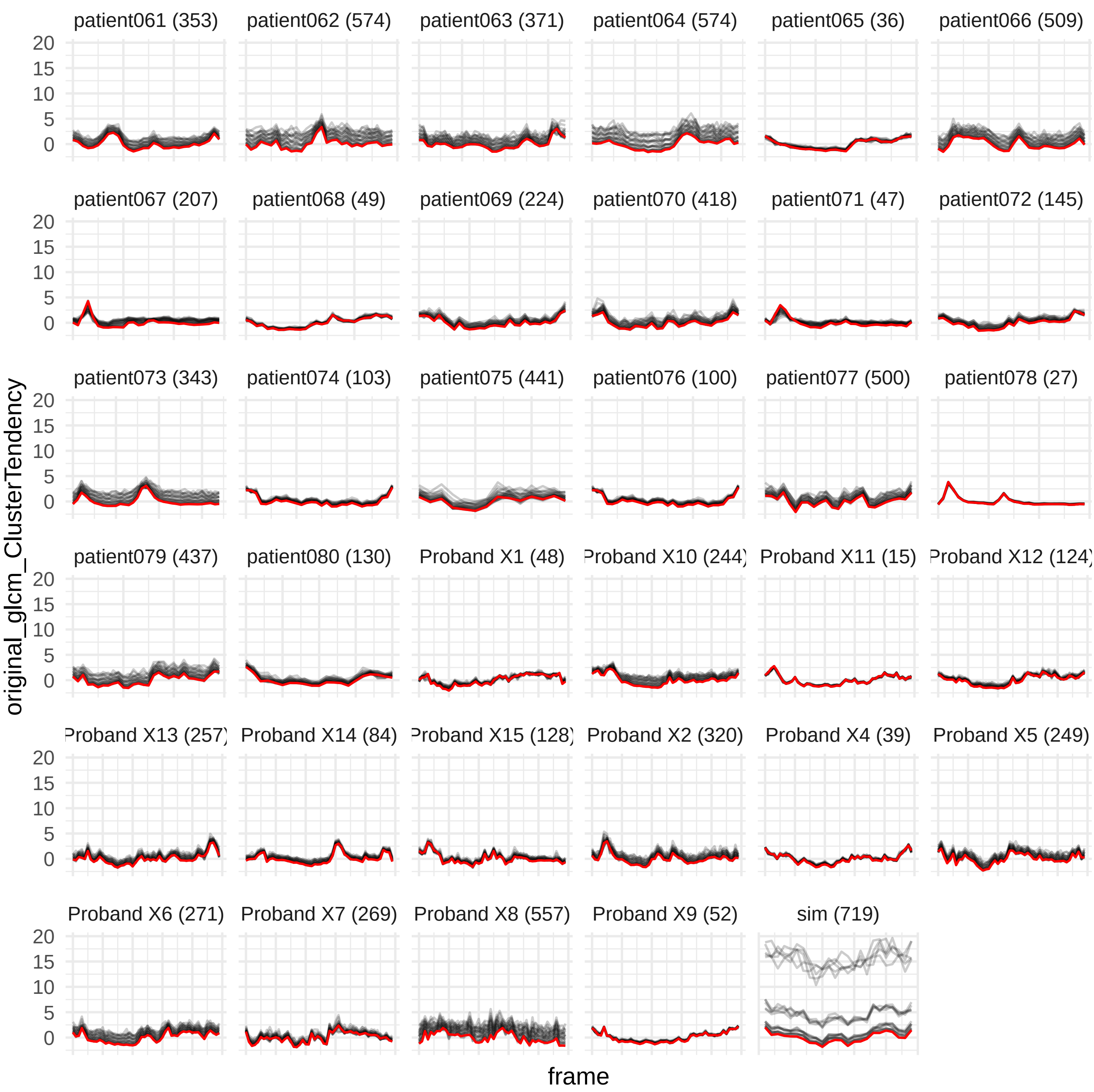
patient061 (353)
patient062 (574)
patient063 (371)
patient064 (574)
patient065 (36)
patient066 (509)
patient067 (207)
patient068 (49)
patient069 (224)
patient070 (418)
patient071 (47)
patient072 (145)
patient073 (343)
patient074 (103)
patient075 (441)
patient076 (100)
patient077 (500)
patient078 (27)
patient079 (437)
patient080 (130)
Proband X1 (48)
Proband X10 (244)
Proband X11 (15)
Proband X12 (124)
Proband X13 (257)
Proband X14 (84)
Proband X15 (128)
Proband X2 (320)
Proband X4 (39)
Proband X5 (249)
Proband X6 (271)
Proband X7 (269)
Proband X8 (557)
Proband X9 (52)
sim (719)
original_glcm_ClusterTendency
frame
0
5
10
15
20

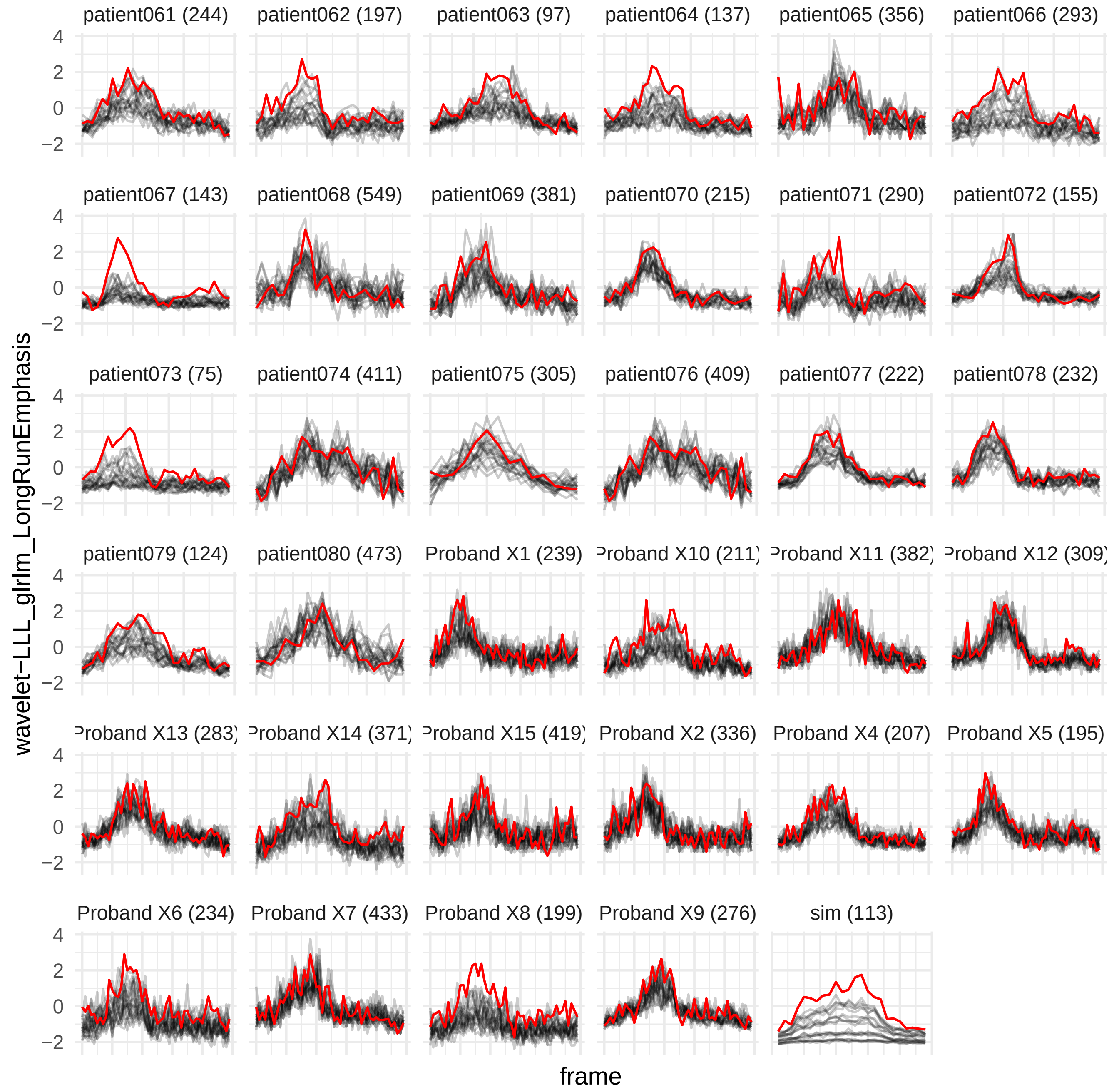

patient061 (244)
patient062 (197)
patient063 (97)
patient064 (137)
patient065 (356)
patient066 (293)
patient067 (143)
patient068 (549)
patient069 (381)
patient070 (215)
patient071 (290)
patient072 (155)
patient073 (75)
patient074 (411)
patient075 (305)
patient076 (409)
patient077 (222)
patient078 (232)
patient079 (124)
patient080 (473)
Proband X1 (239)
Proband X10 (211)
Proband X11 (382)
Proband X12 (309)
Proband X13 (283)
Proband X14 (371)
Proband X15 (419)
Proband X2 (336)
Proband X4 (207)
Proband X5 (195)
Proband X6 (234)
Proband X7 (433)
Proband X8 (199)
Proband X9 (276)
sim (113)
wavelet−LLL_glrlm_LongRunEmphasis
frame

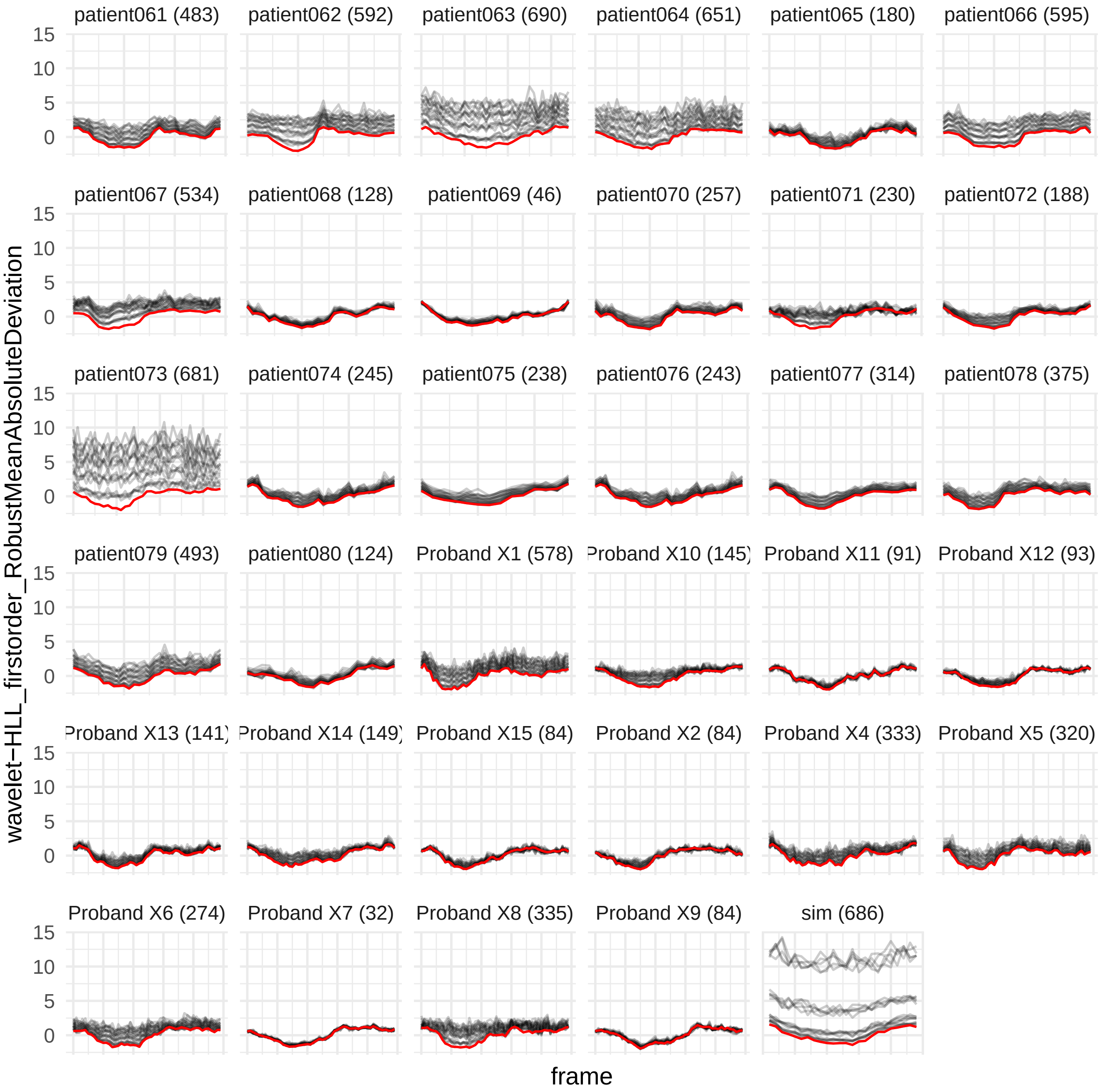

patient061 (483)
patient062 (592)
patient063 (690)
patient064 (651)
patient065 (180)
patient066 (595)
patient067 (534)
patient068 (128)
patient069 (46)
patient070 (257)
patient071 (230)
patient072 (188)
patient073 (681)
patient074 (245)
patient075 (238)
patient076 (243)
patient077 (314)
patient078 (375)
patient079 (493)
patient080 (124)
Proband X1 (578)
Proband X10 (145)
Proband X11 (91)
Proband X12 (93)
Proband X13 (141)
Proband X14 (149)
Proband X15 (84)
Proband X2 (84)
Proband X4 (333)
Proband X5 (320)
Proband X6 (274)
Proband X7 (32)
Proband X8 (335)
Proband X9 (84)
sim (686)
15
10
5
0
wavelet−HLL_firstorder_RobustMeanAbsoluteDeviation
frame

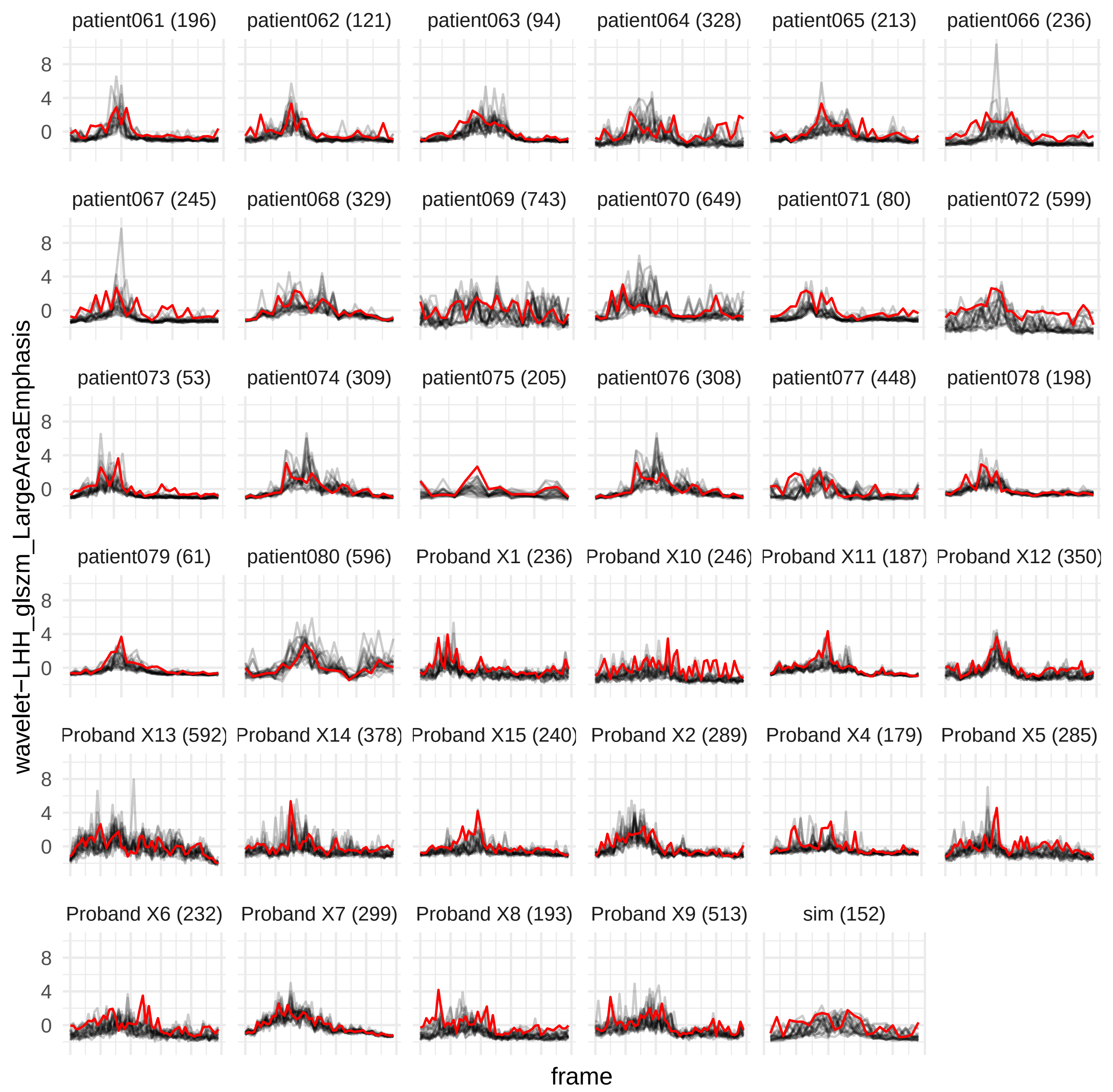
patient061 (196)
patient062 (121)
patient063 (94)
patient064 (328)
patient065 (213)
patient066 (236)
patient067 (245)
patient068 (329)
patient069 (743)
patient070 (649)
patient071 (80)
patient072 (599)
patient073 (53)
patient074 (309)
patient075 (205)
patient076 (308)
patient077 (448)
patient078 (198)
patient079 (61)
patient080 (596)
Proband X1 (236)
Proband X10 (246)
Proband X11 (187)
Proband X12 (350)
Proband X13 (592)
Proband X14 (378)
Proband X15 (240)
Proband X2 (289)
Proband X4 (179)
Proband X5 (285)
Proband X6 (232)
Proband X7 (299)
Proband X8 (193)
Proband X9 (513)
sim (152)
wavelet−LHH_glszm_LargeAreaEmphasis
8
4
0
frame

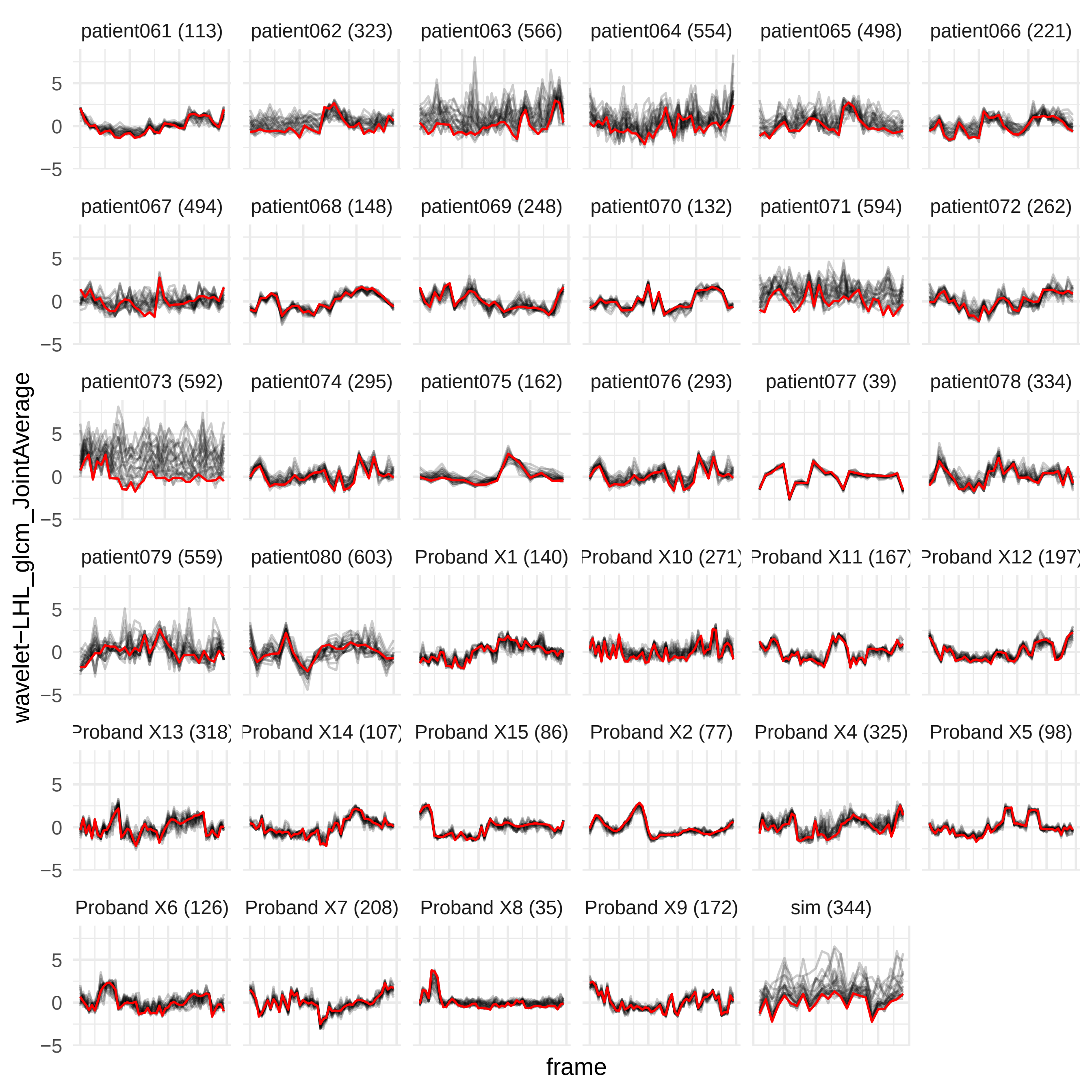
patient061 (113)
patient062 (323)
patient063 (566)
patient064 (554)
patient065 (498)
patient066 (221)
patient067 (494)
patient068 (148)
patient069 (248)
patient070 (132)
patient071 (594)
patient072 (262)
patient073 (592)
patient074 (295)
patient075 (162)
patient076 (293)
patient077 (39)
patient078 (334)
patient079 (559)
patient080 (603)
Proband X1 (140)
Proband X10 (271)
Proband X11 (167)
Proband X12 (197)
Proband X13 (318)
Proband X14 (107)
Proband X15 (86)
Proband X2 (77)
Proband X4 (325)
Proband X5 (98)
Proband X6 (126)
Proband X7 (208)
Proband X8 (35)
Proband X9 (172)
sim (344)
5
0
−5
wavelet−LHL_glcm_JointAverage
frame

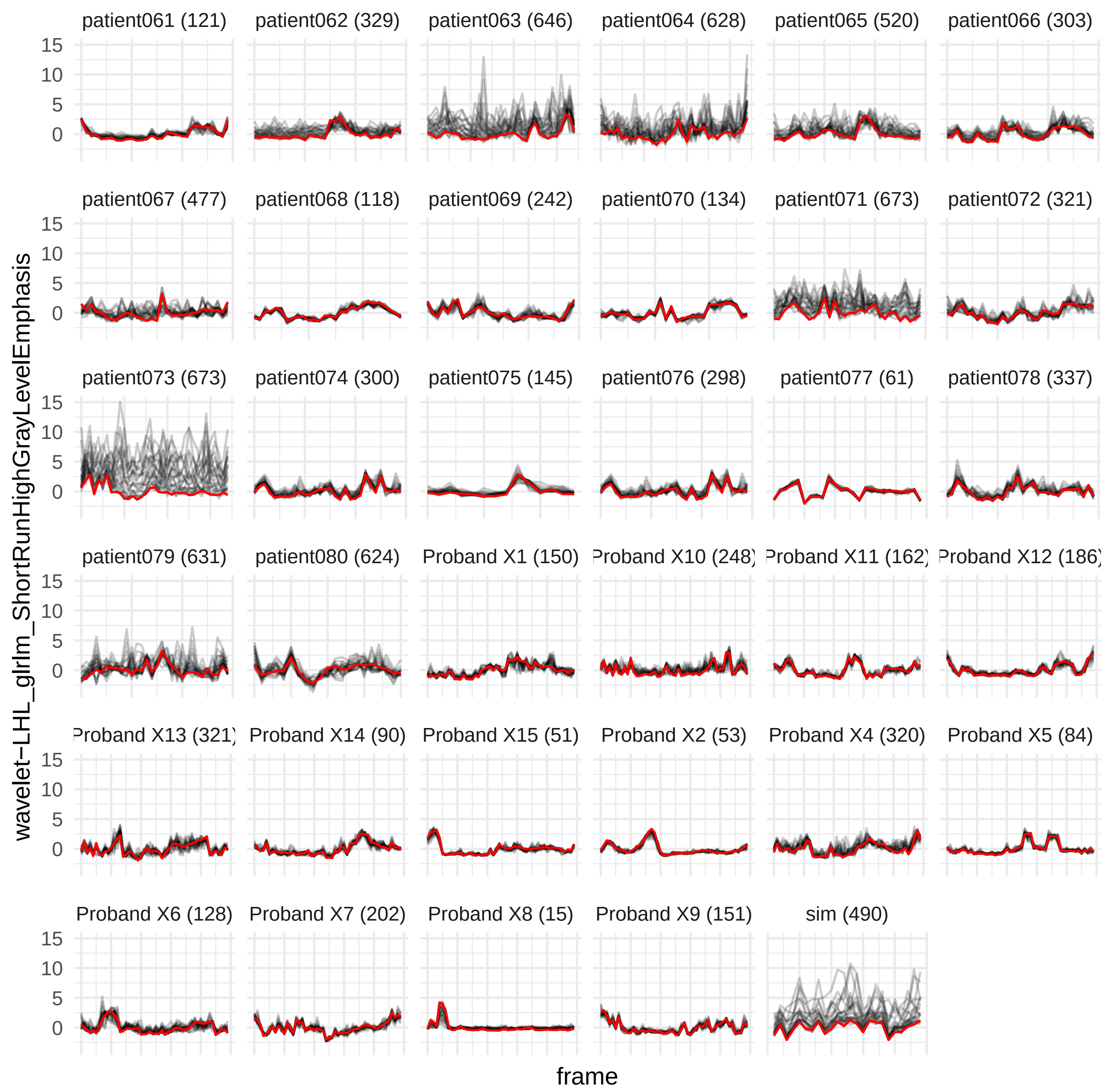
patient061 (121)
patient062 (329)
patient063 (646)
patient064 (628)
patient065 (520)
patient066 (303)
patient067 (477)
patient068 (118)
patient069 (242)
patient070 (134)
patient071 (673)
patient072 (321)
patient073 (673)
patient074 (300)
patient075 (145)
patient076 (298)
patient077 (61)
patient078 (337)
patient079 (631)
patient080 (624)
Proband X1 (150)
Proband X10 (248)
Proband X11 (162)
Proband X12 (186)
Proband X13 (321)
Proband X14 (90)
Proband X15 (51)
Proband X2 (53)
Proband X4 (320)
Proband X5 (84)
Proband X6 (128)
Proband X7 (202)
Proband X8 (15)
Proband X9 (151)
sim (490)
wavelet−LHL_glrlm_ShortRunHighGrayLevelEmphasis
frame

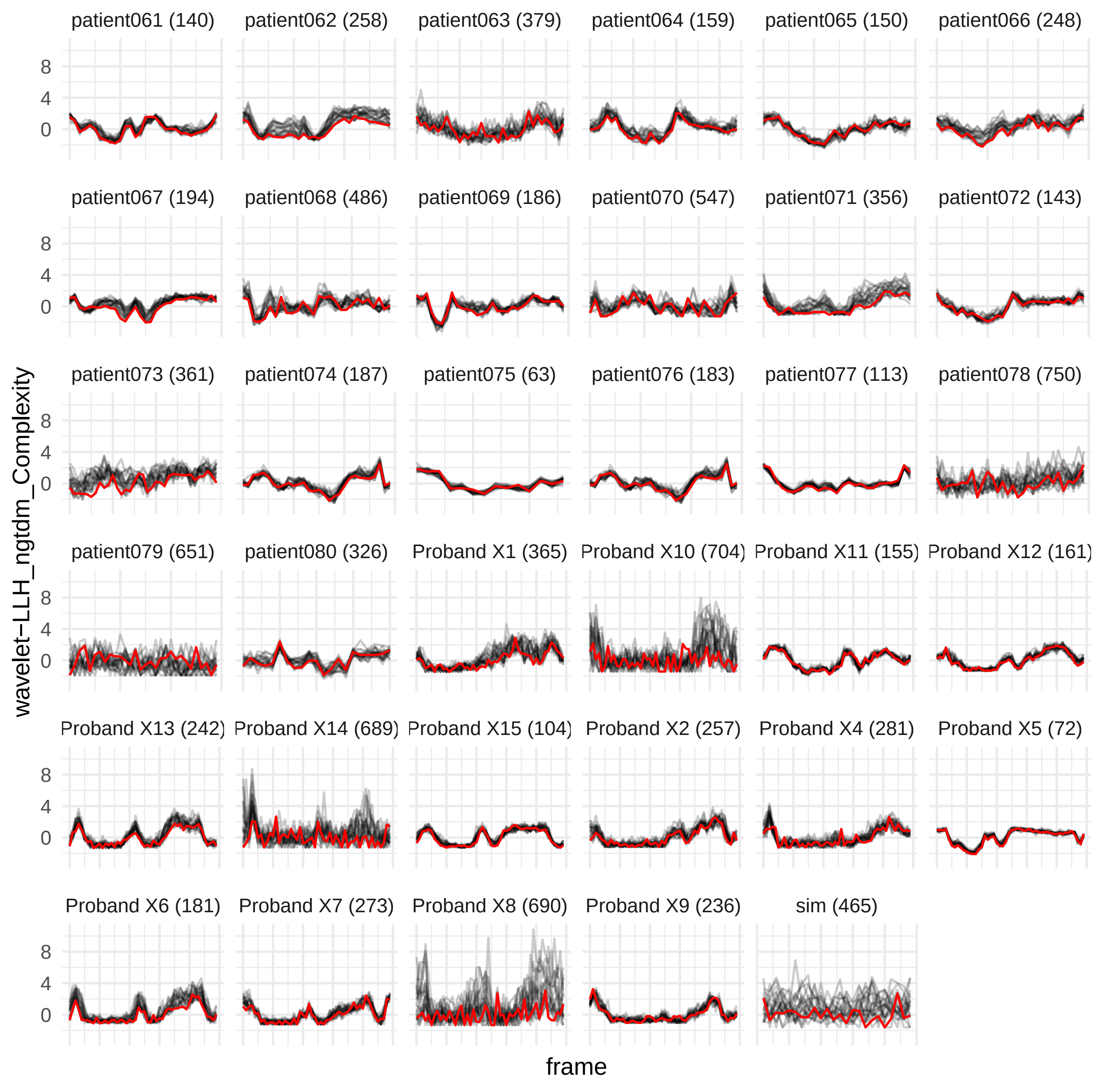
patient061 (140)
patient062 (258)
patient063 (379)
patient064 (159)
patient065 (150)
patient066 (248)
patient067 (194)
patient068 (486)
patient069 (186)
patient070 (547)
patient071 (356)
patient072 (143)
patient073 (361)
patient074 (187)
patient075 (63)
patient076 (183)
patient077 (113)
patient078 (750)
patient079 (651)
patient080 (326)
Proband X1 (365)
Proband X10 (704)
Proband X11 (155)
Proband X12 (161)
Proband X13 (242)
Proband X14 (689)
Proband X15 (104)
Proband X2 (257)
Proband X4 (281)
Proband X5 (72)
Proband X6 (181)
Proband X7 (273)
Proband X8 (690)
Proband X9 (236)
sim (465)
8
4
0
wavelet−LLH_ngtdm_Complexity
frame

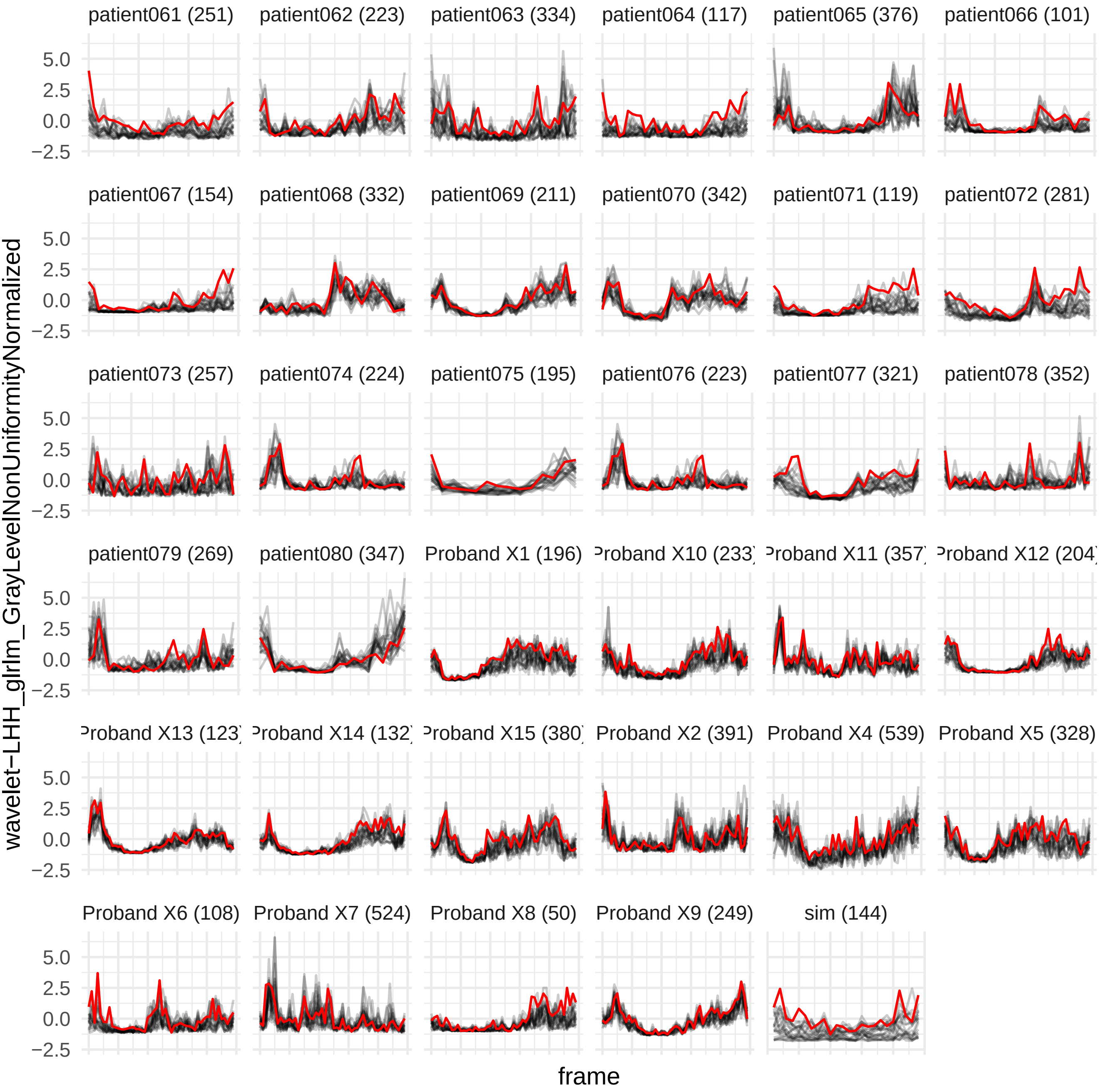

patient061 (251)
patient062 (223)
patient063 (334)
patient064 (117)
patient065 (376)
patient066 (101)
patient067 (154)
patient068 (332)
patient069 (211)
patient070 (342)
patient071 (119)
patient072 (281)
patient073 (257)
patient074 (224)
patient075 (195)
patient076 (223)
patient077 (321)
patient078 (352)
patient079 (269)
patient080 (347)
Proband X1 (196)
Proband X10 (233)
Proband X11 (357)
Proband X12 (204)
Proband X13 (123)
Proband X14 (132)
Proband X15 (380)
Proband X2 (391)
Proband X4 (539)
Proband X5 (328)
Proband X6 (108)
Proband X7 (524)
Proband X8 (50)
Proband X9 (249)
sim (144)
5.0
2.5
0.0
−2.5
wavelet−LHH_glrlm_GrayLevelNonUniformityNormalized
frame

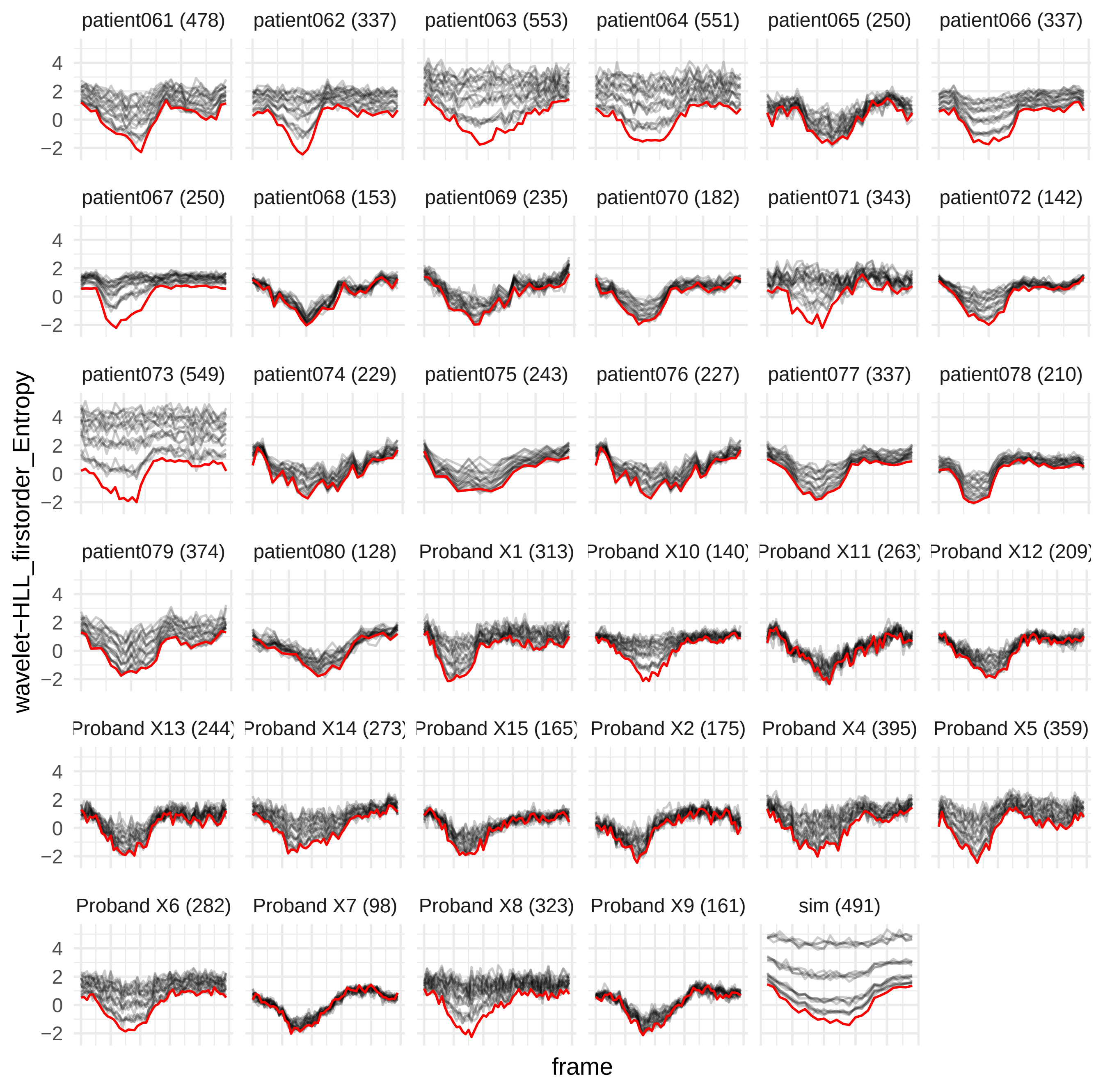

patient061 (478)
patient062 (337)
patient063 (553)
patient064 (551)
patient065 (250)
patient066 (337)
patient067 (250)
patient068 (153)
patient069 (235)
patient070 (182)
patient071 (343)
patient072 (142)
patient073 (549)
patient074 (229)
patient075 (243)
patient076 (227)
patient077 (337)
patient078 (210)
patient079 (374)
patient080 (128)
Proband X1 (313)
Proband X10 (140)
Proband X11 (263)
Proband X12 (209)
Proband X13 (244)
Proband X14 (273)
Proband X15 (165)
Proband X2 (175)
Proband X4 (395)
Proband X5 (359)
Proband X6 (282)
Proband X7 (98)
Proband X8 (323)
Proband X9 (161)
sim (491)
wavelet−HLL_firstorder_Entropy
4
2
0
−2
frame

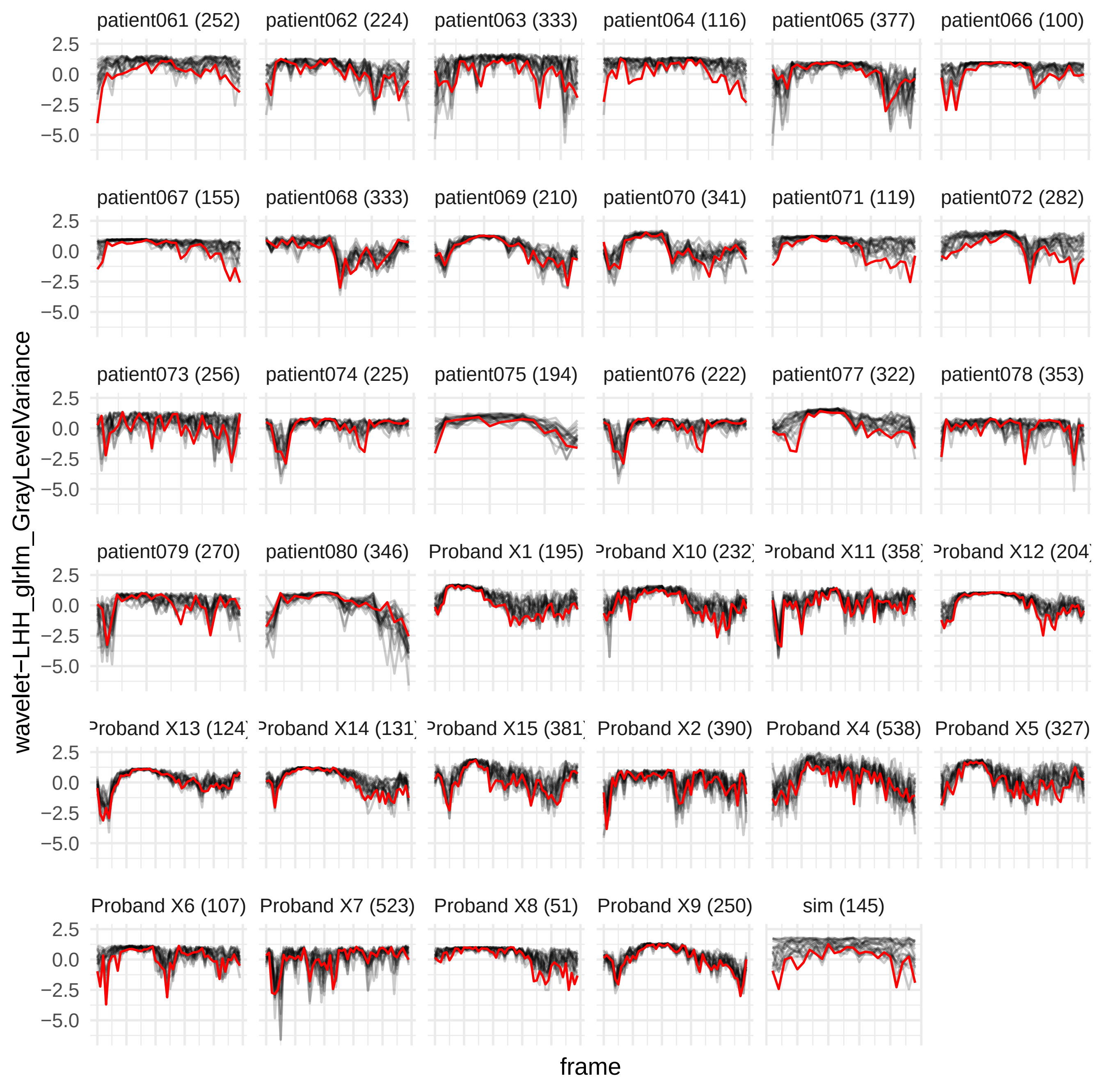

patient061 (252)
patient062 (224)
patient063 (333)
patient064 (116)
patient065 (377)
patient066 (100)
patient067 (155)
patient068 (333)
patient069 (210)
patient070 (341)
patient071 (119)
patient072 (282)
patient073 (256)
patient074 (225)
patient075 (194)
patient076 (222)
patient077 (322)
patient078 (353)
patient079 (270)
patient080 (346)
Proband X1 (195)
Proband X10 (232)
Proband X11 (358)
Proband X12 (204)
Proband X13 (124)
Proband X14 (131)
Proband X15 (381)
Proband X2 (390)
Proband X4 (538)
Proband X5 (327)
Proband X6 (107)
Proband X7 (523)
Proband X8 (51)
Proband X9 (250)
sim (145)
2.5
0.0
−2.5
−5.0
wavelet−LHH_glrlm_GrayLevelVariance
frame

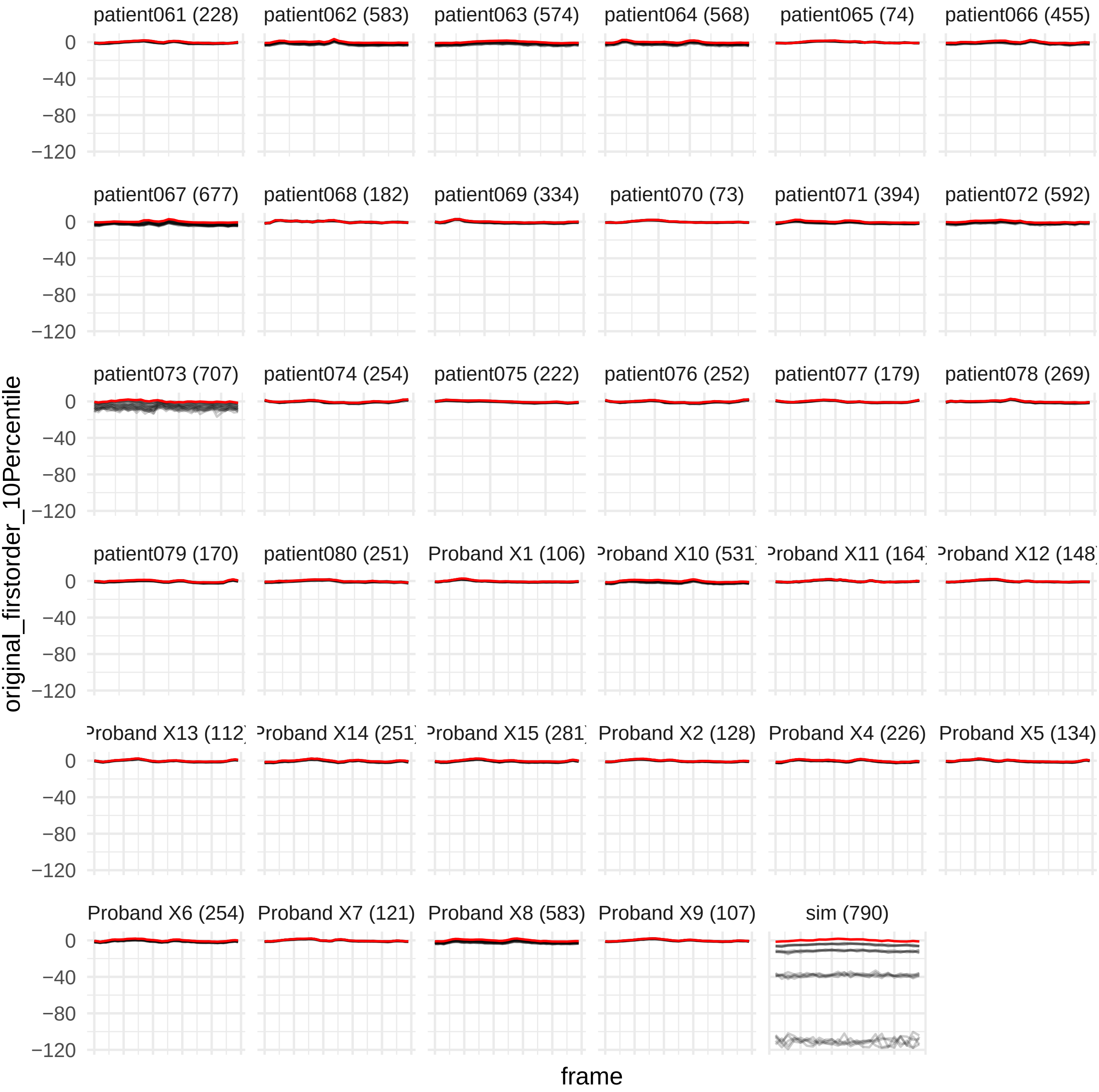

patient061 (228)
patient062 (583)
patient063 (574)
patient064 (568)
patient065 (74)
patient066 (455)
patient067 (677)
patient068 (182)
patient069 (334)
patient070 (73)
patient071 (394)
patient072 (592)
patient073 (707)
patient074 (254)
patient075 (222)
patient076 (252)
patient077 (179)
patient078 (269)
patient079 (170)
patient080 (251)
Proband X1 (106)
Proband X10 (531)
Proband X11 (164)
Proband X12 (148)
Proband X13 (112)
Proband X14 (251)
Proband X15 (281)
Proband X2 (128)
Proband X4 (226)
Proband X5 (134)
Proband X6 (254)
Proband X7 (121)
Proband X8 (583)
Proband X9 (107)
sim (790)
0
−40
−80
−120
original_firstorder_10Percentile
frame

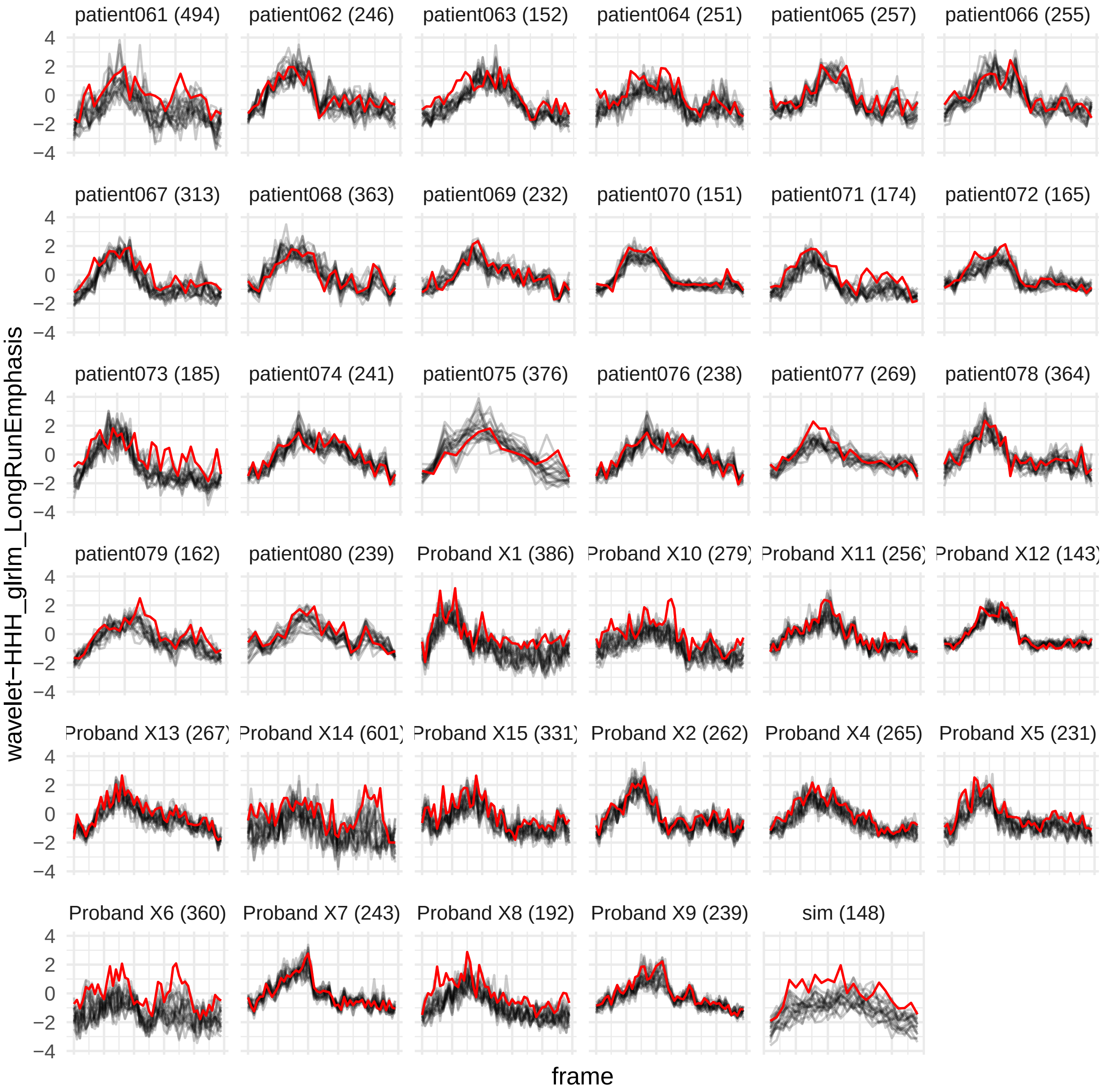

patient061 (494)
patient062 (246)
patient063 (152)
patient064 (251)
patient065 (257)
patient066 (255)
patient067 (313)
patient068 (363)
patient069 (232)
patient070 (151)
patient071 (174)
patient072 (165)
patient073 (185)
patient074 (241)
patient075 (376)
patient076 (238)
patient077 (269)
patient078 (364)
patient079 (162)
patient080 (239)
Proband X1 (386)
Proband X10 (279)
Proband X11 (256)
Proband X12 (143)
Proband X13 (267)
Proband X14 (601)
Proband X15 (331)
Proband X2 (262)
Proband X4 (265)
Proband X5 (231)
Proband X6 (360)
Proband X7 (243)
Proband X8 (192)
Proband X9 (239)
sim (148)
wavelet−HHH_glrlm_LongRunEmphasis
frame
4
2
0
−2
−4

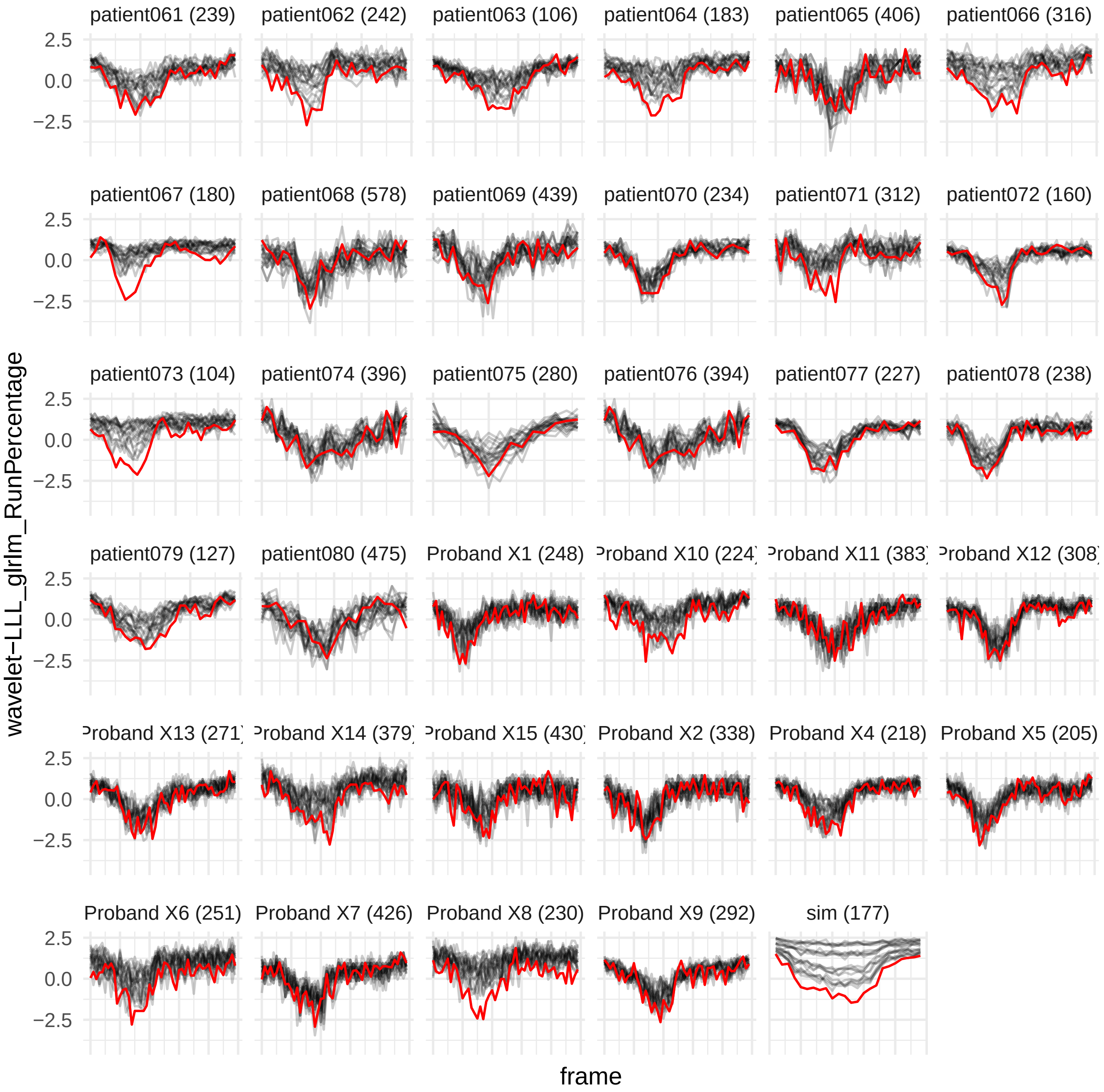

patient061 (239)
patient062 (242)
patient063 (106)
patient064 (183)
patient065 (406)
patient066 (316)
patient067 (180)
patient068 (578)
patient069 (439)
patient070 (234)
patient071 (312)
patient072 (160)
patient073 (104)
patient074 (396)
patient075 (280)
patient076 (394)
patient077 (227)
patient078 (238)
patient079 (127)
patient080 (475)
Proband X1 (248)
Proband X10 (224)
Proband X11 (383)
Proband X12 (308)
Proband X13 (271)
Proband X14 (379)
Proband X15 (430)
Proband X2 (338)
Proband X4 (218)
Proband X5 (205)
Proband X6 (251)
Proband X7 (426)
Proband X8 (230)
Proband X9 (292)
sim (177)
2.5
0.0
−2.5
wavelet−LLL_glrlm_RunPercentage
frame

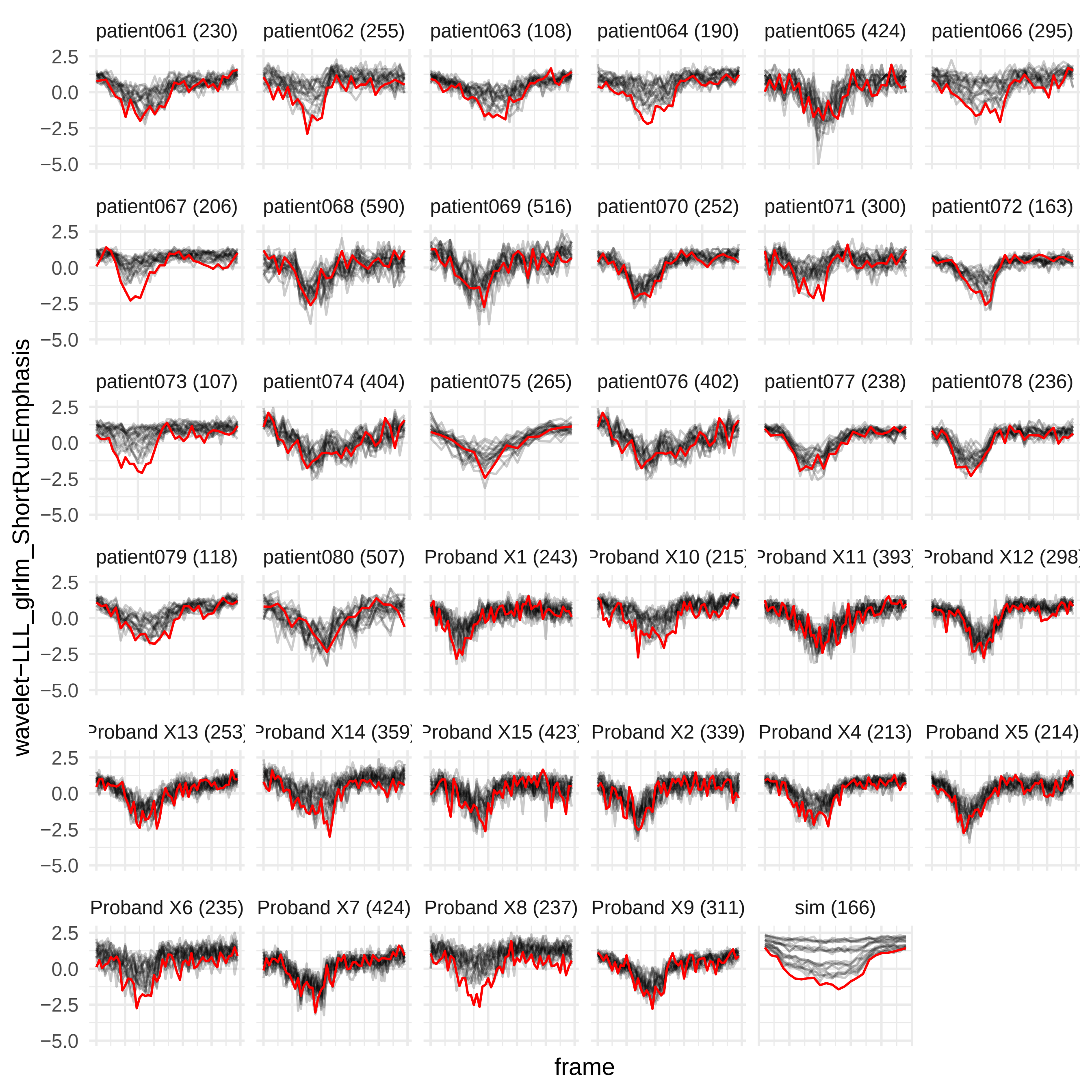

patient061 (230)
patient062 (255)
patient063 (108)
patient064 (190)
patient065 (424)
patient066 (295)
patient067 (206)
patient068 (590)
patient069 (516)
patient070 (252)
patient071 (300)
patient072 (163)
patient073 (107)
patient074 (404)
patient075 (265)
patient076 (402)
patient077 (238)
patient078 (236)
patient079 (118)
patient080 (507)
Proband X1 (243)
Proband X10 (215)
Proband X11 (393)
Proband X12 (298)
Proband X13 (253)
Proband X14 (359)
Proband X15 (423)
Proband X2 (339)
Proband X4 (213)
Proband X5 (214)
Proband X6 (235)
Proband X7 (424)
Proband X8 (237)
Proband X9 (311)
sim (166)
2.5
0.0
−2.5
−5.0
wavelet−LLL_glrlm_ShortRunEmphasis
frame

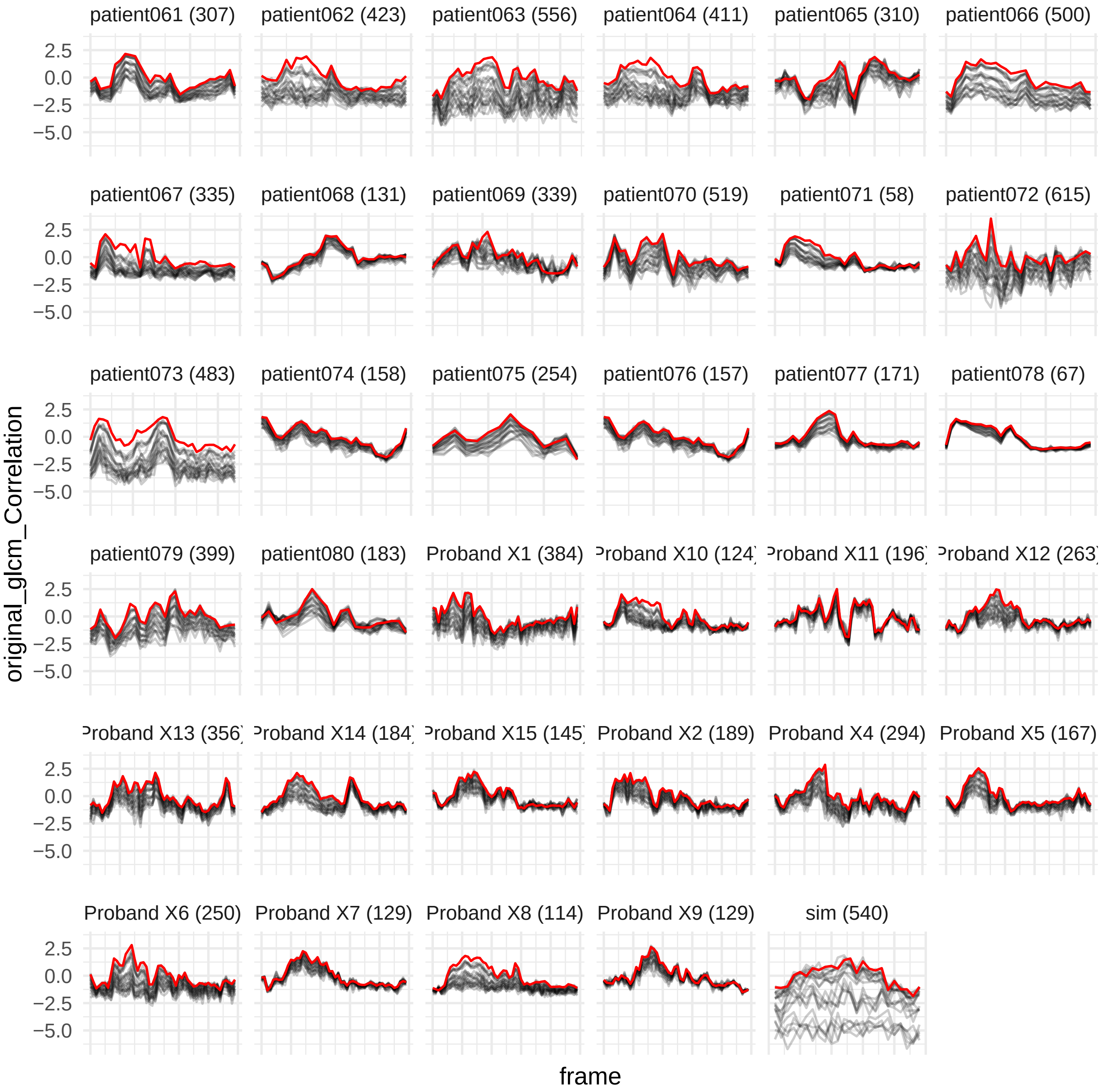

patient061 (307)
patient062 (423)
patient063 (556)
patient064 (411)
patient065 (310)
patient066 (500)
patient067 (335)
patient068 (131)
patient069 (339)
patient070 (519)
patient071 (58)
patient072 (615)
patient073 (483)
patient074 (158)
patient075 (254)
patient076 (157)
patient077 (171)
patient078 (67)
patient079 (399)
patient080 (183)
Proband X1 (384)
Proband X10 (124)
Proband X11 (196)
Proband X12 (263)
Proband X13 (356)
Proband X14 (184)
Proband X15 (145)
Proband X2 (189)
Proband X4 (294)
Proband X5 (167)
Proband X6 (250)
Proband X7 (129)
Proband X8 (114)
Proband X9 (129)
sim (540)
2.5
0.0
−2.5
−5.0
original_glcm_Correlation
frame

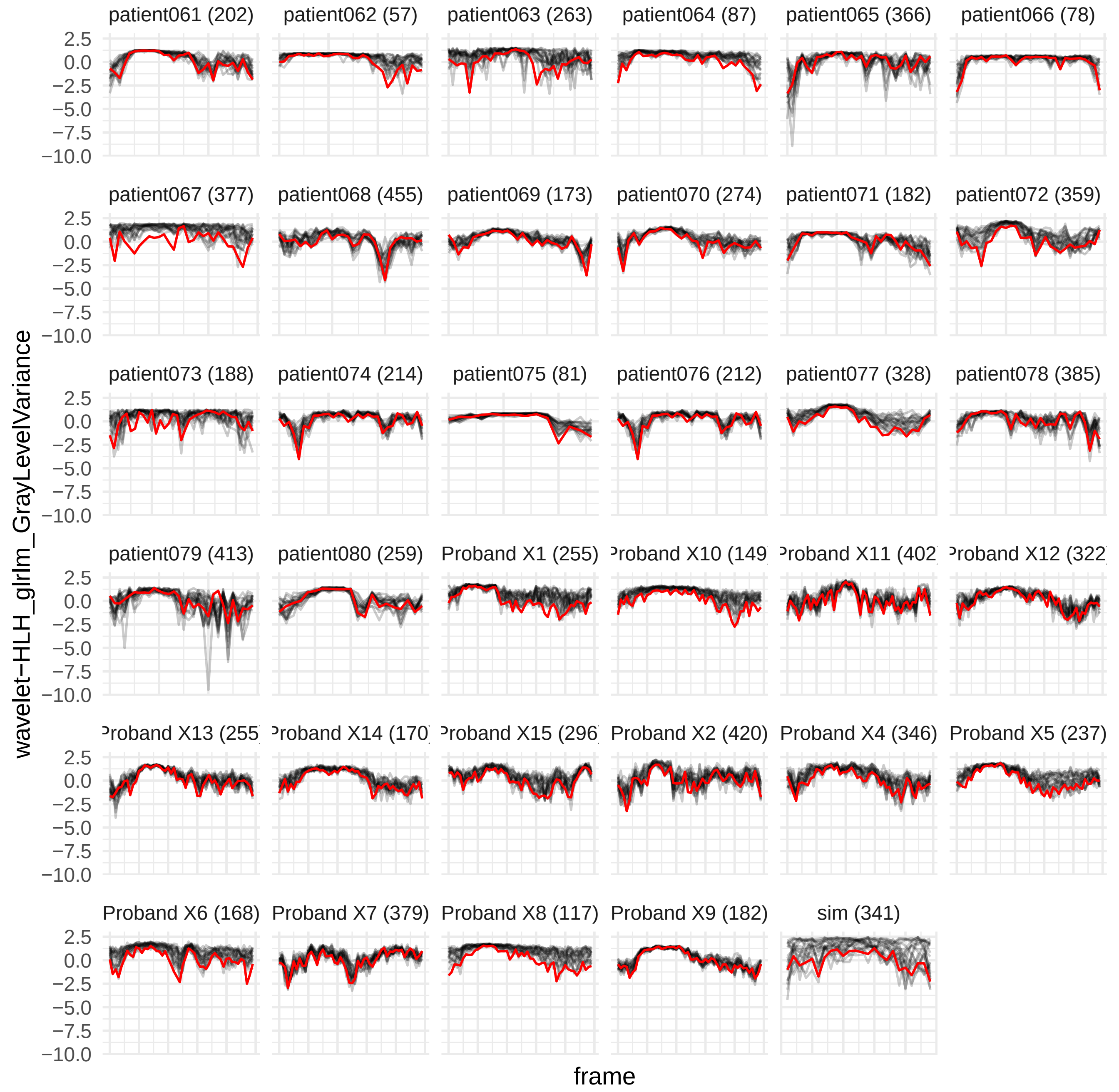

patient061 (202)
patient062 (57)
patient063 (263)
patient064 (87)
patient065 (366)
patient066 (78)
patient067 (377)
patient068 (455)
patient069 (173)
patient070 (274)
patient071 (182)
patient072 (359)
patient073 (188)
patient074 (214)
patient075 (81)
patient076 (212)
patient077 (328)
patient078 (385)
patient079 (413)
patient080 (259)
Proband X1 (255)
Proband X10 (149
Proband X11 (402
Proband X12 (322
Proband X13 (255
Proband X14 (170
Proband X15 (296
Proband X2 (420)
Proband X4 (346)
Proband X5 (237)
Proband X6 (168)
Proband X7 (379)
Proband X8 (117)
Proband X9 (182)
sim (341)
2.5
0.0
−2.5
−5.0
−7.5
−10.0
wavelet−HLH_glrlm_GrayLevelVariance
frame

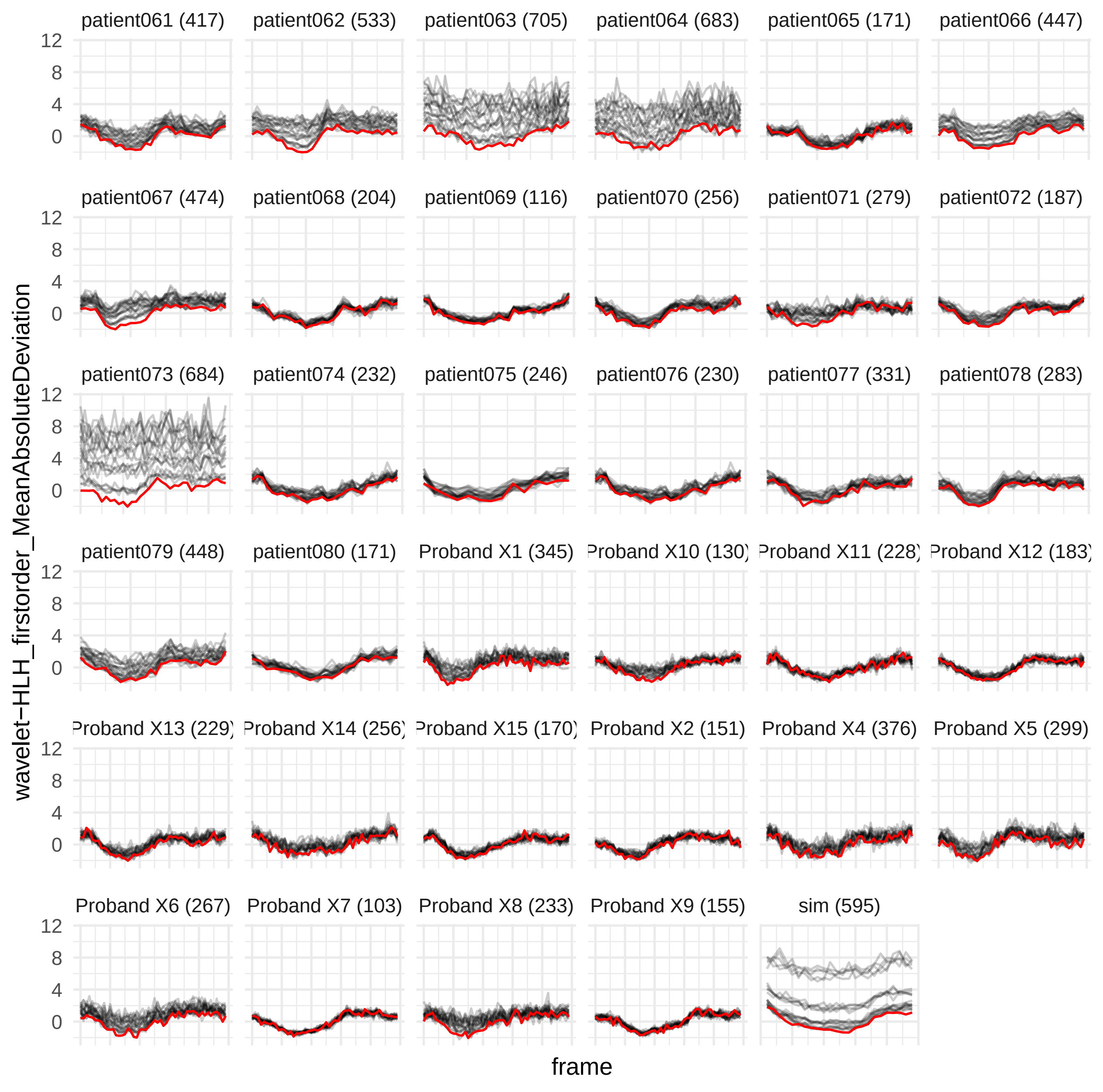

patient061 (417)
patient062 (533)
patient063 (705)
patient064 (683)
patient065 (171)
patient066 (447)
patient067 (474)
patient068 (204)
patient069 (116)
patient070 (256)
patient071 (279)
patient072 (187)
patient073 (684)
patient074 (232)
patient075 (246)
patient076 (230)
patient077 (331)
patient078 (283)
patient079 (448)
patient080 (171)
Proband X1 (345)
Proband X10 (130)
Proband X11 (228)
Proband X12 (183)
Proband X13 (229)
Proband X14 (256)
Proband X15 (170)
Proband X2 (151)
Proband X4 (376)
Proband X5 (299)
Proband X6 (267)
Proband X7 (103)
Proband X8 (233)
Proband X9 (155)
sim (595)
12
8
4
0
wavelet−HLH_firstorder_MeanAbsoluteDeviation
frame

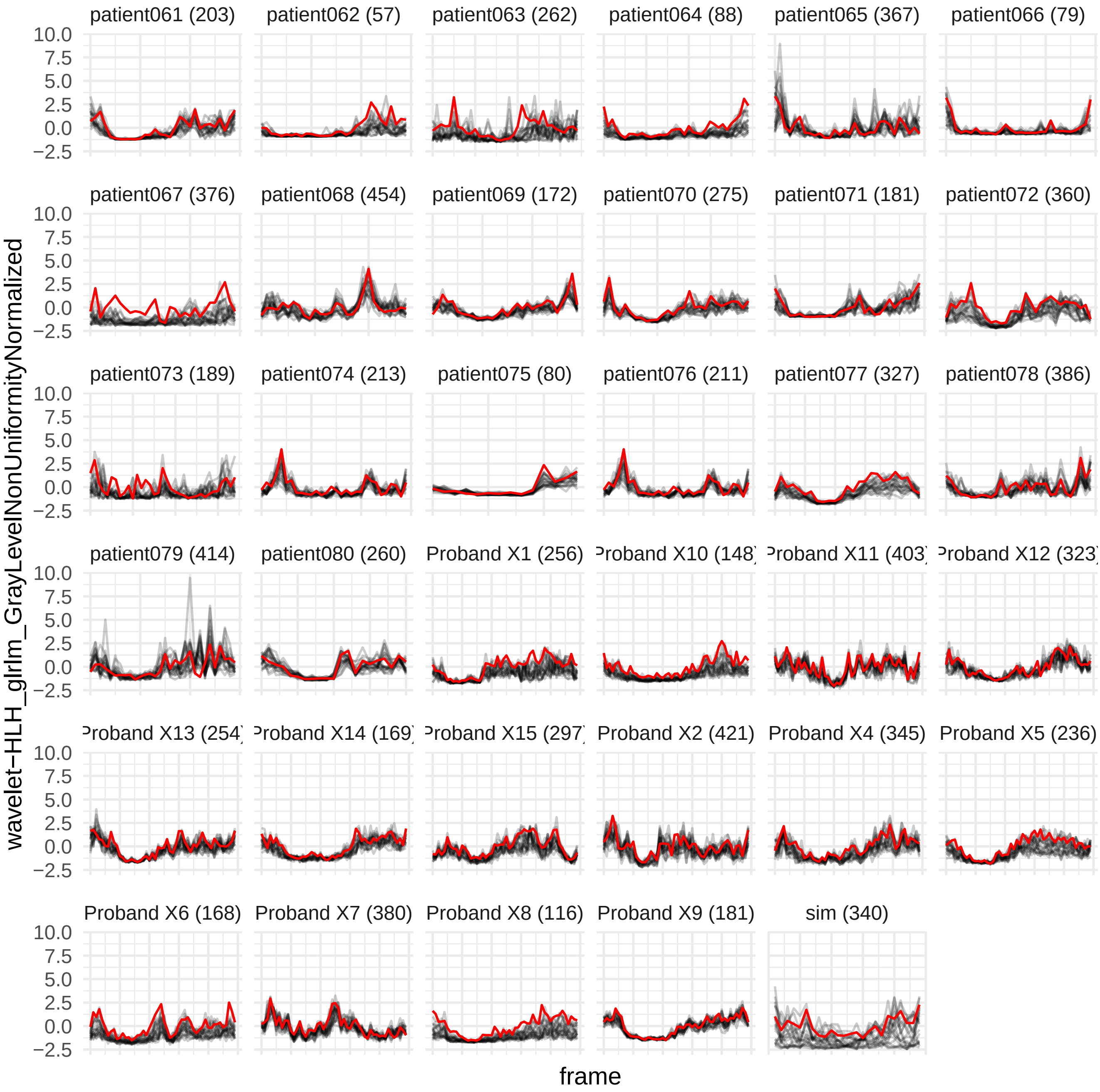

patient061 (203)
patient062 (57)
patient063 (262)
patient064 (88)
patient065 (367)
patient066 (79)
patient067 (376)
patient068 (454)
patient069 (172)
patient070 (275)
patient071 (181)
patient072 (360)
patient073 (189)
patient074 (213)
patient075 (80)
patient076 (211)
patient077 (327)
patient078 (386)
patient079 (414)
patient080 (260)
Proband X1 (256)
Proband X10 (148)
Proband X11 (403)
Proband X12 (323)
Proband X13 (254)
Proband X14 (169)
Proband X15 (297)
Proband X2 (421)
Proband X4 (345)
Proband X5 (236)
Proband X6 (168)
Proband X7 (380)
Proband X8 (116)
Proband X9 (181)
sim (340)
10.0
7.5
5.0
2.5
0.0
−2.5
wavelet−HLH_glrlm_GrayLevelNonUniformityNormalized
frame

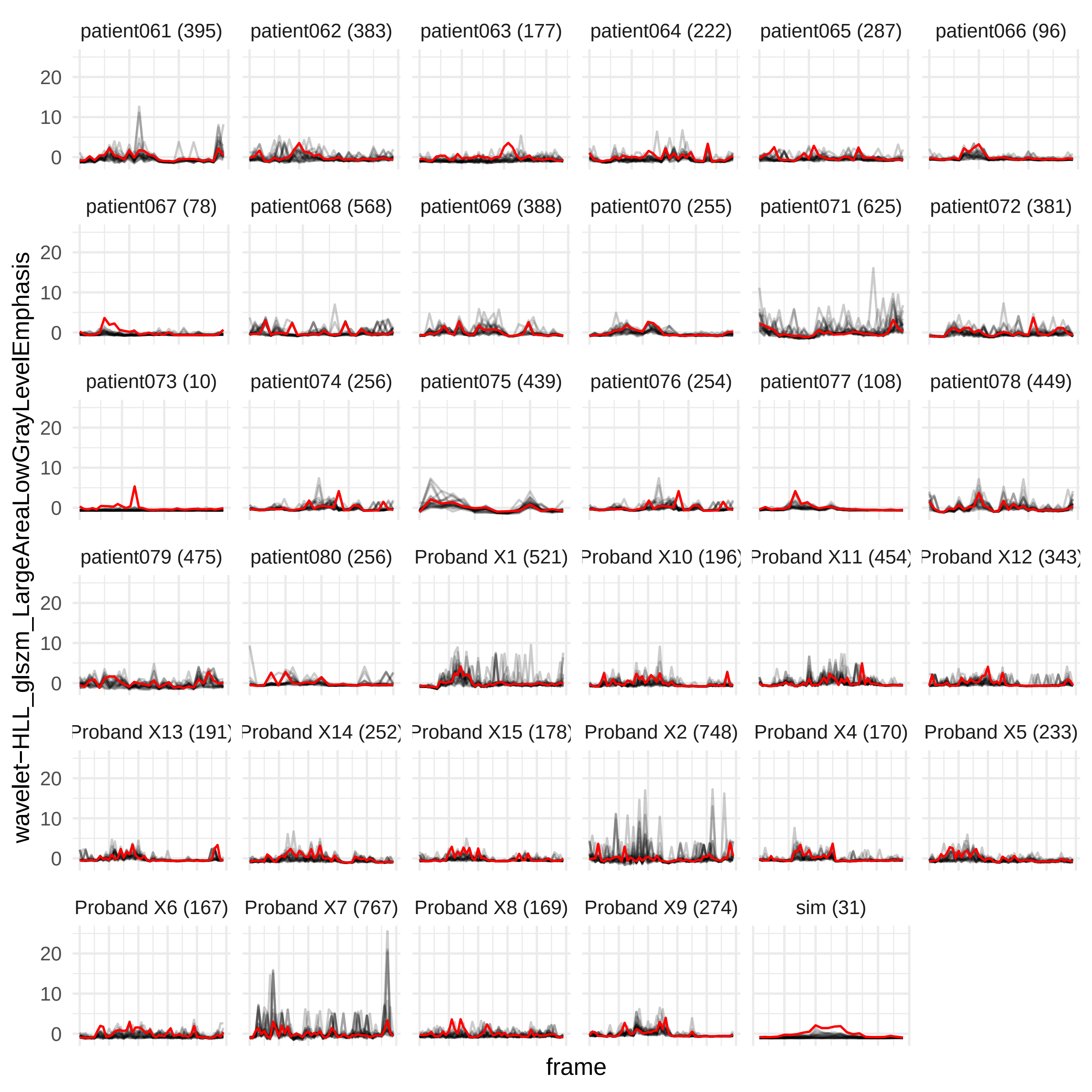
patient061 (395)
patient062 (383)
patient063 (177)
patient064 (222)
patient065 (287)
patient066 (96)
patient067 (78)
patient068 (568)
patient069 (388)
patient070 (255)
patient071 (625)
patient072 (381)
patient073 (10)
patient074 (256)
patient075 (439)
patient076 (254)
patient077 (108)
patient078 (449)
patient079 (475)
patient080 (256)
Proband X1 (521)
Proband X10 (196)
Proband X11 (454)
Proband X12 (343)
Proband X13 (191)
Proband X14 (252)
Proband X15 (178)
Proband X2 (748)
Proband X4 (170)
Proband X5 (233)
Proband X6 (167)
Proband X7 (767)
Proband X8 (169)
Proband X9 (274)
sim (31)
wavelet−HLL_glszm_LargeAreaLowGrayLevelEmphasis
frame
0
10
20

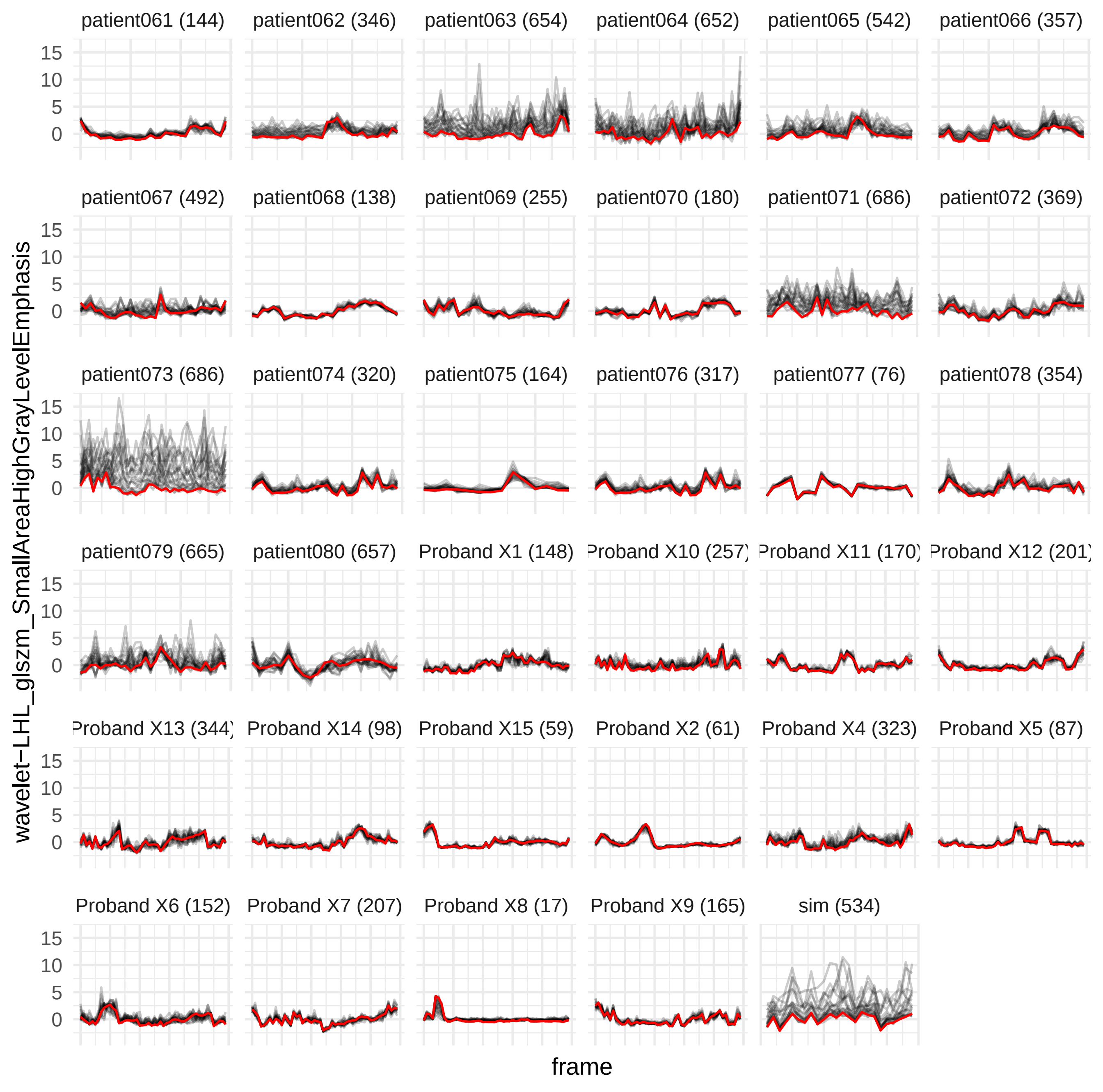

patient061 (144)
patient062 (346)
patient063 (654)
patient064 (652)
patient065 (542)
patient066 (357)
patient067 (492)
patient068 (138)
patient069 (255)
patient070 (180)
patient071 (686)
patient072 (369)
patient073 (686)
patient074 (320)
patient075 (164)
patient076 (317)
patient077 (76)
patient078 (354)
patient079 (665)
patient080 (657)
Proband X1 (148)
Proband X10 (257)
Proband X11 (170)
Proband X12 (201)
Proband X13 (344)
Proband X14 (98)
Proband X15 (59)
Proband X2 (61)
Proband X4 (323)
Proband X5 (87)
Proband X6 (152)
Proband X7 (207)
Proband X8 (17)
Proband X9 (165)
sim (534)
wavelet−LHL_glszm_SmallAreaHighGrayLevelEmphasis
frame
0
5
10
15

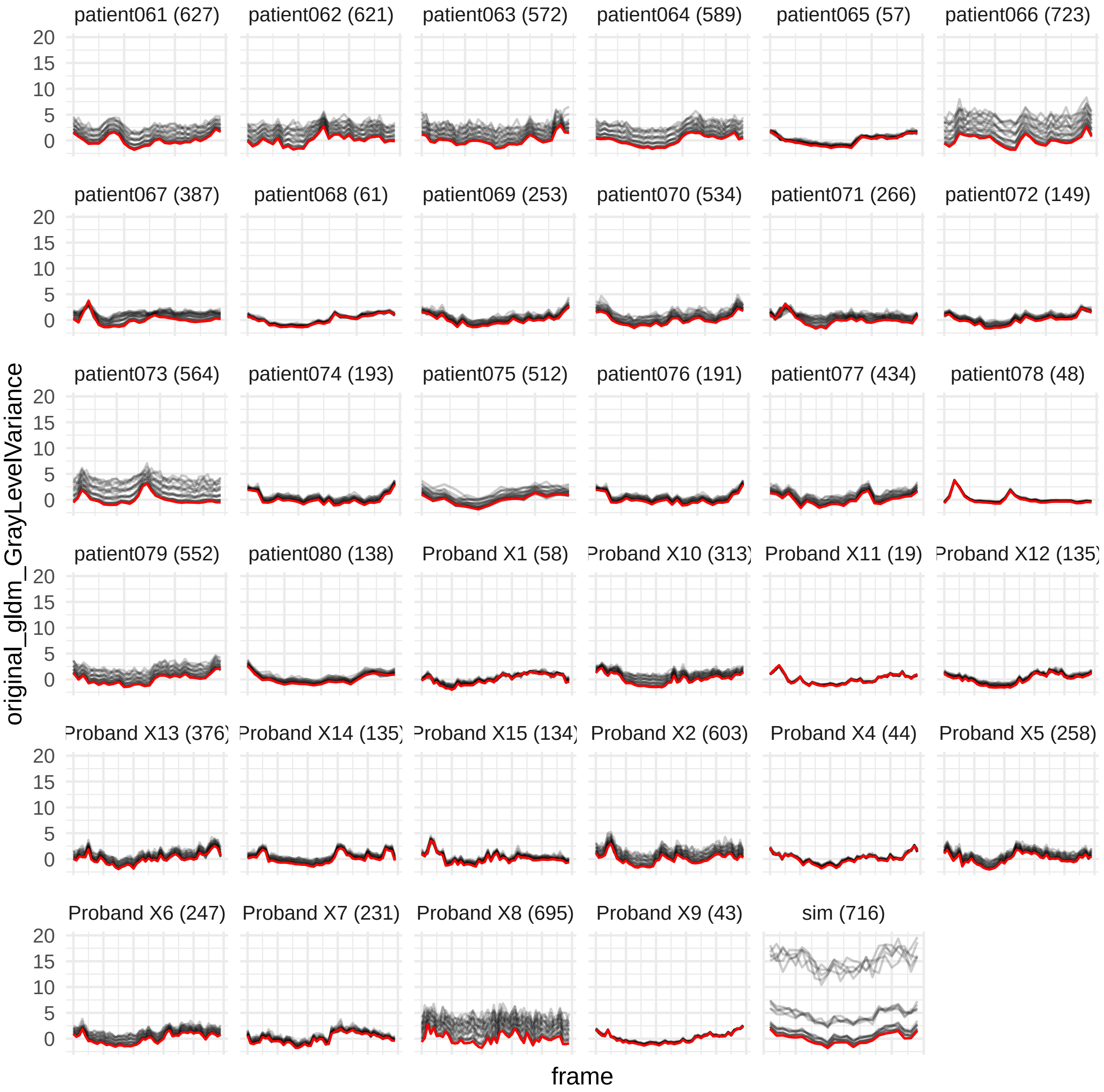

patient061 (627)
patient062 (621)
patient063 (572)
patient064 (589)
patient065 (57)
patient066 (723)
patient067 (387)
patient068 (61)
patient069 (253)
patient070 (534)
patient071 (266)
patient072 (149)
patient073 (564)
patient074 (193)
patient075 (512)
patient076 (191)
patient077 (434)
patient078 (48)
patient079 (552)
patient080 (138)
Proband X1 (58)
Proband X10 (313)
Proband X11 (19)
Proband X12 (135)
Proband X13 (376)
Proband X14 (135)
Proband X15 (134)
Proband X2 (603)
Proband X4 (44)
Proband X5 (258)
Proband X6 (247)
Proband X7 (231)
Proband X8 (695)
Proband X9 (43)
sim (716)
original_gldm_GrayLevelVariance
frame
20
15
10
5
0

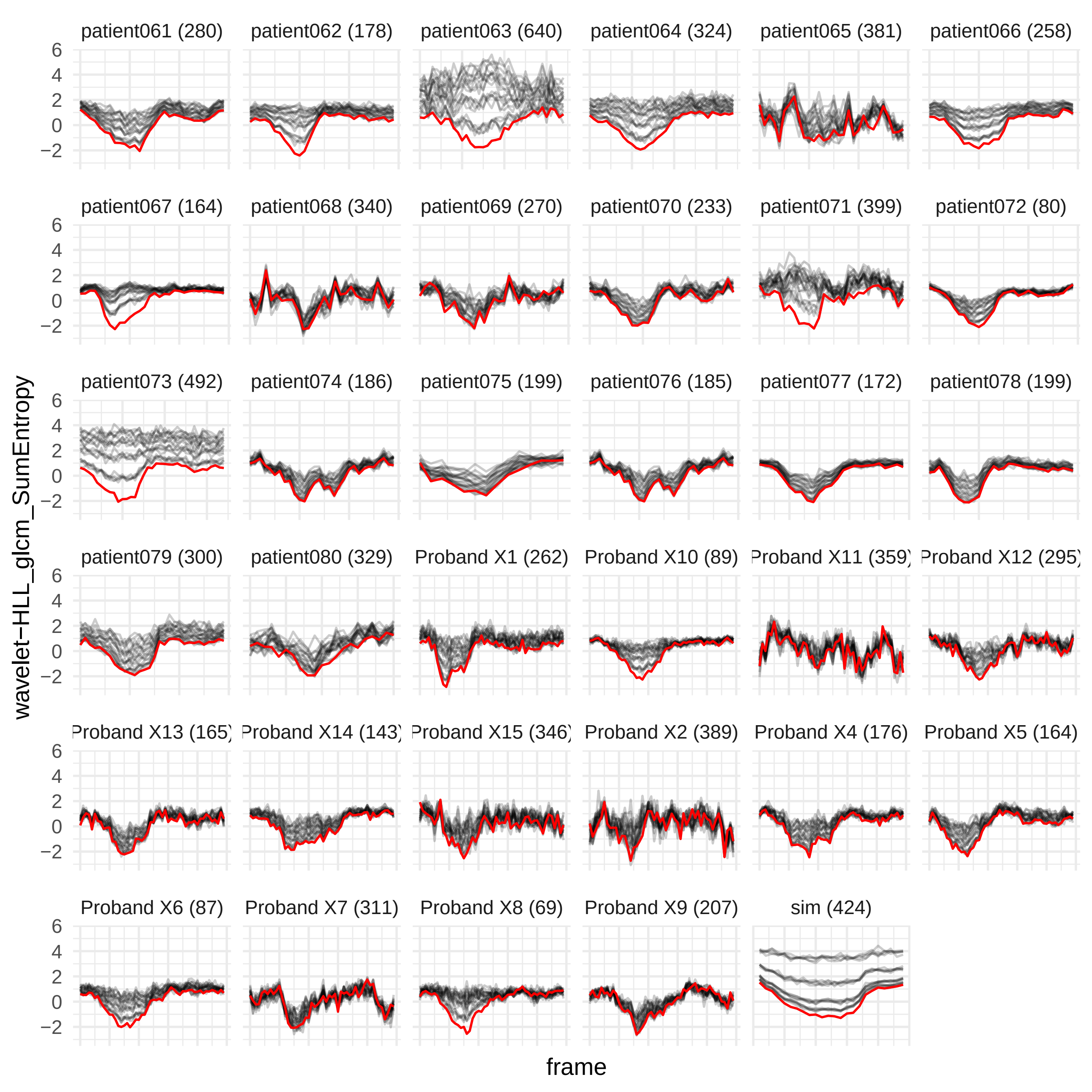

patient061 (280)
patient062 (178)
patient063 (640)
patient064 (324)
patient065 (381)
patient066 (258)
patient067 (164)
patient068 (340)
patient069 (270)
patient070 (233)
patient071 (399)
patient072 (80)
patient073 (492)
patient074 (186)
patient075 (199)
patient076 (185)
patient077 (172)
patient078 (199)
patient079 (300)
patient080 (329)
Proband X1 (262)
Proband X10 (89)
Proband X11 (359)
Proband X12 (295)
Proband X13 (165)
Proband X14 (143)
Proband X15 (346)
Proband X2 (389)
Proband X4 (176)
Proband X5 (164)
Proband X6 (87)
Proband X7 (311)
Proband X8 (69)
Proband X9 (207)
sim (424)
wavelet−HLL_glcm_SumEntropy
frame
6
4
2
0
−2

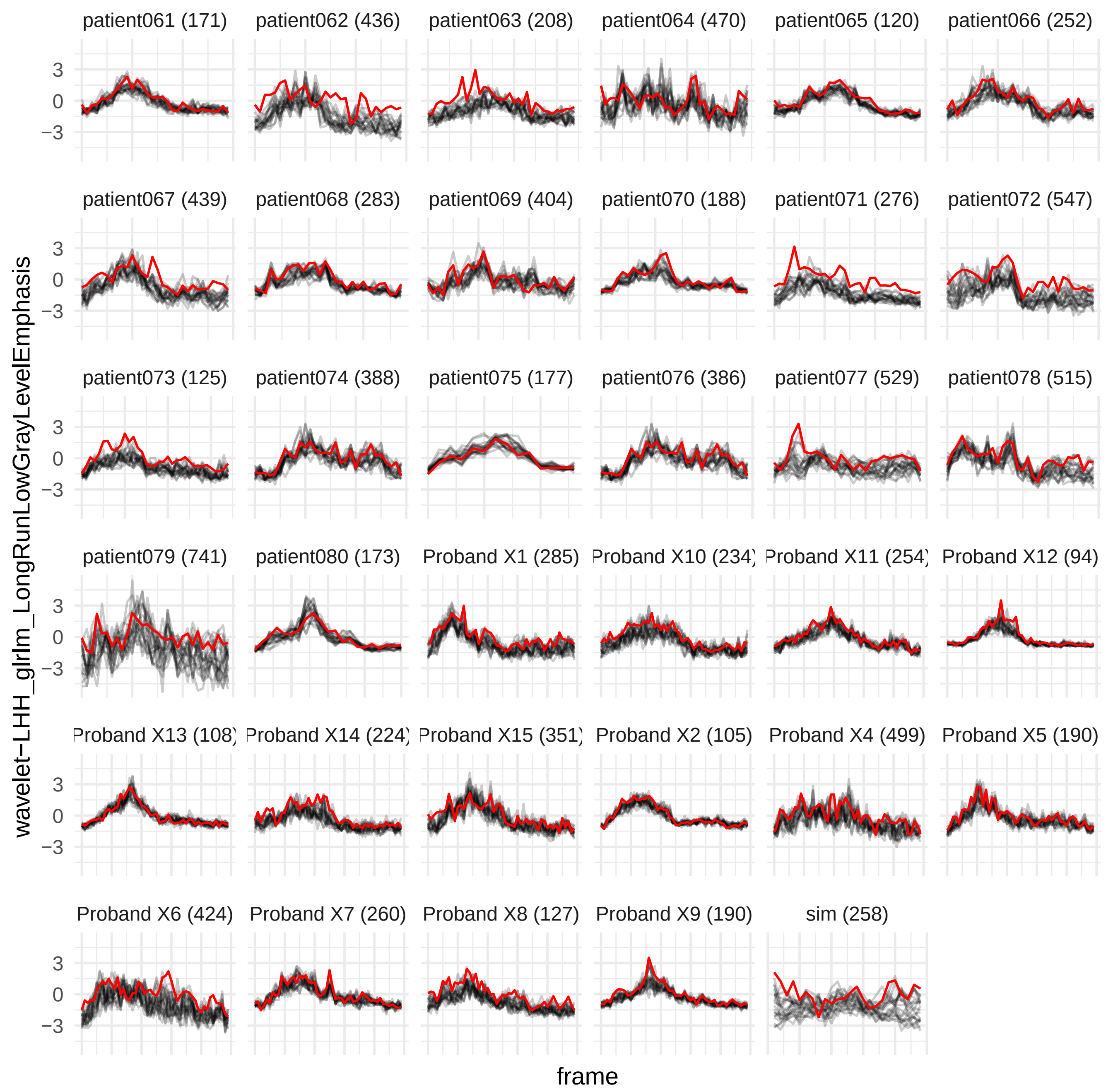
patient061 (171)
patient062 (436)
patient063 (208)
patient064 (470)
patient065 (120)
patient066 (252)
patient067 (439)
patient068 (283)
patient069 (404)
patient070 (188)
patient071 (276)
patient072 (547)
patient073 (125)
patient074 (388)
patient075 (177)
patient076 (386)
patient077 (529)
patient078 (515)
patient079 (741)
patient080 (173)
Proband X1 (285)
Proband X10 (234)
Proband X11 (254)
Proband X12 (94)
Proband X13 (108)
Proband X14 (224)
Proband X15 (351)
Proband X2 (105)
Proband X4 (499)
Proband X5 (190)
Proband X6 (424)
Proband X7 (260)
Proband X8 (127)
Proband X9 (190)
sim (258)
3
0
-3
wavelet-LHH_glrlm_LongRunLowGrayLevelEmphasis
frame

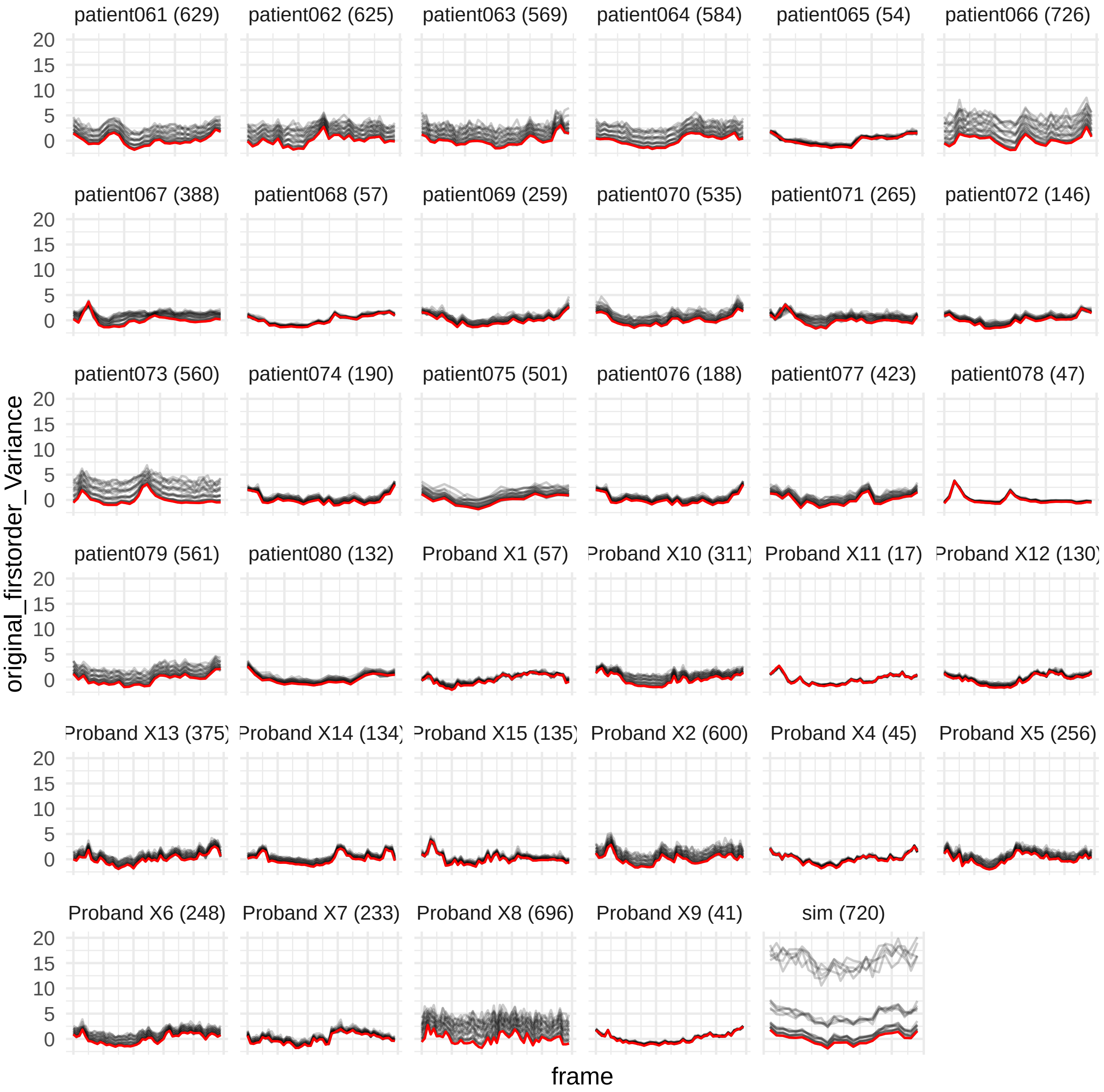

patient061 (629)
patient062 (625)
patient063 (569)
patient064 (584)
patient065 (54)
patient066 (726)
patient067 (388)
patient068 (57)
patient069 (259)
patient070 (535)
patient071 (265)
patient072 (146)
patient073 (560)
patient074 (190)
patient075 (501)
patient076 (188)
patient077 (423)
patient078 (47)
patient079 (561)
patient080 (132)
Proband X1 (57)
Proband X10 (311)
Proband X11 (17)
Proband X12 (130)
Proband X13 (375)
Proband X14 (134)
Proband X15 (135)
Proband X2 (600)
Proband X4 (45)
Proband X5 (256)
Proband X6 (248)
Proband X7 (233)
Proband X8 (696)
Proband X9 (41)
sim (720)
original_firstorder_Variance
0
5
10
15
20
frame

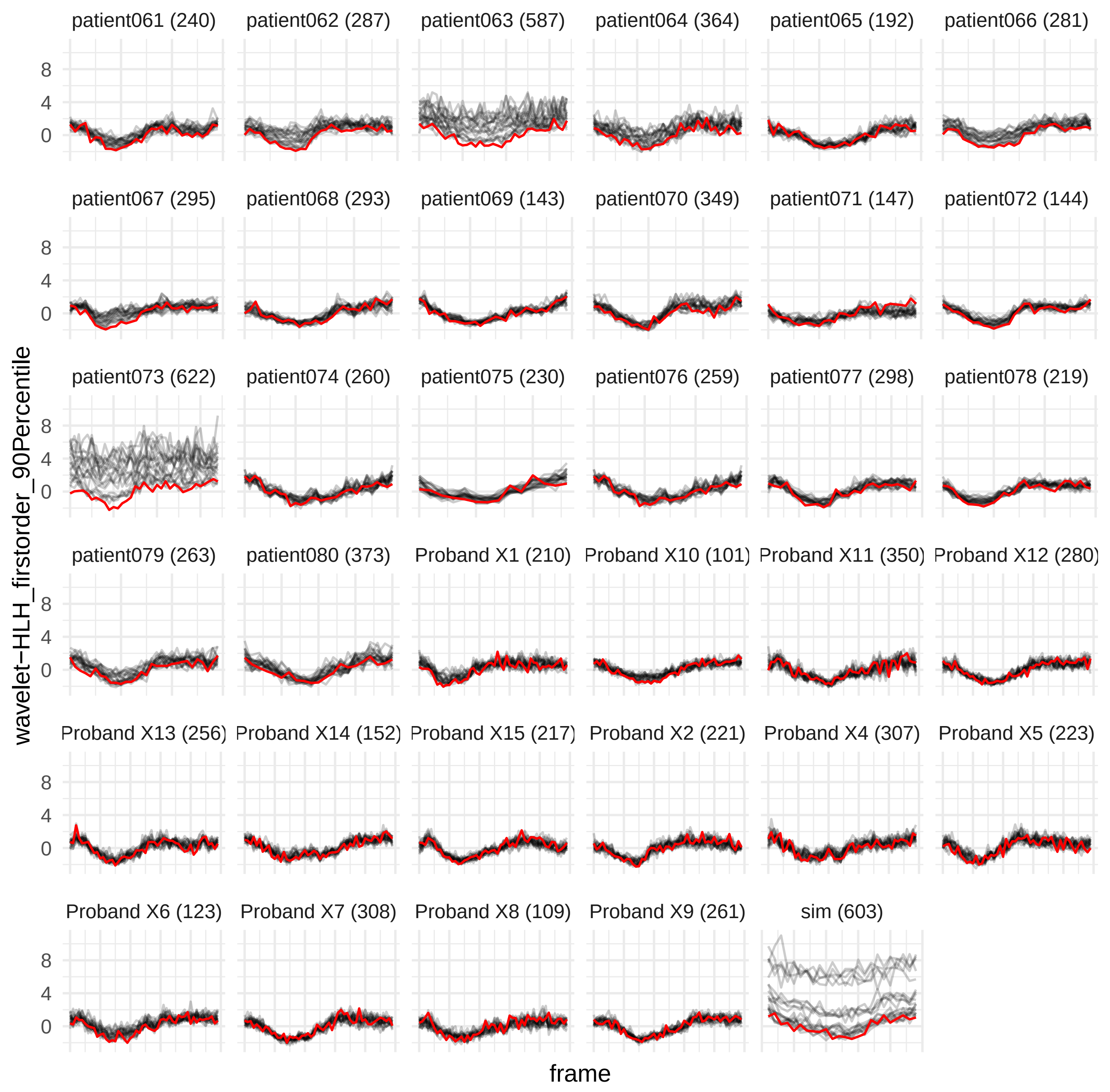

patient061 (240)
patient062 (287)
patient063 (587)
patient064 (364)
patient065 (192)
patient066 (281)
patient067 (295)
patient068 (293)
patient069 (143)
patient070 (349)
patient071 (147)
patient072 (144)
patient073 (622)
patient074 (260)
patient075 (230)
patient076 (259)
patient077 (298)
patient078 (219)
patient079 (263)
patient080 (373)
Proband X1 (210)
Proband X10 (101)
Proband X11 (350)
Proband X12 (280)
Proband X13 (256)
Proband X14 (152)
Proband X15 (217)
Proband X2 (221)
Proband X4 (307)
Proband X5 (223)
Proband X6 (123)
Proband X7 (308)
Proband X8 (109)
Proband X9 (261)
sim (603)
8
4
0
wavelet−HLH_firstorder_90Percentile
frame

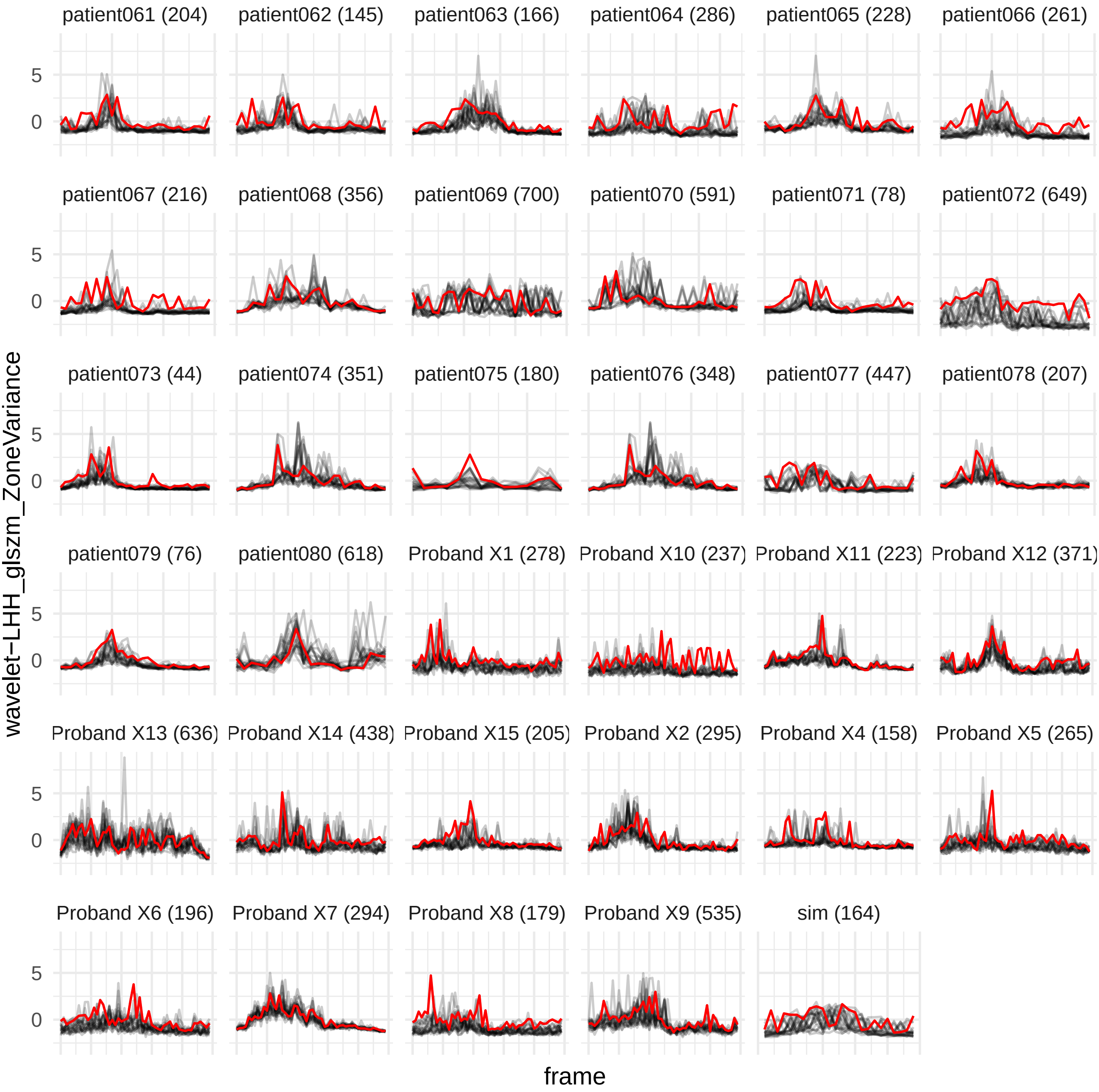

patient061 (204)
patient062 (145)
patient063 (166)
patient064 (286)
patient065 (228)
patient066 (261)
patient067 (216)
patient068 (356)
patient069 (700)
patient070 (591)
patient071 (78)
patient072 (649)
patient073 (44)
patient074 (351)
patient075 (180)
patient076 (348)
patient077 (447)
patient078 (207)
patient079 (76)
patient080 (618)
Proband X1 (278)
Proband X10 (237)
Proband X11 (223)
Proband X12 (371)
Proband X13 (636)
Proband X14 (438)
Proband X15 (205)
Proband X2 (295)
Proband X4 (158)
Proband X5 (265)
Proband X6 (196)
Proband X7 (294)
Proband X8 (179)
Proband X9 (535)
sim (164)
5
0
wavelet−LHH_glszm_ZoneVariance
frame

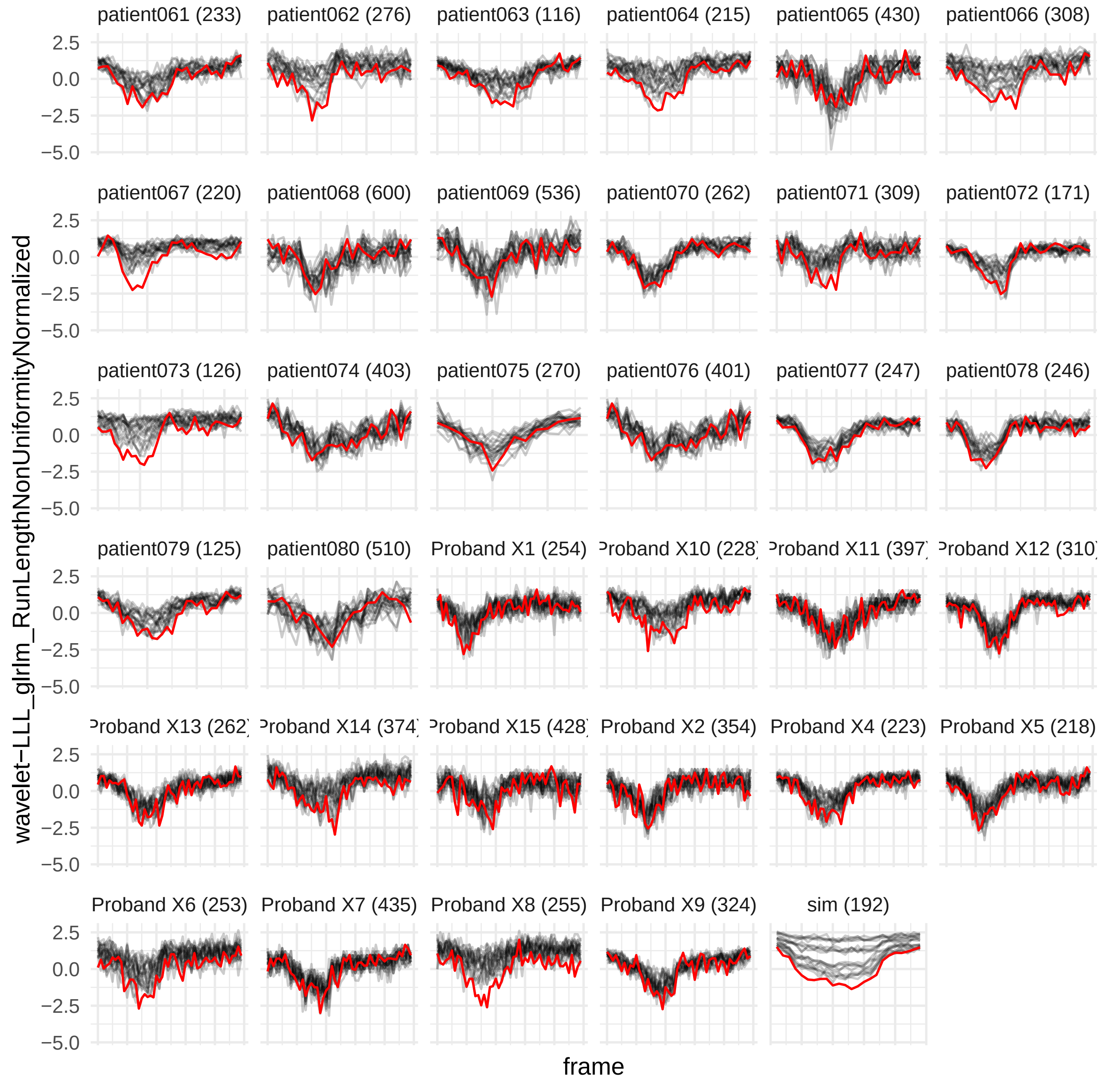

patient061 (233)
patient062 (276)
patient063 (116)
patient064 (215)
patient065 (430)
patient066 (308)
patient067 (220)
patient068 (600)
patient069 (536)
patient070 (262)
patient071 (309)
patient072 (171)
patient073 (126)
patient074 (403)
patient075 (270)
patient076 (401)
patient077 (247)
patient078 (246)
patient079 (125)
patient080 (510)
Proband X1 (254)
Proband X10 (228)
Proband X11 (397)
Proband X12 (310)
Proband X13 (262)
Proband X14 (374)
Proband X15 (428)
Proband X2 (354)
Proband X4 (223)
Proband X5 (218)
Proband X6 (253)
Proband X7 (435)
Proband X8 (255)
Proband X9 (324)
sim (192)
2.5
0.0
−2.5
−5.0
wavelet−LLL_glrlm_RunLengthNonUniformityNormalized
frame

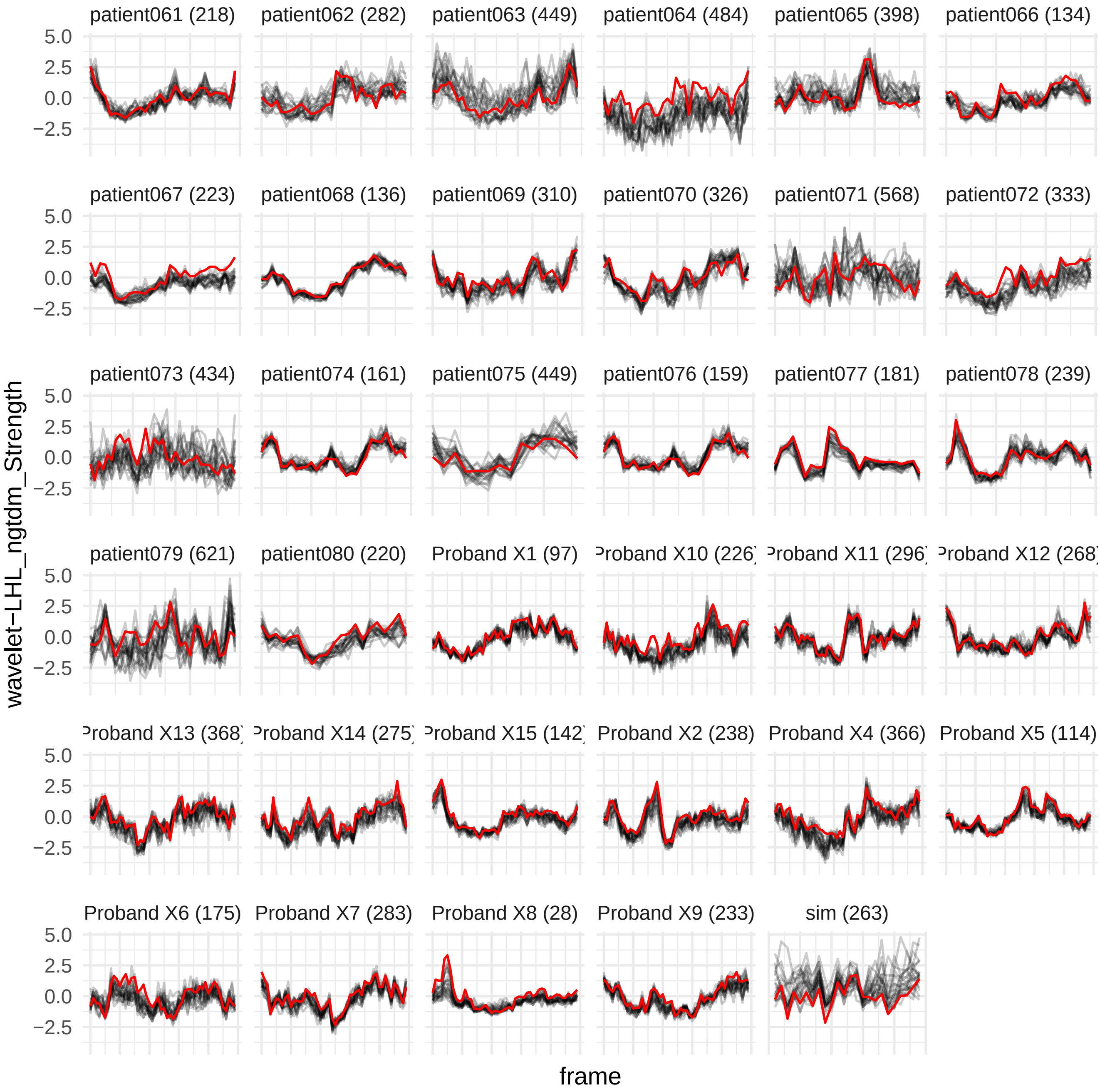
patient061 (218)
patient062 (282)
patient063 (449)
patient064 (484)
patient065 (398)
patient066 (134)
patient067 (223)
patient068 (136)
patient069 (310)
patient070 (326)
patient071 (568)
patient072 (333)
patient073 (434)
patient074 (161)
patient075 (449)
patient076 (159)
patient077 (181)
patient078 (239)
patient079 (621)
patient080 (220)
Proband X1 (97)
Proband X10 (226)
Proband X11 (296)
Proband X12 (268)
Proband X13 (368)
Proband X14 (275)
Proband X15 (142)
Proband X2 (238)
Proband X4 (366)
Proband X5 (114)
Proband X6 (175)
Proband X7 (283)
Proband X8 (28)
Proband X9 (233)
sim (263)
5.0
2.5
0.0
−2.5
wavelet−LHL_ngtdm_Strength
frame

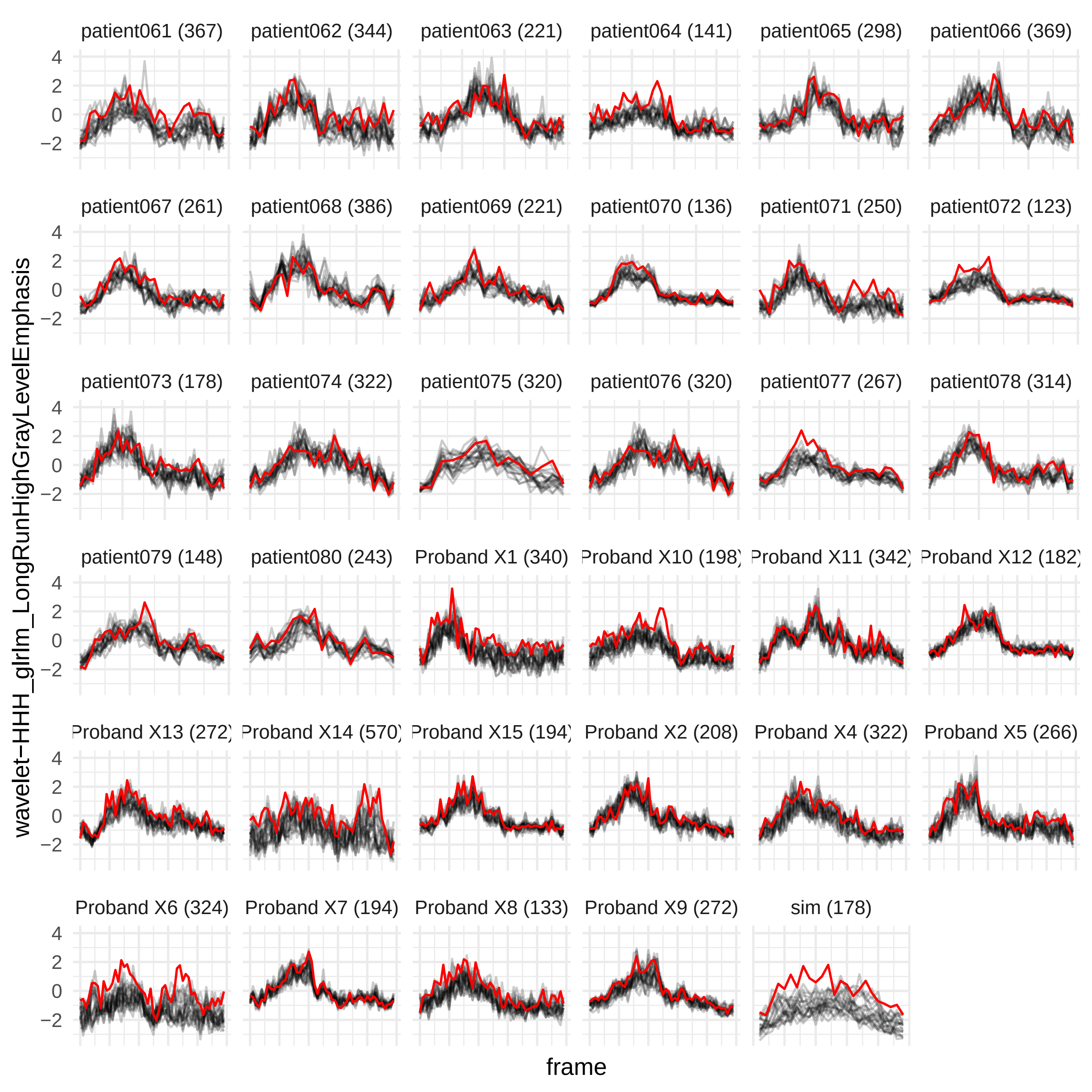
patient061 (367)
patient062 (344)
patient063 (221)
patient064 (141)
patient065 (298)
patient066 (369)
patient067 (261)
patient068 (386)
patient069 (221)
patient070 (136)
patient071 (250)
patient072 (123)
patient073 (178)
patient074 (322)
patient075 (320)
patient076 (320)
patient077 (267)
patient078 (314)
patient079 (148)
patient080 (243)
Proband X1 (340)
Proband X10 (198)
Proband X11 (342)
Proband X12 (182)
Proband X13 (272)
Proband X14 (570)
Proband X15 (194)
Proband X2 (208)
Proband X4 (322)
Proband X5 (266)
Proband X6 (324)
Proband X7 (194)
Proband X8 (133)
Proband X9 (272)
sim (178)
4
2
0
−2
wavelet−HHH_glrlm_LongRunHighGrayLevelEmphasis
frame

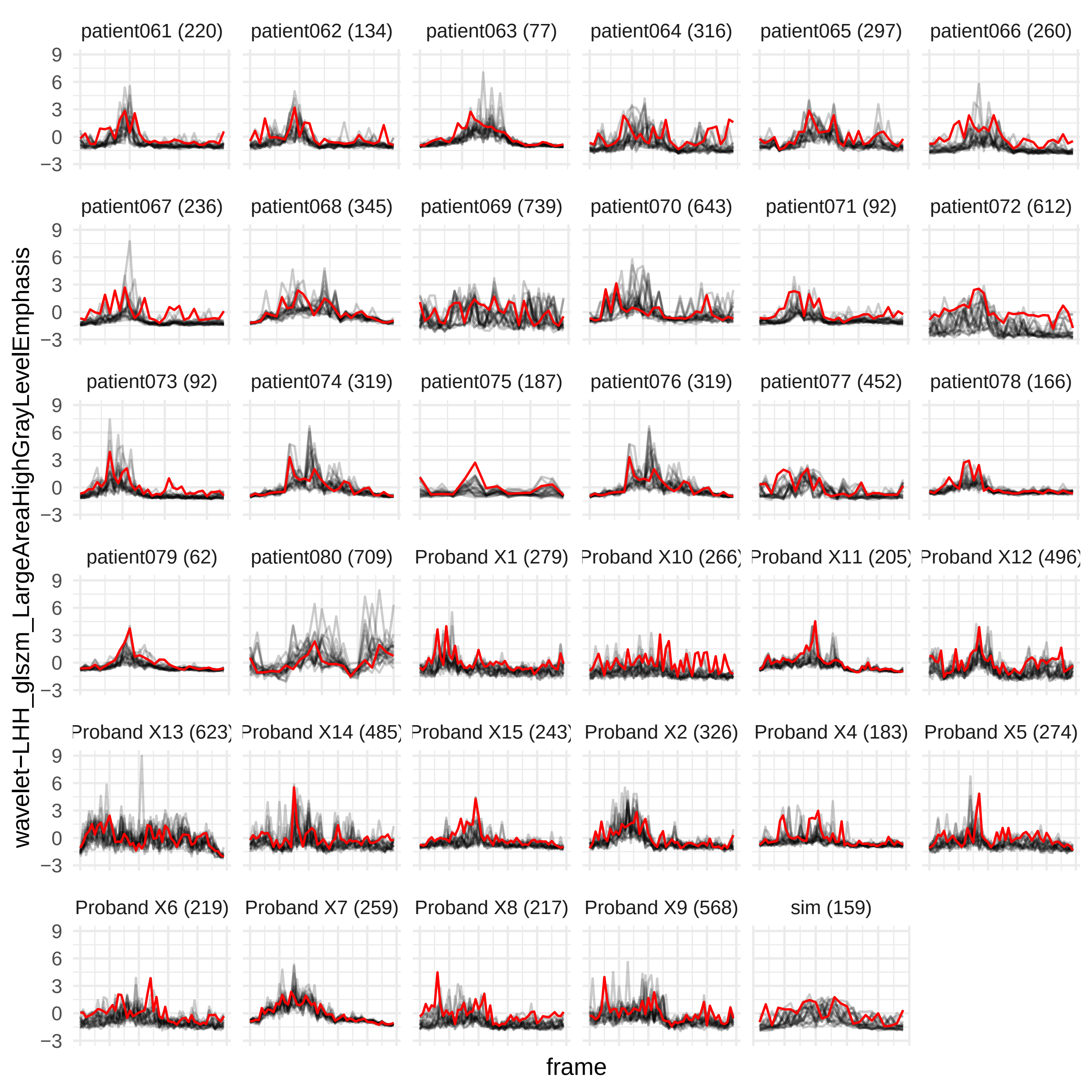

patient061 (220)
patient062 (134)
patient063 (77)
patient064 (316)
patient065 (297)
patient066 (260)
patient067 (236)
patient068 (345)
patient069 (739)
patient070 (643)
patient071 (92)
patient072 (612)
patient073 (92)
patient074 (319)
patient075 (187)
patient076 (319)
patient077 (452)
patient078 (166)
patient079 (62)
patient080 (709)
Proband X1 (279)
Proband X10 (266)
Proband X11 (205)
Proband X12 (496)
Proband X13 (623)
Proband X14 (485)
Proband X15 (243)
Proband X2 (326)
Proband X4 (183)
Proband X5 (274)
Proband X6 (219)
Proband X7 (259)
Proband X8 (217)
Proband X9 (568)
sim (159)
9
6
3
0
−3
wavelet−LHH_glszm_LargeAreaHighGrayLevelEmphasis
frame

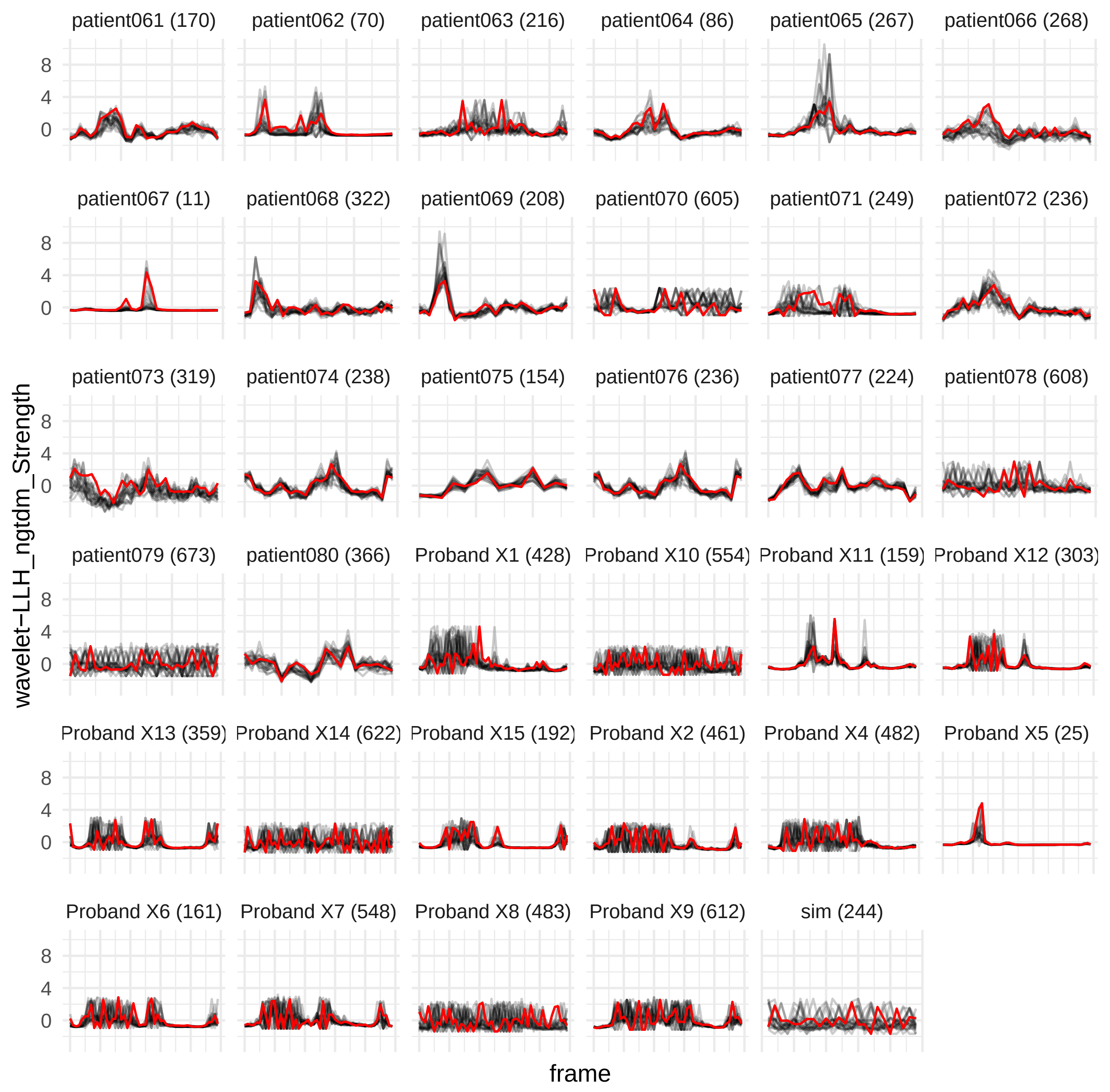

patient061 (170)
patient062 (70)
patient063 (216)
patient064 (86)
patient065 (267)
patient066 (268)
patient067 (11)
patient068 (322)
patient069 (208)
patient070 (605)
patient071 (249)
patient072 (236)
patient073 (319)
patient074 (238)
patient075 (154)
patient076 (236)
patient077 (224)
patient078 (608)
patient079 (673)
patient080 (366)
Proband X1 (428)
Proband X10 (554)
Proband X11 (159)
Proband X12 (303)
Proband X13 (359)
Proband X14 (622)
Proband X15 (192)
Proband X2 (461)
Proband X4 (482)
Proband X5 (25)
Proband X6 (161)
Proband X7 (548)
Proband X8 (483)
Proband X9 (612)
sim (244)
8
4
0
wavelet−LLH_ngtdm_Strength
frame

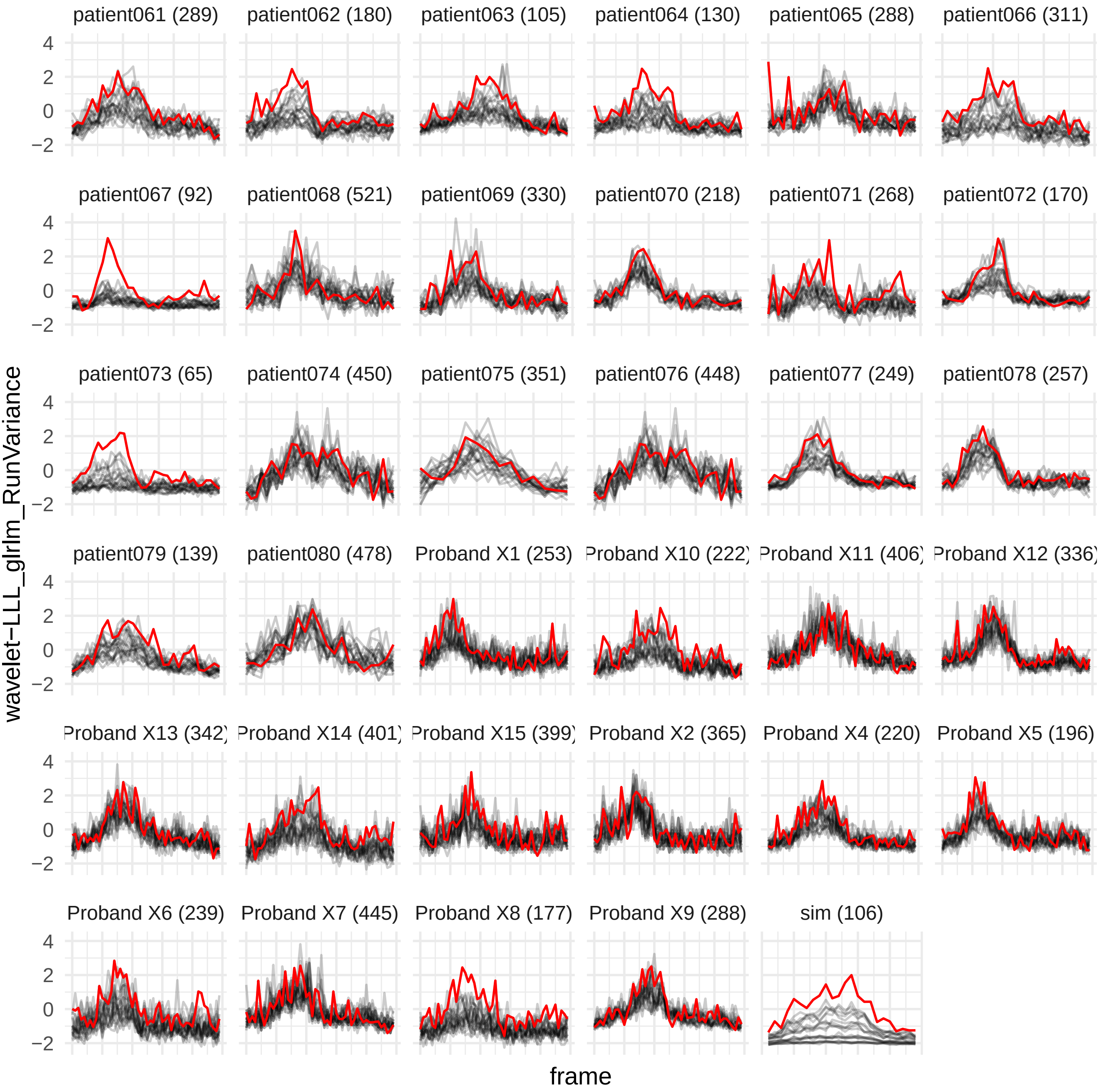

patient061 (289)
patient062 (180)
patient063 (105)
patient064 (130)
patient065 (288)
patient066 (311)
patient067 (92)
patient068 (521)
patient069 (330)
patient070 (218)
patient071 (268)
patient072 (170)
patient073 (65)
patient074 (450)
patient075 (351)
patient076 (448)
patient077 (249)
patient078 (257)
patient079 (139)
patient080 (478)
Proband X1 (253)
Proband X10 (222)
Proband X11 (406)
Proband X12 (336)
Proband X13 (342)
Proband X14 (401)
Proband X15 (399)
Proband X2 (365)
Proband X4 (220)
Proband X5 (196)
Proband X6 (239)
Proband X7 (445)
Proband X8 (177)
Proband X9 (288)
sim (106)
4
2
0
−2
wavelet−LLL_glrlm_RunVariance
frame

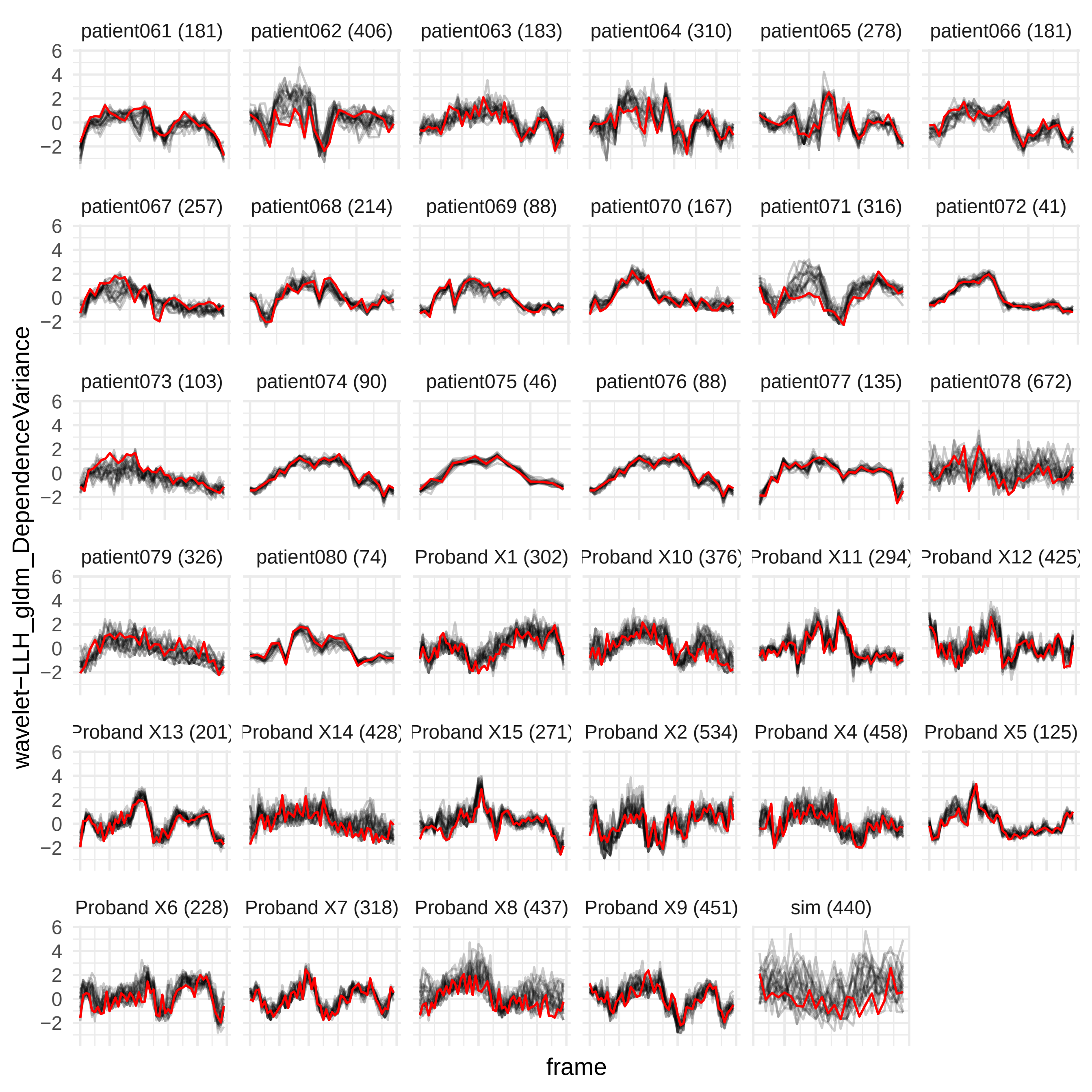

patient061 (181)
patient062 (406)
patient063 (183)
patient064 (310)
patient065 (278)
patient066 (181)
patient067 (257)
patient068 (214)
patient069 (88)
patient070 (167)
patient071 (316)
patient072 (41)
patient073 (103)
patient074 (90)
patient075 (46)
patient076 (88)
patient077 (135)
patient078 (672)
patient079 (326)
patient080 (74)
Proband X1 (302)
Proband X10 (376)
Proband X11 (294)
Proband X12 (425)
Proband X13 (201)
Proband X14 (428)
Proband X15 (271)
Proband X2 (534)
Proband X4 (458)
Proband X5 (125)
Proband X6 (228)
Proband X7 (318)
Proband X8 (437)
Proband X9 (451)
sim (440)
6
4
2
0
−2
wavelet−LLH_gldm_DependenceVariance
frame

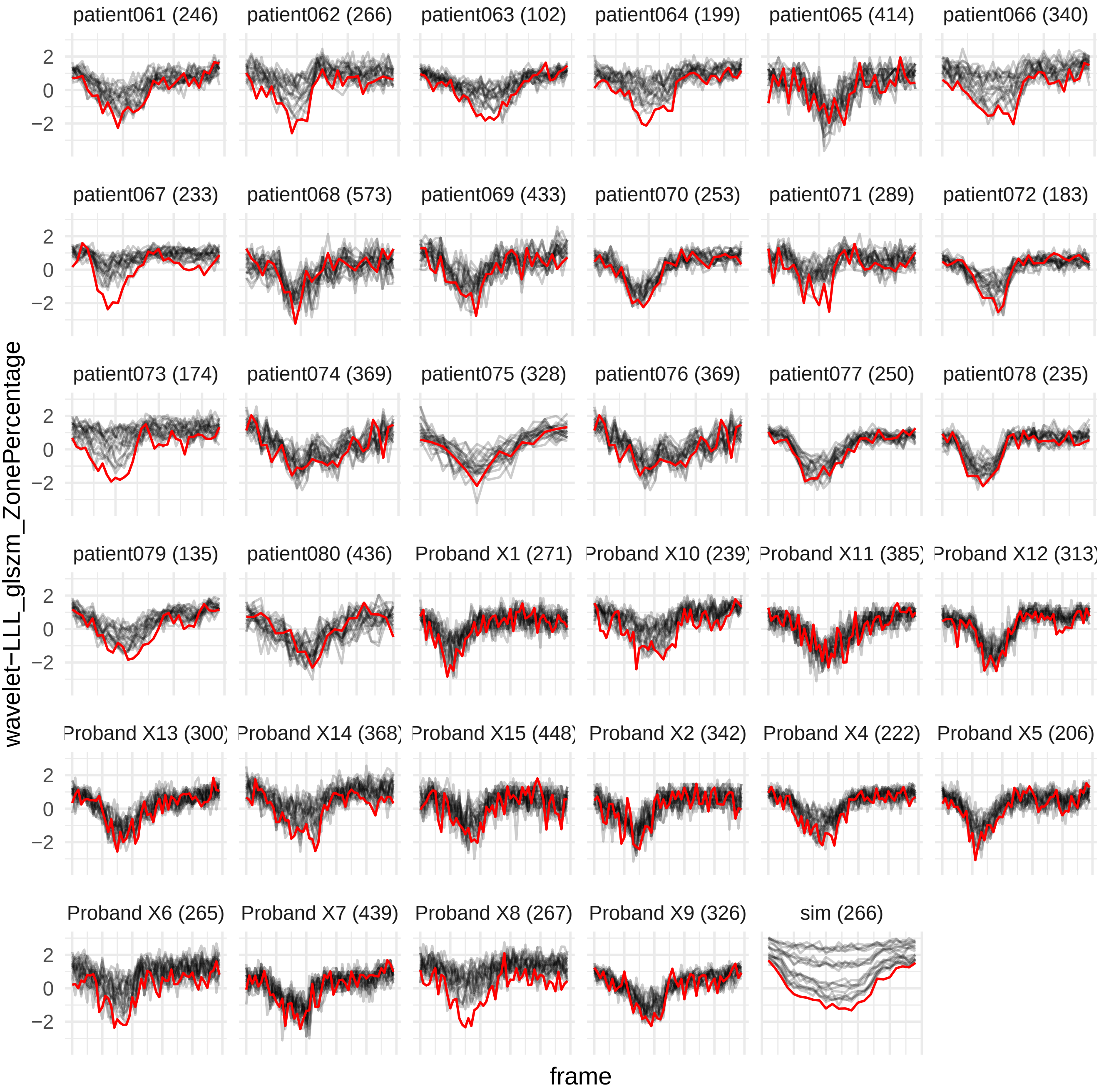

patient061 (246)
patient062 (266)
patient063 (102)
patient064 (199)
patient065 (414)
patient066 (340)
patient067 (233)
patient068 (573)
patient069 (433)
patient070 (253)
patient071 (289)
patient072 (183)
patient073 (174)
patient074 (369)
patient075 (328)
patient076 (369)
patient077 (250)
patient078 (235)
patient079 (135)
patient080 (436)
Proband X1 (271)
Proband X10 (239)
Proband X11 (385)
Proband X12 (313)
Proband X13 (300)
Proband X14 (368)
Proband X15 (448)
Proband X2 (342)
Proband X4 (222)
Proband X5 (206)
Proband X6 (265)
Proband X7 (439)
Proband X8 (267)
Proband X9 (326)
sim (266)
2
0
−2
wavelet−LLL_glszm_ZonePercentage
frame

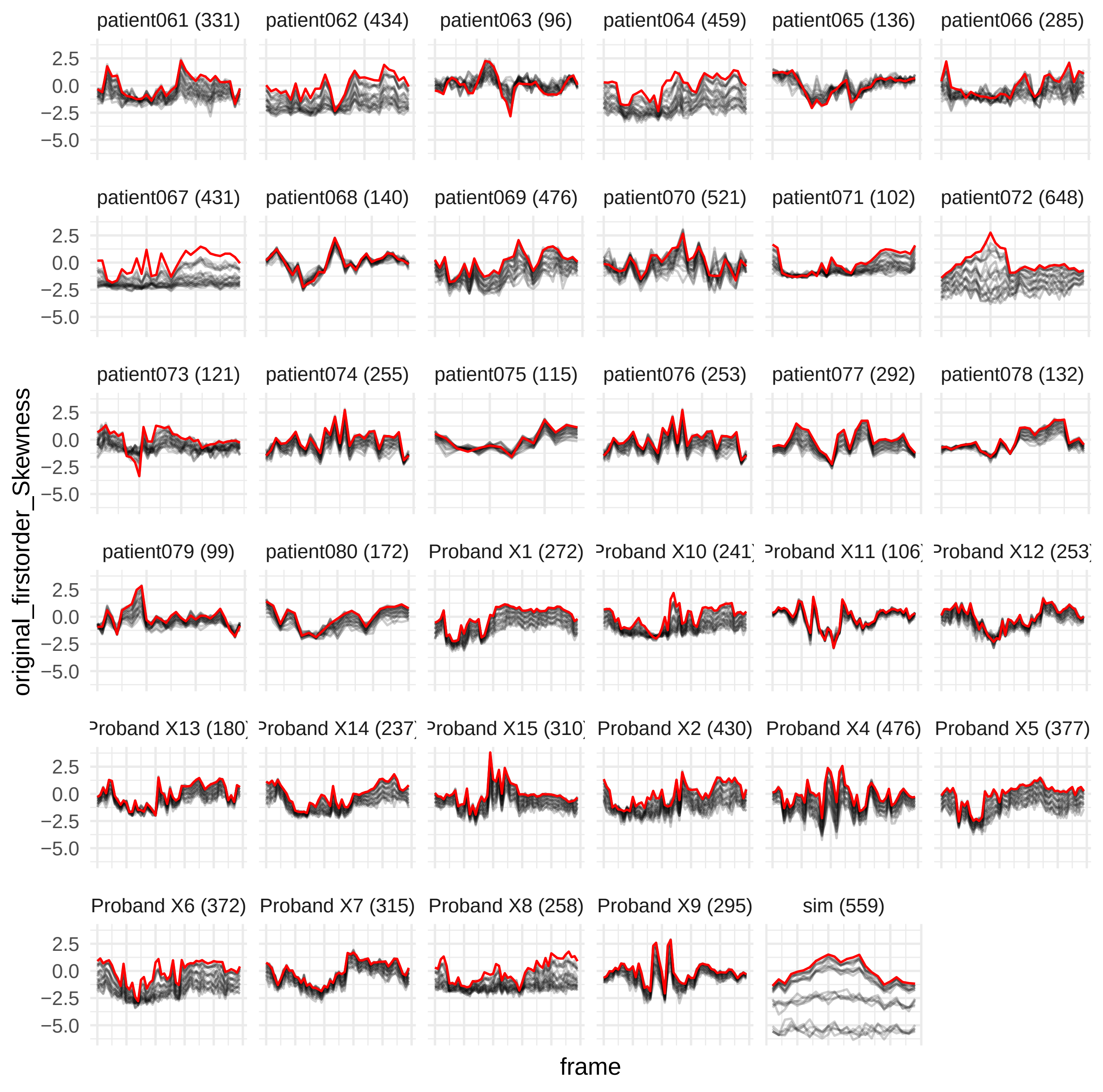
patient061 (331)
patient062 (434)
patient063 (96)
patient064 (459)
patient065 (136)
patient066 (285)
patient067 (431)
patient068 (140)
patient069 (476)
patient070 (521)
patient071 (102)
patient072 (648)
patient073 (121)
patient074 (255)
patient075 (115)
patient076 (253)
patient077 (292)
patient078 (132)
patient079 (99)
patient080 (172)
Proband X1 (272)
Proband X10 (241)
Proband X11 (106)
Proband X12 (253)
Proband X13 (180)
Proband X14 (237)
Proband X15 (310)
Proband X2 (430)
Proband X4 (476)
Proband X5 (377)
Proband X6 (372)
Proband X7 (315)
Proband X8 (258)
Proband X9 (295)
sim (559)
2.5
0.0
−2.5
−5.0
original_firstorder_Skewness
frame

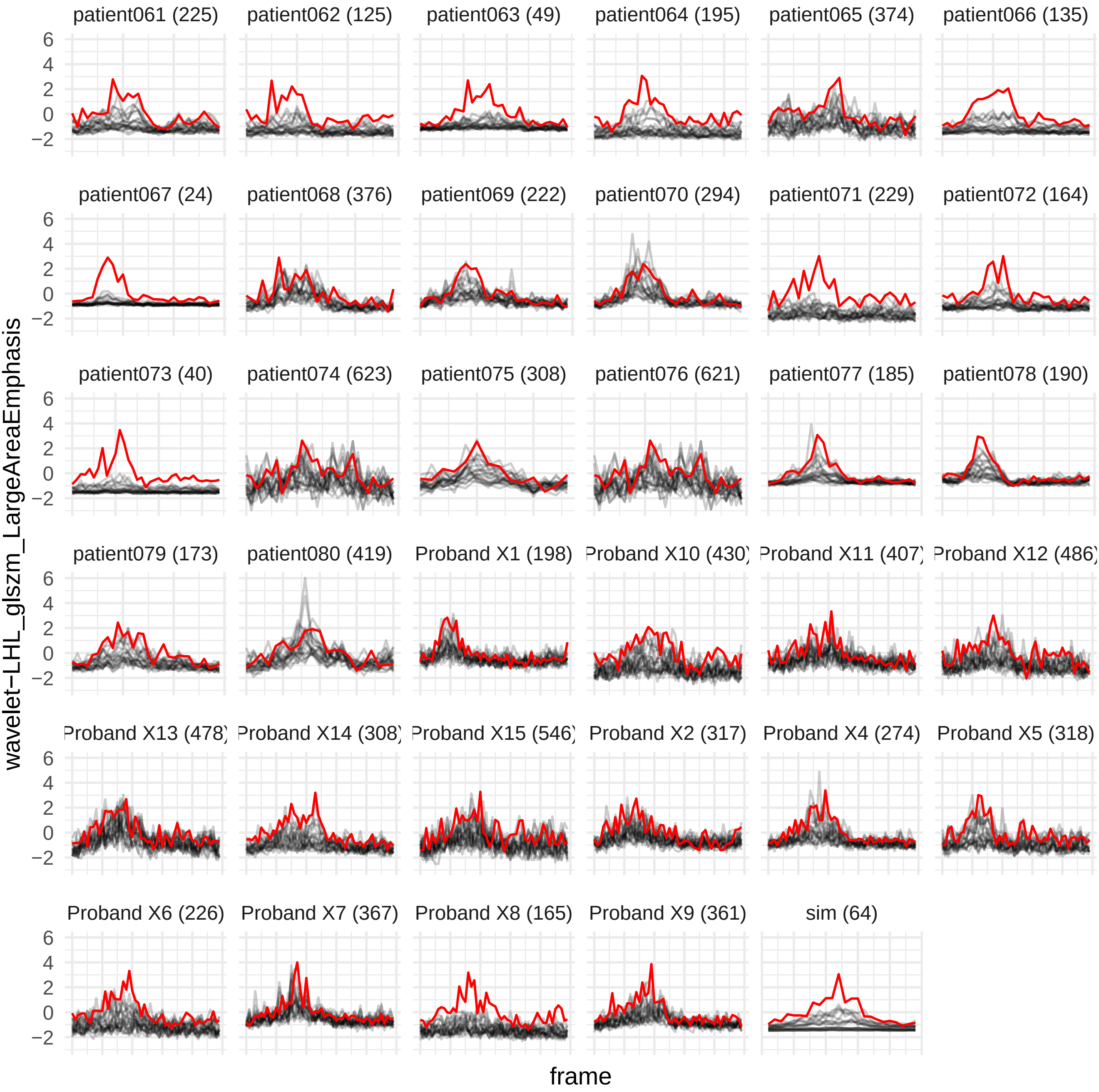

patient061 (225)
patient062 (125)
patient063 (49)
patient064 (195)
patient065 (374)
patient066 (135)
patient067 (24)
patient068 (376)
patient069 (222)
patient070 (294)
patient071 (229)
patient072 (164)
patient073 (40)
patient074 (623)
patient075 (308)
patient076 (621)
patient077 (185)
patient078 (190)
patient079 (173)
patient080 (419)
Proband X1 (198)
Proband X10 (430)
Proband X11 (407)
Proband X12 (486)
Proband X13 (478)
Proband X14 (308)
Proband X15 (546)
Proband X2 (317)
Proband X4 (274)
Proband X5 (318)
Proband X6 (226)
Proband X7 (367)
Proband X8 (165)
Proband X9 (361)
sim (64)
wavelet−LHL_glszm_LargeAreaEmphasis
frame

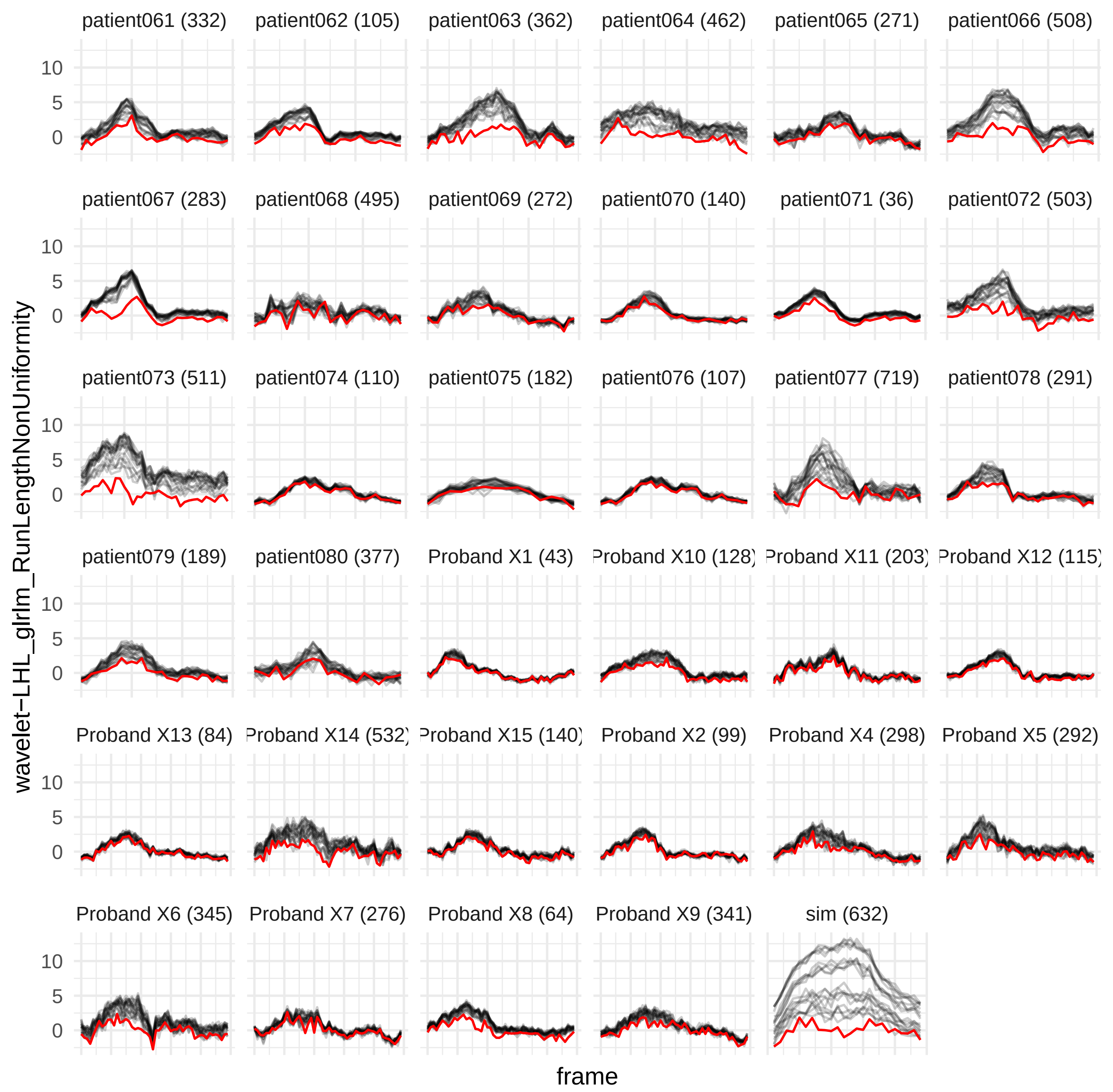
patient061 (332)
patient062 (105)
patient063 (362)
patient064 (462)
patient065 (271)
patient066 (508)
patient067 (283)
patient068 (495)
patient069 (272)
patient070 (140)
patient071 (36)
patient072 (503)
patient073 (511)
patient074 (110)
patient075 (182)
patient076 (107)
patient077 (719)
patient078 (291)
patient079 (189)
patient080 (377)
Proband X1 (43)
Proband X10 (128)
Proband X11 (203)
Proband X12 (115)
Proband X13 (84)
Proband X14 (532)
Proband X15 (140)
Proband X2 (99)
Proband X4 (298)
Proband X5 (292)
Proband X6 (345)
Proband X7 (276)
Proband X8 (64)
Proband X9 (341)
sim (632)
10
5
0
wavelet−LHL_glrlm_RunLengthNonUniformity
frame

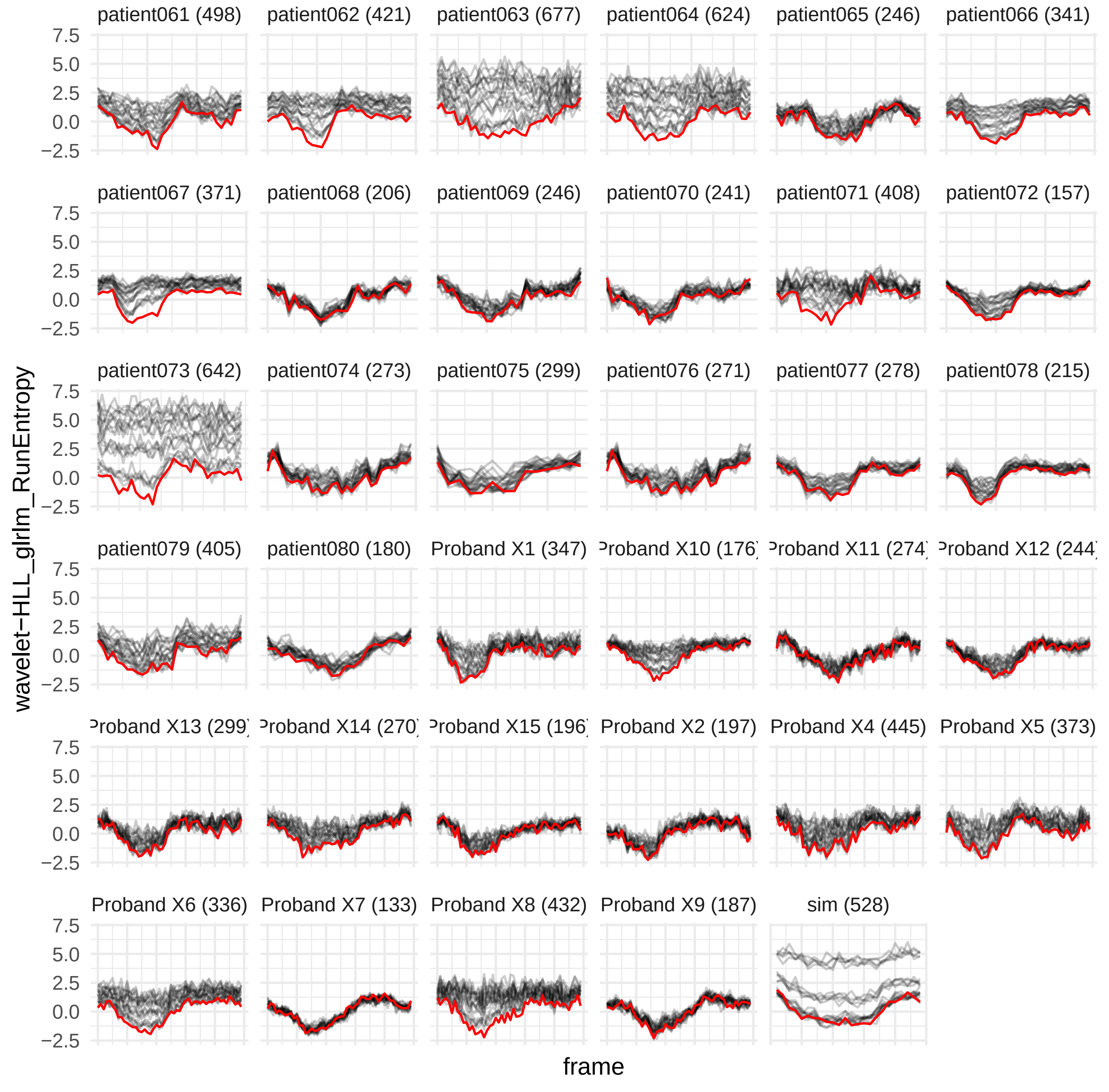

patient061 (498)
patient062 (421)
patient063 (677)
patient064 (624)
patient065 (246)
patient066 (341)
patient067 (371)
patient068 (206)
patient069 (246)
patient070 (241)
patient071 (408)
patient072 (157)
patient073 (642)
patient074 (273)
patient075 (299)
patient076 (271)
patient077 (278)
patient078 (215)
patient079 (405)
patient080 (180)
Proband X1 (347)
Proband X10 (176)
Proband X11 (274)
Proband X12 (244)
Proband X13 (299)
Proband X14 (270)
Proband X15 (196)
Proband X2 (197)
Proband X4 (445)
Proband X5 (373)
Proband X6 (336)
Proband X7 (133)
Proband X8 (432)
Proband X9 (187)
sim (528)
7.5
5.0
2.5
0.0
−2.5
wavelet−HLL_glrlm_RunEntropy
frame

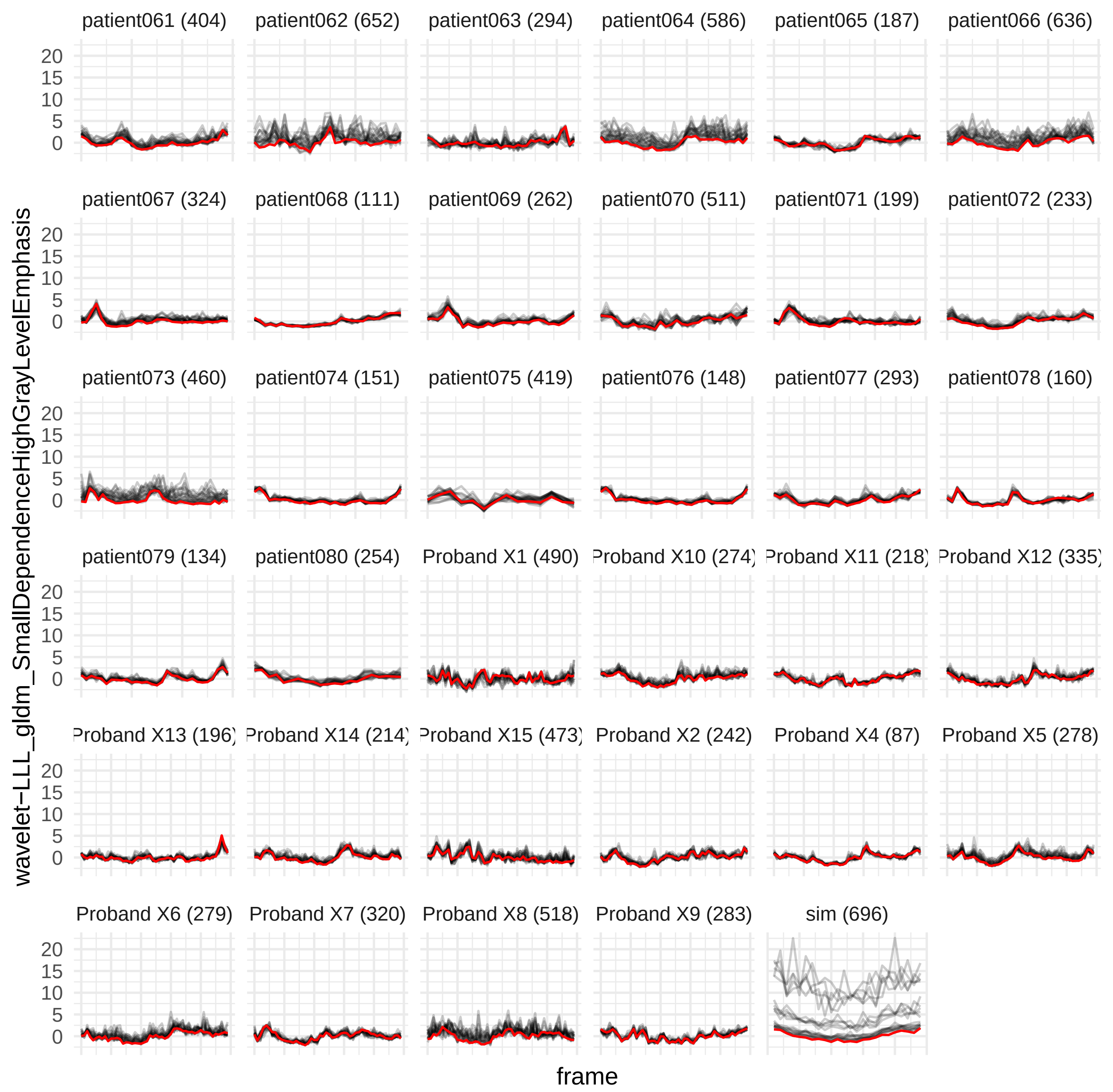
patient061 (404)
patient062 (652)
patient063 (294)
patient064 (586)
patient065 (187)
patient066 (636)
patient067 (324)
patient068 (111)
patient069 (262)
patient070 (511)
patient071 (199)
patient072 (233)
patient073 (460)
patient074 (151)
patient075 (419)
patient076 (148)
patient077 (293)
patient078 (160)
patient079 (134)
patient080 (254)
Proband X1 (490)
Proband X10 (274)
Proband X11 (218)
Proband X12 (335)
Proband X13 (196)
Proband X14 (214)
Proband X15 (473)
Proband X2 (242)
Proband X4 (87)
Proband X5 (278)
Proband X6 (279)
Proband X7 (320)
Proband X8 (518)
Proband X9 (283)
sim (696)
wavelet−LLL_gldm_SmallDependenceHighGrayLevelEmphasis
frame
0
5
10
15
20

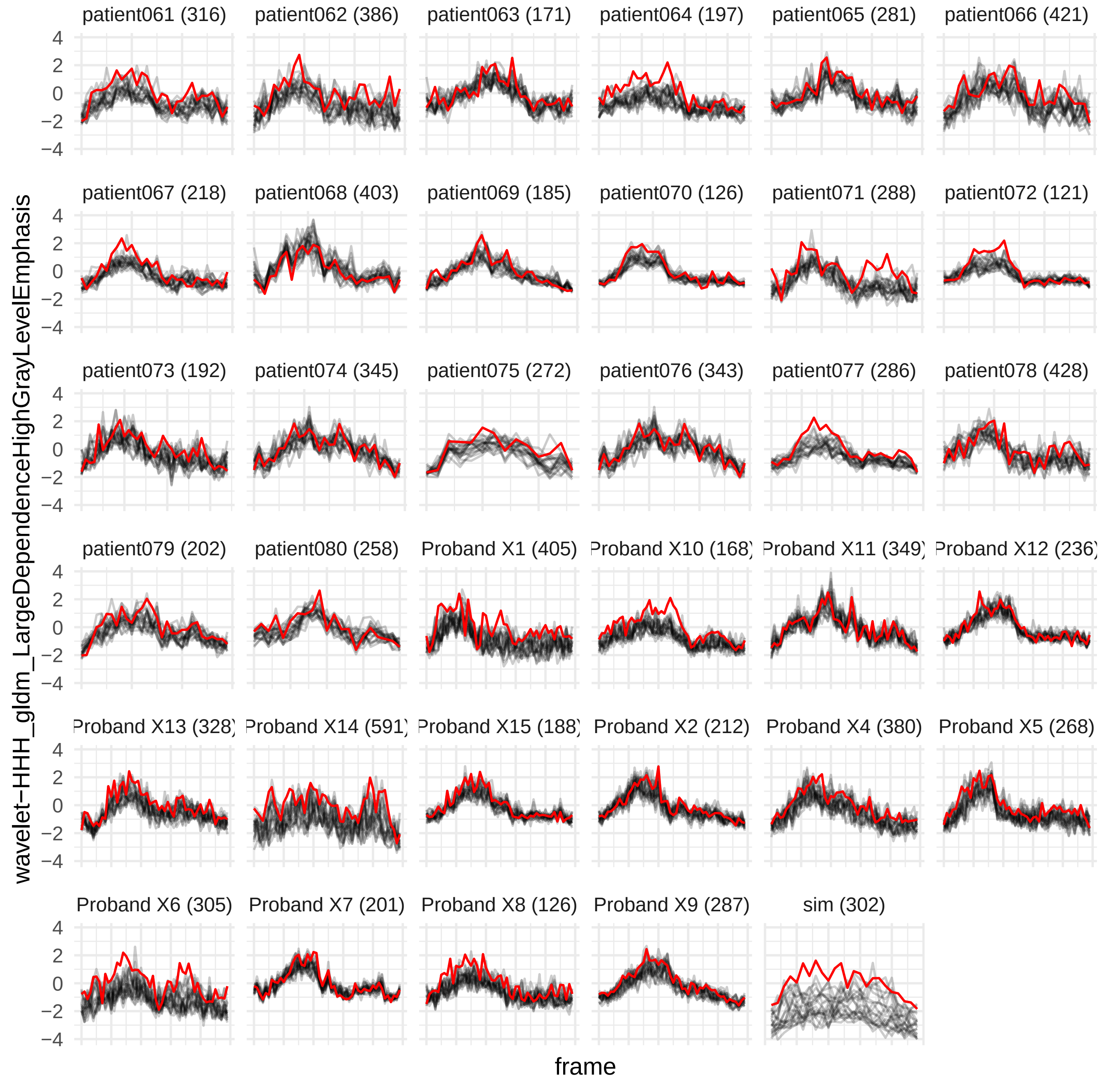

patient061 (316)
patient062 (386)
patient063 (171)
patient064 (197)
patient065 (281)
patient066 (421)
patient067 (218)
patient068 (403)
patient069 (185)
patient070 (126)
patient071 (288)
patient072 (121)
patient073 (192)
patient074 (345)
patient075 (272)
patient076 (343)
patient077 (286)
patient078 (428)
patient079 (202)
patient080 (258)
Proband X1 (405)
Proband X10 (168)
Proband X11 (349)
Proband X12 (236)
Proband X13 (328)
Proband X14 (591)
Proband X15 (188)
Proband X2 (212)
Proband X4 (380)
Proband X5 (268)
Proband X6 (305)
Proband X7 (201)
Proband X8 (126)
Proband X9 (287)
sim (302)
4
2
0
−2
−4
wavelet−HHH_gldm_LargeDependenceHighGrayLevelEmphasis
frame

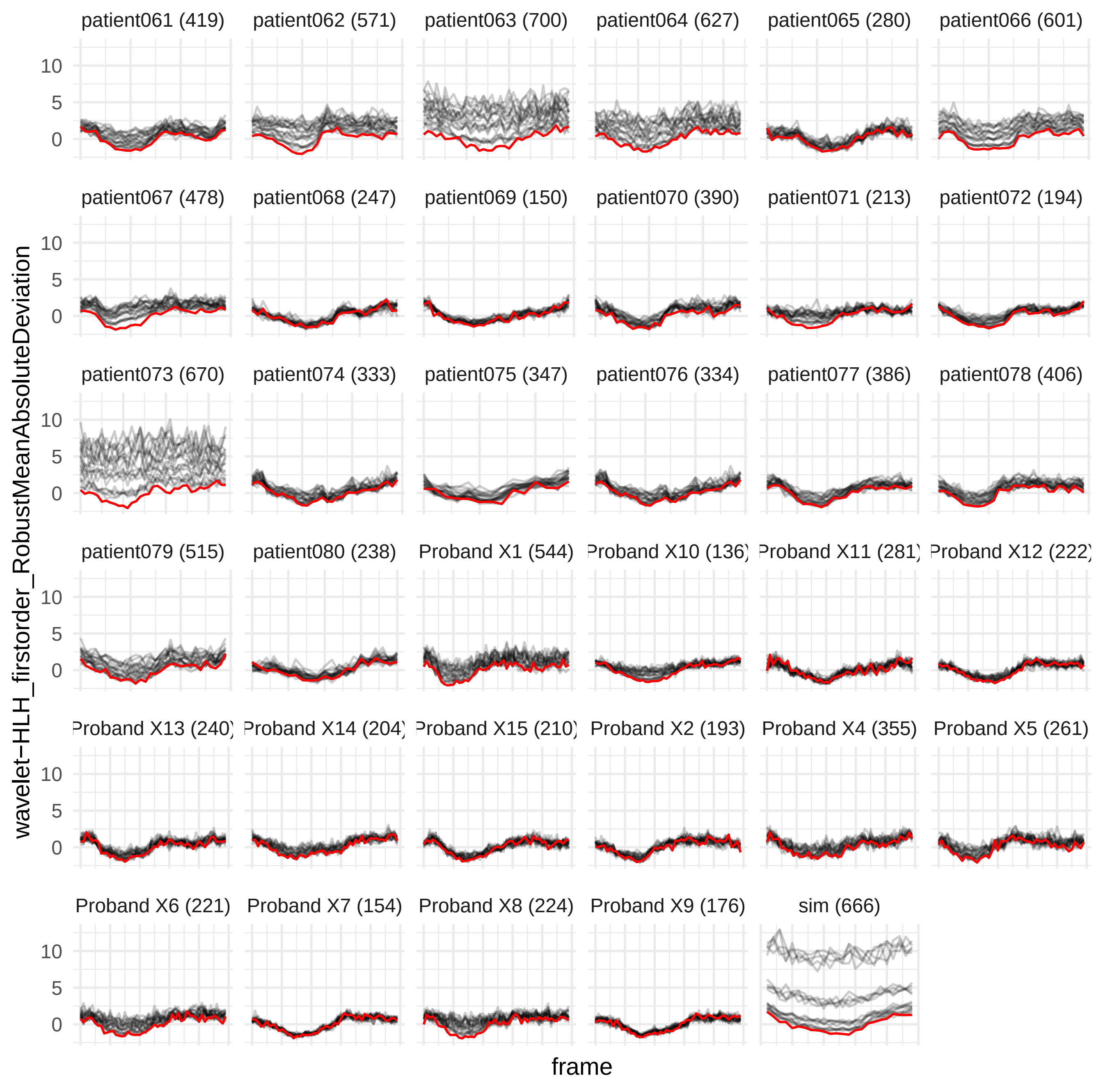
patient061 (419)
patient062 (571)
patient063 (700)
patient064 (627)
patient065 (280)
patient066 (601)
patient067 (478)
patient068 (247)
patient069 (150)
patient070 (390)
patient071 (213)
patient072 (194)
patient073 (670)
patient074 (333)
patient075 (347)
patient076 (334)
patient077 (386)
patient078 (406)
patient079 (515)
patient080 (238)
Proband X1 (544)
Proband X10 (136)
Proband X11 (281)
Proband X12 (222)
Proband X13 (240)
Proband X14 (204)
Proband X15 (210)
Proband X2 (193)
Proband X4 (355)
Proband X5 (261)
Proband X6 (221)
Proband X7 (154)
Proband X8 (224)
Proband X9 (176)
sim (666)
10
5
0
wavelet−HLH_firstorder_RobustMeanAbsoluteDeviation
frame

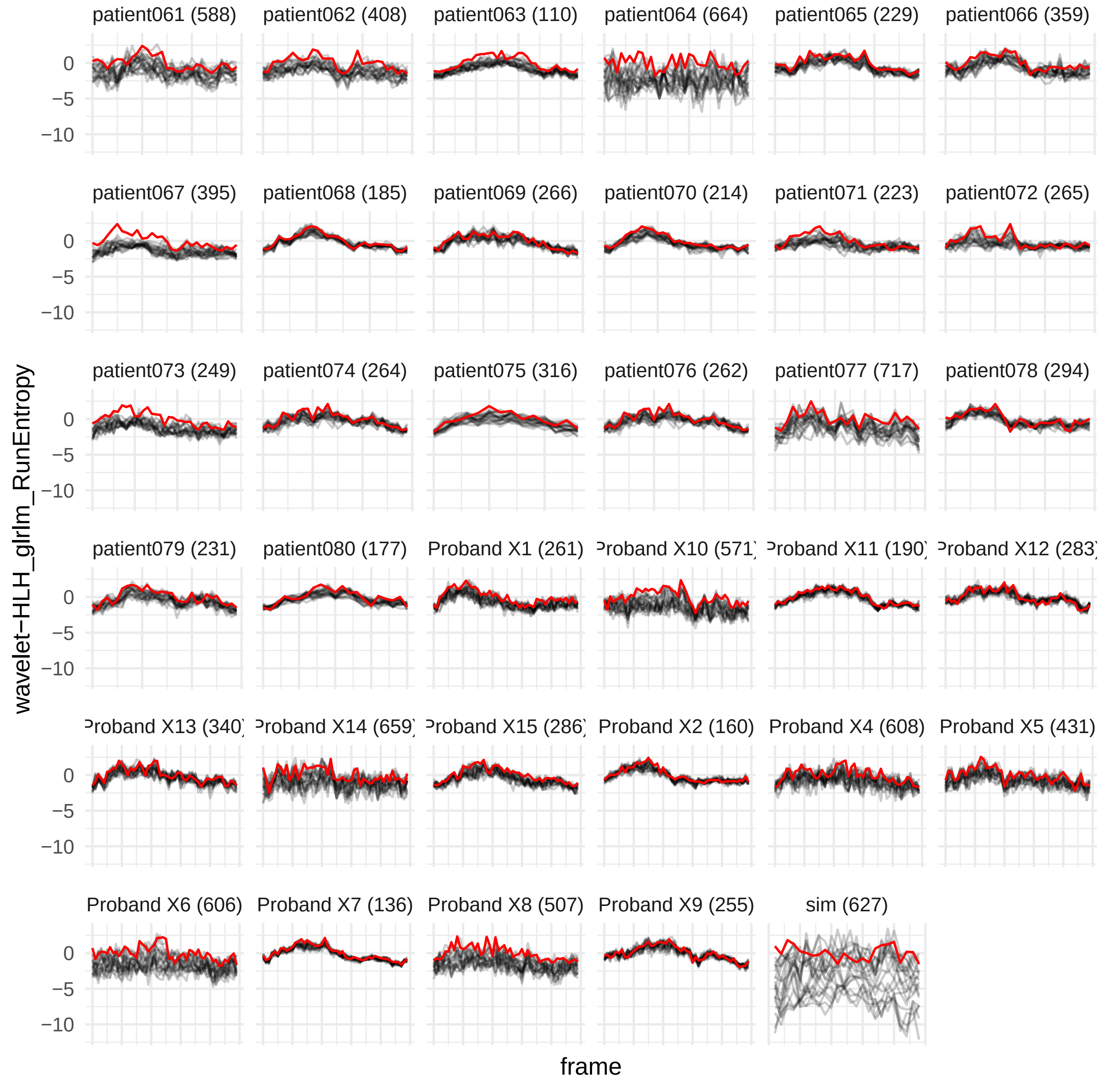

patient061 (588)
patient062 (408)
patient063 (110)
patient064 (664)
patient065 (229)
patient066 (359)
patient067 (395)
patient068 (185)
patient069 (266)
patient070 (214)
patient071 (223)
patient072 (265)
patient073 (249)
patient074 (264)
patient075 (316)
patient076 (262)
patient077 (717)
patient078 (294)
patient079 (231)
patient080 (177)
Proband X1 (261)
Proband X10 (571)
Proband X11 (190)
Proband X12 (283)
Proband X13 (340)
Proband X14 (659)
Proband X15 (286)
Proband X2 (160)
Proband X4 (608)
Proband X5 (431)
Proband X6 (606)
Proband X7 (136)
Proband X8 (507)
Proband X9 (255)
sim (627)
0
−5
−10
wavelet−HLH_glrlm_RunEntropy
frame

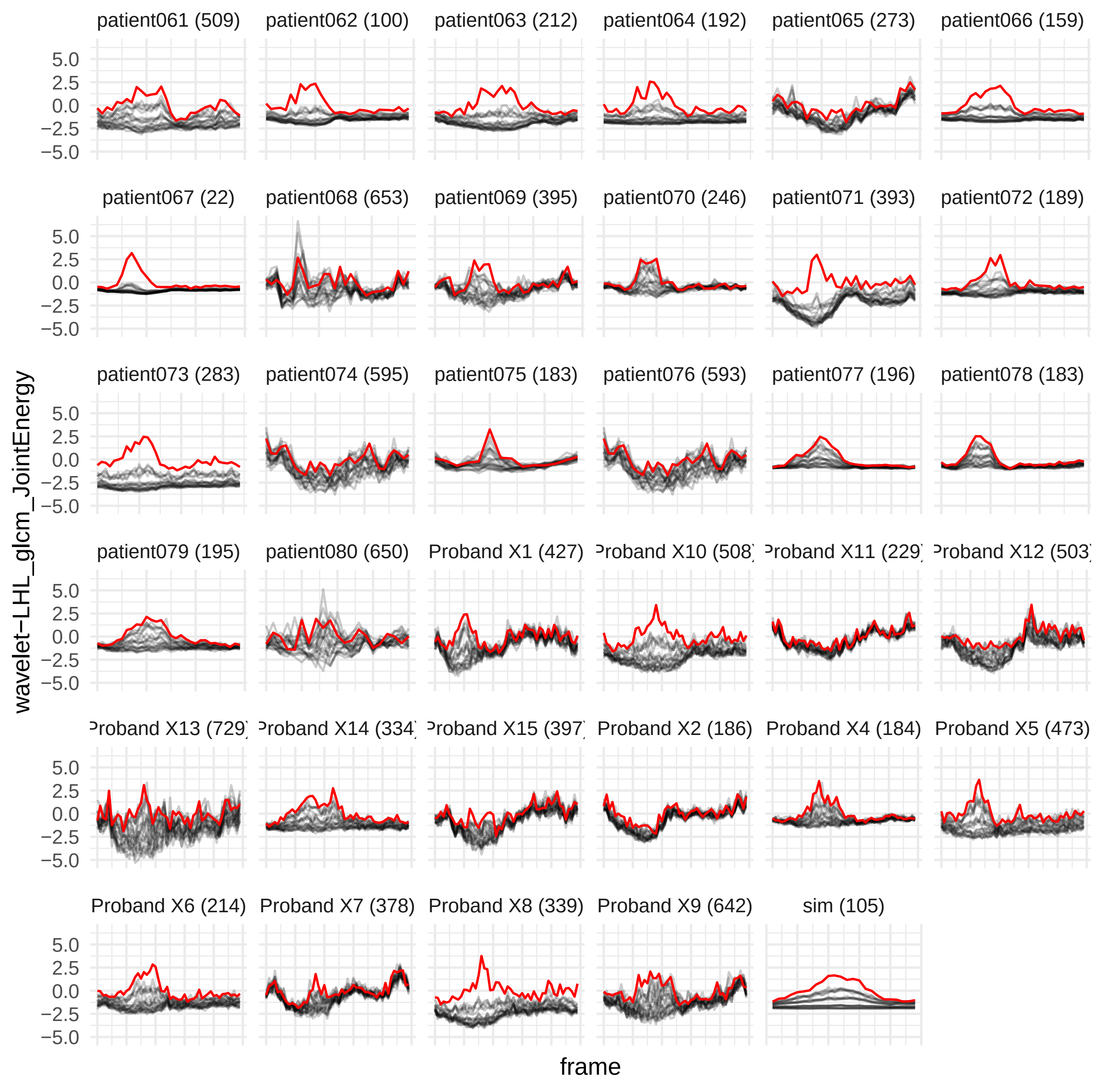
patient061 (509)
patient062 (100)
patient063 (212)
patient064 (192)
patient065 (273)
patient066 (159)
patient067 (22)
patient068 (653)
patient069 (395)
patient070 (246)
patient071 (393)
patient072 (189)
patient073 (283)
patient074 (595)
patient075 (183)
patient076 (593)
patient077 (196)
patient078 (183)
patient079 (195)
patient080 (650)
Proband X1 (427)
Proband X10 (508)
Proband X11 (229)
Proband X12 (503)
Proband X13 (729)
Proband X14 (334)
Proband X15 (397)
Proband X2 (186)
Proband X4 (184)
Proband X5 (473)
Proband X6 (214)
Proband X7 (378)
Proband X8 (339)
Proband X9 (642)
sim (105)
5.0
2.5
0.0
−2.5
−5.0
wavelet−LHL_glcm_JointEnergy
frame

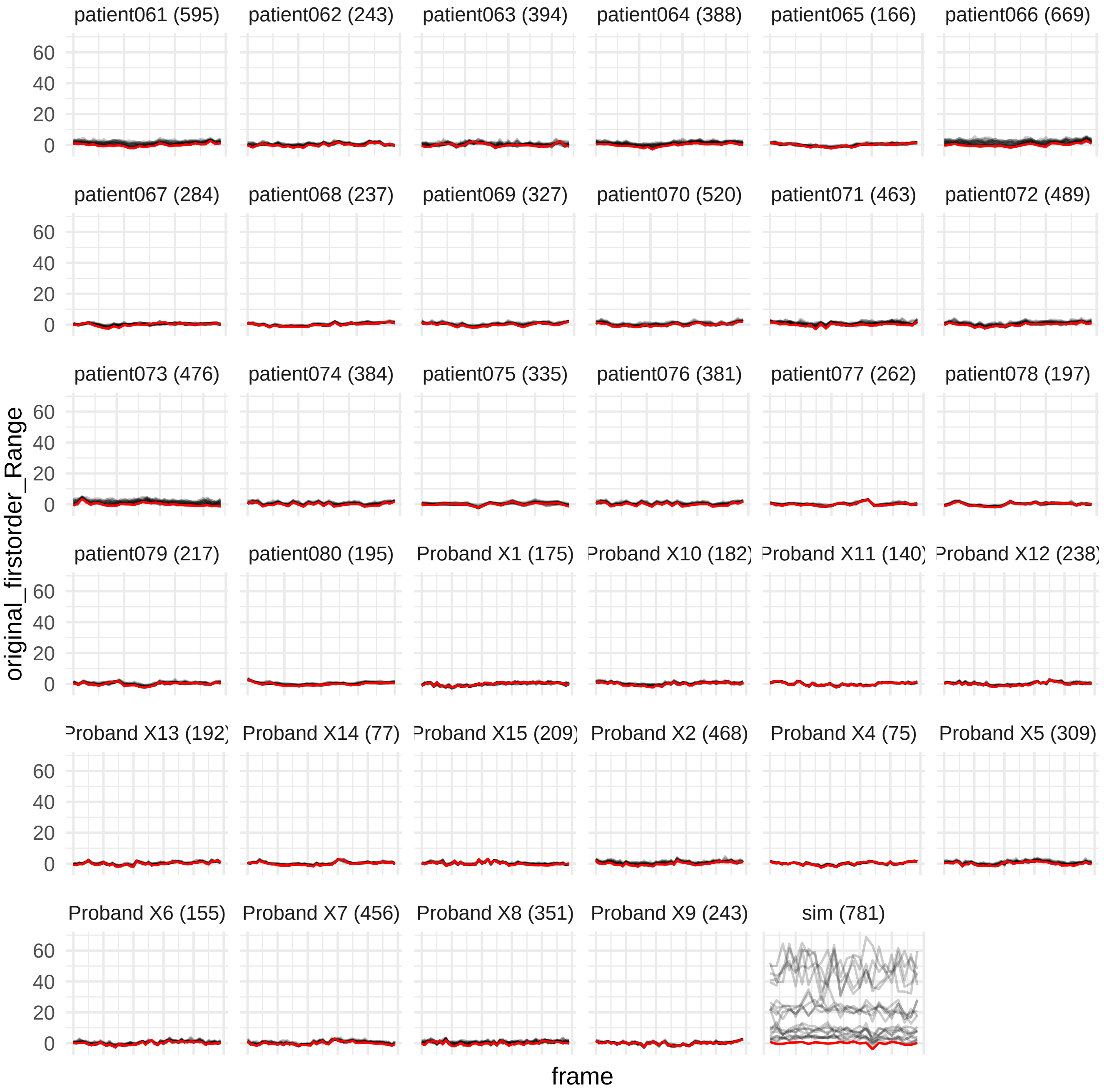

patient061 (595)
patient062 (243)
patient063 (394)
patient064 (388)
patient065 (166)
patient066 (669)
patient067 (284)
patient068 (237)
patient069 (327)
patient070 (520)
patient071 (463)
patient072 (489)
patient073 (476)
patient074 (384)
patient075 (335)
patient076 (381)
patient077 (262)
patient078 (197)
patient079 (217)
patient080 (195)
Proband X1 (175)
Proband X10 (182)
Proband X11 (140)
Proband X12 (238)
Proband X13 (192)
Proband X14 (77)
Proband X15 (209)
Proband X2 (468)
Proband X4 (75)
Proband X5 (309)
Proband X6 (155)
Proband X7 (456)
Proband X8 (351)
Proband X9 (243)
sim (781)
0
20
40
60
original_firstorder_Range
frame

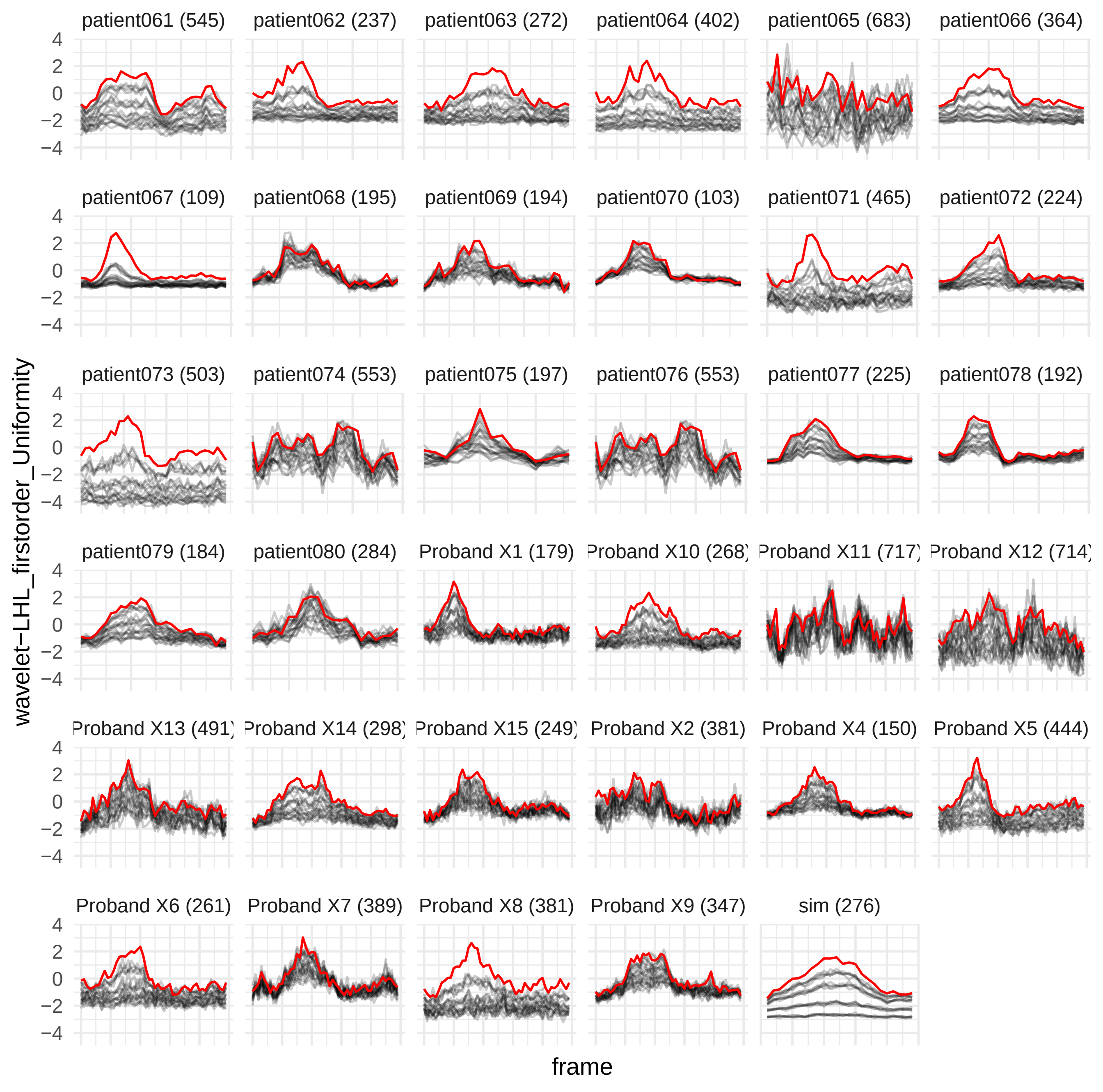

patient061 (545)
patient062 (237)
patient063 (272)
patient064 (402)
patient065 (683)
patient066 (364)
patient067 (109)
patient068 (195)
patient069 (194)
patient070 (103)
patient071 (465)
patient072 (224)
patient073 (503)
patient074 (553)
patient075 (197)
patient076 (553)
patient077 (225)
patient078 (192)
patient079 (184)
patient080 (284)
Proband X1 (179)
Proband X10 (268)
Proband X11 (717)
Proband X12 (714)
Proband X13 (491)
Proband X14 (298)
Proband X15 (249)
Proband X2 (381)
Proband X4 (150)
Proband X5 (444)
Proband X6 (261)
Proband X7 (389)
Proband X8 (381)
Proband X9 (347)
sim (276)
wavelet−LHL_firstorder_Uniformity
frame
4
2
0
−2
−4

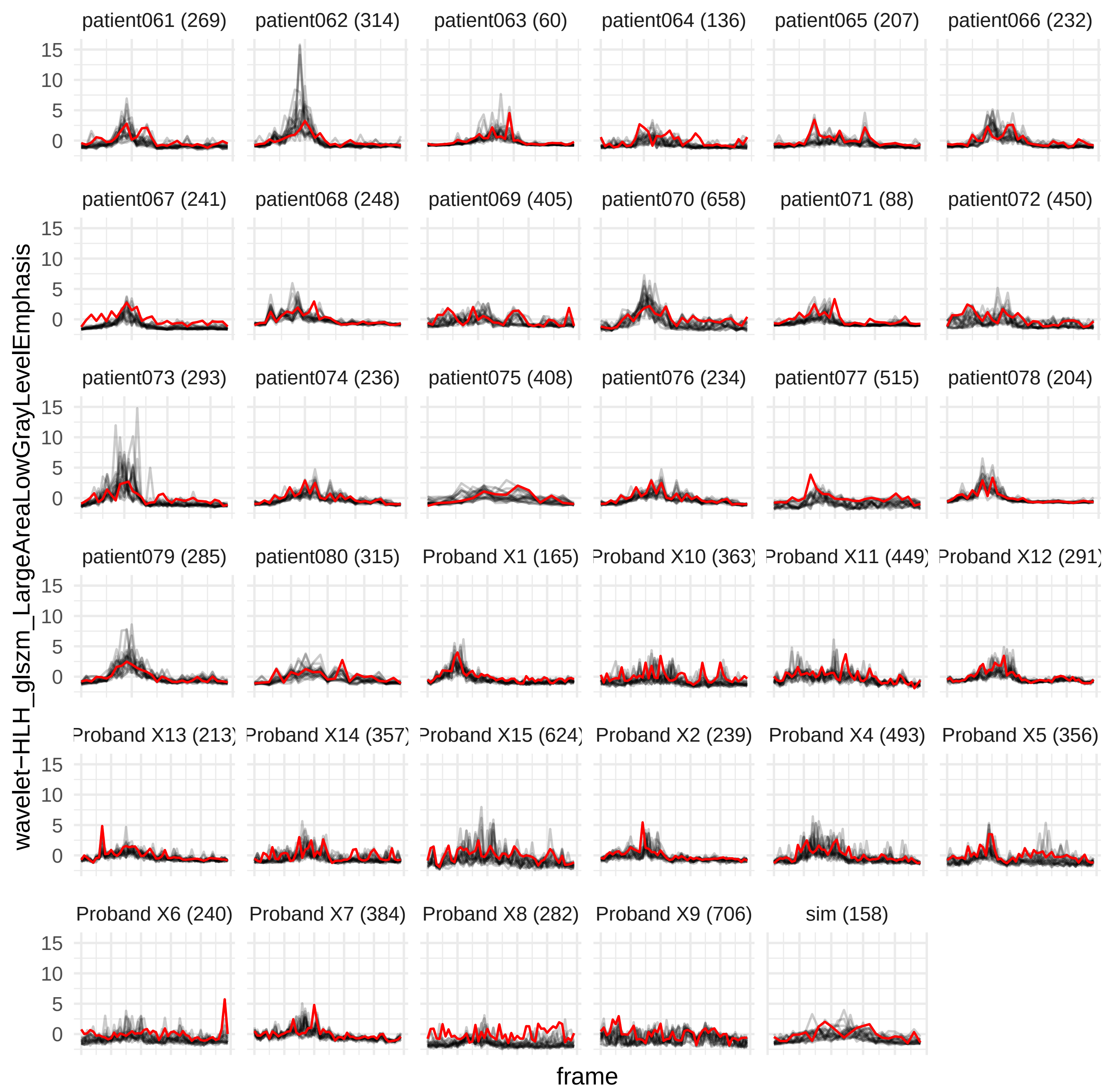

patient061 (269)
patient062 (314)
patient063 (60)
patient064 (136)
patient065 (207)
patient066 (232)
patient067 (241)
patient068 (248)
patient069 (405)
patient070 (658)
patient071 (88)
patient072 (450)
patient073 (293)
patient074 (236)
patient075 (408)
patient076 (234)
patient077 (515)
patient078 (204)
patient079 (285)
patient080 (315)
Proband X1 (165)
Proband X10 (363)
Proband X11 (449)
Proband X12 (291)
Proband X13 (213)
Proband X14 (357)
Proband X15 (624)
Proband X2 (239)
Proband X4 (493)
Proband X5 (356)
Proband X6 (240)
Proband X7 (384)
Proband X8 (282)
Proband X9 (706)
sim (158)
wavelet−HLH_glszm_LargeAreaLowGrayLevelEmphasis
frame
0
5
10
15

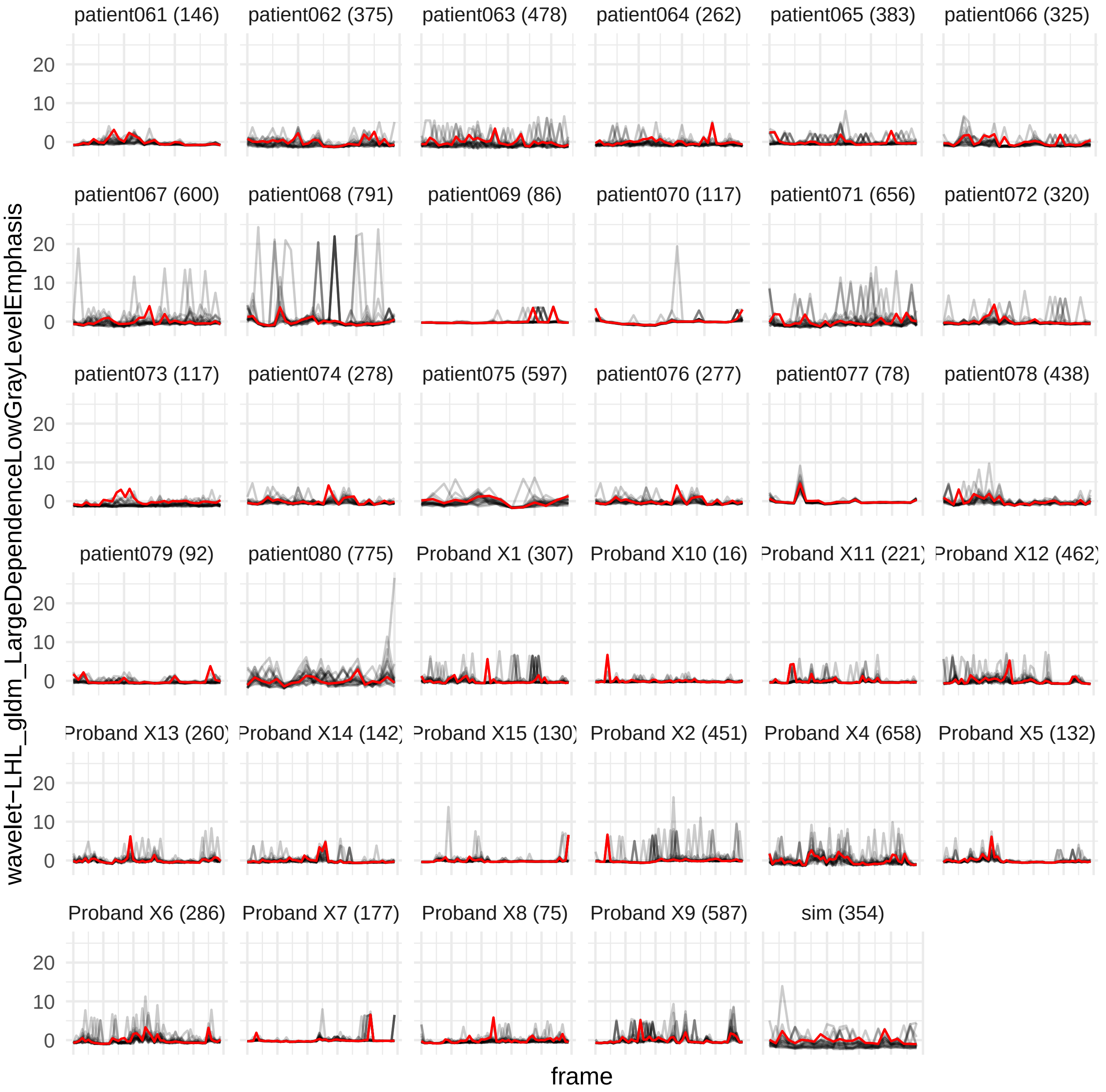
patient061 (146)
patient062 (375)
patient063 (478)
patient064 (262)
patient065 (383)
patient066 (325)
patient067 (600)
patient068 (791)
patient069 (86)
patient070 (117)
patient071 (656)
patient072 (320)
patient073 (117)
patient074 (278)
patient075 (597)
patient076 (277)
patient077 (78)
patient078 (438)
patient079 (92)
patient080 (775)
Proband X1 (307)
Proband X10 (16)
Proband X11 (221)
Proband X12 (462)
Proband X13 (260)
Proband X14 (142)
Proband X15 (130)
Proband X2 (451)
Proband X4 (658)
Proband X5 (132)
Proband X6 (286)
Proband X7 (177)
Proband X8 (75)
Proband X9 (587)
sim (354)
20
10
0
wavelet−LHL_gldm_LargeDependenceLowGrayLevelEmphasis
frame

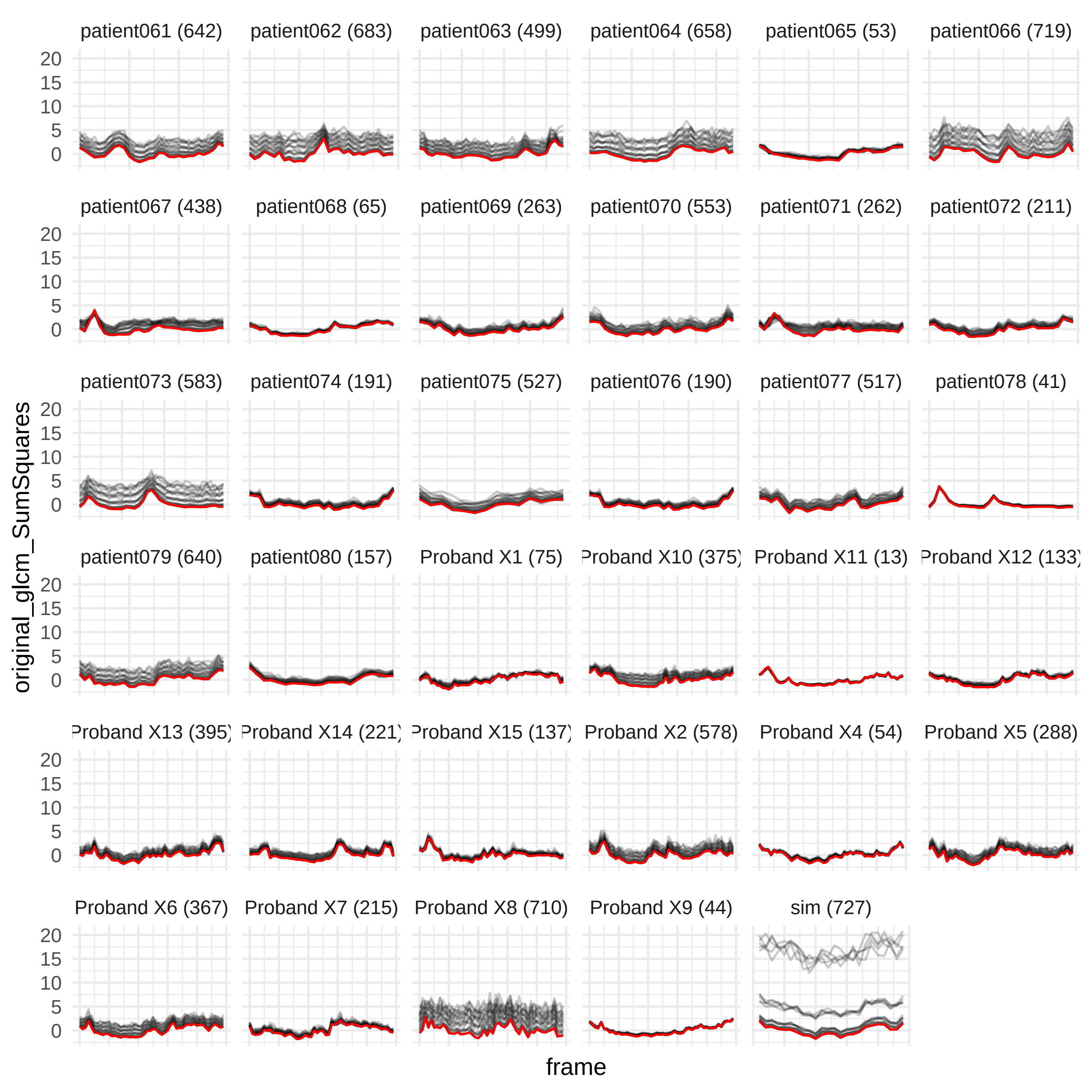
patient061 (642)
patient062 (683)
patient063 (499)
patient064 (658)
patient065 (53)
patient066 (719)
patient067 (438)
patient068 (65)
patient069 (263)
patient070 (553)
patient071 (262)
patient072 (211)
patient073 (583)
patient074 (191)
patient075 (527)
patient076 (190)
patient077 (517)
patient078 (41)
patient079 (640)
patient080 (157)
Proband X1 (75)
Proband X10 (375)
Proband X11 (13)
Proband X12 (133)
Proband X13 (395)
Proband X14 (221)
Proband X15 (137)
Proband X2 (578)
Proband X4 (54)
Proband X5 (288)
Proband X6 (367)
Proband X7 (215)
Proband X8 (710)
Proband X9 (44)
sim (727)
original_glcm_SumSquares
frame
0
5
10
15
20

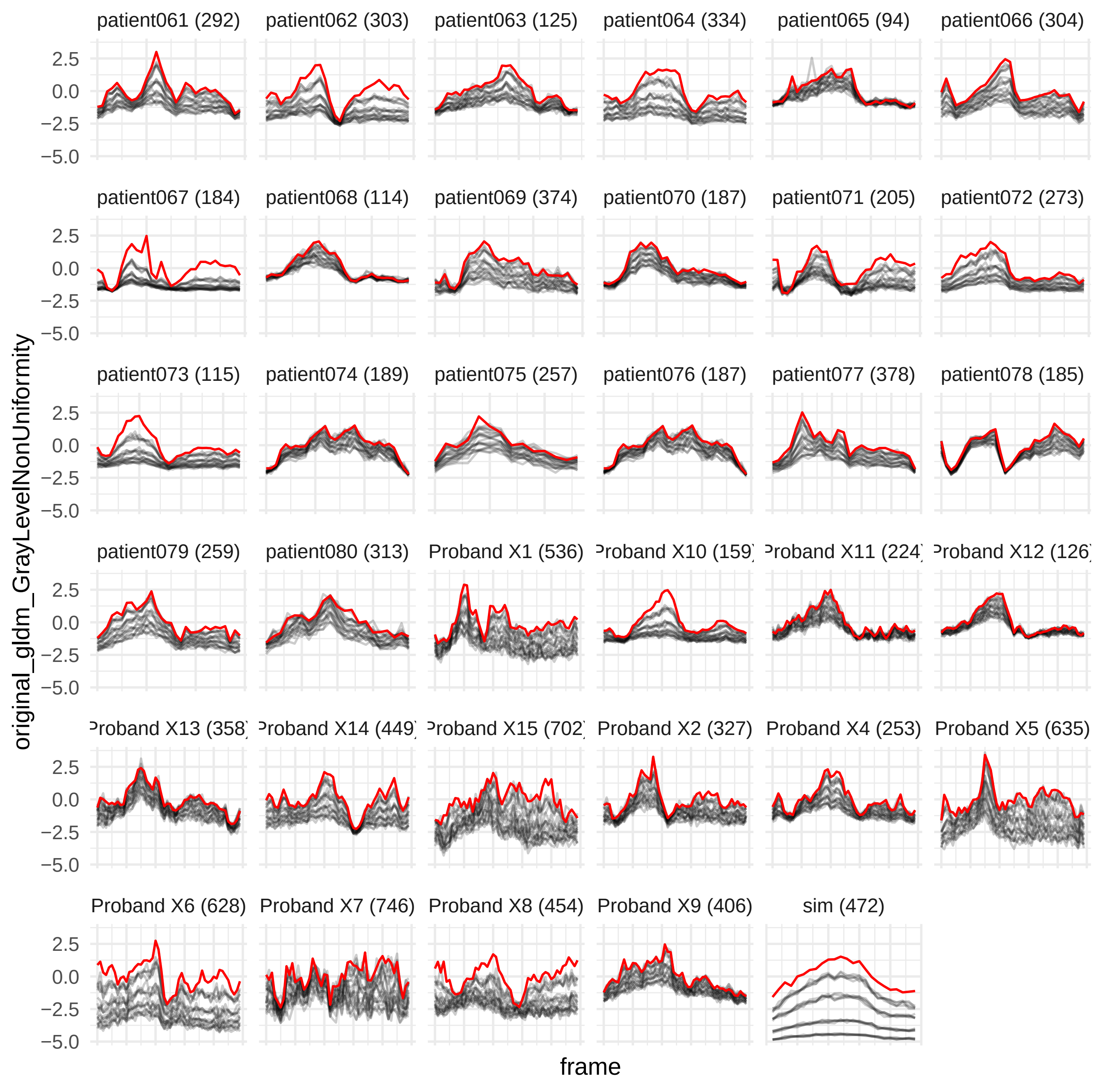

patient061 (292)
patient062 (303)
patient063 (125)
patient064 (334)
patient065 (94)
patient066 (304)
patient067 (184)
patient068 (114)
patient069 (374)
patient070 (187)
patient071 (205)
patient072 (273)
patient073 (115)
patient074 (189)
patient075 (257)
patient076 (187)
patient077 (378)
patient078 (185)
patient079 (259)
patient080 (313)
Proband X1 (536)
Proband X10 (159)
Proband X11 (224)
Proband X12 (126)
Proband X13 (358)
Proband X14 (449)
Proband X15 (702)
Proband X2 (327)
Proband X4 (253)
Proband X5 (635)
Proband X6 (628)
Proband X7 (746)
Proband X8 (454)
Proband X9 (406)
sim (472)
2.5
0.0
−2.5
−5.0
original_gldm_GrayLevelNonUniformity
frame

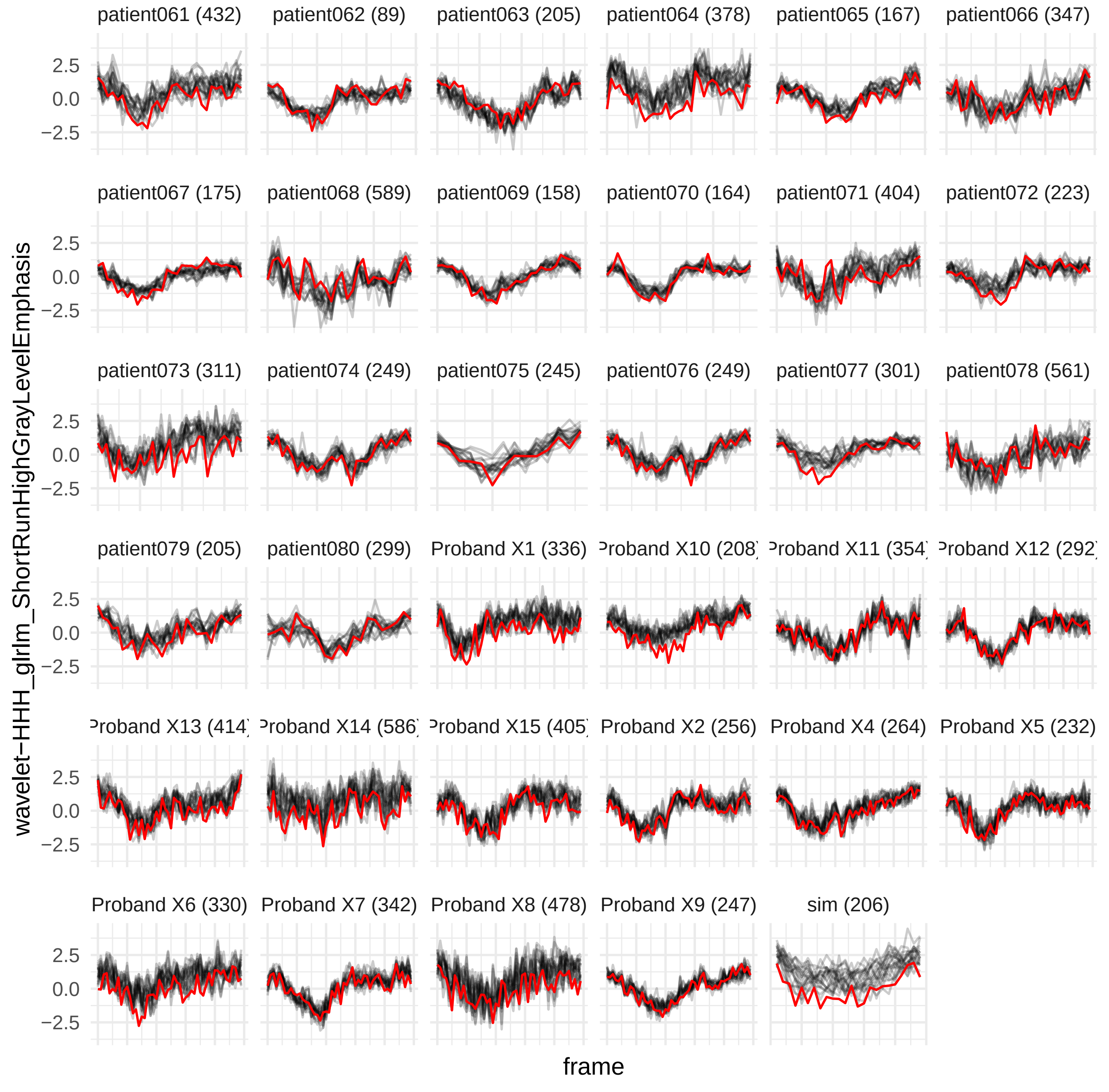

patient061 (432)
patient062 (89)
patient063 (205)
patient064 (378)
patient065 (167)
patient066 (347)
patient067 (175)
patient068 (589)
patient069 (158)
patient070 (164)
patient071 (404)
patient072 (223)
patient073 (311)
patient074 (249)
patient075 (245)
patient076 (249)
patient077 (301)
patient078 (561)
patient079 (205)
patient080 (299)
Proband X1 (336)
Proband X10 (208)
Proband X11 (354)
Proband X12 (292)
Proband X13 (414)
Proband X14 (586)
Proband X15 (405)
Proband X2 (256)
Proband X4 (264)
Proband X5 (232)
Proband X6 (330)
Proband X7 (342)
Proband X8 (478)
Proband X9 (247)
sim (206)
2.5
0.0
−2.5
wavelet−HHH_glrlm_ShortRunHighGrayLevelEmphasis
frame

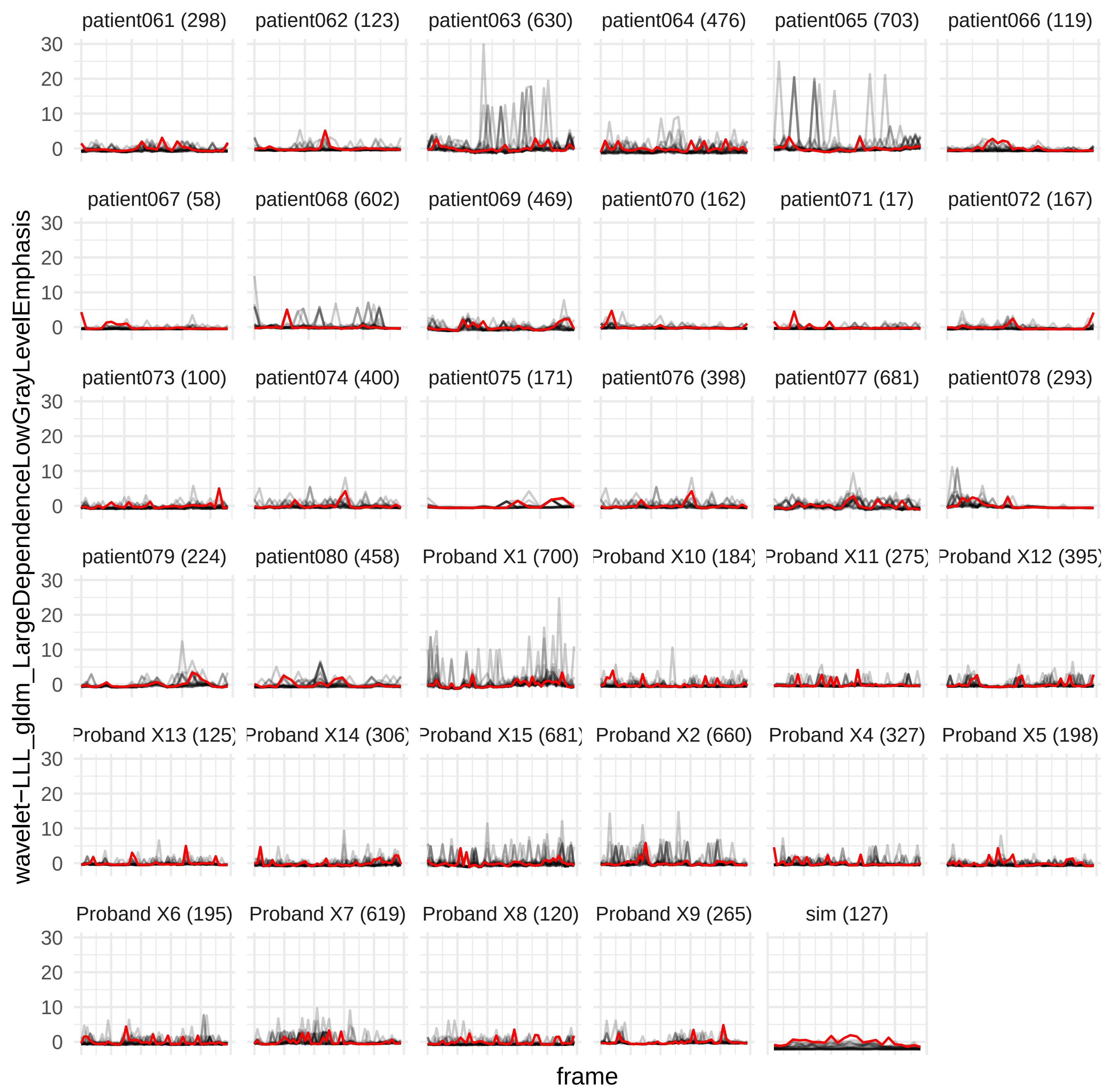

patient061 (298)
patient062 (123)
patient063 (630)
patient064 (476)
patient065 (703)
patient066 (119)
patient067 (58)
patient068 (602)
patient069 (469)
patient070 (162)
patient071 (17)
patient072 (167)
patient073 (100)
patient074 (400)
patient075 (171)
patient076 (398)
patient077 (681)
patient078 (293)
patient079 (224)
patient080 (458)
Proband X1 (700)
Proband X10 (184)
Proband X11 (275)
Proband X12 (395)
Proband X13 (125)
Proband X14 (306)
Proband X15 (681)
Proband X2 (660)
Proband X4 (327)
Proband X5 (198)
Proband X6 (195)
Proband X7 (619)
Proband X8 (120)
Proband X9 (265)
sim (127)
wavelet−LLL_gldm_LargeDependenceLowGrayLevelEmphasis
0
10
20
30
frame

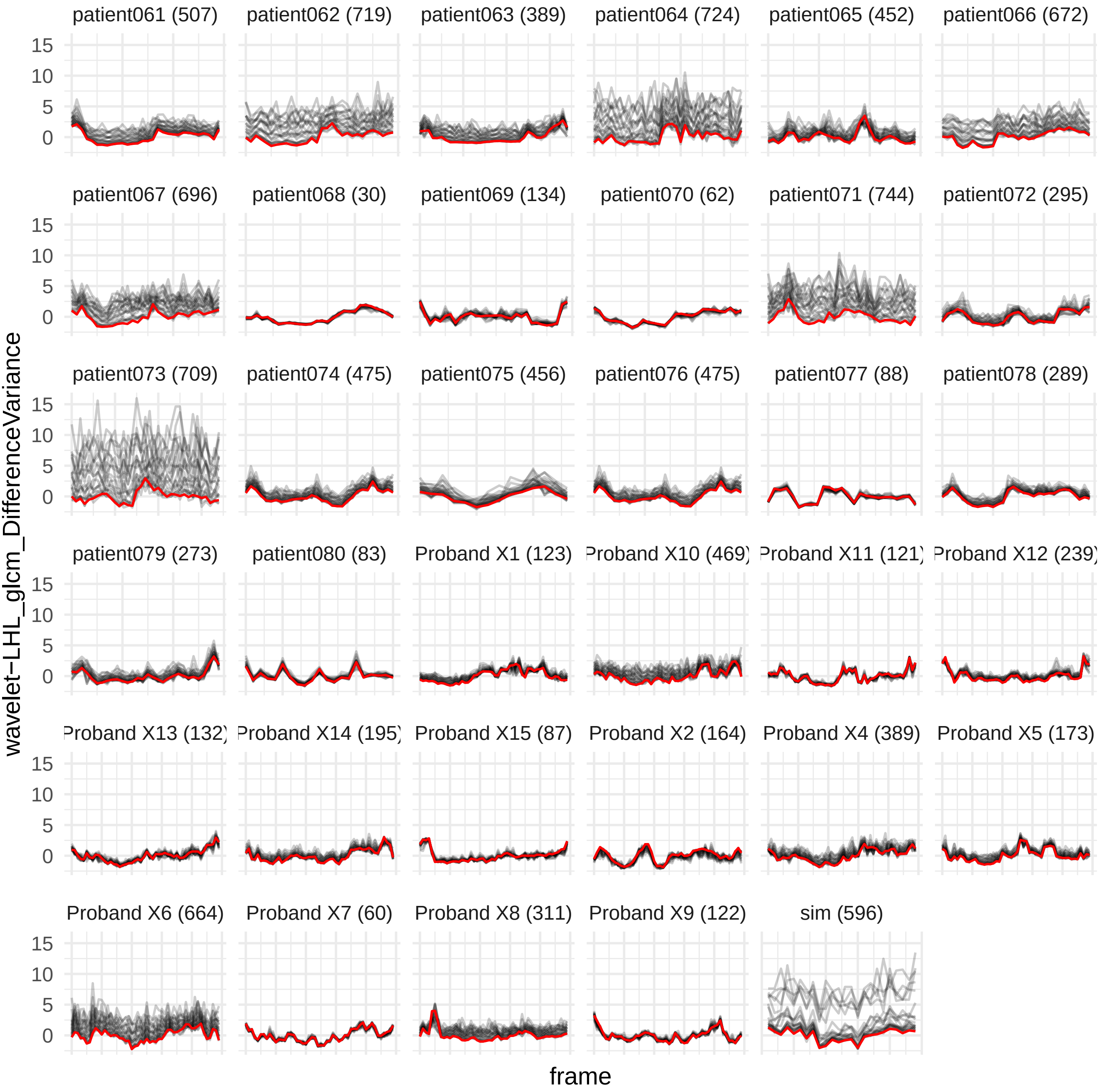
patient061 (507)
patient062 (719)
patient063 (389)
patient064 (724)
patient065 (452)
patient066 (672)
patient067 (696)
patient068 (30)
patient069 (134)
patient070 (62)
patient071 (744)
patient072 (295)
patient073 (709)
patient074 (475)
patient075 (456)
patient076 (475)
patient077 (88)
patient078 (289)
patient079 (273)
patient080 (83)
Proband X1 (123)
Proband X10 (469)
Proband X11 (121)
Proband X12 (239)
Proband X13 (132)
Proband X14 (195)
Proband X15 (87)
Proband X2 (164)
Proband X4 (389)
Proband X5 (173)
Proband X6 (664)
Proband X7 (60)
Proband X8 (311)
Proband X9 (122)
sim (596)
15
10
5
0
wavelet−LHL_glcm_DifferenceVariance
frame

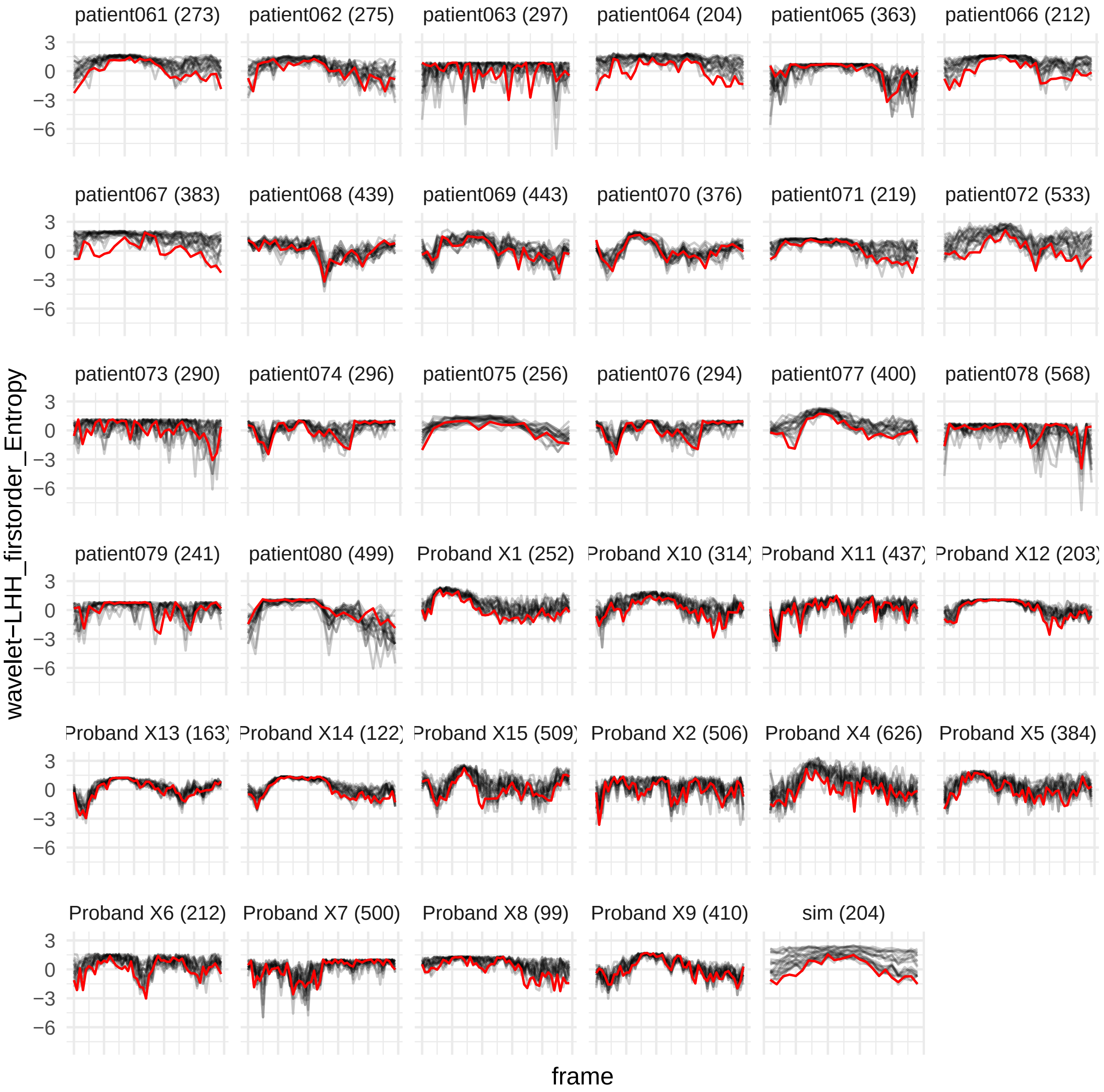
patient061 (273)
patient062 (275)
patient063 (297)
patient064 (204)
patient065 (363)
patient066 (212)
patient067 (383)
patient068 (439)
patient069 (443)
patient070 (376)
patient071 (219)
patient072 (533)
patient073 (290)
patient074 (296)
patient075 (256)
patient076 (294)
patient077 (400)
patient078 (568)
patient079 (241)
patient080 (499)
Proband X1 (252)
Proband X10 (314)
Proband X11 (437)
Proband X12 (203)
Proband X13 (163)
Proband X14 (122)
Proband X15 (509)
Proband X2 (506)
Proband X4 (626)
Proband X5 (384)
Proband X6 (212)
Proband X7 (500)
Proband X8 (99)
Proband X9 (410)
sim (204)
3
0
−3
−6
wavelet−LHH_firstorder_Entropy
frame

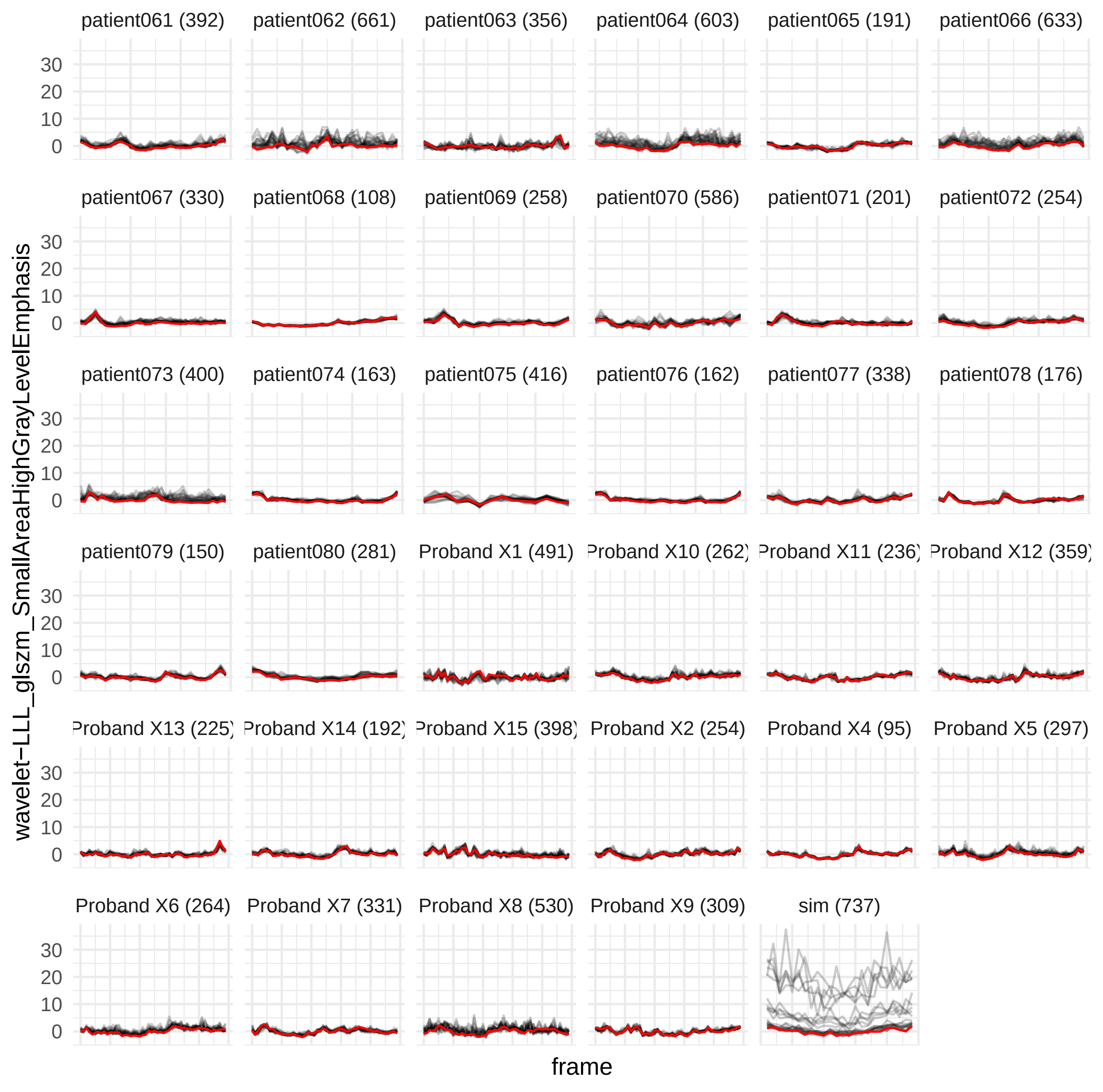

patient061 (392)
patient062 (661)
patient063 (356)
patient064 (603)
patient065 (191)
patient066 (633)
patient067 (330)
patient068 (108)
patient069 (258)
patient070 (586)
patient071 (201)
patient072 (254)
patient073 (400)
patient074 (163)
patient075 (416)
patient076 (162)
patient077 (338)
patient078 (176)
patient079 (150)
patient080 (281)
Proband X1 (491)
Proband X10 (262)
Proband X11 (236)
Proband X12 (359)
Proband X13 (225)
Proband X14 (192)
Proband X15 (398)
Proband X2 (254)
Proband X4 (95)
Proband X5 (297)
Proband X6 (264)
Proband X7 (331)
Proband X8 (530)
Proband X9 (309)
sim (737)
0
10
20
30
wavelet−LLL_glszm_SmallAreaHighGrayLevelEmphasis
frame

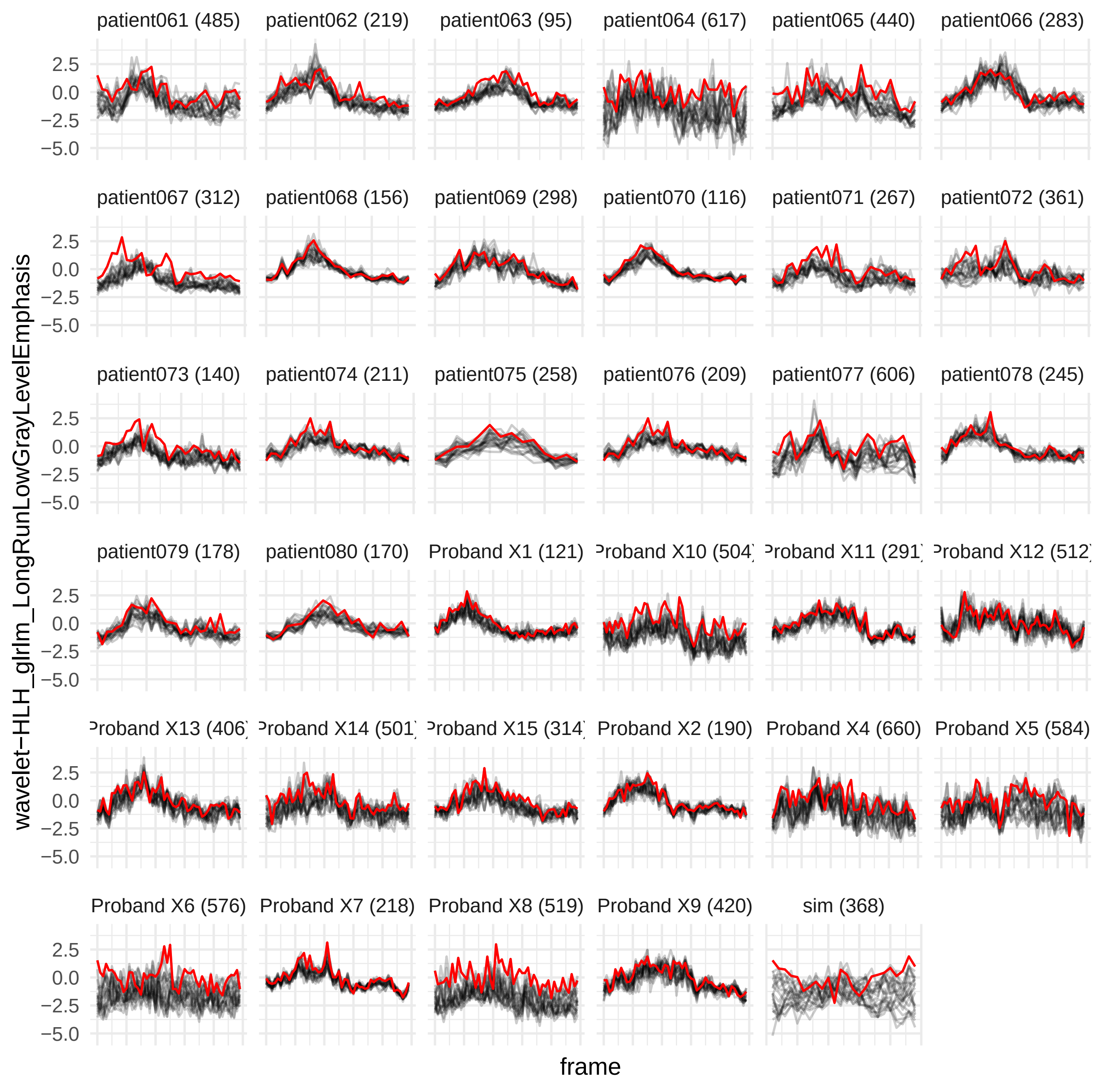

patient061 (485)
patient062 (219)
patient063 (95)
patient064 (617)
patient065 (440)
patient066 (283)
patient067 (312)
patient068 (156)
patient069 (298)
patient070 (116)
patient071 (267)
patient072 (361)
patient073 (140)
patient074 (211)
patient075 (258)
patient076 (209)
patient077 (606)
patient078 (245)
patient079 (178)
patient080 (170)
Proband X1 (121)
Proband X10 (504)
Proband X11 (291)
Proband X12 (512)
Proband X13 (406)
Proband X14 (501)
Proband X15 (314)
Proband X2 (190)
Proband X4 (660)
Proband X5 (584)
Proband X6 (576)
Proband X7 (218)
Proband X8 (519)
Proband X9 (420)
sim (368)
2.5
0.0
−2.5
−5.0
wavelet−HLH_glrlm_LongRunLowGrayLevelEmphasis
frame

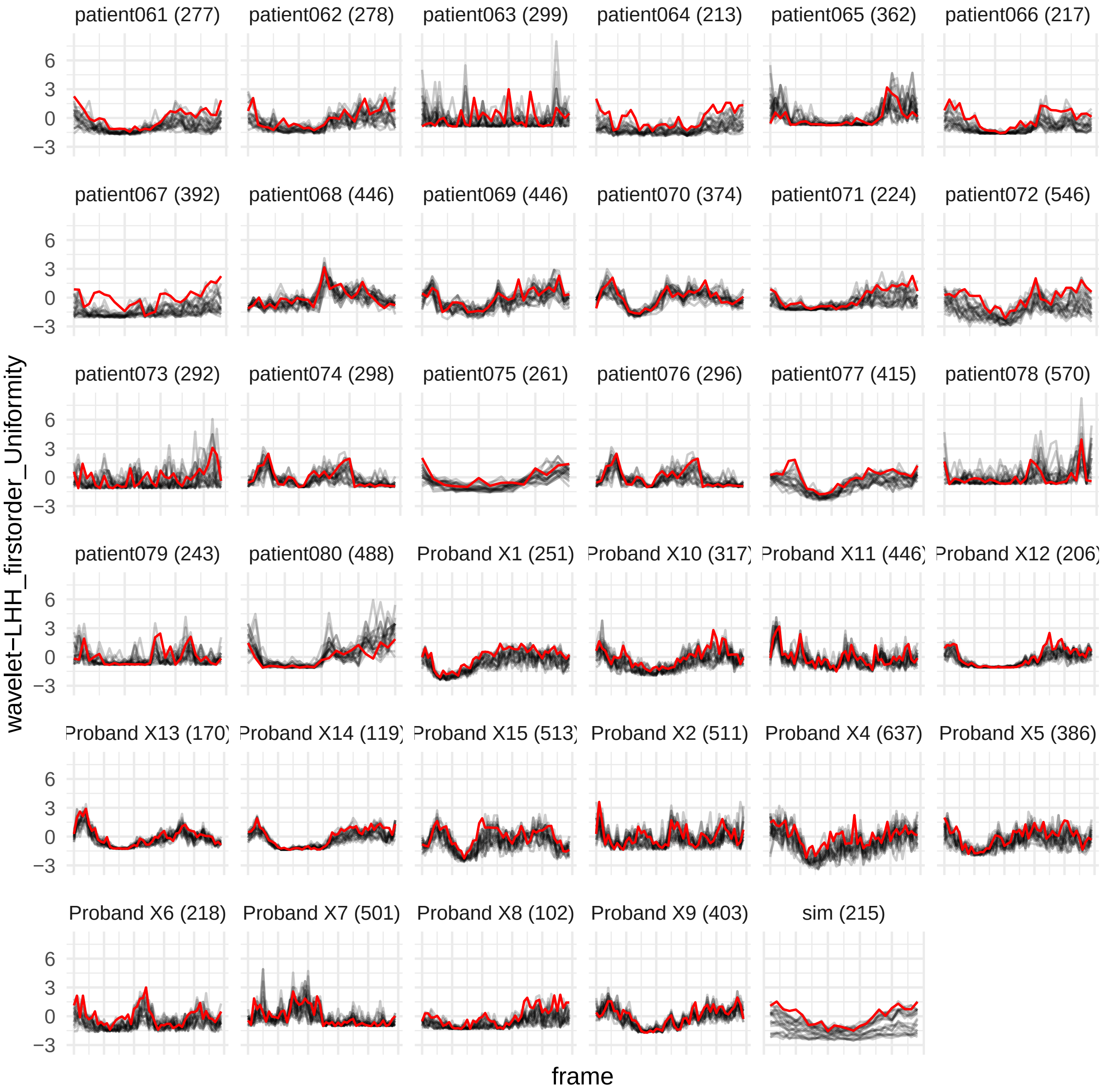

patient061 (277)
patient062 (278)
patient063 (299)
patient064 (213)
patient065 (362)
patient066 (217)
patient067 (392)
patient068 (446)
patient069 (446)
patient070 (374)
patient071 (224)
patient072 (546)
patient073 (292)
patient074 (298)
patient075 (261)
patient076 (296)
patient077 (415)
patient078 (570)
patient079 (243)
patient080 (488)
Proband X1 (251)
Proband X10 (317)
Proband X11 (446)
Proband X12 (206)
Proband X13 (170)
Proband X14 (119)
Proband X15 (513)
Proband X2 (511)
Proband X4 (637)
Proband X5 (386)
Proband X6 (218)
Proband X7 (501)
Proband X8 (102)
Proband X9 (403)
sim (215)
6
3
0
−3
wavelet−LHH_firstorder_Uniformity
frame

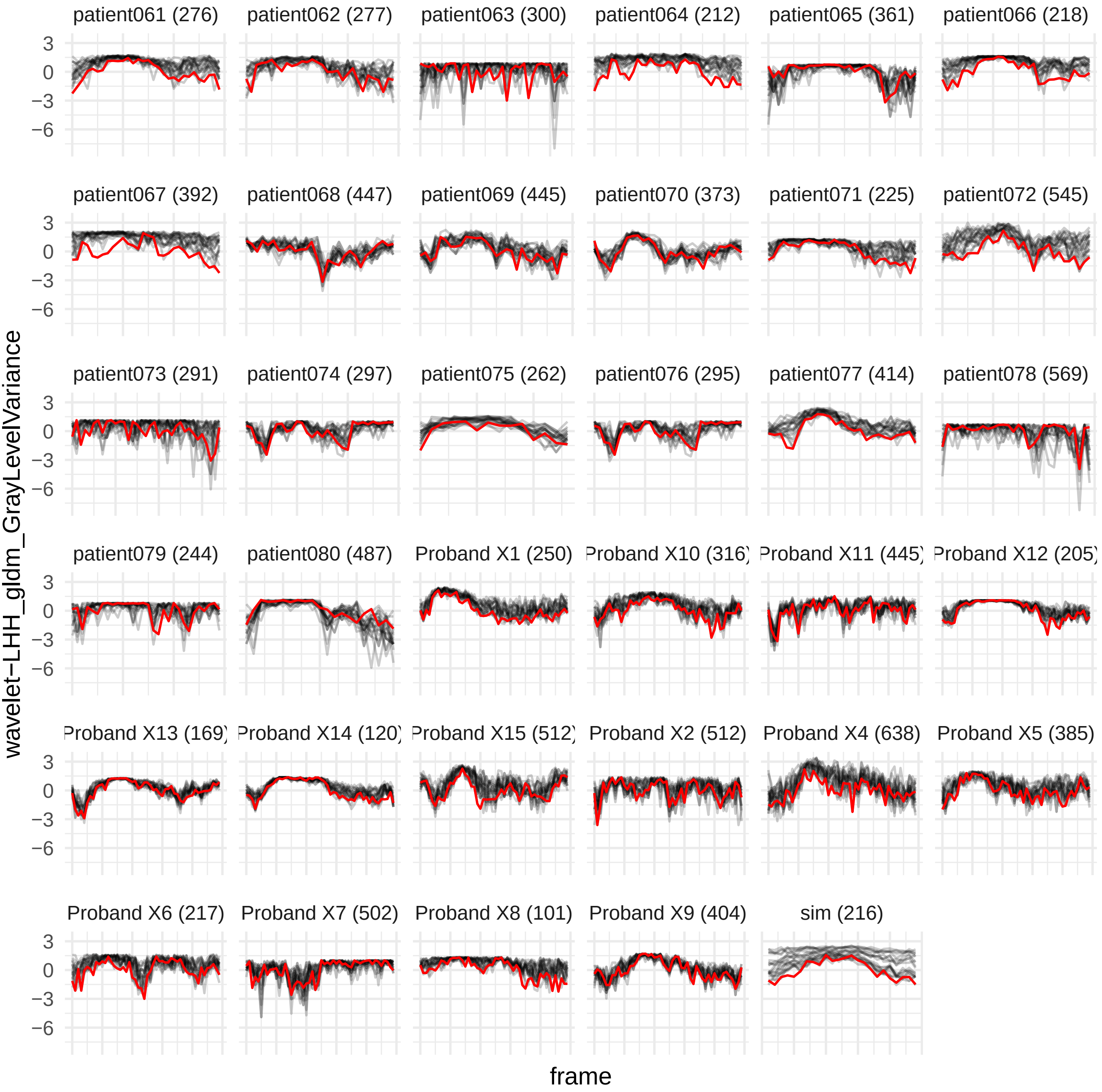
patient061 (276)
patient062 (277)
patient063 (300)
patient064 (212)
patient065 (361)
patient066 (218)
patient067 (392)
patient068 (447)
patient069 (445)
patient070 (373)
patient071 (225)
patient072 (545)
patient073 (291)
patient074 (297)
patient075 (262)
patient076 (295)
patient077 (414)
patient078 (569)
patient079 (244)
patient080 (487)
Proband X1 (250)
Proband X10 (316)
Proband X11 (445)
Proband X12 (205)
Proband X13 (169)
Proband X14 (120)
Proband X15 (512)
Proband X2 (512)
Proband X4 (638)
Proband X5 (385)
Proband X6 (217)
Proband X7 (502)
Proband X8 (101)
Proband X9 (404)
sim (216)
3
0
−3
−6
wavelet−LHH_gldm_GrayLevelVariance
frame

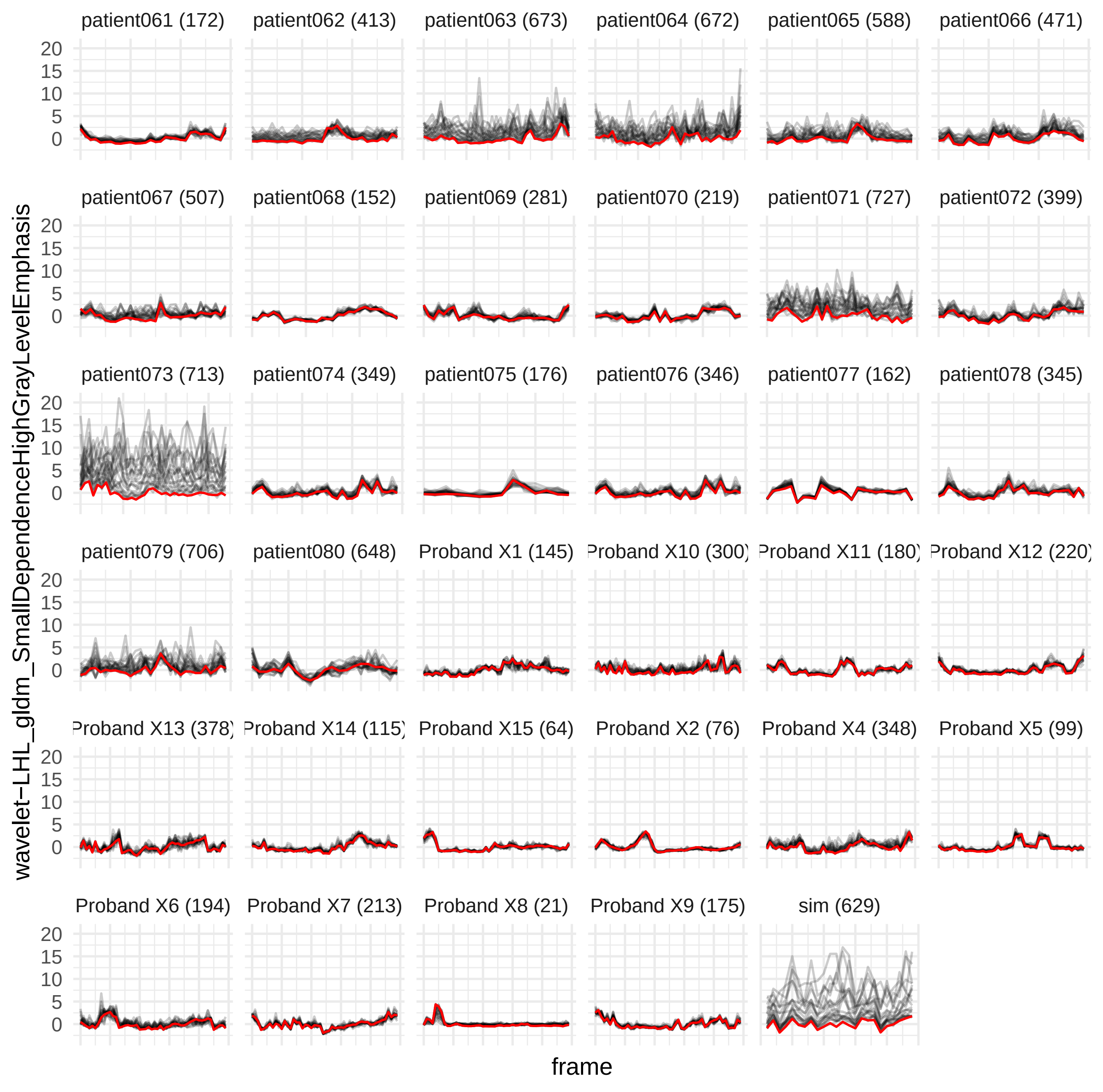
patient061 (172)
patient062 (413)
patient063 (673)
patient064 (672)
patient065 (588)
patient066 (471)
patient067 (507)
patient068 (152)
patient069 (281)
patient070 (219)
patient071 (727)
patient072 (399)
patient073 (713)
patient074 (349)
patient075 (176)
patient076 (346)
patient077 (162)
patient078 (345)
patient079 (706)
patient080 (648)
Proband X1 (145)
Proband X10 (300)
Proband X11 (180)
Proband X12 (220)
Proband X13 (378)
Proband X14 (115)
Proband X15 (64)
Proband X2 (76)
Proband X4 (348)
Proband X5 (99)
Proband X6 (194)
Proband X7 (213)
Proband X8 (21)
Proband X9 (175)
sim (629)
0
5
10
15
20
wavelet−LHL_gldm_SmallDependenceHighGrayLevelEmphasis
frame

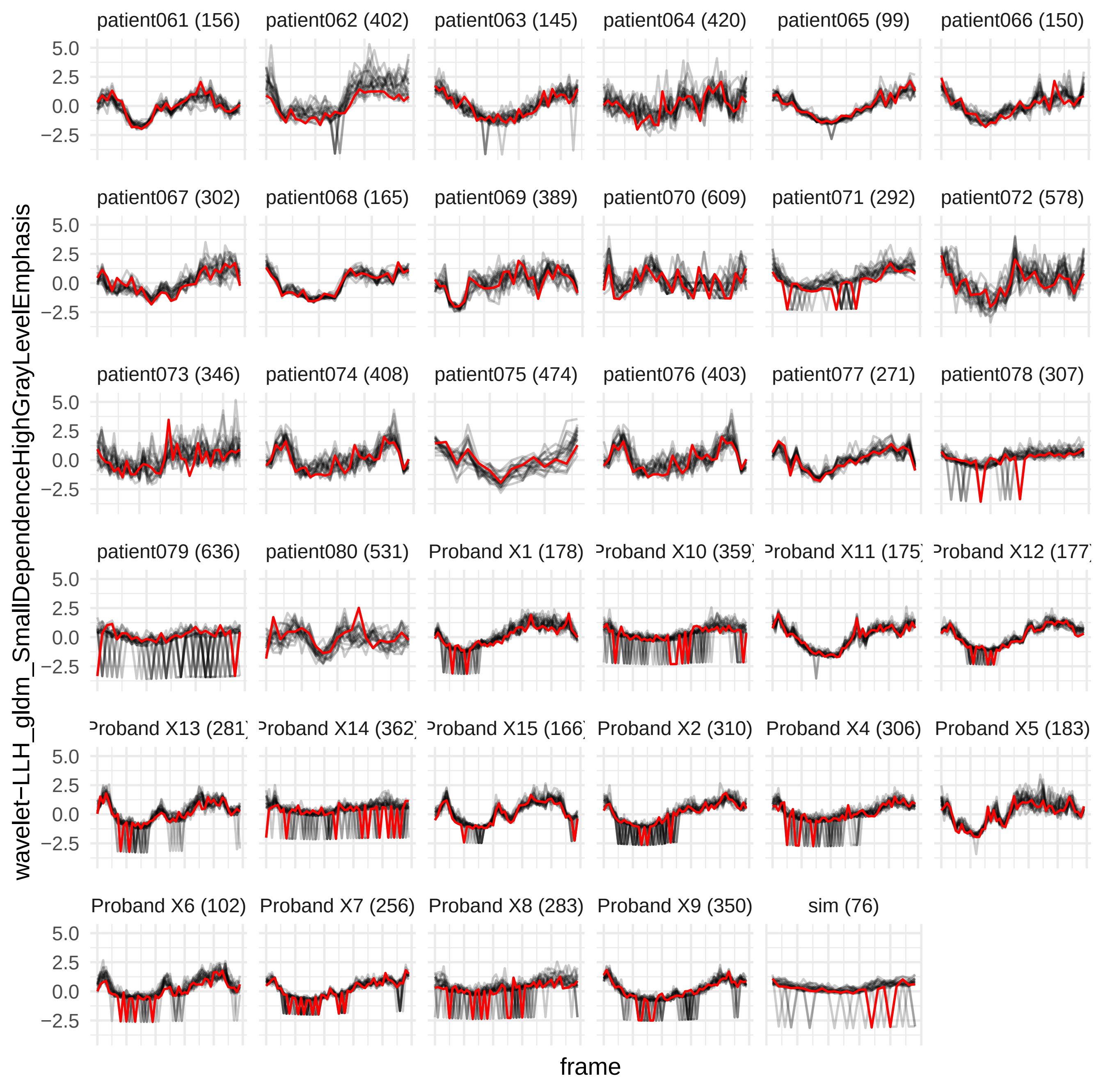

patient061 (156)
patient062 (402)
patient063 (145)
patient064 (420)
patient065 (99)
patient066 (150)
patient067 (302)
patient068 (165)
patient069 (389)
patient070 (609)
patient071 (292)
patient072 (578)
patient073 (346)
patient074 (408)
patient075 (474)
patient076 (403)
patient077 (271)
patient078 (307)
patient079 (636)
patient080 (531)
Proband X1 (178)
Proband X10 (359)
Proband X11 (175)
Proband X12 (177)
Proband X13 (281)
Proband X14 (362)
Proband X15 (166)
Proband X2 (310)
Proband X4 (306)
Proband X5 (183)
Proband X6 (102)
Proband X7 (256)
Proband X8 (283)
Proband X9 (350)
sim (76)
5.0
2.5
0.0
−2.5
wavelet−LLH_gldm_SmallDependenceHighGrayLevelEmphasis
frame

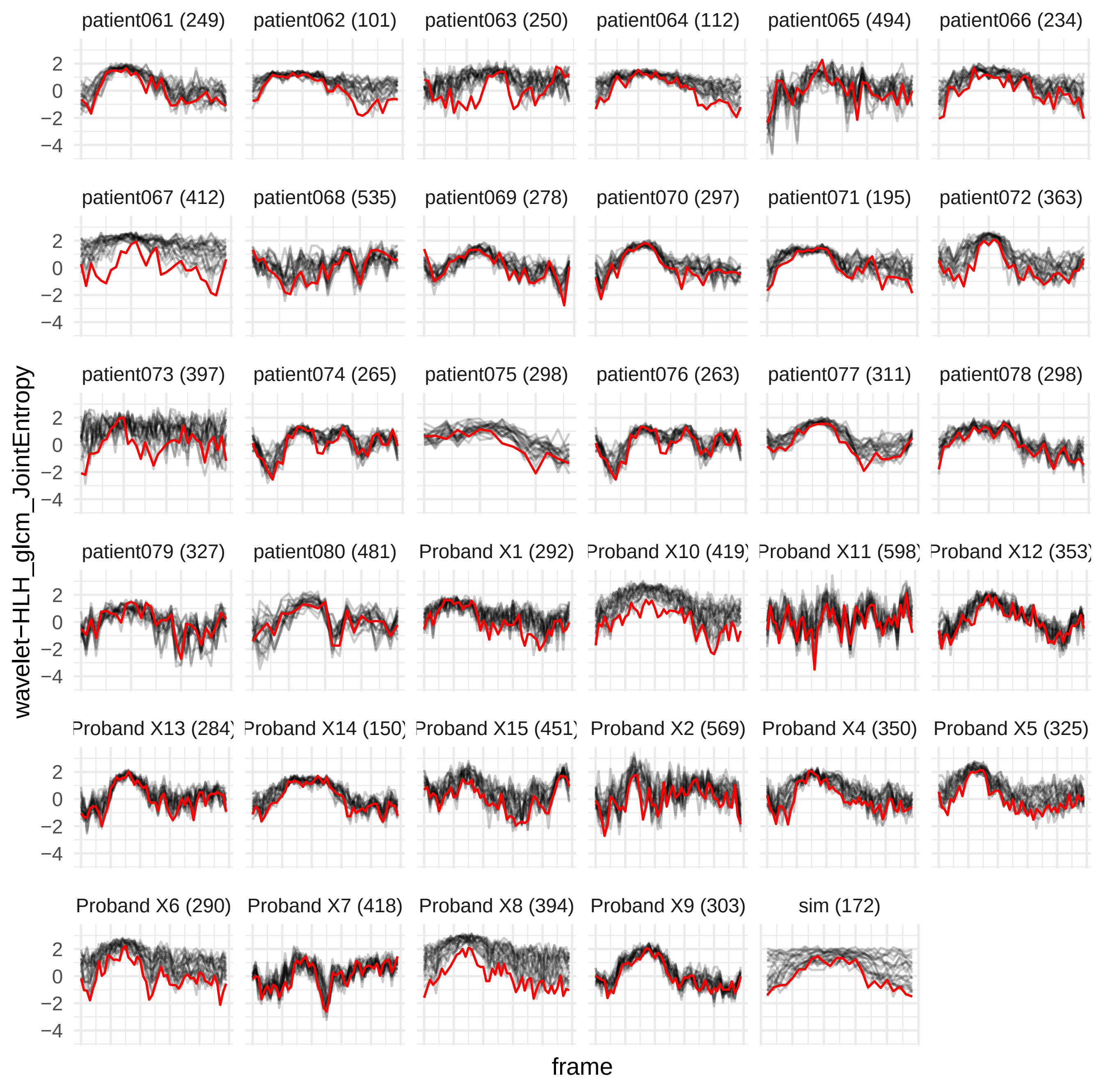
patient061 (249)
patient062 (101)
patient063 (250)
patient064 (112)
patient065 (494)
patient066 (234)
patient067 (412)
patient068 (535)
patient069 (278)
patient070 (297)
patient071 (195)
patient072 (363)
patient073 (397)
patient074 (265)
patient075 (298)
patient076 (263)
patient077 (311)
patient078 (298)
patient079 (327)
patient080 (481)
Proband X1 (292)
Proband X10 (419)
Proband X11 (598)
Proband X12 (353)
Proband X13 (284)
Proband X14 (150)
Proband X15 (451)
Proband X2 (569)
Proband X4 (350)
Proband X5 (325)
Proband X6 (290)
Proband X7 (418)
Proband X8 (394)
Proband X9 (303)
sim (172)
wavelet−HLH_glcm_JointEntropy
frame
2
0
−2
−4

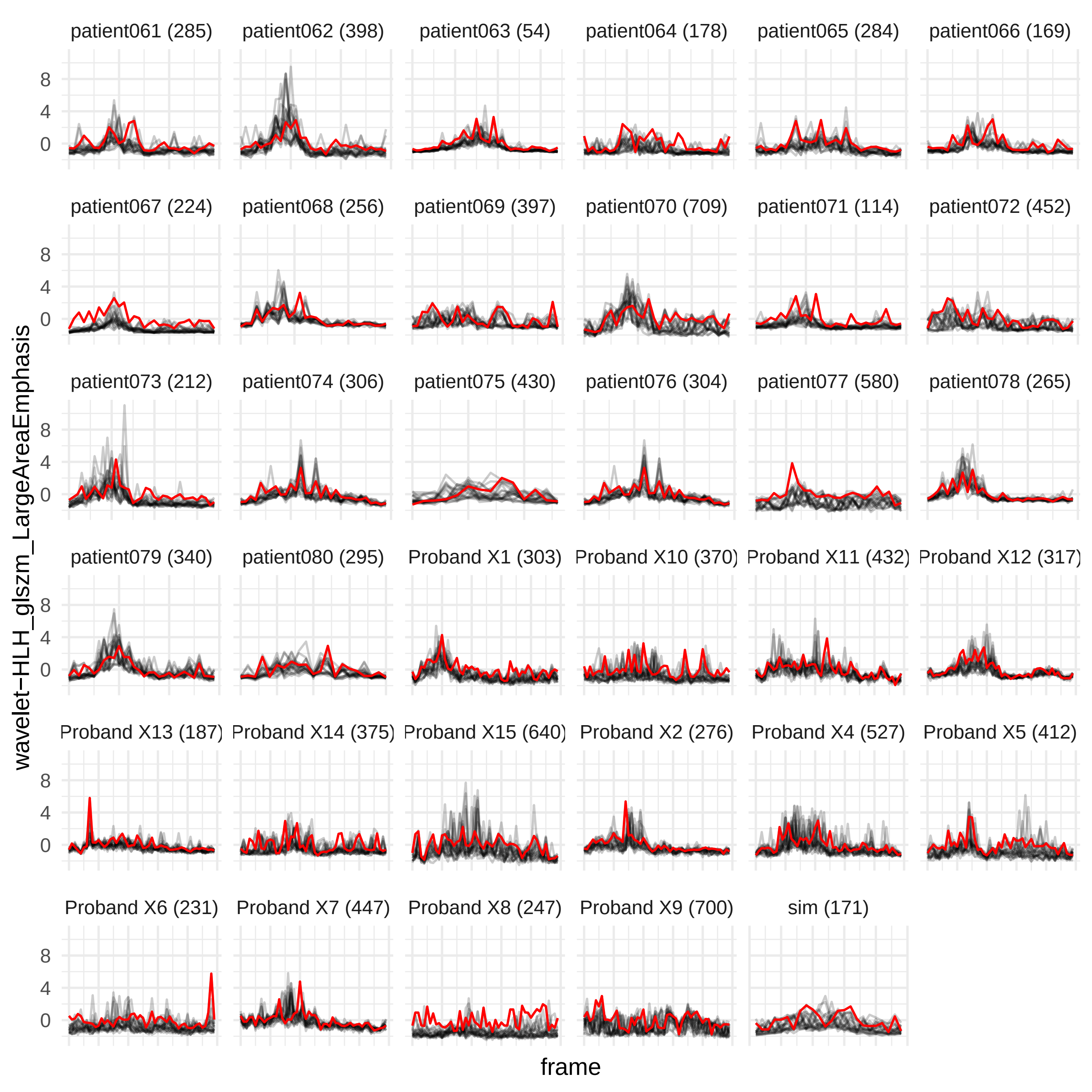
patient061 (285)
patient062 (398)
patient063 (54)
patient064 (178)
patient065 (284)
patient066 (169)
patient067 (224)
patient068 (256)
patient069 (397)
patient070 (709)
patient071 (114)
patient072 (452)
patient073 (212)
patient074 (306)
patient075 (430)
patient076 (304)
patient077 (580)
patient078 (265)
patient079 (340)
patient080 (295)
Proband X1 (303)
Proband X10 (370)
Proband X11 (432)
Proband X12 (317)
Proband X13 (187)
Proband X14 (375)
Proband X15 (640)
Proband X2 (276)
Proband X4 (527)
Proband X5 (412)
Proband X6 (231)
Proband X7 (447)
Proband X8 (247)
Proband X9 (700)
sim (171)
wavelet−HLH_glszm_LargeAreaEmphasis
frame
8
4
0

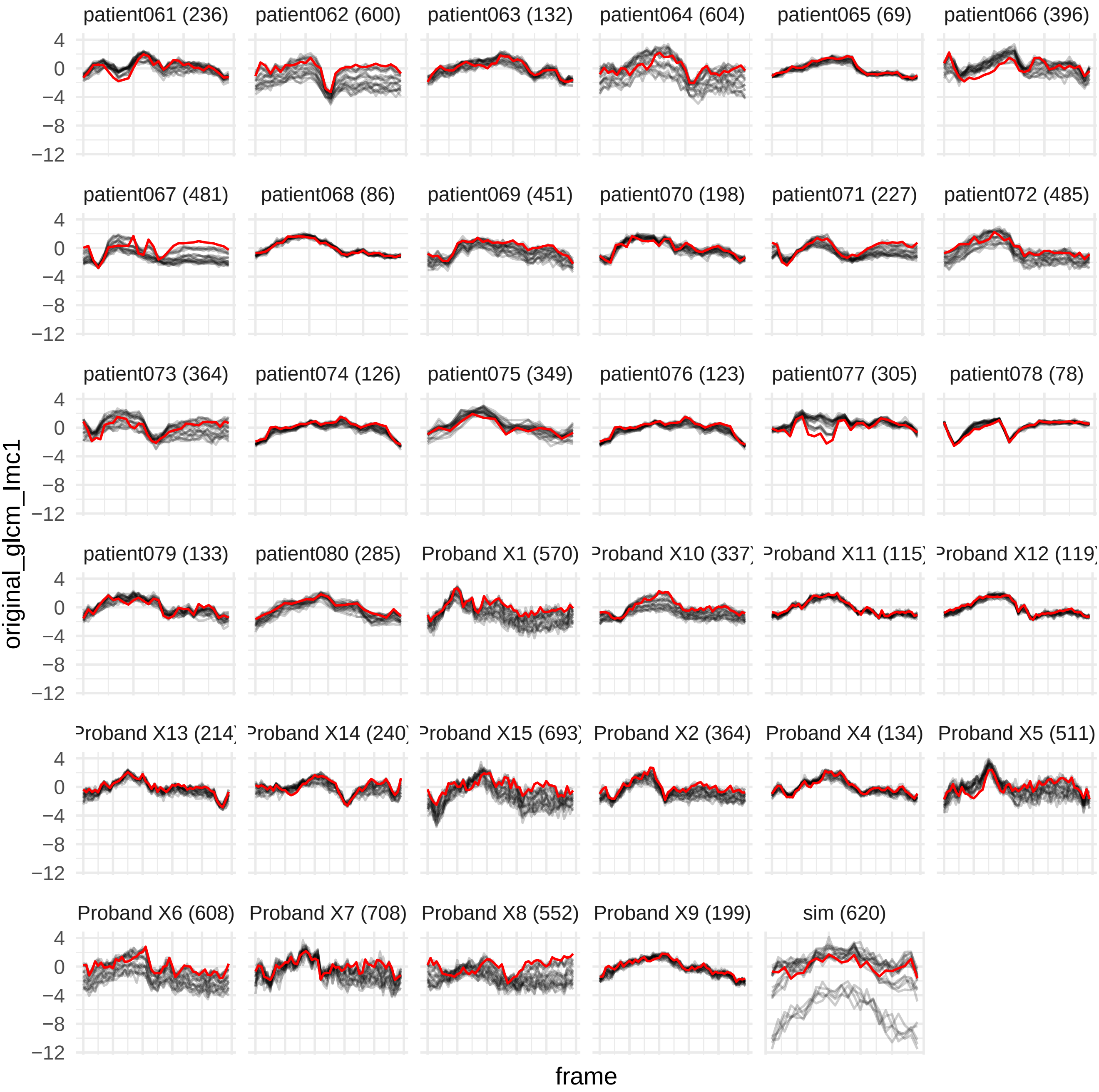

patient061 (236)
patient062 (600)
patient063 (132)
patient064 (604)
patient065 (69)
patient066 (396)
patient067 (481)
patient068 (86)
patient069 (451)
patient070 (198)
patient071 (227)
patient072 (485)
patient073 (364)
patient074 (126)
patient075 (349)
patient076 (123)
patient077 (305)
patient078 (78)
patient079 (133)
patient080 (285)
Proband X1 (570)
Proband X10 (337)
Proband X11 (115)
Proband X12 (119)
Proband X13 (214)
Proband X14 (240)
Proband X15 (693)
Proband X2 (364)
Proband X4 (134)
Proband X5 (511)
Proband X6 (608)
Proband X7 (708)
Proband X8 (552)
Proband X9 (199)
sim (620)
4
0
−4
−8
−12
original_glcm_Imc1
frame

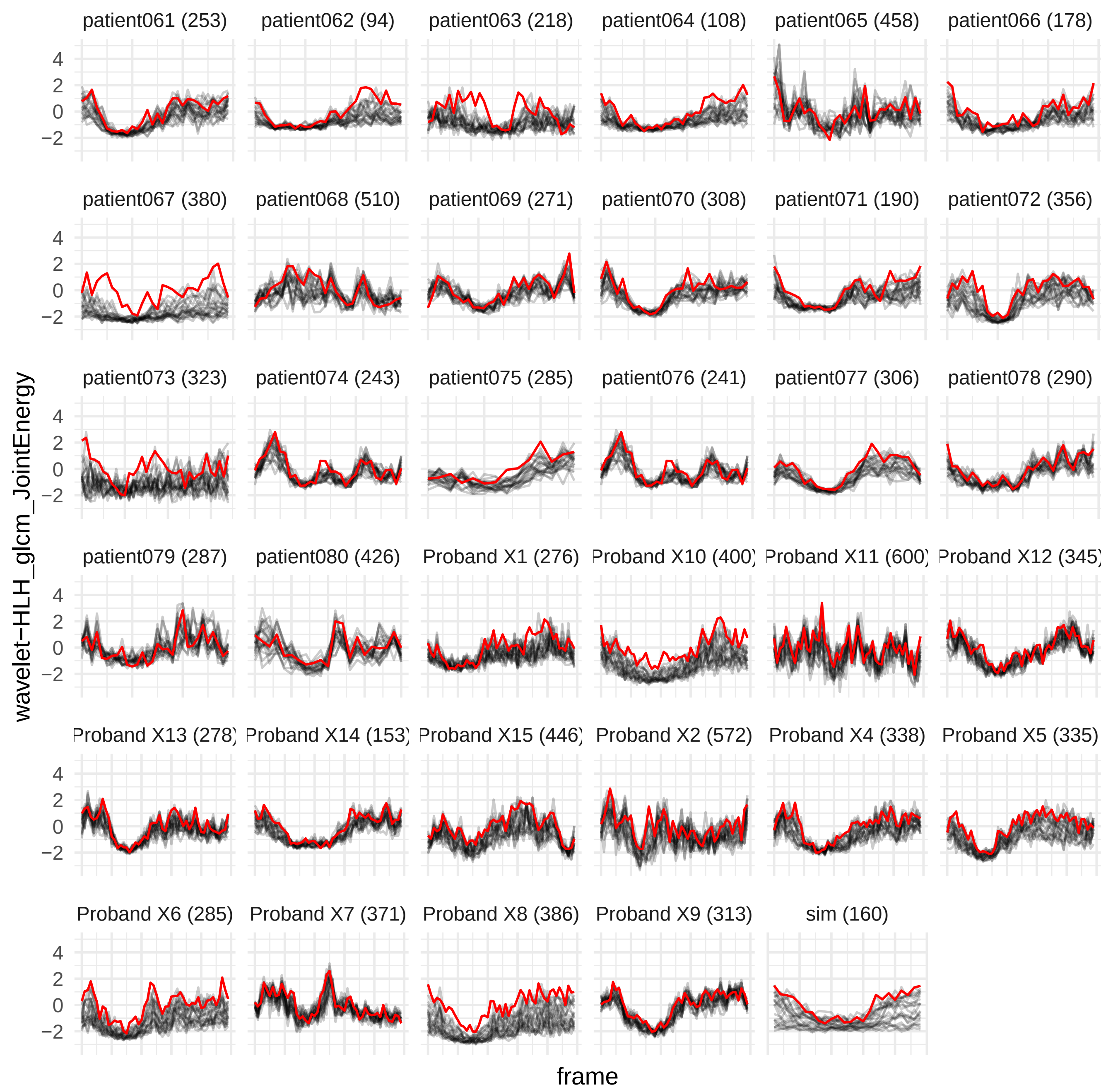

patient061 (253)
patient062 (94)
patient063 (218)
patient064 (108)
patient065 (458)
patient066 (178)
patient067 (380)
patient068 (510)
patient069 (271)
patient070 (308)
patient071 (190)
patient072 (356)
patient073 (323)
patient074 (243)
patient075 (285)
patient076 (241)
patient077 (306)
patient078 (290)
patient079 (287)
patient080 (426)
Proband X1 (276)
Proband X10 (400)
Proband X11 (600)
Proband X12 (345)
Proband X13 (278)
Proband X14 (153)
Proband X15 (446)
Proband X2 (572)
Proband X4 (338)
Proband X5 (335)
Proband X6 (285)
Proband X7 (371)
Proband X8 (386)
Proband X9 (313)
sim (160)
wavelet−HLH_glcm_JointEnergy
frame
4
2
0
−2

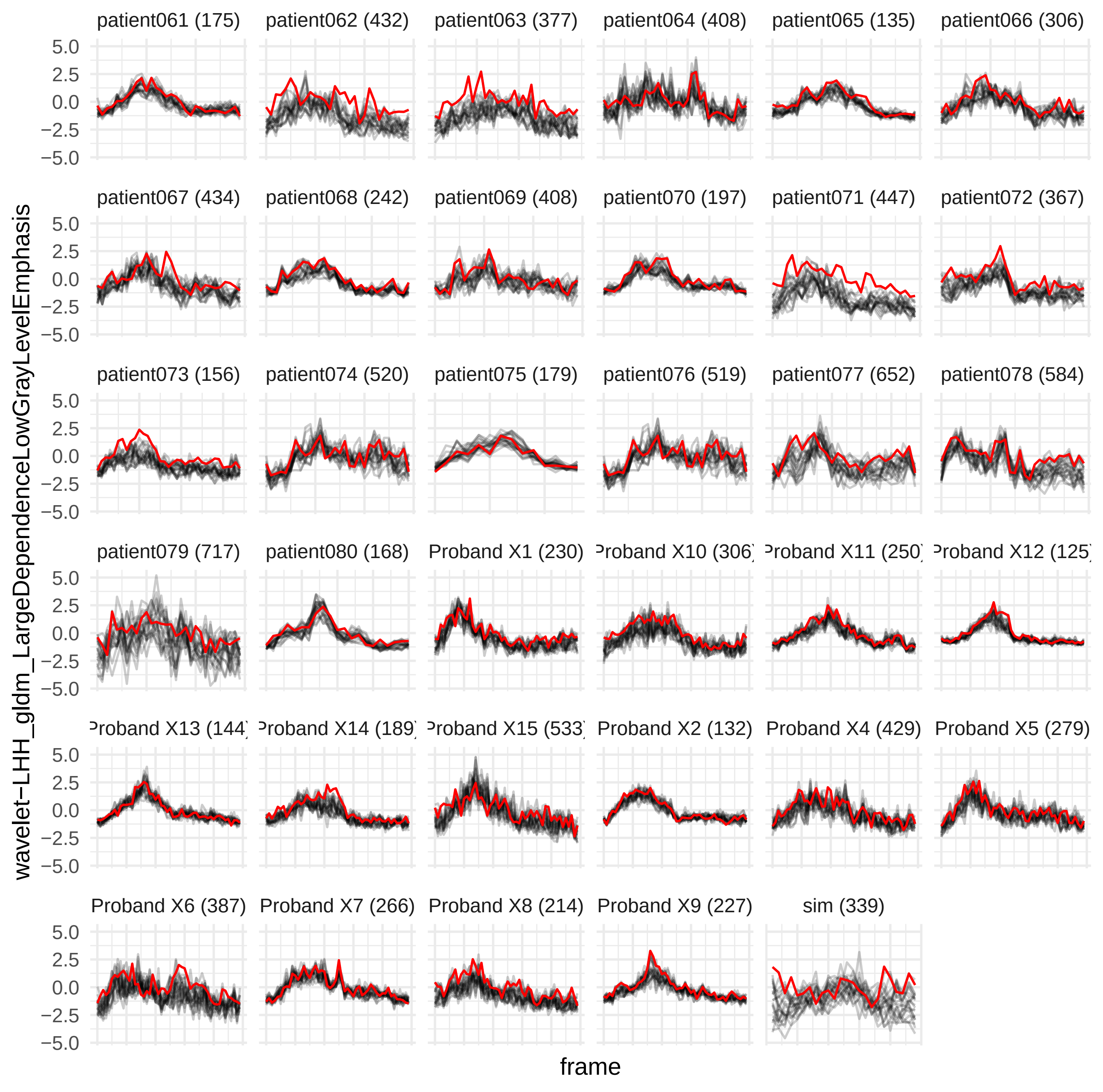
patient061 (175)
patient062 (432)
patient063 (377)
patient064 (408)
patient065 (135)
patient066 (306)
patient067 (434)
patient068 (242)
patient069 (408)
patient070 (197)
patient071 (447)
patient072 (367)
patient073 (156)
patient074 (520)
patient075 (179)
patient076 (519)
patient077 (652)
patient078 (584)
patient079 (717)
patient080 (168)
Proband X1 (230)
Proband X10 (306)
Proband X11 (250)
Proband X12 (125)
Proband X13 (144)
Proband X14 (189)
Proband X15 (533)
Proband X2 (132)
Proband X4 (429)
Proband X5 (279)
Proband X6 (387)
Proband X7 (266)
Proband X8 (214)
Proband X9 (227)
sim (339)
wavelet−LHH_gldm_LargeDependenceLowGrayLevelEmphasis
frame
5.0
2.5
0.0
−2.5
−5.0

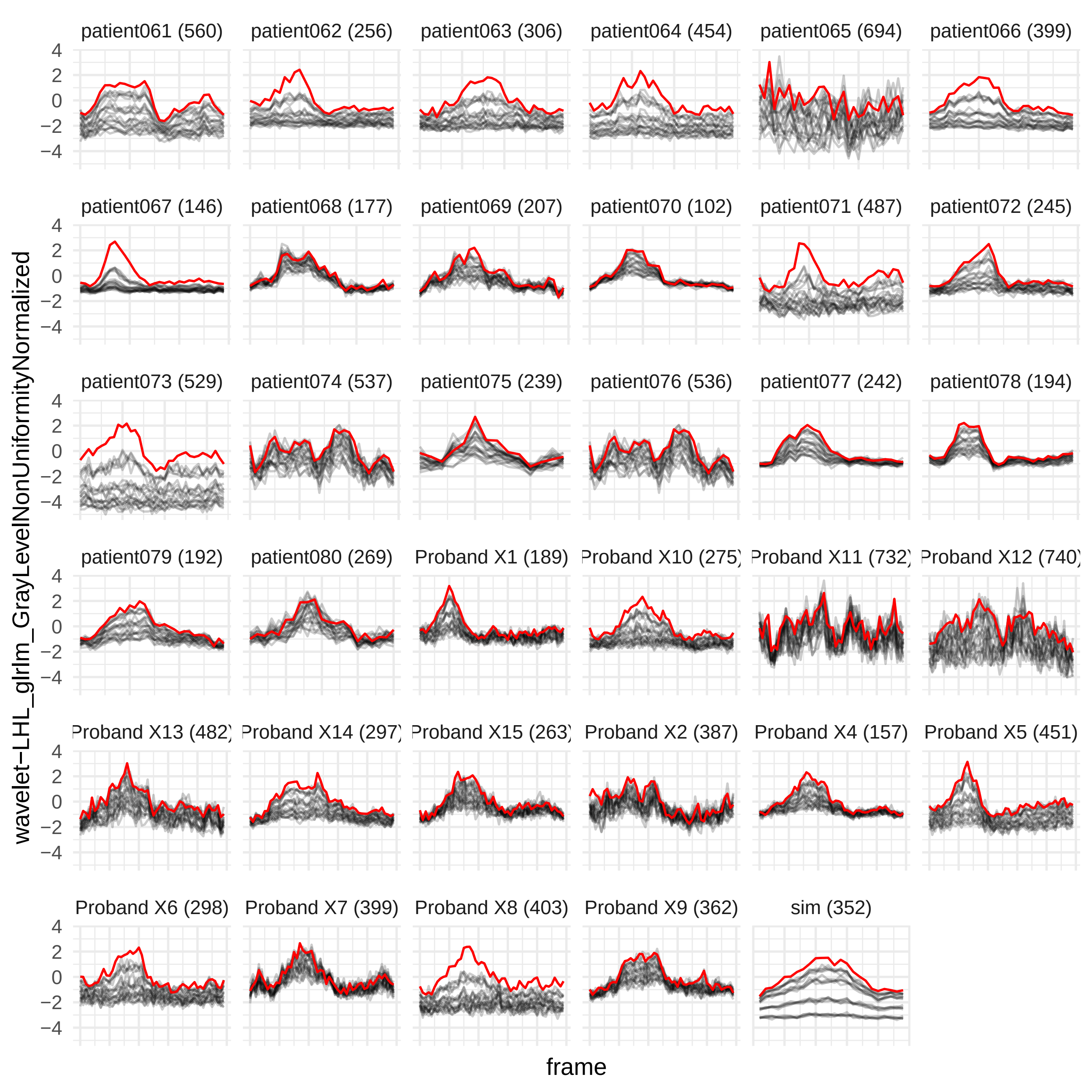
patient061 (560)
patient062 (256)
patient063 (306)
patient064 (454)
patient065 (694)
patient066 (399)
patient067 (146)
patient068 (177)
patient069 (207)
patient070 (102)
patient071 (487)
patient072 (245)
patient073 (529)
patient074 (537)
patient075 (239)
patient076 (536)
patient077 (242)
patient078 (194)
patient079 (192)
patient080 (269)
Proband X1 (189)
Proband X10 (275)
Proband X11 (732)
Proband X12 (740)
Proband X13 (482)
Proband X14 (297)
Proband X15 (263)
Proband X2 (387)
Proband X4 (157)
Proband X5 (451)
Proband X6 (298)
Proband X7 (399)
Proband X8 (403)
Proband X9 (362)
sim (352)
wavelet−LHL_glrlm_GrayLevelNonUniformityNormalized
frame

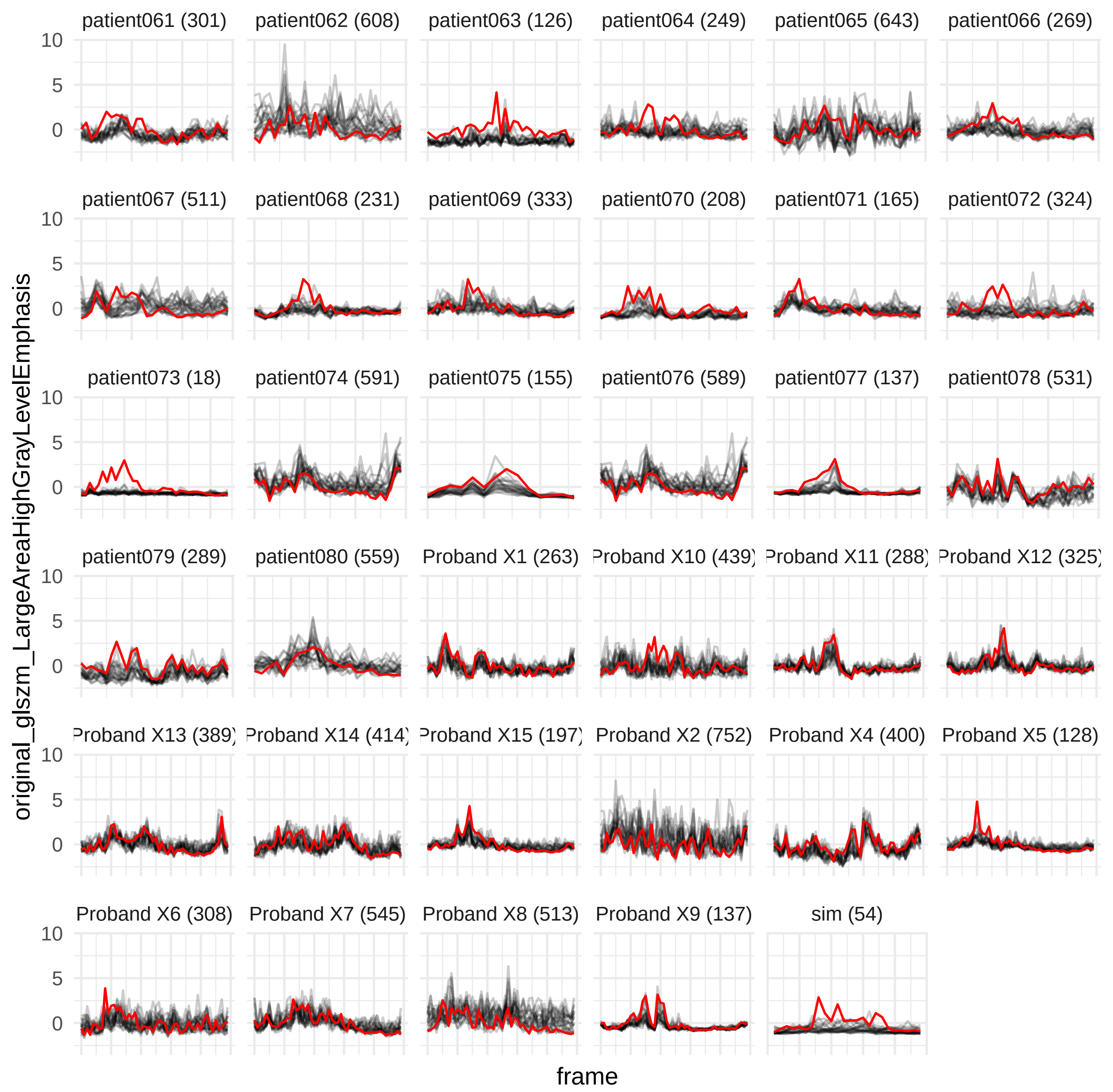

patient061 (301)
patient062 (608)
patient063 (126)
patient064 (249)
patient065 (643)
patient066 (269)
patient067 (511)
patient068 (231)
patient069 (333)
patient070 (208)
patient071 (165)
patient072 (324)
patient073 (18)
patient074 (591)
patient075 (155)
patient076 (589)
patient077 (137)
patient078 (531)
patient079 (289)
patient080 (559)
Proband X1 (263)
Proband X10 (439)
Proband X11 (288)
Proband X12 (325)
Proband X13 (389)
Proband X14 (414)
Proband X15 (197)
Proband X2 (752)
Proband X4 (400)
Proband X5 (128)
Proband X6 (308)
Proband X7 (545)
Proband X8 (513)
Proband X9 (137)
sim (54)
10
5
0
original_glszm_LargeAreaHighGrayLevelEmphasis
frame

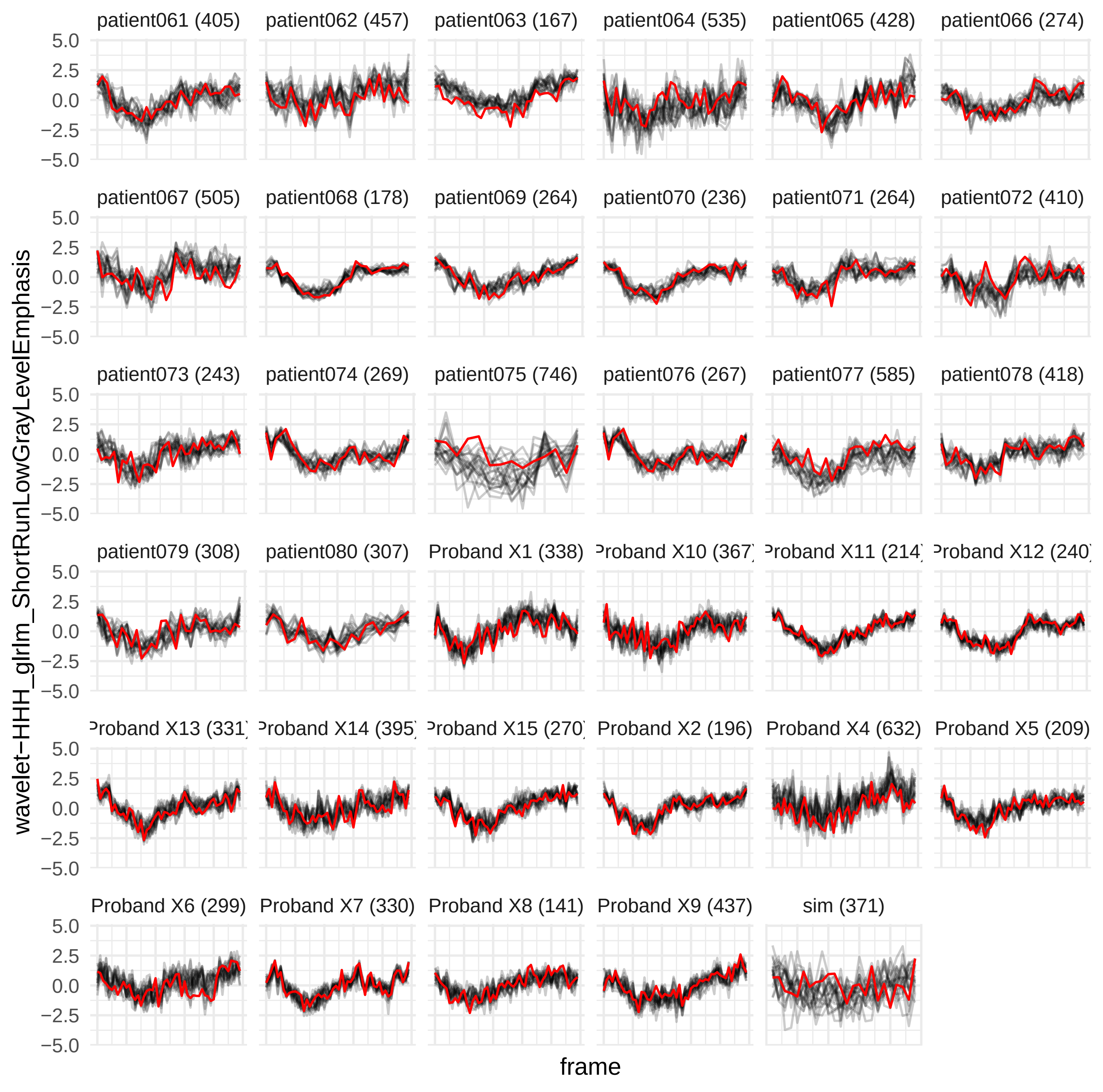

patient061 (405)
patient062 (457)
patient063 (167)
patient064 (535)
patient065 (428)
patient066 (274)
patient067 (505)
patient068 (178)
patient069 (264)
patient070 (236)
patient071 (264)
patient072 (410)
patient073 (243)
patient074 (269)
patient075 (746)
patient076 (267)
patient077 (585)
patient078 (418)
patient079 (308)
patient080 (307)
Proband X1 (338)
Proband X10 (367)
Proband X11 (214)
Proband X12 (240)
Proband X13 (331)
Proband X14 (395)
Proband X15 (270)
Proband X2 (196)
Proband X4 (632)
Proband X5 (209)
Proband X6 (299)
Proband X7 (330)
Proband X8 (141)
Proband X9 (437)
sim (371)
5.0
2.5
0.0
−2.5
−5.0
wavelet−HHH_glrlm_ShortRunLowGrayLevelEmphasis
frame

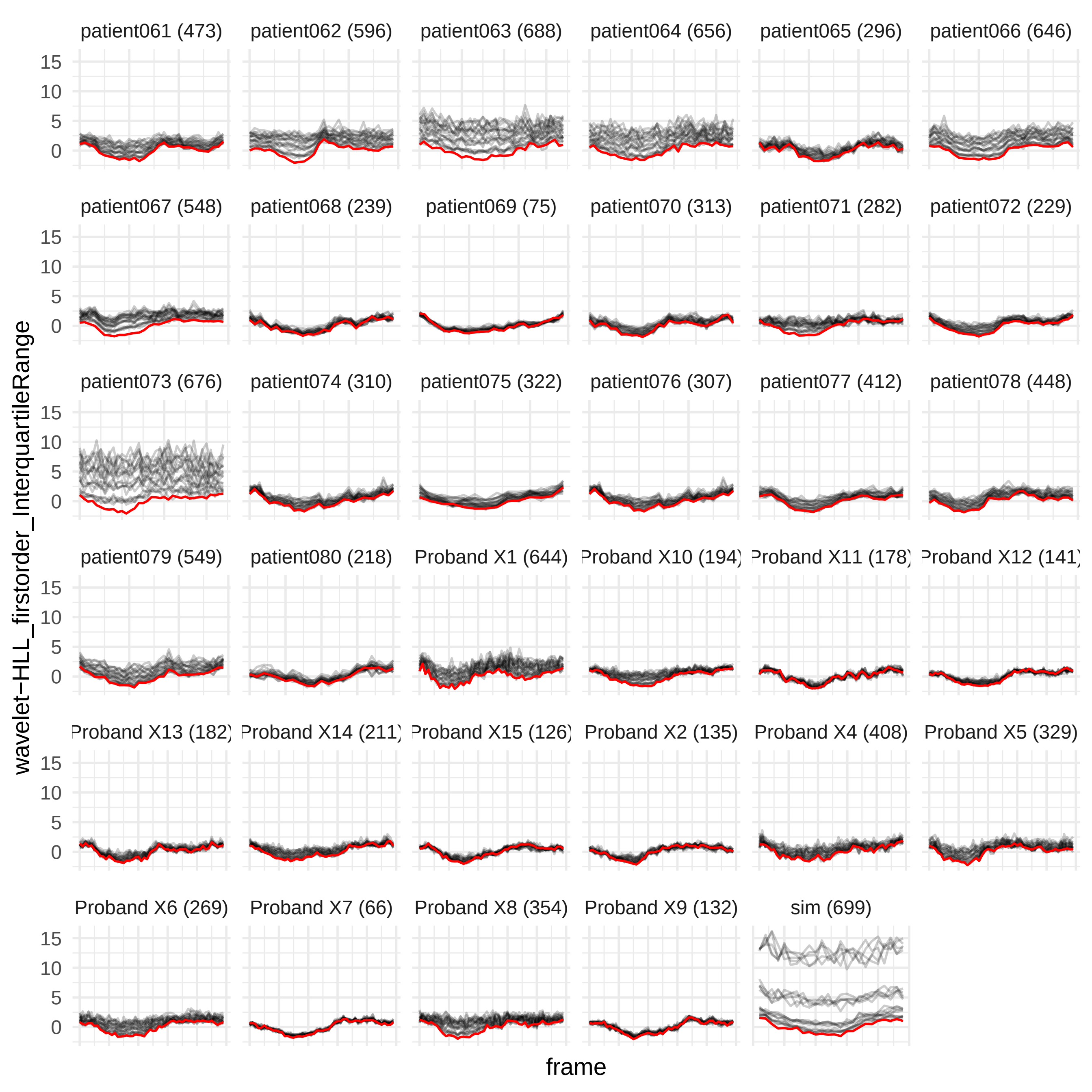

patient061 (473)
patient062 (596)
patient063 (688)
patient064 (656)
patient065 (296)
patient066 (646)
patient067 (548)
patient068 (239)
patient069 (75)
patient070 (313)
patient071 (282)
patient072 (229)
patient073 (676)
patient074 (310)
patient075 (322)
patient076 (307)
patient077 (412)
patient078 (448)
patient079 (549)
patient080 (218)
Proband X1 (644)
Proband X10 (194)
Proband X11 (178)
Proband X12 (141)
Proband X13 (182)
Proband X14 (211)
Proband X15 (126)
Proband X2 (135)
Proband X4 (408)
Proband X5 (329)
Proband X6 (269)
Proband X7 (66)
Proband X8 (354)
Proband X9 (132)
sim (699)
wavelet−HLL_firstorder_InterquartileRange
frame
15
10
5
0

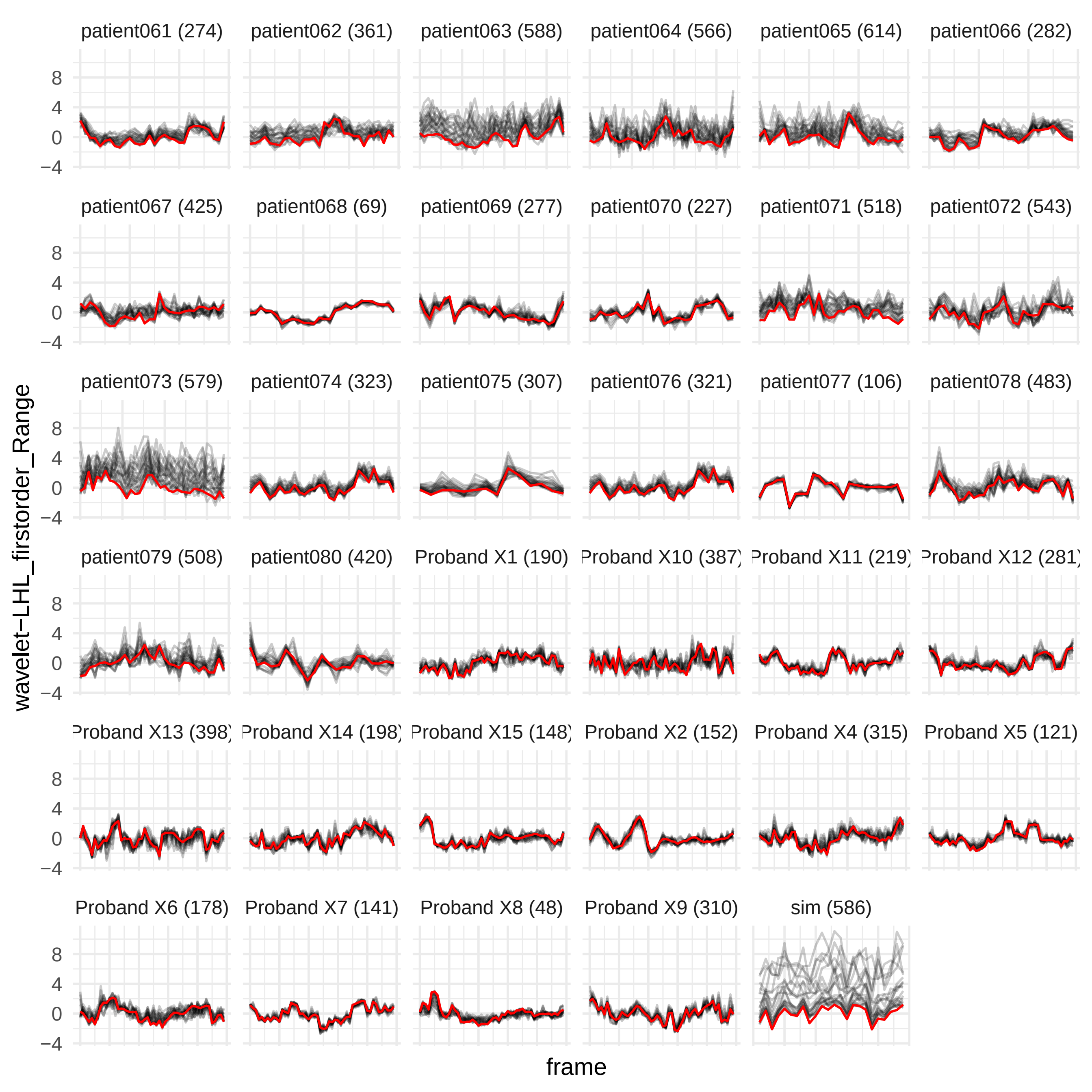

patient061 (274)
patient062 (361)
patient063 (588)
patient064 (566)
patient065 (614)
patient066 (282)
patient067 (425)
patient068 (69)
patient069 (277)
patient070 (227)
patient071 (518)
patient072 (543)
patient073 (579)
patient074 (323)
patient075 (307)
patient076 (321)
patient077 (106)
patient078 (483)
patient079 (508)
patient080 (420)
Proband X1 (190)
Proband X10 (387)
Proband X11 (219)
Proband X12 (281)
Proband X13 (398)
Proband X14 (198)
Proband X15 (148)
Proband X2 (152)
Proband X4 (315)
Proband X5 (121)
Proband X6 (178)
Proband X7 (141)
Proband X8 (48)
Proband X9 (310)
sim (586)
wavelet−LHL_firstorder_Range
8
4
0
−4
frame

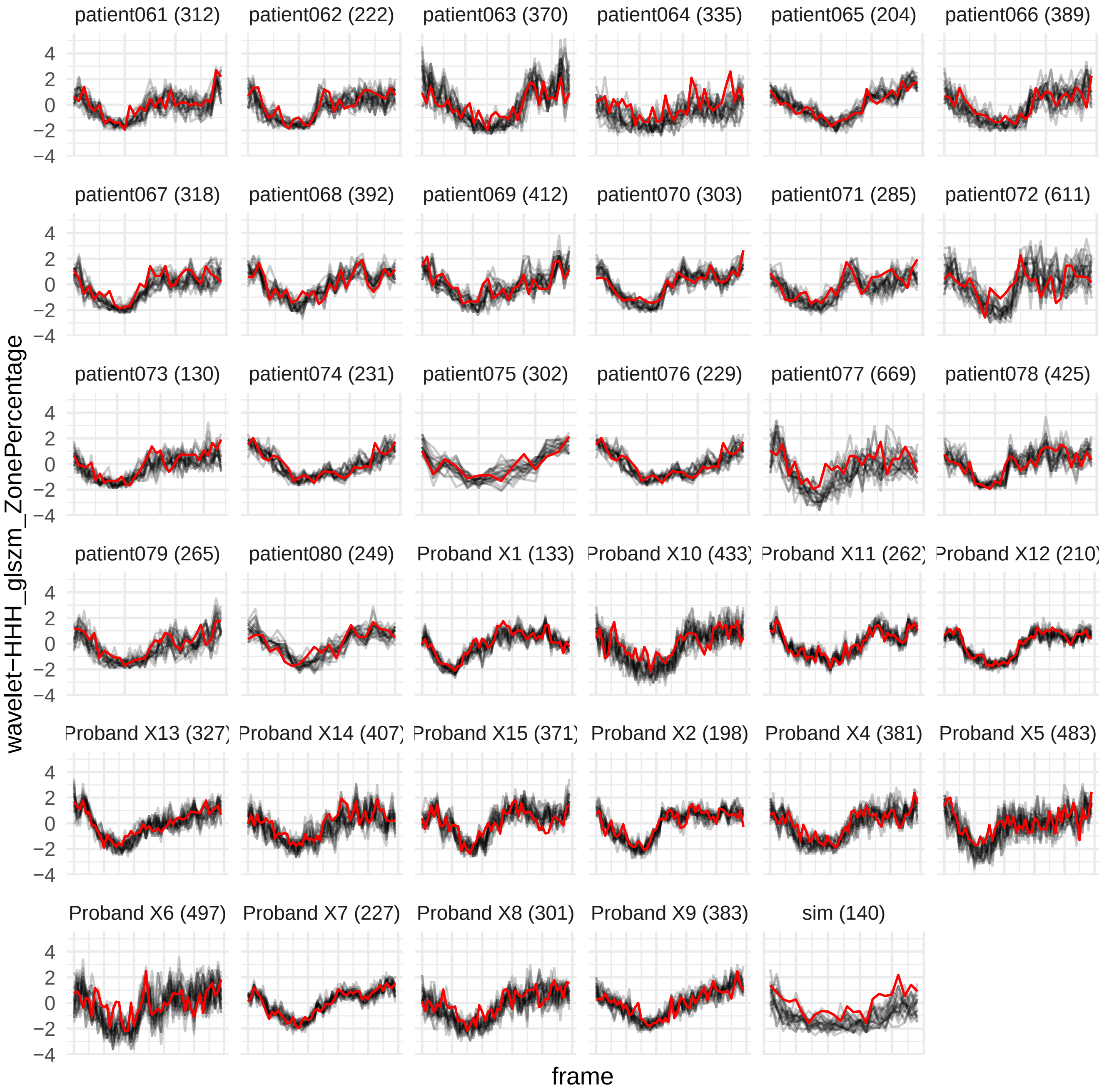

patient061 (312)
patient062 (222)
patient063 (370)
patient064 (335)
patient065 (204)
patient066 (389)
patient067 (318)
patient068 (392)
patient069 (412)
patient070 (303)
patient071 (285)
patient072 (611)
patient073 (130)
patient074 (231)
patient075 (302)
patient076 (229)
patient077 (669)
patient078 (425)
patient079 (265)
patient080 (249)
Proband X1 (133)
Proband X10 (433)
Proband X11 (262)
Proband X12 (210)
Proband X13 (327)
Proband X14 (407)
Proband X15 (371)
Proband X2 (198)
Proband X4 (381)
Proband X5 (483)
Proband X6 (497)
Proband X7 (227)
Proband X8 (301)
Proband X9 (383)
sim (140)
wavelet−HHH_glszm_ZonePercentage
frame
4
2
0
−2
−4

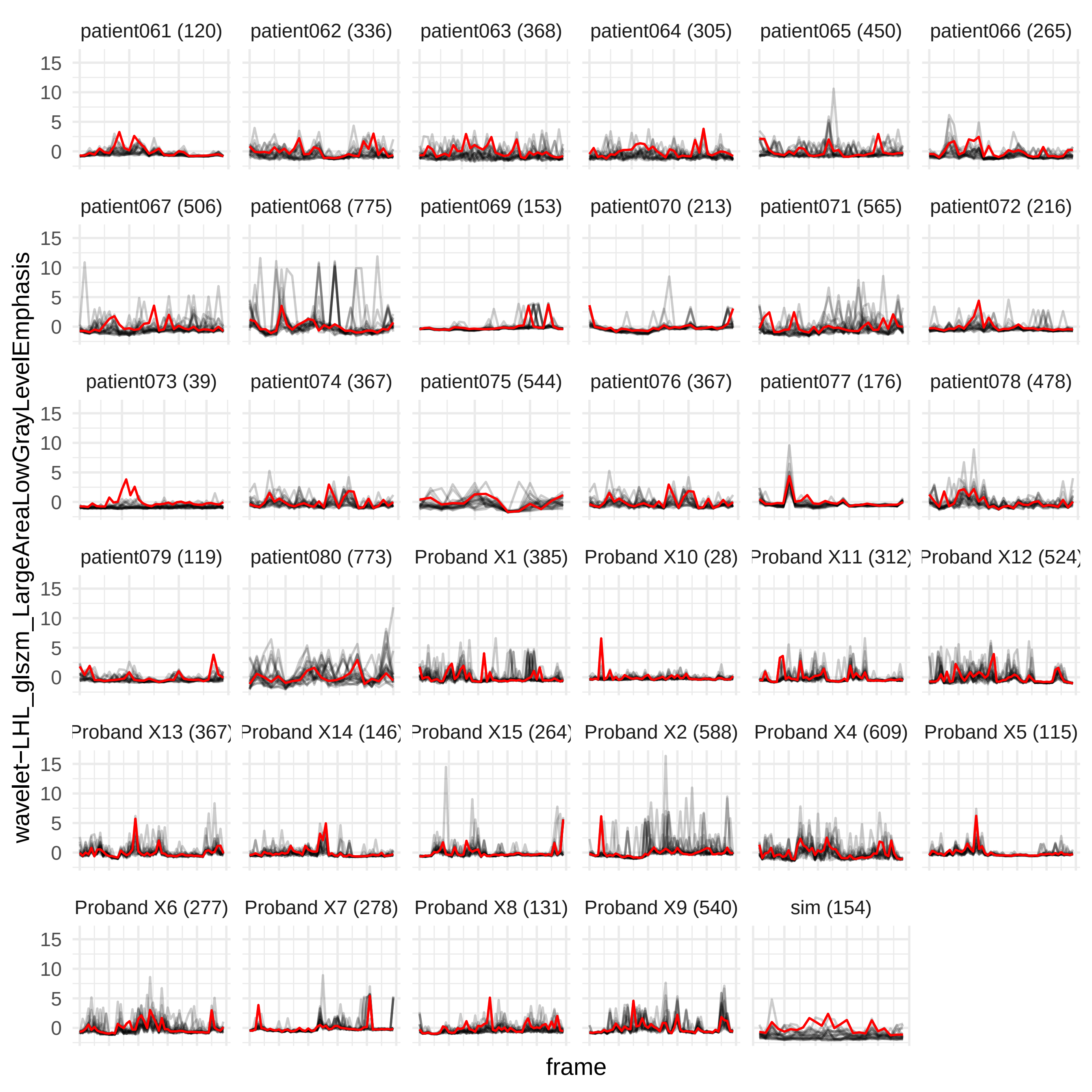
patient061 (120)
patient062 (336)
patient063 (368)
patient064 (305)
patient065 (450)
patient066 (265)
patient067 (506)
patient068 (775)
patient069 (153)
patient070 (213)
patient071 (565)
patient072 (216)
patient073 (39)
patient074 (367)
patient075 (544)
patient076 (367)
patient077 (176)
patient078 (478)
patient079 (119)
patient080 (773)
Proband X1 (385)
Proband X10 (28)
Proband X11 (312)
Proband X12 (524)
Proband X13 (367)
Proband X14 (146)
Proband X15 (264)
Proband X2 (588)
Proband X4 (609)
Proband X5 (115)
Proband X6 (277)
Proband X7 (278)
Proband X8 (131)
Proband X9 (540)
sim (154)
wavelet−LHL_glszm_LargeAreaLowGrayLevelEmphasis
frame
15
10
5
0

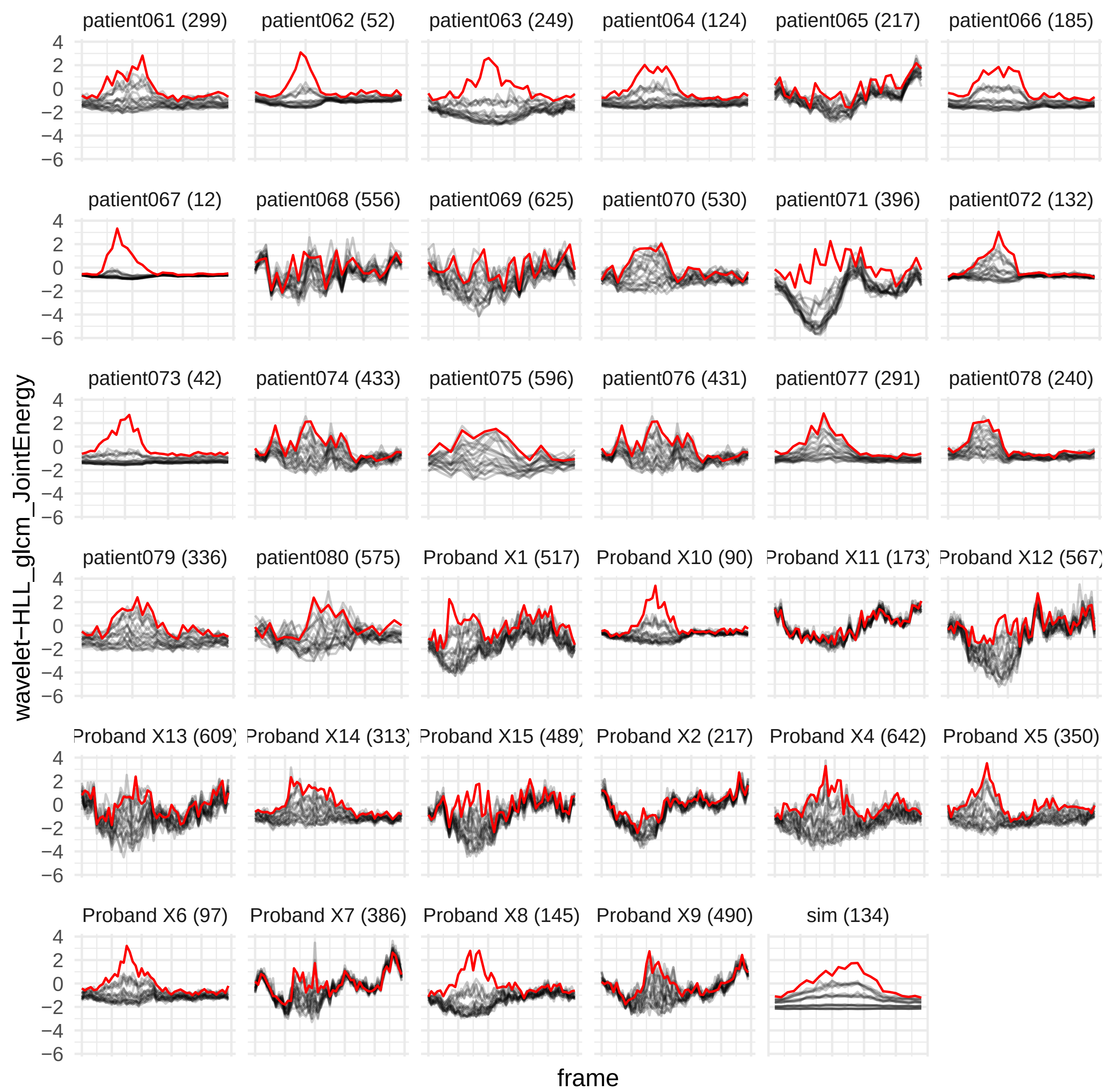

patient061 (299)
patient062 (52)
patient063 (249)
patient064 (124)
patient065 (217)
patient066 (185)
patient067 (12)
patient068 (556)
patient069 (625)
patient070 (530)
patient071 (396)
patient072 (132)
patient073 (42)
patient074 (433)
patient075 (596)
patient076 (431)
patient077 (291)
patient078 (240)
patient079 (336)
patient080 (575)
Proband X1 (517)
Proband X10 (90)
Proband X11 (173)
Proband X12 (567)
Proband X13 (609)
Proband X14 (313)
Proband X15 (489)
Proband X2 (217)
Proband X4 (642)
Proband X5 (350)
Proband X6 (97)
Proband X7 (386)
Proband X8 (145)
Proband X9 (490)
sim (134)
4
2
0
−2
−4
−6
wavelet−HLL_glcm_JointEnergy
frame

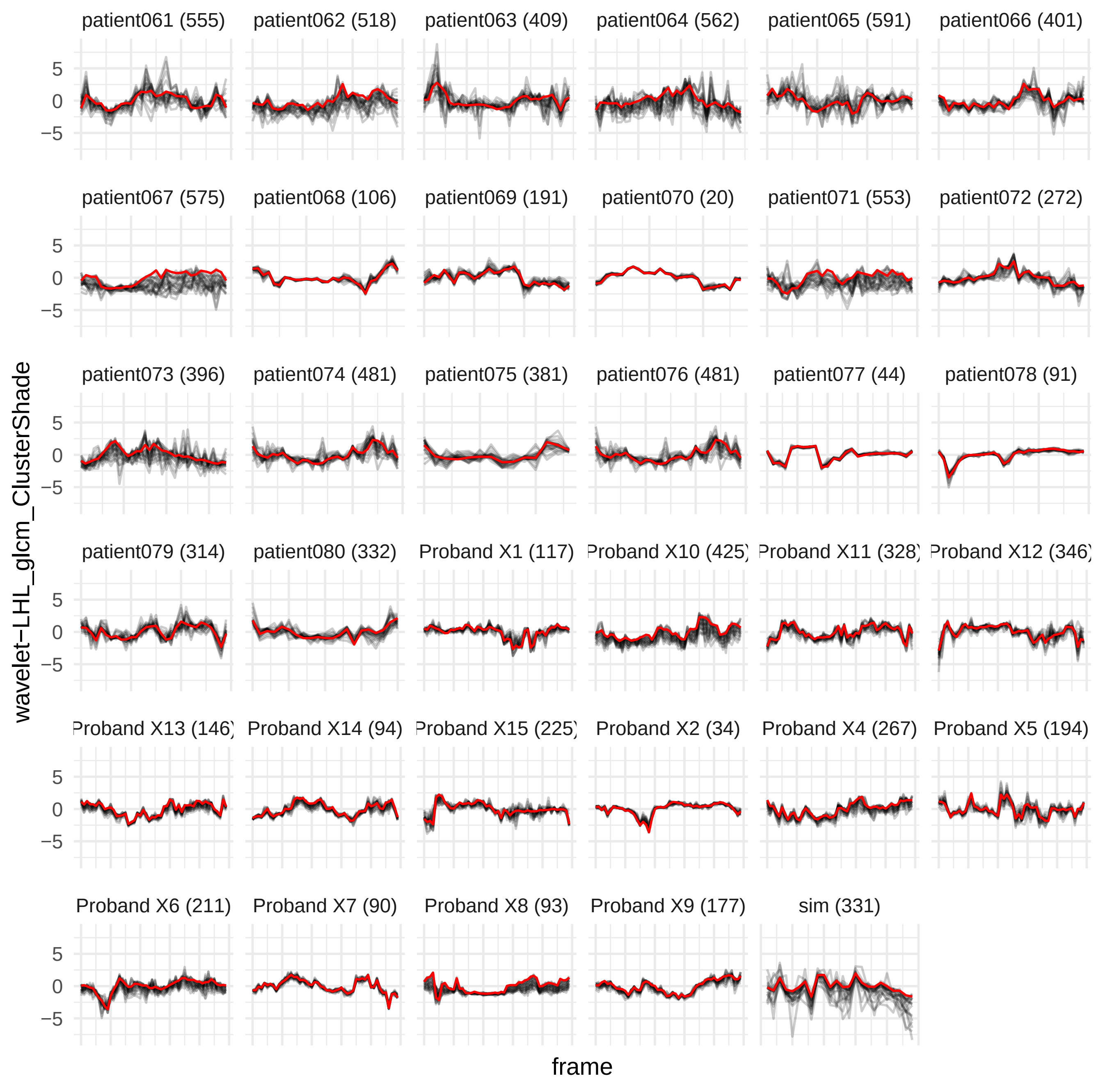
patient061 (555)
patient062 (518)
patient063 (409)
patient064 (562)
patient065 (591)
patient066 (401)
patient067 (575)
patient068 (106)
patient069 (191)
patient070 (20)
patient071 (553)
patient072 (272)
patient073 (396)
patient074 (481)
patient075 (381)
patient076 (481)
patient077 (44)
patient078 (91)
patient079 (314)
patient080 (332)
Proband X1 (117)
Proband X10 (425)
Proband X11 (328)
Proband X12 (346)
Proband X13 (146)
Proband X14 (94)
Proband X15 (225)
Proband X2 (34)
Proband X4 (267)
Proband X5 (194)
Proband X6 (211)
Proband X7 (90)
Proband X8 (93)
Proband X9 (177)
sim (331)
5
0
−5
wavelet−LHL_glcm_ClusterShade
frame

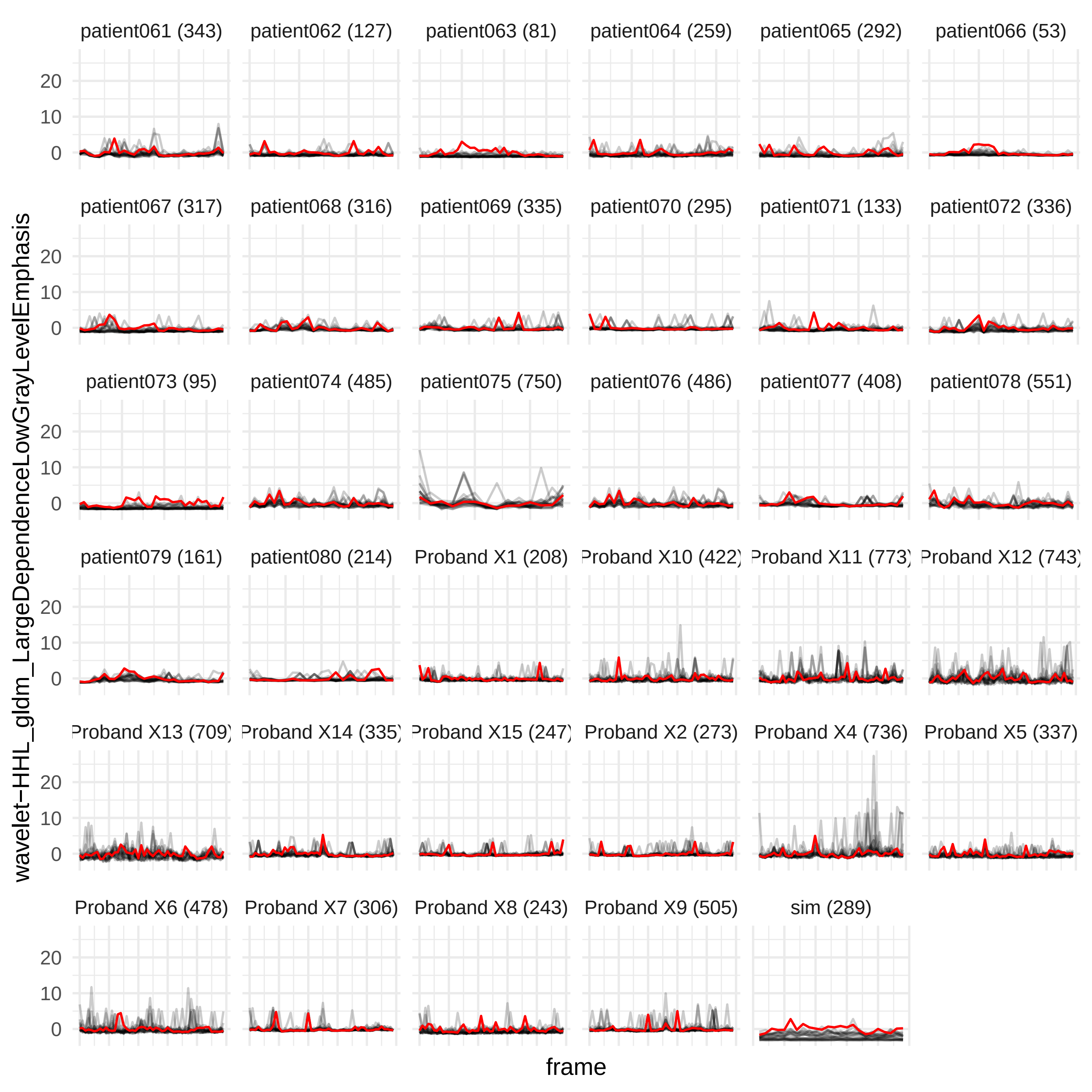

patient061 (343)
patient062 (127)
patient063 (81)
patient064 (259)
patient065 (292)
patient066 (53)
patient067 (317)
patient068 (316)
patient069 (335)
patient070 (295)
patient071 (133)
patient072 (336)
patient073 (95)
patient074 (485)
patient075 (750)
patient076 (486)
patient077 (408)
patient078 (551)
patient079 (161)
patient080 (214)
Proband X1 (208)
Proband X10 (422)
Proband X11 (773)
Proband X12 (743)
Proband X13 (709)
Proband X14 (335)
Proband X15 (247)
Proband X2 (273)
Proband X4 (736)
Proband X5 (337)
Proband X6 (478)
Proband X7 (306)
Proband X8 (243)
Proband X9 (505)
sim (289)
20
10
0
wavelet−HHL_gldm_LargeDependenceLowGrayLevelEmphasis
frame

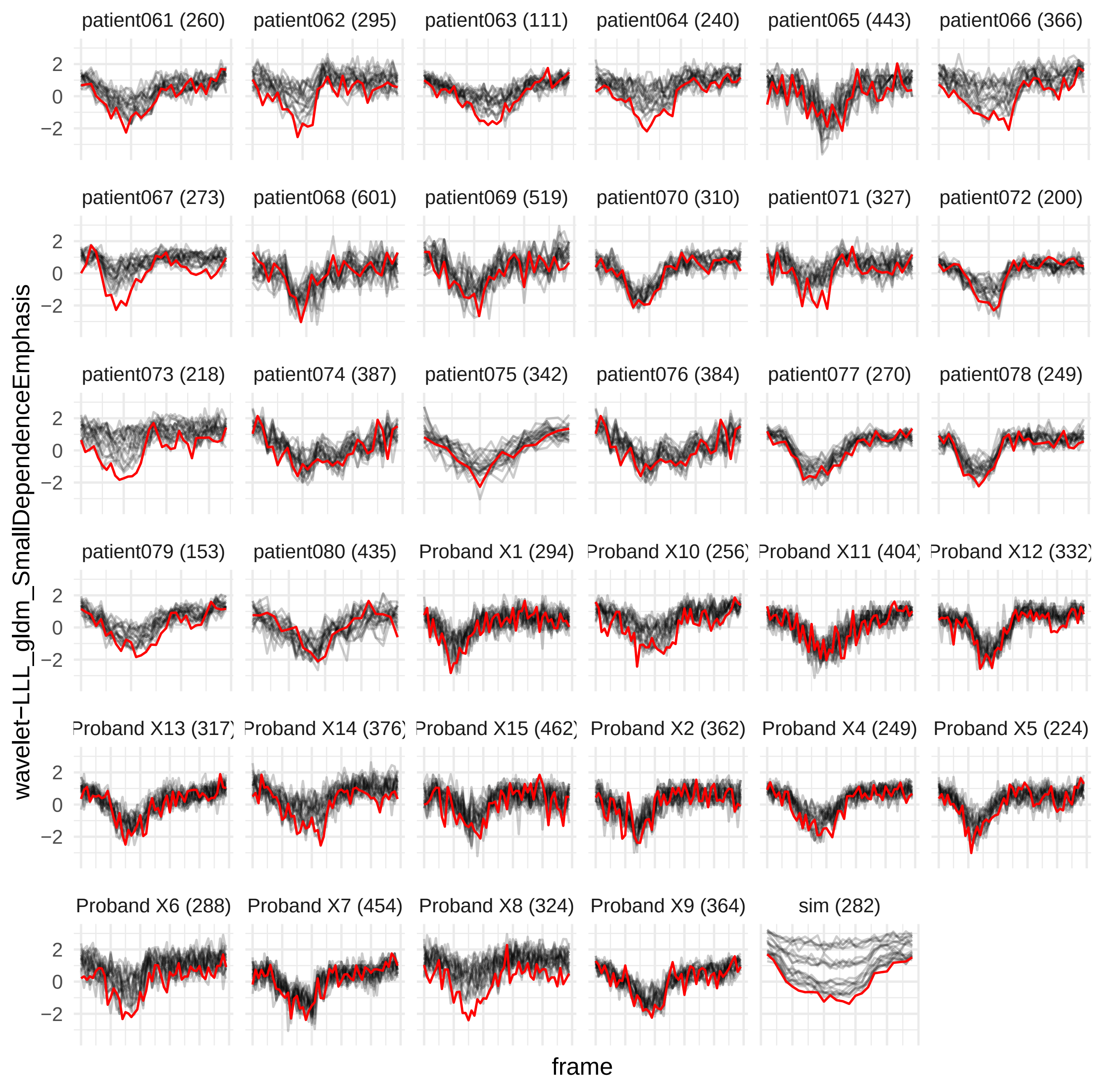

patient061 (260)
patient062 (295)
patient063 (111)
patient064 (240)
patient065 (443)
patient066 (366)
patient067 (273)
patient068 (601)
patient069 (519)
patient070 (310)
patient071 (327)
patient072 (200)
patient073 (218)
patient074 (387)
patient075 (342)
patient076 (384)
patient077 (270)
patient078 (249)
patient079 (153)
patient080 (435)
Proband X1 (294)
Proband X10 (256)
Proband X11 (404)
Proband X12 (332)
Proband X13 (317)
Proband X14 (376)
Proband X15 (462)
Proband X2 (362)
Proband X4 (249)
Proband X5 (224)
Proband X6 (288)
Proband X7 (454)
Proband X8 (324)
Proband X9 (364)
sim (282)
wavelet−LLL_gldm_SmallDependenceEmphasis
frame
2
0
−2

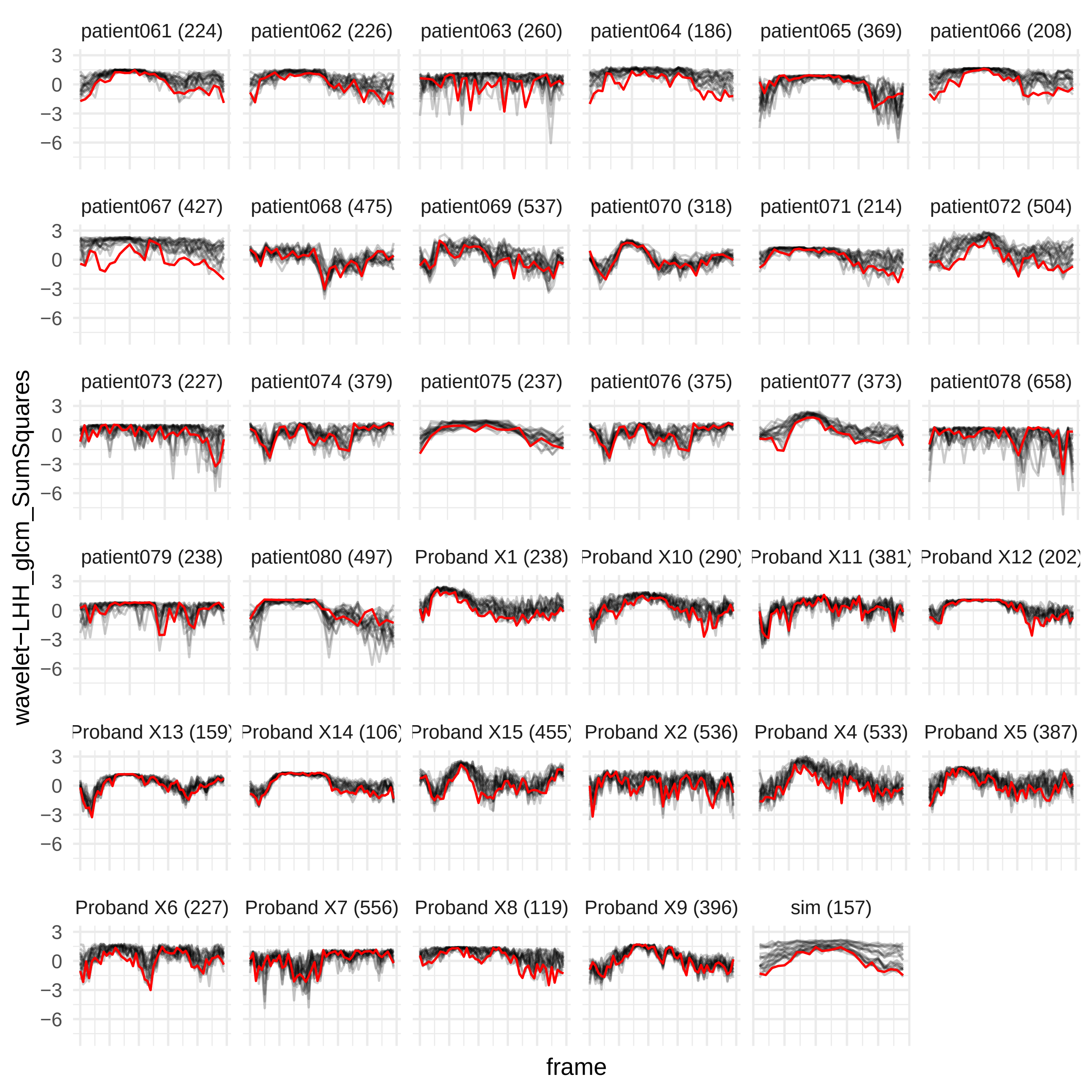

patient061 (224)
patient062 (226)
patient063 (260)
patient064 (186)
patient065 (369)
patient066 (208)
patient067 (427)
patient068 (475)
patient069 (537)
patient070 (318)
patient071 (214)
patient072 (504)
patient073 (227)
patient074 (379)
patient075 (237)
patient076 (375)
patient077 (373)
patient078 (658)
patient079 (238)
patient080 (497)
Proband X1 (238)
Proband X10 (290)
Proband X11 (381)
Proband X12 (202)
Proband X13 (159)
Proband X14 (106)
Proband X15 (455)
Proband X2 (536)
Proband X4 (533)
Proband X5 (387)
Proband X6 (227)
Proband X7 (556)
Proband X8 (119)
Proband X9 (396)
sim (157)
3
0
−3
−6
wavelet−LHH_glcm_SumSquares
frame

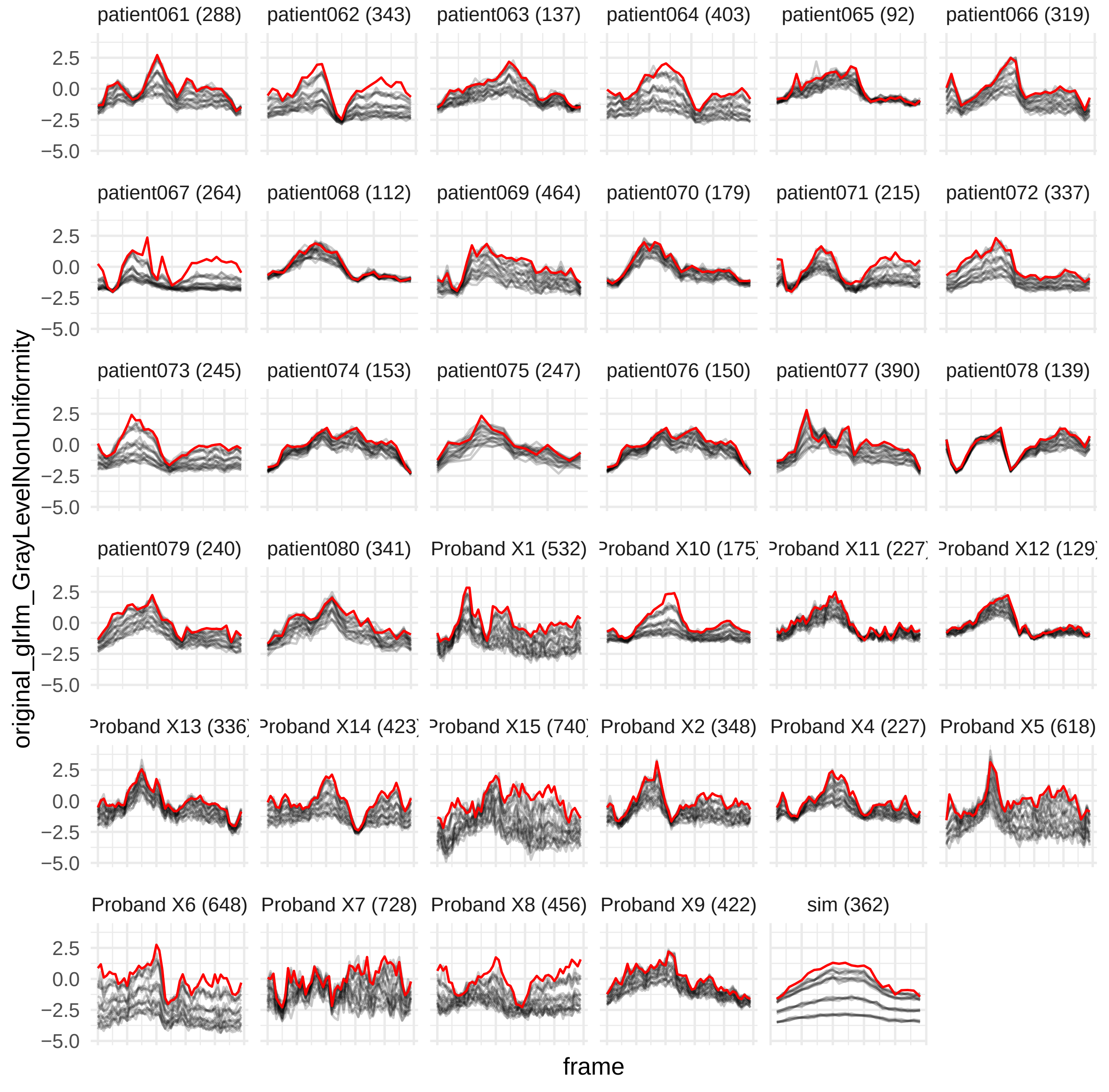
patient061 (288)
patient062 (343)
patient063 (137)
patient064 (403)
patient065 (92)
patient066 (319)
patient067 (264)
patient068 (112)
patient069 (464)
patient070 (179)
patient071 (215)
patient072 (337)
patient073 (245)
patient074 (153)
patient075 (247)
patient076 (150)
patient077 (390)
patient078 (139)
patient079 (240)
patient080 (341)
Proband X1 (532)
Proband X10 (175)
Proband X11 (227)
Proband X12 (129)
Proband X13 (336)
Proband X14 (423)
Proband X15 (740)
Proband X2 (348)
Proband X4 (227)
Proband X5 (618)
Proband X6 (648)
Proband X7 (728)
Proband X8 (456)
Proband X9 (422)
sim (362)
2.5
0.0
−2.5
−5.0
original_glrlm_GrayLevelNonUniformity
frame

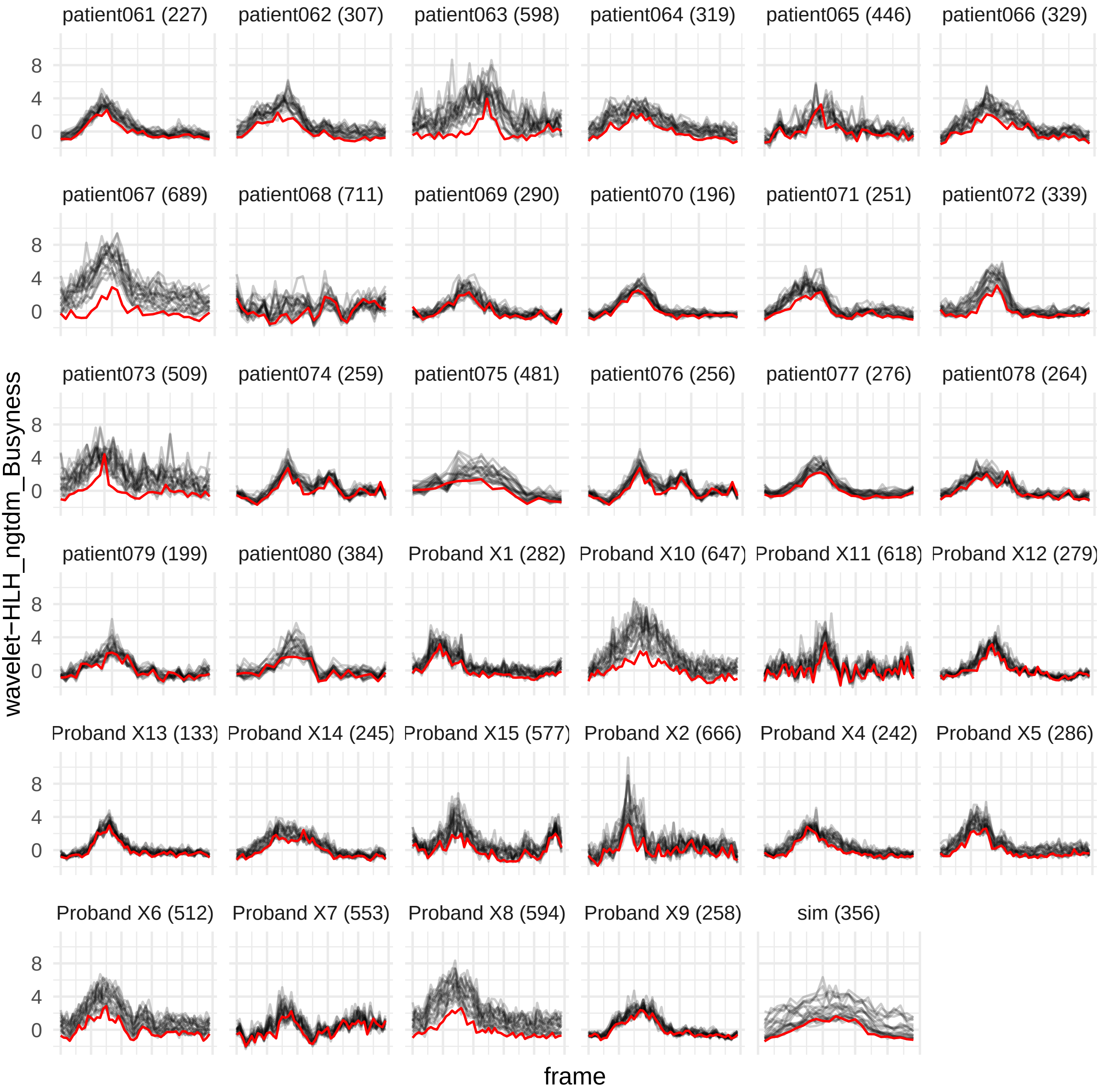
patient061 (227)
patient062 (307)
patient063 (598)
patient064 (319)
patient065 (446)
patient066 (329)
patient067 (689)
patient068 (711)
patient069 (290)
patient070 (196)
patient071 (251)
patient072 (339)
patient073 (509)
patient074 (259)
patient075 (481)
patient076 (256)
patient077 (276)
patient078 (264)
patient079 (199)
patient080 (384)
Proband X1 (282)
Proband X10 (647)
Proband X11 (618)
Proband X12 (279)
Proband X13 (133)
Proband X14 (245)
Proband X15 (577)
Proband X2 (666)
Proband X4 (242)
Proband X5 (286)
Proband X6 (512)
Proband X7 (553)
Proband X8 (594)
Proband X9 (258)
sim (356)
8
4
0
wavelet−HLH_ngtdm_Busyness
frame

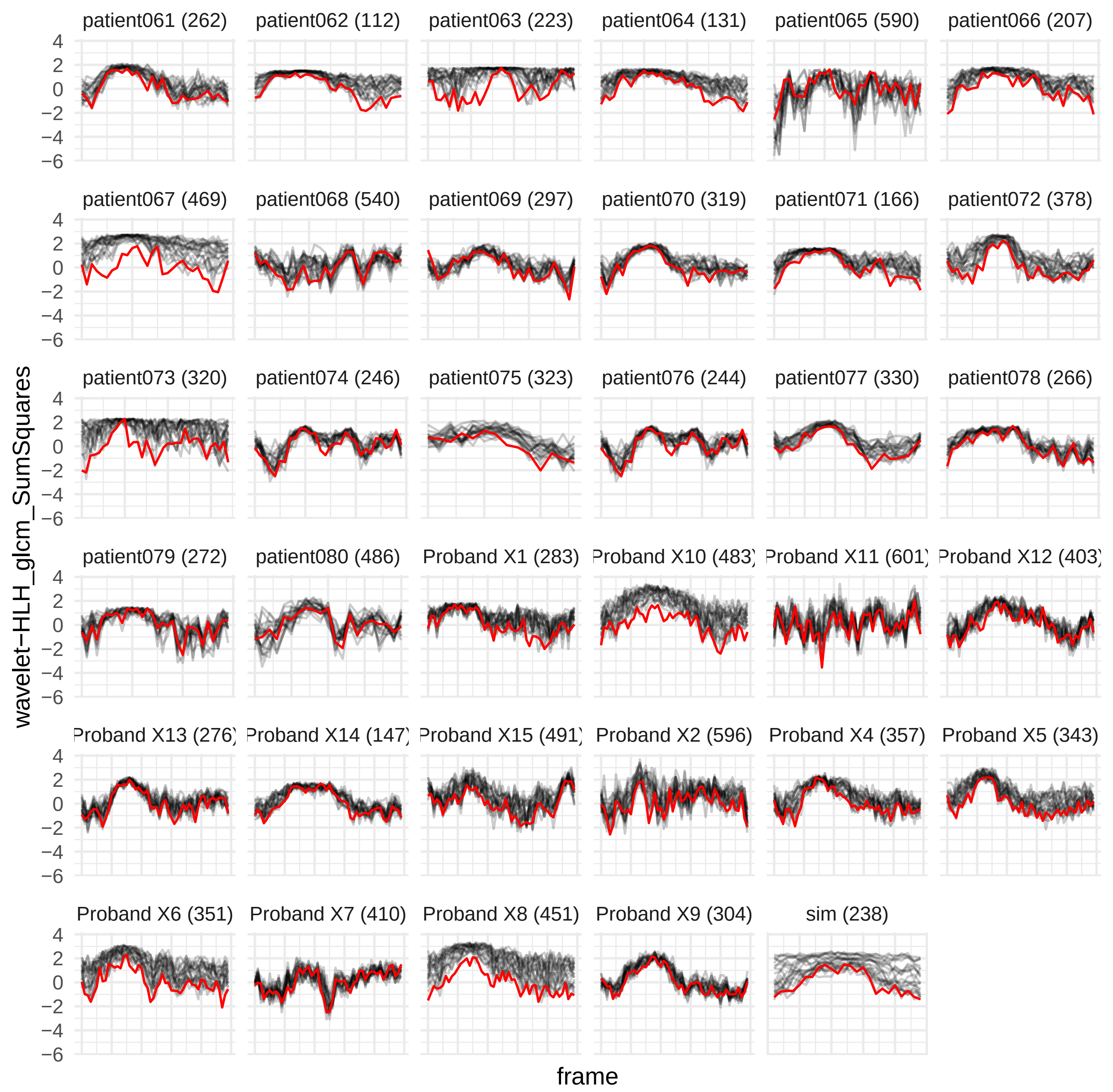
patient061 (262)
patient062 (112)
patient063 (223)
patient064 (131)
patient065 (590)
patient066 (207)
patient067 (469)
patient068 (540)
patient069 (297)
patient070 (319)
patient071 (166)
patient072 (378)
patient073 (320)
patient074 (246)
patient075 (323)
patient076 (244)
patient077 (330)
patient078 (266)
patient079 (272)
patient080 (486)
Proband X1 (283)
Proband X10 (483)
Proband X11 (601)
Proband X12 (403)
Proband X13 (276)
Proband X14 (147)
Proband X15 (491)
Proband X2 (596)
Proband X4 (357)
Proband X5 (343)
Proband X6 (351)
Proband X7 (410)
Proband X8 (451)
Proband X9 (304)
sim (238)
wavelet−HLH_glcm_SumSquares
frame
4
2
0
−2
−4
−6

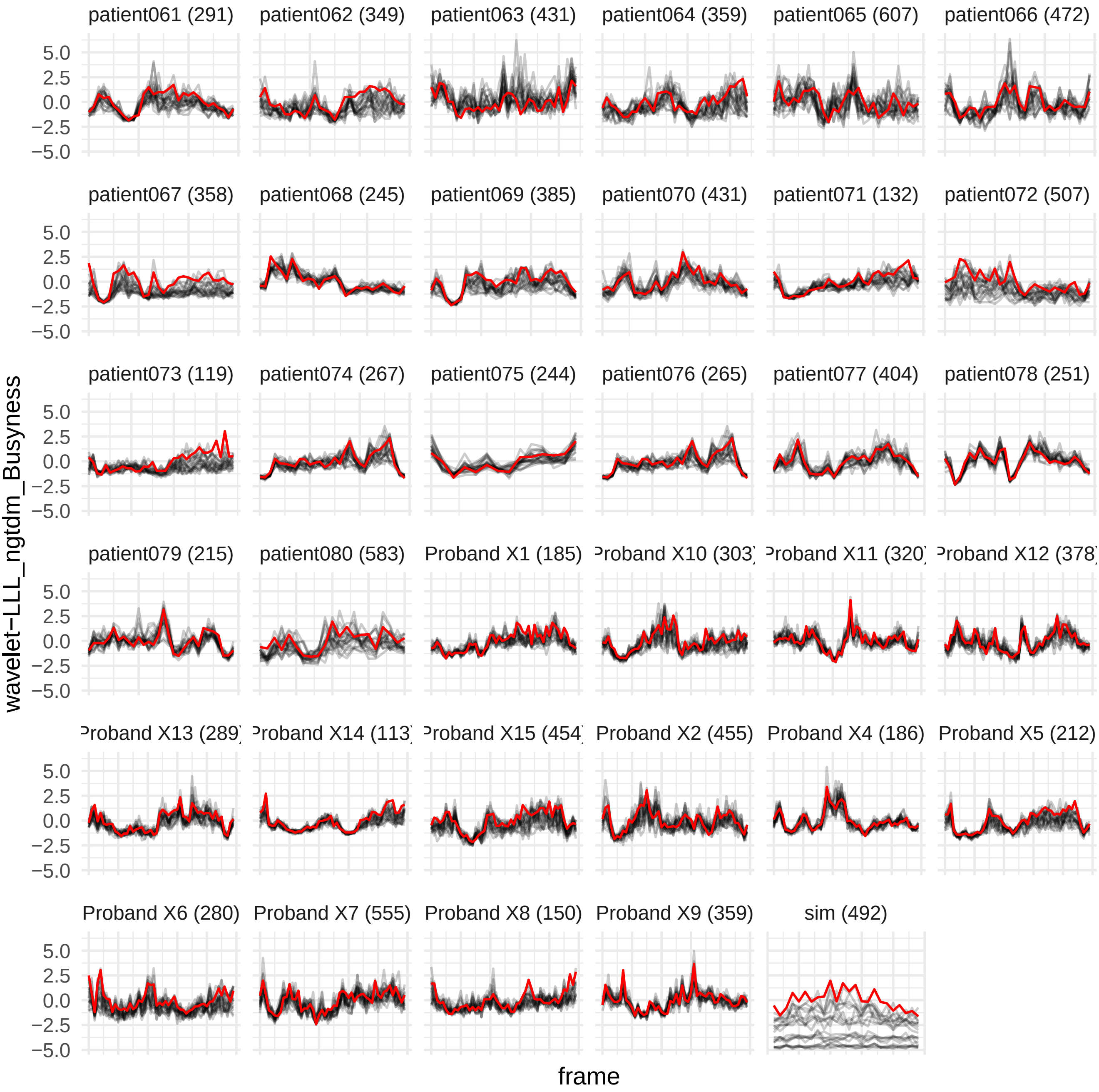
patient061 (291)
patient062 (349)
patient063 (431)
patient064 (359)
patient065 (607)
patient066 (472)
patient067 (358)
patient068 (245)
patient069 (385)
patient070 (431)
patient071 (132)
patient072 (507)
patient073 (119)
patient074 (267)
patient075 (244)
patient076 (265)
patient077 (404)
patient078 (251)
patient079 (215)
patient080 (583)
Proband X1 (185)
Proband X10 (303)
Proband X11 (320)
Proband X12 (378)
Proband X13 (289)
Proband X14 (113)
Proband X15 (454)
Proband X2 (455)
Proband X4 (186)
Proband X5 (212)
Proband X6 (280)
Proband X7 (555)
Proband X8 (150)
Proband X9 (359)
sim (492)
5.0
2.5
0.0
−2.5
−5.0
wavelet−LLL_ngtdm_Busyness
frame

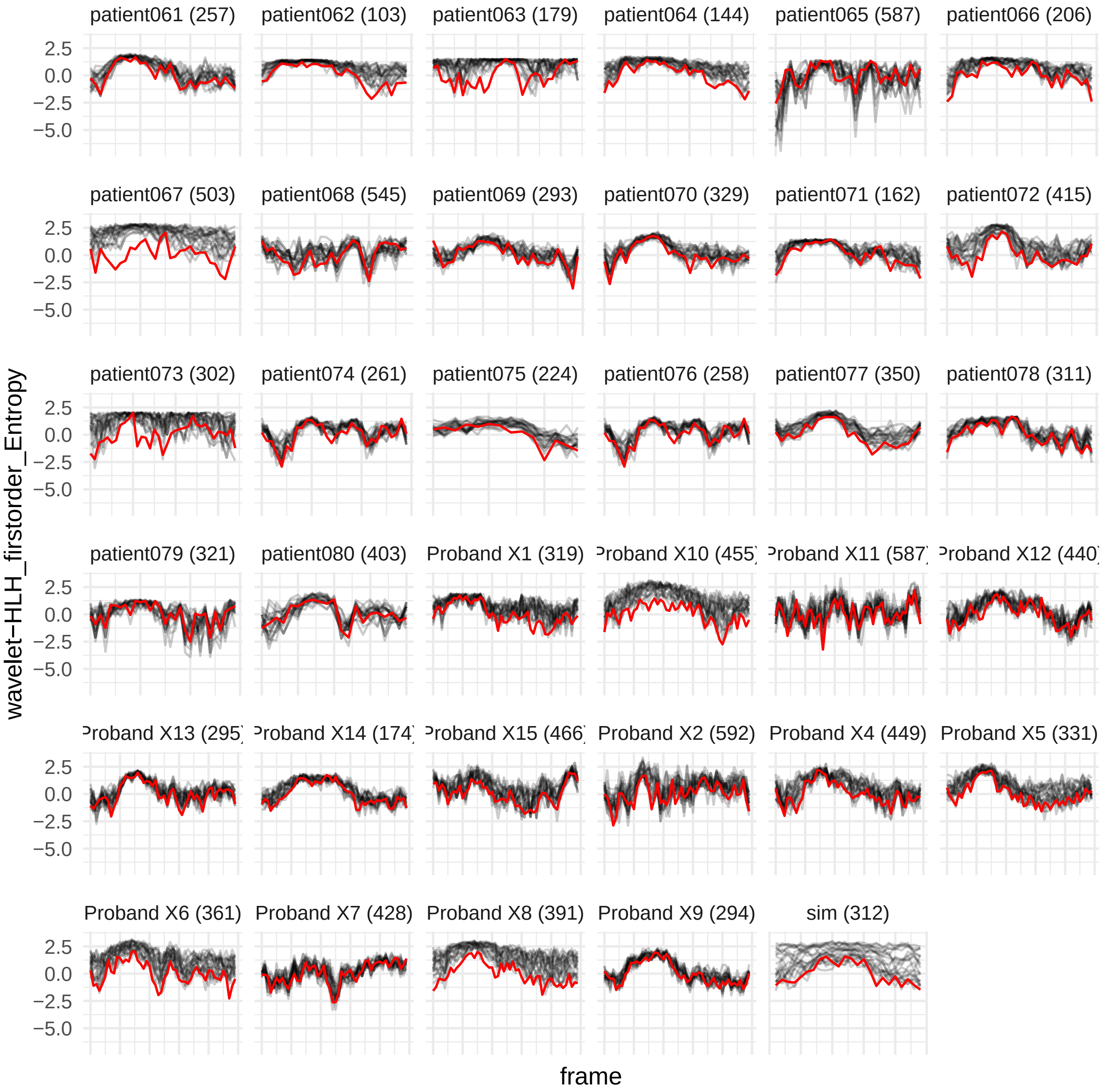
patient061 (257)
patient062 (103)
patient063 (179)
patient064 (144)
patient065 (587)
patient066 (206)
patient067 (503)
patient068 (545)
patient069 (293)
patient070 (329)
patient071 (162)
patient072 (415)
patient073 (302)
patient074 (261)
patient075 (224)
patient076 (258)
patient077 (350)
patient078 (311)
patient079 (321)
patient080 (403)
Proband X1 (319)
Proband X10 (455)
Proband X11 (587)
Proband X12 (440)
Proband X13 (295)
Proband X14 (174)
Proband X15 (466)
Proband X2 (592)
Proband X4 (449)
Proband X5 (331)
Proband X6 (361)
Proband X7 (428)
Proband X8 (391)
Proband X9 (294)
sim (312)
2.5
0.0
−2.5
−5.0
wavelet−HLH_firstorder_Entropy
frame

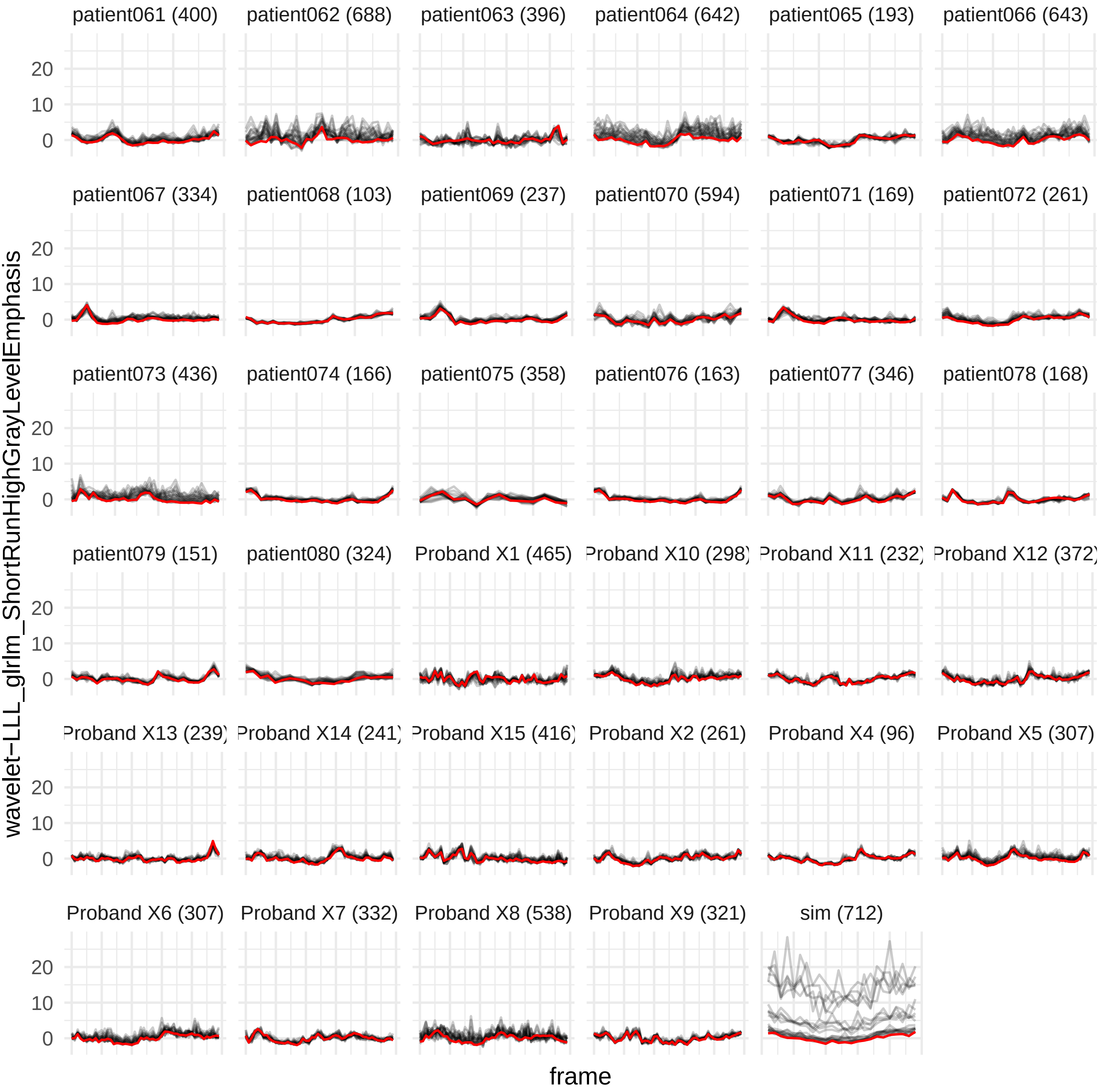

patient061 (400)
patient062 (688)
patient063 (396)
patient064 (642)
patient065 (193)
patient066 (643)
patient067 (334)
patient068 (103)
patient069 (237)
patient070 (594)
patient071 (169)
patient072 (261)
patient073 (436)
patient074 (166)
patient075 (358)
patient076 (163)
patient077 (346)
patient078 (168)
patient079 (151)
patient080 (324)
Proband X1 (465)
Proband X10 (298)
Proband X11 (232)
Proband X12 (372)
Proband X13 (239)
Proband X14 (241)
Proband X15 (416)
Proband X2 (261)
Proband X4 (96)
Proband X5 (307)
Proband X6 (307)
Proband X7 (332)
Proband X8 (538)
Proband X9 (321)
sim (712)
0
10
20
wavelet−LLL_glrlm_ShortRunHighGrayLevelEmphasis
frame

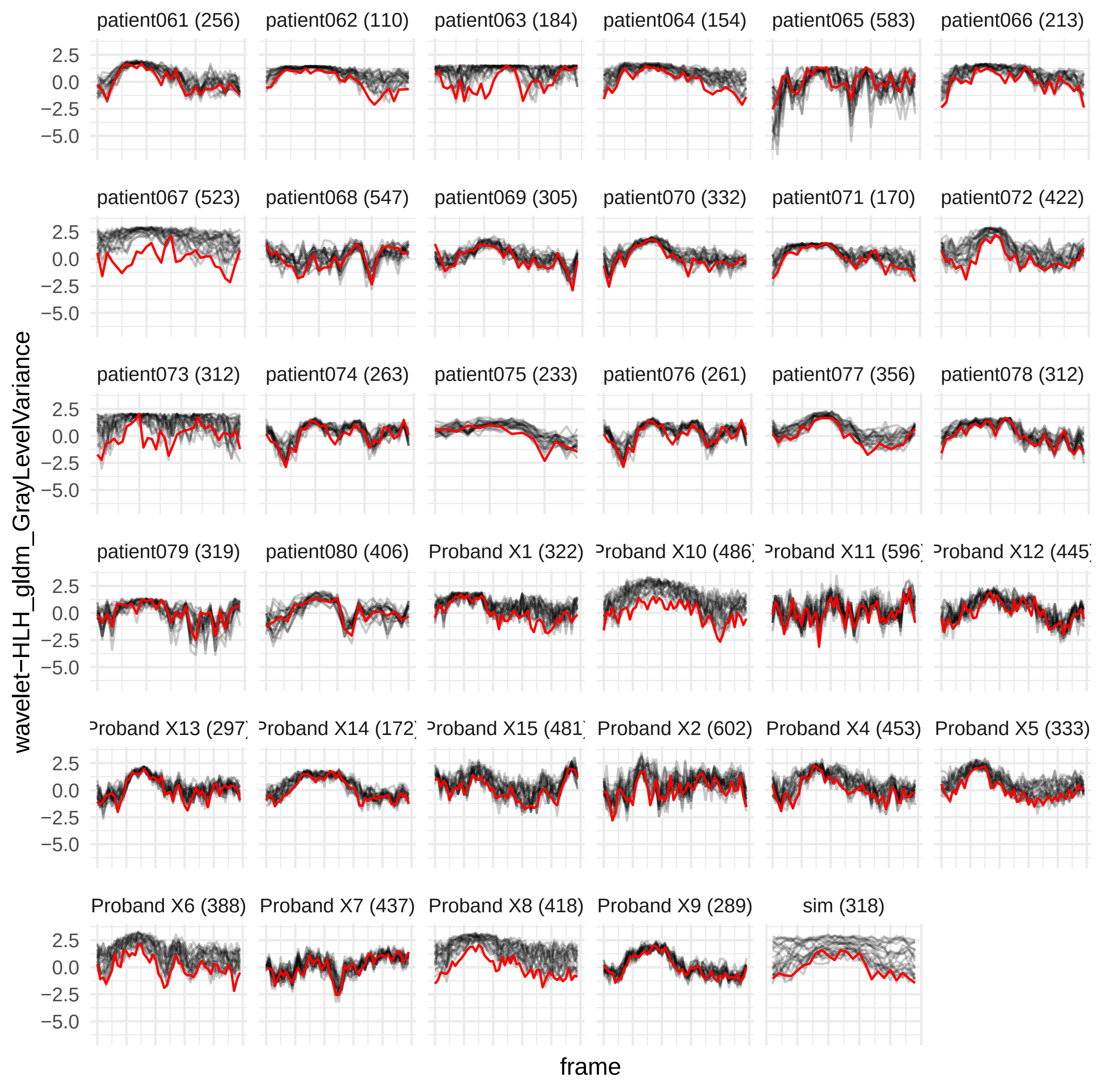
patient061 (256)
patient062 (110)
patient063 (184)
patient064 (154)
patient065 (583)
patient066 (213)
patient067 (523)
patient068 (547)
patient069 (305)
patient070 (332)
patient071 (170)
patient072 (422)
patient073 (312)
patient074 (263)
patient075 (233)
patient076 (261)
patient077 (356)
patient078 (312)
patient079 (319)
patient080 (406)
Proband X1 (322)
Proband X10 (486)
Proband X11 (596)
Proband X12 (445)
Proband X13 (297)
Proband X14 (172)
Proband X15 (481)
Proband X2 (602)
Proband X4 (453)
Proband X5 (333)
Proband X6 (388)
Proband X7 (437)
Proband X8 (418)
Proband X9 (289)
sim (318)
2.5
0.0
−2.5
−5.0
wavelet−HLH_gldm_GrayLevelVariance
frame

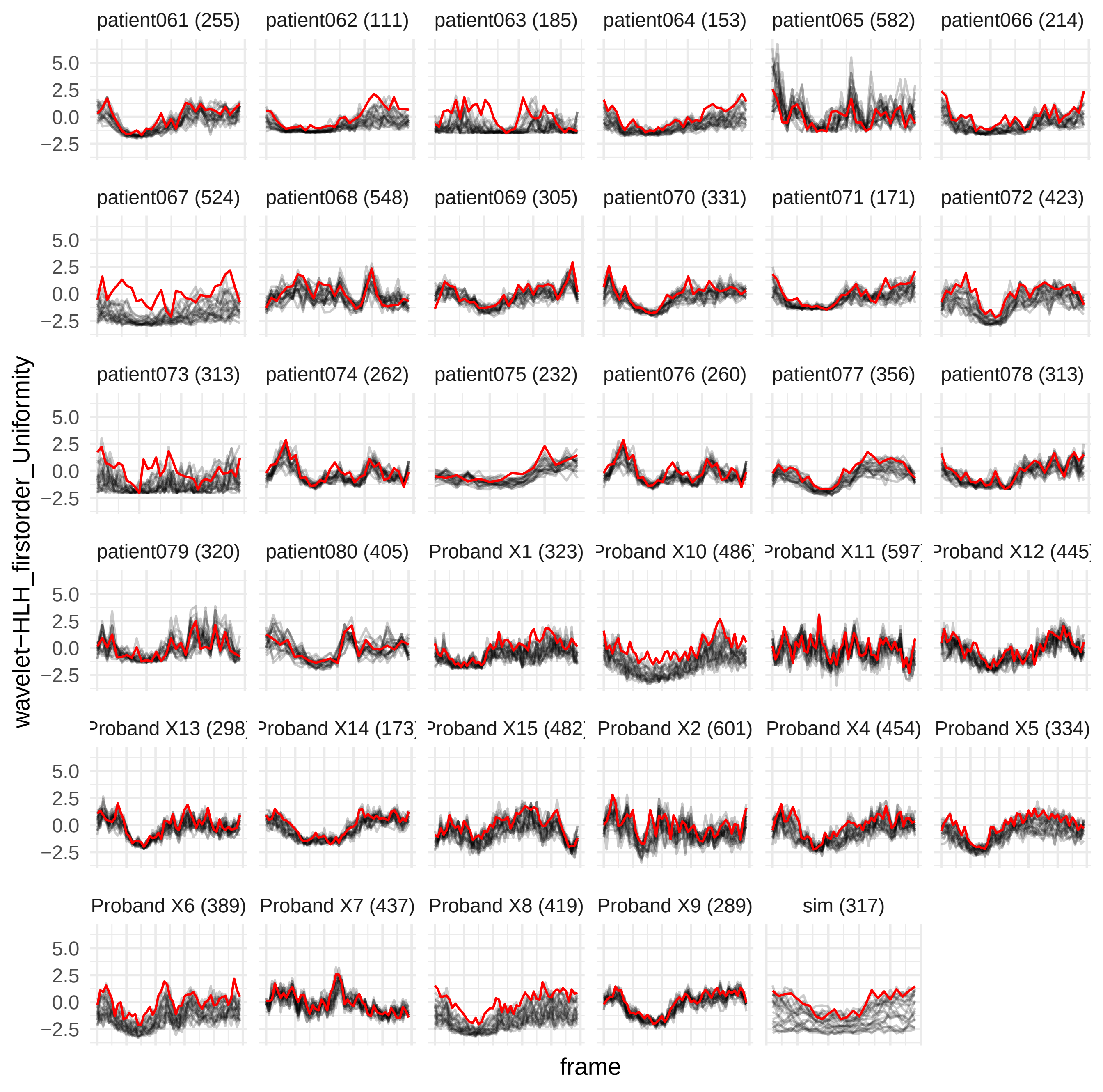

patient061 (255)
patient062 (111)
patient063 (185)
patient064 (153)
patient065 (582)
patient066 (214)
patient067 (524)
patient068 (548)
patient069 (305)
patient070 (331)
patient071 (171)
patient072 (423)
patient073 (313)
patient074 (262)
patient075 (232)
patient076 (260)
patient077 (356)
patient078 (313)
patient079 (320)
patient080 (405)
Proband X1 (323)
roband X10 (486)
roband X11 (597)
roband X12 (445)
roband X13 (298)
roband X14 (173)
roband X15 (482)
Proband X2 (601)
Proband X4 (454)
Proband X5 (334)
Proband X6 (389)
Proband X7 (437)
Proband X8 (419)
Proband X9 (289)
sim (317)
5.0
2.5
0.0
−2.5
wavelet−HLH_firstorder_Uniformity
frame

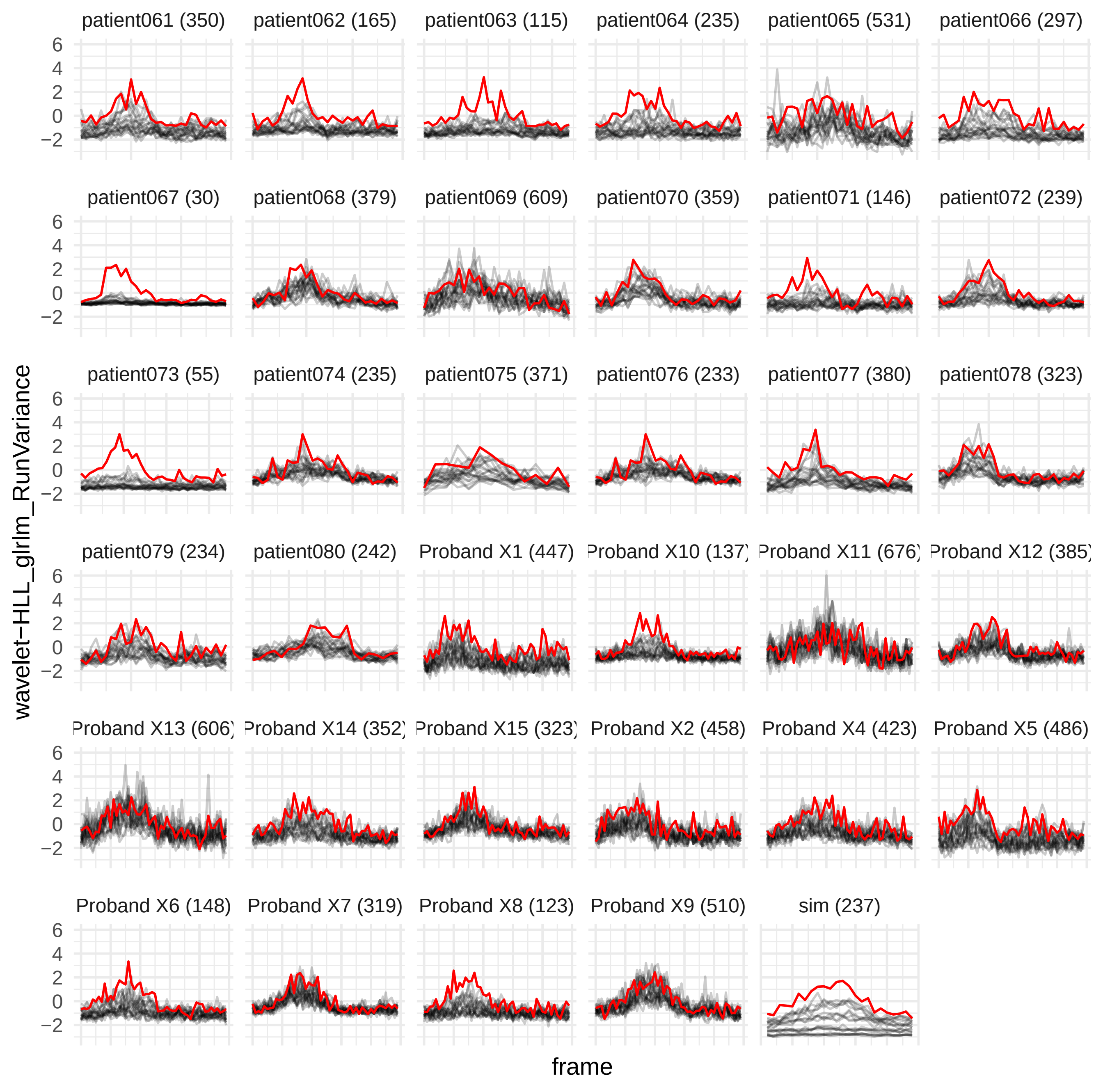
patient061 (350)
patient062 (165)
patient063 (115)
patient064 (235)
patient065 (531)
patient066 (297)
patient067 (30)
patient068 (379)
patient069 (609)
patient070 (359)
patient071 (146)
patient072 (239)
patient073 (55)
patient074 (235)
patient075 (371)
patient076 (233)
patient077 (380)
patient078 (323)
patient079 (234)
patient080 (242)
Proband X1 (447)
Proband X10 (137)
Proband X11 (676)
Proband X12 (385)
Proband X13 (606)
Proband X14 (352)
Proband X15 (323)
Proband X2 (458)
Proband X4 (423)
Proband X5 (486)
Proband X6 (148)
Proband X7 (319)
Proband X8 (123)
Proband X9 (510)
sim (237)
wavelet−HLL_glrlm_RunVariance
frame

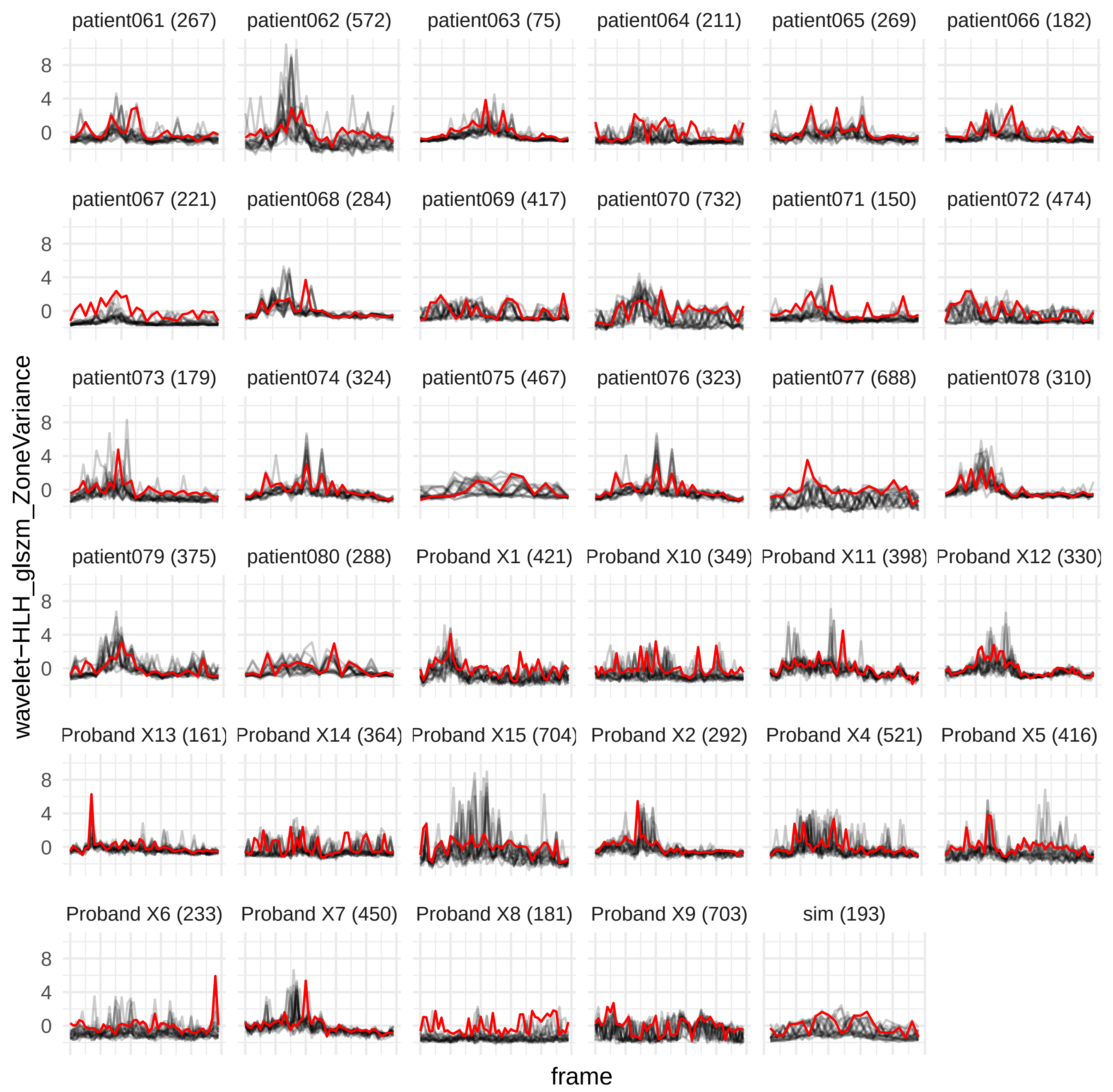

patient061 (267)
patient062 (572)
patient063 (75)
patient064 (211)
patient065 (269)
patient066 (182)
patient067 (221)
patient068 (284)
patient069 (417)
patient070 (732)
patient071 (150)
patient072 (474)
patient073 (179)
patient074 (324)
patient075 (467)
patient076 (323)
patient077 (688)
patient078 (310)
patient079 (375)
patient080 (288)
Proband X1 (421)
Proband X10 (349)
Proband X11 (398)
Proband X12 (330)
Proband X13 (161)
Proband X14 (364)
Proband X15 (704)
Proband X2 (292)
Proband X4 (521)
Proband X5 (416)
Proband X6 (233)
Proband X7 (450)
Proband X8 (181)
Proband X9 (703)
sim (193)
wavelet−HLH_glszm_ZoneVariance
8
4
0
frame

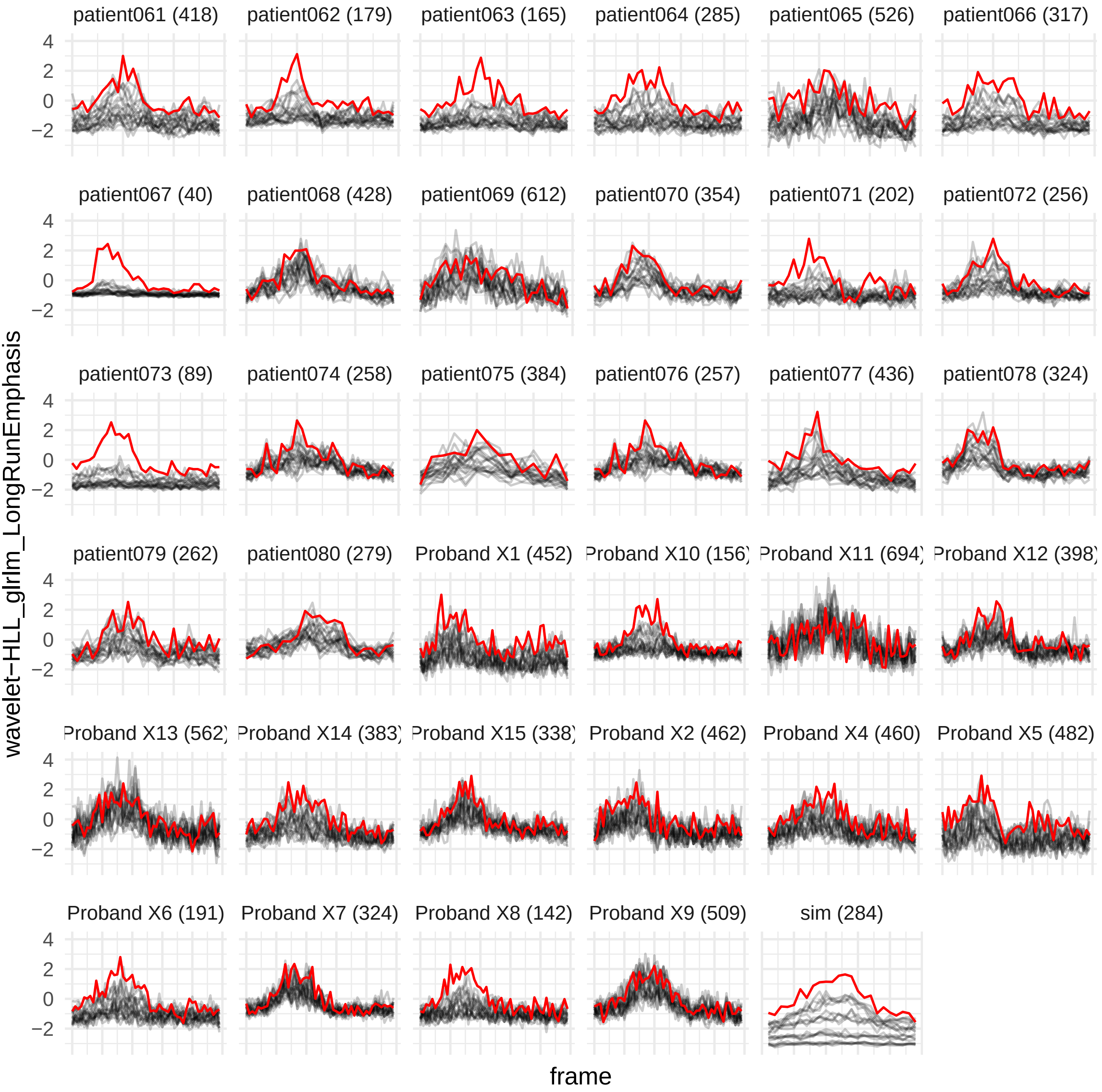

patient061 (418)
patient062 (179)
patient063 (165)
patient064 (285)
patient065 (526)
patient066 (317)
patient067 (40)
patient068 (428)
patient069 (612)
patient070 (354)
patient071 (202)
patient072 (256)
patient073 (89)
patient074 (258)
patient075 (384)
patient076 (257)
patient077 (436)
patient078 (324)
patient079 (262)
patient080 (279)
Proband X1 (452)
Proband X10 (156)
Proband X11 (694)
Proband X12 (398)
Proband X13 (562)
Proband X14 (383)
Proband X15 (338)
Proband X2 (462)
Proband X4 (460)
Proband X5 (482)
Proband X6 (191)
Proband X7 (324)
Proband X8 (142)
Proband X9 (509)
sim (284)
4
2
0
−2
wavelet−HLL_glrlm_LongRunEmphasis
frame

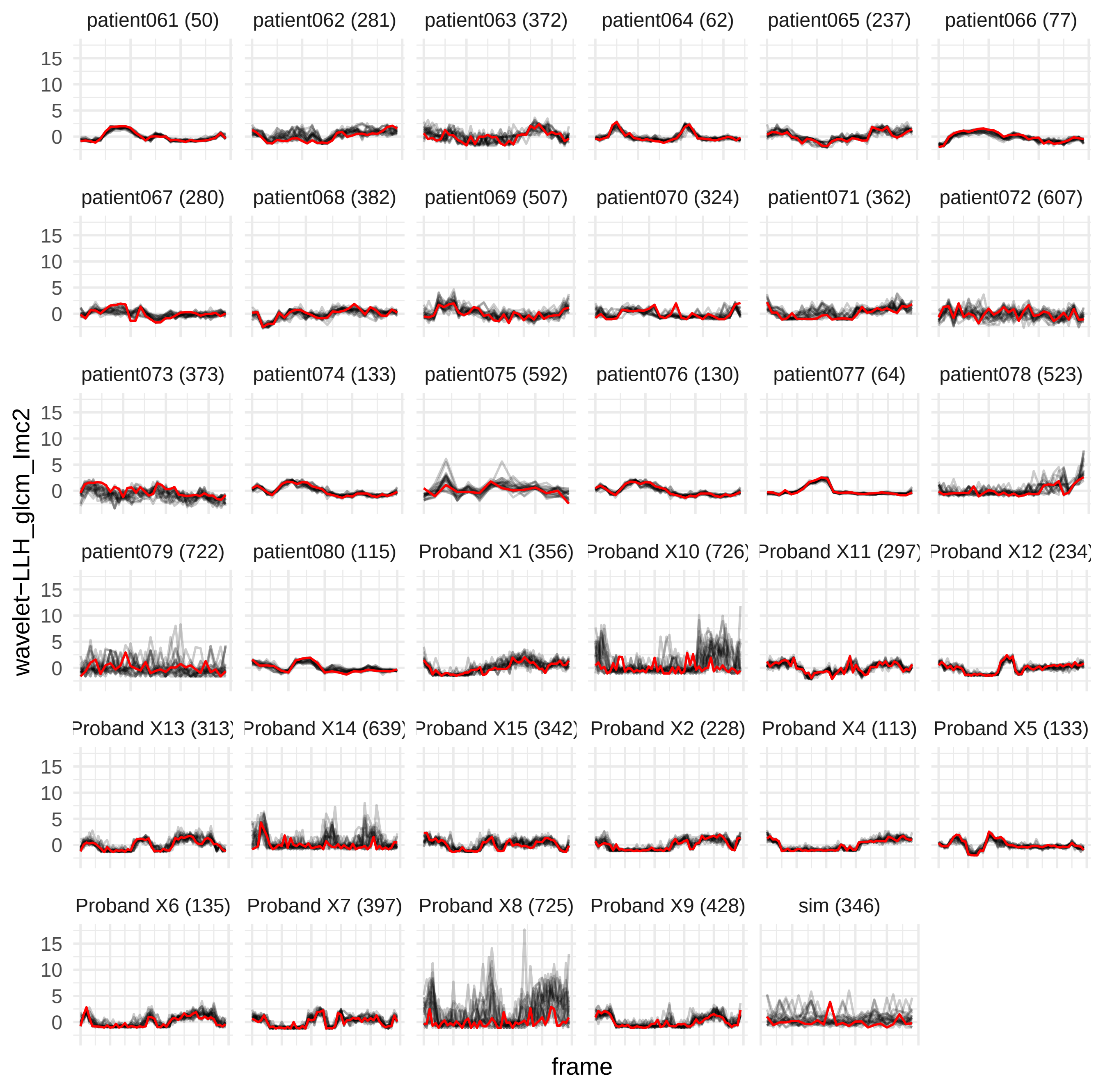

patient061 (50)
patient062 (281)
patient063 (372)
patient064 (62)
patient065 (237)
patient066 (77)
patient067 (280)
patient068 (382)
patient069 (507)
patient070 (324)
patient071 (362)
patient072 (607)
patient073 (373)
patient074 (133)
patient075 (592)
patient076 (130)
patient077 (64)
patient078 (523)
patient079 (722)
patient080 (115)
Proband X1 (356)
Proband X10 (726)
Proband X11 (297)
Proband X12 (234)
Proband X13 (313)
Proband X14 (639)
Proband X15 (342)
Proband X2 (228)
Proband X4 (113)
Proband X5 (133)
Proband X6 (135)
Proband X7 (397)
Proband X8 (725)
Proband X9 (428)
sim (346)
wavelet−LLH_glcm_Imc2
15
10
5
0
frame

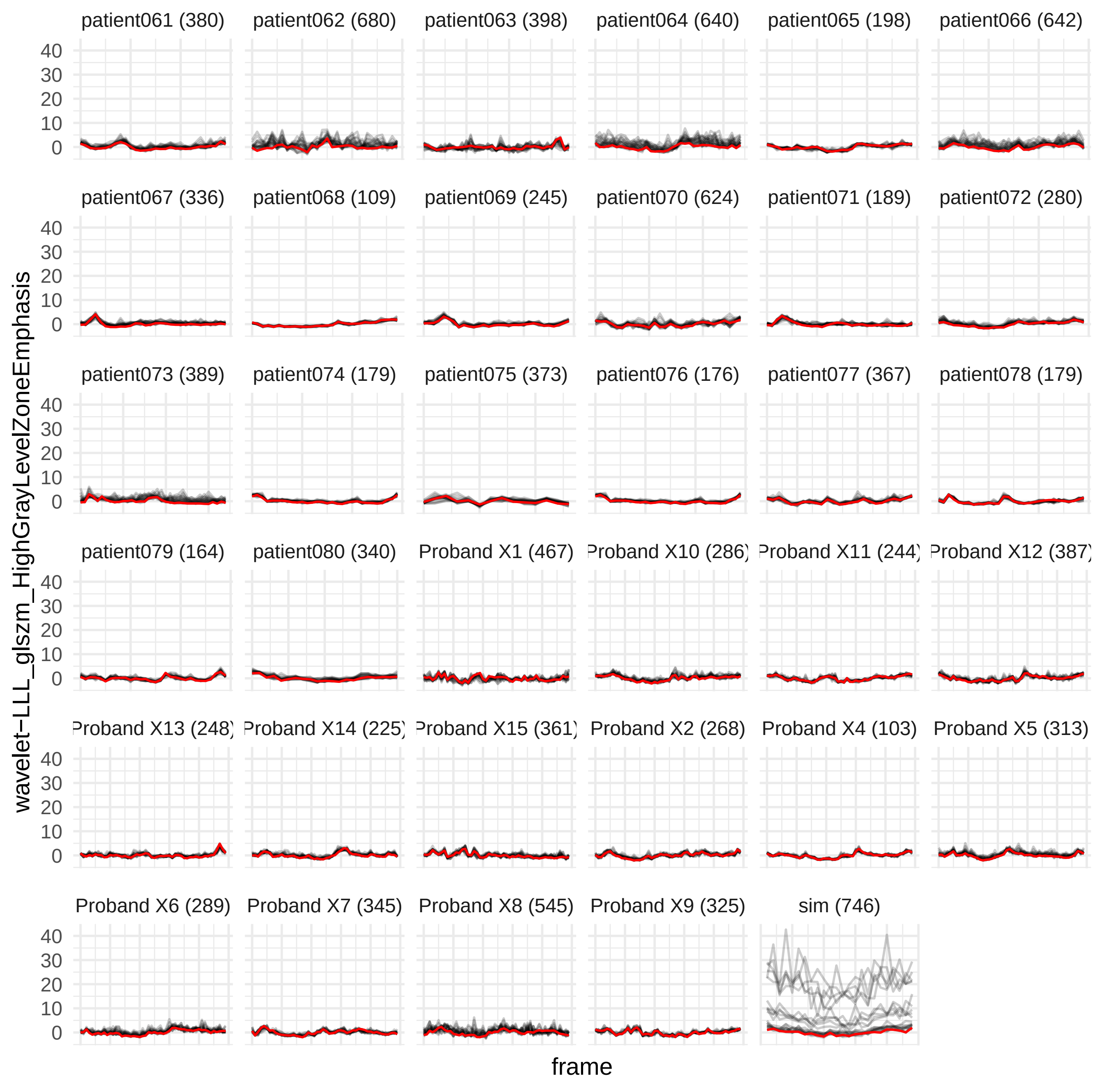

patient061 (380)
patient062 (680)
patient063 (398)
patient064 (640)
patient065 (198)
patient066 (642)
patient067 (336)
patient068 (109)
patient069 (245)
patient070 (624)
patient071 (189)
patient072 (280)
patient073 (389)
patient074 (179)
patient075 (373)
patient076 (176)
patient077 (367)
patient078 (179)
patient079 (164)
patient080 (340)
Proband X1 (467)
Proband X10 (286)
Proband X11 (244)
Proband X12 (387)
Proband X13 (248)
Proband X14 (225)
Proband X15 (361)
Proband X2 (268)
Proband X4 (103)
Proband X5 (313)
Proband X6 (289)
Proband X7 (345)
Proband X8 (545)
Proband X9 (325)
sim (746)
0
10
20
30
40
wavelet–LLL_glszm_HighGrayLevelZoneEmphasis
frame

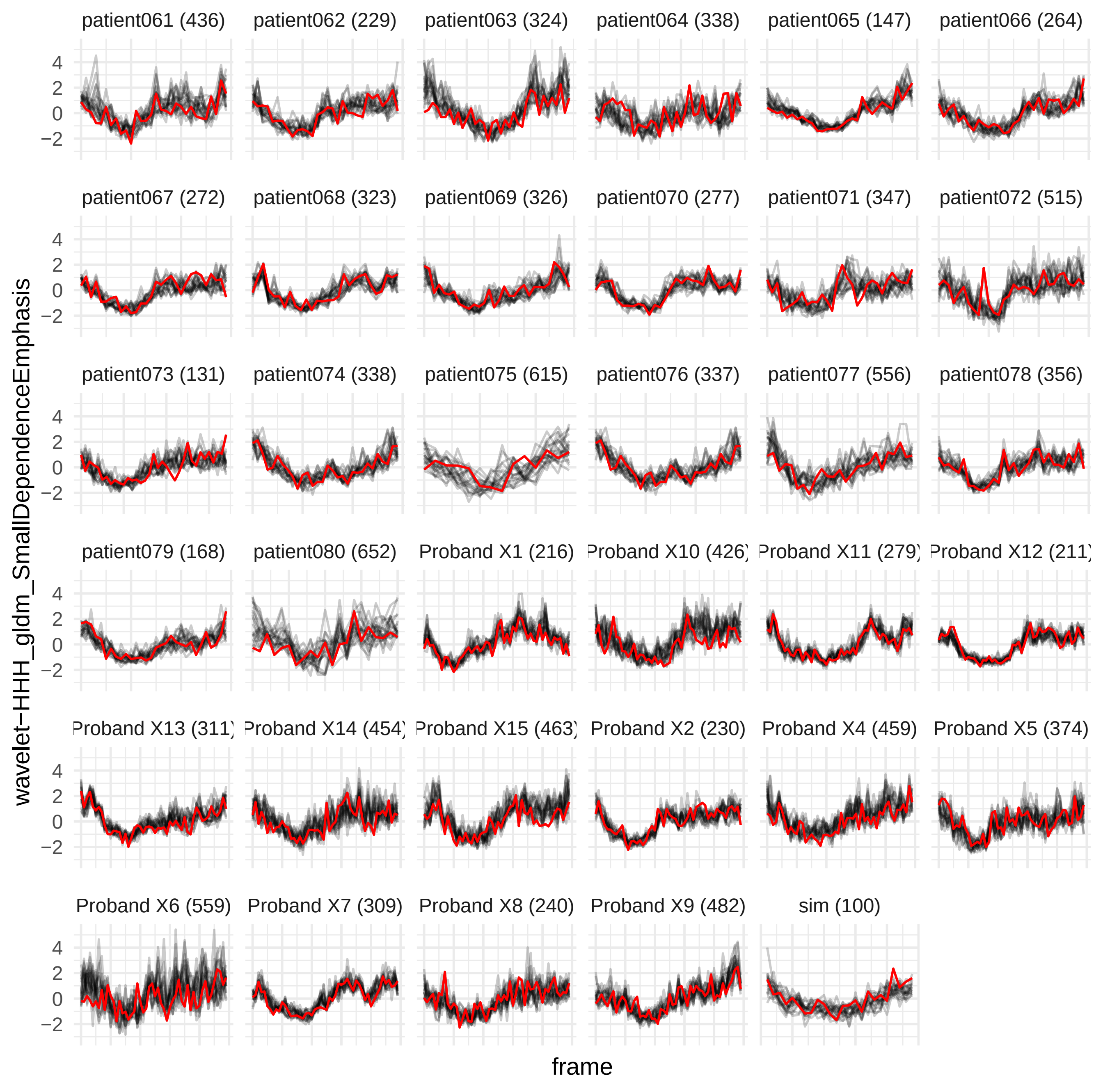

patient061 (436)
patient062 (229)
patient063 (324)
patient064 (338)
patient065 (147)
patient066 (264)
patient067 (272)
patient068 (323)
patient069 (326)
patient070 (277)
patient071 (347)
patient072 (515)
patient073 (131)
patient074 (338)
patient075 (615)
patient076 (337)
patient077 (556)
patient078 (356)
patient079 (168)
patient080 (652)
Proband X1 (216)
Proband X10 (426)
Proband X11 (279)
Proband X12 (211)
Proband X13 (311)
Proband X14 (454)
Proband X15 (463)
Proband X2 (230)
Proband X4 (459)
Proband X5 (374)
Proband X6 (559)
Proband X7 (309)
Proband X8 (240)
Proband X9 (482)
sim (100)
wavelet−HHH_gldm_SmallDependenceEmphasis
frame

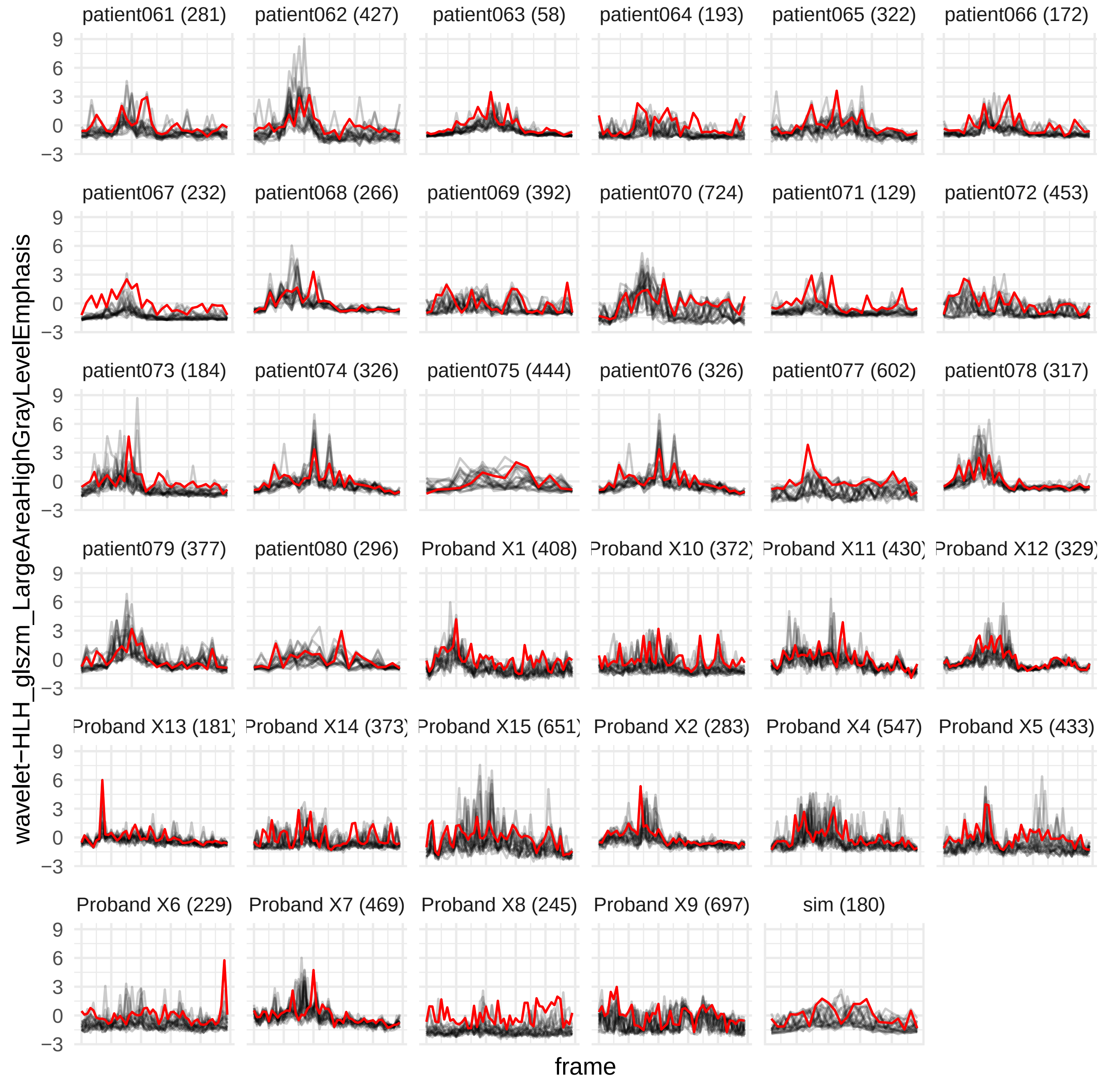
patient061 (281)
patient062 (427)
patient063 (58)
patient064 (193)
patient065 (322)
patient066 (172)
patient067 (232)
patient068 (266)
patient069 (392)
patient070 (724)
patient071 (129)
patient072 (453)
patient073 (184)
patient074 (326)
patient075 (444)
patient076 (326)
patient077 (602)
patient078 (317)
patient079 (377)
patient080 (296)
Proband X1 (408)
Proband X10 (372)
Proband X11 (430)
Proband X12 (329)
Proband X13 (181)
Proband X14 (373)
Proband X15 (651)
Proband X2 (283)
Proband X4 (547)
Proband X5 (433)
Proband X6 (229)
Proband X7 (469)
Proband X8 (245)
Proband X9 (697)
sim (180)
9
6
3
0
−3
wavelet−HLH_glszm_LargeAreaHighGrayLevelEmphasis
frame

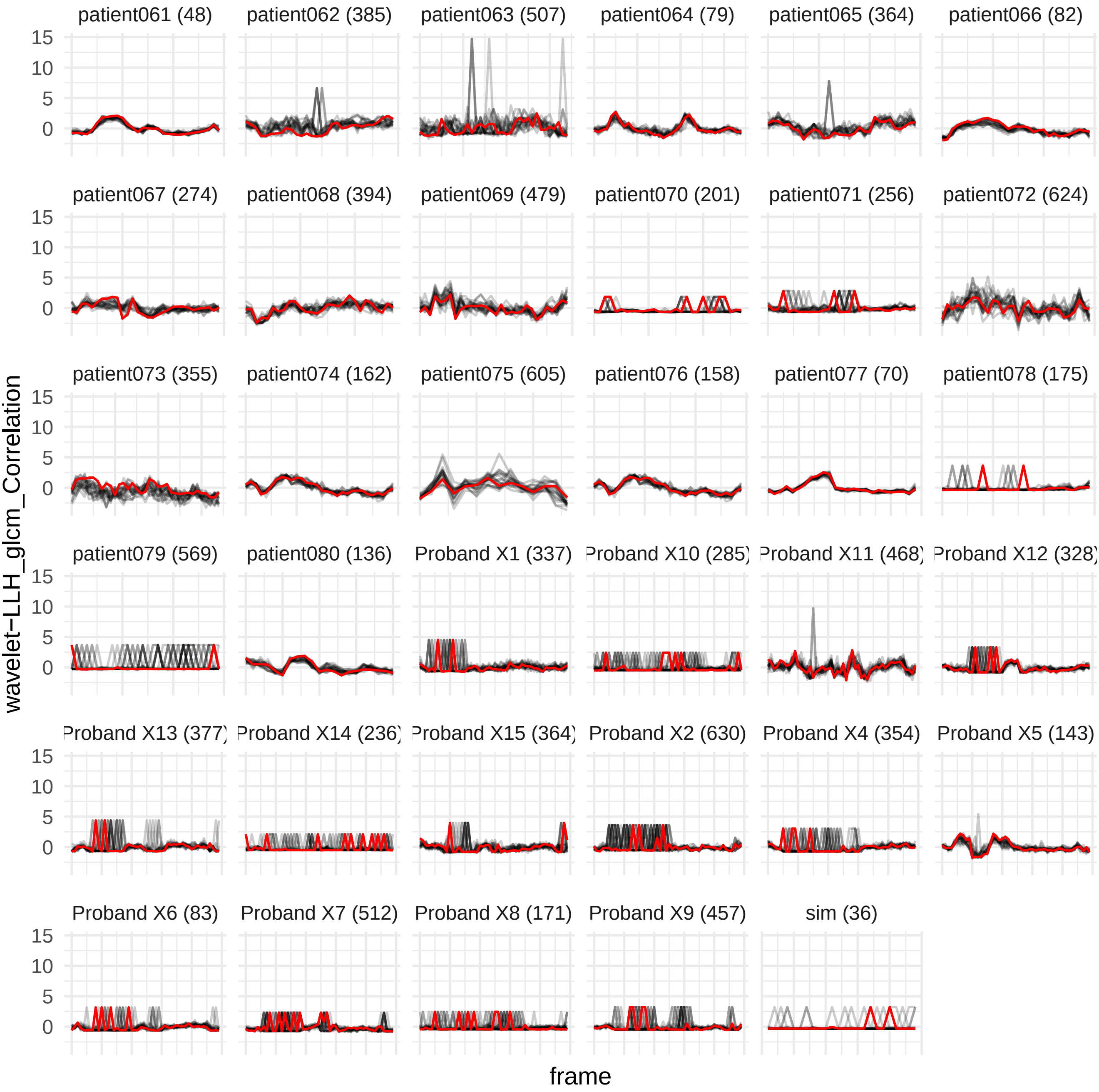
patient061 (48)
patient062 (385)
patient063 (507)
patient064 (79)
patient065 (364)
patient066 (82)
patient067 (274)
patient068 (394)
patient069 (479)
patient070 (201)
patient071 (256)
patient072 (624)
patient073 (355)
patient074 (162)
patient075 (605)
patient076 (158)
patient077 (70)
patient078 (175)
patient079 (569)
patient080 (136)
Proband X1 (337)
Proband X10 (285)
Proband X11 (468)
Proband X12 (328)
Proband X13 (377)
Proband X14 (236)
Proband X15 (364)
Proband X2 (630)
Proband X4 (354)
Proband X5 (143)
Proband X6 (83)
Proband X7 (512)
Proband X8 (171)
Proband X9 (457)
sim (36)
wavelet−LLH_glcm_Correlation
frame
15
10
5
0

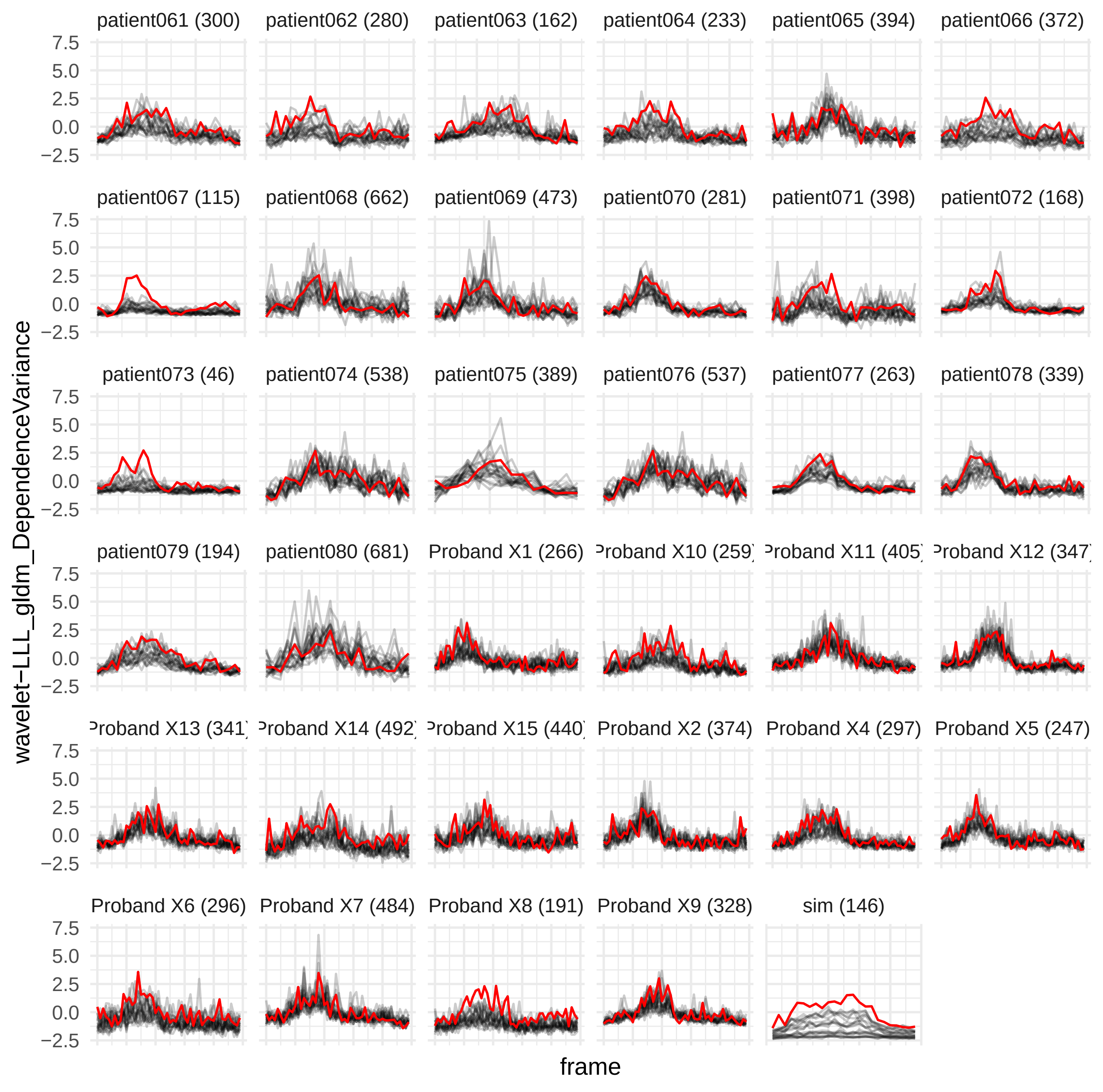

patient061 (300)
patient062 (280)
patient063 (162)
patient064 (233)
patient065 (394)
patient066 (372)
patient067 (115)
patient068 (662)
patient069 (473)
patient070 (281)
patient071 (398)
patient072 (168)
patient073 (46)
patient074 (538)
patient075 (389)
patient076 (537)
patient077 (263)
patient078 (339)
patient079 (194)
patient080 (681)
Proband X1 (266)
Proband X10 (259)
Proband X11 (405)
Proband X12 (347)
Proband X13 (341)
Proband X14 (492)
Proband X15 (440)
Proband X2 (374)
Proband X4 (297)
Proband X5 (247)
Proband X6 (296)
Proband X7 (484)
Proband X8 (191)
Proband X9 (328)
sim (146)
7.5
5.0
2.5
0.0
−2.5
wavelet−LLL_gldm_DependenceVariance
frame

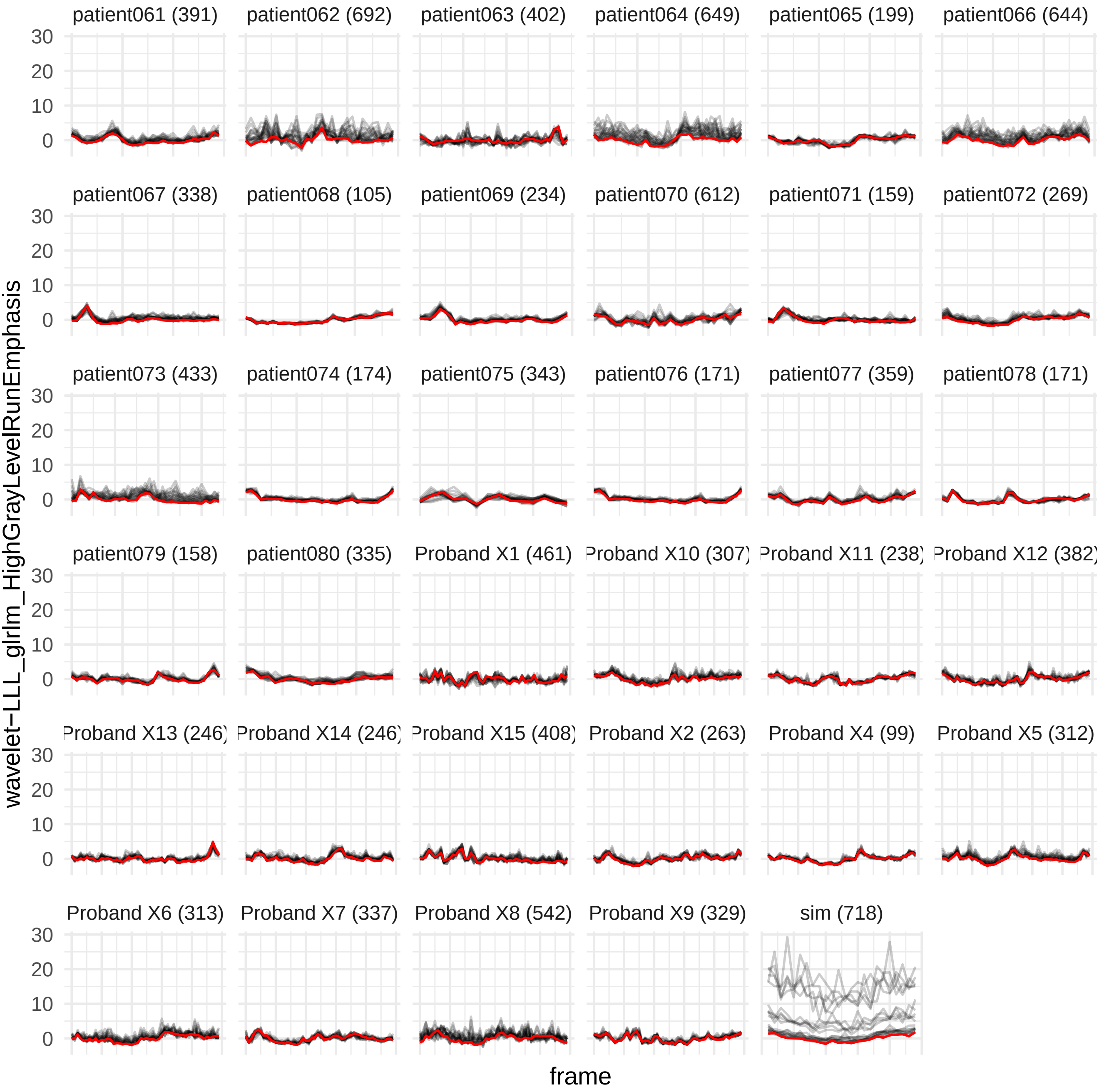

patient061 (391)
patient062 (692)
patient063 (402)
patient064 (649)
patient065 (199)
patient066 (644)
patient067 (338)
patient068 (105)
patient069 (234)
patient070 (612)
patient071 (159)
patient072 (269)
patient073 (433)
patient074 (174)
patient075 (343)
patient076 (171)
patient077 (359)
patient078 (171)
patient079 (158)
patient080 (335)
Proband X1 (461)
Proband X10 (307)
Proband X11 (238)
Proband X12 (382)
Proband X13 (246)
Proband X14 (246)
Proband X15 (408)
Proband X2 (263)
Proband X4 (99)
Proband X5 (312)
Proband X6 (313)
Proband X7 (337)
Proband X8 (542)
Proband X9 (329)
sim (718)
wavelet−LLL_glrlm_HighGrayLevelRunEmphasis
frame
0
10
20
30

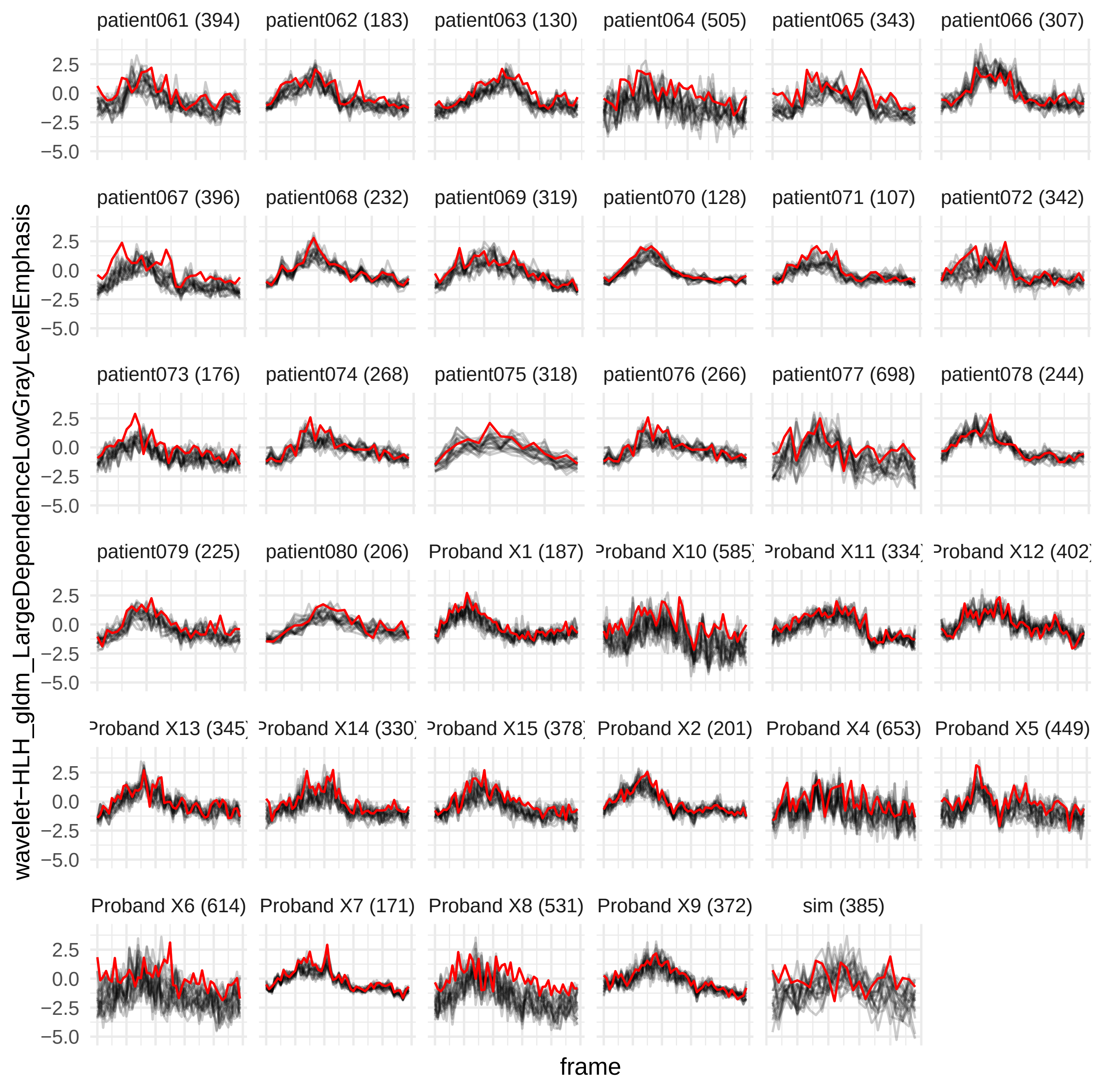

patient061 (394)
patient062 (183)
patient063 (130)
patient064 (505)
patient065 (343)
patient066 (307)
patient067 (396)
patient068 (232)
patient069 (319)
patient070 (128)
patient071 (107)
patient072 (342)
patient073 (176)
patient074 (268)
patient075 (318)
patient076 (266)
patient077 (698)
patient078 (244)
patient079 (225)
patient080 (206)
Proband X1 (187)
Proband X10 (585)
Proband X11 (334)
Proband X12 (402)
Proband X13 (345)
Proband X14 (330)
Proband X15 (378)
Proband X2 (201)
Proband X4 (653)
Proband X5 (449)
Proband X6 (614)
Proband X7 (171)
Proband X8 (531)
Proband X9 (372)
sim (385)
2.5
0.0
−2.5
−5.0
wavelet−HLH_gldm_LargeDependenceLowGrayLevelEmphasis
frame

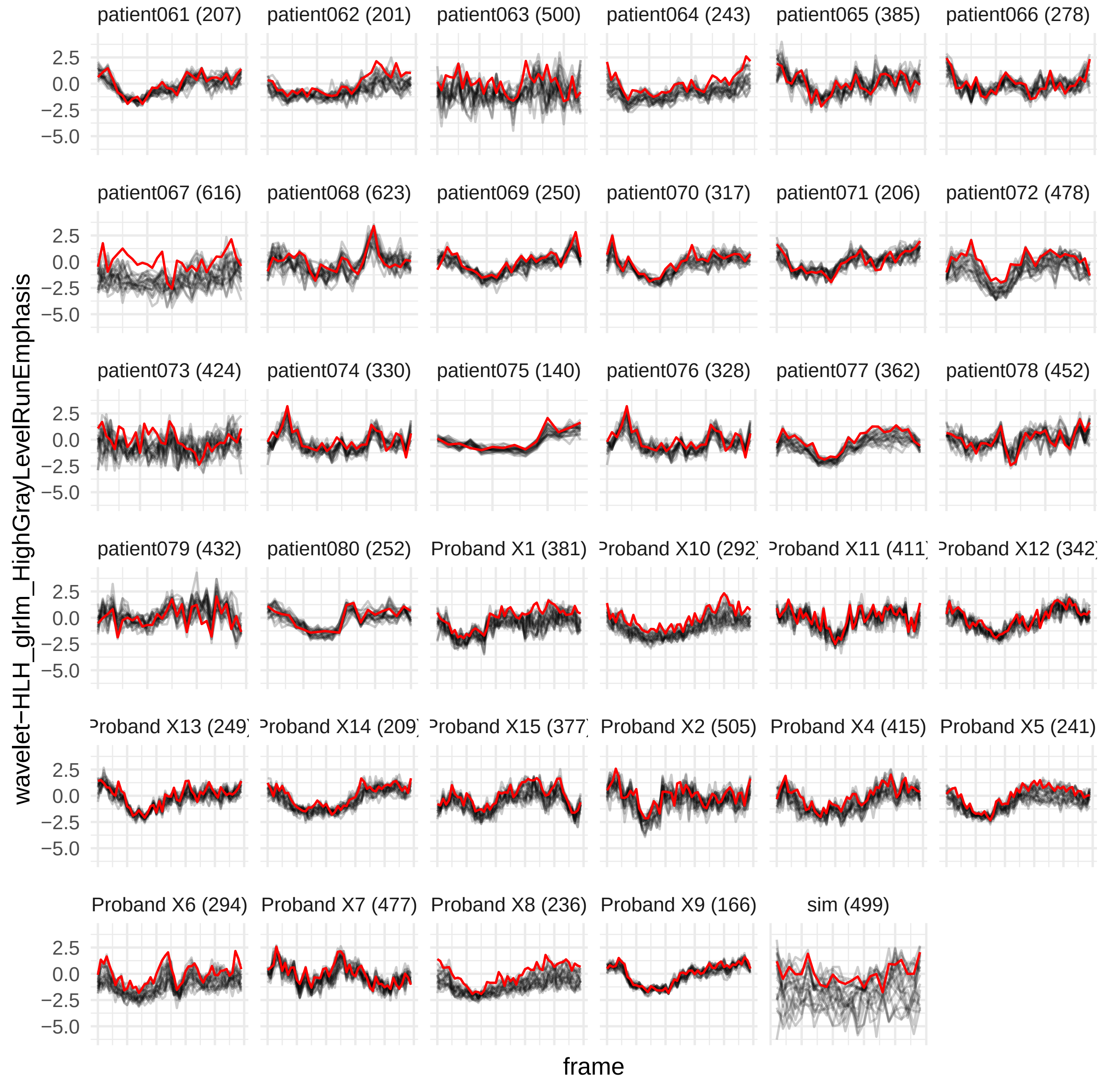
patient061 (207)
patient062 (201)
patient063 (500)
patient064 (243)
patient065 (385)
patient066 (278)
patient067 (616)
patient068 (623)
patient069 (250)
patient070 (317)
patient071 (206)
patient072 (478)
patient073 (424)
patient074 (330)
patient075 (140)
patient076 (328)
patient077 (362)
patient078 (452)
patient079 (432)
patient080 (252)
Proband X1 (381)
Proband X10 (292)
Proband X11 (411)
Proband X12 (342)
Proband X13 (249)
Proband X14 (209)
Proband X15 (377)
Proband X2 (505)
Proband X4 (415)
Proband X5 (241)
Proband X6 (294)
Proband X7 (477)
Proband X8 (236)
Proband X9 (166)
sim (499)
2.5
0.0
−2.5
−5.0
wavelet−HLH_glrlm_HighGrayLevelRunEmphasis
frame

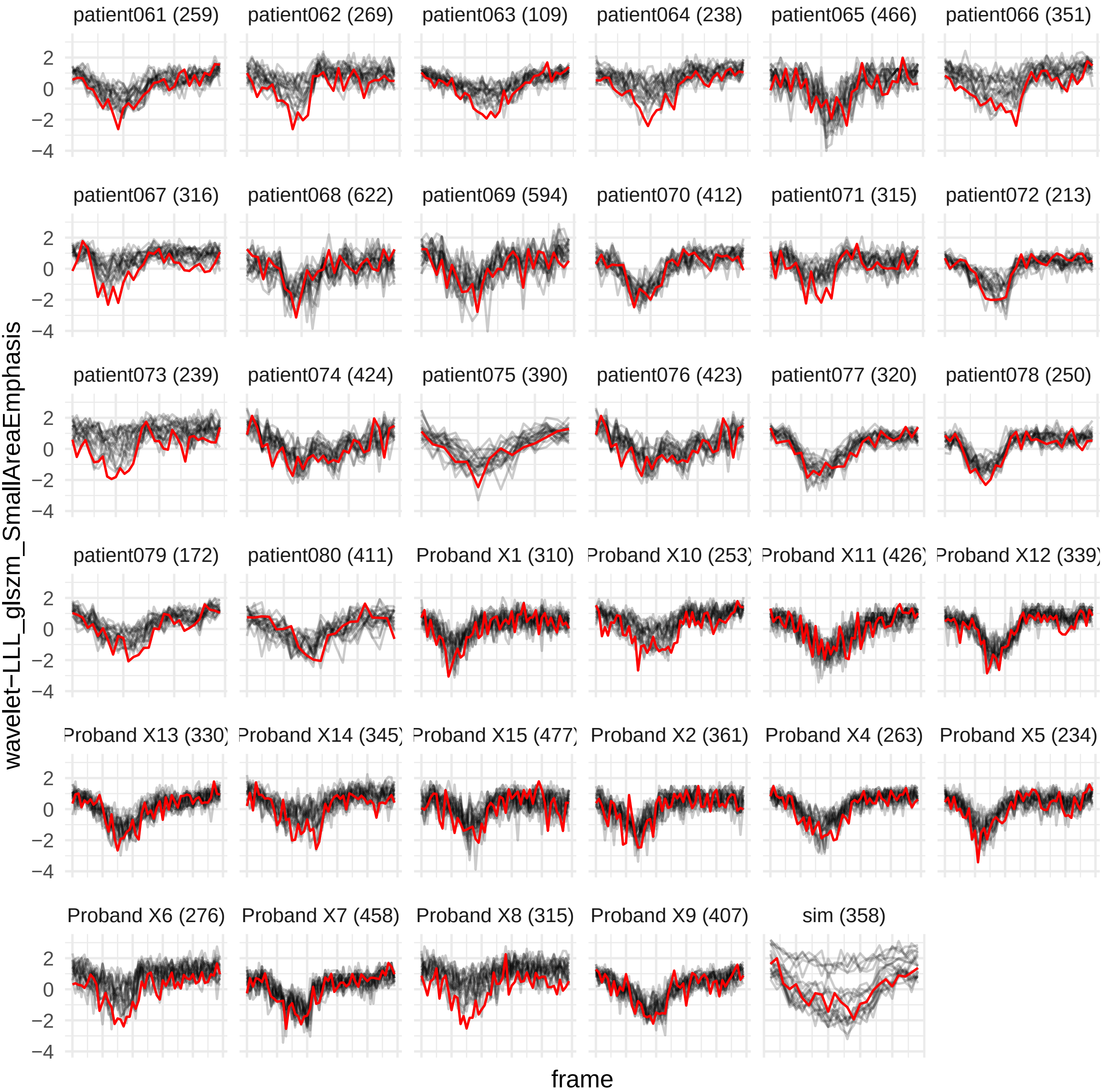
patient061 (259)
patient062 (269)
patient063 (109)
patient064 (238)
patient065 (466)
patient066 (351)
patient067 (316)
patient068 (622)
patient069 (594)
patient070 (412)
patient071 (315)
patient072 (213)
patient073 (239)
patient074 (424)
patient075 (390)
patient076 (423)
patient077 (320)
patient078 (250)
patient079 (172)
patient080 (411)
Proband X1 (310)
Proband X10 (253)
Proband X11 (426)
Proband X12 (339)
Proband X13 (330)
Proband X14 (345)
Proband X15 (477)
Proband X2 (361)
Proband X4 (263)
Proband X5 (234)
Proband X6 (276)
Proband X7 (458)
Proband X8 (315)
Proband X9 (407)
sim (358)
wavelet−LLL_glszm_SmallAreaEmphasis
frame
2
0
−2
−4

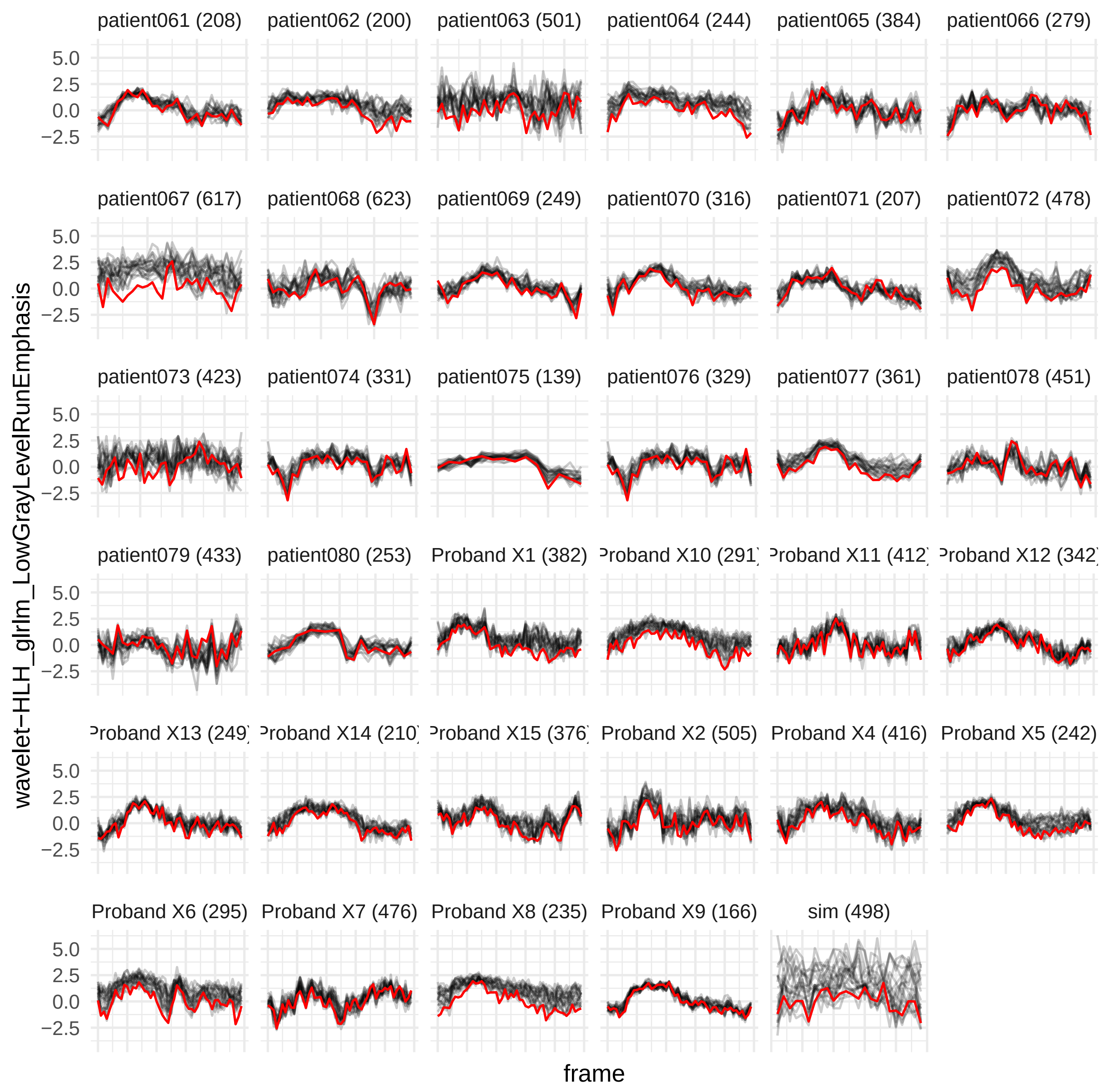
patient061 (208)
patient062 (200)
patient063 (501)
patient064 (244)
patient065 (384)
patient066 (279)
patient067 (617)
patient068 (623)
patient069 (249)
patient070 (316)
patient071 (207)
patient072 (478)
patient073 (423)
patient074 (331)
patient075 (139)
patient076 (329)
patient077 (361)
patient078 (451)
patient079 (433)
patient080 (253)
Proband X1 (382)
Proband X10 (291)
Proband X11 (412)
Proband X12 (342)
Proband X13 (249)
Proband X14 (210)
Proband X15 (376)
Proband X2 (505)
Proband X4 (416)
Proband X5 (242)
Proband X6 (295)
Proband X7 (476)
Proband X8 (235)
Proband X9 (166)
sim (498)
5.0
2.5
0.0
−2.5
wavelet−HLH_glrlm_LowGrayLevelRunEmphasis
frame

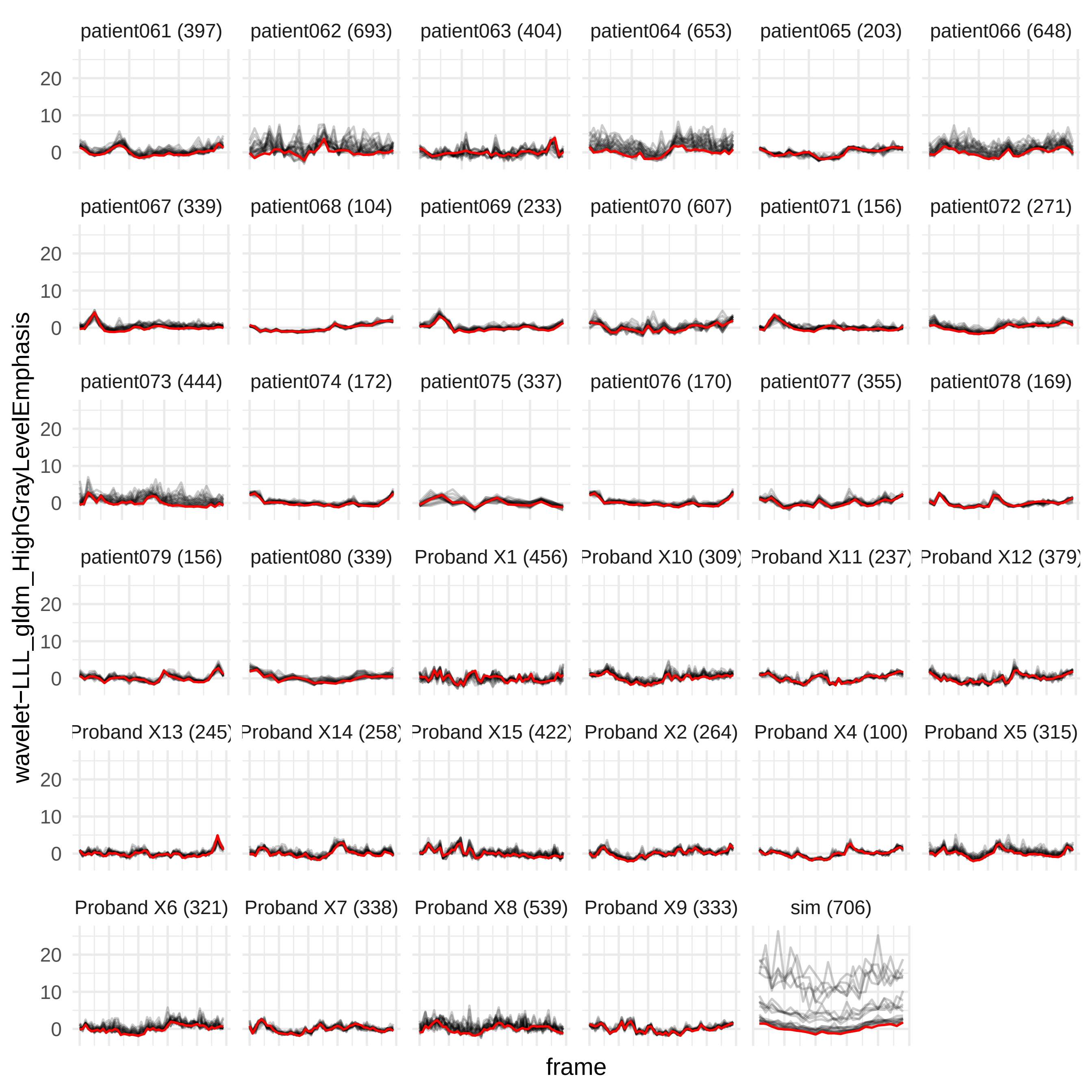

patient061 (397)
patient062 (693)
patient063 (404)
patient064 (653)
patient065 (203)
patient066 (648)
patient067 (339)
patient068 (104)
patient069 (233)
patient070 (607)
patient071 (156)
patient072 (271)
patient073 (444)
patient074 (172)
patient075 (337)
patient076 (170)
patient077 (355)
patient078 (169)
patient079 (156)
patient080 (339)
Proband X1 (456)
Proband X10 (309)
Proband X11 (237)
Proband X12 (379)
Proband X13 (245)
Proband X14 (258)
Proband X15 (422)
Proband X2 (264)
Proband X4 (100)
Proband X5 (315)
Proband X6 (321)
Proband X7 (338)
Proband X8 (539)
Proband X9 (333)
sim (706)
0
10
20
wavelet−LLL_gldm_HighGrayLevelEmphasis
frame

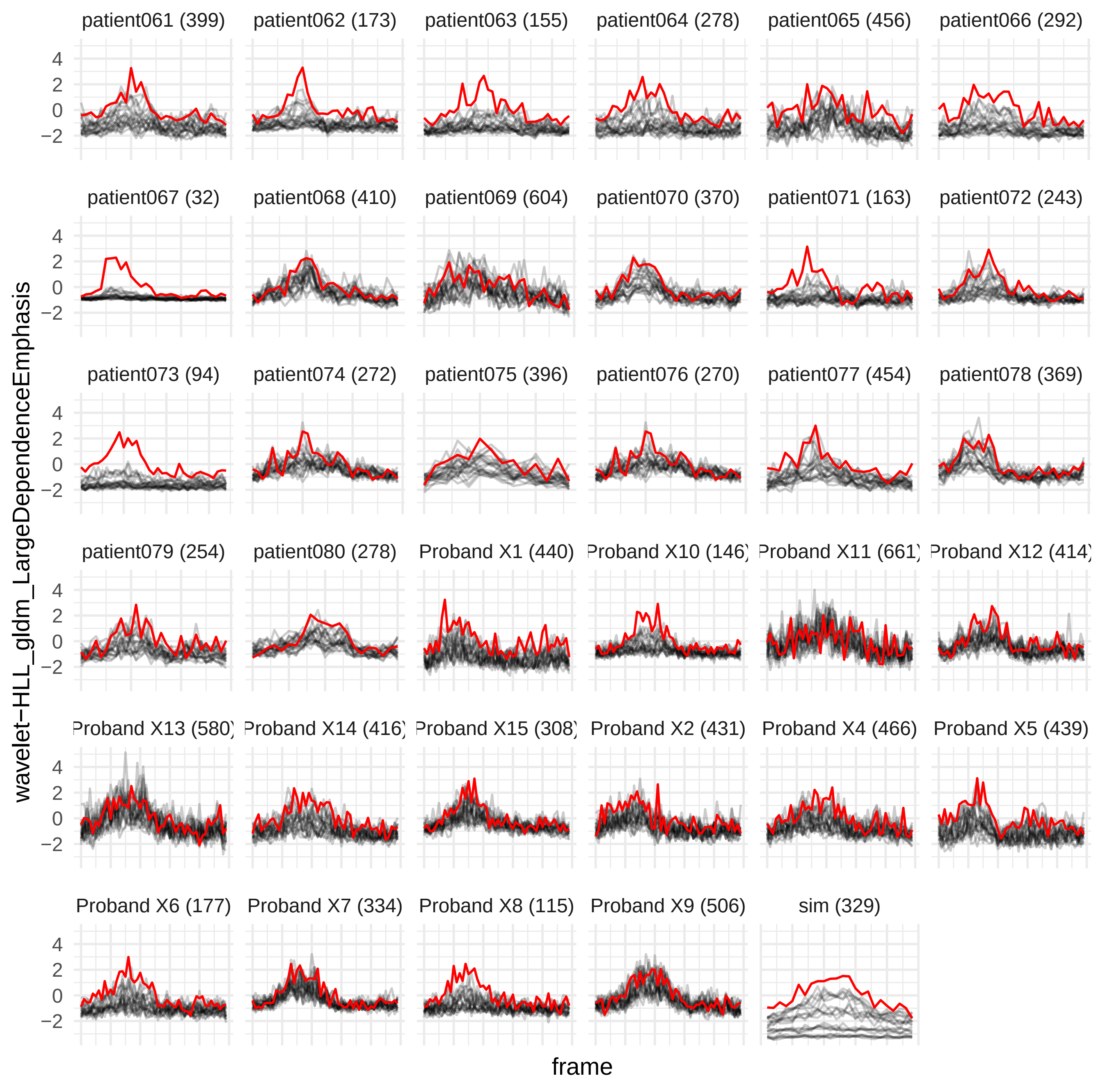

patient061 (399)
patient062 (173)
patient063 (155)
patient064 (278)
patient065 (456)
patient066 (292)
patient067 (32)
patient068 (410)
patient069 (604)
patient070 (370)
patient071 (163)
patient072 (243)
patient073 (94)
patient074 (272)
patient075 (396)
patient076 (270)
patient077 (454)
patient078 (369)
patient079 (254)
patient080 (278)
Proband X1 (440)
Proband X10 (146)
Proband X11 (661)
Proband X12 (414)
Proband X13 (580)
Proband X14 (416)
Proband X15 (308)
Proband X2 (431)
Proband X4 (466)
Proband X5 (439)
Proband X6 (177)
Proband X7 (334)
Proband X8 (115)
Proband X9 (506)
sim (329)
4
2
0
−2
wavelet−HLL_gldm_LargeDependenceEmphasis
frame

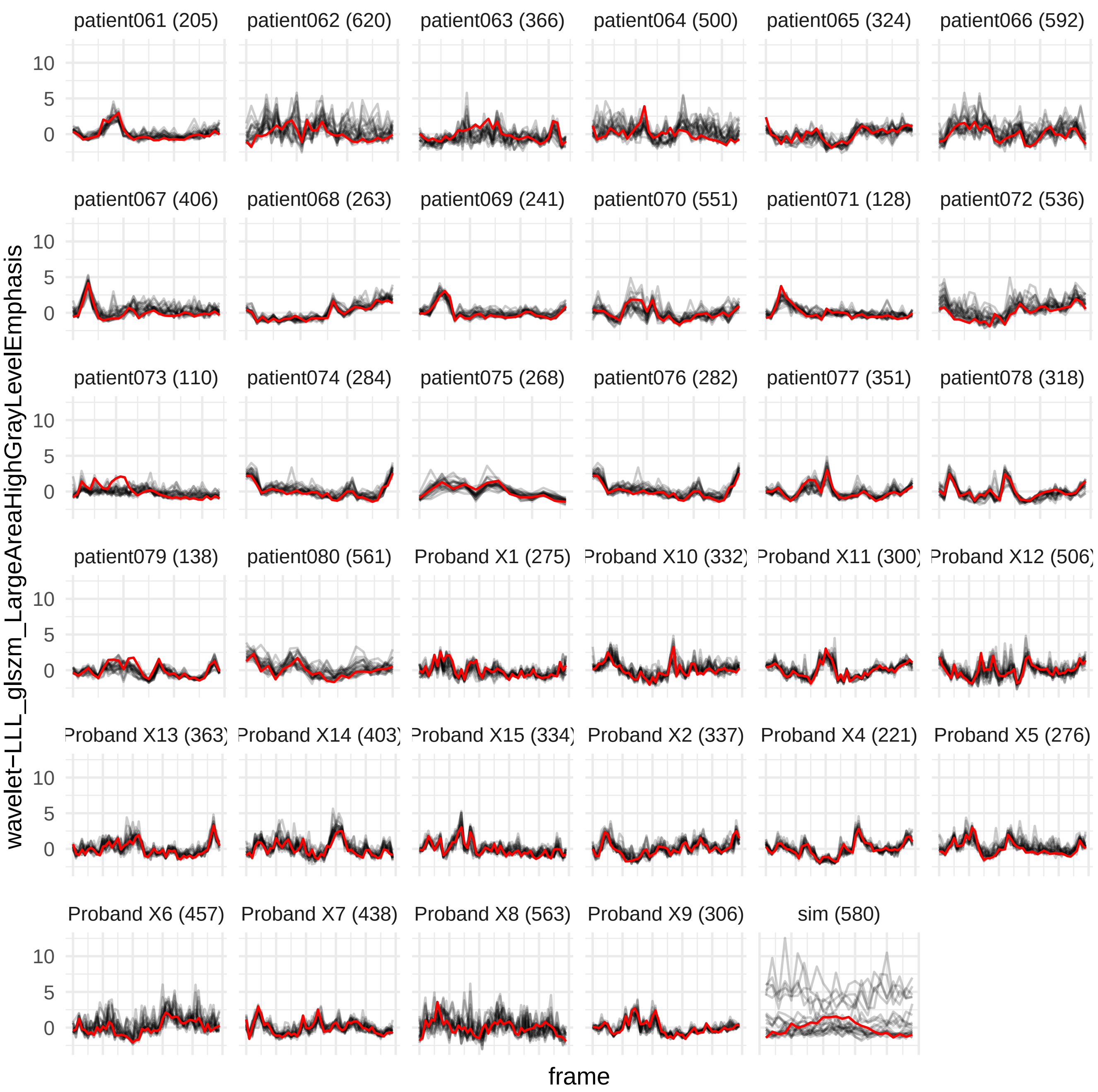

patient061 (205)
patient062 (620)
patient063 (366)
patient064 (500)
patient065 (324)
patient066 (592)
patient067 (406)
patient068 (263)
patient069 (241)
patient070 (551)
patient071 (128)
patient072 (536)
patient073 (110)
patient074 (284)
patient075 (268)
patient076 (282)
patient077 (351)
patient078 (318)
patient079 (138)
patient080 (561)
Proband X1 (275)
Proband X10 (332)
Proband X11 (300)
Proband X12 (506)
Proband X13 (363)
Proband X14 (403)
Proband X15 (334)
Proband X2 (337)
Proband X4 (221)
Proband X5 (276)
Proband X6 (457)
Proband X7 (438)
Proband X8 (563)
Proband X9 (306)
sim (580)
10
5
0
wavelet−LLL_glszm_LargeAreaHighGrayLevelEmphasis
frame

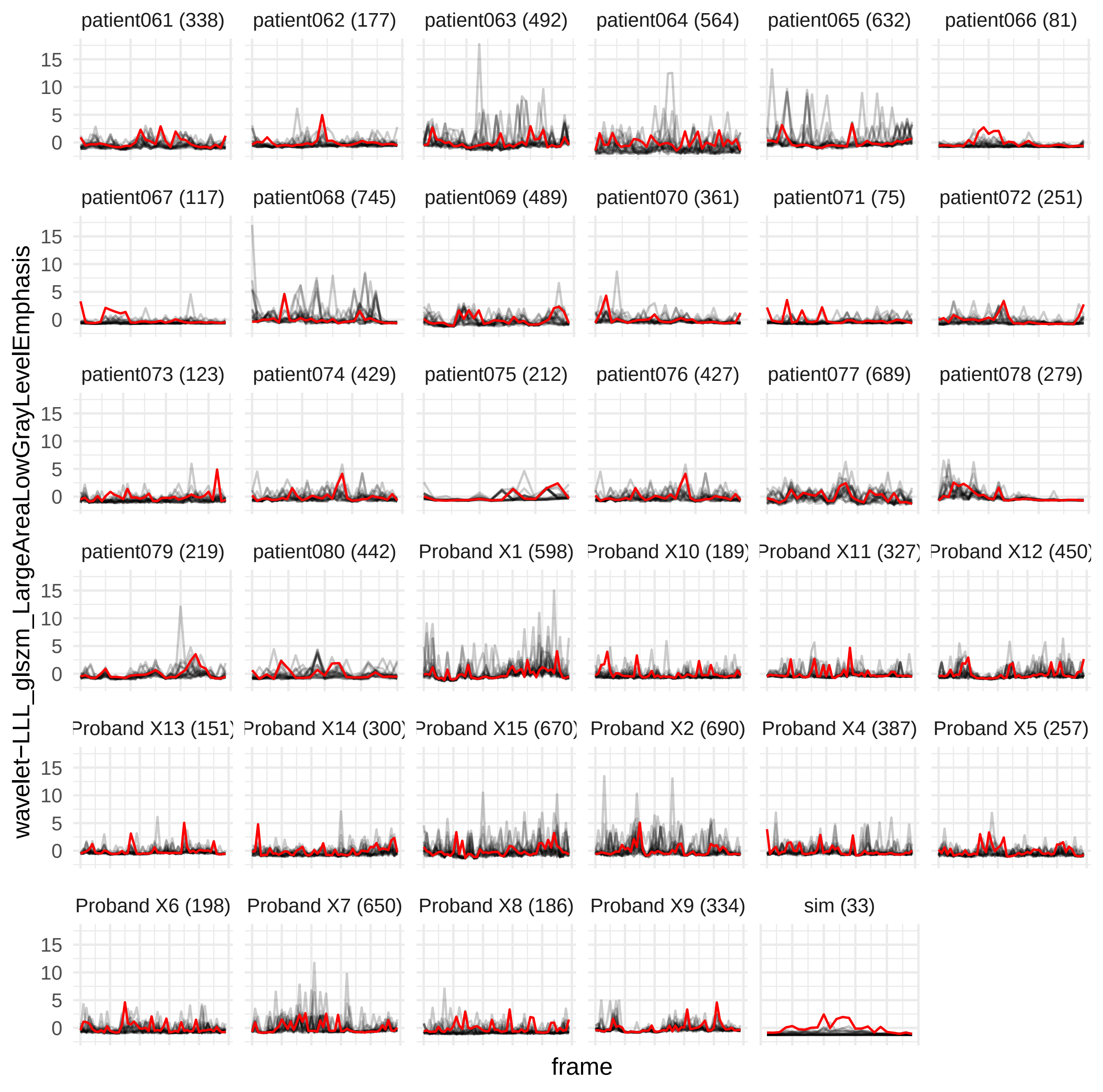

patient061 (338)
patient062 (177)
patient063 (492)
patient064 (564)
patient065 (632)
patient066 (81)
patient067 (117)
patient068 (745)
patient069 (489)
patient070 (361)
patient071 (75)
patient072 (251)
patient073 (123)
patient074 (429)
patient075 (212)
patient076 (427)
patient077 (689)
patient078 (279)
patient079 (219)
patient080 (442)
Proband X1 (598)
Proband X10 (189)
Proband X11 (327)
Proband X12 (450)
Proband X13 (151)
Proband X14 (300)
Proband X15 (670)
Proband X2 (690)
Proband X4 (387)
Proband X5 (257)
Proband X6 (198)
Proband X7 (650)
Proband X8 (186)
Proband X9 (334)
sim (33)
15
10
5
0
wavelet−LLL_glszm_LargeAreaLowGrayLevelEmphasis
frame

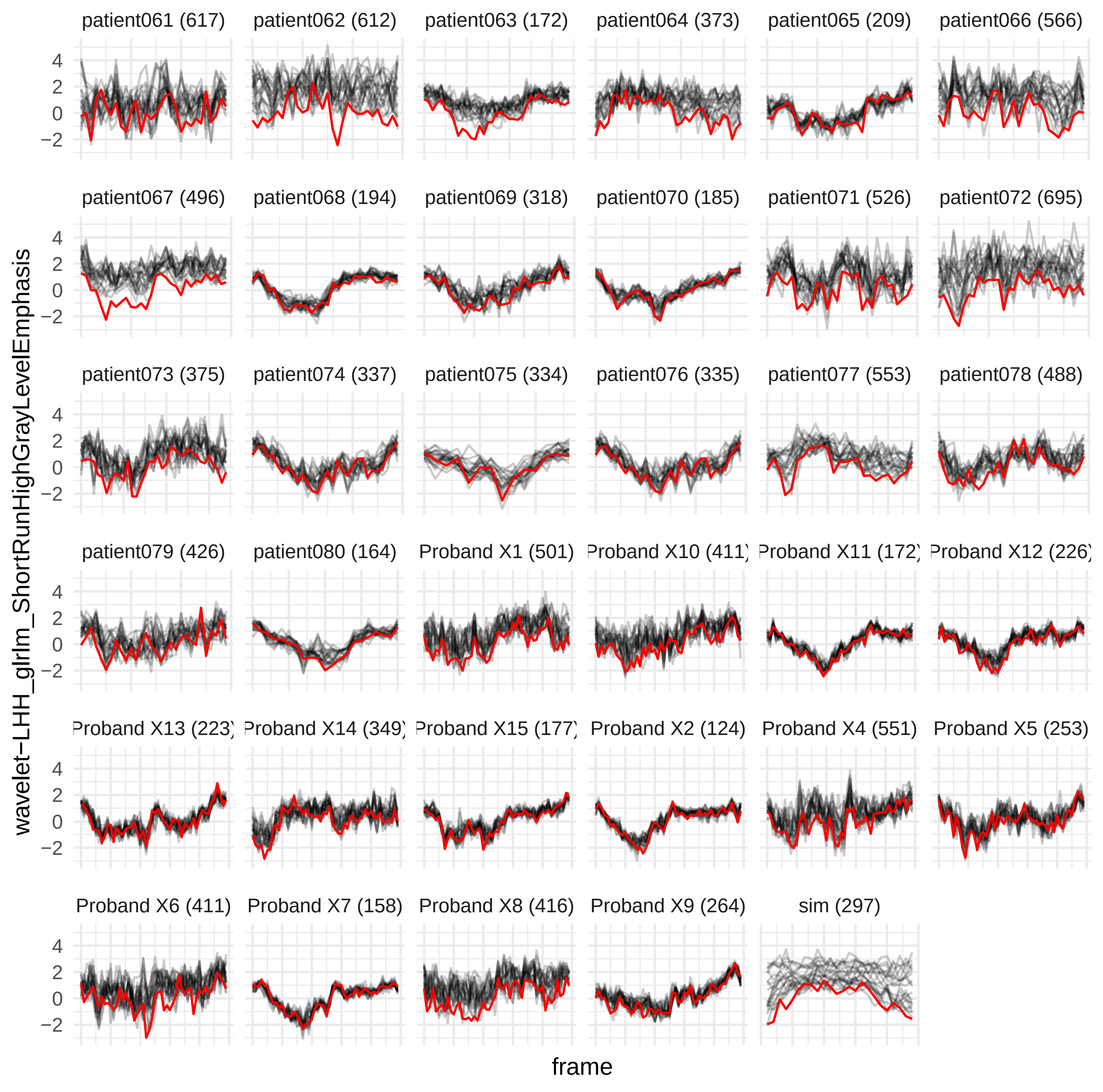
patient061 (617)
patient062 (612)
patient063 (172)
patient064 (373)
patient065 (209)
patient066 (566)
patient067 (496)
patient068 (194)
patient069 (318)
patient070 (185)
patient071 (526)
patient072 (695)
patient073 (375)
patient074 (337)
patient075 (334)
patient076 (335)
patient077 (553)
patient078 (488)
patient079 (426)
patient080 (164)
Proband X1 (501)
Proband X10 (411)
Proband X11 (172)
Proband X12 (226)
Proband X13 (223)
Proband X14 (349)
Proband X15 (177)
Proband X2 (124)
Proband X4 (551)
Proband X5 (253)
Proband X6 (411)
Proband X7 (158)
Proband X8 (416)
Proband X9 (264)
sim (297)
4
2
0
−2
wavelet−LHH_glrlm_ShortRunHighGrayLevelEmphasis
frame

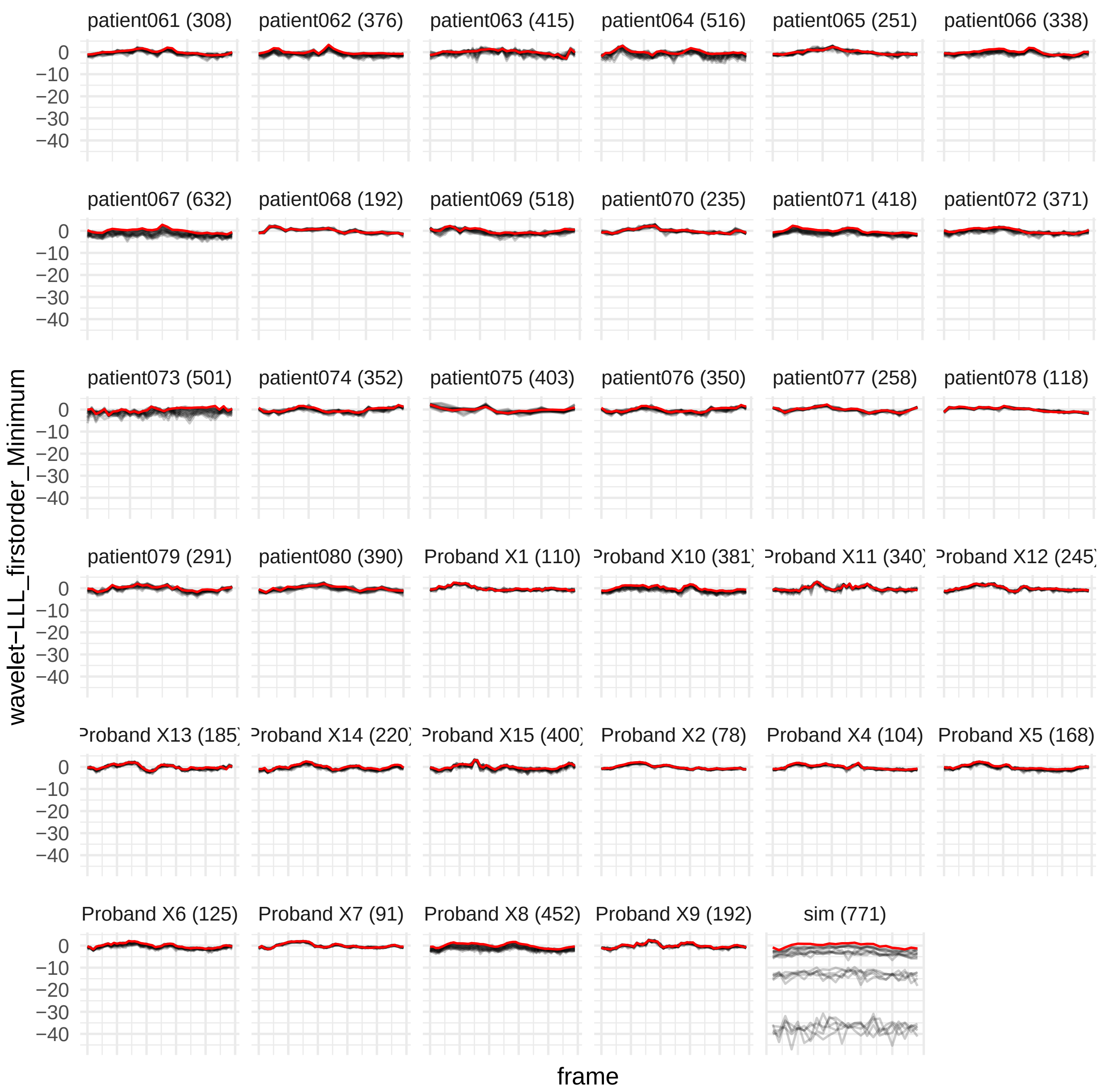

patient061 (308)
patient062 (376)
patient063 (415)
patient064 (516)
patient065 (251)
patient066 (338)
patient067 (632)
patient068 (192)
patient069 (518)
patient070 (235)
patient071 (418)
patient072 (371)
patient073 (501)
patient074 (352)
patient075 (403)
patient076 (350)
patient077 (258)
patient078 (118)
patient079 (291)
patient080 (390)
Proband X1 (110)
roband X10 (381
roband X11 (340
roband X12 (245
roband X13 (185
roband X14 (220
roband X15 (400
Proband X2 (78)
Proband X4 (104)
Proband X5 (168)
Proband X6 (125)
Proband X7 (91)
Proband X8 (452)
Proband X9 (192)
sim (771)
0
−10
−20
−30
−40
wavelet−LLL_firstorder_Minimum
frame

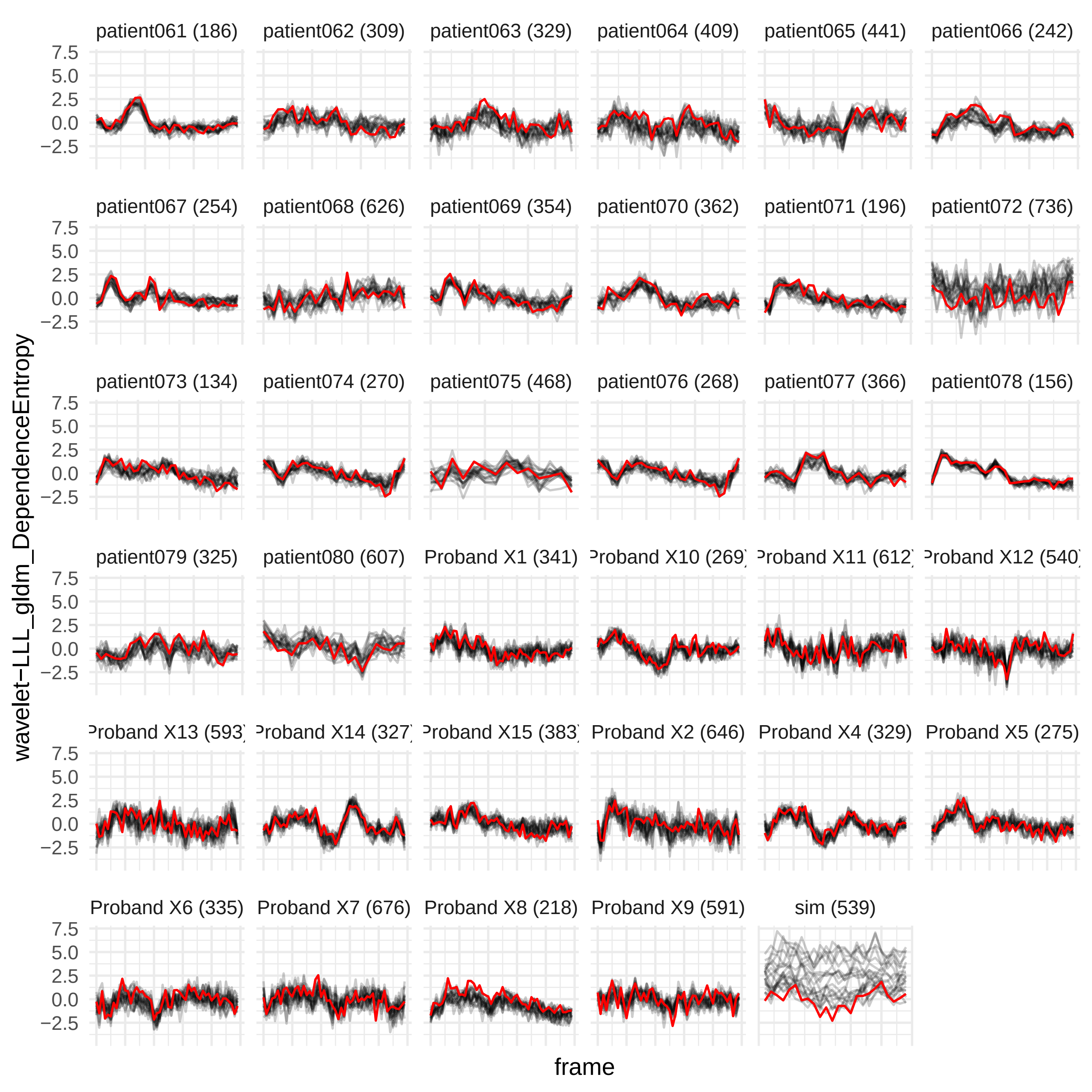

patient061 (186)
patient062 (309)
patient063 (329)
patient064 (409)
patient065 (441)
patient066 (242)
patient067 (254)
patient068 (626)
patient069 (354)
patient070 (362)
patient071 (196)
patient072 (736)
patient073 (134)
patient074 (270)
patient075 (468)
patient076 (268)
patient077 (366)
patient078 (156)
patient079 (325)
patient080 (607)
Proband X1 (341)
Proband X10 (269)
Proband X11 (612)
Proband X12 (540)
Proband X13 (593)
Proband X14 (327)
Proband X15 (383)
Proband X2 (646)
Proband X4 (329)
Proband X5 (275)
Proband X6 (335)
Proband X7 (676)
Proband X8 (218)
Proband X9 (591)
sim (539)
7.5
5.0
2.5
0.0
−2.5
wavelet−LLL_gldm_DependenceEntropy
frame

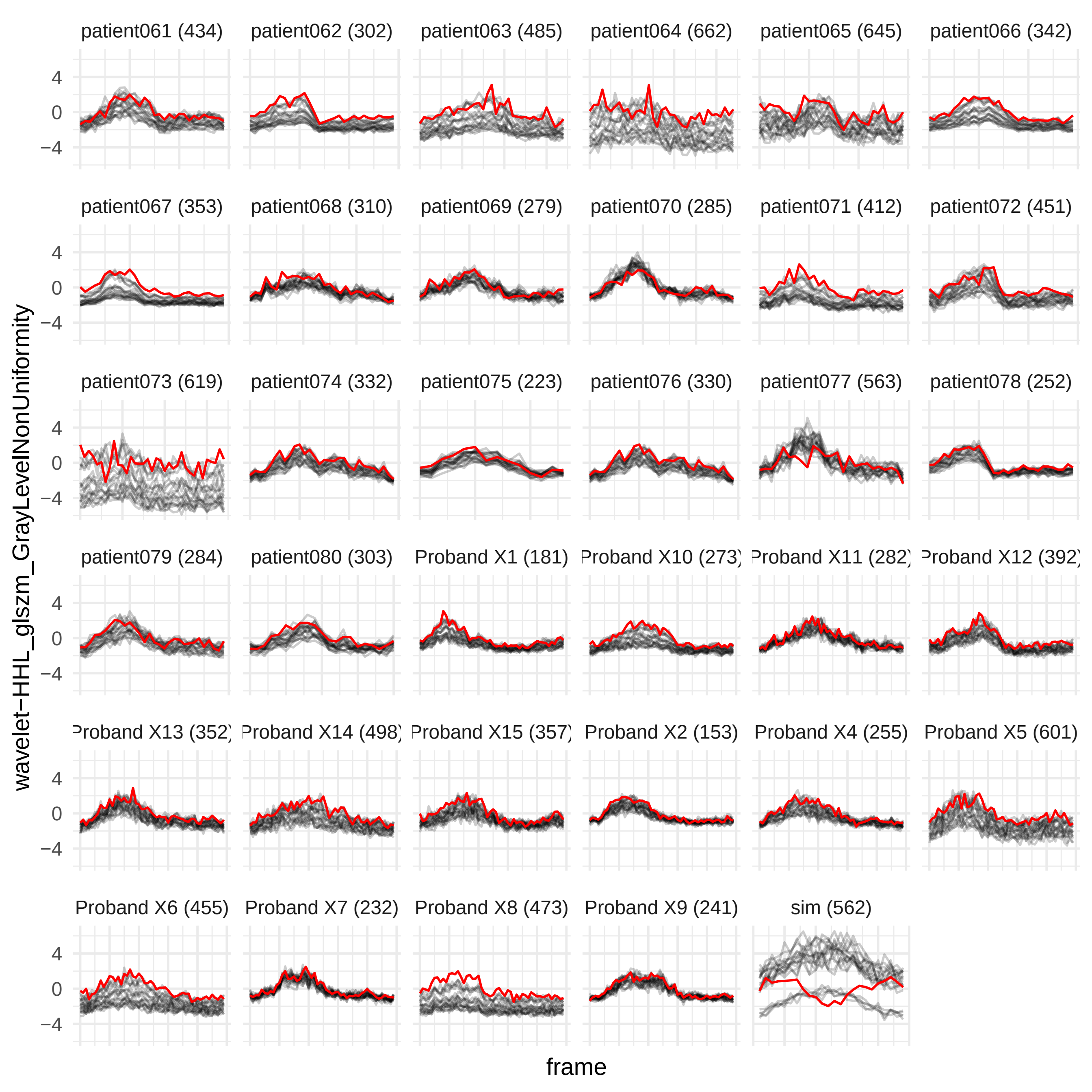
patient061 (434)
patient062 (302)
patient063 (485)
patient064 (662)
patient065 (645)
patient066 (342)
patient067 (353)
patient068 (310)
patient069 (279)
patient070 (285)
patient071 (412)
patient072 (451)
patient073 (619)
patient074 (332)
patient075 (223)
patient076 (330)
patient077 (563)
patient078 (252)
patient079 (284)
patient080 (303)
Proband X1 (181)
Proband X10 (273)
Proband X11 (282)
Proband X12 (392)
Proband X13 (352)
Proband X14 (498)
Proband X15 (357)
Proband X2 (153)
Proband X4 (255)
Proband X5 (601)
Proband X6 (455)
Proband X7 (232)
Proband X8 (473)
Proband X9 (241)
sim (562)
4
0
−4
wavelet−HHL_glszm_GrayLevelNonUniformity
frame

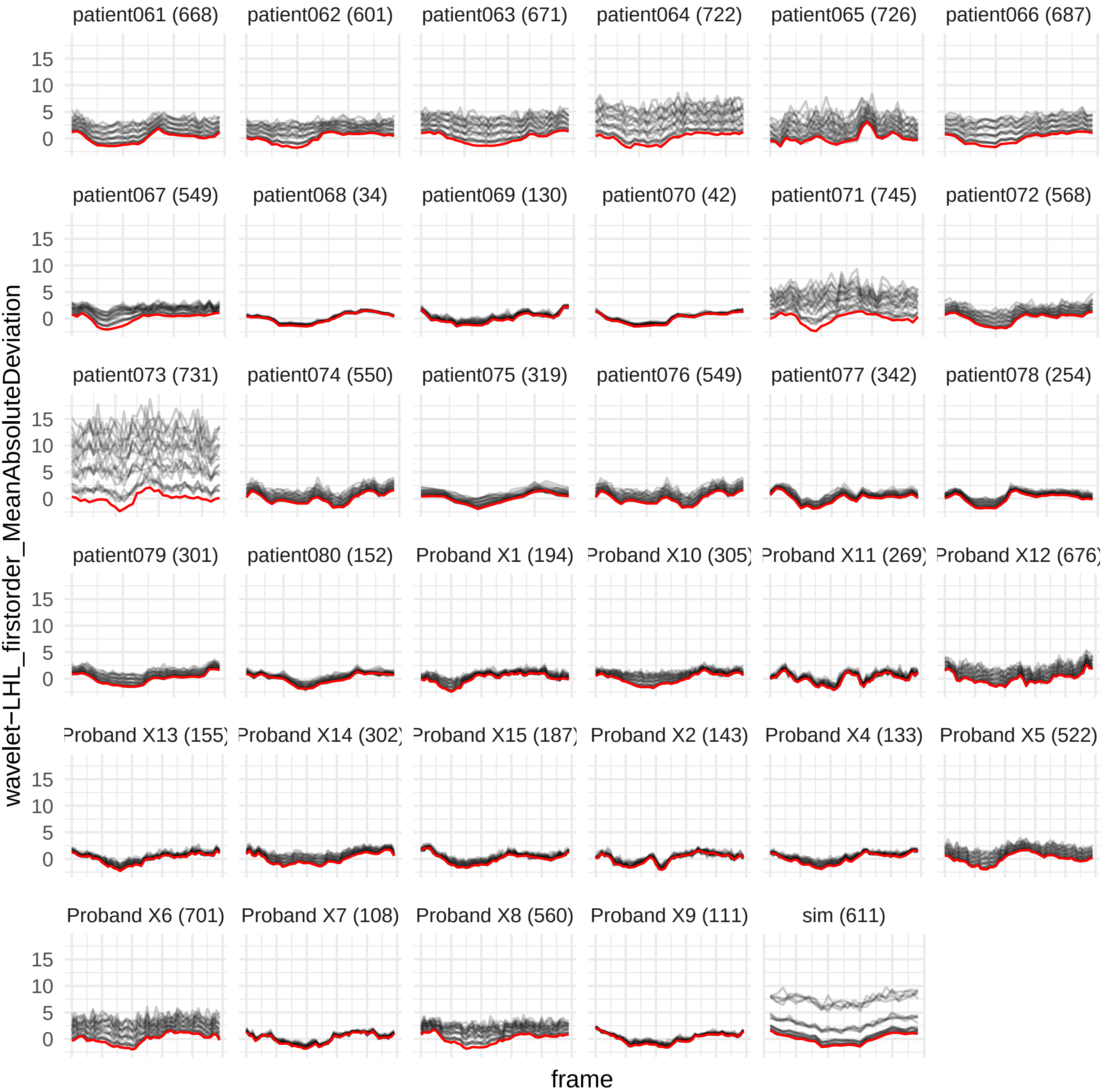

patient061 (668)
patient062 (601)
patient063 (671)
patient064 (722)
patient065 (726)
patient066 (687)
patient067 (549)
patient068 (34)
patient069 (130)
patient070 (42)
patient071 (745)
patient072 (568)
patient073 (731)
patient074 (550)
patient075 (319)
patient076 (549)
patient077 (342)
patient078 (254)
patient079 (301)
patient080 (152)
Proband X1 (194)
Proband X10 (305)
Proband X11 (269)
Proband X12 (676)
Proband X13 (155)
Proband X14 (302)
Proband X15 (187)
Proband X2 (143)
Proband X4 (133)
Proband X5 (522)
Proband X6 (701)
Proband X7 (108)
Proband X8 (560)
Proband X9 (111)
sim (611)
wavelet−LHL_firstorder_MeanAbsoluteDeviation
0
5
10
15
frame

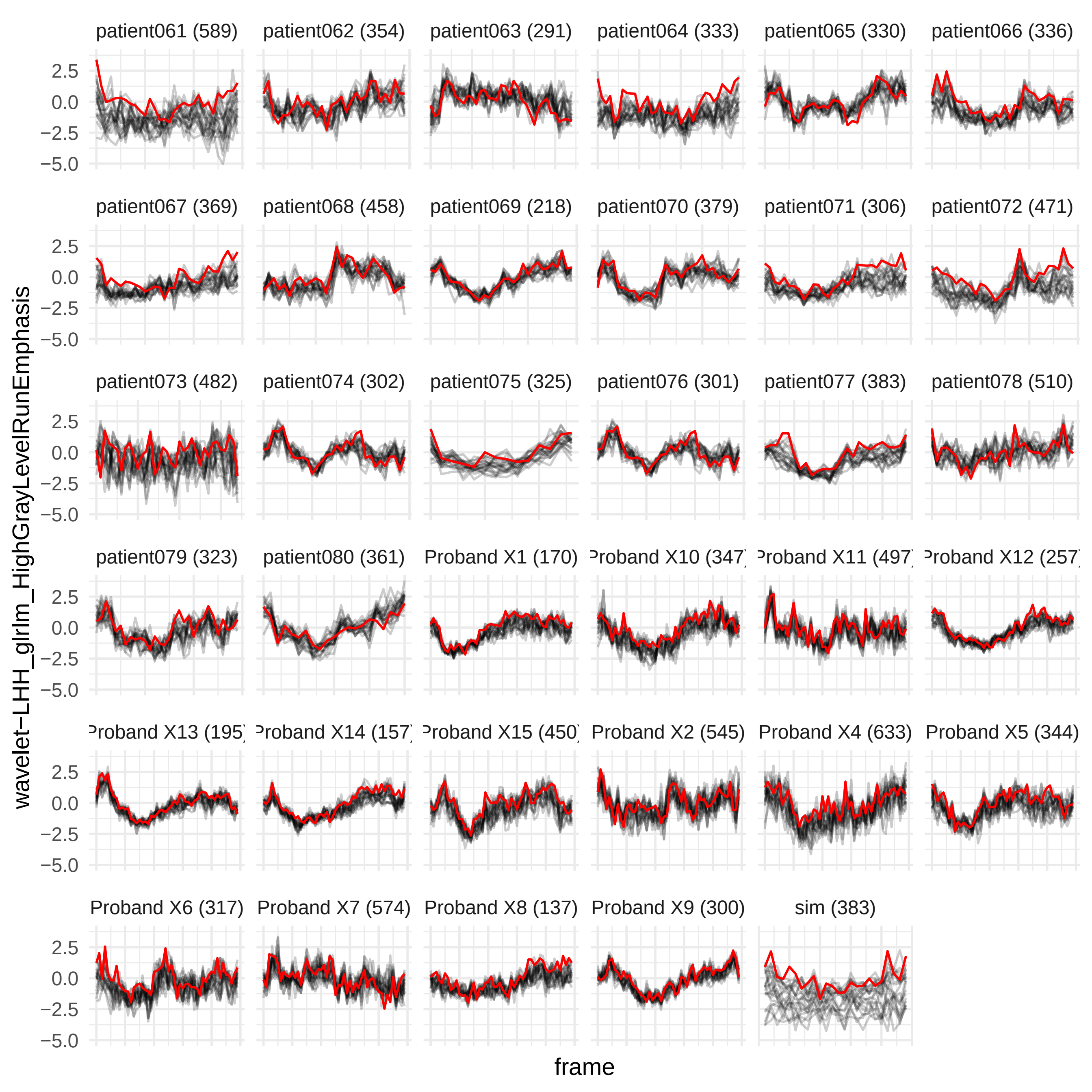

patient061 (589)
patient062 (354)
patient063 (291)
patient064 (333)
patient065 (330)
patient066 (336)
patient067 (369)
patient068 (458)
patient069 (218)
patient070 (379)
patient071 (306)
patient072 (471)
patient073 (482)
patient074 (302)
patient075 (325)
patient076 (301)
patient077 (383)
patient078 (510)
patient079 (323)
patient080 (361)
Proband X1 (170)
Proband X10 (347)
Proband X11 (497)
Proband X12 (257)
Proband X13 (195)
Proband X14 (157)
Proband X15 (450)
Proband X2 (545)
Proband X4 (633)
Proband X5 (344)
Proband X6 (317)
Proband X7 (574)
Proband X8 (137)
Proband X9 (300)
sim (383)
2.5
0.0
−2.5
−5.0
wavelet−LHH_glrlm_HighGrayLevelRunEmphasis
frame

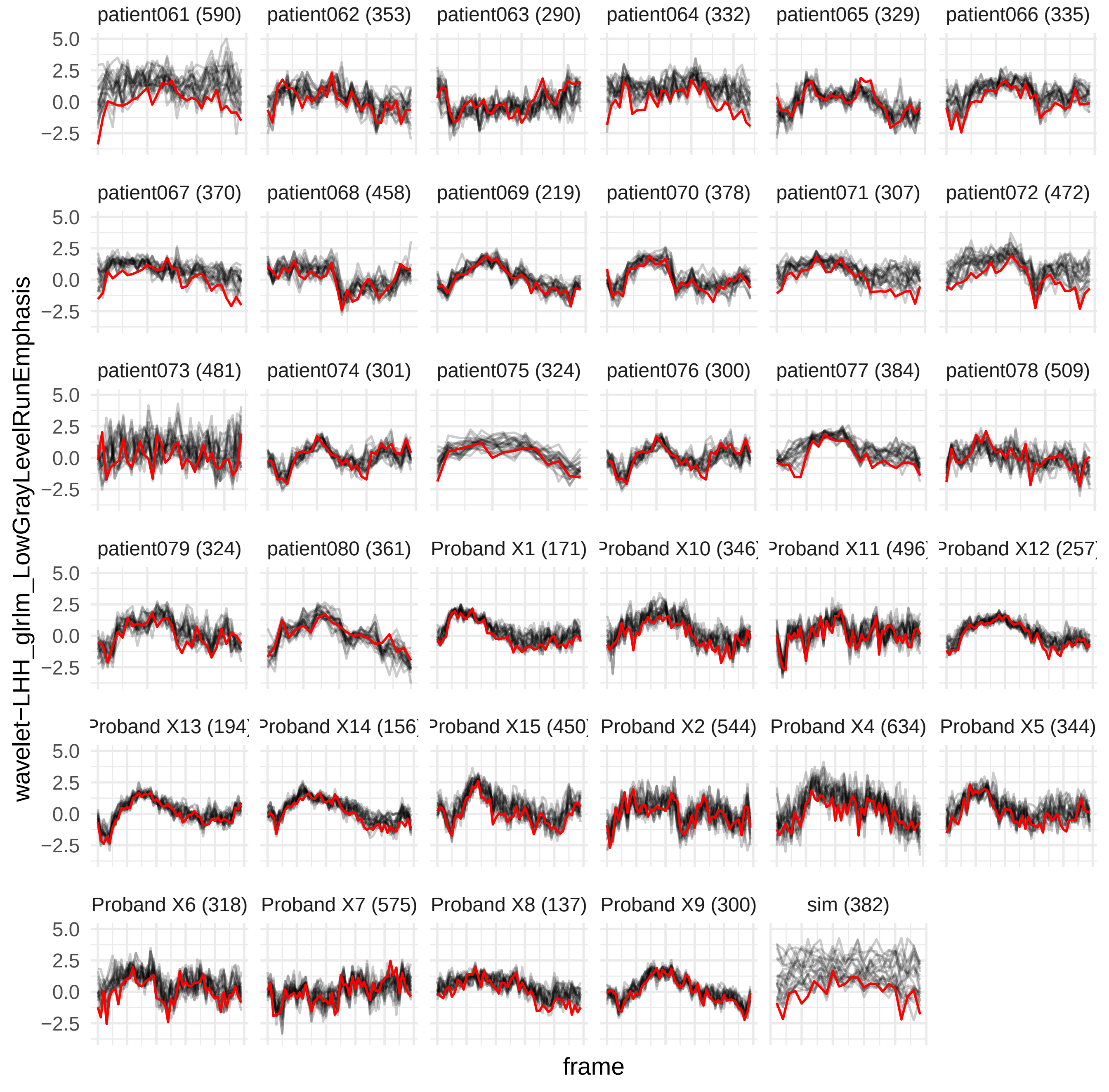

patient061 (590)
patient062 (353)
patient063 (290)
patient064 (332)
patient065 (329)
patient066 (335)
patient067 (370)
patient068 (458)
patient069 (219)
patient070 (378)
patient071 (307)
patient072 (472)
patient073 (481)
patient074 (301)
patient075 (324)
patient076 (300)
patient077 (384)
patient078 (509)
patient079 (324)
patient080 (361)
Proband X1 (171)
Proband X2 (544)
Proband X4 (634)
Proband X5 (344)
Proband X6 (318)
Proband X7 (575)
Proband X8 (137)
Proband X9 (300)
sim (382)
5.0
2.5
0.0
−2.5
wavelet−LHH_glrlm_LowGrayLevelRunEmphasis
frame

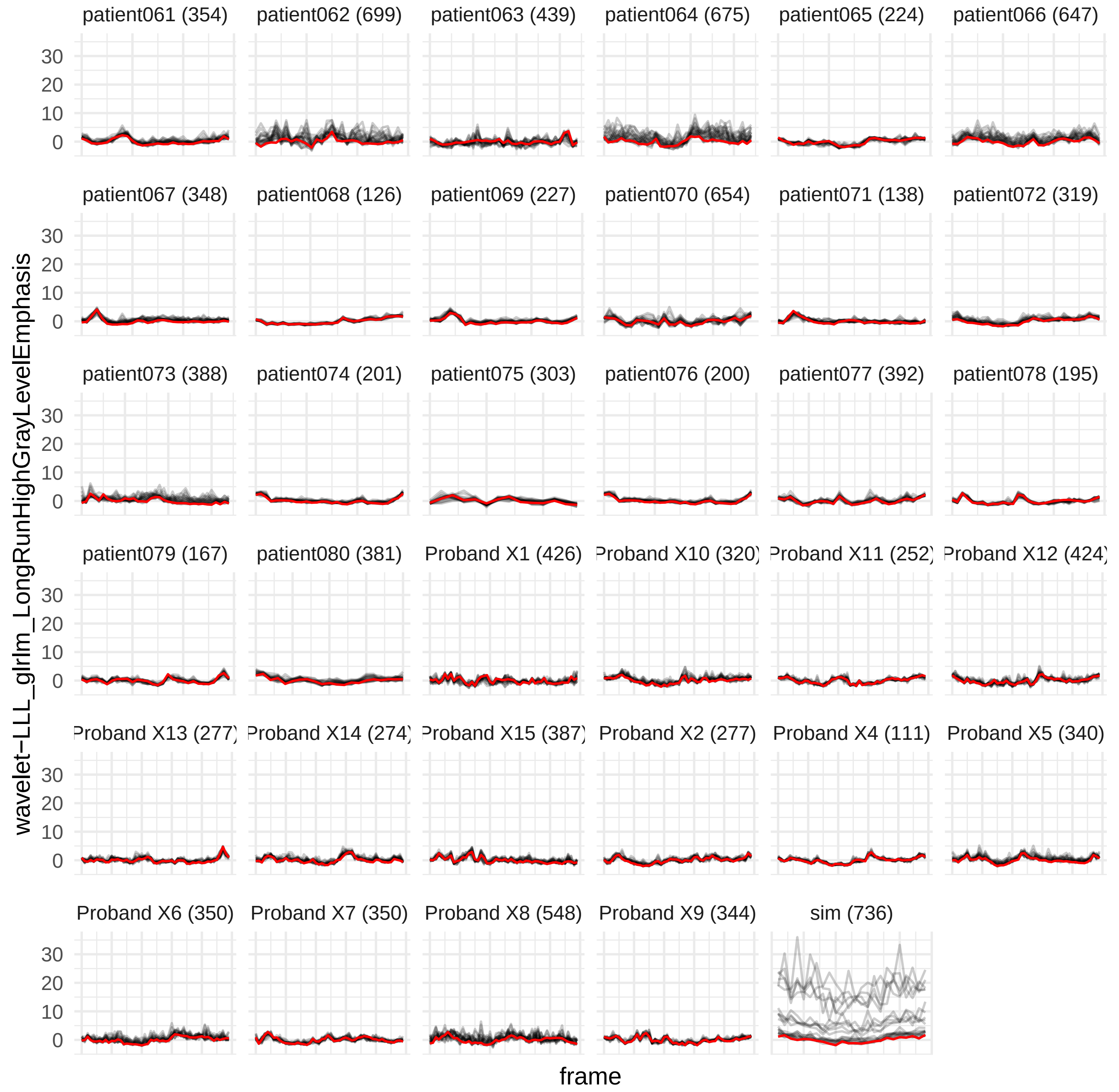

patient061 (354)
patient062 (699)
patient063 (439)
patient064 (675)
patient065 (224)
patient066 (647)
patient067 (348)
patient068 (126)
patient069 (227)
patient070 (654)
patient071 (138)
patient072 (319)
patient073 (388)
patient074 (201)
patient075 (303)
patient076 (200)
patient077 (392)
patient078 (195)
patient079 (167)
patient080 (381)
Proband X1 (426)
Proband X10 (320)
Proband X11 (252)
Proband X12 (424)
Proband X13 (277)
Proband X14 (274)
Proband X15 (387)
Proband X2 (277)
Proband X4 (111)
Proband X5 (340)
Proband X6 (350)
Proband X7 (350)
Proband X8 (548)
Proband X9 (344)
sim (736)
30
20
10
0
wavelet−LLL_glrlm_LongRunHighGrayLevelEmphasis
frame

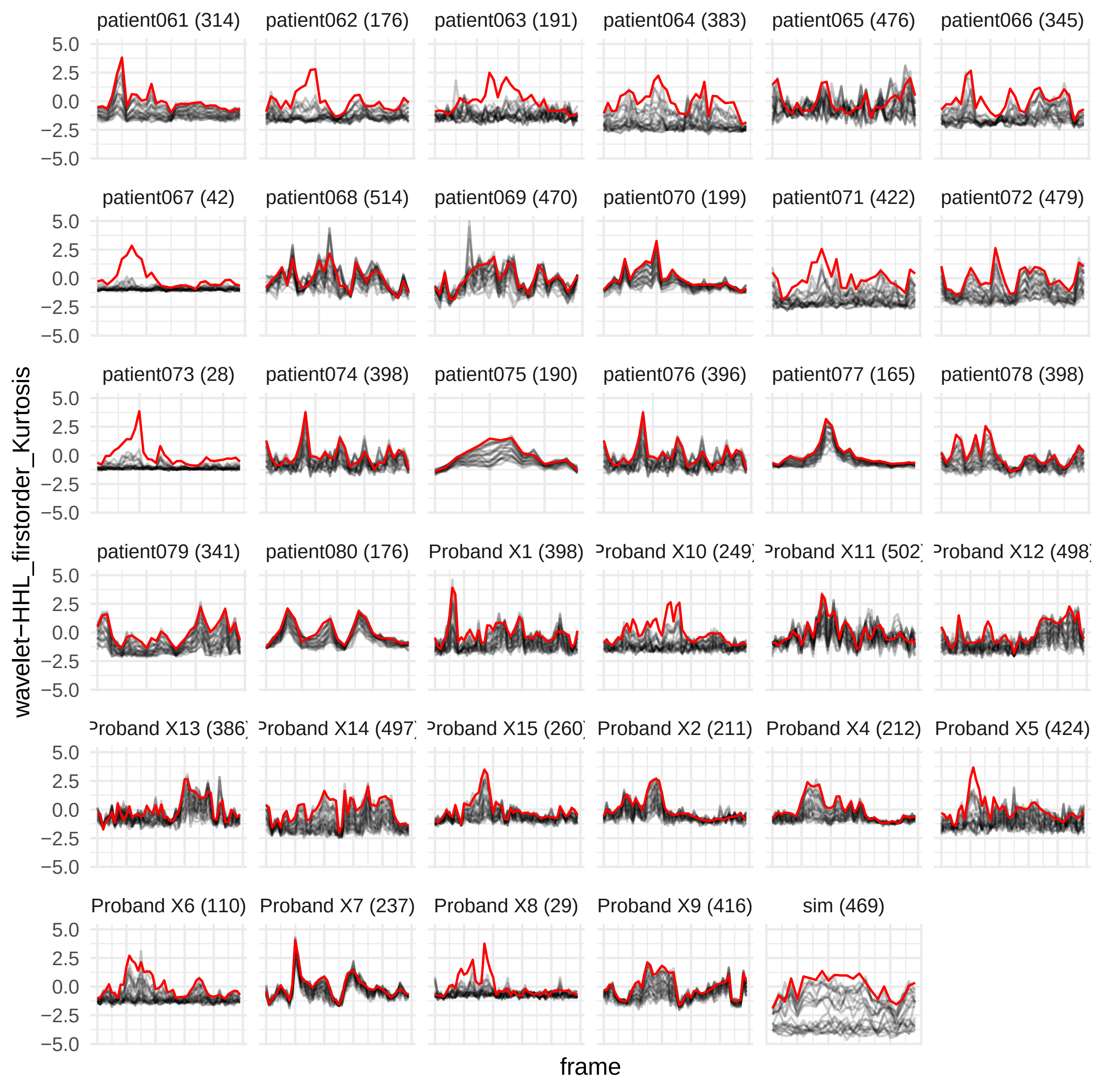
patient061 (314)
patient062 (176)
patient063 (191)
patient064 (383)
patient065 (476)
patient066 (345)
patient067 (42)
patient068 (514)
patient069 (470)
patient070 (199)
patient071 (422)
patient072 (479)
patient073 (28)
patient074 (398)
patient075 (190)
patient076 (396)
patient077 (165)
patient078 (398)
patient079 (341)
patient080 (176)
Proband X1 (398)
Proband X10 (249)
Proband X11 (502)
Proband X12 (498)
Proband X13 (386)
Proband X14 (497)
Proband X15 (260)
Proband X2 (211)
Proband X4 (212)
Proband X5 (424)
Proband X6 (110)
Proband X7 (237)
Proband X8 (29)
Proband X9 (416)
sim (469)
5.0
2.5
0.0
−2.5
−5.0
wavelet−HHL_firstorder_Kurtosis
frame

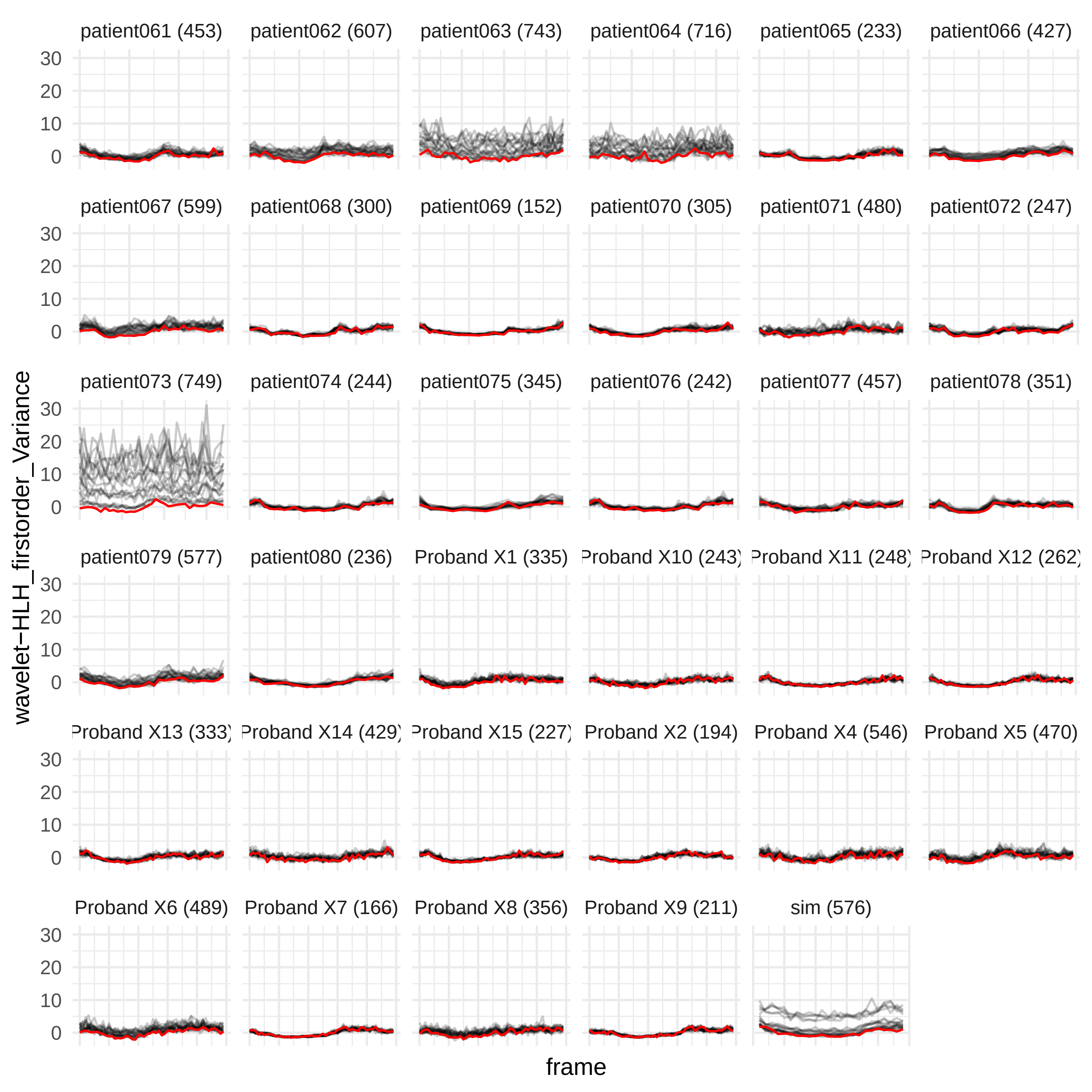

patient061 (453)
patient062 (607)
patient063 (743)
patient064 (716)
patient065 (233)
patient066 (427)
patient067 (599)
patient068 (300)
patient069 (152)
patient070 (305)
patient071 (480)
patient072 (247)
patient073 (749)
patient074 (244)
patient075 (345)
patient076 (242)
patient077 (457)
patient078 (351)
patient079 (577)
patient080 (236)
Proband X1 (335)
Proband X10 (243)
Proband X11 (248)
Proband X12 (262)
Proband X13 (333)
Proband X14 (429)
Proband X15 (227)
Proband X2 (194)
Proband X4 (546)
Proband X5 (470)
Proband X6 (489)
Proband X7 (166)
Proband X8 (356)
Proband X9 (211)
sim (576)
wavelet−HLH_firstorder_Variance
0
10
20
30
frame

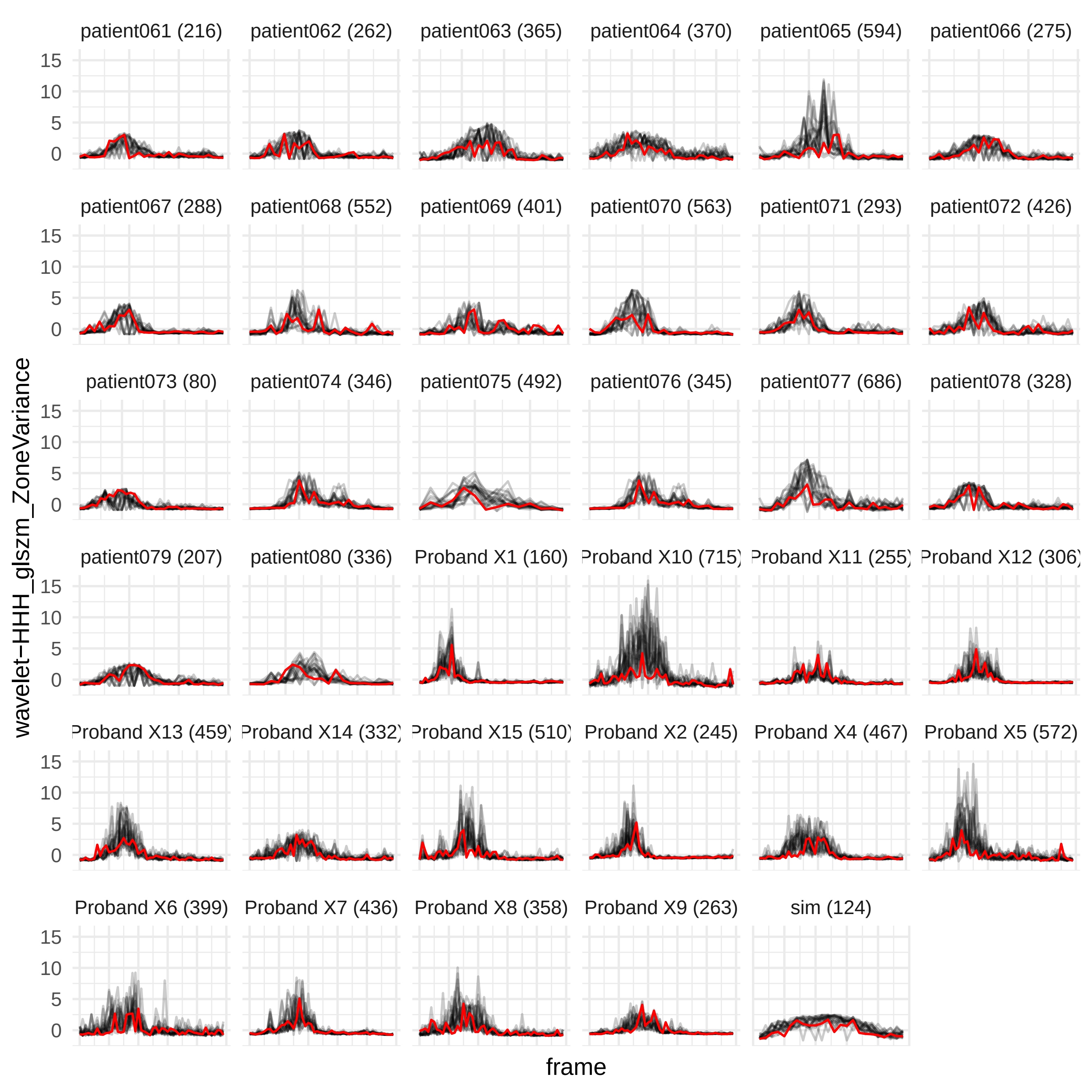
patient061 (216)
patient062 (262)
patient063 (365)
patient064 (370)
patient065 (594)
patient066 (275)
patient067 (288)
patient068 (552)
patient069 (401)
patient070 (563)
patient071 (293)
patient072 (426)
patient073 (80)
patient074 (346)
patient075 (492)
patient076 (345)
patient077 (686)
patient078 (328)
patient079 (207)
patient080 (336)
Proband X1 (160)
Proband X10 (715)
Proband X11 (255)
Proband X12 (306)
Proband X13 (459)
Proband X14 (332)
Proband X15 (510)
Proband X2 (245)
Proband X4 (467)
Proband X5 (572)
Proband X6 (399)
Proband X7 (436)
Proband X8 (358)
Proband X9 (263)
sim (124)
wavelet−HHH_glszm_ZoneVariance
frame
15
10
5
0

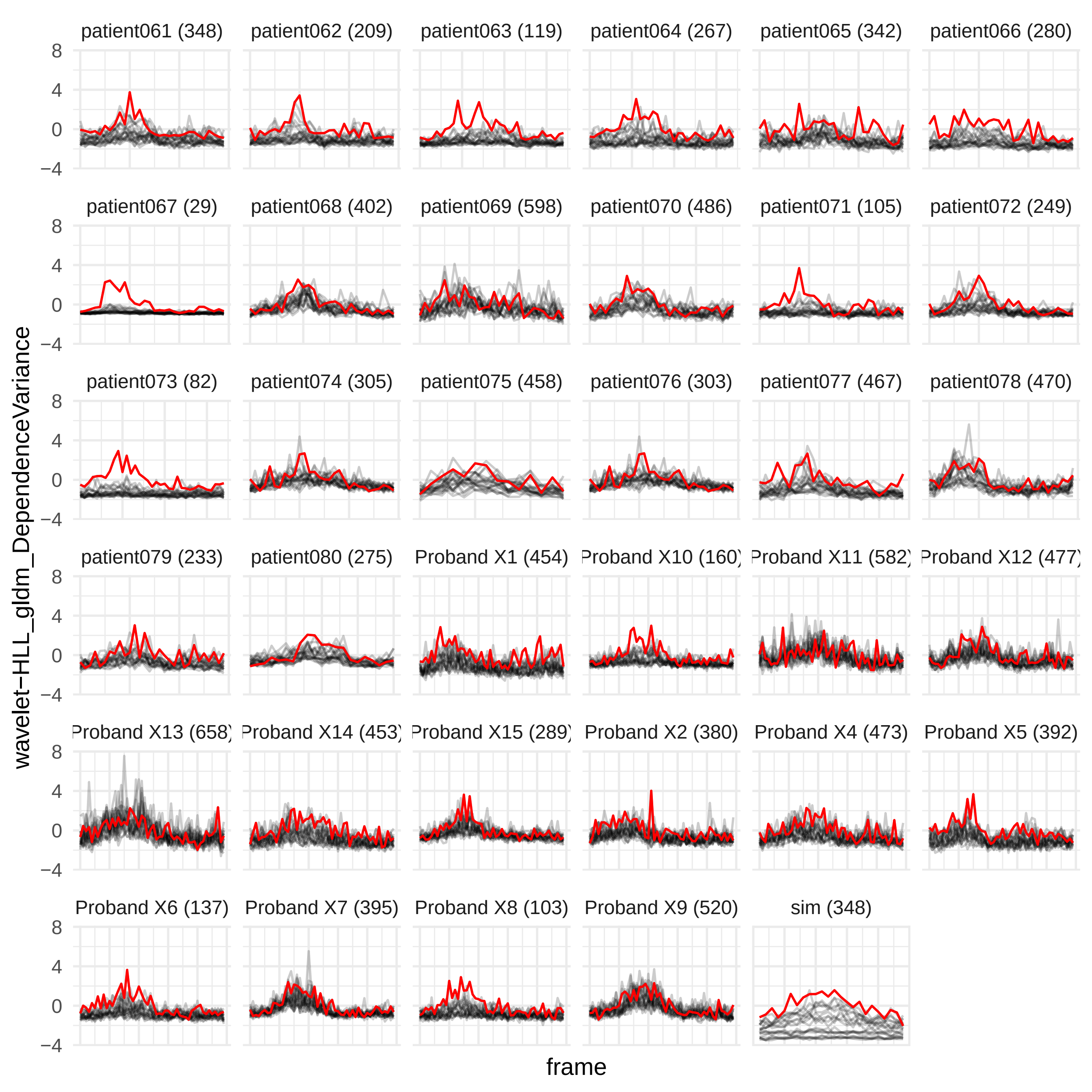

patient061 (348)
patient062 (209)
patient063 (119)
patient064 (267)
patient065 (342)
patient066 (280)
patient067 (29)
patient068 (402)
patient069 (598)
patient070 (486)
patient071 (105)
patient072 (249)
patient073 (82)
patient074 (305)
patient075 (458)
patient076 (303)
patient077 (467)
patient078 (470)
patient079 (233)
patient080 (275)
Proband X1 (454)
Proband X10 (160)
Proband X11 (582)
Proband X12 (477)
Proband X13 (658)
Proband X14 (453)
Proband X15 (289)
Proband X2 (380)
Proband X4 (473)
Proband X5 (392)
Proband X6 (137)
Proband X7 (395)
Proband X8 (103)
Proband X9 (520)
sim (348)
wavelet−HLL_gldm_DependenceVariance
frame
8
4
0
−4

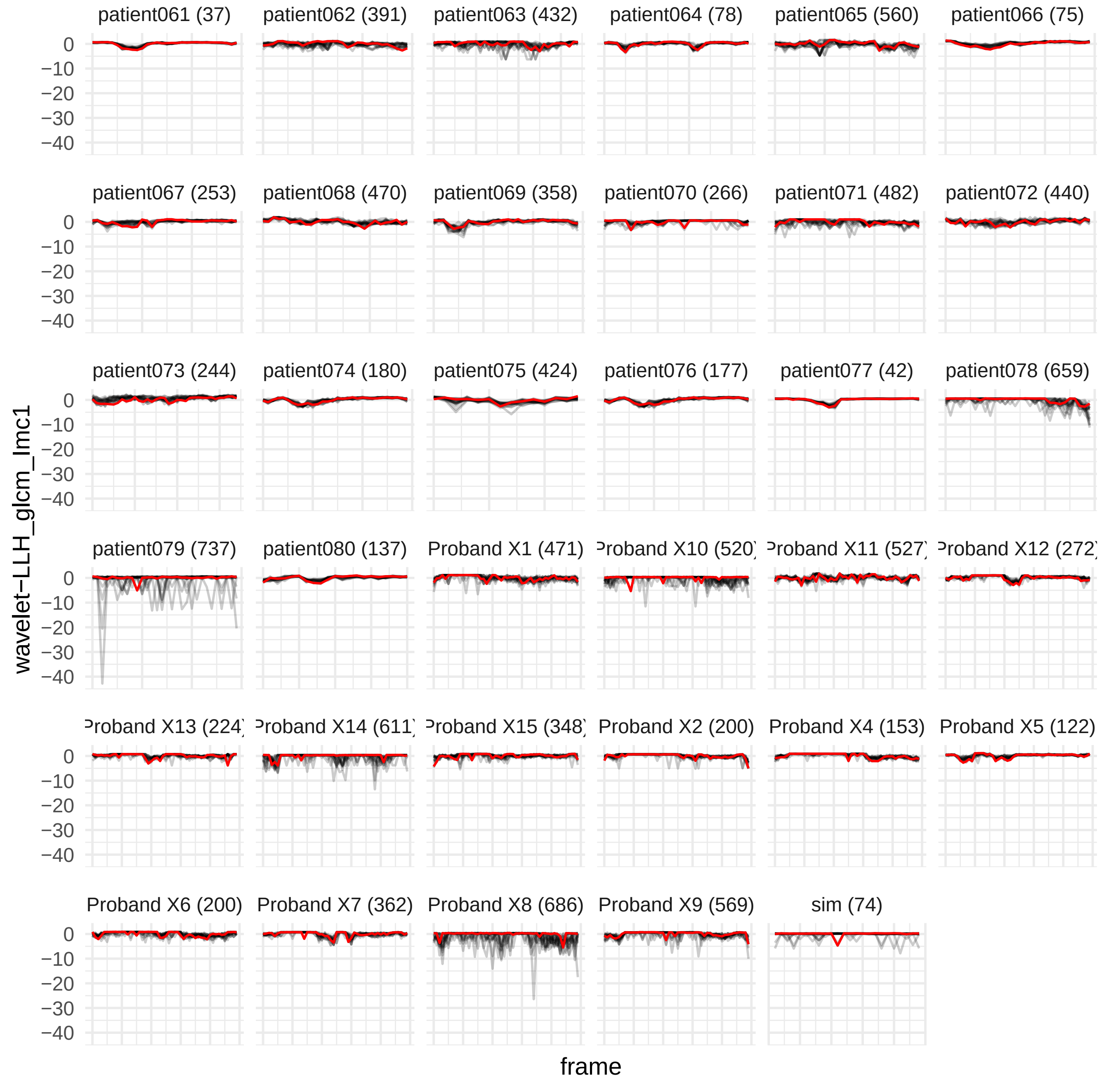

patient061 (37)
patient062 (391)
patient063 (432)
patient064 (78)
patient065 (560)
patient066 (75)
patient067 (253)
patient068 (470)
patient069 (358)
patient070 (266)
patient071 (482)
patient072 (440)
patient073 (244)
patient074 (180)
patient075 (424)
patient076 (177)
patient077 (42)
patient078 (659)
patient079 (737)
patient080 (137)
Proband X1 (471)
Proband X10 (520)
Proband X11 (527)
Proband X12 (272)
Proband X13 (224)
Proband X14 (611)
Proband X15 (348)
Proband X2 (200)
Proband X4 (153)
Proband X5 (122)
Proband X6 (200)
Proband X7 (362)
Proband X8 (686)
Proband X9 (569)
sim (74)
0
−10
−20
−30
−40
wavelet−LLH_glcm_Imc1
frame

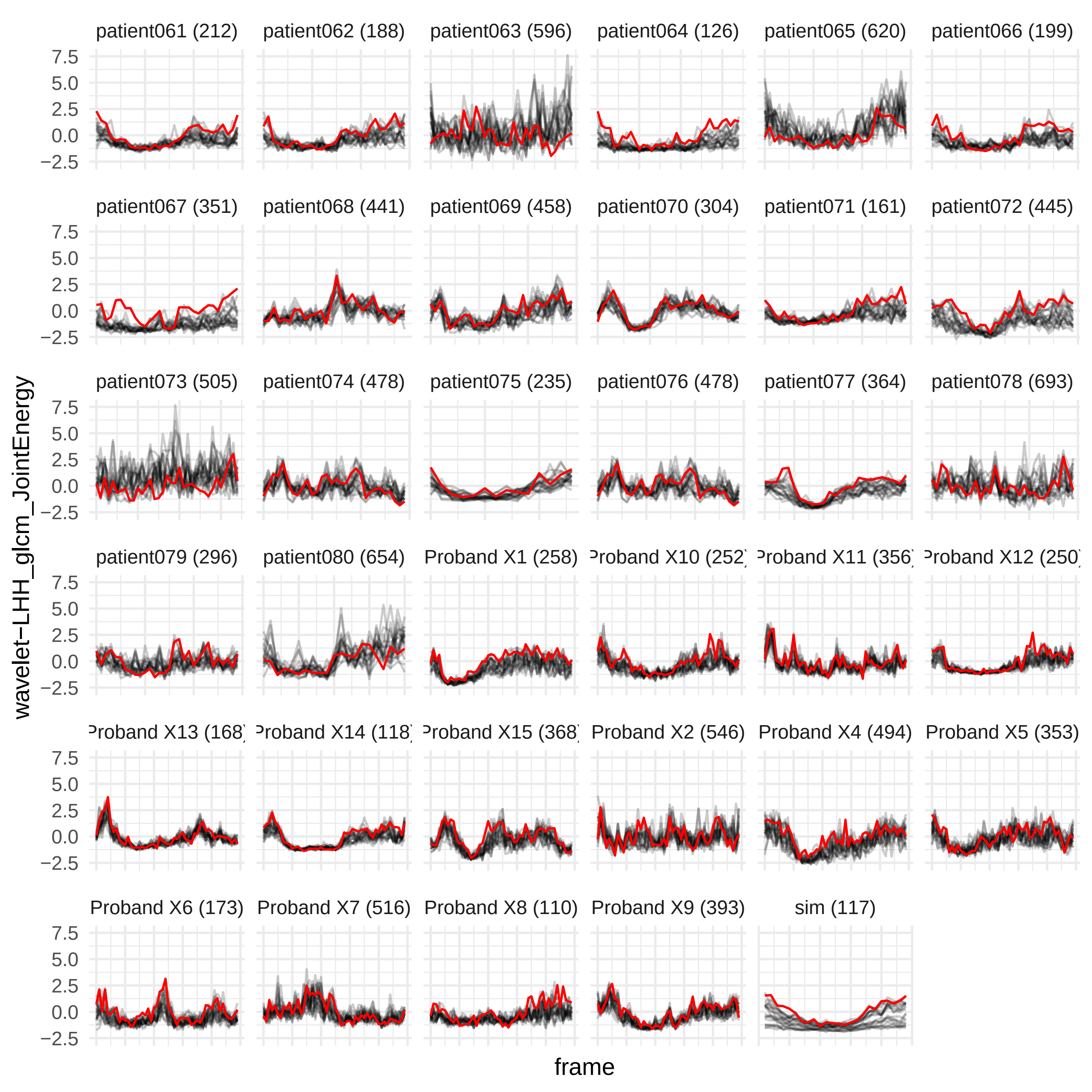
patient061 (212)
patient062 (188)
patient063 (596)
patient064 (126)
patient065 (620)
patient066 (199)
patient067 (351)
patient068 (441)
patient069 (458)
patient070 (304)
patient071 (161)
patient072 (445)
patient073 (505)
patient074 (478)
patient075 (235)
patient076 (478)
patient077 (364)
patient078 (693)
patient079 (296)
patient080 (654)
Proband X1 (258)
Proband X10 (252)
Proband X11 (356)
Proband X12 (250)
Proband X13 (168)
Proband X14 (118)
Proband X15 (368)
Proband X2 (546)
Proband X4 (494)
Proband X5 (353)
Proband X6 (173)
Proband X7 (516)
Proband X8 (110)
Proband X9 (393)
sim (117)
7.5
5.0
2.5
0.0
−2.5
wavelet−LHH_glcm_JointEnergy
frame

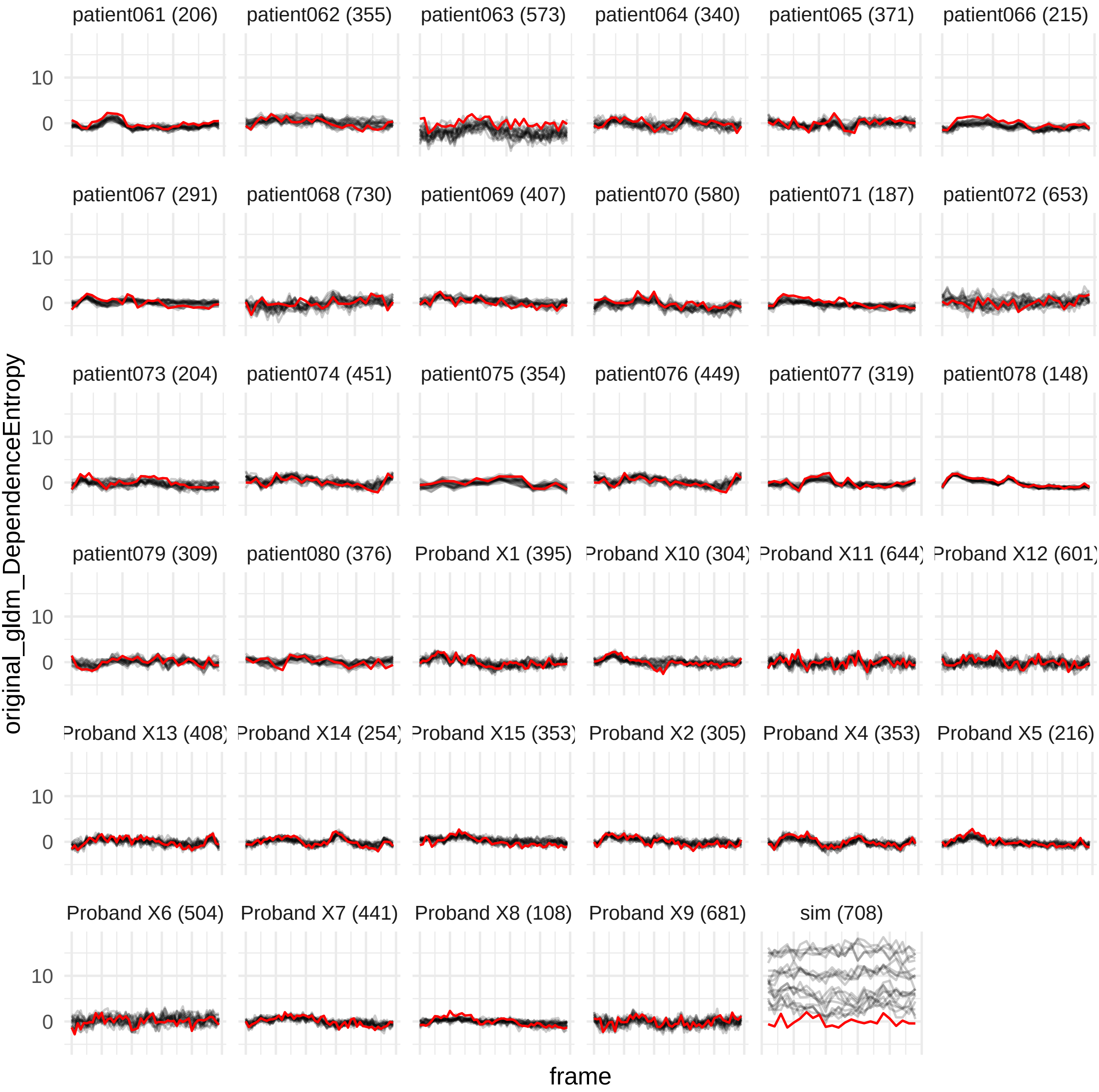

patient061 (206)
patient062 (355)
patient063 (573)
patient064 (340)
patient065 (371)
patient066 (215)
patient067 (291)
patient068 (730)
patient069 (407)
patient070 (580)
patient071 (187)
patient072 (653)
patient073 (204)
patient074 (451)
patient075 (354)
patient076 (449)
patient077 (319)
patient078 (148)
patient079 (309)
patient080 (376)
Proband X1 (395)
Proband X10 (304)
Proband X11 (644)
Proband X12 (601)
Proband X13 (408)
Proband X14 (254)
Proband X15 (353)
Proband X2 (305)
Proband X4 (353)
Proband X5 (216)
Proband X6 (504)
Proband X7 (441)
Proband X8 (108)
Proband X9 (681)
sim (708)
10
0
original_gldm_DependenceEntropy
frame

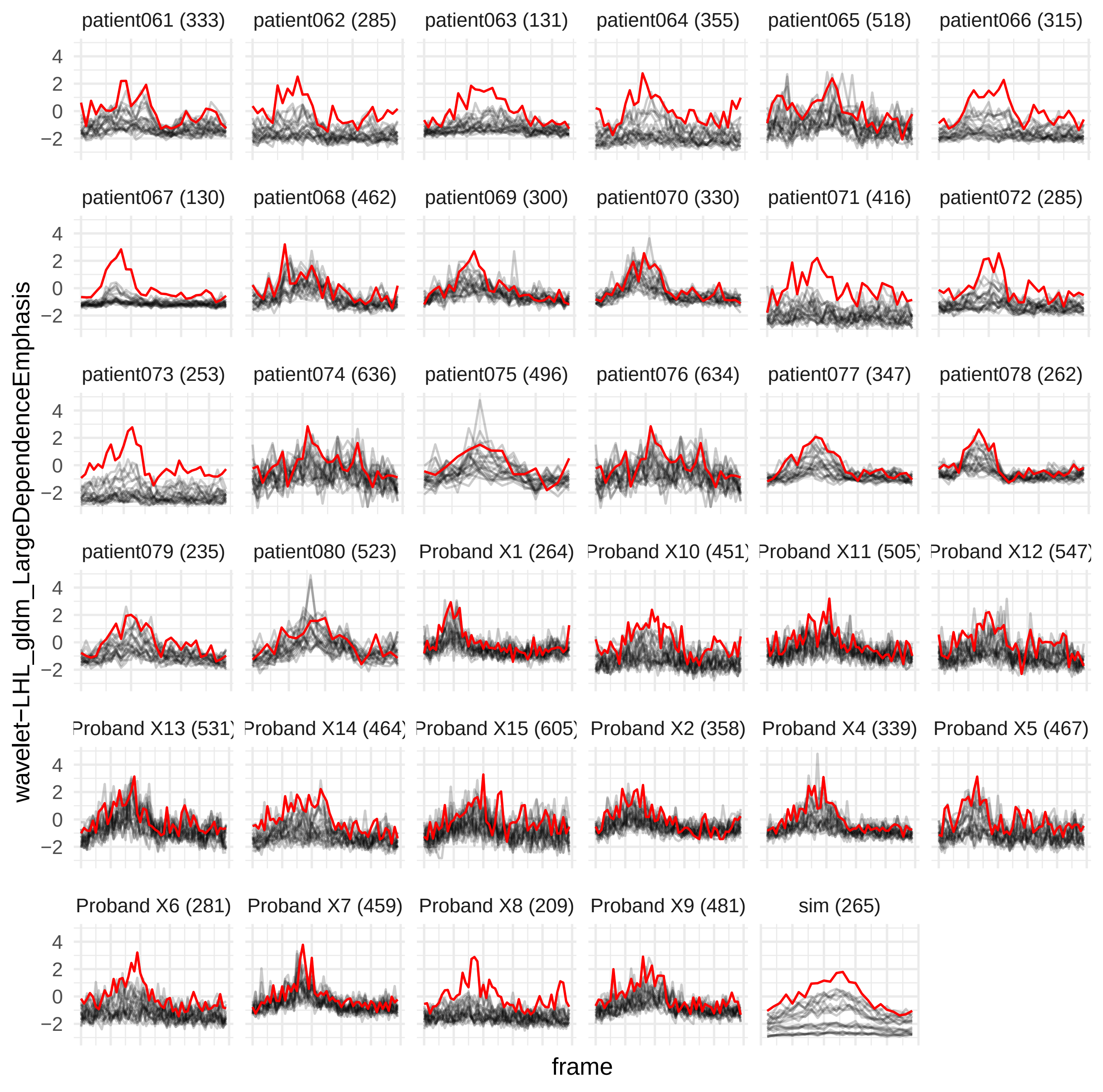
patient061 (333)
patient062 (285)
patient063 (131)
patient064 (355)
patient065 (518)
patient066 (315)
patient067 (130)
patient068 (462)
patient069 (300)
patient070 (330)
patient071 (416)
patient072 (285)
patient073 (253)
patient074 (636)
patient075 (496)
patient076 (634)
patient077 (347)
patient078 (262)
patient079 (235)
patient080 (523)
Proband X1 (264)
Proband X10 (451)
Proband X11 (505)
Proband X12 (547)
Proband X13 (531)
Proband X14 (464)
Proband X15 (605)
Proband X2 (358)
Proband X4 (339)
Proband X5 (467)
Proband X6 (281)
Proband X7 (459)
Proband X8 (209)
Proband X9 (481)
sim (265)
4
2
0
−2
wavelet−LHL_gldm_LargeDependenceEmphasis
frame

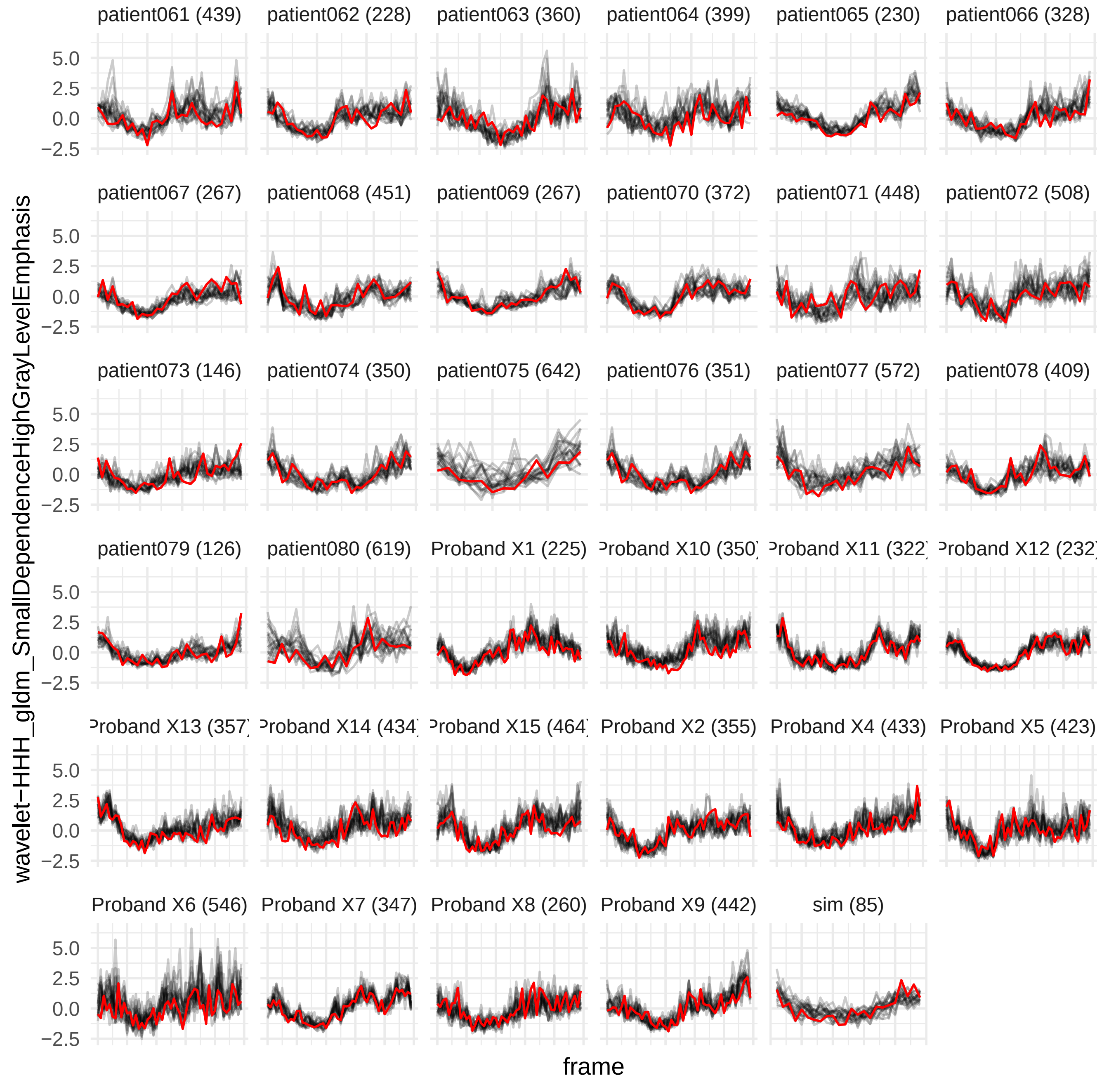

patient061 (439)
patient062 (228)
patient063 (360)
patient064 (399)
patient065 (230)
patient066 (328)
patient067 (267)
patient068 (451)
patient069 (267)
patient070 (372)
patient071 (448)
patient072 (508)
patient073 (146)
patient074 (350)
patient075 (642)
patient076 (351)
patient077 (572)
patient078 (409)
patient079 (126)
patient080 (619)
Proband X1 (225)
Proband X10 (350)
Proband X11 (322)
Proband X12 (232)
Proband X13 (357)
Proband X14 (434)
Proband X15 (464)
Proband X2 (355)
Proband X4 (433)
Proband X5 (423)
Proband X6 (546)
Proband X7 (347)
Proband X8 (260)
Proband X9 (442)
sim (85)
5.0
2.5
0.0
−2.5
wavelet−HHH_gldm_SmallDependenceHighGrayLevelEmphasis
frame

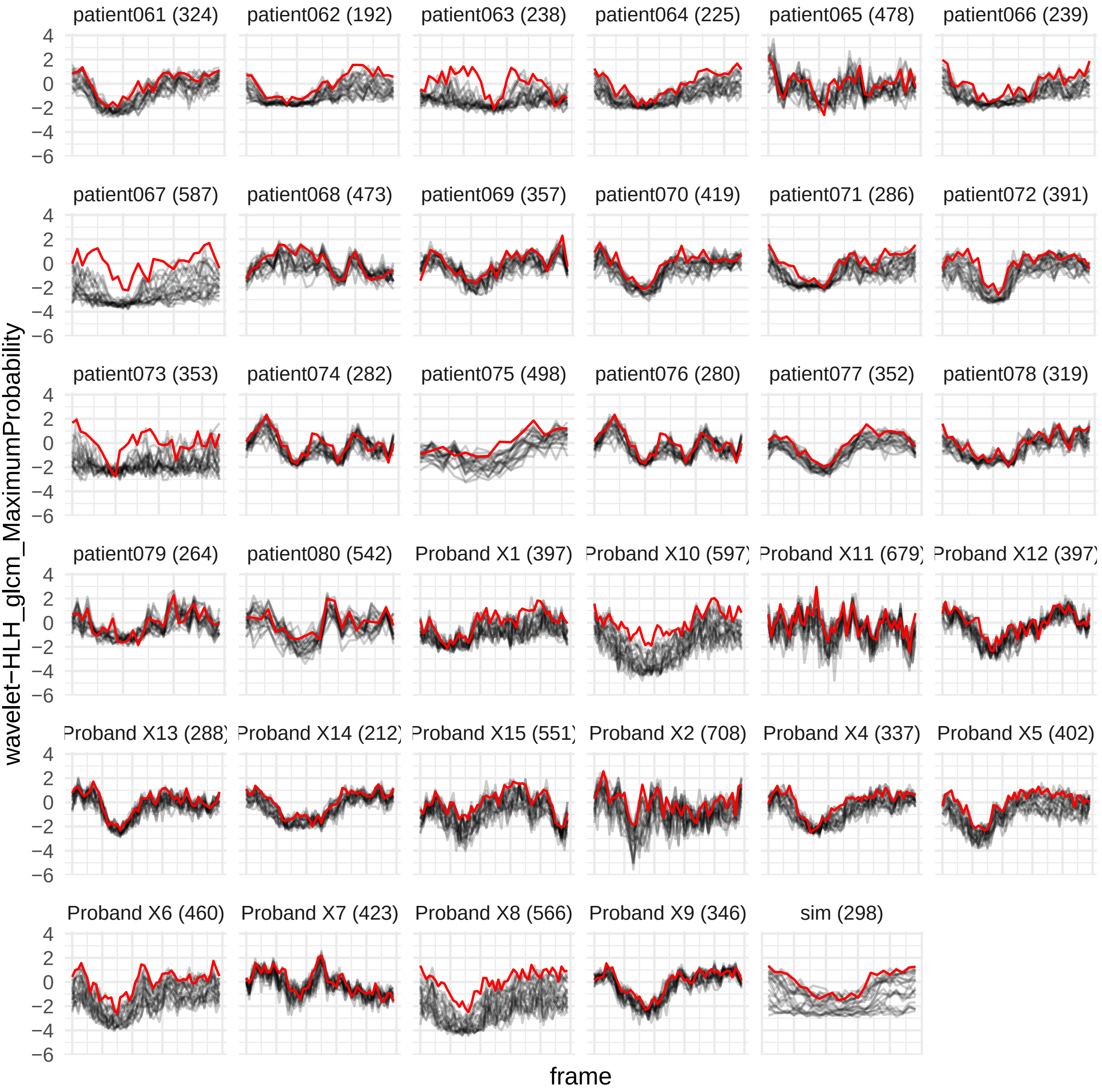

patient061 (324)
patient062 (192)
patient063 (238)
patient064 (225)
patient065 (478)
patient066 (239)
patient067 (587)
patient068 (473)
patient069 (357)
patient070 (419)
patient071 (286)
patient072 (391)
patient073 (353)
patient074 (282)
patient075 (498)
patient076 (280)
patient077 (352)
patient078 (319)
patient079 (264)
patient080 (542)
Proband X1 (397)
Proband X10 (597)
Proband X11 (679)
Proband X12 (397)
Proband X13 (288)
Proband X14 (212)
Proband X15 (551)
Proband X2 (708)
Proband X4 (337)
Proband X5 (402)
Proband X6 (460)
Proband X7 (423)
Proband X8 (566)
Proband X9 (346)
sim (298)
4
2
0
−2
−4
−6
wavelet−HLH_glcm_MaximumProbability
frame

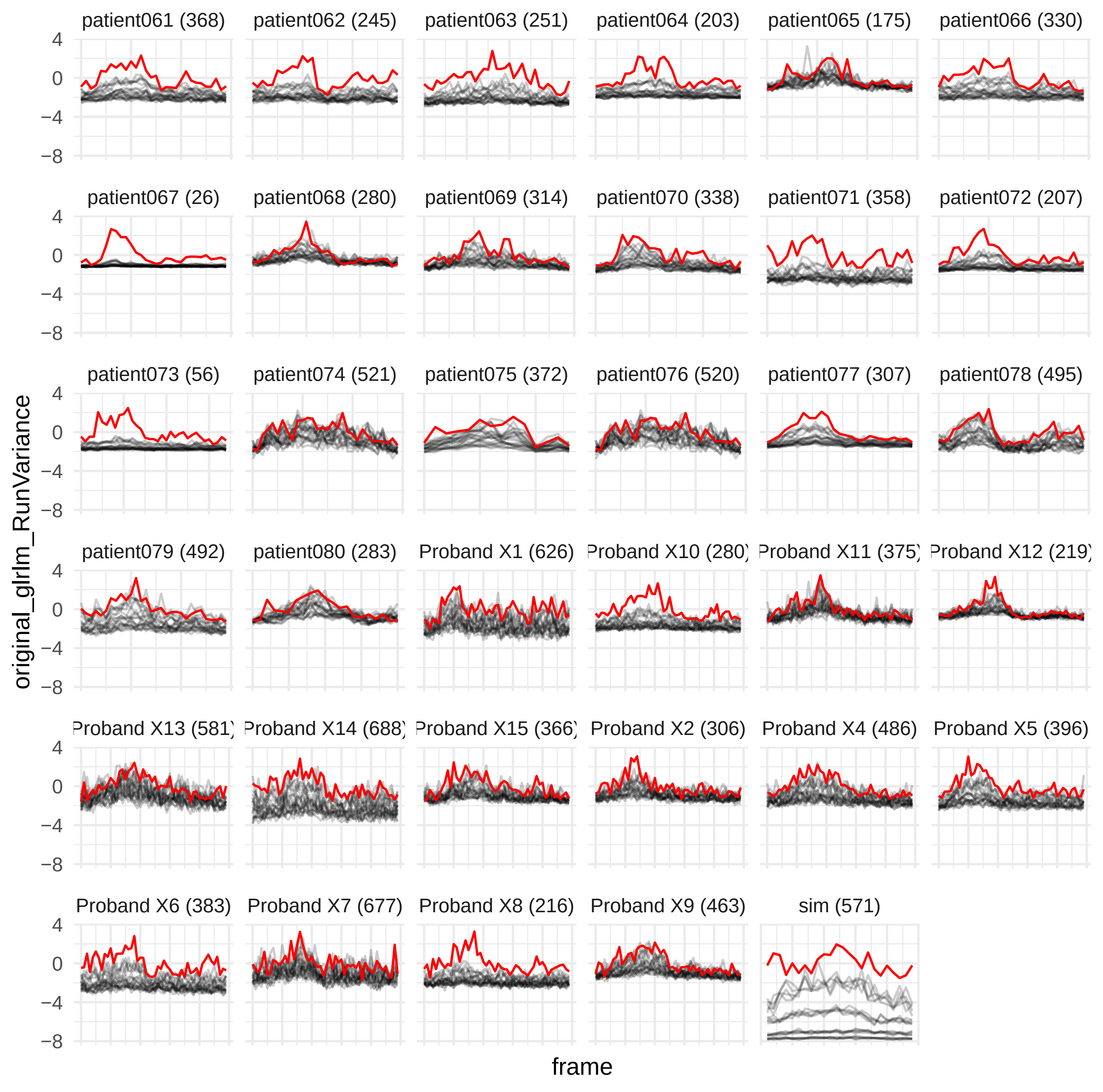
patient061 (368)
patient062 (245)
patient063 (251)
patient064 (203)
patient065 (175)
patient066 (330)
patient067 (26)
patient068 (280)
patient069 (314)
patient070 (338)
patient071 (358)
patient072 (207)
patient073 (56)
patient074 (521)
patient075 (372)
patient076 (520)
patient077 (307)
patient078 (495)
patient079 (492)
patient080 (283)
Proband X1 (626)
Proband X10 (280)
Proband X11 (375)
Proband X12 (219)
Proband X13 (581)
Proband X14 (688)
Proband X15 (366)
Proband X2 (306)
Proband X4 (486)
Proband X5 (396)
Proband X6 (383)
Proband X7 (677)
Proband X8 (216)
Proband X9 (463)
sim (571)
4
0
−4
−8
original_glrlm_RunVariance
frame

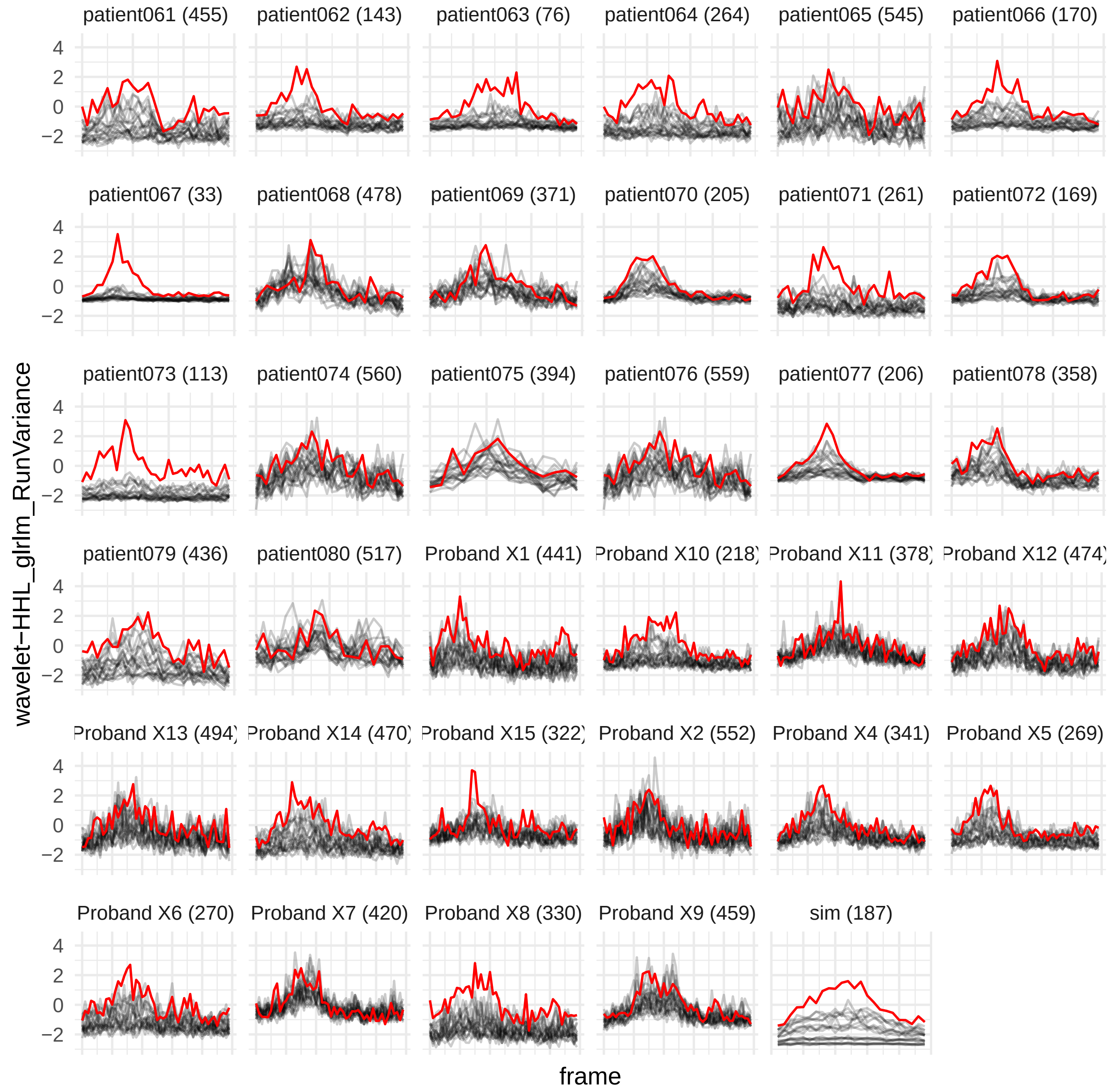
patient061 (455)
patient062 (143)
patient063 (76)
patient064 (264)
patient065 (545)
patient066 (170)
patient067 (33)
patient068 (478)
patient069 (371)
patient070 (205)
patient071 (261)
patient072 (169)
patient073 (113)
patient074 (560)
patient075 (394)
patient076 (559)
patient077 (206)
patient078 (358)
patient079 (436)
patient080 (517)
Proband X1 (441)
Proband X10 (218)
Proband X11 (378)
Proband X12 (474)
Proband X13 (494)
Proband X14 (470)
Proband X15 (322)
Proband X2 (552)
Proband X4 (341)
Proband X5 (269)
Proband X6 (270)
Proband X7 (420)
Proband X8 (330)
Proband X9 (459)
sim (187)
wavelet−HHL_glrlm_RunVariance
frame
4
2
0
−2

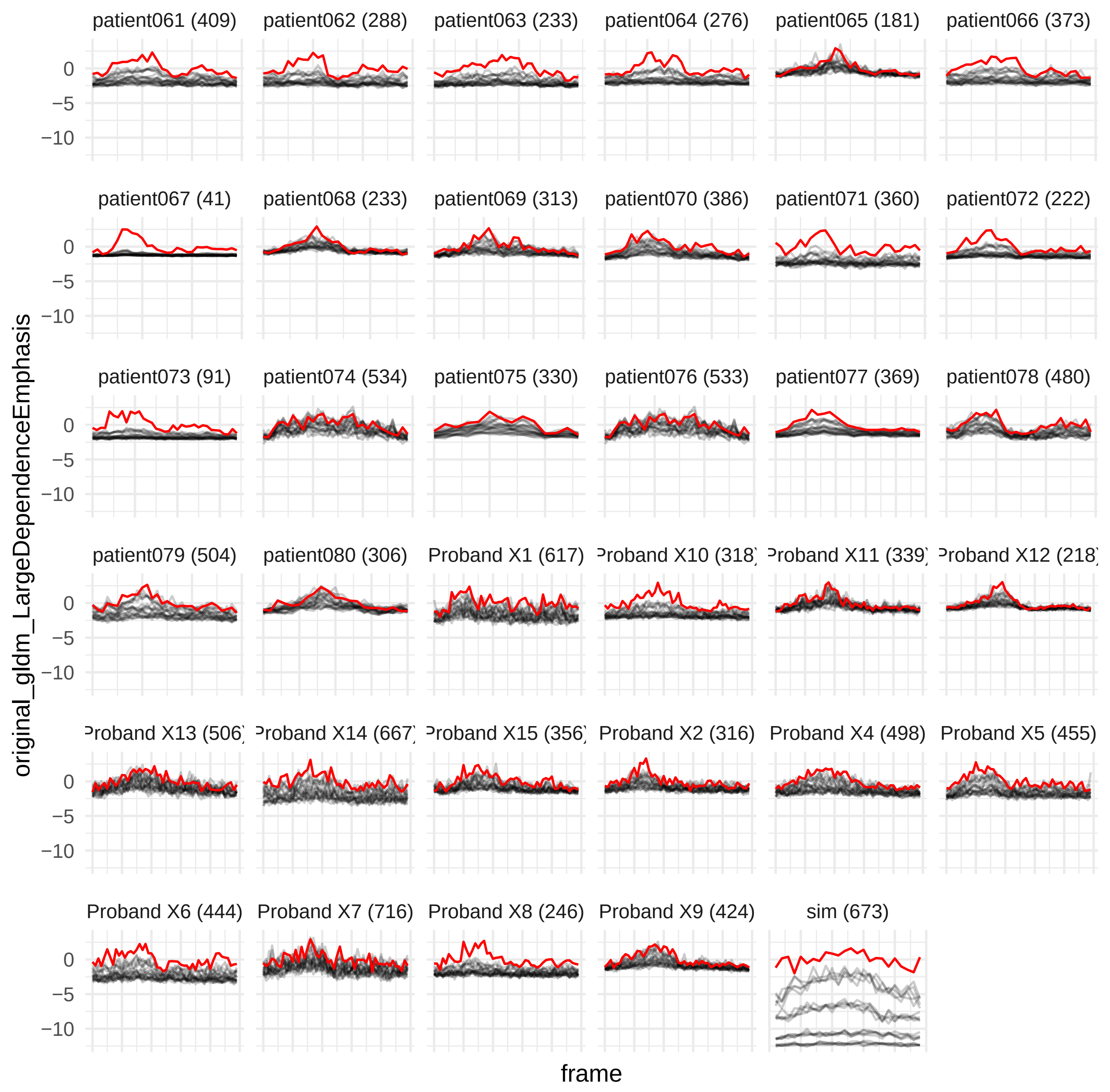

patient061 (409)
patient062 (288)
patient063 (233)
patient064 (276)
patient065 (181)
patient066 (373)
patient067 (41)
patient068 (233)
patient069 (313)
patient070 (386)
patient071 (360)
patient072 (222)
patient073 (91)
patient074 (534)
patient075 (330)
patient076 (533)
patient077 (369)
patient078 (480)
patient079 (504)
patient080 (306)
Proband X1 (617)
Proband X10 (318)
Proband X11 (339)
Proband X12 (218)
Proband X13 (506)
Proband X14 (667)
Proband X15 (356)
Proband X2 (316)
Proband X4 (498)
Proband X5 (455)
Proband X6 (444)
Proband X7 (716)
Proband X8 (246)
Proband X9 (424)
sim (673)
0
−5
−10
original_gldm_LargeDependenceEmphasis
frame

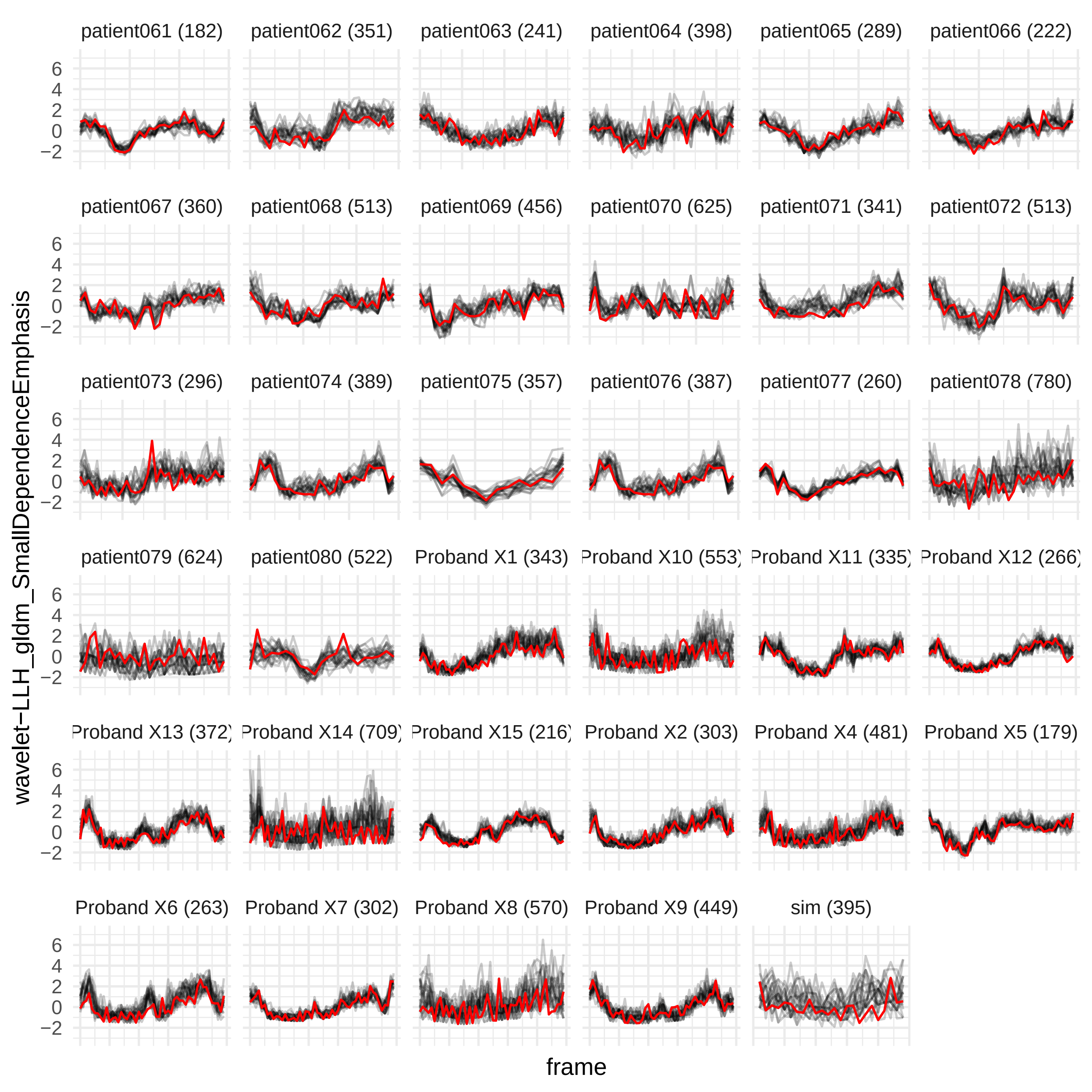
patient061 (182)
patient062 (351)
patient063 (241)
patient064 (398)
patient065 (289)
patient066 (222)
patient067 (360)
patient068 (513)
patient069 (456)
patient070 (625)
patient071 (341)
patient072 (513)
patient073 (296)
patient074 (389)
patient075 (357)
patient076 (387)
patient077 (260)
patient078 (780)
patient079 (624)
patient080 (522)
Proband X1 (343)
Proband X10 (553)
Proband X11 (335)
Proband X12 (266)
Proband X13 (372)
Proband X14 (709)
Proband X15 (216)
Proband X2 (303)
Proband X4 (481)
Proband X5 (179)
Proband X6 (263)
Proband X7 (302)
Proband X8 (570)
Proband X9 (449)
sim (395)
6
4
2
0
−2
wavelet−LLH_gldm_SmallDependenceEmphasis
frame

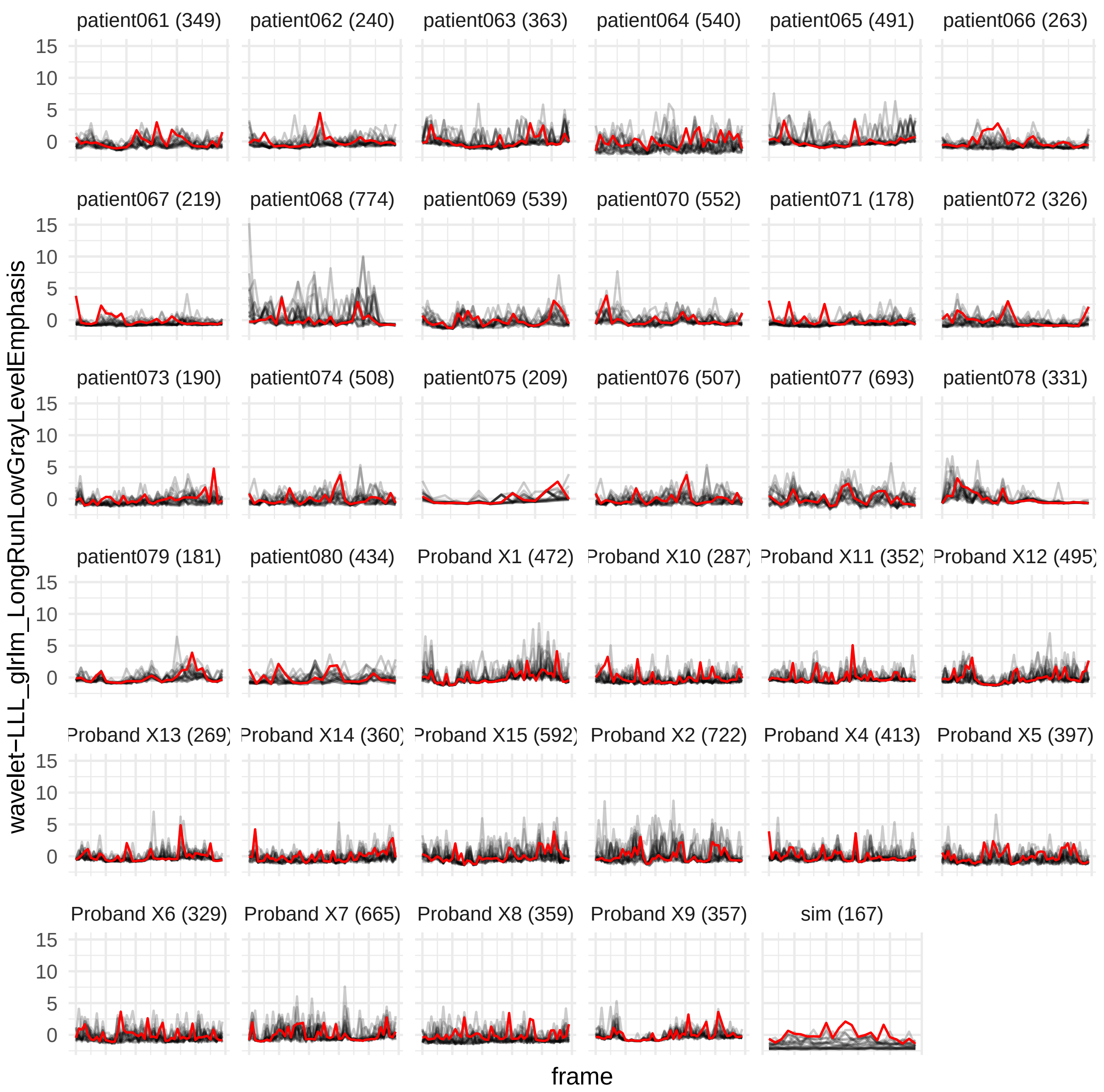
patient061 (349)
patient062 (240)
patient063 (363)
patient064 (540)
patient065 (491)
patient066 (263)
patient067 (219)
patient068 (774)
patient069 (539)
patient070 (552)
patient071 (178)
patient072 (326)
patient073 (190)
patient074 (508)
patient075 (209)
patient076 (507)
patient077 (693)
patient078 (331)
patient079 (181)
patient080 (434)
Proband X1 (472)
Proband X10 (287)
Proband X11 (352)
Proband X12 (495)
Proband X13 (269)
Proband X14 (360)
Proband X15 (592)
Proband X2 (722)
Proband X4 (413)
Proband X5 (397)
Proband X6 (329)
Proband X7 (665)
Proband X8 (359)
Proband X9 (357)
sim (167)
wavelet−LLL_glrlm_LongRunLowGrayLevelEmphasis
frame
0
5
10
15

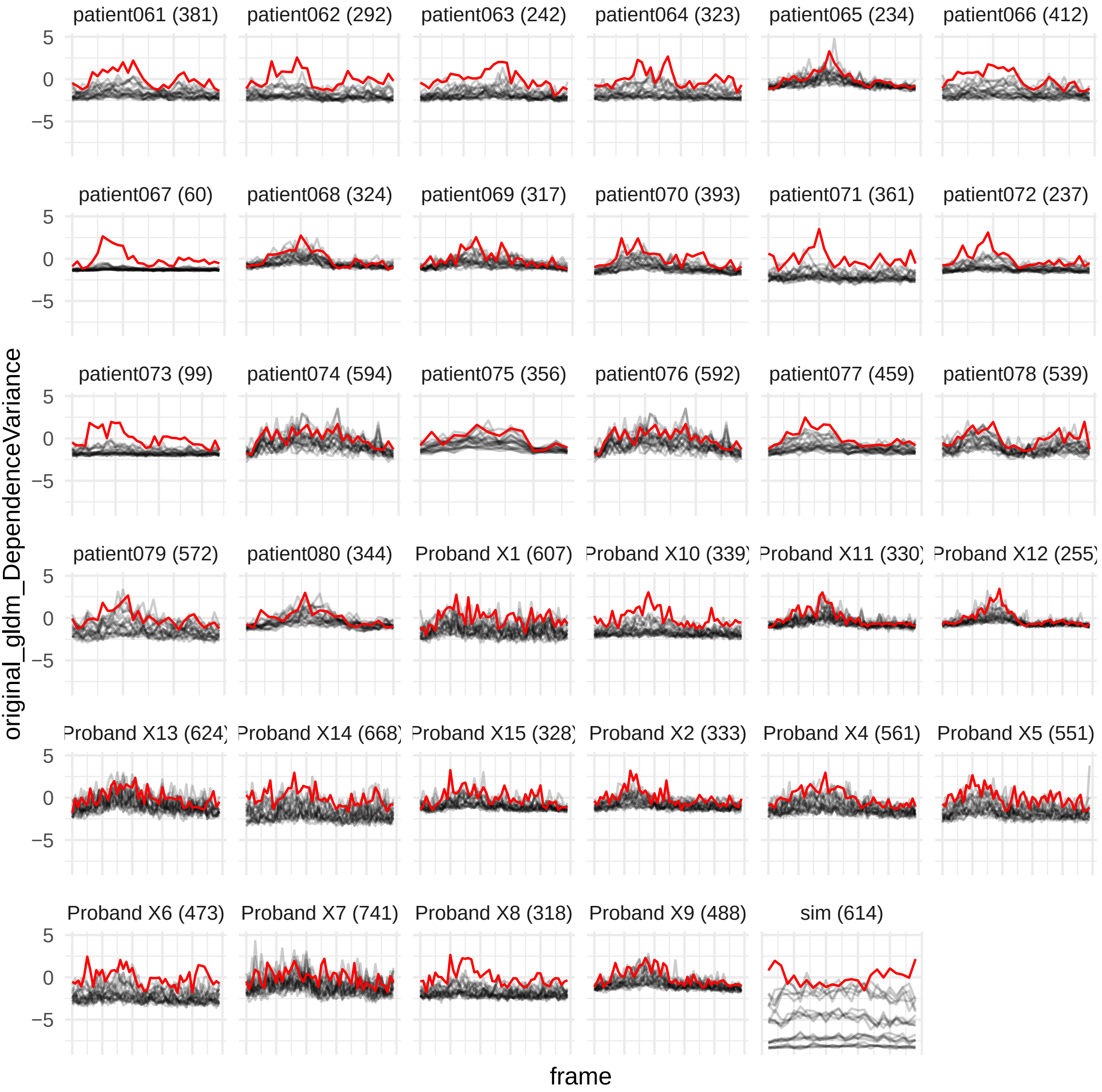

patient061 (381)
patient062 (292)
patient063 (242)
patient064 (323)
patient065 (234)
patient066 (412)
patient067 (60)
patient068 (324)
patient069 (317)
patient070 (393)
patient071 (361)
patient072 (237)
patient073 (99)
patient074 (594)
patient075 (356)
patient076 (592)
patient077 (459)
patient078 (539)
patient079 (572)
patient080 (344)
Proband X1 (607)
Proband X10 (339)
Proband X11 (330)
Proband X12 (255)
Proband X13 (624)
Proband X14 (668)
Proband X15 (328)
Proband X2 (333)
Proband X4 (561)
Proband X5 (551)
Proband X6 (473)
Proband X7 (741)
Proband X8 (318)
Proband X9 (488)
sim (614)
5
0
−5
original_gldm_DependenceVariance
frame

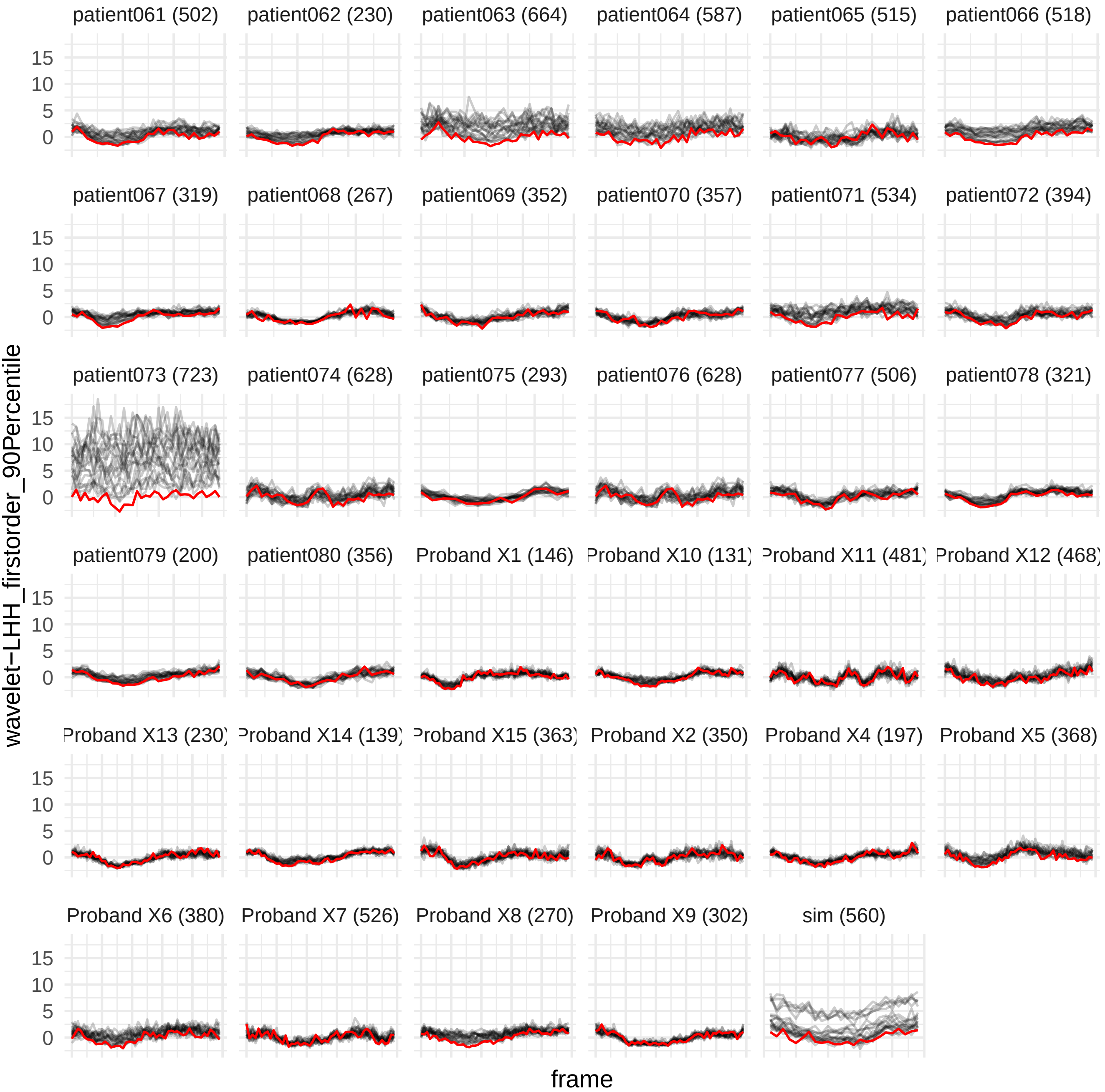
patient061 (502)
patient062 (230)
patient063 (664)
patient064 (587)
patient065 (515)
patient066 (518)
patient067 (319)
patient068 (267)
patient069 (352)
patient070 (357)
patient071 (534)
patient072 (394)
patient073 (723)
patient074 (628)
patient075 (293)
patient076 (628)
patient077 (506)
patient078 (321)
patient079 (200)
patient080 (356)
Proband X1 (146)
Proband X10 (131)
Proband X11 (481)
Proband X12 (468)
Proband X13 (230)
Proband X14 (139)
Proband X15 (363)
Proband X2 (350)
Proband X4 (197)
Proband X5 (368)
Proband X6 (380)
Proband X7 (526)
Proband X8 (270)
Proband X9 (302)
sim (560)
wavelet−LHH_firstorder_90Percentile
frame
0
5
10
15

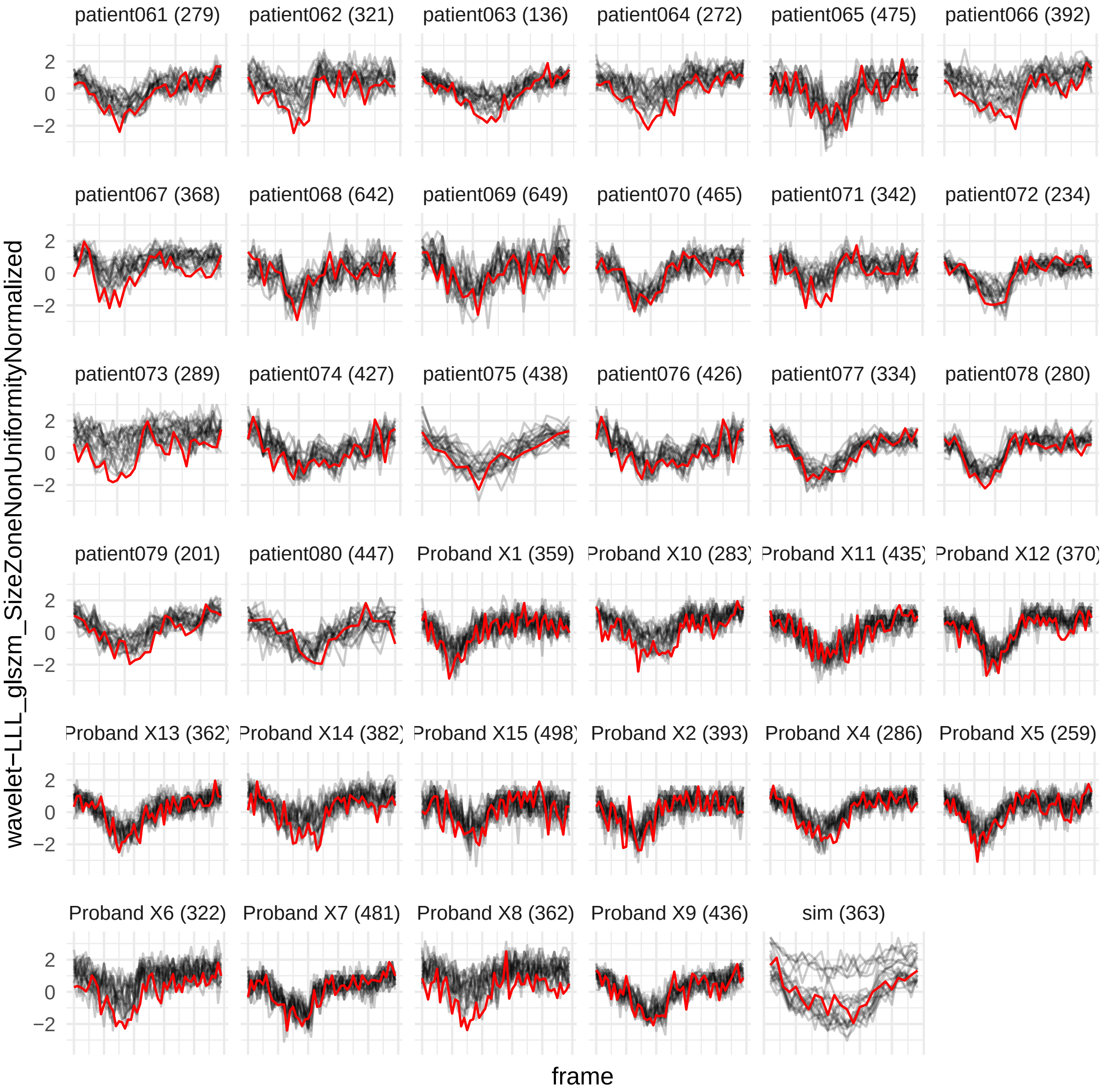

patient061 (279)
patient062 (321)
patient063 (136)
patient064 (272)
patient065 (475)
patient066 (392)
patient067 (368)
patient068 (642)
patient069 (649)
patient070 (465)
patient071 (342)
patient072 (234)
patient073 (289)
patient074 (427)
patient075 (438)
patient076 (426)
patient077 (334)
patient078 (280)
patient079 (201)
patient080 (447)
Proband X1 (359)
Proband X10 (283)
Proband X11 (435)
Proband X12 (370)
Proband X13 (362)
Proband X14 (382)
Proband X15 (498)
Proband X2 (393)
Proband X4 (286)
Proband X5 (259)
Proband X6 (322)
Proband X7 (481)
Proband X8 (362)
Proband X9 (436)
sim (363)
2
0
−2
wavelet−LLL_glszm_SizeZoneNonUniformityNormalized
frame

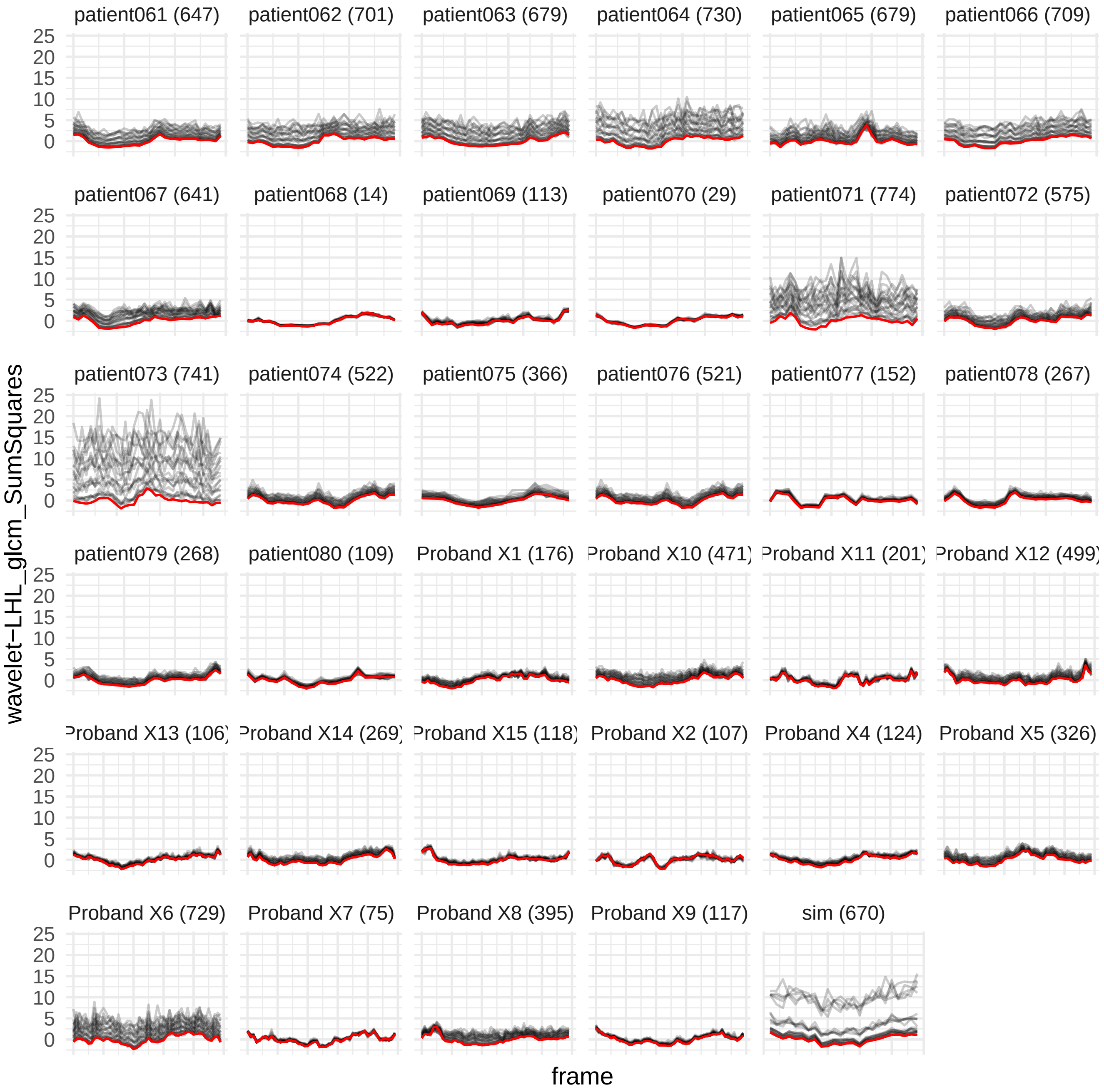
patient061 (647)
patient062 (701)
patient063 (679)
patient064 (730)
patient065 (679)
patient066 (709)
patient067 (641)
patient068 (14)
patient069 (113)
patient070 (29)
patient071 (774)
patient072 (575)
patient073 (741)
patient074 (522)
patient075 (366)
patient076 (521)
patient077 (152)
patient078 (267)
patient079 (268)
patient080 (109)
Proband X1 (176)
Proband X10 (471)
Proband X11 (201)
Proband X12 (499)
Proband X13 (106)
Proband X14 (269)
Proband X15 (118)
Proband X2 (107)
Proband X4 (124)
Proband X5 (326)
Proband X6 (729)
Proband X7 (75)
Proband X8 (395)
Proband X9 (117)
sim (670)
25
20
15
10
5
0
wavelet−LHL_glcm_SumSquares
frame

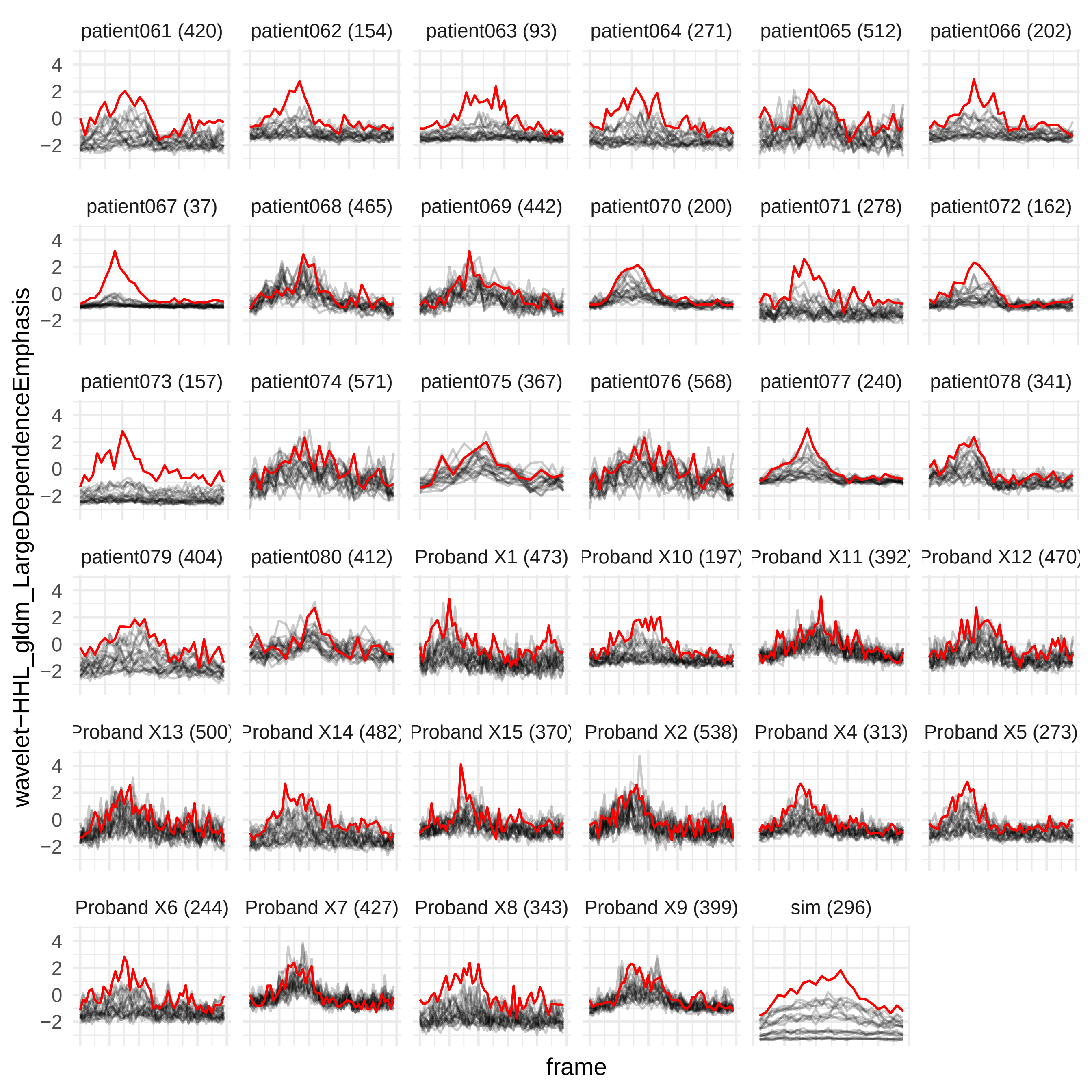

patient061 (420)
patient062 (154)
patient063 (93)
patient064 (271)
patient065 (512)
patient066 (202)
patient067 (37)
patient068 (465)
patient069 (442)
patient070 (200)
patient071 (278)
patient072 (162)
patient073 (157)
patient074 (571)
patient075 (367)
patient076 (568)
patient077 (240)
patient078 (341)
patient079 (404)
patient080 (412)
Proband X1 (473)
Proband X10 (197)
Proband X11 (392)
Proband X12 (470)
Proband X13 (500)
Proband X14 (482)
Proband X15 (370)
Proband X2 (538)
Proband X4 (313)
Proband X5 (273)
Proband X6 (244)
Proband X7 (427)
Proband X8 (343)
Proband X9 (399)
sim (296)
wavelet−HHL_gldm_LargeDependenceEmphasis
frame

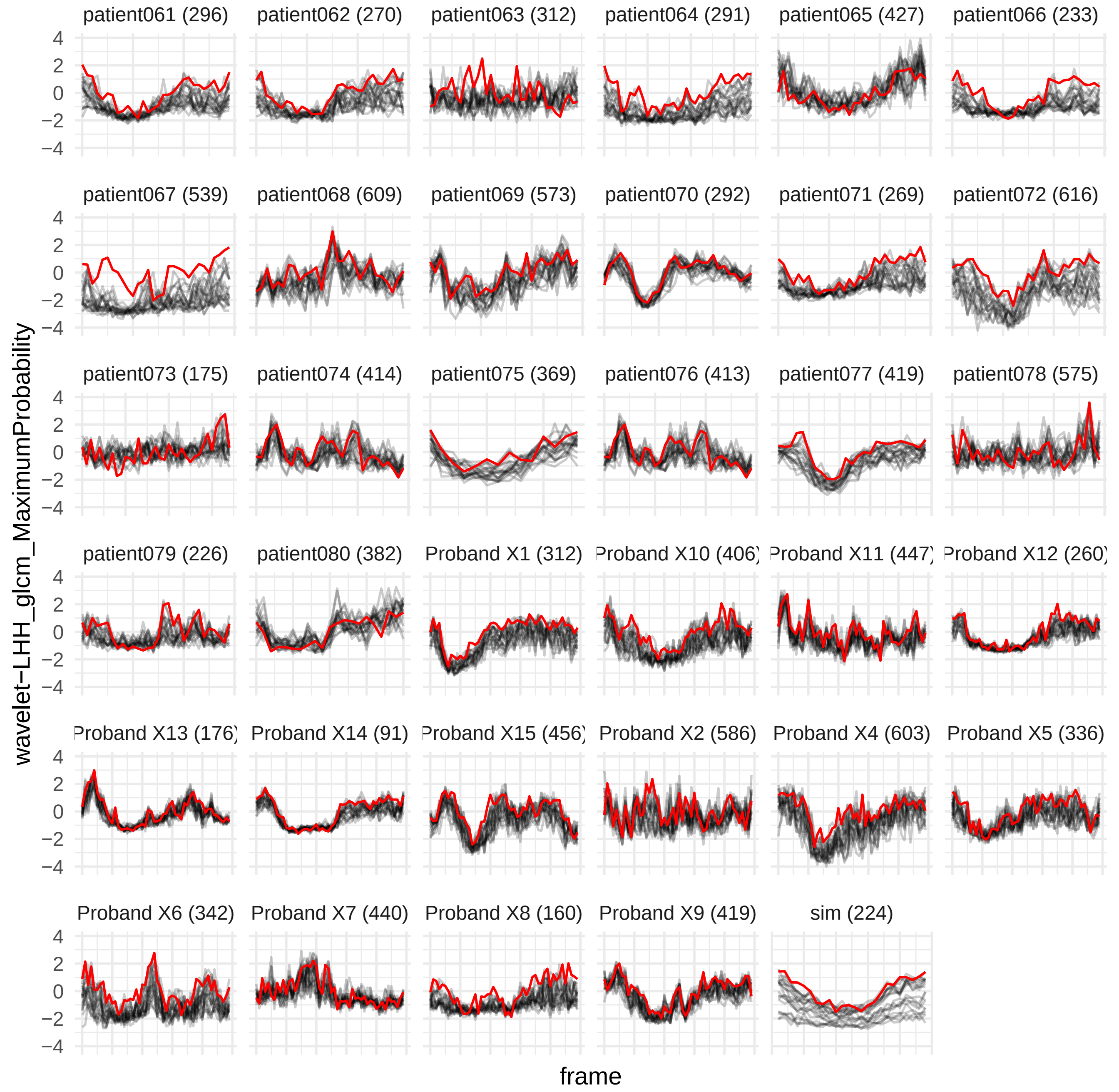

patient061 (296)
patient062 (270)
patient063 (312)
patient064 (291)
patient065 (427)
patient066 (233)
patient067 (539)
patient068 (609)
patient069 (573)
patient070 (292)
patient071 (269)
patient072 (616)
patient073 (175)
patient074 (414)
patient075 (369)
patient076 (413)
patient077 (419)
patient078 (575)
patient079 (226)
patient080 (382)
Proband X1 (312)
Proband X10 (406)
Proband X11 (447)
Proband X12 (260)
Proband X13 (176)
Proband X14 (91)
Proband X15 (456)
Proband X2 (586)
Proband X4 (603)
Proband X5 (336)
Proband X6 (342)
Proband X7 (440)
Proband X8 (160)
Proband X9 (419)
sim (224)
4
2
0
−2
−4
wavelet−LHH_glcm_MaximumProbability
frame

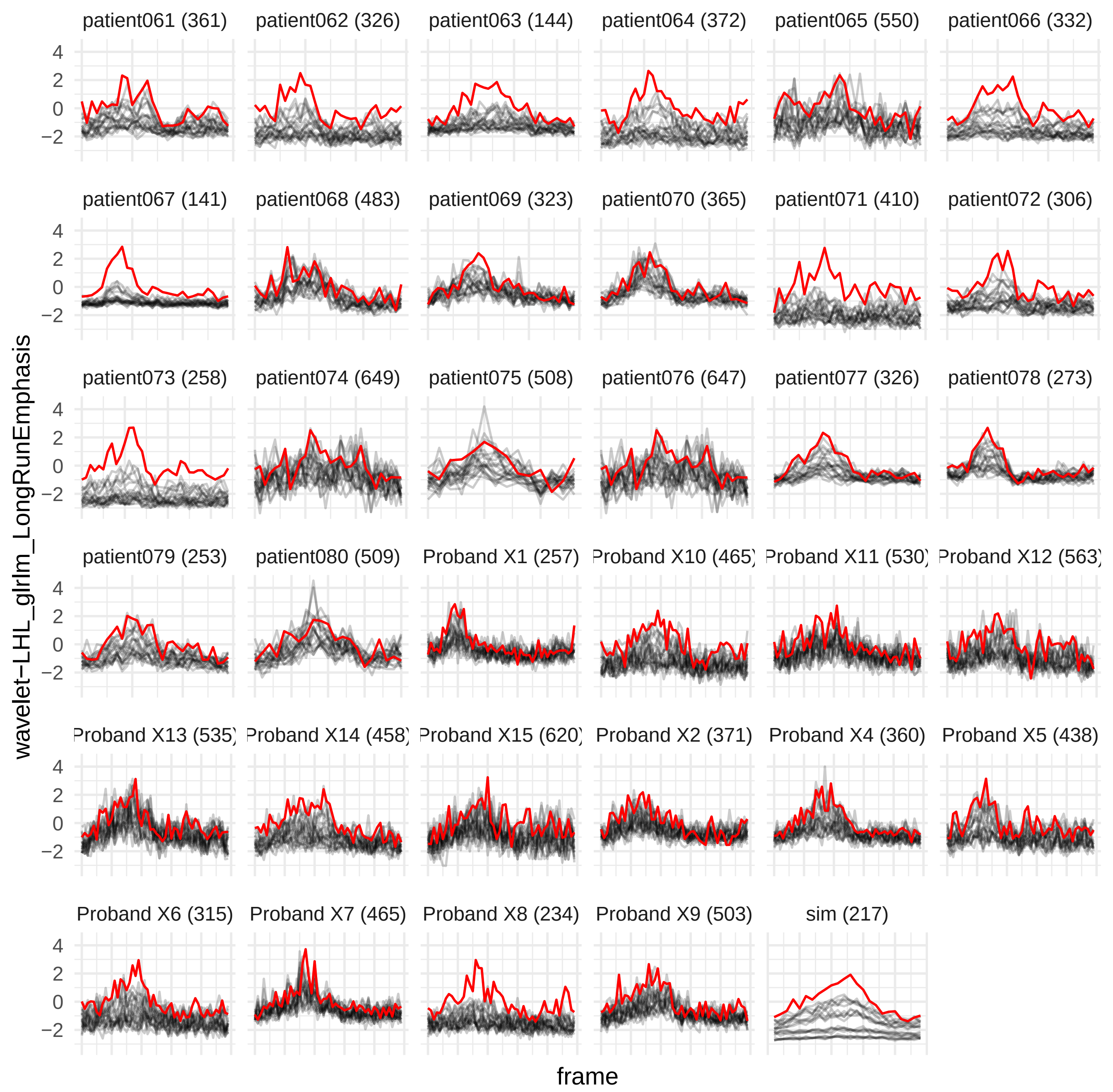

patient061 (361)
patient062 (326)
patient063 (144)
patient064 (372)
patient065 (550)
patient066 (332)
patient067 (141)
patient068 (483)
patient069 (323)
patient070 (365)
patient071 (410)
patient072 (306)
patient073 (258)
patient074 (649)
patient075 (508)
patient076 (647)
patient077 (326)
patient078 (273)
patient079 (253)
patient080 (509)
Proband X1 (257)
Proband X10 (465)
Proband X11 (530)
Proband X12 (563)
Proband X13 (535)
Proband X14 (458)
Proband X15 (620)
Proband X2 (371)
Proband X4 (360)
Proband X5 (438)
Proband X6 (315)
Proband X7 (465)
Proband X8 (234)
Proband X9 (503)
sim (217)
wavelet−LHL_glrlm_LongRunEmphasis
frame

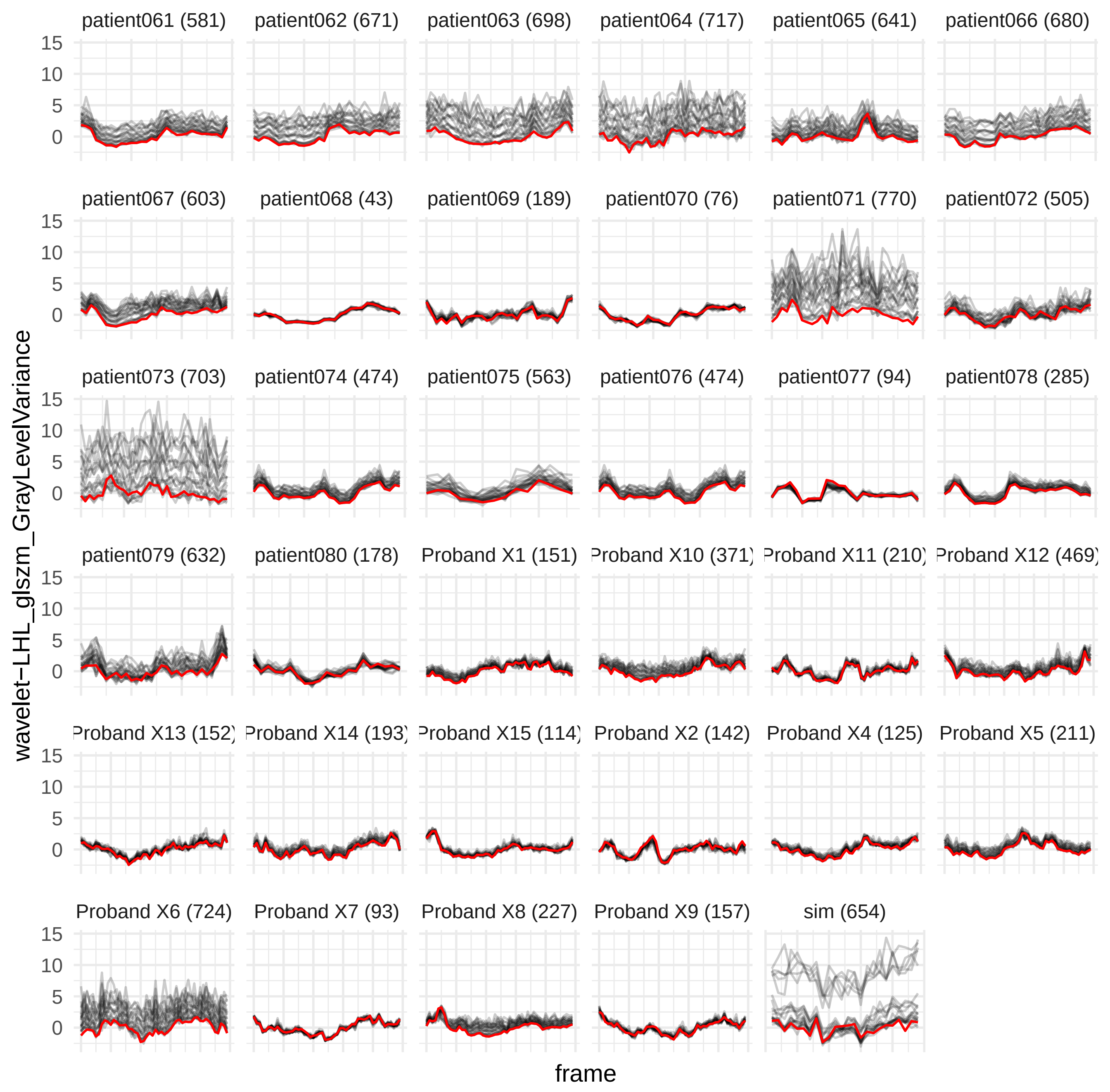

patient061 (581)
patient062 (671)
patient063 (698)
patient064 (717)
patient065 (641)
patient066 (680)
patient067 (603)
patient068 (43)
patient069 (189)
patient070 (76)
patient071 (770)
patient072 (505)
patient073 (703)
patient074 (474)
patient075 (563)
patient076 (474)
patient077 (94)
patient078 (285)
patient079 (632)
patient080 (178)
Proband X1 (151)
Proband X10 (371)
Proband X11 (210)
Proband X12 (469)
Proband X13 (152)
Proband X14 (193)
Proband X15 (114)
Proband X2 (142)
Proband X4 (125)
Proband X5 (211)
Proband X6 (724)
Proband X7 (93)
Proband X8 (227)
Proband X9 (157)
sim (654)
15
10
5
0
wavelet−LHL_glszm_GrayLevelVariance
frame

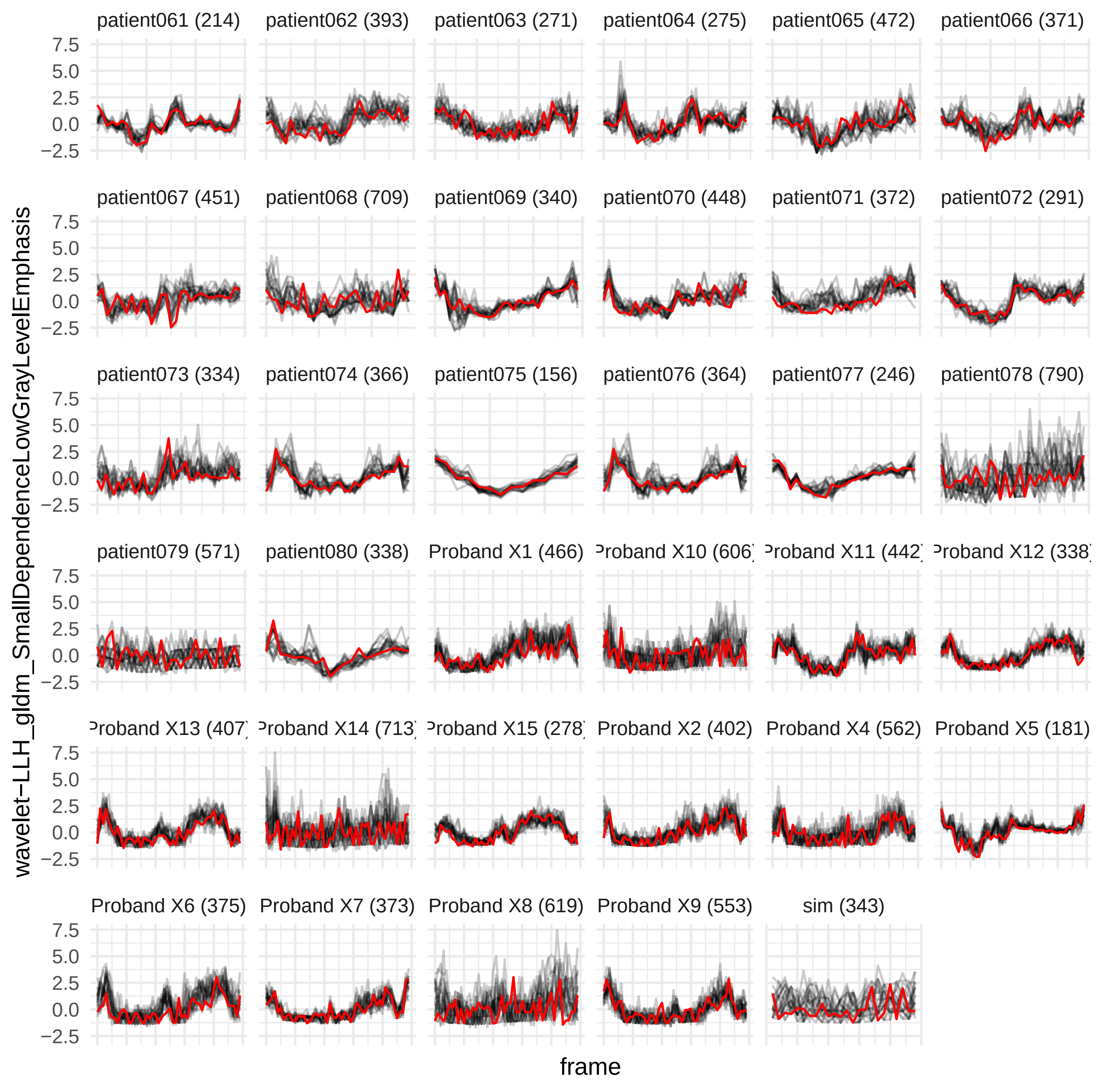

patient061 (214)
patient062 (393)
patient063 (271)
patient064 (275)
patient065 (472)
patient066 (371)
patient067 (451)
patient068 (709)
patient069 (340)
patient070 (448)
patient071 (372)
patient072 (291)
patient073 (334)
patient074 (366)
patient075 (156)
patient076 (364)
patient077 (246)
patient078 (790)
patient079 (571)
patient080 (338)
Proband X1 (466)
Proband X10 (606
Proband X11 (442
Proband X12 (338
Proband X13 (407
Proband X14 (713
Proband X15 (278
Proband X2 (402)
Proband X4 (562)
Proband X5 (181)
Proband X6 (375)
Proband X7 (373)
Proband X8 (619)
Proband X9 (553)
sim (343)
7.5
5.0
2.5
0.0
−2.5
wavelet−LLH_gldm_SmallDependenceLowGrayLevelEmphasis
frame

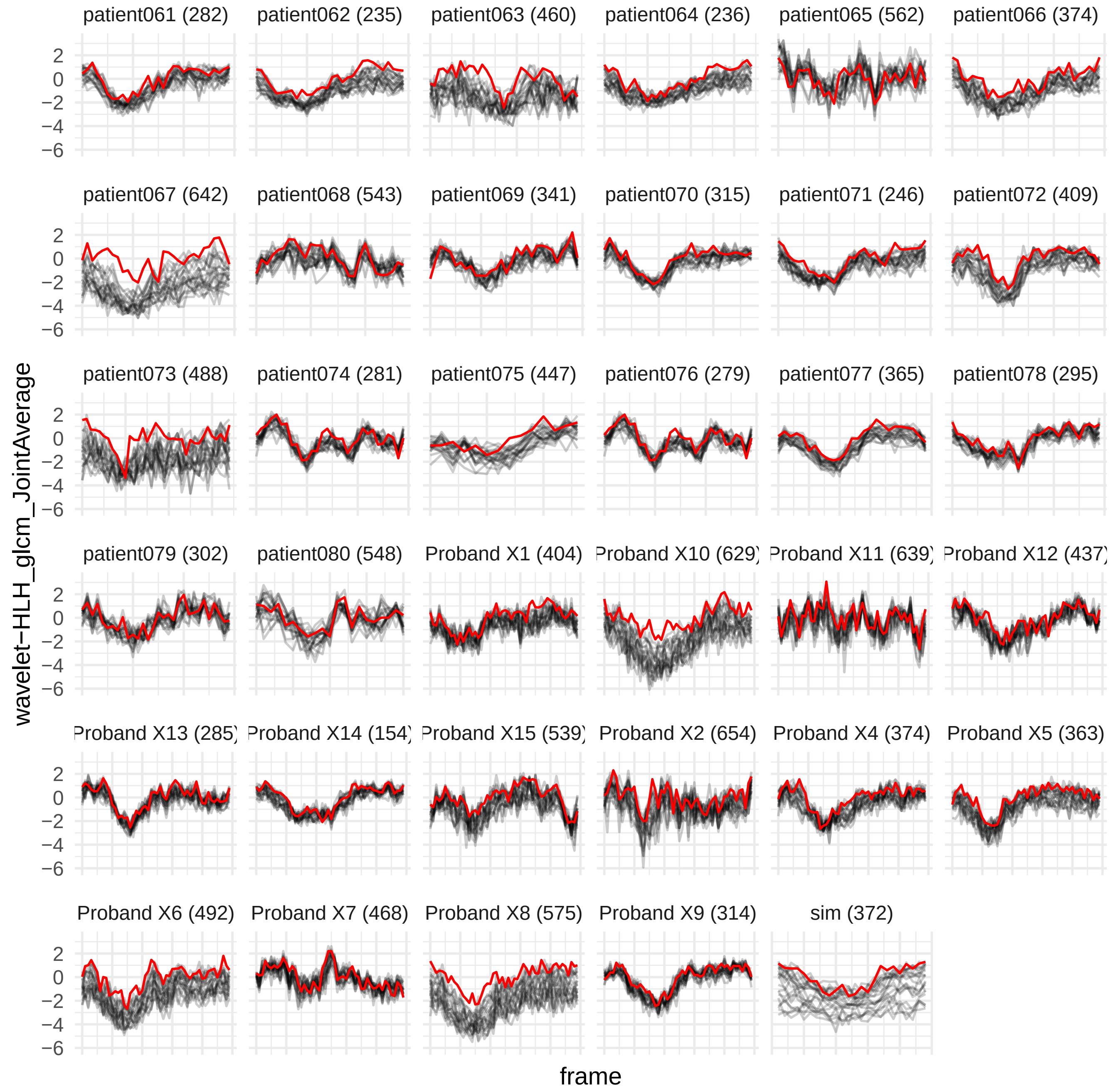
patient061 (282)
patient062 (235)
patient063 (460)
patient064 (236)
patient065 (562)
patient066 (374)
patient067 (642)
patient068 (543)
patient069 (341)
patient070 (315)
patient071 (246)
patient072 (409)
patient073 (488)
patient074 (281)
patient075 (447)
patient076 (279)
patient077 (365)
patient078 (295)
patient079 (302)
patient080 (548)
Proband X1 (404)
Proband X10 (629)
Proband X11 (639)
Proband X12 (437)
Proband X13 (285)
Proband X14 (154)
Proband X15 (539)
Proband X2 (654)
Proband X4 (374)
Proband X5 (363)
Proband X6 (492)
Proband X7 (468)
Proband X8 (575)
Proband X9 (314)
sim (372)
wavelet−HLH_glcm_JointAverage
frame
2
0
−2
−4
−6

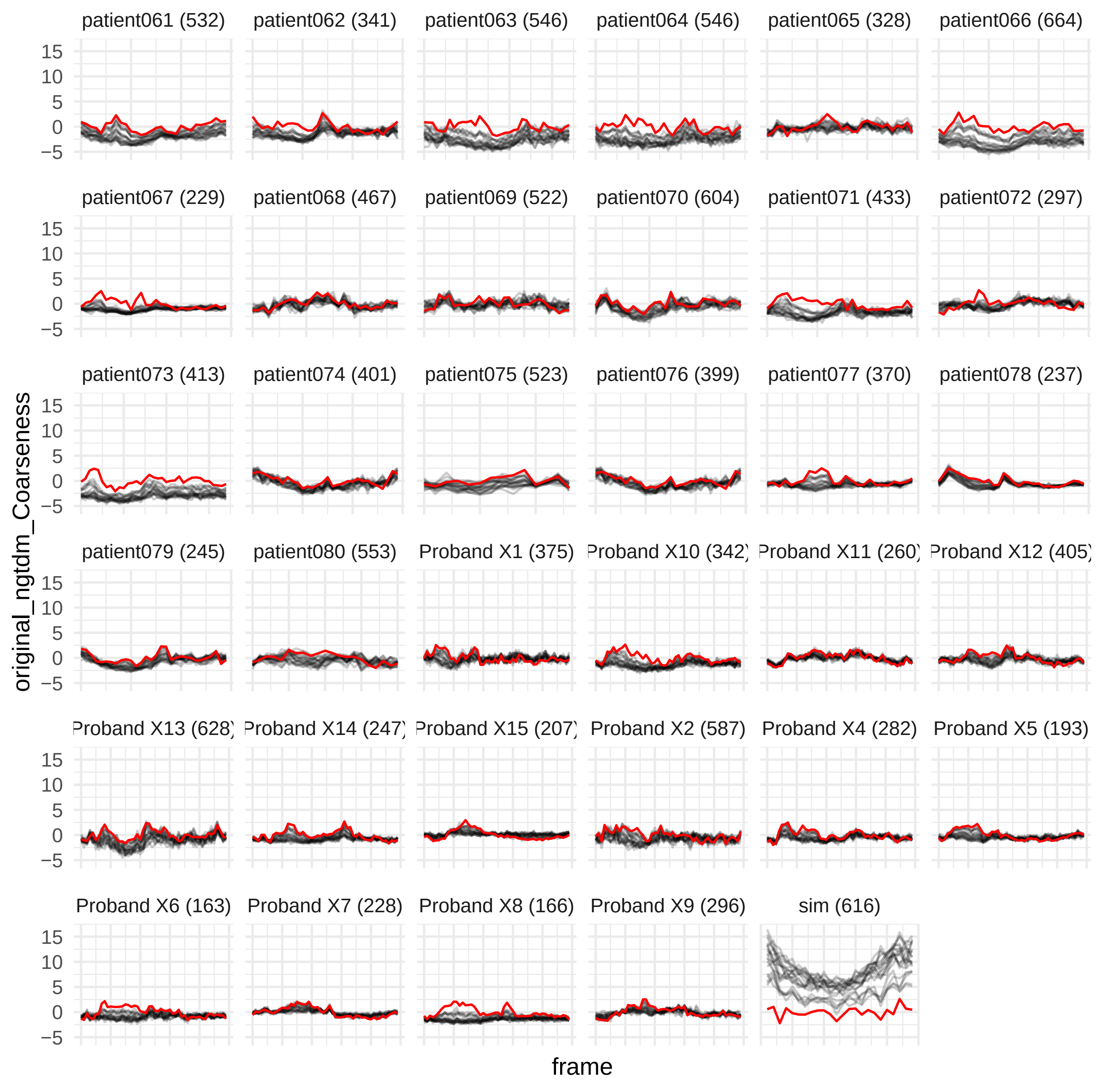

patient061 (532)
patient062 (341)
patient063 (546)
patient064 (546)
patient065 (328)
patient066 (664)
patient067 (229)
patient068 (467)
patient069 (522)
patient070 (604)
patient071 (433)
patient072 (297)
patient073 (413)
patient074 (401)
patient075 (523)
patient076 (399)
patient077 (370)
patient078 (237)
patient079 (245)
patient080 (553)
Proband X1 (375)
Proband X10 (342)
Proband X11 (260)
Proband X12 (405)
Proband X13 (628)
Proband X14 (247)
Proband X15 (207)
Proband X2 (587)
Proband X4 (282)
Proband X5 (193)
Proband X6 (163)
Proband X7 (228)
Proband X8 (166)
Proband X9 (296)
sim (616)
15
10
5
0
−5
original_ngtdm_Coarseness
frame

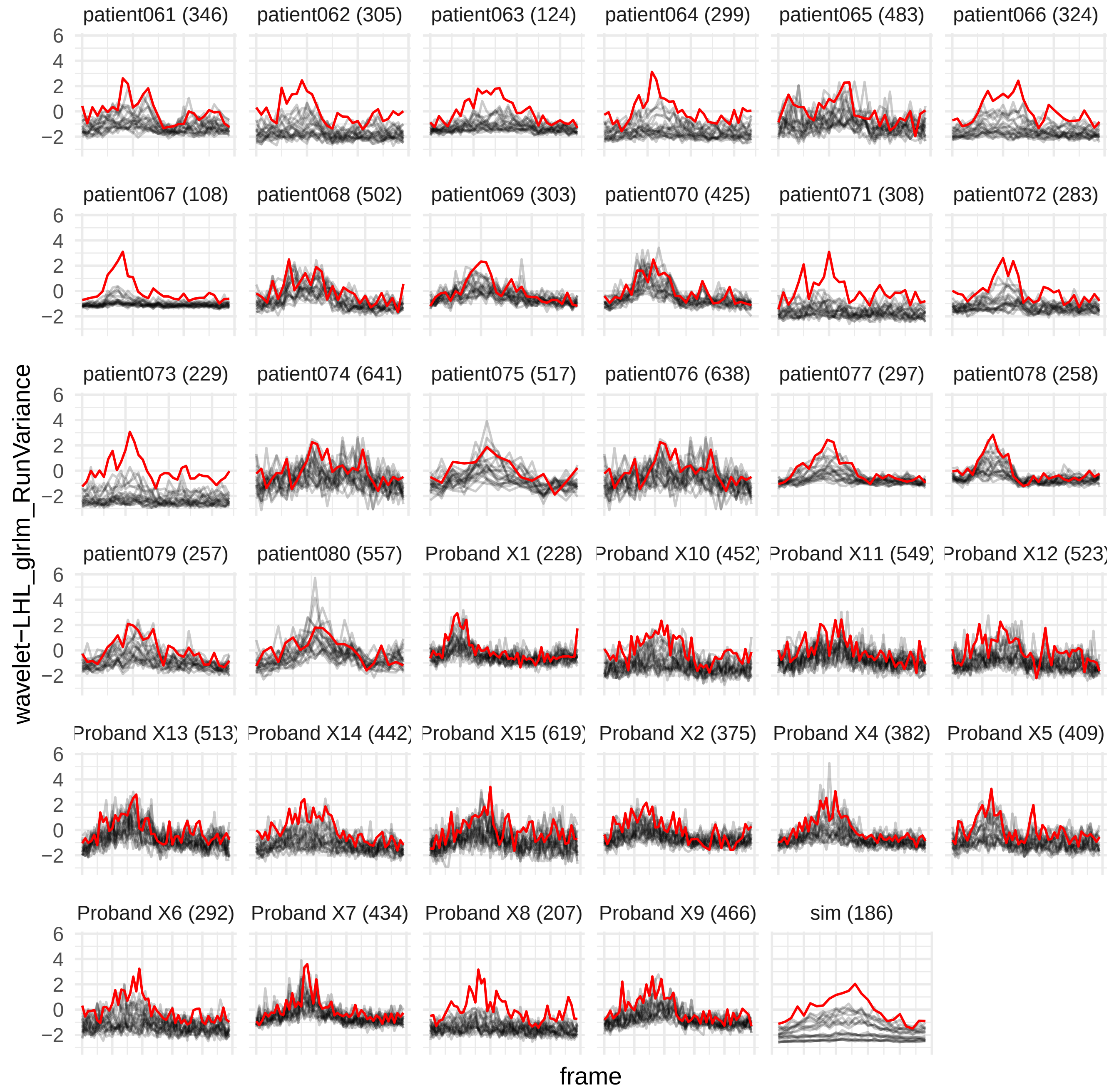

patient061 (346)
patient062 (305)
patient063 (124)
patient064 (299)
patient065 (483)
patient066 (324)
patient067 (108)
patient068 (502)
patient069 (303)
patient070 (425)
patient071 (308)
patient072 (283)
patient073 (229)
patient074 (641)
patient075 (517)
patient076 (638)
patient077 (297)
patient078 (258)
patient079 (257)
patient080 (557)
Proband X1 (228)
Proband X10 (452)
Proband X11 (549)
Proband X12 (523)
Proband X13 (513)
Proband X14 (442)
Proband X15 (619)
Proband X2 (375)
Proband X4 (382)
Proband X5 (409)
Proband X6 (292)
Proband X7 (434)
Proband X8 (207)
Proband X9 (466)
sim (186)
6
4
2
0
−2
wavelet−LHL_glrlm_RunVariance
frame

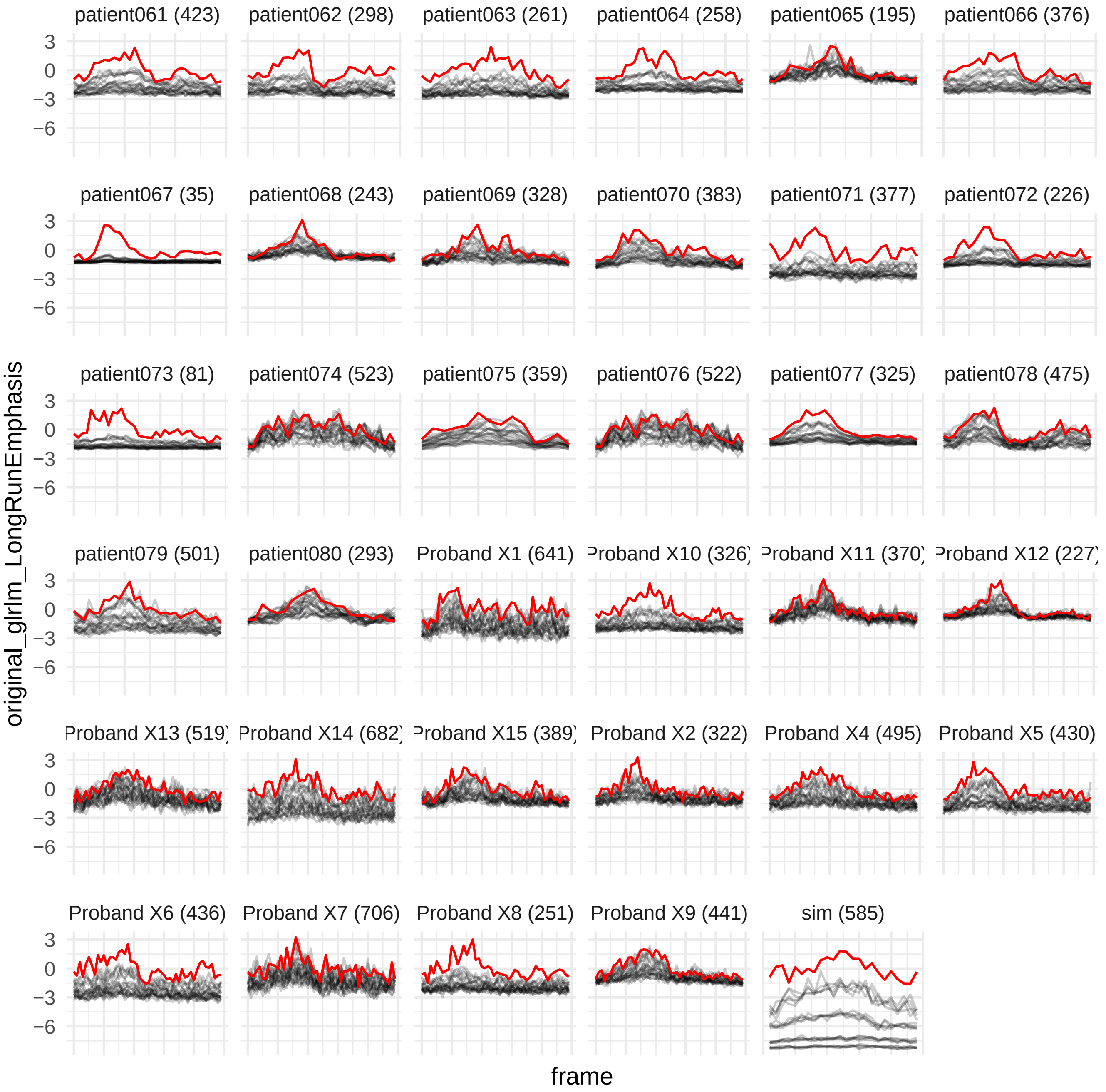
patient061 (423)
patient062 (298)
patient063 (261)
patient064 (258)
patient065 (195)
patient066 (376)
patient067 (35)
patient068 (243)
patient069 (328)
patient070 (383)
patient071 (377)
patient072 (226)
patient073 (81)
patient074 (523)
patient075 (359)
patient076 (522)
patient077 (325)
patient078 (475)
patient079 (501)
patient080 (293)
Proband X1 (641)
Proband X10 (326)
Proband X11 (370)
Proband X12 (227)
Proband X13 (519)
Proband X14 (682)
Proband X15 (389)
Proband X2 (322)
Proband X4 (495)
Proband X5 (430)
Proband X6 (436)
Proband X7 (706)
Proband X8 (251)
Proband X9 (441)
sim (585)
3
0
−3
−6
original_glrlm_LongRunEmphasis
frame

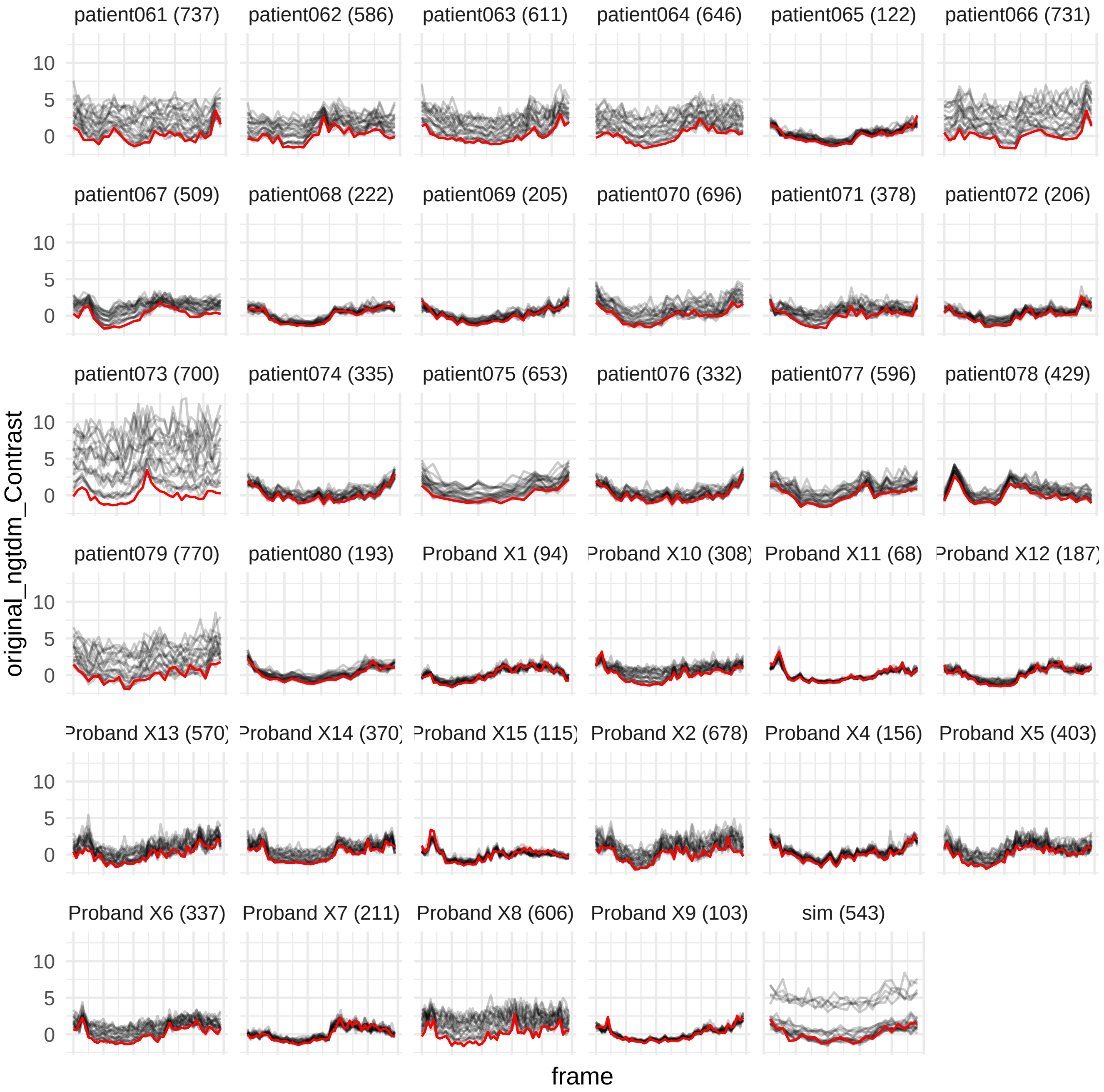
patient061 (737)
patient062 (586)
patient063 (611)
patient064 (646)
patient065 (122)
patient066 (731)
patient067 (509)
patient068 (222)
patient069 (205)
patient070 (696)
patient071 (378)
patient072 (206)
patient073 (700)
patient074 (335)
patient075 (653)
patient076 (332)
patient077 (596)
patient078 (429)
patient079 (770)
patient080 (193)
Proband X1 (94)
Proband X10 (308)
Proband X11 (68)
Proband X12 (187)
Proband X13 (570)
Proband X14 (370)
Proband X15 (115)
Proband X2 (678)
Proband X4 (156)
Proband X5 (403)
Proband X6 (337)
Proband X7 (211)
Proband X8 (606)
Proband X9 (103)
sim (543)
original_ngtdm_Contrast
10
5
0
frame

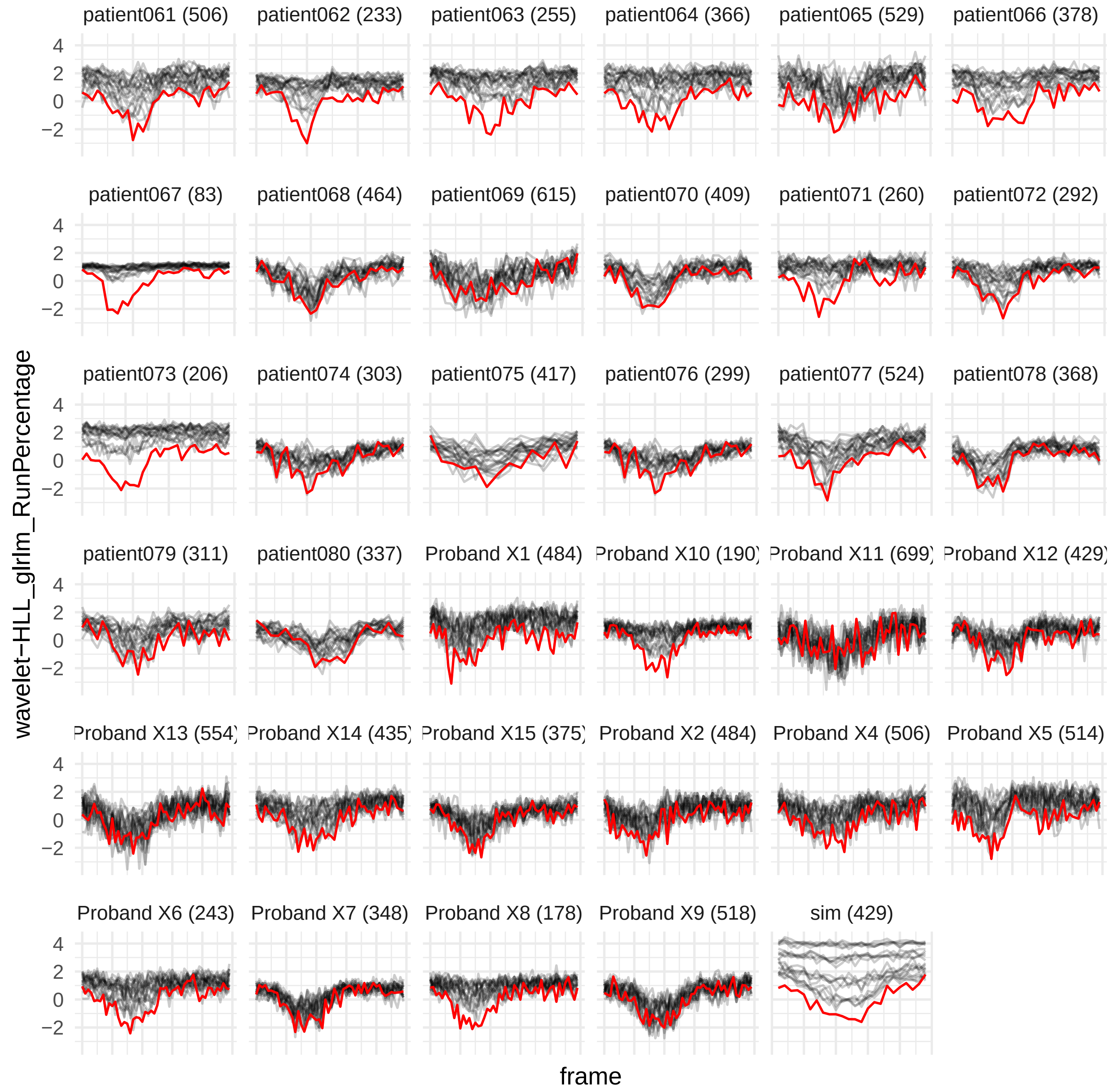
patient061 (506)
patient062 (233)
patient063 (255)
patient064 (366)
patient065 (529)
patient066 (378)
patient067 (83)
patient068 (464)
patient069 (615)
patient070 (409)
patient071 (260)
patient072 (292)
patient073 (206)
patient074 (303)
patient075 (417)
patient076 (299)
patient077 (524)
patient078 (368)
patient079 (311)
patient080 (337)
Proband X1 (484)
Proband X10 (190)
Proband X11 (699)
Proband X12 (429)
Proband X13 (554)
Proband X14 (435)
Proband X15 (375)
Proband X2 (484)
Proband X4 (506)
Proband X5 (514)
Proband X6 (243)
Proband X7 (348)
Proband X8 (178)
Proband X9 (518)
sim (429)
wavelet−HLL_glrlm_RunPercentage
frame

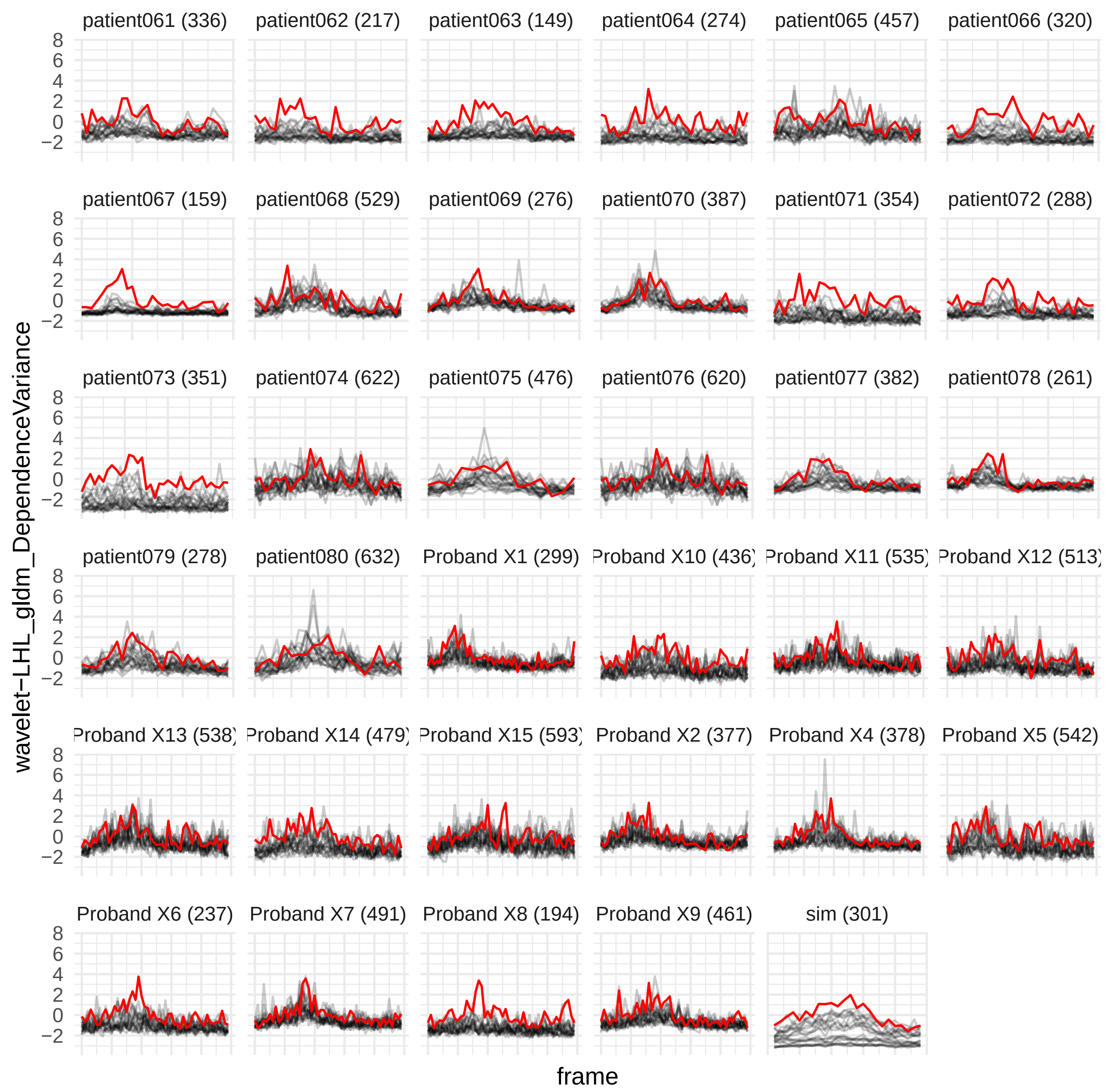
patient061 (336)
patient062 (217)
patient063 (149)
patient064 (274)
patient065 (457)
patient066 (320)
patient067 (159)
patient068 (529)
patient069 (276)
patient070 (387)
patient071 (354)
patient072 (288)
patient073 (351)
patient074 (622)
patient075 (476)
patient076 (620)
patient077 (382)
patient078 (261)
patient079 (278)
patient080 (632)
Proband X1 (299)
Proband X10 (436)
Proband X11 (535)
Proband X12 (513)
Proband X13 (538)
Proband X14 (479)
Proband X15 (593)
Proband X2 (377)
Proband X4 (378)
Proband X5 (542)
Proband X6 (237)
Proband X7 (491)
Proband X8 (194)
Proband X9 (461)
sim (301)
8
6
4
2
0
−2
wavelet−LHL_gldm_DependenceVariance
frame

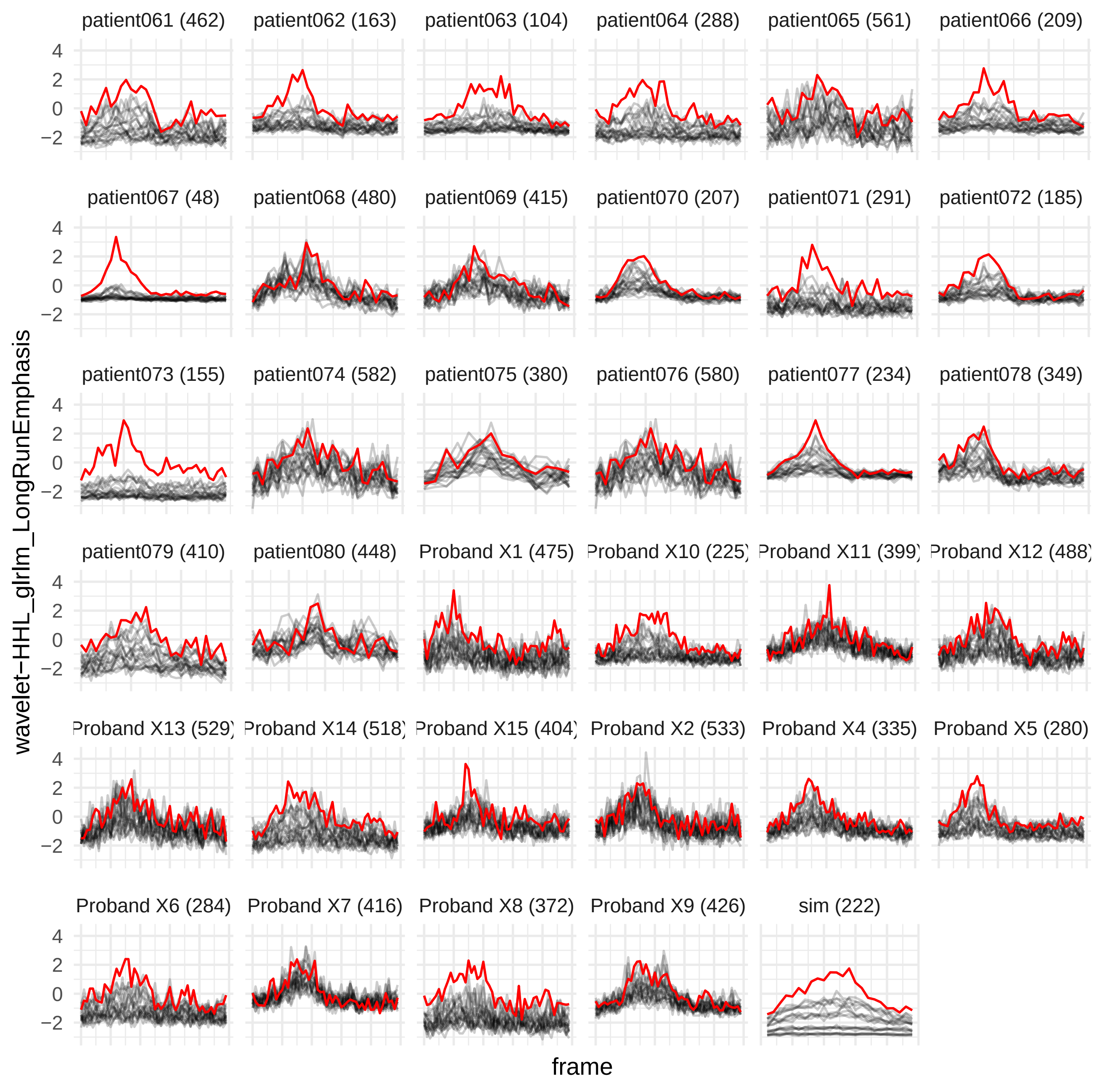
patient061 (462)
patient062 (163)
patient063 (104)
patient064 (288)
patient065 (561)
patient066 (209)
patient067 (48)
patient068 (480)
patient069 (415)
patient070 (207)
patient071 (291)
patient072 (185)
patient073 (155)
patient074 (582)
patient075 (380)
patient076 (580)
patient077 (234)
patient078 (349)
patient079 (410)
patient080 (448)
Proband X1 (475)
Proband X10 (225)
Proband X11 (399)
Proband X12 (488)
Proband X13 (529)
Proband X14 (518)
Proband X15 (404)
Proband X2 (533)
Proband X4 (335)
Proband X5 (280)
Proband X6 (284)
Proband X7 (416)
Proband X8 (372)
Proband X9 (426)
sim (222)
4
2
0
−2
wavelet−HHL_glrlm_LongRunEmphasis
frame

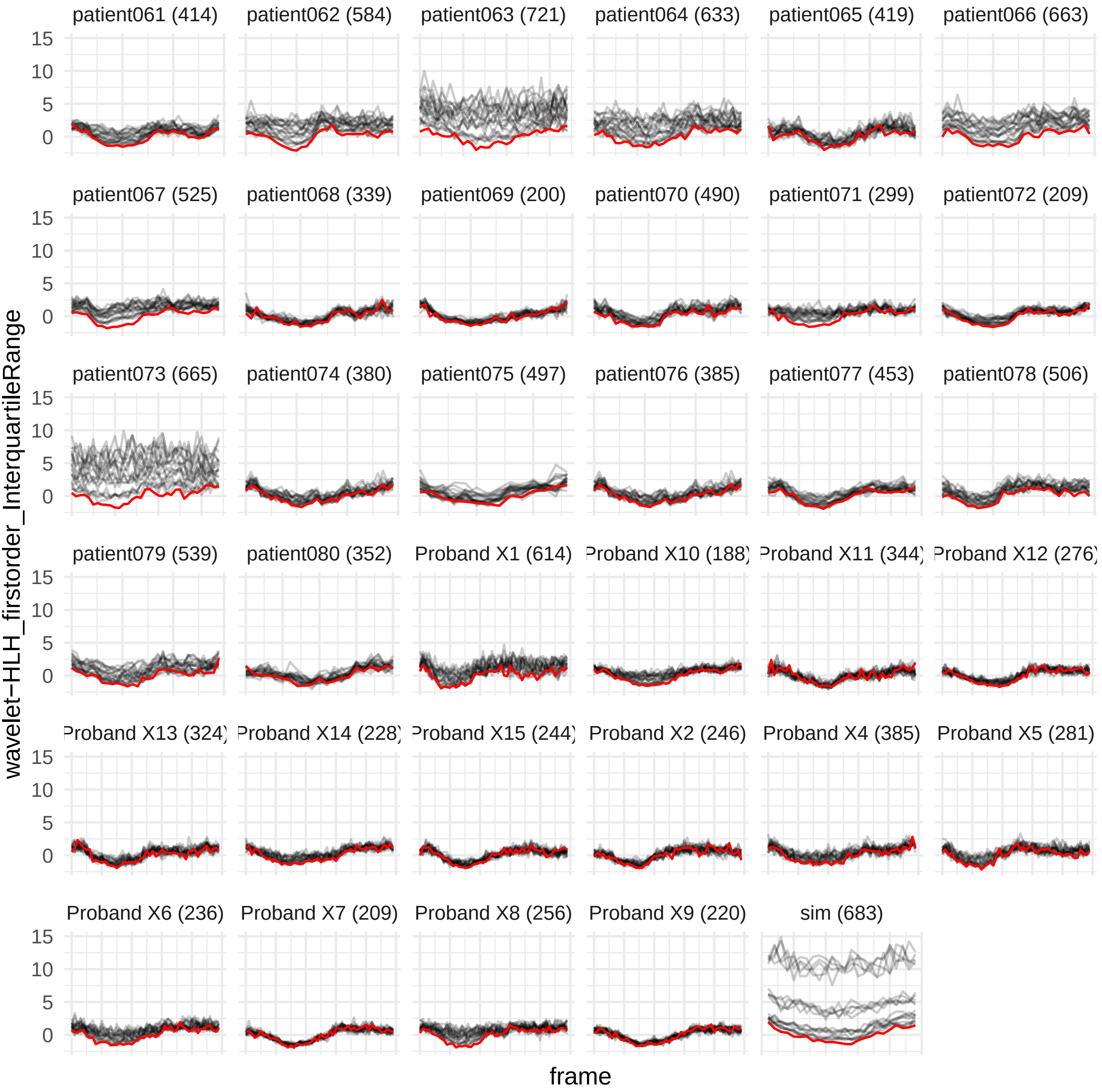

patient061 (414)
patient062 (584)
patient063 (721)
patient064 (633)
patient065 (419)
patient066 (663)
patient067 (525)
patient068 (339)
patient069 (200)
patient070 (490)
patient071 (299)
patient072 (209)
patient073 (665)
patient074 (380)
patient075 (497)
patient076 (385)
patient077 (453)
patient078 (506)
patient079 (539)
patient080 (352)
Proband X1 (614)
Proband X10 (188)
Proband X11 (344)
Proband X12 (276)
Proband X13 (324)
Proband X14 (228)
Proband X15 (244)
Proband X2 (246)
Proband X4 (385)
Proband X5 (281)
Proband X6 (236)
Proband X7 (209)
Proband X8 (256)
Proband X9 (220)
sim (683)
15
10
5
0
wavelet−HLH_firstorder_InterquartileRange
frame

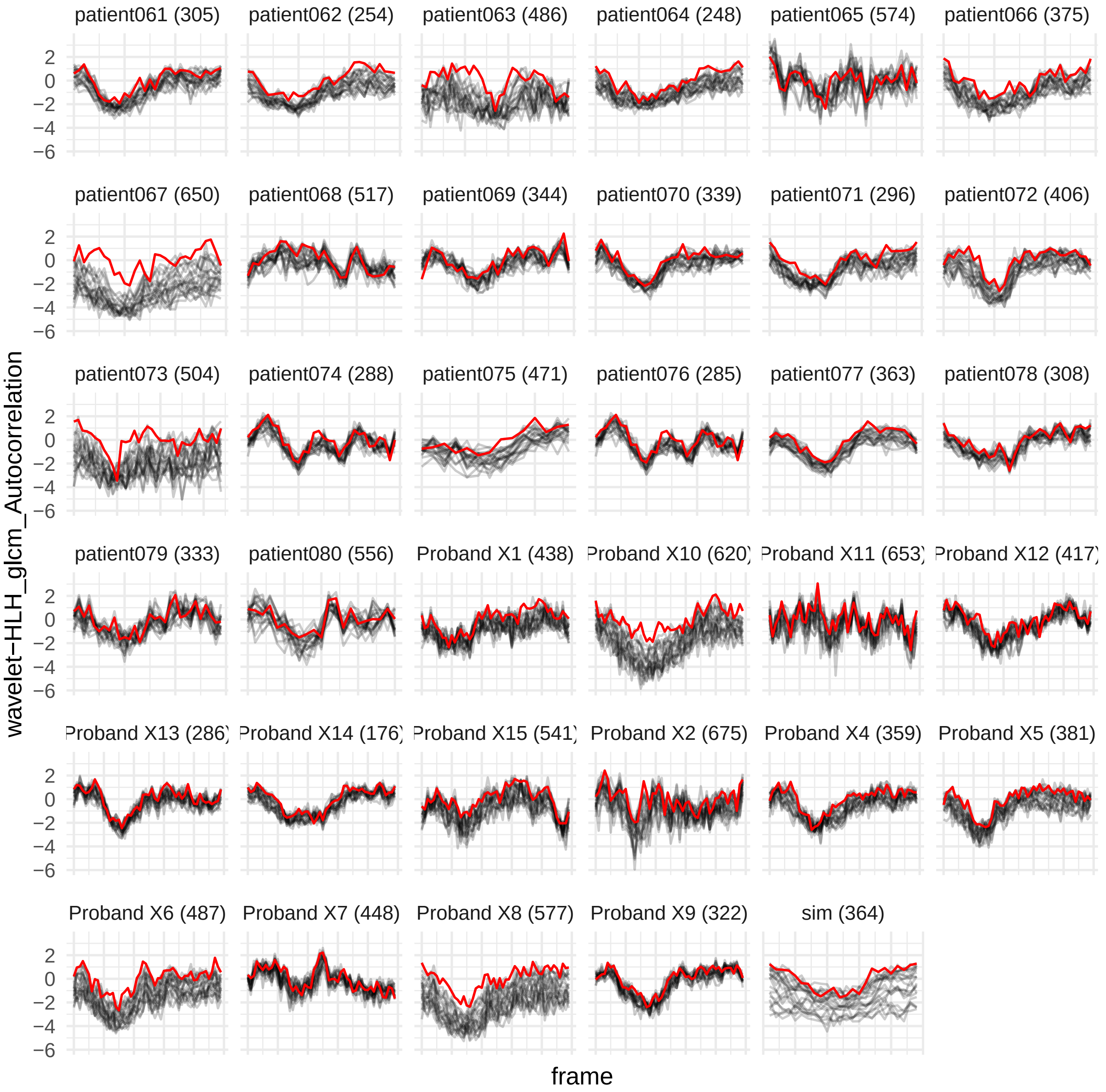

patient061 (305)
patient062 (254)
patient063 (486)
patient064 (248)
patient065 (574)
patient066 (375)
patient067 (650)
patient068 (517)
patient069 (344)
patient070 (339)
patient071 (296)
patient072 (406)
patient073 (504)
patient074 (288)
patient075 (471)
patient076 (285)
patient077 (363)
patient078 (308)
patient079 (333)
patient080 (556)
Proband X1 (438)
Proband X10 (620)
Proband X11 (653)
Proband X12 (417)
Proband X13 (286)
Proband X14 (176)
Proband X15 (541)
Proband X2 (675)
Proband X4 (359)
Proband X5 (381)
Proband X6 (487)
Proband X7 (448)
Proband X8 (577)
Proband X9 (322)
sim (364)
2
0
−2
−4
−6
wavelet−HLH_glcm_Autocorrelation
frame

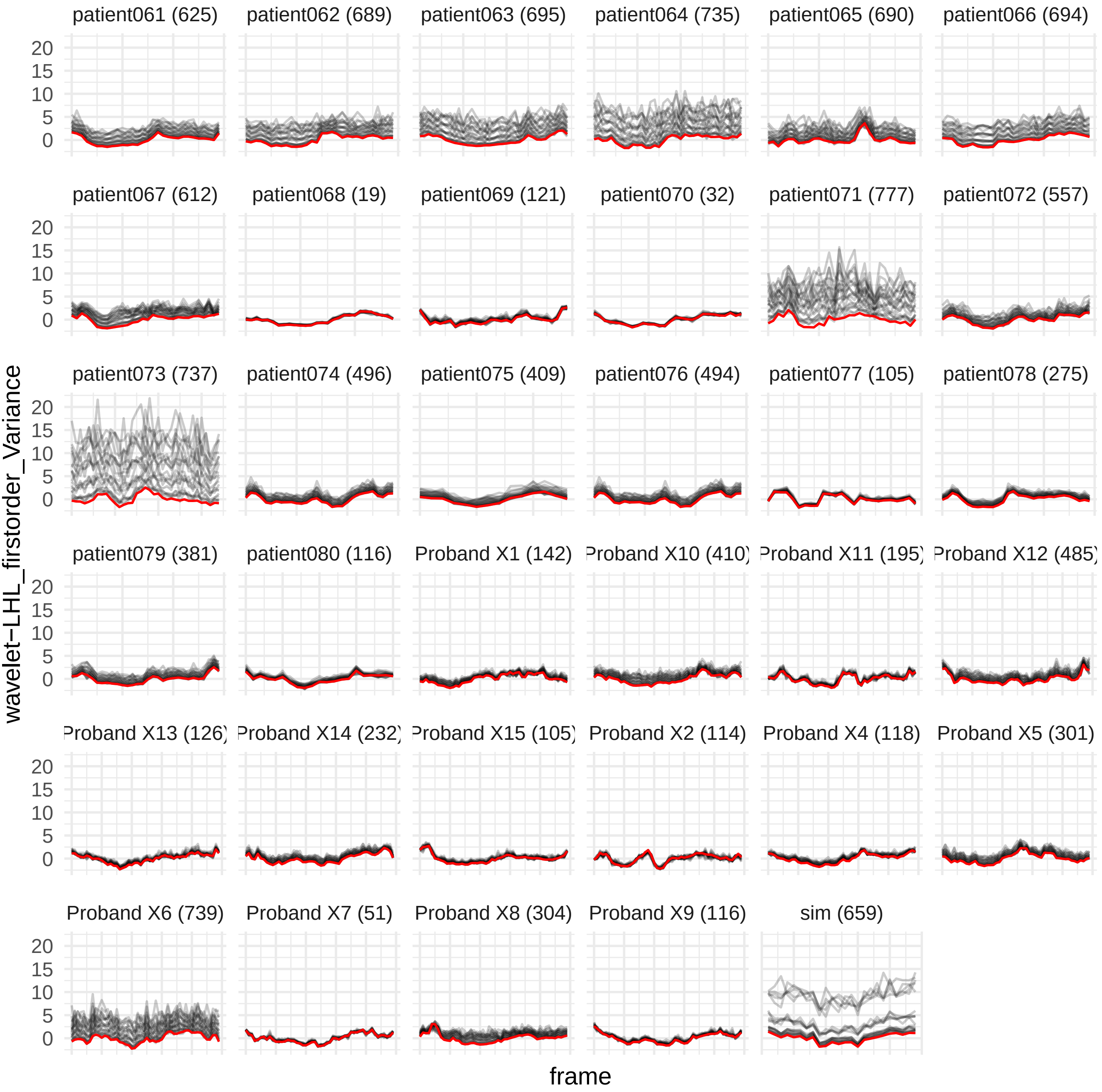

patient061 (625)
patient062 (689)
patient063 (695)
patient064 (735)
patient065 (690)
patient066 (694)
patient067 (612)
patient068 (19)
patient069 (121)
patient070 (32)
patient071 (777)
patient072 (557)
patient073 (737)
patient074 (496)
patient075 (409)
patient076 (494)
patient077 (105)
patient078 (275)
patient079 (381)
patient080 (116)
Proband X1 (142)
Proband X10 (410)
Proband X11 (195)
Proband X12 (485)
Proband X13 (126)
Proband X14 (232)
Proband X15 (105)
Proband X2 (114)
Proband X4 (118)
Proband X5 (301)
Proband X6 (739)
Proband X7 (51)
Proband X8 (304)
Proband X9 (116)
sim (659)
wavelet−LHL_firstorder_Variance
frame
0
5
10
15
20

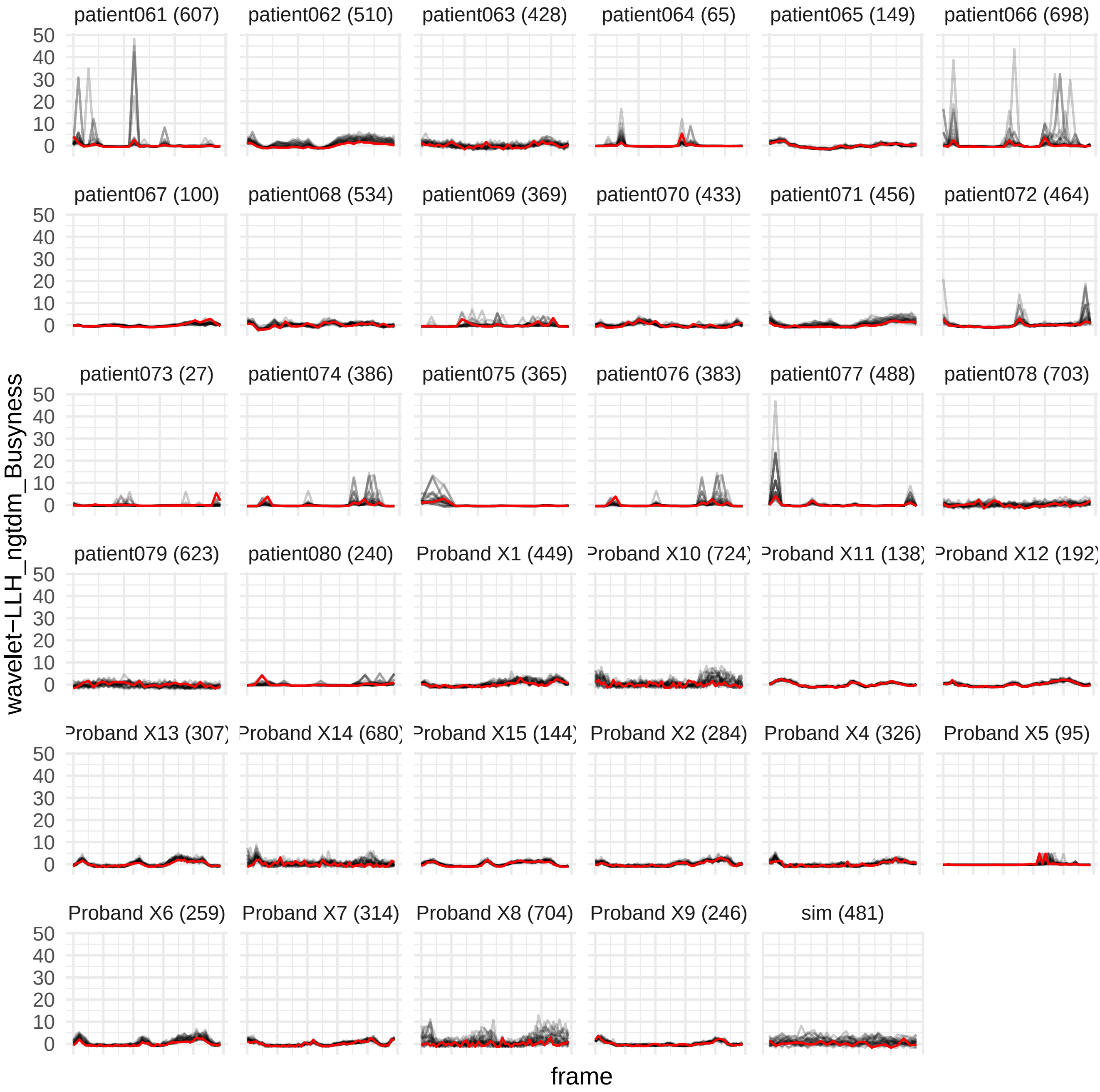
patient061 (607)
patient062 (510)
patient063 (428)
patient064 (65)
patient065 (149)
patient066 (698)
patient067 (100)
patient068 (534)
patient069 (369)
patient070 (433)
patient071 (456)
patient072 (464)
patient073 (27)
patient074 (386)
patient075 (365)
patient076 (383)
patient077 (488)
patient078 (703)
patient079 (623)
patient080 (240)
Proband X1 (449)
Proband X10 (724)
Proband X11 (138)
Proband X12 (192)
Proband X13 (307)
Proband X14 (680)
Proband X15 (144)
Proband X2 (284)
Proband X4 (326)
Proband X5 (95)
Proband X6 (259)
Proband X7 (314)
Proband X8 (704)
Proband X9 (246)
sim (481)
0
10
20
30
40
50
wavelet−LLH_ngtdm_Busyness
frame

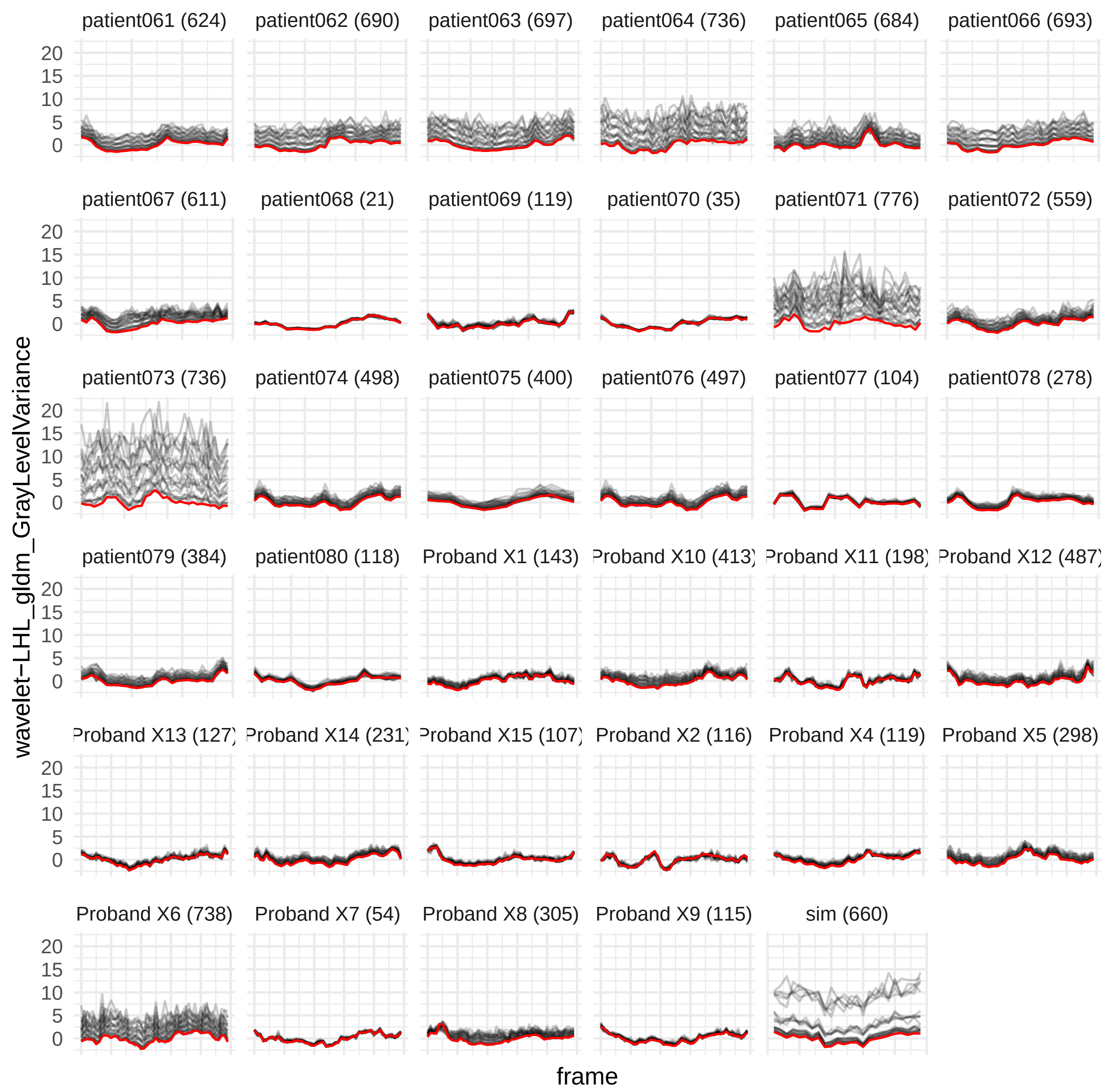
patient061 (624)
patient062 (690)
patient063 (697)
patient064 (736)
patient065 (684)
patient066 (693)
patient067 (611)
patient068 (21)
patient069 (119)
patient070 (35)
patient071 (776)
patient072 (559)
patient073 (736)
patient074 (498)
patient075 (400)
patient076 (497)
patient077 (104)
patient078 (278)
patient079 (384)
patient080 (118)
Proband X1 (143)
Proband X10 (413)
Proband X11 (198)
Proband X12 (487)
Proband X13 (127)
Proband X14 (231)
Proband X15 (107)
Proband X2 (116)
Proband X4 (119)
Proband X5 (298)
Proband X6 (738)
Proband X7 (54)
Proband X8 (305)
Proband X9 (115)
sim (660)
wavelet−LHL_gldm_GrayLevelVariance
20
15
10
5
0
frame

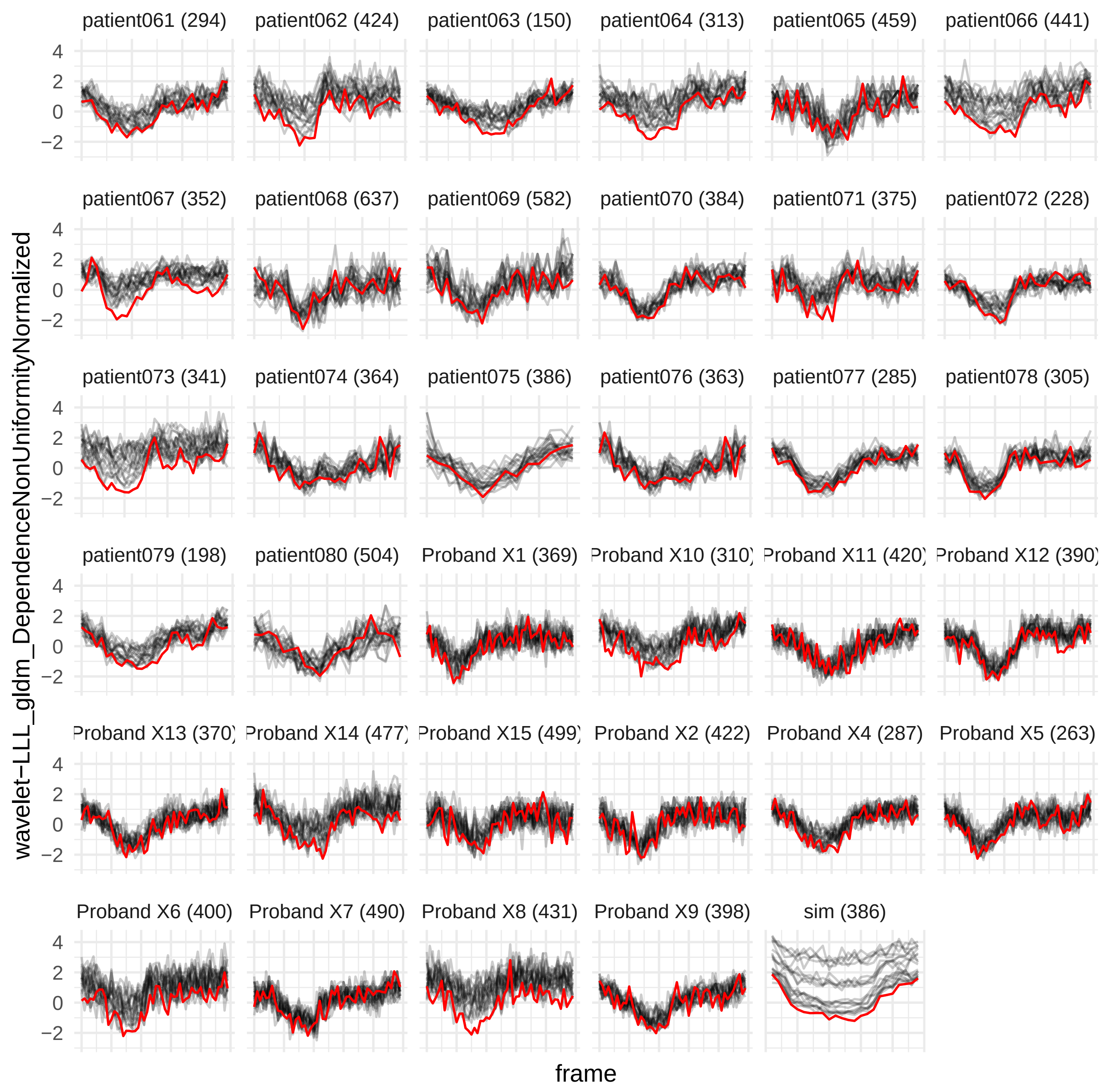
patient061 (294)
patient062 (424)
patient063 (150)
patient064 (313)
patient065 (459)
patient066 (441)
patient067 (352)
patient068 (637)
patient069 (582)
patient070 (384)
patient071 (375)
patient072 (228)
patient073 (341)
patient074 (364)
patient075 (386)
patient076 (363)
patient077 (285)
patient078 (305)
patient079 (198)
patient080 (504)
Proband X1 (369)
Proband X10 (310)
Proband X11 (420)
Proband X12 (390)
Proband X13 (370)
Proband X14 (477)
Proband X15 (499)
Proband X2 (422)
Proband X4 (287)
Proband X5 (263)
Proband X6 (400)
Proband X7 (490)
Proband X8 (431)
Proband X9 (398)
sim (386)
4
2
0
−2
wavelet−LLL_gldm_DependenceNonUniformityNormalized
frame

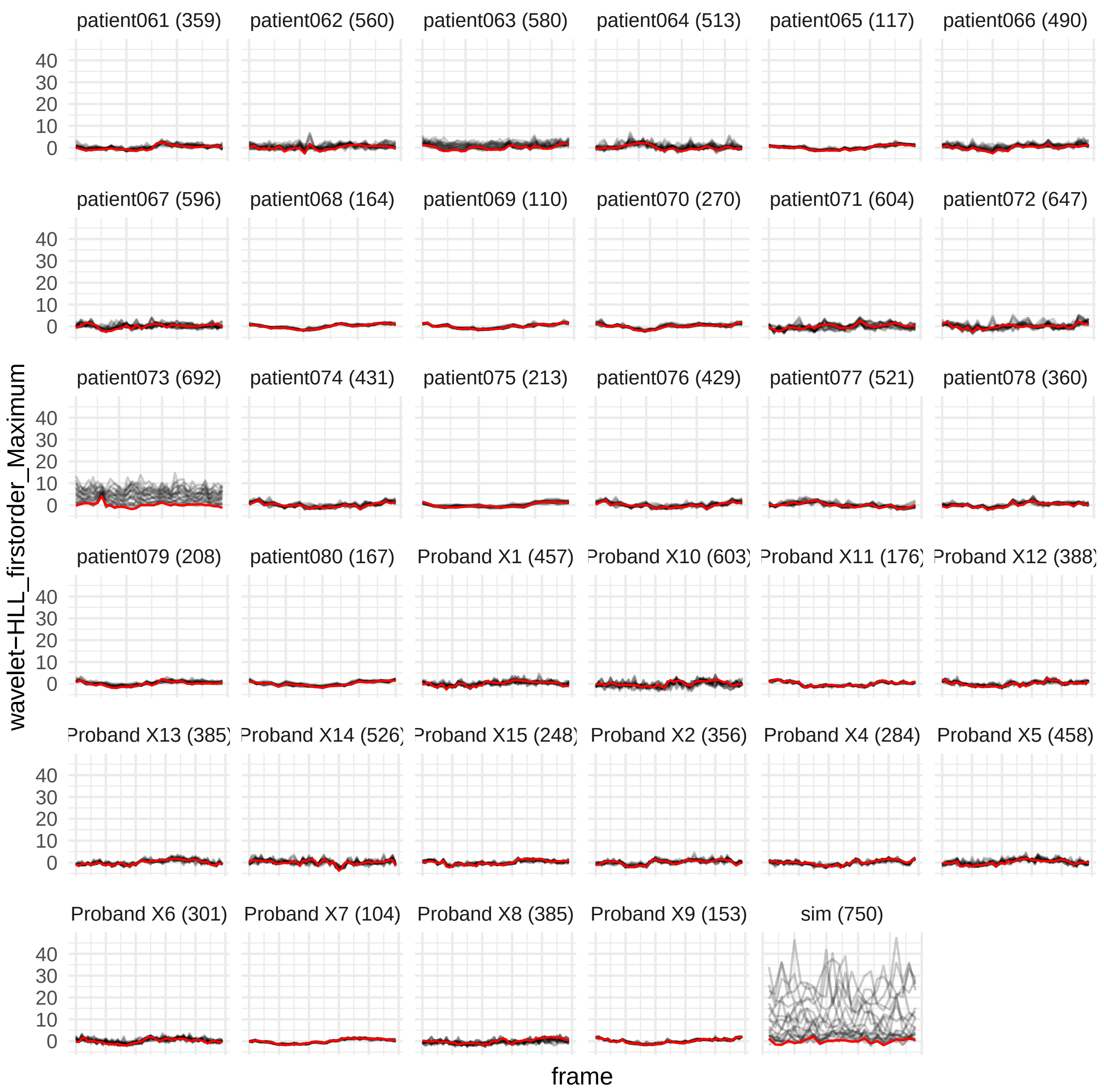
patient061 (359)
patient062 (560)
patient063 (580)
patient064 (513)
patient065 (117)
patient066 (490)
patient067 (596)
patient068 (164)
patient069 (110)
patient070 (270)
patient071 (604)
patient072 (647)
patient073 (692)
patient074 (431)
patient075 (213)
patient076 (429)
patient077 (521)
patient078 (360)
patient079 (208)
patient080 (167)
Proband X1 (457)
Proband X10 (603)
Proband X11 (176)
Proband X12 (388)
Proband X13 (385)
Proband X14 (526)
Proband X15 (248)
Proband X2 (356)
Proband X4 (284)
Proband X5 (458)
Proband X6 (301)
Proband X7 (104)
Proband X8 (385)
Proband X9 (153)
sim (750)
0
10
20
30
40
wavelet−HLL_firstorder_Maximum
frame

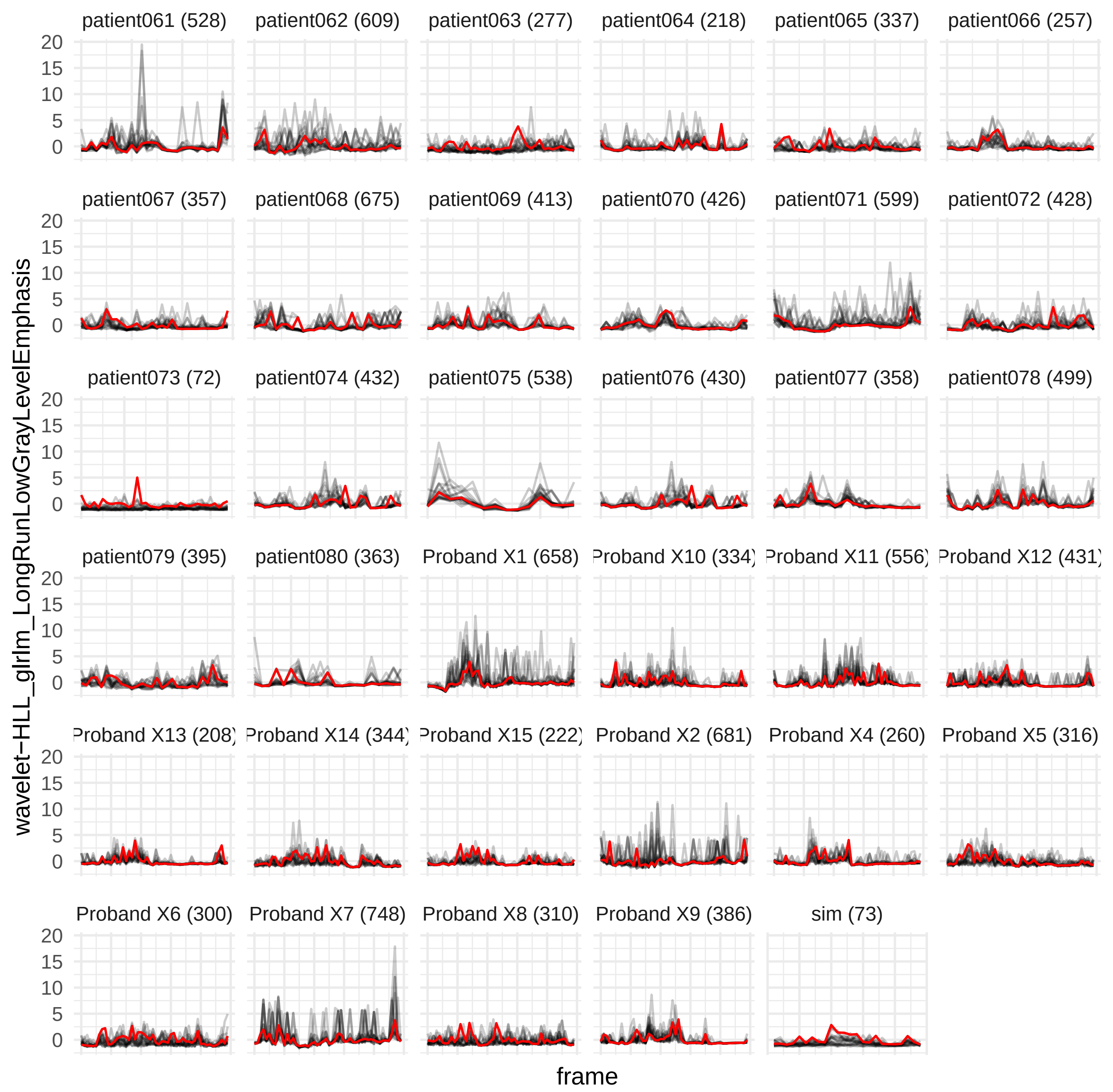
patient061 (528)
patient062 (609)
patient063 (277)
patient064 (218)
patient065 (337)
patient066 (257)
patient067 (357)
patient068 (675)
patient069 (413)
patient070 (426)
patient071 (599)
patient072 (428)
patient073 (72)
patient074 (432)
patient075 (538)
patient076 (430)
patient077 (358)
patient078 (499)
patient079 (395)
patient080 (363)
Proband X1 (658)
Proband X10 (334)
Proband X11 (556)
Proband X12 (431)
Proband X13 (208)
Proband X14 (344)
Proband X15 (222)
Proband X2 (681)
Proband X4 (260)
Proband X5 (316)
Proband X6 (300)
Proband X7 (748)
Proband X8 (310)
Proband X9 (386)
sim (73)
0
5
10
15
20
wavelet−HLL_glrlm_LongRunLowGrayLevelEmphasis
frame

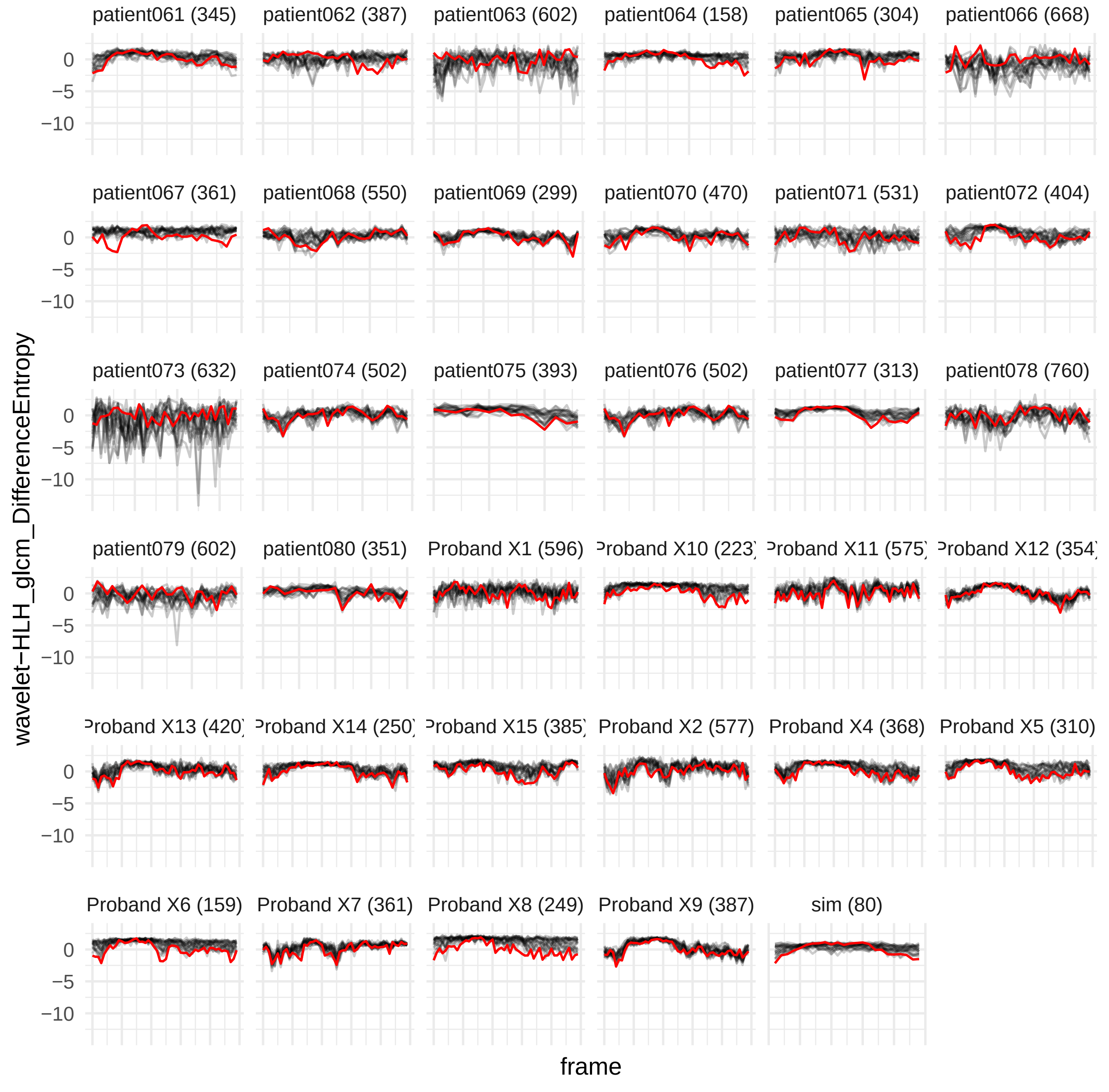
patient061 (345)
patient062 (387)
patient063 (602)
patient064 (158)
patient065 (304)
patient066 (668)
patient067 (361)
patient068 (550)
patient069 (299)
patient070 (470)
patient071 (531)
patient072 (404)
patient073 (632)
patient074 (502)
patient075 (393)
patient076 (502)
patient077 (313)
patient078 (760)
patient079 (602)
patient080 (351)
Proband X1 (596)
Proband X10 (223)
Proband X11 (575)
Proband X12 (354)
Proband X13 (420)
Proband X14 (250)
Proband X15 (385)
Proband X2 (577)
Proband X4 (368)
Proband X5 (310)
Proband X6 (159)
Proband X7 (361)
Proband X8 (249)
Proband X9 (387)
sim (80)
0
−5
−10
wavelet−HLH_glcm_DifferenceEntropy
frame

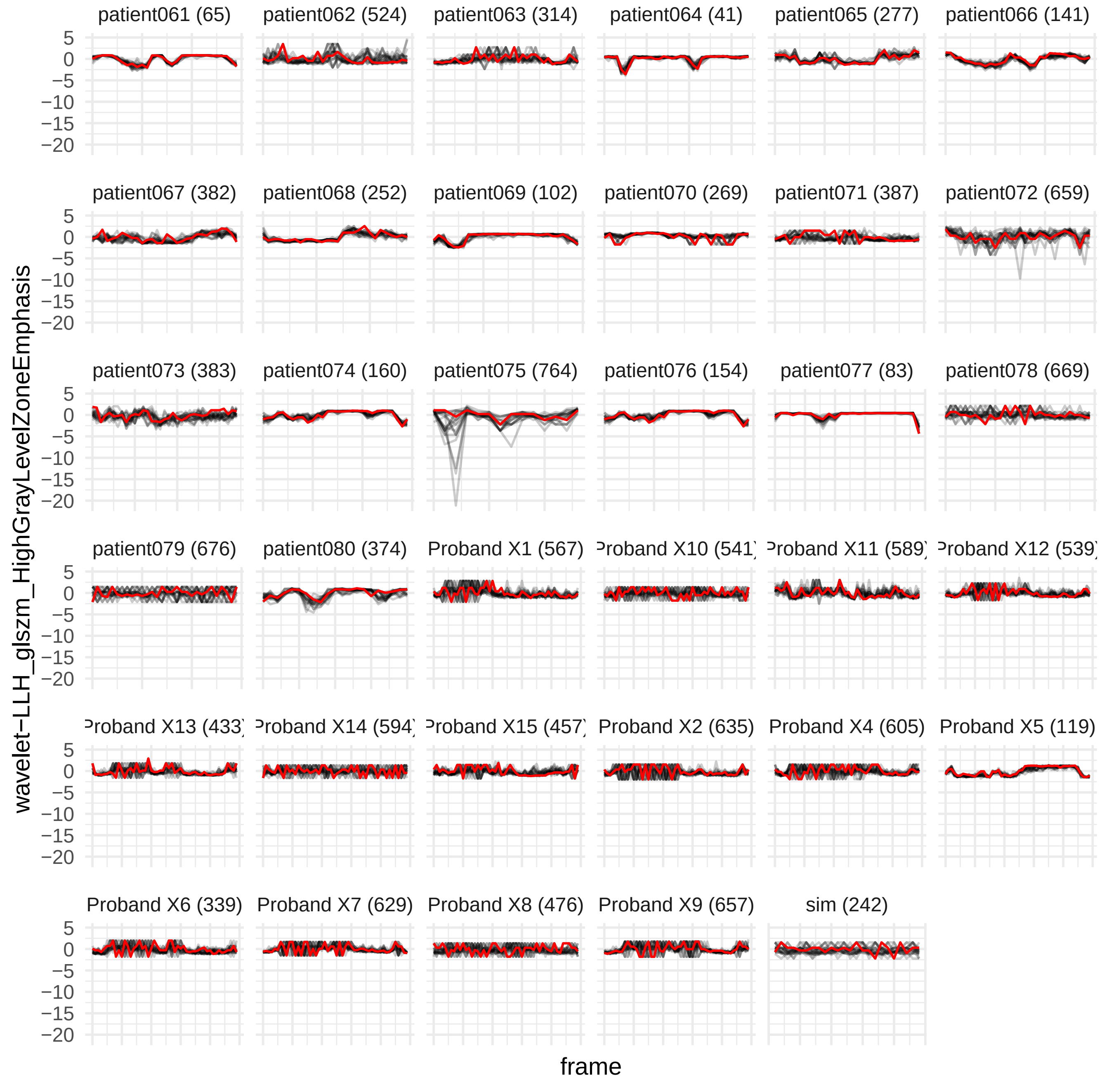

patient061 (65)
patient062 (524)
patient063 (314)
patient064 (41)
patient065 (277)
patient066 (141)
patient067 (382)
patient068 (252)
patient069 (102)
patient070 (269)
patient071 (387)
patient072 (659)
patient073 (383)
patient074 (160)
patient075 (764)
patient076 (154)
patient077 (83)
patient078 (669)
patient079 (676)
patient080 (374)
Proband X1 (567)
Proband X10 (541)
Proband X11 (589)
Proband X12 (539)
Proband X13 (433)
Proband X14 (594)
Proband X15 (457)
Proband X2 (635)
Proband X4 (605)
Proband X5 (119)
Proband X6 (339)
Proband X7 (629)
Proband X8 (476)
Proband X9 (657)
sim (242)
5
0
−5
−10
−15
−20
wavelet−LLH_glszm_HighGrayLevelZoneEmphasis
frame

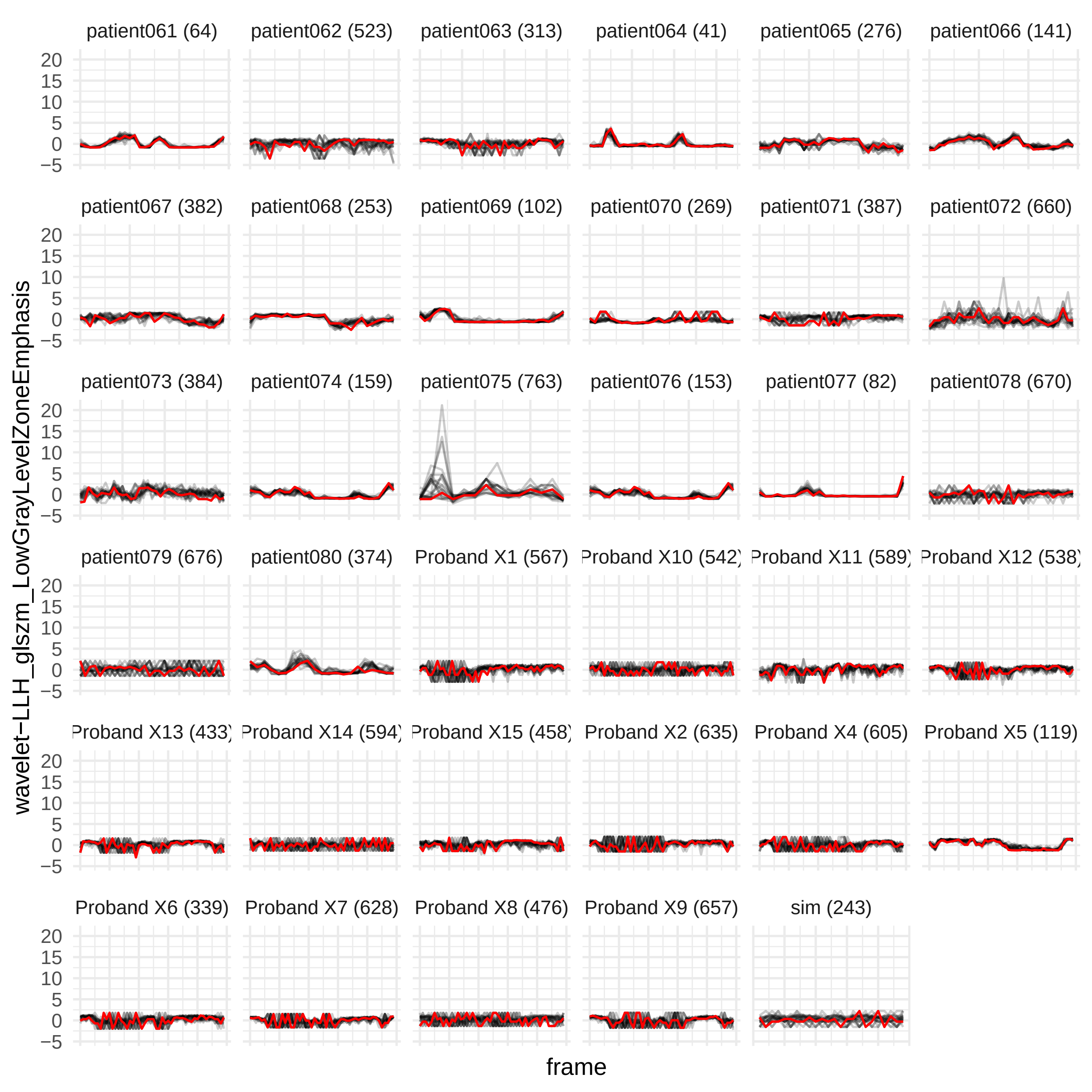
patient061 (64)
patient062 (523)
patient063 (313)
patient064 (41)
patient065 (276)
patient066 (141)
patient067 (382)
patient068 (253)
patient069 (102)
patient070 (269)
patient071 (387)
patient072 (660)
patient073 (384)
patient074 (159)
patient075 (763)
patient076 (153)
patient077 (82)
patient078 (670)
patient079 (676)
patient080 (374)
Proband X1 (567)
Proband X10 (542)
Proband X11 (589)
Proband X12 (538)
Proband X13 (433)
Proband X14 (594)
Proband X15 (458)
Proband X2 (635)
Proband X4 (605)
Proband X5 (119)
Proband X6 (339)
Proband X7 (628)
Proband X8 (476)
Proband X9 (657)
sim (243)
20
15
10
5
0
−5
wavelet−LLH_glszm_LowGrayLevelZoneEmphasis
frame

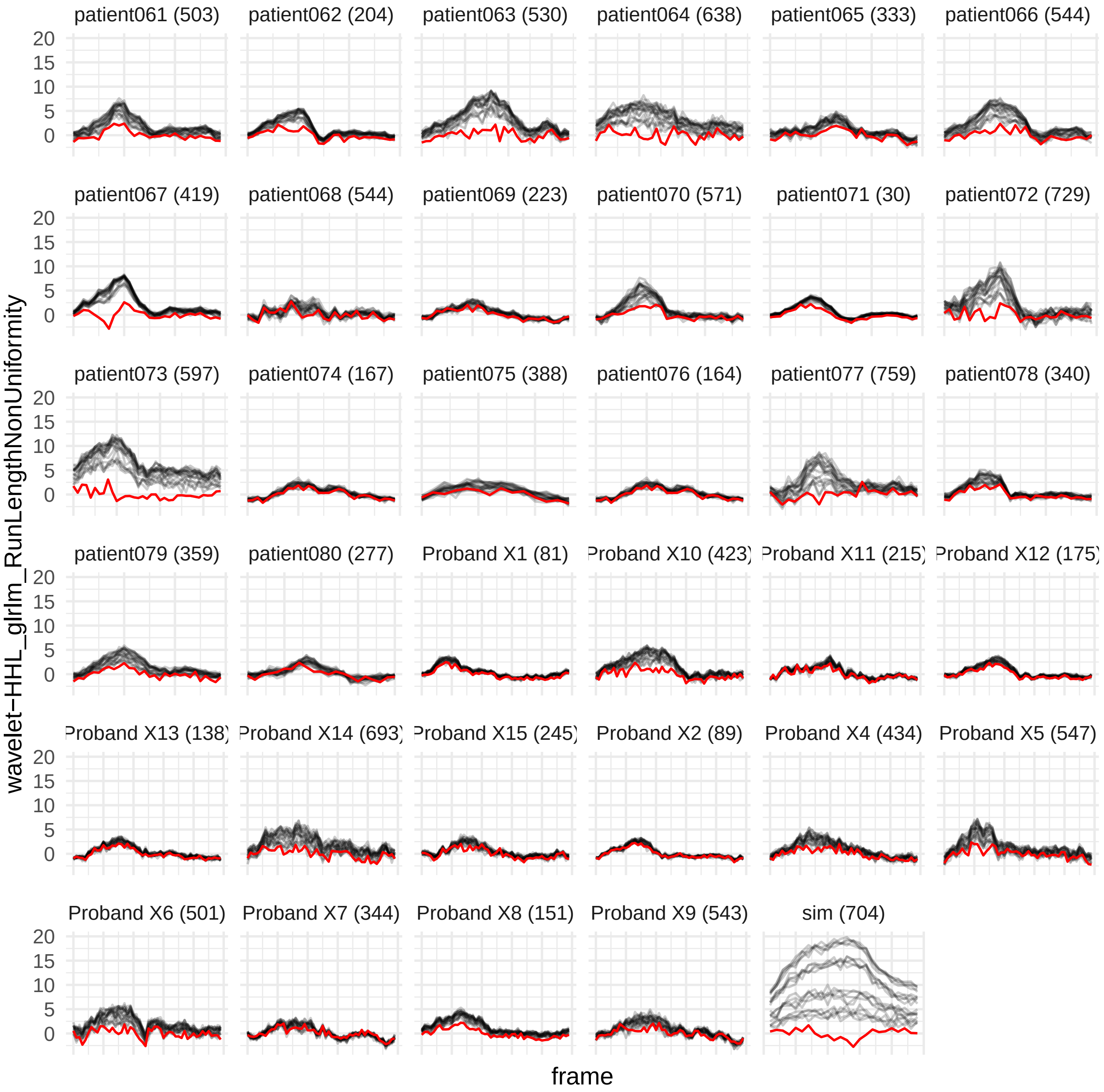

patient061 (503)
patient062 (204)
patient063 (530)
patient064 (638)
patient065 (333)
patient066 (544)
patient067 (419)
patient068 (544)
patient069 (223)
patient070 (571)
patient071 (30)
patient072 (729)
patient073 (597)
patient074 (167)
patient075 (388)
patient076 (164)
patient077 (759)
patient078 (340)
patient079 (359)
patient080 (277)
Proband X1 (81)
Proband X10 (423)
Proband X11 (215)
Proband X12 (175)
Proband X13 (138)
Proband X14 (693)
Proband X15 (245)
Proband X2 (89)
Proband X4 (434)
Proband X5 (547)
Proband X6 (501)
Proband X7 (344)
Proband X8 (151)
Proband X9 (543)
sim (704)
20
15
10
5
0
wavelet−HHL_glrlm_RunLengthNonUniformity
frame

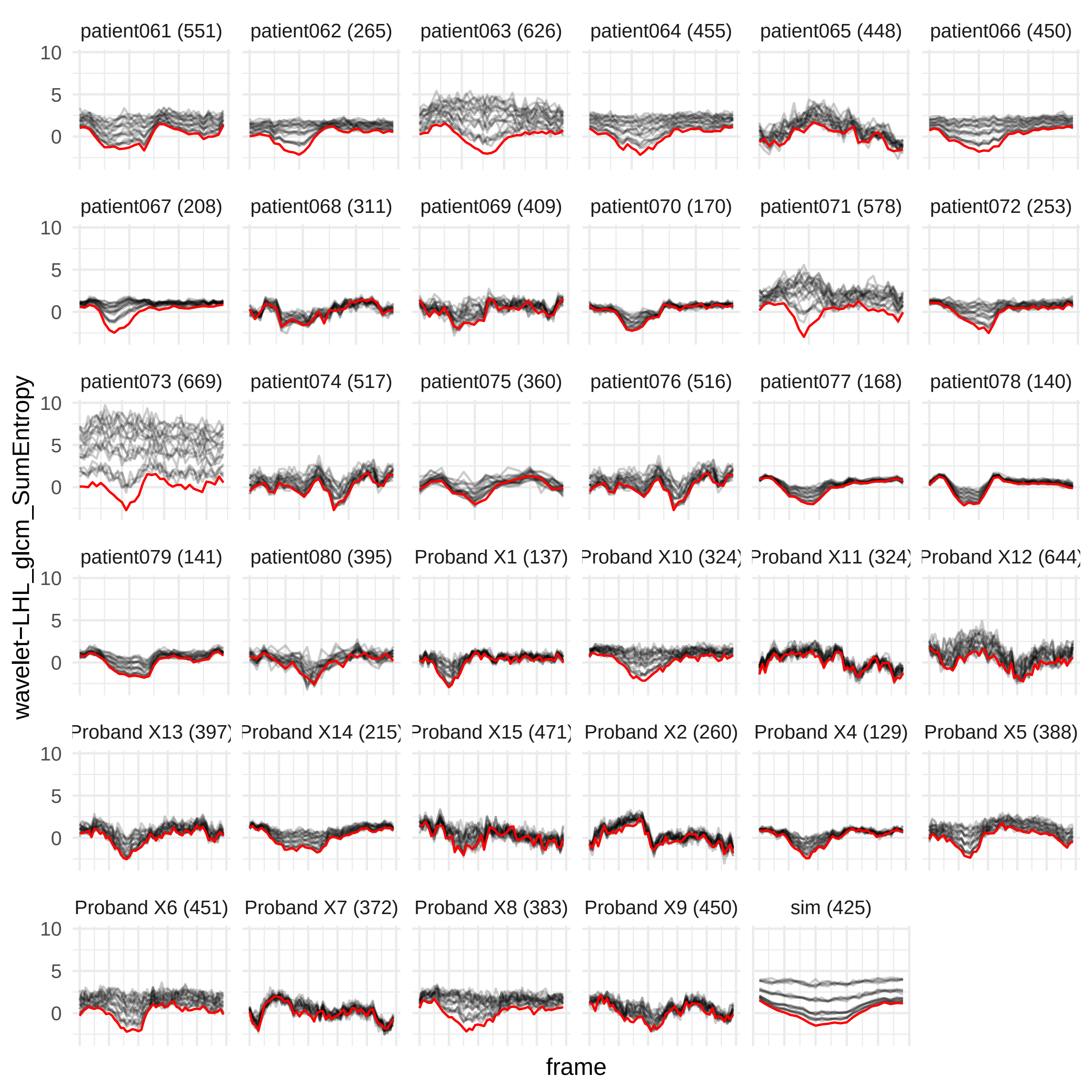
patient061 (551)
patient062 (265)
patient063 (626)
patient064 (455)
patient065 (448)
patient066 (450)
patient067 (208)
patient068 (311)
patient069 (409)
patient070 (170)
patient071 (578)
patient072 (253)
patient073 (669)
patient074 (517)
patient075 (360)
patient076 (516)
patient077 (168)
patient078 (140)
patient079 (141)
patient080 (395)
Proband X1 (137)
Proband X10 (324)
Proband X11 (324)
Proband X12 (644)
Proband X13 (397)
Proband X14 (215)
Proband X15 (471)
Proband X2 (260)
Proband X4 (129)
Proband X5 (388)
Proband X6 (451)
Proband X7 (372)
Proband X8 (383)
Proband X9 (450)
sim (425)
10
5
0
wavelet−LHL_glcm_SumEntropy
frame

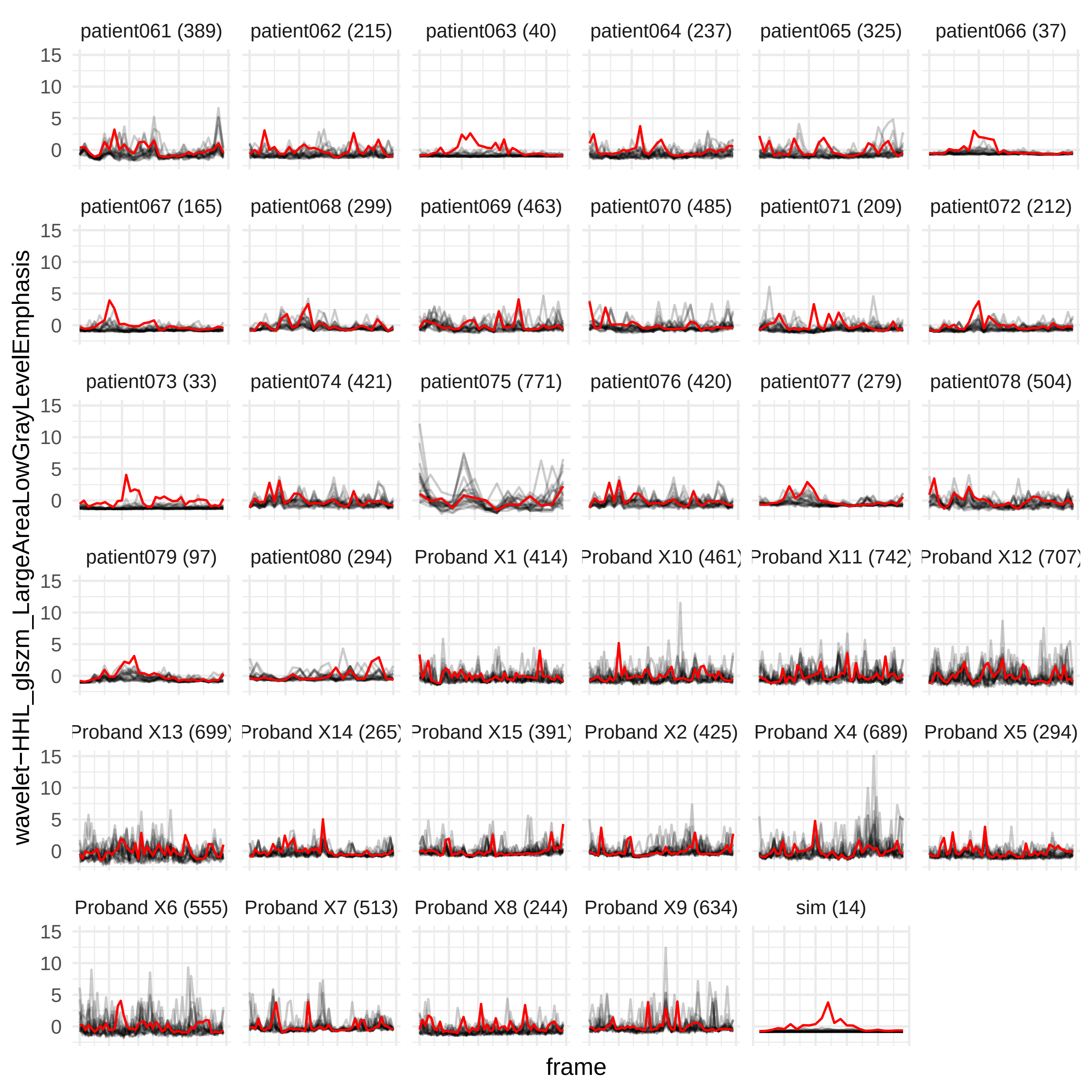
patient061 (389)
patient062 (215)
patient063 (40)
patient064 (237)
patient065 (325)
patient066 (37)
patient067 (165)
patient068 (299)
patient069 (463)
patient070 (485)
patient071 (209)
patient072 (212)
patient073 (33)
patient074 (421)
patient075 (771)
patient076 (420)
patient077 (279)
patient078 (504)
patient079 (97)
patient080 (294)
Proband X1 (414)
Proband X10 (461)
Proband X11 (742)
Proband X12 (707)
Proband X13 (699)
Proband X14 (265)
Proband X15 (391)
Proband X2 (425)
Proband X4 (689)
Proband X5 (294)
Proband X6 (555)
Proband X7 (513)
Proband X8 (244)
Proband X9 (634)
sim (14)
wavelet−HHL_glszm_LargeAreaLowGrayLevelEmphasis
frame
0
5
10
15

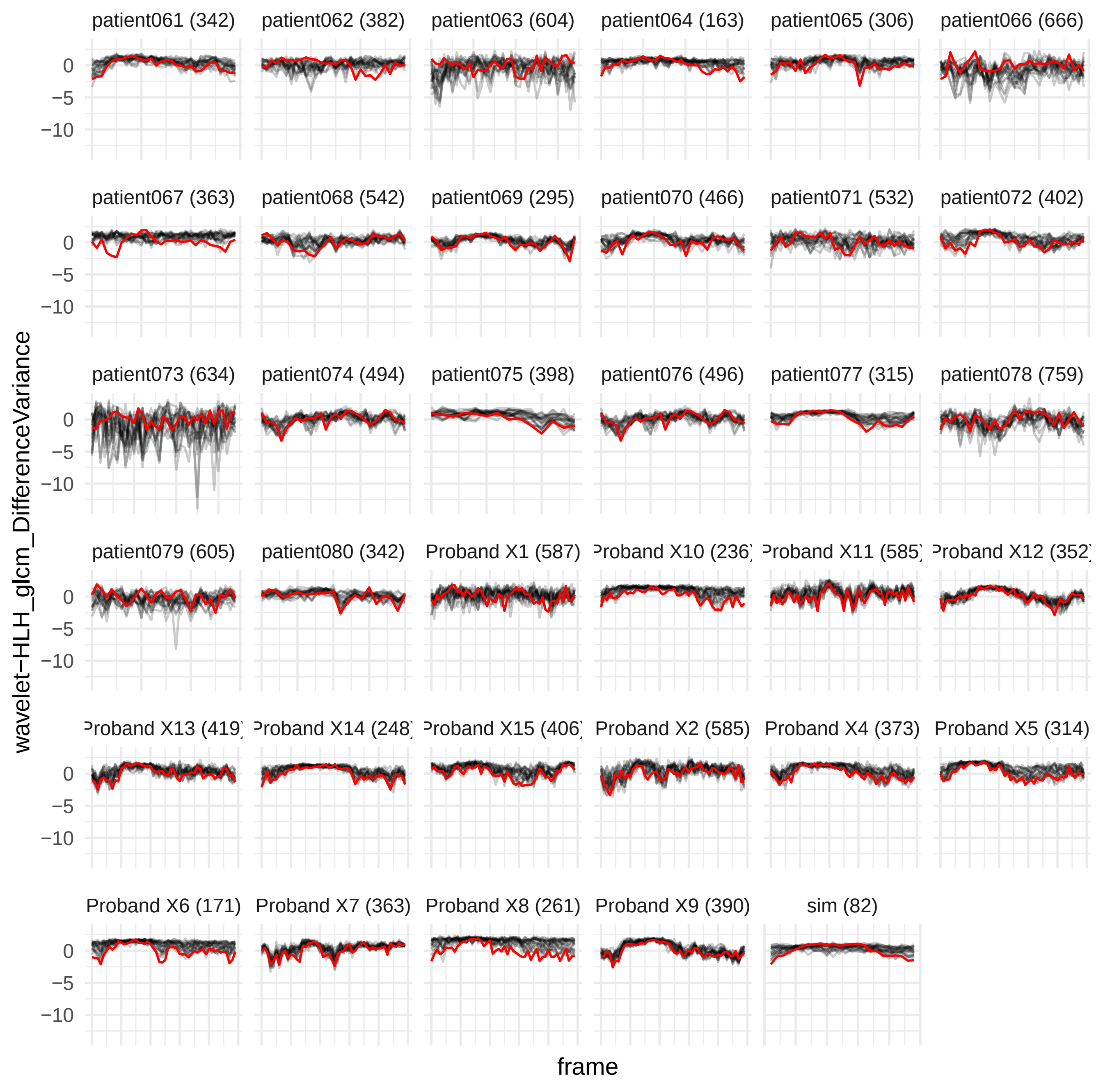

patient061 (342)
patient062 (382)
patient063 (604)
patient064 (163)
patient065 (306)
patient066 (666)
patient067 (363)
patient068 (542)
patient069 (295)
patient070 (466)
patient071 (532)
patient072 (402)
patient073 (634)
patient074 (494)
patient075 (398)
patient076 (496)
patient077 (315)
patient078 (759)
patient079 (605)
patient080 (342)
Proband X1 (587)
Proband X10 (236)
Proband X11 (585)
Proband X12 (352)
Proband X13 (419)
Proband X14 (248)
Proband X15 (406)
Proband X2 (585)
Proband X4 (373)
Proband X5 (314)
Proband X6 (171)
Proband X7 (363)
Proband X8 (261)
Proband X9 (390)
sim (82)
0
−5
−10
wavelet−HLH_glcm_DifferenceVariance
frame

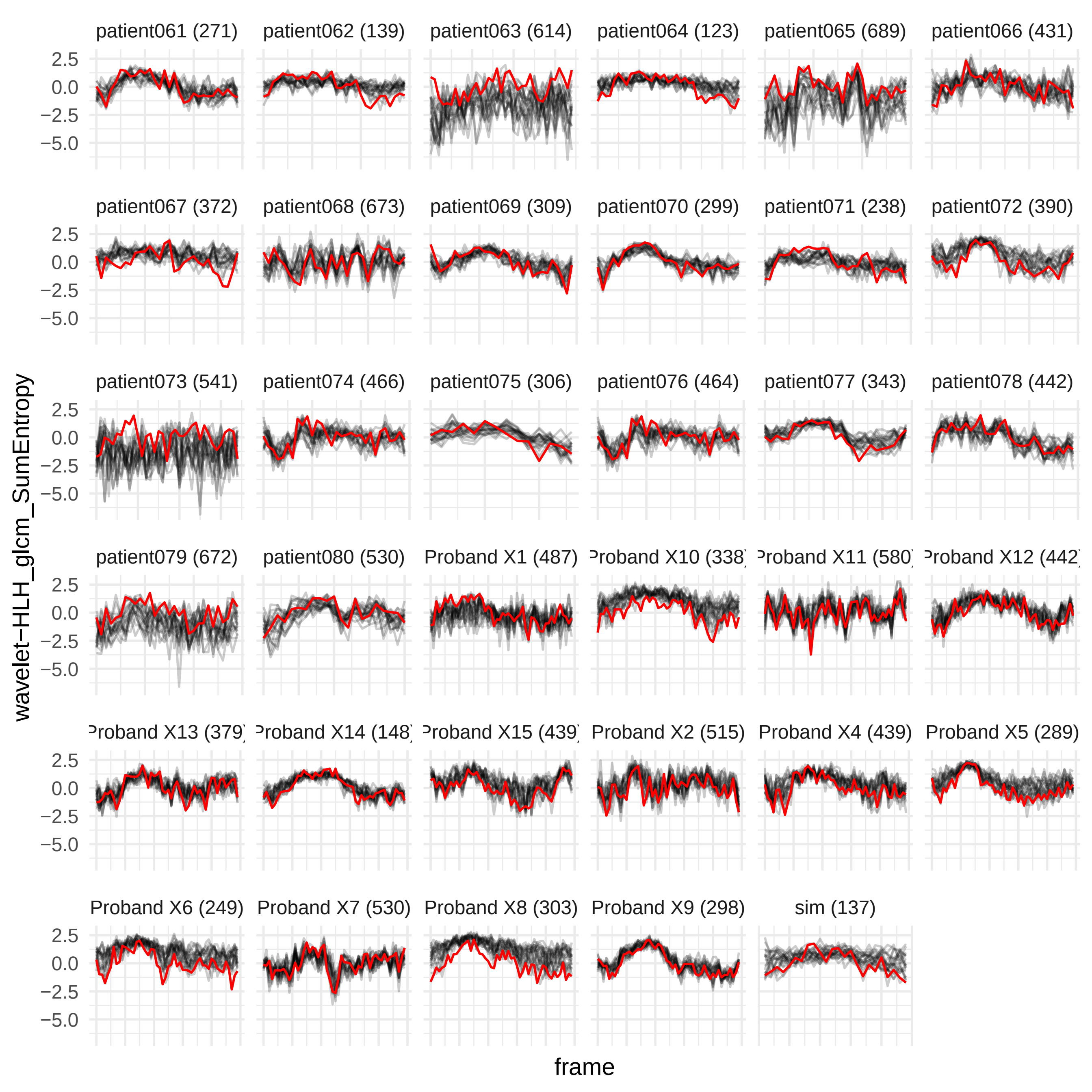
patient061 (271)
patient062 (139)
patient063 (614)
patient064 (123)
patient065 (689)
patient066 (431)
patient067 (372)
patient068 (673)
patient069 (309)
patient070 (299)
patient071 (238)
patient072 (390)
patient073 (541)
patient074 (466)
patient075 (306)
patient076 (464)
patient077 (343)
patient078 (442)
patient079 (672)
patient080 (530)
Proband X1 (487)
Proband X10 (338)
Proband X11 (580)
Proband X12 (442)
Proband X13 (379)
Proband X14 (148)
Proband X15 (439)
Proband X2 (515)
Proband X4 (439)
Proband X5 (289)
Proband X6 (249)
Proband X7 (530)
Proband X8 (303)
Proband X9 (298)
sim (137)
2.5
0.0
−2.5
−5.0
wavelet−HLH_glcm_SumEntropy
frame

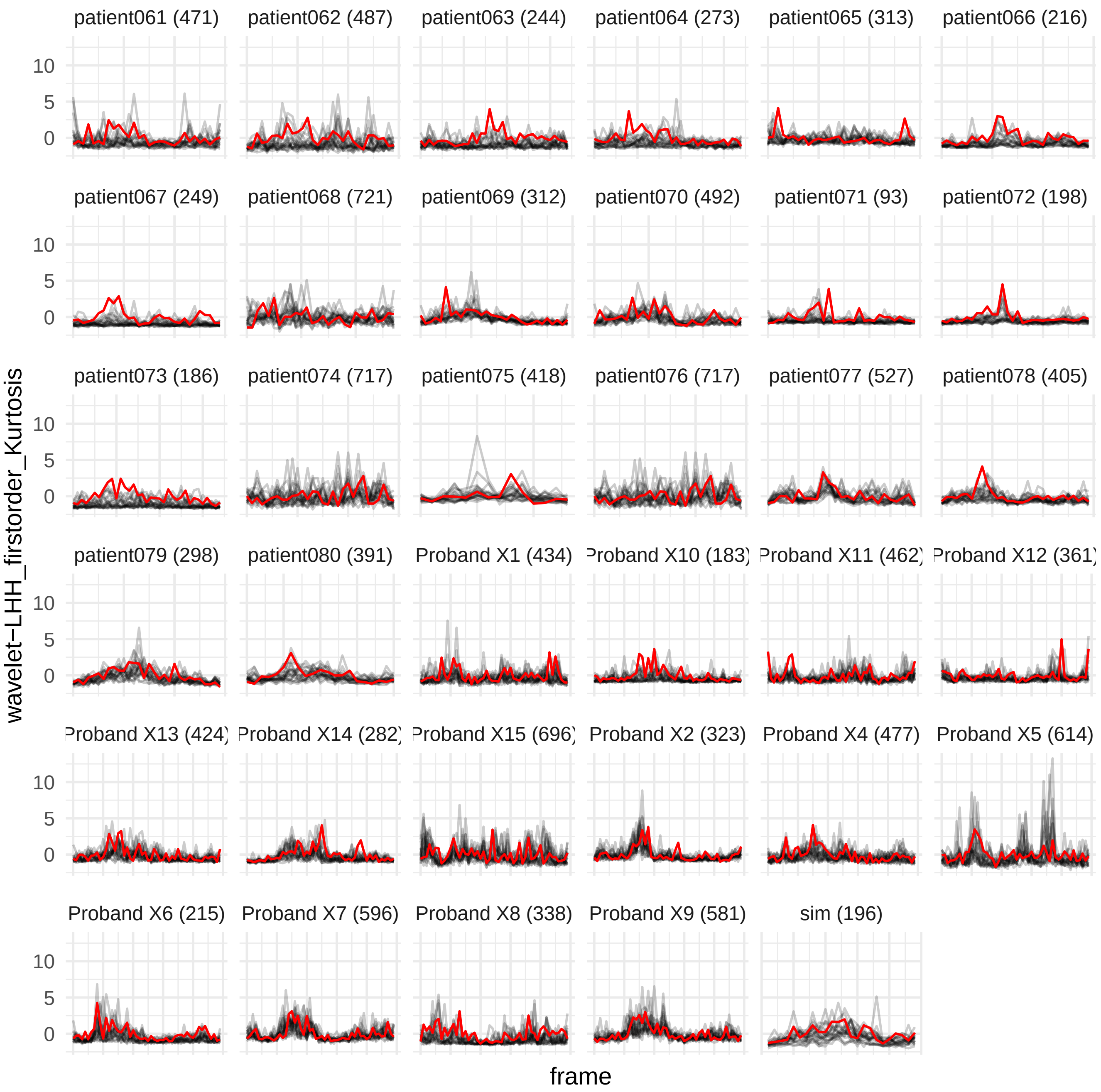

patient061 (471)
patient062 (487)
patient063 (244)
patient064 (273)
patient065 (313)
patient066 (216)
patient067 (249)
patient068 (721)
patient069 (312)
patient070 (492)
patient071 (93)
patient072 (198)
patient073 (186)
patient074 (717)
patient075 (418)
patient076 (717)
patient077 (527)
patient078 (405)
patient079 (298)
patient080 (391)
Proband X1 (434)
Proband X10 (183)
Proband X11 (462)
Proband X12 (361)
Proband X13 (424)
Proband X14 (282)
Proband X15 (696)
Proband X2 (323)
Proband X4 (477)
Proband X5 (614)
Proband X6 (215)
Proband X7 (596)
Proband X8 (338)
Proband X9 (581)
sim (196)
10
5
0
wavelet−LHH_firstorder_Kurtosis
frame

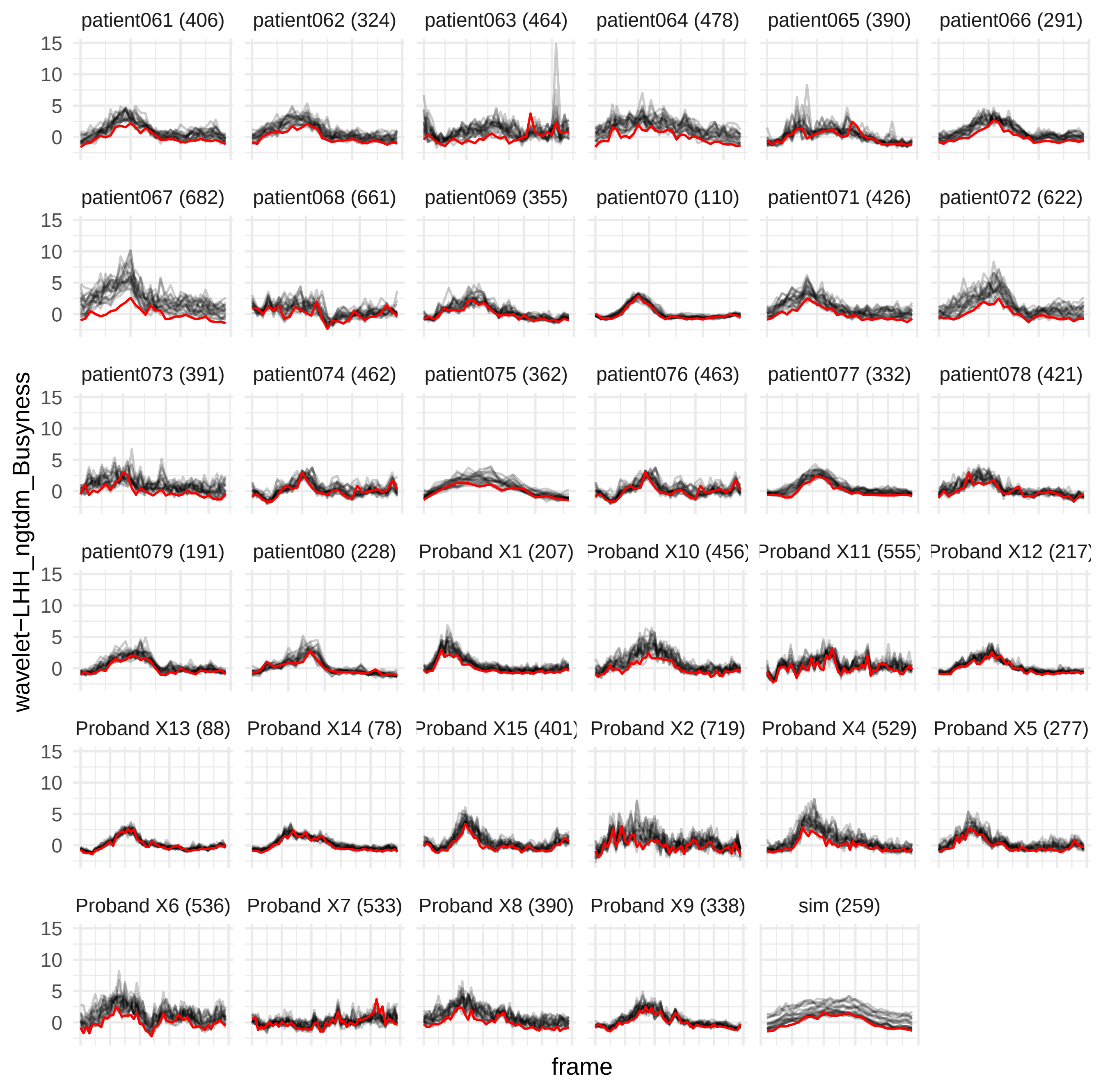

patient061 (406)
patient062 (324)
patient063 (464)
patient064 (478)
patient065 (390)
patient066 (291)
patient067 (682)
patient068 (661)
patient069 (355)
patient070 (110)
patient071 (426)
patient072 (622)
patient073 (391)
patient074 (462)
patient075 (362)
patient076 (463)
patient077 (332)
patient078 (421)
patient079 (191)
patient080 (228)
Proband X1 (207)
Proband X10 (456)
Proband X11 (555)
Proband X12 (217)
Proband X13 (88)
Proband X14 (78)
Proband X15 (401)
Proband X2 (719)
Proband X4 (529)
Proband X5 (277)
Proband X6 (536)
Proband X7 (533)
Proband X8 (390)
Proband X9 (338)
sim (259)
wavelet−LHH_ngtdm_Busyness
frame
15
10
5
0

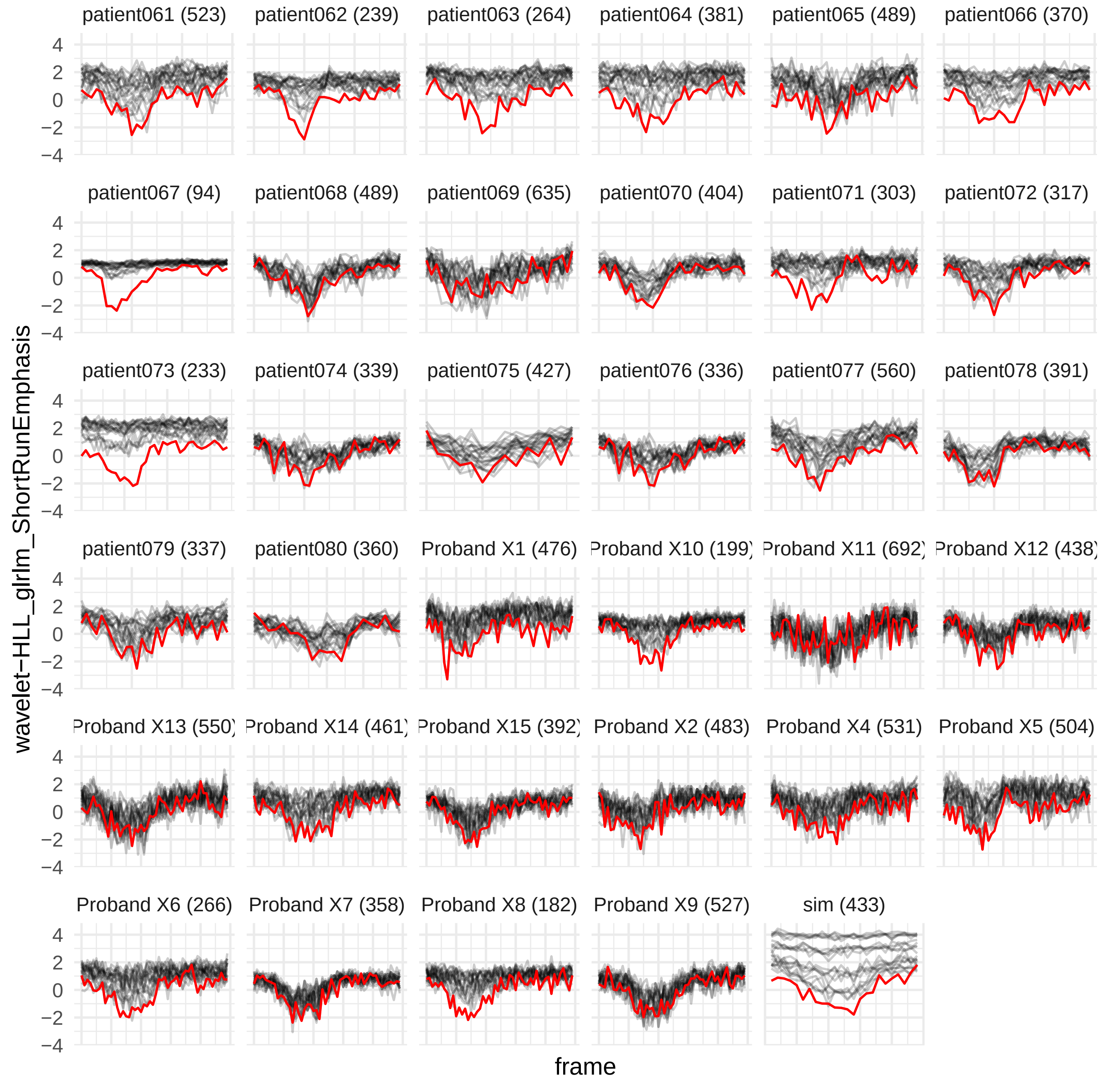
patient061 (523)
patient062 (239)
patient063 (264)
patient064 (381)
patient065 (489)
patient066 (370)
patient067 (94)
patient068 (489)
patient069 (635)
patient070 (404)
patient071 (303)
patient072 (317)
patient073 (233)
patient074 (339)
patient075 (427)
patient076 (336)
patient077 (560)
patient078 (391)
patient079 (337)
patient080 (360)
Proband X1 (476)
Proband X10 (199)
Proband X11 (692)
Proband X12 (438)
Proband X13 (550)
Proband X14 (461)
Proband X15 (392)
Proband X2 (483)
Proband X4 (531)
Proband X5 (504)
Proband X6 (266)
Proband X7 (358)
Proband X8 (182)
Proband X9 (527)
sim (433)
wavelet−HLL_glrlm_ShortRunEmphasis
frame
4
2
0
−2
−4

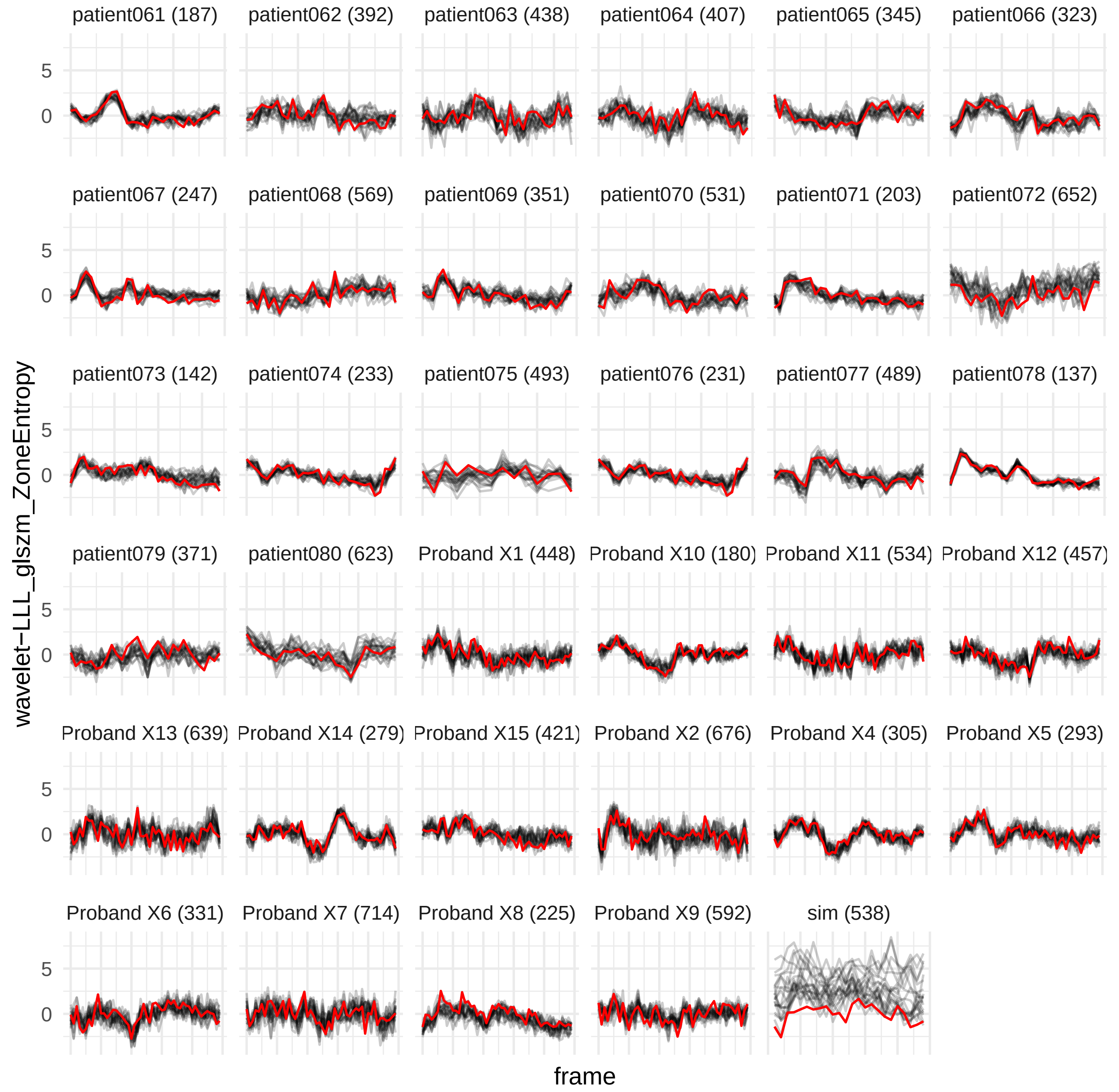

patient061 (187)
patient062 (392)
patient063 (438)
patient064 (407)
patient065 (345)
patient066 (323)
patient067 (247)
patient068 (569)
patient069 (351)
patient070 (531)
patient071 (203)
patient072 (652)
patient073 (142)
patient074 (233)
patient075 (493)
patient076 (231)
patient077 (489)
patient078 (137)
patient079 (371)
patient080 (623)
Proband X1 (448)
Proband X10 (180)
Proband X11 (534)
Proband X12 (457)
Proband X13 (639)
Proband X14 (279)
Proband X15 (421)
Proband X2 (676)
Proband X4 (305)
Proband X5 (293)
Proband X6 (331)
Proband X7 (714)
Proband X8 (225)
Proband X9 (592)
sim (538)
5
0
wavelet−LLL_glszm_ZoneEntropy
frame

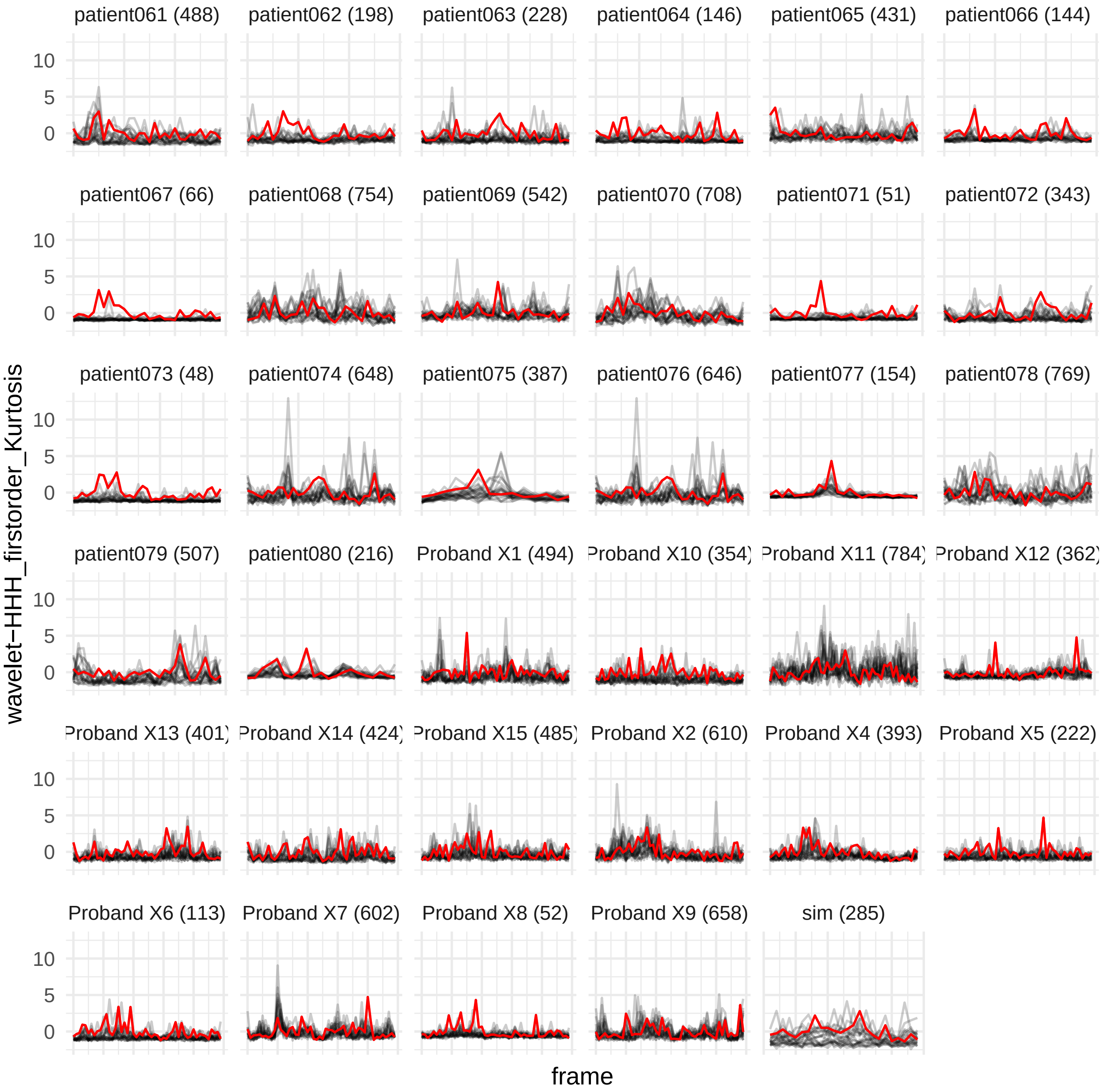

patient061 (488)
patient062 (198)
patient063 (228)
patient064 (146)
patient065 (431)
patient066 (144)
patient067 (66)
patient068 (754)
patient069 (542)
patient070 (708)
patient071 (51)
patient072 (343)
patient073 (48)
patient074 (648)
patient075 (387)
patient076 (646)
patient077 (154)
patient078 (769)
patient079 (507)
patient080 (216)
Proband X1 (494)
Proband X10 (354)
Proband X11 (784)
Proband X12 (362)
Proband X13 (401)
Proband X14 (424)
Proband X15 (485)
Proband X2 (610)
Proband X4 (393)
Proband X5 (222)
Proband X6 (113)
Proband X7 (602)
Proband X8 (52)
Proband X9 (658)
sim (285)
10
5
0
wavelet−HHH_firstorder_Kurtosis
frame

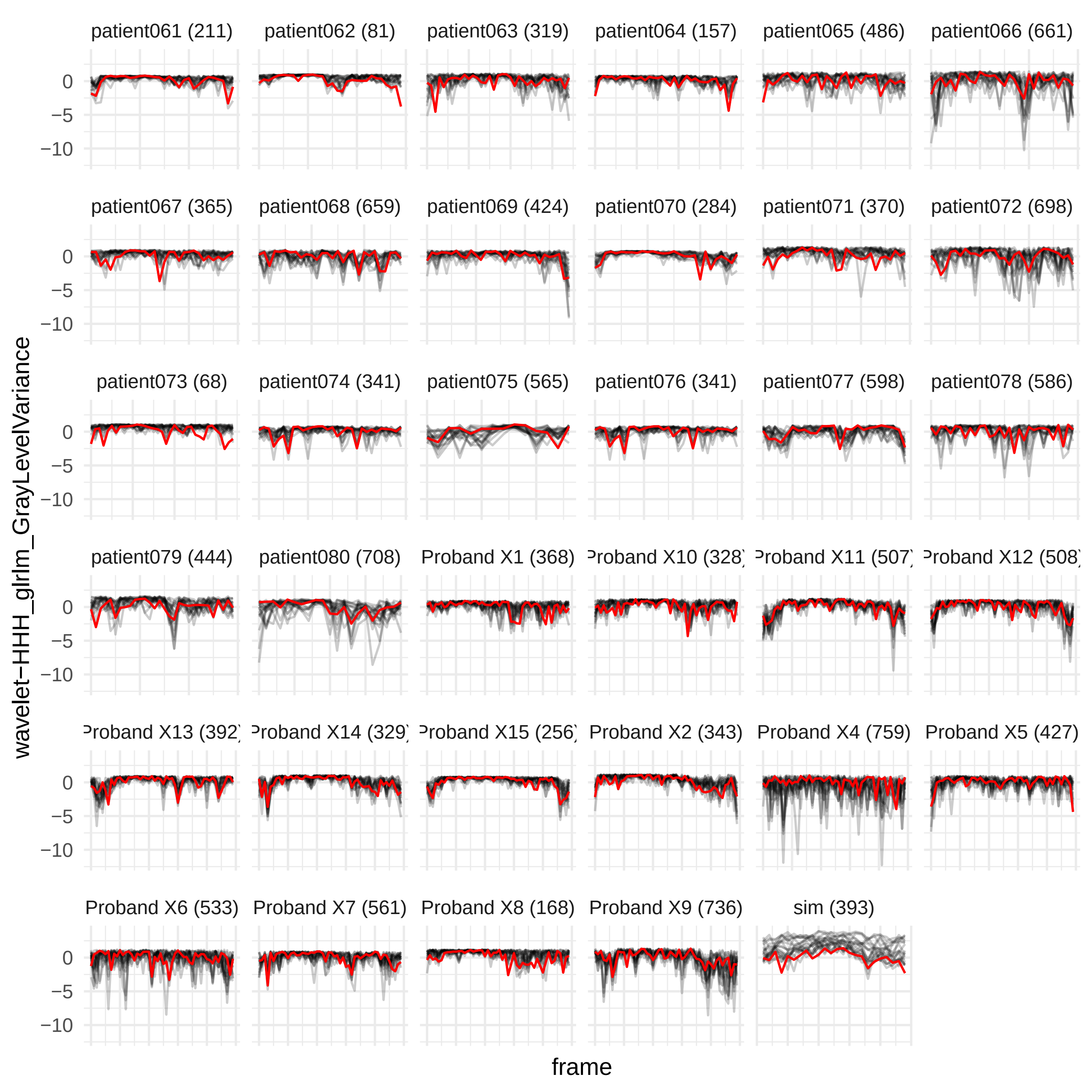
patient061 (211)
patient062 (81)
patient063 (319)
patient064 (157)
patient065 (486)
patient066 (661)
patient067 (365)
patient068 (659)
patient069 (424)
patient070 (284)
patient071 (370)
patient072 (698)
patient073 (68)
patient074 (341)
patient075 (565)
patient076 (341)
patient077 (598)
patient078 (586)
patient079 (444)
patient080 (708)
Proband X1 (368)
Proband X10 (328)
Proband X11 (507)
Proband X12 (508)
Proband X13 (392)
Proband X14 (329)
Proband X15 (256)
Proband X2 (343)
Proband X4 (759)
Proband X5 (427)
Proband X6 (533)
Proband X7 (561)
Proband X8 (168)
Proband X9 (736)
sim (393)
0
−5
−10
wavelet−HHH_glrlm_GrayLevelVariance
frame

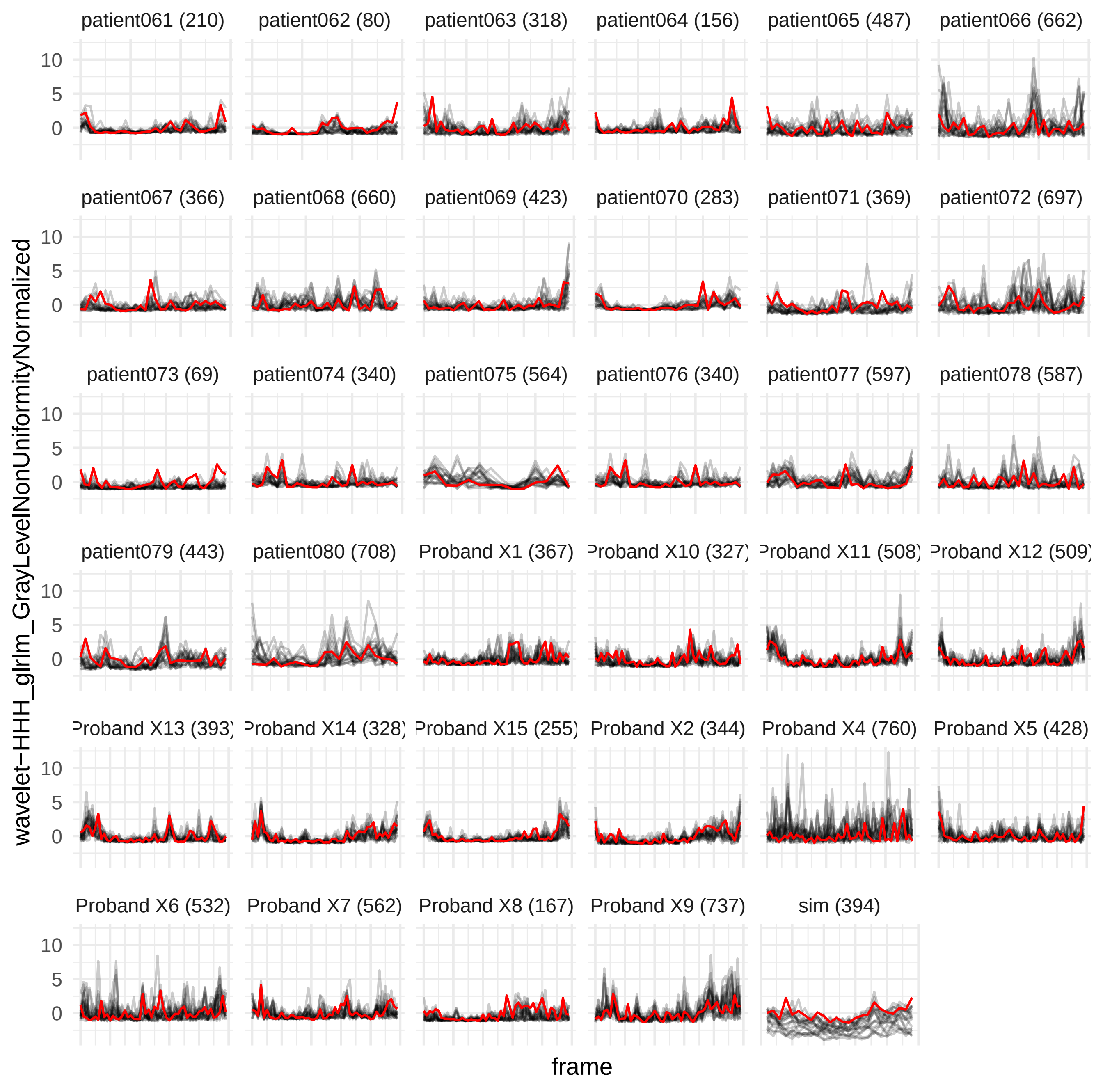
patient061 (210)
patient062 (80)
patient063 (318)
patient064 (156)
patient065 (487)
patient066 (662)
patient067 (366)
patient068 (660)
patient069 (423)
patient070 (283)
patient071 (369)
patient072 (697)
patient073 (69)
patient074 (340)
patient075 (564)
patient076 (340)
patient077 (597)
patient078 (587)
patient079 (443)
patient080 (708)
Proband X1 (367)
Proband X10 (327)
Proband X11 (508)
Proband X12 (509)
Proband X13 (393)
Proband X14 (328)
Proband X15 (255)
Proband X2 (344)
Proband X4 (760)
Proband X5 (428)
Proband X6 (532)
Proband X7 (562)
Proband X8 (167)
Proband X9 (737)
sim (394)
10
5
0
wavelet−HHH_glrlm_GrayLevelNonUniformityNormalized
frame

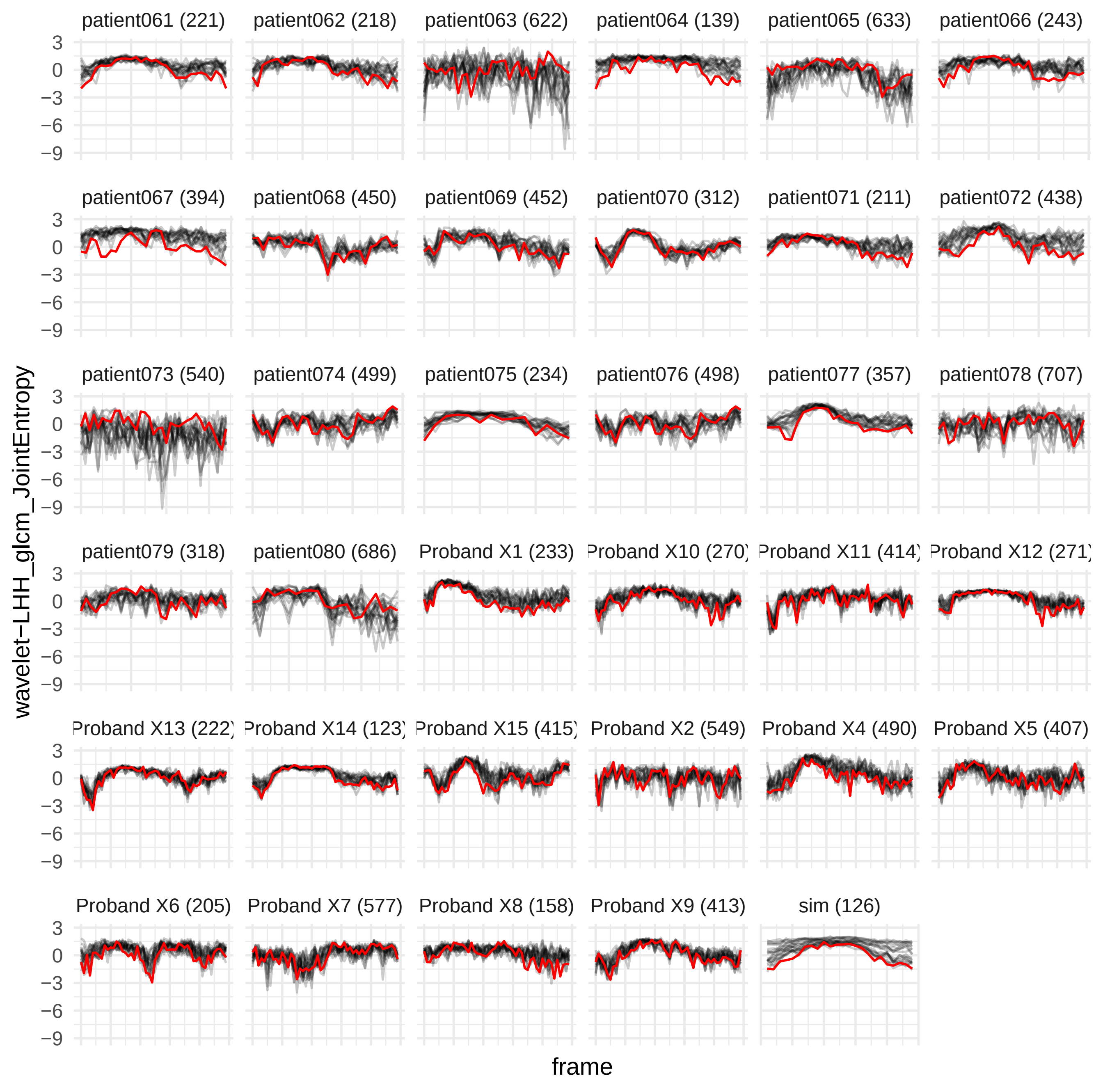
patient061 (221)
patient062 (218)
patient063 (622)
patient064 (139)
patient065 (633)
patient066 (243)
patient067 (394)
patient068 (450)
patient069 (452)
patient070 (312)
patient071 (211)
patient072 (438)
patient073 (540)
patient074 (499)
patient075 (234)
patient076 (498)
patient077 (357)
patient078 (707)
patient079 (318)
patient080 (686)
Proband X1 (233)
Proband X10 (270)
Proband X11 (414)
Proband X12 (271)
Proband X13 (222)
Proband X14 (123)
Proband X15 (415)
Proband X2 (549)
Proband X4 (490)
Proband X5 (407)
Proband X6 (205)
Proband X7 (577)
Proband X8 (158)
Proband X9 (413)
sim (126)
3
0
−3
−6
−9
wavelet−LHH_glcm_JointEntropy
frame

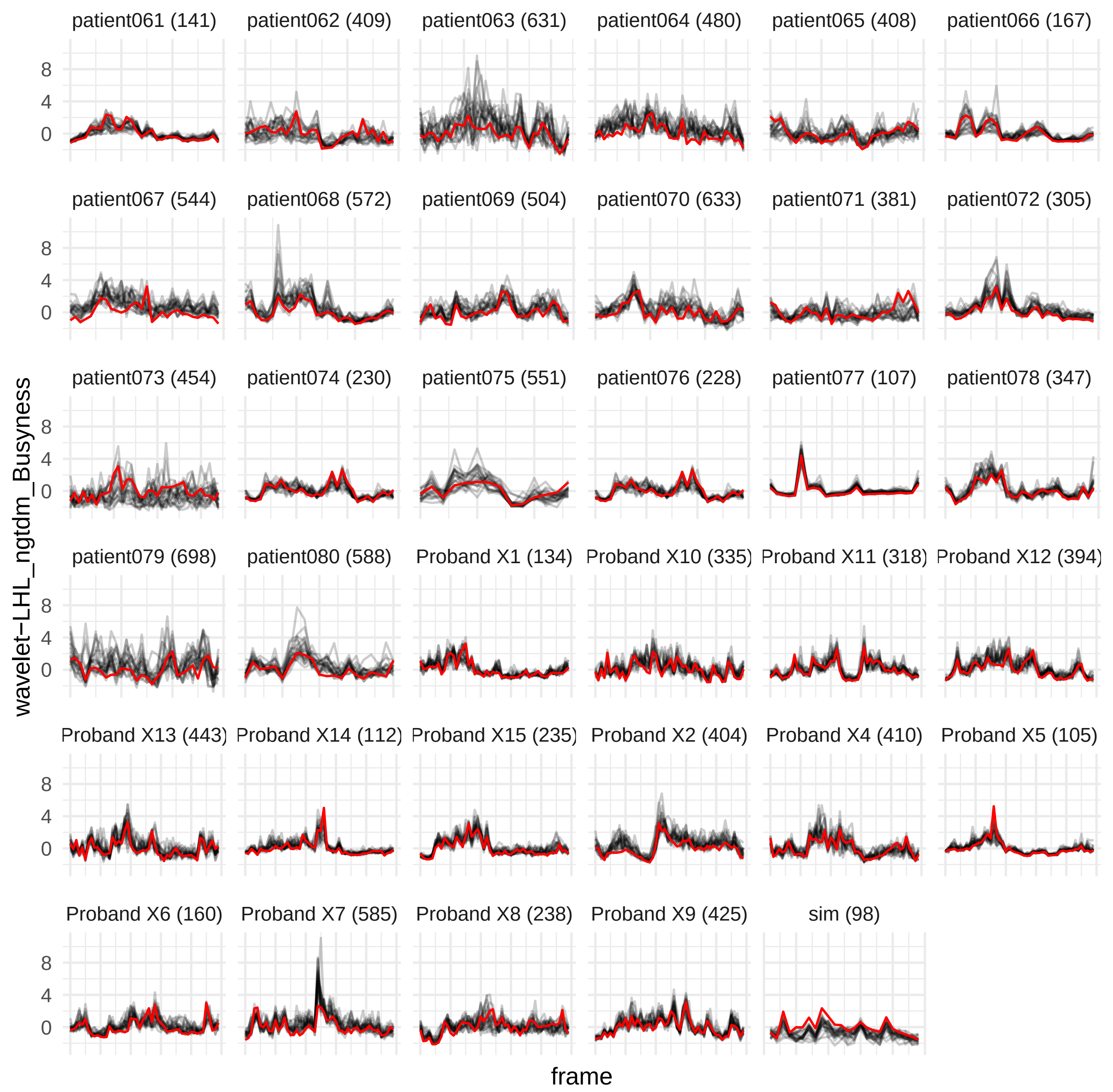

patient061 (141)
patient062 (409)
patient063 (631)
patient064 (480)
patient065 (408)
patient066 (167)
patient067 (544)
patient068 (572)
patient069 (504)
patient070 (633)
patient071 (381)
patient072 (305)
patient073 (454)
patient074 (230)
patient075 (551)
patient076 (228)
patient077 (107)
patient078 (347)
patient079 (698)
patient080 (588)
Proband X1 (134)
Proband X10 (335)
Proband X11 (318)
Proband X12 (394)
Proband X13 (443)
Proband X14 (112)
Proband X15 (235)
Proband X2 (404)
Proband X4 (410)
Proband X5 (105)
Proband X6 (160)
Proband X7 (585)
Proband X8 (238)
Proband X9 (425)
sim (98)
8
4
0
wavelet−LHL_ngtdm_Busyness
frame

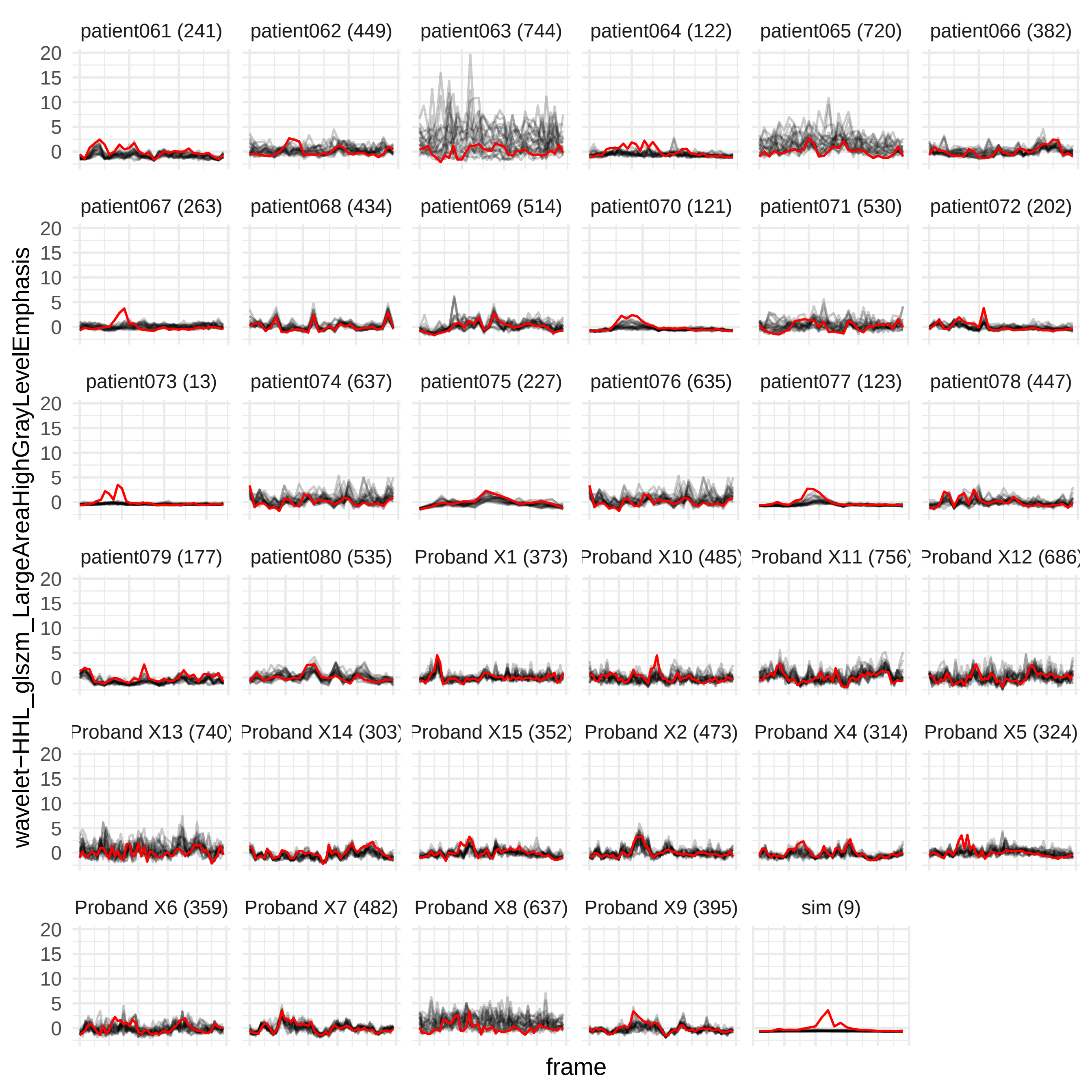

patient061 (241)
patient062 (449)
patient063 (744)
patient064 (122)
patient065 (720)
patient066 (382)
patient067 (263)
patient068 (434)
patient069 (514)
patient070 (121)
patient071 (530)
patient072 (202)
patient073 (13)
patient074 (637)
patient075 (227)
patient076 (635)
patient077 (123)
patient078 (447)
patient079 (177)
patient080 (535)
Proband X1 (373)
Proband X10 (485)
Proband X11 (756)
Proband X12 (686)
Proband X13 (740)
Proband X14 (303)
Proband X15 (352)
Proband X2 (473)
Proband X4 (314)
Proband X5 (324)
Proband X6 (359)
Proband X7 (482)
Proband X8 (637)
Proband X9 (395)
sim (9)
20
15
10
5
0
wavelet−HHL_glszm_LargeAreaHighGrayLevelEmphasis
frame

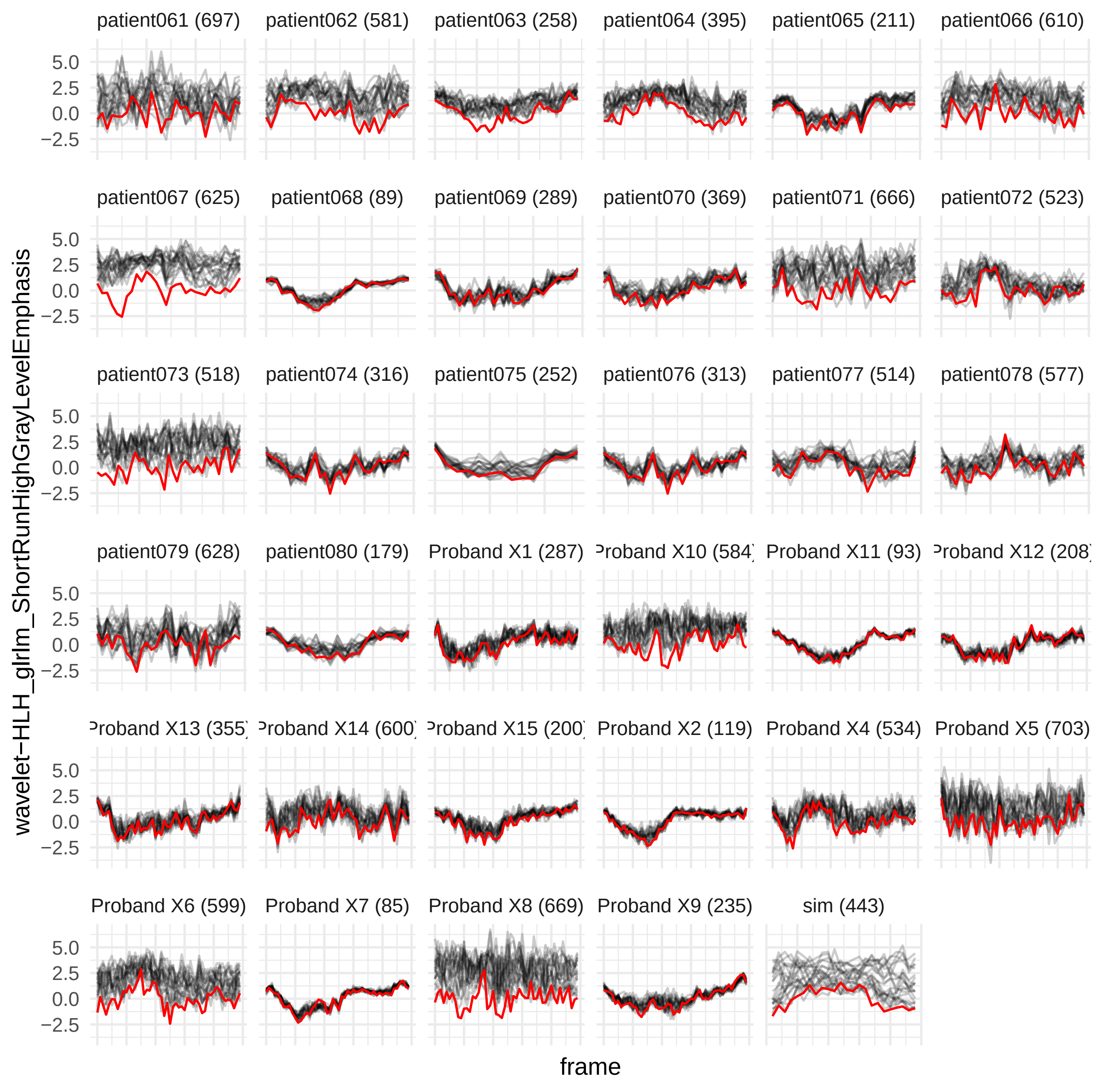
patient061 (697)
patient062 (581)
patient063 (258)
patient064 (395)
patient065 (211)
patient066 (610)
patient067 (625)
patient068 (89)
patient069 (289)
patient070 (369)
patient071 (666)
patient072 (523)
patient073 (518)
patient074 (316)
patient075 (252)
patient076 (313)
patient077 (514)
patient078 (577)
patient079 (628)
patient080 (179)
Proband X1 (287)
Proband X10 (584)
Proband X11 (93)
Proband X12 (208)
Proband X13 (355)
Proband X14 (600)
Proband X15 (200)
Proband X2 (119)
Proband X4 (534)
Proband X5 (703)
Proband X6 (599)
Proband X7 (85)
Proband X8 (669)
Proband X9 (235)
sim (443)
5.0
2.5
0.0
−2.5
wavelet−HLH_glrlm_ShortRunHighGrayLevelEmphasis
frame

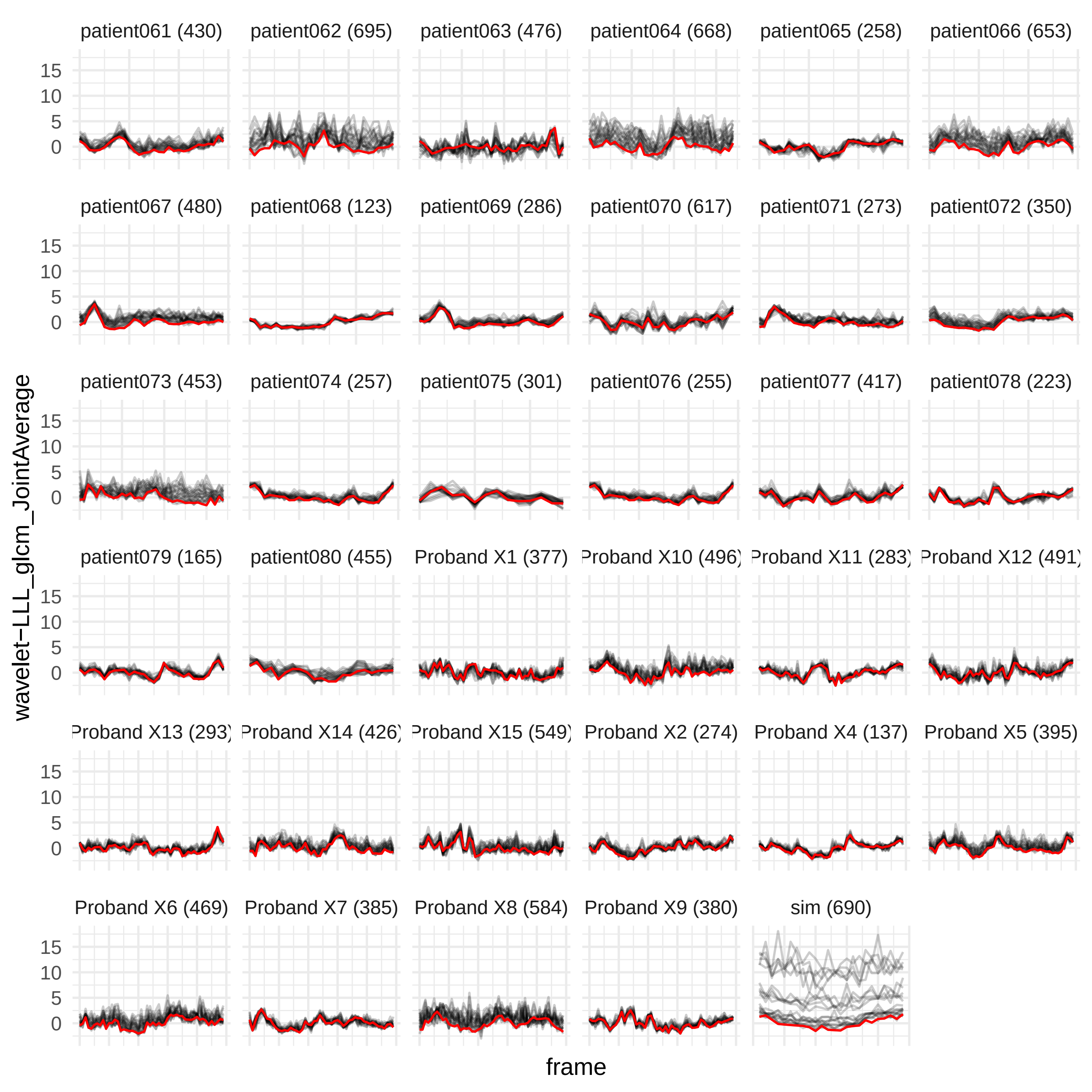

patient061 (430)
patient062 (695)
patient063 (476)
patient064 (668)
patient065 (258)
patient066 (653)
patient067 (480)
patient068 (123)
patient069 (286)
patient070 (617)
patient071 (273)
patient072 (350)
patient073 (453)
patient074 (257)
patient075 (301)
patient076 (255)
patient077 (417)
patient078 (223)
patient079 (165)
patient080 (455)
Proband X1 (377)
Proband X10 (496)
Proband X11 (283)
Proband X12 (491)
Proband X13 (293)
Proband X14 (426)
Proband X15 (549)
Proband X2 (274)
Proband X4 (137)
Proband X5 (395)
Proband X6 (469)
Proband X7 (385)
Proband X8 (584)
Proband X9 (380)
sim (690)
15
10
5
0
wavelet−LLL_glcm_JointAverage
frame

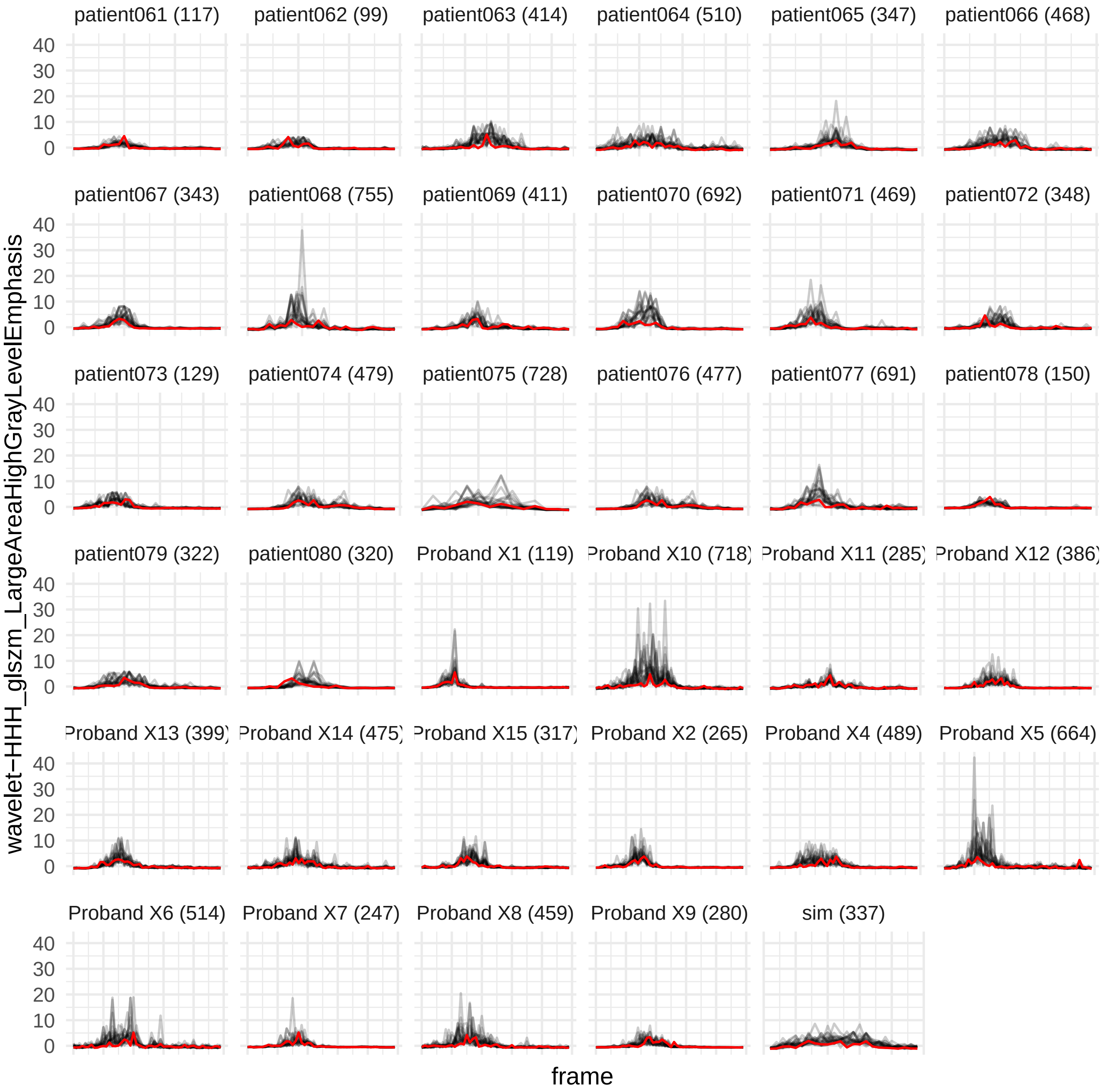

patient061 (117)
patient062 (99)
patient063 (414)
patient064 (510)
patient065 (347)
patient066 (468)
patient067 (343)
patient068 (755)
patient069 (411)
patient070 (692)
patient071 (469)
patient072 (348)
patient073 (129)
patient074 (479)
patient075 (728)
patient076 (477)
patient077 (691)
patient078 (150)
patient079 (322)
patient080 (320)
Proband X1 (119)
Proband X10 (718)
Proband X11 (285)
Proband X12 (386)
Proband X13 (399)
Proband X14 (475)
Proband X15 (317)
Proband X2 (265)
Proband X4 (489)
Proband X5 (664)
Proband X6 (514)
Proband X7 (247)
Proband X8 (459)
Proband X9 (280)
sim (337)
0
10
20
30
40
wavelet−HHH_glszm_LargeAreaHighGrayLevelEmphasis
frame

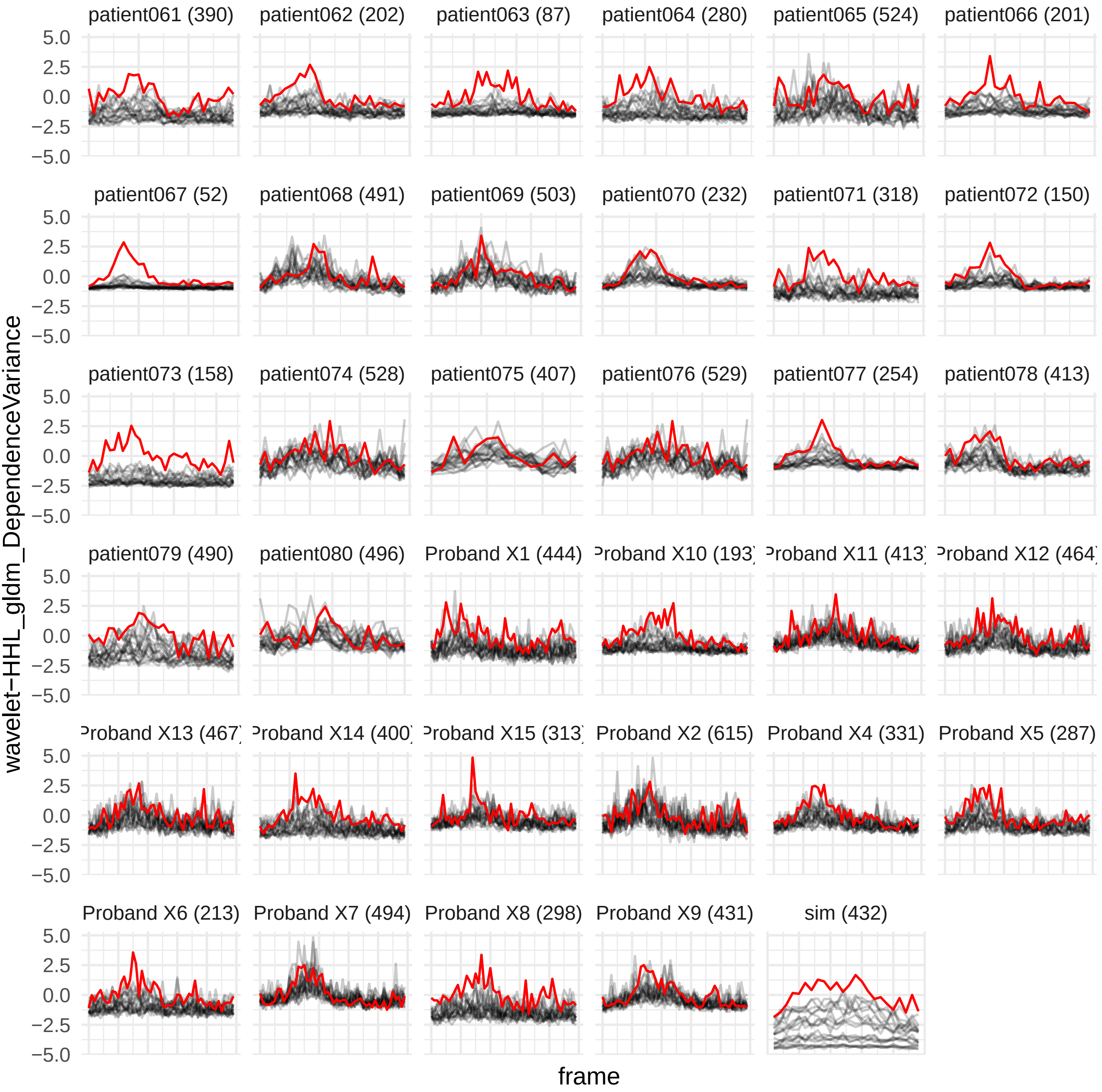
patient061 (390)
patient062 (202)
patient063 (87)
patient064 (280)
patient065 (524)
patient066 (201)
patient067 (52)
patient068 (491)
patient069 (503)
patient070 (232)
patient071 (318)
patient072 (150)
patient073 (158)
patient074 (528)
patient075 (407)
patient076 (529)
patient077 (254)
patient078 (413)
patient079 (490)
patient080 (496)
Proband X1 (444)
Proband X10 (193)
Proband X11 (413)
Proband X12 (464)
Proband X13 (467)
Proband X14 (400)
Proband X15 (313)
Proband X2 (615)
Proband X4 (331)
Proband X5 (287)
Proband X6 (213)
Proband X7 (494)
Proband X8 (298)
Proband X9 (431)
sim (432)
5.0
2.5
0.0
−2.5
−5.0
wavelet−HHL_gldm_DependenceVariance
frame

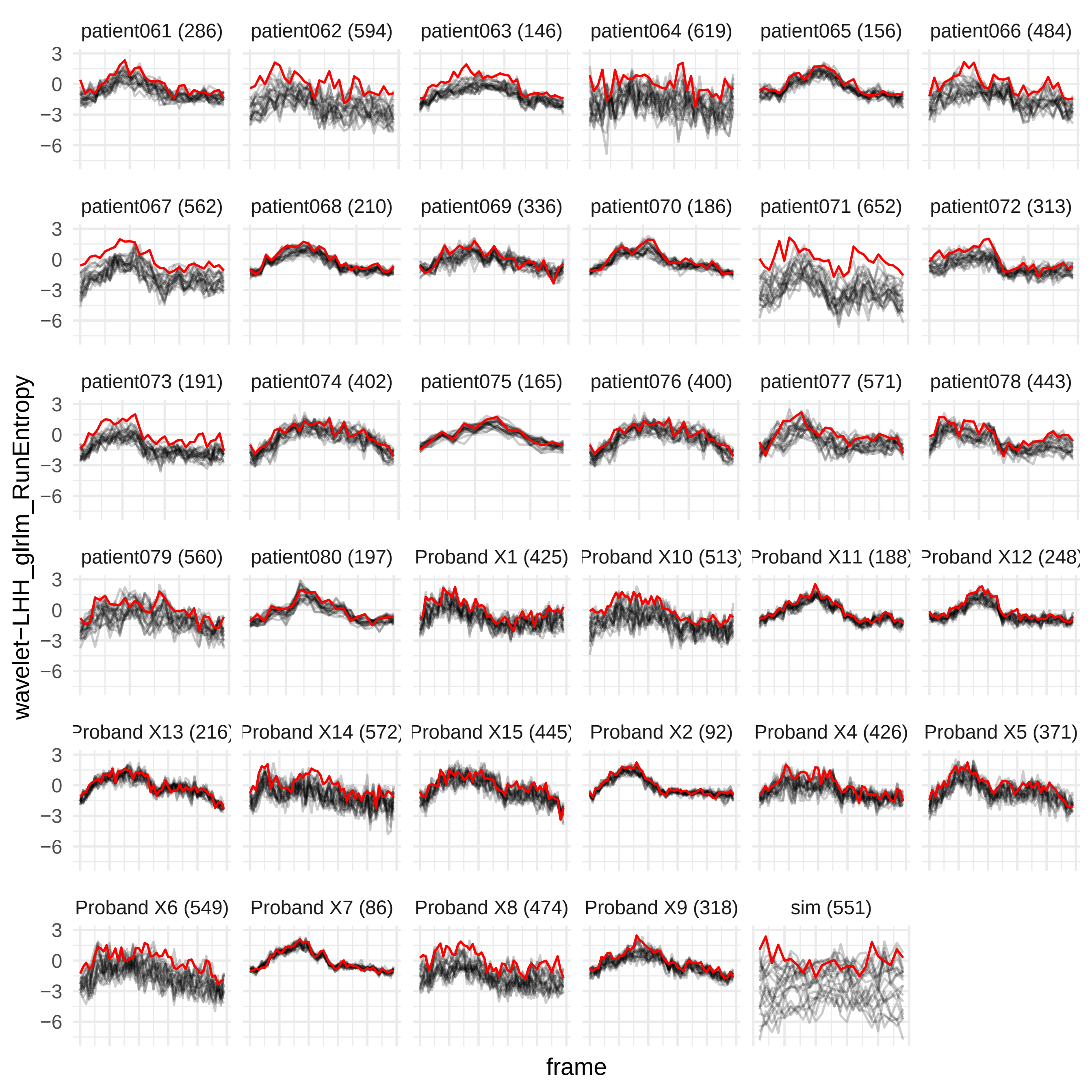
patient061 (286)
patient062 (594)
patient063 (146)
patient064 (619)
patient065 (156)
patient066 (484)
patient067 (562)
patient068 (210)
patient069 (336)
patient070 (186)
patient071 (652)
patient072 (313)
patient073 (191)
patient074 (402)
patient075 (165)
patient076 (400)
patient077 (571)
patient078 (443)
patient079 (560)
patient080 (197)
Proband X1 (425)
Proband X10 (513)
Proband X11 (188)
Proband X12 (248)
Proband X13 (216)
Proband X14 (572)
Proband X15 (445)
Proband X2 (92)
Proband X4 (426)
Proband X5 (371)
Proband X6 (549)
Proband X7 (86)
Proband X8 (474)
Proband X9 (318)
sim (551)
3
0
−3
−6
wavelet−LHH_glrlm_RunEntropy
frame

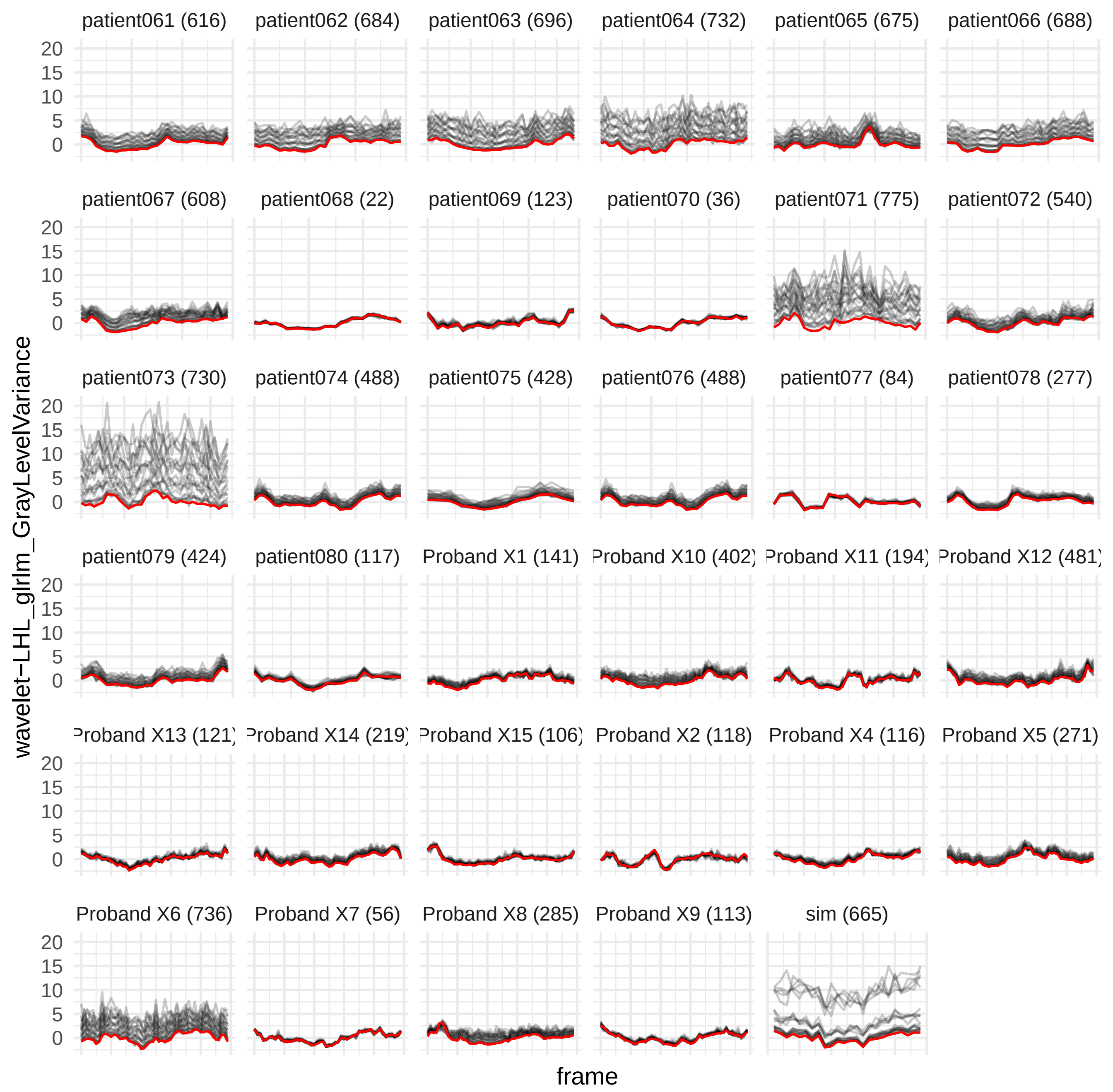
patient061 (616)
patient062 (684)
patient063 (696)
patient064 (732)
patient065 (675)
patient066 (688)
patient067 (608)
patient068 (22)
patient069 (123)
patient070 (36)
patient071 (775)
patient072 (540)
patient073 (730)
patient074 (488)
patient075 (428)
patient076 (488)
patient077 (84)
patient078 (277)
patient079 (424)
patient080 (117)
Proband X1 (141)
Proband X10 (402)
Proband X11 (194)
Proband X12 (481)
Proband X13 (121)
Proband X14 (219)
Proband X15 (106)
Proband X2 (118)
Proband X4 (116)
Proband X5 (271)
Proband X6 (736)
Proband X7 (56)
Proband X8 (285)
Proband X9 (113)
sim (665)
wavelet−LHL_glrlm_GrayLevelVariance
frame
0
5
10
15
20

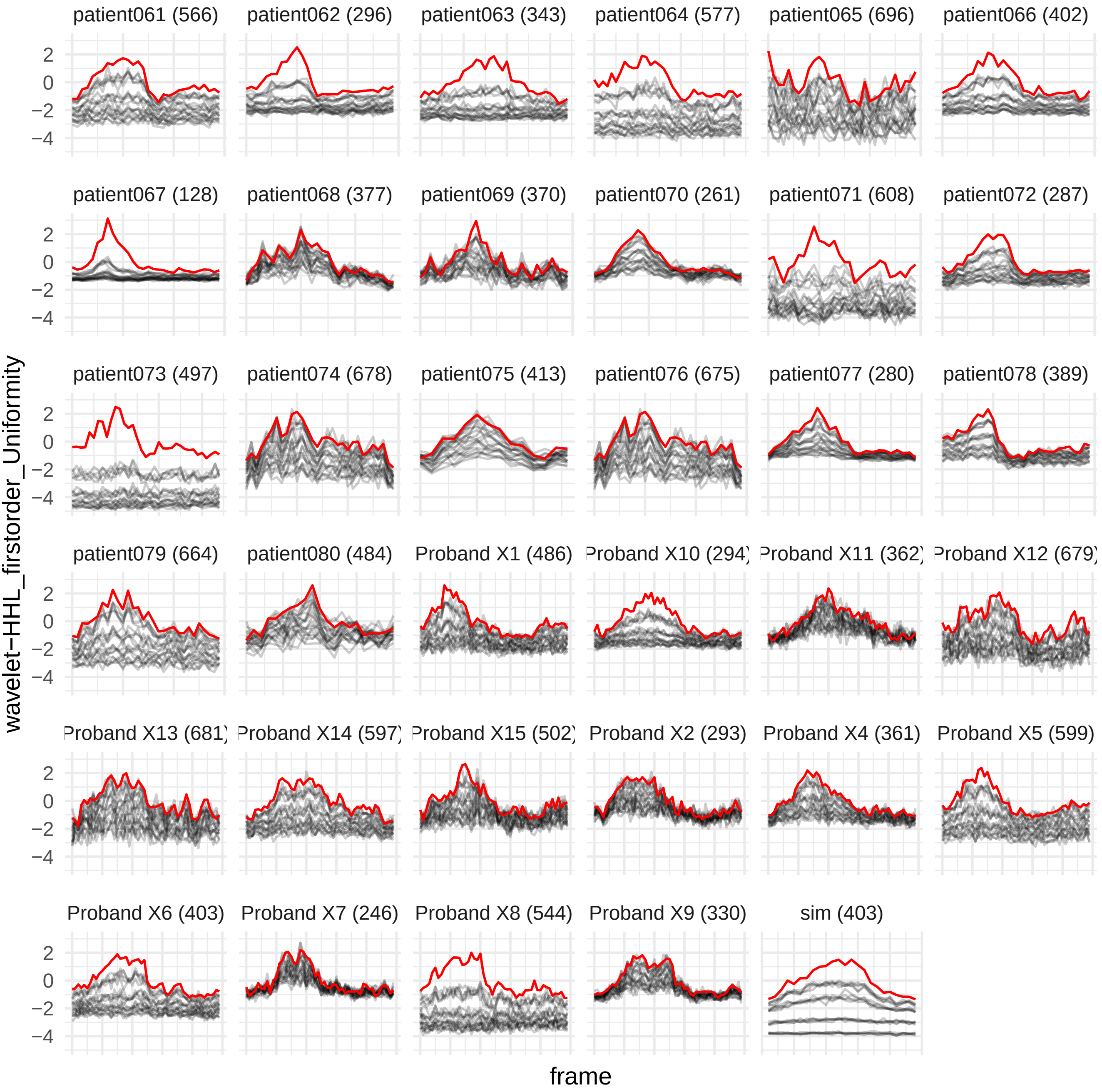

patient061 (566)
patient062 (296)
patient063 (343)
patient064 (577)
patient065 (696)
patient066 (402)
patient067 (128)
patient068 (377)
patient069 (370)
patient070 (261)
patient071 (608)
patient072 (287)
patient073 (497)
patient074 (678)
patient075 (413)
patient076 (675)
patient077 (280)
patient078 (389)
patient079 (664)
patient080 (484)
Proband X1 (486)
Proband X10 (294)
Proband X11 (362)
Proband X12 (679)
Proband X13 (681)
Proband X14 (597)
Proband X15 (502)
Proband X2 (293)
Proband X4 (361)
Proband X5 (599)
Proband X6 (403)
Proband X7 (246)
Proband X8 (544)
Proband X9 (330)
sim (403)
2
0
−2
−4
wavelet−HHL_firstorder_Uniformity
frame

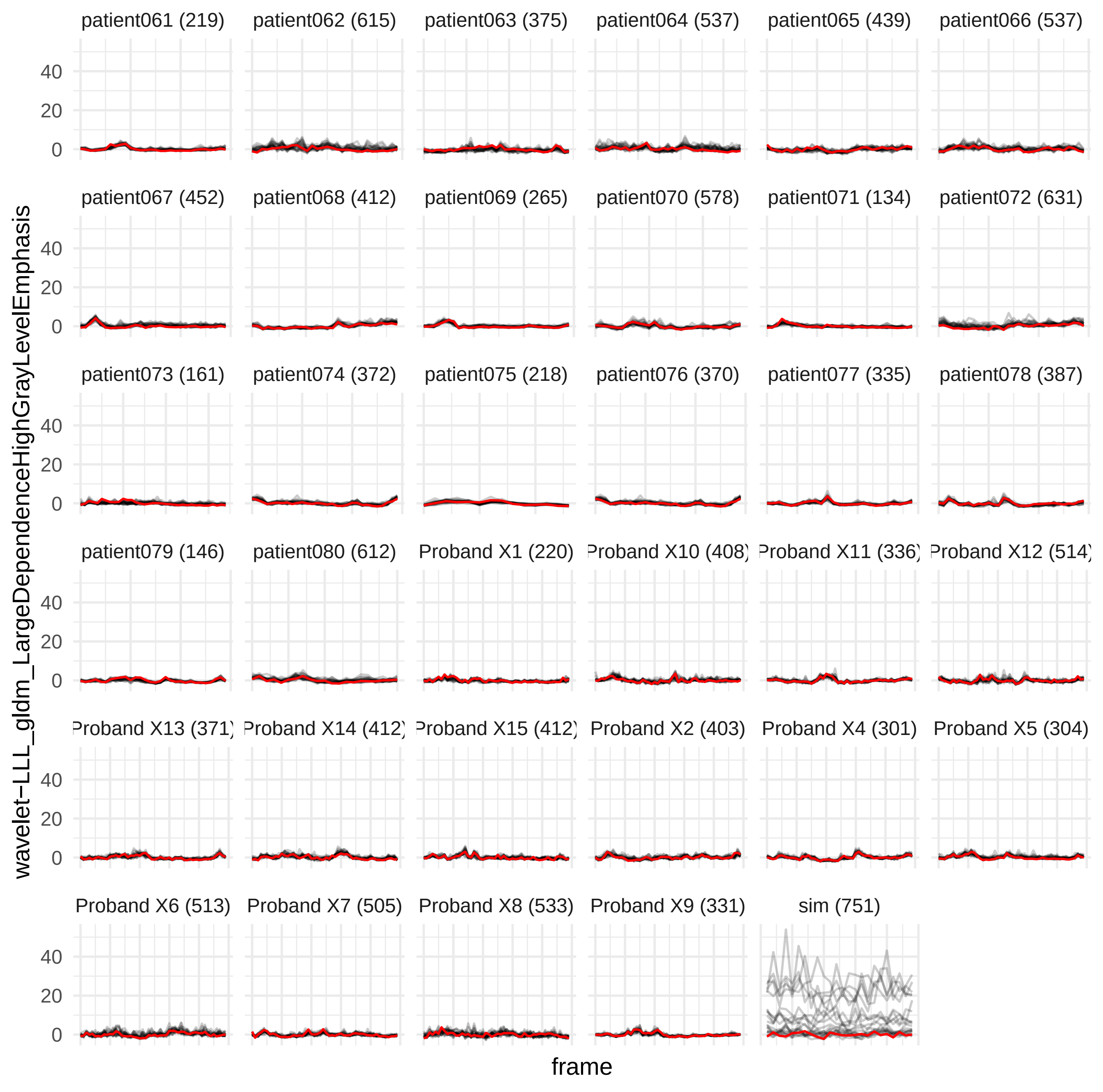

patient061 (219)
patient062 (615)
patient063 (375)
patient064 (537)
patient065 (439)
patient066 (537)
patient067 (452)
patient068 (412)
patient069 (265)
patient070 (578)
patient071 (134)
patient072 (631)
patient073 (161)
patient074 (372)
patient075 (218)
patient076 (370)
patient077 (335)
patient078 (387)
patient079 (146)
patient080 (612)
Proband X1 (220)
Proband X10 (408)
Proband X11 (336)
Proband X12 (514)
Proband X13 (371)
Proband X14 (412)
Proband X15 (412)
Proband X2 (403)
Proband X4 (301)
Proband X5 (304)
Proband X6 (513)
Proband X7 (505)
Proband X8 (533)
Proband X9 (331)
sim (751)
0
20
40
wavelet−LLL_gldm_LargeDependenceHighGrayLevelEmphasis
frame

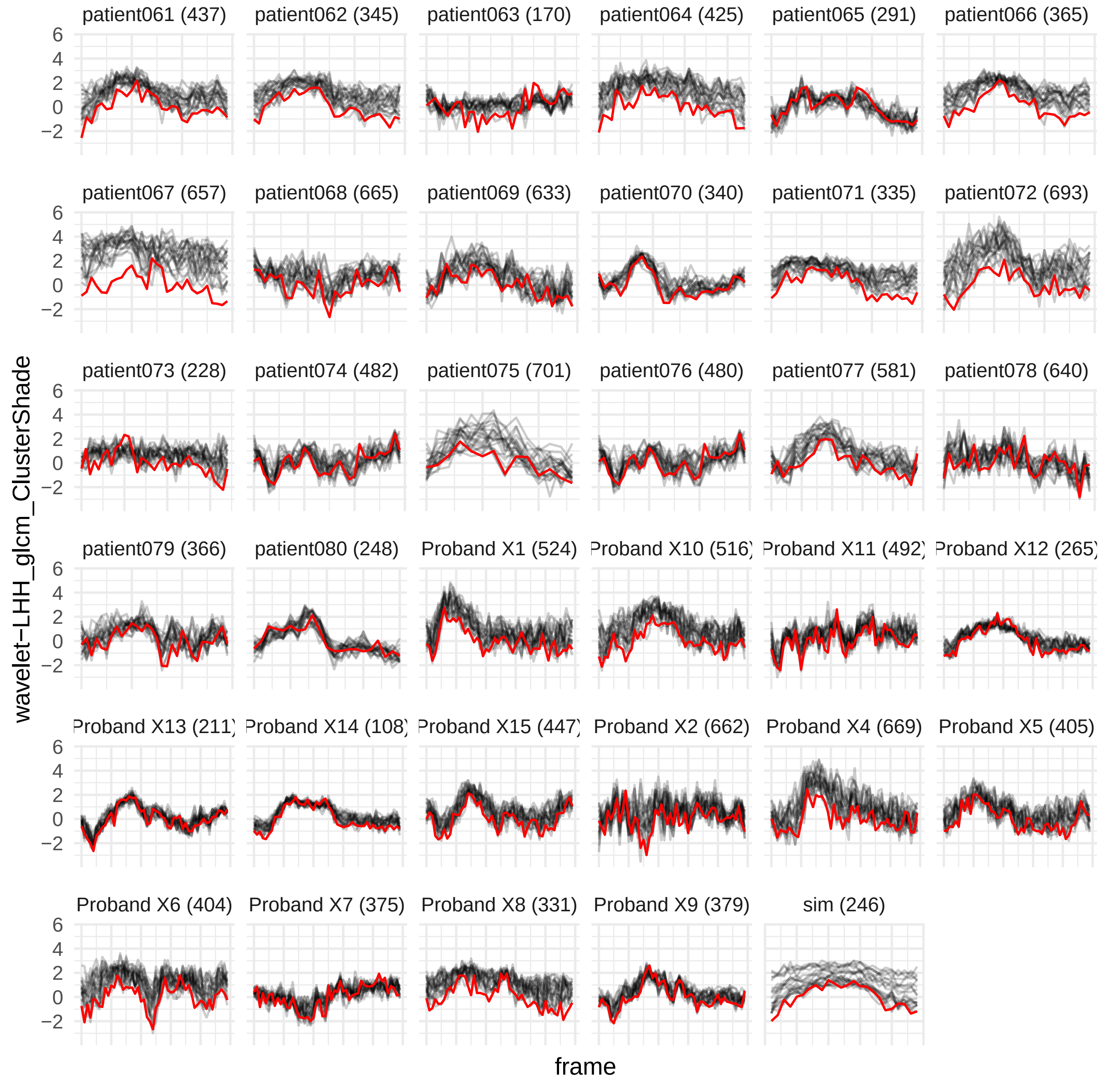
patient061 (437)
patient062 (345)
patient063 (170)
patient064 (425)
patient065 (291)
patient066 (365)
patient067 (657)
patient068 (665)
patient069 (633)
patient070 (340)
patient071 (335)
patient072 (693)
patient073 (228)
patient074 (482)
patient075 (701)
patient076 (480)
patient077 (581)
patient078 (640)
patient079 (366)
patient080 (248)
Proband X1 (524)
Proband X10 (516)
Proband X11 (492)
Proband X12 (265)
Proband X13 (211)
Proband X14 (108)
Proband X15 (447)
Proband X2 (662)
Proband X4 (669)
Proband X5 (405)
Proband X6 (404)
Proband X7 (375)
Proband X8 (331)
Proband X9 (379)
sim (246)
6
4
2
0
−2
wavelet−LHH_glcm_ClusterShade
frame

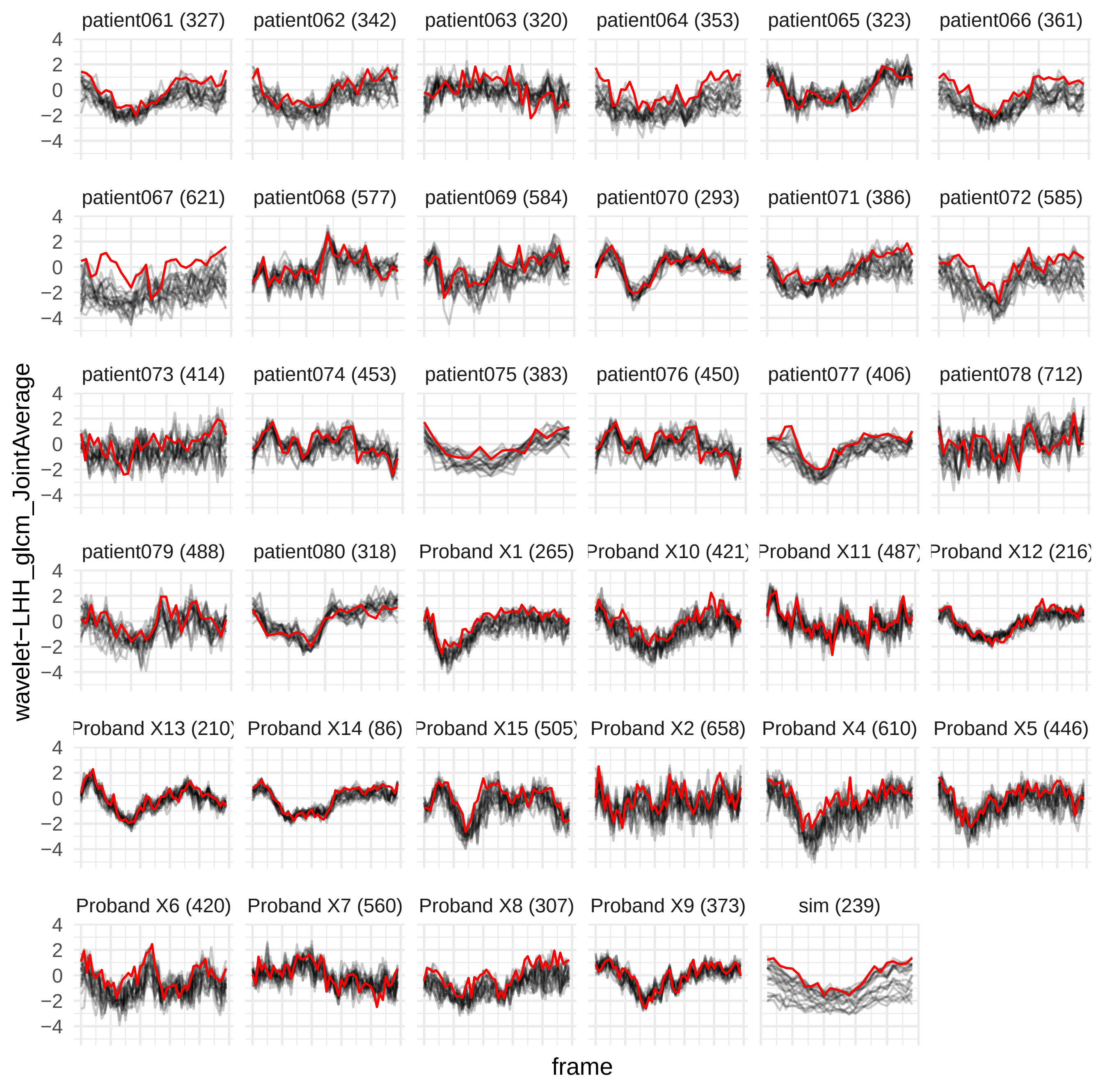
patient061 (327)
patient062 (342)
patient063 (320)
patient064 (353)
patient065 (323)
patient066 (361)
patient067 (621)
patient068 (577)
patient069 (584)
patient070 (293)
patient071 (386)
patient072 (585)
patient073 (414)
patient074 (453)
patient075 (383)
patient076 (450)
patient077 (406)
patient078 (712)
patient079 (488)
patient080 (318)
Proband X1 (265)
Proband X10 (421)
Proband X11 (487)
Proband X12 (216)
Proband X13 (210)
Proband X14 (86)
Proband X15 (505)
Proband X2 (658)
Proband X4 (610)
Proband X5 (446)
Proband X6 (420)
Proband X7 (560)
Proband X8 (307)
Proband X9 (373)
sim (239)
4
2
0
−2
−4
wavelet−LHH_glcm_JointAverage
frame

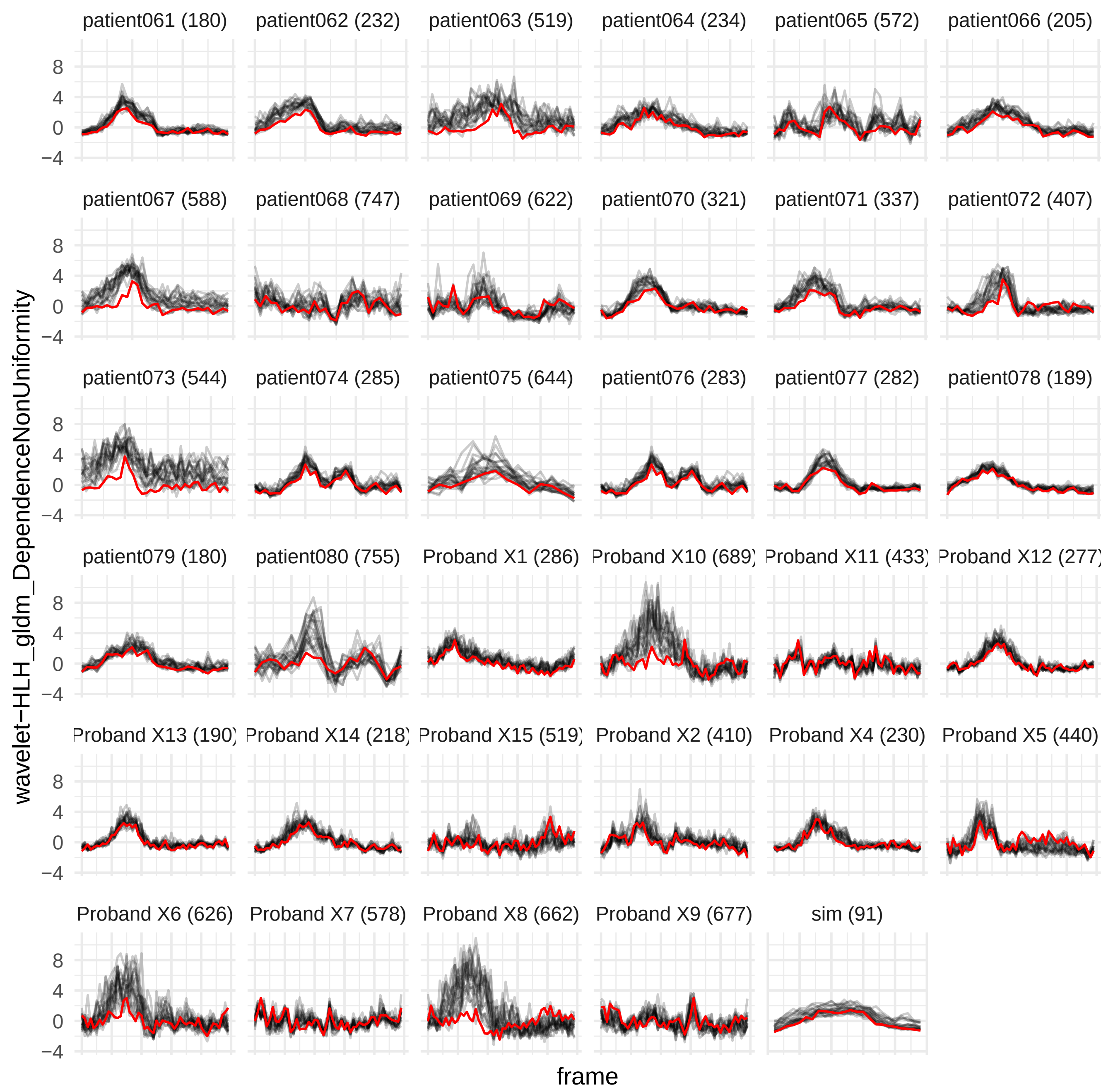
patient061 (180)
patient062 (232)
patient063 (519)
patient064 (234)
patient065 (572)
patient066 (205)
patient067 (588)
patient068 (747)
patient069 (622)
patient070 (321)
patient071 (337)
patient072 (407)
patient073 (544)
patient074 (285)
patient075 (644)
patient076 (283)
patient077 (282)
patient078 (189)
patient079 (180)
patient080 (755)
Proband X1 (286)
Proband X10 (689)
Proband X11 (433)
Proband X12 (277)
Proband X13 (190)
Proband X14 (218)
Proband X15 (519)
Proband X2 (410)
Proband X4 (230)
Proband X5 (440)
Proband X6 (626)
Proband X7 (578)
Proband X8 (662)
Proband X9 (677)
sim (91)
8
4
0
−4
wavelet−HLH_gldm_DependenceNonUniformity
frame

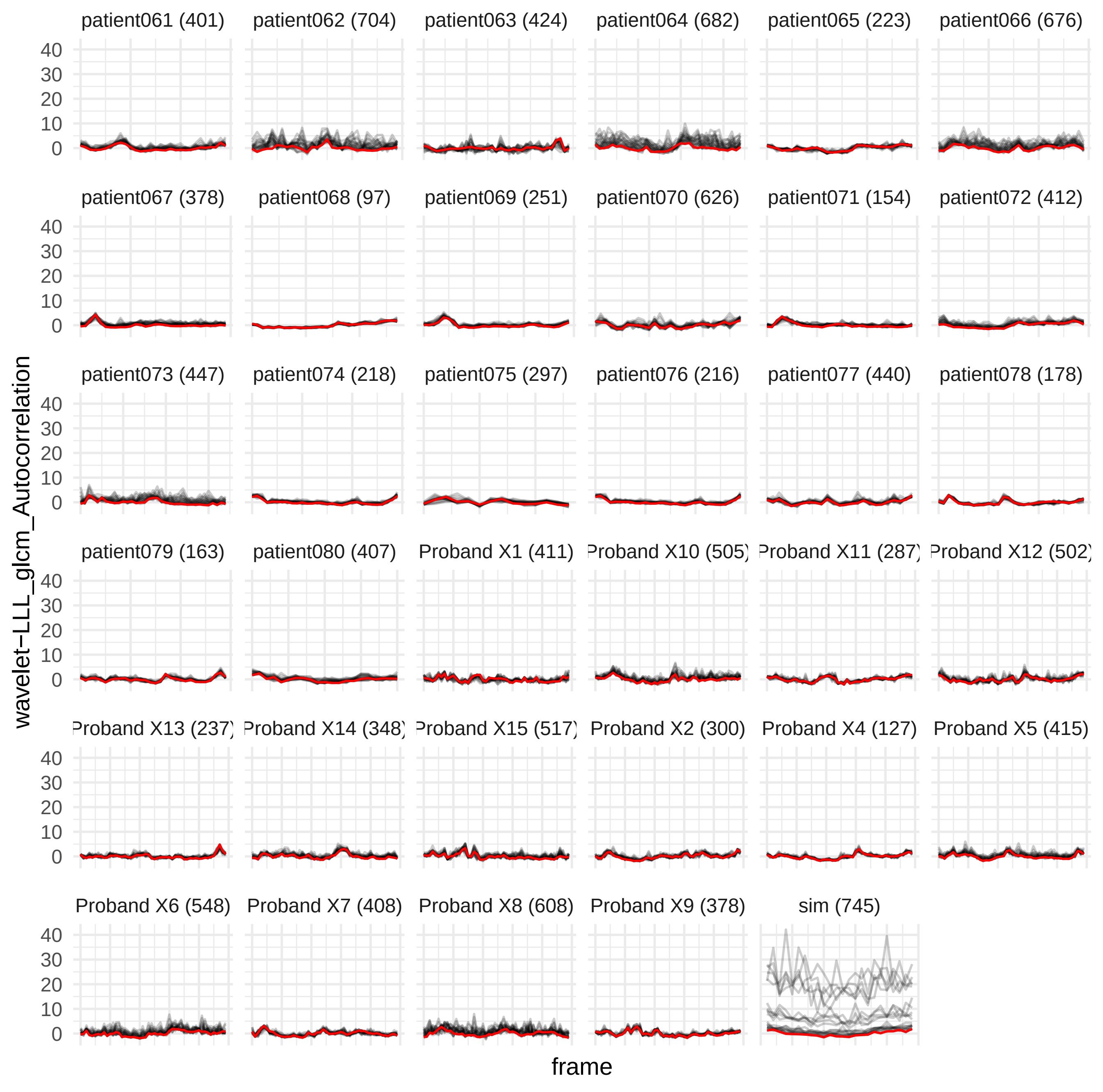

patient061 (401)
patient062 (704)
patient063 (424)
patient064 (682)
patient065 (223)
patient066 (676)
patient067 (378)
patient068 (97)
patient069 (251)
patient070 (626)
patient071 (154)
patient072 (412)
patient073 (447)
patient074 (218)
patient075 (297)
patient076 (216)
patient077 (440)
patient078 (178)
patient079 (163)
patient080 (407)
Proband X1 (411)
Proband X10 (505)
Proband X11 (287)
Proband X12 (502)
Proband X13 (237)
Proband X14 (348)
Proband X15 (517)
Proband X2 (300)
Proband X4 (127)
Proband X5 (415)
Proband X6 (548)
Proband X7 (408)
Proband X8 (608)
Proband X9 (378)
sim (745)
0
10
20
30
40
wavelet−LLL_glcm_Autocorrelation
frame

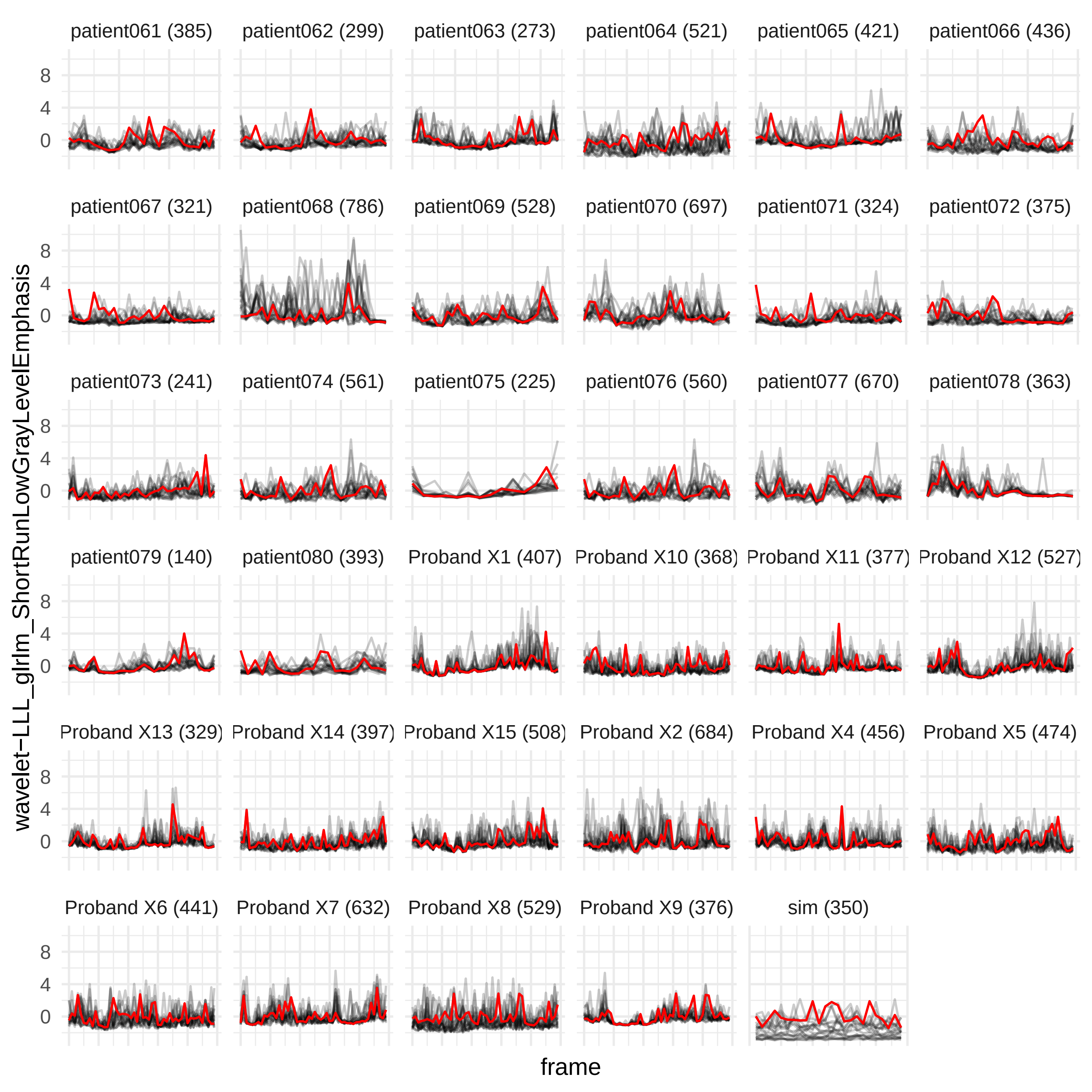
patient061 (385)
patient062 (299)
patient063 (273)
patient064 (521)
patient065 (421)
patient066 (436)
patient067 (321)
patient068 (786)
patient069 (528)
patient070 (697)
patient071 (324)
patient072 (375)
patient073 (241)
patient074 (561)
patient075 (225)
patient076 (560)
patient077 (670)
patient078 (363)
patient079 (140)
patient080 (393)
Proband X1 (407)
Proband X10 (368)
Proband X11 (377)
Proband X12 (527)
Proband X13 (329)
Proband X14 (397)
Proband X15 (508)
Proband X2 (684)
Proband X4 (456)
Proband X5 (474)
Proband X6 (441)
Proband X7 (632)
Proband X8 (529)
Proband X9 (376)
sim (350)
wavelet−LLL_glrlm_ShortRunLowGrayLevelEmphasis
8
4
0
frame

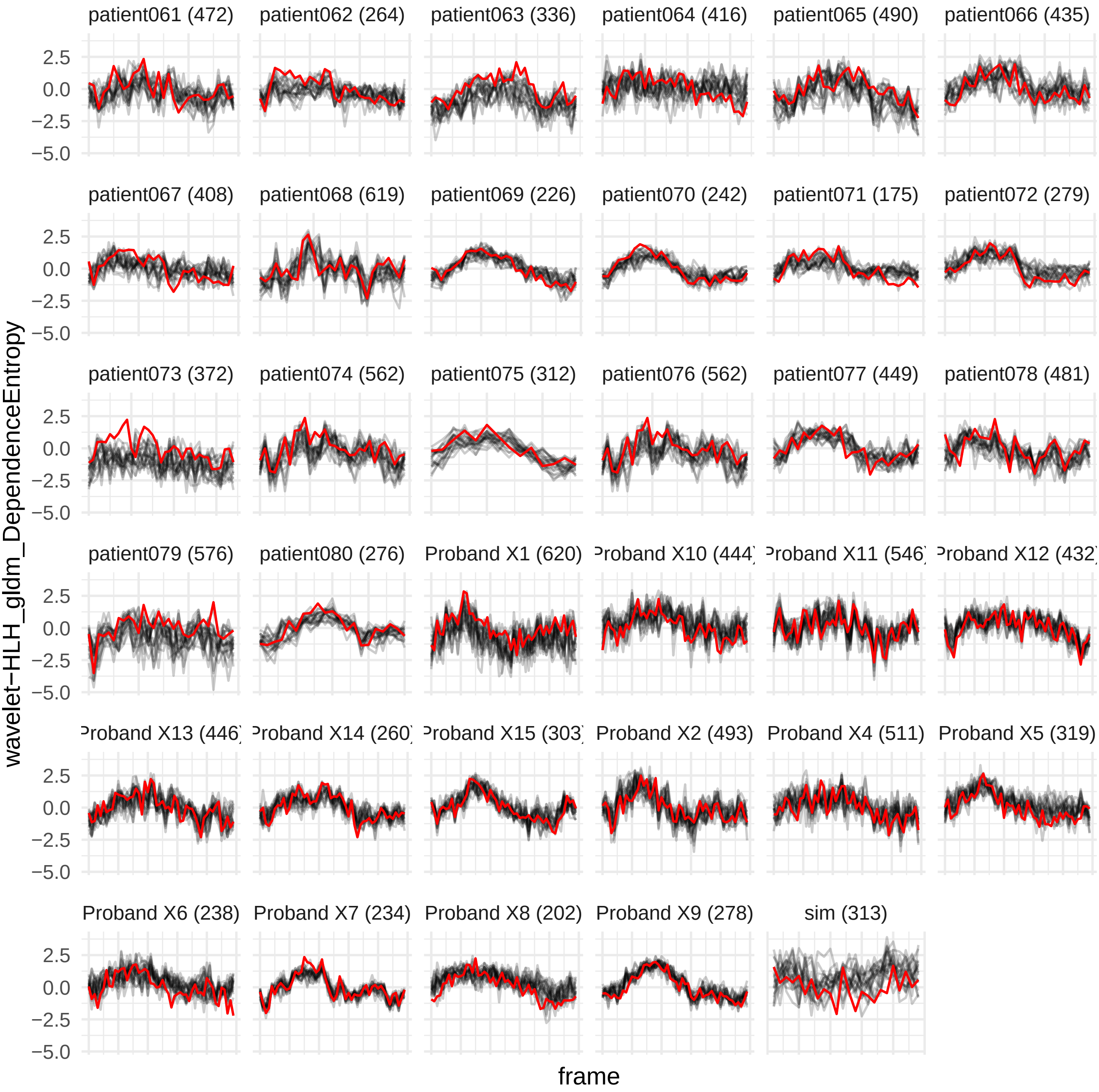

patient061 (472)
patient062 (264)
patient063 (336)
patient064 (416)
patient065 (490)
patient066 (435)
patient067 (408)
patient068 (619)
patient069 (226)
patient070 (242)
patient071 (175)
patient072 (279)
patient073 (372)
patient074 (562)
patient075 (312)
patient076 (562)
patient077 (449)
patient078 (481)
patient079 (576)
patient080 (276)
Proband X1 (620)
Proband X10 (444)
Proband X11 (546)
Proband X12 (432)
Proband X13 (446)
Proband X14 (260)
Proband X15 (303)
Proband X2 (493)
Proband X4 (511)
Proband X5 (319)
Proband X6 (238)
Proband X7 (234)
Proband X8 (202)
Proband X9 (278)
sim (313)
2.5
0.0
−2.5
−5.0
wavelet−HLH_gldm_DependenceEntropy
frame

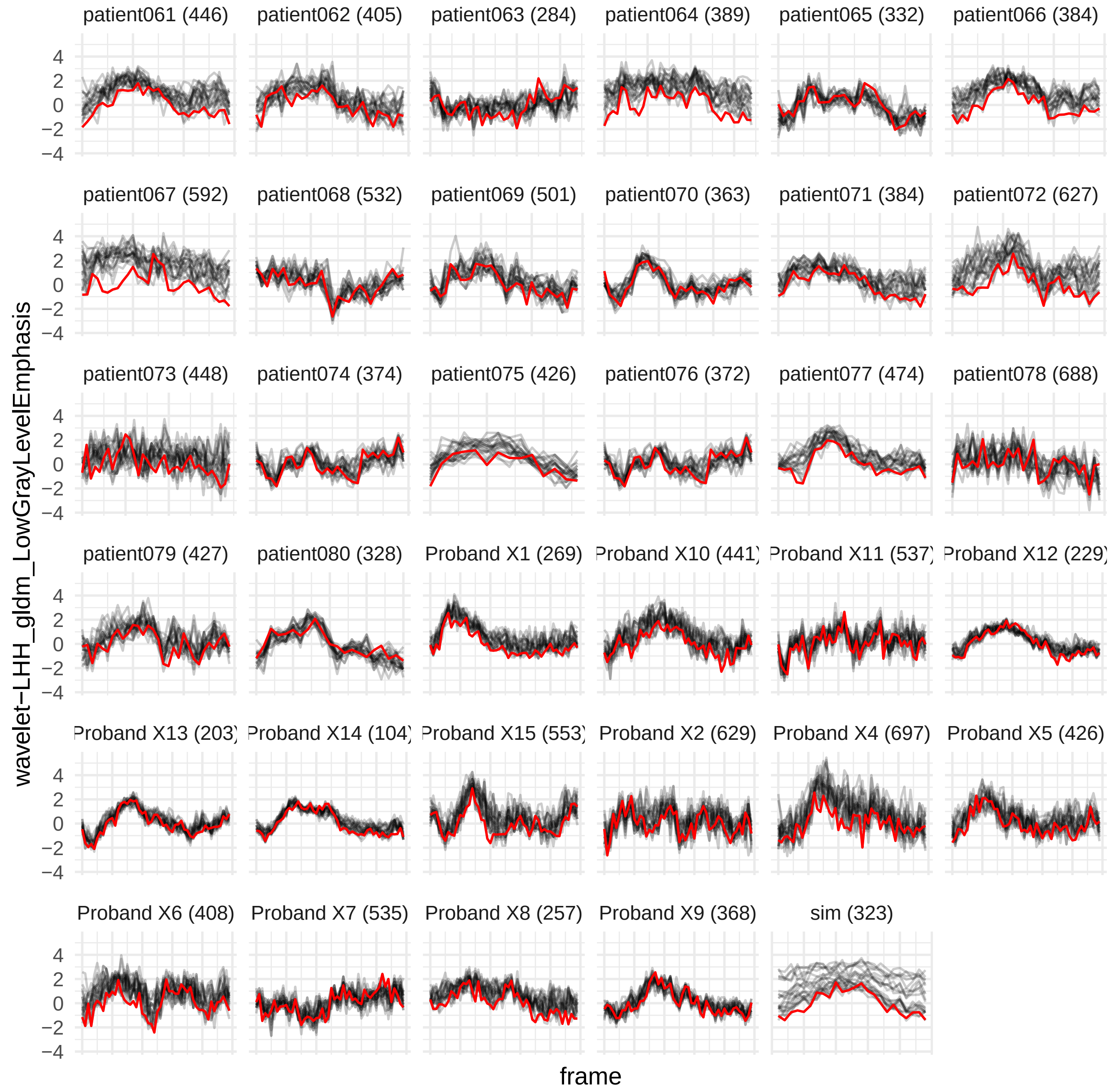
patient061 (446)
patient062 (405)
patient063 (284)
patient064 (389)
patient065 (332)
patient066 (384)
patient067 (592)
patient068 (532)
patient069 (501)
patient070 (363)
patient071 (384)
patient072 (627)
patient073 (448)
patient074 (374)
patient075 (426)
patient076 (372)
patient077 (474)
patient078 (688)
patient079 (427)
patient080 (328)
Proband X1 (269)
Proband X10 (441)
Proband X11 (537)
Proband X12 (229)
Proband X13 (203)
Proband X14 (104)
Proband X15 (553)
Proband X2 (629)
Proband X4 (697)
Proband X5 (426)
Proband X6 (408)
Proband X7 (535)
Proband X8 (257)
Proband X9 (368)
sim (323)
wavelet−LHH_gldm_LowGrayLevelEmphasis
frame
4
2
0
−2
−4

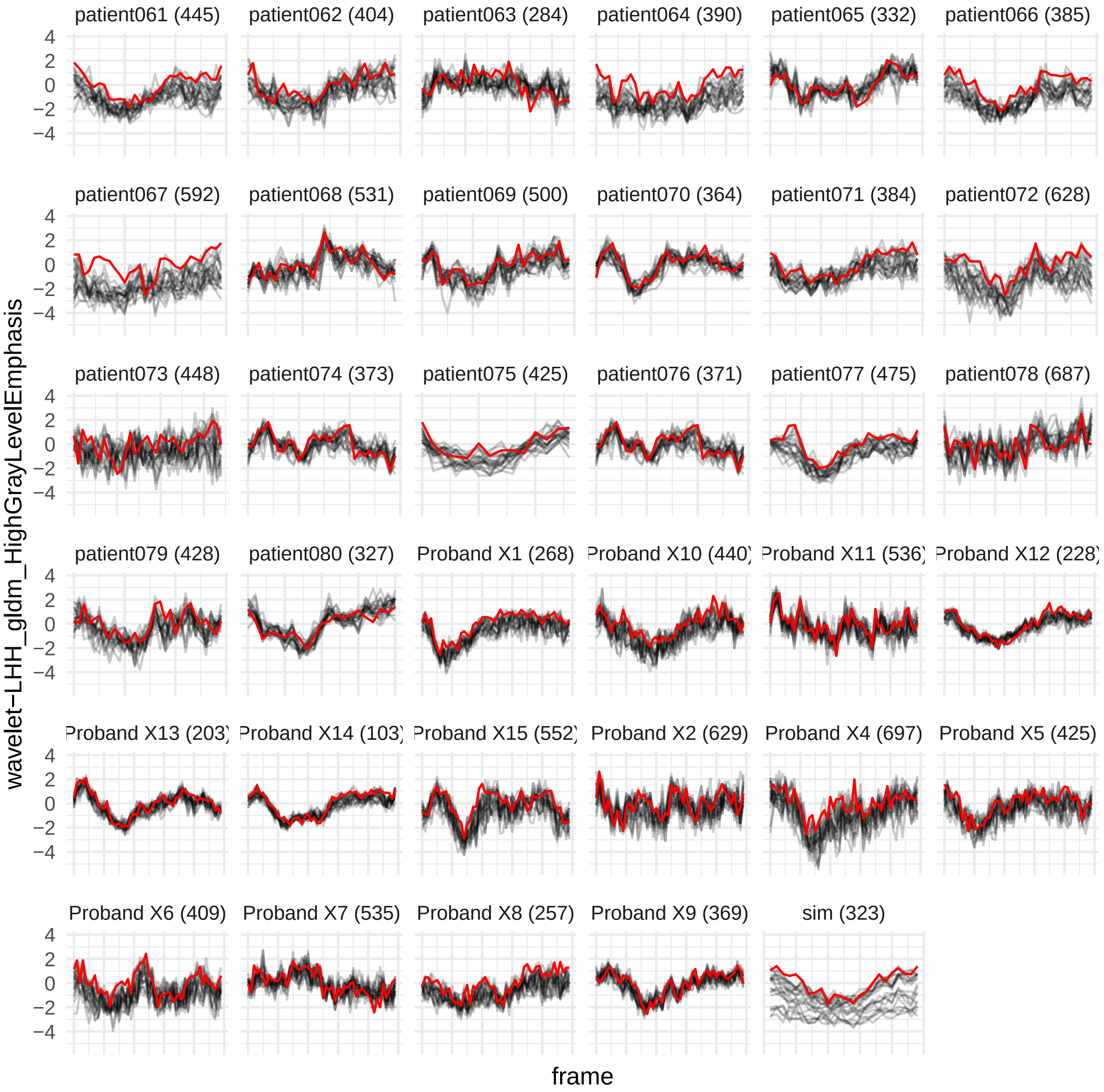
patient061 (445)
patient062 (404)
patient063 (284)
patient064 (390)
patient065 (332)
patient066 (385)
patient067 (592)
patient068 (531)
patient069 (500)
patient070 (364)
patient071 (384)
patient072 (628)
patient073 (448)
patient074 (373)
patient075 (425)
patient076 (371)
patient077 (475)
patient078 (687)
patient079 (428)
patient080 (327)
Proband X1 (268)
Proband X10 (440)
Proband X11 (536)
Proband X12 (228)
Proband X13 (203)
Proband X14 (103)
Proband X15 (552)
Proband X2 (629)
Proband X4 (697)
Proband X5 (425)
Proband X6 (409)
Proband X7 (535)
Proband X8 (257)
Proband X9 (369)
sim (323)
4
2
0
−2
−4
wavelet−LHH_gldm_HighGrayLevelEmphasis
frame

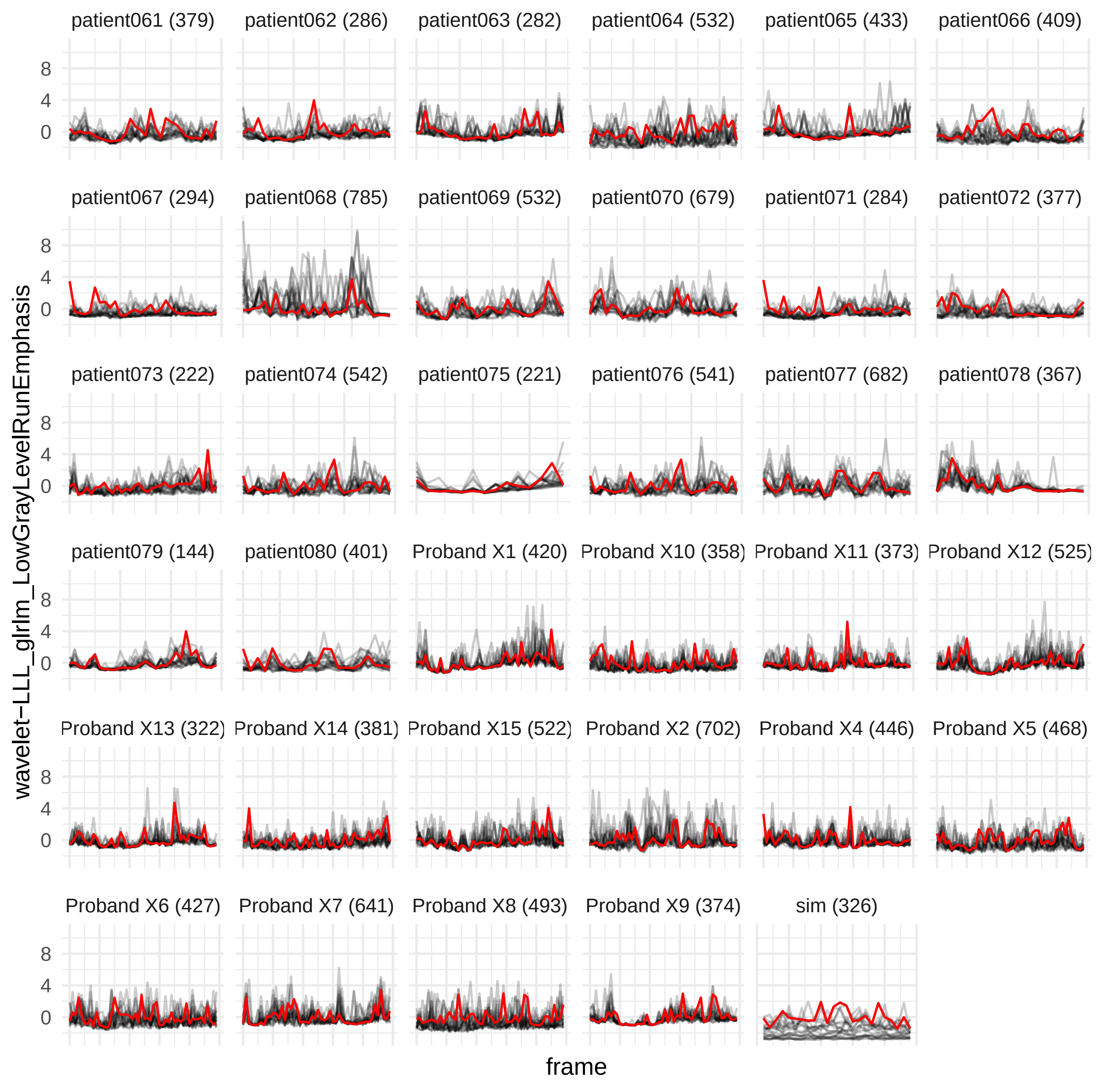
patient061 (379)
patient062 (286)
patient063 (282)
patient064 (532)
patient065 (433)
patient066 (409)
patient067 (294)
patient068 (785)
patient069 (532)
patient070 (679)
patient071 (284)
patient072 (377)
patient073 (222)
patient074 (542)
patient075 (221)
patient076 (541)
patient077 (682)
patient078 (367)
patient079 (144)
patient080 (401)
Proband X1 (420)
Proband X10 (358)
Proband X11 (373)
Proband X12 (525)
Proband X13 (322)
Proband X14 (381)
Proband X15 (522)
Proband X2 (702)
Proband X4 (446)
Proband X5 (468)
Proband X6 (427)
Proband X7 (641)
Proband X8 (493)
Proband X9 (374)
sim (326)
wavelet−LLL_glrlm_LowGrayLevelRunEmphasis
frame
8
4
0

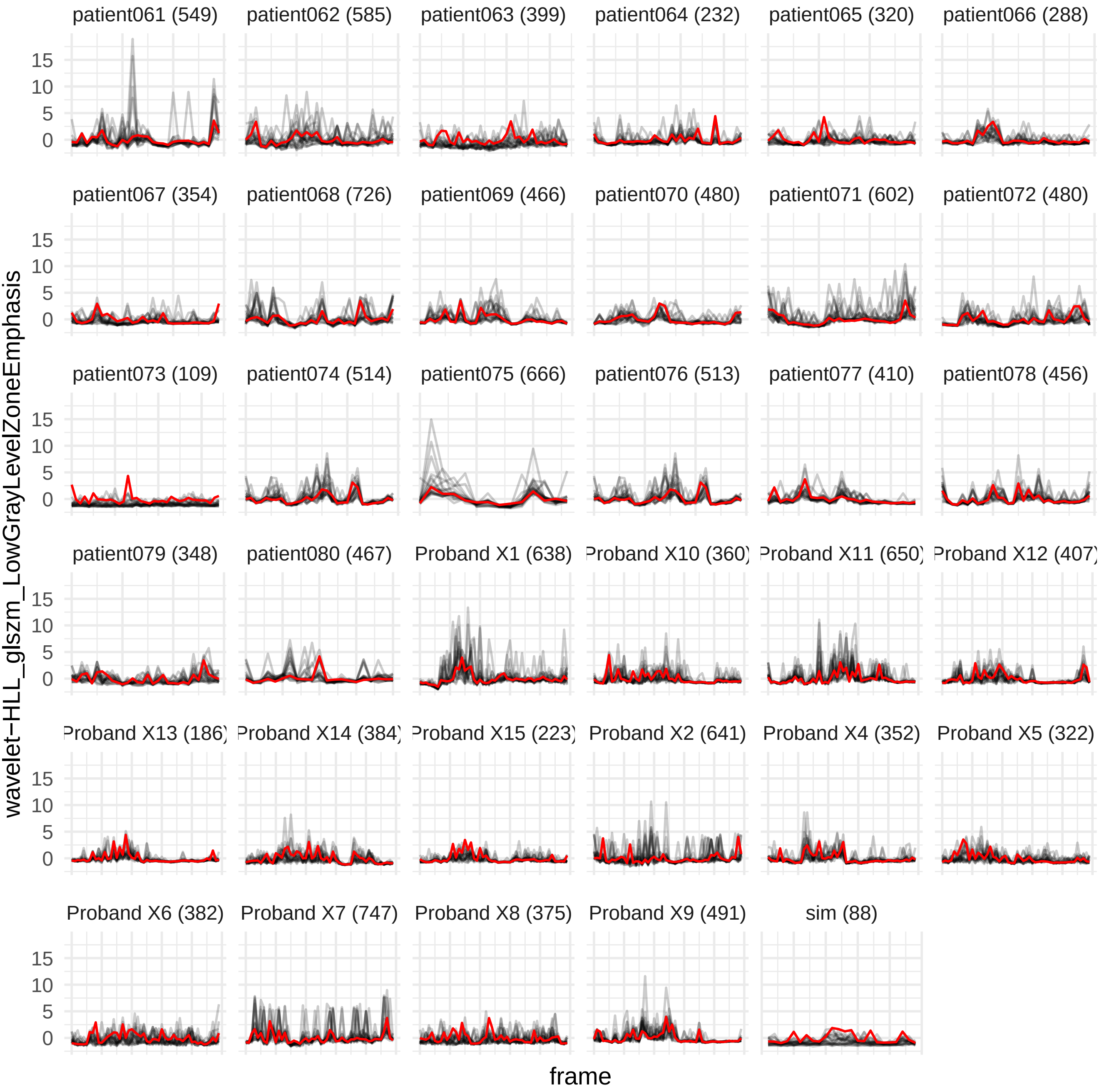

patient061 (549)
patient062 (585)
patient063 (399)
patient064 (232)
patient065 (320)
patient066 (288)
patient067 (354)
patient068 (726)
patient069 (466)
patient070 (480)
patient071 (602)
patient072 (480)
patient073 (109)
patient074 (514)
patient075 (666)
patient076 (513)
patient077 (410)
patient078 (456)
patient079 (348)
patient080 (467)
Proband X1 (638)
Proband X10 (360)
Proband X11 (650)
Proband X12 (407)
Proband X13 (186)
Proband X14 (384)
Proband X15 (223)
Proband X2 (641)
Proband X4 (352)
Proband X5 (322)
Proband X6 (382)
Proband X7 (747)
Proband X8 (375)
Proband X9 (491)
sim (88)
wavelet−HLL_glszm_LowGrayLevelZoneEmphasis
frame
0
5
10
15

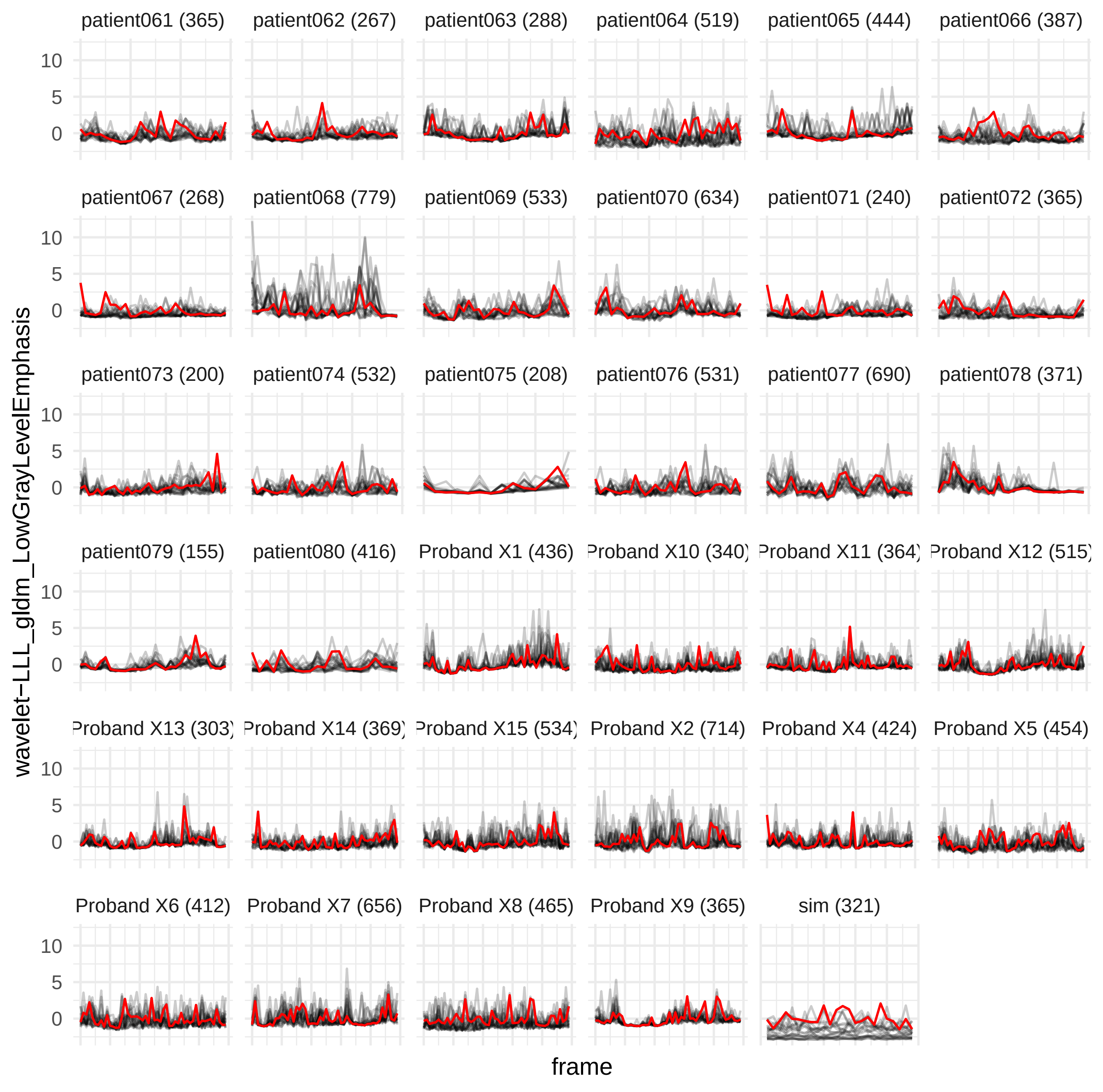

patient061 (365)
patient062 (267)
patient063 (288)
patient064 (519)
patient065 (444)
patient066 (387)
patient067 (268)
patient068 (779)
patient069 (533)
patient070 (634)
patient071 (240)
patient072 (365)
patient073 (200)
patient074 (532)
patient075 (208)
patient076 (531)
patient077 (690)
patient078 (371)
patient079 (155)
patient080 (416)
Proband X1 (436)
Proband X10 (340)
Proband X11 (364)
Proband X12 (515)
Proband X13 (303)
Proband X14 (369)
Proband X15 (534)
Proband X2 (714)
Proband X4 (424)
Proband X5 (454)
Proband X6 (412)
Proband X7 (656)
Proband X8 (465)
Proband X9 (365)
sim (321)
10
5
0
wavelet−LLL_gldm_LowGrayLevelEmphasis
frame

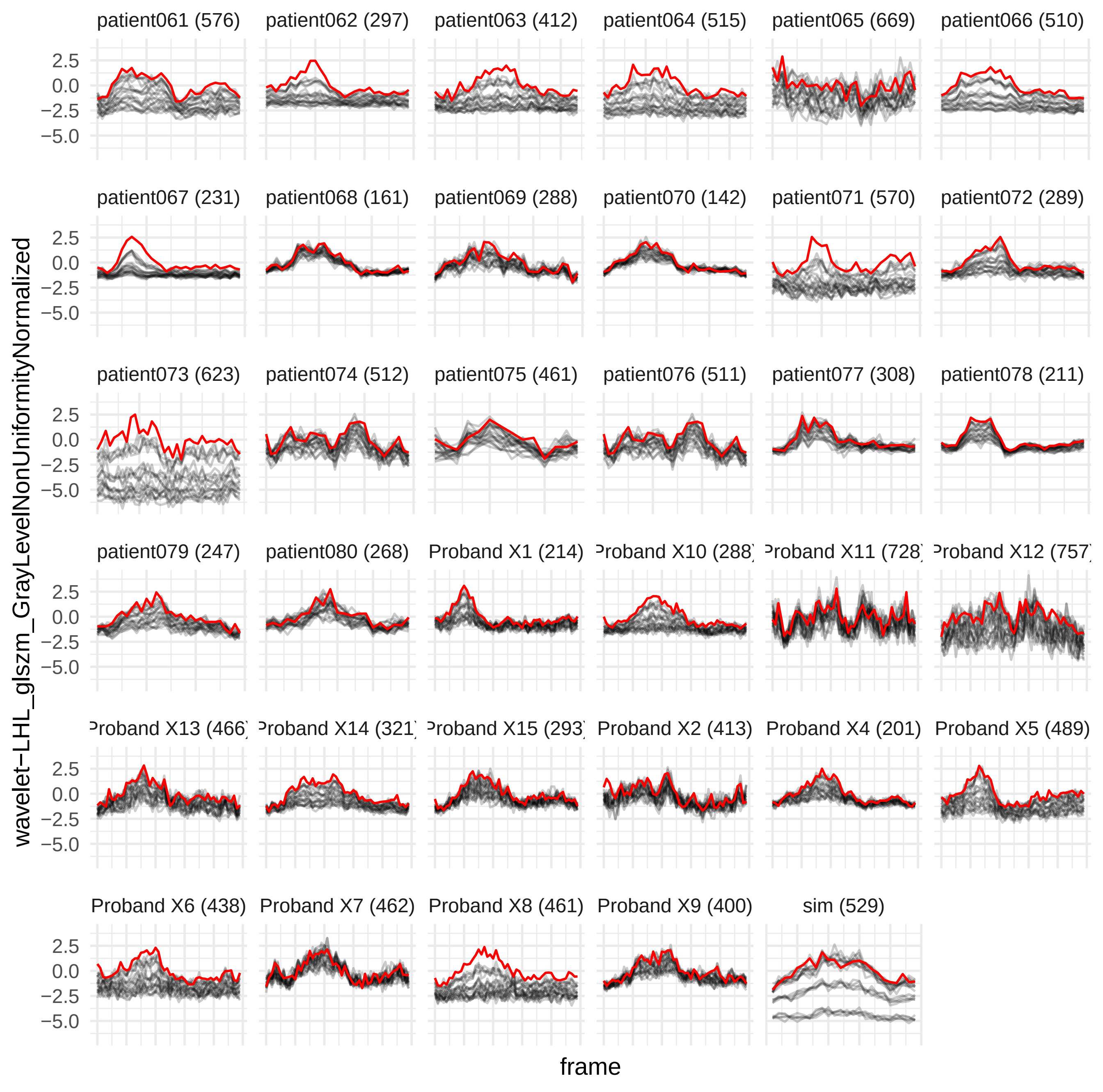

patient061 (576)
patient062 (297)
patient063 (412)
patient064 (515)
patient065 (669)
patient066 (510)
patient067 (231)
patient068 (161)
patient069 (288)
patient070 (142)
patient071 (570)
patient072 (289)
patient073 (623)
patient074 (512)
patient075 (461)
patient076 (511)
patient077 (308)
patient078 (211)
patient079 (247)
patient080 (268)
Proband X1 (214)
Proband X10 (288)
Proband X11 (728)
Proband X12 (757)
Proband X13 (466)
Proband X14 (321)
Proband X15 (293)
Proband X2 (413)
Proband X4 (201)
Proband X5 (489)
Proband X6 (438)
Proband X7 (462)
Proband X8 (461)
Proband X9 (400)
sim (529)
2.5
0.0
−2.5
−5.0
wavelet−LHL_glszm_GrayLevelNonUniformityNormalized
frame

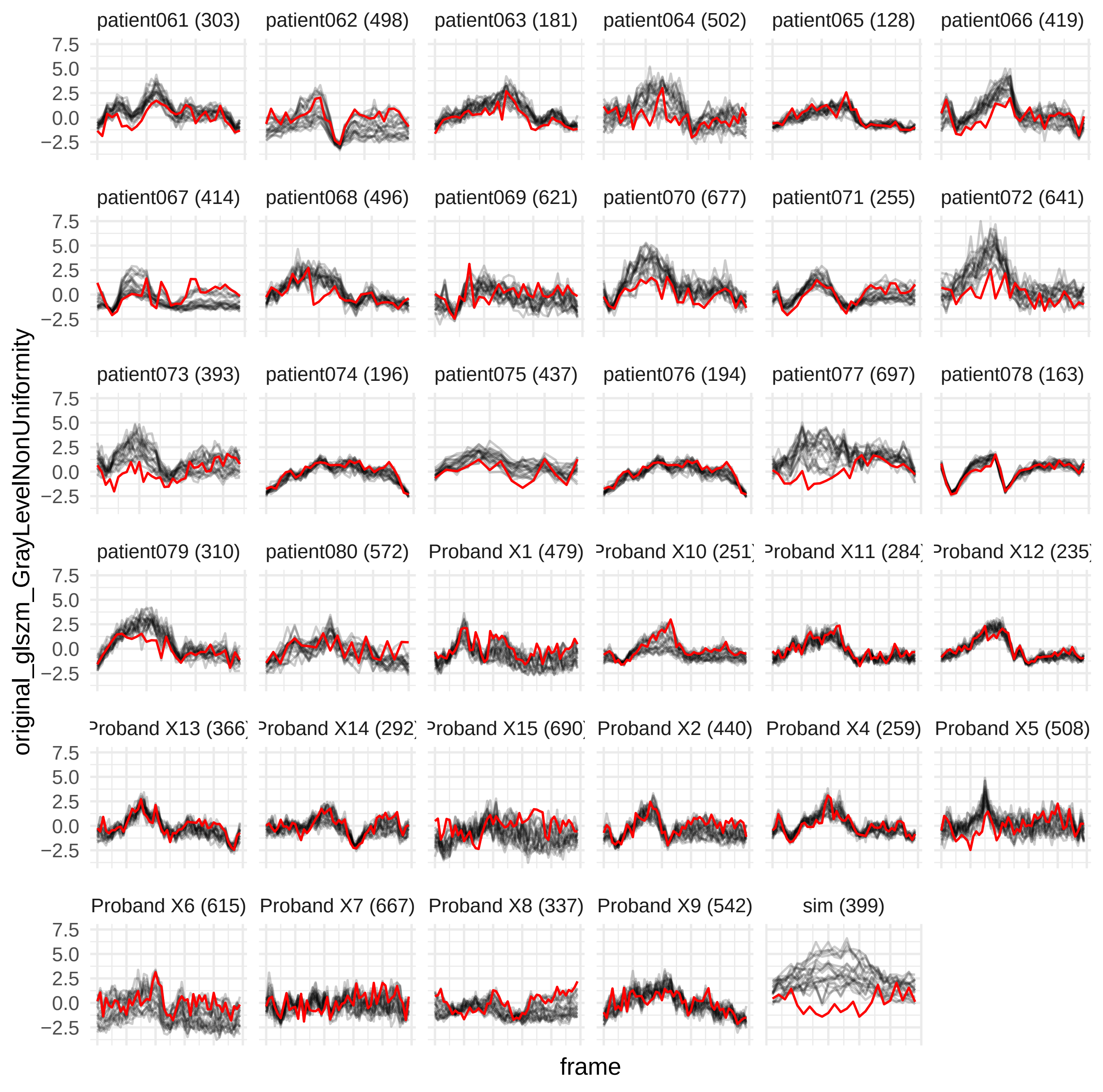

patient061 (303)
patient062 (498)
patient063 (181)
patient064 (502)
patient065 (128)
patient066 (419)
patient067 (414)
patient068 (496)
patient069 (621)
patient070 (677)
patient071 (255)
patient072 (641)
patient073 (393)
patient074 (196)
patient075 (437)
patient076 (194)
patient077 (697)
patient078 (163)
patient079 (310)
patient080 (572)
Proband X1 (479)
Proband X10 (251)
Proband X11 (284)
Proband X12 (235)
Proband X13 (366)
Proband X14 (292)
Proband X15 (690)
Proband X2 (440)
Proband X4 (259)
Proband X5 (508)
Proband X6 (615)
Proband X7 (667)
Proband X8 (337)
Proband X9 (542)
sim (399)
7.5
5.0
2.5
0.0
−2.5
original_glszm_GrayLevelNonUniformity
frame

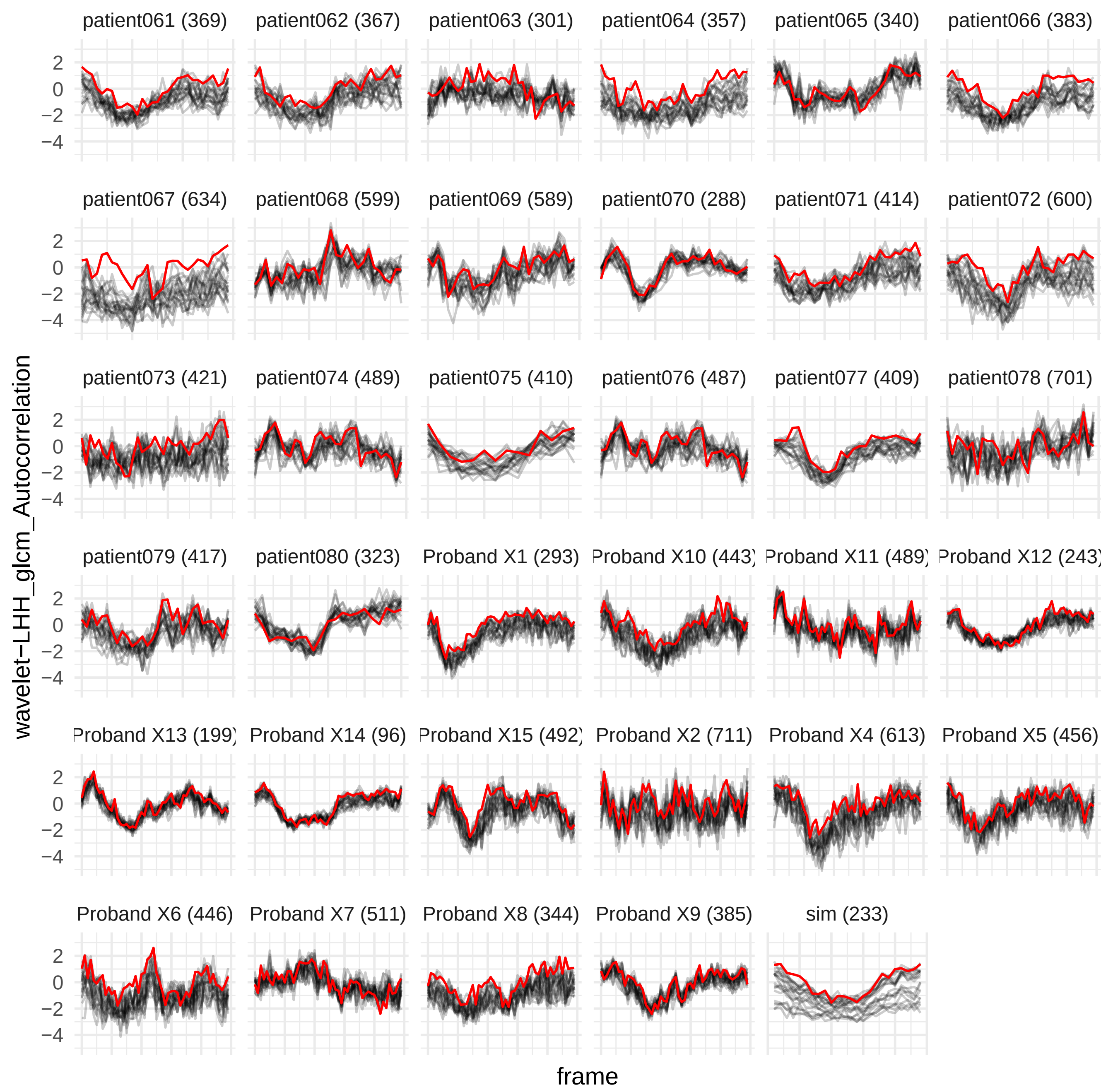

patient061 (369)
patient062 (367)
patient063 (301)
patient064 (357)
patient065 (340)
patient066 (383)
patient067 (634)
patient068 (599)
patient069 (589)
patient070 (288)
patient071 (414)
patient072 (600)
patient073 (421)
patient074 (489)
patient075 (410)
patient076 (487)
patient077 (409)
patient078 (701)
patient079 (417)
patient080 (323)
Proband X1 (293)
Proband X10 (443)
Proband X11 (489)
Proband X12 (243)
Proband X13 (199)
Proband X14 (96)
Proband X15 (492)
Proband X2 (711)
Proband X4 (613)
Proband X5 (456)
Proband X6 (446)
Proband X7 (511)
Proband X8 (344)
Proband X9 (385)
sim (233)
wavelet−LHH_glcm_Autocorrelation
frame
2
0
−2
−4

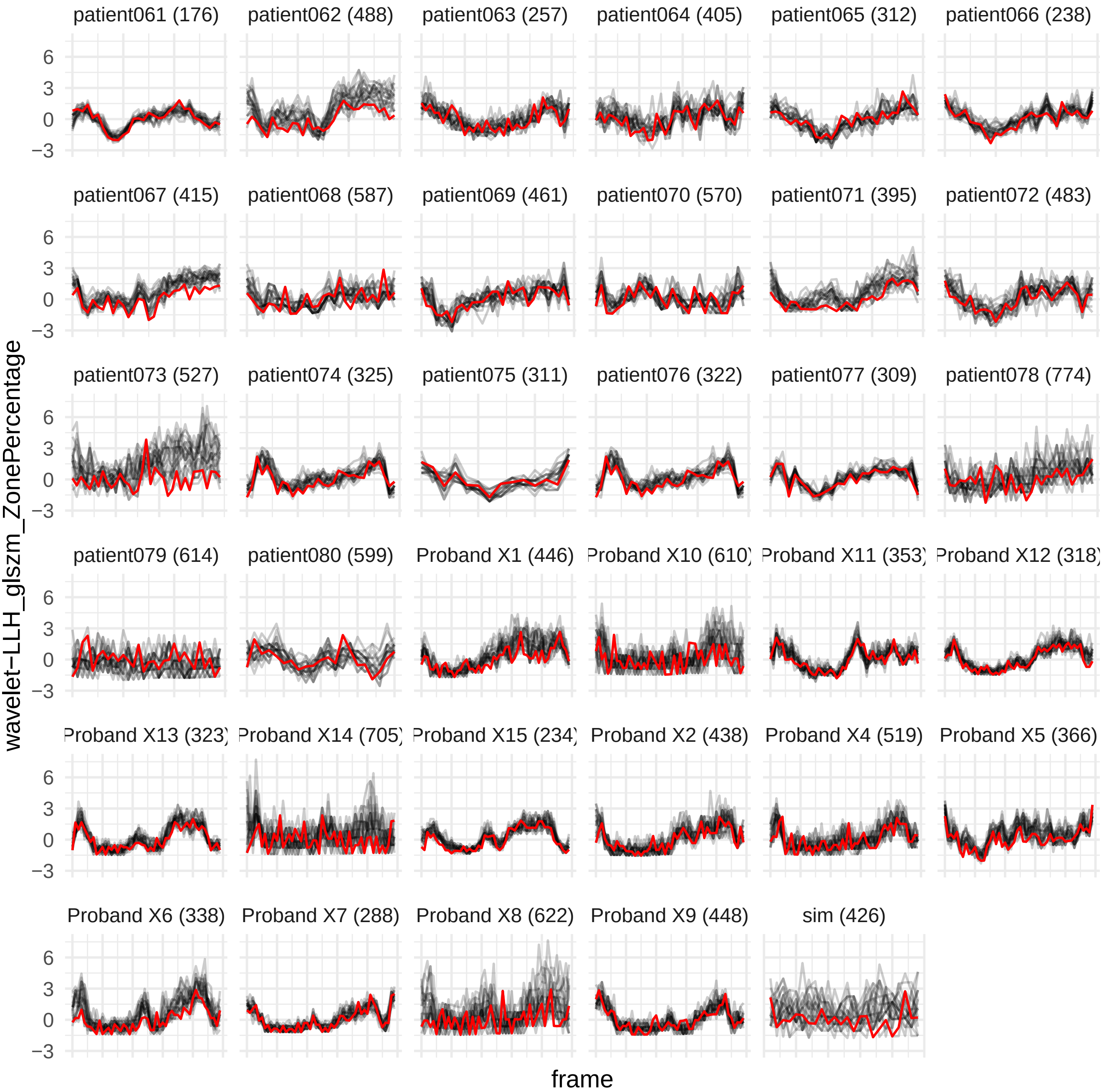

patient061 (176)
patient062 (488)
patient063 (257)
patient064 (405)
patient065 (312)
patient066 (238)
patient067 (415)
patient068 (587)
patient069 (461)
patient070 (570)
patient071 (395)
patient072 (483)
patient073 (527)
patient074 (325)
patient075 (311)
patient076 (322)
patient077 (309)
patient078 (774)
patient079 (614)
patient080 (599)
Proband X1 (446)
Proband X10 (610)
Proband X11 (353)
Proband X12 (318)
Proband X13 (323)
Proband X14 (705)
Proband X15 (234)
Proband X2 (438)
Proband X4 (519)
Proband X5 (366)
Proband X6 (338)
Proband X7 (288)
Proband X8 (622)
Proband X9 (448)
sim (426)
6
3
0
−3
wavelet−LLH_glszm_ZonePercentage
frame

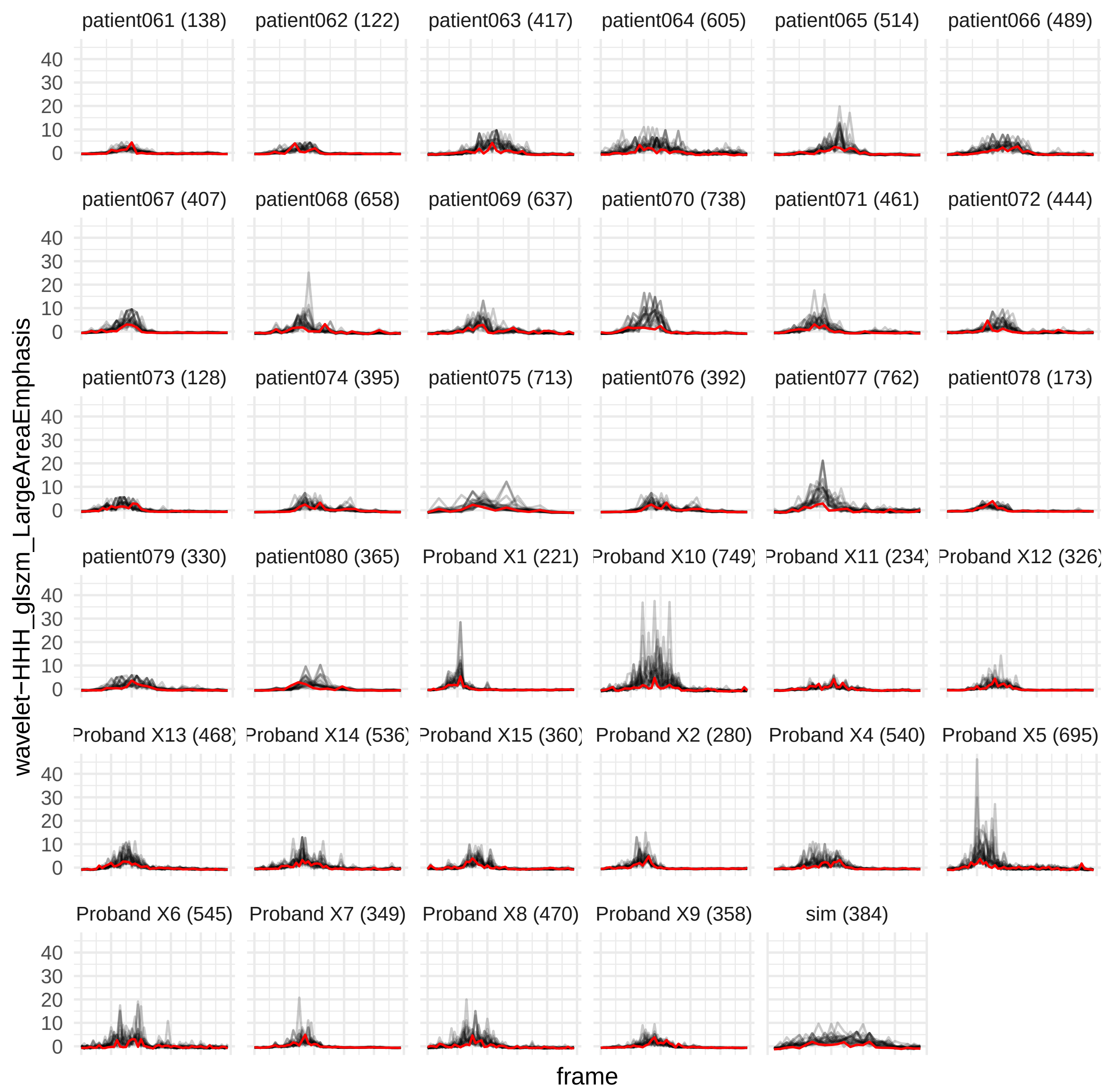

patient061 (138)
patient062 (122)
patient063 (417)
patient064 (605)
patient065 (514)
patient066 (489)
patient067 (407)
patient068 (658)
patient069 (637)
patient070 (738)
patient071 (461)
patient072 (444)
patient073 (128)
patient074 (395)
patient075 (713)
patient076 (392)
patient077 (762)
patient078 (173)
patient079 (330)
patient080 (365)
Proband X1 (221)
Proband X10 (749)
Proband X11 (234)
Proband X12 (326)
Proband X13 (468)
Proband X14 (536)
Proband X15 (360)
Proband X2 (280)
Proband X4 (540)
Proband X5 (695)
Proband X6 (545)
Proband X7 (349)
Proband X8 (470)
Proband X9 (358)
sim (384)
wavelet−HHH_glszm_LargeAreaEmphasis
0
10
20
30
40
frame

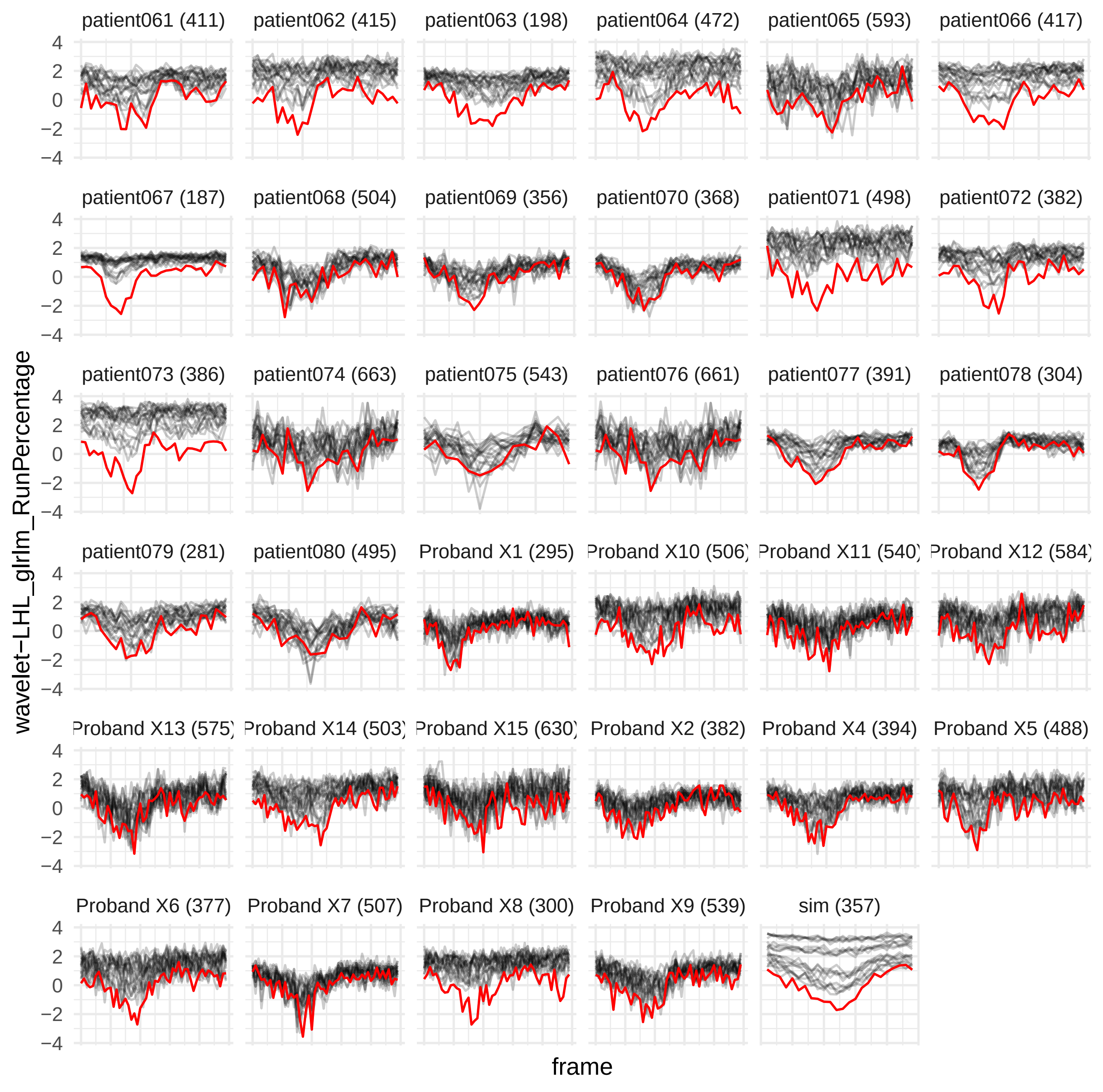

patient061 (411)
patient062 (415)
patient063 (198)
patient064 (472)
patient065 (593)
patient066 (417)
patient067 (187)
patient068 (504)
patient069 (356)
patient070 (368)
patient071 (498)
patient072 (382)
patient073 (386)
patient074 (663)
patient075 (543)
patient076 (661)
patient077 (391)
patient078 (304)
patient079 (281)
patient080 (495)
Proband X1 (295)
Proband X10 (506)
Proband X11 (540)
Proband X12 (584)
Proband X13 (575)
Proband X14 (503)
Proband X15 (630)
Proband X2 (382)
Proband X4 (394)
Proband X5 (488)
Proband X6 (377)
Proband X7 (507)
Proband X8 (300)
Proband X9 (539)
sim (357)
4
2
0
−2
−4
wavelet−LHL_glrlm_RunPercentage
frame

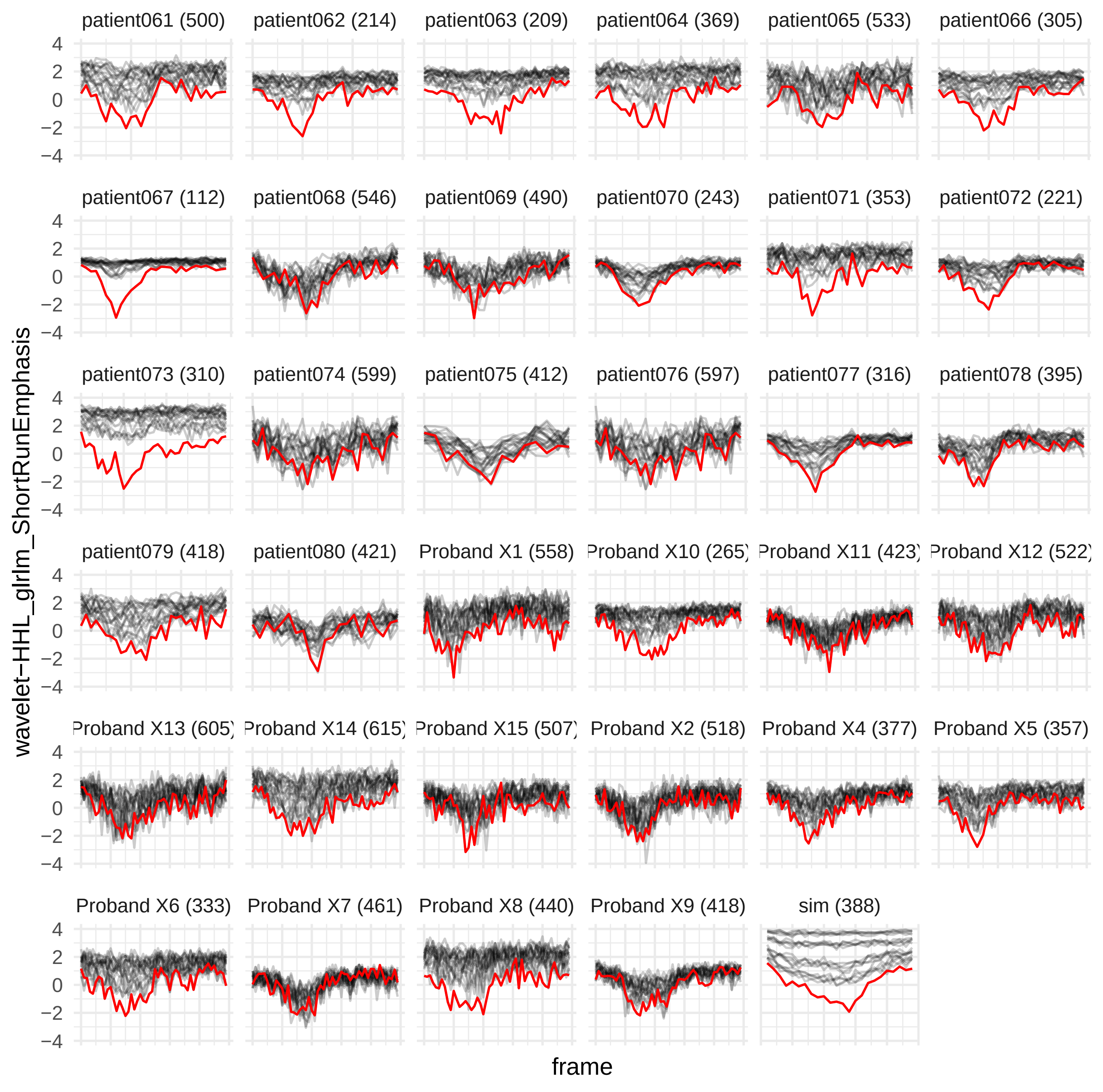
patient061 (500)
patient062 (214)
patient063 (209)
patient064 (369)
patient065 (533)
patient066 (305)
patient067 (112)
patient068 (546)
patient069 (490)
patient070 (243)
patient071 (353)
patient072 (221)
patient073 (310)
patient074 (599)
patient075 (412)
patient076 (597)
patient077 (316)
patient078 (395)
patient079 (418)
patient080 (421)
Proband X1 (558)
Proband X10 (265)
Proband X11 (423)
Proband X12 (522)
Proband X13 (605)
Proband X14 (615)
Proband X15 (507)
Proband X2 (518)
Proband X4 (377)
Proband X5 (357)
Proband X6 (333)
Proband X7 (461)
Proband X8 (440)
Proband X9 (418)
sim (388)
wavelet−HHL_glrlm_ShortRunEmphasis
frame
4
2
0
−2
−4

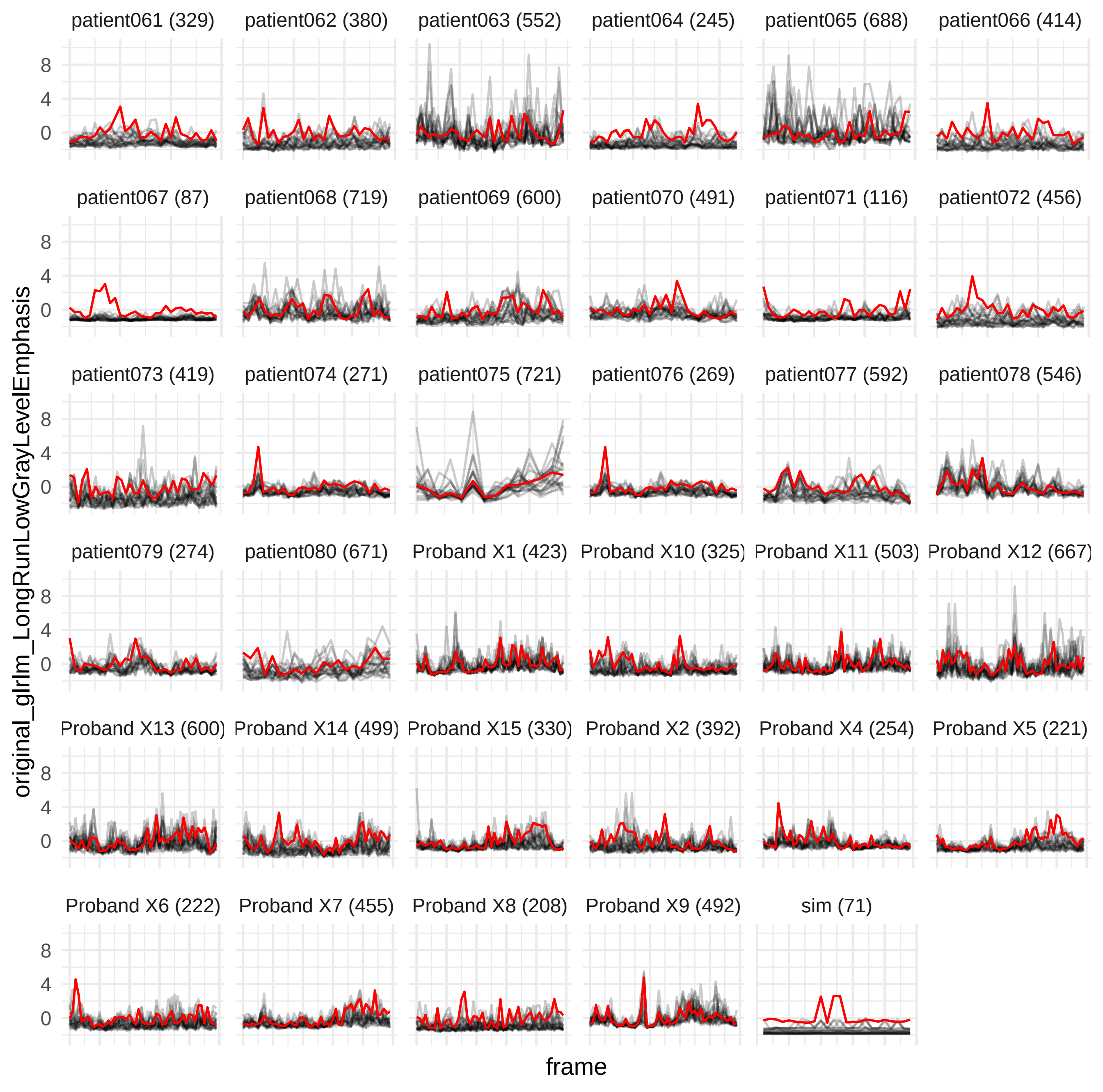

patient061 (329)
patient062 (380)
patient063 (552)
patient064 (245)
patient065 (688)
patient066 (414)
patient067 (87)
patient068 (719)
patient069 (600)
patient070 (491)
patient071 (116)
patient072 (456)
patient073 (419)
patient074 (271)
patient075 (721)
patient076 (269)
patient077 (592)
patient078 (546)
patient079 (274)
patient080 (671)
Proband X1 (423)
Proband X10 (325)
Proband X11 (503)
Proband X12 (667)
Proband X13 (600)
Proband X14 (499)
Proband X15 (330)
Proband X2 (392)
Proband X4 (254)
Proband X5 (221)
Proband X6 (222)
Proband X7 (455)
Proband X8 (208)
Proband X9 (492)
sim (71)
original_glrlm_LongRunLowGrayLevelEmphasis
frame
8
4
0

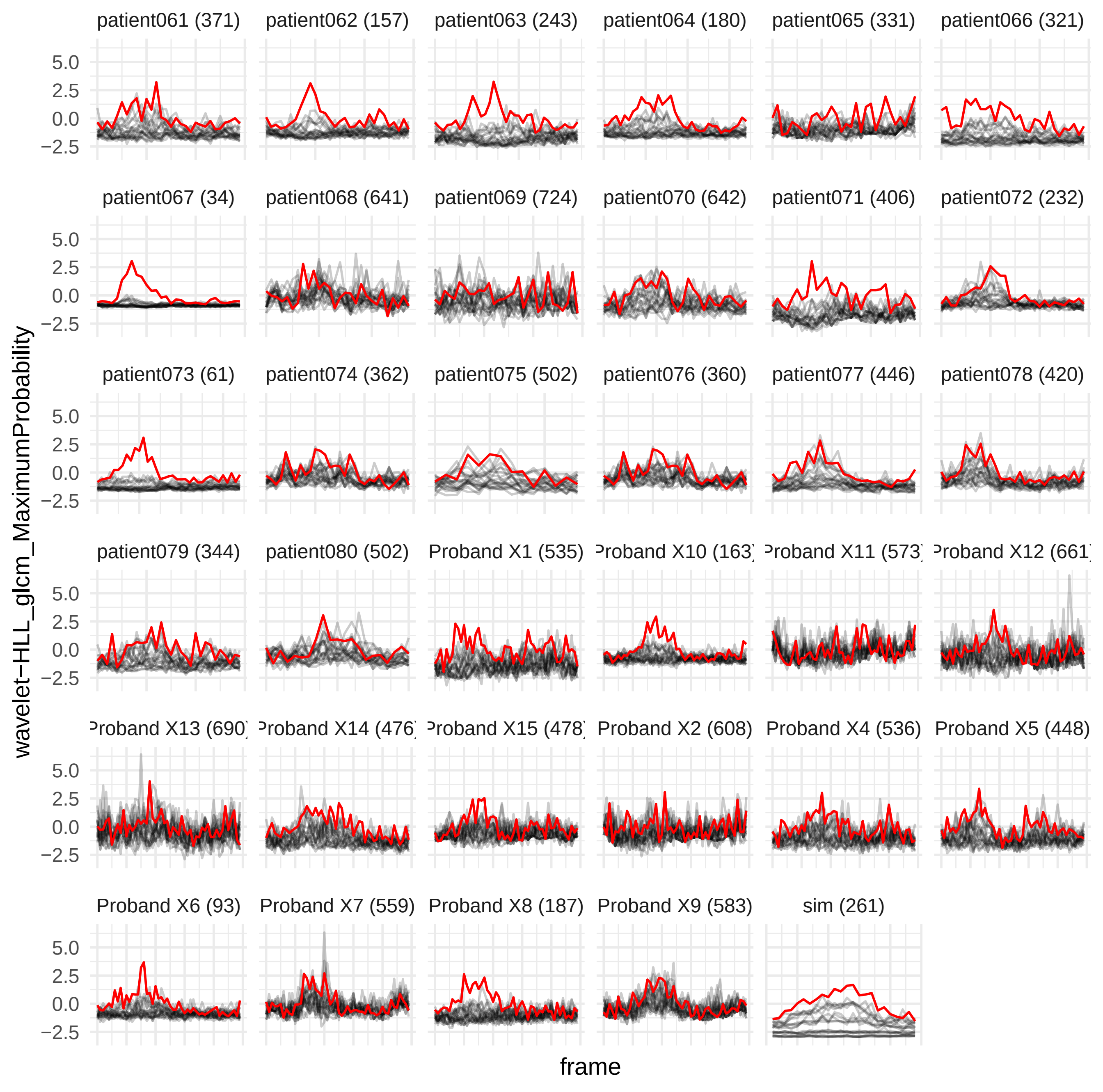

patient061 (371)
patient062 (157)
patient063 (243)
patient064 (180)
patient065 (331)
patient066 (321)
patient067 (34)
patient068 (641)
patient069 (724)
patient070 (642)
patient071 (406)
patient072 (232)
patient073 (61)
patient074 (362)
patient075 (502)
patient076 (360)
patient077 (446)
patient078 (420)
patient079 (344)
patient080 (502)
Proband X1 (535)
Proband X10 (163)
Proband X11 (573)
Proband X12 (661)
Proband X13 (690)
Proband X14 (476)
Proband X15 (478)
Proband X2 (608)
Proband X4 (536)
Proband X5 (448)
Proband X6 (93)
Proband X7 (559)
Proband X8 (187)
Proband X9 (583)
sim (261)
5.0
2.5
0.0
−2.5
wavelet−HLL_glcm_MaximumProbability
frame

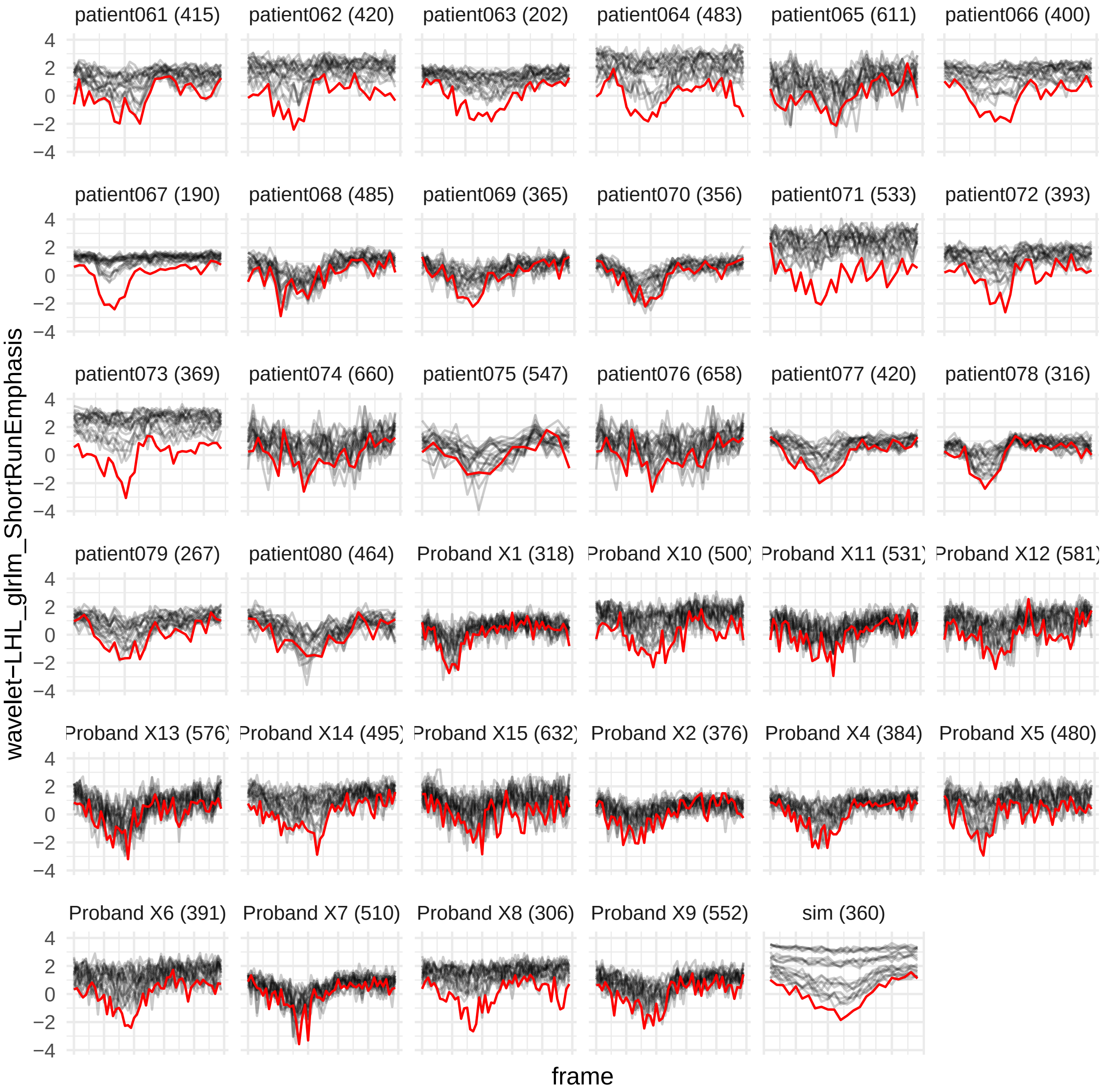
patient061 (415)
patient062 (420)
patient063 (202)
patient064 (483)
patient065 (611)
patient066 (400)
patient067 (190)
patient068 (485)
patient069 (365)
patient070 (356)
patient071 (533)
patient072 (393)
patient073 (369)
patient074 (660)
patient075 (547)
patient076 (658)
patient077 (420)
patient078 (316)
patient079 (267)
patient080 (464)
Proband X1 (318)
Proband X10 (500)
Proband X11 (531)
Proband X12 (581)
Proband X13 (576)
Proband X14 (495)
Proband X15 (632)
Proband X2 (376)
Proband X4 (384)
Proband X5 (480)
Proband X6 (391)
Proband X7 (510)
Proband X8 (306)
Proband X9 (552)
sim (360)
wavelet−LHL_glrlm_ShortRunEmphasis
frame
4
2
0
−2
−4

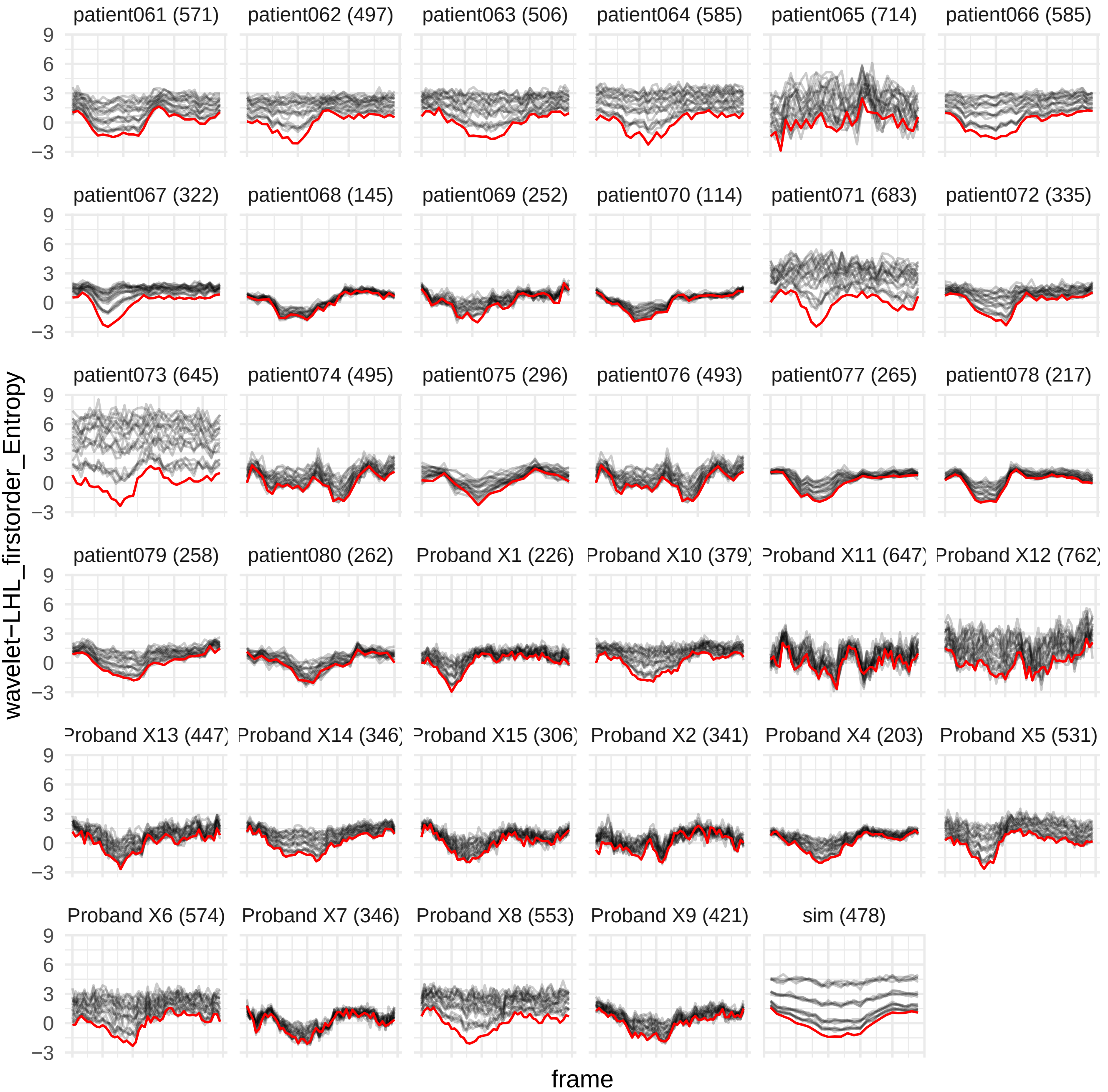

patient061 (571)
patient062 (497)
patient063 (506)
patient064 (585)
patient065 (714)
patient066 (585)
patient067 (322)
patient068 (145)
patient069 (252)
patient070 (114)
patient071 (683)
patient072 (335)
patient073 (645)
patient074 (495)
patient075 (296)
patient076 (493)
patient077 (265)
patient078 (217)
patient079 (258)
patient080 (262)
Proband X1 (226)
Proband X10 (379)
Proband X11 (647)
Proband X12 (762)
Proband X13 (447)
Proband X14 (346)
Proband X15 (306)
Proband X2 (341)
Proband X4 (203)
Proband X5 (531)
Proband X6 (574)
Proband X7 (346)
Proband X8 (553)
Proband X9 (421)
sim (478)
wavelet−LHL_firstorder_Entropy
frame
9
6
3
0
−3

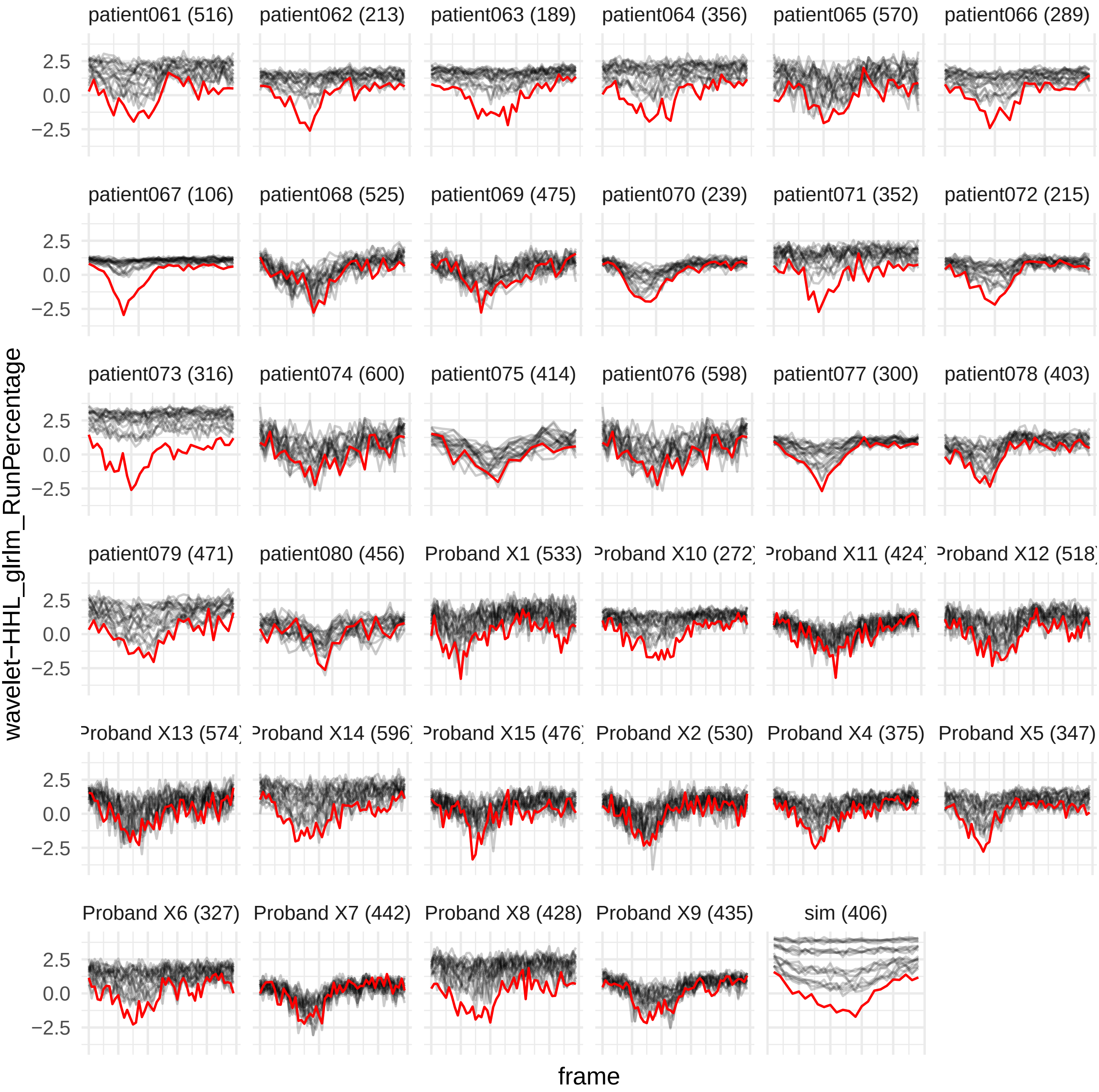

patient061 (516)
patient062 (213)
patient063 (189)
patient064 (356)
patient065 (570)
patient066 (289)
patient067 (106)
patient068 (525)
patient069 (475)
patient070 (239)
patient071 (352)
patient072 (215)
patient073 (316)
patient074 (600)
patient075 (414)
patient076 (598)
patient077 (300)
patient078 (403)
patient079 (471)
patient080 (456)
Proband X1 (533)
Proband X10 (272)
Proband X11 (424)
Proband X12 (518)
Proband X13 (574)
Proband X14 (596)
Proband X15 (476)
Proband X2 (530)
Proband X4 (375)
Proband X5 (347)
Proband X6 (327)
Proband X7 (442)
Proband X8 (428)
Proband X9 (435)
sim (406)
2.5
0.0
−2.5
wavelet−HHL_glrlm_RunPercentage
frame

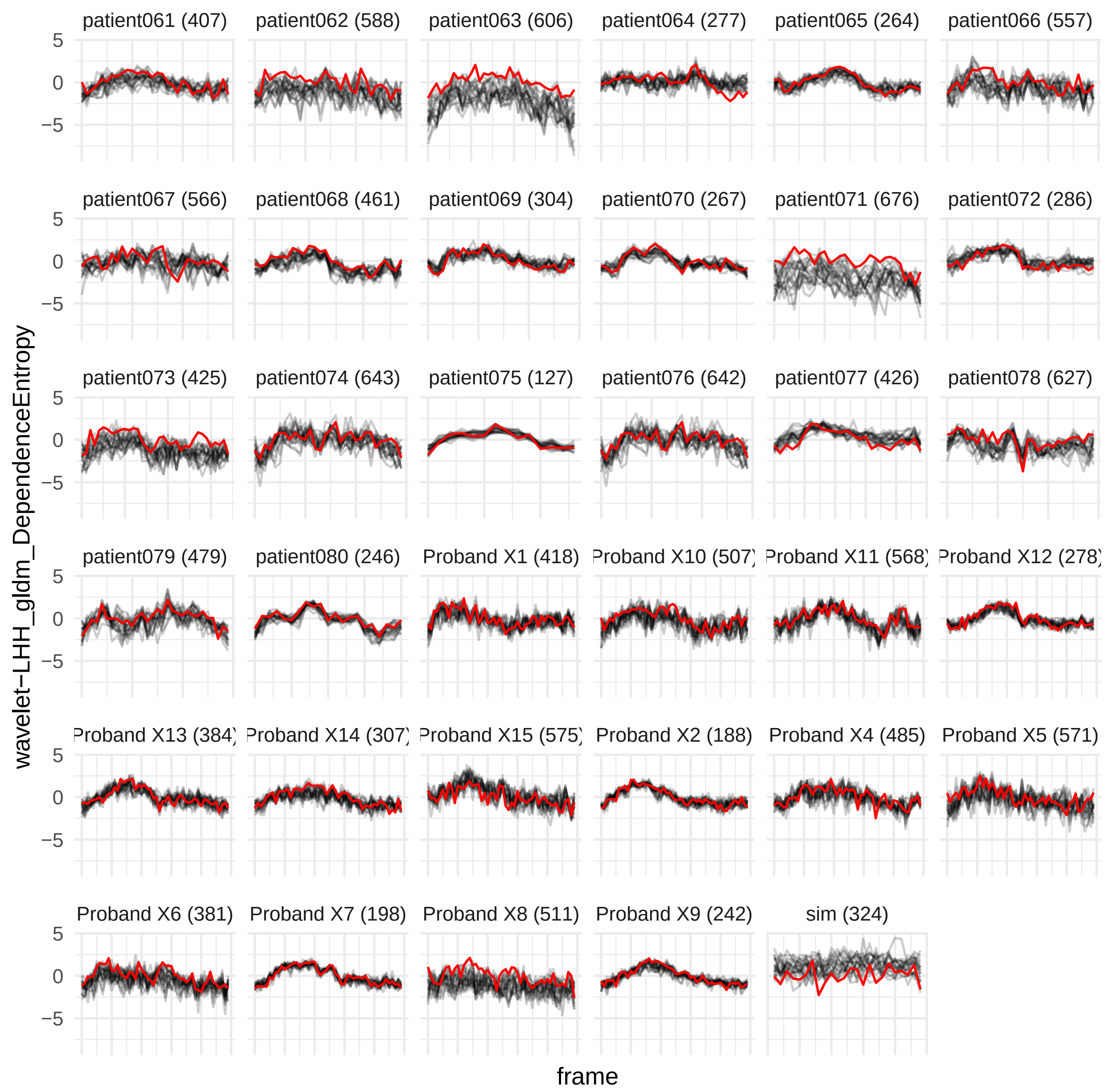
patient061 (407)
patient062 (588)
patient063 (606)
patient064 (277)
patient065 (264)
patient066 (557)
patient067 (566)
patient068 (461)
patient069 (304)
patient070 (267)
patient071 (676)
patient072 (286)
patient073 (425)
patient074 (643)
patient075 (127)
patient076 (642)
patient077 (426)
patient078 (627)
patient079 (479)
patient080 (246)
Proband X1 (418)
Proband X10 (507)
Proband X11 (568)
Proband X12 (278)
Proband X13 (384)
Proband X14 (307)
Proband X15 (575)
Proband X2 (188)
Proband X4 (485)
Proband X5 (571)
Proband X6 (381)
Proband X7 (198)
Proband X8 (511)
Proband X9 (242)
sim (324)
5
0
−5
wavelet−LHH_gldm_DependenceEntropy
frame

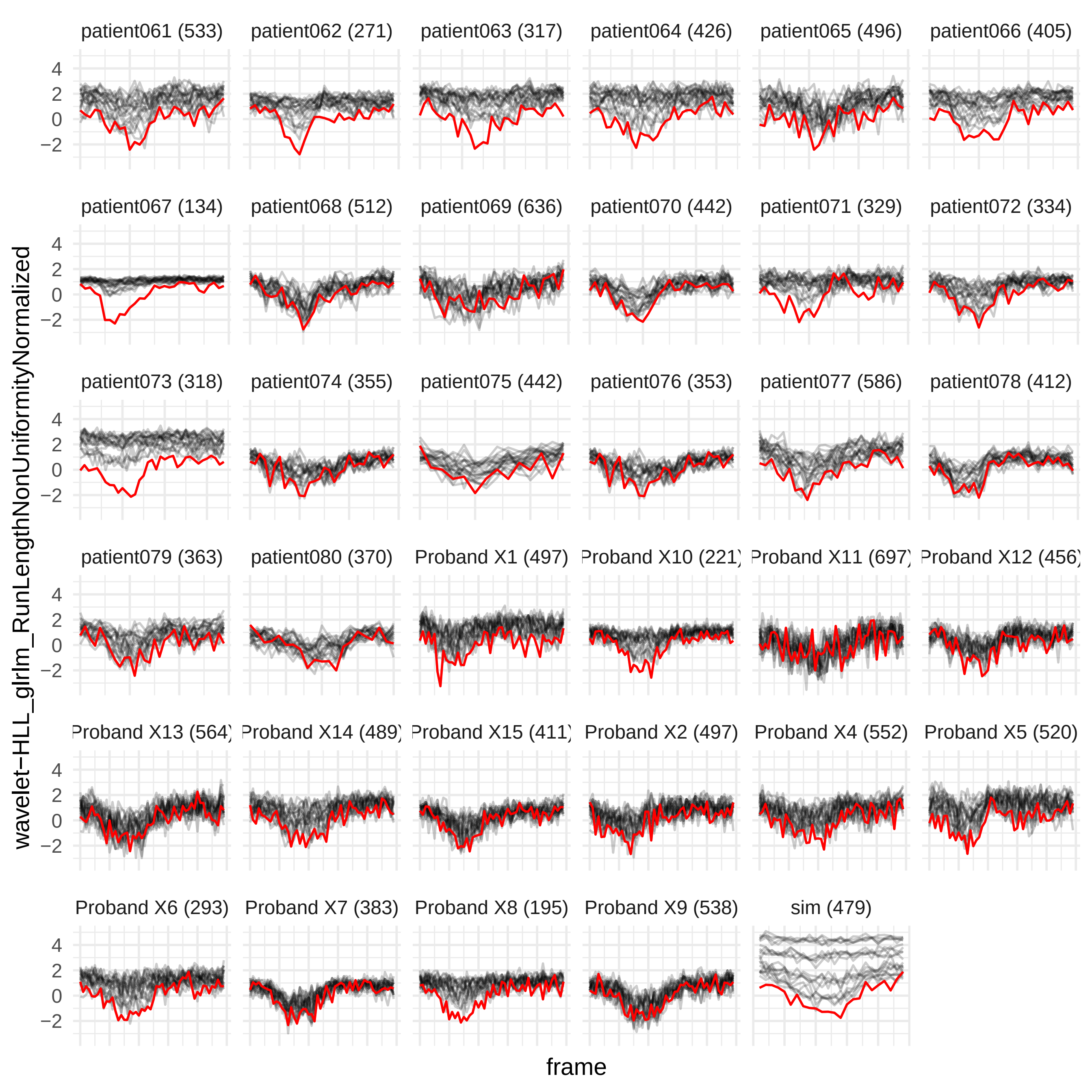
patient061 (533)
patient062 (271)
patient063 (317)
patient064 (426)
patient065 (496)
patient066 (405)
patient067 (134)
patient068 (512)
patient069 (636)
patient070 (442)
patient071 (329)
patient072 (334)
patient073 (318)
patient074 (355)
patient075 (442)
patient076 (353)
patient077 (586)
patient078 (412)
patient079 (363)
patient080 (370)
Proband X1 (497)
Proband X10 (221)
Proband X11 (697)
Proband X12 (456)
Proband X13 (564)
Proband X14 (489)
Proband X15 (411)
Proband X2 (497)
Proband X4 (552)
Proband X5 (520)
Proband X6 (293)
Proband X7 (383)
Proband X8 (195)
Proband X9 (538)
sim (479)
4
2
0
−2
wavelet−HLL_glrlm_RunLengthNonUniformityNormalized
frame

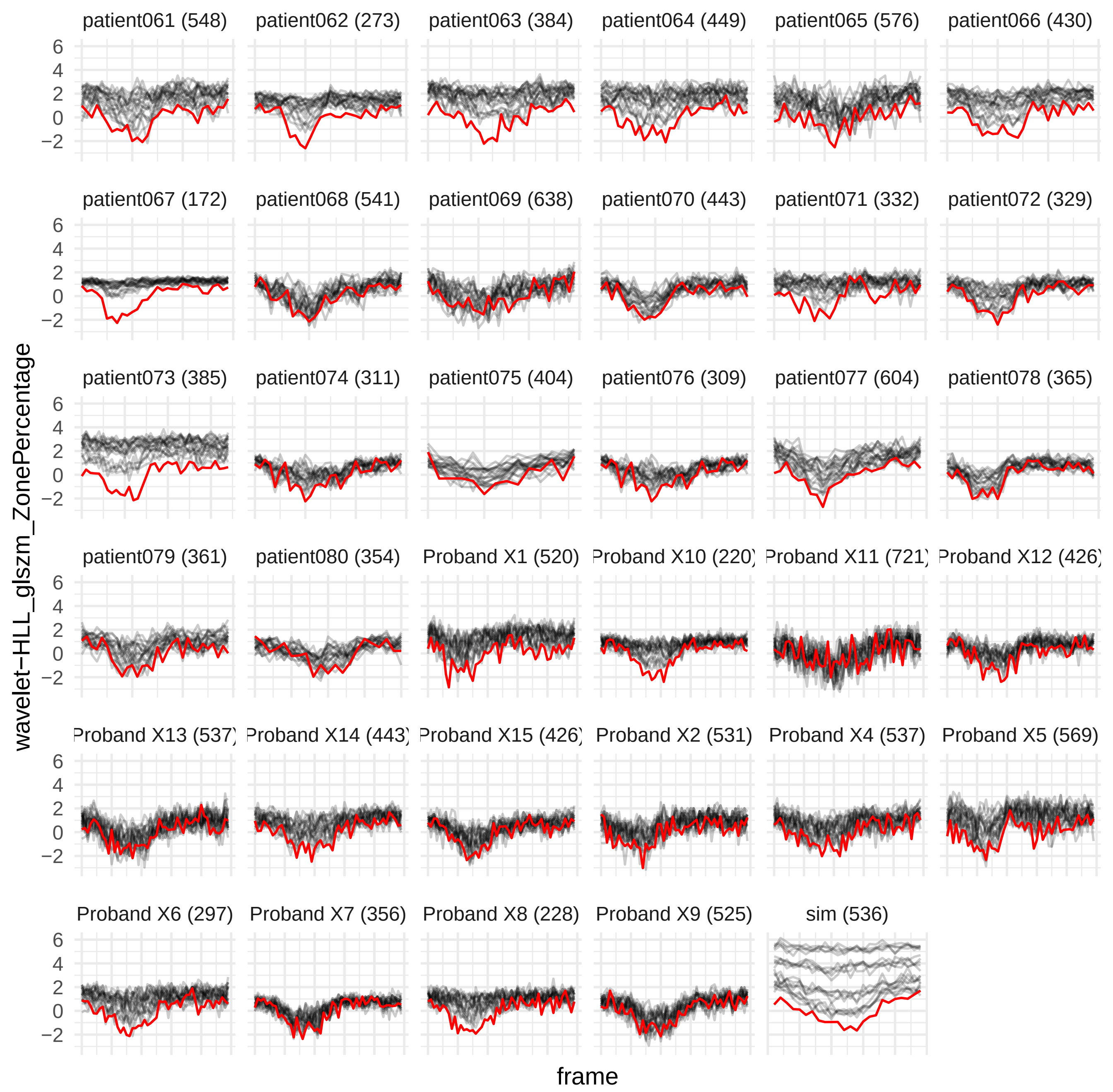
patient061 (548)
patient062 (273)
patient063 (384)
patient064 (449)
patient065 (576)
patient066 (430)
patient067 (172)
patient068 (541)
patient069 (638)
patient070 (443)
patient071 (332)
patient072 (329)
patient073 (385)
patient074 (311)
patient075 (404)
patient076 (309)
patient077 (604)
patient078 (365)
patient079 (361)
patient080 (354)
Proband X1 (520)
Proband X10 (220)
Proband X11 (721)
Proband X12 (426)
Proband X13 (537)
Proband X14 (443)
Proband X15 (426)
Proband X2 (531)
Proband X4 (537)
Proband X5 (569)
Proband X6 (297)
Proband X7 (356)
Proband X8 (228)
Proband X9 (525)
sim (536)
6
4
2
0
−2
wavelet−HLL_glszm_ZonePercentage
frame

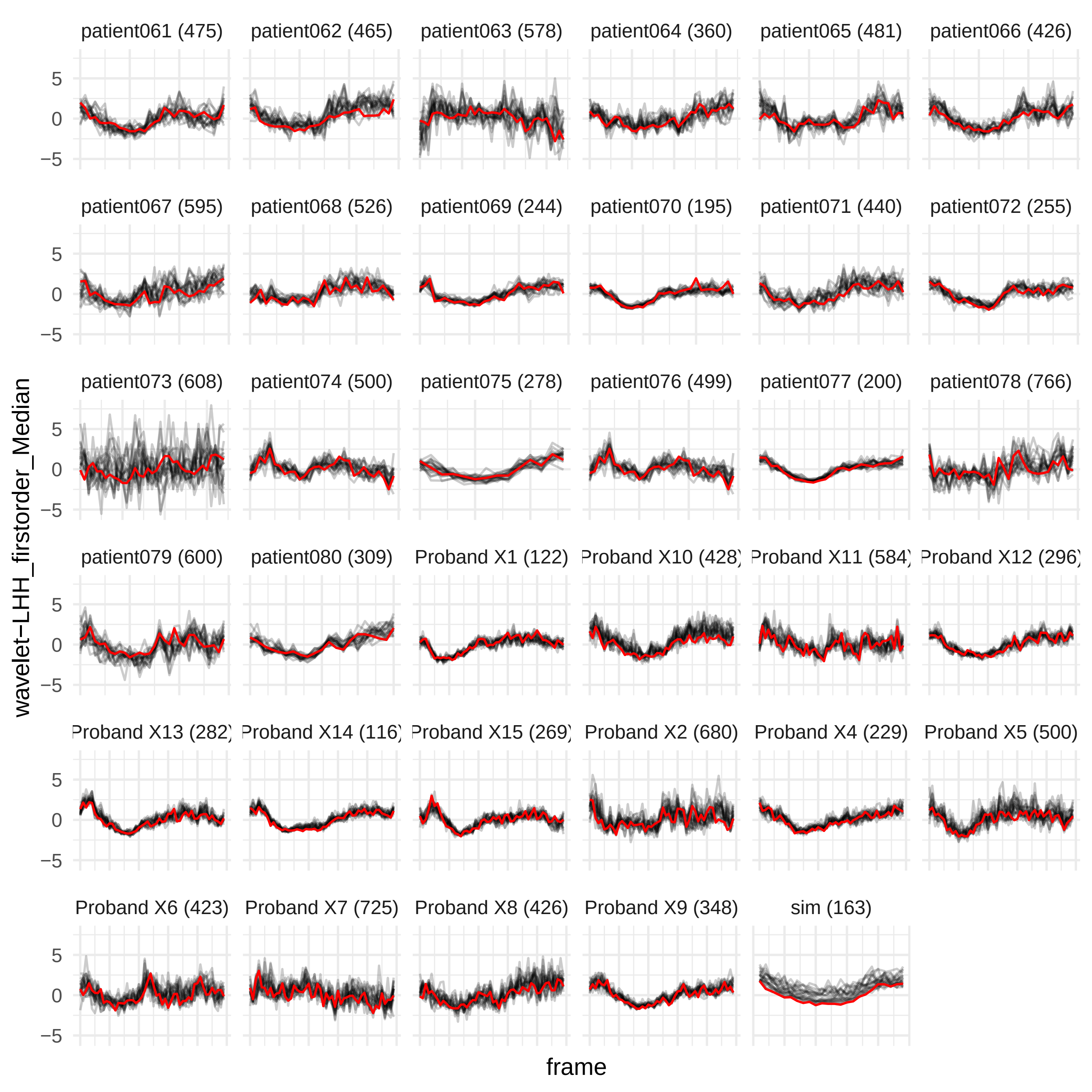
patient061 (475)
patient062 (465)
patient063 (578)
patient064 (360)
patient065 (481)
patient066 (426)
patient067 (595)
patient068 (526)
patient069 (244)
patient070 (195)
patient071 (440)
patient072 (255)
patient073 (608)
patient074 (500)
patient075 (278)
patient076 (499)
patient077 (200)
patient078 (766)
patient079 (600)
patient080 (309)
Proband X1 (122)
Proband X10 (428)
Proband X11 (584)
Proband X12 (296)
Proband X13 (282)
Proband X14 (116)
Proband X15 (269)
Proband X2 (680)
Proband X4 (229)
Proband X5 (500)
Proband X6 (423)
Proband X7 (725)
Proband X8 (426)
Proband X9 (348)
sim (163)
wavelet−LHH_firstorder_Median
frame
5
0
−5

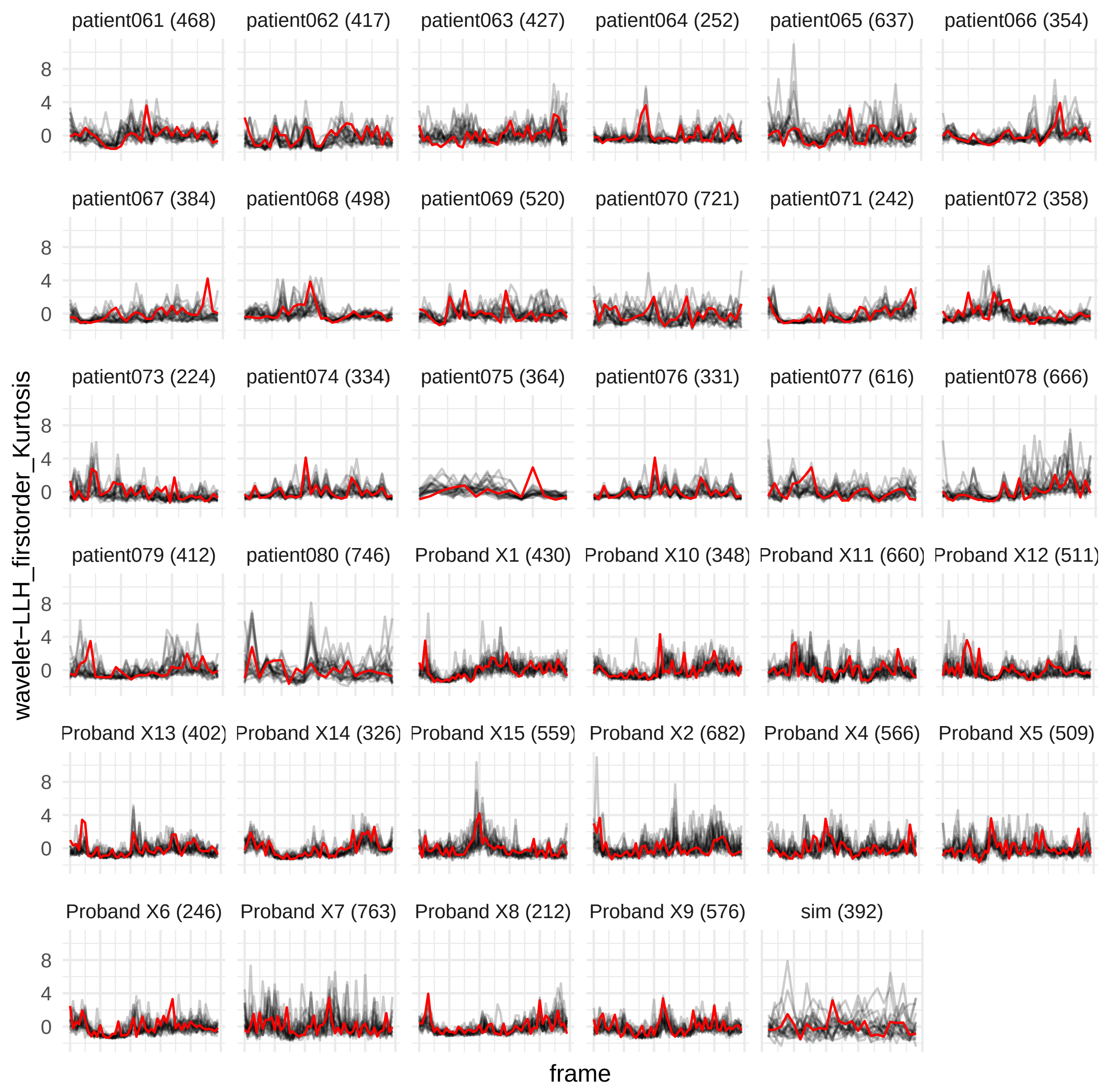
patient061 (468)
patient062 (417)
patient063 (427)
patient064 (252)
patient065 (637)
patient066 (354)
patient067 (384)
patient068 (498)
patient069 (520)
patient070 (721)
patient071 (242)
patient072 (358)
patient073 (224)
patient074 (334)
patient075 (364)
patient076 (331)
patient077 (616)
patient078 (666)
patient079 (412)
patient080 (746)
Proband X1 (430)
Proband X10 (348)
Proband X11 (660)
Proband X12 (511)
Proband X13 (402)
Proband X14 (326)
Proband X15 (559)
Proband X2 (682)
Proband X4 (566)
Proband X5 (509)
Proband X6 (246)
Proband X7 (763)
Proband X8 (212)
Proband X9 (576)
sim (392)
8
4
0
wavelet−LLH_firstorder_Kurtosis
frame

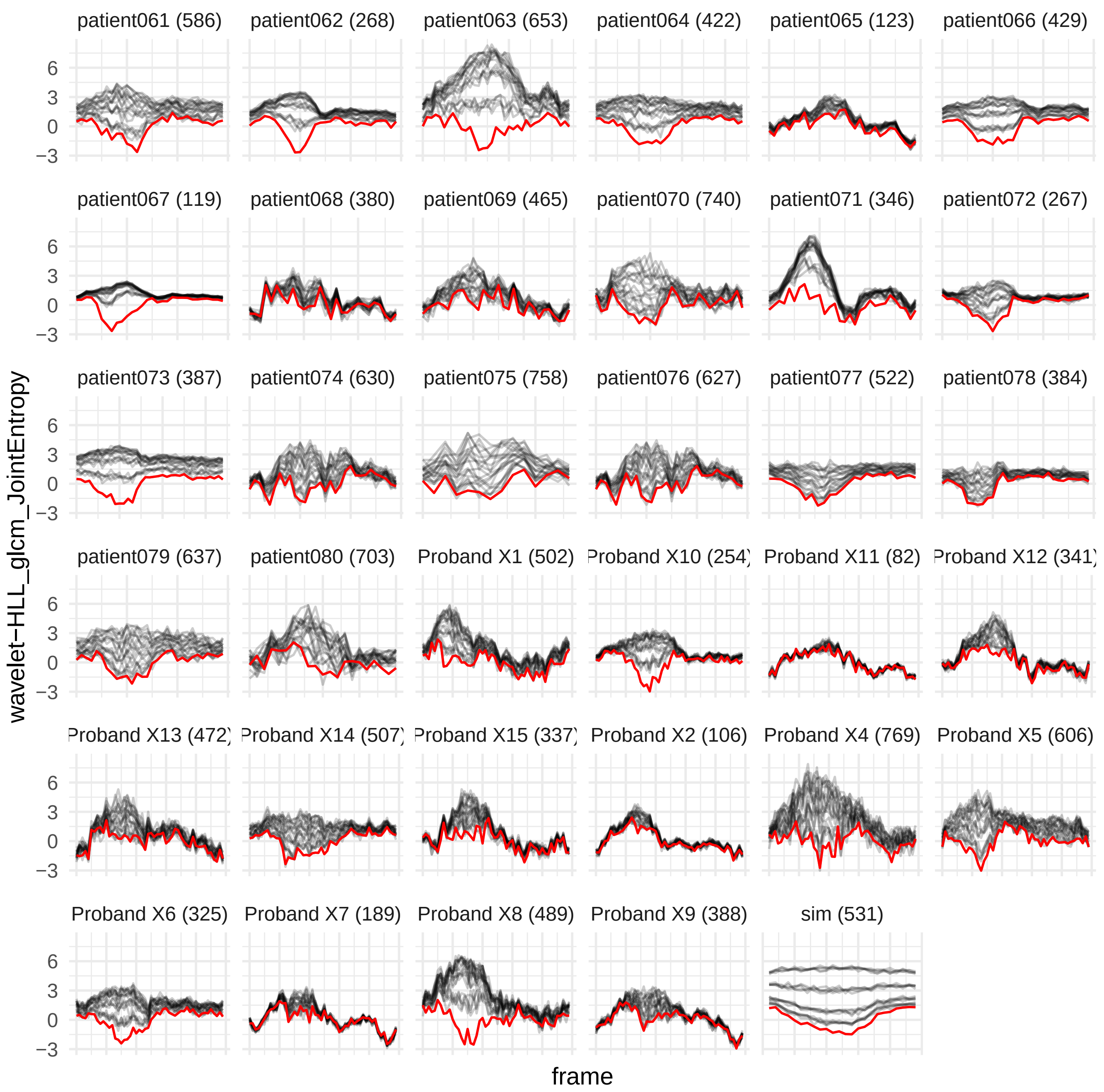
patient061 (586)
patient062 (268)
patient063 (653)
patient064 (422)
patient065 (123)
patient066 (429)
patient067 (119)
patient068 (380)
patient069 (465)
patient070 (740)
patient071 (346)
patient072 (267)
patient073 (387)
patient074 (630)
patient075 (758)
patient076 (627)
patient077 (522)
patient078 (384)
patient079 (637)
patient080 (703)
Proband X1 (502)
Proband X10 (254)
Proband X11 (82)
Proband X12 (341)
Proband X13 (472)
Proband X14 (507)
Proband X15 (337)
Proband X2 (106)
Proband X4 (769)
Proband X5 (606)
Proband X6 (325)
Proband X7 (189)
Proband X8 (489)
Proband X9 (388)
sim (531)
6
3
0
−3
wavelet−HLL_glcm_JointEntropy
frame

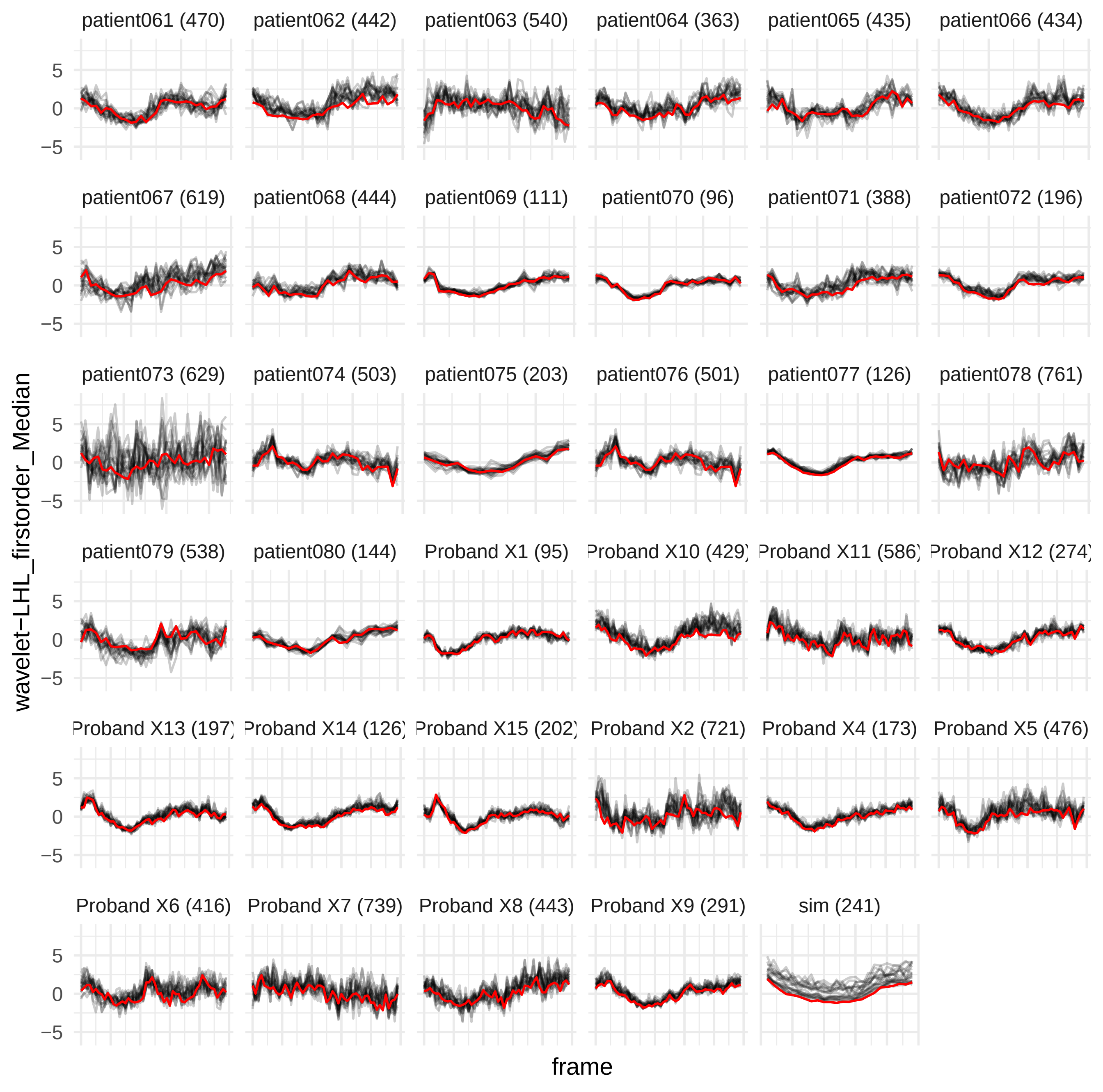

patient061 (470)
patient062 (442)
patient063 (540)
patient064 (363)
patient065 (435)
patient066 (434)
patient067 (619)
patient068 (444)
patient069 (111)
patient070 (96)
patient071 (388)
patient072 (196)
patient073 (629)
patient074 (503)
patient075 (203)
patient076 (501)
patient077 (126)
patient078 (761)
patient079 (538)
patient080 (144)
Proband X1 (95)
Proband X10 (429)
Proband X11 (586)
Proband X12 (274)
Proband X13 (197)
Proband X14 (126)
Proband X15 (202)
Proband X2 (721)
Proband X4 (173)
Proband X5 (476)
Proband X6 (416)
Proband X7 (739)
Proband X8 (443)
Proband X9 (291)
sim (241)
5
0
−5
wavelet−LHL_firstorder_Median
frame

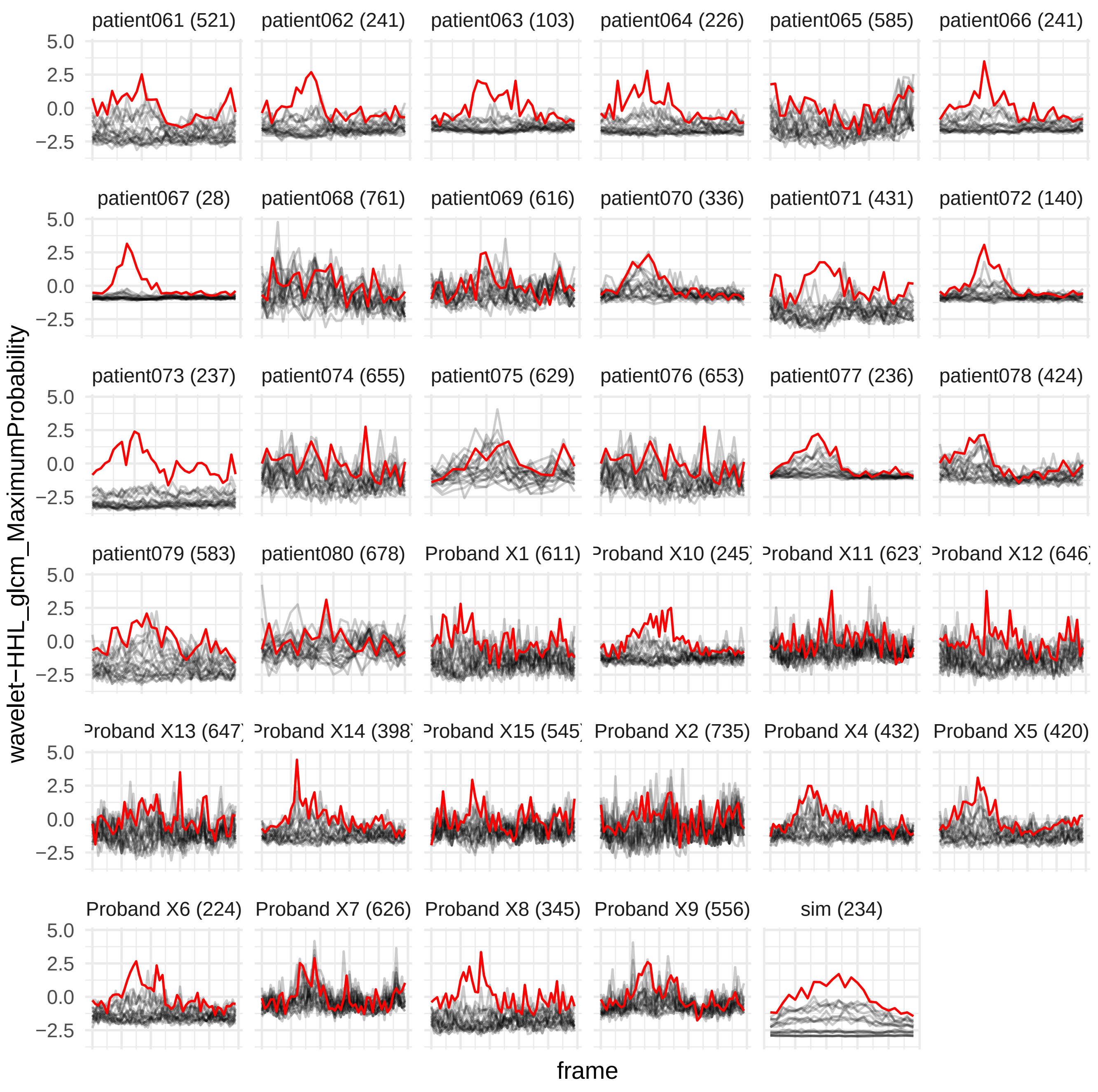

patient061 (521)
patient062 (241)
patient063 (103)
patient064 (226)
patient065 (585)
patient066 (241)
patient067 (28)
patient068 (761)
patient069 (616)
patient070 (336)
patient071 (431)
patient072 (140)
patient073 (237)
patient074 (655)
patient075 (629)
patient076 (653)
patient077 (236)
patient078 (424)
patient079 (583)
patient080 (678)
Proband X1 (611)
Proband X10 (245)
Proband X11 (623)
Proband X12 (646)
Proband X13 (647)
Proband X14 (398)
Proband X15 (545)
Proband X2 (735)
Proband X4 (432)
Proband X5 (420)
Proband X6 (224)
Proband X7 (626)
Proband X8 (345)
Proband X9 (556)
sim (234)
5.0
2.5
0.0
−2.5
wavelet−HHL_glcm_MaximumProbability
frame

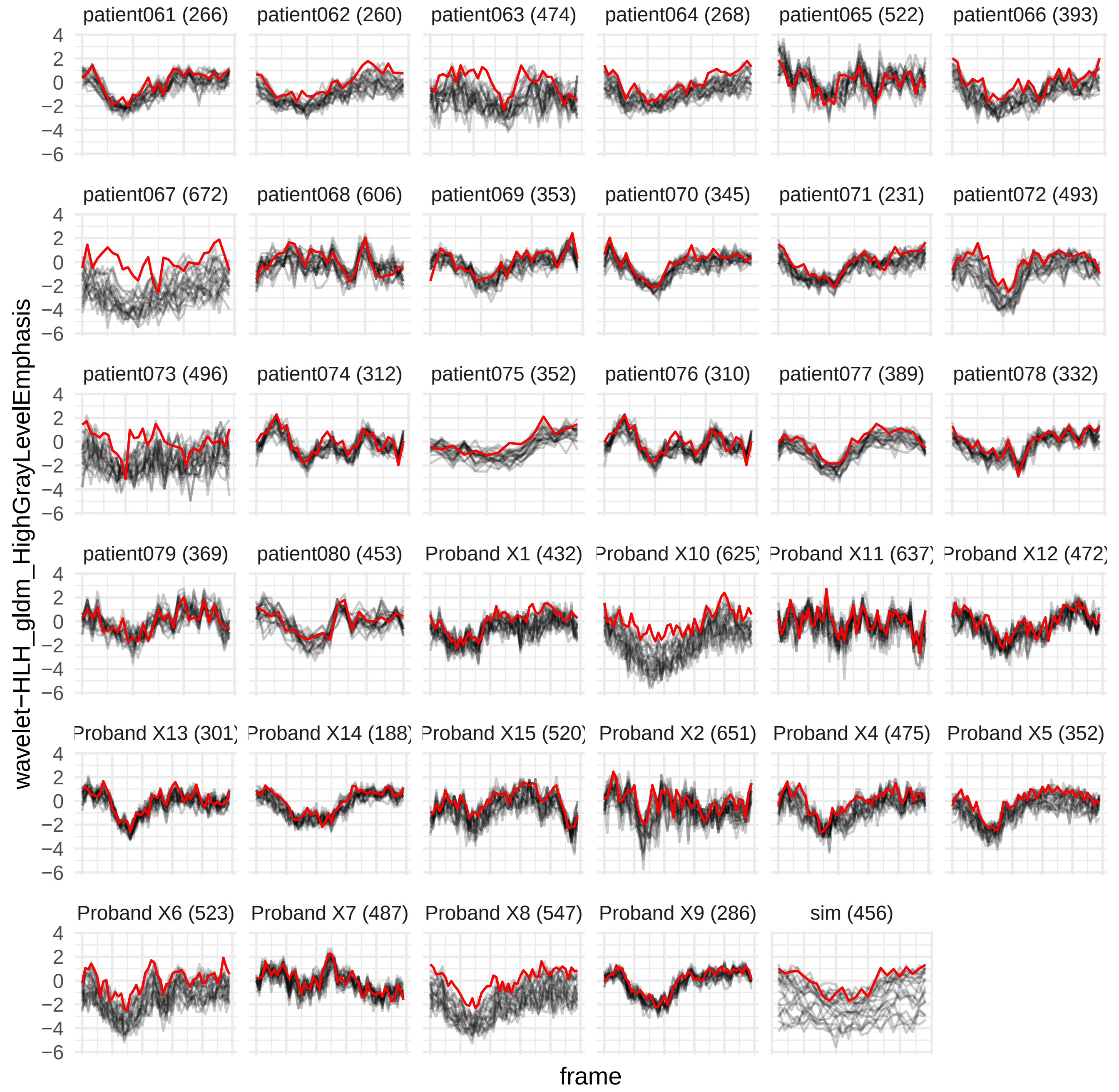

patient061 (266)
patient062 (260)
patient063 (474)
patient064 (268)
patient065 (522)
patient066 (393)
patient067 (672)
patient068 (606)
patient069 (353)
patient070 (345)
patient071 (231)
patient072 (493)
patient073 (496)
patient074 (312)
patient075 (352)
patient076 (310)
patient077 (389)
patient078 (332)
patient079 (369)
patient080 (453)
Proband X1 (432)
Proband X10 (625)
Proband X11 (637)
Proband X12 (472)
Proband X13 (301)
Proband X14 (188)
Proband X15 (520)
Proband X2 (651)
Proband X4 (475)
Proband X5 (352)
Proband X6 (523)
Proband X7 (487)
Proband X8 (547)
Proband X9 (286)
sim (456)
4
2
0
−2
−4
−6
wavelet−HLH_gldm_HighGrayLevelEmphasis
frame

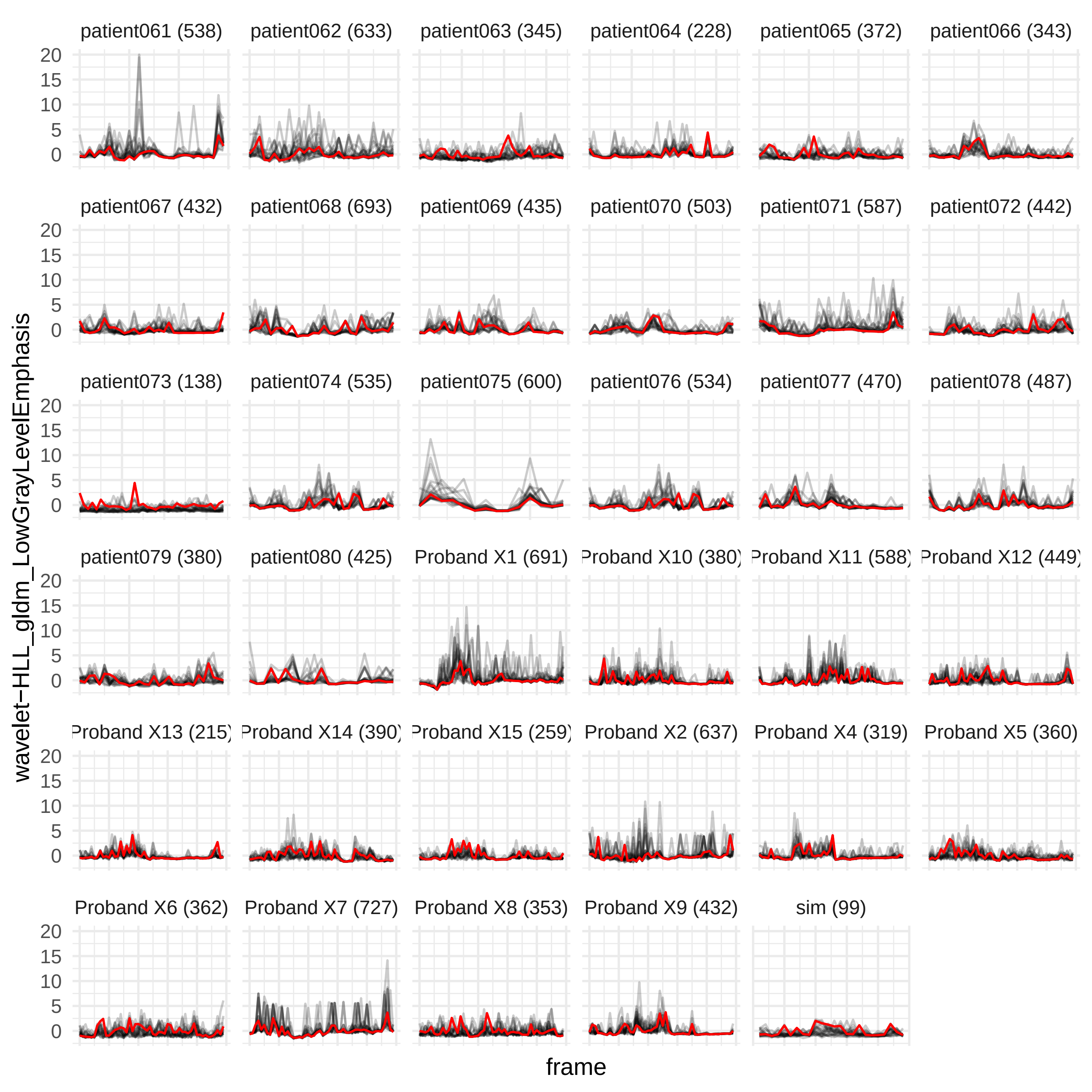

patient061 (538)
patient062 (633)
patient063 (345)
patient064 (228)
patient065 (372)
patient066 (343)
patient067 (432)
patient068 (693)
patient069 (435)
patient070 (503)
patient071 (587)
patient072 (442)
patient073 (138)
patient074 (535)
patient075 (600)
patient076 (534)
patient077 (470)
patient078 (487)
patient079 (380)
patient080 (425)
Proband X1 (691)
Proband X10 (380)
Proband X11 (588)
Proband X12 (449)
Proband X13 (215)
Proband X14 (390)
Proband X15 (259)
Proband X2 (637)
Proband X4 (319)
Proband X5 (360)
Proband X6 (362)
Proband X7 (727)
Proband X8 (353)
Proband X9 (432)
sim (99)
wavelet−HLL_gldm_LowGrayLevelEmphasis
0
5
10
15
20
frame

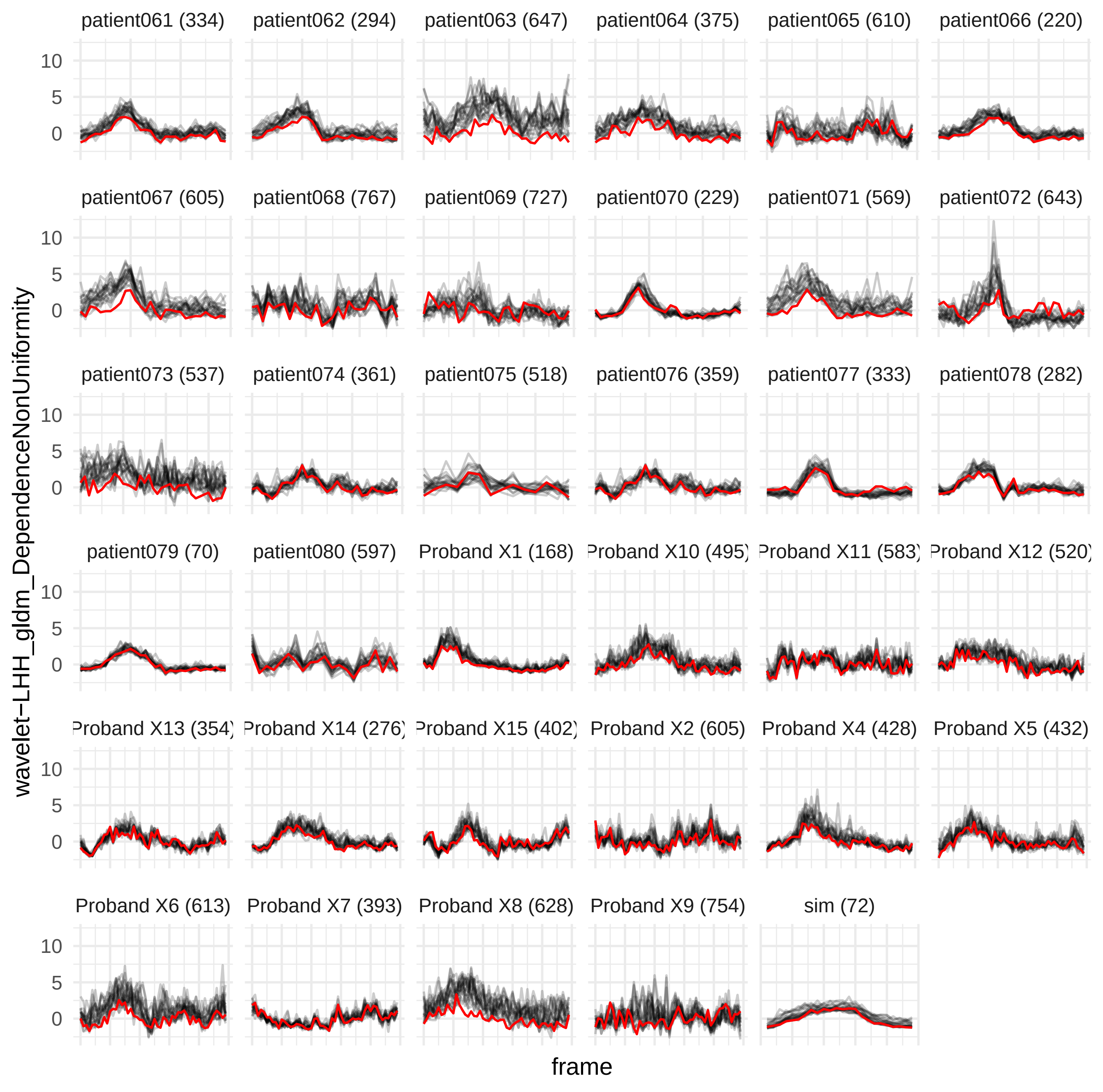

patient061 (334)
patient062 (294)
patient063 (647)
patient064 (375)
patient065 (610)
patient066 (220)
patient067 (605)
patient068 (767)
patient069 (727)
patient070 (229)
patient071 (569)
patient072 (643)
patient073 (537)
patient074 (361)
patient075 (518)
patient076 (359)
patient077 (333)
patient078 (282)
patient079 (70)
patient080 (597)
Proband X1 (168)
Proband X10 (495)
Proband X11 (583)
Proband X12 (520)
Proband X13 (354)
Proband X14 (276)
Proband X15 (402)
Proband X2 (605)
Proband X4 (428)
Proband X5 (432)
Proband X6 (613)
Proband X7 (393)
Proband X8 (628)
Proband X9 (754)
sim (72)
10
5
0
wavelet−LHH_gldm_DependenceNonUniformity
frame

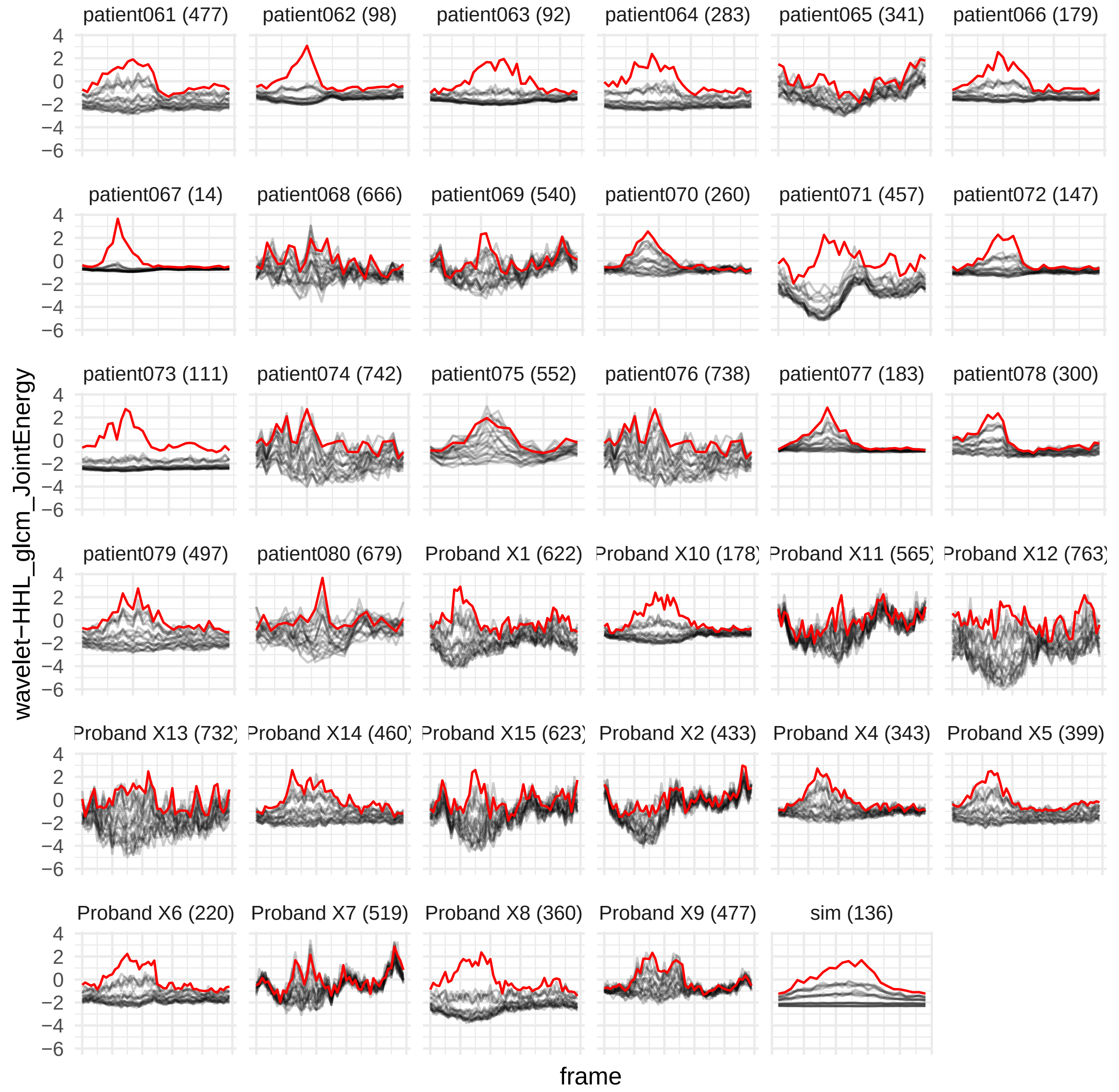
patient061 (477)
patient062 (98)
patient063 (92)
patient064 (283)
patient065 (341)
patient066 (179)
patient067 (14)
patient068 (666)
patient069 (540)
patient070 (260)
patient071 (457)
patient072 (147)
patient073 (111)
patient074 (742)
patient075 (552)
patient076 (738)
patient077 (183)
patient078 (300)
patient079 (497)
patient080 (679)
Proband X1 (622)
Proband X10 (178)
Proband X11 (565)
Proband X12 (763)
Proband X13 (732)
Proband X14 (460)
Proband X15 (623)
Proband X2 (433)
Proband X4 (343)
Proband X5 (399)
Proband X6 (220)
Proband X7 (519)
Proband X8 (360)
Proband X9 (477)
sim (136)
4
2
0
−2
−4
−6
wavelet−HHL_glcm_JointEnergy
frame

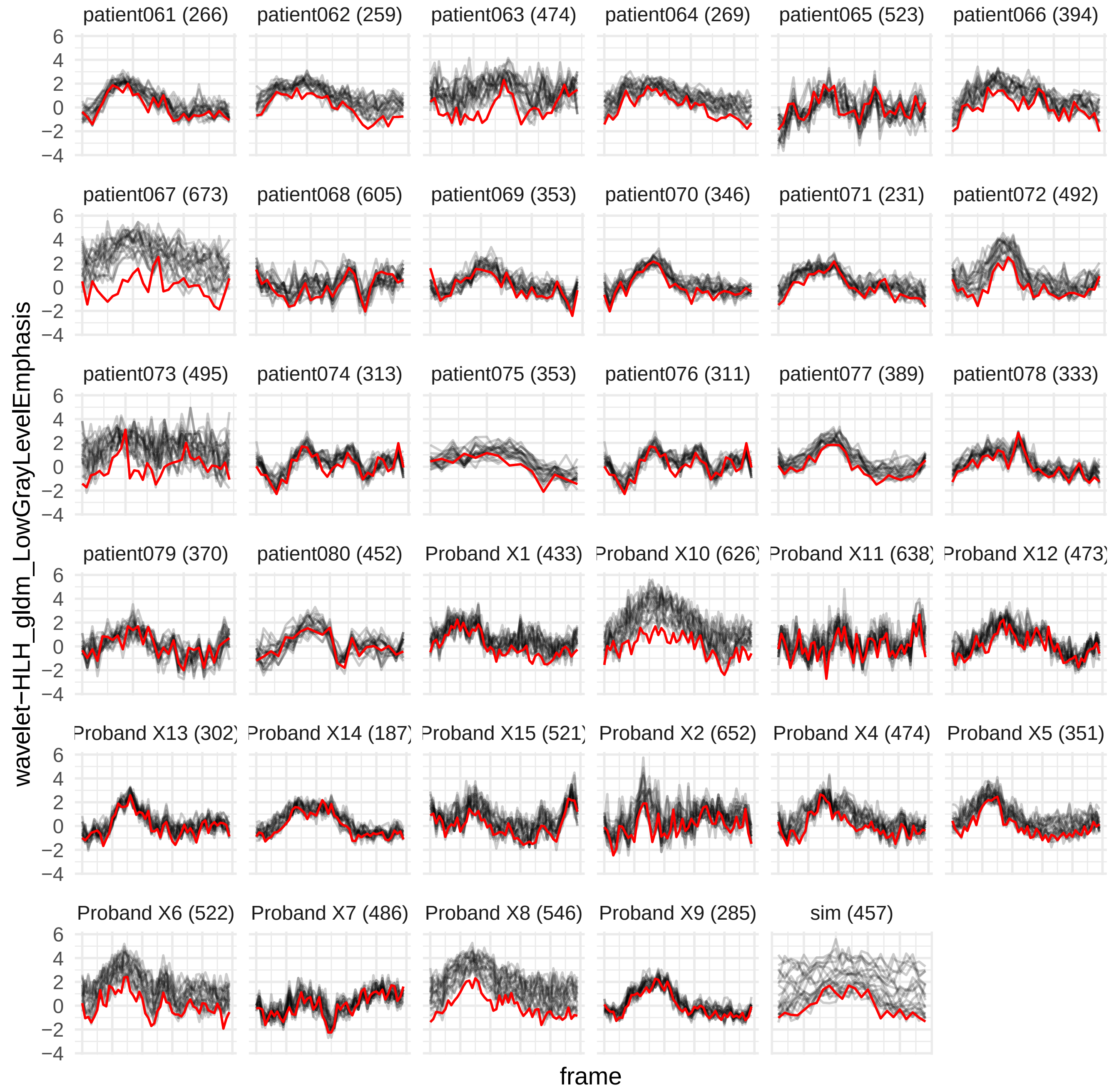

patient061 (266)
patient062 (259)
patient063 (474)
patient064 (269)
patient065 (523)
patient066 (394)
patient067 (673)
patient068 (605)
patient069 (353)
patient070 (346)
patient071 (231)
patient072 (492)
patient073 (495)
patient074 (313)
patient075 (353)
patient076 (311)
patient077 (389)
patient078 (333)
patient079 (370)
patient080 (452)
Proband X1 (433)
Proband X10 (626)
Proband X11 (638)
Proband X12 (473)
Proband X13 (302)
Proband X14 (187)
Proband X15 (521)
Proband X2 (652)
Proband X4 (474)
Proband X5 (351)
Proband X6 (522)
Proband X7 (486)
Proband X8 (546)
Proband X9 (285)
sim (457)
6
4
2
0
−2
−4
wavelet−HLH_gldm_LowGrayLevelEmphasis
frame

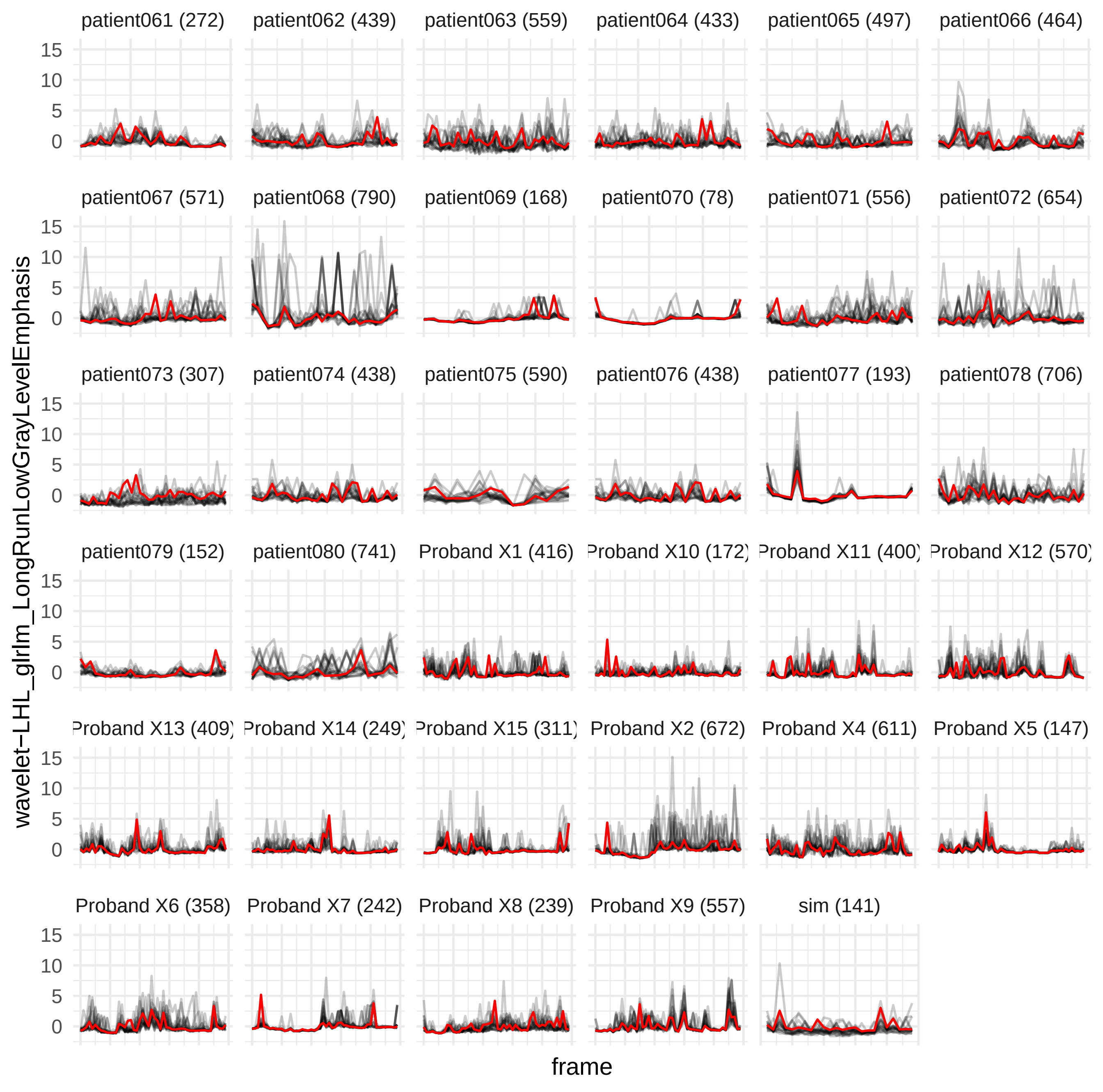
patient061 (272)
patient062 (439)
patient063 (559)
patient064 (433)
patient065 (497)
patient066 (464)
patient067 (571)
patient068 (790)
patient069 (168)
patient070 (78)
patient071 (556)
patient072 (654)
patient073 (307)
patient074 (438)
patient075 (590)
patient076 (438)
patient077 (193)
patient078 (706)
patient079 (152)
patient080 (741)
Proband X1 (416)
Proband X10 (172)
Proband X11 (400)
Proband X12 (570)
Proband X13 (409)
Proband X14 (249)
Proband X15 (311)
Proband X2 (672)
Proband X4 (611)
Proband X5 (147)
Proband X6 (358)
Proband X7 (242)
Proband X8 (239)
Proband X9 (557)
sim (141)
wavelet−LHL_glrlm_LongRunLowGrayLevelEmphasis
15
10
5
0
frame

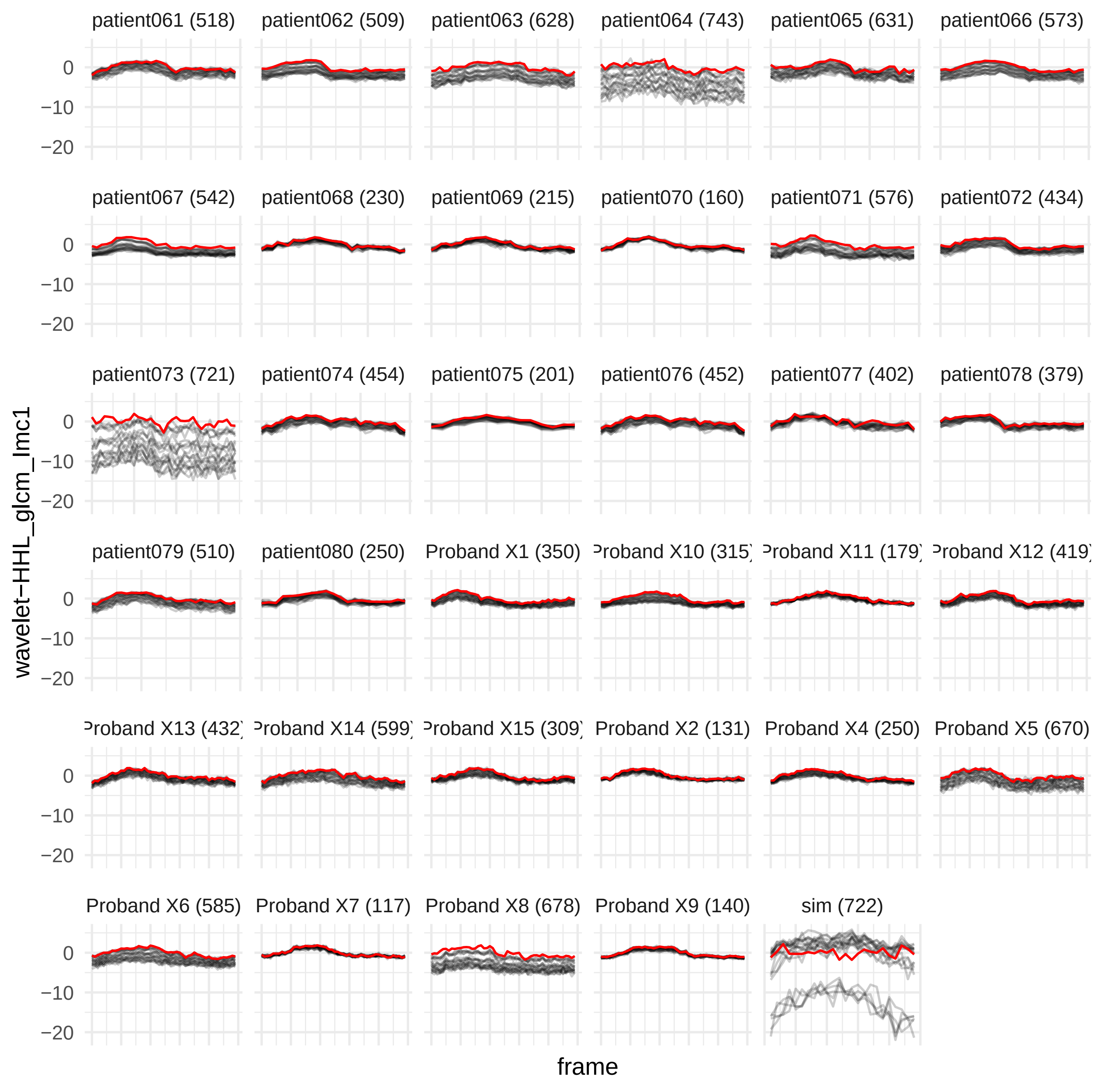
patient061 (518)
patient062 (509)
patient063 (628)
patient064 (743)
patient065 (631)
patient066 (573)
patient067 (542)
patient068 (230)
patient069 (215)
patient070 (160)
patient071 (576)
patient072 (434)
patient073 (721)
patient074 (454)
patient075 (201)
patient076 (452)
patient077 (402)
patient078 (379)
patient079 (510)
patient080 (250)
Proband X1 (350)
Proband X10 (315)
Proband X11 (179)
Proband X12 (419)
Proband X13 (432)
Proband X14 (599)
Proband X15 (309)
Proband X2 (131)
Proband X4 (250)
Proband X5 (670)
Proband X6 (585)
Proband X7 (117)
Proband X8 (678)
Proband X9 (140)
sim (722)
0
−10
−20
wavelet−HHL_glcm_Imc1
frame

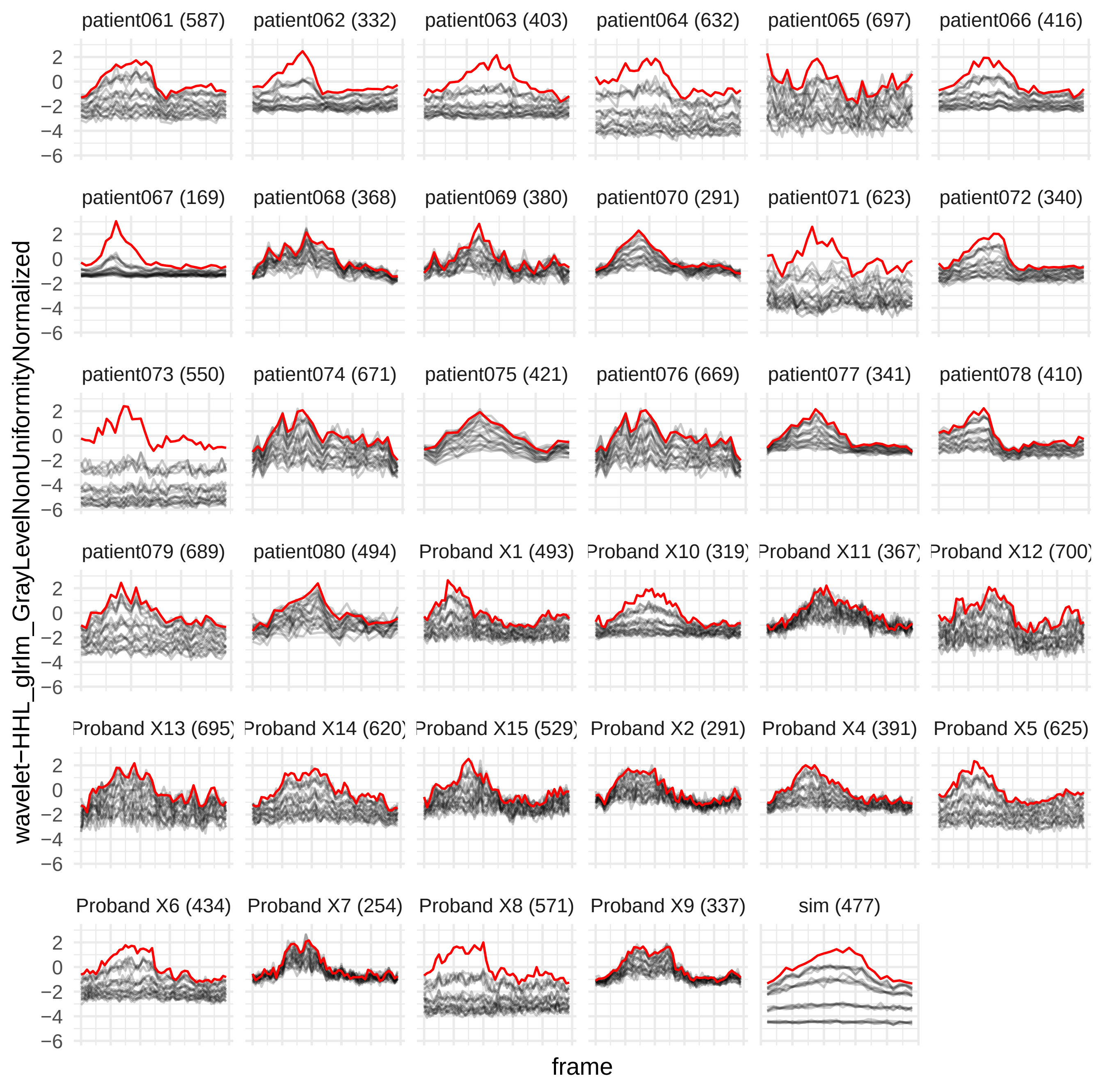

patient061 (587)
patient062 (332)
patient063 (403)
patient064 (632)
patient065 (697)
patient066 (416)
patient067 (169)
patient068 (368)
patient069 (380)
patient070 (291)
patient071 (623)
patient072 (340)
patient073 (550)
patient074 (671)
patient075 (421)
patient076 (669)
patient077 (341)
patient078 (410)
patient079 (689)
patient080 (494)
Proband X1 (493)
Proband X10 (319)
Proband X11 (367)
Proband X12 (700)
Proband X13 (695)
Proband X14 (620)
Proband X15 (529)
Proband X2 (291)
Proband X4 (391)
Proband X5 (625)
Proband X6 (434)
Proband X7 (254)
Proband X8 (571)
Proband X9 (337)
sim (477)
2
0
−2
−4
−6
wavelet−HHL_glrlm_GrayLevelNonUniformityNormalized
frame

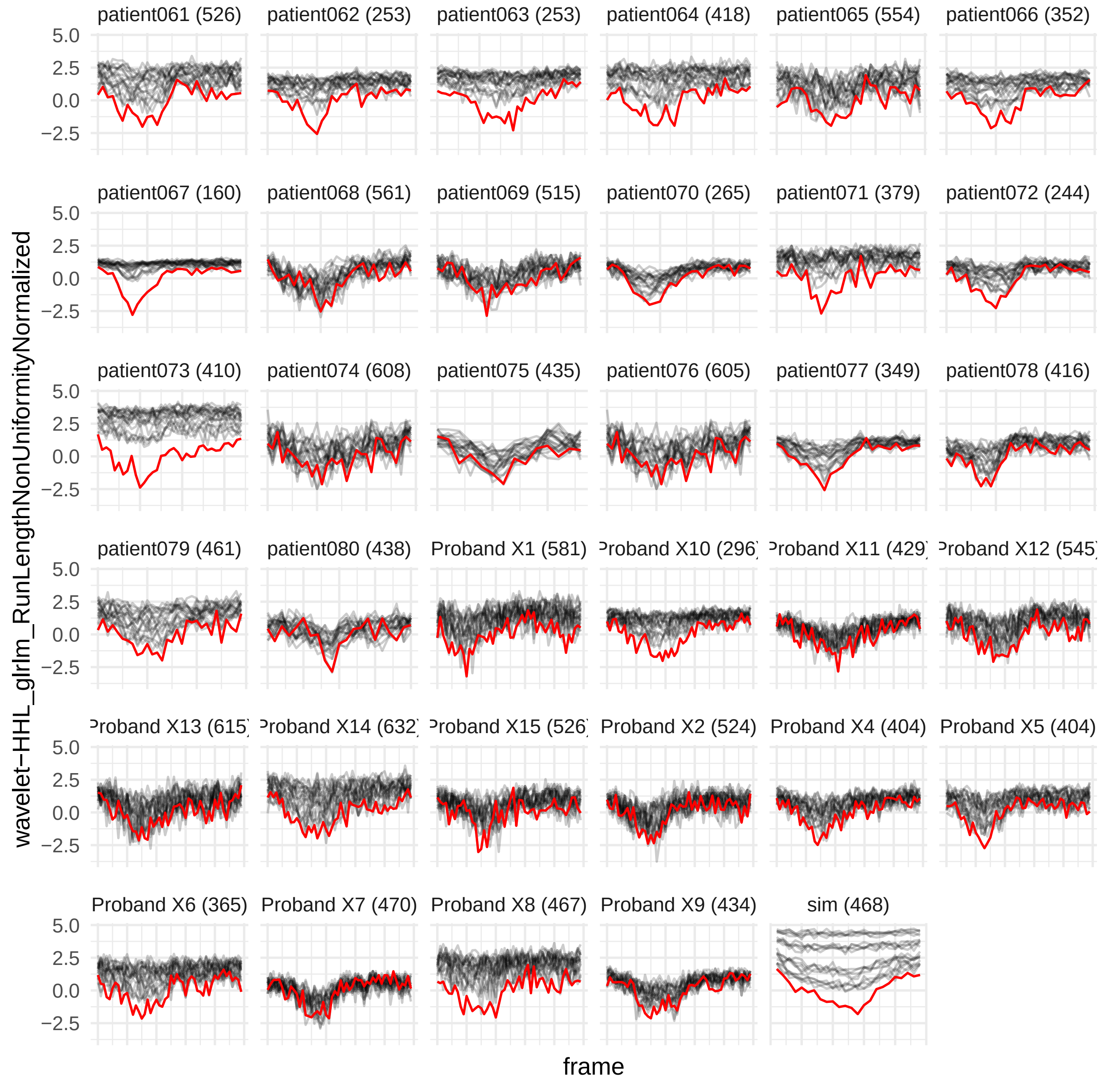
patient061 (526)
patient062 (253)
patient063 (253)
patient064 (418)
patient065 (554)
patient066 (352)
patient067 (160)
patient068 (561)
patient069 (515)
patient070 (265)
patient071 (379)
patient072 (244)
patient073 (410)
patient074 (608)
patient075 (435)
patient076 (605)
patient077 (349)
patient078 (416)
patient079 (461)
patient080 (438)
Proband X1 (581)
Proband X10 (296)
Proband X11 (429)
Proband X12 (545)
Proband X13 (615)
Proband X14 (632)
Proband X15 (526)
Proband X2 (524)
Proband X4 (404)
Proband X5 (404)
Proband X6 (365)
Proband X7 (470)
Proband X8 (467)
Proband X9 (434)
sim (468)
5.0
2.5
0.0
−2.5
wavelet−HHL_glrlm_RunLengthNonUniformityNormalized
frame

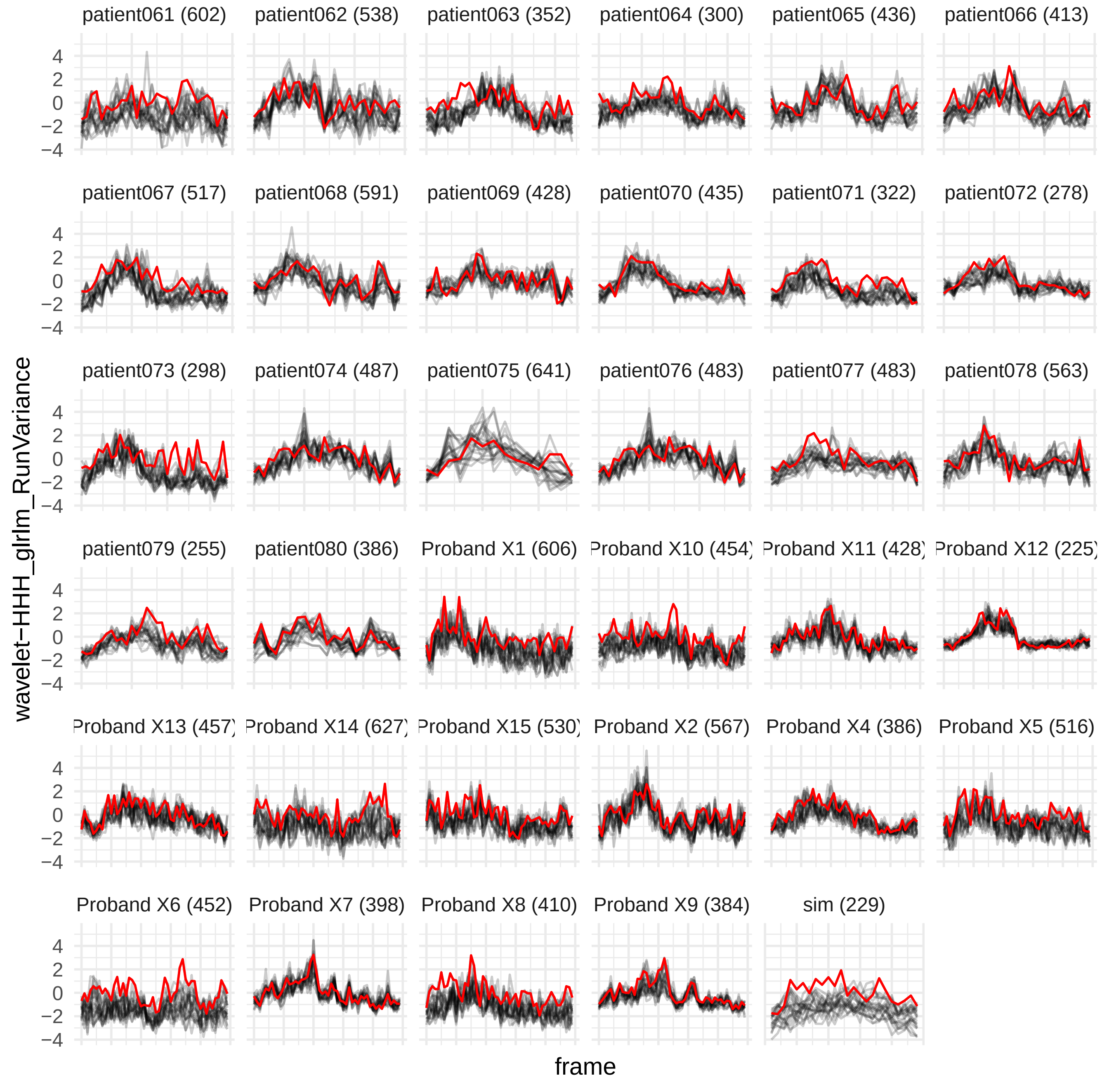

patient061 (602)
patient062 (538)
patient063 (352)
patient064 (300)
patient065 (436)
patient066 (413)
patient067 (517)
patient068 (591)
patient069 (428)
patient070 (435)
patient071 (322)
patient072 (278)
patient073 (298)
patient074 (487)
patient075 (641)
patient076 (483)
patient077 (483)
patient078 (563)
patient079 (255)
patient080 (386)
Proband X1 (606)
Proband X10 (454)
Proband X11 (428)
Proband X12 (225)
Proband X13 (457)
Proband X14 (627)
Proband X15 (530)
Proband X2 (567)
Proband X4 (386)
Proband X5 (516)
Proband X6 (452)
Proband X7 (398)
Proband X8 (410)
Proband X9 (384)
sim (229)
wavelet−HHH_glrlm_RunVariance
frame
4
2
0
−2
−4

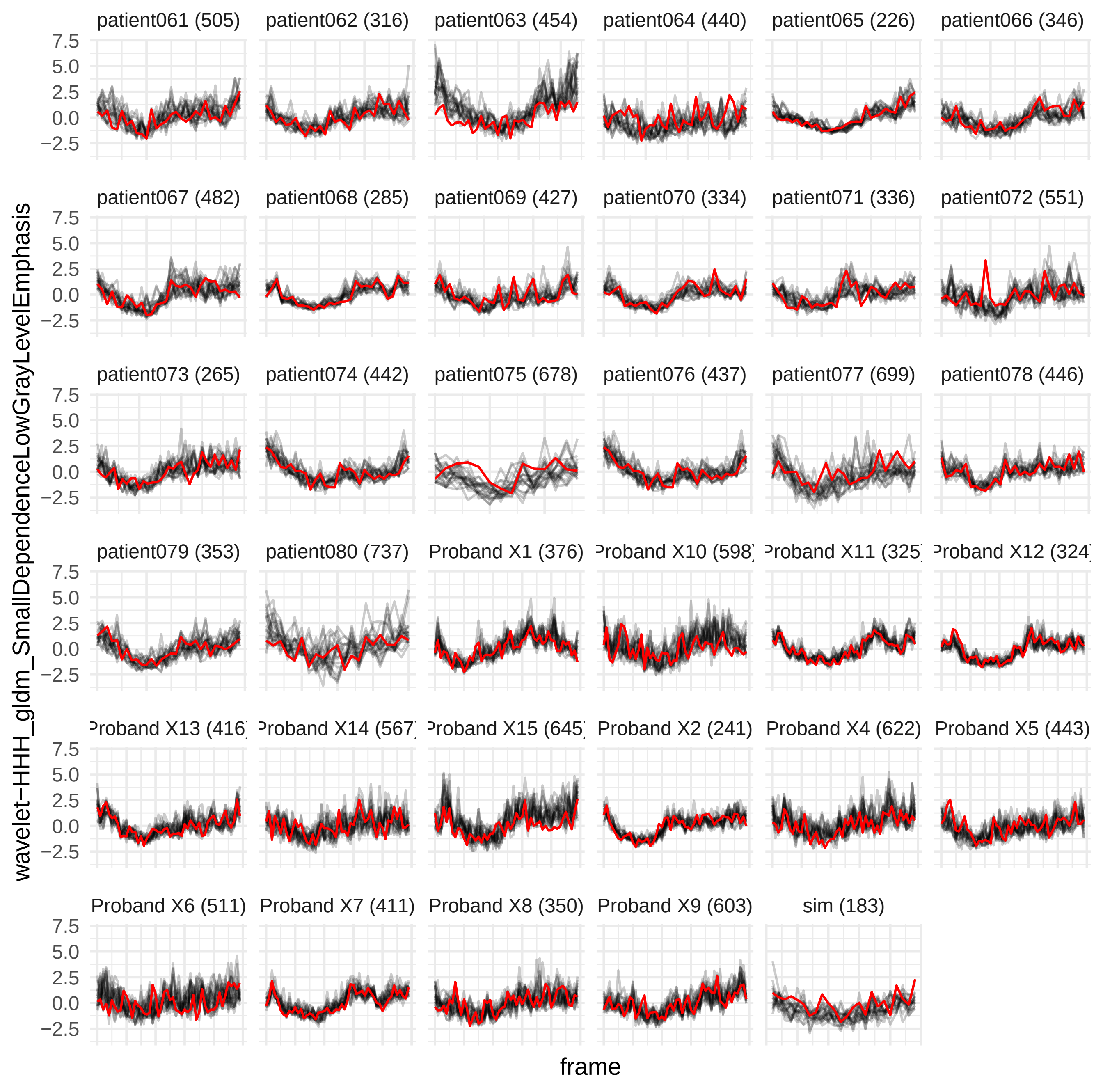

patient061 (505)
patient062 (316)
patient063 (454)
patient064 (440)
patient065 (226)
patient066 (346)
patient067 (482)
patient068 (285)
patient069 (427)
patient070 (334)
patient071 (336)
patient072 (551)
patient073 (265)
patient074 (442)
patient075 (678)
patient076 (437)
patient077 (699)
patient078 (446)
patient079 (353)
patient080 (737)
Proband X1 (376)
Proband X10 (598)
Proband X11 (325)
Proband X12 (324)
Proband X13 (416)
Proband X14 (567)
Proband X15 (645)
Proband X2 (241)
Proband X4 (622)
Proband X5 (443)
Proband X6 (511)
Proband X7 (411)
Proband X8 (350)
Proband X9 (603)
sim (183)
7.5
5.0
2.5
0.0
−2.5
wavelet−HHH_gldm_SmallDependenceLowGrayLevelEmphasis
frame

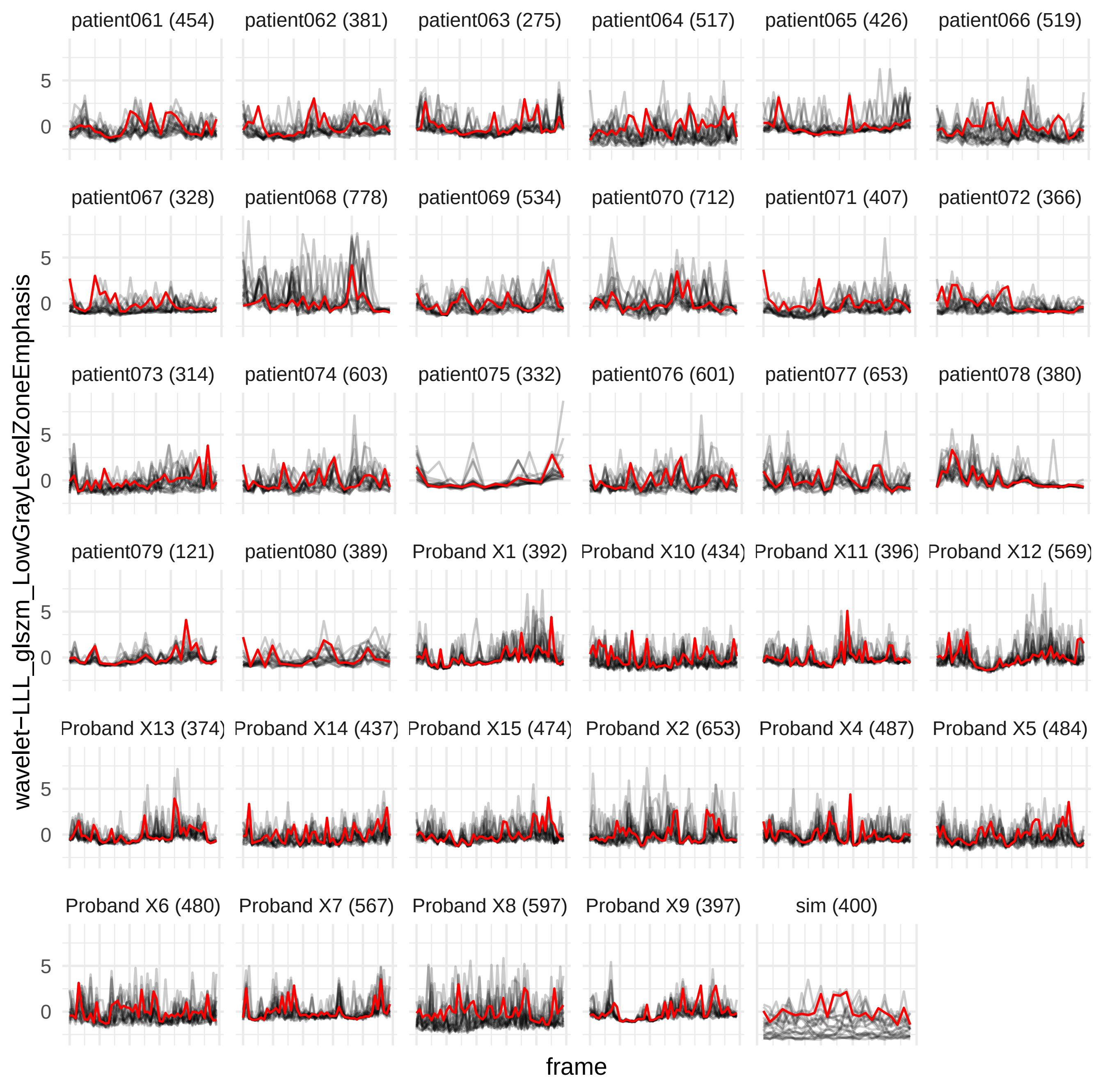

patient061 (454)
patient062 (381)
patient063 (275)
patient064 (517)
patient065 (426)
patient066 (519)
patient067 (328)
patient068 (778)
patient069 (534)
patient070 (712)
patient071 (407)
patient072 (366)
patient073 (314)
patient074 (603)
patient075 (332)
patient076 (601)
patient077 (653)
patient078 (380)
patient079 (121)
patient080 (389)
Proband X1 (392)
Proband X10 (434)
Proband X11 (396)
Proband X12 (569)
Proband X13 (374)
Proband X14 (437)
Proband X15 (474)
Proband X2 (653)
Proband X4 (487)
Proband X5 (484)
Proband X6 (480)
Proband X7 (567)
Proband X8 (597)
Proband X9 (397)
sim (400)
5
0
wavelet−LLL_glszm_LowGrayLevelZoneEmphasis
frame

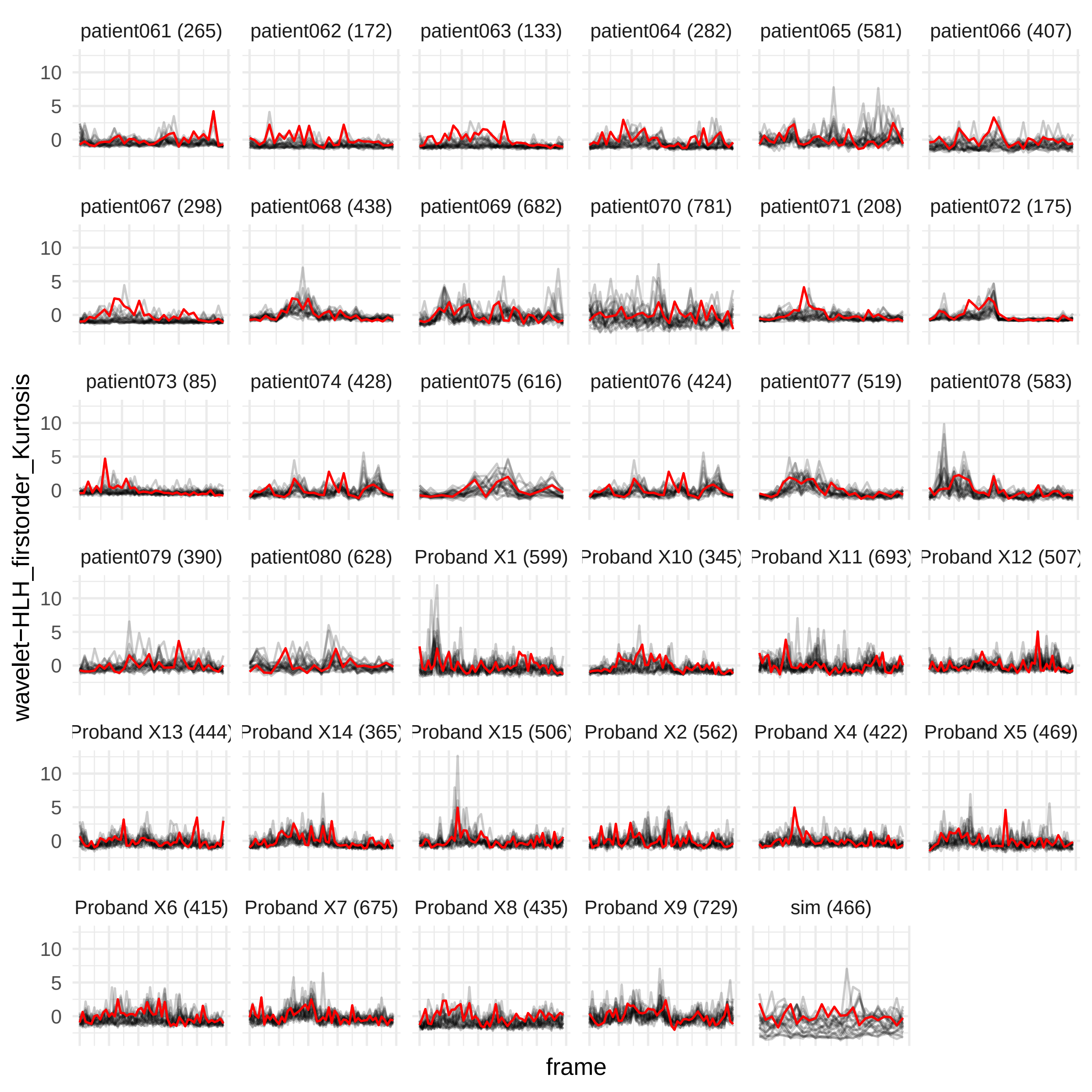

patient061 (265)
patient062 (172)
patient063 (133)
patient064 (282)
patient065 (581)
patient066 (407)
patient067 (298)
patient068 (438)
patient069 (682)
patient070 (781)
patient071 (208)
patient072 (175)
patient073 (85)
patient074 (428)
patient075 (616)
patient076 (424)
patient077 (519)
patient078 (583)
patient079 (390)
patient080 (628)
Proband X1 (599)
Proband X10 (345)
Proband X11 (693)
Proband X12 (507)
Proband X13 (444)
Proband X14 (365)
Proband X15 (506)
Proband X2 (562)
Proband X4 (422)
Proband X5 (469)
Proband X6 (415)
Proband X7 (675)
Proband X8 (435)
Proband X9 (729)
sim (466)
10
5
0
wavelet−HLH_firstorder_Kurtosis
frame

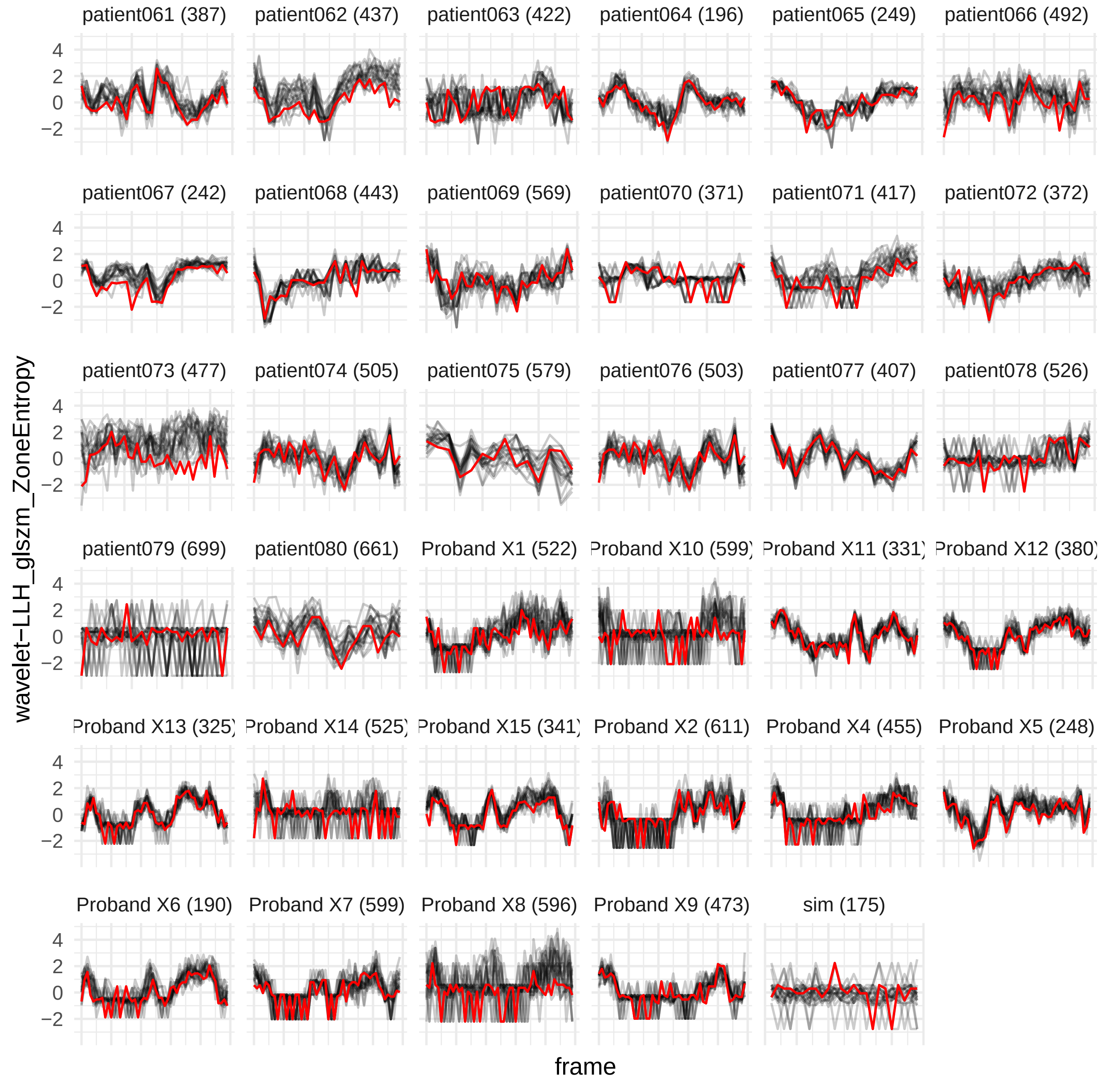

patient061 (387)
patient062 (437)
patient063 (422)
patient064 (196)
patient065 (249)
patient066 (492)
patient067 (242)
patient068 (443)
patient069 (569)
patient070 (371)
patient071 (417)
patient072 (372)
patient073 (477)
patient074 (505)
patient075 (579)
patient076 (503)
patient077 (407)
patient078 (526)
patient079 (699)
patient080 (661)
Proband X1 (522)
Proband X10 (599)
Proband X11 (331)
Proband X12 (380)
Proband X13 (325)
Proband X14 (525)
Proband X15 (341)
Proband X2 (611)
Proband X4 (455)
Proband X5 (248)
Proband X6 (190)
Proband X7 (599)
Proband X8 (596)
Proband X9 (473)
sim (175)
wavelet−LLH_glszm_ZoneEntropy
frame
4
2
0
−2

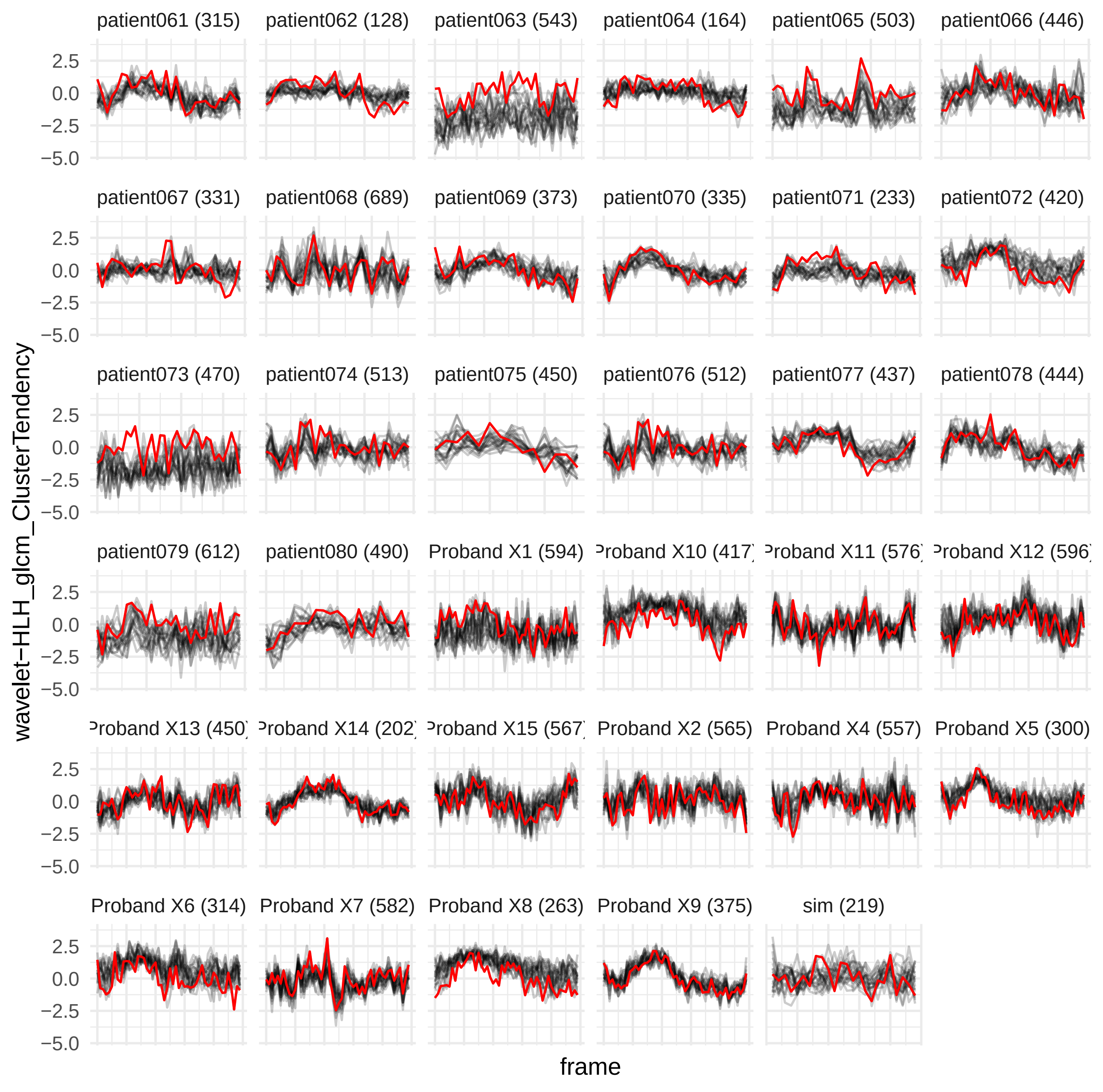
patient061 (315)
patient062 (128)
patient063 (543)
patient064 (164)
patient065 (503)
patient066 (446)
patient067 (331)
patient068 (689)
patient069 (373)
patient070 (335)
patient071 (233)
patient072 (420)
patient073 (470)
patient074 (513)
patient075 (450)
patient076 (512)
patient077 (437)
patient078 (444)
patient079 (612)
patient080 (490)
Proband X1 (594)
roband X10 (417
roband X11 (576
roband X12 (596
roband X13 (450
roband X14 (202
roband X15 (567
Proband X2 (565)
Proband X4 (557)
Proband X5 (300)
Proband X6 (314)
Proband X7 (582)
Proband X8 (263)
Proband X9 (375)
sim (219)
2.5
0.0
−2.5
−5.0
wavelet−HLH_glcm_ClusterTendency
frame

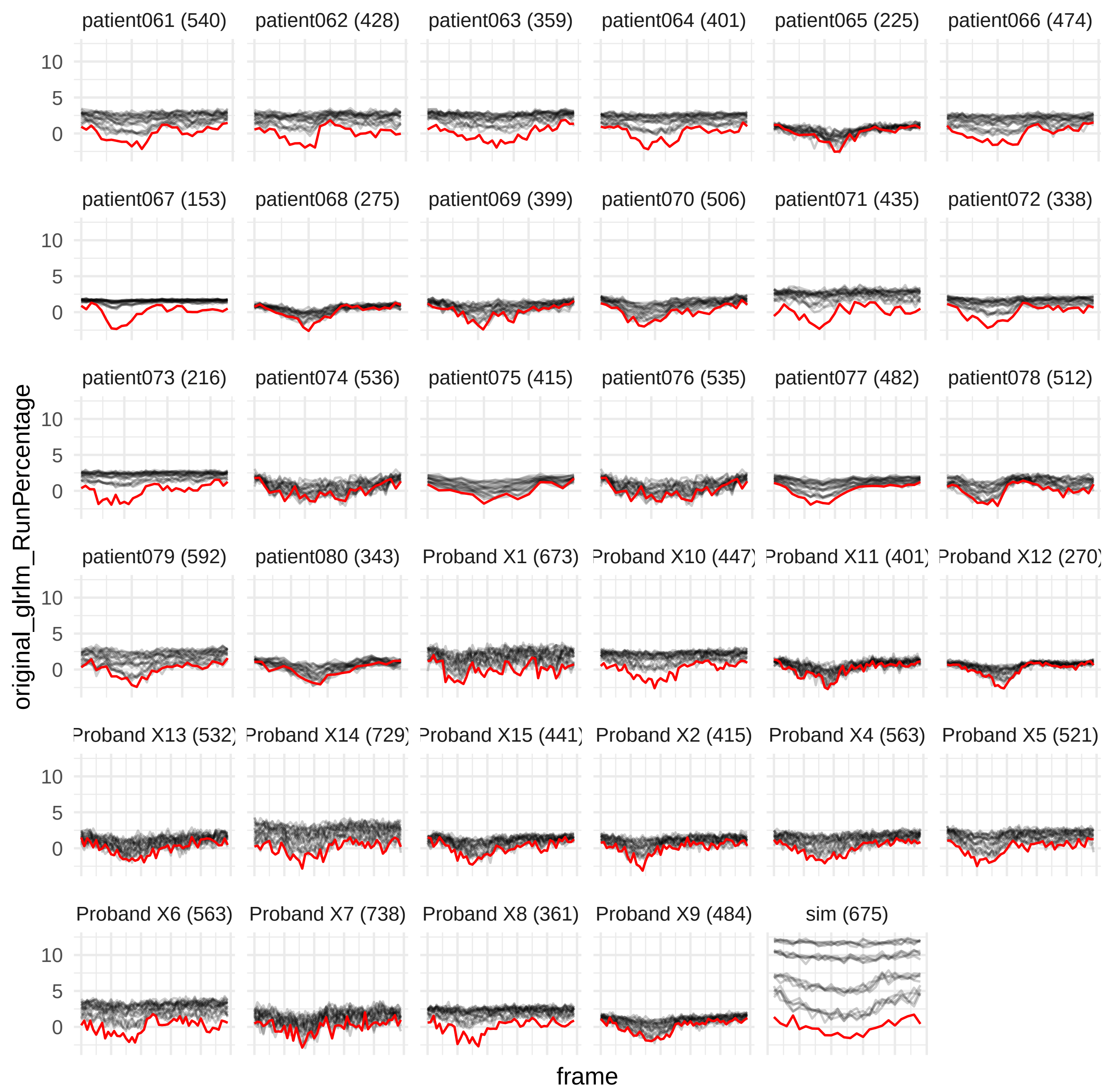
patient061 (540)
patient062 (428)
patient063 (359)
patient064 (401)
patient065 (225)
patient066 (474)
patient067 (153)
patient068 (275)
patient069 (399)
patient070 (506)
patient071 (435)
patient072 (338)
patient073 (216)
patient074 (536)
patient075 (415)
patient076 (535)
patient077 (482)
patient078 (512)
patient079 (592)
patient080 (343)
Proband X1 (673)
Proband X10 (447)
Proband X11 (401)
Proband X12 (270)
Proband X13 (532)
Proband X14 (729)
Proband X15 (441)
Proband X2 (415)
Proband X4 (563)
Proband X5 (521)
Proband X6 (563)
Proband X7 (738)
Proband X8 (361)
Proband X9 (484)
sim (675)
original_glrlm_RunPercentage
frame
0
5
10

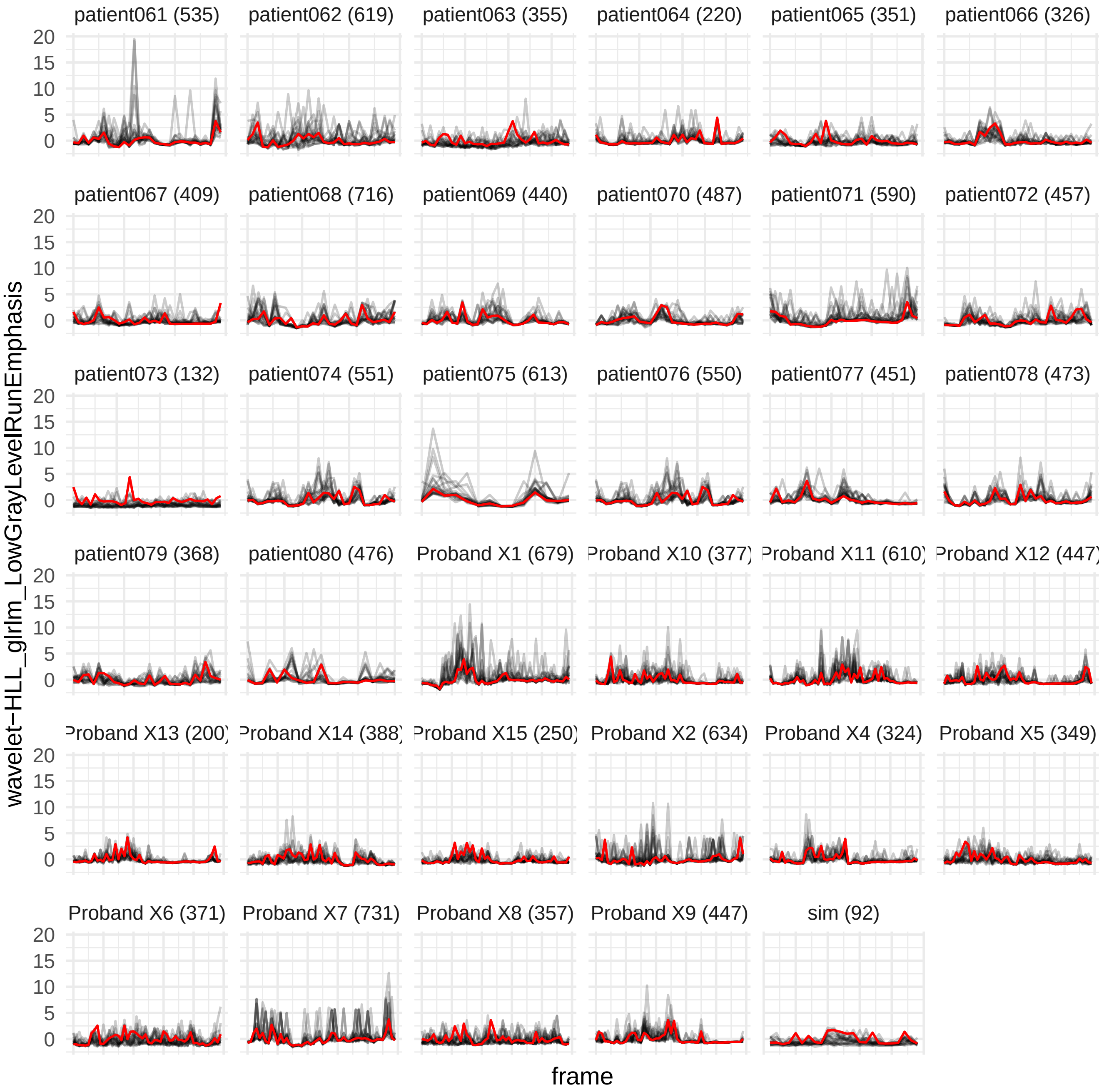

patient061 (535)
patient062 (619)
patient063 (355)
patient064 (220)
patient065 (351)
patient066 (326)
patient067 (409)
patient068 (716)
patient069 (440)
patient070 (487)
patient071 (590)
patient072 (457)
patient073 (132)
patient074 (551)
patient075 (613)
patient076 (550)
patient077 (451)
patient078 (473)
patient079 (368)
patient080 (476)
Proband X1 (679)
Proband X10 (377)
Proband X11 (610)
Proband X12 (447)
Proband X13 (200)
Proband X14 (388)
Proband X15 (250)
Proband X2 (634)
Proband X4 (324)
Proband X5 (349)
Proband X6 (371)
Proband X7 (731)
Proband X8 (357)
Proband X9 (447)
sim (92)
0
5
10
15
20
wavelet−HLL_glrlm_LowGrayLevelRunEmphasis
frame

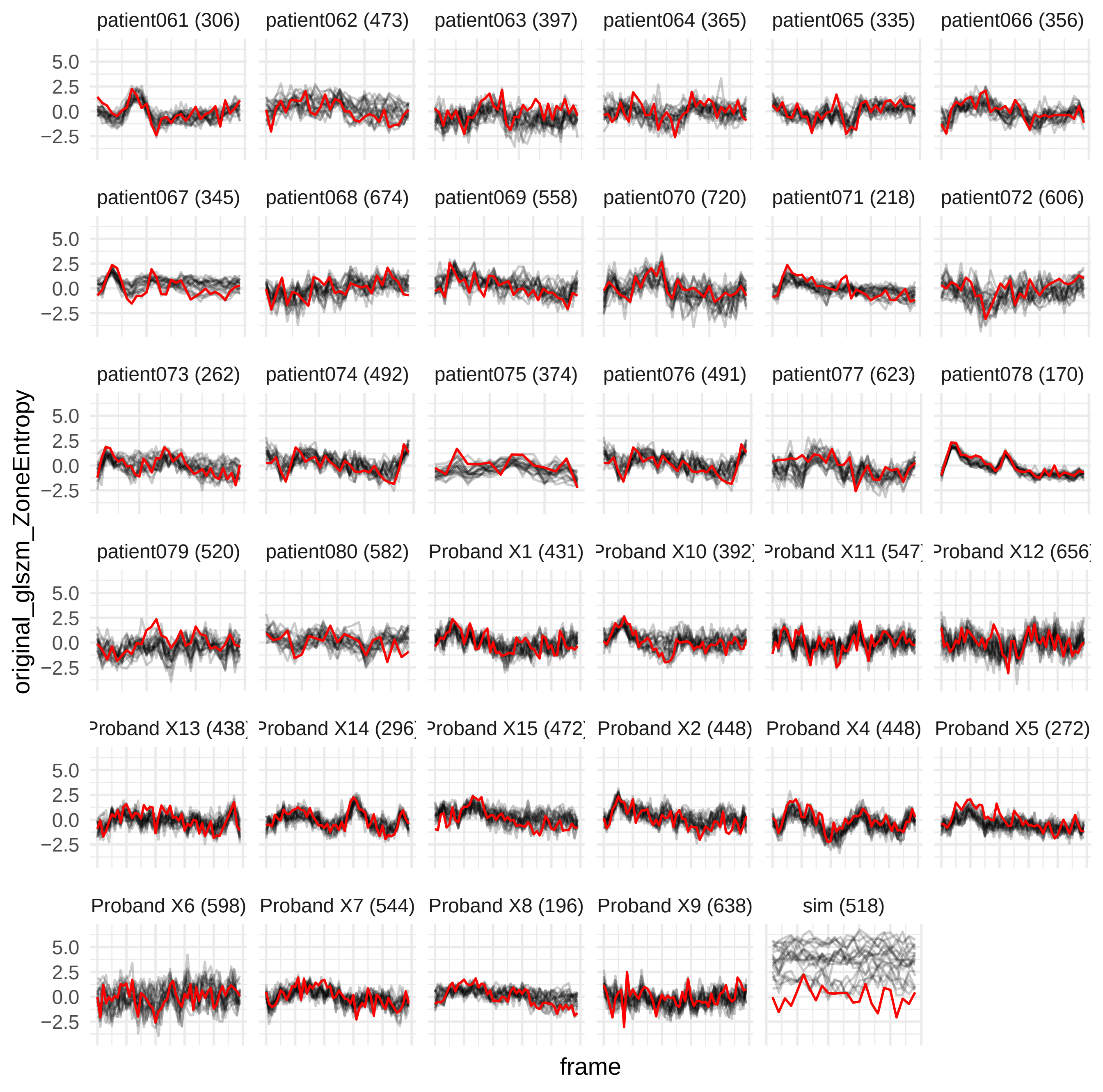

patient061 (306)
patient062 (473)
patient063 (397)
patient064 (365)
patient065 (335)
patient066 (356)
patient067 (345)
patient068 (674)
patient069 (558)
patient070 (720)
patient071 (218)
patient072 (606)
patient073 (262)
patient074 (492)
patient075 (374)
patient076 (491)
patient077 (623)
patient078 (170)
patient079 (520)
patient080 (582)
Proband X1 (431)
Proband X10 (392)
Proband X11 (547)
Proband X12 (656)
Proband X13 (438)
Proband X14 (296)
Proband X15 (472)
Proband X2 (448)
Proband X4 (448)
Proband X5 (272)
Proband X6 (598)
Proband X7 (544)
Proband X8 (196)
Proband X9 (638)
sim (518)
5.0
2.5
0.0
−2.5
original_glszm_ZoneEntropy
frame

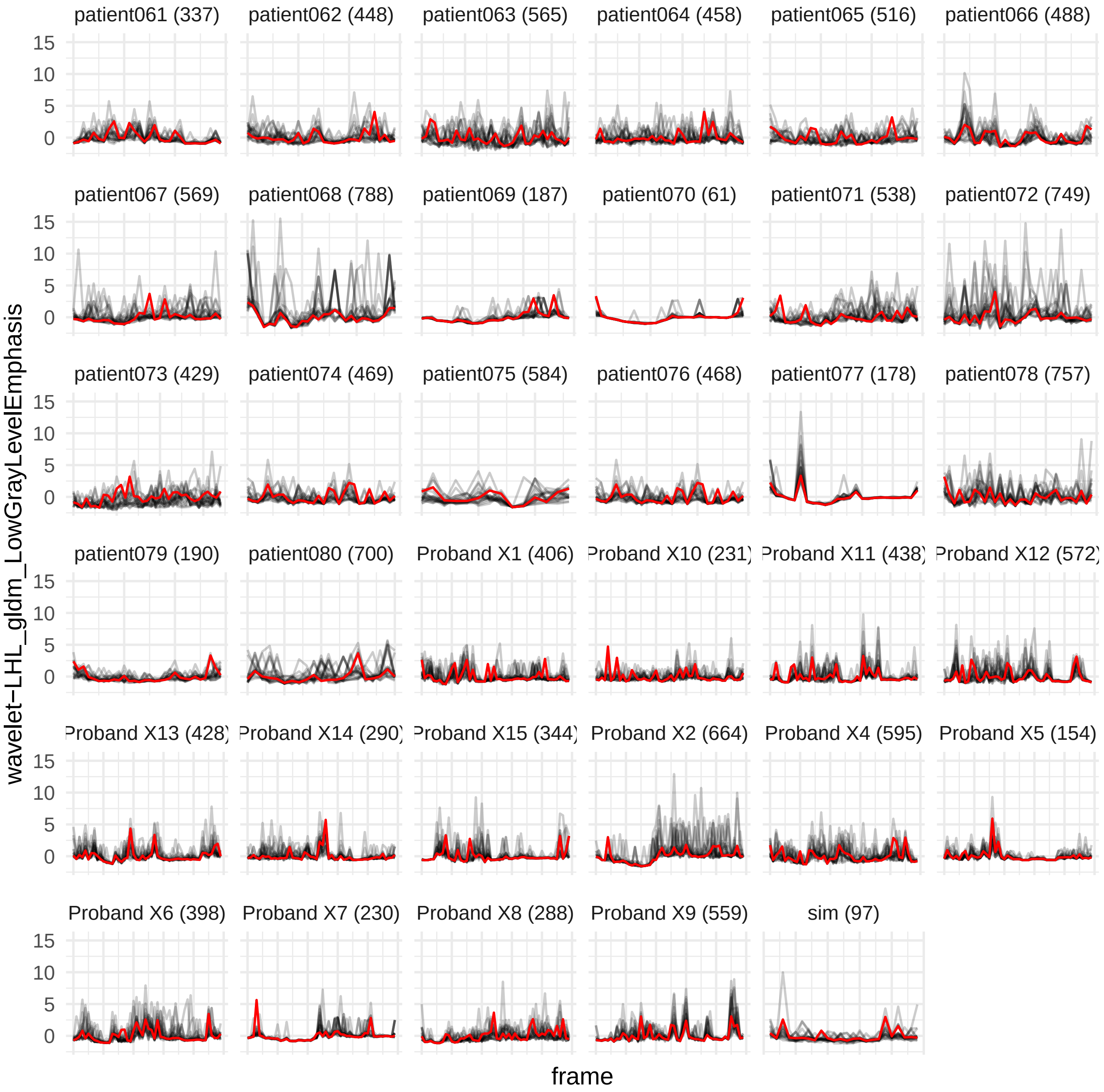
patient061 (337)
patient062 (448)
patient063 (565)
patient064 (458)
patient065 (516)
patient066 (488)
patient067 (569)
patient068 (788)
patient069 (187)
patient070 (61)
patient071 (538)
patient072 (749)
patient073 (429)
patient074 (469)
patient075 (584)
patient076 (468)
patient077 (178)
patient078 (757)
patient079 (190)
patient080 (700)
Proband X1 (406)
Proband X10 (231)
Proband X11 (438)
Proband X12 (572)
Proband X13 (428)
Proband X14 (290)
Proband X15 (344)
Proband X2 (664)
Proband X4 (595)
Proband X5 (154)
Proband X6 (398)
Proband X7 (230)
Proband X8 (288)
Proband X9 (559)
sim (97)
15
10
5
0
wavelet−LHL_gldm_LowGrayLevelEmphasis
frame

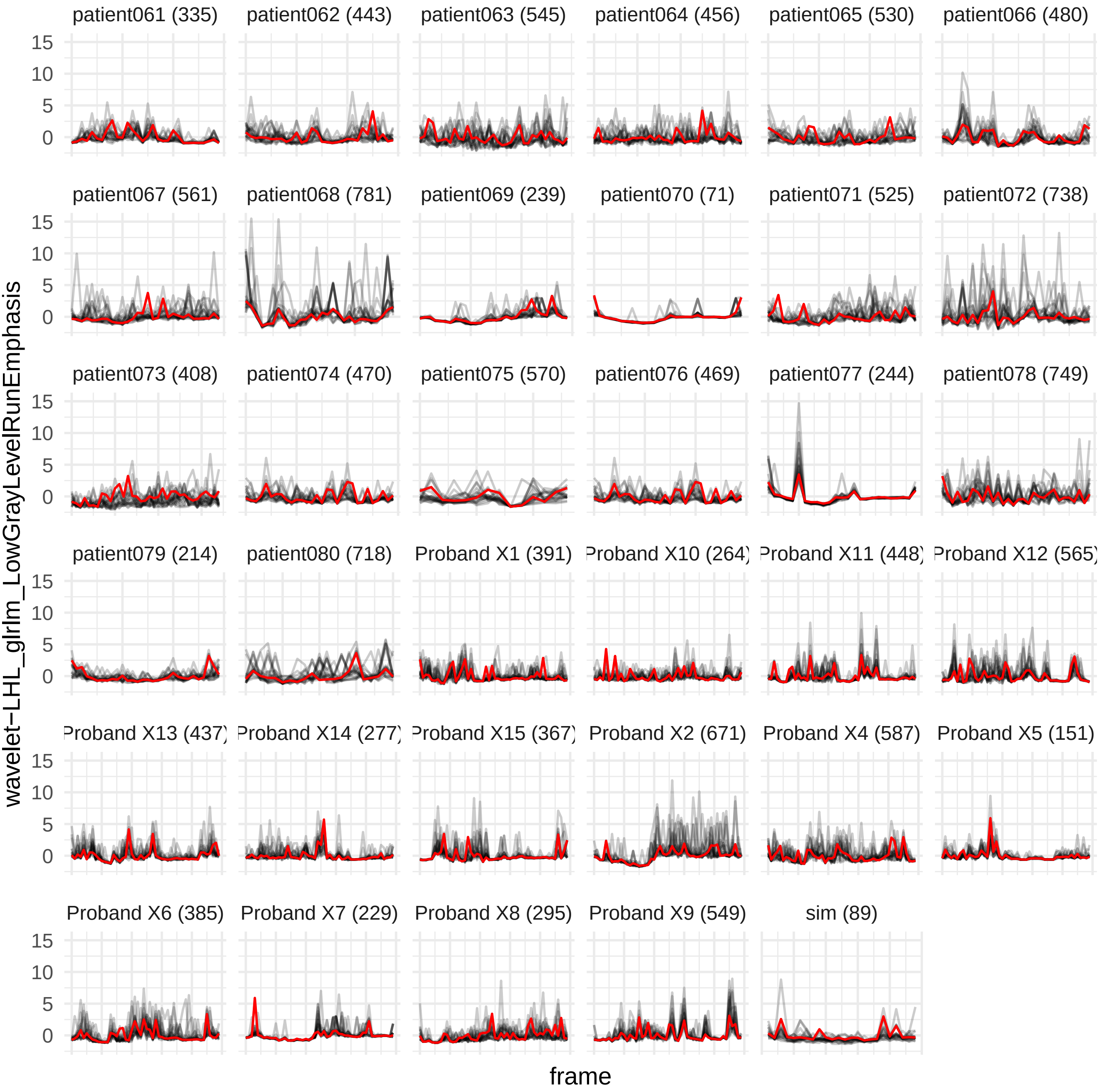

patient061 (335)
patient062 (443)
patient063 (545)
patient064 (456)
patient065 (530)
patient066 (480)
patient067 (561)
patient068 (781)
patient069 (239)
patient070 (71)
patient071 (525)
patient072 (738)
patient073 (408)
patient074 (470)
patient075 (570)
patient076 (469)
patient077 (244)
patient078 (749)
patient079 (214)
patient080 (718)
Proband X1 (391)
Proband X10 (264)
Proband X11 (448)
Proband X12 (565)
Proband X13 (437)
Proband X14 (277)
Proband X15 (367)
Proband X2 (671)
Proband X4 (587)
Proband X5 (151)
Proband X6 (385)
Proband X7 (229)
Proband X8 (295)
Proband X9 (549)
sim (89)
wavelet−LHL_glrlm_LowGrayLevelRunEmphasis
frame
0
5
10
15

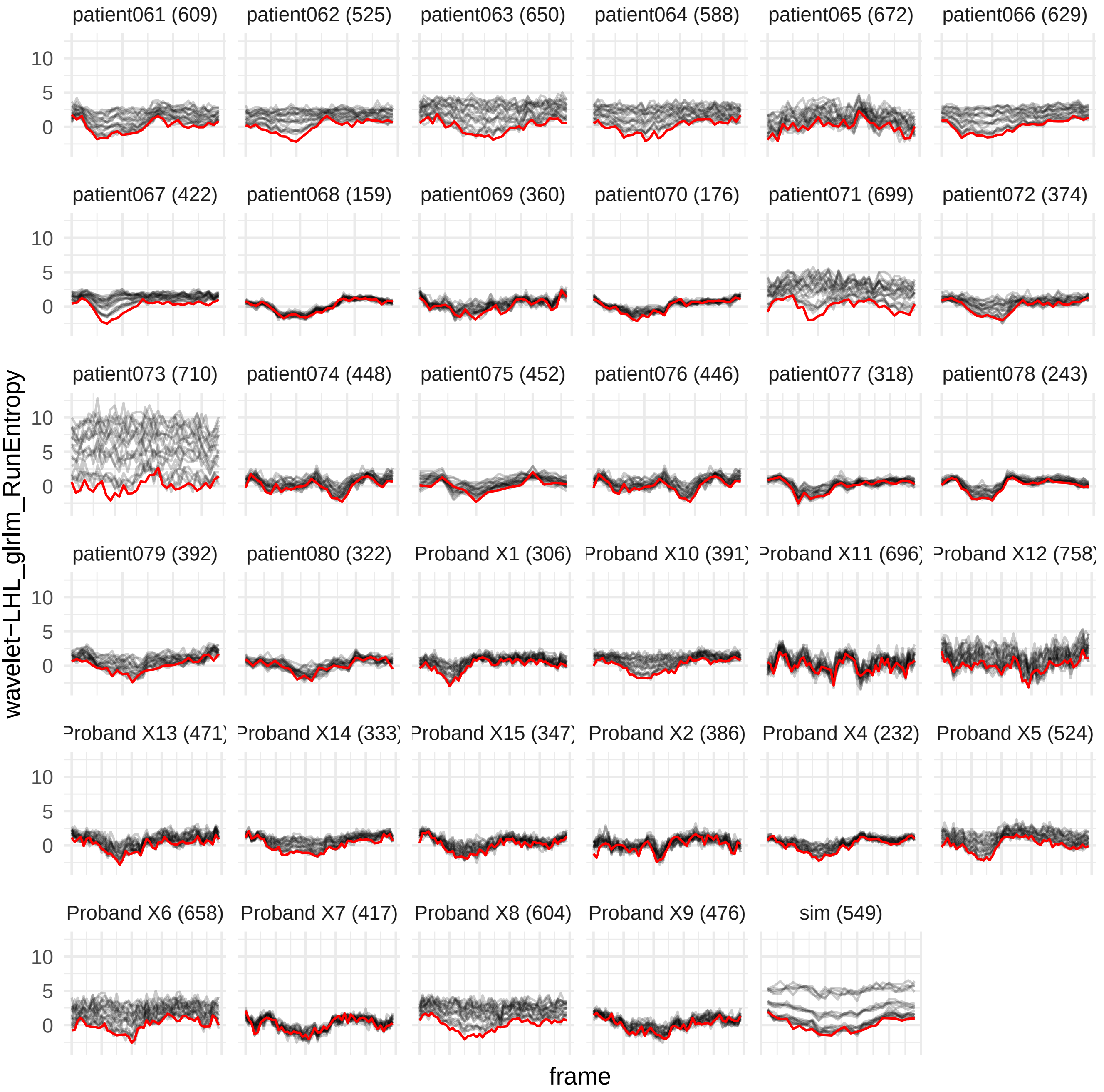

patient061 (609)
patient062 (525)
patient063 (650)
patient064 (588)
patient065 (672)
patient066 (629)
patient067 (422)
patient068 (159)
patient069 (360)
patient070 (176)
patient071 (699)
patient072 (374)
patient073 (710)
patient074 (448)
patient075 (452)
patient076 (446)
patient077 (318)
patient078 (243)
patient079 (392)
patient080 (322)
Proband X1 (306)
Proband X10 (391)
Proband X11 (696)
Proband X12 (758)
Proband X13 (471)
Proband X14 (333)
Proband X15 (347)
Proband X2 (386)
Proband X4 (232)
Proband X5 (524)
Proband X6 (658)
Proband X7 (417)
Proband X8 (604)
Proband X9 (476)
sim (549)
10
5
0
wavelet−LHL_glrlm_RunEntropy
frame

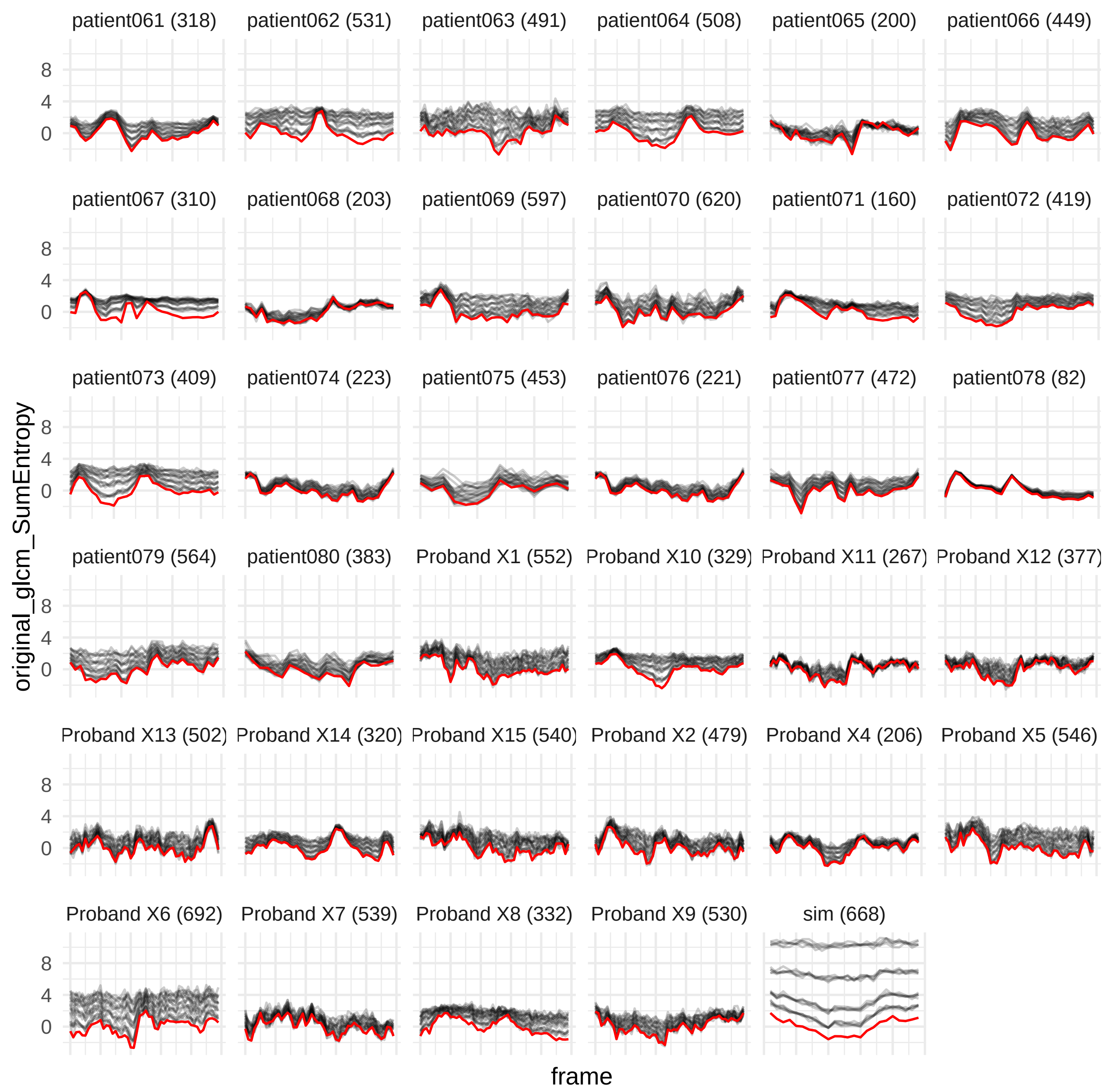
patient061 (318)
patient062 (531)
patient063 (491)
patient064 (508)
patient065 (200)
patient066 (449)
patient067 (310)
patient068 (203)
patient069 (597)
patient070 (620)
patient071 (160)
patient072 (419)
patient073 (409)
patient074 (223)
patient075 (453)
patient076 (221)
patient077 (472)
patient078 (82)
patient079 (564)
patient080 (383)
Proband X1 (552)
Proband X10 (329)
Proband X11 (267)
Proband X12 (377)
Proband X13 (502)
Proband X14 (320)
Proband X15 (540)
Proband X2 (479)
Proband X4 (206)
Proband X5 (546)
Proband X6 (692)
Proband X7 (539)
Proband X8 (332)
Proband X9 (530)
sim (668)
original_glcm_SumEntropy
frame
8
4
0

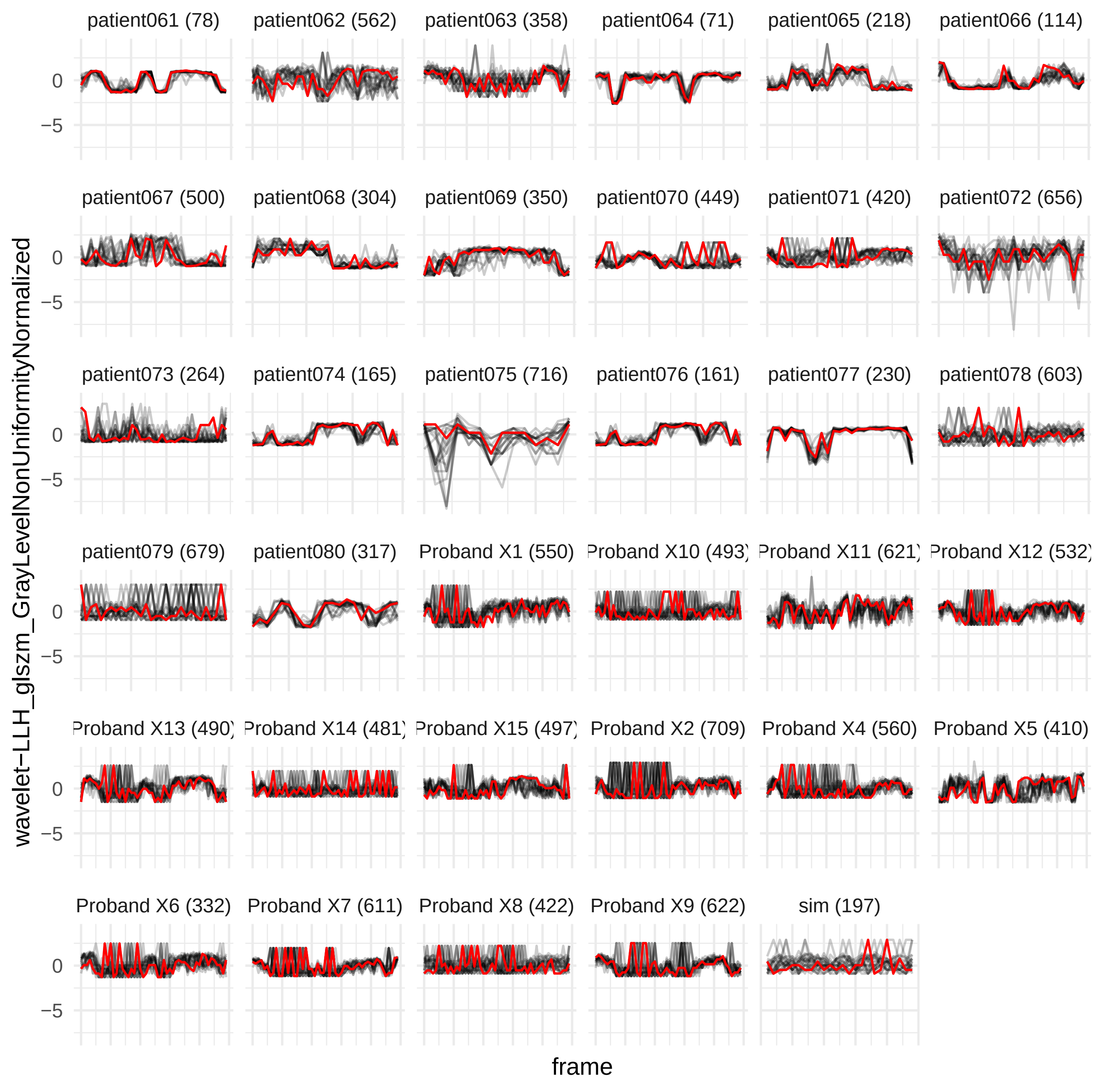

patient061 (78)
patient062 (562)
patient063 (358)
patient064 (71)
patient065 (218)
patient066 (114)
patient067 (500)
patient068 (304)
patient069 (350)
patient070 (449)
patient071 (420)
patient072 (656)
patient073 (264)
patient074 (165)
patient075 (716)
patient076 (161)
patient077 (230)
patient078 (603)
patient079 (679)
patient080 (317)
Proband X1 (550)
Proband X10 (493)
Proband X11 (621)
Proband X12 (532)
Proband X13 (490)
Proband X14 (481)
Proband X15 (497)
Proband X2 (709)
Proband X4 (560)
Proband X5 (410)
Proband X6 (332)
Proband X7 (611)
Proband X8 (422)
Proband X9 (622)
sim (197)
0
−5
wavelet−LLH_glszm_GrayLevelNonUniformityNormalized
frame

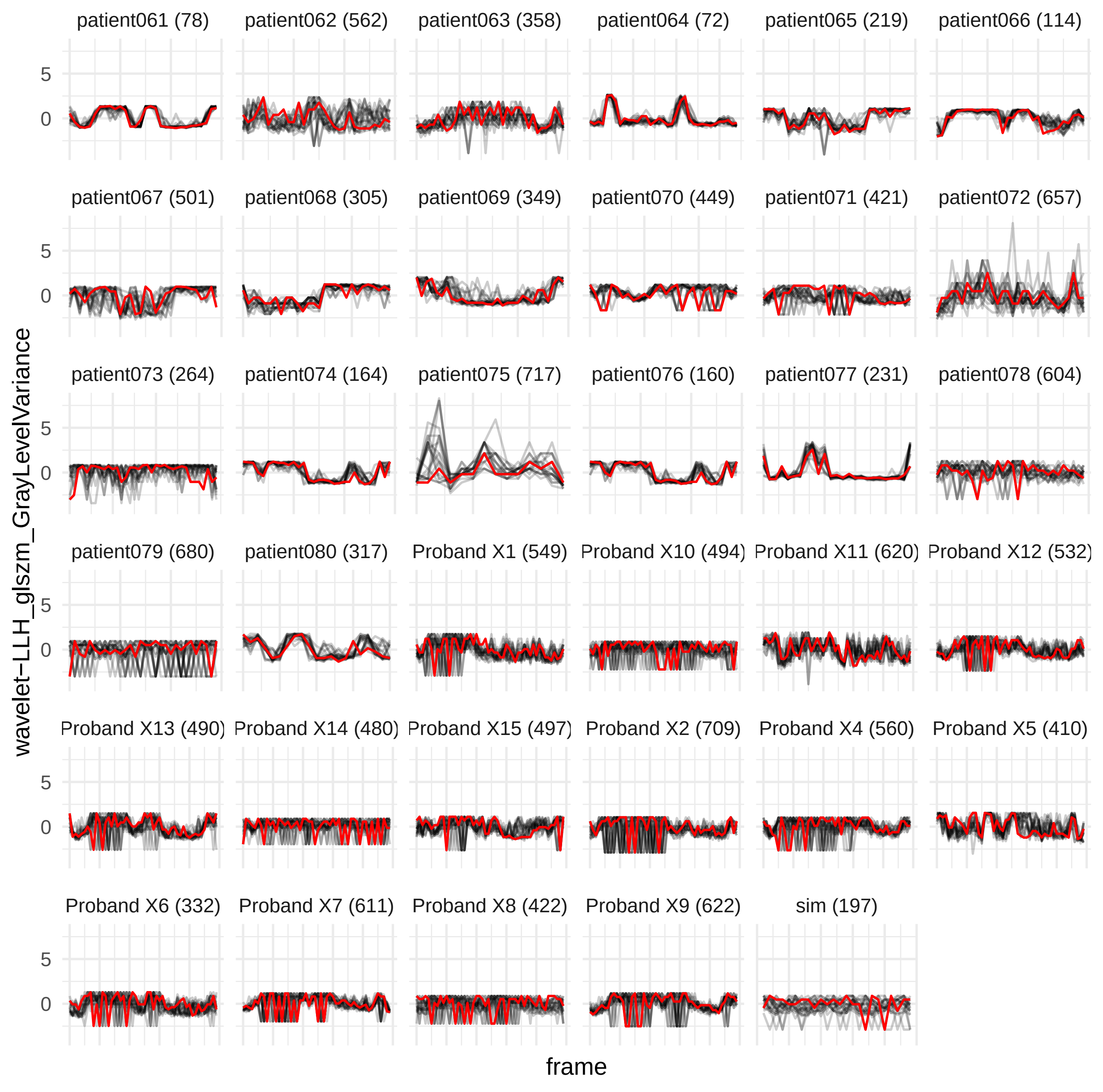

patient061 (78)
patient062 (562)
patient063 (358)
patient064 (72)
patient065 (219)
patient066 (114)
patient067 (501)
patient068 (305)
patient069 (349)
patient070 (449)
patient071 (421)
patient072 (657)
patient073 (264)
patient074 (164)
patient075 (717)
patient076 (160)
patient077 (231)
patient078 (604)
patient079 (680)
patient080 (317)
Proband X1 (549)
Proband X10 (494)
Proband X11 (620)
Proband X12 (532)
Proband X13 (490)
Proband X14 (480)
Proband X15 (497)
Proband X2 (709)
Proband X4 (560)
Proband X5 (410)
Proband X6 (332)
Proband X7 (611)
Proband X8 (422)
Proband X9 (622)
sim (197)
wavelet−LLH_glszm_GrayLevelVariance
5
0
frame

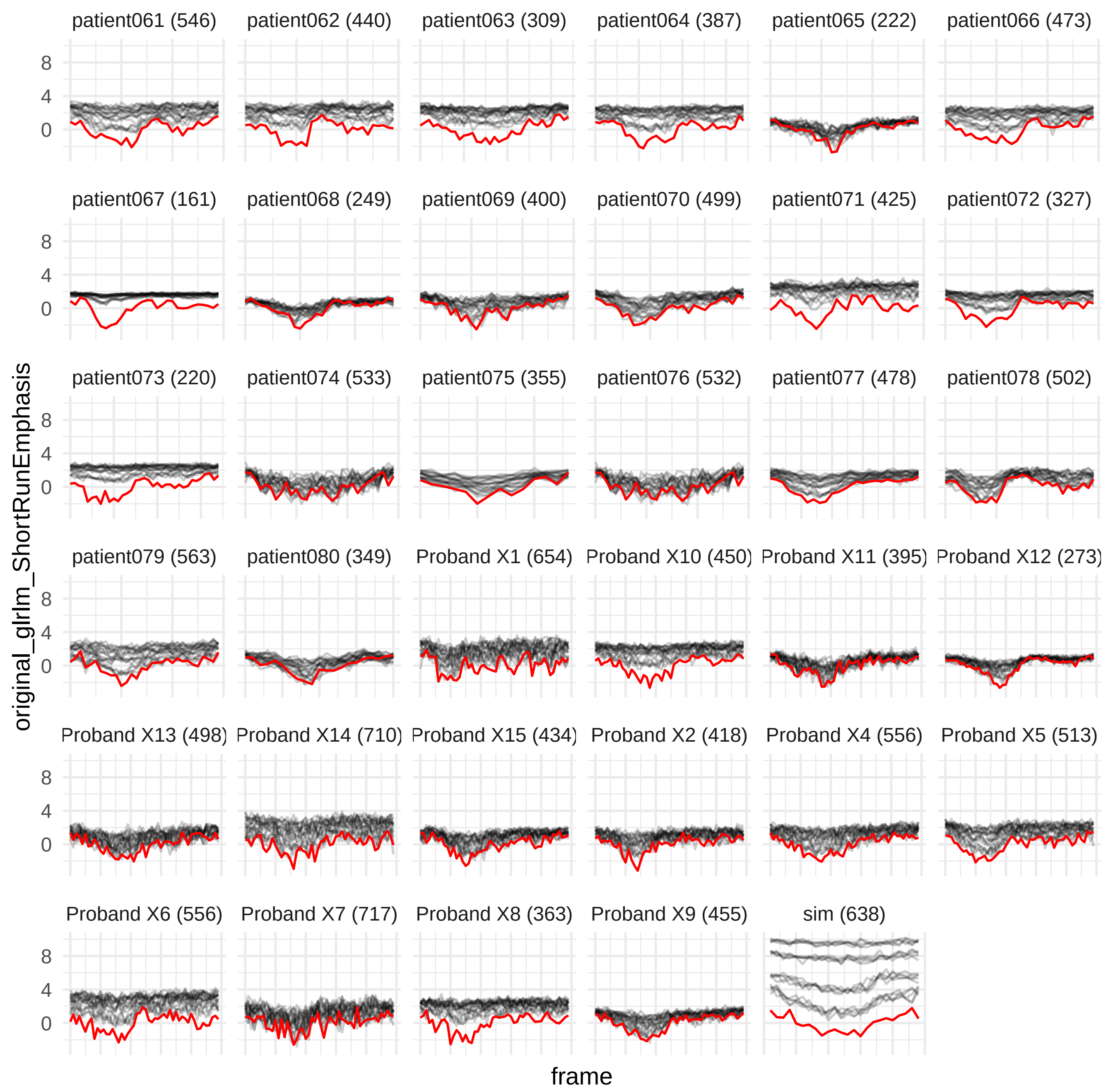

patient061 (546)
patient062 (440)
patient063 (309)
patient064 (387)
patient065 (222)
patient066 (473)
patient067 (161)
patient068 (249)
patient069 (400)
patient070 (499)
patient071 (425)
patient072 (327)
patient073 (220)
patient074 (533)
patient075 (355)
patient076 (532)
patient077 (478)
patient078 (502)
patient079 (563)
patient080 (349)
Proband X1 (654)
Proband X10 (450)
Proband X11 (395)
Proband X12 (273)
Proband X13 (498)
Proband X14 (710)
Proband X15 (434)
Proband X2 (418)
Proband X4 (556)
Proband X5 (513)
Proband X6 (556)
Proband X7 (717)
Proband X8 (363)
Proband X9 (455)
sim (638)
8
4
0
original_glrlm_ShortRunEmphasis
frame

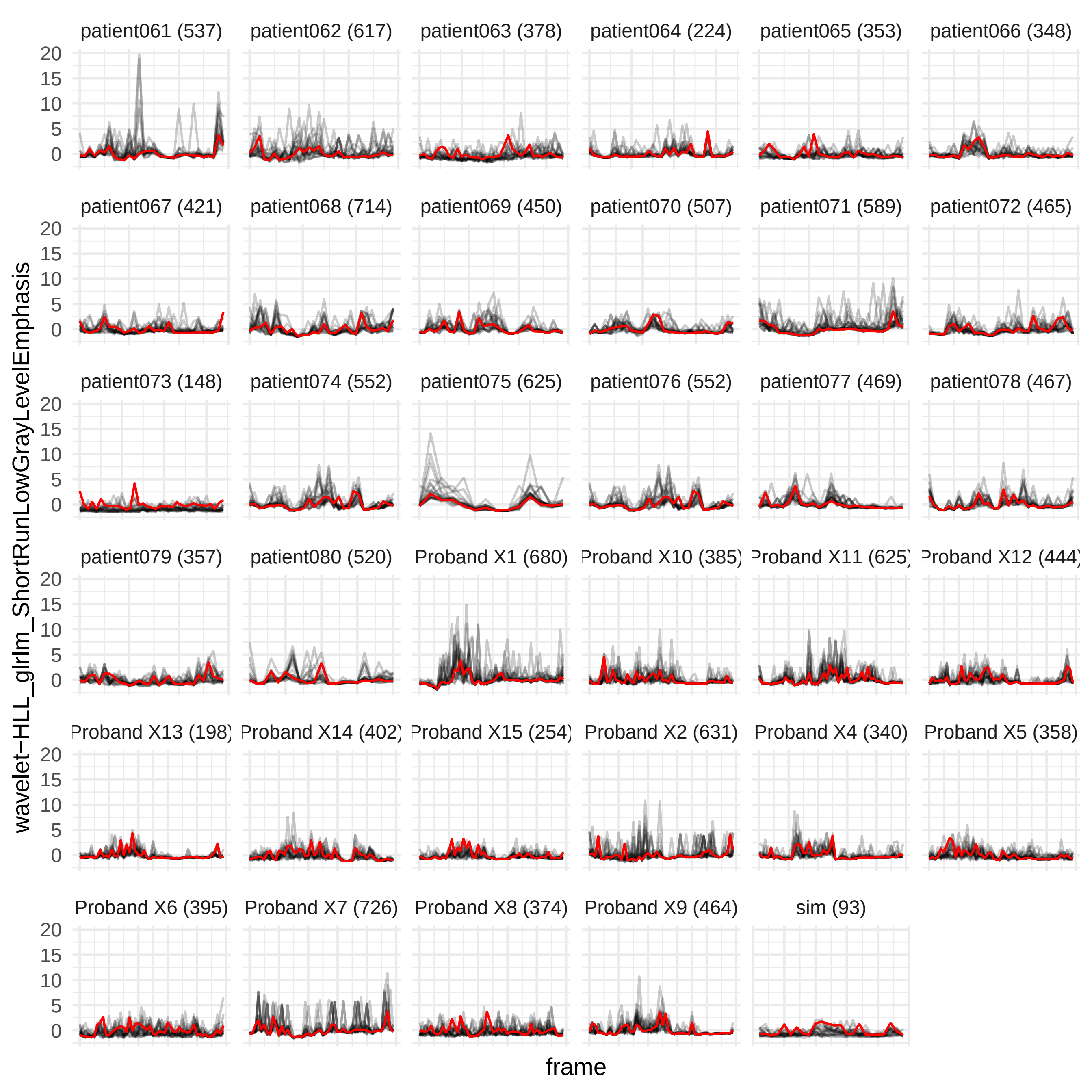

patient061 (537)
patient062 (617)
patient063 (378)
patient064 (224)
patient065 (353)
patient066 (348)
patient067 (421)
patient068 (714)
patient069 (450)
patient070 (507)
patient071 (589)
patient072 (465)
patient073 (148)
patient074 (552)
patient075 (625)
patient076 (552)
patient077 (469)
patient078 (467)
patient079 (357)
patient080 (520)
Proband X1 (680)
Proband X10 (385)
Proband X11 (625)
Proband X12 (444)
Proband X13 (198)
Proband X14 (402)
Proband X15 (254)
Proband X2 (631)
Proband X4 (340)
Proband X5 (358)
Proband X6 (395)
Proband X7 (726)
Proband X8 (374)
Proband X9 (464)
sim (93)
20
15
10
5
0
wavelet−HLL_glrlm_ShortRunLowGrayLevelEmphasis
frame

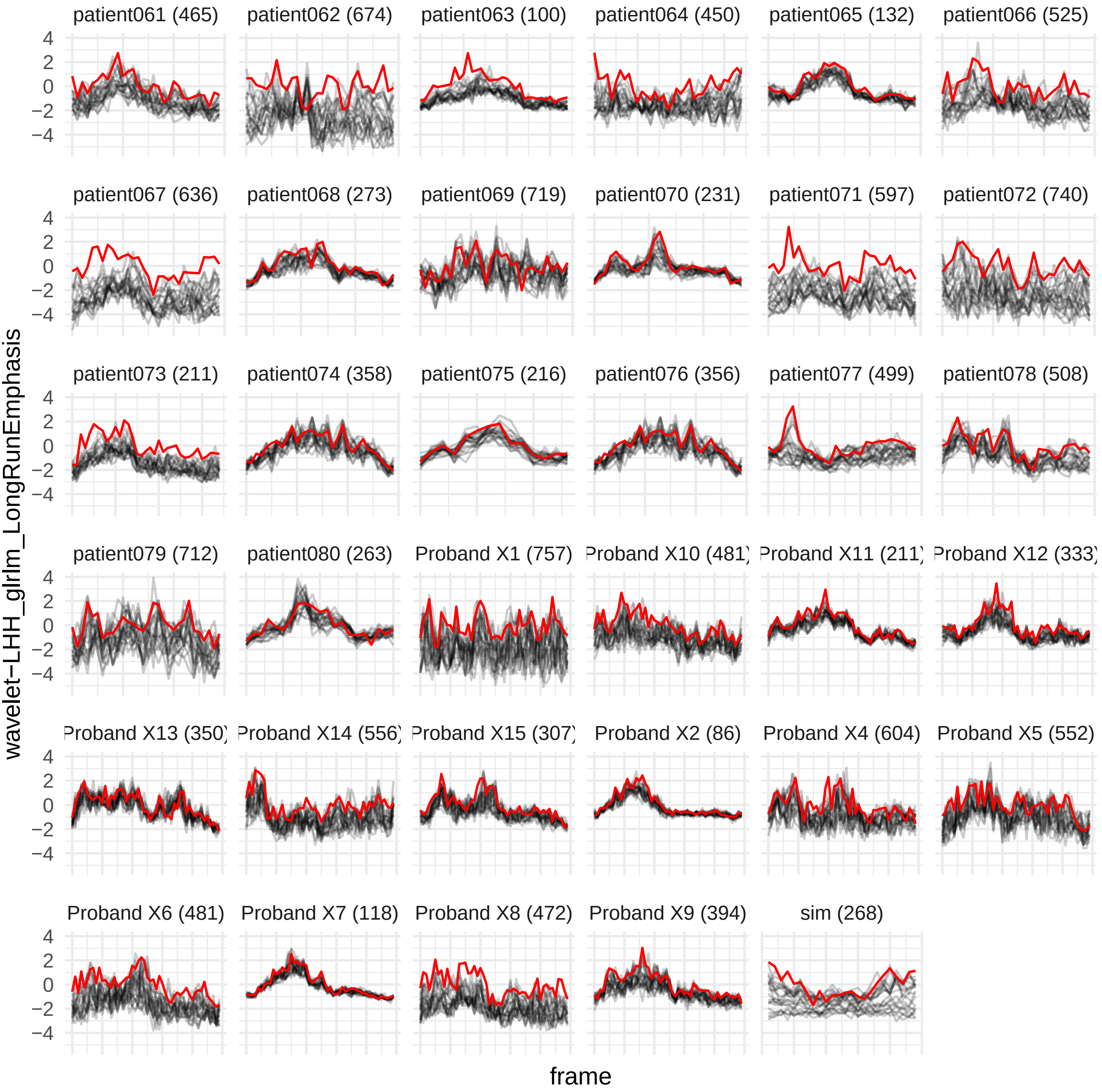

patient061 (465)
patient062 (674)
patient063 (100)
patient064 (450)
patient065 (132)
patient066 (525)
patient067 (636)
patient068 (273)
patient069 (719)
patient070 (231)
patient071 (597)
patient072 (740)
patient073 (211)
patient074 (358)
patient075 (216)
patient076 (356)
patient077 (499)
patient078 (508)
patient079 (712)
patient080 (263)
Proband X1 (757)
Proband X10 (481)
Proband X11 (211)
Proband X12 (333)
Proband X13 (350)
Proband X14 (556)
Proband X15 (307)
Proband X2 (86)
Proband X4 (604)
Proband X5 (552)
Proband X6 (481)
Proband X7 (118)
Proband X8 (472)
Proband X9 (394)
sim (268)
wavelet−LHH_glrlm_LongRunEmphasis
frame
4
2
0
−2
−4

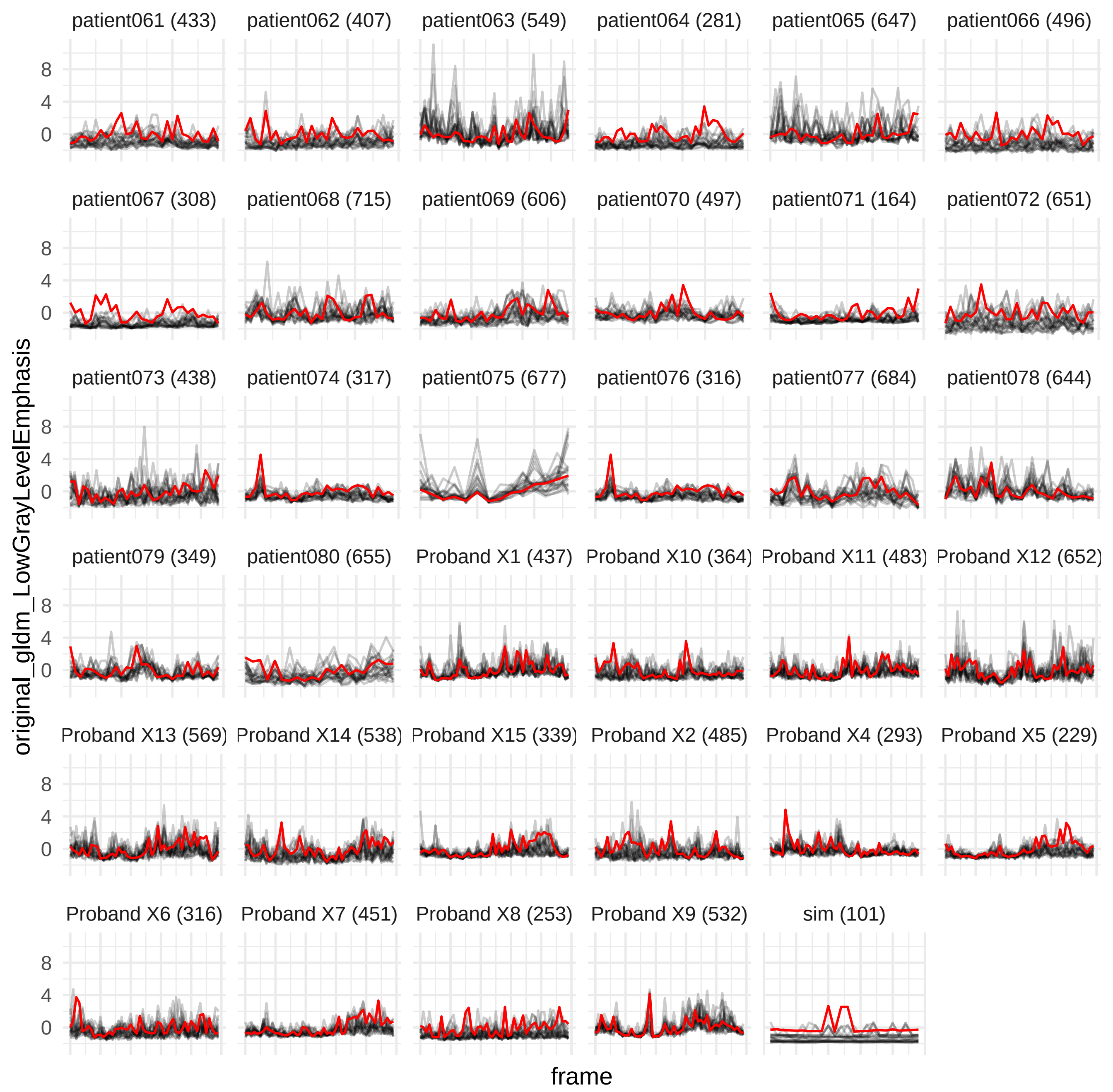

patient061 (433)
patient062 (407)
patient063 (549)
patient064 (281)
patient065 (647)
patient066 (496)
patient067 (308)
patient068 (715)
patient069 (606)
patient070 (497)
patient071 (164)
patient072 (651)
patient073 (438)
patient074 (317)
patient075 (677)
patient076 (316)
patient077 (684)
patient078 (644)
patient079 (349)
patient080 (655)
Proband X1 (437)
Proband X10 (364)
Proband X11 (483)
Proband X12 (652)
Proband X13 (569)
Proband X14 (538)
Proband X15 (339)
Proband X2 (485)
Proband X4 (293)
Proband X5 (229)
Proband X6 (316)
Proband X7 (451)
Proband X8 (253)
Proband X9 (532)
sim (101)
original_gldm_LowGrayLevelEmphasis
frame
0
4
8

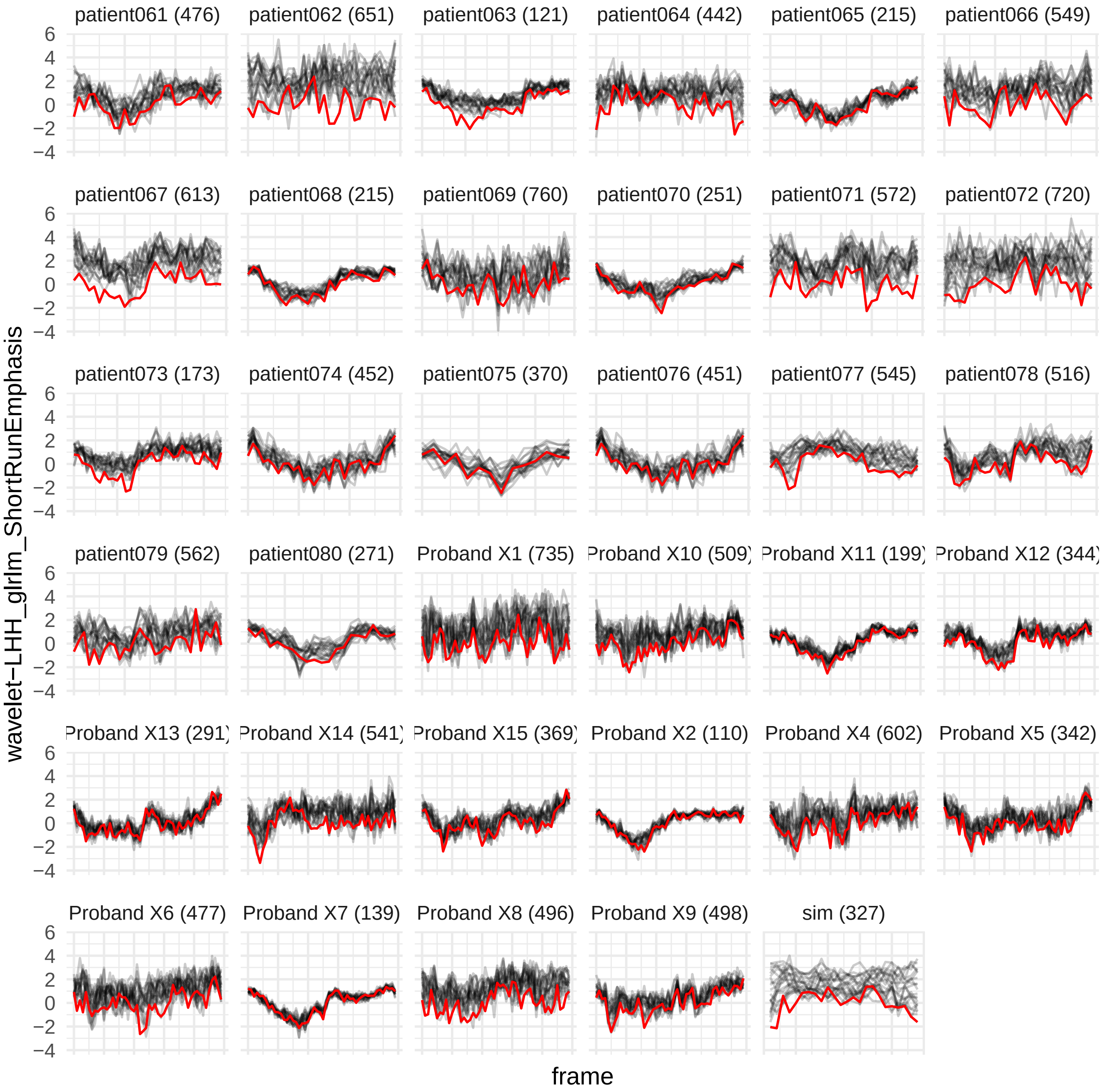
patient061 (476)
patient062 (651)
patient063 (121)
patient064 (442)
patient065 (215)
patient066 (549)
patient067 (613)
patient068 (215)
patient069 (760)
patient070 (251)
patient071 (572)
patient072 (720)
patient073 (173)
patient074 (452)
patient075 (370)
patient076 (451)
patient077 (545)
patient078 (516)
patient079 (562)
patient080 (271)
Proband X1 (735)
Proband X10 (509)
Proband X11 (199)
Proband X12 (344)
Proband X13 (291)
Proband X14 (541)
Proband X15 (369)
Proband X2 (110)
Proband X4 (602)
Proband X5 (342)
Proband X6 (477)
Proband X7 (139)
Proband X8 (496)
Proband X9 (498)
sim (327)
wavelet−LHH_glrlm_ShortRunEmphasis
frame
6
4
2
0
−2
−4

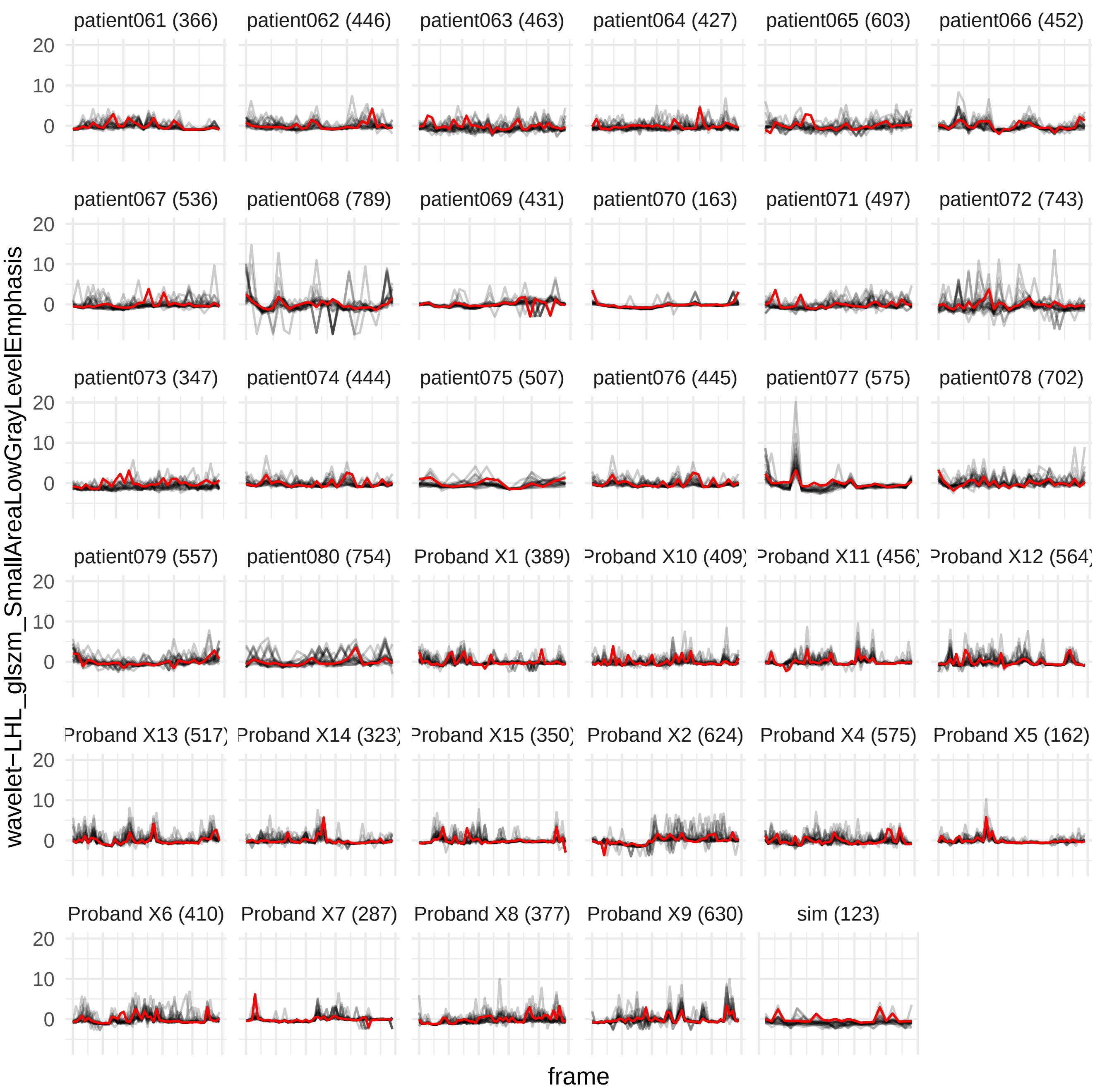

patient061 (366)
patient062 (446)
patient063 (463)
patient064 (427)
patient065 (603)
patient066 (452)
patient067 (536)
patient068 (789)
patient069 (431)
patient070 (163)
patient071 (497)
patient072 (743)
patient073 (347)
patient074 (444)
patient075 (507)
patient076 (445)
patient077 (575)
patient078 (702)
patient079 (557)
patient080 (754)
Proband X1 (389)
Proband X10 (409)
Proband X11 (456)
Proband X12 (564)
Proband X13 (517)
Proband X14 (323)
Proband X15 (350)
Proband X2 (624)
Proband X4 (575)
Proband X5 (162)
Proband X6 (410)
Proband X7 (287)
Proband X8 (377)
Proband X9 (630)
sim (123)
20
10
0
wavelet−LHL_glszm_SmallAreaLowGrayLevelEmphasis
frame

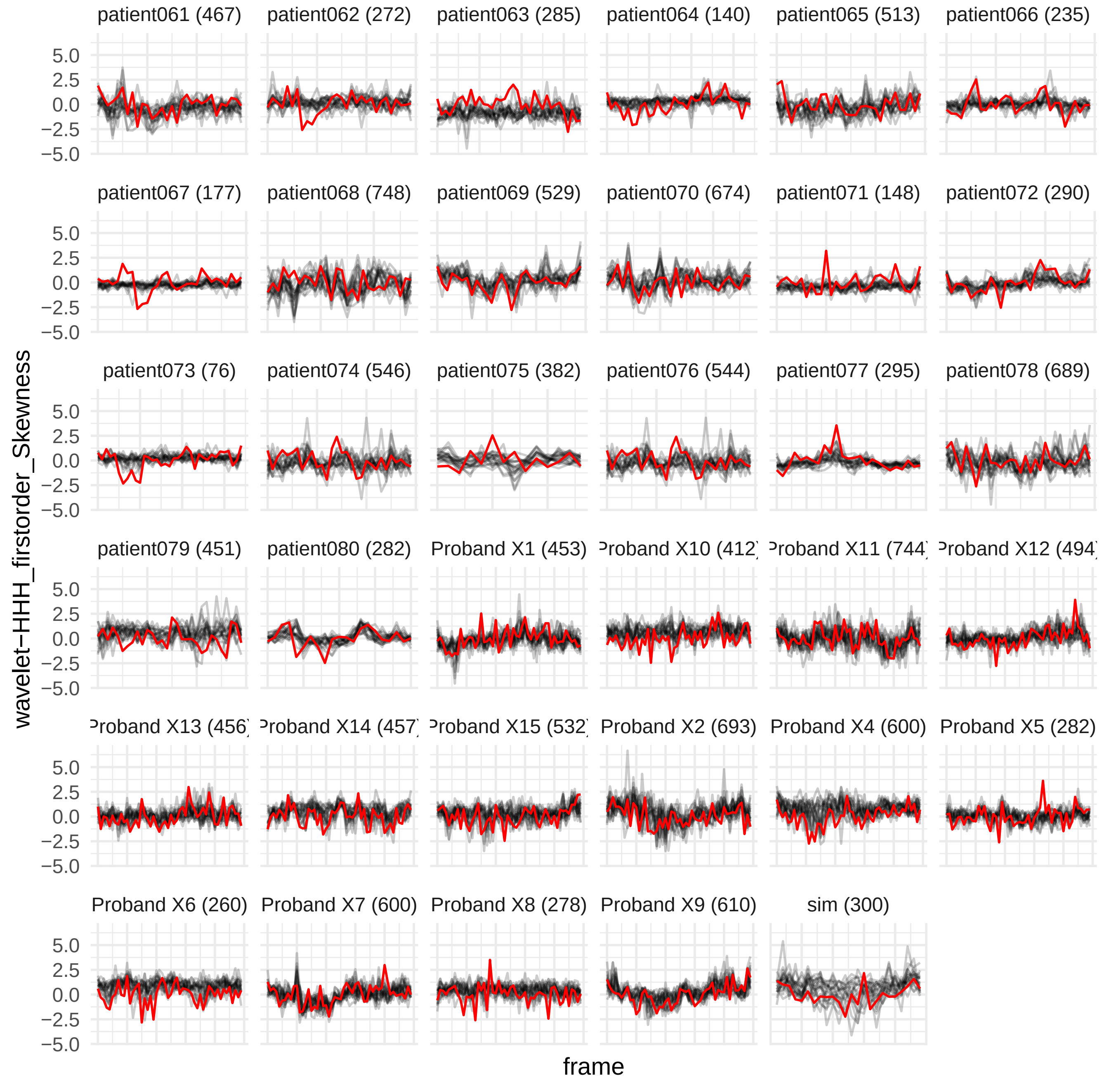
patient061 (467)
patient062 (272)
patient063 (285)
patient064 (140)
patient065 (513)
patient066 (235)
patient067 (177)
patient068 (748)
patient069 (529)
patient070 (674)
patient071 (148)
patient072 (290)
patient073 (76)
patient074 (546)
patient075 (382)
patient076 (544)
patient077 (295)
patient078 (689)
patient079 (451)
patient080 (282)
Proband X1 (453)
Proband X10 (412)
Proband X11 (744)
Proband X12 (494)
Proband X13 (456)
Proband X14 (457)
Proband X15 (532)
Proband X2 (693)
Proband X4 (600)
Proband X5 (282)
Proband X6 (260)
Proband X7 (600)
Proband X8 (278)
Proband X9 (610)
sim (300)
5.0
2.5
0.0
−2.5
−5.0
wavelet−HHH_firstorder_Skewness
frame

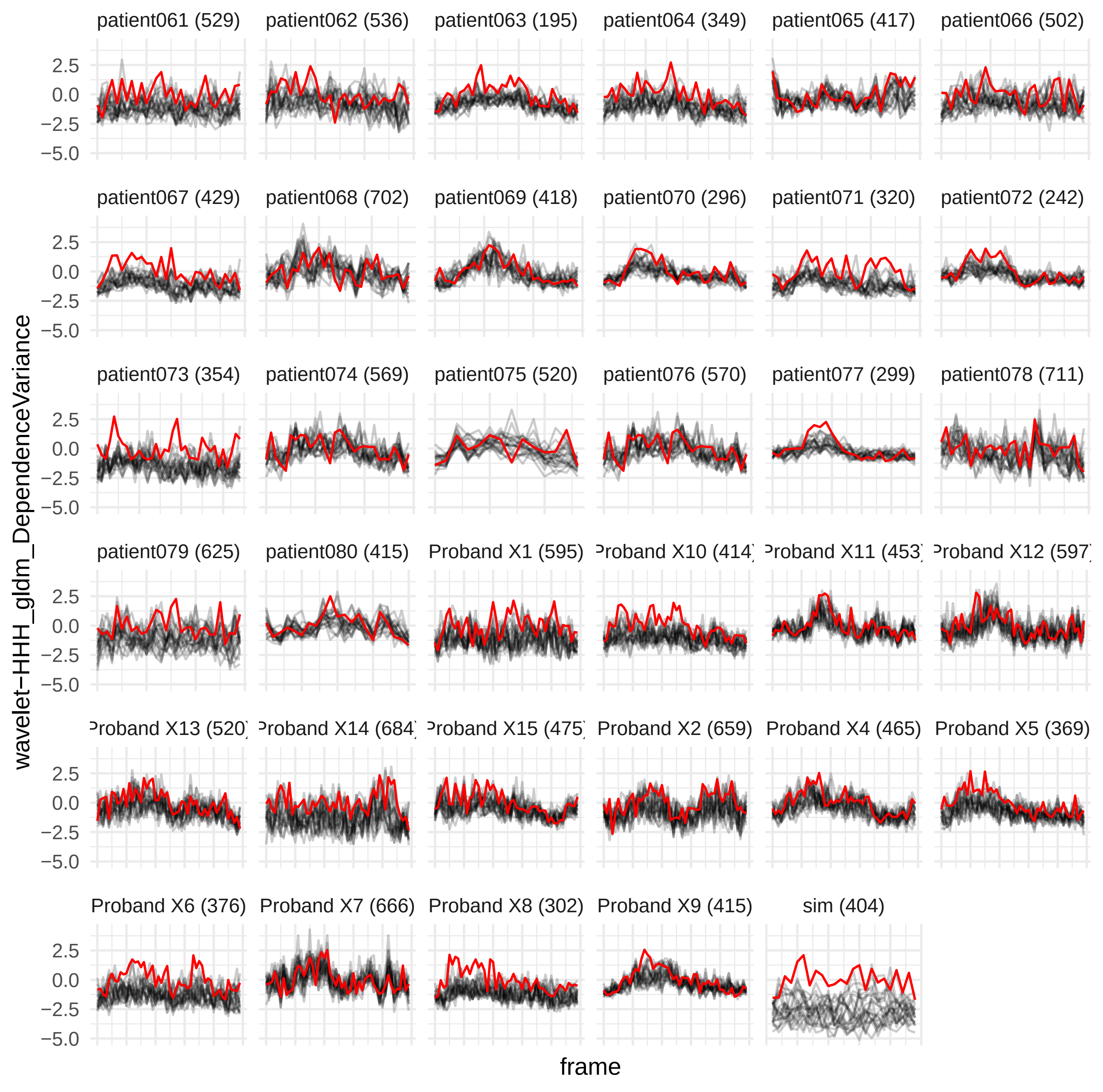
patient061 (529)
patient062 (536)
patient063 (195)
patient064 (349)
patient065 (417)
patient066 (502)
patient067 (429)
patient068 (702)
patient069 (418)
patient070 (296)
patient071 (320)
patient072 (242)
patient073 (354)
patient074 (569)
patient075 (520)
patient076 (570)
patient077 (299)
patient078 (711)
patient079 (625)
patient080 (415)
Proband X1 (595)
Proband X10 (414)
Proband X11 (453)
Proband X12 (597)
Proband X13 (520)
Proband X14 (684)
Proband X15 (475)
Proband X2 (659)
Proband X4 (465)
Proband X5 (369)
Proband X6 (376)
Proband X7 (666)
Proband X8 (302)
Proband X9 (415)
sim (404)
2.5
0.0
−2.5
−5.0
wavelet−HHH_gldm_DependenceVariance
frame

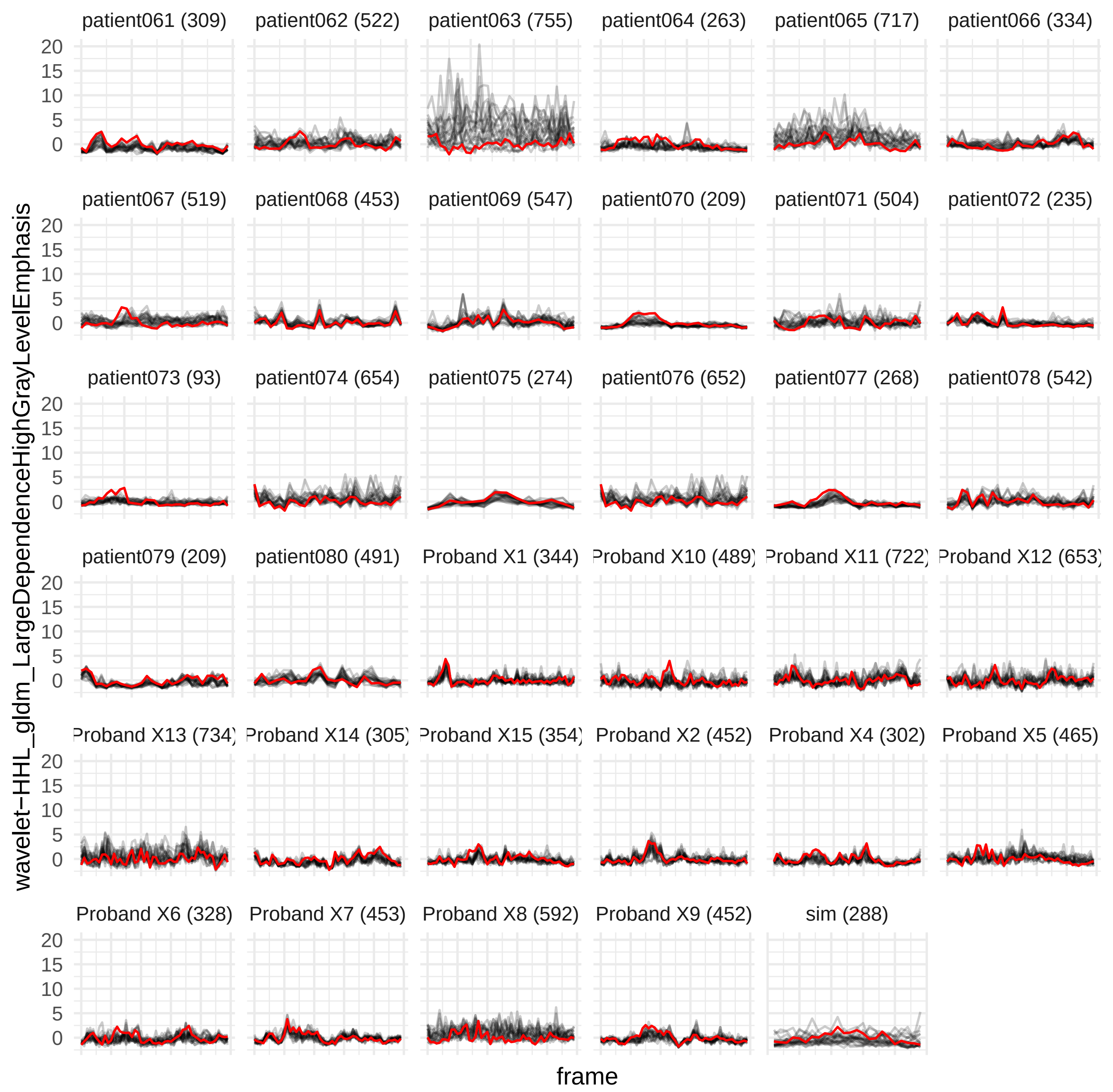
patient061 (309)
patient062 (522)
patient063 (755)
patient064 (263)
patient065 (717)
patient066 (334)
patient067 (519)
patient068 (453)
patient069 (547)
patient070 (209)
patient071 (504)
patient072 (235)
patient073 (93)
patient074 (654)
patient075 (274)
patient076 (652)
patient077 (268)
patient078 (542)
patient079 (209)
patient080 (491)
Proband X1 (344)
Proband X10 (489)
Proband X11 (722)
Proband X12 (653)
Proband X13 (734)
Proband X14 (305)
Proband X15 (354)
Proband X2 (452)
Proband X4 (302)
Proband X5 (465)
Proband X6 (328)
Proband X7 (453)
Proband X8 (592)
Proband X9 (452)
sim (288)
20
15
10
5
0
wavelet−HHL_gldm_LargeDependenceHighGrayLevelEmphasis
frame

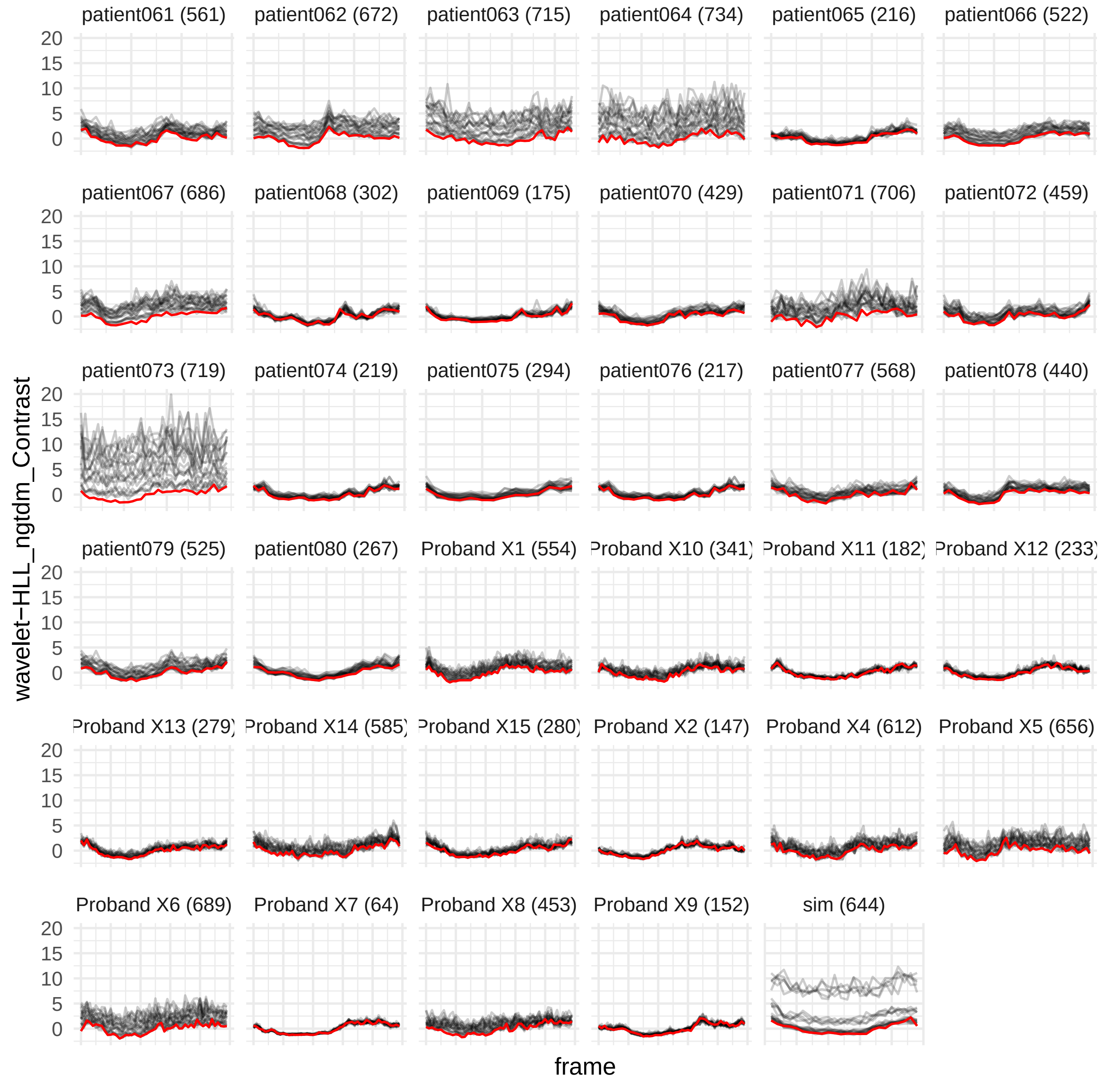

patient061 (561)
patient062 (672)
patient063 (715)
patient064 (734)
patient065 (216)
patient066 (522)
patient067 (686)
patient068 (302)
patient069 (175)
patient070 (429)
patient071 (706)
patient072 (459)
patient073 (719)
patient074 (219)
patient075 (294)
patient076 (217)
patient077 (568)
patient078 (440)
patient079 (525)
patient080 (267)
Proband X1 (554)
Proband X10 (341)
Proband X11 (182)
Proband X12 (233)
Proband X13 (279)
Proband X14 (585)
Proband X15 (280)
Proband X2 (147)
Proband X4 (612)
Proband X5 (656)
Proband X6 (689)
Proband X7 (64)
Proband X8 (453)
Proband X9 (152)
sim (644)
wavelet−HLL_ngtdm_Contrast
frame
0
5
10
15
20

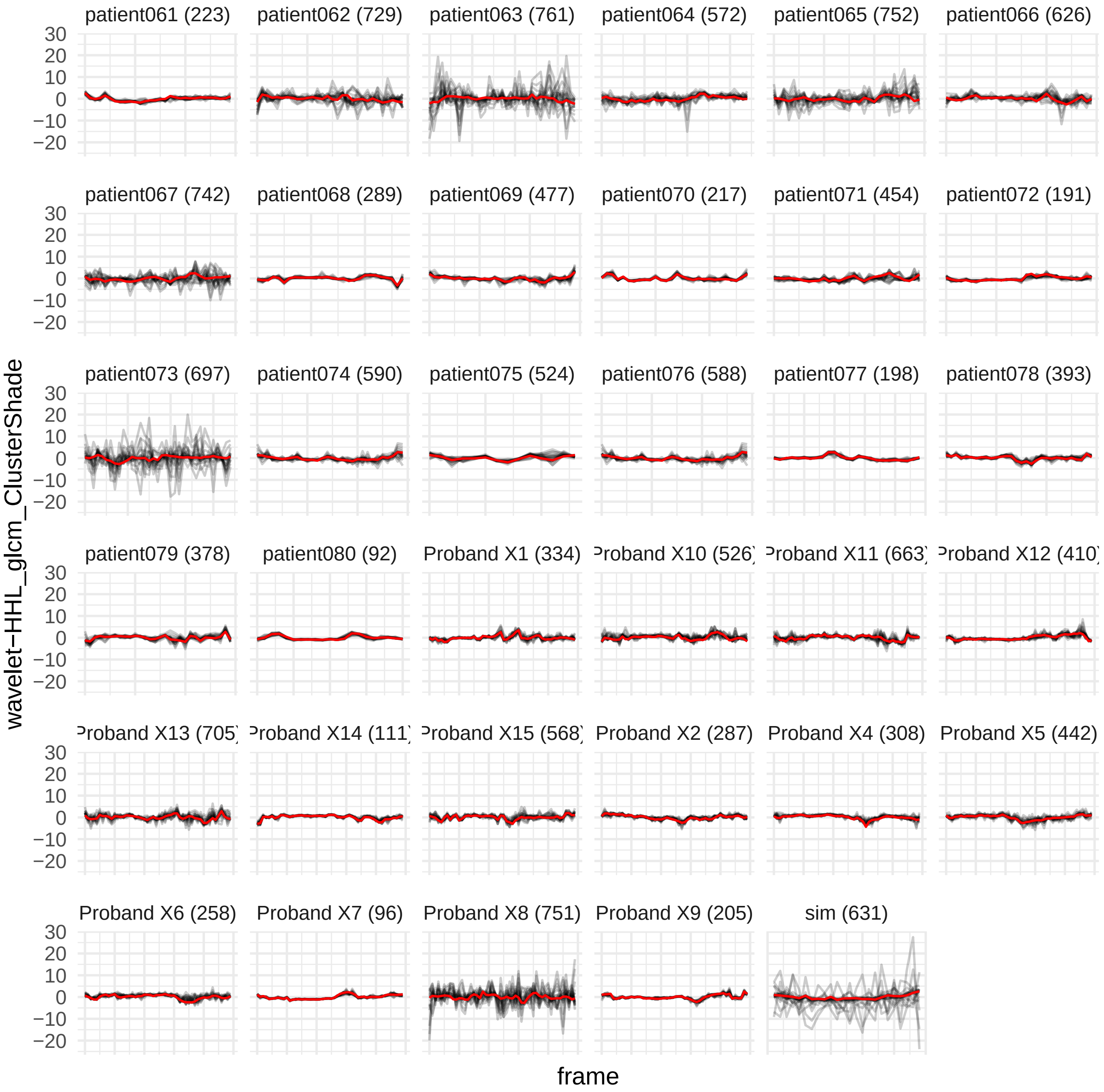

patient061 (223)
patient062 (729)
patient063 (761)
patient064 (572)
patient065 (752)
patient066 (626)
patient067 (742)
patient068 (289)
patient069 (477)
patient070 (217)
patient071 (454)
patient072 (191)
patient073 (697)
patient074 (590)
patient075 (524)
patient076 (588)
patient077 (198)
patient078 (393)
patient079 (378)
patient080 (92)
Proband X1 (334)
Proband X10 (526)
Proband X11 (663)
Proband X12 (410)
Proband X13 (705)
Proband X14 (111)
Proband X15 (568)
Proband X2 (287)
Proband X4 (308)
Proband X5 (442)
Proband X6 (258)
Proband X7 (96)
Proband X8 (751)
Proband X9 (205)
sim (631)
30
20
10
0
−10
−20
wavelet−HHL_glcm_ClusterShade
frame

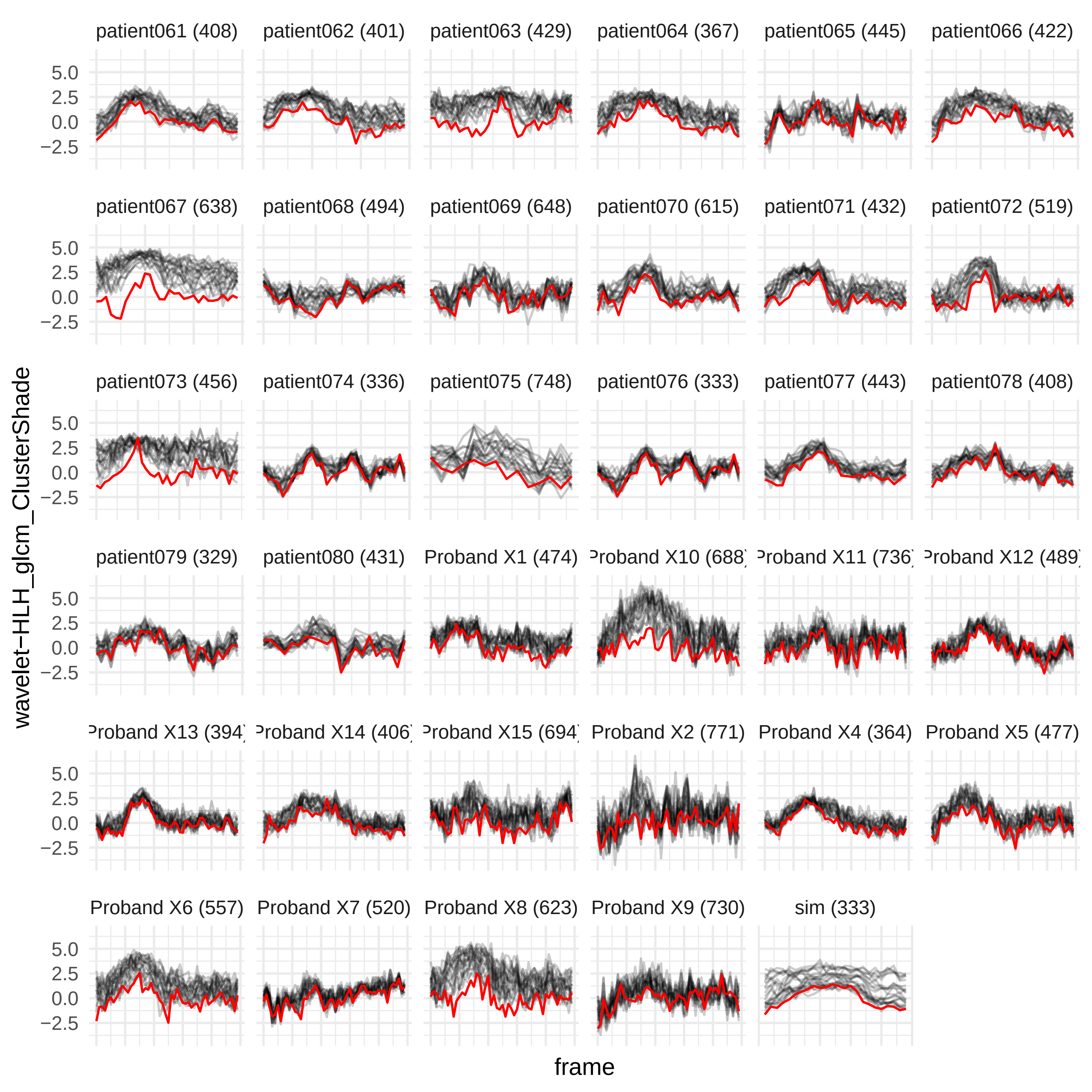

patient061 (408)
patient062 (401)
patient063 (429)
patient064 (367)
patient065 (445)
patient066 (422)
patient067 (638)
patient068 (494)
patient069 (648)
patient070 (615)
patient071 (432)
patient072 (519)
patient073 (456)
patient074 (336)
patient075 (748)
patient076 (333)
patient077 (443)
patient078 (408)
patient079 (329)
patient080 (431)
Proband X1 (474)
Proband X10 (688)
Proband X11 (736)
Proband X12 (489)
Proband X13 (394)
Proband X14 (406)
Proband X15 (694)
Proband X2 (771)
Proband X4 (364)
Proband X5 (477)
Proband X6 (557)
Proband X7 (520)
Proband X8 (623)
Proband X9 (730)
sim (333)
5.0
2.5
0.0
−2.5
wavelet−HLH_glcm_ClusterShade
frame

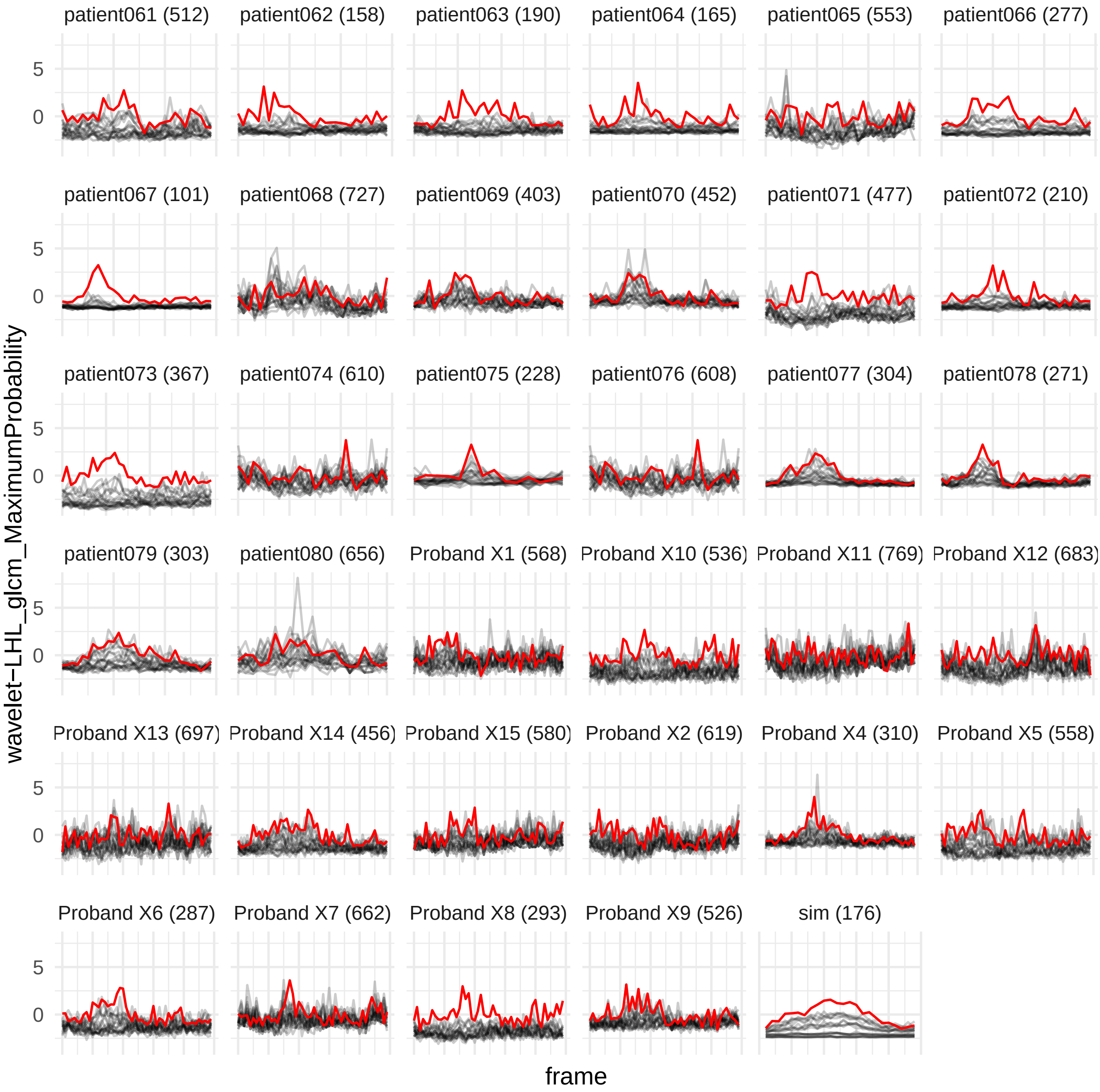

patient061 (512)
patient062 (158)
patient063 (190)
patient064 (165)
patient065 (553)
patient066 (277)
patient067 (101)
patient068 (727)
patient069 (403)
patient070 (452)
patient071 (477)
patient072 (210)
patient073 (367)
patient074 (610)
patient075 (228)
patient076 (608)
patient077 (304)
patient078 (271)
patient079 (303)
patient080 (656)
Proband X1 (568)
Proband X10 (536)
Proband X11 (769)
Proband X12 (683)
Proband X13 (697)
Proband X14 (456)
Proband X15 (580)
Proband X2 (619)
Proband X4 (310)
Proband X5 (558)
Proband X6 (287)
Proband X7 (662)
Proband X8 (293)
Proband X9 (526)
sim (176)
5
0
wavelet−LHL_glcm_MaximumProbability
frame

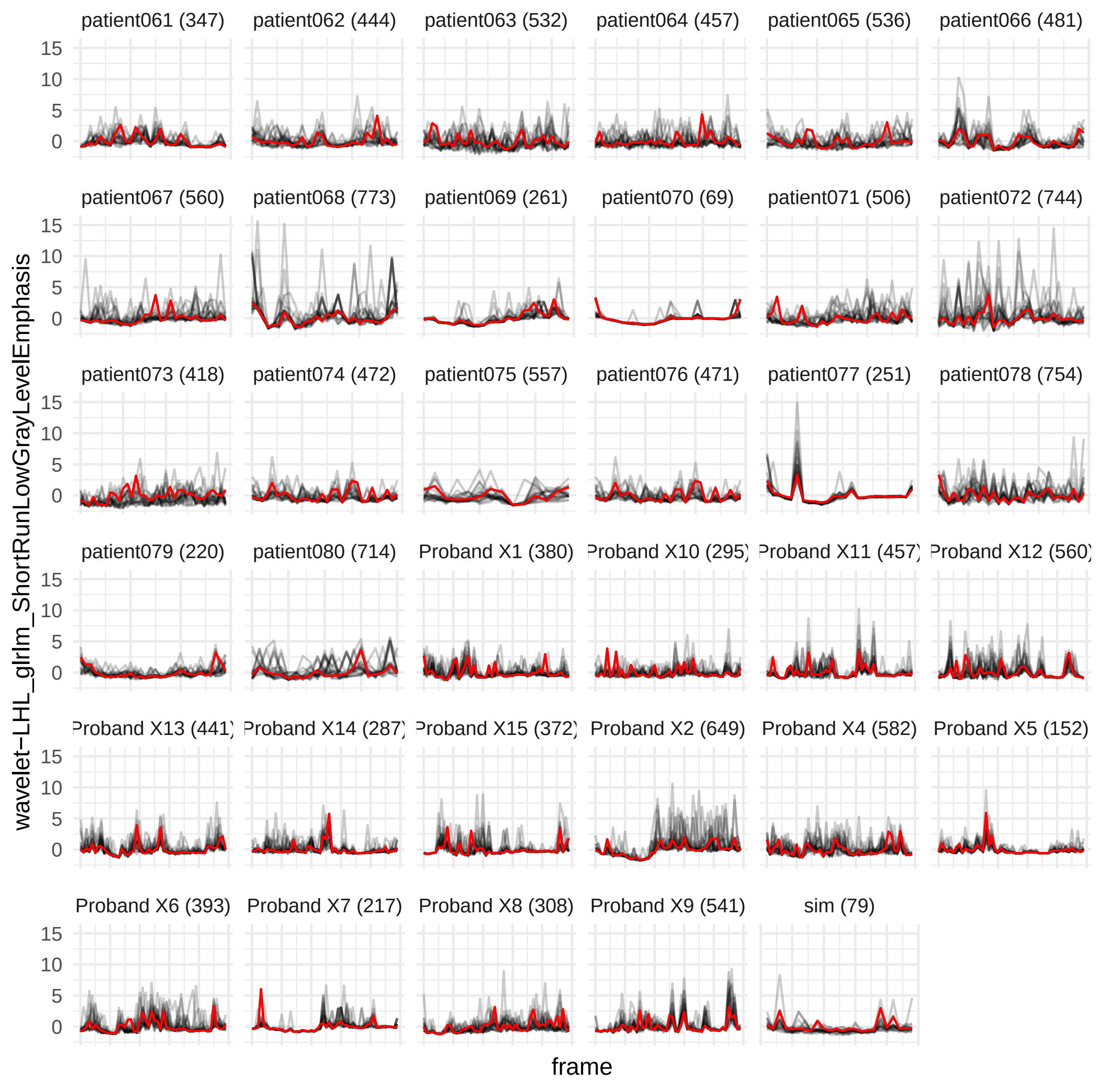

patient061 (347)
patient062 (444)
patient063 (532)
patient064 (457)
patient065 (536)
patient066 (481)
patient067 (560)
patient068 (773)
patient069 (261)
patient070 (69)
patient071 (506)
patient072 (744)
patient073 (418)
patient074 (472)
patient075 (557)
patient076 (471)
patient077 (251)
patient078 (754)
patient079 (220)
patient080 (714)
Proband X1 (380)
Proband X10 (295)
Proband X11 (457)
Proband X12 (560)
Proband X13 (441)
Proband X14 (287)
Proband X15 (372)
Proband X2 (649)
Proband X4 (582)
Proband X5 (152)
Proband X6 (393)
Proband X7 (217)
Proband X8 (308)
Proband X9 (541)
sim (79)
wavelet−LHL_glrlm_ShortRunLowGrayLevelEmphasis
frame
0
5
10
15

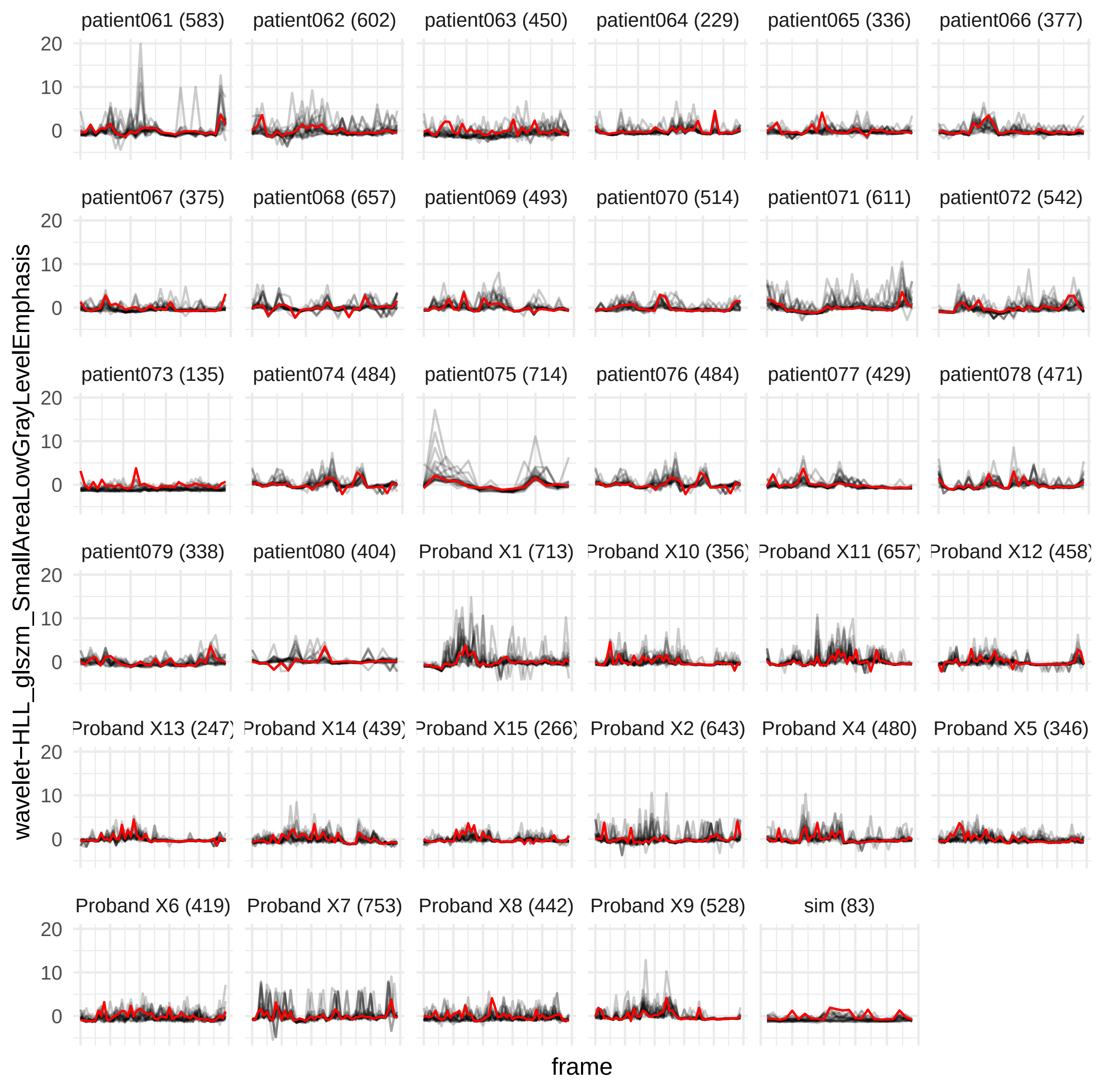
patient061 (583)
patient062 (602)
patient063 (450)
patient064 (229)
patient065 (336)
patient066 (377)
patient067 (375)
patient068 (657)
patient069 (493)
patient070 (514)
patient071 (611)
patient072 (542)
patient073 (135)
patient074 (484)
patient075 (714)
patient076 (484)
patient077 (429)
patient078 (471)
patient079 (338)
patient080 (404)
Proband X1 (713)
Proband X10 (356)
Proband X11 (657)
Proband X12 (458)
Proband X13 (247)
Proband X14 (439)
Proband X15 (266)
Proband X2 (643)
Proband X4 (480)
Proband X5 (346)
Proband X6 (419)
Proband X7 (753)
Proband X8 (442)
Proband X9 (528)
sim (83)
wavelet−HLL_glszm_SmallAreaLowGrayLevelEmphasis
20
10
0
frame

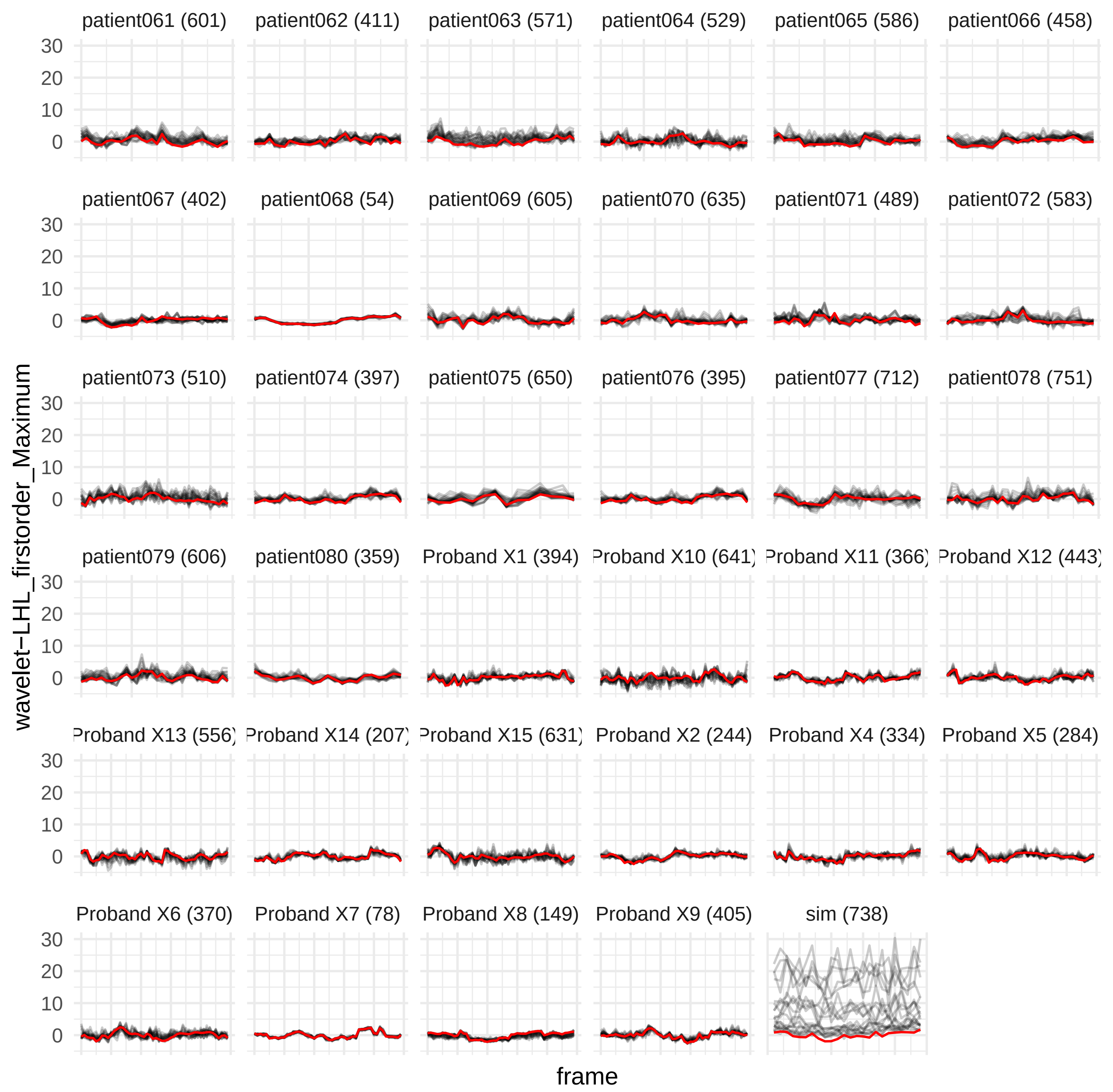

patient061 (601)
patient062 (411)
patient063 (571)
patient064 (529)
patient065 (586)
patient066 (458)
patient067 (402)
patient068 (54)
patient069 (605)
patient070 (635)
patient071 (489)
patient072 (583)
patient073 (510)
patient074 (397)
patient075 (650)
patient076 (395)
patient077 (712)
patient078 (751)
patient079 (606)
patient080 (359)
Proband X1 (394)
Proband X10 (641)
Proband X11 (366)
Proband X12 (443)
Proband X13 (556)
Proband X14 (207)
Proband X15 (631)
Proband X2 (244)
Proband X4 (334)
Proband X5 (284)
Proband X6 (370)
Proband X7 (78)
Proband X8 (149)
Proband X9 (405)
sim (738)
30
20
10
0
wavelet−LHL_firstorder_Maximum
frame

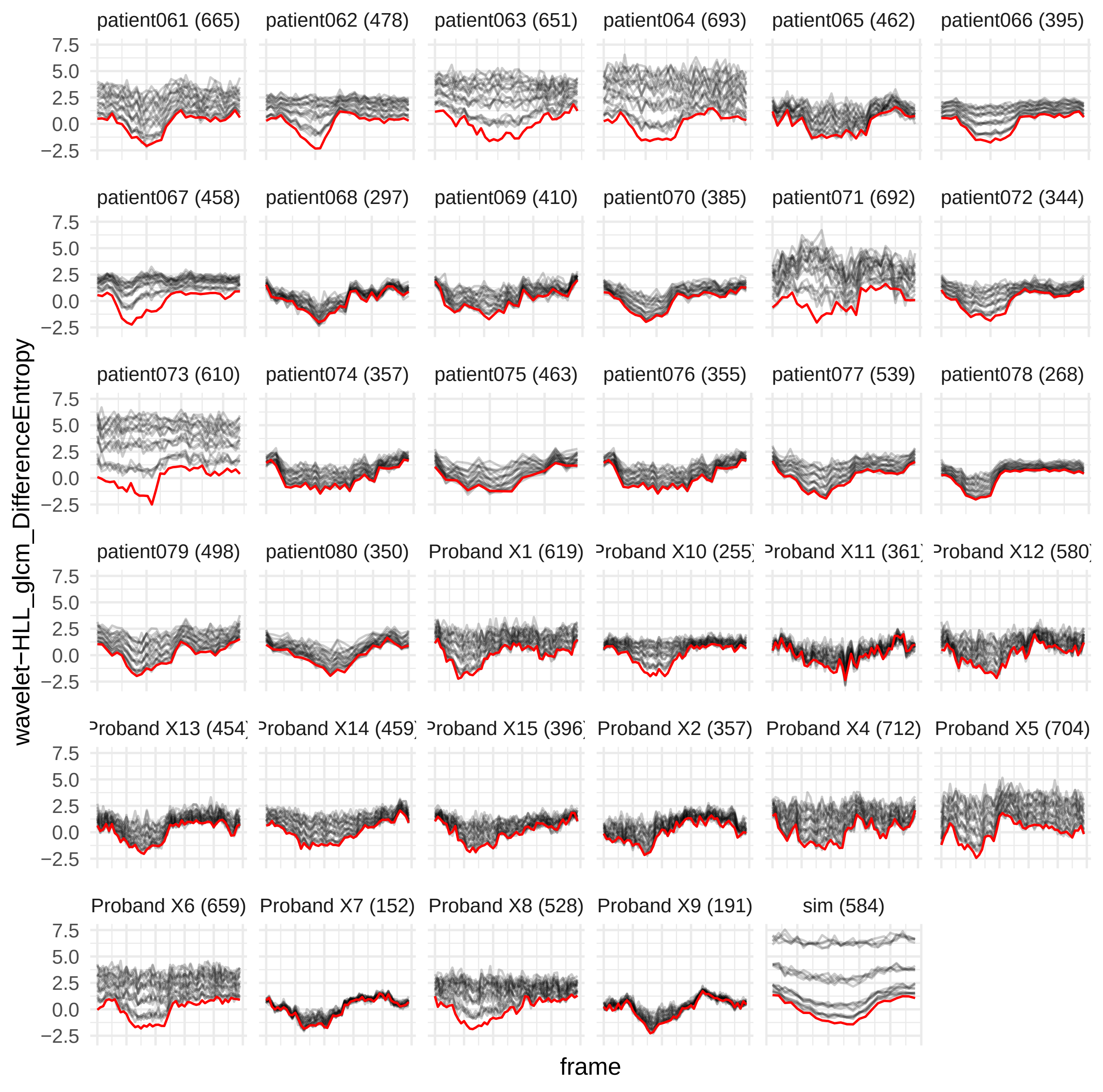

patient061 (665)
patient062 (478)
patient063 (651)
patient064 (693)
patient065 (462)
patient066 (395)
patient067 (458)
patient068 (297)
patient069 (410)
patient070 (385)
patient071 (692)
patient072 (344)
patient073 (610)
patient074 (357)
patient075 (463)
patient076 (355)
patient077 (539)
patient078 (268)
patient079 (498)
patient080 (350)
Proband X1 (619)
Proband X10 (255)
Proband X11 (361)
Proband X12 (580)
Proband X13 (454)
Proband X14 (459)
Proband X15 (396)
Proband X2 (357)
Proband X4 (712)
Proband X5 (704)
Proband X6 (659)
Proband X7 (152)
Proband X8 (528)
Proband X9 (191)
sim (584)
7.5
5.0
2.5
0.0
−2.5
wavelet−HLL_glcm_DifferenceEntropy
frame

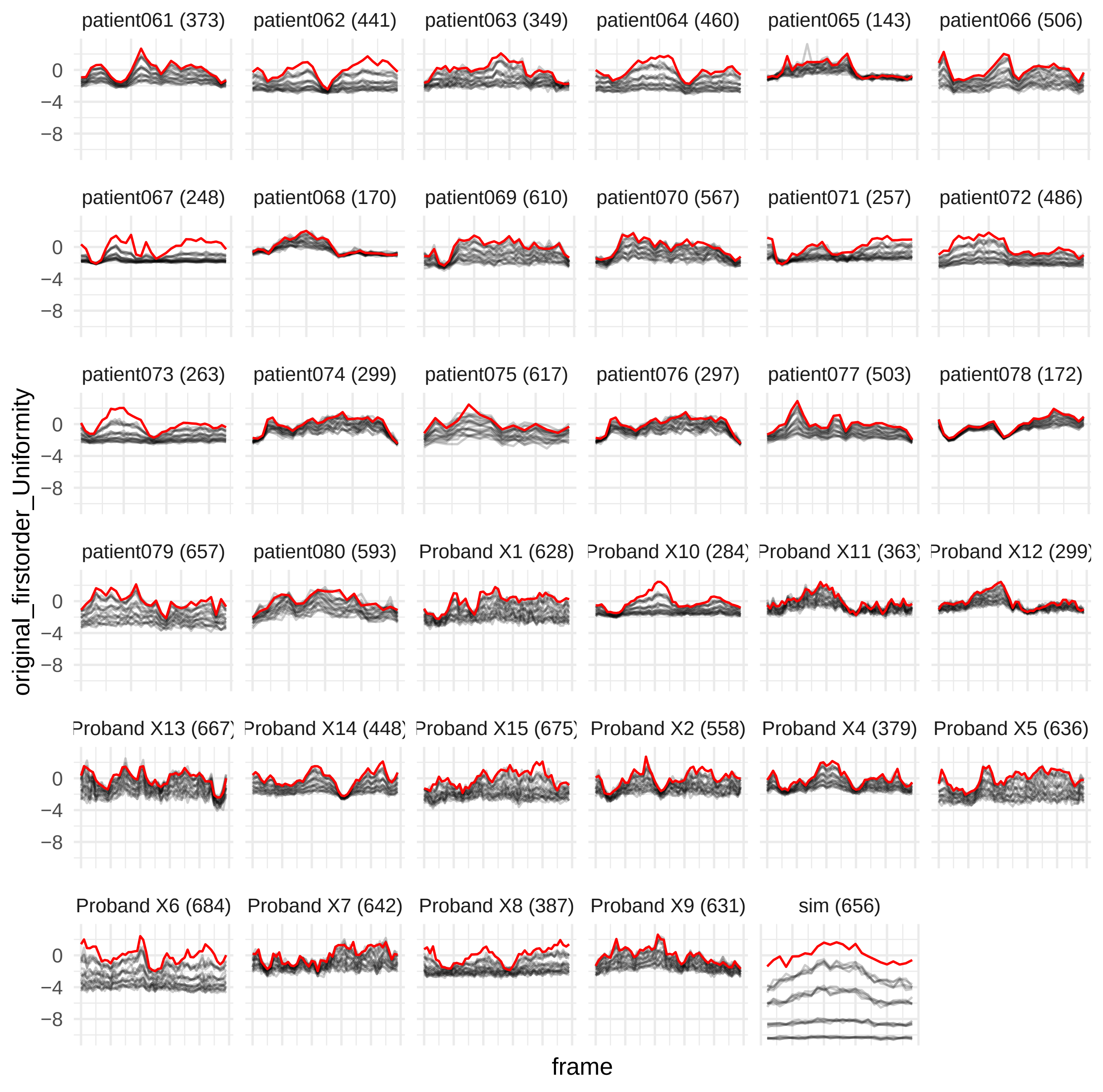
patient061 (373)
patient062 (441)
patient063 (349)
patient064 (460)
patient065 (143)
patient066 (506)
patient067 (248)
patient068 (170)
patient069 (610)
patient070 (567)
patient071 (257)
patient072 (486)
patient073 (263)
patient074 (299)
patient075 (617)
patient076 (297)
patient077 (503)
patient078 (172)
patient079 (657)
patient080 (593)
Proband X1 (628)
Proband X10 (284)
Proband X11 (363)
Proband X12 (299)
Proband X13 (667)
Proband X14 (448)
Proband X15 (675)
Proband X2 (558)
Proband X4 (379)
Proband X5 (636)
Proband X6 (684)
Proband X7 (642)
Proband X8 (387)
Proband X9 (631)
sim (656)
0
−4
−8
original_firstorder_Uniformity
frame

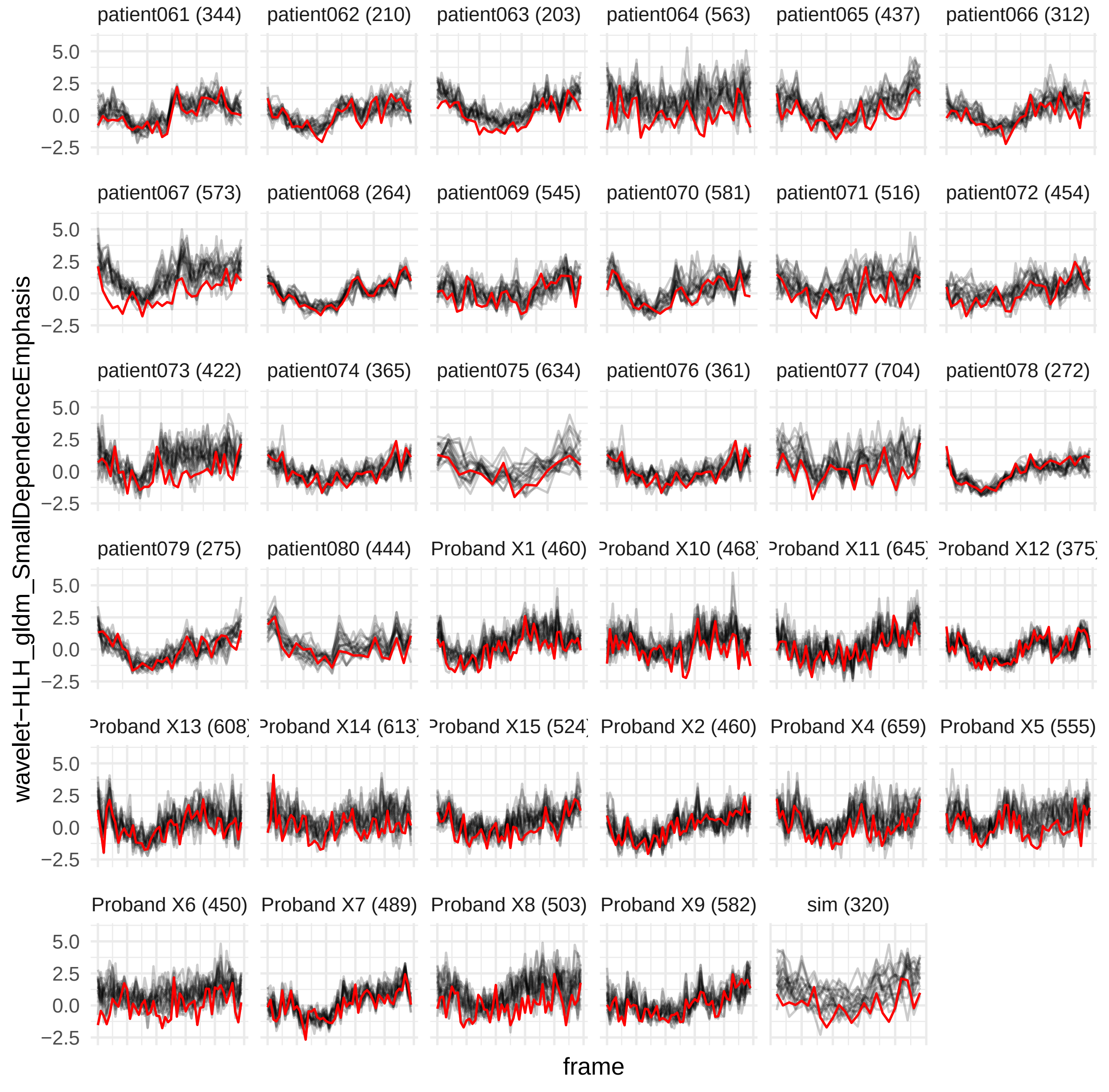

patient061 (344)
patient062 (210)
patient063 (203)
patient064 (563)
patient065 (437)
patient066 (312)
patient067 (573)
patient068 (264)
patient069 (545)
patient070 (581)
patient071 (516)
patient072 (454)
patient073 (422)
patient074 (365)
patient075 (634)
patient076 (361)
patient077 (704)
patient078 (272)
patient079 (275)
patient080 (444)
Proband X1 (460)
Proband X10 (468)
Proband X11 (645)
Proband X12 (375)
Proband X13 (608)
Proband X14 (613)
Proband X15 (524)
Proband X2 (460)
Proband X4 (659)
Proband X5 (555)
Proband X6 (450)
Proband X7 (489)
Proband X8 (503)
Proband X9 (582)
sim (320)
5.0
2.5
0.0
−2.5
wavelet−HLH_gldm_SmallDependenceEmphasis
frame

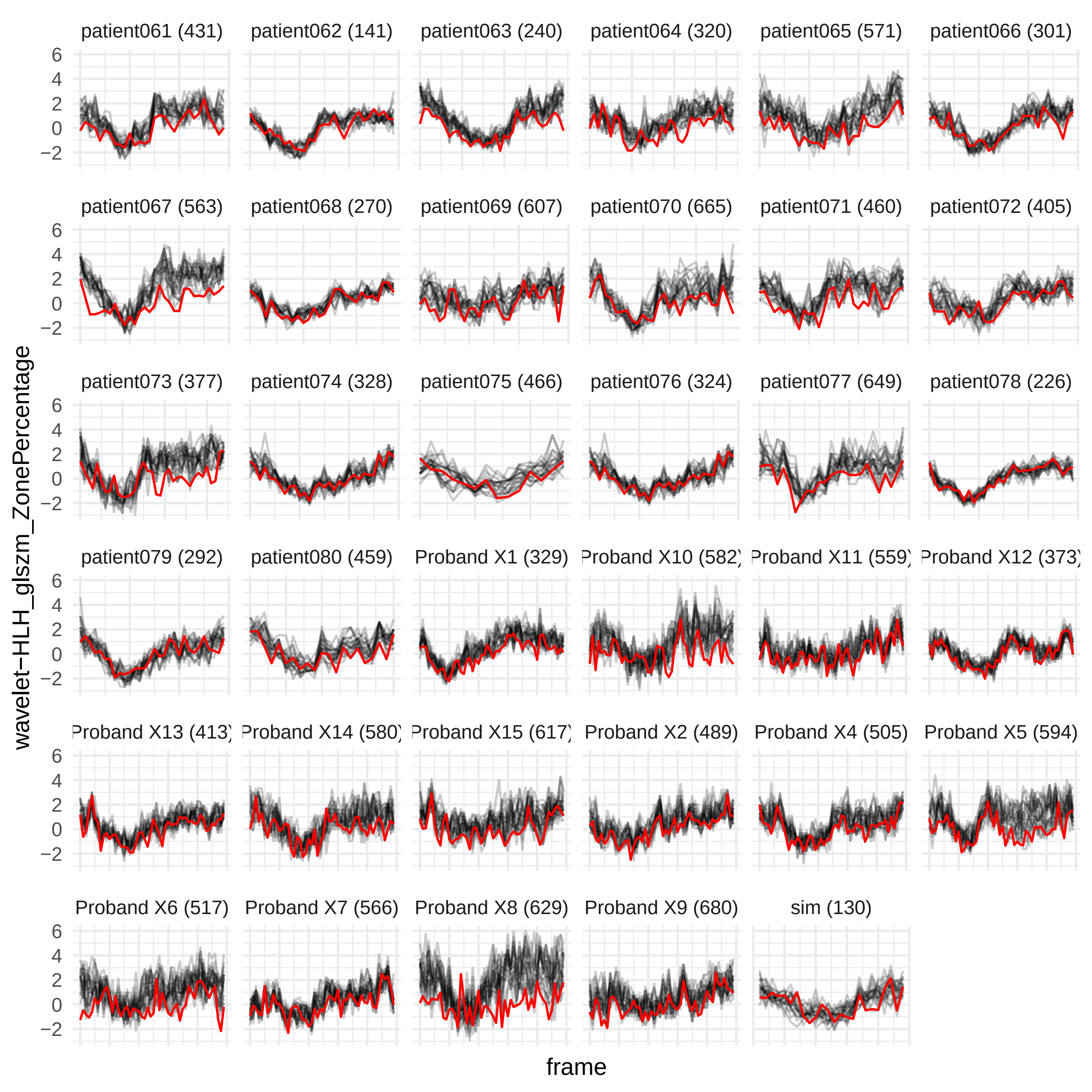

patient061 (431)
patient062 (141)
patient063 (240)
patient064 (320)
patient065 (571)
patient066 (301)
patient067 (563)
patient068 (270)
patient069 (607)
patient070 (665)
patient071 (460)
patient072 (405)
patient073 (377)
patient074 (328)
patient075 (466)
patient076 (324)
patient077 (649)
patient078 (226)
patient079 (292)
patient080 (459)
Proband X1 (329)
Proband X10 (582)
Proband X11 (559)
Proband X12 (373)
Proband X13 (413)
Proband X14 (580)
Proband X15 (617)
Proband X2 (489)
Proband X4 (505)
Proband X5 (594)
Proband X6 (517)
Proband X7 (566)
Proband X8 (629)
Proband X9 (680)
sim (130)
6
4
2
0
−2
wavelet−HLH_glszm_ZonePercentage
frame

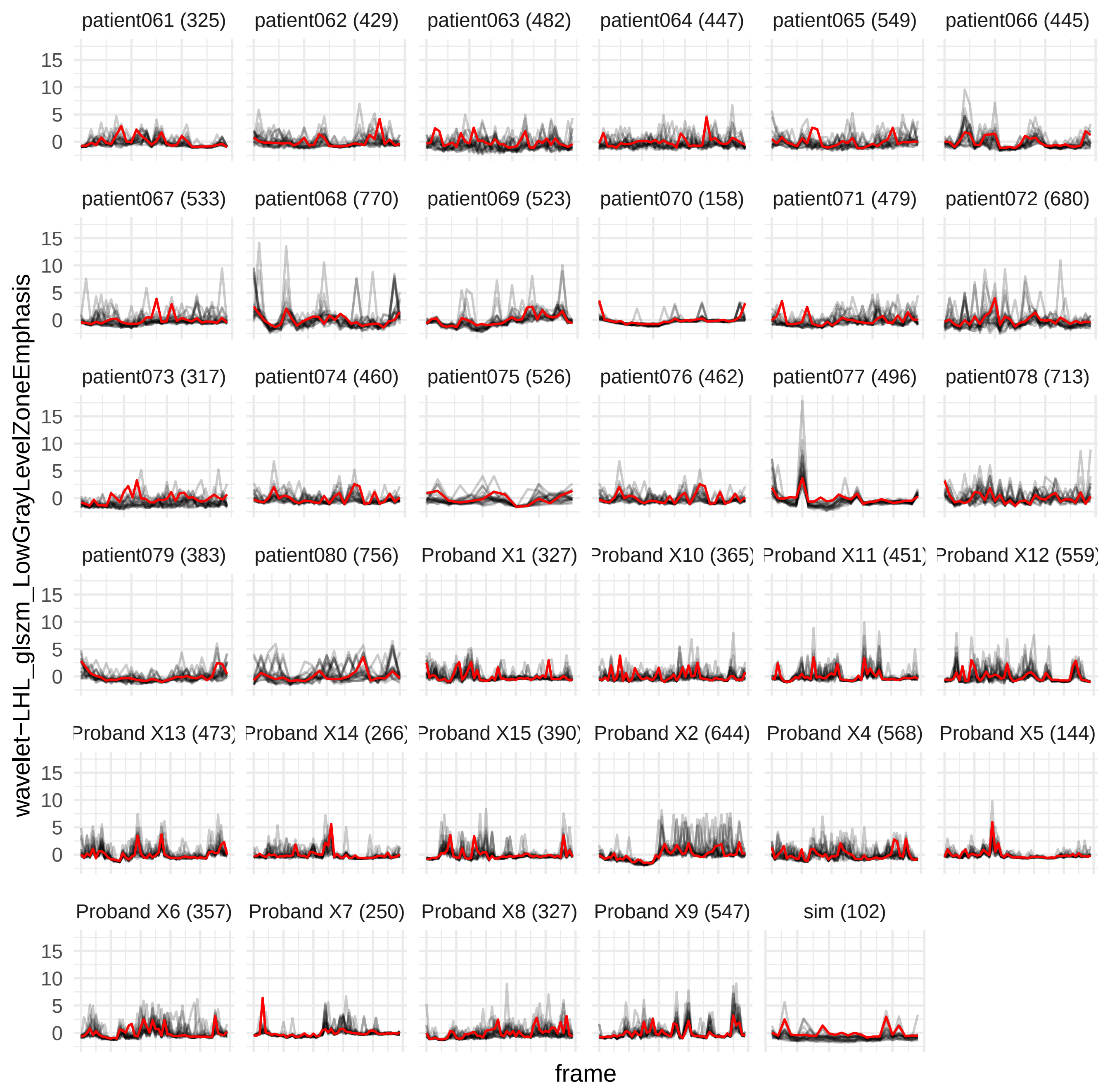
patient061 (325)
patient062 (429)
patient063 (482)
patient064 (447)
patient065 (549)
patient066 (445)
patient067 (533)
patient068 (770)
patient069 (523)
patient070 (158)
patient071 (479)
patient072 (680)
patient073 (317)
patient074 (460)
patient075 (526)
patient076 (462)
patient077 (496)
patient078 (713)
patient079 (383)
patient080 (756)
Proband X1 (327)
Proband X10 (365)
Proband X11 (451)
Proband X12 (559)
Proband X13 (473)
Proband X14 (266)
Proband X15 (390)
Proband X2 (644)
Proband X4 (568)
Proband X5 (144)
Proband X6 (357)
Proband X7 (250)
Proband X8 (327)
Proband X9 (547)
sim (102)
wavelet–LHL_glszm_LowGrayLevelZoneEmphasis
frame
15
10
5
0

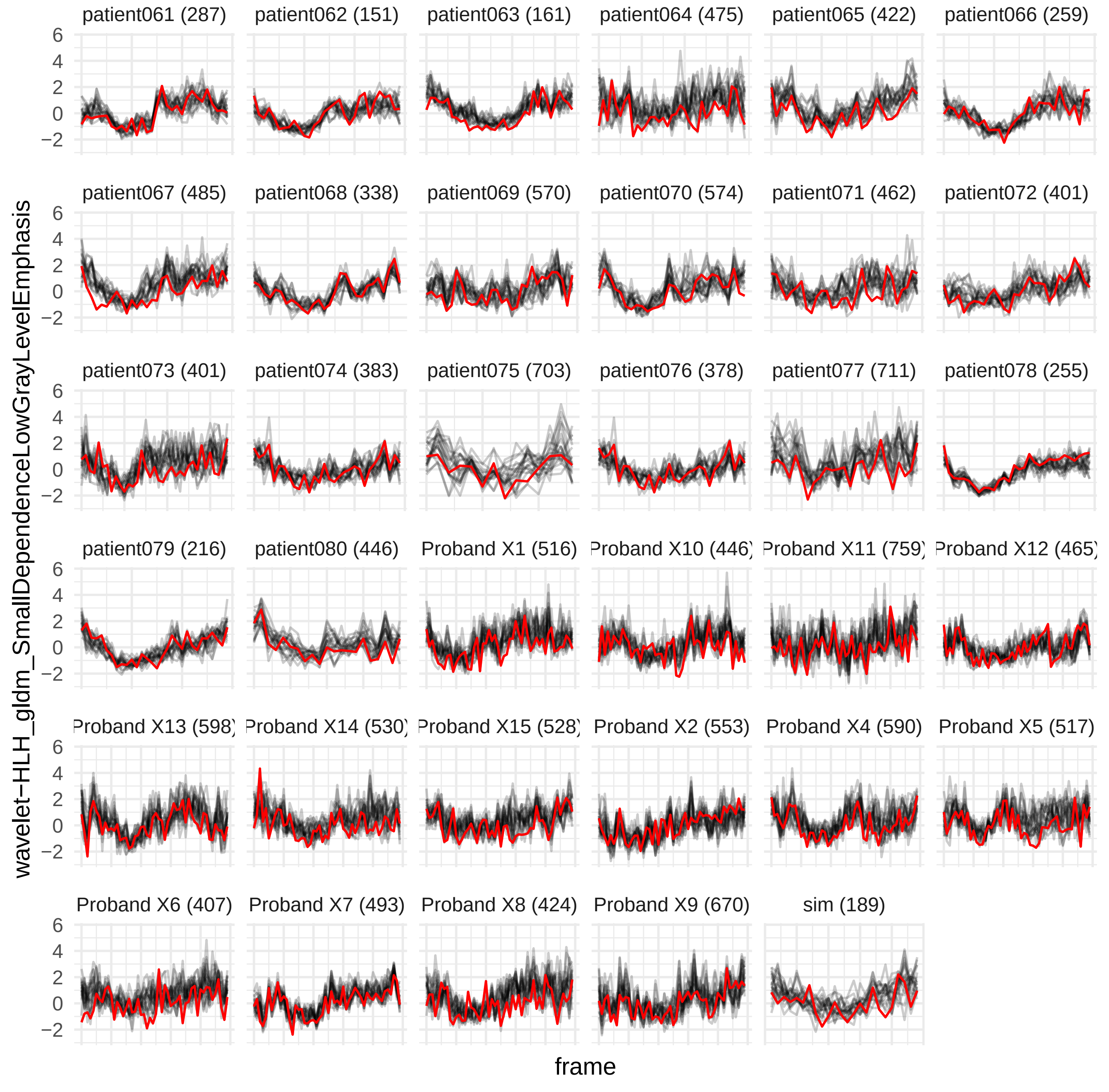

patient061 (287)
patient062 (151)
patient063 (161)
patient064 (475)
patient065 (422)
patient066 (259)
patient067 (485)
patient068 (338)
patient069 (570)
patient070 (574)
patient071 (462)
patient072 (401)
patient073 (401)
patient074 (383)
patient075 (703)
patient076 (378)
patient077 (711)
patient078 (255)
patient079 (216)
patient080 (446)
Proband X1 (516)
Proband X10 (446)
Proband X11 (759)
Proband X12 (465)
Proband X13 (598)
Proband X14 (530)
Proband X15 (528)
Proband X2 (553)
Proband X4 (590)
Proband X5 (517)
Proband X6 (407)
Proband X7 (493)
Proband X8 (424)
Proband X9 (670)
sim (189)
wavelet−HLH_gldm_SmallDependenceLowGrayLevelEmphasis
frame
6
4
2
0
−2

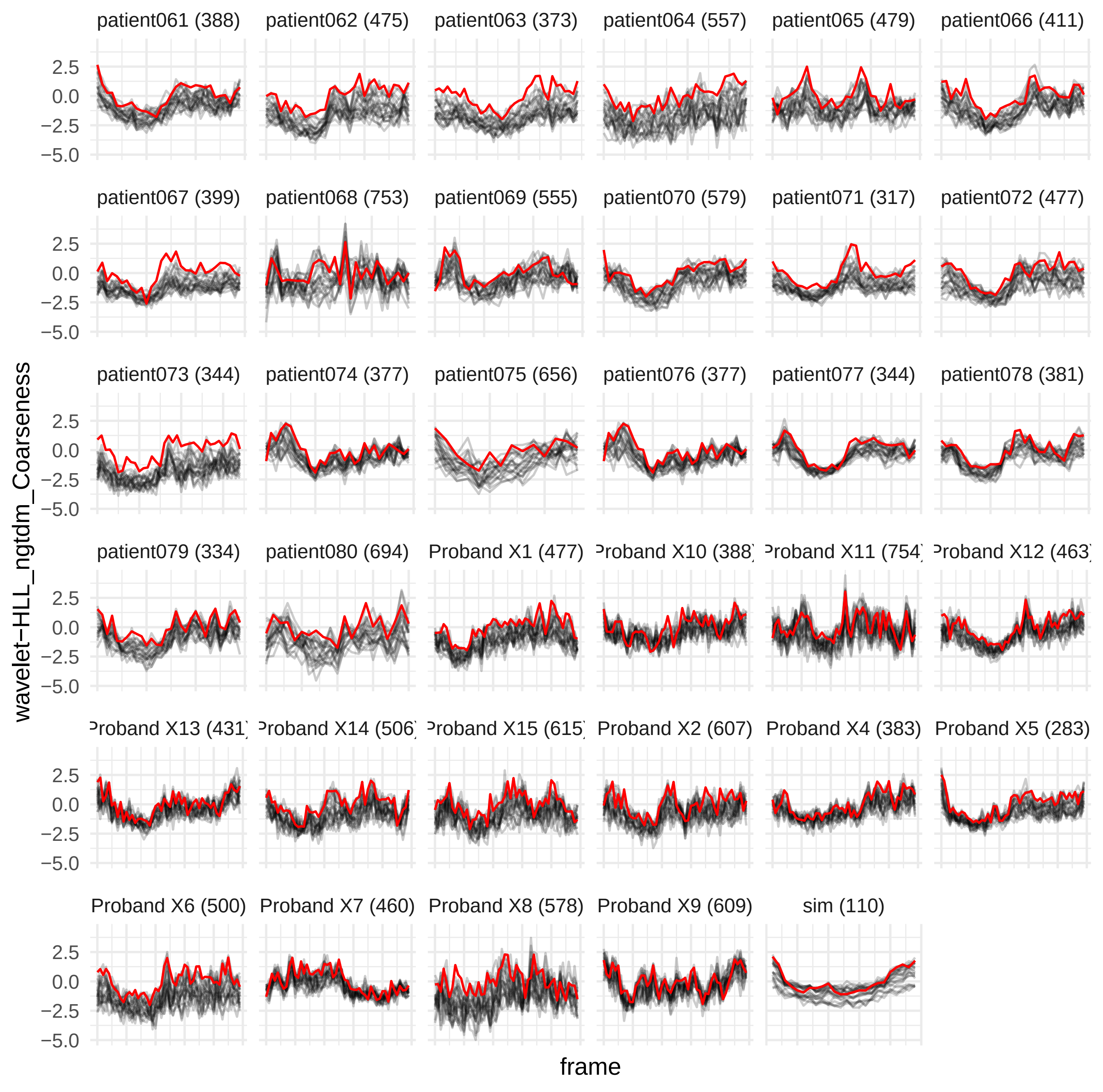

patient061 (388)
patient062 (475)
patient063 (373)
patient064 (557)
patient065 (479)
patient066 (411)
patient067 (399)
patient068 (753)
patient069 (555)
patient070 (579)
patient071 (317)
patient072 (477)
patient073 (344)
patient074 (377)
patient075 (656)
patient076 (377)
patient077 (344)
patient078 (381)
patient079 (334)
patient080 (694)
Proband X1 (477)
Proband X10 (388)
Proband X11 (754)
Proband X12 (463)
Proband X13 (431)
Proband X14 (506)
Proband X15 (615)
Proband X2 (607)
Proband X4 (383)
Proband X5 (283)
Proband X6 (500)
Proband X7 (460)
Proband X8 (578)
Proband X9 (609)
sim (110)
2.5
0.0
−2.5
−5.0
wavelet−HLL_ngtdm_Coarseness
frame

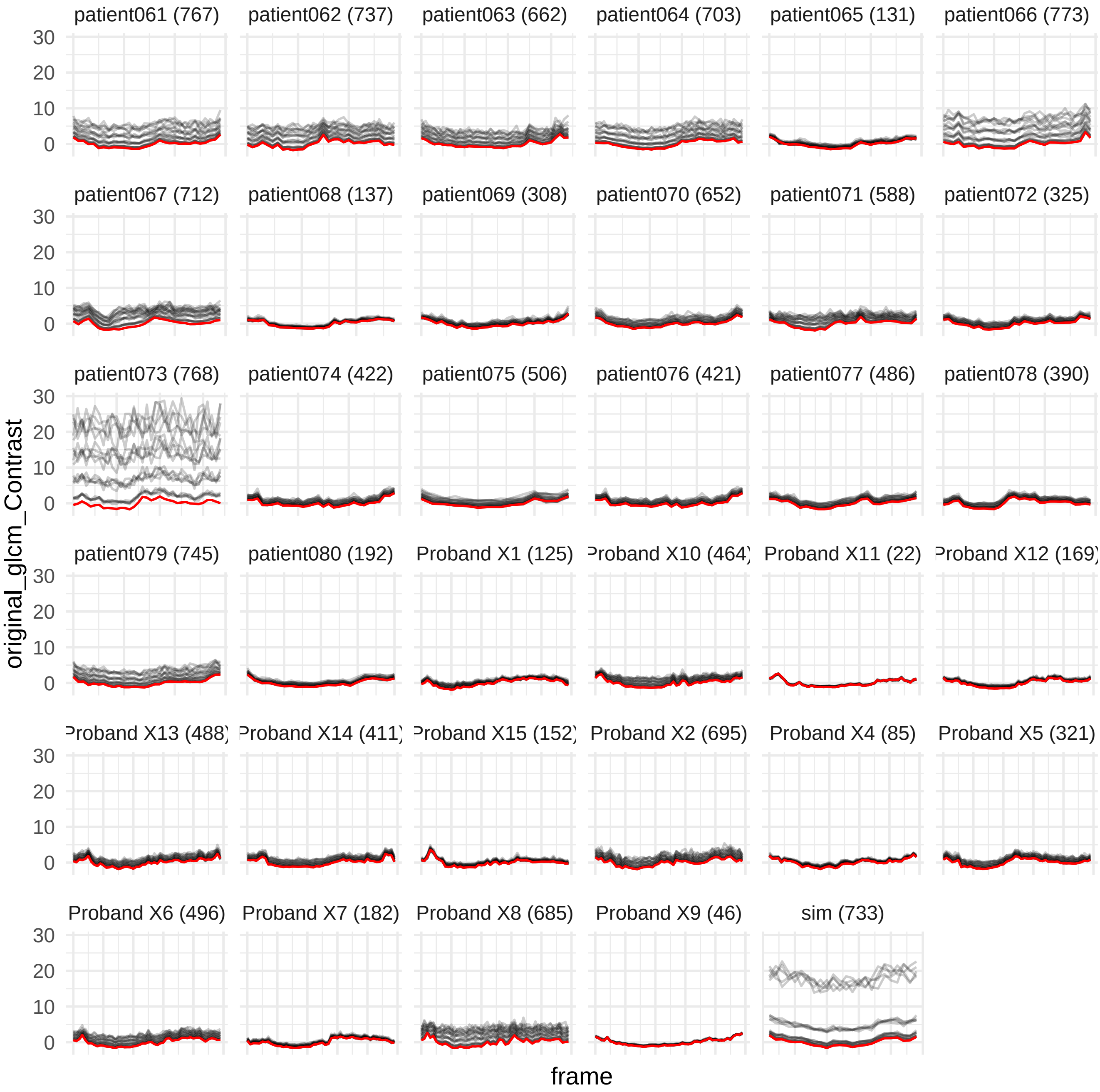
patient061 (767)
patient062 (737)
patient063 (662)
patient064 (703)
patient065 (131)
patient066 (773)
patient067 (712)
patient068 (137)
patient069 (308)
patient070 (652)
patient071 (588)
patient072 (325)
patient073 (768)
patient074 (422)
patient075 (506)
patient076 (421)
patient077 (486)
patient078 (390)
patient079 (745)
patient080 (192)
Proband X1 (125)
Proband X10 (464)
Proband X11 (22)
Proband X12 (169)
Proband X13 (488)
Proband X14 (411)
Proband X15 (152)
Proband X2 (695)
Proband X4 (85)
Proband X5 (321)
Proband X6 (496)
Proband X7 (182)
Proband X8 (685)
Proband X9 (46)
sim (733)
original_glcm_Contrast
frame
0
10
20
30

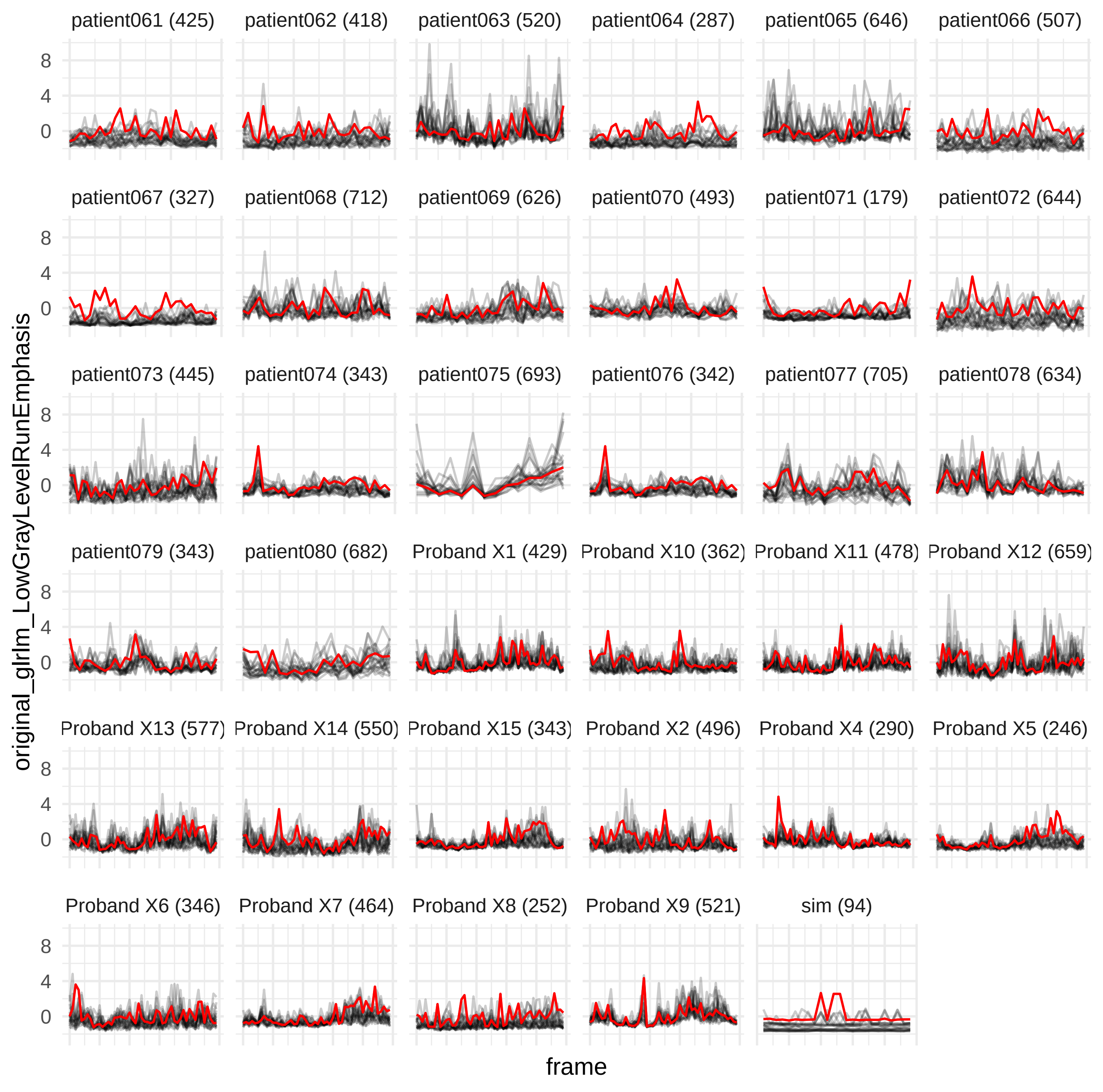

patient061 (425)
patient062 (418)
patient063 (520)
patient064 (287)
patient065 (646)
patient066 (507)
patient067 (327)
patient068 (712)
patient069 (626)
patient070 (493)
patient071 (179)
patient072 (644)
patient073 (445)
patient074 (343)
patient075 (693)
patient076 (342)
patient077 (705)
patient078 (634)
patient079 (343)
patient080 (682)
Proband X1 (429)
Proband X10 (362)
Proband X11 (478)
Proband X12 (659)
Proband X13 (577)
Proband X14 (550)
Proband X15 (343)
Proband X2 (496)
Proband X4 (290)
Proband X5 (246)
Proband X6 (346)
Proband X7 (464)
Proband X8 (252)
Proband X9 (521)
sim (94)
8
4
0
original_glrlm_LowGrayLevelRunEmphasis
frame

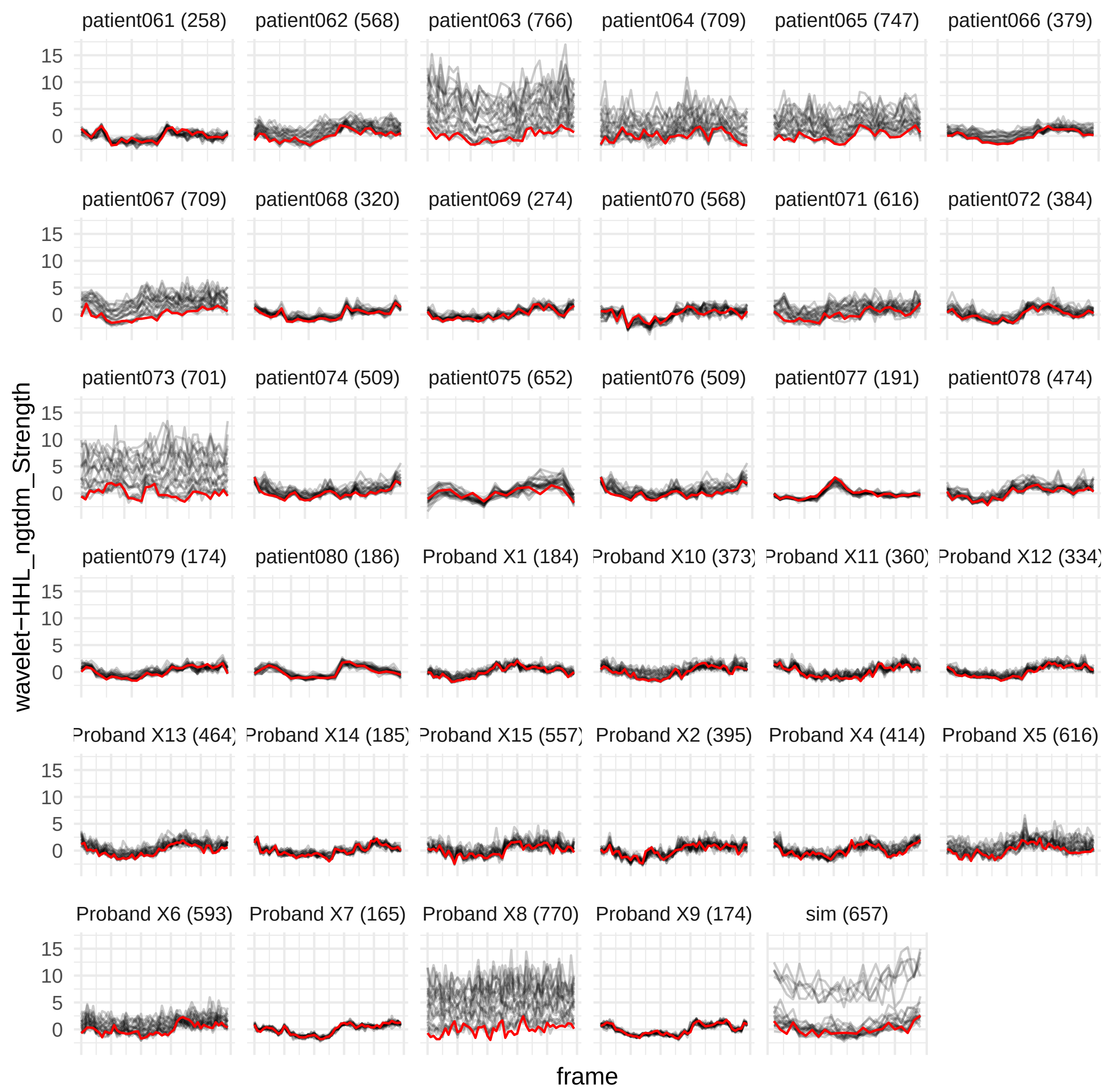
patient061 (258)
patient062 (568)
patient063 (766)
patient064 (709)
patient065 (747)
patient066 (379)
patient067 (709)
patient068 (320)
patient069 (274)
patient070 (568)
patient071 (616)
patient072 (384)
patient073 (701)
patient074 (509)
patient075 (652)
patient076 (509)
patient077 (191)
patient078 (474)
patient079 (174)
patient080 (186)
Proband X1 (184)
Proband X10 (373)
Proband X11 (360)
Proband X12 (334)
Proband X13 (464)
Proband X14 (185)
Proband X15 (557)
Proband X2 (395)
Proband X4 (414)
Proband X5 (616)
Proband X6 (593)
Proband X7 (165)
Proband X8 (770)
Proband X9 (174)
sim (657)
15
10
5
0
wavelet−HHL_ngtdm_Strength
frame

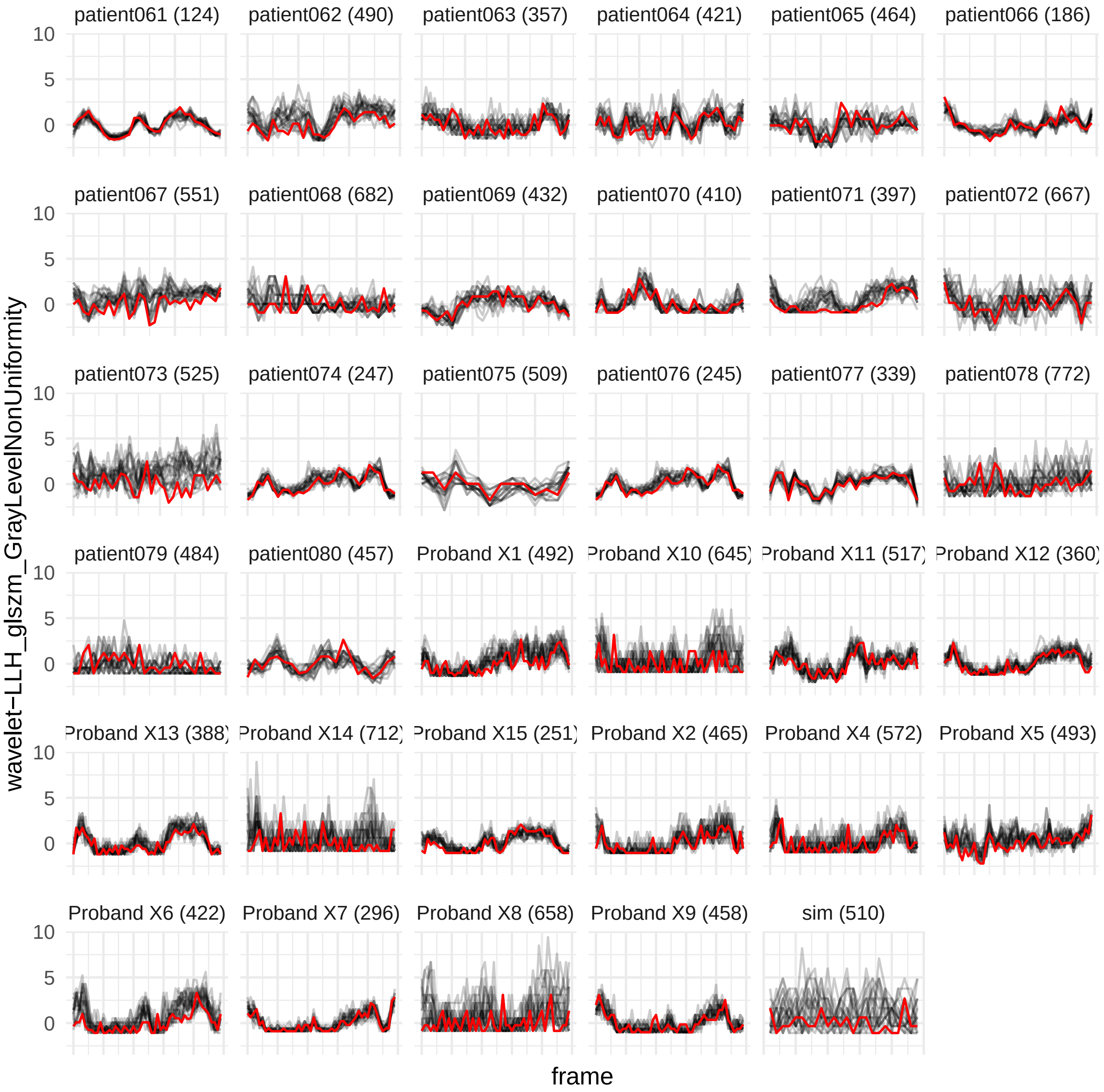

patient061 (124)
patient062 (490)
patient063 (357)
patient064 (421)
patient065 (464)
patient066 (186)
patient067 (551)
patient068 (682)
patient069 (432)
patient070 (410)
patient071 (397)
patient072 (667)
patient073 (525)
patient074 (247)
patient075 (509)
patient076 (245)
patient077 (339)
patient078 (772)
patient079 (484)
patient080 (457)
Proband X1 (492)
Proband X10 (645)
Proband X11 (517)
Proband X12 (360)
Proband X13 (388)
Proband X14 (712)
Proband X15 (251)
Proband X2 (465)
Proband X4 (572)
Proband X5 (493)
Proband X6 (422)
Proband X7 (296)
Proband X8 (658)
Proband X9 (458)
sim (510)
10
5
0
wavelet−LLH_glszm_GrayLevelNonUniformity
frame

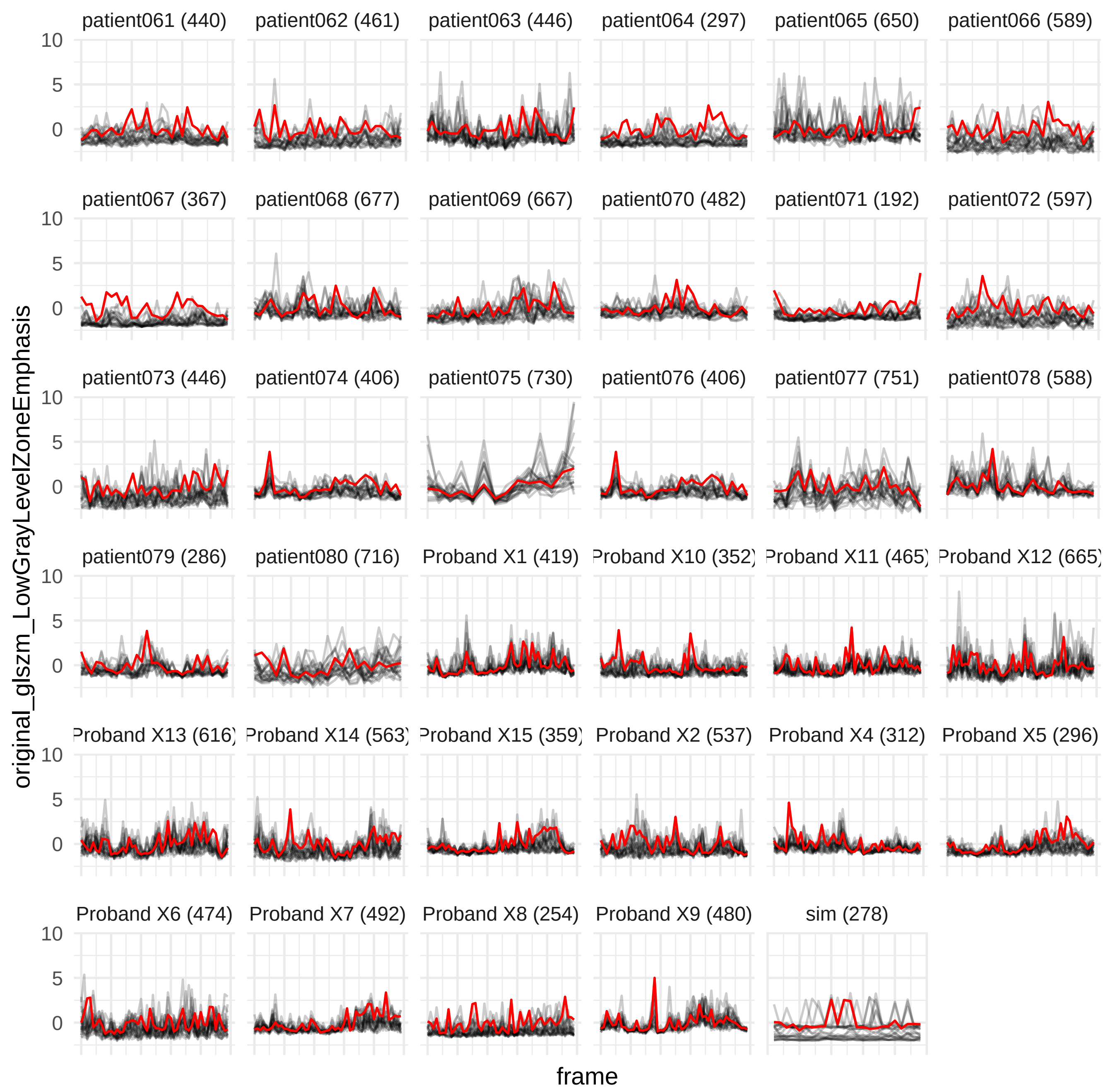

patient061 (440)
patient062 (461)
patient063 (446)
patient064 (297)
patient065 (650)
patient066 (589)
patient067 (367)
patient068 (677)
patient069 (667)
patient070 (482)
patient071 (192)
patient072 (597)
patient073 (446)
patient074 (406)
patient075 (730)
patient076 (406)
patient077 (751)
patient078 (588)
patient079 (286)
patient080 (716)
Proband X1 (419)
Proband X10 (352)
Proband X11 (465)
Proband X12 (665)
Proband X13 (616)
Proband X14 (563)
Proband X15 (359)
Proband X2 (537)
Proband X4 (312)
Proband X5 (296)
Proband X6 (474)
Proband X7 (492)
Proband X8 (254)
Proband X9 (480)
sim (278)
10
5
0
original_glszm_LowGrayLevelZoneEmphasis
frame

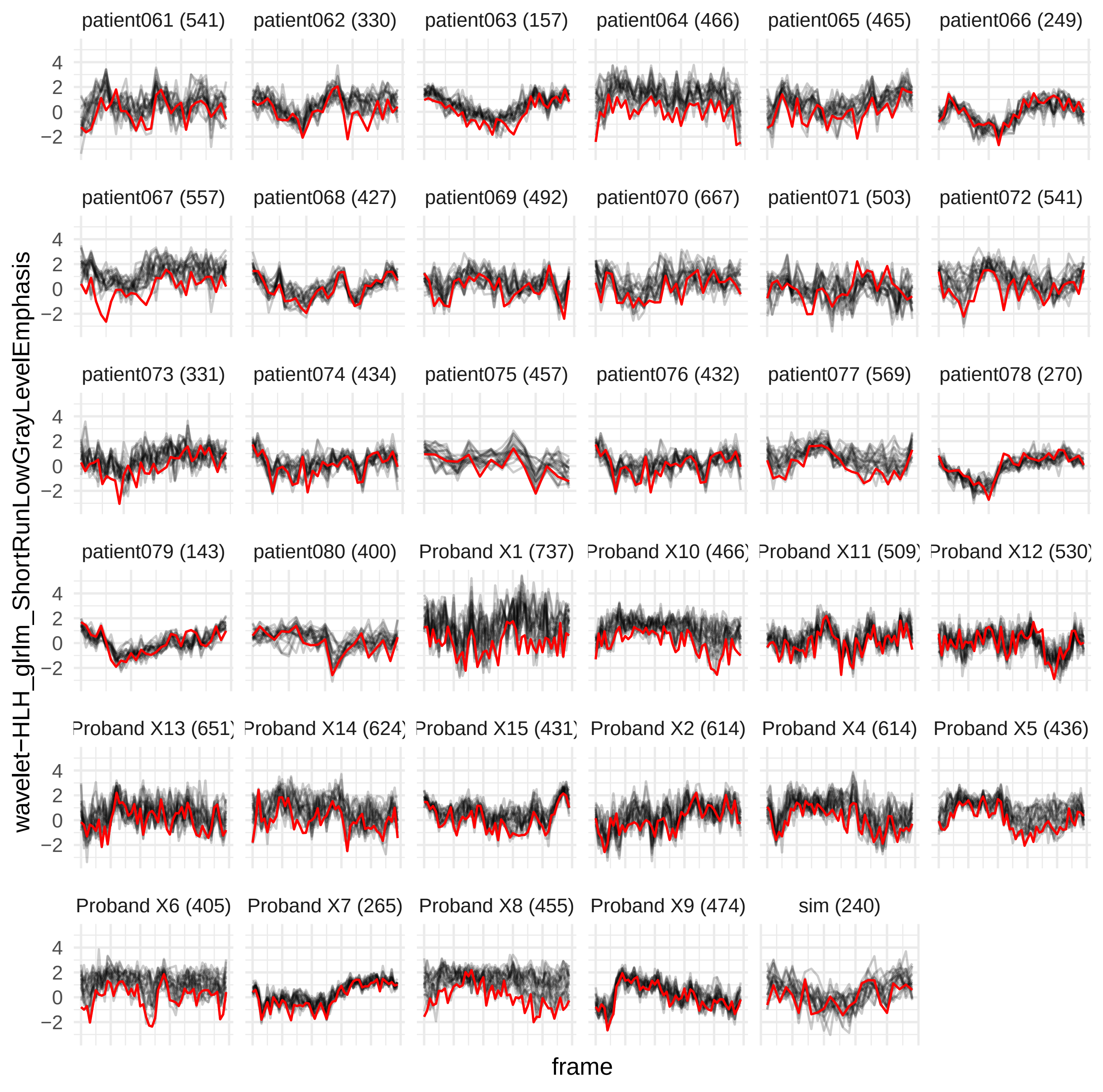
patient061 (541)
patient062 (330)
patient063 (157)
patient064 (466)
patient065 (465)
patient066 (249)
patient067 (557)
patient068 (427)
patient069 (492)
patient070 (667)
patient071 (503)
patient072 (541)
patient073 (331)
patient074 (434)
patient075 (457)
patient076 (432)
patient077 (569)
patient078 (270)
patient079 (143)
patient080 (400)
Proband X1 (737)
Proband X10 (466)
Proband X11 (509)
Proband X12 (530)
Proband X13 (651)
Proband X14 (624)
Proband X15 (431)
Proband X2 (614)
Proband X4 (614)
Proband X5 (436)
Proband X6 (405)
Proband X7 (265)
Proband X8 (455)
Proband X9 (474)
sim (240)
4
2
0
−2
wavelet−HLH_glrlm_ShortRunLowGrayLevelEmphasis
frame

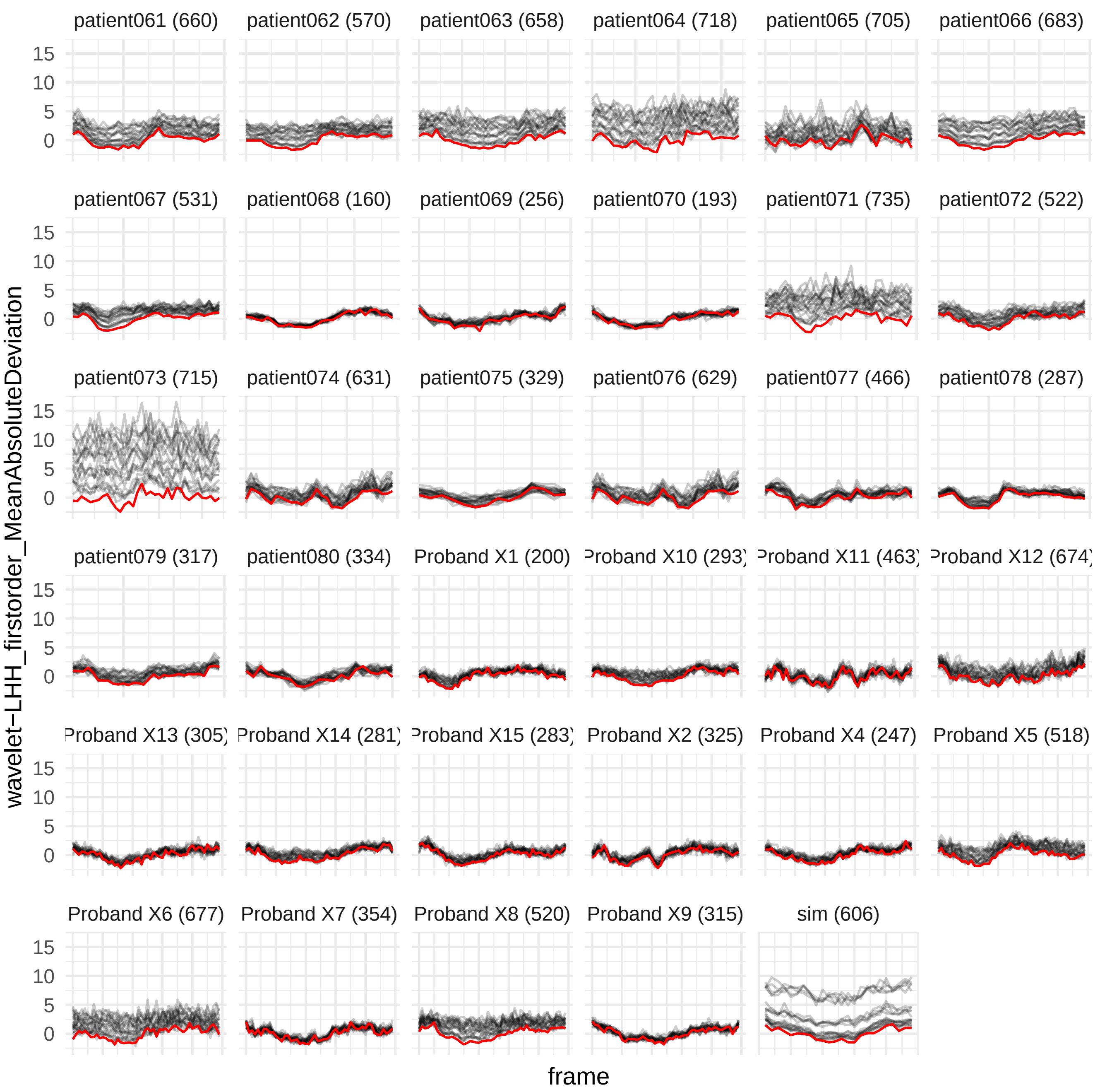
patient061 (660)
patient062 (570)
patient063 (658)
patient064 (718)
patient065 (705)
patient066 (683)
patient067 (531)
patient068 (160)
patient069 (256)
patient070 (193)
patient071 (735)
patient072 (522)
patient073 (715)
patient074 (631)
patient075 (329)
patient076 (629)
patient077 (466)
patient078 (287)
patient079 (317)
patient080 (334)
Proband X1 (200)
Proband X10 (293)
Proband X11 (463)
Proband X12 (674)
Proband X13 (305)
Proband X14 (281)
Proband X15 (283)
Proband X2 (325)
Proband X4 (247)
Proband X5 (518)
Proband X6 (677)
Proband X7 (354)
Proband X8 (520)
Proband X9 (315)
sim (606)
15
10
5
0
wavelet−LHH_firstorder_MeanAbsoluteDeviation
frame

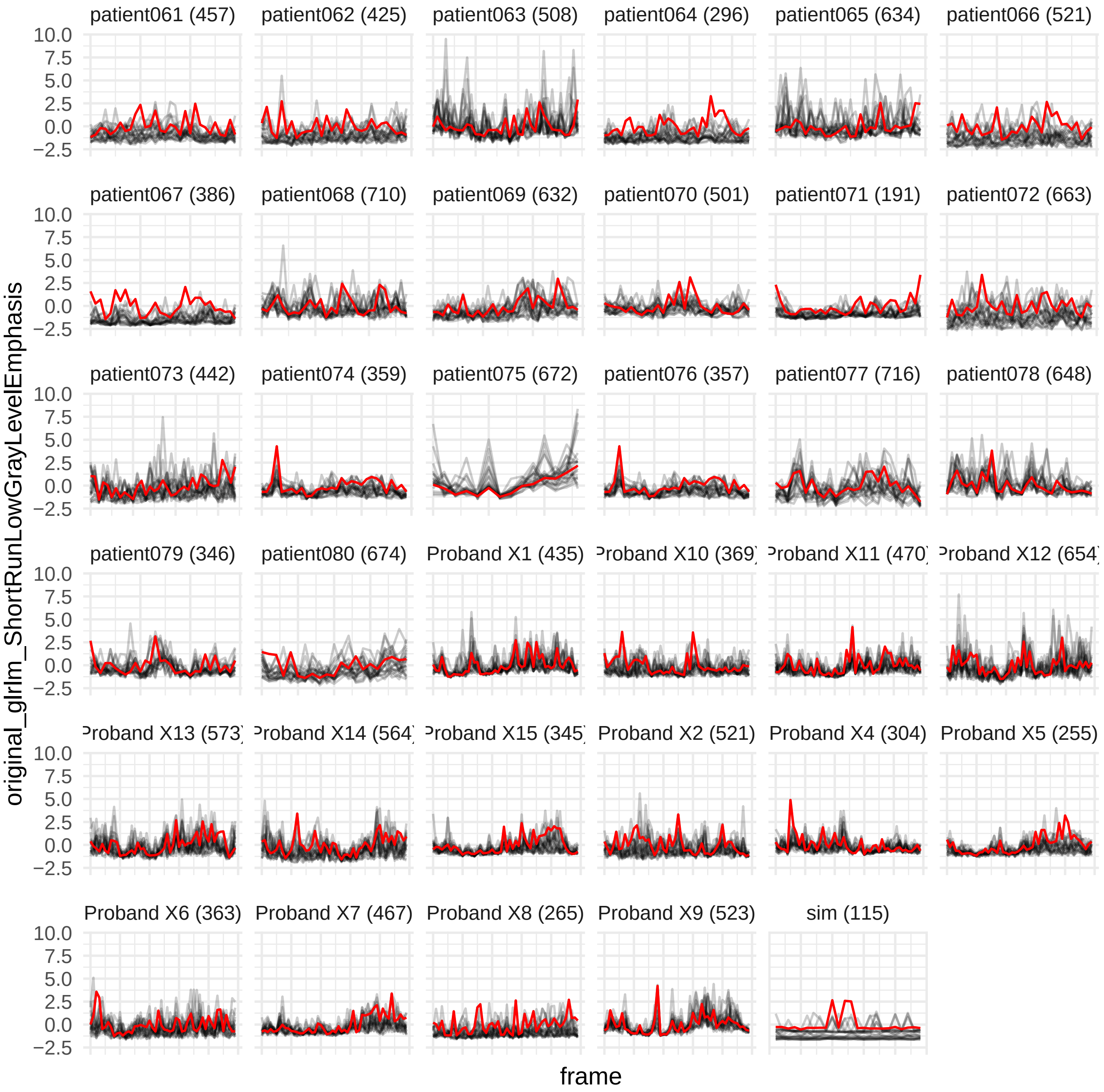
patient061 (457)
patient062 (425)
patient063 (508)
patient064 (296)
patient065 (634)
patient066 (521)
patient067 (386)
patient068 (710)
patient069 (632)
patient070 (501)
patient071 (191)
patient072 (663)
patient073 (442)
patient074 (359)
patient075 (672)
patient076 (357)
patient077 (716)
patient078 (648)
patient079 (346)
patient080 (674)
Proband X1 (435)
Proband X10 (369)
Proband X11 (470)
Proband X12 (654)
Proband X13 (573)
Proband X14 (564)
Proband X15 (345)
Proband X2 (521)
Proband X4 (304)
Proband X5 (255)
Proband X6 (363)
Proband X7 (467)
Proband X8 (265)
Proband X9 (523)
sim (115)
original_glrlm_ShortRunLowGrayLevelEmphasis
frame
10.0
7.5
5.0
2.5
0.0
−2.5

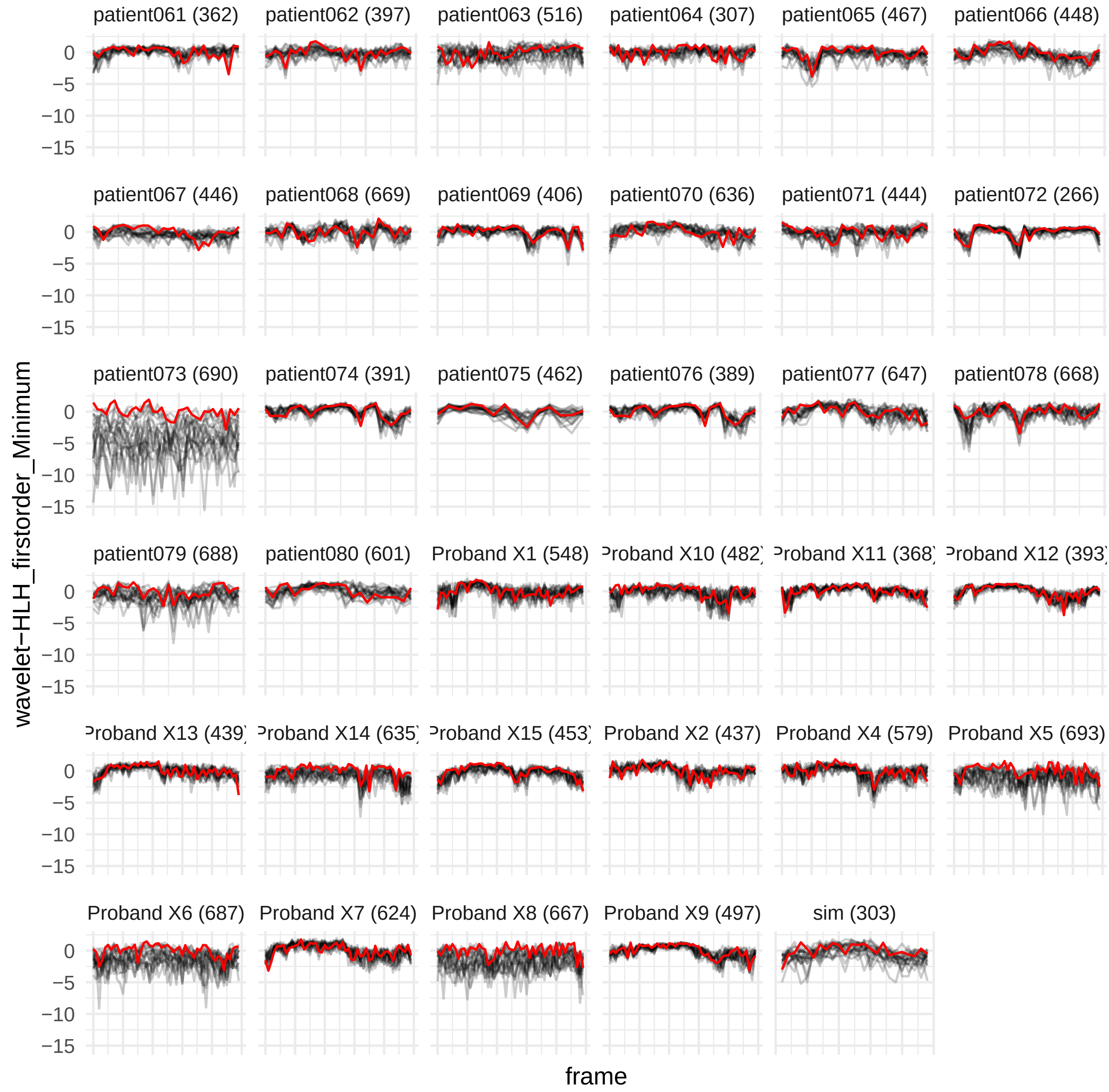

patient061 (362)
patient062 (397)
patient063 (516)
patient064 (307)
patient065 (467)
patient066 (448)
patient067 (446)
patient068 (669)
patient069 (406)
patient070 (636)
patient071 (444)
patient072 (266)
patient073 (690)
patient074 (391)
patient075 (462)
patient076 (389)
patient077 (647)
patient078 (668)
patient079 (688)
patient080 (601)
Proband X1 (548)
roband X10 (482
roband X11 (368
roband X12 (393
roband X13 (439
roband X14 (635
roband X15 (453
Proband X2 (437)
Proband X4 (579)
Proband X5 (693)
Proband X6 (687)
Proband X7 (624)
Proband X8 (667)
Proband X9 (497)
sim (303)
0
−5
−10
−15
wavelet−HLH_firstorder_Minimum
frame

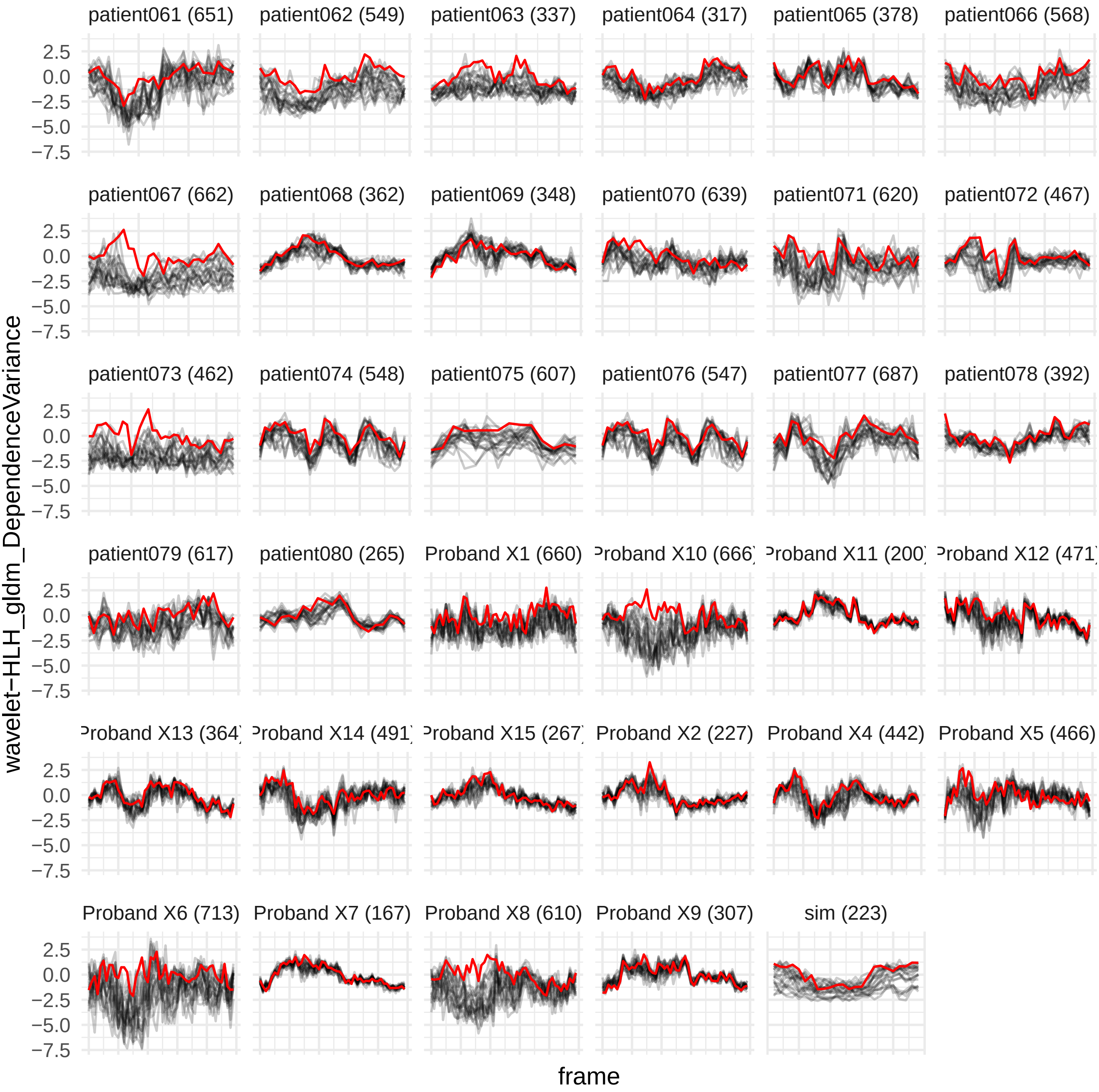

patient061 (651)
patient062 (549)
patient063 (337)
patient064 (317)
patient065 (378)
patient066 (568)
patient067 (662)
patient068 (362)
patient069 (348)
patient070 (639)
patient071 (620)
patient072 (467)
patient073 (462)
patient074 (548)
patient075 (607)
patient076 (547)
patient077 (687)
patient078 (392)
patient079 (617)
patient080 (265)
Proband X1 (660)
Proband X10 (666)
Proband X11 (200)
Proband X12 (471)
Proband X13 (364)
Proband X14 (491)
Proband X15 (267)
Proband X2 (227)
Proband X4 (442)
Proband X5 (466)
Proband X6 (713)
Proband X7 (167)
Proband X8 (610)
Proband X9 (307)
sim (223)
2.5
0.0
−2.5
−5.0
−7.5
wavelet−HLH_gldm_DependenceVariance
frame

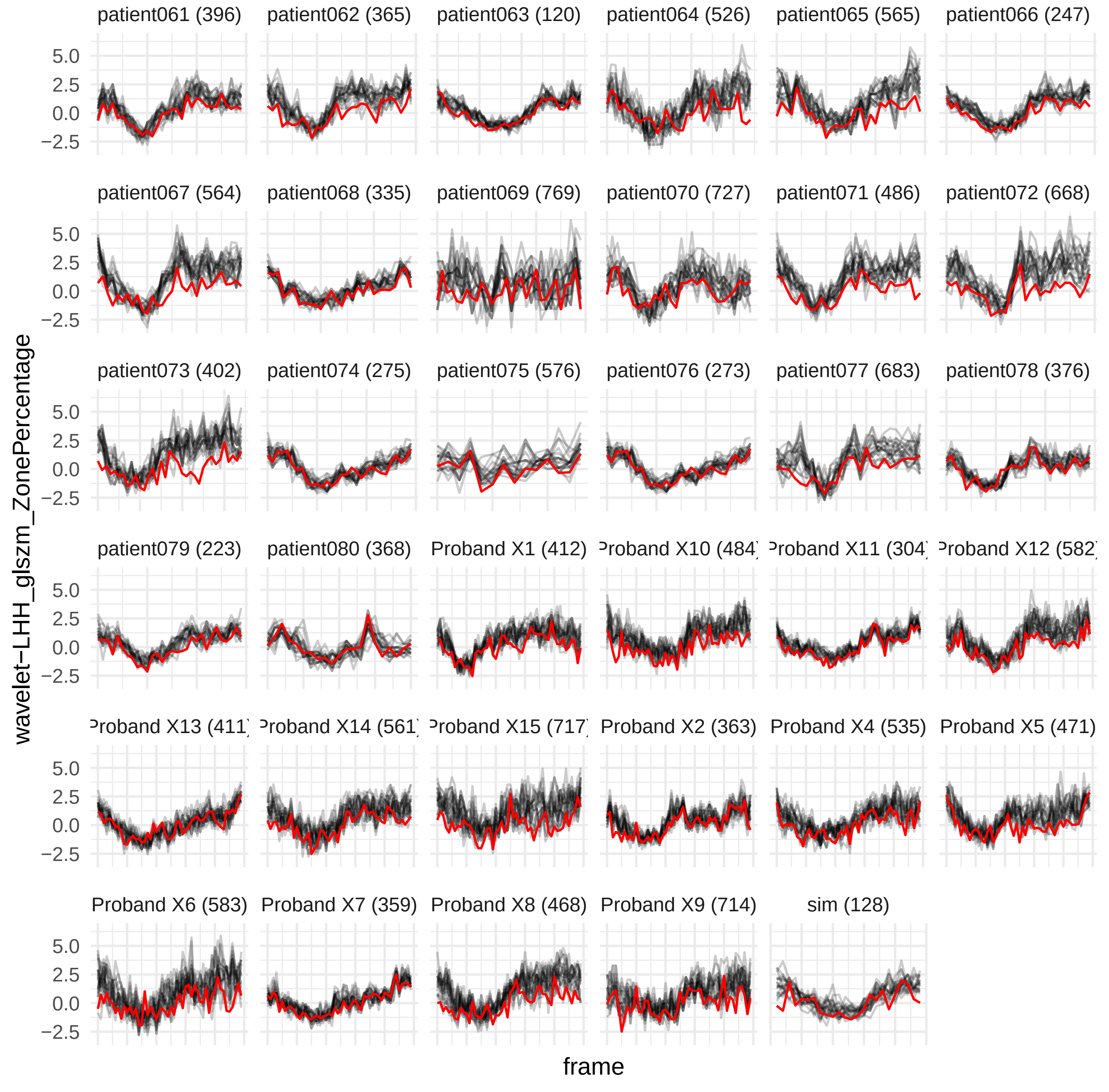

patient061 (396)
patient062 (365)
patient063 (120)
patient064 (526)
patient065 (565)
patient066 (247)
patient067 (564)
patient068 (335)
patient069 (769)
patient070 (727)
patient071 (486)
patient072 (668)
patient073 (402)
patient074 (275)
patient075 (576)
patient076 (273)
patient077 (683)
patient078 (376)
patient079 (223)
patient080 (368)
Proband X1 (412)
Proband X10 (484)
Proband X11 (304)
Proband X12 (582)
Proband X13 (411)
Proband X14 (561)
Proband X15 (717)
Proband X2 (363)
Proband X4 (535)
Proband X5 (471)
Proband X6 (583)
Proband X7 (359)
Proband X8 (468)
Proband X9 (714)
sim (128)
5.0
2.5
0.0
−2.5
wavelet−LHH_glszm_ZonePercentage
frame

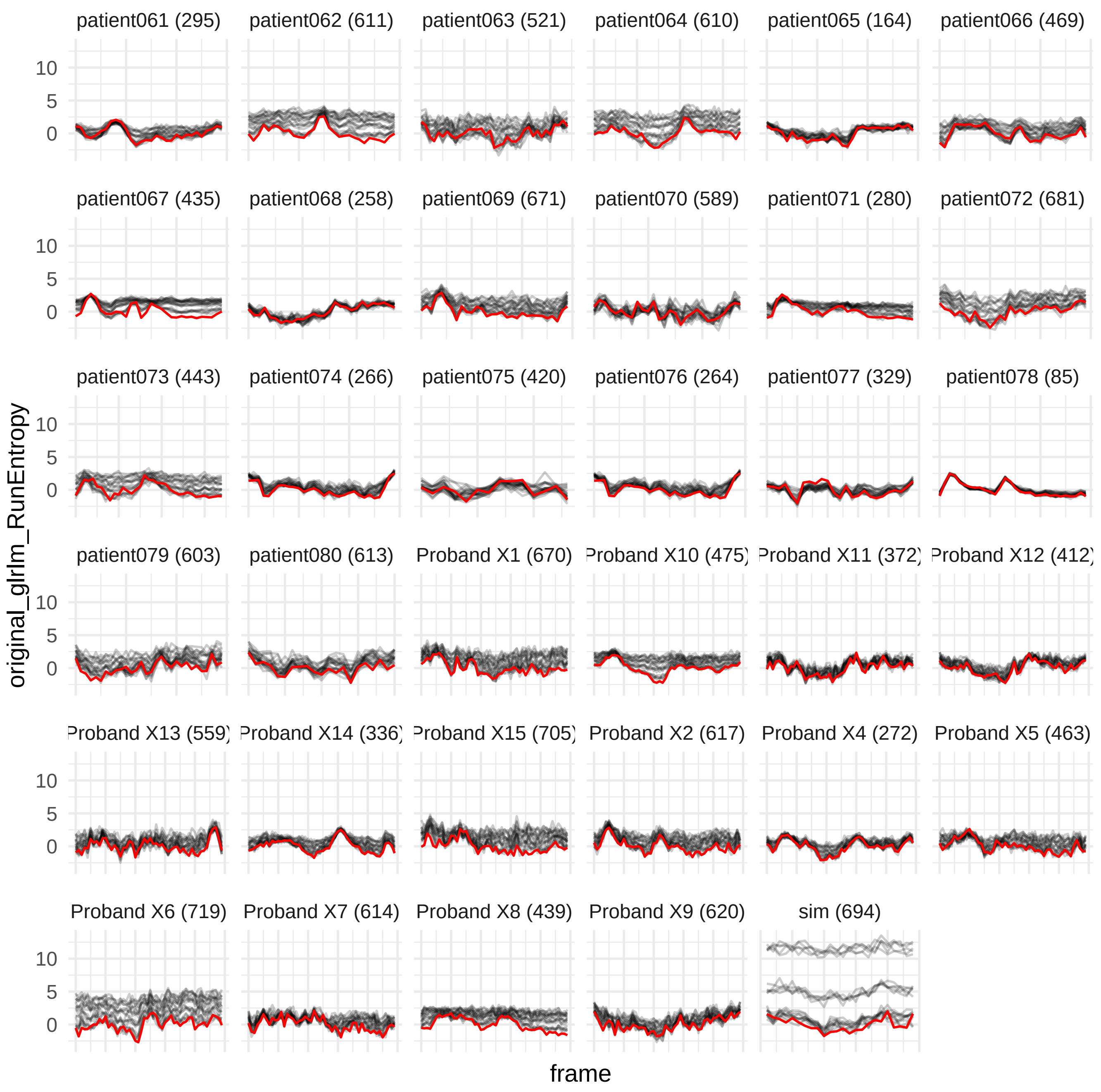
patient061 (295)
patient062 (611)
patient063 (521)
patient064 (610)
patient065 (164)
patient066 (469)
patient067 (435)
patient068 (258)
patient069 (671)
patient070 (589)
patient071 (280)
patient072 (681)
patient073 (443)
patient074 (266)
patient075 (420)
patient076 (264)
patient077 (329)
patient078 (85)
patient079 (603)
patient080 (613)
Proband X1 (670)
Proband X10 (475)
Proband X11 (372)
Proband X12 (412)
Proband X13 (559)
Proband X14 (336)
Proband X15 (705)
Proband X2 (617)
Proband X4 (272)
Proband X5 (463)
Proband X6 (719)
Proband X7 (614)
Proband X8 (439)
Proband X9 (620)
sim (694)
original_glrlm_RunEntropy
10
5
0
frame

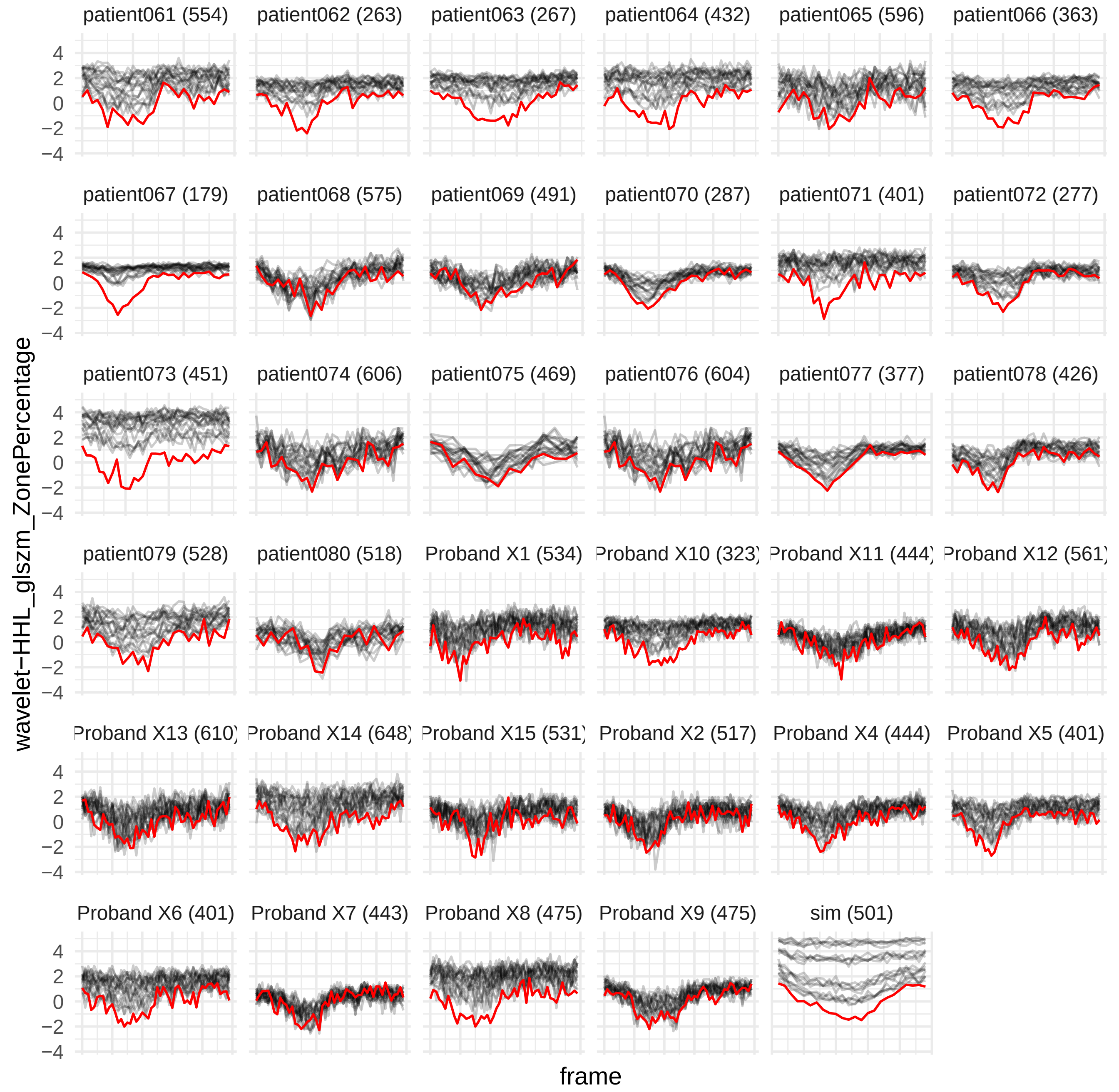

patient061 (554)
patient062 (263)
patient063 (267)
patient064 (432)
patient065 (596)
patient066 (363)
patient067 (179)
patient068 (575)
patient069 (491)
patient070 (287)
patient071 (401)
patient072 (277)
patient073 (451)
patient074 (606)
patient075 (469)
patient076 (604)
patient077 (377)
patient078 (426)
patient079 (528)
patient080 (518)
Proband X1 (534)
Proband X10 (323)
Proband X11 (444)
Proband X12 (561)
Proband X13 (610)
Proband X14 (648)
Proband X15 (531)
Proband X2 (517)
Proband X4 (444)
Proband X5 (401)
Proband X6 (401)
Proband X7 (443)
Proband X8 (475)
Proband X9 (475)
sim (501)
wavelet−HHL_glszm_ZonePercentage
4
2
0
−2
−4
frame

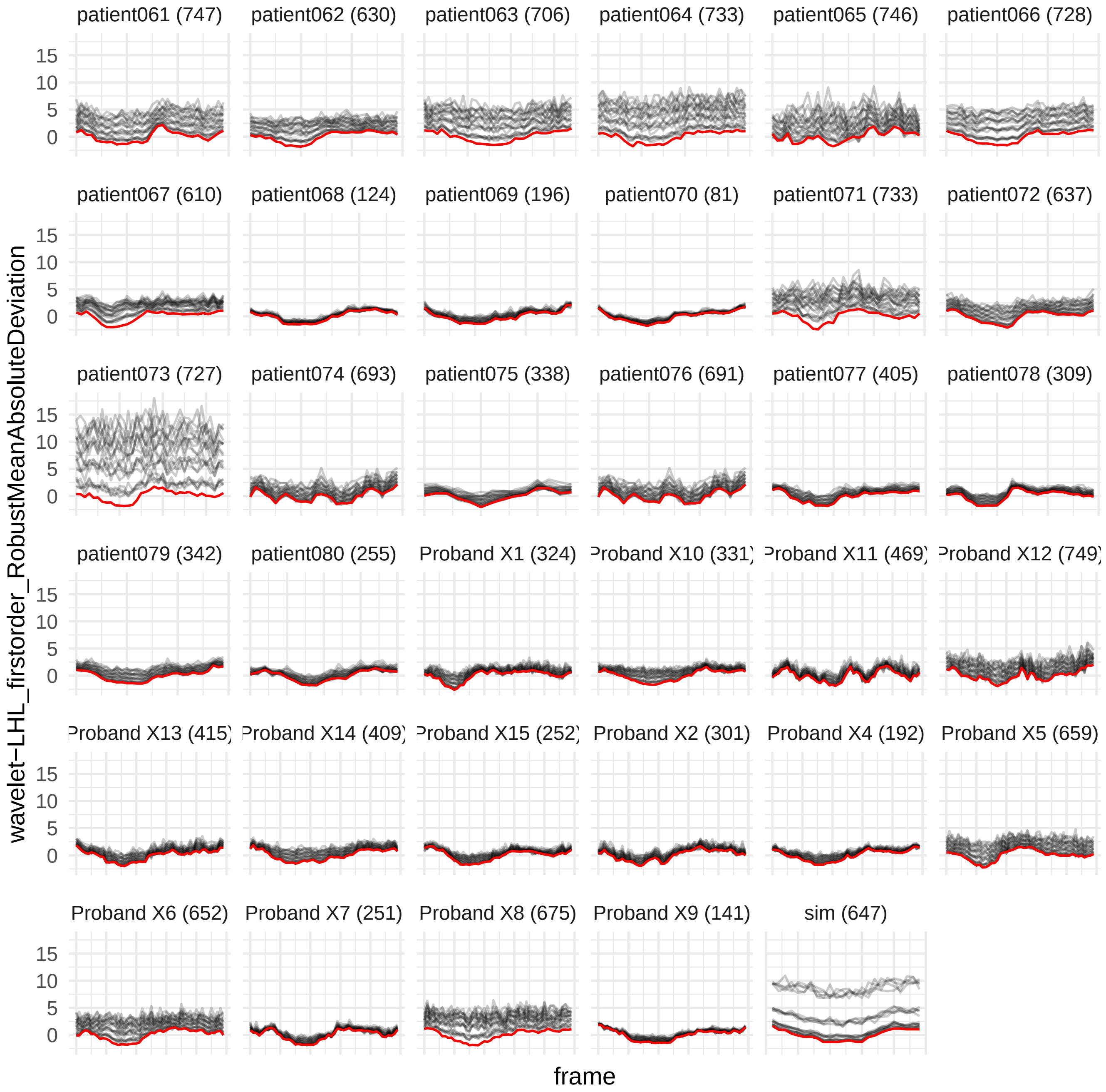

patient061 (747)
patient062 (630)
patient063 (706)
patient064 (733)
patient065 (746)
patient066 (728)
patient067 (610)
patient068 (124)
patient069 (196)
patient070 (81)
patient071 (733)
patient072 (637)
patient073 (727)
patient074 (693)
patient075 (338)
patient076 (691)
patient077 (405)
patient078 (309)
patient079 (342)
patient080 (255)
Proband X1 (324)
Proband X10 (331)
Proband X11 (469)
Proband X12 (749)
Proband X13 (415)
Proband X14 (409)
Proband X15 (252)
Proband X2 (301)
Proband X4 (192)
Proband X5 (659)
Proband X6 (652)
Proband X7 (251)
Proband X8 (675)
Proband X9 (141)
sim (647)
wavelet−LHL_firstorder_RobustMeanAbsoluteDeviation
frame
15
10
5
0

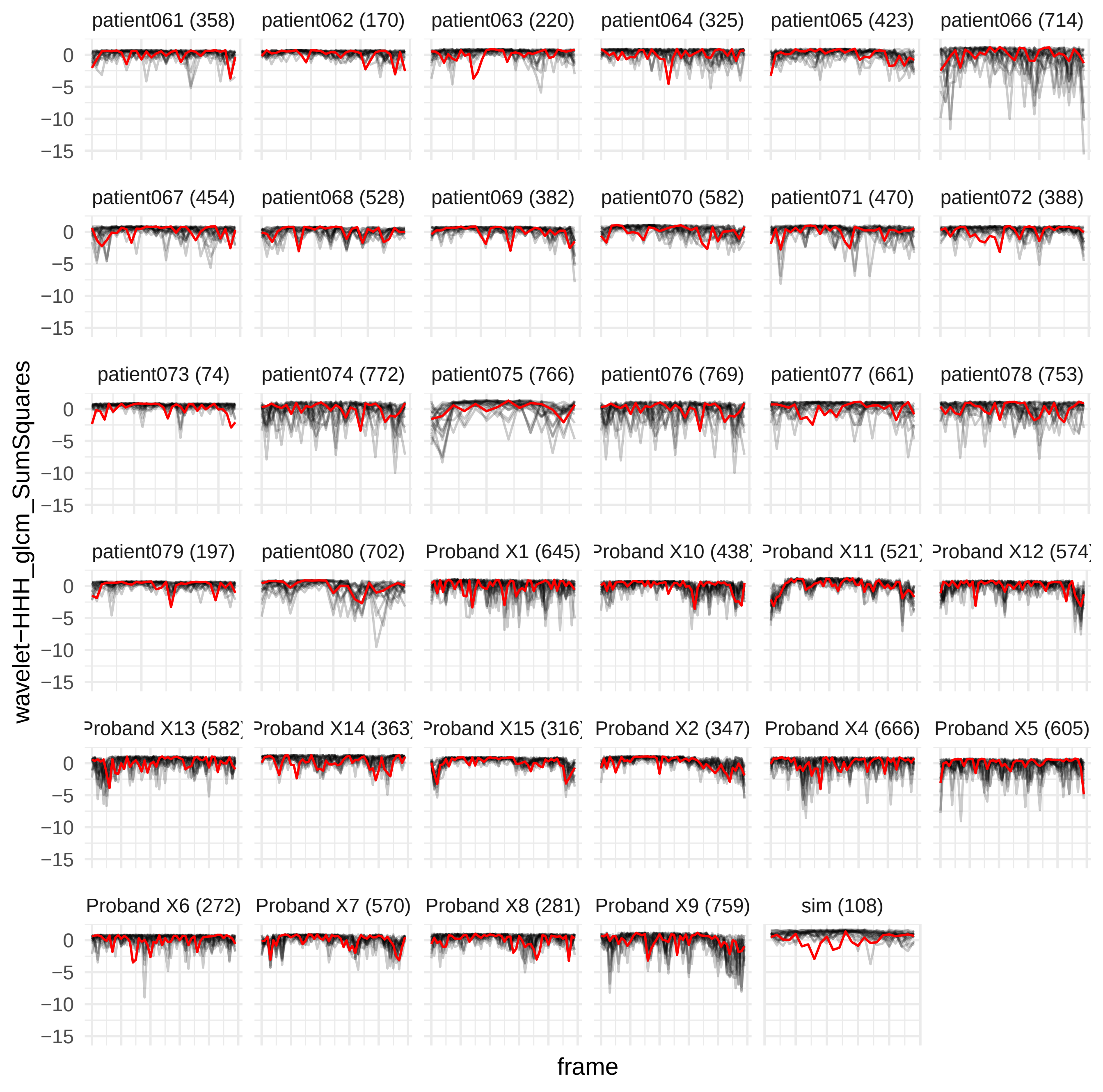
patient061 (358)
patient062 (170)
patient063 (220)
patient064 (325)
patient065 (423)
patient066 (714)
patient067 (454)
patient068 (528)
patient069 (382)
patient070 (582)
patient071 (470)
patient072 (388)
patient073 (74)
patient074 (772)
patient075 (766)
patient076 (769)
patient077 (661)
patient078 (753)
patient079 (197)
patient080 (702)
Proband X1 (645)
Proband X10 (438)
Proband X11 (521)
Proband X12 (574)
Proband X13 (582)
Proband X14 (363)
Proband X15 (316)
Proband X2 (347)
Proband X4 (666)
Proband X5 (605)
Proband X6 (272)
Proband X7 (570)
Proband X8 (281)
Proband X9 (759)
sim (108)
0
−5
−10
−15
wavelet−HHH_glcm_SumSquares
frame

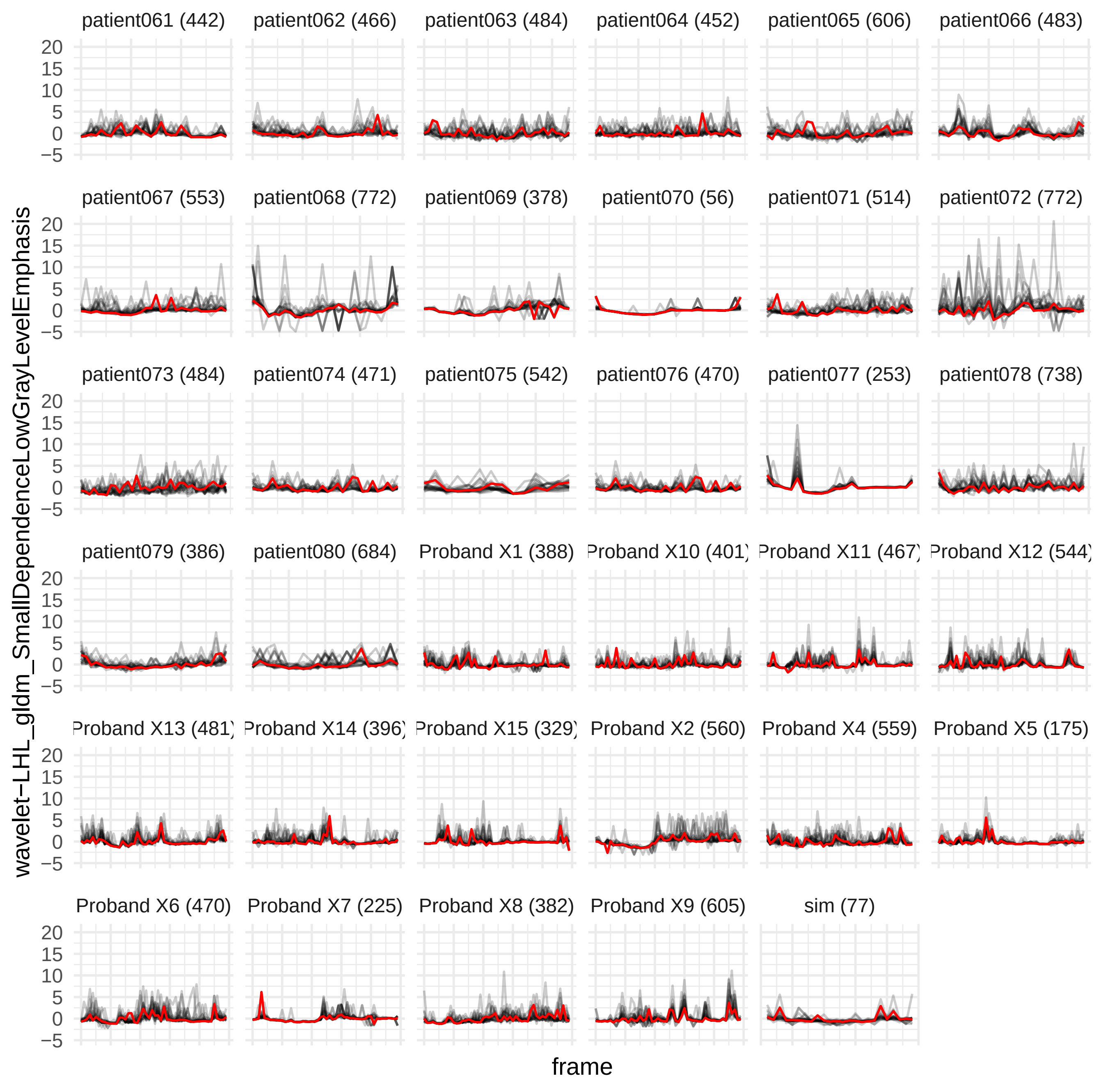

patient061 (442)
patient062 (466)
patient063 (484)
patient064 (452)
patient065 (606)
patient066 (483)
patient067 (553)
patient068 (772)
patient069 (378)
patient070 (56)
patient071 (514)
patient072 (772)
patient073 (484)
patient074 (471)
patient075 (542)
patient076 (470)
patient077 (253)
patient078 (738)
patient079 (386)
patient080 (684)
Proband X1 (388)
Proband X10 (401)
Proband X11 (467)
Proband X12 (544)
Proband X13 (481)
Proband X14 (396)
Proband X15 (329)
Proband X2 (560)
Proband X4 (559)
Proband X5 (175)
Proband X6 (470)
Proband X7 (225)
Proband X8 (382)
Proband X9 (605)
sim (77)
20
15
10
5
0
−5
wavelet−LHL_gldm_SmallDependenceLowGrayLevelEmphasis
frame

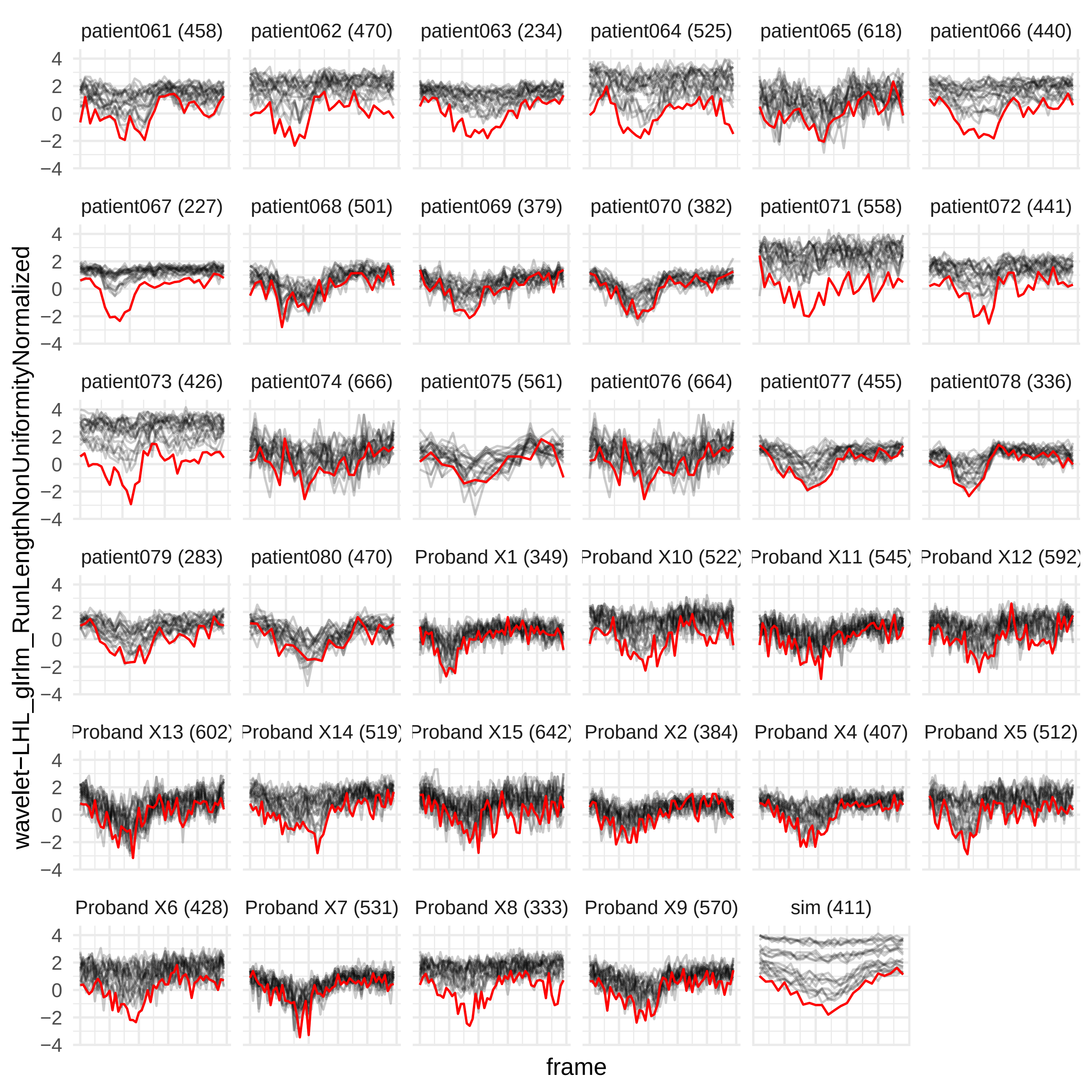

patient061 (458)
patient062 (470)
patient063 (234)
patient064 (525)
patient065 (618)
patient066 (440)
patient067 (227)
patient068 (501)
patient069 (379)
patient070 (382)
patient071 (558)
patient072 (441)
patient073 (426)
patient074 (666)
patient075 (561)
patient076 (664)
patient077 (455)
patient078 (336)
patient079 (283)
patient080 (470)
Proband X1 (349)
Proband X10 (522)
Proband X11 (545)
Proband X12 (592)
Proband X13 (602)
Proband X14 (519)
Proband X15 (642)
Proband X2 (384)
Proband X4 (407)
Proband X5 (512)
Proband X6 (428)
Proband X7 (531)
Proband X8 (333)
Proband X9 (570)
sim (411)
4
2
0
−2
−4
wavelet−LHL_glrlm_RunLengthNonUniformityNormalized
frame

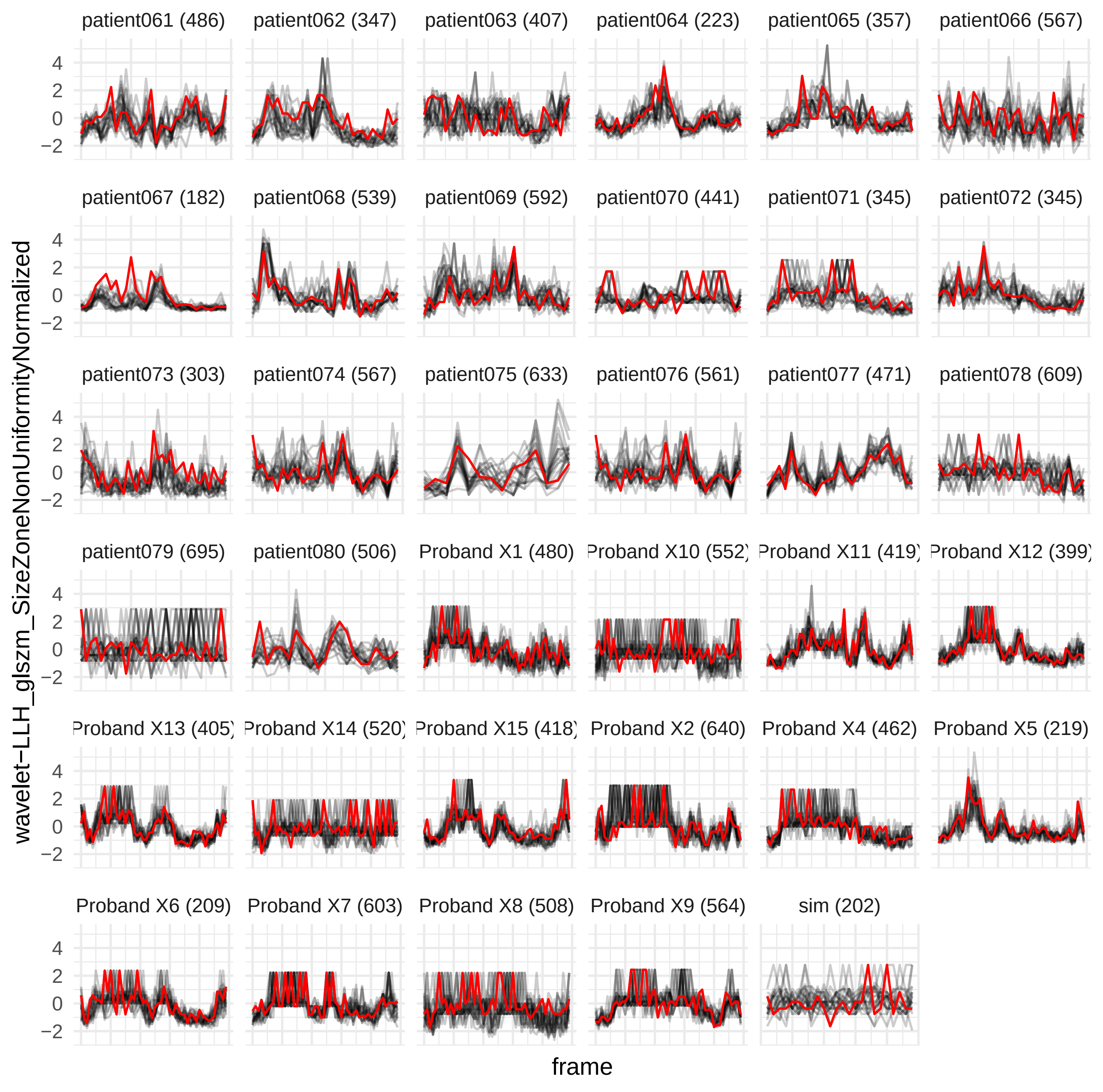

patient061 (486)
patient062 (347)
patient063 (407)
patient064 (223)
patient065 (357)
patient066 (567)
patient067 (182)
patient068 (539)
patient069 (592)
patient070 (441)
patient071 (345)
patient072 (345)
patient073 (303)
patient074 (567)
patient075 (633)
patient076 (561)
patient077 (471)
patient078 (609)
patient079 (695)
patient080 (506)
Proband X1 (480)
Proband X10 (552)
Proband X11 (419)
Proband X12 (399)
Proband X13 (405)
Proband X14 (520)
Proband X15 (418)
Proband X2 (640)
Proband X4 (462)
Proband X5 (219)
Proband X6 (209)
Proband X7 (603)
Proband X8 (508)
Proband X9 (564)
sim (202)
wavelet−LLH_glszm_SizeZoneNonUniformityNormalized
frame

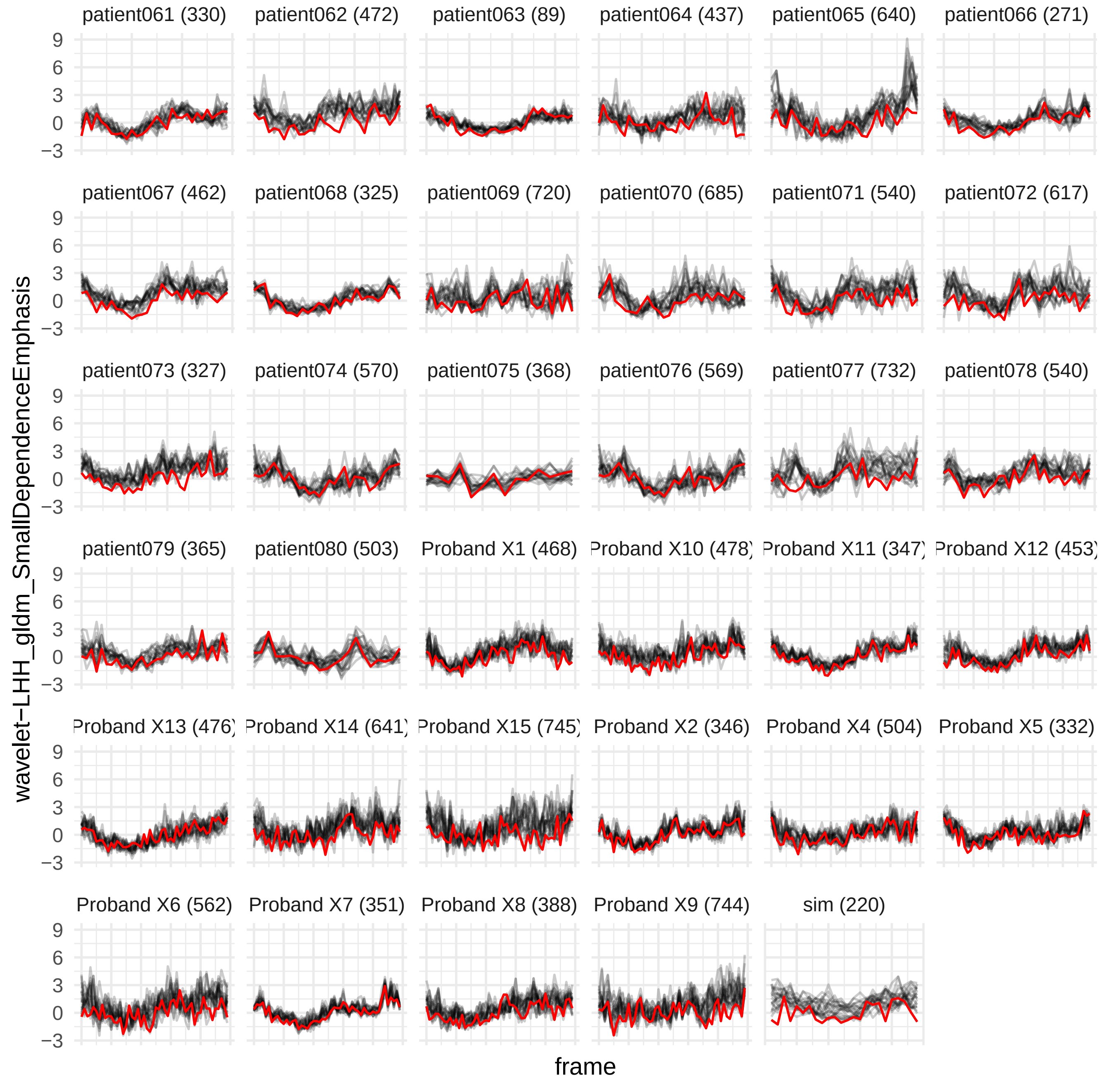
patient061 (330)
patient062 (472)
patient063 (89)
patient064 (437)
patient065 (640)
patient066 (271)
patient067 (462)
patient068 (325)
patient069 (720)
patient070 (685)
patient071 (540)
patient072 (617)
patient073 (327)
patient074 (570)
patient075 (368)
patient076 (569)
patient077 (732)
patient078 (540)
patient079 (365)
patient080 (503)
Proband X1 (468)
Proband X10 (478)
Proband X11 (347)
Proband X12 (453)
Proband X13 (476)
Proband X14 (641)
Proband X15 (745)
Proband X2 (346)
Proband X4 (504)
Proband X5 (332)
Proband X6 (562)
Proband X7 (351)
Proband X8 (388)
Proband X9 (744)
sim (220)
9
6
3
0
−3
wavelet−LHH_gldm_SmallDependenceEmphasis
frame

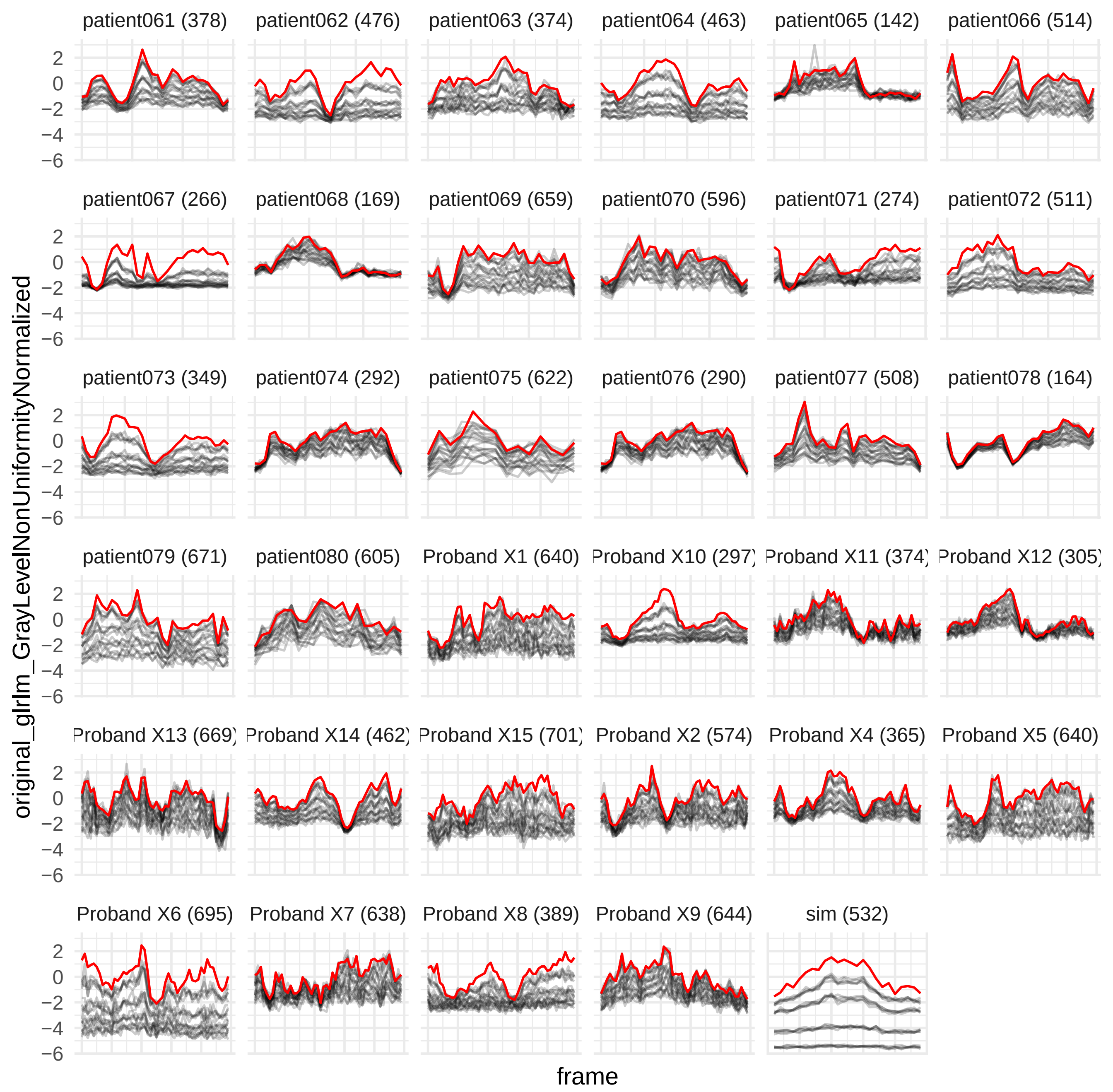

patient061 (378)
patient062 (476)
patient063 (374)
patient064 (463)
patient065 (142)
patient066 (514)
patient067 (266)
patient068 (169)
patient069 (659)
patient070 (596)
patient071 (274)
patient072 (511)
patient073 (349)
patient074 (292)
patient075 (622)
patient076 (290)
patient077 (508)
patient078 (164)
patient079 (671)
patient080 (605)
Proband X1 (640)
Proband X10 (297)
Proband X11 (374)
Proband X12 (305)
Proband X13 (669)
Proband X14 (462)
Proband X15 (701)
Proband X2 (574)
Proband X4 (365)
Proband X5 (640)
Proband X6 (695)
Proband X7 (638)
Proband X8 (389)
Proband X9 (644)
sim (532)
original_glrlm_GrayLevelNonUniformityNormalized
frame
2
0
−2
−4
−6

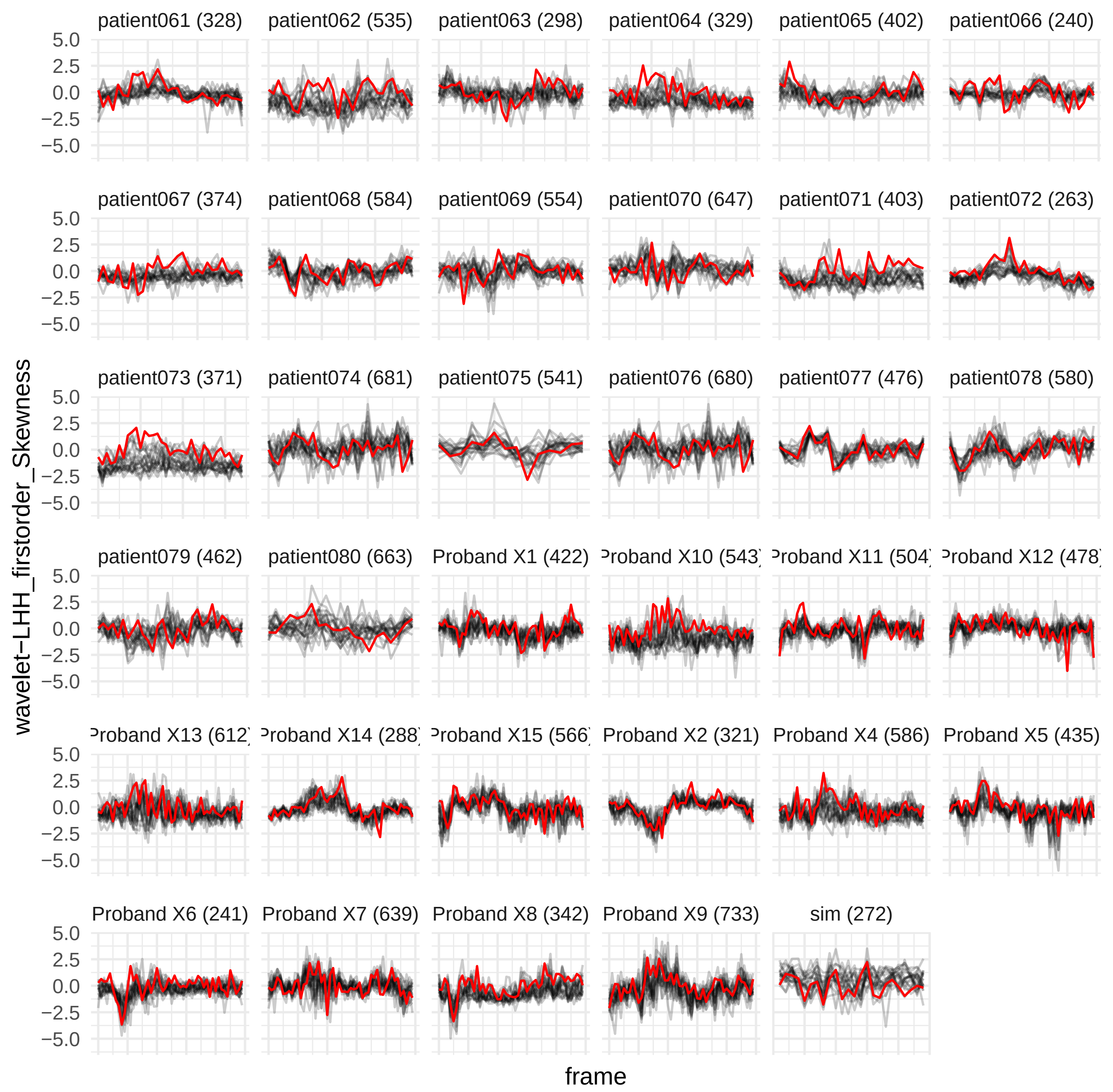

patient061 (328)
patient062 (535)
patient063 (298)
patient064 (329)
patient065 (402)
patient066 (240)
patient067 (374)
patient068 (584)
patient069 (554)
patient070 (647)
patient071 (403)
patient072 (263)
patient073 (371)
patient074 (681)
patient075 (541)
patient076 (680)
patient077 (476)
patient078 (580)
patient079 (462)
patient080 (663)
Proband X1 (422)
Proband X10 (543)
Proband X11 (504)
Proband X12 (478)
Proband X13 (612)
Proband X14 (288)
Proband X15 (566)
Proband X2 (321)
Proband X4 (586)
Proband X5 (435)
Proband X6 (241)
Proband X7 (639)
Proband X8 (342)
Proband X9 (733)
sim (272)
5.0
2.5
0.0
−2.5
−5.0
wavelet−LHH_firstorder_Skewness
frame

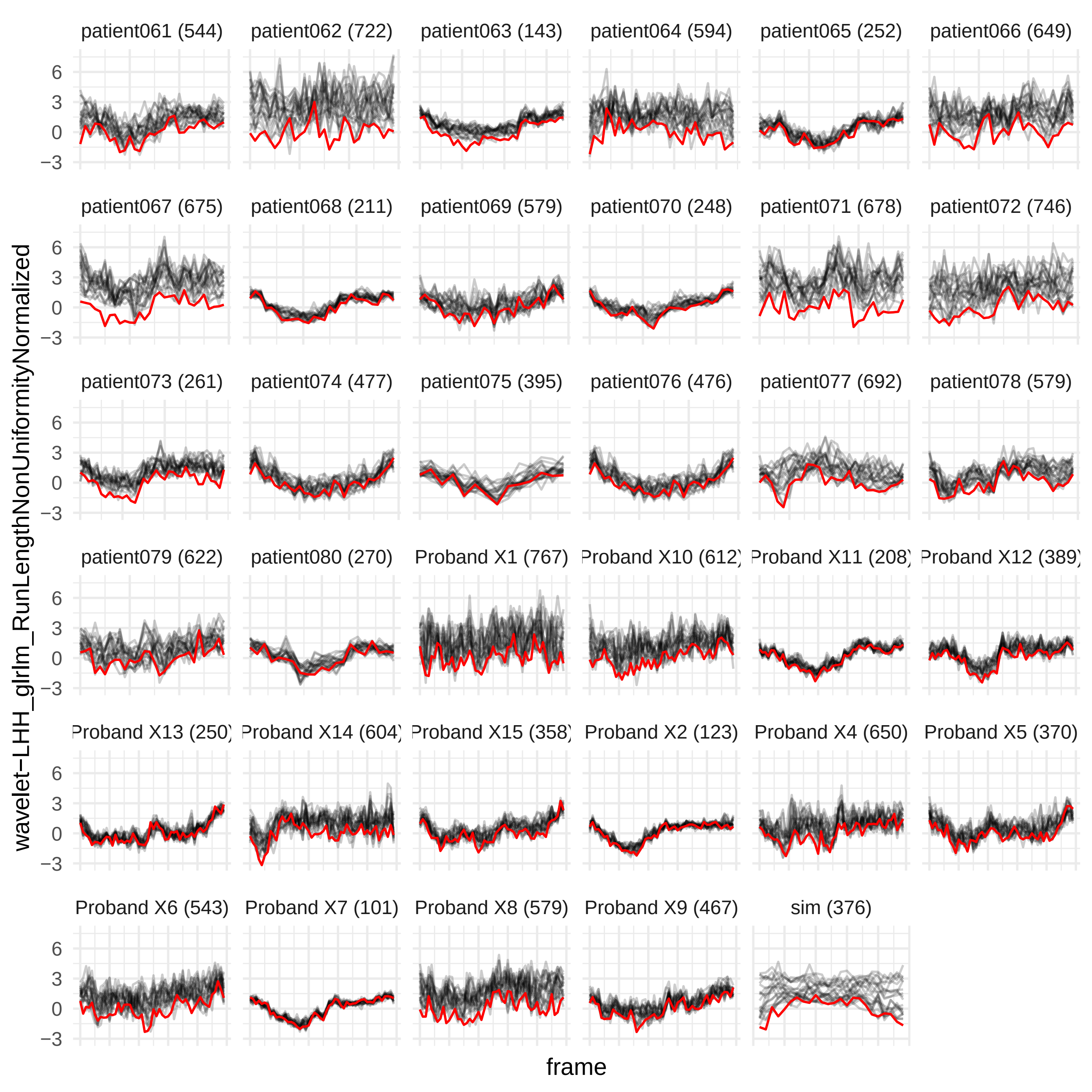
patient061 (544)
patient062 (722)
patient063 (143)
patient064 (594)
patient065 (252)
patient066 (649)
patient067 (675)
patient068 (211)
patient069 (579)
patient070 (248)
patient071 (678)
patient072 (746)
patient073 (261)
patient074 (477)
patient075 (395)
patient076 (476)
patient077 (692)
patient078 (579)
patient079 (622)
patient080 (270)
Proband X1 (767)
Proband X10 (612)
Proband X11 (208)
Proband X12 (389)
Proband X13 (250)
Proband X14 (604)
Proband X15 (358)
Proband X2 (123)
Proband X4 (650)
Proband X5 (370)
Proband X6 (543)
Proband X7 (101)
Proband X8 (579)
Proband X9 (467)
sim (376)
6
3
0
−3
wavelet−LHH_glrlm_RunLengthNonUniformityNormalized
frame

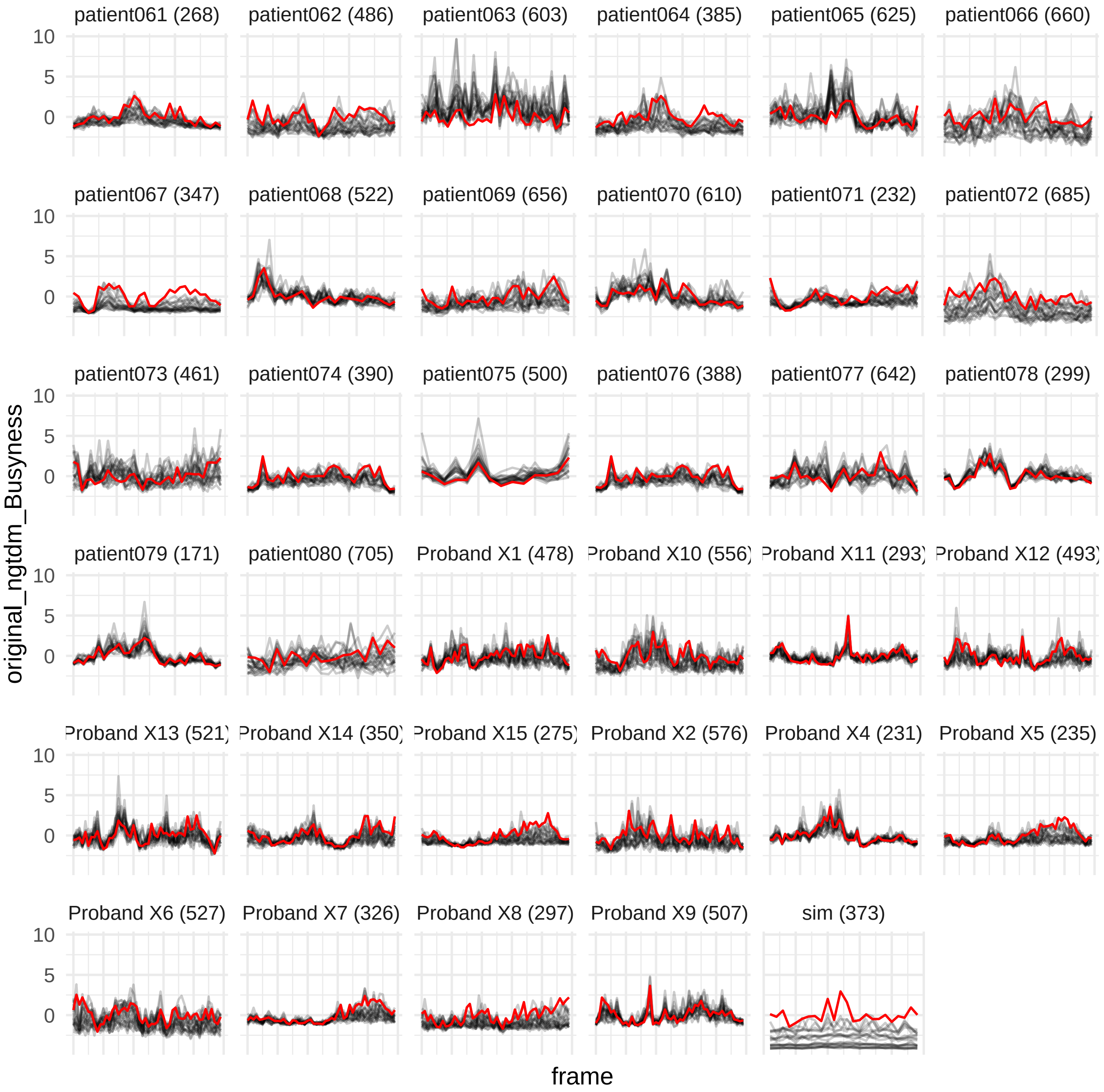

patient061 (268)
patient062 (486)
patient063 (603)
patient064 (385)
patient065 (625)
patient066 (660)
patient067 (347)
patient068 (522)
patient069 (656)
patient070 (610)
patient071 (232)
patient072 (685)
patient073 (461)
patient074 (390)
patient075 (500)
patient076 (388)
patient077 (642)
patient078 (299)
patient079 (171)
patient080 (705)
Proband X1 (478)
Proband X10 (556)
Proband X11 (293)
Proband X12 (493)
Proband X13 (521)
Proband X14 (350)
Proband X15 (275)
Proband X2 (576)
Proband X4 (231)
Proband X5 (235)
Proband X6 (527)
Proband X7 (326)
Proband X8 (297)
Proband X9 (507)
sim (373)
original_ngtdm_Busyness
10
5
0
frame

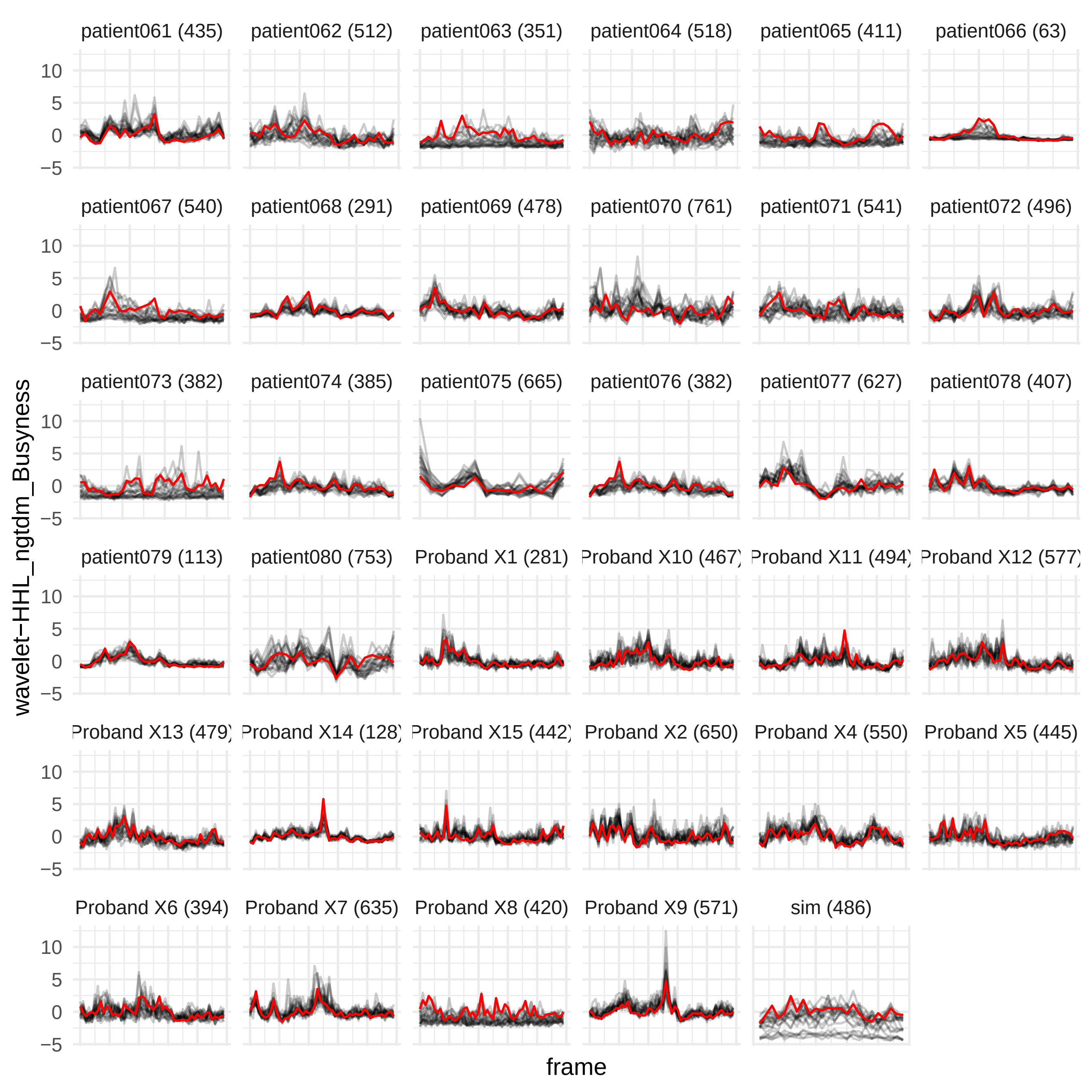

patient061 (435)
patient062 (512)
patient063 (351)
patient064 (518)
patient065 (411)
patient066 (63)
patient067 (540)
patient068 (291)
patient069 (478)
patient070 (761)
patient071 (541)
patient072 (496)
patient073 (382)
patient074 (385)
patient075 (665)
patient076 (382)
patient077 (627)
patient078 (407)
patient079 (113)
patient080 (753)
Proband X1 (281)
Proband X10 (467)
Proband X11 (494)
Proband X12 (577)
Proband X13 (479)
Proband X14 (128)
Proband X15 (442)
Proband X2 (650)
Proband X4 (550)
Proband X5 (445)
Proband X6 (394)
Proband X7 (635)
Proband X8 (420)
Proband X9 (571)
sim (486)
wavelet−HHL_ngtdm_Busyness
frame
10
5
0
−5

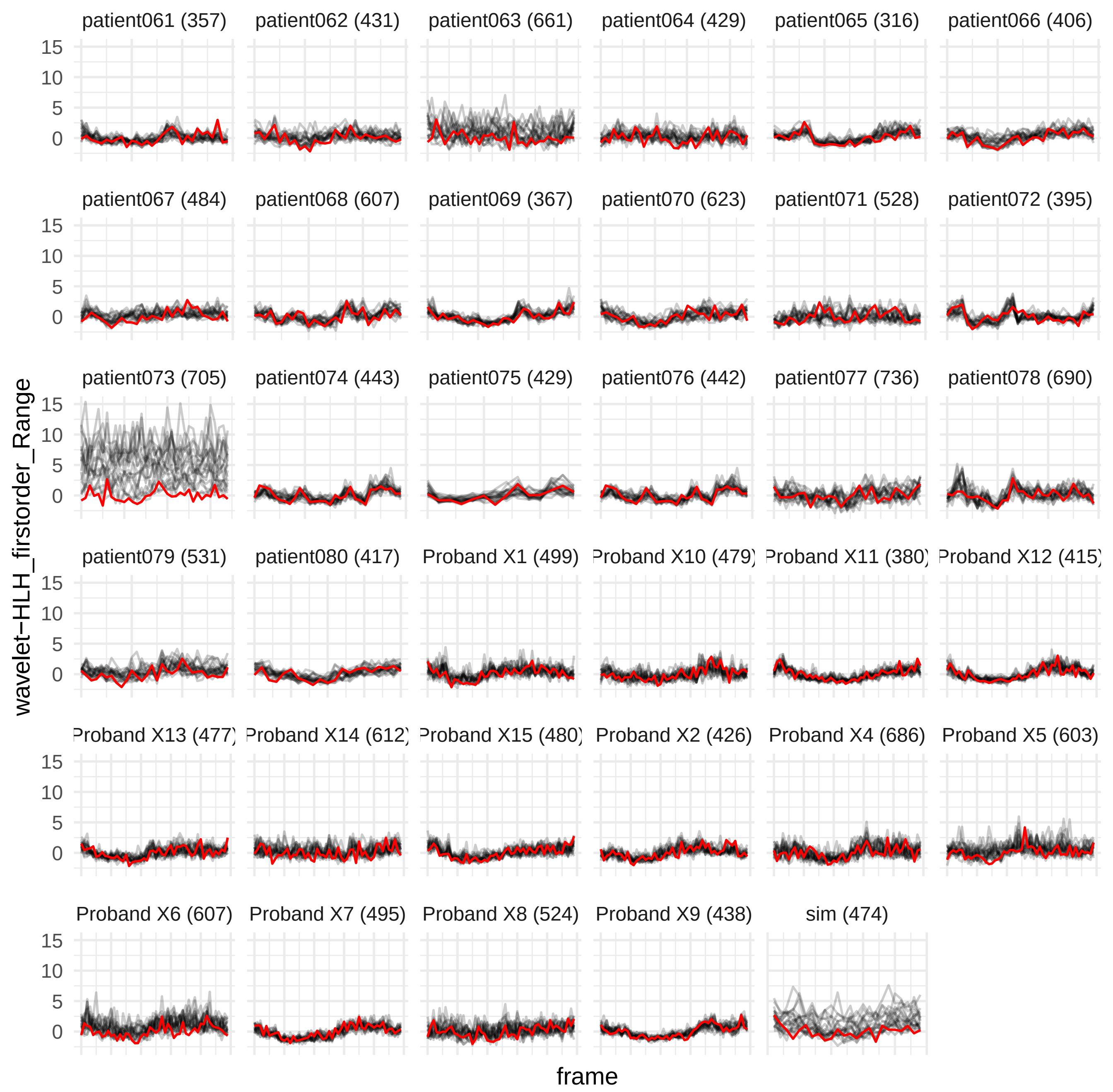

patient061 (357)
patient062 (431)
patient063 (661)
patient064 (429)
patient065 (316)
patient066 (406)
patient067 (484)
patient068 (607)
patient069 (367)
patient070 (623)
patient071 (528)
patient072 (395)
patient073 (705)
patient074 (443)
patient075 (429)
patient076 (442)
patient077 (736)
patient078 (690)
patient079 (531)
patient080 (417)
Proband X1 (499)
Proband X10 (479)
Proband X11 (380)
Proband X12 (415)
Proband X13 (477)
Proband X14 (612)
Proband X15 (480)
Proband X2 (426)
Proband X4 (686)
Proband X5 (603)
Proband X6 (607)
Proband X7 (495)
Proband X8 (524)
Proband X9 (438)
sim (474)
wavelet−HLH_firstorder_Range
0
5
10
15
frame

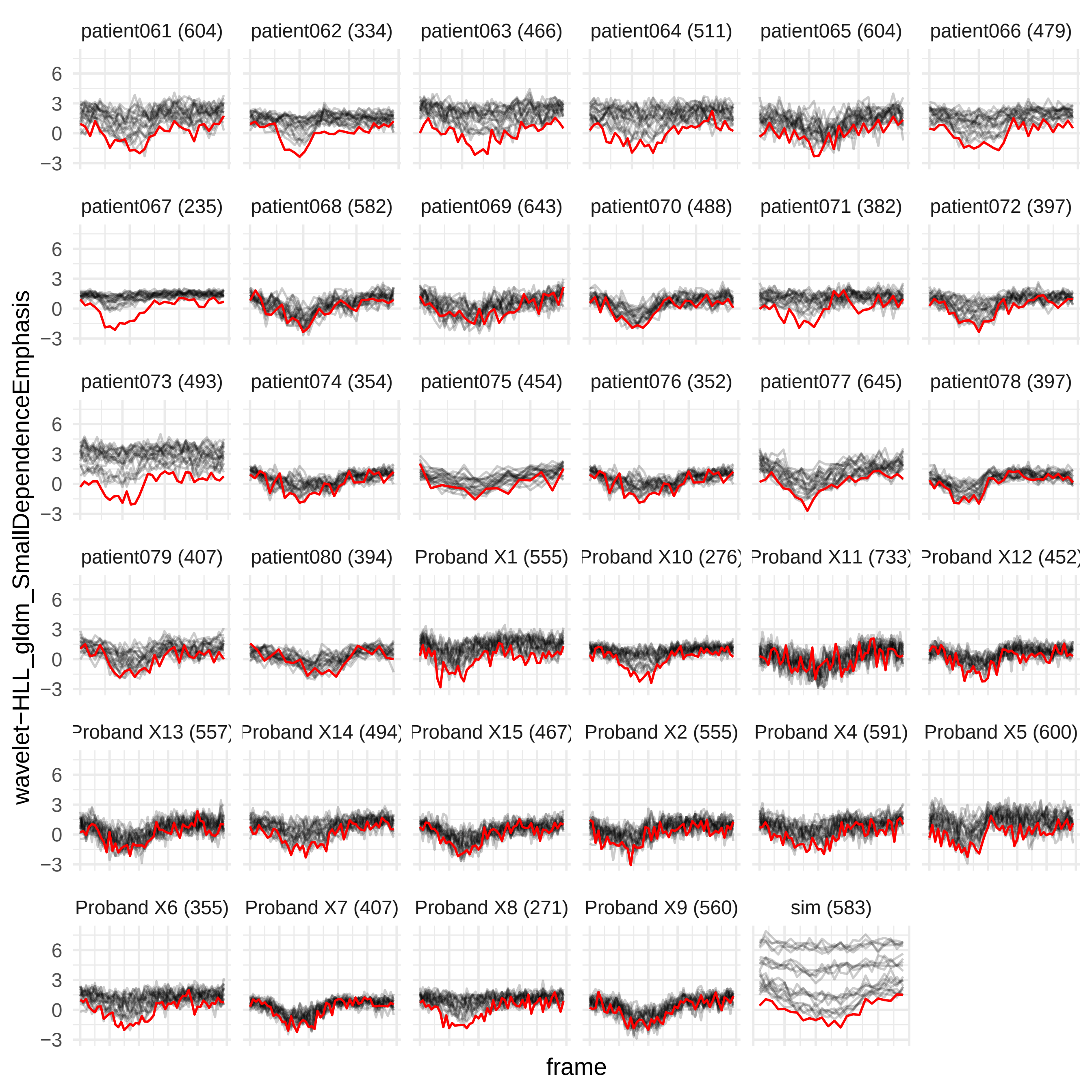

patient061 (604)
patient062 (334)
patient063 (466)
patient064 (511)
patient065 (604)
patient066 (479)
patient067 (235)
patient068 (582)
patient069 (643)
patient070 (488)
patient071 (382)
patient072 (397)
patient073 (493)
patient074 (354)
patient075 (454)
patient076 (352)
patient077 (645)
patient078 (397)
patient079 (407)
patient080 (394)
Proband X1 (555)
Proband X10 (276)
Proband X11 (733)
Proband X12 (452)
Proband X13 (557)
Proband X14 (494)
Proband X15 (467)
Proband X2 (555)
Proband X4 (591)
Proband X5 (600)
Proband X6 (355)
Proband X7 (407)
Proband X8 (271)
Proband X9 (560)
sim (583)
6
3
0
−3
wavelet−HLL_gldm_SmallDependenceEmphasis
frame

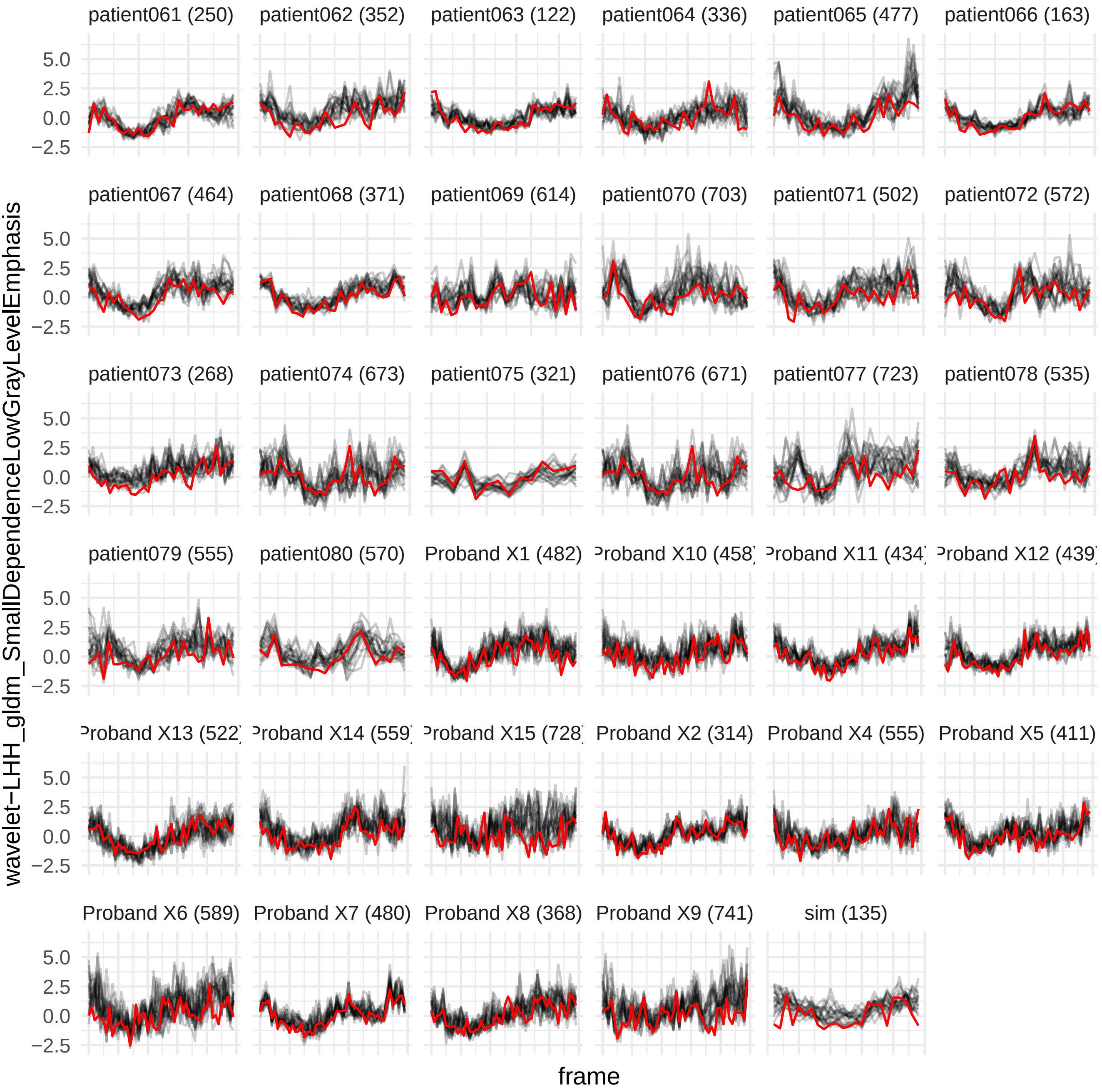

patient061 (250)
patient062 (352)
patient063 (122)
patient064 (336)
patient065 (477)
patient066 (163)
patient067 (464)
patient068 (371)
patient069 (614)
patient070 (703)
patient071 (502)
patient072 (572)
patient073 (268)
patient074 (673)
patient075 (321)
patient076 (671)
patient077 (723)
patient078 (535)
patient079 (555)
patient080 (570)
Proband X1 (482)
Proband X10 (458)
Proband X11 (434)
Proband X12 (439)
Proband X13 (522)
Proband X14 (559)
Proband X15 (728)
Proband X2 (314)
Proband X4 (555)
Proband X5 (411)
Proband X6 (589)
Proband X7 (480)
Proband X8 (368)
Proband X9 (741)
sim (135)
5.0
2.5
0.0
−2.5
wavelet−LHH_gldm_SmallDependenceLowGrayLevelEmphasis
frame

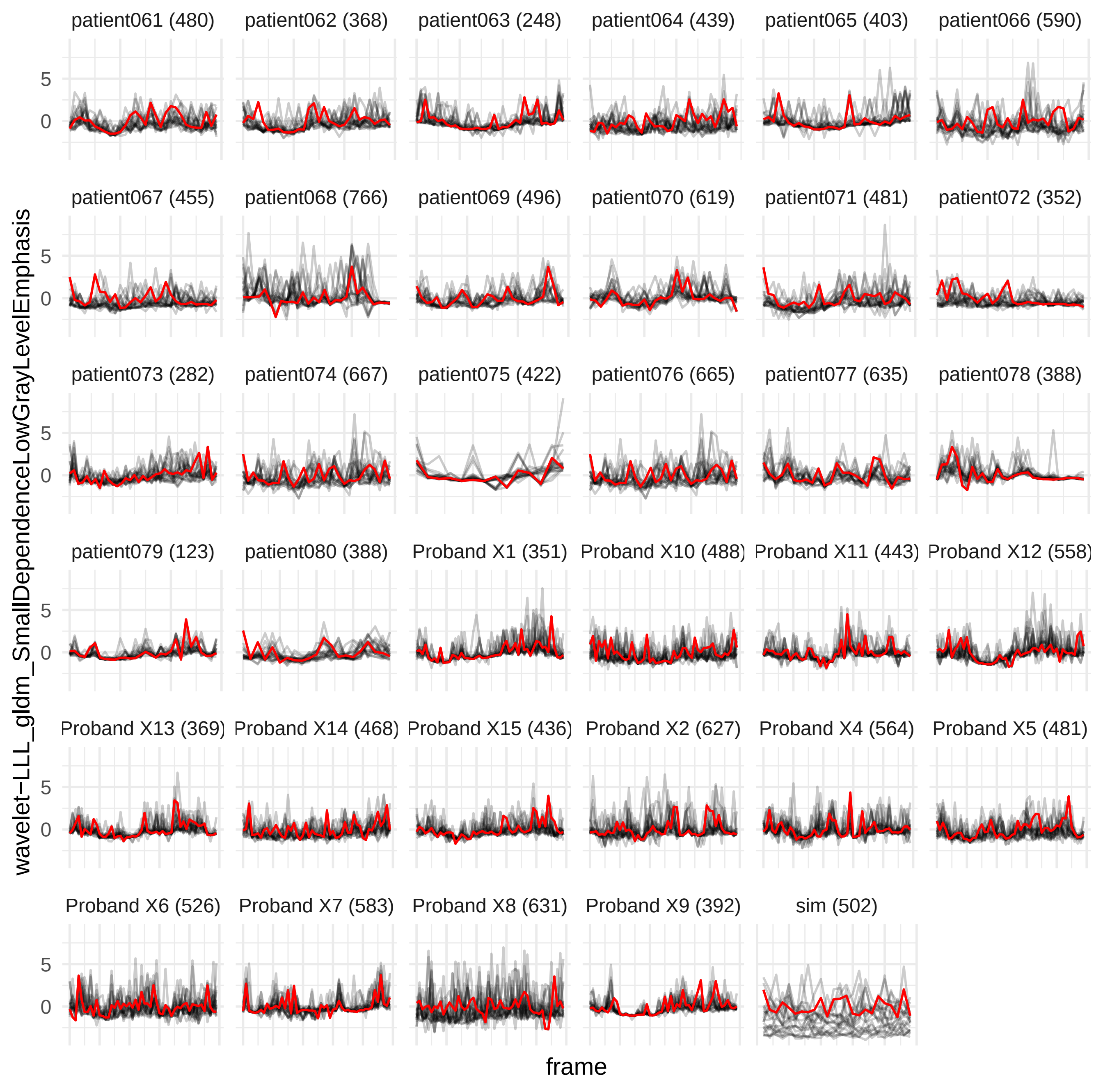

patient061 (480)
patient062 (368)
patient063 (248)
patient064 (439)
patient065 (403)
patient066 (590)
patient067 (455)
patient068 (766)
patient069 (496)
patient070 (619)
patient071 (481)
patient072 (352)
patient073 (282)
patient074 (667)
patient075 (422)
patient076 (665)
patient077 (635)
patient078 (388)
patient079 (123)
patient080 (388)
Proband X1 (351)
Proband X10 (488)
Proband X11 (443)
Proband X12 (558)
Proband X13 (369)
Proband X14 (468)
Proband X15 (436)
Proband X2 (627)
Proband X4 (564)
Proband X5 (481)
Proband X6 (526)
Proband X7 (583)
Proband X8 (631)
Proband X9 (392)
sim (502)
5
0
wavelet−LLL_gldm_SmallDependenceLowGrayLevelEmphasis
frame

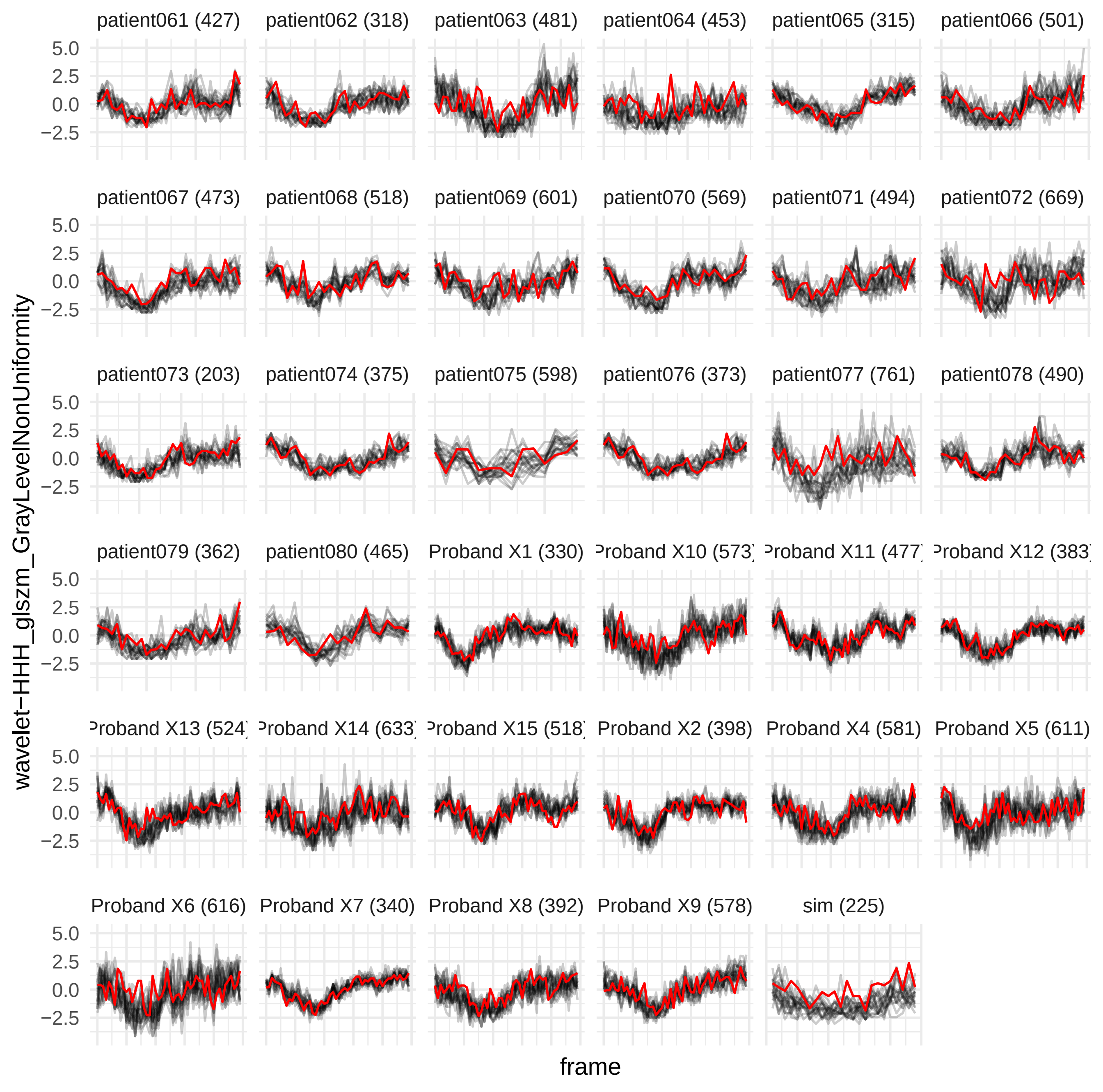

patient061 (427)
patient062 (318)
patient063 (481)
patient064 (453)
patient065 (315)
patient066 (501)
patient067 (473)
patient068 (518)
patient069 (601)
patient070 (569)
patient071 (494)
patient072 (669)
patient073 (203)
patient074 (375)
patient075 (598)
patient076 (373)
patient077 (761)
patient078 (490)
patient079 (362)
patient080 (465)
Proband X1 (330)
Proband X10 (573)
Proband X11 (477)
Proband X12 (383)
Proband X13 (524)
Proband X14 (633)
Proband X15 (518)
Proband X2 (398)
Proband X4 (581)
Proband X5 (611)
Proband X6 (616)
Proband X7 (340)
Proband X8 (392)
Proband X9 (578)
sim (225)
5.0
2.5
0.0
−2.5
wavelet−HHH_glszm_GrayLevelNonUniformity
frame

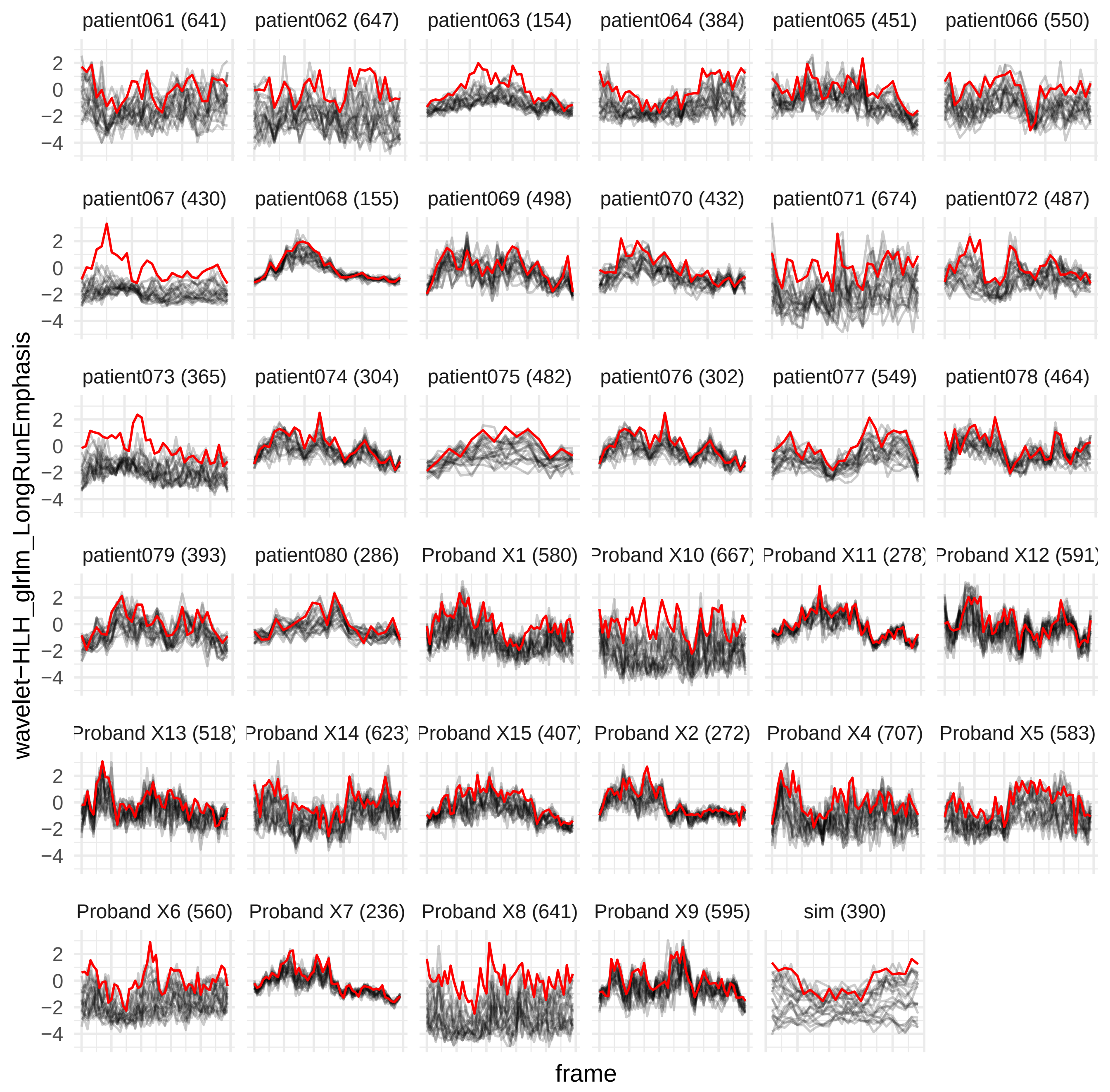

patient061 (641)
patient062 (647)
patient063 (154)
patient064 (384)
patient065 (451)
patient066 (550)
patient067 (430)
patient068 (155)
patient069 (498)
patient070 (432)
patient071 (674)
patient072 (487)
patient073 (365)
patient074 (304)
patient075 (482)
patient076 (302)
patient077 (549)
patient078 (464)
patient079 (393)
patient080 (286)
Proband X1 (580)
Proband X10 (667)
Proband X11 (278)
Proband X12 (591)
Proband X13 (518)
Proband X14 (623)
Proband X15 (407)
Proband X2 (272)
Proband X4 (707)
Proband X5 (583)
Proband X6 (560)
Proband X7 (236)
Proband X8 (641)
Proband X9 (595)
sim (390)
2
0
−2
−4
wavelet−HLH_glrlm_LongRunEmphasis
frame

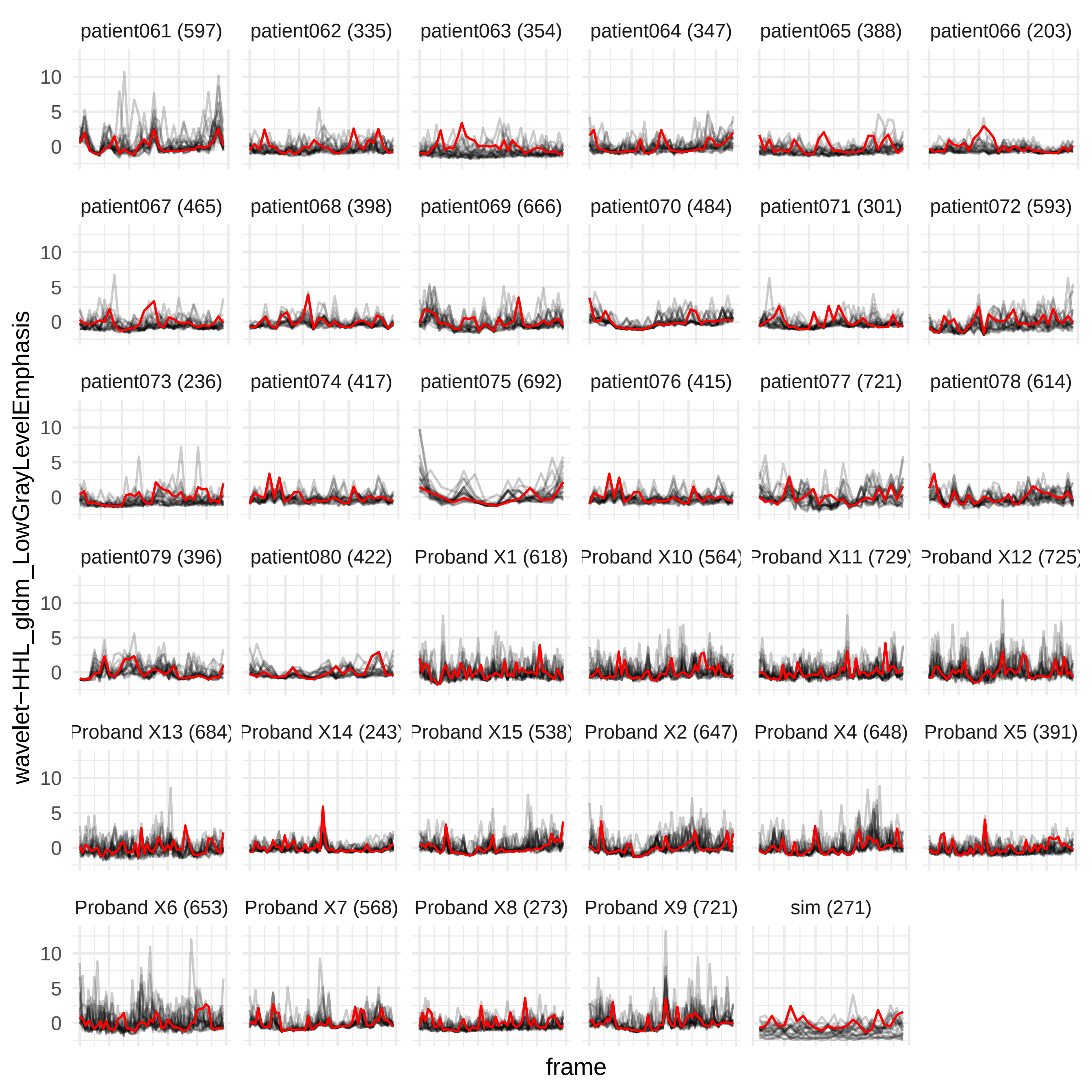
patient061 (597)
patient062 (335)
patient063 (354)
patient064 (347)
patient065 (388)
patient066 (203)
patient067 (465)
patient068 (398)
patient069 (666)
patient070 (484)
patient071 (301)
patient072 (593)
patient073 (236)
patient074 (417)
patient075 (692)
patient076 (415)
patient077 (721)
patient078 (614)
patient079 (396)
patient080 (422)
Proband X1 (618)
Proband X10 (564)
Proband X11 (729)
Proband X12 (725)
Proband X13 (684)
Proband X14 (243)
Proband X15 (538)
Proband X2 (647)
Proband X4 (648)
Proband X5 (391)
Proband X6 (653)
Proband X7 (568)
Proband X8 (273)
Proband X9 (721)
sim (271)
wavelet−HHL_gldm_LowGrayLevelEmphasis
0
5
10
frame

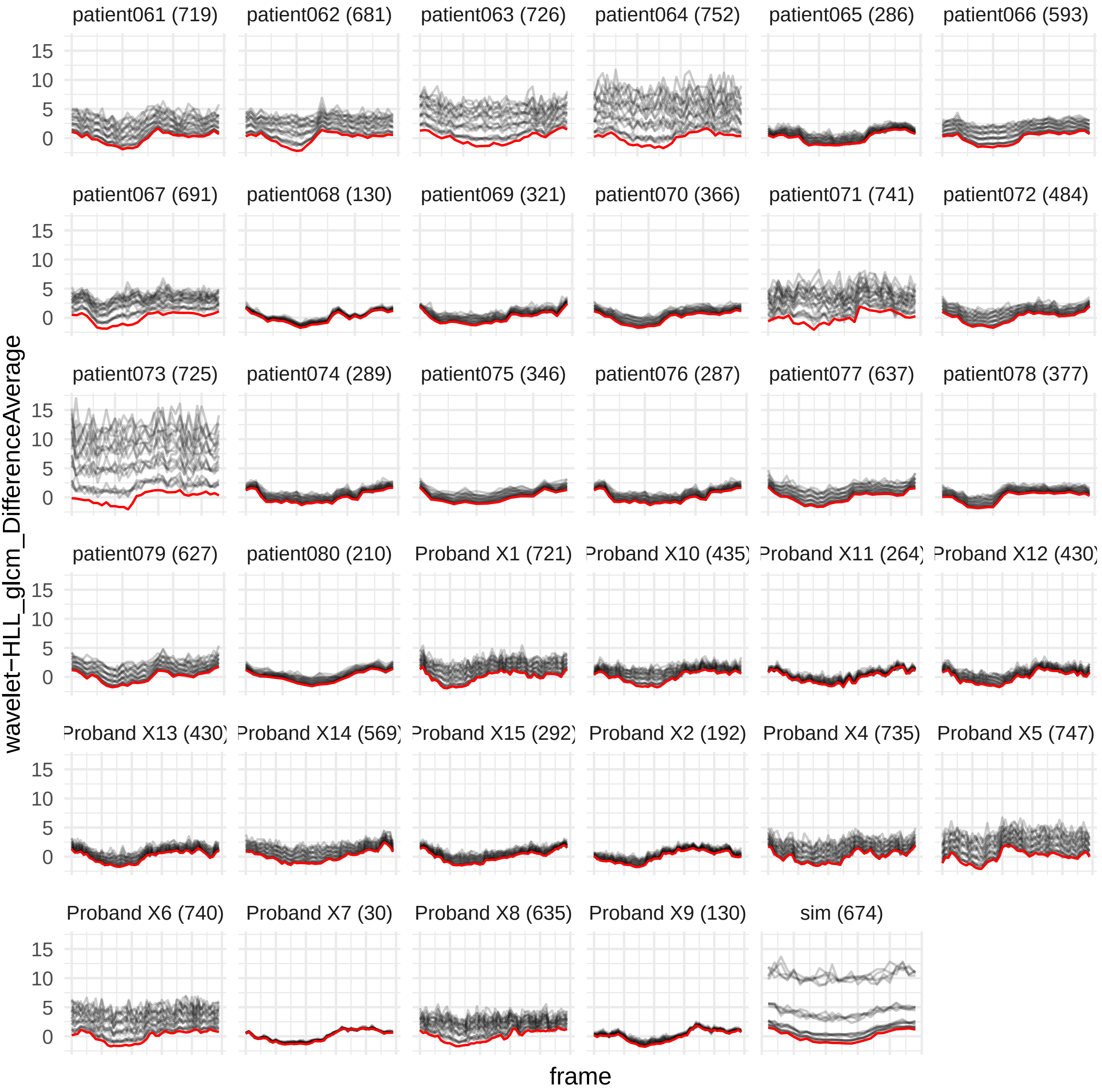

patient061 (719)
patient062 (681)
patient063 (726)
patient064 (752)
patient065 (286)
patient066 (593)
patient067 (691)
patient068 (130)
patient069 (321)
patient070 (366)
patient071 (741)
patient072 (484)
patient073 (725)
patient074 (289)
patient075 (346)
patient076 (287)
patient077 (637)
patient078 (377)
patient079 (627)
patient080 (210)
Proband X1 (721)
Proband X10 (435)
Proband X11 (264)
Proband X12 (430)
Proband X13 (430)
Proband X14 (569)
Proband X15 (292)
Proband X2 (192)
Proband X4 (735)
Proband X5 (747)
Proband X6 (740)
Proband X7 (30)
Proband X8 (635)
Proband X9 (130)
sim (674)
wavelet−HLL_glcm_DifferenceAverage
frame
15
10
5
0

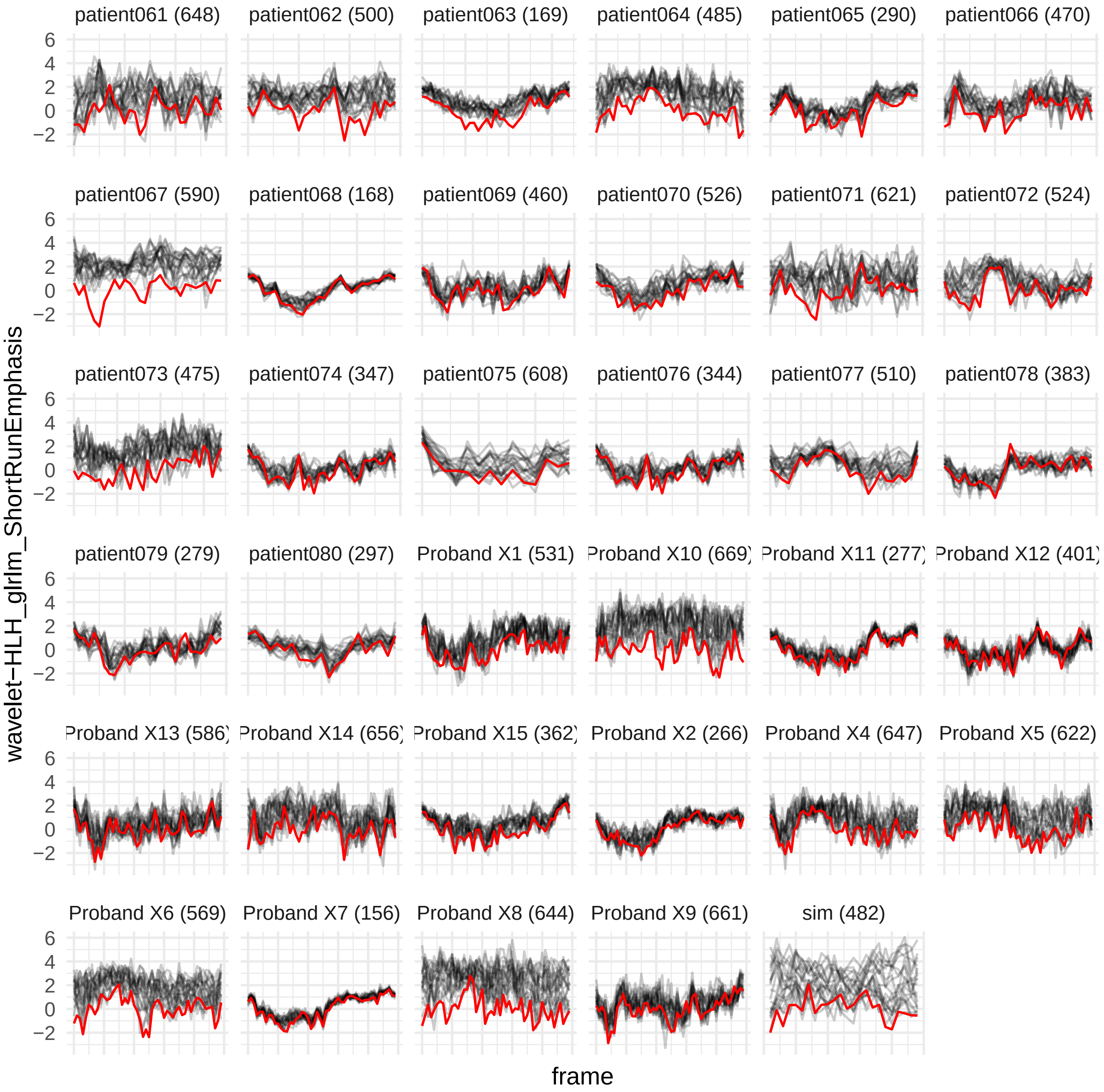

patient061 (648)
patient062 (500)
patient063 (169)
patient064 (485)
patient065 (290)
patient066 (470)
patient067 (590)
patient068 (168)
patient069 (460)
patient070 (526)
patient071 (621)
patient072 (524)
patient073 (475)
patient074 (347)
patient075 (608)
patient076 (344)
patient077 (510)
patient078 (383)
patient079 (279)
patient080 (297)
Proband X1 (531)
Proband X10 (669)
Proband X11 (277)
Proband X12 (401)
Proband X13 (586)
Proband X14 (656)
Proband X15 (362)
Proband X2 (266)
Proband X4 (647)
Proband X5 (622)
Proband X6 (569)
Proband X7 (156)
Proband X8 (644)
Proband X9 (661)
sim (482)
6
4
2
0
−2
wavelet−HLH_glrlm_ShortRunEmphasis
frame

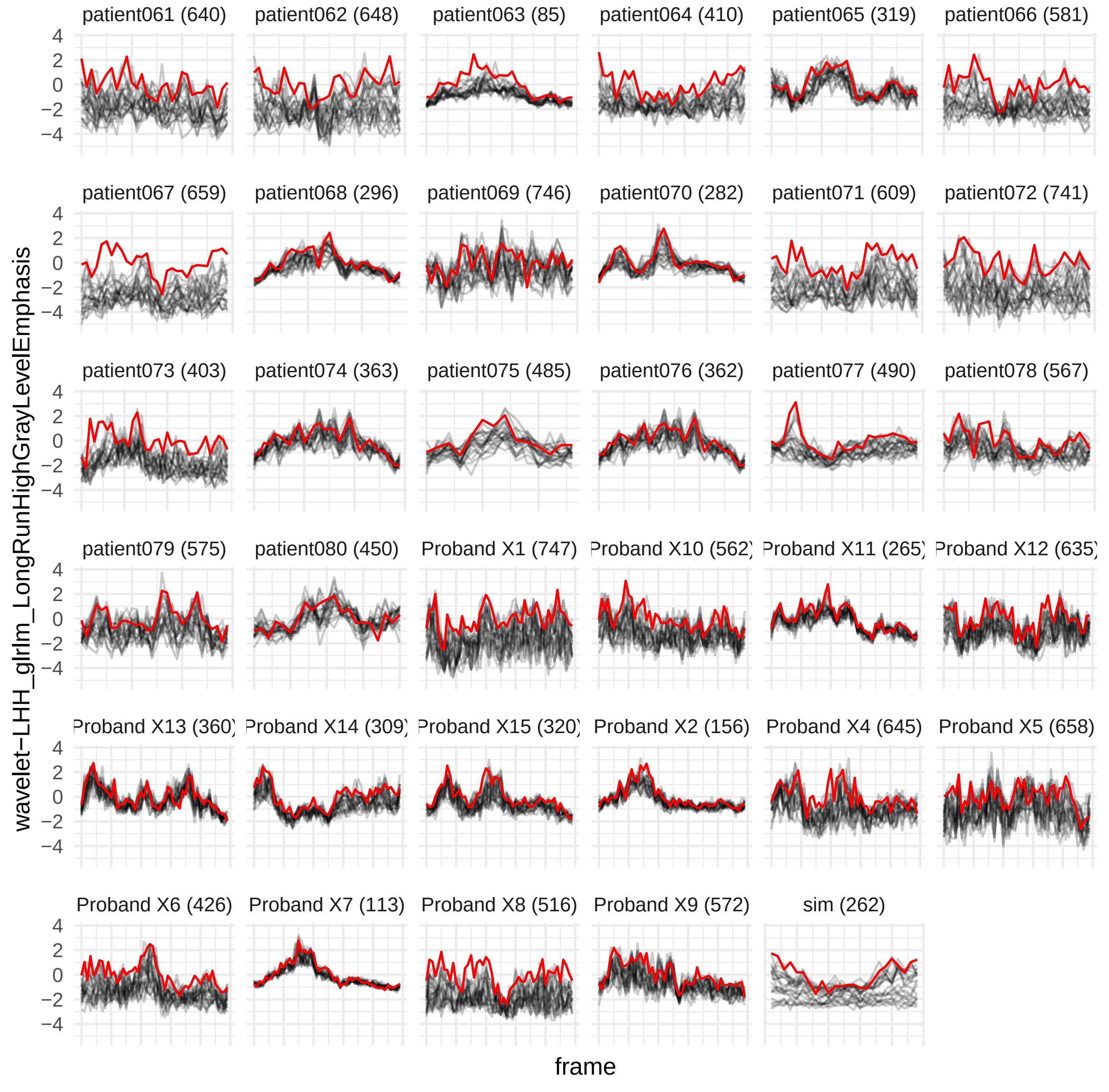

patient061 (640)
patient062 (648)
patient063 (85)
patient064 (410)
patient065 (319)
patient066 (581)
patient067 (659)
patient068 (296)
patient069 (746)
patient070 (282)
patient071 (609)
patient072 (741)
patient073 (403)
patient074 (363)
patient075 (485)
patient076 (362)
patient077 (490)
patient078 (567)
patient079 (575)
patient080 (450)
Proband X1 (747)
Proband X10 (562)
Proband X11 (265)
Proband X12 (635)
Proband X13 (360)
Proband X14 (309)
Proband X15 (320)
Proband X2 (156)
Proband X4 (645)
Proband X5 (658)
Proband X6 (426)
Proband X7 (113)
Proband X8 (516)
Proband X9 (572)
sim (262)
4
2
0
−2
−4
wavelet−LHH_glrlm_LongRunHighGrayLevelEmphasis
frame

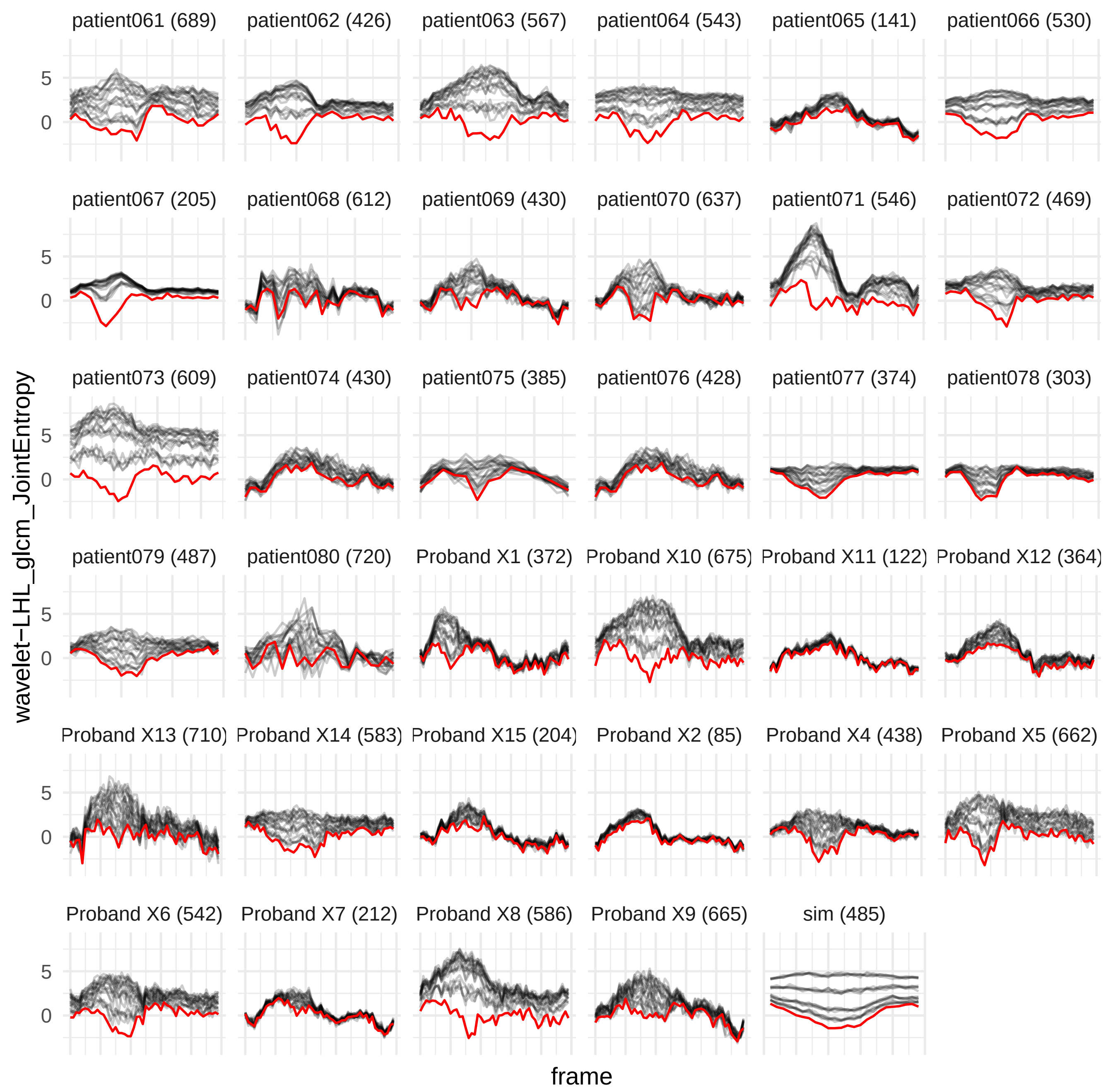

patient061 (689)
patient062 (426)
patient063 (567)
patient064 (543)
patient065 (141)
patient066 (530)
patient067 (205)
patient068 (612)
patient069 (430)
patient070 (637)
patient071 (546)
patient072 (469)
patient073 (609)
patient074 (430)
patient075 (385)
patient076 (428)
patient077 (374)
patient078 (303)
patient079 (487)
patient080 (720)
Proband X1 (372)
Proband X10 (675)
Proband X11 (122)
Proband X12 (364)
Proband X13 (710)
Proband X14 (583)
Proband X15 (204)
Proband X2 (85)
Proband X4 (438)
Proband X5 (662)
Proband X6 (542)
Proband X7 (212)
Proband X8 (586)
Proband X9 (665)
sim (485)
wavelet−LHL_glcm_JointEntropy
frame
5
0

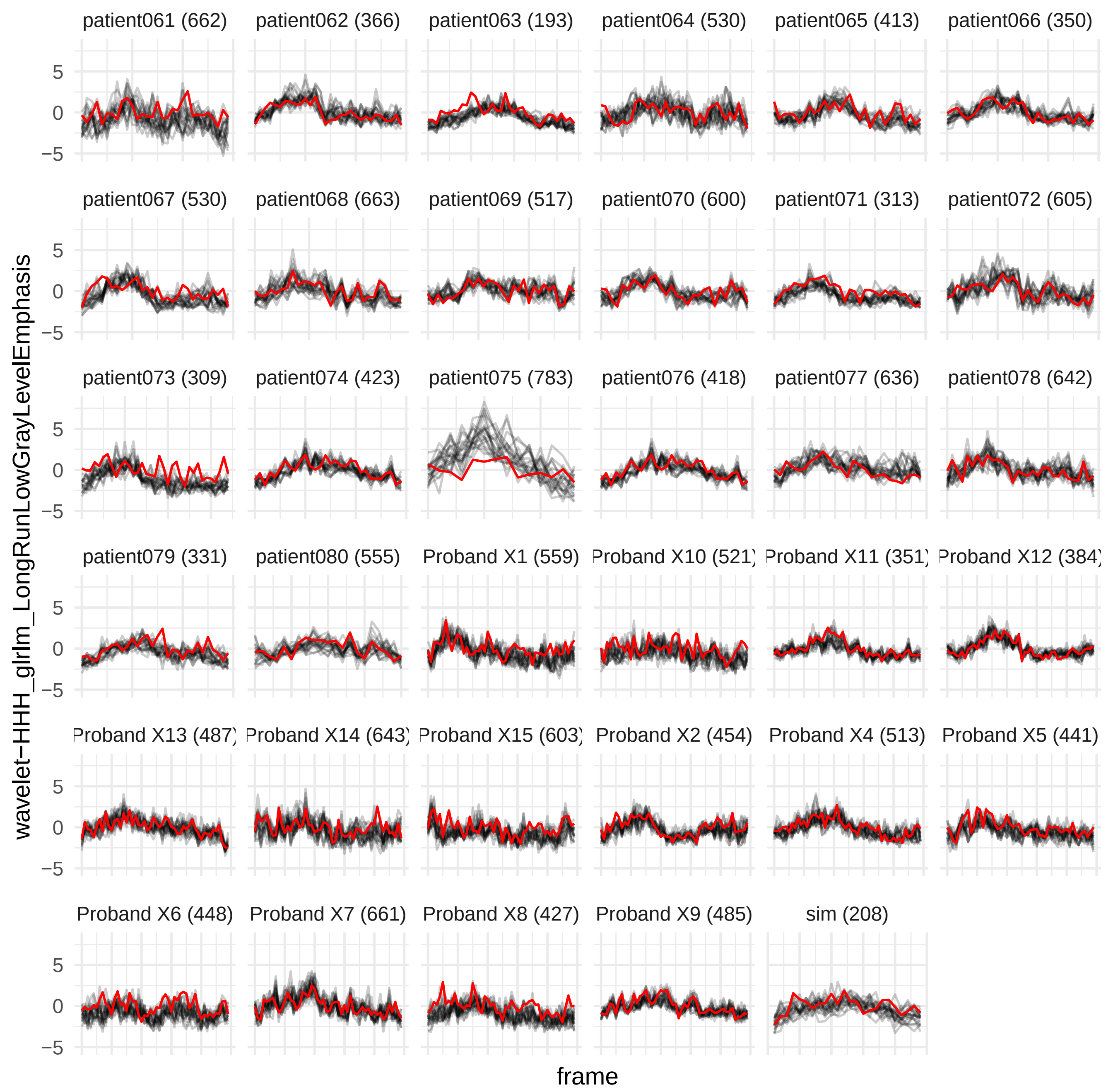
patient061 (662)
patient062 (366)
patient063 (193)
patient064 (530)
patient065 (413)
patient066 (350)
patient067 (530)
patient068 (663)
patient069 (517)
patient070 (600)
patient071 (313)
patient072 (605)
patient073 (309)
patient074 (423)
patient075 (783)
patient076 (418)
patient077 (636)
patient078 (642)
patient079 (331)
patient080 (555)
Proband X1 (559)
Proband X10 (521)
Proband X11 (351)
Proband X12 (384)
Proband X13 (487)
Proband X14 (643)
Proband X15 (603)
Proband X2 (454)
Proband X4 (513)
Proband X5 (441)
Proband X6 (448)
Proband X7 (661)
Proband X8 (427)
Proband X9 (485)
sim (208)
wavelet−HHH_glrlm_LongRunLowGrayLevelEmphasis
frame
5
0
−5

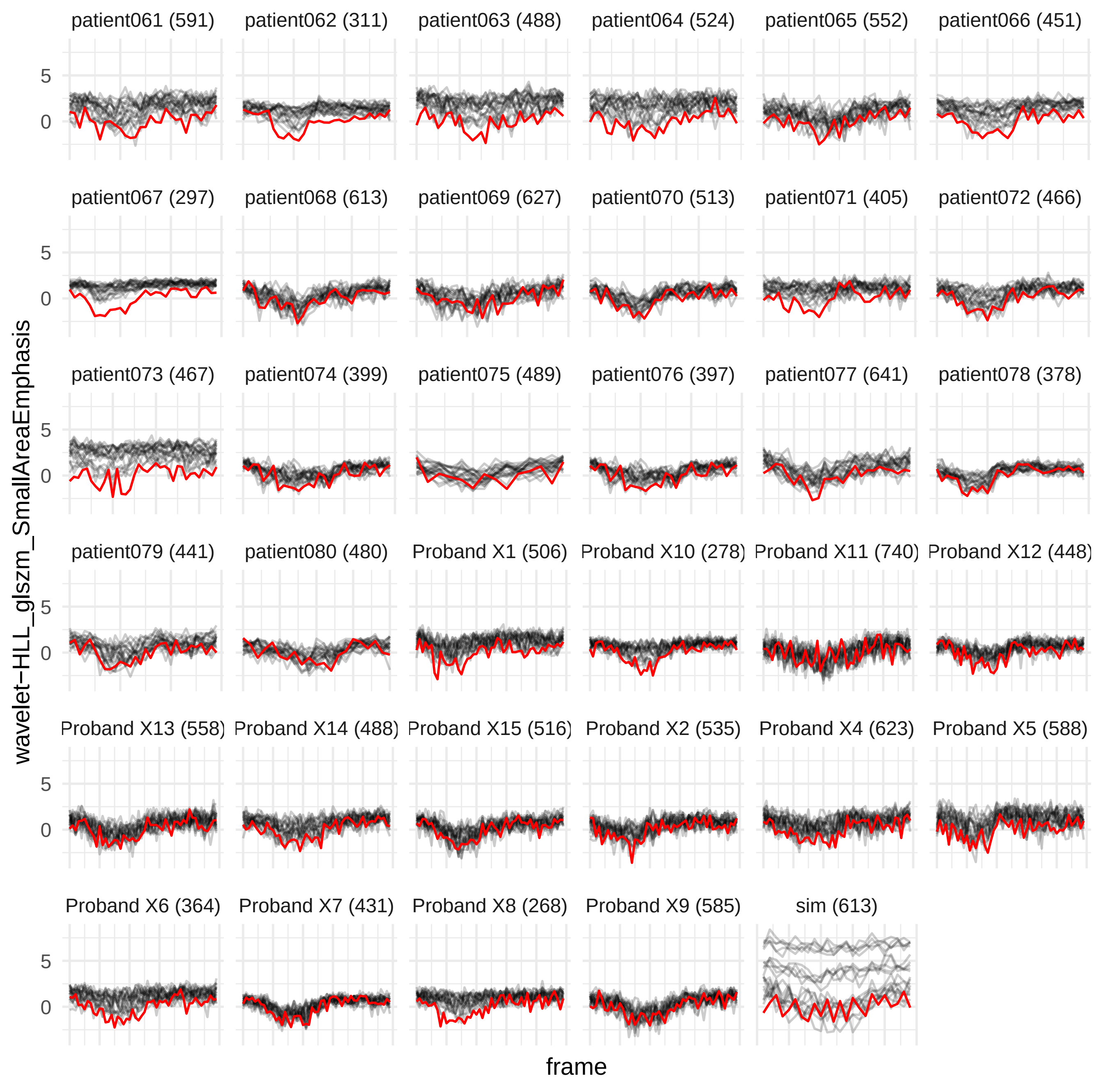

patient061 (591)
patient062 (311)
patient063 (488)
patient064 (524)
patient065 (552)
patient066 (451)
patient067 (297)
patient068 (613)
patient069 (627)
patient070 (513)
patient071 (405)
patient072 (466)
patient073 (467)
patient074 (399)
patient075 (489)
patient076 (397)
patient077 (641)
patient078 (378)
patient079 (441)
patient080 (480)
Proband X1 (506)
Proband X10 (278)
Proband X11 (740)
Proband X12 (448)
Proband X13 (558)
Proband X14 (488)
Proband X15 (516)
Proband X2 (535)
Proband X4 (623)
Proband X5 (588)
Proband X6 (364)
Proband X7 (431)
Proband X8 (268)
Proband X9 (585)
sim (613)
5
0
wavelet−HLL_glszm_SmallAreaEmphasis
frame

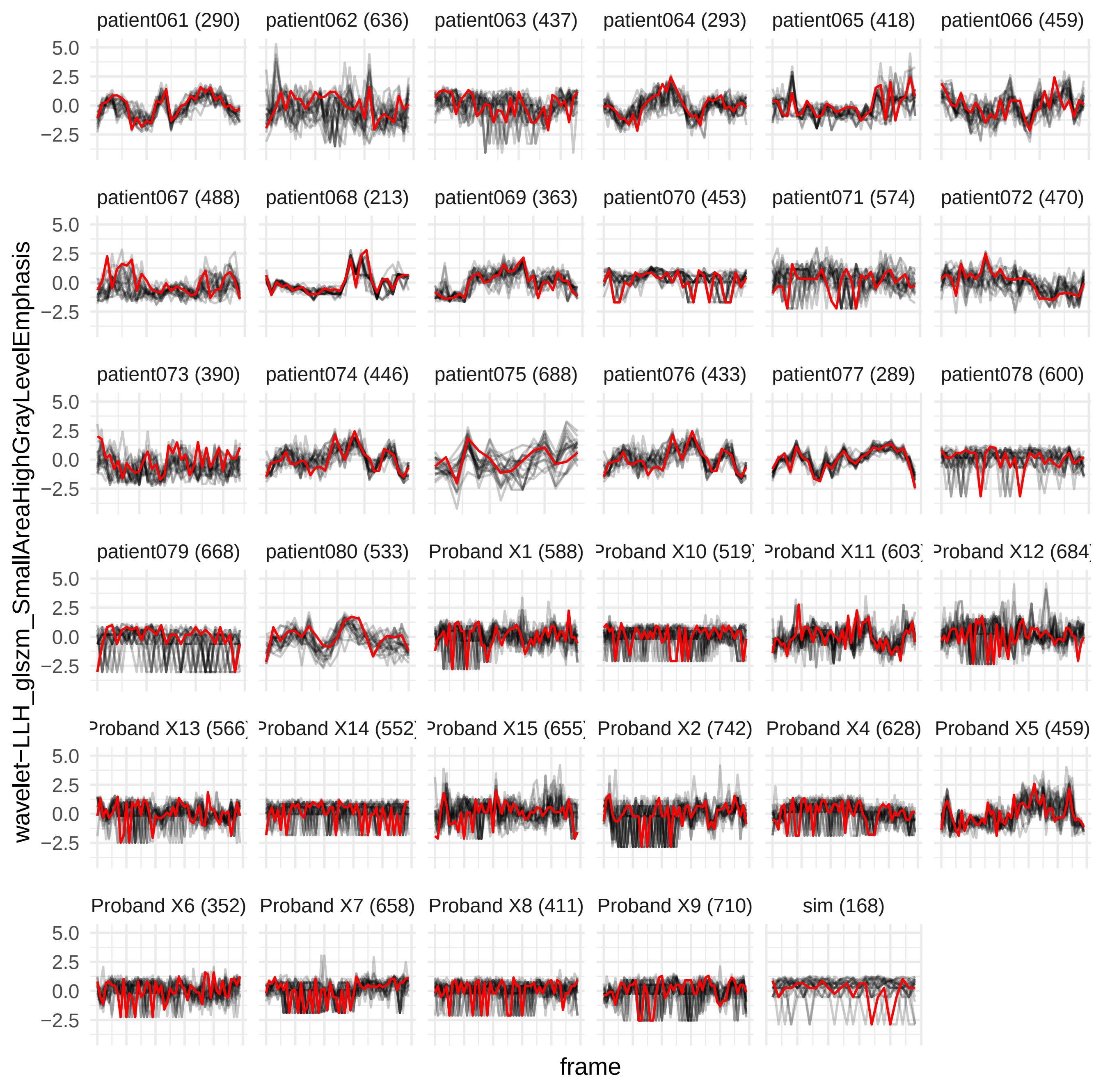
patient061 (290)
patient062 (636)
patient063 (437)
patient064 (293)
patient065 (418)
patient066 (459)
patient067 (488)
patient068 (213)
patient069 (363)
patient070 (453)
patient071 (574)
patient072 (470)
patient073 (390)
patient074 (446)
patient075 (688)
patient076 (433)
patient077 (289)
patient078 (600)
patient079 (668)
patient080 (533)
Proband X1 (588)
Proband X10 (519)
Proband X11 (603)
Proband X12 (684)
Proband X13 (566)
Proband X14 (552)
Proband X15 (655)
Proband X2 (742)
Proband X4 (628)
Proband X5 (459)
Proband X6 (352)
Proband X7 (658)
Proband X8 (411)
Proband X9 (710)
sim (168)
5.0
2.5
0.0
−2.5
wavelet−LLH_glszm_SmallAreaHighGrayLevelEmphasis
frame

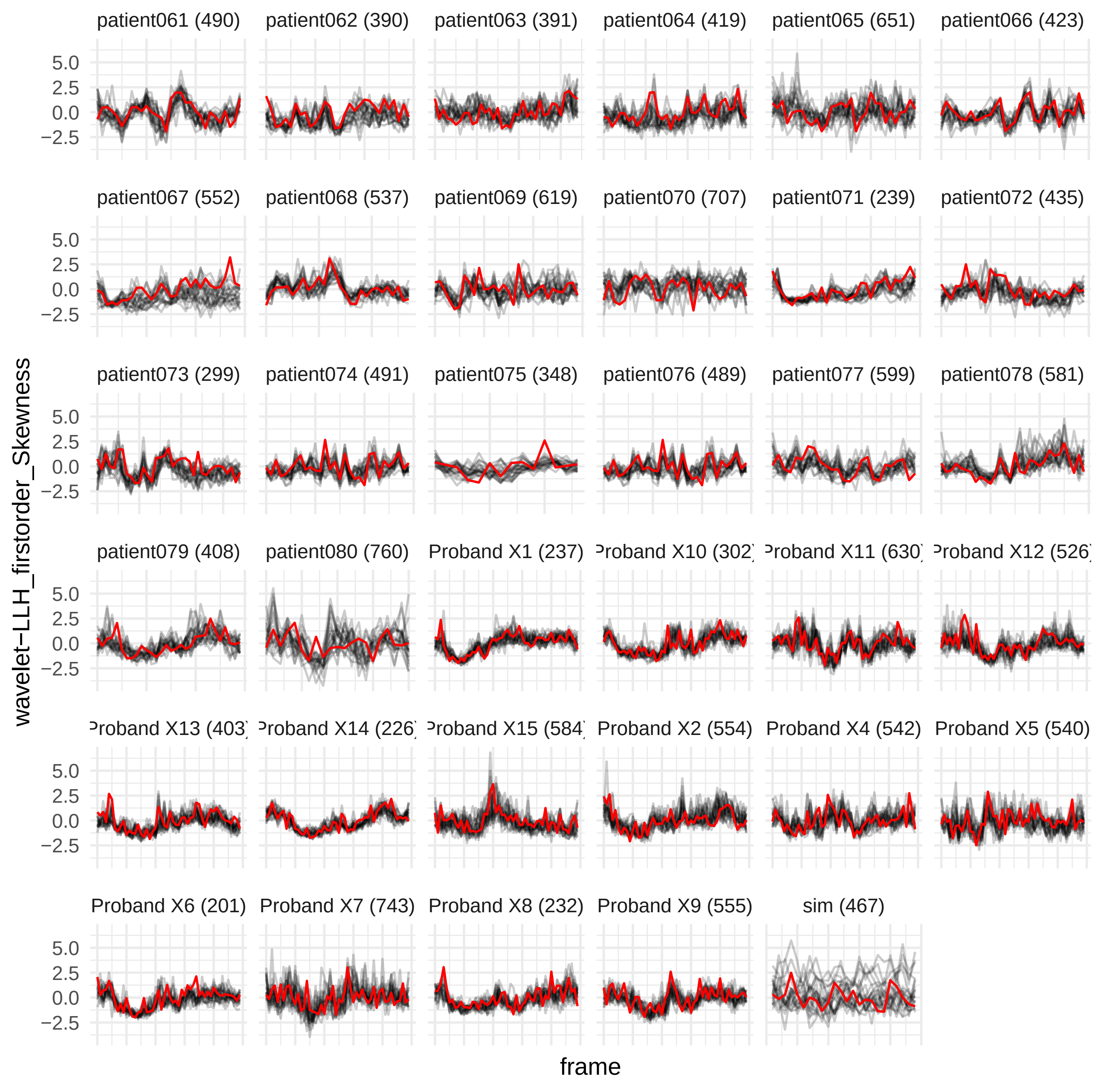

patient061 (490)
patient062 (390)
patient063 (391)
patient064 (419)
patient065 (651)
patient066 (423)
patient067 (552)
patient068 (537)
patient069 (619)
patient070 (707)
patient071 (239)
patient072 (435)
patient073 (299)
patient074 (491)
patient075 (348)
patient076 (489)
patient077 (599)
patient078 (581)
patient079 (408)
patient080 (760)
Proband X1 (237)
Proband X10 (302)
Proband X11 (630)
Proband X12 (526)
Proband X13 (403)
Proband X14 (226)
Proband X15 (584)
Proband X2 (554)
Proband X4 (542)
Proband X5 (540)
Proband X6 (201)
Proband X7 (743)
Proband X8 (232)
Proband X9 (555)
sim (467)
5.0
2.5
0.0
−2.5
wavelet−LLH_firstorder_Skewness
frame

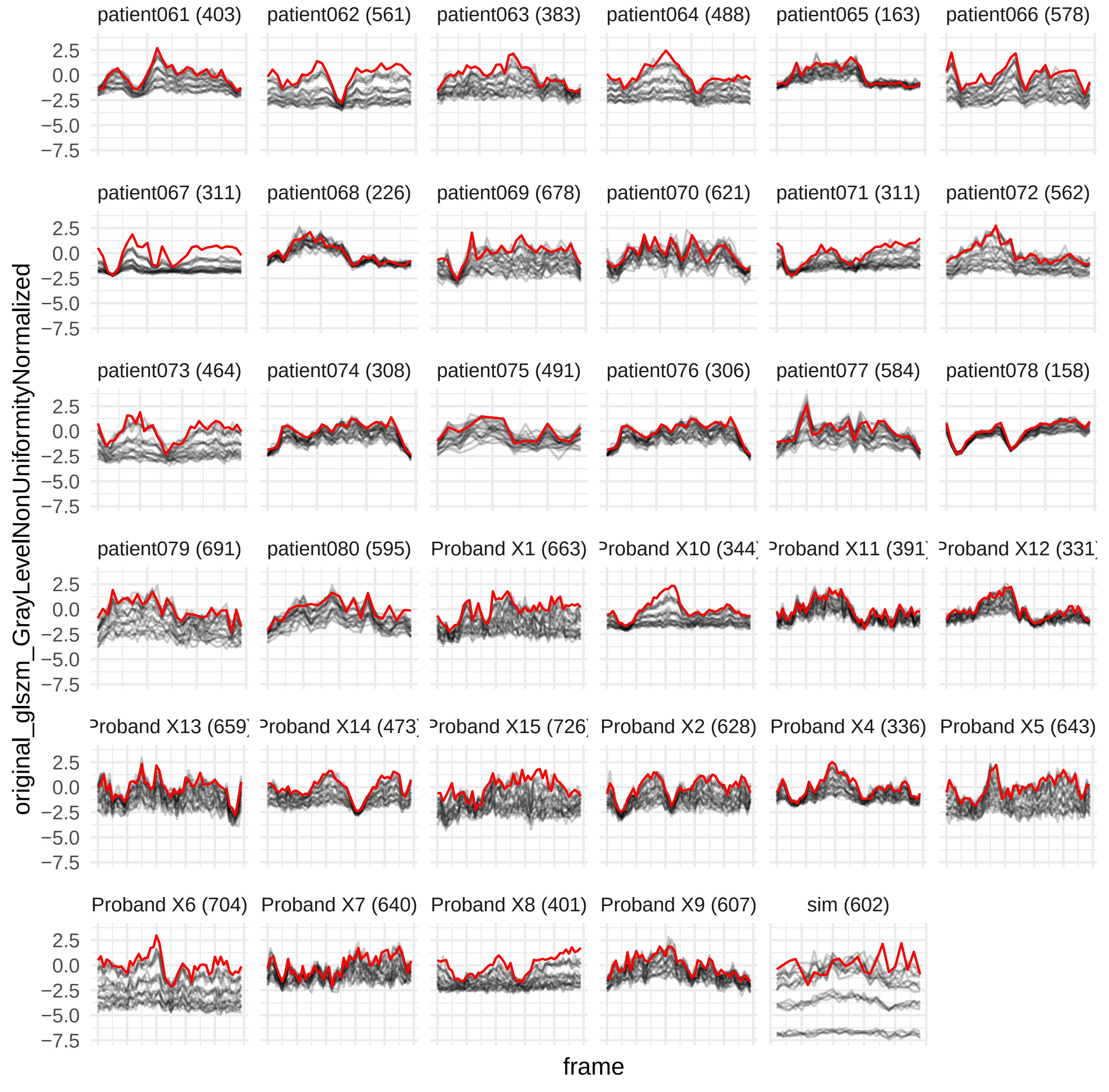
patient061 (403)
patient062 (561)
patient063 (383)
patient064 (488)
patient065 (163)
patient066 (578)
patient067 (311)
patient068 (226)
patient069 (678)
patient070 (621)
patient071 (311)
patient072 (562)
patient073 (464)
patient074 (308)
patient075 (491)
patient076 (306)
patient077 (584)
patient078 (158)
patient079 (691)
patient080 (595)
Proband X1 (663)
Proband X10 (344)
Proband X11 (391)
Proband X12 (331)
Proband X13 (659)
Proband X14 (473)
Proband X15 (726)
Proband X2 (628)
Proband X4 (336)
Proband X5 (643)
Proband X6 (704)
Proband X7 (640)
Proband X8 (401)
Proband X9 (607)
sim (602)
2.5
0.0
−2.5
−5.0
−7.5
original_glszm_GrayLevelNonUniformityNormalized
frame

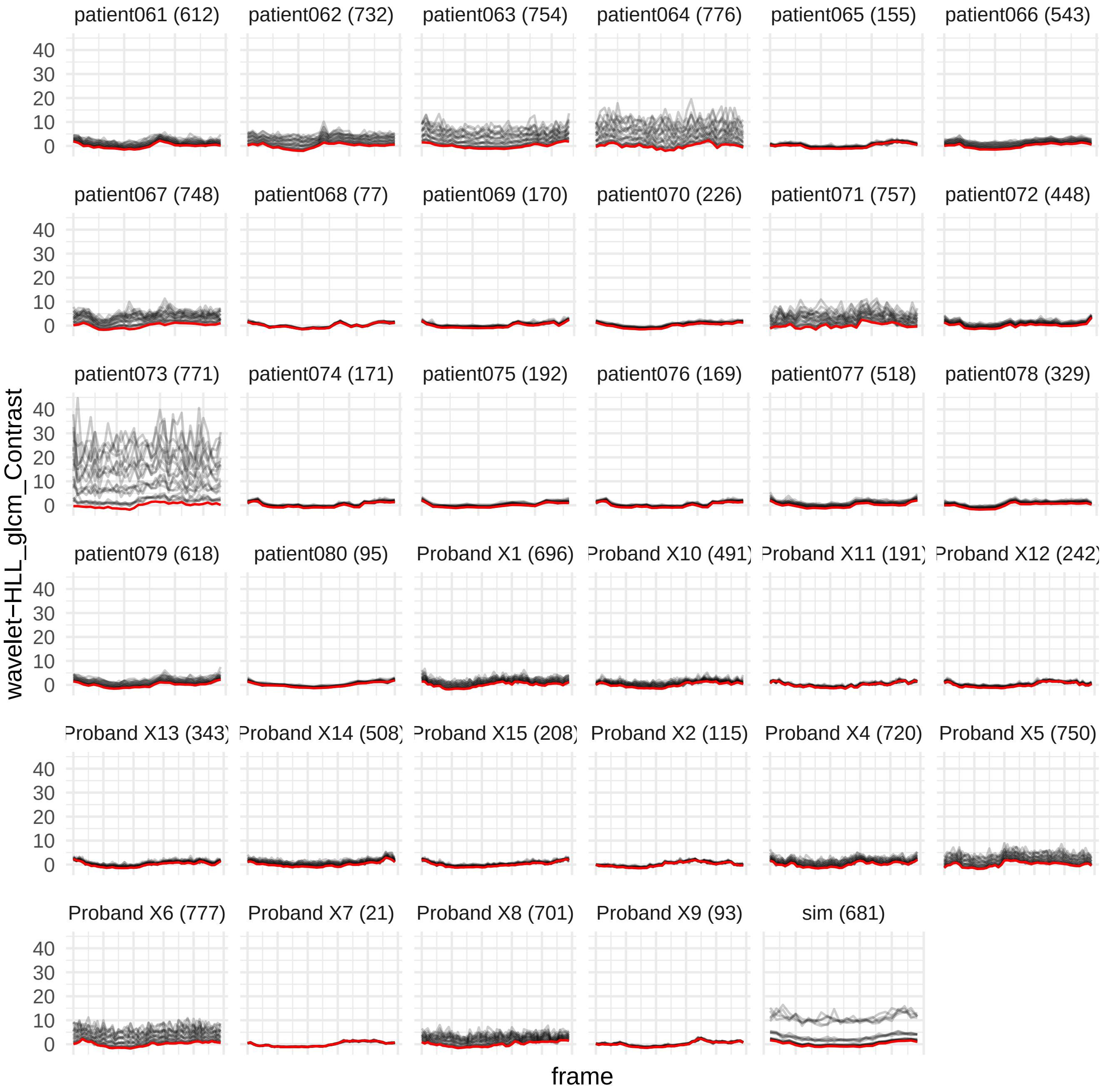
patient061 (612)
patient062 (732)
patient063 (754)
patient064 (776)
patient065 (155)
patient066 (543)
patient067 (748)
patient068 (77)
patient069 (170)
patient070 (226)
patient071 (757)
patient072 (448)
patient073 (771)
patient074 (171)
patient075 (192)
patient076 (169)
patient077 (518)
patient078 (329)
patient079 (618)
patient080 (95)
Proband X1 (696)
Proband X10 (491)
Proband X11 (191)
Proband X12 (242)
Proband X13 (343)
Proband X14 (508)
Proband X15 (208)
Proband X2 (115)
Proband X4 (720)
Proband X5 (750)
Proband X6 (777)
Proband X7 (21)
Proband X8 (701)
Proband X9 (93)
sim (681)
wavelet−HLL_glcm_Contrast
0
10
20
30
40
frame

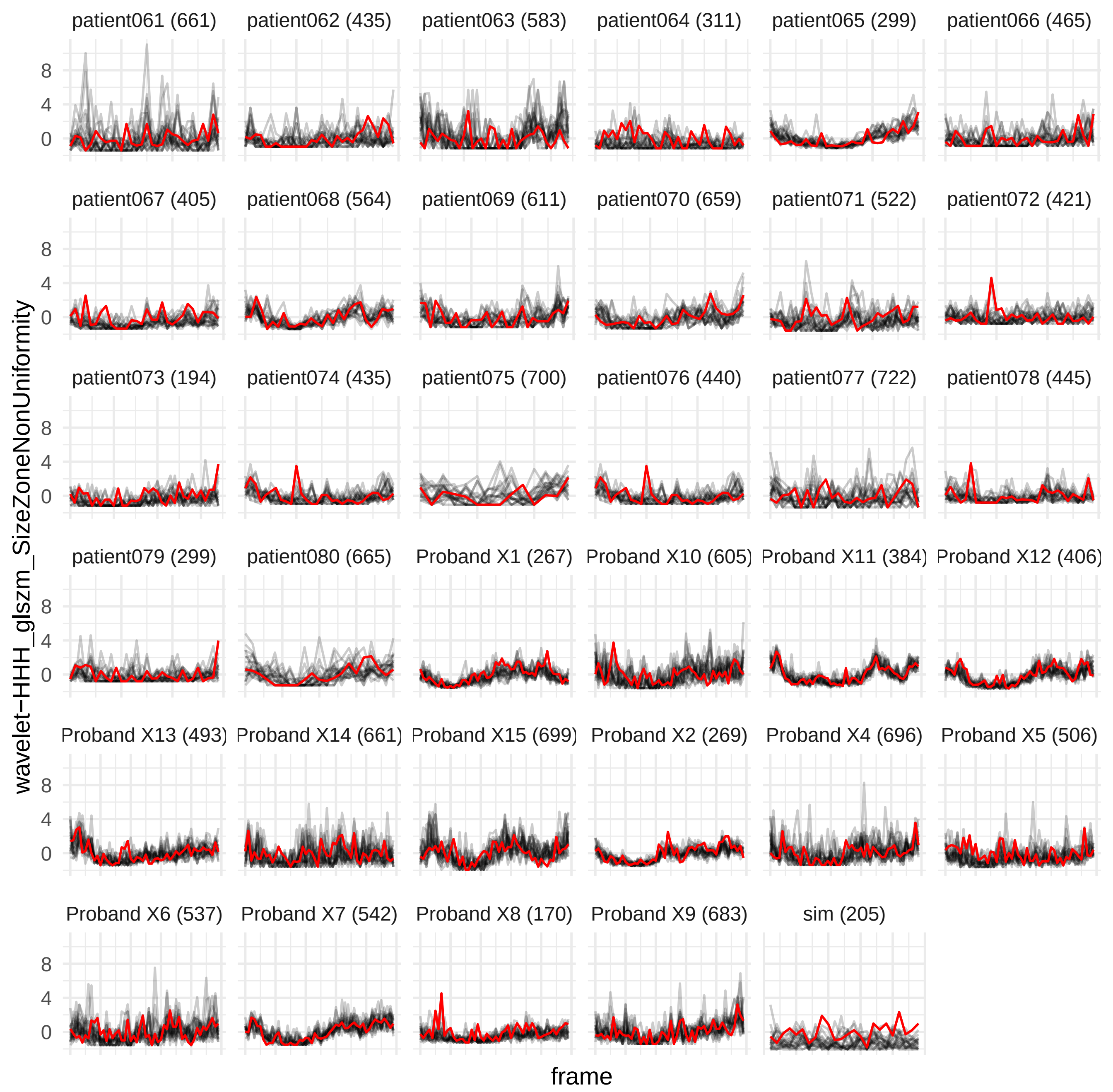
patient061 (661)
patient062 (435)
patient063 (583)
patient064 (311)
patient065 (299)
patient066 (465)
patient067 (405)
patient068 (564)
patient069 (611)
patient070 (659)
patient071 (522)
patient072 (421)
patient073 (194)
patient074 (435)
patient075 (700)
patient076 (440)
patient077 (722)
patient078 (445)
patient079 (299)
patient080 (665)
Proband X1 (267)
Proband X10 (605)
Proband X11 (384)
Proband X12 (406)
Proband X13 (493)
Proband X14 (661)
Proband X15 (699)
Proband X2 (269)
Proband X4 (696)
Proband X5 (506)
Proband X6 (537)
Proband X7 (542)
Proband X8 (170)
Proband X9 (683)
sim (205)
wavelet−HHH_glszm_SizeZoneNonUniformity
frame
8
4
0

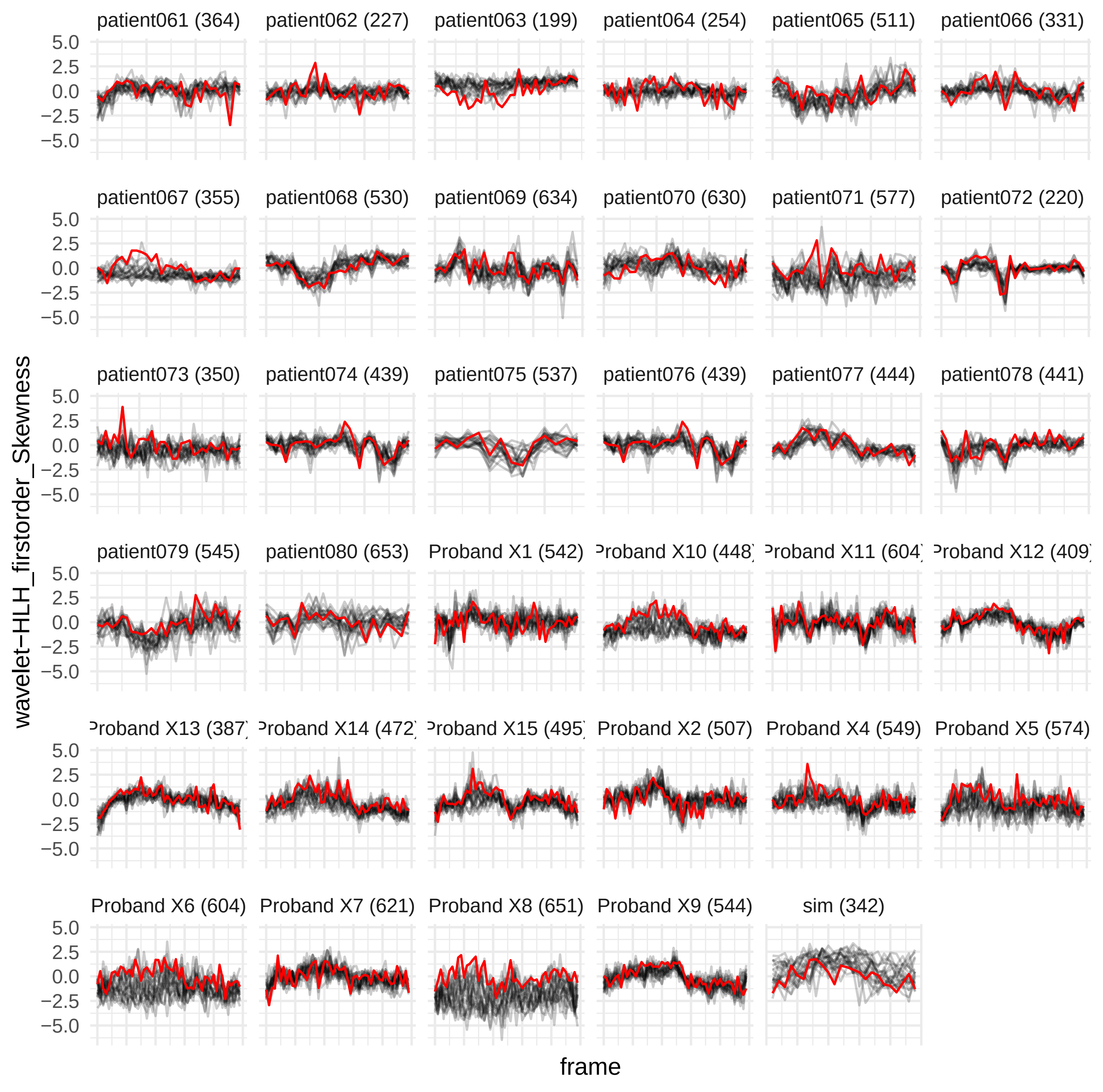

patient061 (364)
patient062 (227)
patient063 (199)
patient064 (254)
patient065 (511)
patient066 (331)
patient067 (355)
patient068 (530)
patient069 (634)
patient070 (630)
patient071 (577)
patient072 (220)
patient073 (350)
patient074 (439)
patient075 (537)
patient076 (439)
patient077 (444)
patient078 (441)
patient079 (545)
patient080 (653)
Proband X1 (542)
Proband X10 (448)
Proband X11 (604)
Proband X12 (409)
Proband X13 (387)
Proband X14 (472)
Proband X15 (495)
Proband X2 (507)
Proband X4 (549)
Proband X5 (574)
Proband X6 (604)
Proband X7 (621)
Proband X8 (651)
Proband X9 (544)
sim (342)
5.0
2.5
0.0
−2.5
−5.0
wavelet−HLH_firstorder_Skewness
frame

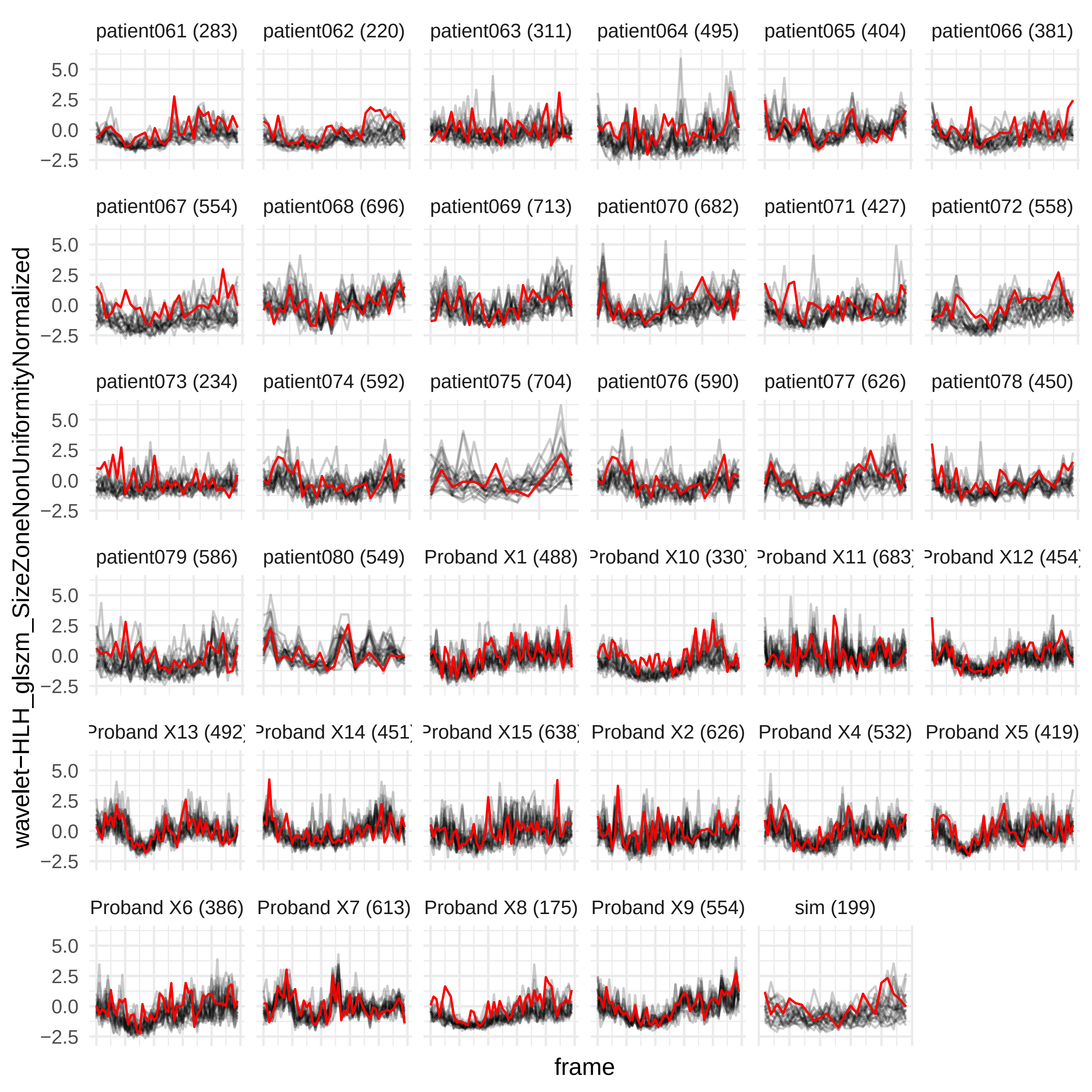
patient061 (283)
patient062 (220)
patient063 (311)
patient064 (495)
patient065 (404)
patient066 (381)
patient067 (554)
patient068 (696)
patient069 (713)
patient070 (682)
patient071 (427)
patient072 (558)
patient073 (234)
patient074 (592)
patient075 (704)
patient076 (590)
patient077 (626)
patient078 (450)
patient079 (586)
patient080 (549)
Proband X1 (488)
Proband X10 (330)
Proband X11 (683)
Proband X12 (454)
Proband X13 (492)
Proband X14 (451)
Proband X15 (638)
Proband X2 (626)
Proband X4 (532)
Proband X5 (419)
Proband X6 (386)
Proband X7 (613)
Proband X8 (175)
Proband X9 (554)
sim (199)
5.0
2.5
0.0
−2.5
wavelet−HLH_glszm_SizeZoneNonUniformityNormalized
frame

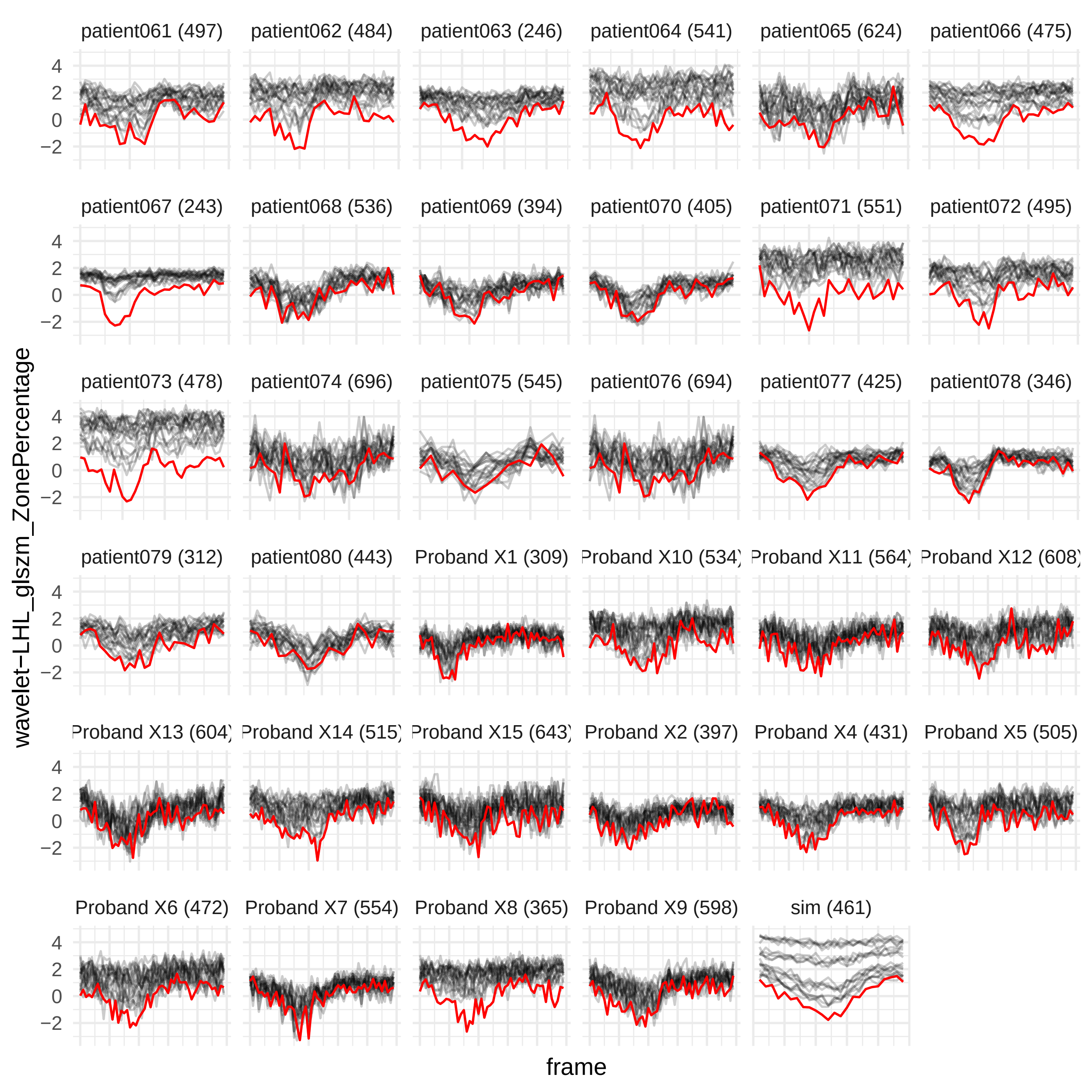
patient061 (497)
patient062 (484)
patient063 (246)
patient064 (541)
patient065 (624)
patient066 (475)
patient067 (243)
patient068 (536)
patient069 (394)
patient070 (405)
patient071 (551)
patient072 (495)
patient073 (478)
patient074 (696)
patient075 (545)
patient076 (694)
patient077 (425)
patient078 (346)
patient079 (312)
patient080 (443)
Proband X1 (309)
Proband X10 (534)
Proband X11 (564)
Proband X12 (608)
Proband X13 (604)
Proband X14 (515)
Proband X15 (643)
Proband X2 (397)
Proband X4 (431)
Proband X5 (505)
Proband X6 (472)
Proband X7 (554)
Proband X8 (365)
Proband X9 (598)
sim (461)
4
2
0
−2
wavelet−LHL_glszm_ZonePercentage
frame

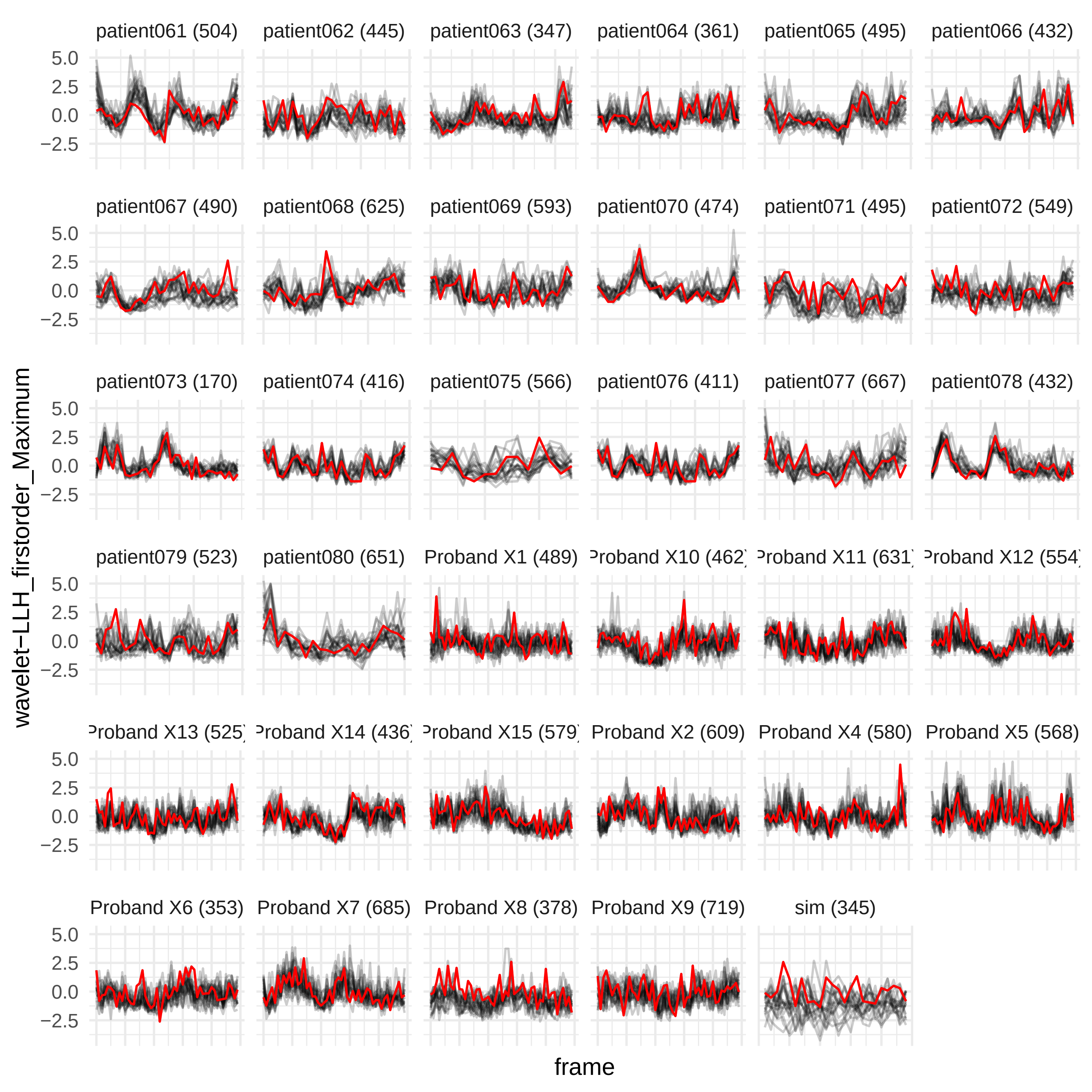

patient061 (504)
patient062 (445)
patient063 (347)
patient064 (361)
patient065 (495)
patient066 (432)
patient067 (490)
patient068 (625)
patient069 (593)
patient070 (474)
patient071 (495)
patient072 (549)
patient073 (170)
patient074 (416)
patient075 (566)
patient076 (411)
patient077 (667)
patient078 (432)
patient079 (523)
patient080 (651)
Proband X1 (489)
Proband X10 (462
Proband X11 (631
Proband X12 (554
Proband X13 (525
Proband X14 (436
Proband X15 (579
Proband X2 (609)
Proband X4 (580)
Proband X5 (568)
Proband X6 (353)
Proband X7 (685)
Proband X8 (378)
Proband X9 (719)
sim (345)
5.0
2.5
0.0
−2.5
wavelet−LLH_firstorder_Maximum
frame

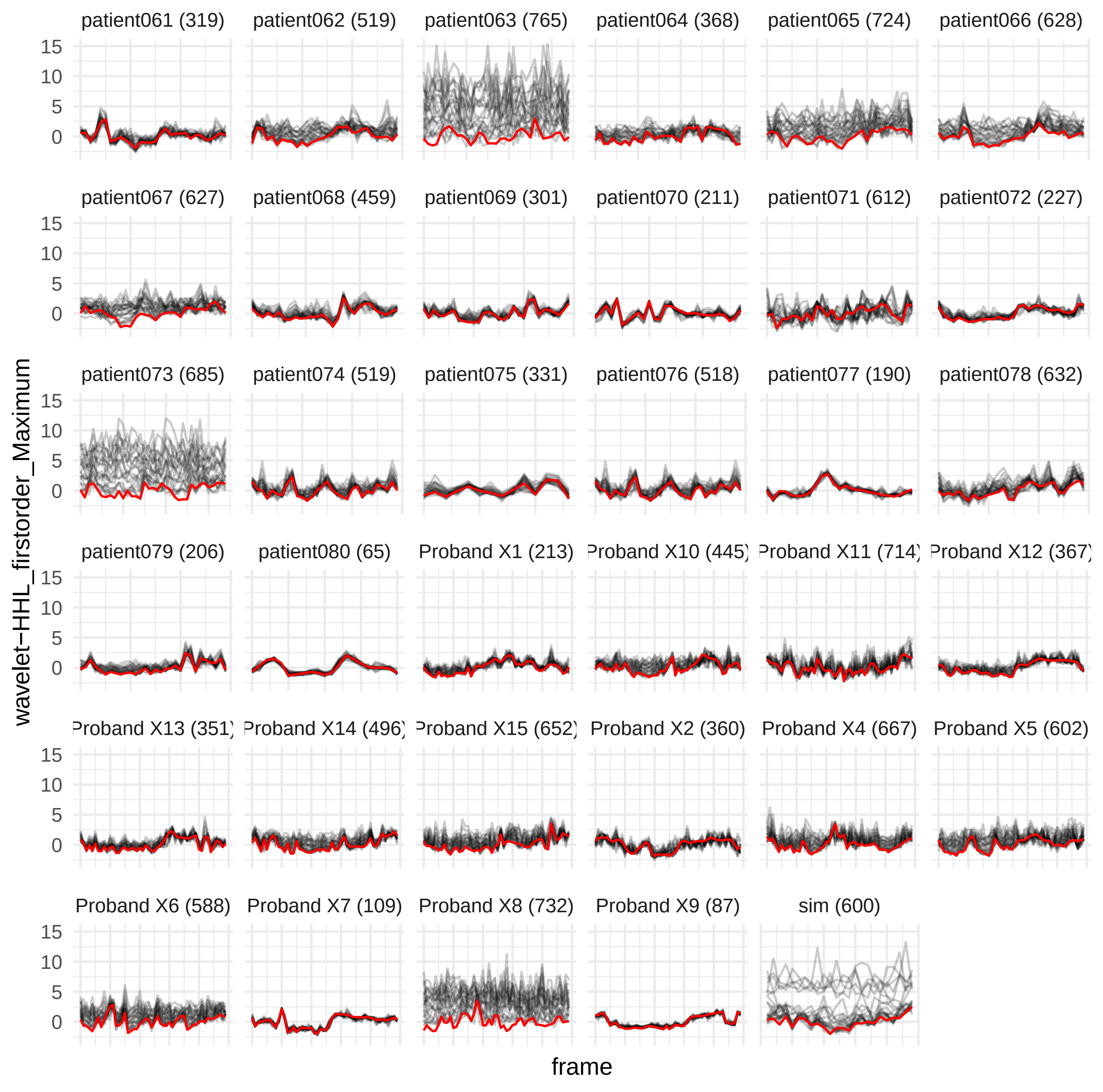
patient061 (319)
patient062 (519)
patient063 (765)
patient064 (368)
patient065 (724)
patient066 (628)
patient067 (627)
patient068 (459)
patient069 (301)
patient070 (211)
patient071 (612)
patient072 (227)
patient073 (685)
patient074 (519)
patient075 (331)
patient076 (518)
patient077 (190)
patient078 (632)
patient079 (206)
patient080 (65)
Proband X1 (213)
Proband X10 (445)
Proband X11 (714)
Proband X12 (367)
Proband X13 (351)
Proband X14 (496)
Proband X15 (652)
Proband X2 (360)
Proband X4 (667)
Proband X5 (602)
Proband X6 (588)
Proband X7 (109)
Proband X8 (732)
Proband X9 (87)
sim (600)
15
10
5
0
wavelet−HHL_firstorder_Maximum
frame

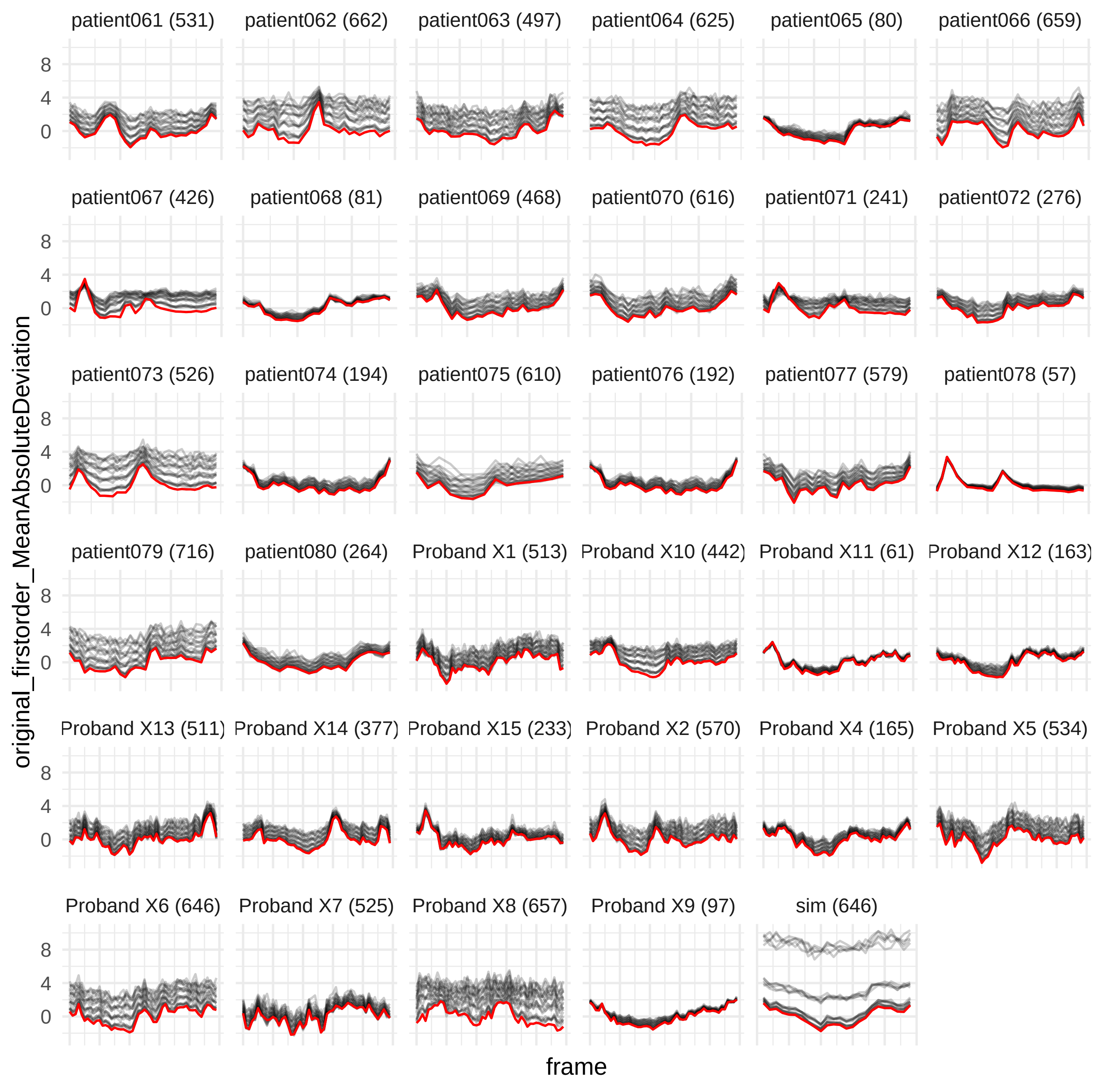

patient061 (531)
patient062 (662)
patient063 (497)
patient064 (625)
patient065 (80)
patient066 (659)
patient067 (426)
patient068 (81)
patient069 (468)
patient070 (616)
patient071 (241)
patient072 (276)
patient073 (526)
patient074 (194)
patient075 (610)
patient076 (192)
patient077 (579)
patient078 (57)
patient079 (716)
patient080 (264)
Proband X1 (513)
Proband X10 (442)
Proband X11 (61)
Proband X12 (163)
Proband X13 (511)
Proband X14 (377)
Proband X15 (233)
Proband X2 (570)
Proband X4 (165)
Proband X5 (534)
Proband X6 (646)
Proband X7 (525)
Proband X8 (657)
Proband X9 (97)
sim (646)
original_firstorder_MeanAbsoluteDeviation
0
4
8
frame

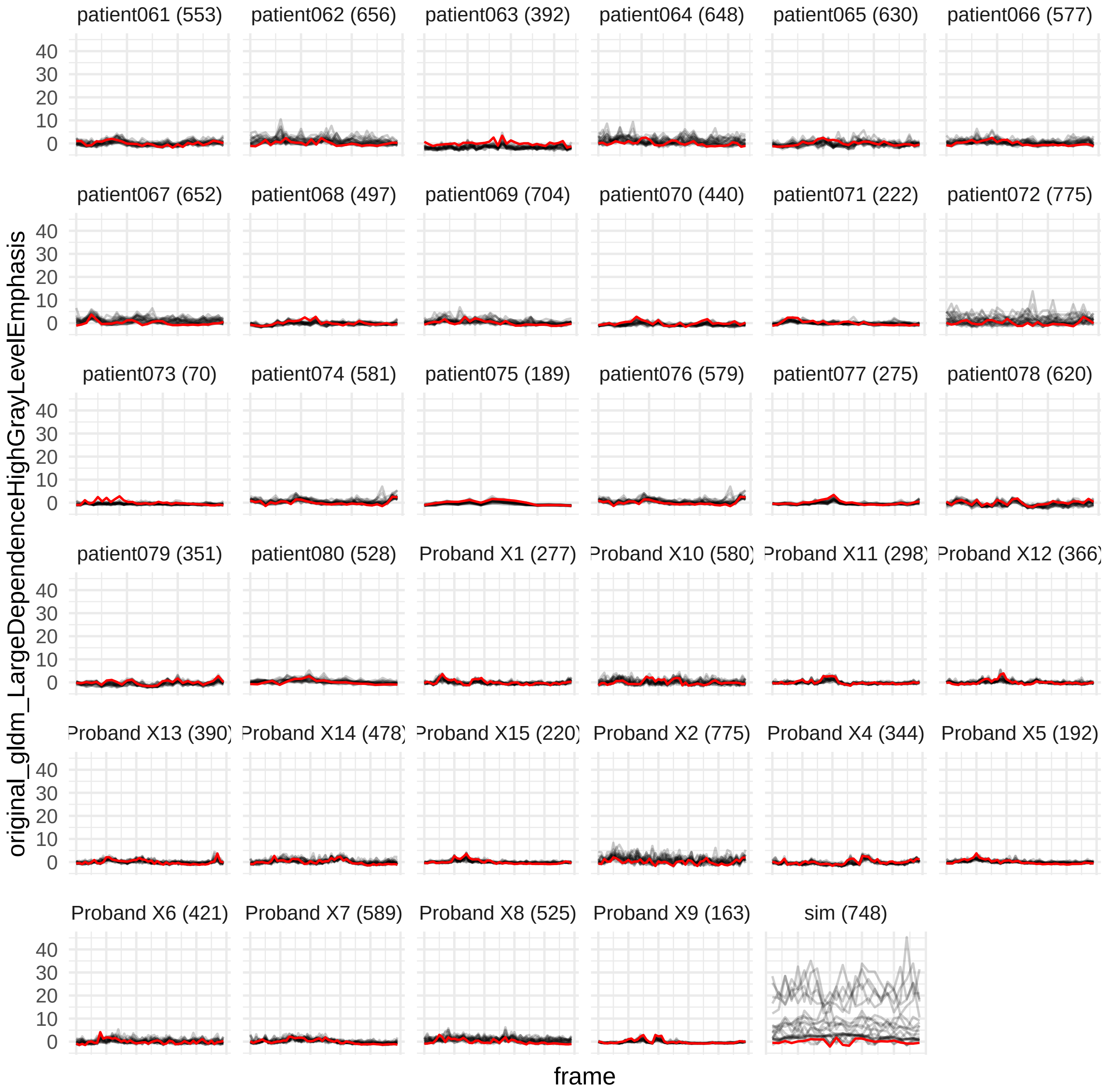

patient061 (553)
patient062 (656)
patient063 (392)
patient064 (648)
patient065 (630)
patient066 (577)
patient067 (652)
patient068 (497)
patient069 (704)
patient070 (440)
patient071 (222)
patient072 (775)
patient073 (70)
patient074 (581)
patient075 (189)
patient076 (579)
patient077 (275)
patient078 (620)
patient079 (351)
patient080 (528)
Proband X1 (277)
Proband X10 (580)
Proband X11 (298)
Proband X12 (366)
Proband X13 (390)
Proband X14 (478)
Proband X15 (220)
Proband X2 (775)
Proband X4 (344)
Proband X5 (192)
Proband X6 (421)
Proband X7 (589)
Proband X8 (525)
Proband X9 (163)
sim (748)
40
30
20
10
0
original_gldm_LargeDependenceHighGrayLevelEmphasis
frame

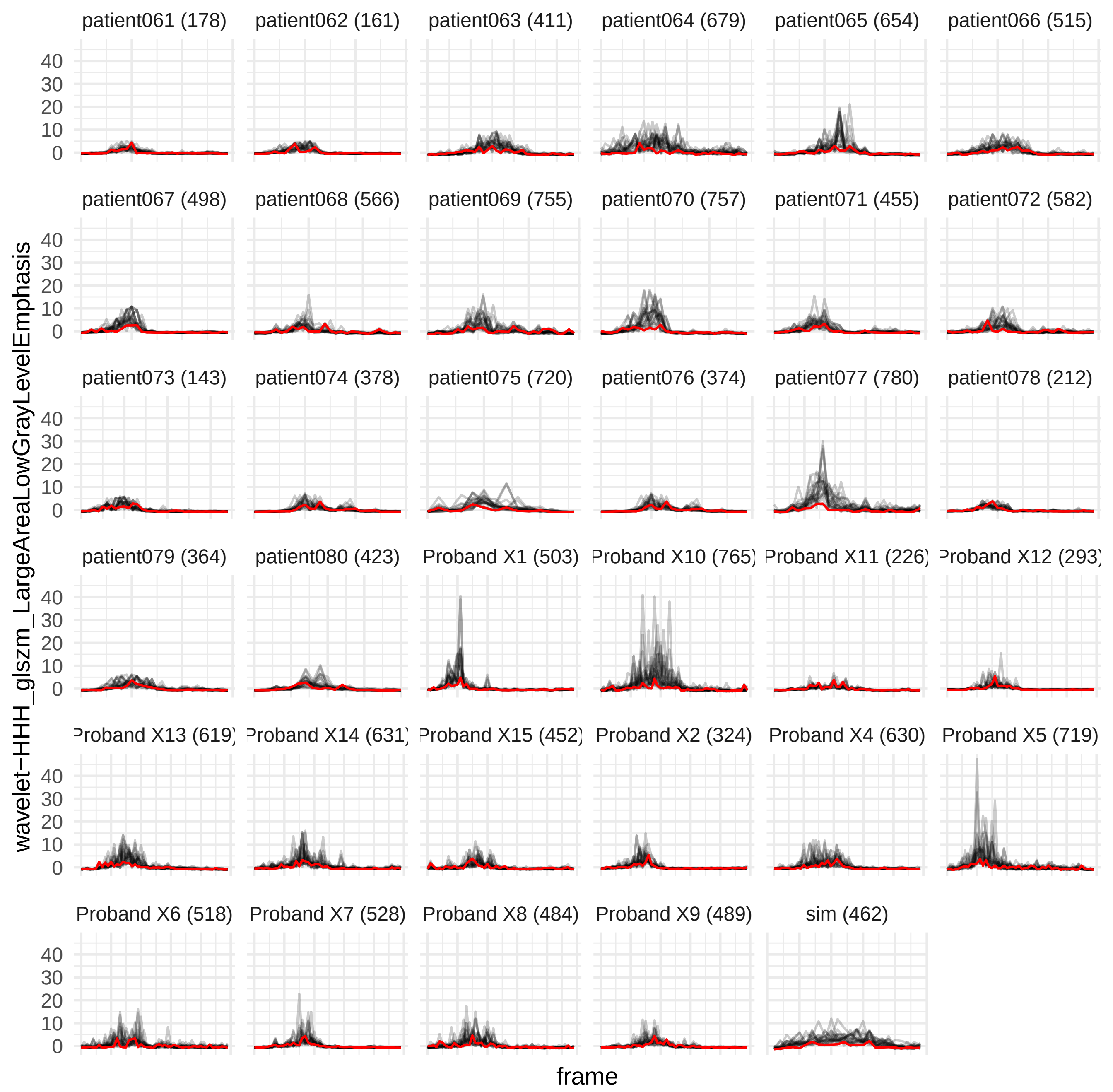

patient061 (178)
patient062 (161)
patient063 (411)
patient064 (679)
patient065 (654)
patient066 (515)
patient067 (498)
patient068 (566)
patient069 (755)
patient070 (757)
patient071 (455)
patient072 (582)
patient073 (143)
patient074 (378)
patient075 (720)
patient076 (374)
patient077 (780)
patient078 (212)
patient079 (364)
patient080 (423)
Proband X1 (503)
Proband X10 (765)
Proband X11 (226)
Proband X12 (293)
Proband X13 (619)
Proband X14 (631)
Proband X15 (452)
Proband X2 (324)
Proband X4 (630)
Proband X5 (719)
Proband X6 (518)
Proband X7 (528)
Proband X8 (484)
Proband X9 (489)
sim (462)
0
10
20
30
40
wavelet−HHH_glszm_LargeAreaLowGrayLevelEmphasis
frame

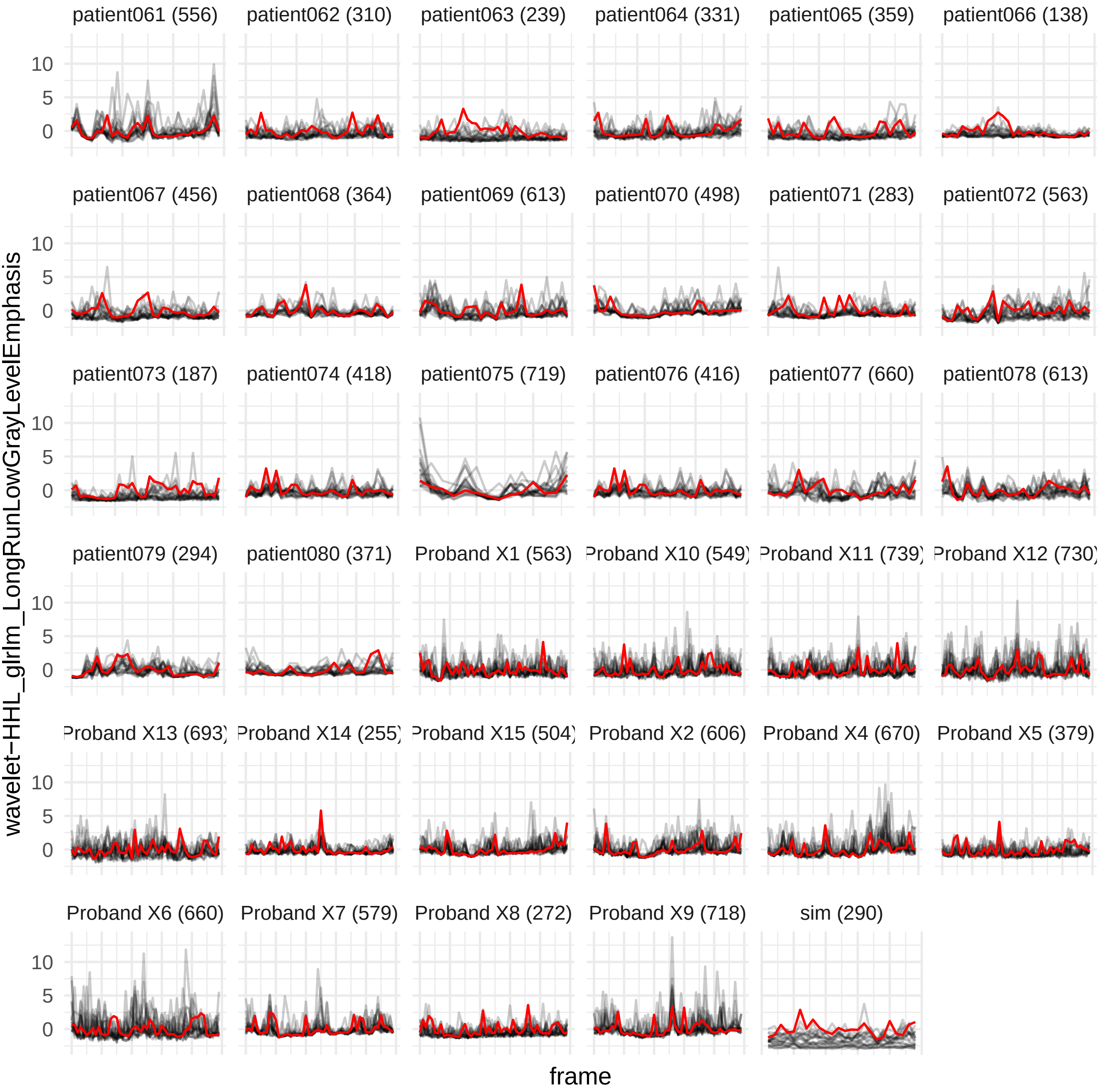

patient061 (556)
patient062 (310)
patient063 (239)
patient064 (331)
patient065 (359)
patient066 (138)
patient067 (456)
patient068 (364)
patient069 (613)
patient070 (498)
patient071 (283)
patient072 (563)
patient073 (187)
patient074 (418)
patient075 (719)
patient076 (416)
patient077 (660)
patient078 (613)
patient079 (294)
patient080 (371)
Proband X1 (563)
Proband X10 (549)
Proband X11 (739)
Proband X12 (730)
Proband X13 (693)
Proband X14 (255)
Proband X15 (504)
Proband X2 (606)
Proband X4 (670)
Proband X5 (379)
Proband X6 (660)
Proband X7 (579)
Proband X8 (272)
Proband X9 (718)
sim (290)
10
5
0
wavelet−HHL_glrlm_LongRunLowGrayLevelEmphasis
frame

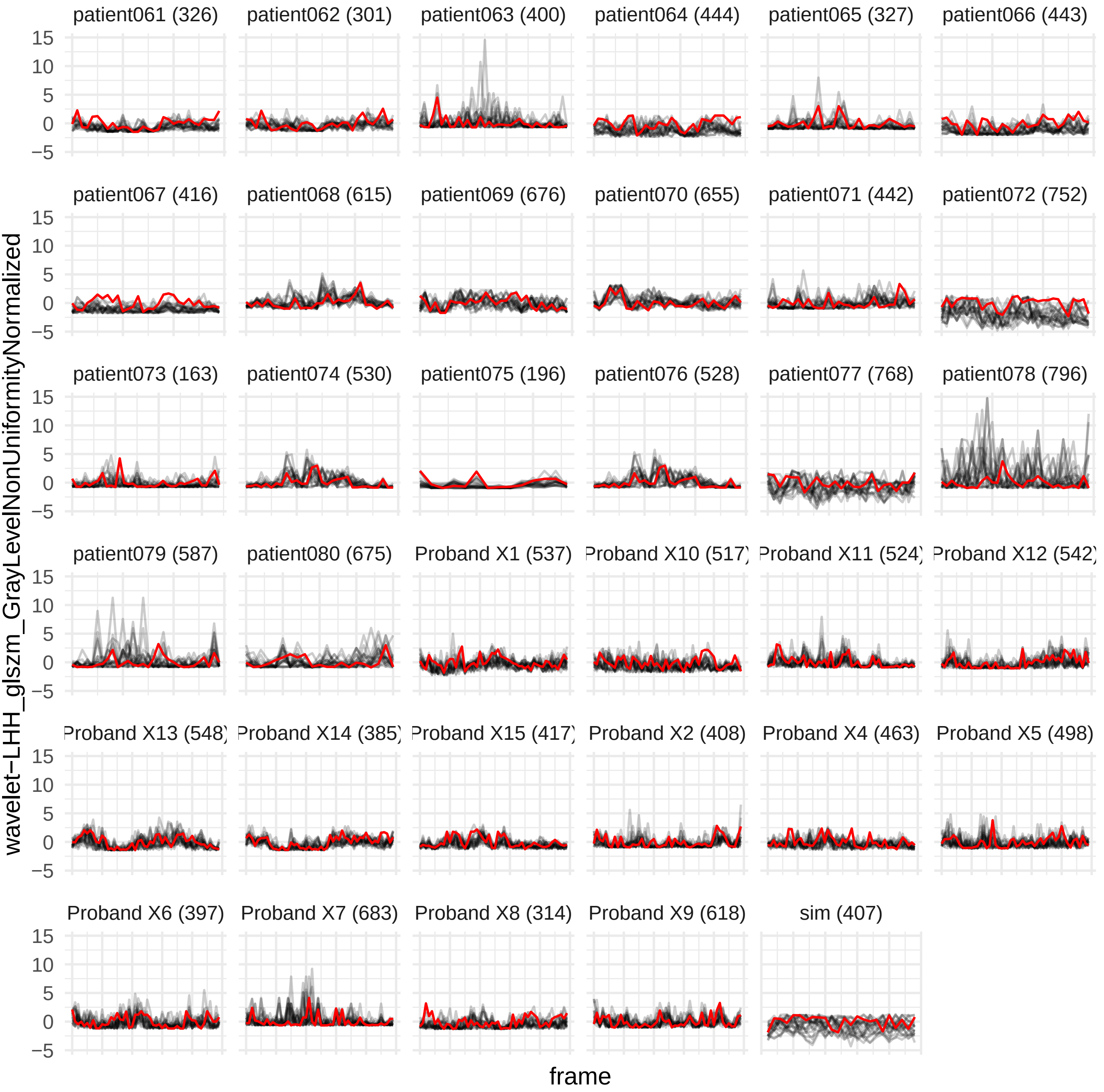

patient061 (326)
patient062 (301)
patient063 (400)
patient064 (444)
patient065 (327)
patient066 (443)
patient067 (416)
patient068 (615)
patient069 (676)
patient070 (655)
patient071 (442)
patient072 (752)
patient073 (163)
patient074 (530)
patient075 (196)
patient076 (528)
patient077 (768)
patient078 (796)
patient079 (587)
patient080 (675)
Proband X1 (537)
Proband X10 (517)
Proband X11 (524)
Proband X12 (542)
Proband X13 (548)
Proband X14 (385)
Proband X15 (417)
Proband X2 (408)
Proband X4 (463)
Proband X5 (498)
Proband X6 (397)
Proband X7 (683)
Proband X8 (314)
Proband X9 (618)
sim (407)
15
10
5
0
−5
wavelet−LHH_glszm_GrayLevelNonUniformityNormalized
frame

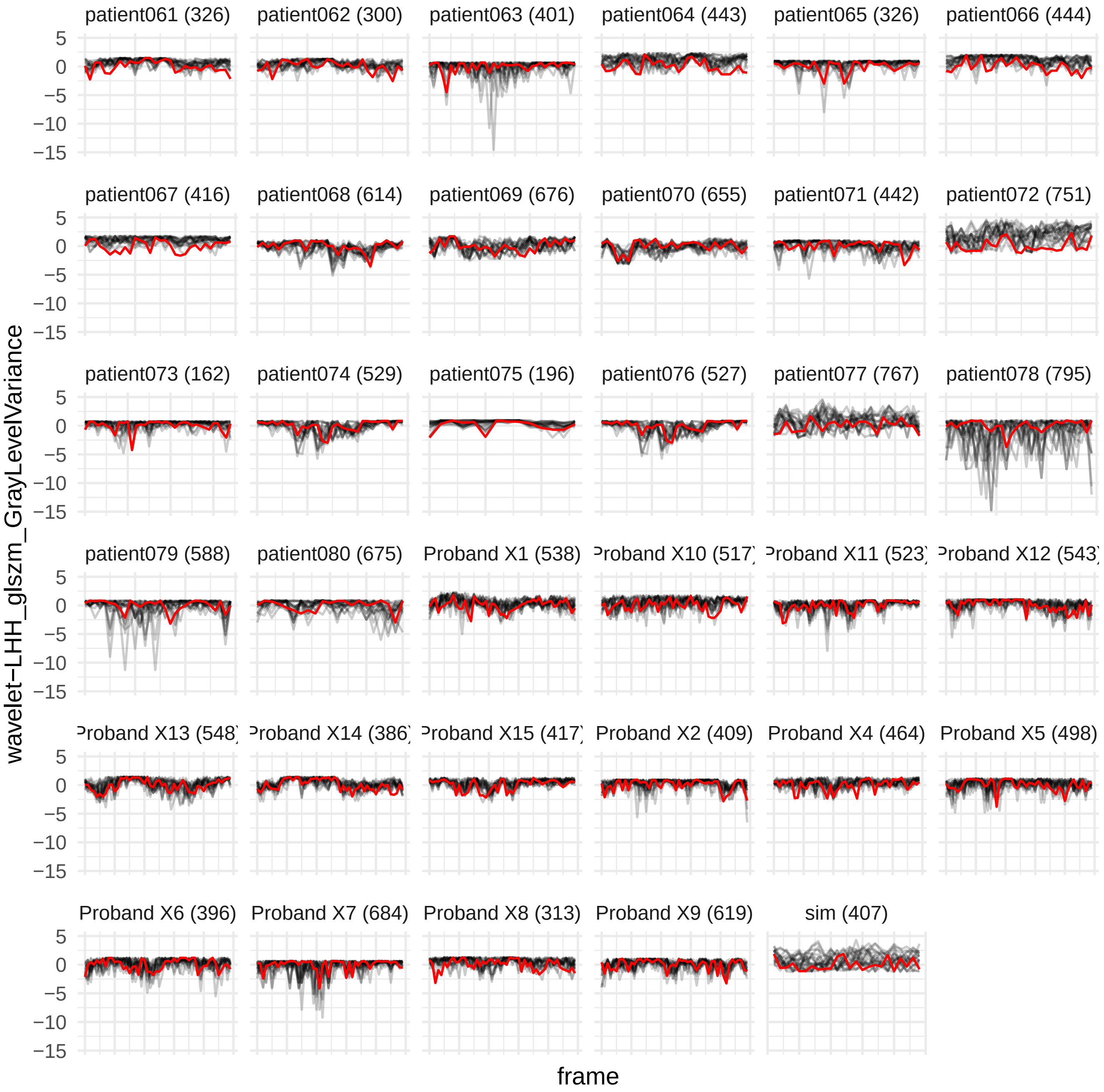

patient061 (326)
patient062 (300)
patient063 (401)
patient064 (443)
patient065 (326)
patient066 (444)
patient067 (416)
patient068 (614)
patient069 (676)
patient070 (655)
patient071 (442)
patient072 (751)
patient073 (162)
patient074 (529)
patient075 (196)
patient076 (527)
patient077 (767)
patient078 (795)
patient079 (588)
patient080 (675)
Proband X1 (538)
Proband X10 (517)
Proband X11 (523)
Proband X12 (543)
Proband X13 (548)
Proband X14 (386)
Proband X15 (417)
Proband X2 (409)
Proband X4 (464)
Proband X5 (498)
Proband X6 (396)
Proband X7 (684)
Proband X8 (313)
Proband X9 (619)
sim (407)
5
0
−5
−10
−15
wavelet−LHH_glszm_GrayLevelVariance
frame

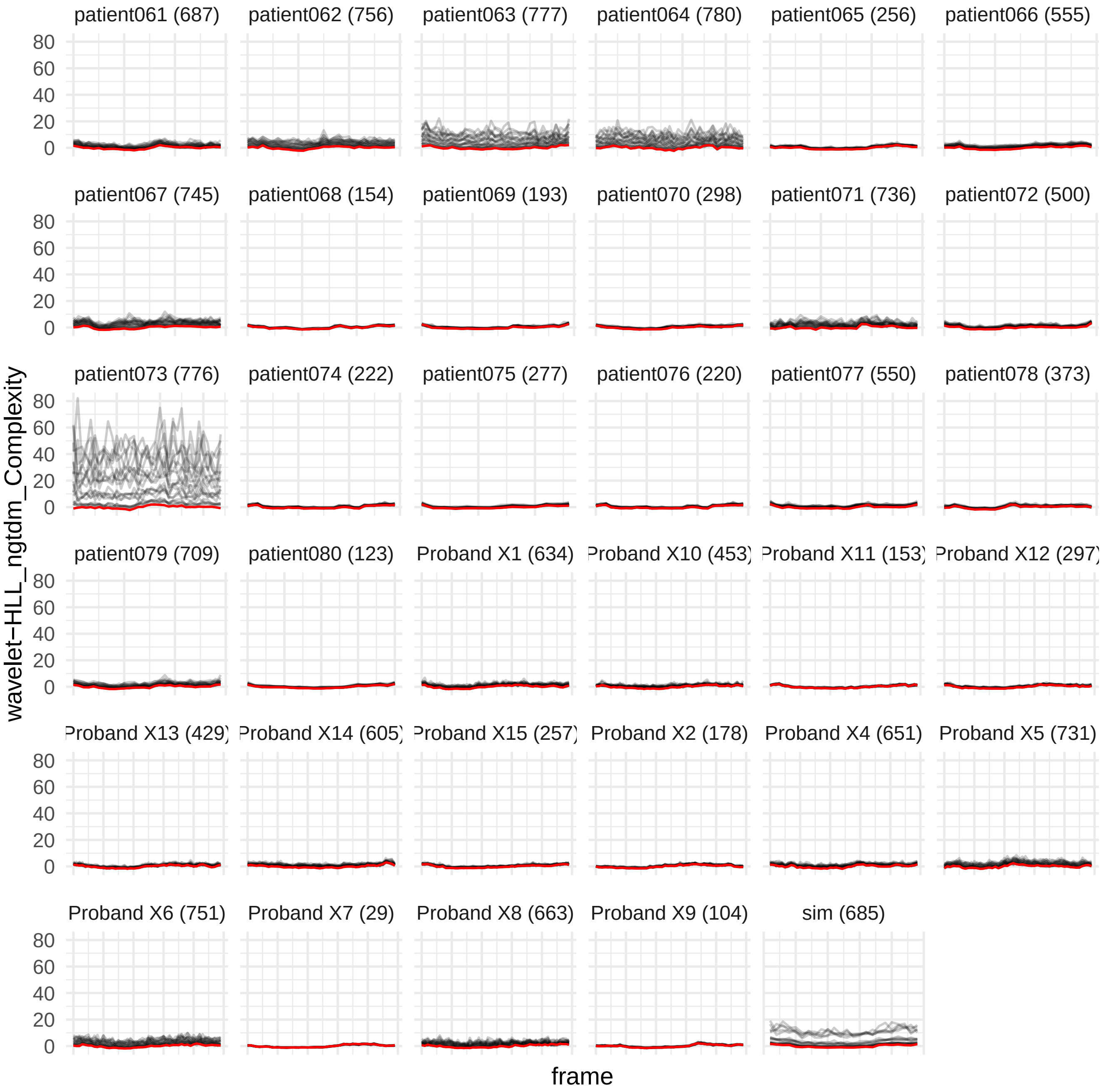

patient061 (687)
patient062 (756)
patient063 (777)
patient064 (780)
patient065 (256)
patient066 (555)
patient067 (745)
patient068 (154)
patient069 (193)
patient070 (298)
patient071 (736)
patient072 (500)
patient073 (776)
patient074 (222)
patient075 (277)
patient076 (220)
patient077 (550)
patient078 (373)
patient079 (709)
patient080 (123)
Proband X1 (634)
Proband X10 (453)
Proband X11 (153)
Proband X12 (297)
Proband X13 (429)
Proband X14 (605)
Proband X15 (257)
Proband X2 (178)
Proband X4 (651)
Proband X5 (731)
Proband X6 (751)
Proband X7 (29)
Proband X8 (663)
Proband X9 (104)
sim (685)
80
60
40
20
0
wavelet−HLL_ngtdm_Complexity
frame

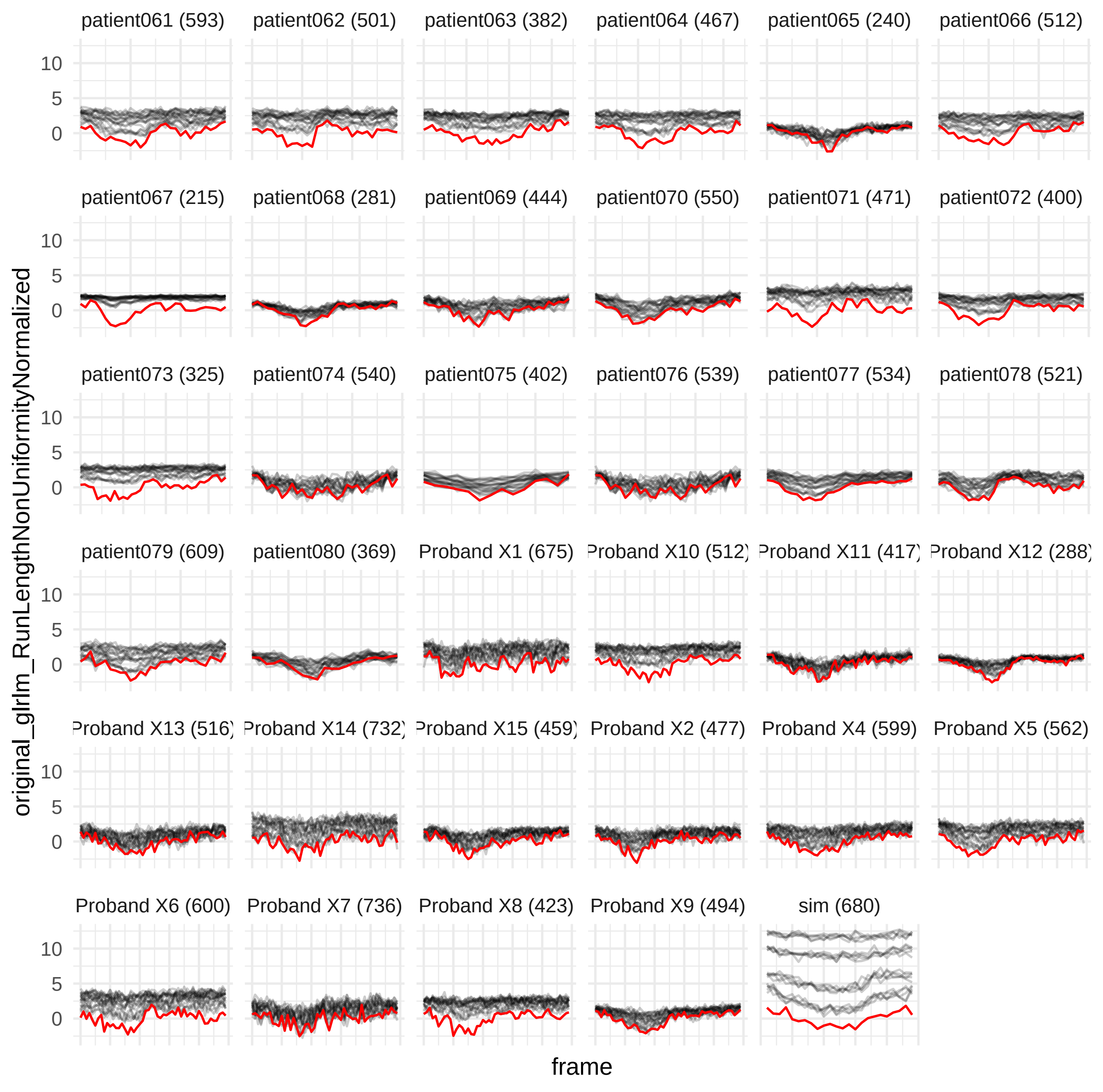
patient061 (593)
patient062 (501)
patient063 (382)
patient064 (467)
patient065 (240)
patient066 (512)
patient067 (215)
patient068 (281)
patient069 (444)
patient070 (550)
patient071 (471)
patient072 (400)
patient073 (325)
patient074 (540)
patient075 (402)
patient076 (539)
patient077 (534)
patient078 (521)
patient079 (609)
patient080 (369)
Proband X1 (675)
Proband X10 (512)
Proband X11 (417)
Proband X12 (288)
Proband X13 (516)
Proband X14 (732)
Proband X15 (459)
Proband X2 (477)
Proband X4 (599)
Proband X5 (562)
Proband X6 (600)
Proband X7 (736)
Proband X8 (423)
Proband X9 (494)
sim (680)
original_glrlm_RunLengthNonUniformityNormalized
frame
10
5
0

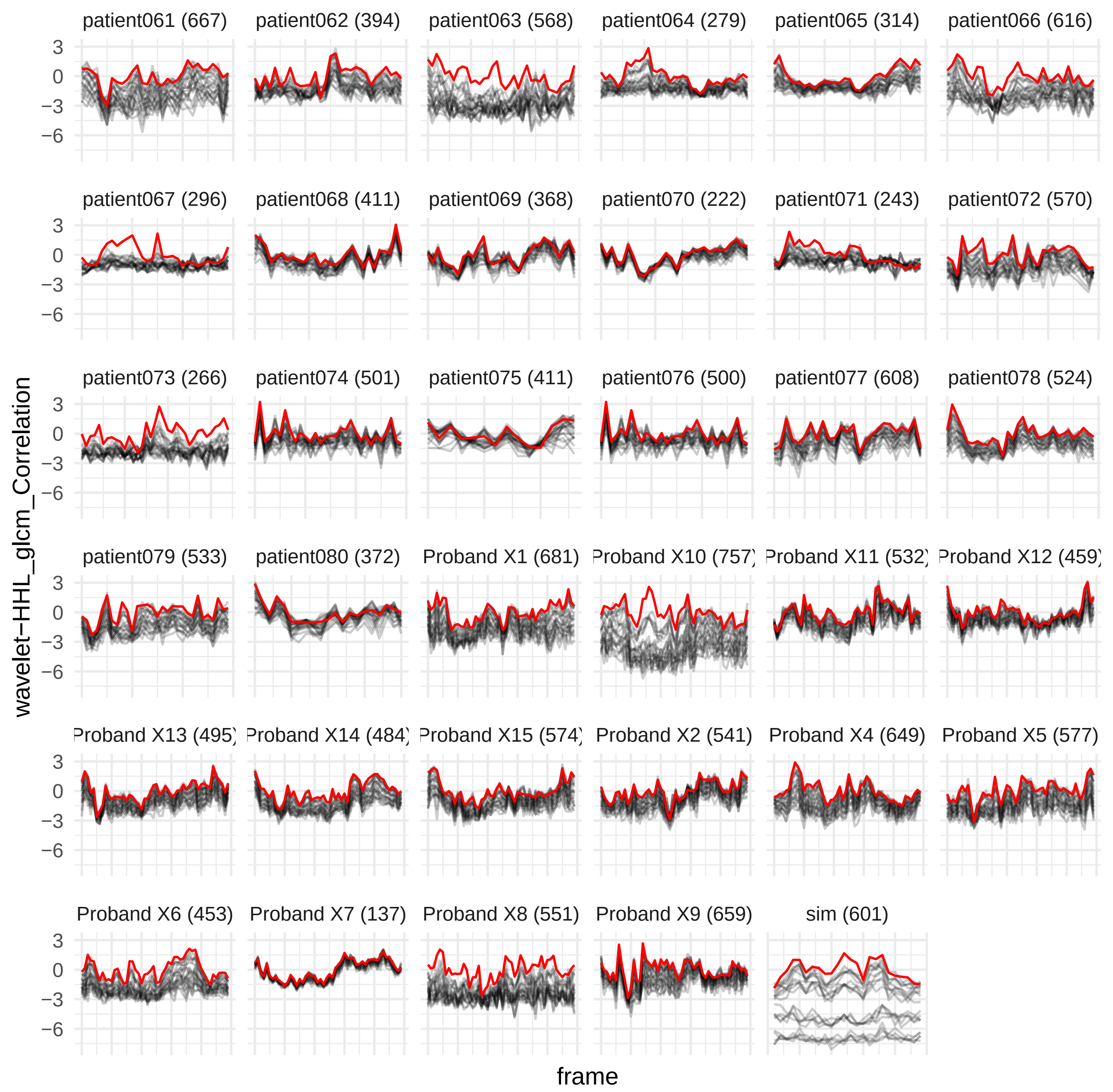

patient061 (667)
patient062 (394)
patient063 (568)
patient064 (279)
patient065 (314)
patient066 (616)
patient067 (296)
patient068 (411)
patient069 (368)
patient070 (222)
patient071 (243)
patient072 (570)
patient073 (266)
patient074 (501)
patient075 (411)
patient076 (500)
patient077 (608)
patient078 (524)
patient079 (533)
patient080 (372)
Proband X1 (681)
Proband X10 (757)
Proband X11 (532)
Proband X12 (459)
Proband X13 (495)
Proband X14 (484)
Proband X15 (574)
Proband X2 (541)
Proband X4 (649)
Proband X5 (577)
Proband X6 (453)
Proband X7 (137)
Proband X8 (551)
Proband X9 (659)
sim (601)
3
0
−3
−6
wavelet−HHL_glcm_Correlation
frame

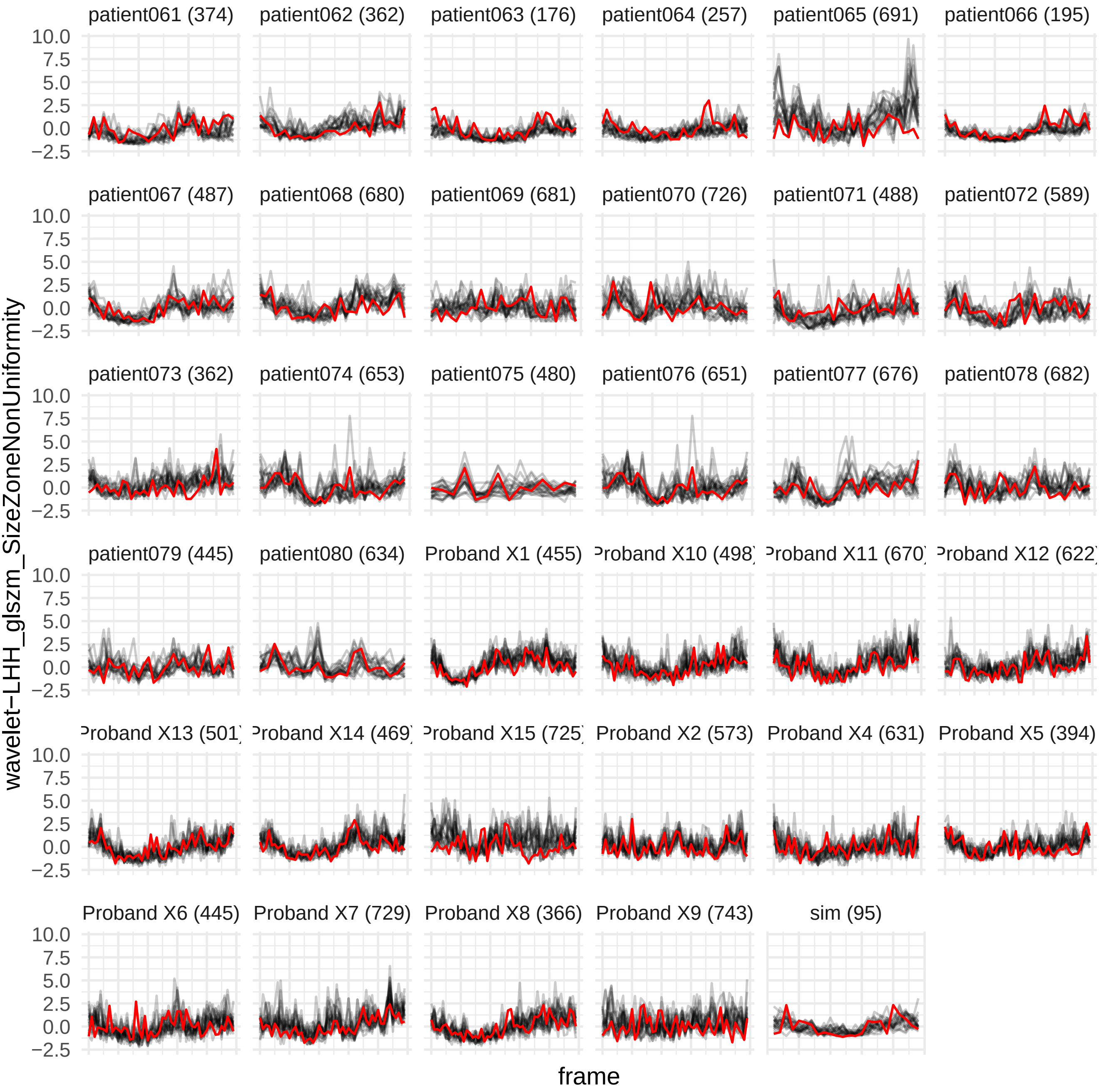

patient061 (374)
patient062 (362)
patient063 (176)
patient064 (257)
patient065 (691)
patient066 (195)
patient067 (487)
patient068 (680)
patient069 (681)
patient070 (726)
patient071 (488)
patient072 (589)
patient073 (362)
patient074 (653)
patient075 (480)
patient076 (651)
patient077 (676)
patient078 (682)
patient079 (445)
patient080 (634)
Proband X1 (455)
Proband X10 (498)
Proband X11 (670)
Proband X12 (622)
Proband X13 (501)
Proband X14 (469)
Proband X15 (725)
Proband X2 (573)
Proband X4 (631)
Proband X5 (394)
Proband X6 (445)
Proband X7 (729)
Proband X8 (366)
Proband X9 (743)
sim (95)
10.0
7.5
5.0
2.5
0.0
−2.5
wavelet−LHH_glszm_SizeZoneNonUniformity
frame

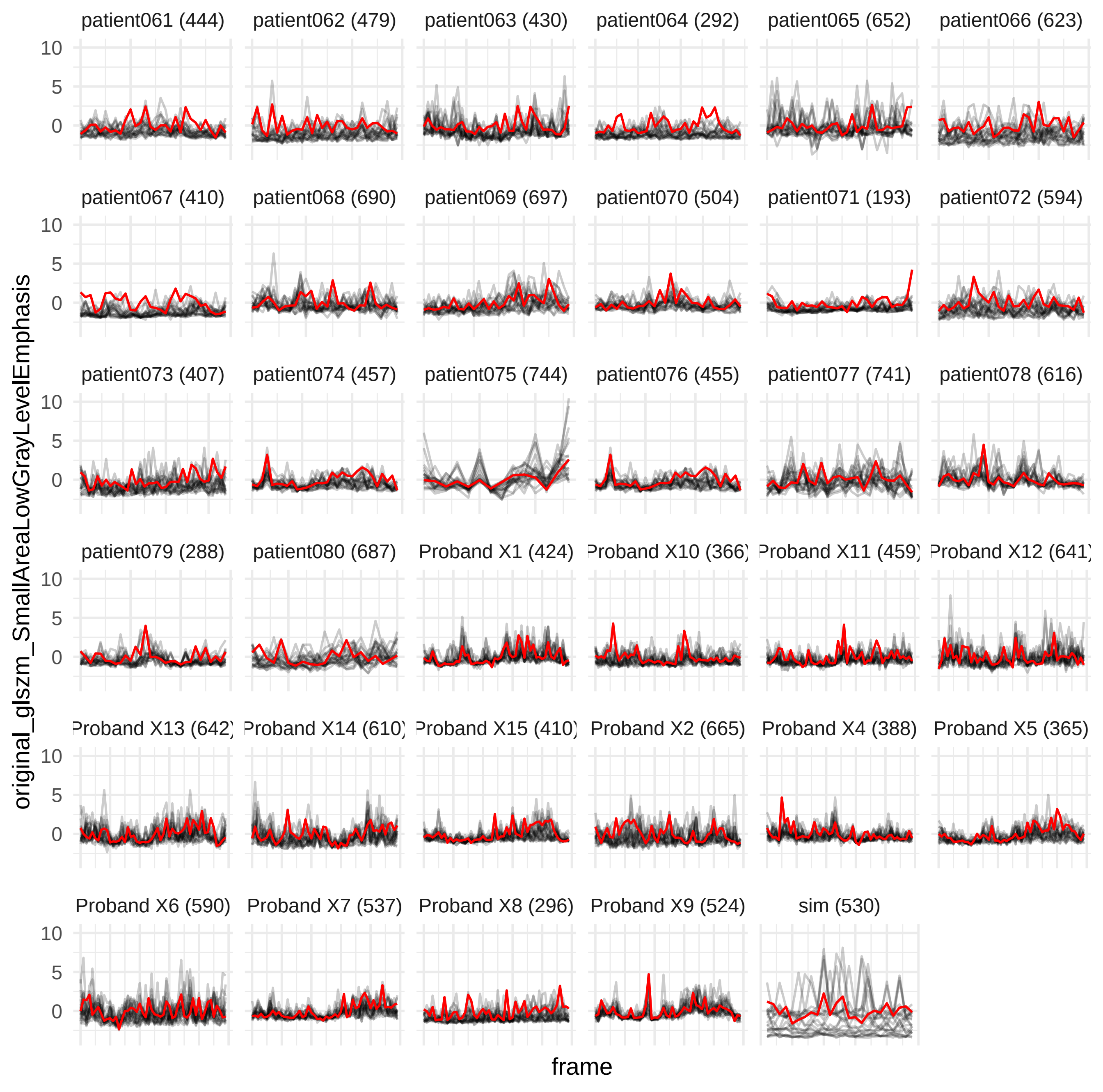
patient061 (444)
patient062 (479)
patient063 (430)
patient064 (292)
patient065 (652)
patient066 (623)
patient067 (410)
patient068 (690)
patient069 (697)
patient070 (504)
patient071 (193)
patient072 (594)
patient073 (407)
patient074 (457)
patient075 (744)
patient076 (455)
patient077 (741)
patient078 (616)
patient079 (288)
patient080 (687)
Proband X1 (424)
Proband X10 (366)
Proband X11 (459)
Proband X12 (641)
Proband X13 (642)
Proband X14 (610)
Proband X15 (410)
Proband X2 (665)
Proband X4 (388)
Proband X5 (365)
Proband X6 (590)
Proband X7 (537)
Proband X8 (296)
Proband X9 (524)
sim (530)
original_glszm_SmallAreaLowGrayLevelEmphasis
frame
10
5
0

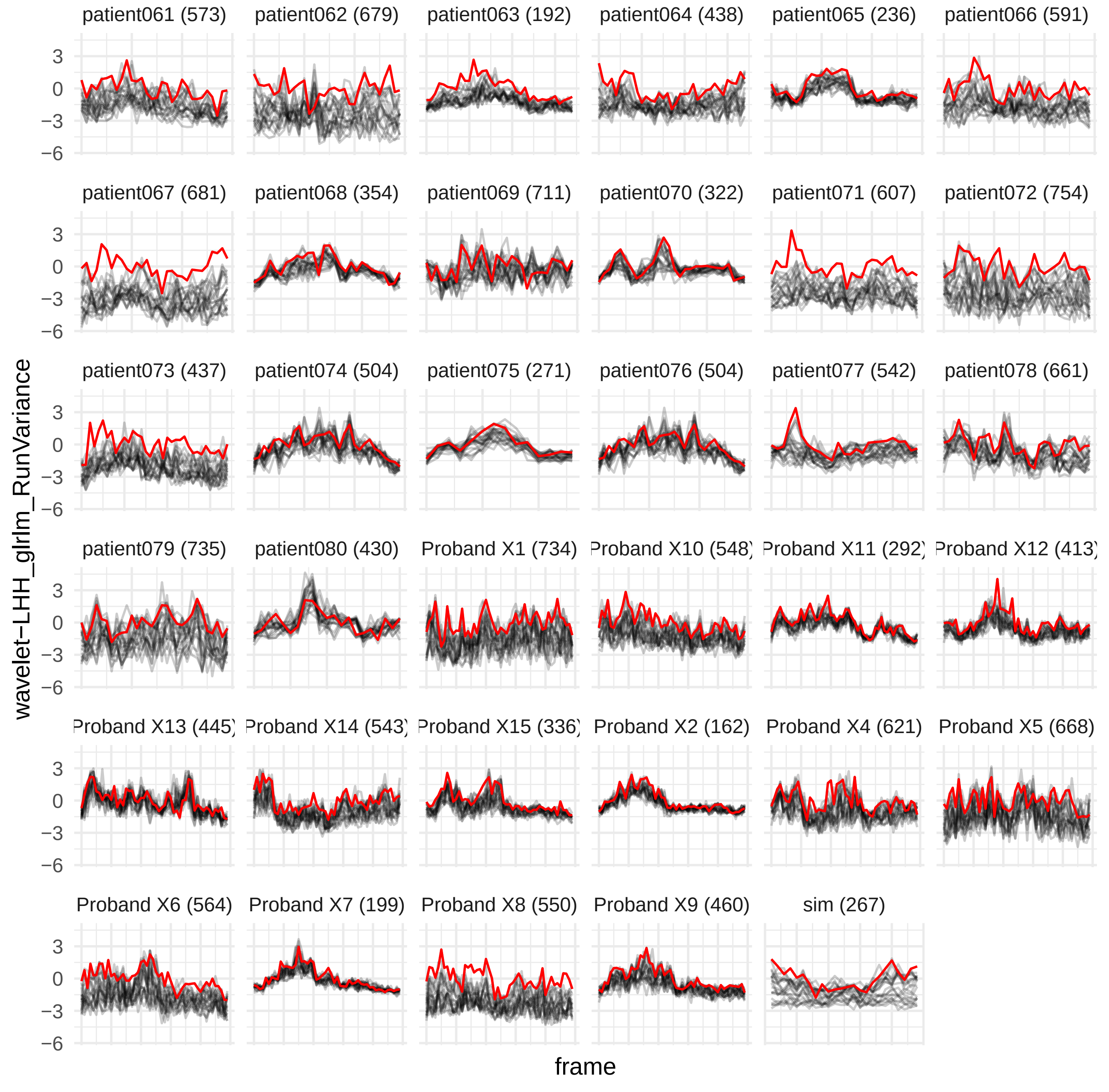

patient061 (573)
patient062 (679)
patient063 (192)
patient064 (438)
patient065 (236)
patient066 (591)
patient067 (681)
patient068 (354)
patient069 (711)
patient070 (322)
patient071 (607)
patient072 (754)
patient073 (437)
patient074 (504)
patient075 (271)
patient076 (504)
patient077 (542)
patient078 (661)
patient079 (735)
patient080 (430)
Proband X1 (734)
Proband X10 (548)
Proband X11 (292)
Proband X12 (413)
Proband X13 (445)
Proband X14 (543)
Proband X15 (336)
Proband X2 (162)
Proband X4 (621)
Proband X5 (668)
Proband X6 (564)
Proband X7 (199)
Proband X8 (550)
Proband X9 (460)
sim (267)
3
0
−3
−6
wavelet−LHH_glrlm_RunVariance
frame

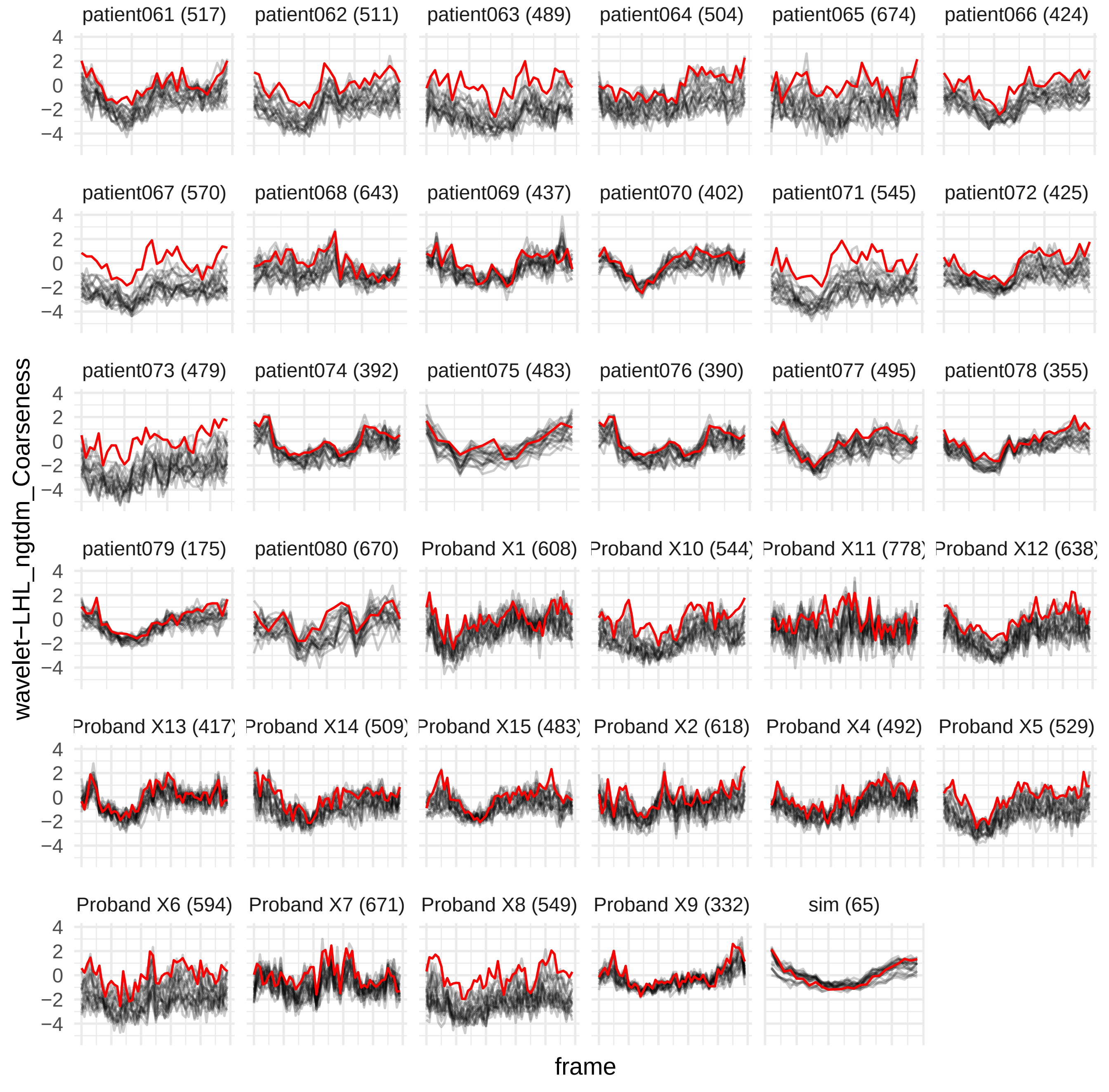
patient061 (517)
patient062 (511)
patient063 (489)
patient064 (504)
patient065 (674)
patient066 (424)
patient067 (570)
patient068 (643)
patient069 (437)
patient070 (402)
patient071 (545)
patient072 (425)
patient073 (479)
patient074 (392)
patient075 (483)
patient076 (390)
patient077 (495)
patient078 (355)
patient079 (175)
patient080 (670)
Proband X1 (608)
Proband X10 (544)
Proband X11 (778)
Proband X12 (638)
Proband X13 (417)
Proband X14 (509)
Proband X15 (483)
Proband X2 (618)
Proband X4 (492)
Proband X5 (529)
Proband X6 (594)
Proband X7 (671)
Proband X8 (549)
Proband X9 (332)
sim (65)
4
2
0
−2
−4
wavelet−LHL_ngtdm_Coarseness
frame

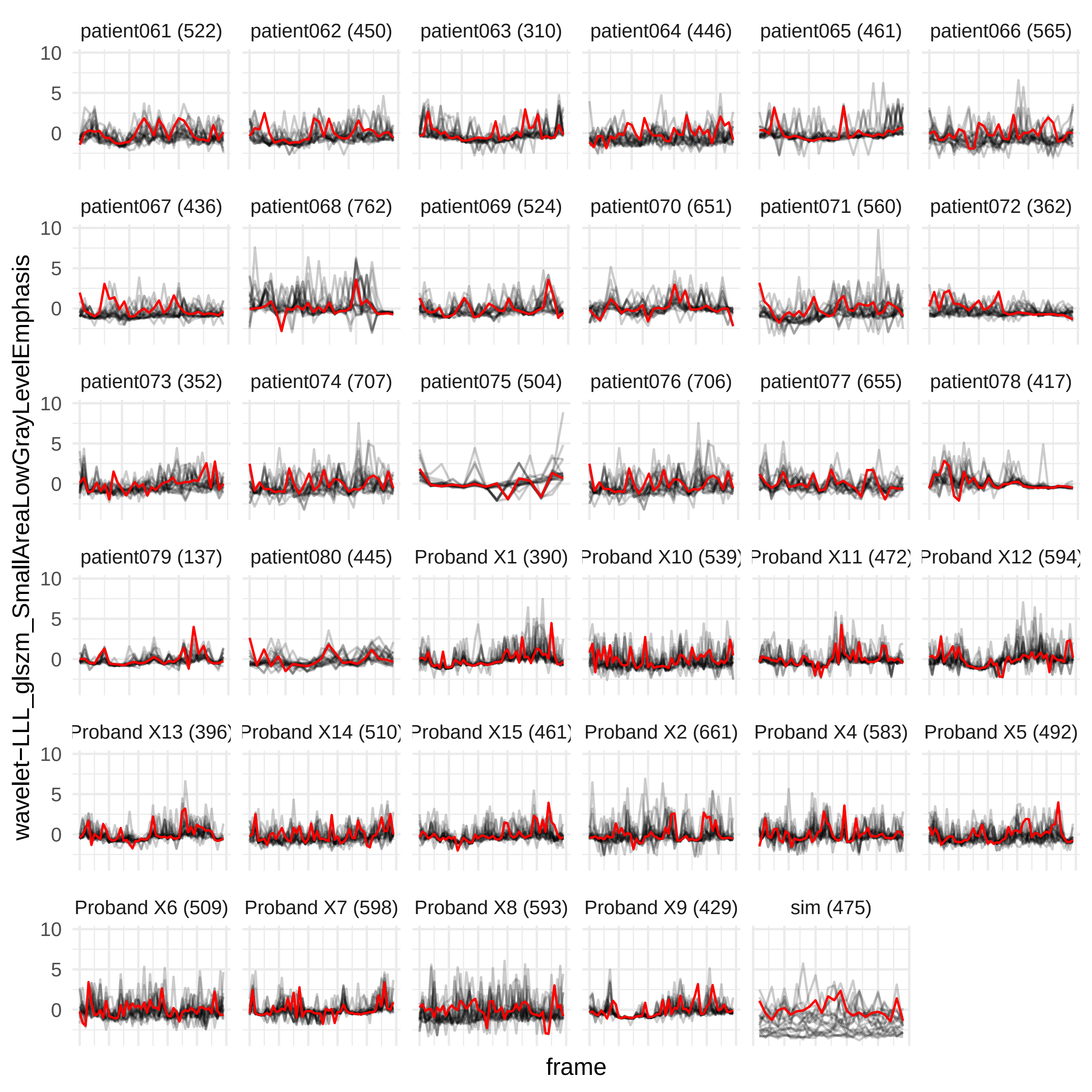

patient061 (522)
patient062 (450)
patient063 (310)
patient064 (446)
patient065 (461)
patient066 (565)
patient067 (436)
patient068 (762)
patient069 (524)
patient070 (651)
patient071 (560)
patient072 (362)
patient073 (352)
patient074 (707)
patient075 (504)
patient076 (706)
patient077 (655)
patient078 (417)
patient079 (137)
patient080 (445)
Proband X1 (390)
Proband X10 (539)
Proband X11 (472)
Proband X12 (594)
Proband X13 (396)
Proband X14 (510)
Proband X15 (461)
Proband X2 (661)
Proband X4 (583)
Proband X5 (492)
Proband X6 (509)
Proband X7 (598)
Proband X8 (593)
Proband X9 (429)
sim (475)
10
5
0
wavelet−LLL_glszm_SmallAreaLowGrayLevelEmphasis
frame

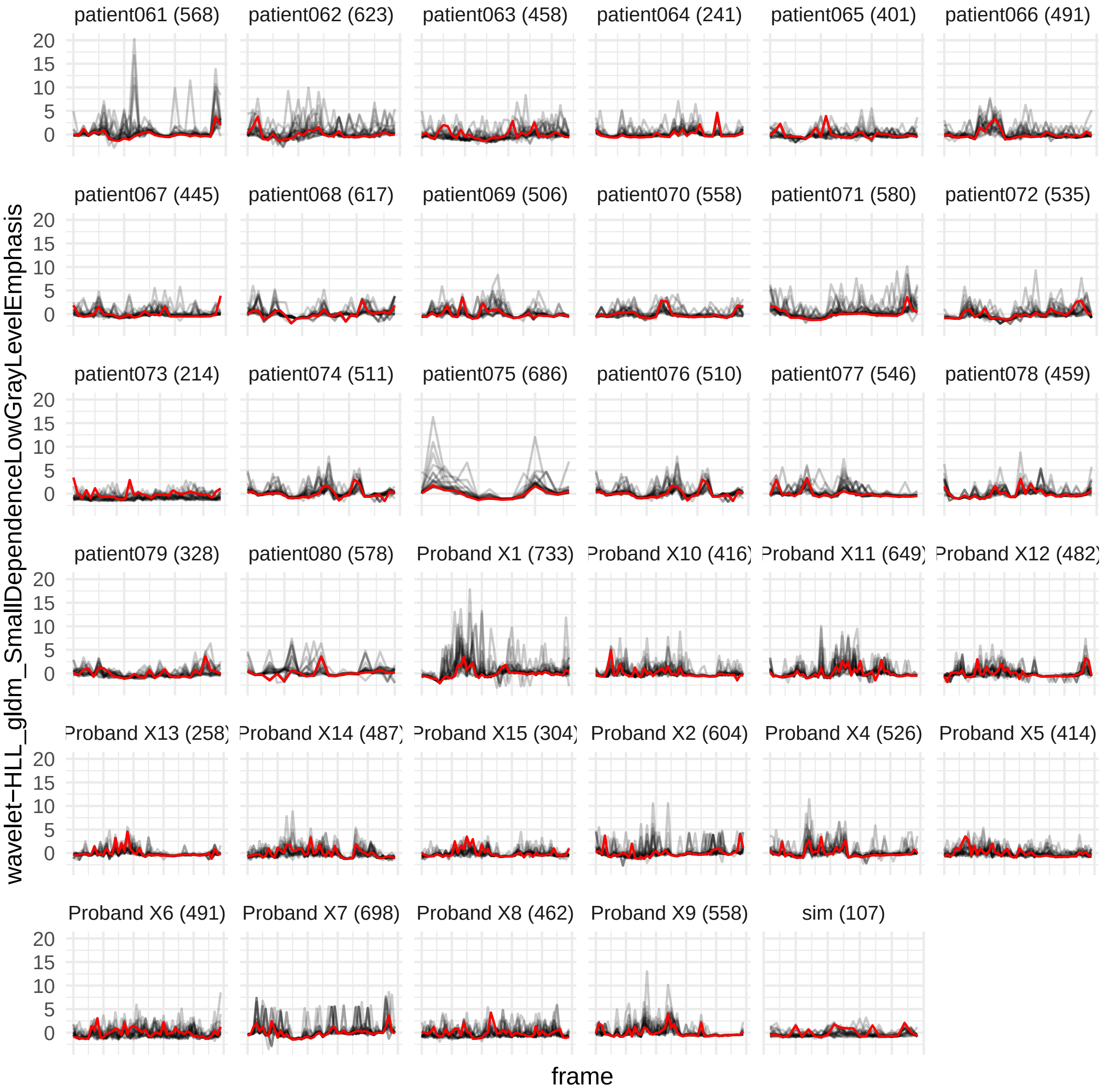
patient061 (568)
patient062 (623)
patient063 (458)
patient064 (241)
patient065 (401)
patient066 (491)
patient067 (445)
patient068 (617)
patient069 (506)
patient070 (558)
patient071 (580)
patient072 (535)
patient073 (214)
patient074 (511)
patient075 (686)
patient076 (510)
patient077 (546)
patient078 (459)
patient079 (328)
patient080 (578)
Proband X1 (733)
Proband X10 (416)
Proband X11 (649)
Proband X12 (482)
Proband X13 (258)
Proband X14 (487)
Proband X15 (304)
Proband X2 (604)
Proband X4 (526)
Proband X5 (414)
Proband X6 (491)
Proband X7 (698)
Proband X8 (462)
Proband X9 (558)
sim (107)
wavelet−HLL_gldm_SmallDependenceLowGrayLevelEmphasis
frame
0
5
10
15
20

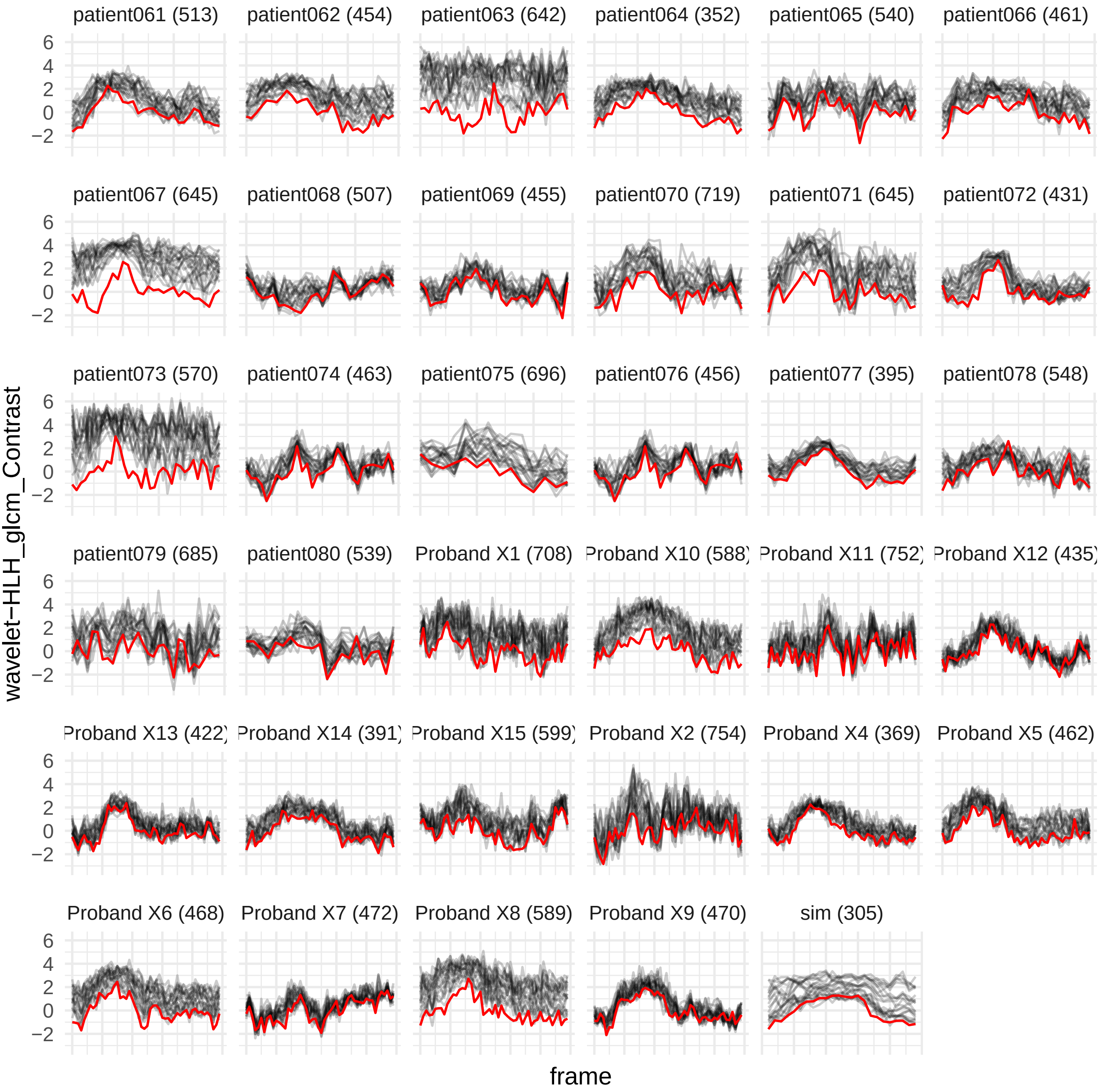

patient061 (513)
patient062 (454)
patient063 (642)
patient064 (352)
patient065 (540)
patient066 (461)
patient067 (645)
patient068 (507)
patient069 (455)
patient070 (719)
patient071 (645)
patient072 (431)
patient073 (570)
patient074 (463)
patient075 (696)
patient076 (456)
patient077 (395)
patient078 (548)
patient079 (685)
patient080 (539)
Proband X1 (708)
Proband X10 (588)
Proband X11 (752)
Proband X12 (435)
Proband X13 (422)
Proband X14 (391)
Proband X15 (599)
Proband X2 (754)
Proband X4 (369)
Proband X5 (462)
Proband X6 (468)
Proband X7 (472)
Proband X8 (589)
Proband X9 (470)
sim (305)
wavelet−HLH_glcm_Contrast
frame
6
4
2
0
−2

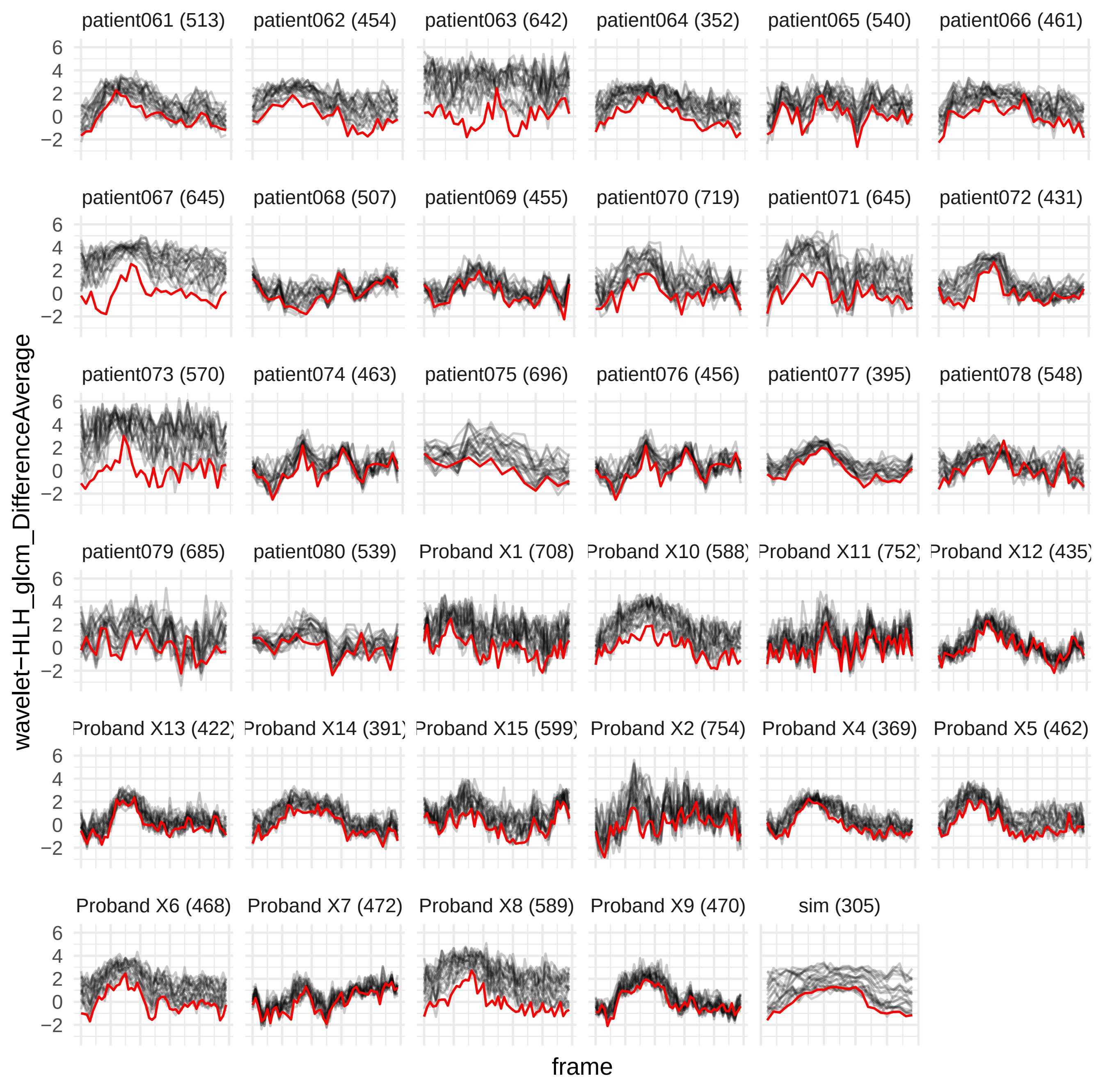

patient061 (513)
patient062 (454)
patient063 (642)
patient064 (352)
patient065 (540)
patient066 (461)
patient067 (645)
patient068 (507)
patient069 (455)
patient070 (719)
patient071 (645)
patient072 (431)
patient073 (570)
patient074 (463)
patient075 (696)
patient076 (456)
patient077 (395)
patient078 (548)
patient079 (685)
patient080 (539)
Proband X1 (708)
Proband X10 (588)
Proband X11 (752)
Proband X12 (435)
Proband X13 (422)
Proband X14 (391)
Proband X15 (599)
Proband X2 (754)
Proband X4 (369)
Proband X5 (462)
Proband X6 (468)
Proband X7 (472)
Proband X8 (589)
Proband X9 (470)
sim (305)
6
4
2
0
−2
wavelet−HLH_glcm_DifferenceAverage
frame

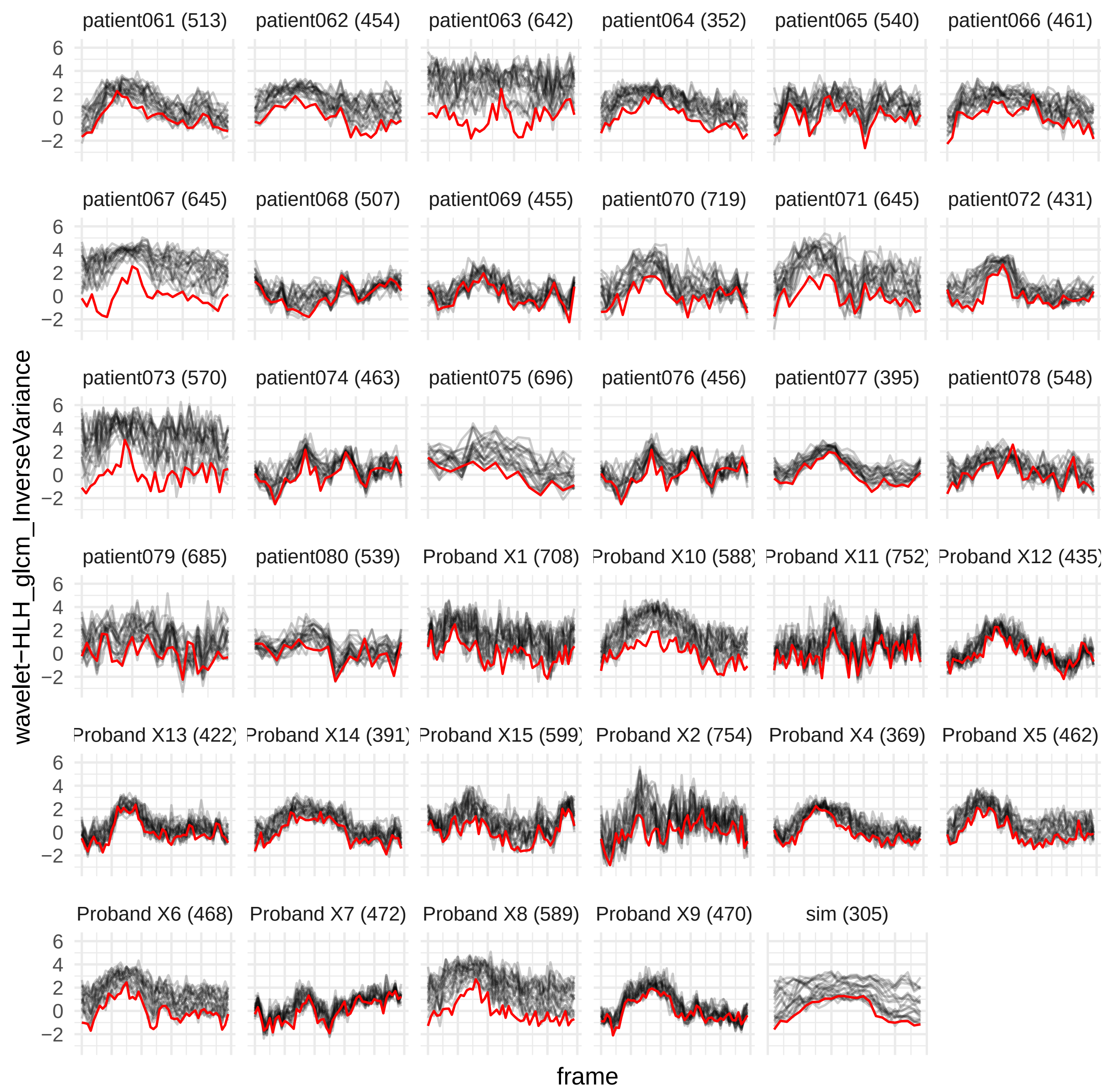
patient061 (513)
patient062 (454)
patient063 (642)
patient064 (352)
patient065 (540)
patient066 (461)
patient067 (645)
patient068 (507)
patient069 (455)
patient070 (719)
patient071 (645)
patient072 (431)
patient073 (570)
patient074 (463)
patient075 (696)
patient076 (456)
patient077 (395)
patient078 (548)
patient079 (685)
patient080 (539)
Proband X1 (708)
Proband X10 (588)
Proband X11 (752)
Proband X12 (435)
Proband X13 (422)
Proband X14 (391)
Proband X15 (599)
Proband X2 (754)
Proband X4 (369)
Proband X5 (462)
Proband X6 (468)
Proband X7 (472)
Proband X8 (589)
Proband X9 (470)
sim (305)
wavelet−HLH_glcm_InverseVariance
frame

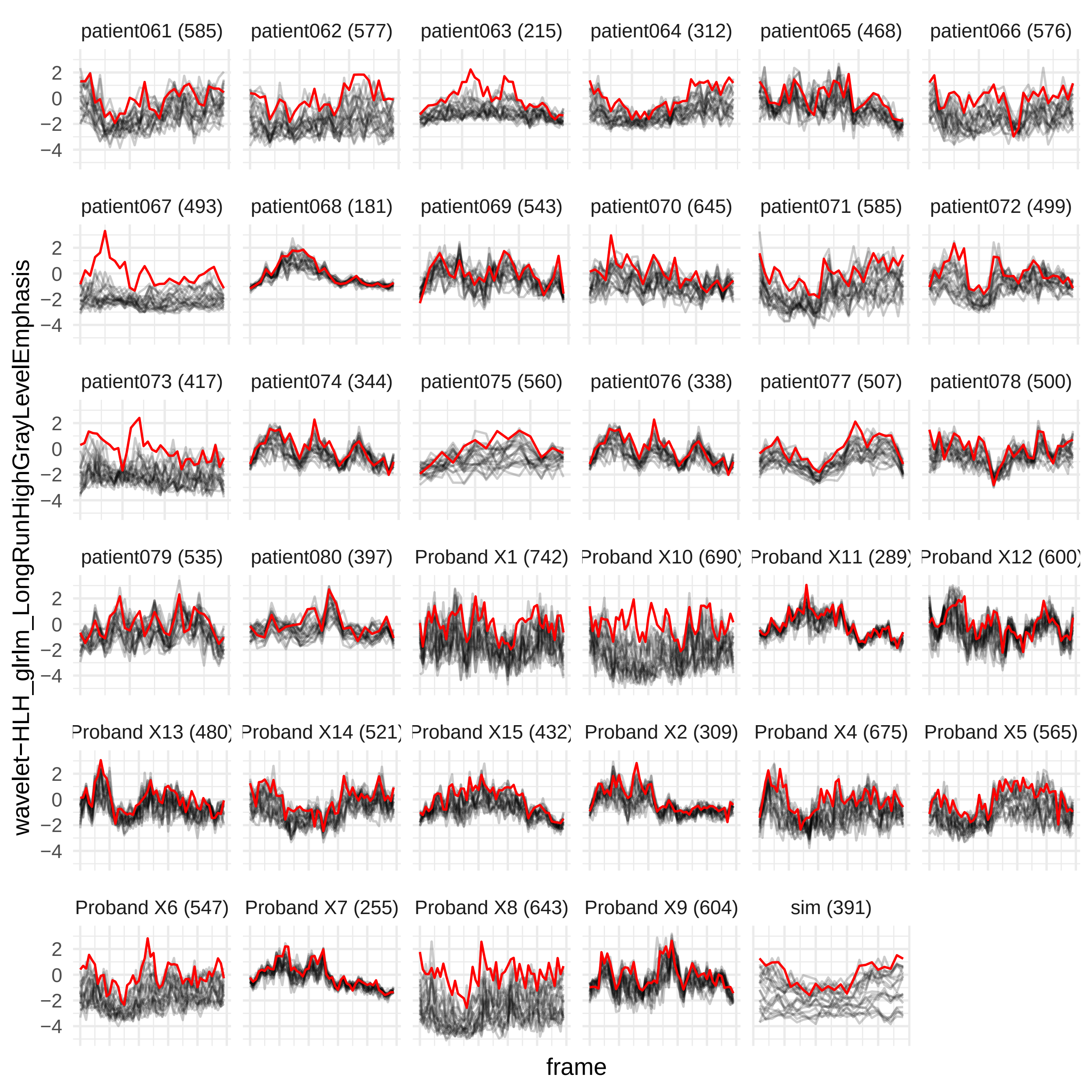
patient061 (585)
patient062 (577)
patient063 (215)
patient064 (312)
patient065 (468)
patient066 (576)
patient067 (493)
patient068 (181)
patient069 (543)
patient070 (645)
patient071 (585)
patient072 (499)
patient073 (417)
patient074 (344)
patient075 (560)
patient076 (338)
patient077 (507)
patient078 (500)
patient079 (535)
patient080 (397)
Proband X1 (742)
Proband X10 (690)
Proband X11 (289)
Proband X12 (600)
Proband X13 (480)
Proband X14 (521)
Proband X15 (432)
Proband X2 (309)
Proband X4 (675)
Proband X5 (565)
Proband X6 (547)
Proband X7 (255)
Proband X8 (643)
Proband X9 (604)
sim (391)
wavelet−HLH_glrlm_LongRunHighGrayLevelEmphasis
frame
2
0
−2
−4

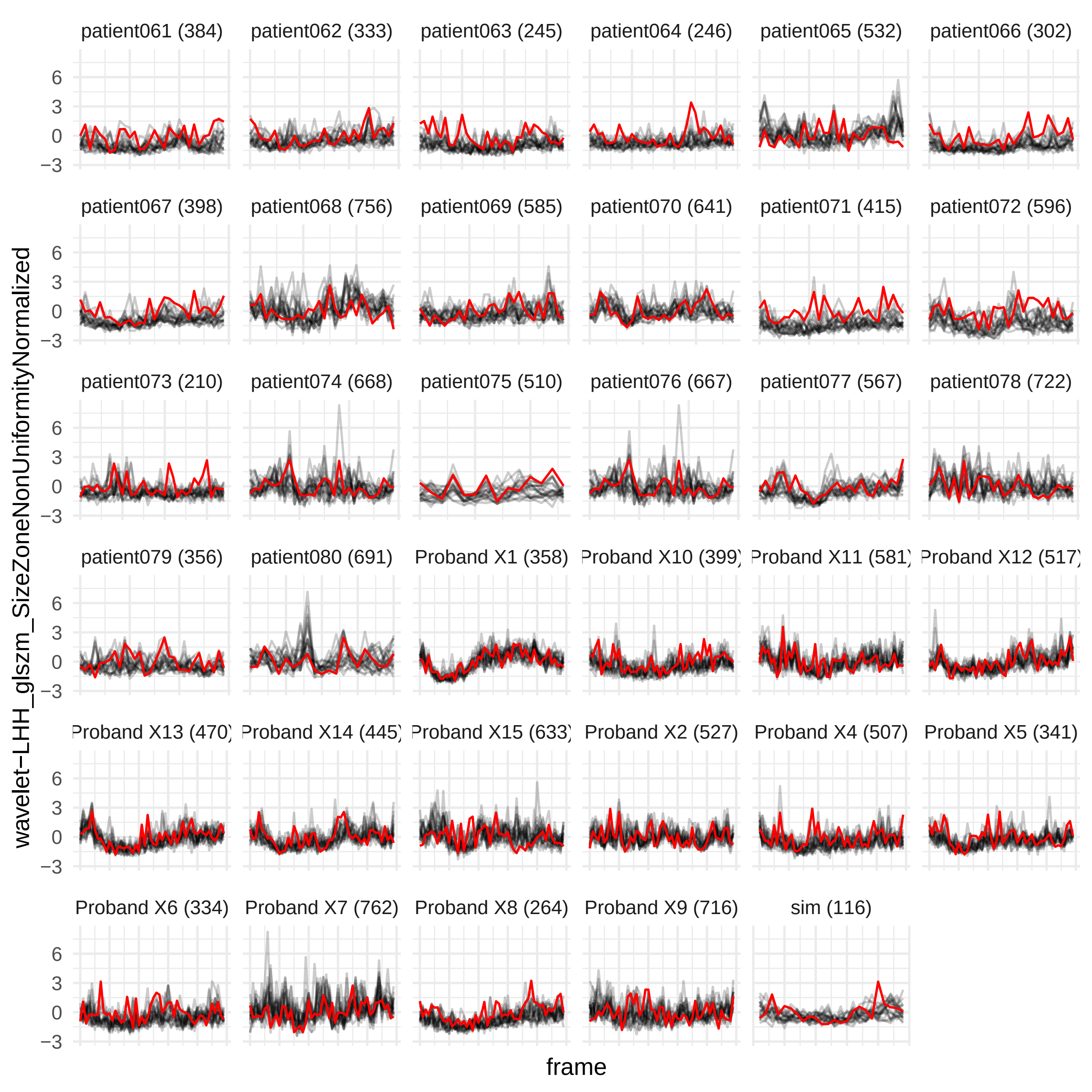

patient061 (384)
patient062 (333)
patient063 (245)
patient064 (246)
patient065 (532)
patient066 (302)
patient067 (398)
patient068 (756)
patient069 (585)
patient070 (641)
patient071 (415)
patient072 (596)
patient073 (210)
patient074 (668)
patient075 (510)
patient076 (667)
patient077 (567)
patient078 (722)
patient079 (356)
patient080 (691)
Proband X1 (358)
Proband X10 (399)
Proband X11 (581)
Proband X12 (517)
Proband X13 (470)
Proband X14 (445)
Proband X15 (633)
Proband X2 (527)
Proband X4 (507)
Proband X5 (341)
Proband X6 (334)
Proband X7 (762)
Proband X8 (264)
Proband X9 (716)
sim (116)
6
3
0
−3
wavelet−LHH_glszm_SizeZoneNonUniformityNormalized
frame

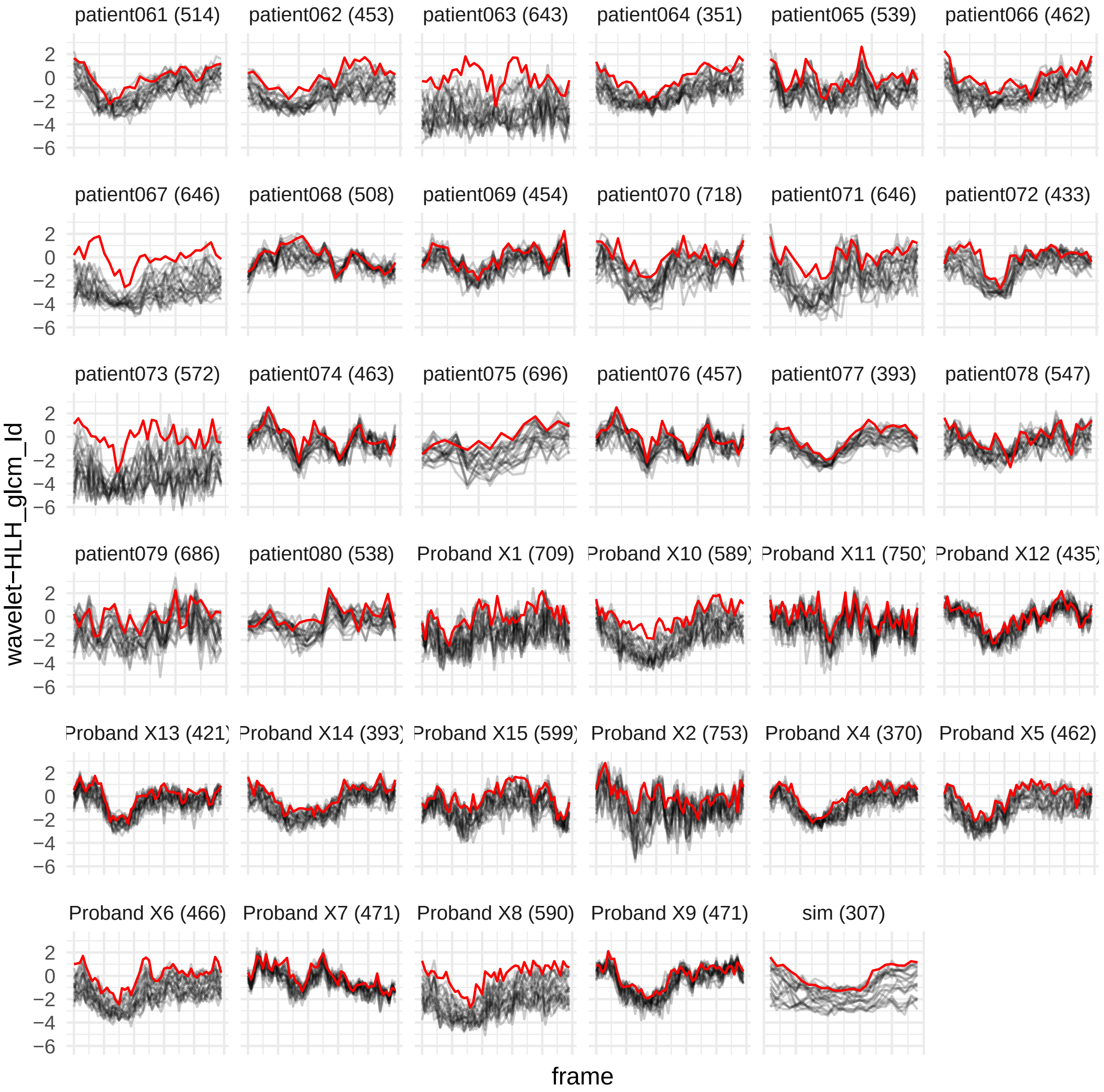

patient061 (514)
patient062 (453)
patient063 (643)
patient064 (351)
patient065 (539)
patient066 (462)
patient067 (646)
patient068 (508)
patient069 (454)
patient070 (718)
patient071 (646)
patient072 (433)
patient073 (572)
patient074 (463)
patient075 (696)
patient076 (457)
patient077 (393)
patient078 (547)
patient079 (686)
patient080 (538)
Proband X1 (709)
Proband X10 (589)
Proband X11 (750)
Proband X12 (435)
Proband X13 (421)
Proband X14 (393)
Proband X15 (599)
Proband X2 (753)
Proband X4 (370)
Proband X5 (462)
Proband X6 (466)
Proband X7 (471)
Proband X8 (590)
Proband X9 (471)
sim (307)
2
0
−2
−4
−6
wavelet−HLH_glcm_Id
frame

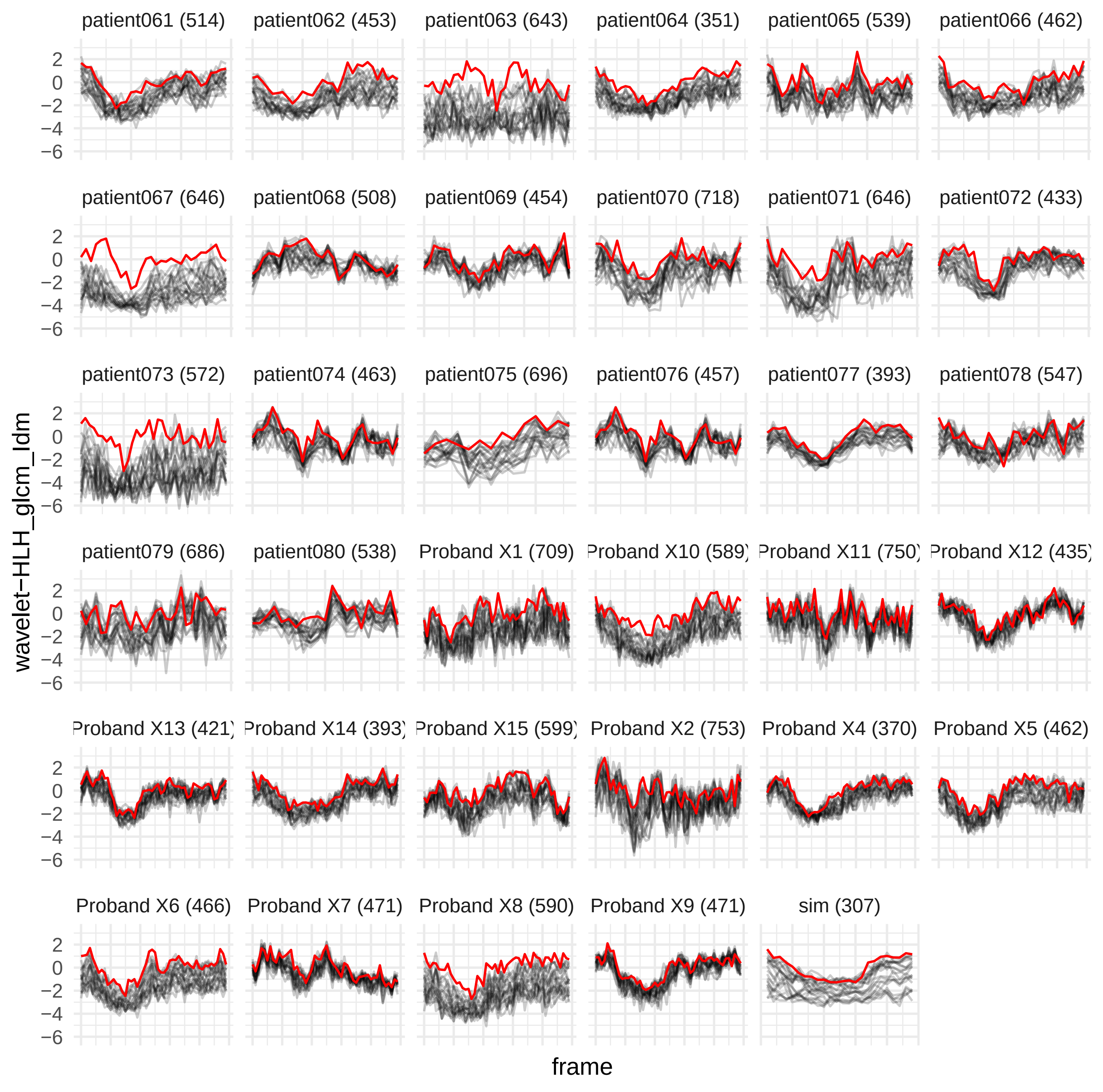

patient061 (514)
patient062 (453)
patient063 (643)
patient064 (351)
patient065 (539)
patient066 (462)
patient067 (646)
patient068 (508)
patient069 (454)
patient070 (718)
patient071 (646)
patient072 (433)
patient073 (572)
patient074 (463)
patient075 (696)
patient076 (457)
patient077 (393)
patient078 (547)
patient079 (686)
patient080 (538)
Proband X1 (709)
Proband X10 (589)
Proband X11 (750)
Proband X12 (435)
Proband X13 (421)
Proband X14 (393)
Proband X15 (599)
Proband X2 (753)
Proband X4 (370)
Proband X5 (462)
Proband X6 (466)
Proband X7 (471)
Proband X8 (590)
Proband X9 (471)
sim (307)
wavelet−HLH_glcm_Idm
frame
2
0
−2
−4
−6

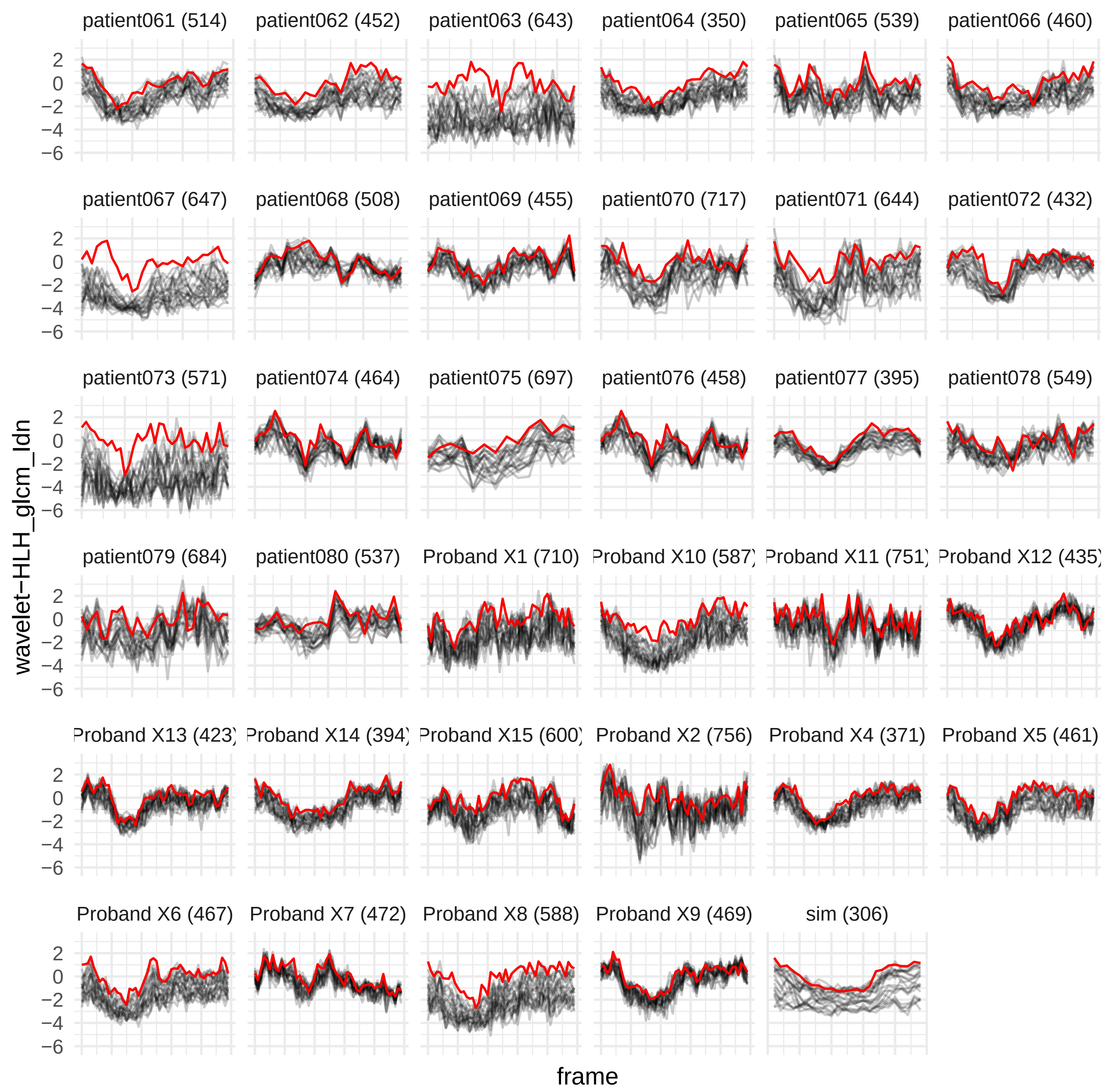

patient061 (514)
patient062 (452)
patient063 (643)
patient064 (350)
patient065 (539)
patient066 (460)
patient067 (647)
patient068 (508)
patient069 (455)
patient070 (717)
patient071 (644)
patient072 (432)
patient073 (571)
patient074 (464)
patient075 (697)
patient076 (458)
patient077 (395)
patient078 (549)
patient079 (684)
patient080 (537)
Proband X1 (710)
Proband X10 (587)
Proband X11 (751)
Proband X12 (435)
Proband X13 (423)
Proband X14 (394)
Proband X15 (600)
Proband X2 (756)
Proband X4 (371)
Proband X5 (461)
Proband X6 (467)
Proband X7 (472)
Proband X8 (588)
Proband X9 (469)
sim (306)
2
0
−2
−4
−6
wavelet−HLH_glcm_Idn
frame

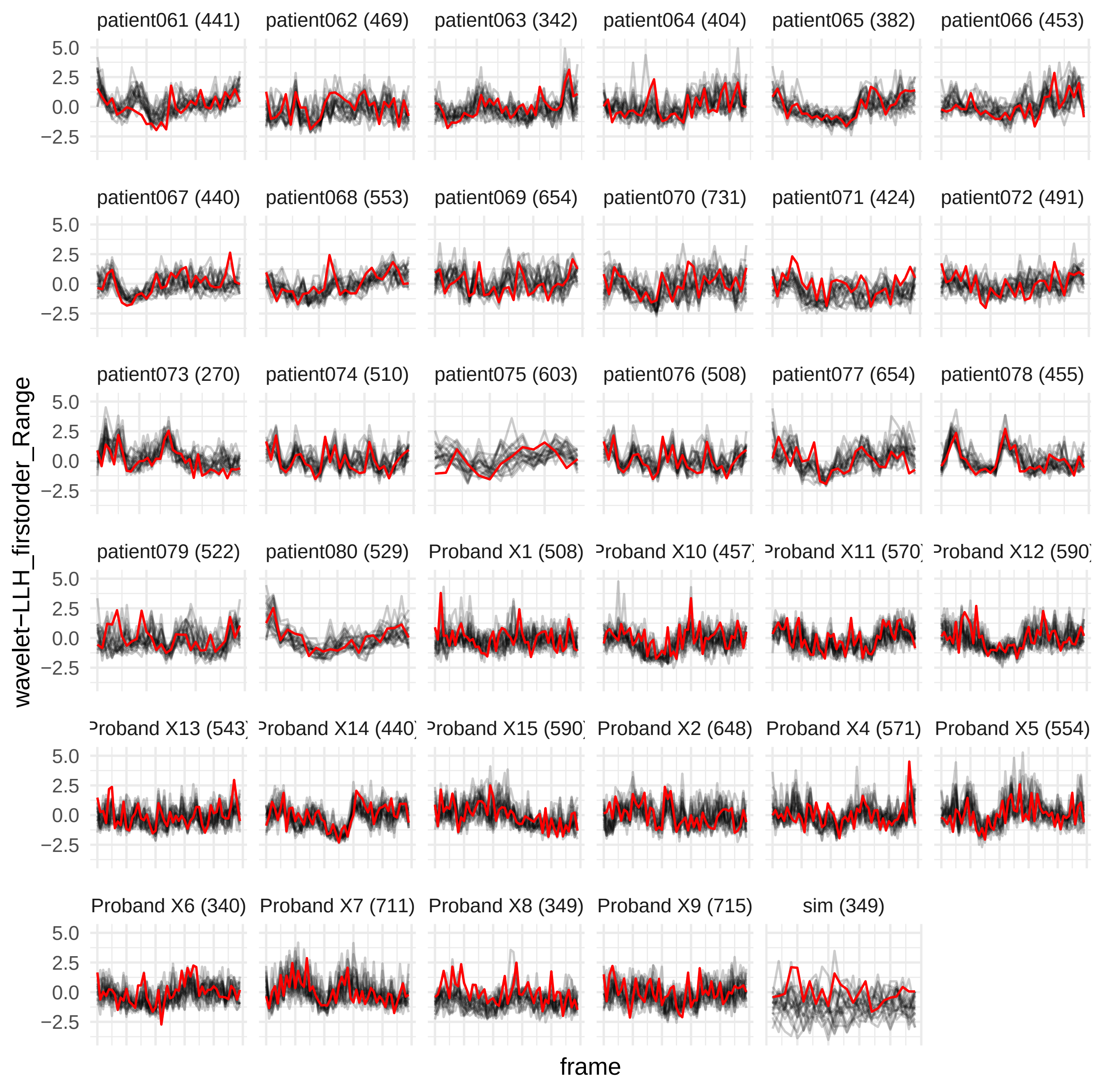
patient061 (441)
patient062 (469)
patient063 (342)
patient064 (404)
patient065 (382)
patient066 (453)
patient067 (440)
patient068 (553)
patient069 (654)
patient070 (731)
patient071 (424)
patient072 (491)
patient073 (270)
patient074 (510)
patient075 (603)
patient076 (508)
patient077 (654)
patient078 (455)
patient079 (522)
patient080 (529)
Proband X1 (508)
Proband X10 (457)
Proband X11 (570)
Proband X12 (590)
Proband X13 (543)
Proband X14 (440)
Proband X15 (590)
Proband X2 (648)
Proband X4 (571)
Proband X5 (554)
Proband X6 (340)
Proband X7 (711)
Proband X8 (349)
Proband X9 (715)
sim (349)
5.0
2.5
0.0
−2.5
wavelet−LLH_firstorder_Range
frame

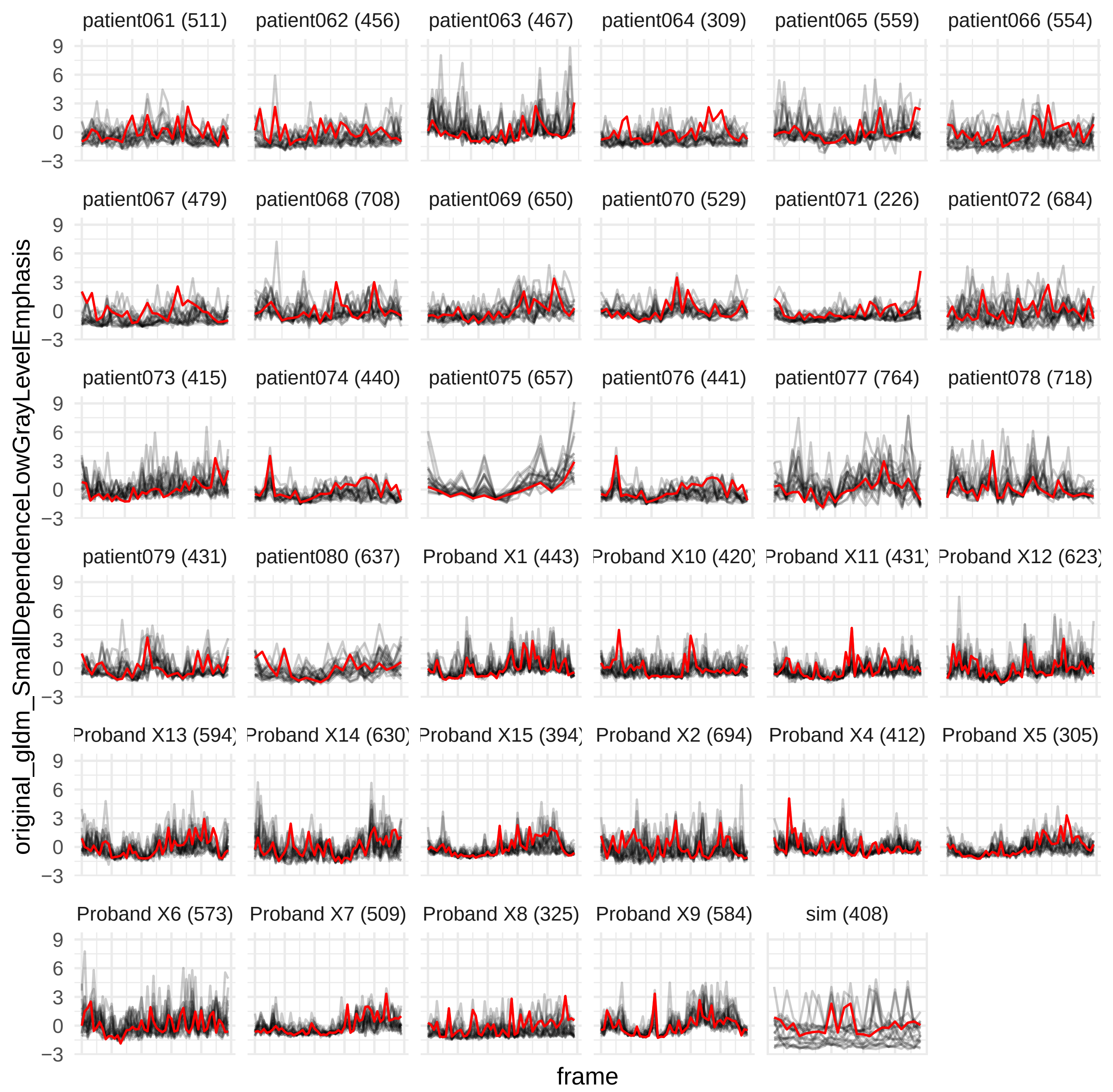
patient061 (511)
patient062 (456)
patient063 (467)
patient064 (309)
patient065 (559)
patient066 (554)
patient067 (479)
patient068 (708)
patient069 (650)
patient070 (529)
patient071 (226)
patient072 (684)
patient073 (415)
patient074 (440)
patient075 (657)
patient076 (441)
patient077 (764)
patient078 (718)
patient079 (431)
patient080 (637)
Proband X1 (443)
Proband X10 (420)
Proband X11 (431)
Proband X12 (623)
Proband X13 (594)
Proband X14 (630)
Proband X15 (394)
Proband X2 (694)
Proband X4 (412)
Proband X5 (305)
Proband X6 (573)
Proband X7 (509)
Proband X8 (325)
Proband X9 (584)
sim (408)
original_gldm_SmallDependenceLowGrayLevelEmphasis
frame
9
6
3
0
−3

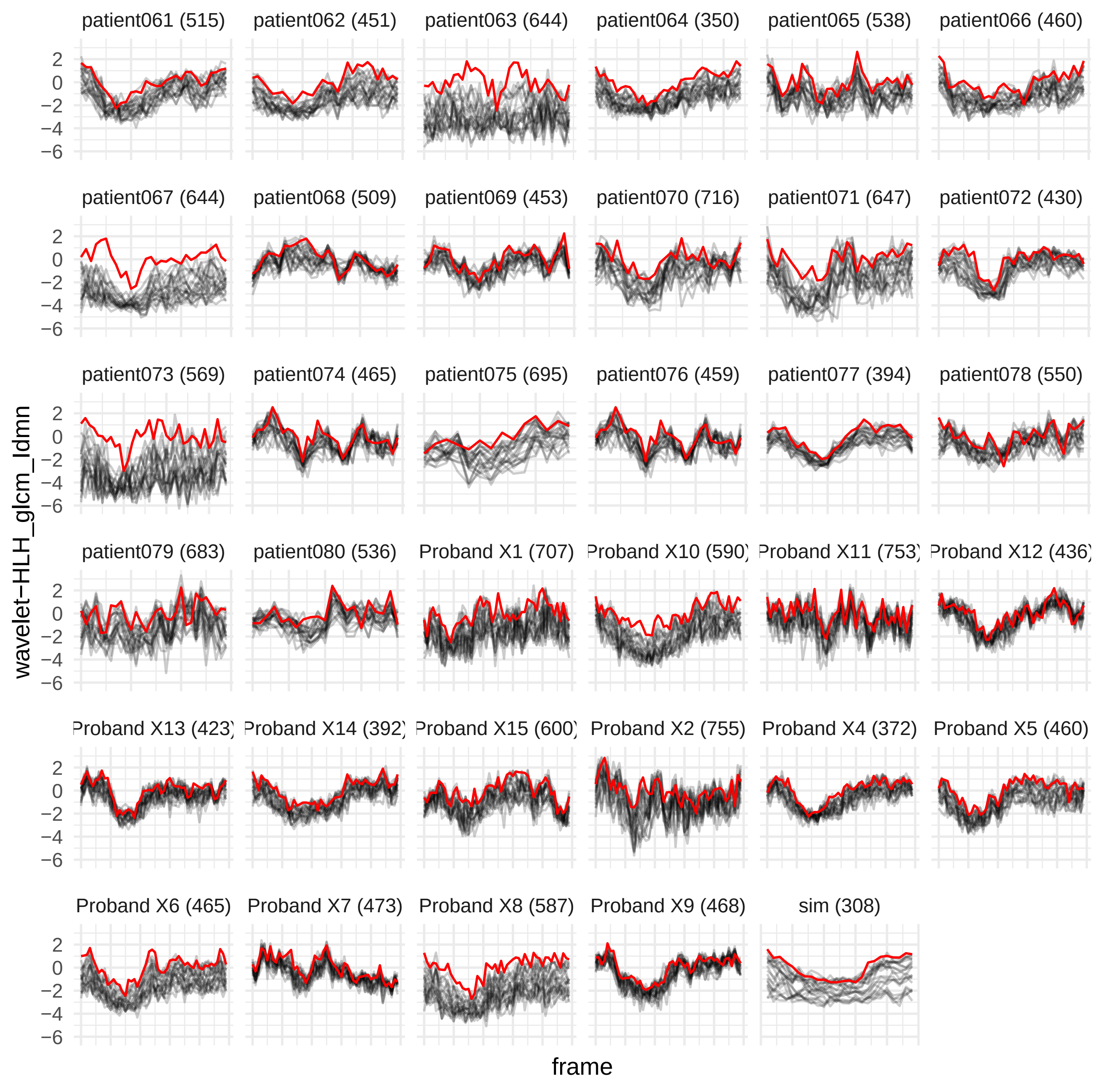

patient061 (515)
patient062 (451)
patient063 (644)
patient064 (350)
patient065 (538)
patient066 (460)
patient067 (644)
patient068 (509)
patient069 (453)
patient070 (716)
patient071 (647)
patient072 (430)
patient073 (569)
patient074 (465)
patient075 (695)
patient076 (459)
patient077 (394)
patient078 (550)
patient079 (683)
patient080 (536)
Proband X1 (707)
Proband X10 (590)
Proband X11 (753)
Proband X12 (436)
Proband X13 (423)
Proband X14 (392)
Proband X15 (600)
Proband X2 (755)
Proband X4 (372)
Proband X5 (460)
Proband X6 (465)
Proband X7 (473)
Proband X8 (587)
Proband X9 (468)
sim (308)
wavelet−HLH_glcm_Idmn
2
0
−2
−4
−6
frame

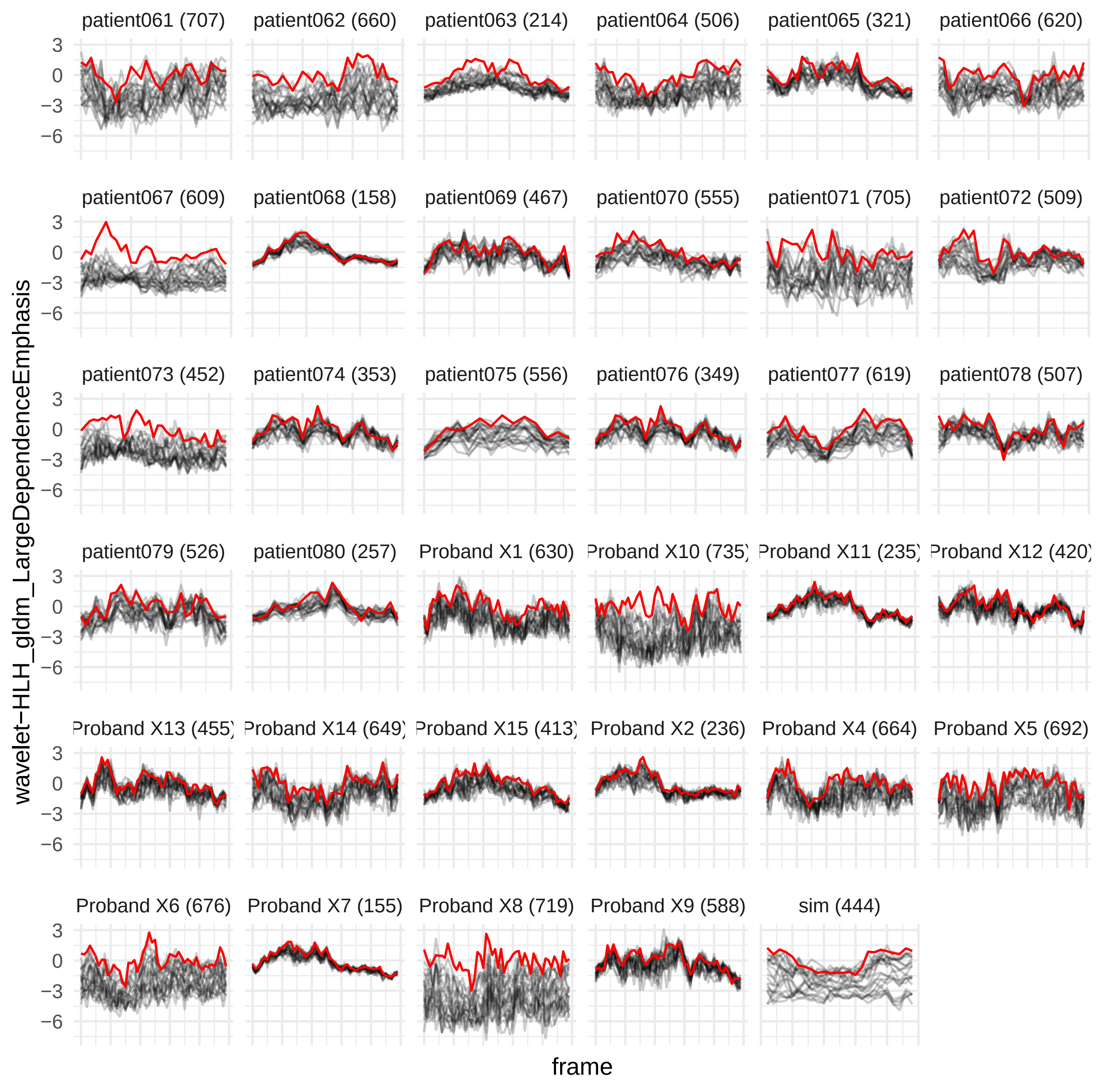
patient061 (707)
patient062 (660)
patient063 (214)
patient064 (506)
patient065 (321)
patient066 (620)
patient067 (609)
patient068 (158)
patient069 (467)
patient070 (555)
patient071 (705)
patient072 (509)
patient073 (452)
patient074 (353)
patient075 (556)
patient076 (349)
patient077 (619)
patient078 (507)
patient079 (526)
patient080 (257)
Proband X1 (630)
Proband X10 (735)
Proband X11 (235)
Proband X12 (420)
Proband X13 (455)
Proband X14 (649)
Proband X15 (413)
Proband X2 (236)
Proband X4 (664)
Proband X5 (692)
Proband X6 (676)
Proband X7 (155)
Proband X8 (719)
Proband X9 (588)
sim (444)
3
0
−3
−6
wavelet−HLH_gldm_LargeDependenceEmphasis
frame

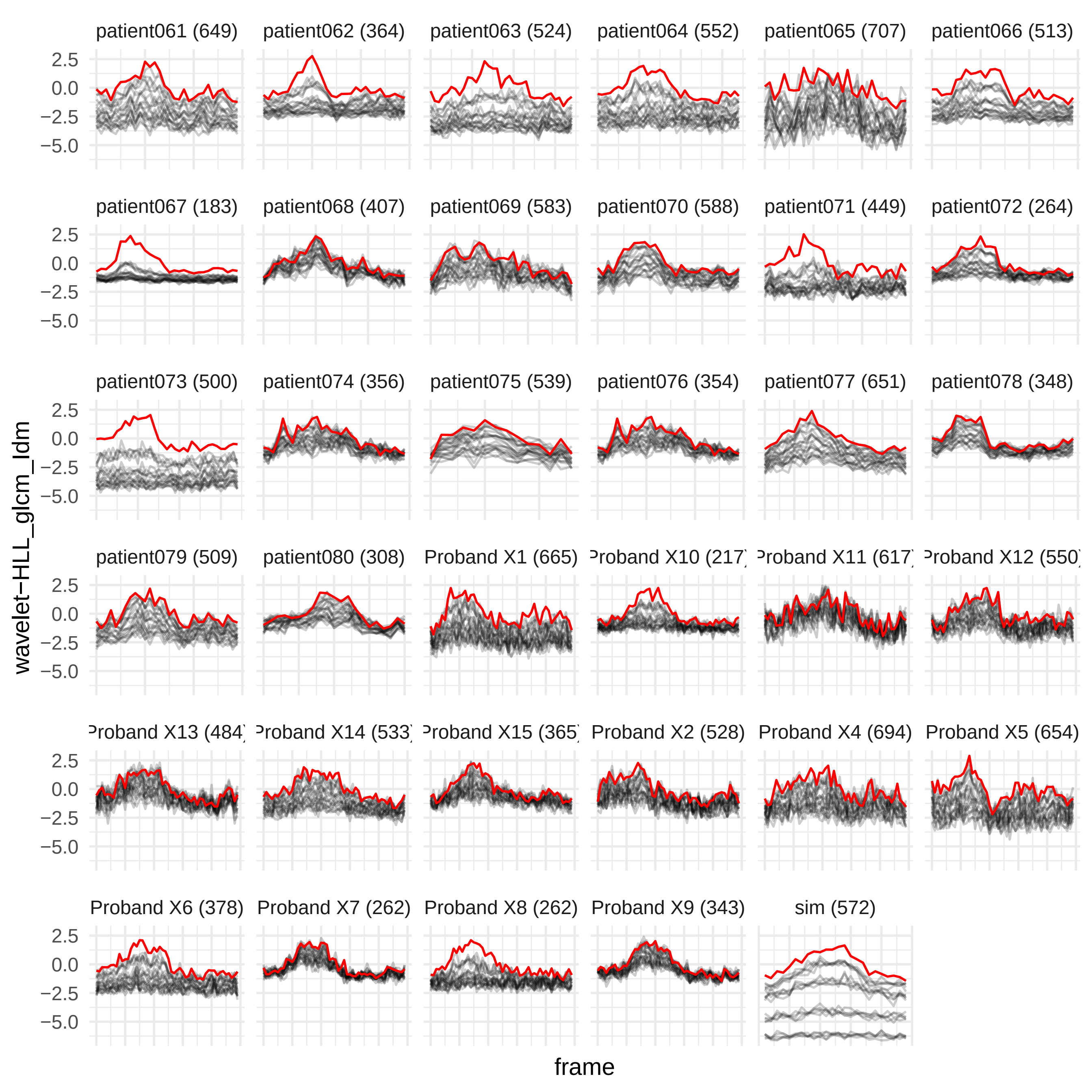
patient061 (649)
patient062 (364)
patient063 (524)
patient064 (552)
patient065 (707)
patient066 (513)
patient067 (183)
patient068 (407)
patient069 (583)
patient070 (588)
patient071 (449)
patient072 (264)
patient073 (500)
patient074 (356)
patient075 (539)
patient076 (354)
patient077 (651)
patient078 (348)
patient079 (509)
patient080 (308)
Proband X1 (665)
Proband X10 (217)
Proband X11 (617)
Proband X12 (550)
Proband X13 (484)
Proband X14 (533)
Proband X15 (365)
Proband X2 (528)
Proband X4 (694)
Proband X5 (654)
Proband X6 (378)
Proband X7 (262)
Proband X8 (262)
Proband X9 (343)
sim (572)
2.5
0.0
−2.5
−5.0
wavelet−HLL_glcm_Idm
frame

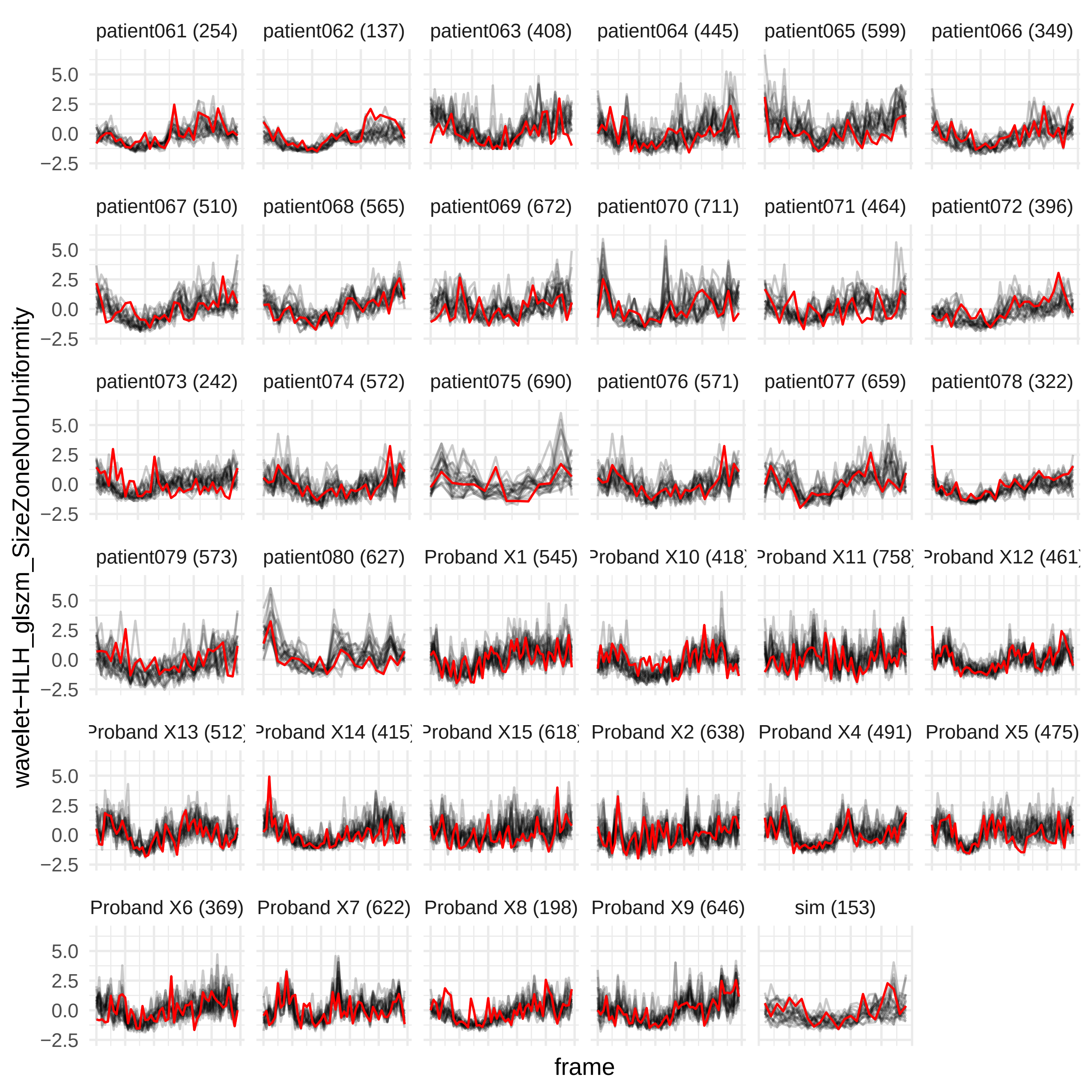
patient061 (254)
patient062 (137)
patient063 (408)
patient064 (445)
patient065 (599)
patient066 (349)
patient067 (510)
patient068 (565)
patient069 (672)
patient070 (711)
patient071 (464)
patient072 (396)
patient073 (242)
patient074 (572)
patient075 (690)
patient076 (571)
patient077 (659)
patient078 (322)
patient079 (573)
patient080 (627)
Proband X1 (545)
Proband X10 (418)
Proband X11 (758)
Proband X12 (461)
Proband X13 (512)
Proband X14 (415)
Proband X15 (618)
Proband X2 (638)
Proband X4 (491)
Proband X5 (475)
Proband X6 (369)
Proband X7 (622)
Proband X8 (198)
Proband X9 (646)
sim (153)
5.0
2.5
0.0
−2.5
wavelet−HLH_glszm_SizeZoneNonUniformity
frame

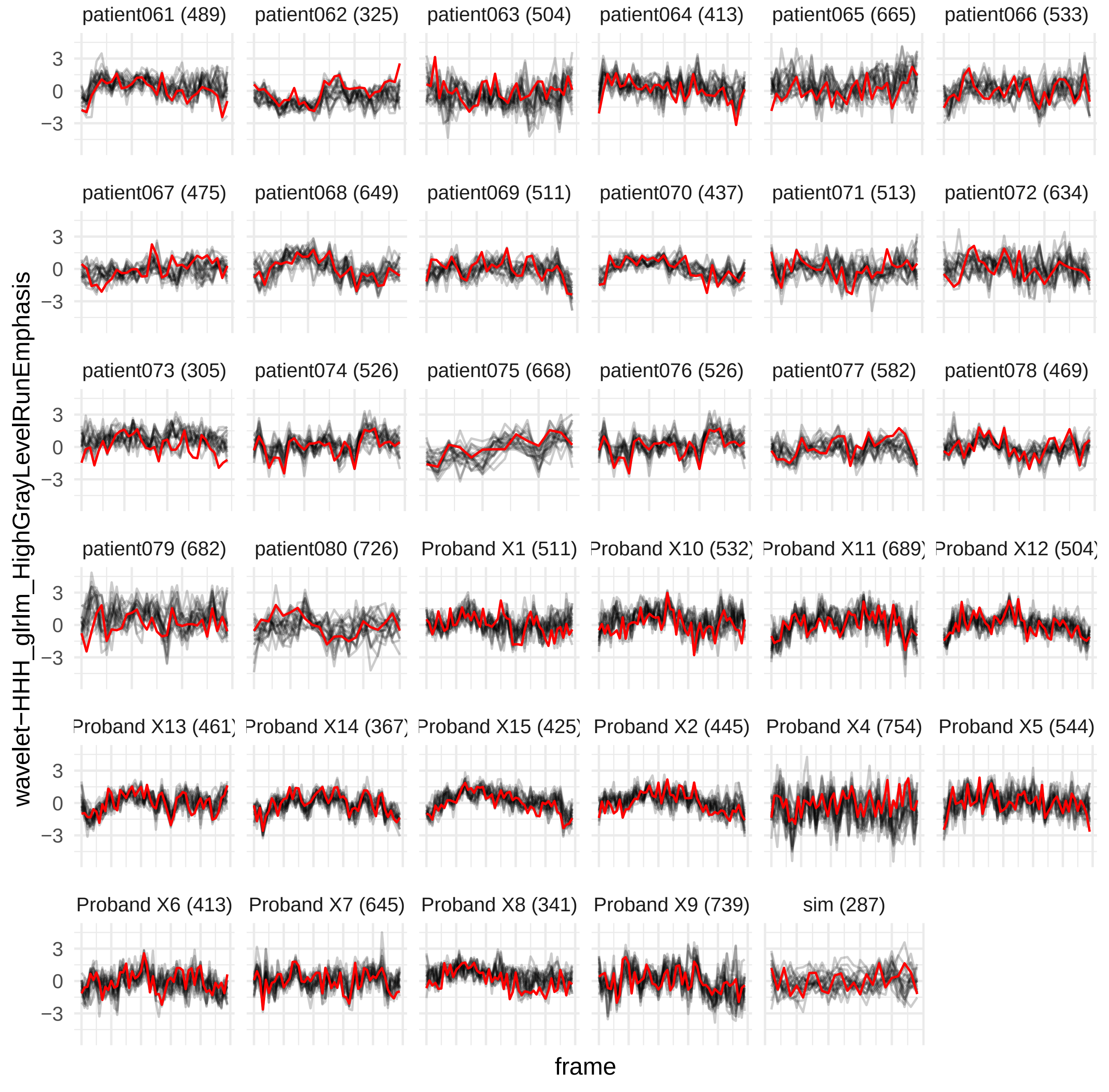

patient061 (489)
patient062 (325)
patient063 (504)
patient064 (413)
patient065 (665)
patient066 (533)
patient067 (475)
patient068 (649)
patient069 (511)
patient070 (437)
patient071 (513)
patient072 (634)
patient073 (305)
patient074 (526)
patient075 (668)
patient076 (526)
patient077 (582)
patient078 (469)
patient079 (682)
patient080 (726)
Proband X1 (511)
Proband X10 (532)
Proband X11 (689)
Proband X12 (504)
Proband X13 (461)
Proband X14 (367)
Proband X15 (425)
Proband X2 (445)
Proband X4 (754)
Proband X5 (544)
Proband X6 (413)
Proband X7 (645)
Proband X8 (341)
Proband X9 (739)
sim (287)
3
0
−3
wavelet−HHH_glrlm_HighGrayLevelRunEmphasis
frame

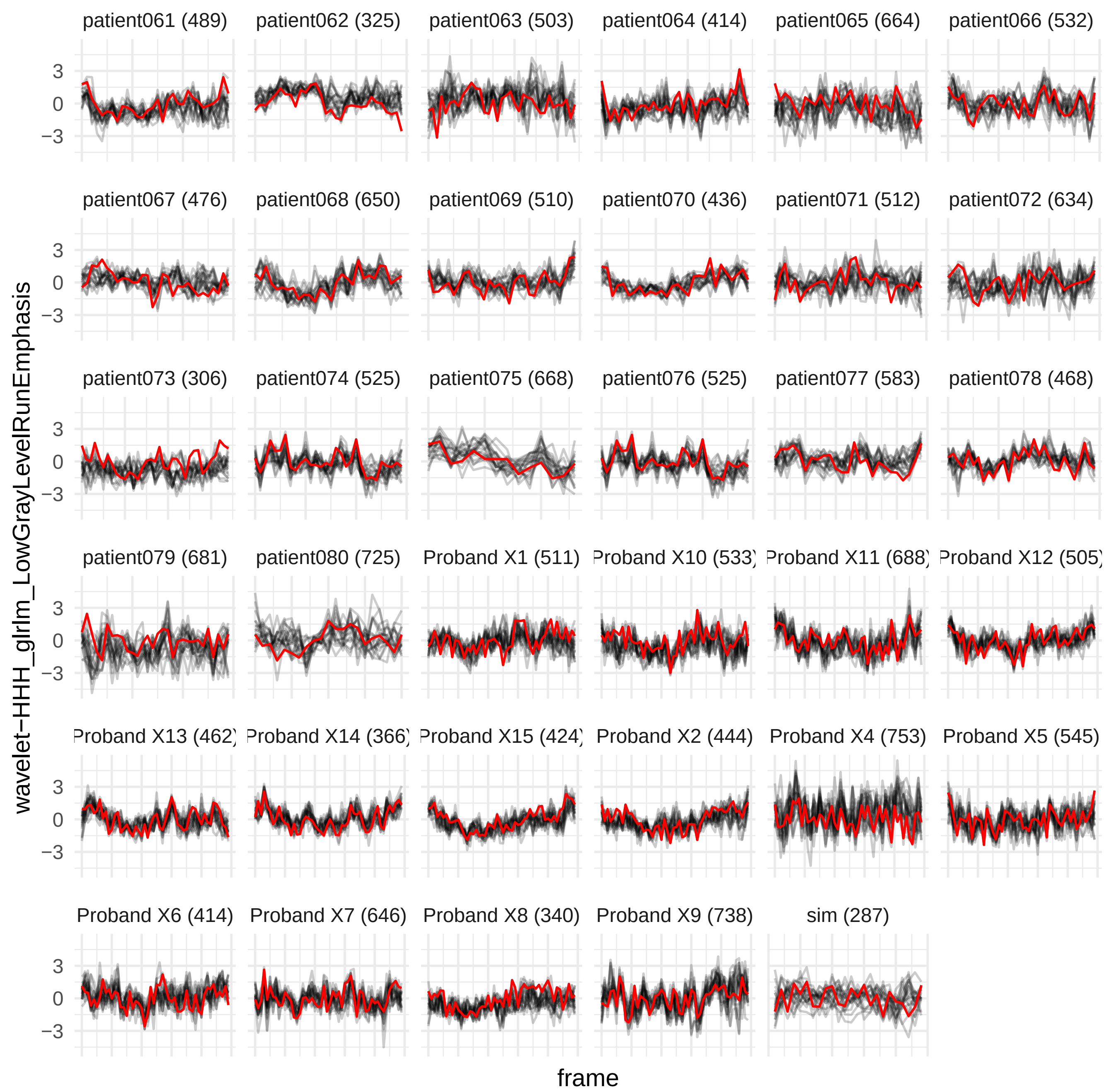
patient061 (489)
patient062 (325)
patient063 (503)
patient064 (414)
patient065 (664)
patient066 (532)
patient067 (476)
patient068 (650)
patient069 (510)
patient070 (436)
patient071 (512)
patient072 (634)
patient073 (306)
patient074 (525)
patient075 (668)
patient076 (525)
patient077 (583)
patient078 (468)
patient079 (681)
patient080 (725)
Proband X1 (511)
Proband X10 (533)
Proband X11 (688)
Proband X12 (505)
Proband X13 (462)
Proband X14 (366)
Proband X15 (424)
Proband X2 (444)
Proband X4 (753)
Proband X5 (545)
Proband X6 (414)
Proband X7 (646)
Proband X8 (340)
Proband X9 (738)
sim (287)
3
0
−3
wavelet−HHH_glrlm_LowGrayLevelRunEmphasis
frame

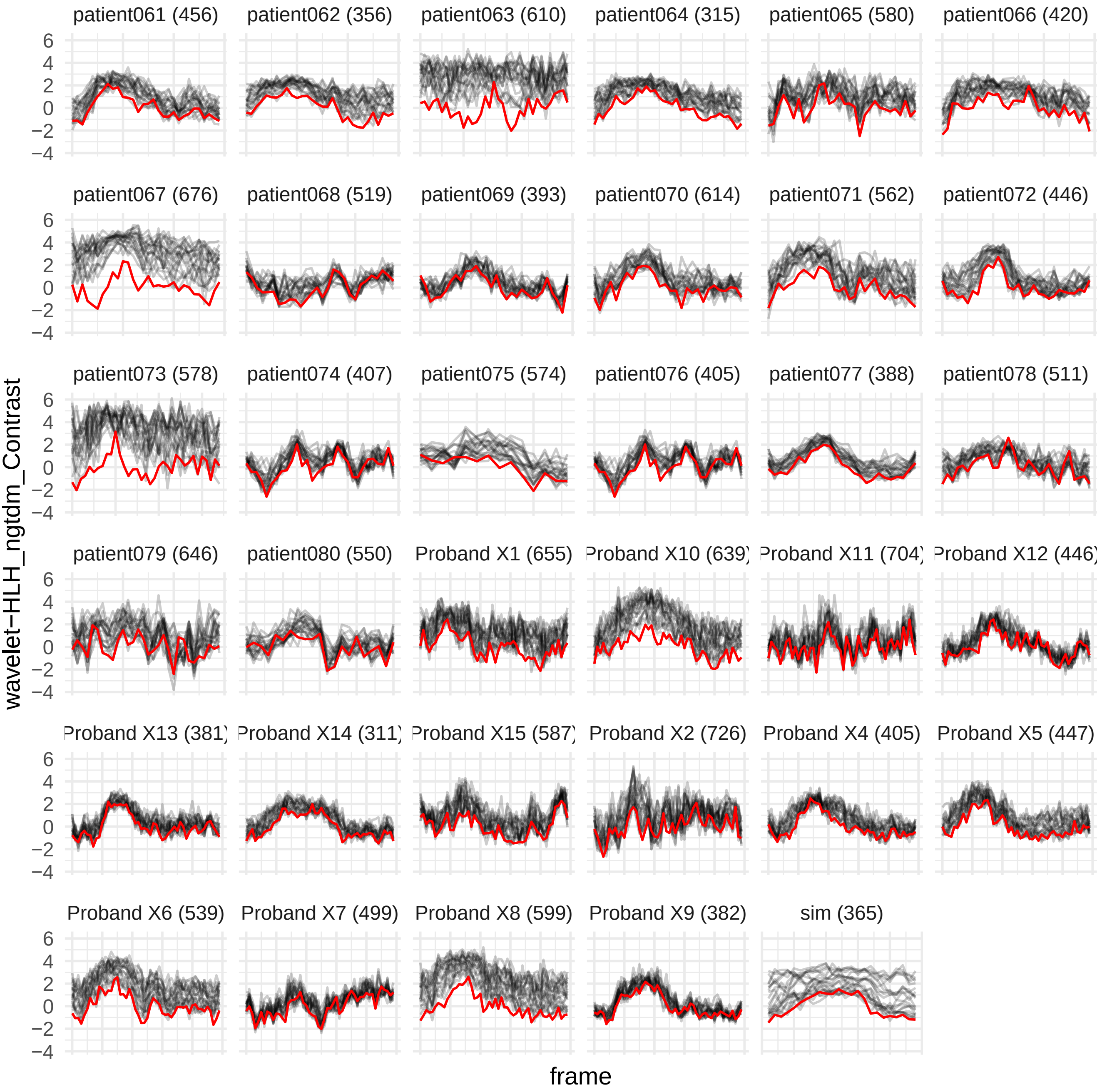
patient061 (456)
patient062 (356)
patient063 (610)
patient064 (315)
patient065 (580)
patient066 (420)
patient067 (676)
patient068 (519)
patient069 (393)
patient070 (614)
patient071 (562)
patient072 (446)
patient073 (578)
patient074 (407)
patient075 (574)
patient076 (405)
patient077 (388)
patient078 (511)
patient079 (646)
patient080 (550)
Proband X1 (655)
Proband X10 (639)
Proband X11 (704)
Proband X12 (446)
Proband X13 (381)
Proband X14 (311)
Proband X15 (587)
Proband X2 (726)
Proband X4 (405)
Proband X5 (447)
Proband X6 (539)
Proband X7 (499)
Proband X8 (599)
Proband X9 (382)
sim (365)
wavelet−HLH_ngtdm_Contrast
frame
6
4
2
0
−2
−4

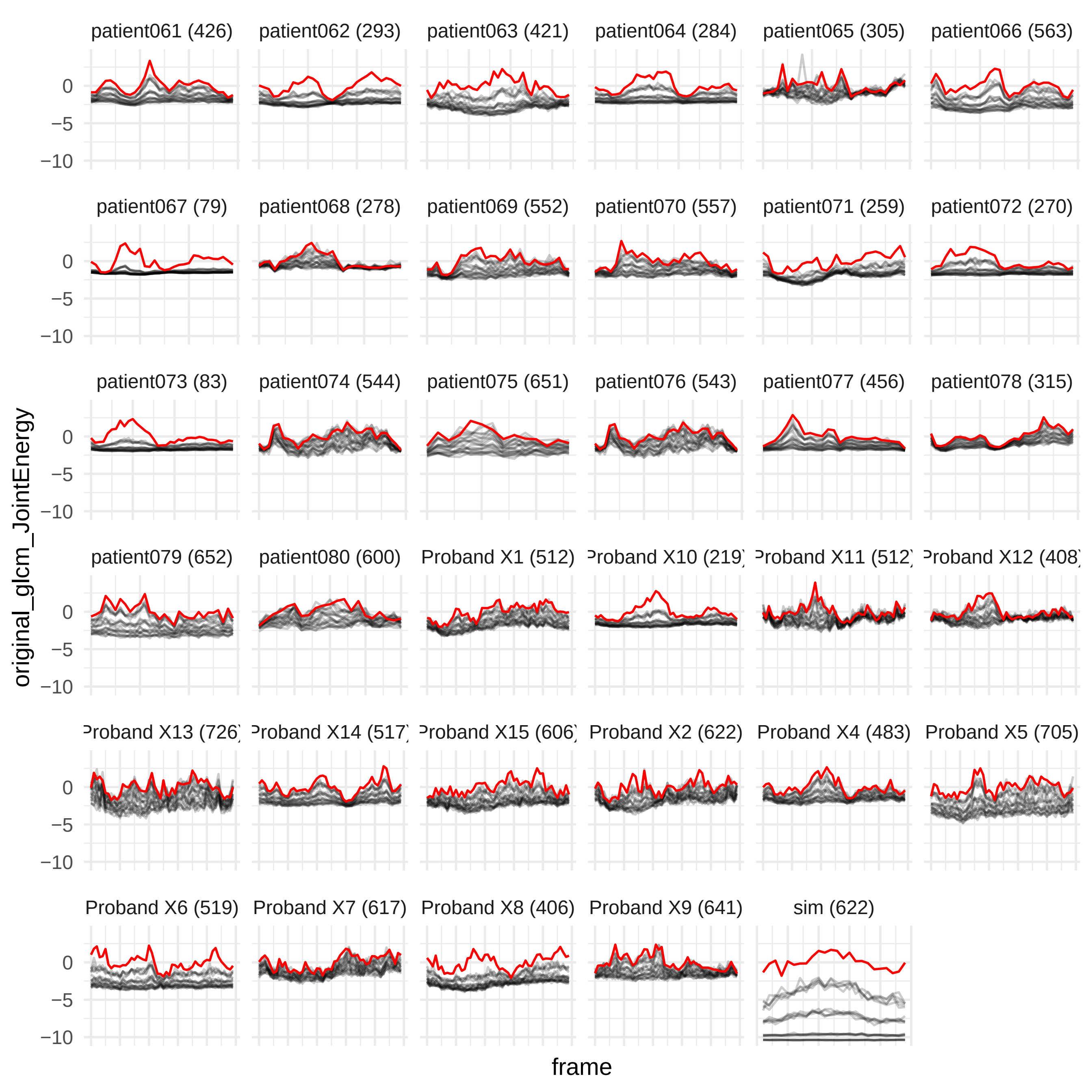
patient061 (426)
patient062 (293)
patient063 (421)
patient064 (284)
patient065 (305)
patient066 (563)
patient067 (79)
patient068 (278)
patient069 (552)
patient070 (557)
patient071 (259)
patient072 (270)
patient073 (83)
patient074 (544)
patient075 (651)
patient076 (543)
patient077 (456)
patient078 (315)
patient079 (652)
patient080 (600)
Proband X1 (512)
Proband X10 (219)
Proband X11 (512)
Proband X12 (408)
Proband X13 (726)
Proband X14 (517)
Proband X15 (606)
Proband X2 (622)
Proband X4 (483)
Proband X5 (705)
Proband X6 (519)
Proband X7 (617)
Proband X8 (406)
Proband X9 (641)
sim (622)
0
−5
−10
original_glcm_JointEnergy
frame

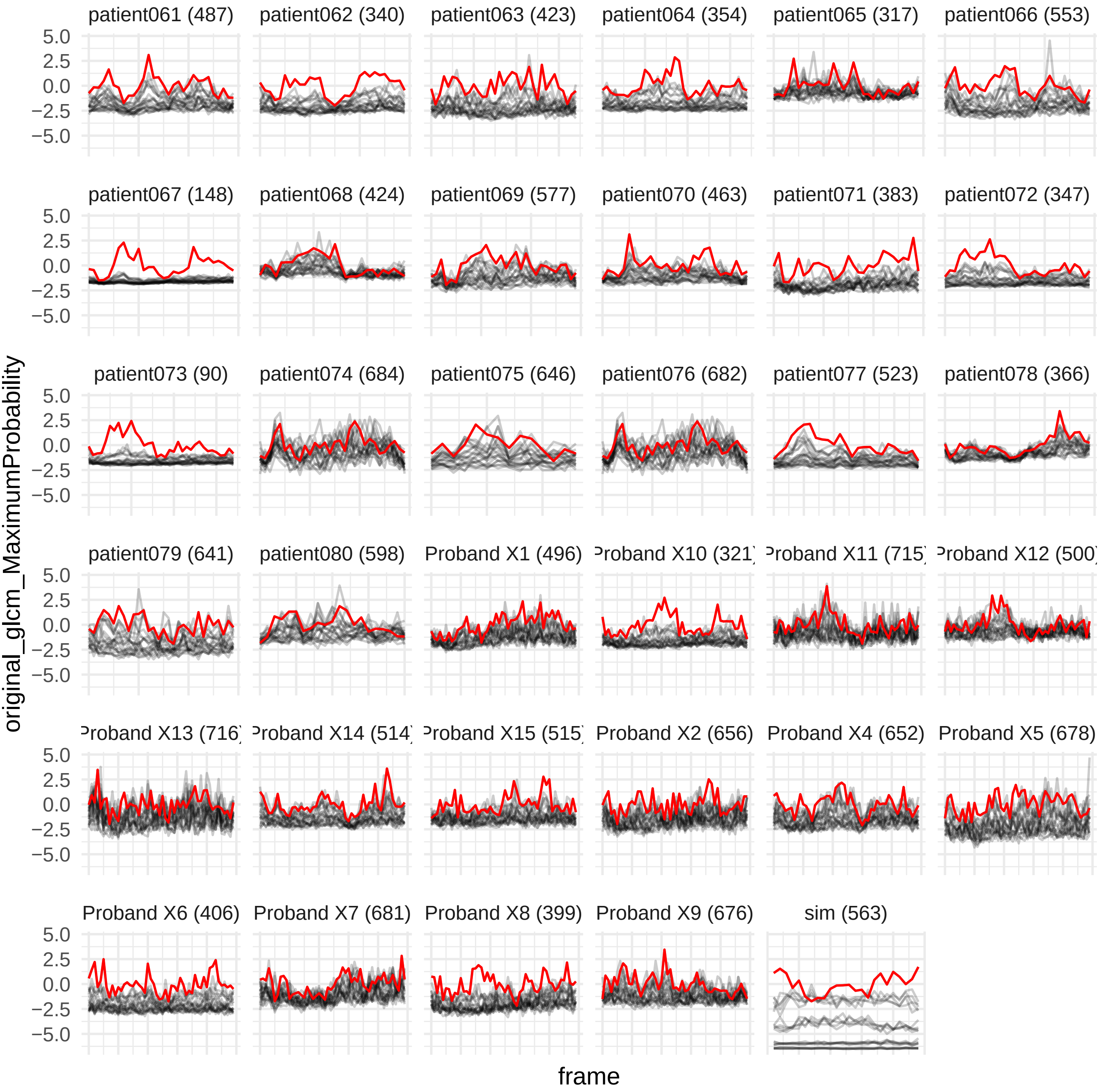

patient061 (487)
patient062 (340)
patient063 (423)
patient064 (354)
patient065 (317)
patient066 (553)
patient067 (148)
patient068 (424)
patient069 (577)
patient070 (463)
patient071 (383)
patient072 (347)
patient073 (90)
patient074 (684)
patient075 (646)
patient076 (682)
patient077 (523)
patient078 (366)
patient079 (641)
patient080 (598)
Proband X1 (496)
Proband X10 (321)
Proband X11 (715)
Proband X12 (500)
Proband X13 (716)
Proband X14 (514)
Proband X15 (515)
Proband X2 (656)
Proband X4 (652)
Proband X5 (678)
Proband X6 (406)
Proband X7 (681)
Proband X8 (399)
Proband X9 (676)
sim (563)
5.0
2.5
0.0
−2.5
−5.0
original_glcm_MaximumProbability
frame

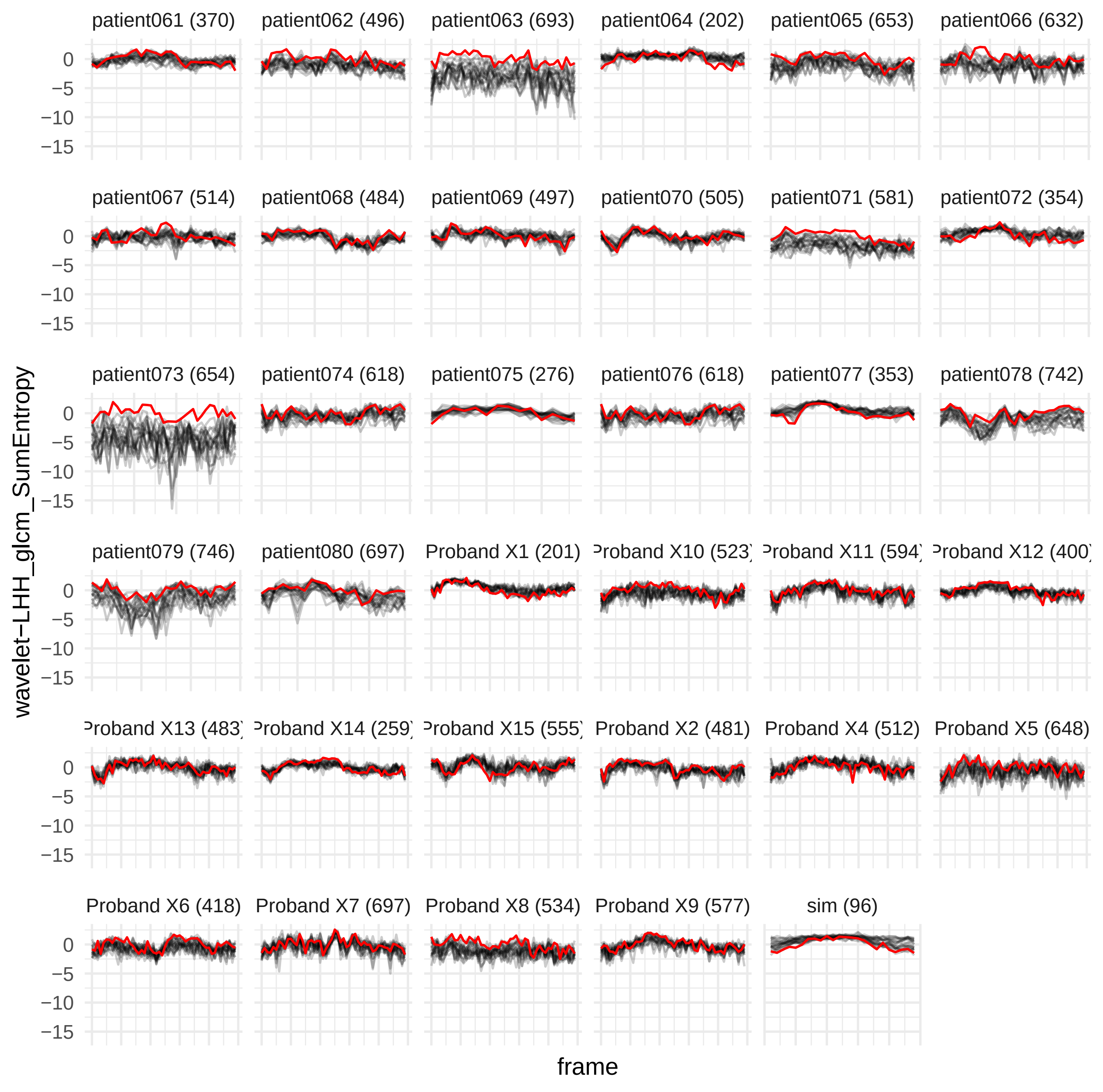

patient061 (370)
patient062 (496)
patient063 (693)
patient064 (202)
patient065 (653)
patient066 (632)
patient067 (514)
patient068 (484)
patient069 (497)
patient070 (505)
patient071 (581)
patient072 (354)
patient073 (654)
patient074 (618)
patient075 (276)
patient076 (618)
patient077 (353)
patient078 (742)
patient079 (746)
patient080 (697)
Proband X1 (201)
Proband X10 (523)
Proband X11 (594)
Proband X12 (400)
Proband X13 (483)
Proband X14 (259)
Proband X15 (555)
Proband X2 (481)
Proband X4 (512)
Proband X5 (648)
Proband X6 (418)
Proband X7 (697)
Proband X8 (534)
Proband X9 (577)
sim (96)
0
−5
−10
−15
wavelet−LHH_glcm_SumEntropy
frame

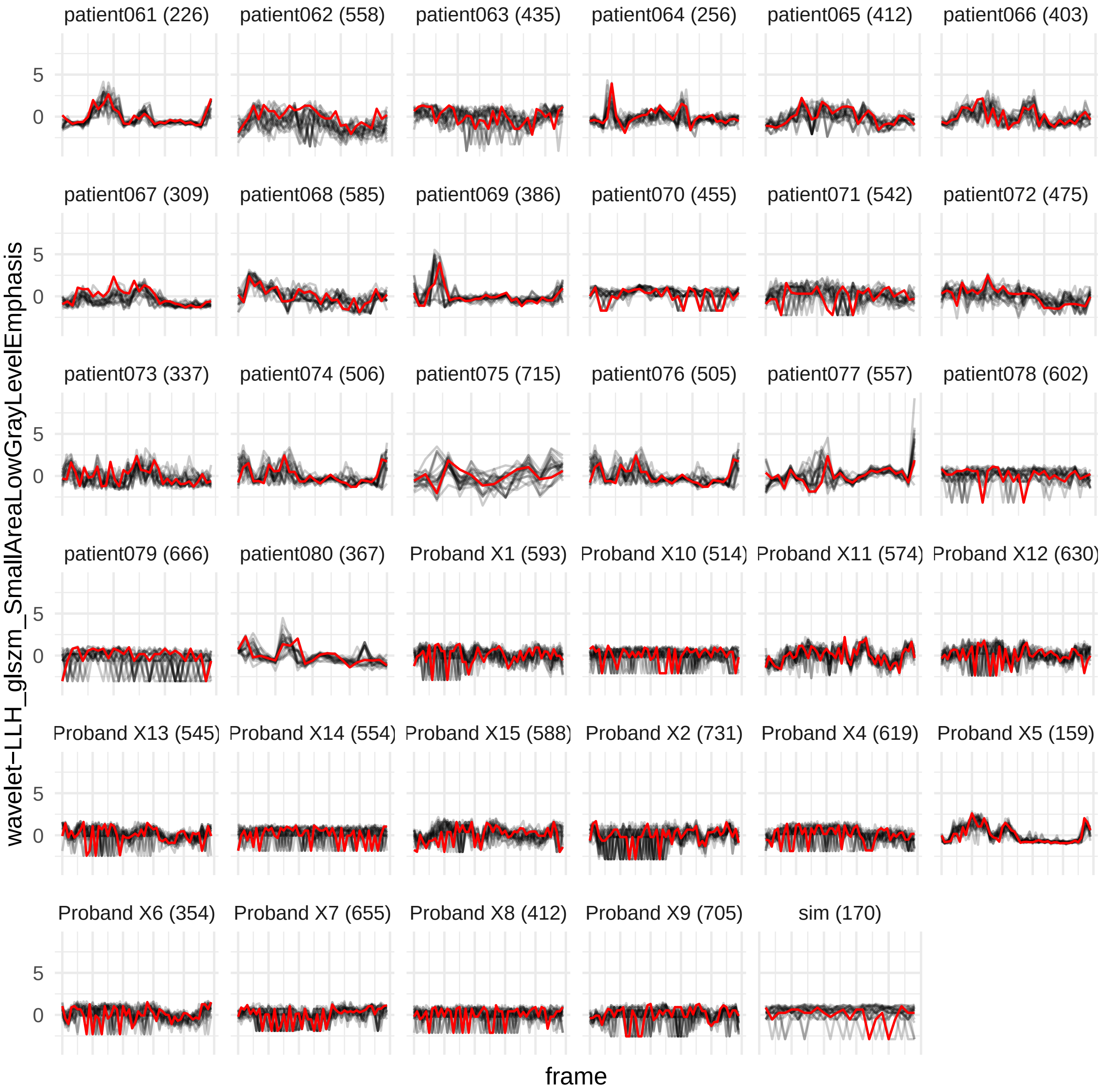

patient061 (226)
patient062 (558)
patient063 (435)
patient064 (256)
patient065 (412)
patient066 (403)
patient067 (309)
patient068 (585)
patient069 (386)
patient070 (455)
patient071 (542)
patient072 (475)
patient073 (337)
patient074 (506)
patient075 (715)
patient076 (505)
patient077 (557)
patient078 (602)
patient079 (666)
patient080 (367)
Proband X1 (593)
Proband X10 (514)
Proband X11 (574)
Proband X12 (630)
Proband X13 (545)
Proband X14 (554)
Proband X15 (588)
Proband X2 (731)
Proband X4 (619)
Proband X5 (159)
Proband X6 (354)
Proband X7 (655)
Proband X8 (412)
Proband X9 (705)
sim (170)
5
0
wavelet−LLH_glszm_SmallAreaLowGrayLevelEmphasis
frame

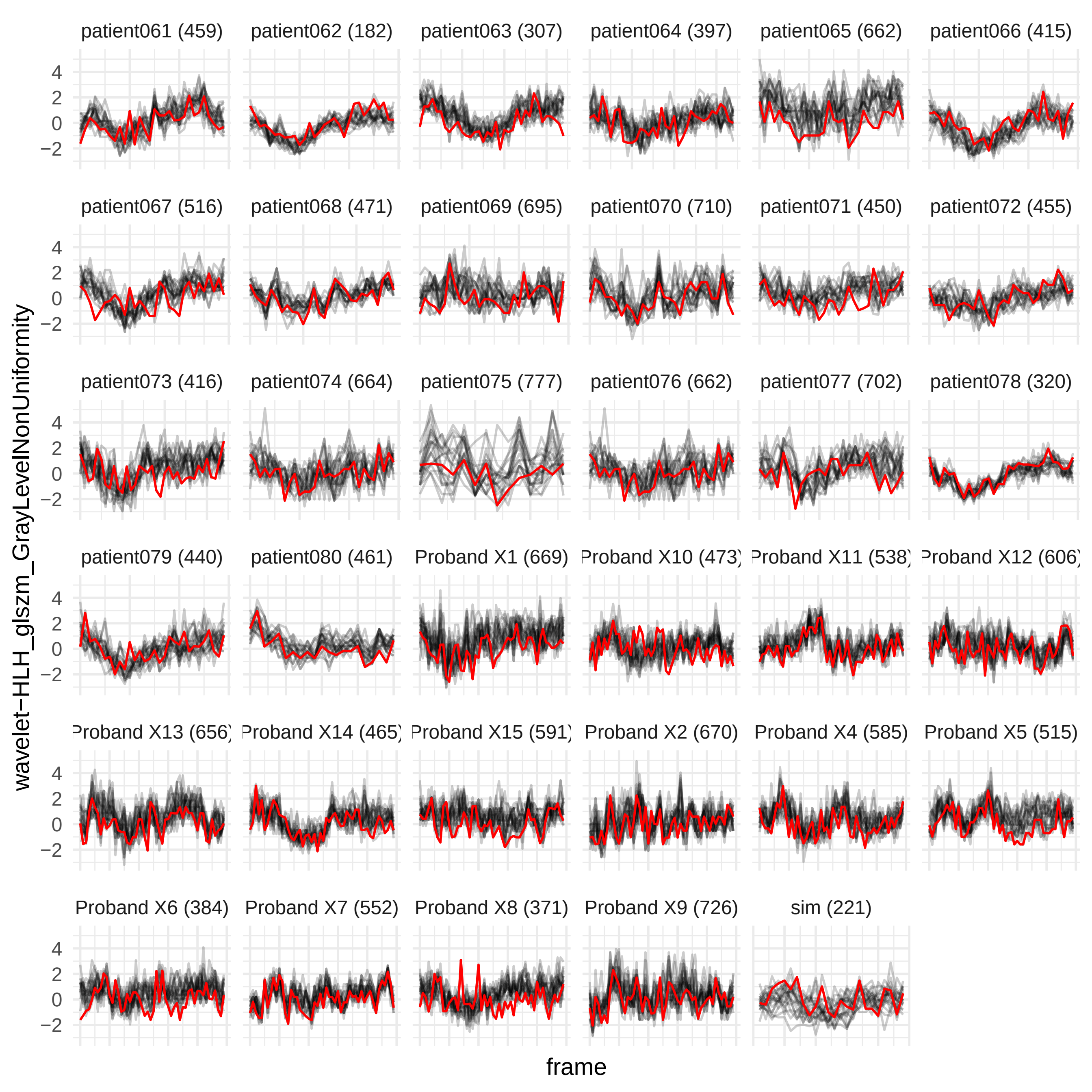
patient061 (459)
patient062 (182)
patient063 (307)
patient064 (397)
patient065 (662)
patient066 (415)
patient067 (516)
patient068 (471)
patient069 (695)
patient070 (710)
patient071 (450)
patient072 (455)
patient073 (416)
patient074 (664)
patient075 (777)
patient076 (662)
patient077 (702)
patient078 (320)
patient079 (440)
patient080 (461)
Proband X1 (669)
Proband X10 (473)
Proband X11 (538)
Proband X12 (606)
Proband X13 (656)
Proband X14 (465)
Proband X15 (591)
Proband X2 (670)
Proband X4 (585)
Proband X5 (515)
Proband X6 (384)
Proband X7 (552)
Proband X8 (371)
Proband X9 (726)
sim (221)
4
2
0
−2
wavelet−HLH_glszm_GrayLevelNonUniformity
frame

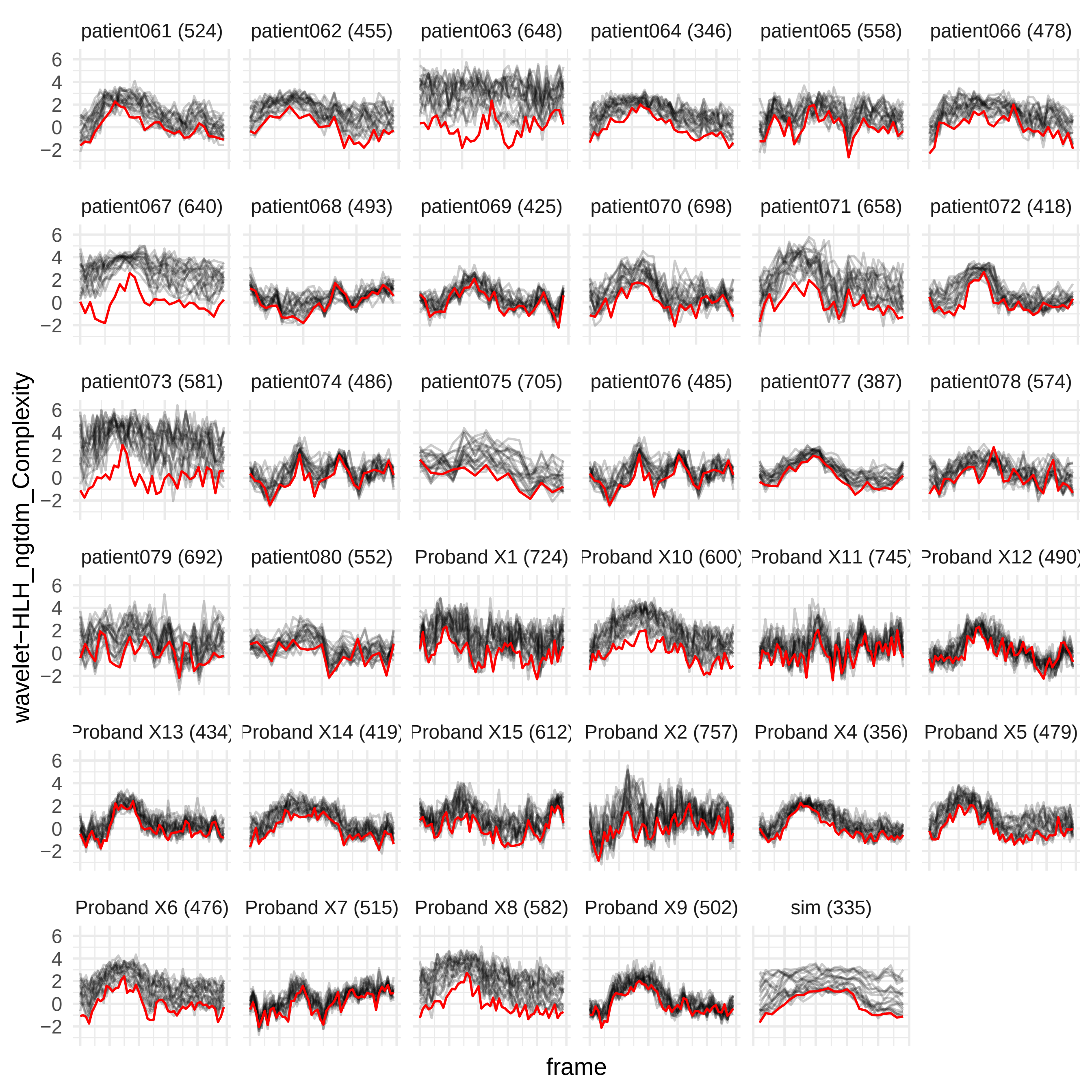

patient061 (524)
patient062 (455)
patient063 (648)
patient064 (346)
patient065 (558)
patient066 (478)
patient067 (640)
patient068 (493)
patient069 (425)
patient070 (698)
patient071 (658)
patient072 (418)
patient073 (581)
patient074 (486)
patient075 (705)
patient076 (485)
patient077 (387)
patient078 (574)
patient079 (692)
patient080 (552)
Proband X1 (724)
Proband X10 (600)
Proband X11 (745)
Proband X12 (490)
Proband X13 (434)
Proband X14 (419)
Proband X15 (612)
Proband X2 (757)
Proband X4 (356)
Proband X5 (479)
Proband X6 (476)
Proband X7 (515)
Proband X8 (582)
Proband X9 (502)
sim (335)
wavelet−HLH_ngtdm_Complexity
frame
6
4
2
0
−2

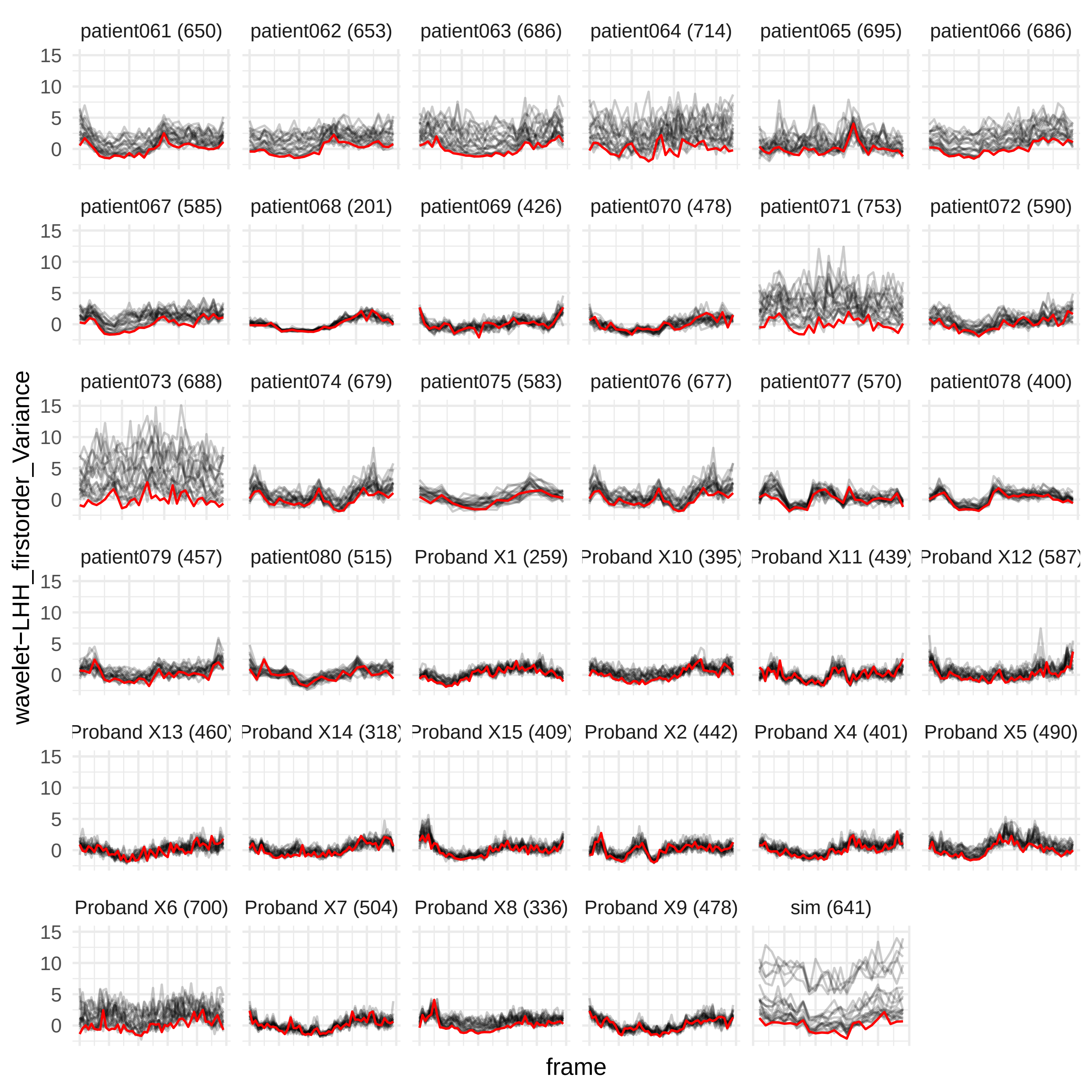
patient061 (650)
patient062 (653)
patient063 (686)
patient064 (714)
patient065 (695)
patient066 (686)
patient067 (585)
patient068 (201)
patient069 (426)
patient070 (478)
patient071 (753)
patient072 (590)
patient073 (688)
patient074 (679)
patient075 (583)
patient076 (677)
patient077 (570)
patient078 (400)
patient079 (457)
patient080 (515)
Proband X1 (259)
Proband X10 (395)
Proband X11 (439)
Proband X12 (587)
Proband X13 (460)
Proband X14 (318)
Proband X15 (409)
Proband X2 (442)
Proband X4 (401)
Proband X5 (490)
Proband X6 (700)
Proband X7 (504)
Proband X8 (336)
Proband X9 (478)
sim (641)
wavelet−LHH_firstorder_Variance
frame
0
5
10
15

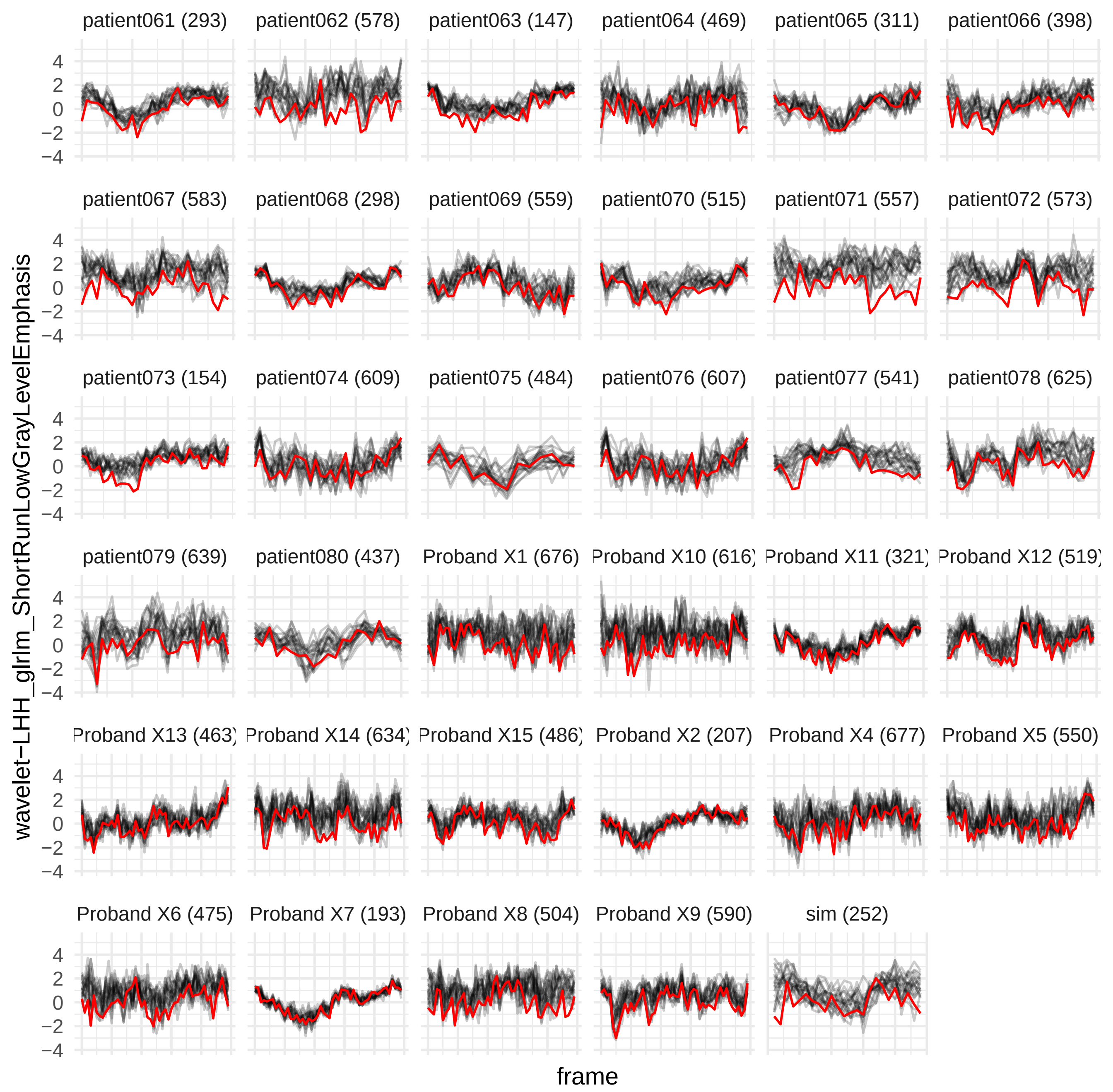

patient061 (293)
patient062 (578)
patient063 (147)
patient064 (469)
patient065 (311)
patient066 (398)
patient067 (583)
patient068 (298)
patient069 (559)
patient070 (515)
patient071 (557)
patient072 (573)
patient073 (154)
patient074 (609)
patient075 (484)
patient076 (607)
patient077 (541)
patient078 (625)
patient079 (639)
patient080 (437)
Proband X1 (676)
Proband X10 (616)
Proband X11 (321)
Proband X12 (519)
Proband X13 (463)
Proband X14 (634)
Proband X15 (486)
Proband X2 (207)
Proband X4 (677)
Proband X5 (550)
Proband X6 (475)
Proband X7 (193)
Proband X8 (504)
Proband X9 (590)
sim (252)
4
2
0
−2
−4
wavelet−LHH_glrlm_ShortRunLowGrayLevelEmphasis
frame

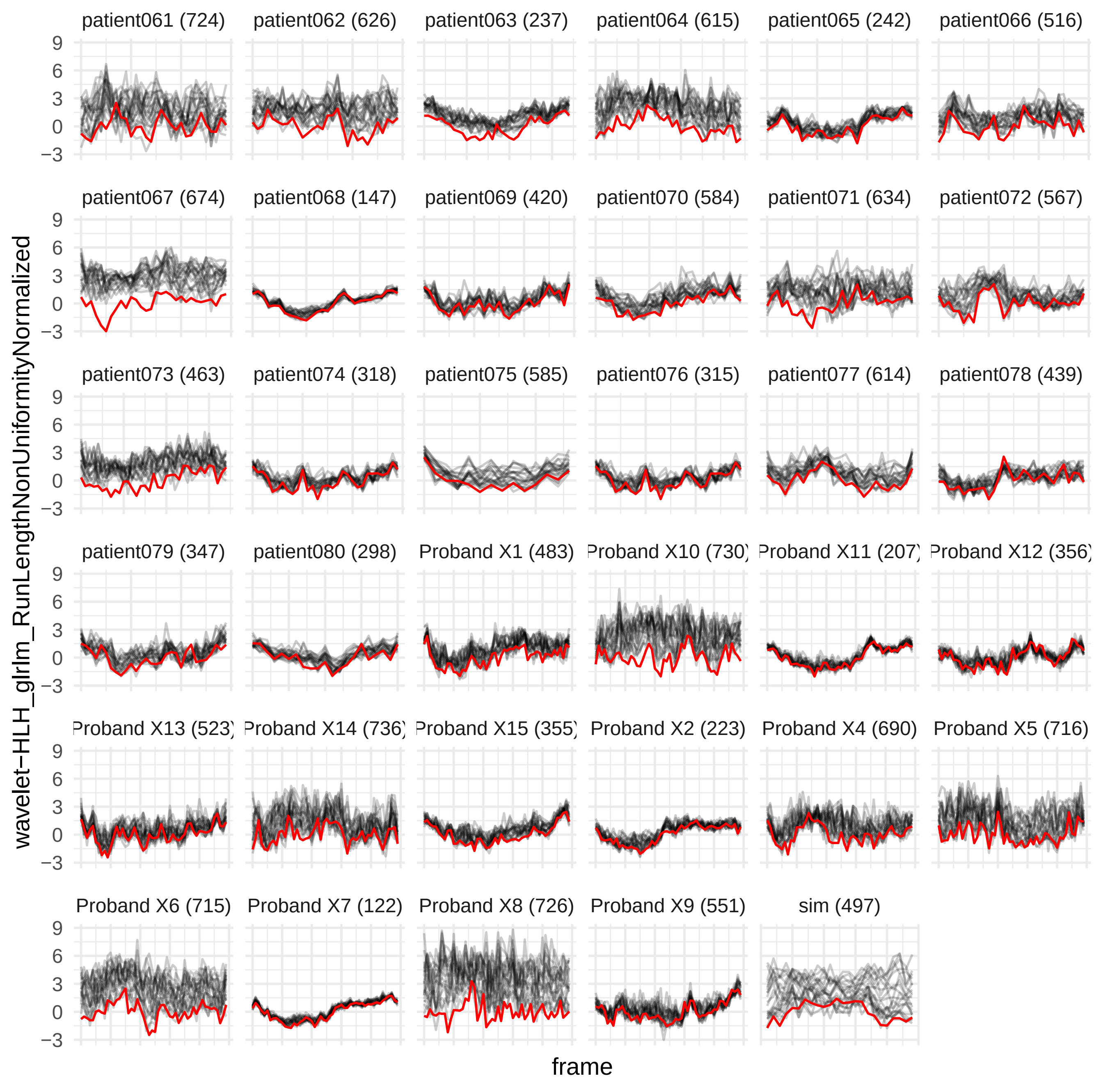
patient061 (724)
patient062 (626)
patient063 (237)
patient064 (615)
patient065 (242)
patient066 (516)
patient067 (674)
patient068 (147)
patient069 (420)
patient070 (584)
patient071 (634)
patient072 (567)
patient073 (463)
patient074 (318)
patient075 (585)
patient076 (315)
patient077 (614)
patient078 (439)
patient079 (347)
patient080 (298)
Proband X1 (483)
Proband X10 (730)
Proband X11 (207)
Proband X12 (356)
Proband X13 (523)
Proband X14 (736)
Proband X15 (355)
Proband X2 (223)
Proband X4 (690)
Proband X5 (716)
Proband X6 (715)
Proband X7 (122)
Proband X8 (726)
Proband X9 (551)
sim (497)
9
6
3
0
−3
wavelet−HLH_glrlm_RunLengthNonUniformityNormalized
frame

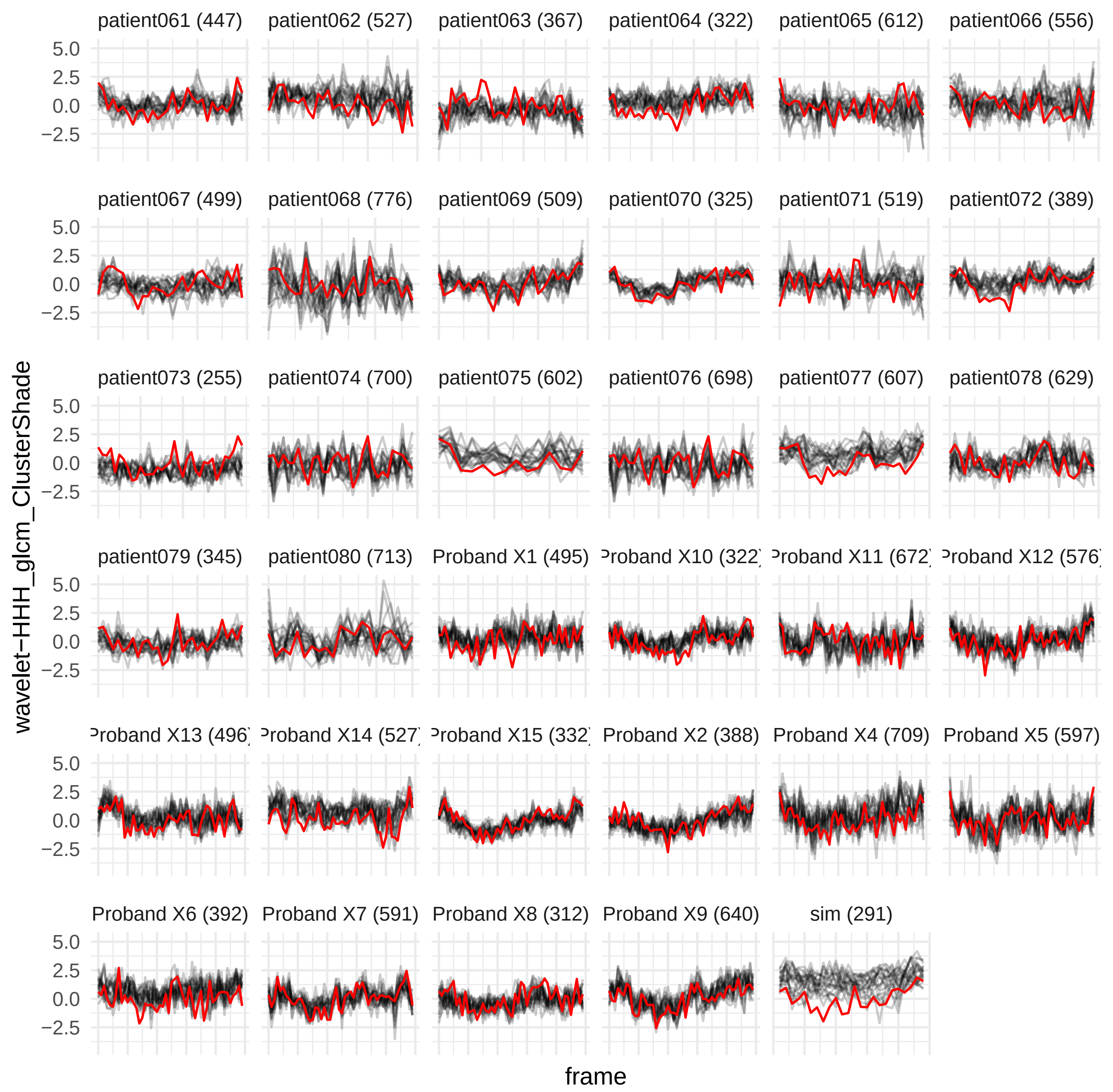

patient061 (447)
patient062 (527)
patient063 (367)
patient064 (322)
patient065 (612)
patient066 (556)
patient067 (499)
patient068 (776)
patient069 (509)
patient070 (325)
patient071 (519)
patient072 (389)
patient073 (255)
patient074 (700)
patient075 (602)
patient076 (698)
patient077 (607)
patient078 (629)
patient079 (345)
patient080 (713)
Proband X1 (495)
Proband X10 (322)
Proband X11 (672)
Proband X12 (576)
Proband X13 (496)
Proband X14 (527)
Proband X15 (332)
Proband X2 (388)
Proband X4 (709)
Proband X5 (597)
Proband X6 (392)
Proband X7 (591)
Proband X8 (312)
Proband X9 (640)
sim (291)
5.0
2.5
0.0
−2.5
wavelet−HHH_glcm_ClusterShade
frame

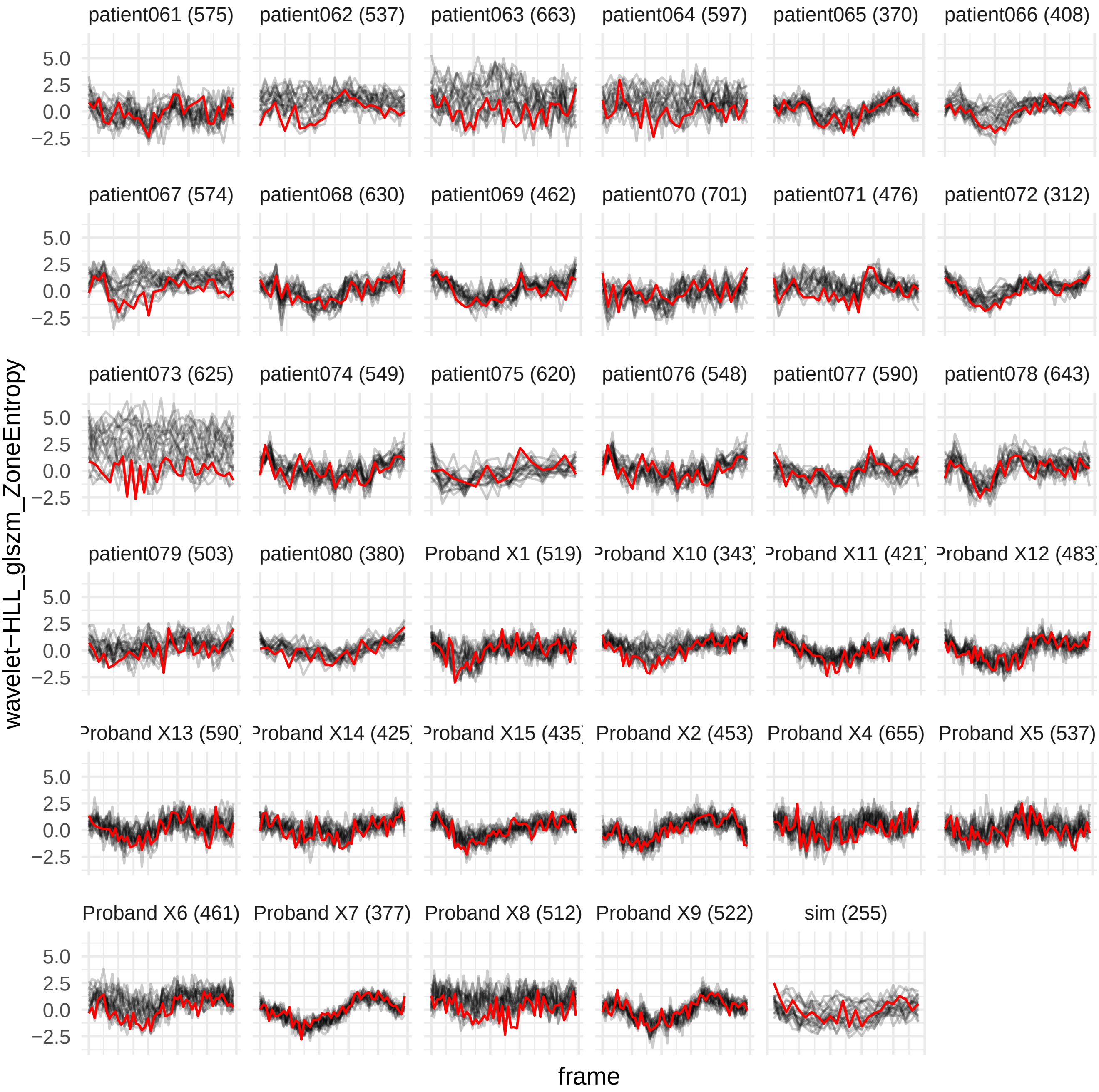

patient061 (575)
patient062 (537)
patient063 (663)
patient064 (597)
patient065 (370)
patient066 (408)
patient067 (574)
patient068 (630)
patient069 (462)
patient070 (701)
patient071 (476)
patient072 (312)
patient073 (625)
patient074 (549)
patient075 (620)
patient076 (548)
patient077 (590)
patient078 (643)
patient079 (503)
patient080 (380)
Proband X1 (519)
Proband X10 (343)
Proband X11 (421)
Proband X12 (483)
Proband X13 (590)
Proband X14 (425)
Proband X15 (435)
Proband X2 (453)
Proband X4 (655)
Proband X5 (537)
Proband X6 (461)
Proband X7 (377)
Proband X8 (512)
Proband X9 (522)
sim (255)
5.0
2.5
0.0
−2.5
wavelet−HLL_glszm_ZoneEntropy
frame

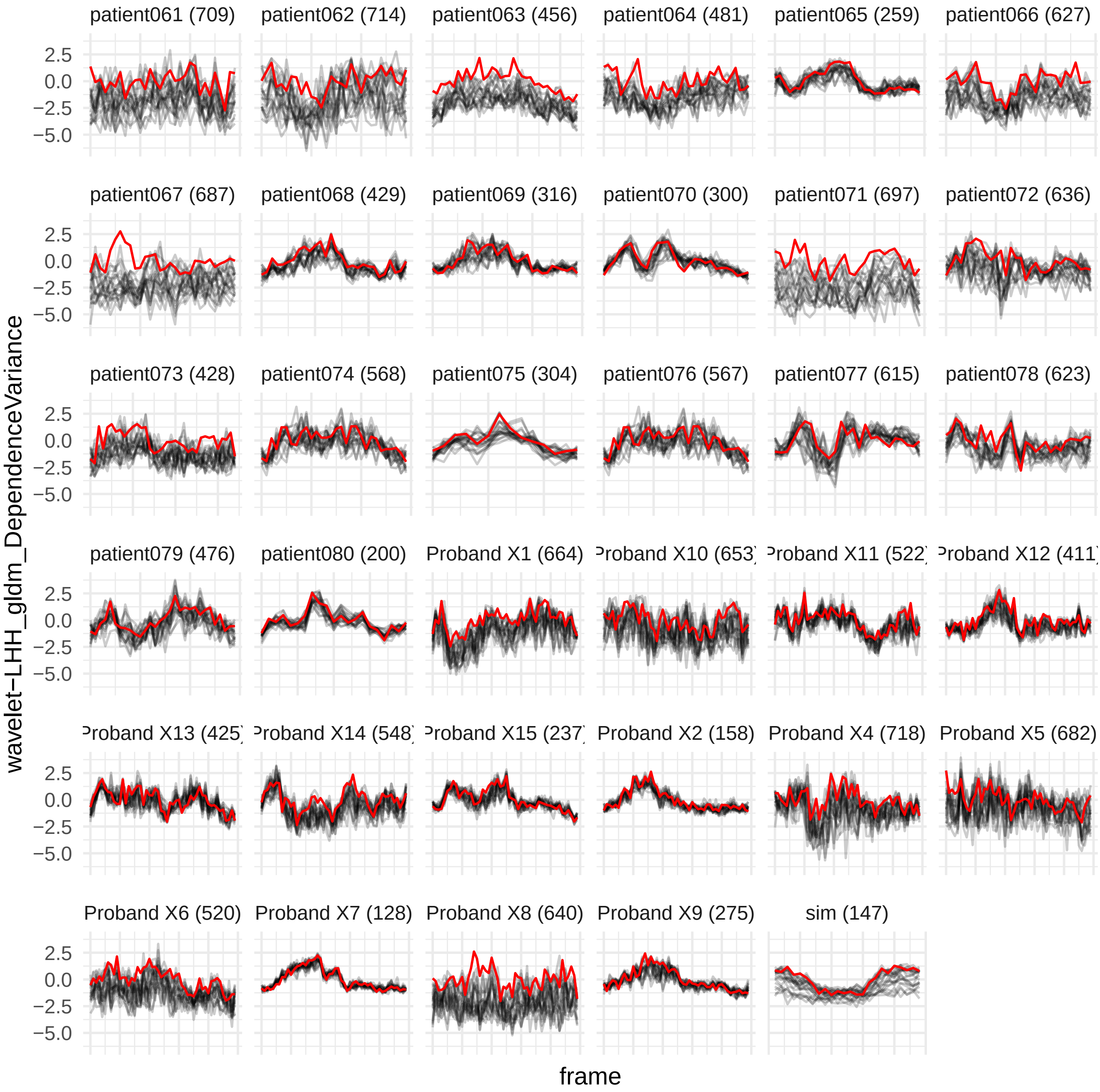

patient061 (709)
patient062 (714)
patient063 (456)
patient064 (481)
patient065 (259)
patient066 (627)
patient067 (687)
patient068 (429)
patient069 (316)
patient070 (300)
patient071 (697)
patient072 (636)
patient073 (428)
patient074 (568)
patient075 (304)
patient076 (567)
patient077 (615)
patient078 (623)
patient079 (476)
patient080 (200)
Proband X1 (664)
Proband X10 (653)
Proband X11 (522)
Proband X12 (411)
Proband X13 (425)
Proband X14 (548)
Proband X15 (237)
Proband X2 (158)
Proband X4 (718)
Proband X5 (682)
Proband X6 (520)
Proband X7 (128)
Proband X8 (640)
Proband X9 (275)
sim (147)
2.5
0.0
−2.5
−5.0
wavelet−LHH_gldm_DependenceVariance
frame

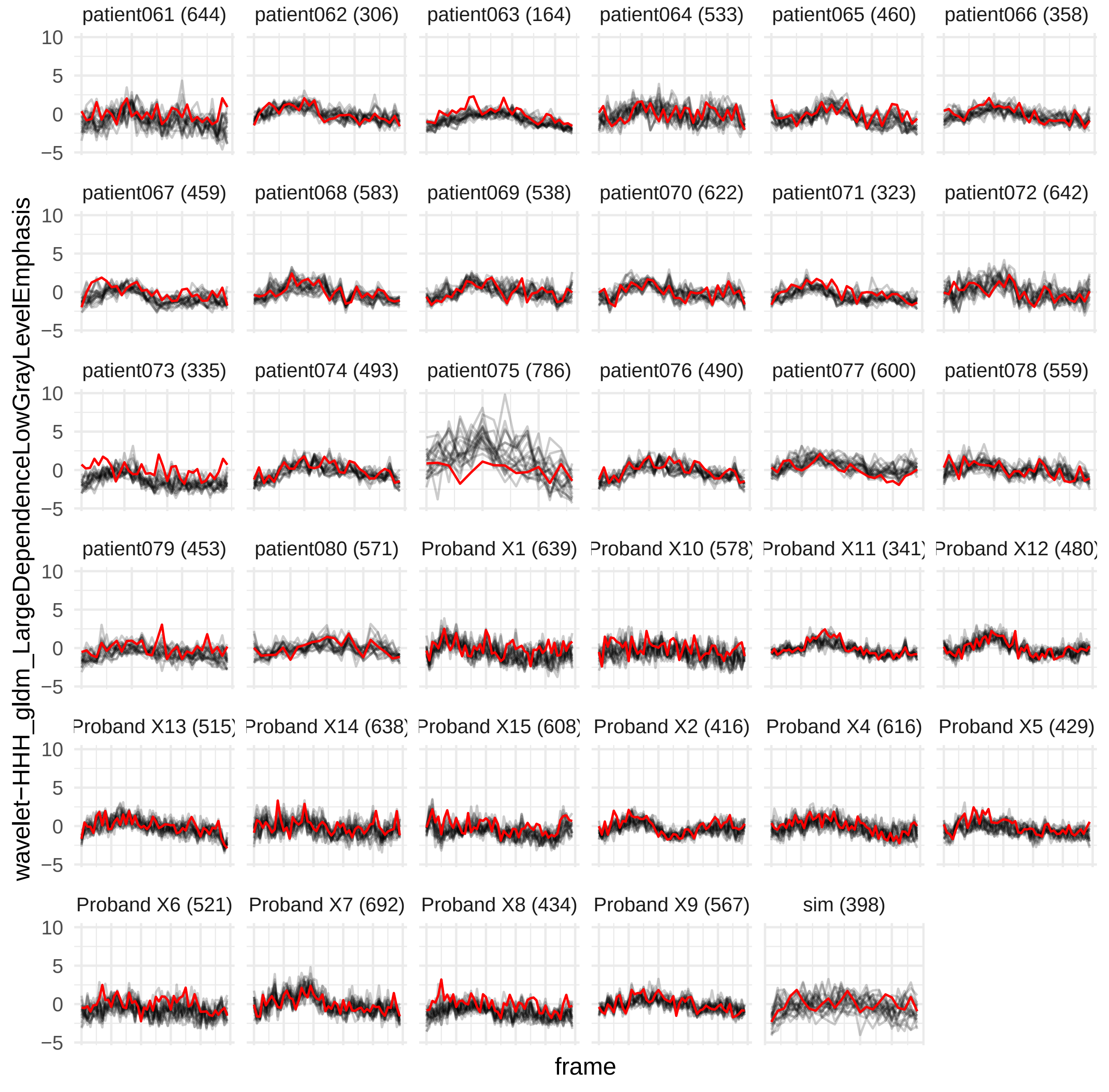

patient061 (644)
patient062 (306)
patient063 (164)
patient064 (533)
patient065 (460)
patient066 (358)
patient067 (459)
patient068 (583)
patient069 (538)
patient070 (622)
patient071 (323)
patient072 (642)
patient073 (335)
patient074 (493)
patient075 (786)
patient076 (490)
patient077 (600)
patient078 (559)
patient079 (453)
patient080 (571)
Proband X1 (639)
Proband X10 (578)
Proband X11 (341)
Proband X12 (480)
Proband X13 (515)
Proband X14 (638)
Proband X15 (608)
Proband X2 (416)
Proband X4 (616)
Proband X5 (429)
Proband X6 (521)
Proband X7 (692)
Proband X8 (434)
Proband X9 (567)
sim (398)
10
5
0
−5
wavelet−HHH_gldm_LargeDependenceLowGrayLevelEmphasis
frame

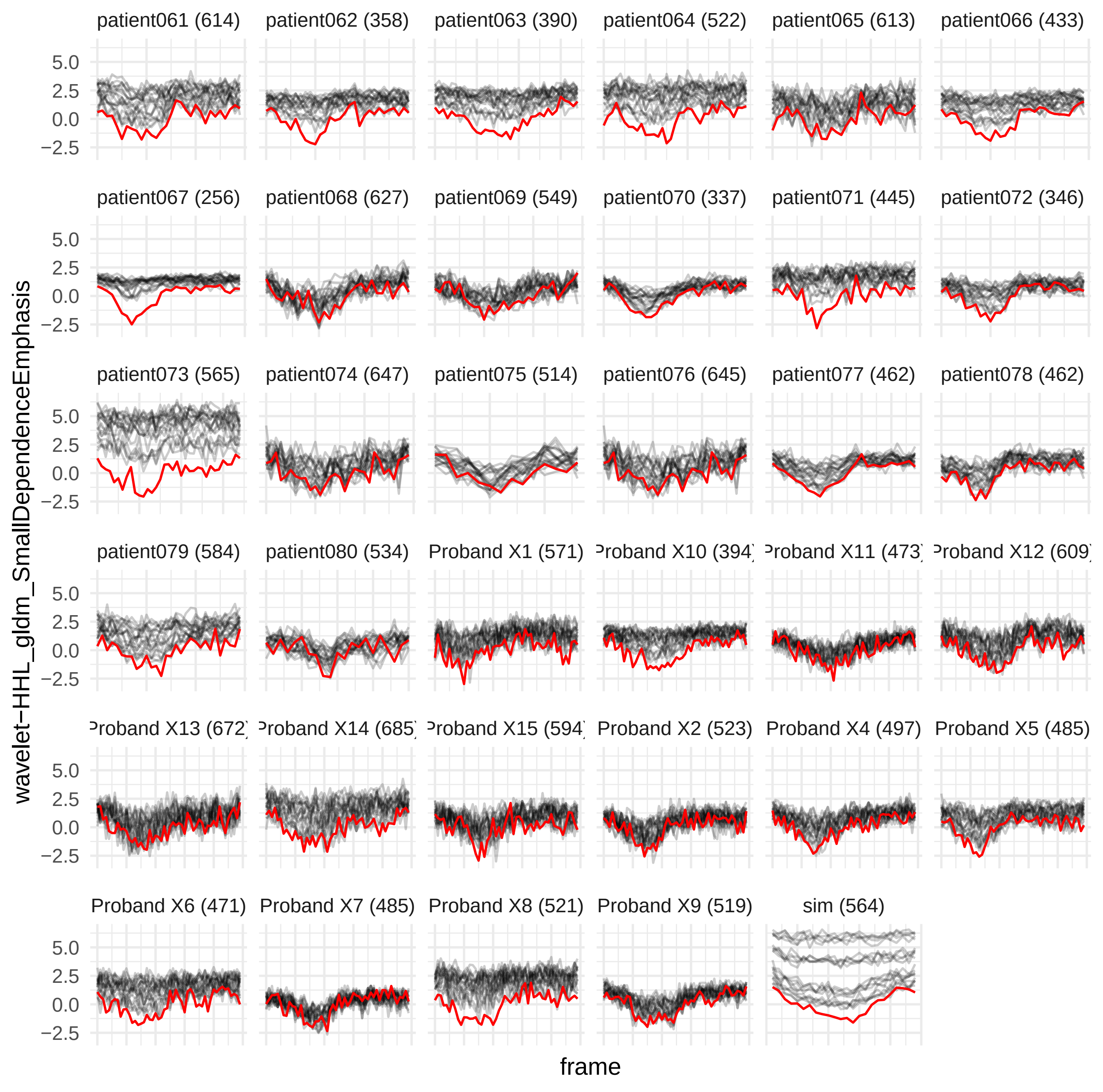

patient061 (614)
patient062 (358)
patient063 (390)
patient064 (522)
patient065 (613)
patient066 (433)
patient067 (256)
patient068 (627)
patient069 (549)
patient070 (337)
patient071 (445)
patient072 (346)
patient073 (565)
patient074 (647)
patient075 (514)
patient076 (645)
patient077 (462)
patient078 (462)
patient079 (584)
patient080 (534)
Proband X1 (571)
Proband X10 (394)
Proband X11 (473)
Proband X12 (609)
Proband X13 (672)
Proband X14 (685)
Proband X15 (594)
Proband X2 (523)
Proband X4 (497)
Proband X5 (485)
Proband X6 (471)
Proband X7 (485)
Proband X8 (521)
Proband X9 (519)
sim (564)
5.0
2.5
0.0
−2.5
wavelet−HHL_gldm_SmallDependenceEmphasis
frame

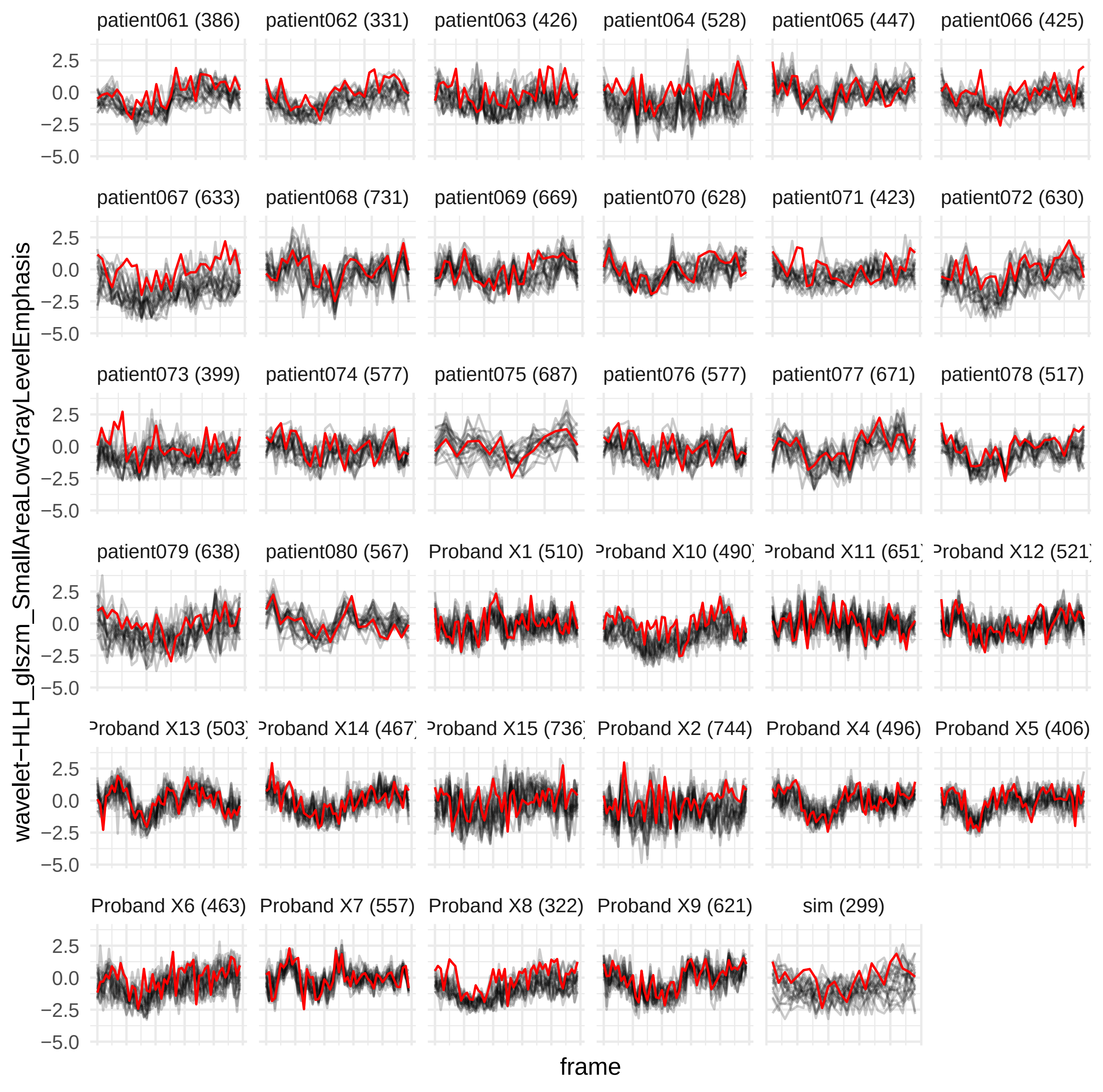
patient061 (386)
patient062 (331)
patient063 (426)
patient064 (528)
patient065 (447)
patient066 (425)
patient067 (633)
patient068 (731)
patient069 (669)
patient070 (628)
patient071 (423)
patient072 (630)
patient073 (399)
patient074 (577)
patient075 (687)
patient076 (577)
patient077 (671)
patient078 (517)
patient079 (638)
patient080 (567)
Proband X1 (510)
Proband X10 (490)
Proband X11 (651)
Proband X12 (521)
Proband X13 (503)
Proband X14 (467)
Proband X15 (736)
Proband X2 (744)
Proband X4 (496)
Proband X5 (406)
Proband X6 (463)
Proband X7 (557)
Proband X8 (322)
Proband X9 (621)
sim (299)
2.5
0.0
−2.5
−5.0
wavelet−HLH_glszm_SmallAreaLowGrayLevelEmphasis
frame

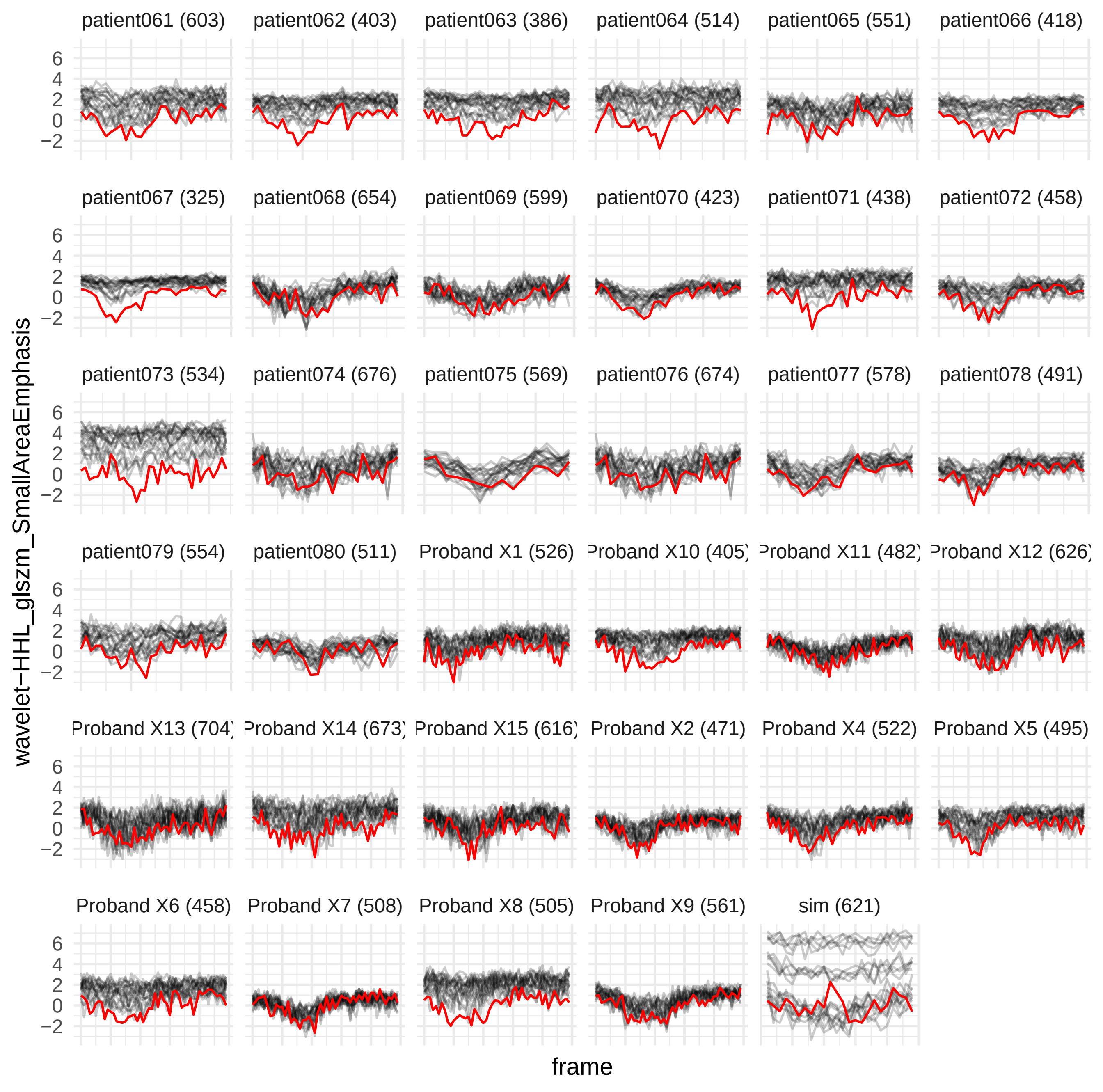
patient061 (603)
patient062 (403)
patient063 (386)
patient064 (514)
patient065 (551)
patient066 (418)
patient067 (325)
patient068 (654)
patient069 (599)
patient070 (423)
patient071 (438)
patient072 (458)
patient073 (534)
patient074 (676)
patient075 (569)
patient076 (674)
patient077 (578)
patient078 (491)
patient079 (554)
patient080 (511)
Proband X1 (526)
Proband X10 (405)
Proband X11 (482)
Proband X12 (626)
Proband X13 (704)
Proband X14 (673)
Proband X15 (616)
Proband X2 (471)
Proband X4 (522)
Proband X5 (495)
Proband X6 (458)
Proband X7 (508)
Proband X8 (505)
Proband X9 (561)
sim (621)
wavelet−HHL_glszm_SmallAreaEmphasis
frame

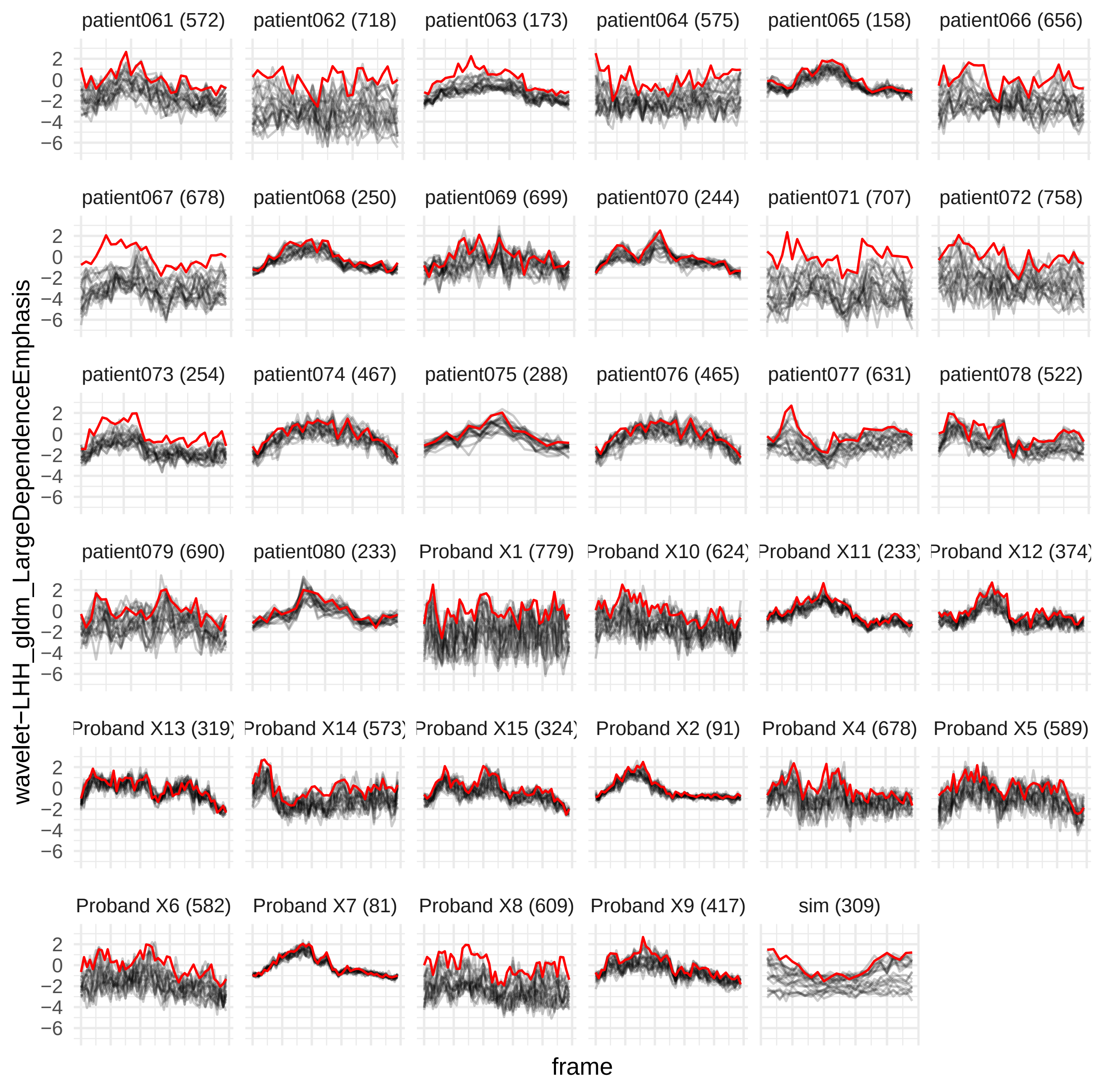

patient061 (572)
patient062 (718)
patient063 (173)
patient064 (575)
patient065 (158)
patient066 (656)
patient067 (678)
patient068 (250)
patient069 (699)
patient070 (244)
patient071 (707)
patient072 (758)
patient073 (254)
patient074 (467)
patient075 (288)
patient076 (465)
patient077 (631)
patient078 (522)
patient079 (690)
patient080 (233)
Proband X1 (779)
Proband X10 (624)
Proband X11 (233)
Proband X12 (374)
Proband X13 (319)
Proband X14 (573)
Proband X15 (324)
Proband X2 (91)
Proband X4 (678)
Proband X5 (589)
Proband X6 (582)
Proband X7 (81)
Proband X8 (609)
Proband X9 (417)
sim (309)
2
0
−2
−4
−6
wavelet−LHH_gldm_LargeDependenceEmphasis
frame

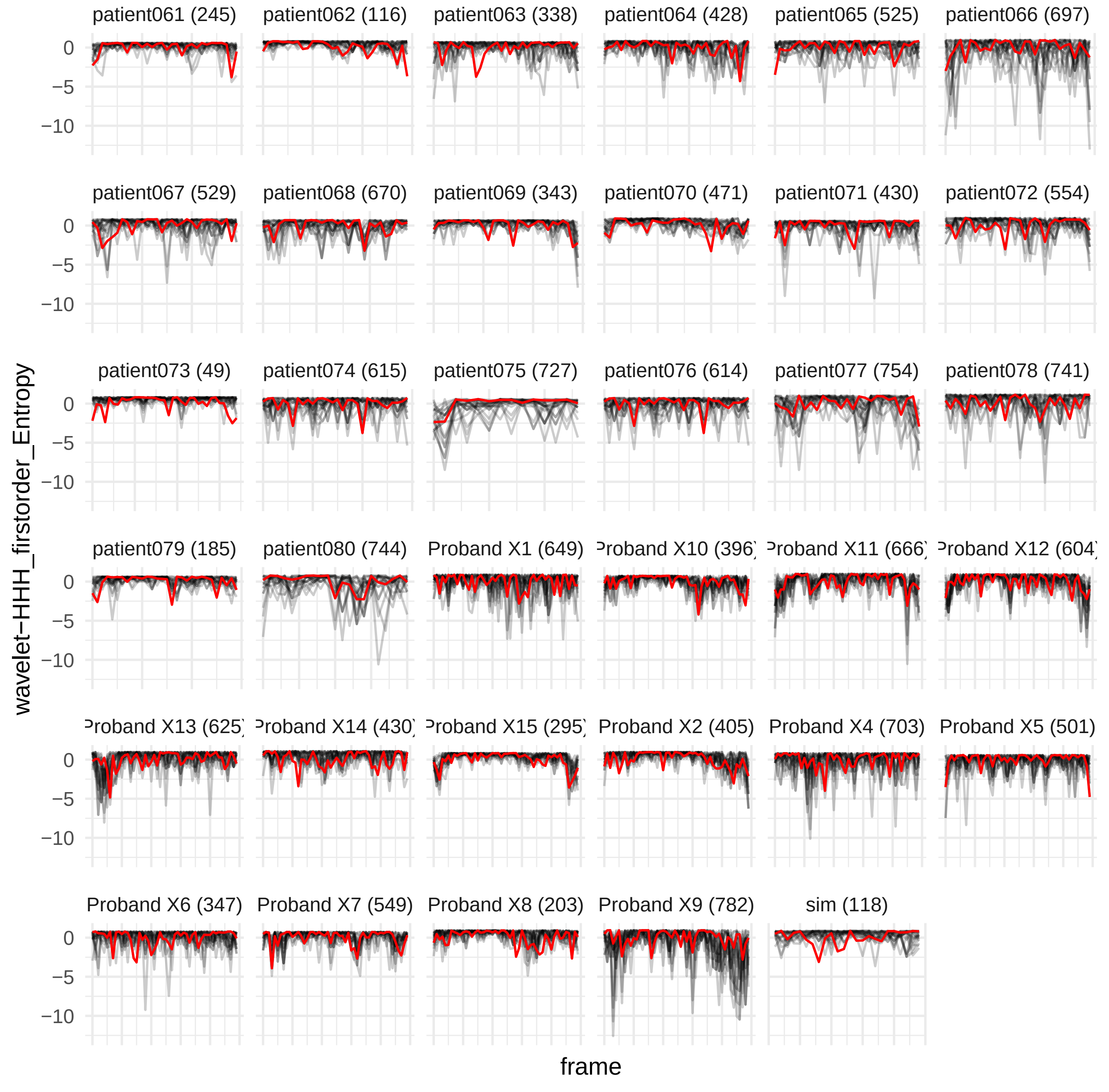
patient061 (245)
patient062 (116)
patient063 (338)
patient064 (428)
patient065 (525)
patient066 (697)
patient067 (529)
patient068 (670)
patient069 (343)
patient070 (471)
patient071 (430)
patient072 (554)
patient073 (49)
patient074 (615)
patient075 (727)
patient076 (614)
patient077 (754)
patient078 (741)
patient079 (185)
patient080 (744)
Proband X1 (649)
Proband X10 (396)
Proband X11 (666)
Proband X12 (604)
Proband X13 (625)
Proband X14 (430)
Proband X15 (295)
Proband X2 (405)
Proband X4 (703)
Proband X5 (501)
Proband X6 (347)
Proband X7 (549)
Proband X8 (203)
Proband X9 (782)
sim (118)
0
−5
−10
wavelet−HHH_firstorder_Entropy
frame

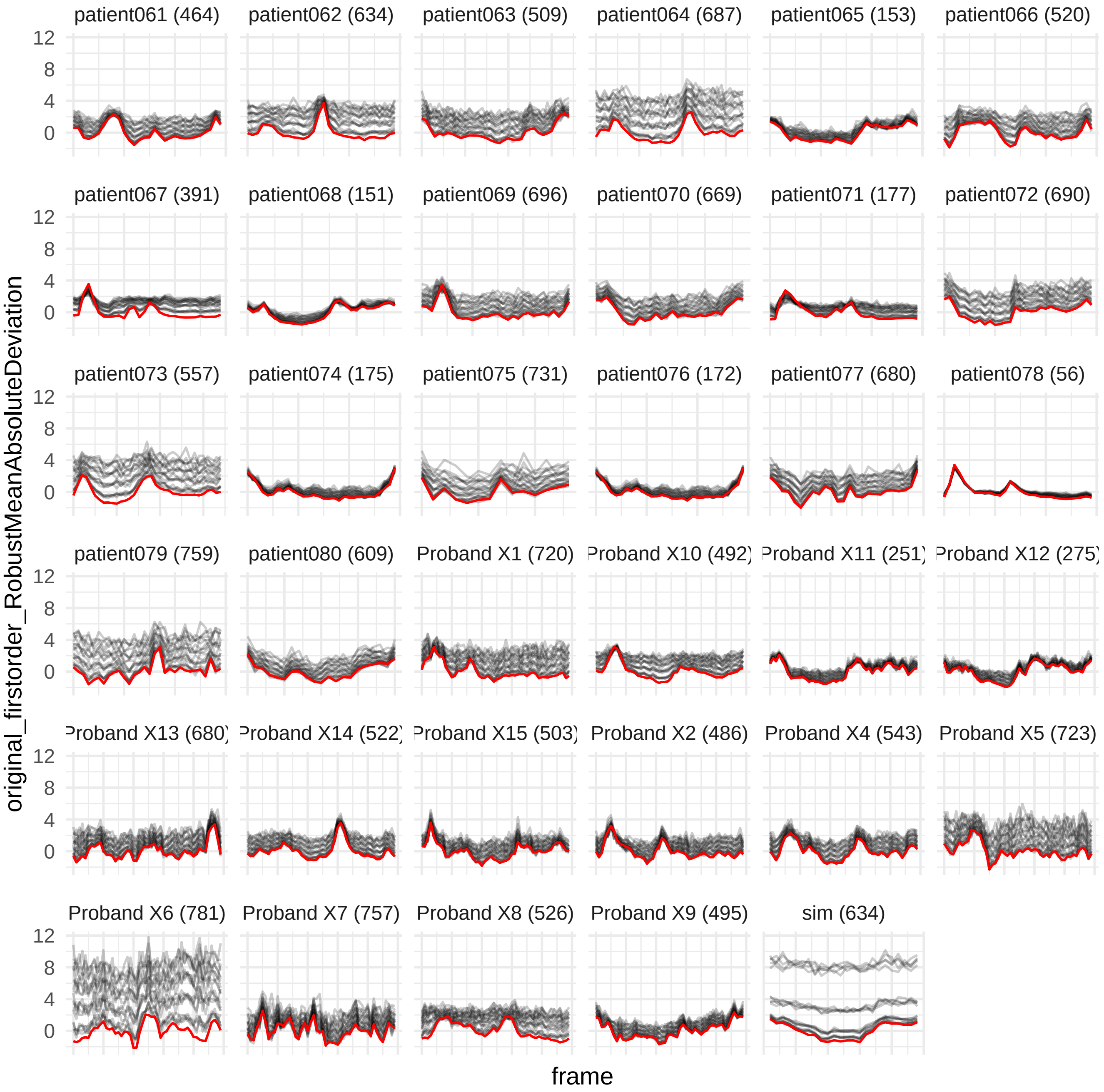
patient061 (464)
patient062 (634)
patient063 (509)
patient064 (687)
patient065 (153)
patient066 (520)
patient067 (391)
patient068 (151)
patient069 (696)
patient070 (669)
patient071 (177)
patient072 (690)
patient073 (557)
patient074 (175)
patient075 (731)
patient076 (172)
patient077 (680)
patient078 (56)
patient079 (759)
patient080 (609)
Proband X1 (720)
Proband X10 (492)
Proband X11 (251)
Proband X12 (275)
Proband X13 (680)
Proband X14 (522)
Proband X15 (503)
Proband X2 (486)
Proband X4 (543)
Proband X5 (723)
Proband X6 (781)
Proband X7 (757)
Proband X8 (526)
Proband X9 (495)
sim (634)
12
8
4
0
original_firstorder_RobustMeanAbsoluteDeviation
frame

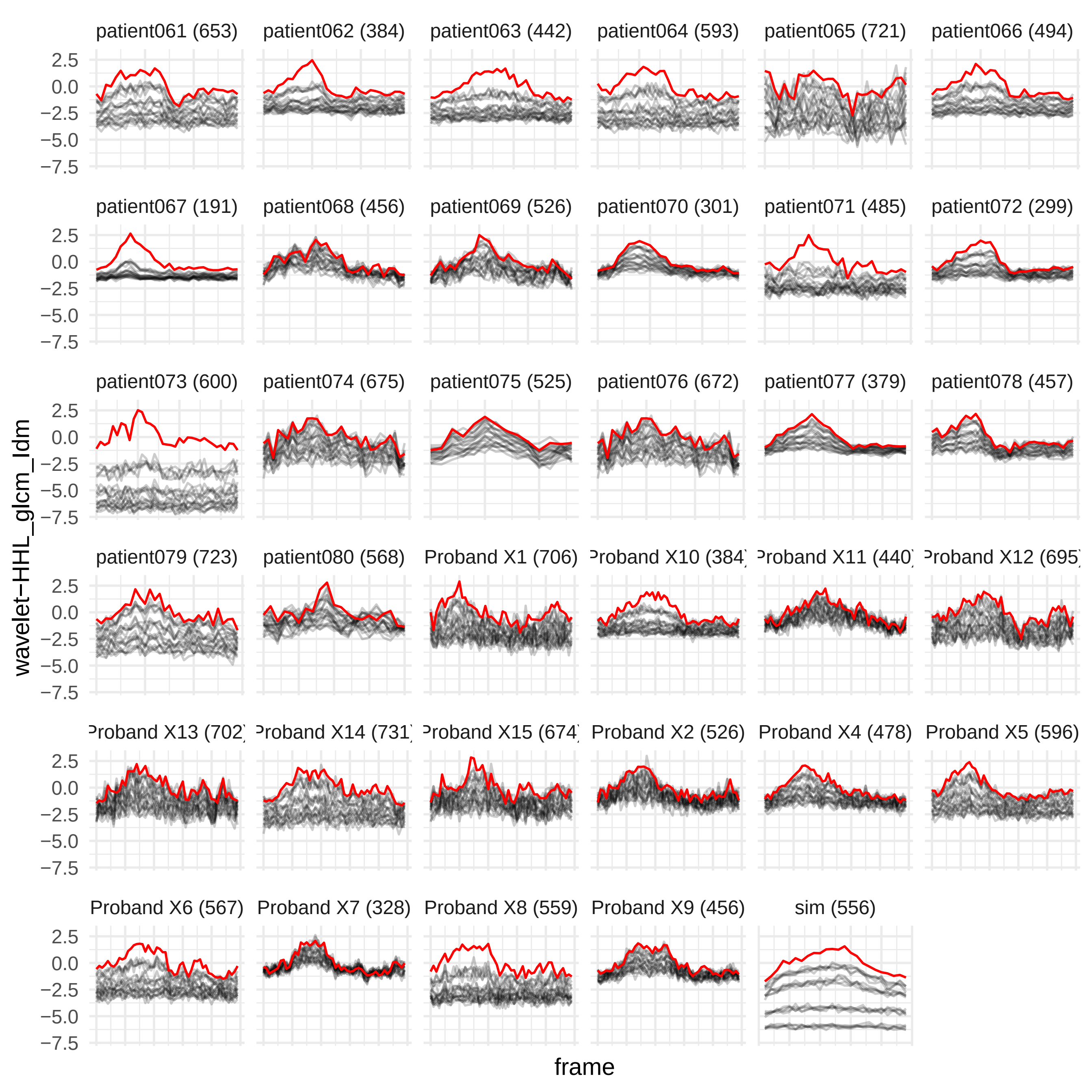

patient061 (653)
patient062 (384)
patient063 (442)
patient064 (593)
patient065 (721)
patient066 (494)
patient067 (191)
patient068 (456)
patient069 (526)
patient070 (301)
patient071 (485)
patient072 (299)
patient073 (600)
patient074 (675)
patient075 (525)
patient076 (672)
patient077 (379)
patient078 (457)
patient079 (723)
patient080 (568)
Proband X1 (706)
Proband X10 (384)
Proband X11 (440)
Proband X12 (695)
Proband X13 (702)
Proband X14 (731)
Proband X15 (674)
Proband X2 (526)
Proband X4 (478)
Proband X5 (596)
Proband X6 (567)
Proband X7 (328)
Proband X8 (559)
Proband X9 (456)
sim (556)
2.5
0.0
−2.5
−5.0
−7.5
wavelet−HHL_glcm_Idm
frame

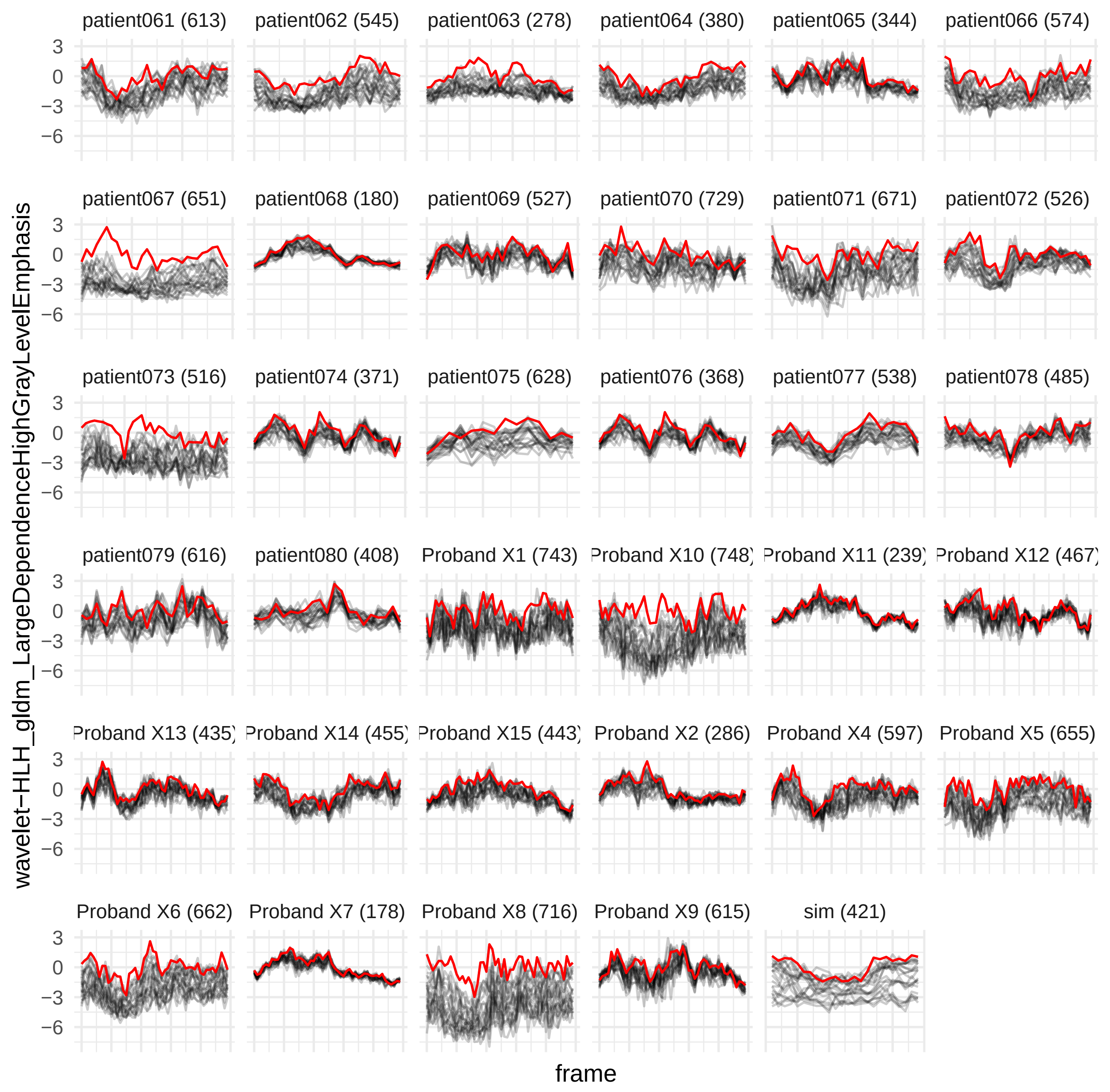

patient061 (613)
patient062 (545)
patient063 (278)
patient064 (380)
patient065 (344)
patient066 (574)
patient067 (651)
patient068 (180)
patient069 (527)
patient070 (729)
patient071 (671)
patient072 (526)
patient073 (516)
patient074 (371)
patient075 (628)
patient076 (368)
patient077 (538)
patient078 (485)
patient079 (616)
patient080 (408)
Proband X1 (743)
Proband X10 (748)
Proband X11 (239)
Proband X12 (467)
Proband X13 (435)
Proband X14 (455)
Proband X15 (443)
Proband X2 (286)
Proband X4 (597)
Proband X5 (655)
Proband X6 (662)
Proband X7 (178)
Proband X8 (716)
Proband X9 (615)
sim (421)
3
0
−3
−6
wavelet−HLH_gldm_LargeDependenceHighGrayLevelEmphasis
frame

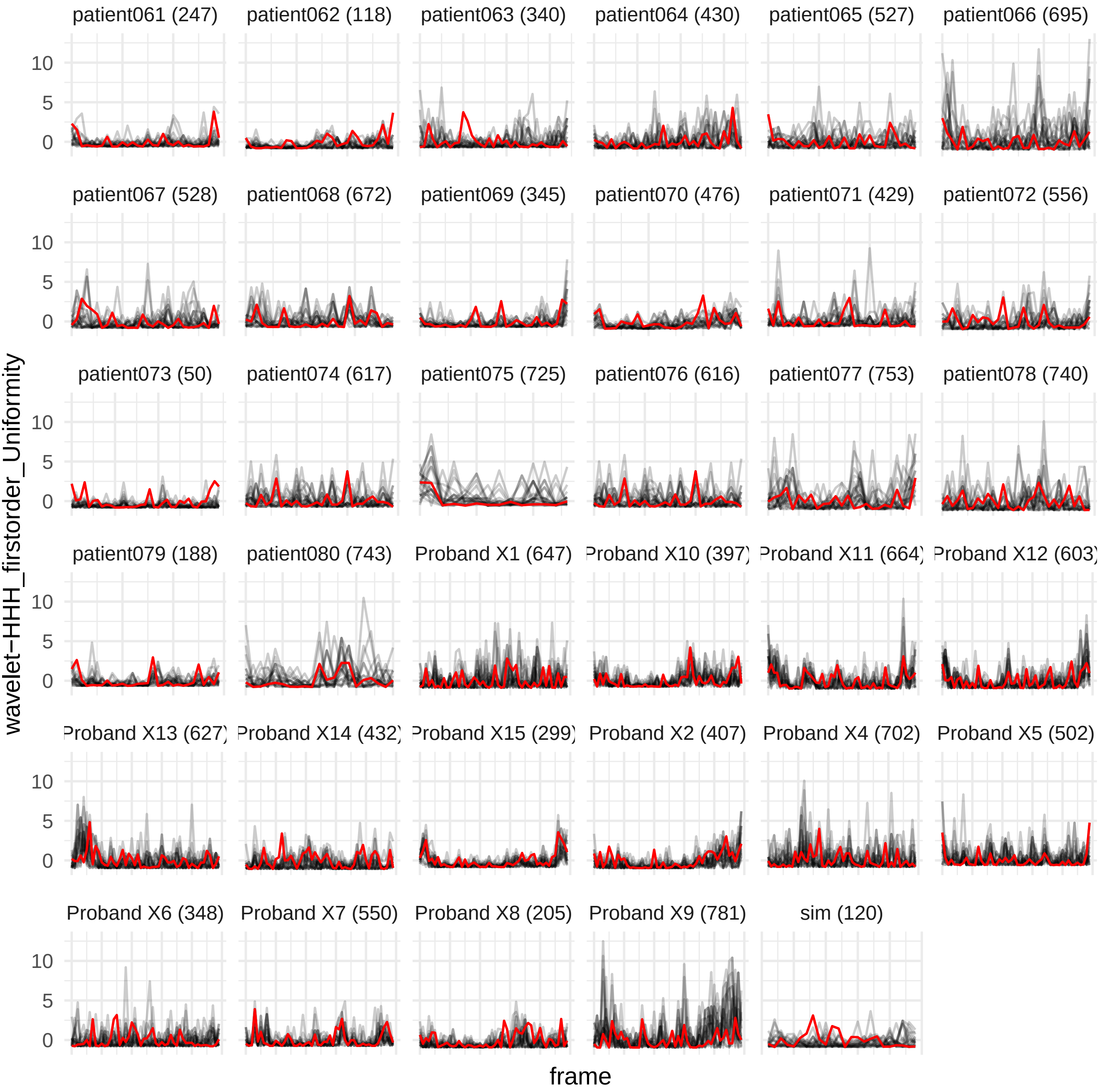

patient061 (247)
patient062 (118)
patient063 (340)
patient064 (430)
patient065 (527)
patient066 (695)
patient067 (528)
patient068 (672)
patient069 (345)
patient070 (476)
patient071 (429)
patient072 (556)
patient073 (50)
patient074 (617)
patient075 (725)
patient076 (616)
patient077 (753)
patient078 (740)
patient079 (188)
patient080 (743)
Proband X1 (647)
Proband X10 (397)
Proband X11 (664)
Proband X12 (603)
Proband X13 (627)
Proband X14 (432)
Proband X15 (299)
Proband X2 (407)
Proband X4 (702)
Proband X5 (502)
Proband X6 (348)
Proband X7 (550)
Proband X8 (205)
Proband X9 (781)
sim (120)
wavelet−HHH_firstorder_Uniformity
frame
0
5
10

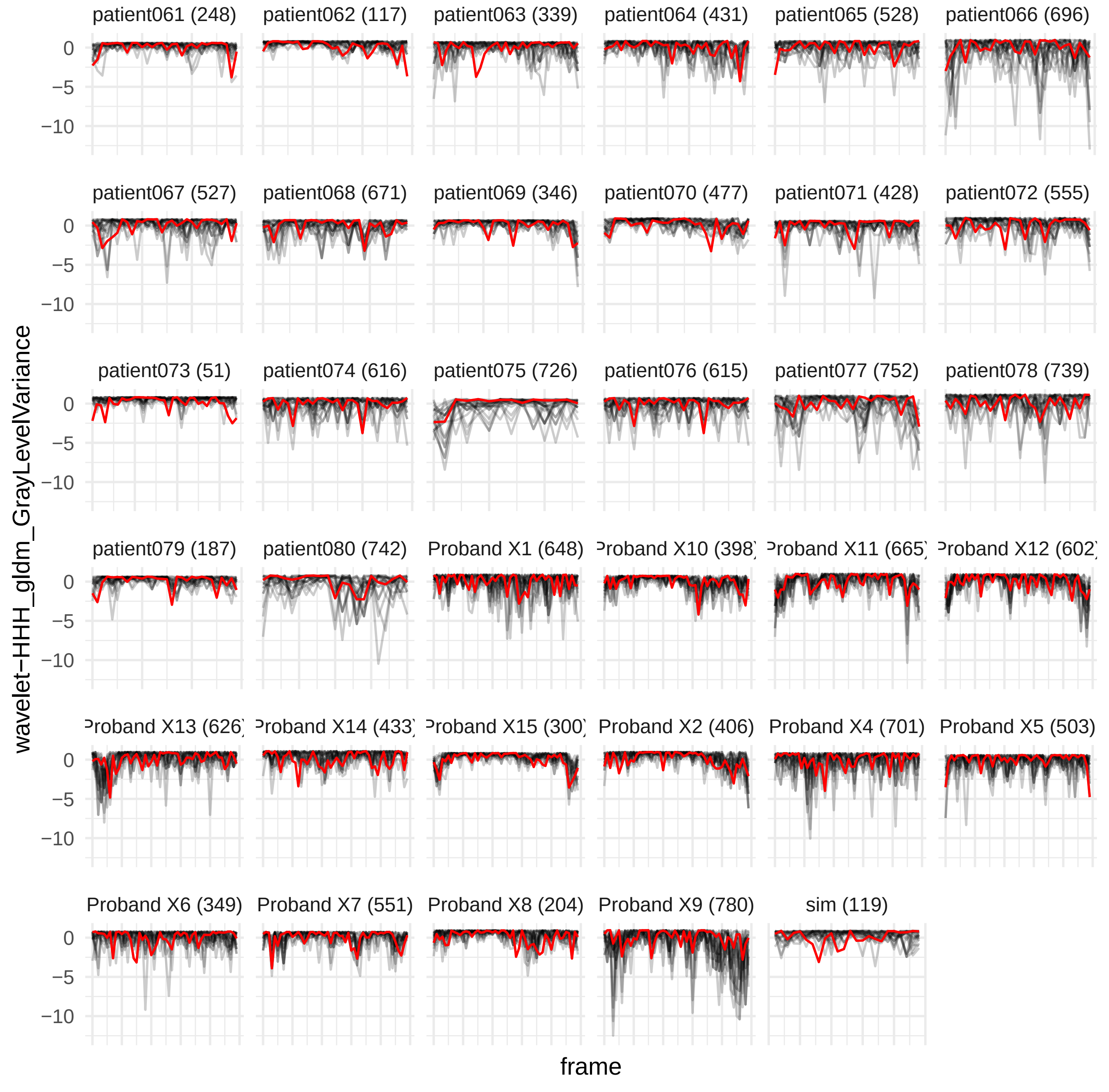

patient061 (248)
patient062 (117)
patient063 (339)
patient064 (431)
patient065 (528)
patient066 (696)
patient067 (527)
patient068 (671)
patient069 (346)
patient070 (477)
patient071 (428)
patient072 (555)
patient073 (51)
patient074 (616)
patient075 (726)
patient076 (615)
patient077 (752)
patient078 (739)
patient079 (187)
patient080 (742)
Proband X1 (648)
Proband X10 (398)
Proband X11 (665)
Proband X12 (602)
Proband X13 (626)
Proband X14 (433)
Proband X15 (300)
Proband X2 (406)
Proband X4 (701)
Proband X5 (503)
Proband X6 (349)
Proband X7 (551)
Proband X8 (204)
Proband X9 (780)
sim (119)
0
−5
−10
wavelet−HHH_gldm_GrayLevelVariance
frame

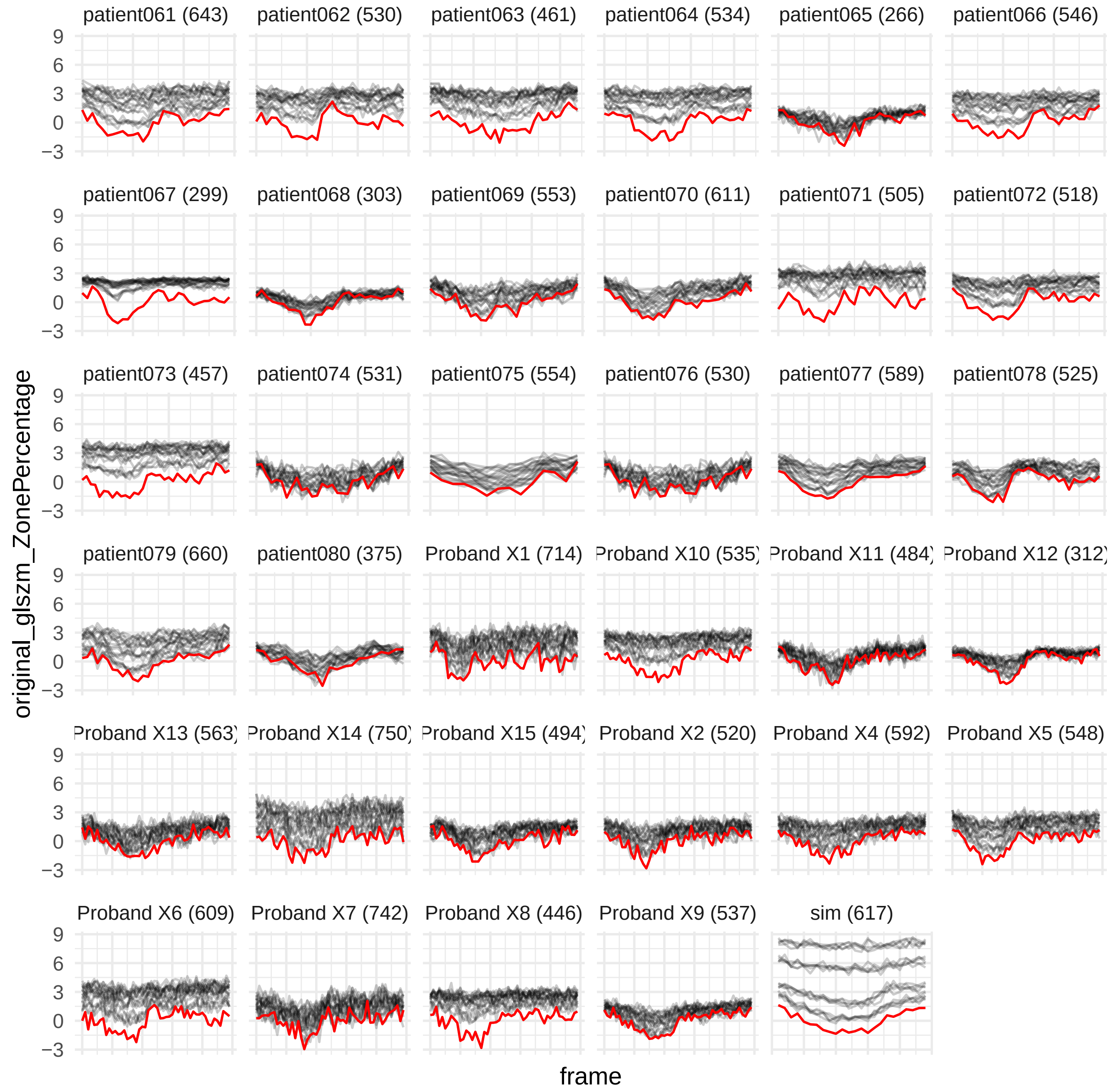

patient061 (643)
patient062 (530)
patient063 (461)
patient064 (534)
patient065 (266)
patient066 (546)
patient067 (299)
patient068 (303)
patient069 (553)
patient070 (611)
patient071 (505)
patient072 (518)
patient073 (457)
patient074 (531)
patient075 (554)
patient076 (530)
patient077 (589)
patient078 (525)
patient079 (660)
patient080 (375)
Proband X1 (714)
Proband X10 (535)
Proband X11 (484)
Proband X12 (312)
Proband X13 (563)
Proband X14 (750)
Proband X15 (494)
Proband X2 (520)
Proband X4 (592)
Proband X5 (548)
Proband X6 (609)
Proband X7 (742)
Proband X8 (446)
Proband X9 (537)
sim (617)
9
6
3
0
−3
original_glszm_ZonePercentage
frame

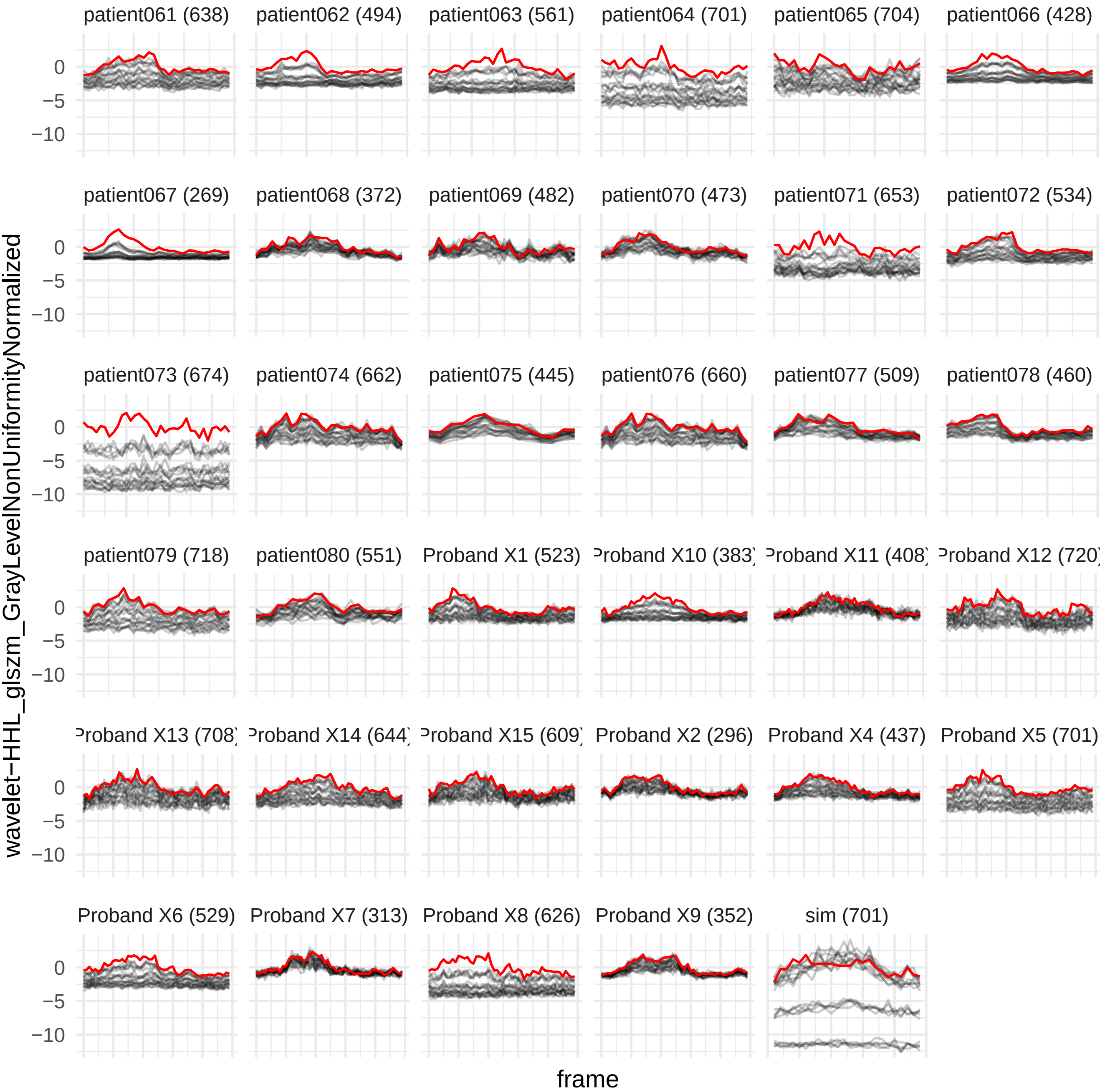

patient061 (638)
patient062 (494)
patient063 (561)
patient064 (701)
patient065 (704)
patient066 (428)
patient067 (269)
patient068 (372)
patient069 (482)
patient070 (473)
patient071 (653)
patient072 (534)
patient073 (674)
patient074 (662)
patient075 (445)
patient076 (660)
patient077 (509)
patient078 (460)
patient079 (718)
patient080 (551)
Proband X1 (523)
Proband X10 (383)
Proband X11 (408)
Proband X12 (720)
Proband X13 (708)
Proband X14 (644)
Proband X15 (609)
Proband X2 (296)
Proband X4 (437)
Proband X5 (701)
Proband X6 (529)
Proband X7 (313)
Proband X8 (626)
Proband X9 (352)
sim (701)
0
−5
−10
wavelet−HHL_glszm_GrayLevelNonUniformityNormalized
frame

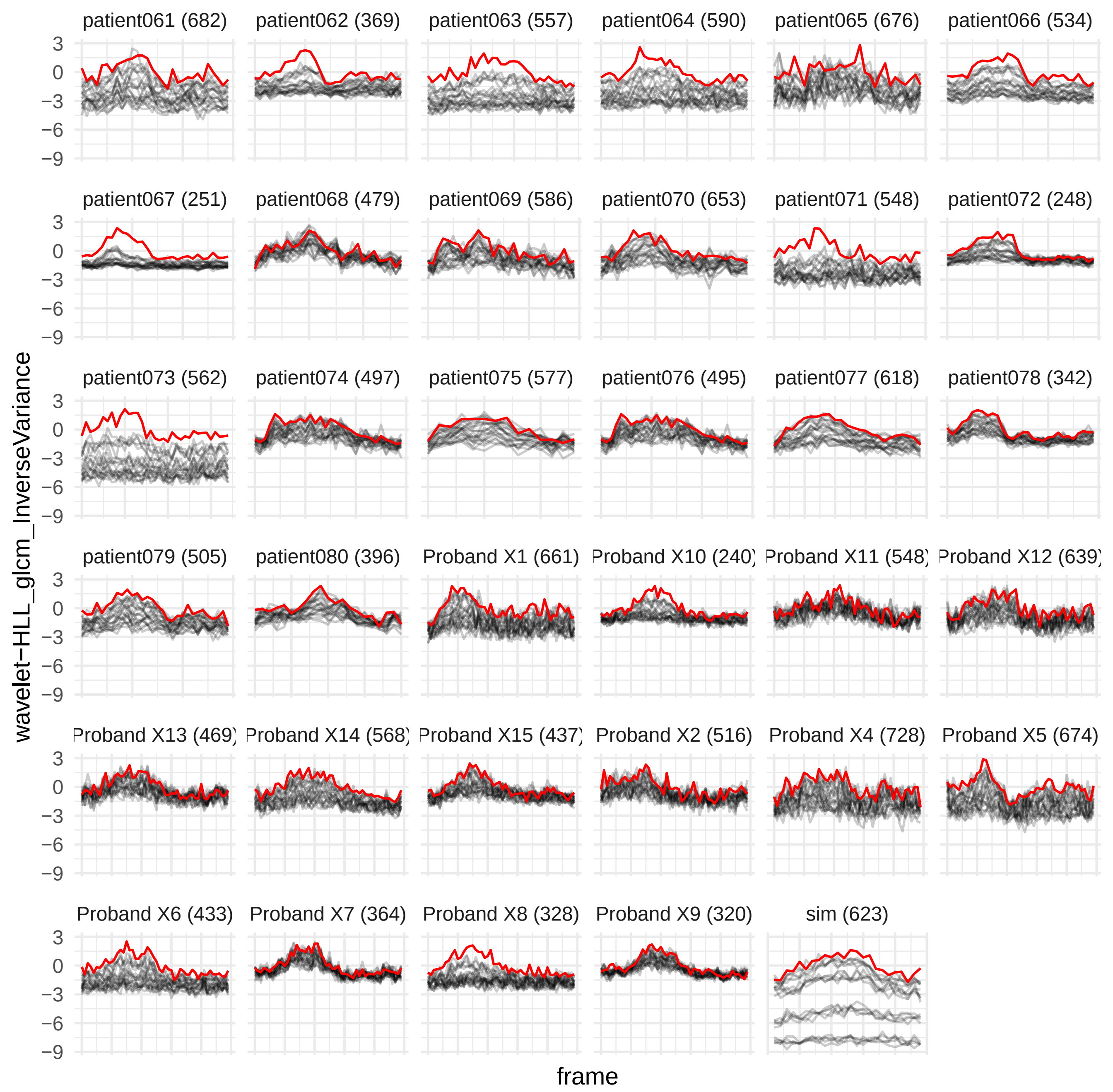

patient061 (682)
patient062 (369)
patient063 (557)
patient064 (590)
patient065 (676)
patient066 (534)
patient067 (251)
patient068 (479)
patient069 (586)
patient070 (653)
patient071 (548)
patient072 (248)
patient073 (562)
patient074 (497)
patient075 (577)
patient076 (495)
patient077 (618)
patient078 (342)
patient079 (505)
patient080 (396)
Proband X1 (661)
Proband X10 (240)
Proband X11 (548)
Proband X12 (639)
Proband X13 (469)
Proband X14 (568)
Proband X15 (437)
Proband X2 (516)
Proband X4 (728)
Proband X5 (674)
Proband X6 (433)
Proband X7 (364)
Proband X8 (328)
Proband X9 (320)
sim (623)
3
0
−3
−6
−9
wavelet−HLL_glcm_InverseVariance
frame

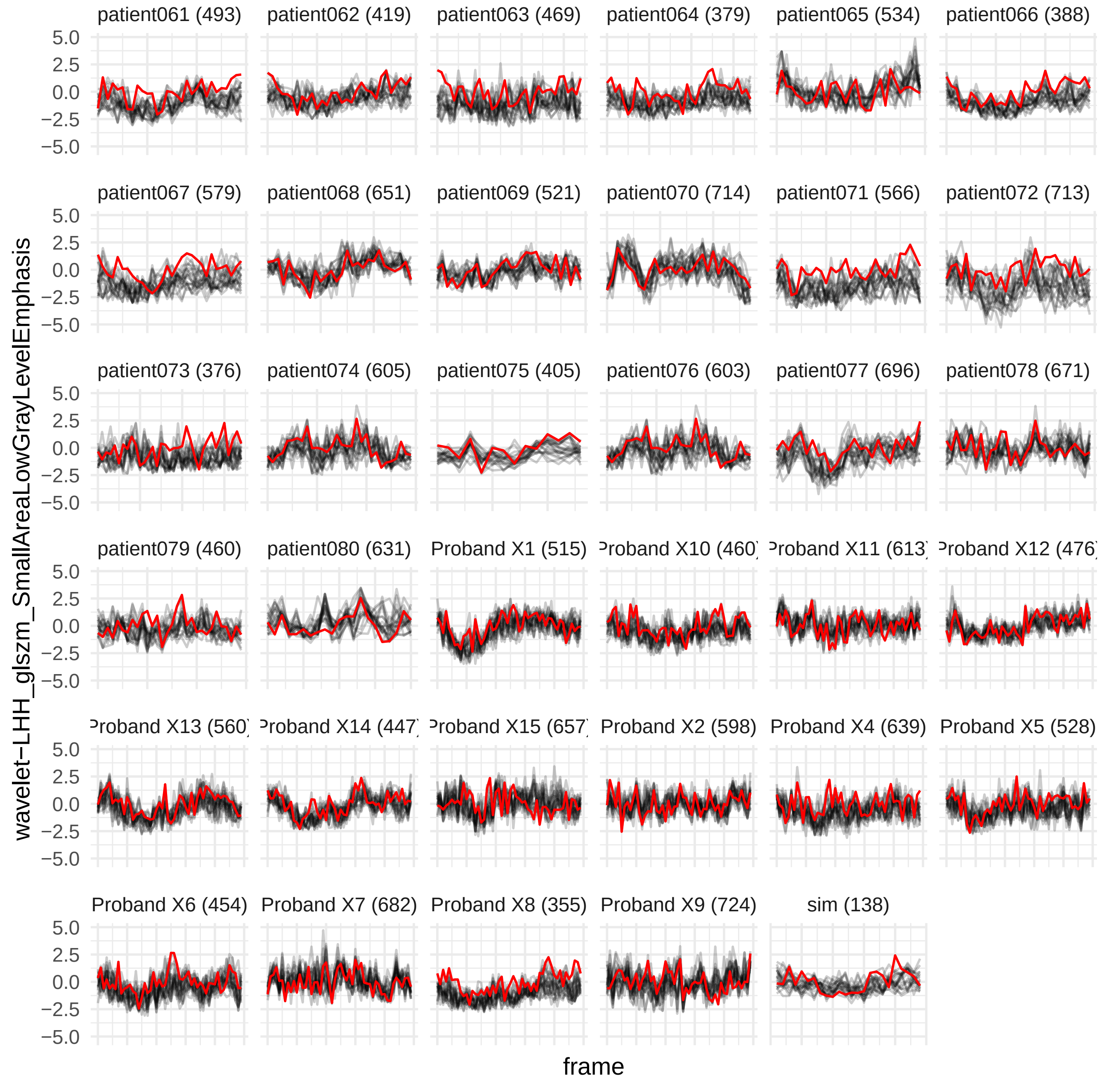

patient061 (493)
patient062 (419)
patient063 (469)
patient064 (379)
patient065 (534)
patient066 (388)
patient067 (579)
patient068 (651)
patient069 (521)
patient070 (714)
patient071 (566)
patient072 (713)
patient073 (376)
patient074 (605)
patient075 (405)
patient076 (603)
patient077 (696)
patient078 (671)
patient079 (460)
patient080 (631)
Proband X1 (515)
Proband X10 (460)
Proband X11 (613)
Proband X12 (476)
Proband X13 (560)
Proband X14 (447)
Proband X15 (657)
Proband X2 (598)
Proband X4 (639)
Proband X5 (528)
Proband X6 (454)
Proband X7 (682)
Proband X8 (355)
Proband X9 (724)
sim (138)
5.0
2.5
0.0
−2.5
−5.0
wavelet−LHH_glszm_SmallAreaLowGrayLevelEmphasis
frame

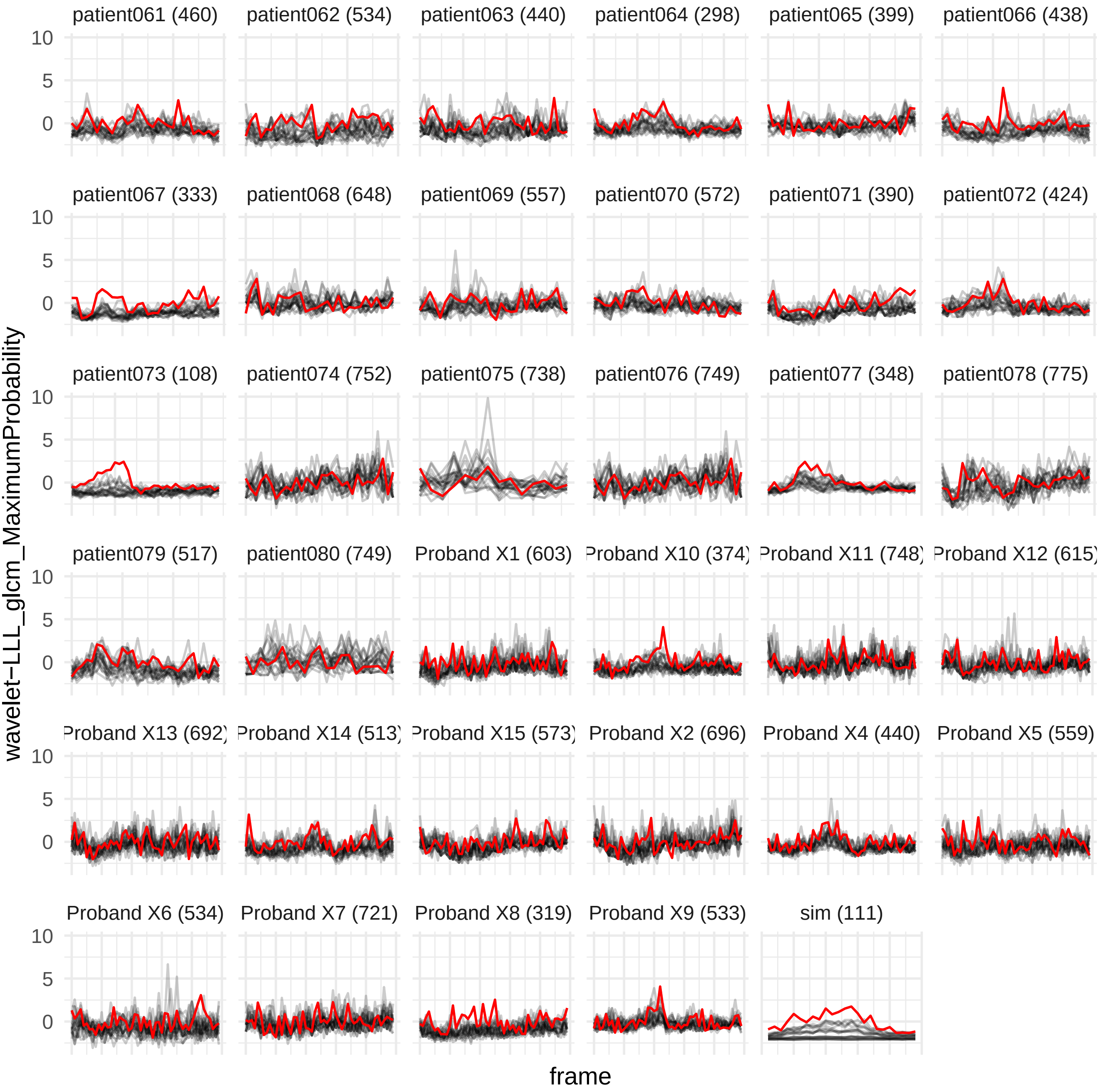

patient061 (460)
patient062 (534)
patient063 (440)
patient064 (298)
patient065 (399)
patient066 (438)
patient067 (333)
patient068 (648)
patient069 (557)
patient070 (572)
patient071 (390)
patient072 (424)
patient073 (108)
patient074 (752)
patient075 (738)
patient076 (749)
patient077 (348)
patient078 (775)
patient079 (517)
patient080 (749)
Proband X1 (603)
Proband X10 (374)
Proband X11 (748)
Proband X12 (615)
Proband X13 (692)
Proband X14 (513)
Proband X15 (573)
Proband X2 (696)
Proband X4 (440)
Proband X5 (559)
Proband X6 (534)
Proband X7 (721)
Proband X8 (319)
Proband X9 (533)
sim (111)
10
5
0
wavelet−LLL_glcm_MaximumProbability
frame

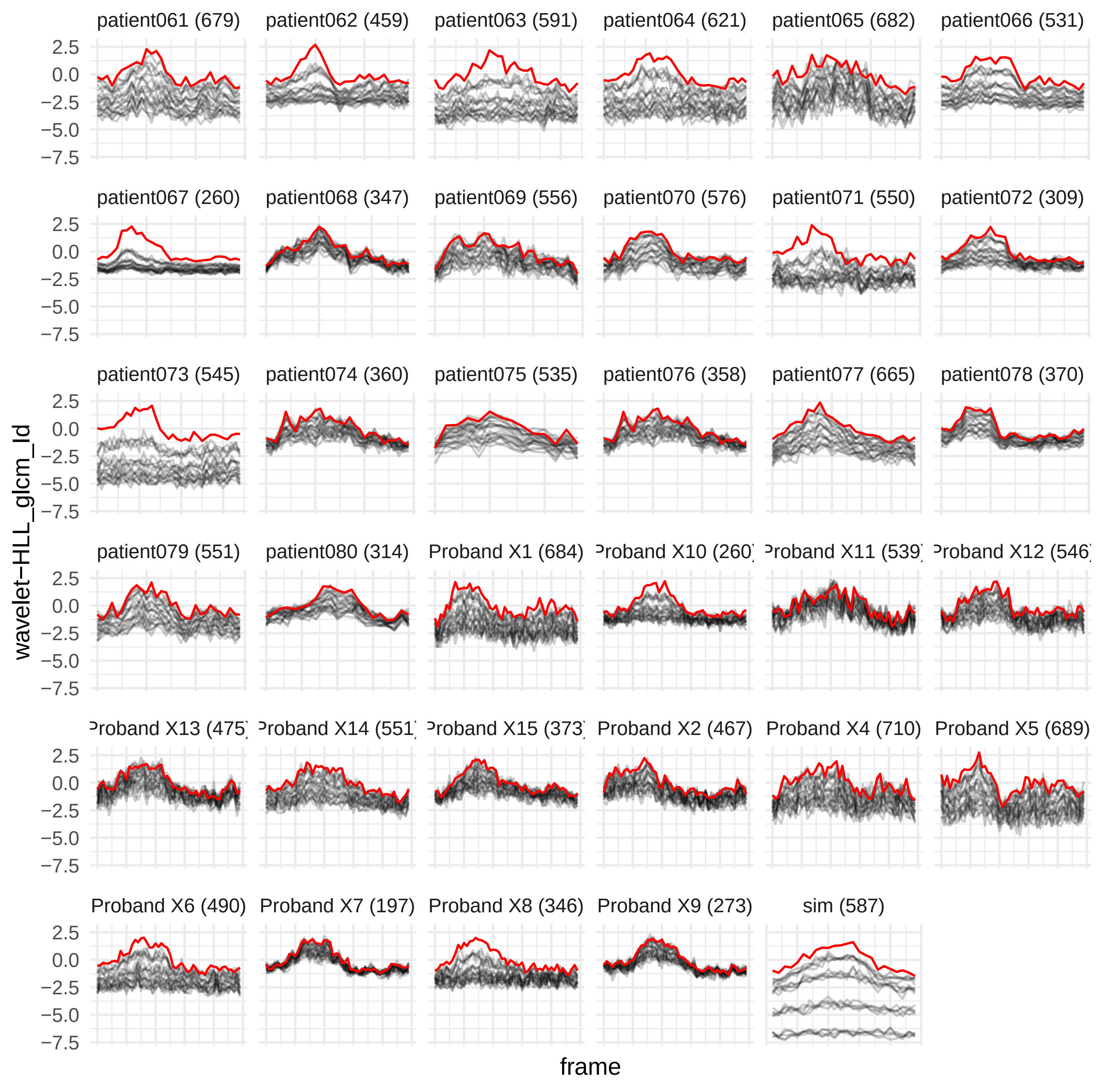
patient061 (679)
patient062 (459)
patient063 (591)
patient064 (621)
patient065 (682)
patient066 (531)
patient067 (260)
patient068 (347)
patient069 (556)
patient070 (576)
patient071 (550)
patient072 (309)
patient073 (545)
patient074 (360)
patient075 (535)
patient076 (358)
patient077 (665)
patient078 (370)
patient079 (551)
patient080 (314)
Proband X1 (684)
Proband X10 (260)
Proband X11 (539)
Proband X12 (546)
Proband X13 (475)
Proband X14 (551)
Proband X15 (373)
Proband X2 (467)
Proband X4 (710)
Proband X5 (689)
Proband X6 (490)
Proband X7 (197)
Proband X8 (346)
Proband X9 (273)
sim (587)
2.5
0.0
−2.5
−5.0
−7.5
wavelet−HLL_glcm_Id
frame

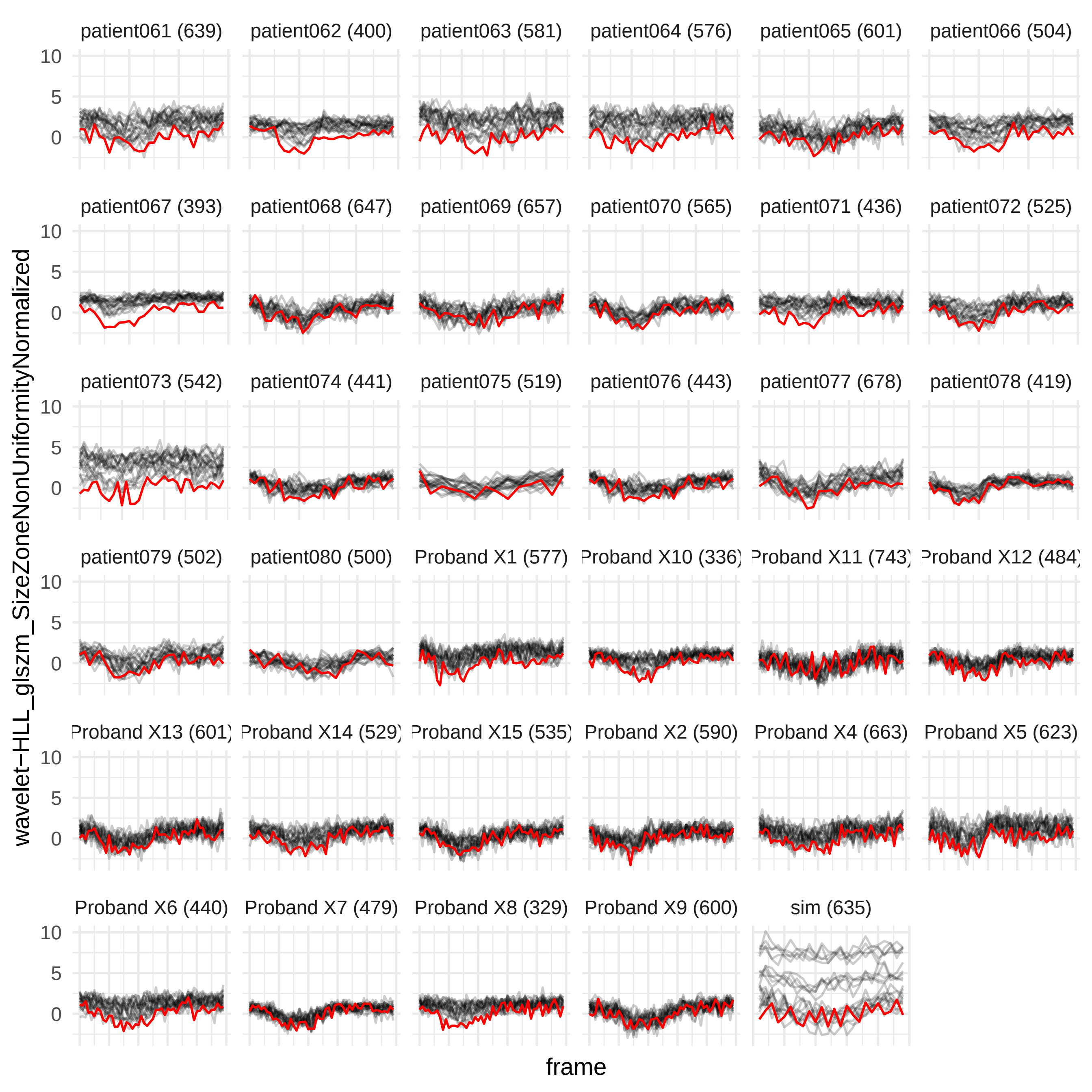

patient061 (639)
patient062 (400)
patient063 (581)
patient064 (576)
patient065 (601)
patient066 (504)
patient067 (393)
patient068 (647)
patient069 (657)
patient070 (565)
patient071 (436)
patient072 (525)
patient073 (542)
patient074 (441)
patient075 (519)
patient076 (443)
patient077 (678)
patient078 (419)
patient079 (502)
patient080 (500)
Proband X1 (577)
Proband X10 (336)
Proband X11 (743)
Proband X12 (484)
Proband X13 (601)
Proband X14 (529)
Proband X15 (535)
Proband X2 (590)
Proband X4 (663)
Proband X5 (623)
Proband X6 (440)
Proband X7 (479)
Proband X8 (329)
Proband X9 (600)
sim (635)
10
5
0
wavelet−HLL_glszm_SizeZoneNonUniformityNormalized
frame

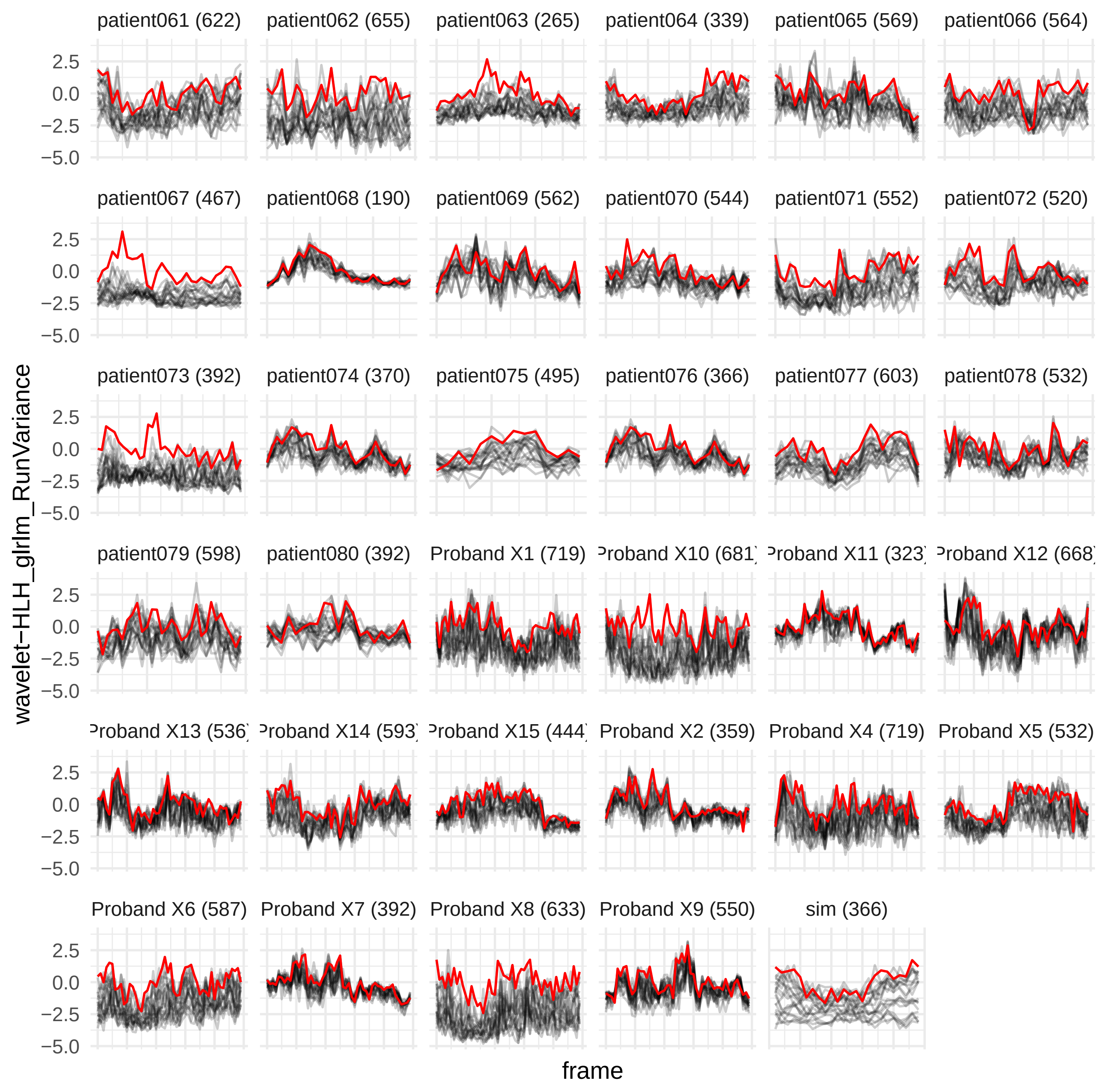

patient061 (622)
patient062 (655)
patient063 (265)
patient064 (339)
patient065 (569)
patient066 (564)
patient067 (467)
patient068 (190)
patient069 (562)
patient070 (544)
patient071 (552)
patient072 (520)
patient073 (392)
patient074 (370)
patient075 (495)
patient076 (366)
patient077 (603)
patient078 (532)
patient079 (598)
patient080 (392)
Proband X1 (719)
Proband X10 (681)
Proband X11 (323)
Proband X12 (668)
Proband X13 (536)
Proband X14 (593)
Proband X15 (444)
Proband X2 (359)
Proband X4 (719)
Proband X5 (532)
Proband X6 (587)
Proband X7 (392)
Proband X8 (633)
Proband X9 (550)
sim (366)
2.5
0.0
−2.5
−5.0
wavelet−HLH_glrlm_RunVariance
frame

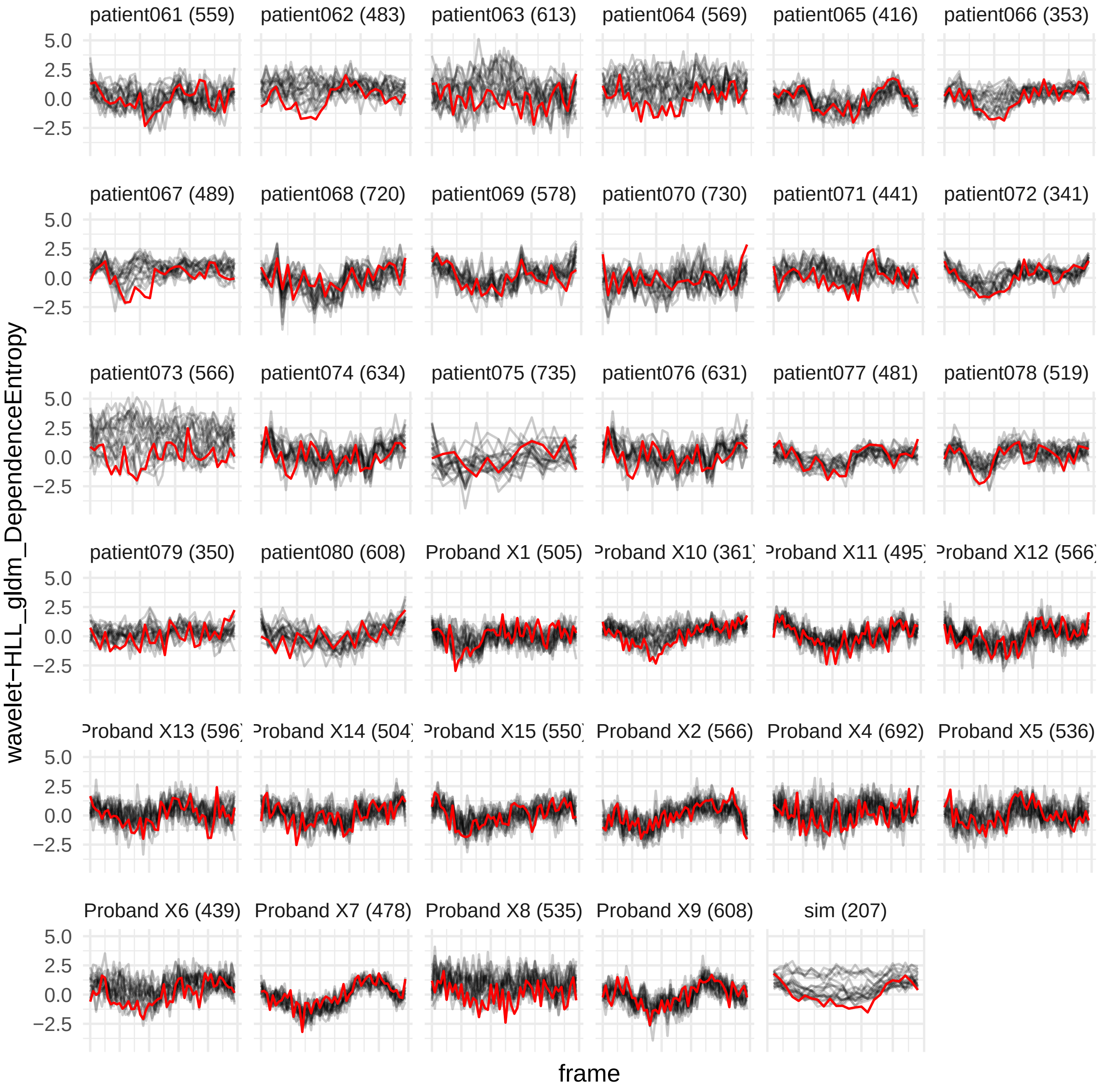

patient061 (559)
patient062 (483)
patient063 (613)
patient064 (569)
patient065 (416)
patient066 (353)
patient067 (489)
patient068 (720)
patient069 (578)
patient070 (730)
patient071 (441)
patient072 (341)
patient073 (566)
patient074 (634)
patient075 (735)
patient076 (631)
patient077 (481)
patient078 (519)
patient079 (350)
patient080 (608)
Proband X1 (505)
Proband X10 (361)
Proband X11 (495)
Proband X12 (566)
Proband X13 (596)
Proband X14 (504)
Proband X15 (550)
Proband X2 (566)
Proband X4 (692)
Proband X5 (536)
Proband X6 (439)
Proband X7 (478)
Proband X8 (535)
Proband X9 (608)
sim (207)
5.0
2.5
0.0
−2.5
wavelet−HLL_gldm_DependenceEntropy
frame

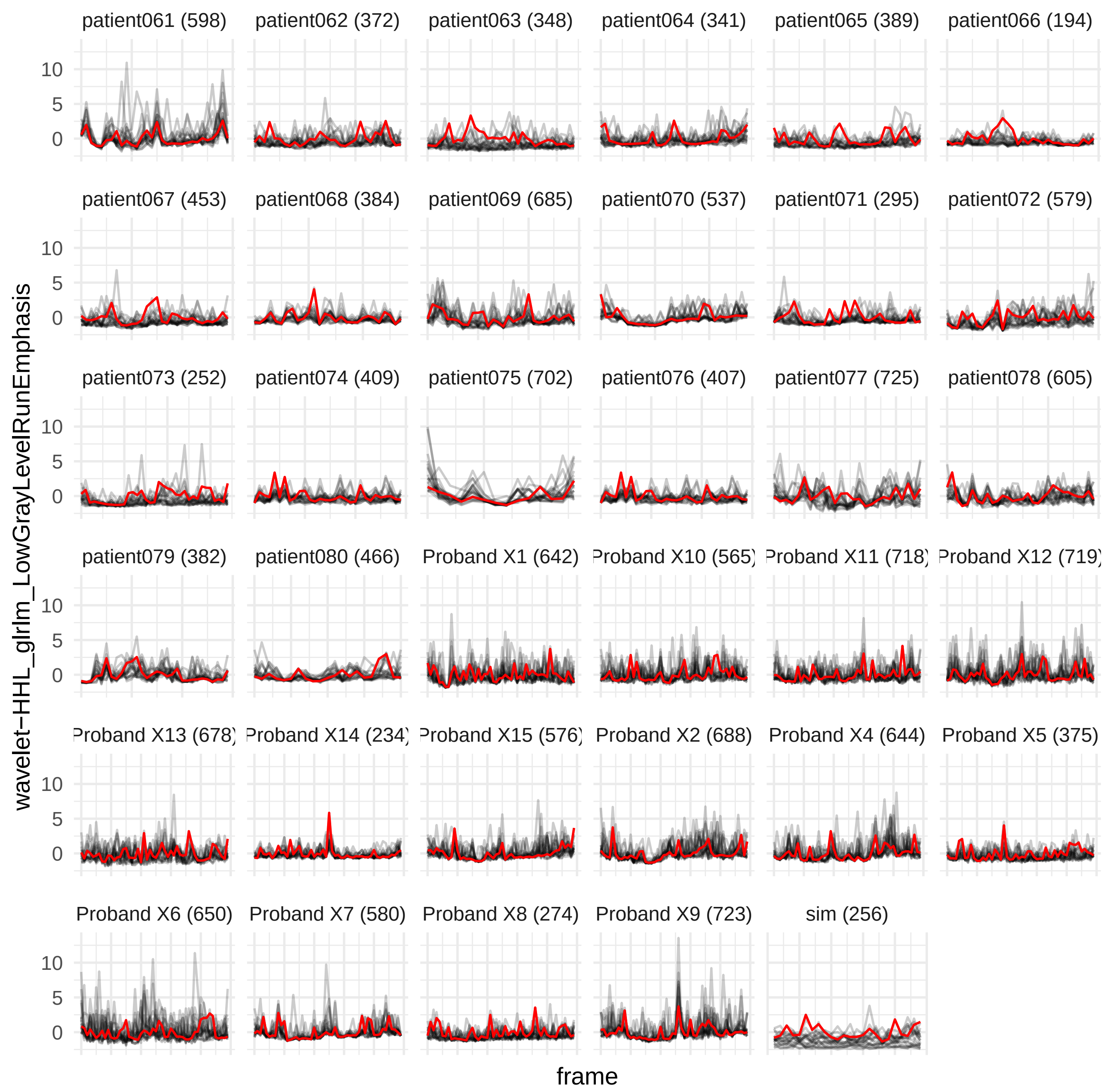

patient061 (598)
patient062 (372)
patient063 (348)
patient064 (341)
patient065 (389)
patient066 (194)
patient067 (453)
patient068 (384)
patient069 (685)
patient070 (537)
patient071 (295)
patient072 (579)
patient073 (252)
patient074 (409)
patient075 (702)
patient076 (407)
patient077 (725)
patient078 (605)
patient079 (382)
patient080 (466)
Proband X1 (642)
Proband X10 (565)
Proband X11 (718)
Proband X12 (719)
Proband X13 (678)
Proband X14 (234)
Proband X15 (576)
Proband X2 (688)
Proband X4 (644)
Proband X5 (375)
Proband X6 (650)
Proband X7 (580)
Proband X8 (274)
Proband X9 (723)
sim (256)
wavelet−HHL_glrlm_LowGrayLevelRunEmphasis
frame
10
5
0

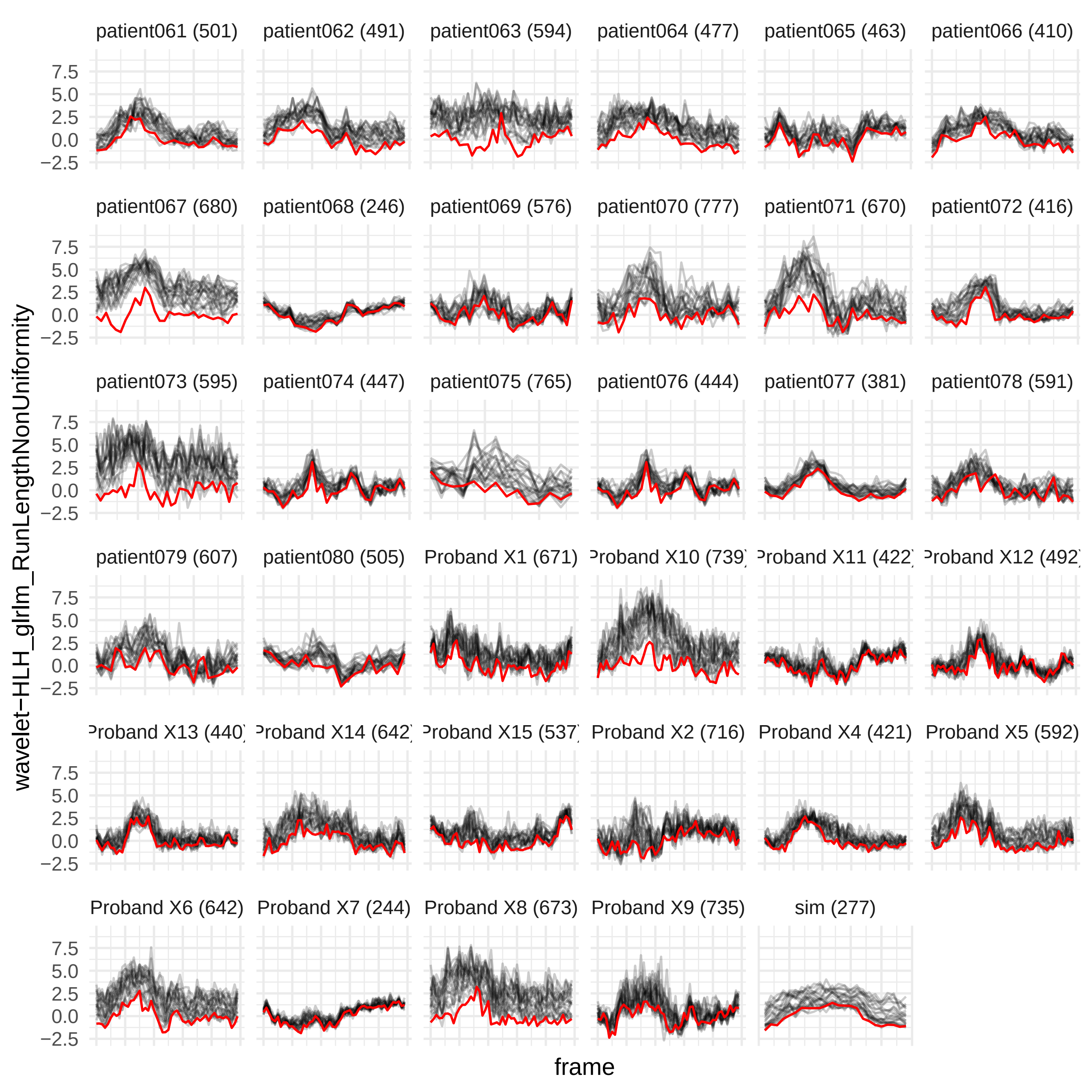

patient061 (501)
patient062 (491)
patient063 (594)
patient064 (477)
patient065 (463)
patient066 (410)
patient067 (680)
patient068 (246)
patient069 (576)
patient070 (777)
patient071 (670)
patient072 (416)
patient073 (595)
patient074 (447)
patient075 (765)
patient076 (444)
patient077 (381)
patient078 (591)
patient079 (607)
patient080 (505)
Proband X1 (671)
Proband X10 (739)
Proband X11 (422)
Proband X12 (492)
Proband X13 (440)
Proband X14 (642)
Proband X15 (537)
Proband X2 (716)
Proband X4 (421)
Proband X5 (592)
Proband X6 (642)
Proband X7 (244)
Proband X8 (673)
Proband X9 (735)
sim (277)
7.5
5.0
2.5
0.0
−2.5
wavelet−HLH_glrlm_RunLengthNonUniformity
frame

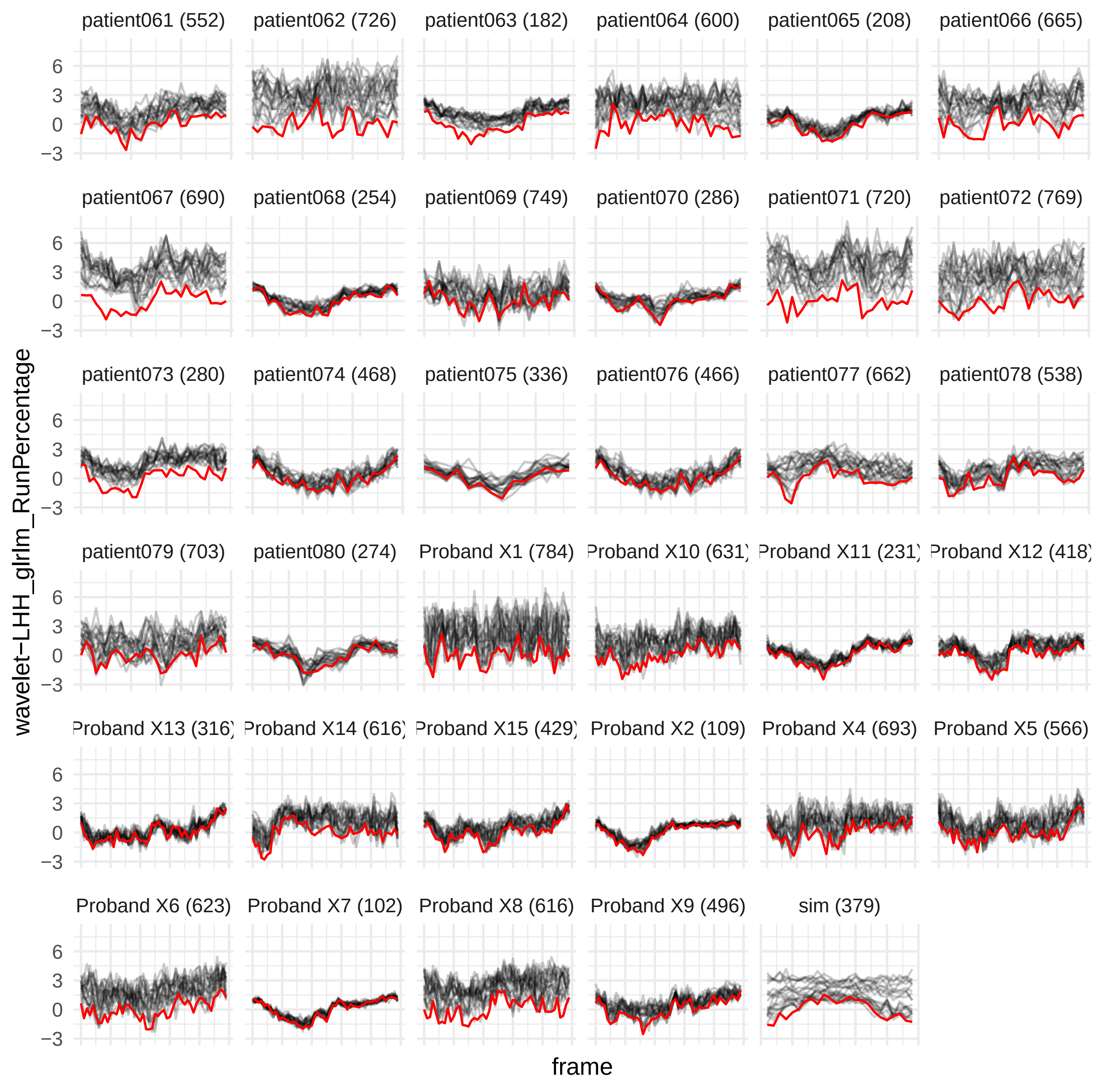
patient061 (552)
patient062 (726)
patient063 (182)
patient064 (600)
patient065 (208)
patient066 (665)
patient067 (690)
patient068 (254)
patient069 (749)
patient070 (286)
patient071 (720)
patient072 (769)
patient073 (280)
patient074 (468)
patient075 (336)
patient076 (466)
patient077 (662)
patient078 (538)
patient079 (703)
patient080 (274)
Proband X1 (784)
Proband X10 (631)
Proband X11 (231)
Proband X12 (418)
Proband X13 (316)
Proband X14 (616)
Proband X15 (429)
Proband X2 (109)
Proband X4 (693)
Proband X5 (566)
Proband X6 (623)
Proband X7 (102)
Proband X8 (616)
Proband X9 (496)
sim (379)
6
3
0
−3
wavelet−LHH_glrlm_RunPercentage
frame

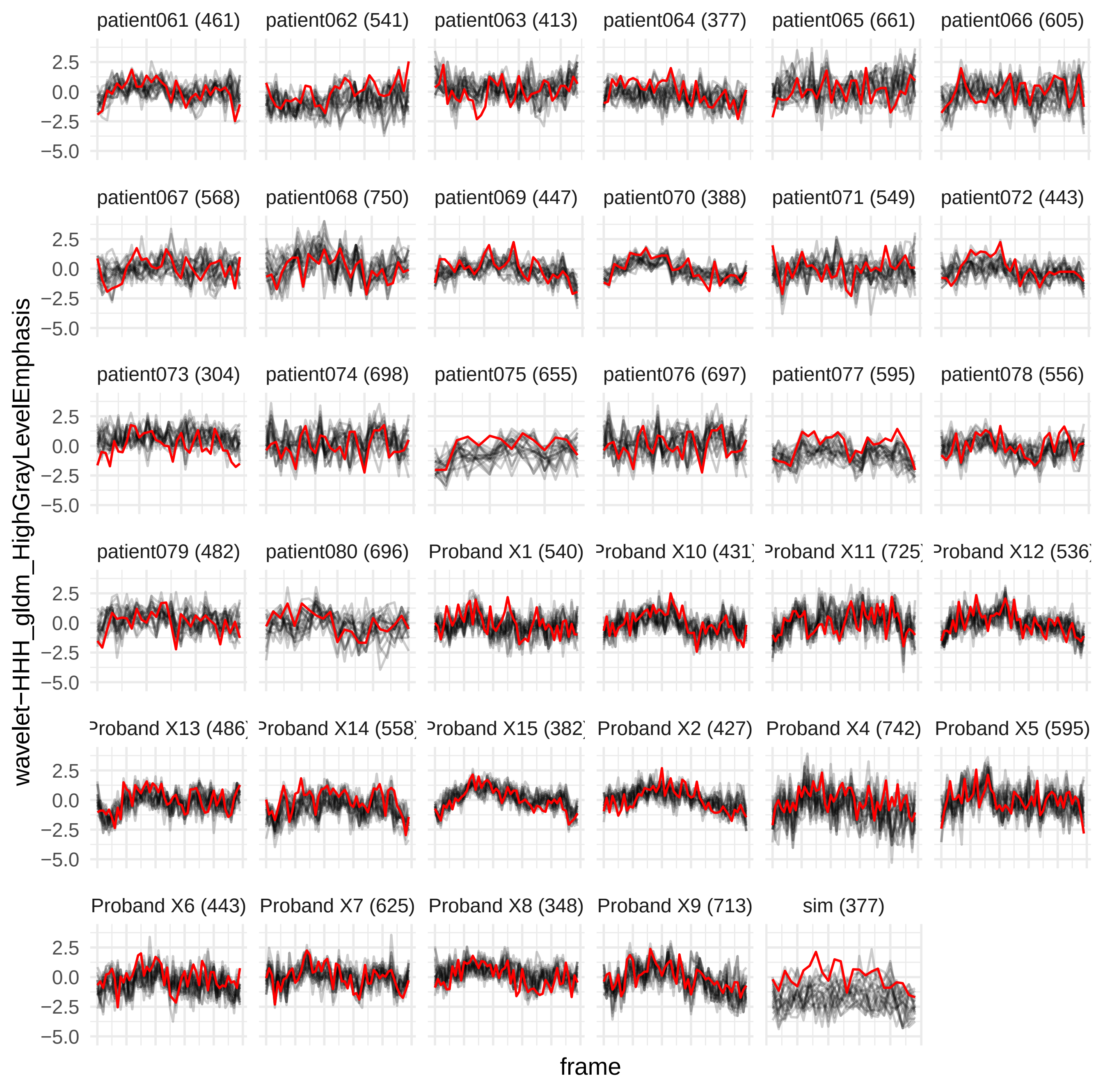
patient061 (461)
patient062 (541)
patient063 (413)
patient064 (377)
patient065 (661)
patient066 (605)
patient067 (568)
patient068 (750)
patient069 (447)
patient070 (388)
patient071 (549)
patient072 (443)
patient073 (304)
patient074 (698)
patient075 (655)
patient076 (697)
patient077 (595)
patient078 (556)
patient079 (482)
patient080 (696)
Proband X1 (540)
Proband X10 (431)
Proband X11 (725)
Proband X12 (536)
Proband X13 (486)
Proband X14 (558)
Proband X15 (382)
Proband X2 (427)
Proband X4 (742)
Proband X5 (595)
Proband X6 (443)
Proband X7 (625)
Proband X8 (348)
Proband X9 (713)
sim (377)
2.5
0.0
−2.5
−5.0
wavelet−HHH_gldm_HighGrayLevelEmphasis
frame

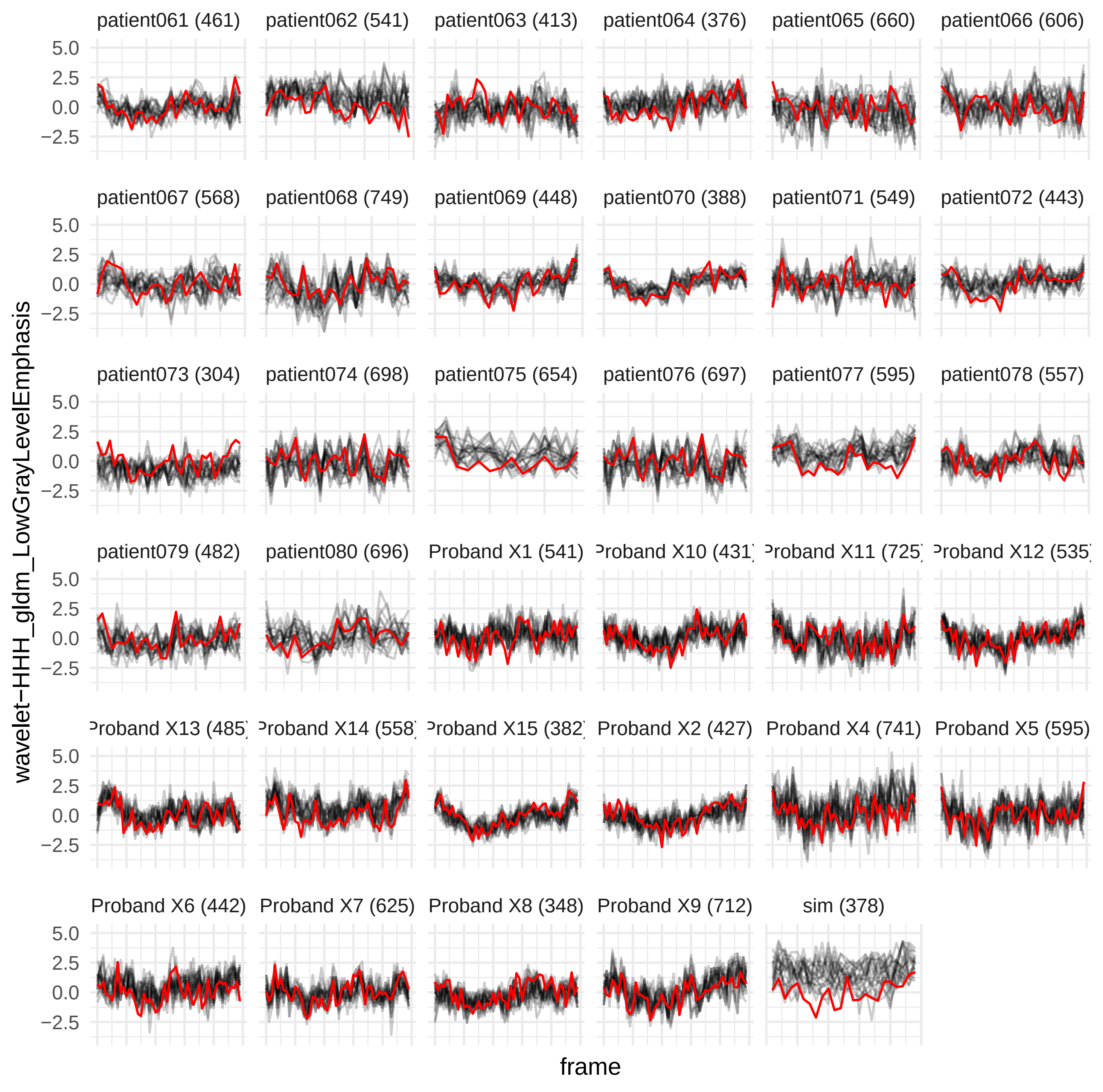
patient061 (461)
patient062 (541)
patient063 (413)
patient064 (376)
patient065 (660)
patient066 (606)
patient067 (568)
patient068 (749)
patient069 (448)
patient070 (388)
patient071 (549)
patient072 (443)
patient073 (304)
patient074 (698)
patient075 (654)
patient076 (697)
patient077 (595)
patient078 (557)
patient079 (482)
patient080 (696)
Proband X1 (541)
Proband X10 (431)
Proband X11 (725)
Proband X12 (535)
Proband X13 (485)
Proband X14 (558)
Proband X15 (382)
Proband X2 (427)
Proband X4 (741)
Proband X5 (595)
Proband X6 (442)
Proband X7 (625)
Proband X8 (348)
Proband X9 (712)
sim (378)
5.0
2.5
0.0
−2.5
wavelet−HHH_gldm_LowGrayLevelEmphasis
frame

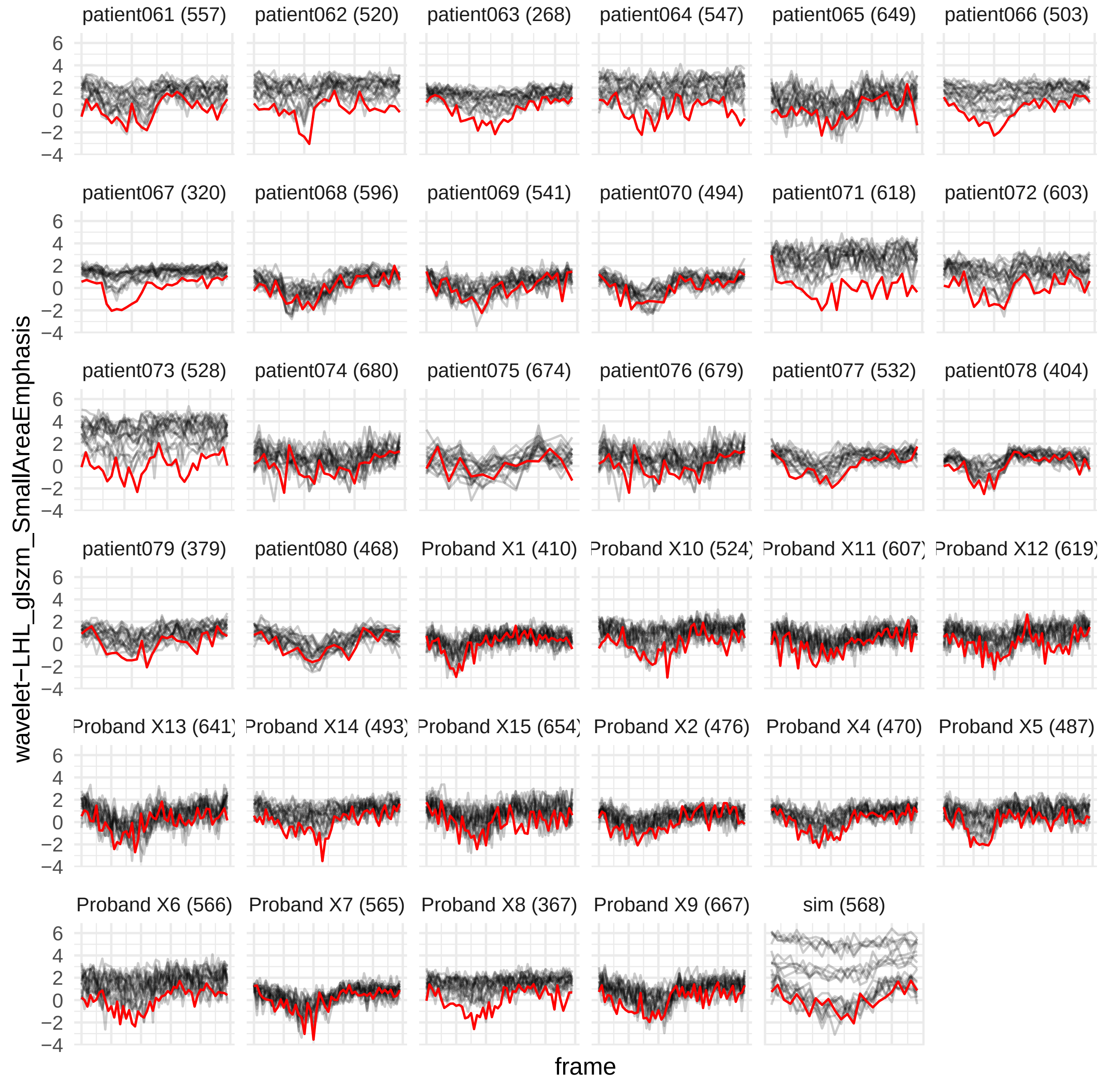

patient061 (557)
patient062 (520)
patient063 (268)
patient064 (547)
patient065 (649)
patient066 (503)
patient067 (320)
patient068 (596)
patient069 (541)
patient070 (494)
patient071 (618)
patient072 (603)
patient073 (528)
patient074 (680)
patient075 (674)
patient076 (679)
patient077 (532)
patient078 (404)
patient079 (379)
patient080 (468)
Proband X1 (410)
Proband X10 (524)
Proband X11 (607)
Proband X12 (619)
Proband X13 (641)
Proband X14 (493)
Proband X15 (654)
Proband X2 (476)
Proband X4 (470)
Proband X5 (487)
Proband X6 (566)
Proband X7 (565)
Proband X8 (367)
Proband X9 (667)
sim (568)
6
4
2
0
−2
−4
wavelet−LHL_glszm_SmallAreaEmphasis
frame

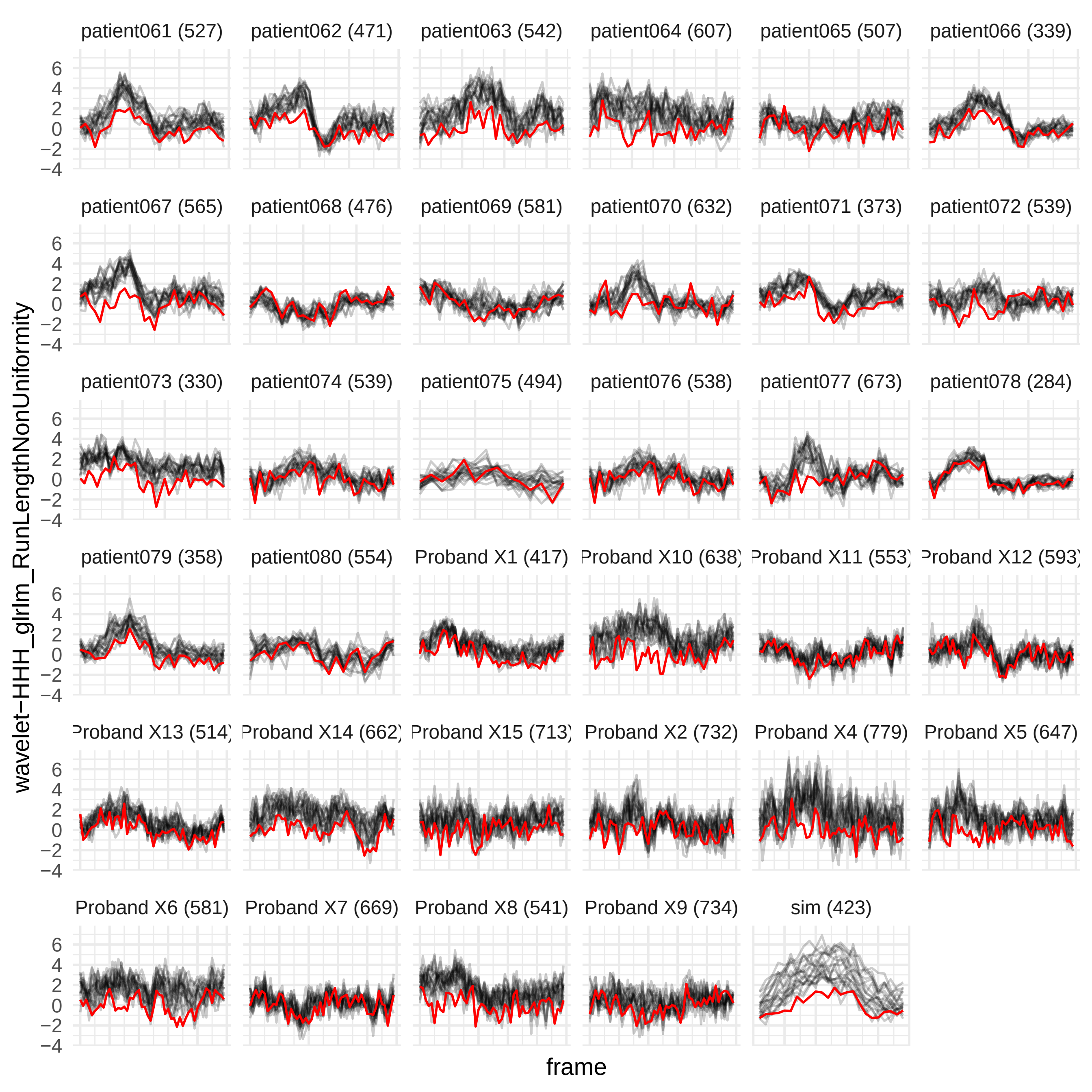
patient061 (527)
patient062 (471)
patient063 (542)
patient064 (607)
patient065 (507)
patient066 (339)
patient067 (565)
patient068 (476)
patient069 (581)
patient070 (632)
patient071 (373)
patient072 (539)
patient073 (330)
patient074 (539)
patient075 (494)
patient076 (538)
patient077 (673)
patient078 (284)
patient079 (358)
patient080 (554)
Proband X1 (417)
Proband X10 (638)
Proband X11 (553)
Proband X12 (593)
Proband X13 (514)
Proband X14 (662)
Proband X15 (713)
Proband X2 (732)
Proband X4 (779)
Proband X5 (647)
Proband X6 (581)
Proband X7 (669)
Proband X8 (541)
Proband X9 (734)
sim (423)
wavelet−HHH_glrlm_RunLengthNonUniformity
frame
6
4
2
0
−2
−4

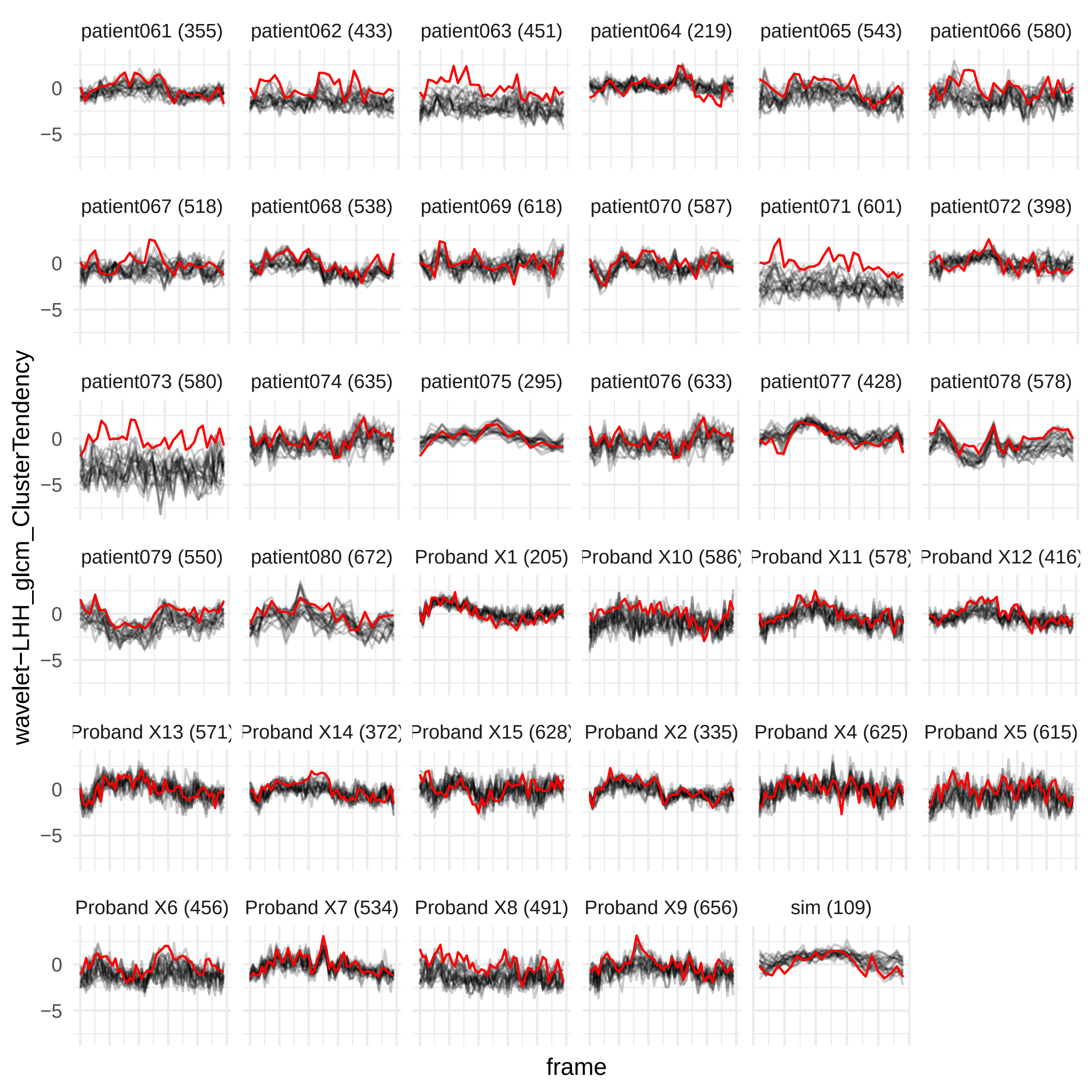
patient061 (355)
patient062 (433)
patient063 (451)
patient064 (219)
patient065 (543)
patient066 (580)
patient067 (518)
patient068 (538)
patient069 (618)
patient070 (587)
patient071 (601)
patient072 (398)
patient073 (580)
patient074 (635)
patient075 (295)
patient076 (633)
patient077 (428)
patient078 (578)
patient079 (550)
patient080 (672)
Proband X1 (205)
Proband X10 (586)
Proband X11 (578)
Proband X12 (416)
Proband X13 (571)
Proband X14 (372)
Proband X15 (628)
Proband X2 (335)
Proband X4 (625)
Proband X5 (615)
Proband X6 (456)
Proband X7 (534)
Proband X8 (491)
Proband X9 (656)
sim (109)
0
−5
wavelet−LHH_glcm_ClusterTendency
frame

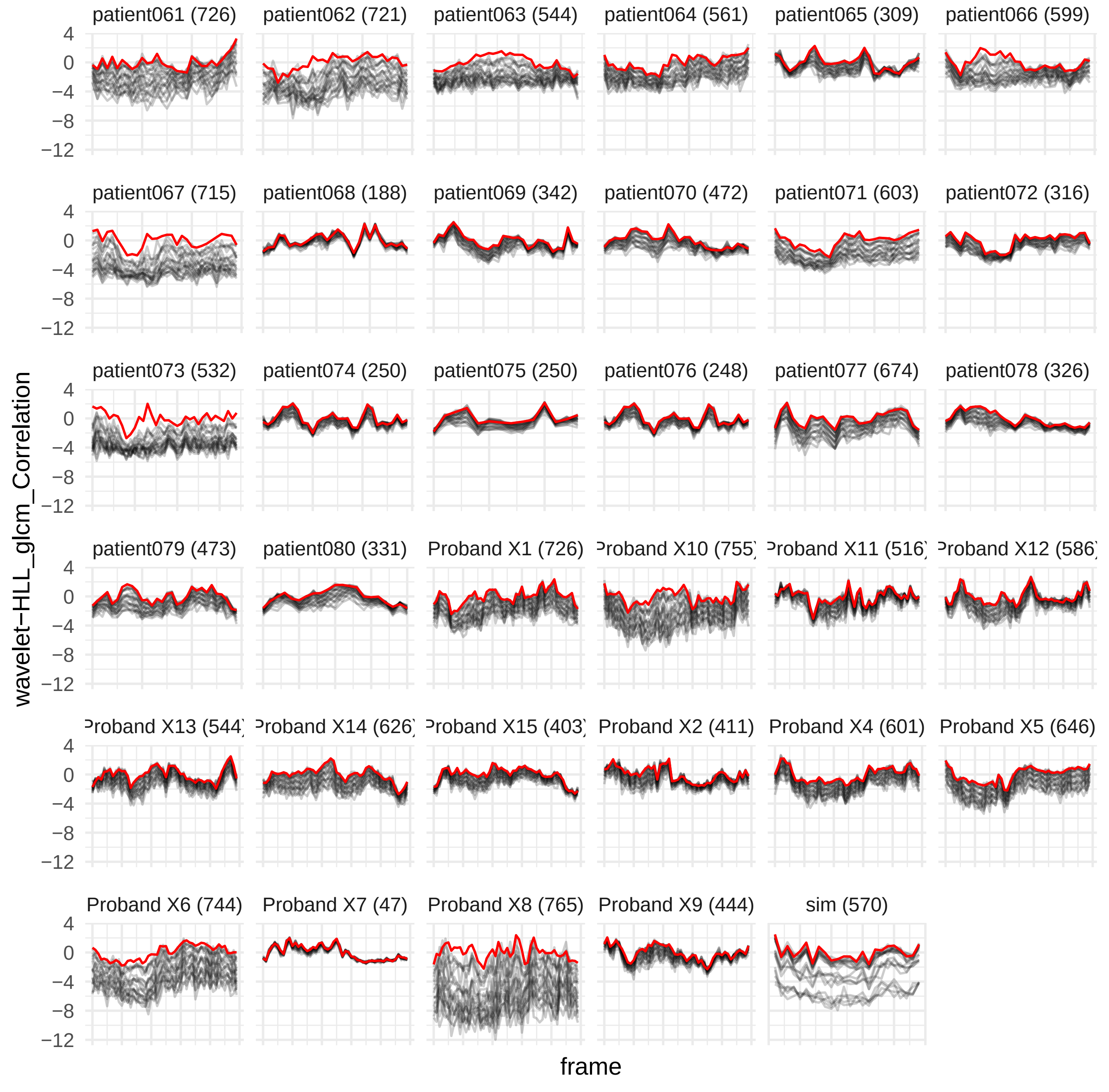

patient061 (726)
patient062 (721)
patient063 (544)
patient064 (561)
patient065 (309)
patient066 (599)
patient067 (715)
patient068 (188)
patient069 (342)
patient070 (472)
patient071 (603)
patient072 (316)
patient073 (532)
patient074 (250)
patient075 (250)
patient076 (248)
patient077 (674)
patient078 (326)
patient079 (473)
patient080 (331)
Proband X1 (726)
Proband X10 (755)
Proband X11 (516)
Proband X12 (586)
Proband X13 (544)
Proband X14 (626)
Proband X15 (403)
Proband X2 (411)
Proband X4 (601)
Proband X5 (646)
Proband X6 (744)
Proband X7 (47)
Proband X8 (765)
Proband X9 (444)
sim (570)
4
0
−4
−8
−12
wavelet−HLL_glcm_Correlation
frame

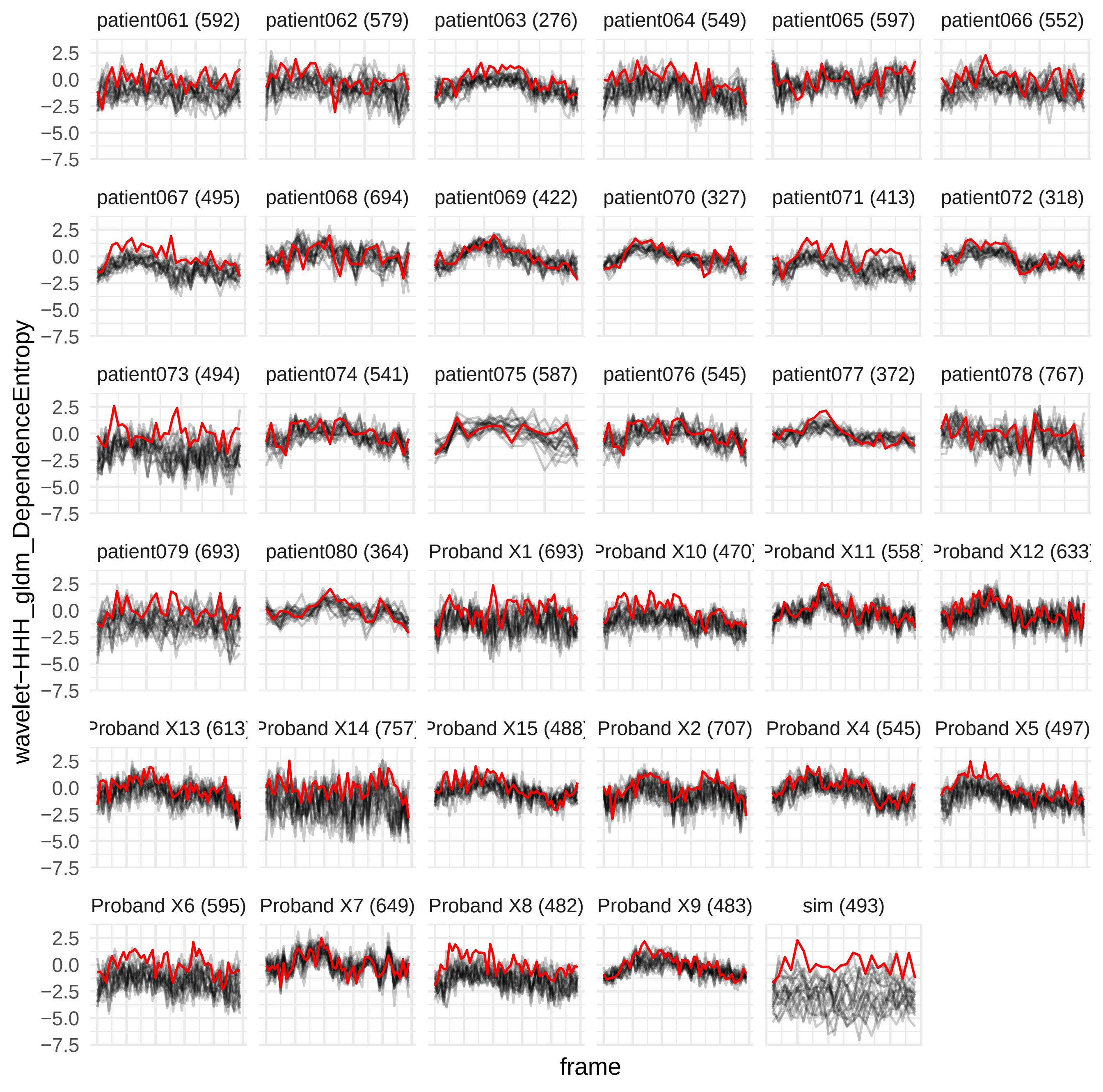
patient061 (592)
patient062 (579)
patient063 (276)
patient064 (549)
patient065 (597)
patient066 (552)
patient067 (495)
patient068 (694)
patient069 (422)
patient070 (327)
patient071 (413)
patient072 (318)
patient073 (494)
patient074 (541)
patient075 (587)
patient076 (545)
patient077 (372)
patient078 (767)
patient079 (693)
patient080 (364)
Proband X1 (693)
Proband X10 (470)
Proband X11 (558)
Proband X12 (633)
Proband X13 (613)
Proband X14 (757)
Proband X15 (488)
Proband X2 (707)
Proband X4 (545)
Proband X5 (497)
Proband X6 (595)
Proband X7 (649)
Proband X8 (482)
Proband X9 (483)
sim (493)
2.5
0.0
−2.5
−5.0
−7.5
wavelet−HHH_gldm_DependenceEntropy
frame

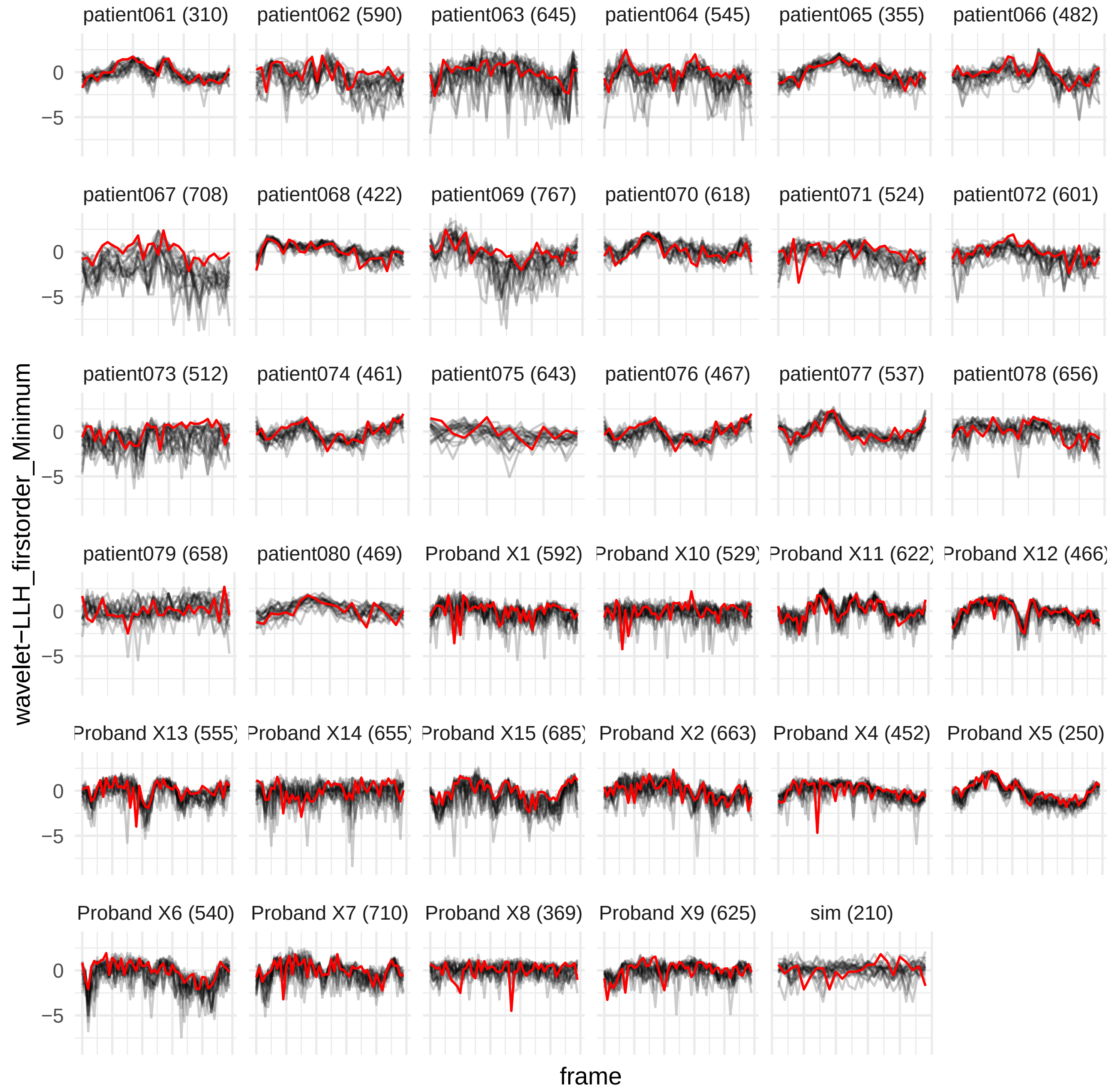
patient061 (310)
patient062 (590)
patient063 (645)
patient064 (545)
patient065 (355)
patient066 (482)
patient067 (708)
patient068 (422)
patient069 (767)
patient070 (618)
patient071 (524)
patient072 (601)
patient073 (512)
patient074 (461)
patient075 (643)
patient076 (467)
patient077 (537)
patient078 (656)
patient079 (658)
patient080 (469)
Proband X1 (592)
Proband X10 (529)
Proband X11 (622)
Proband X12 (466)
Proband X13 (555)
Proband X14 (655)
Proband X15 (685)
Proband X2 (663)
Proband X4 (452)
Proband X5 (250)
Proband X6 (540)
Proband X7 (710)
Proband X8 (369)
Proband X9 (625)
sim (210)
0
−5
wavelet−LLH_firstorder_Minimum
frame

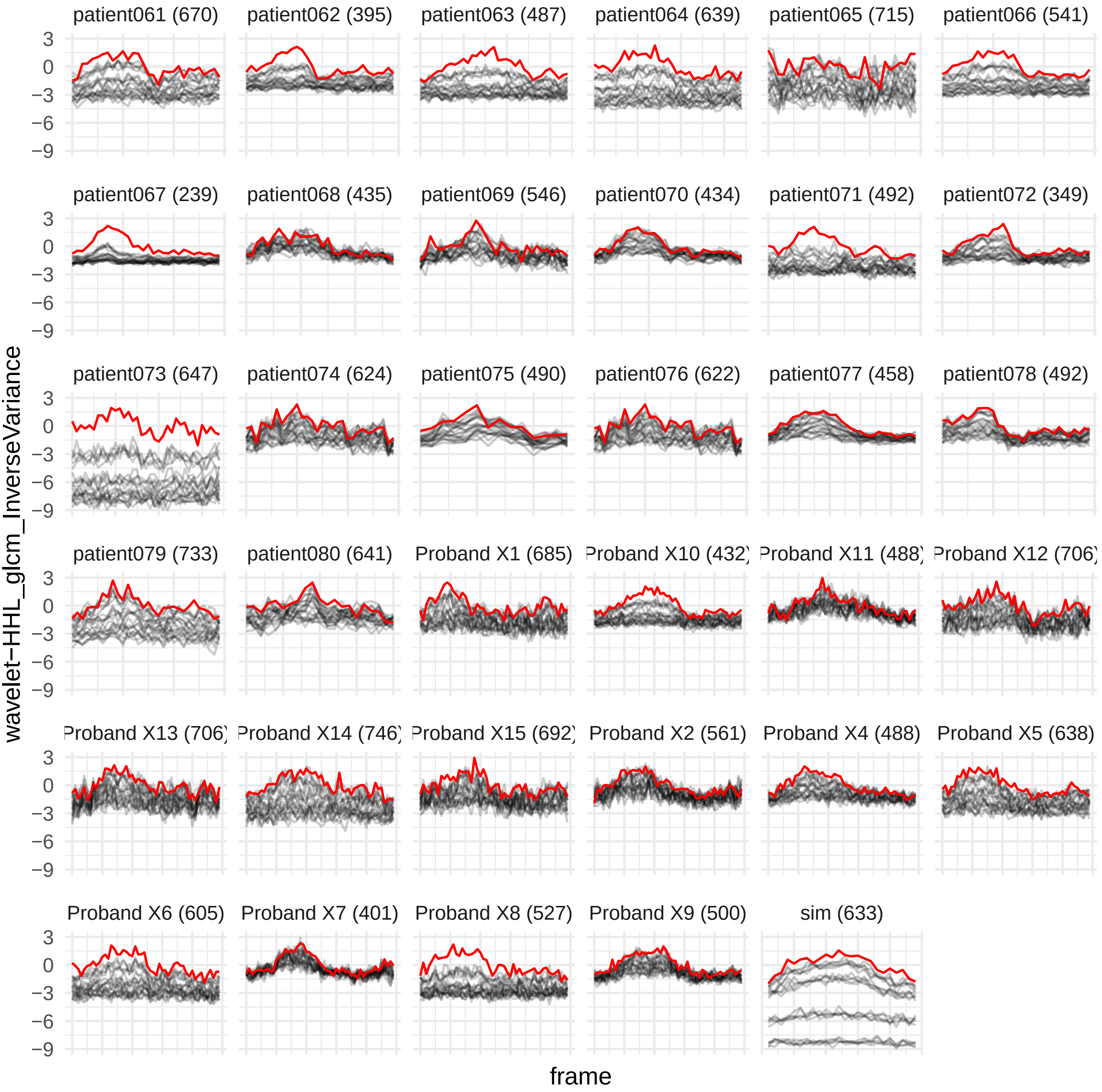

patient061 (670)
patient062 (395)
patient063 (487)
patient064 (639)
patient065 (715)
patient066 (541)
patient067 (239)
patient068 (435)
patient069 (546)
patient070 (434)
patient071 (492)
patient072 (349)
patient073 (647)
patient074 (624)
patient075 (490)
patient076 (622)
patient077 (458)
patient078 (492)
patient079 (733)
patient080 (641)
Proband X1 (685)
Proband X10 (432)
Proband X11 (488)
Proband X12 (706)
Proband X13 (706)
Proband X14 (746)
Proband X15 (692)
Proband X2 (561)
Proband X4 (488)
Proband X5 (638)
Proband X6 (605)
Proband X7 (401)
Proband X8 (527)
Proband X9 (500)
sim (633)
3
0
−3
−6
−9
wavelet−HHL_glcm_InverseVariance
frame

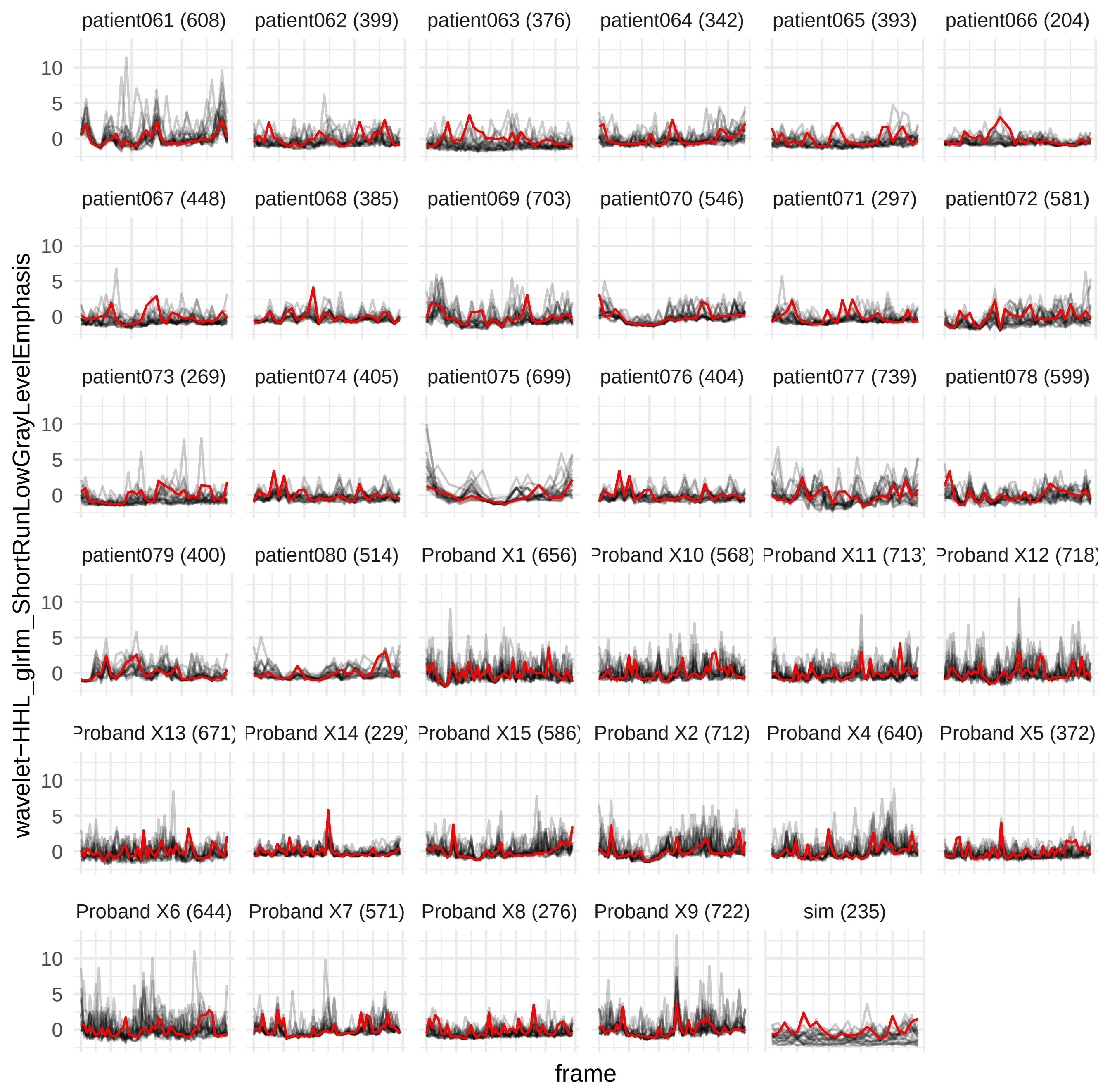
patient061 (608)
patient062 (399)
patient063 (376)
patient064 (342)
patient065 (393)
patient066 (204)
patient067 (448)
patient068 (385)
patient069 (703)
patient070 (546)
patient071 (297)
patient072 (581)
patient073 (269)
patient074 (405)
patient075 (699)
patient076 (404)
patient077 (739)
patient078 (599)
patient079 (400)
patient080 (514)
Proband X1 (656)
Proband X10 (568)
Proband X11 (713)
Proband X12 (718)
Proband X13 (671)
Proband X14 (229)
Proband X15 (586)
Proband X2 (712)
Proband X4 (640)
Proband X5 (372)
Proband X6 (644)
Proband X7 (571)
Proband X8 (276)
Proband X9 (722)
sim (235)
10
5
0
wavelet−HHL_glrlm_ShortRunLowGrayLevelEmphasis
frame

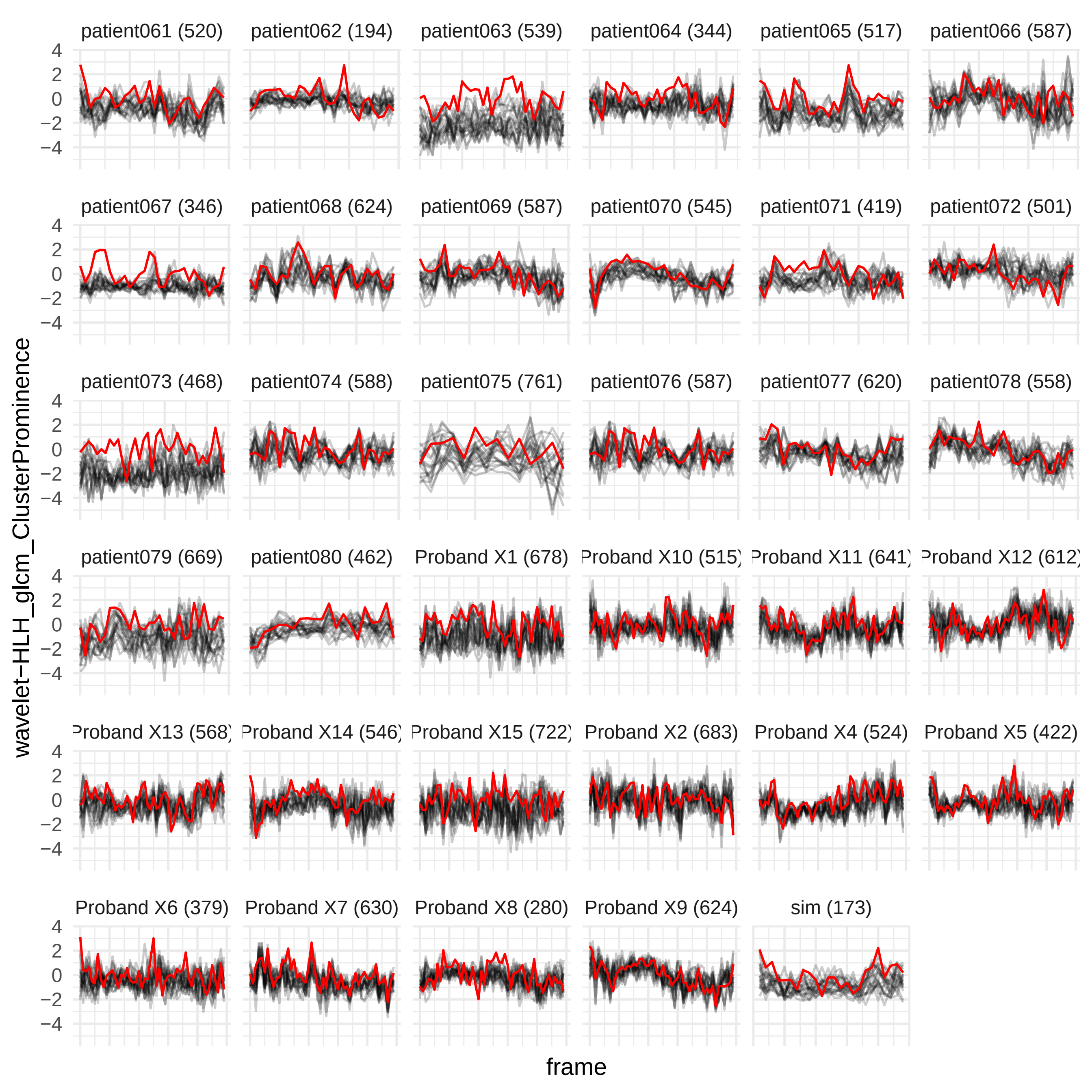

patient061 (520)
patient062 (194)
patient063 (539)
patient064 (344)
patient065 (517)
patient066 (587)
patient067 (346)
patient068 (624)
patient069 (587)
patient070 (545)
patient071 (419)
patient072 (501)
patient073 (468)
patient074 (588)
patient075 (761)
patient076 (587)
patient077 (620)
patient078 (558)
patient079 (669)
patient080 (462)
Proband X1 (678)
Proband X10 (515)
Proband X11 (641)
Proband X12 (612)
Proband X13 (568)
Proband X14 (546)
Proband X15 (722)
Proband X2 (683)
Proband X4 (524)
Proband X5 (422)
Proband X6 (379)
Proband X7 (630)
Proband X8 (280)
Proband X9 (624)
sim (173)
wavelet-HLH_glcm_ClusterProminence
frame

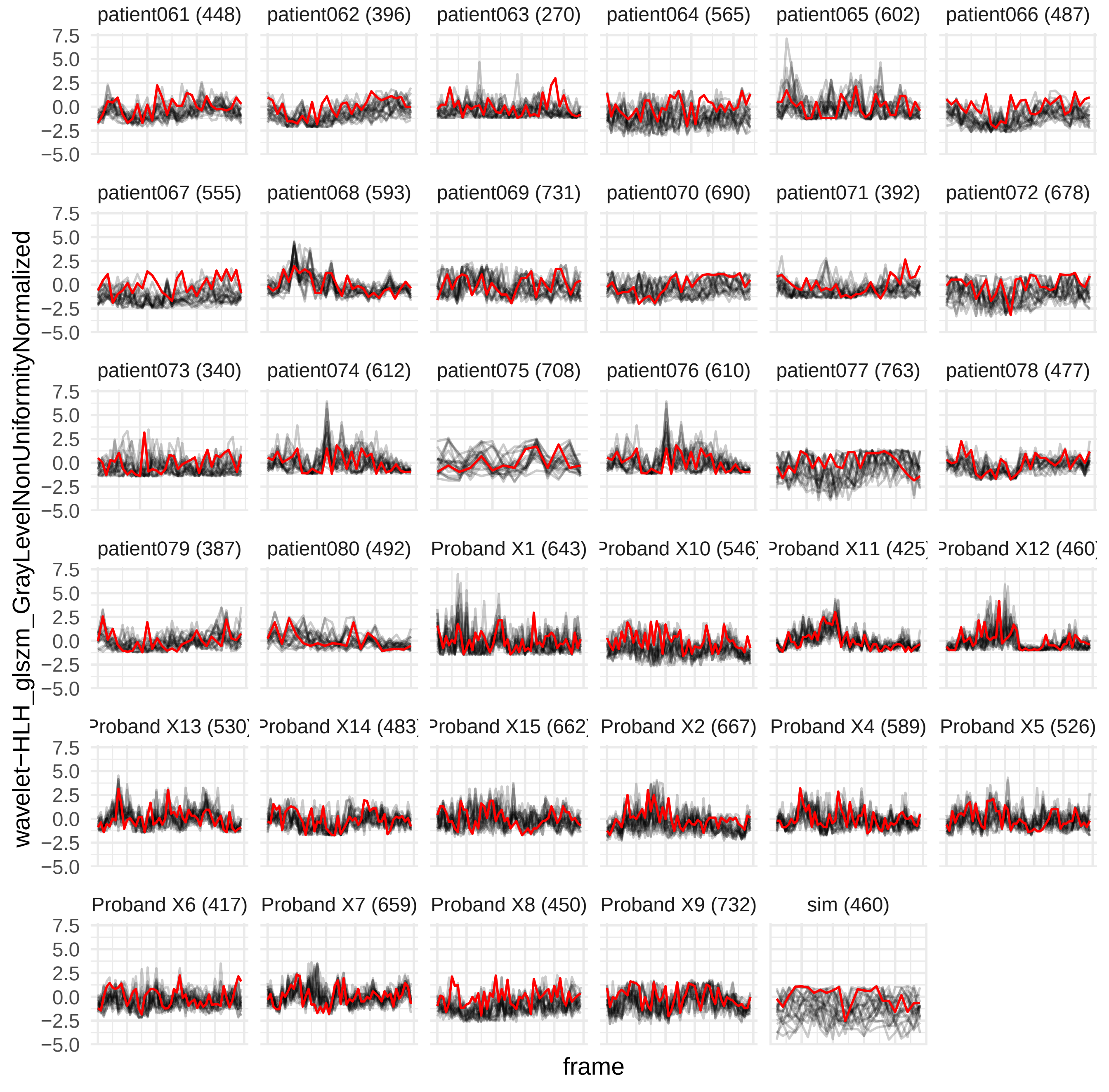

patient061 (448)
patient062 (396)
patient063 (270)
patient064 (565)
patient065 (602)
patient066 (487)
patient067 (555)
patient068 (593)
patient069 (731)
patient070 (690)
patient071 (392)
patient072 (678)
patient073 (340)
patient074 (612)
patient075 (708)
patient076 (610)
patient077 (763)
patient078 (477)
patient079 (387)
patient080 (492)
Proband X1 (643)
Proband X10 (546)
Proband X11 (425)
Proband X12 (460)
Proband X13 (530)
Proband X14 (483)
Proband X15 (662)
Proband X2 (667)
Proband X4 (589)
Proband X5 (526)
Proband X6 (417)
Proband X7 (659)
Proband X8 (450)
Proband X9 (732)
sim (460)
7.5
5.0
2.5
0.0
−2.5
−5.0
wavelet−HLH_glszm_GrayLevelNonUniformityNormalized
frame

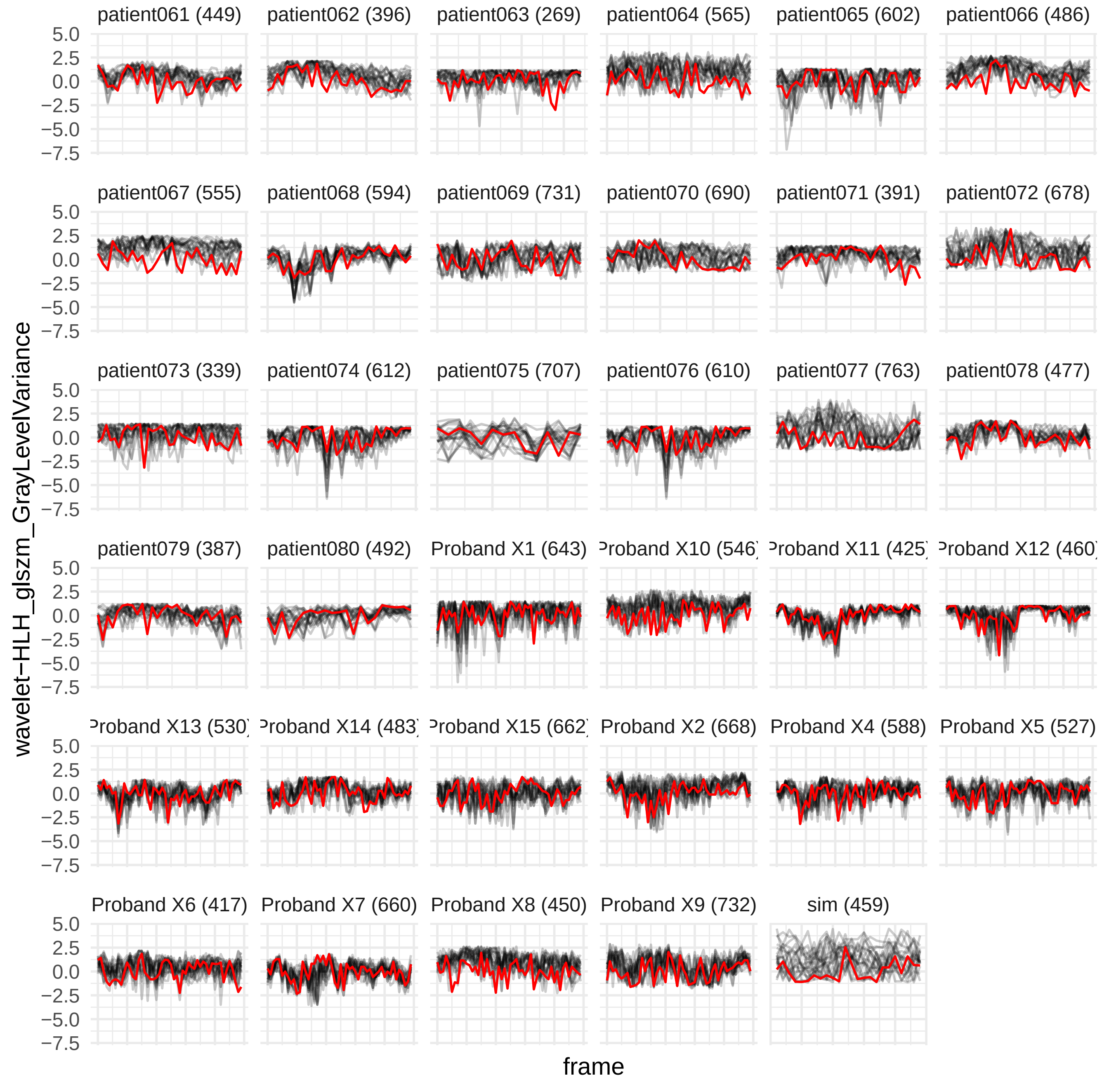
patient061 (449)
patient062 (396)
patient063 (269)
patient064 (565)
patient065 (602)
patient066 (486)
patient067 (555)
patient068 (594)
patient069 (731)
patient070 (690)
patient071 (391)
patient072 (678)
patient073 (339)
patient074 (612)
patient075 (707)
patient076 (610)
patient077 (763)
patient078 (477)
patient079 (387)
patient080 (492)
Proband X1 (643)
Proband X10 (546)
Proband X11 (425)
Proband X12 (460)
Proband X13 (530)
Proband X14 (483)
Proband X15 (662)
Proband X2 (668)
Proband X4 (588)
Proband X5 (527)
Proband X6 (417)
Proband X7 (660)
Proband X8 (450)
Proband X9 (732)
sim (459)
5.0
2.5
0.0
−2.5
−5.0
−7.5
wavelet−HLH_glszm_GrayLevelVariance
frame

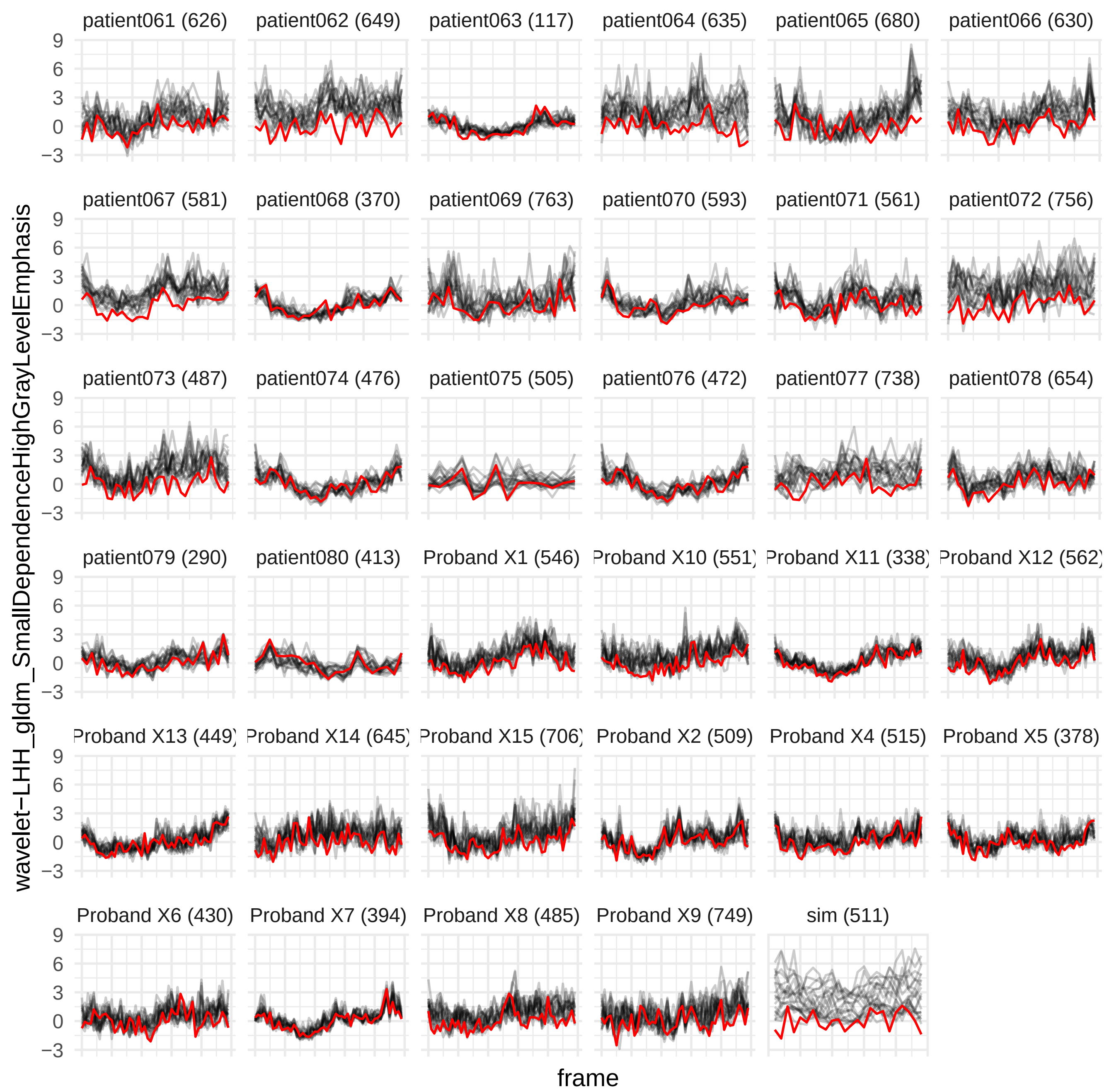

patient061 (626)
patient062 (649)
patient063 (117)
patient064 (635)
patient065 (680)
patient066 (630)
patient067 (581)
patient068 (370)
patient069 (763)
patient070 (593)
patient071 (561)
patient072 (756)
patient073 (487)
patient074 (476)
patient075 (505)
patient076 (472)
patient077 (738)
patient078 (654)
patient079 (290)
patient080 (413)
Proband X1 (546)
Proband X10 (551)
Proband X11 (338)
Proband X12 (562)
Proband X13 (449)
Proband X14 (645)
Proband X15 (706)
Proband X2 (509)
Proband X4 (515)
Proband X5 (378)
Proband X6 (430)
Proband X7 (394)
Proband X8 (485)
Proband X9 (749)
sim (511)
9
6
3
0
−3
wavelet−LHH_gldm_SmallDependenceHighGrayLevelEmphasis
frame

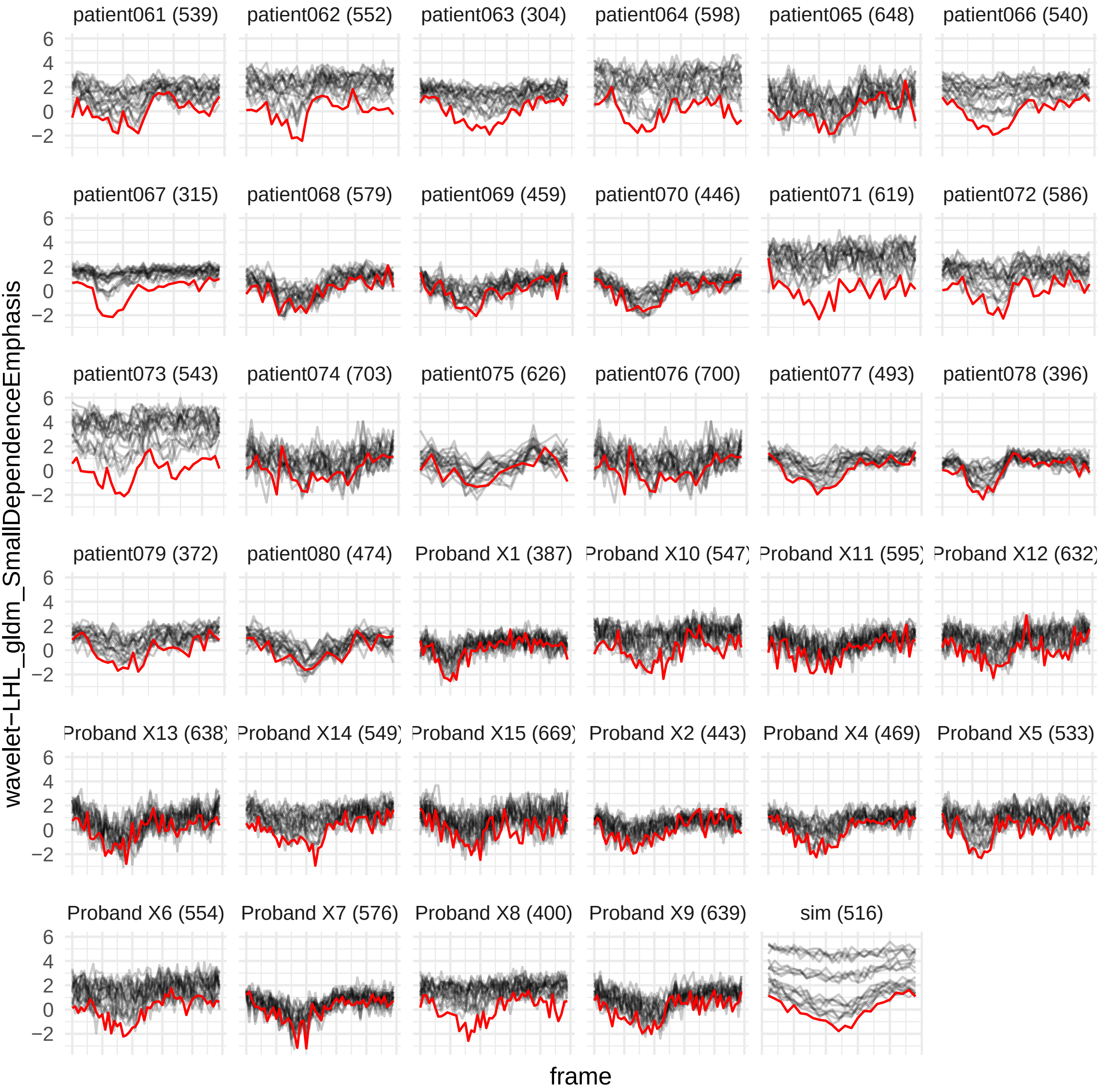
patient061 (539)
patient062 (552)
patient063 (304)
patient064 (598)
patient065 (648)
patient066 (540)
patient067 (315)
patient068 (579)
patient069 (459)
patient070 (446)
patient071 (619)
patient072 (586)
patient073 (543)
patient074 (703)
patient075 (626)
patient076 (700)
patient077 (493)
patient078 (396)
patient079 (372)
patient080 (474)
Proband X1 (387)
Proband X10 (547)
Proband X11 (595)
Proband X12 (632)
Proband X13 (638)
Proband X14 (549)
Proband X15 (669)
Proband X2 (443)
Proband X4 (469)
Proband X5 (533)
Proband X6 (554)
Proband X7 (576)
Proband X8 (400)
Proband X9 (639)
sim (516)
wavelet−LHL_gldm_SmallDependenceEmphasis
frame

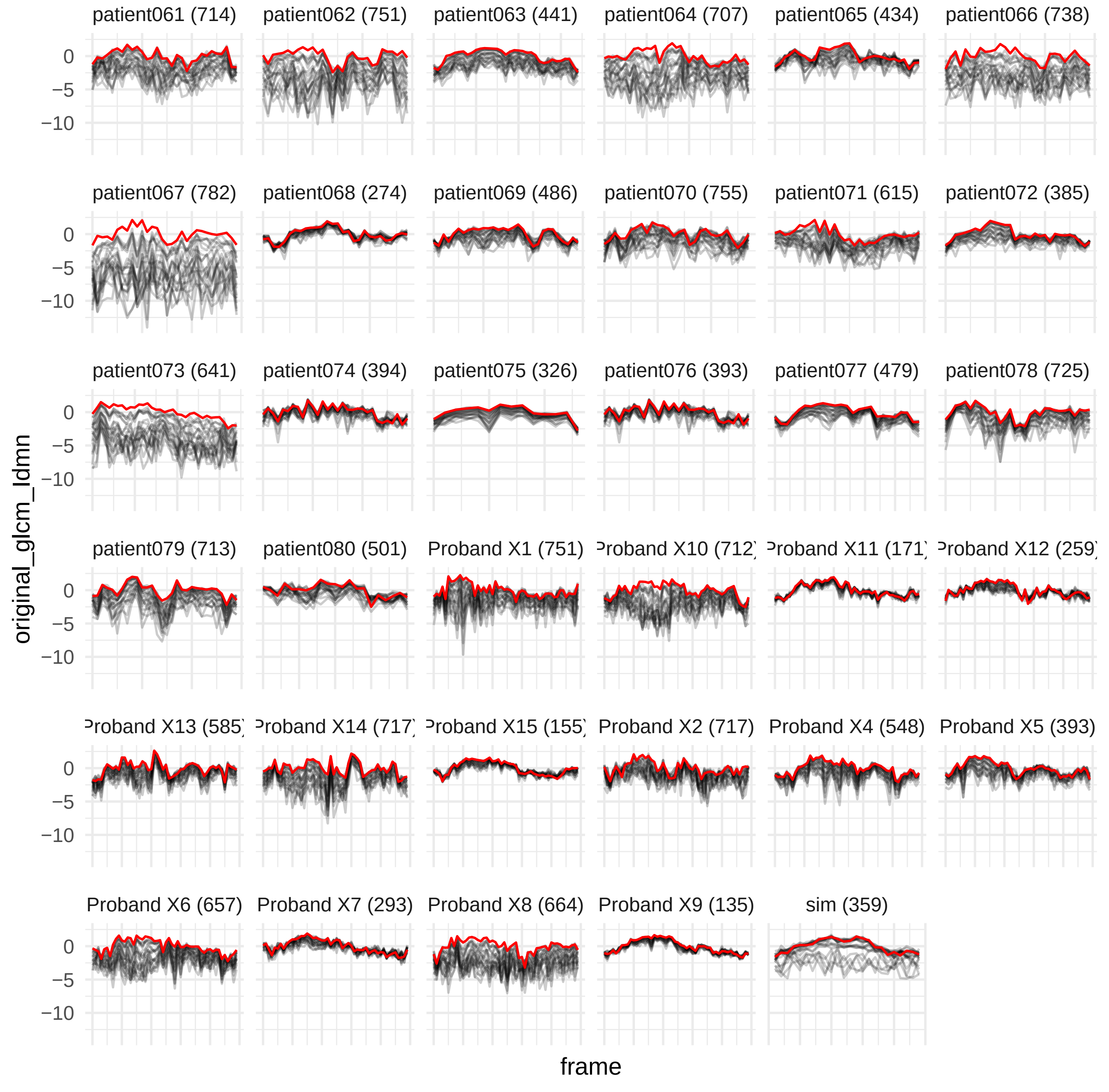

patient061 (714)
patient062 (751)
patient063 (441)
patient064 (707)
patient065 (434)
patient066 (738)
patient067 (782)
patient068 (274)
patient069 (486)
patient070 (755)
patient071 (615)
patient072 (385)
patient073 (641)
patient074 (394)
patient075 (326)
patient076 (393)
patient077 (479)
patient078 (725)
patient079 (713)
patient080 (501)
Proband X1 (751)
Proband X10 (712)
Proband X11 (171)
Proband X12 (259)
Proband X13 (585)
Proband X14 (717)
Proband X15 (155)
Proband X2 (717)
Proband X4 (548)
Proband X5 (393)
Proband X6 (657)
Proband X7 (293)
Proband X8 (664)
Proband X9 (135)
sim (359)
original_glcm_ldmn
frame
0
−5
−10

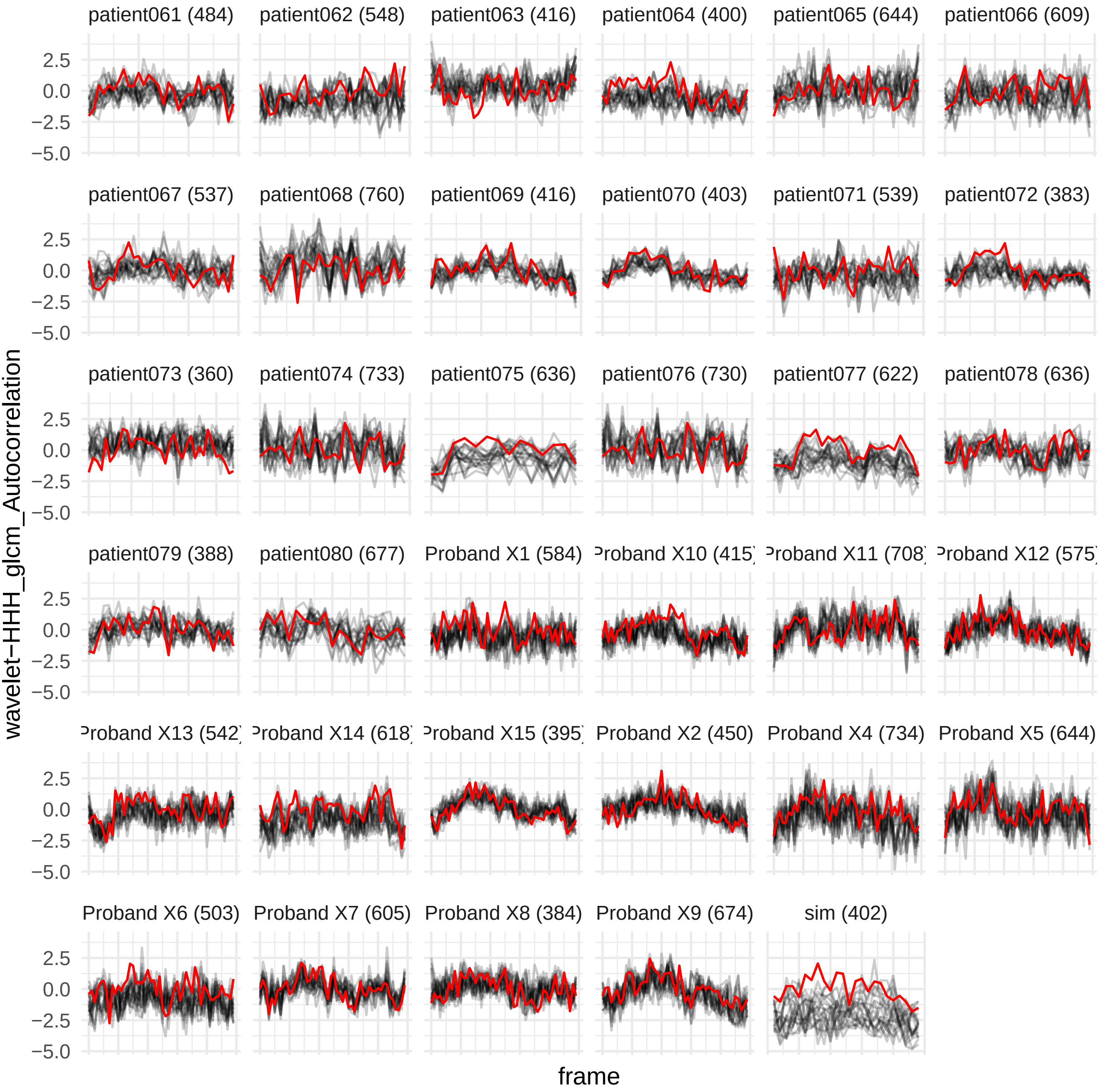

patient061 (484)
patient062 (548)
patient063 (416)
patient064 (400)
patient065 (644)
patient066 (609)
patient067 (537)
patient068 (760)
patient069 (416)
patient070 (403)
patient071 (539)
patient072 (383)
patient073 (360)
patient074 (733)
patient075 (636)
patient076 (730)
patient077 (622)
patient078 (636)
patient079 (388)
patient080 (677)
Proband X1 (584)
Proband X10 (415)
Proband X11 (708)
Proband X12 (575)
Proband X13 (542)
Proband X14 (618)
Proband X15 (395)
Proband X2 (450)
Proband X4 (734)
Proband X5 (644)
Proband X6 (503)
Proband X7 (605)
Proband X8 (384)
Proband X9 (674)
sim (402)
2.5
0.0
−2.5
−5.0
wavelet−HHH_glcm_Autocorrelation
frame

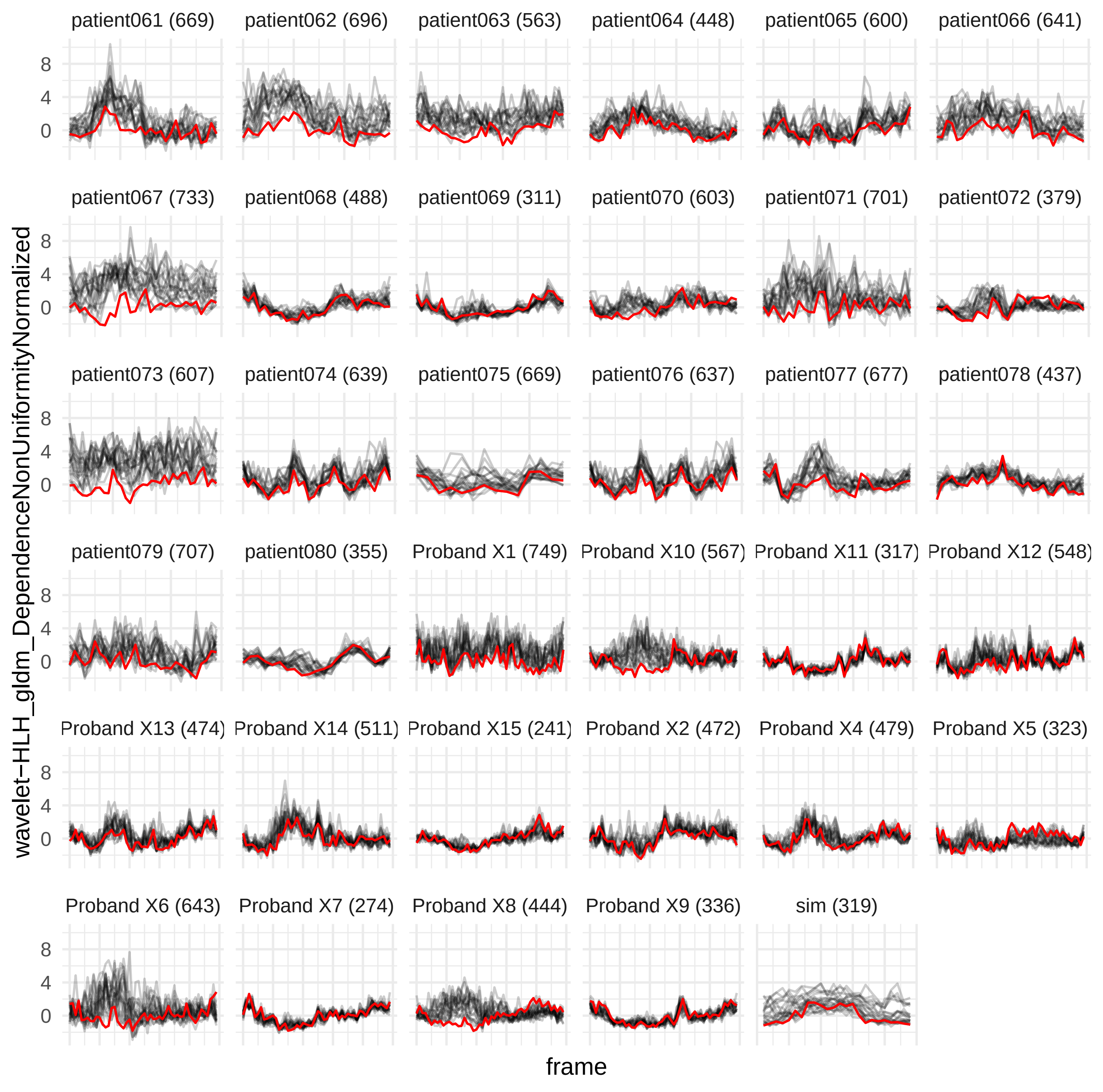
patient061 (669)
patient062 (696)
patient063 (563)
patient064 (448)
patient065 (600)
patient066 (641)
patient067 (733)
patient068 (488)
patient069 (311)
patient070 (603)
patient071 (701)
patient072 (379)
patient073 (607)
patient074 (639)
patient075 (669)
patient076 (637)
patient077 (677)
patient078 (437)
patient079 (707)
patient080 (355)
Proband X1 (749)
Proband X10 (567)
Proband X11 (317)
Proband X12 (548)
Proband X13 (474)
Proband X14 (511)
Proband X15 (241)
Proband X2 (472)
Proband X4 (479)
Proband X5 (323)
Proband X6 (643)
Proband X7 (274)
Proband X8 (444)
Proband X9 (336)
sim (319)
8
4
0
wavelet−HLH_gldm_DependenceNonUniformityNormalized
frame

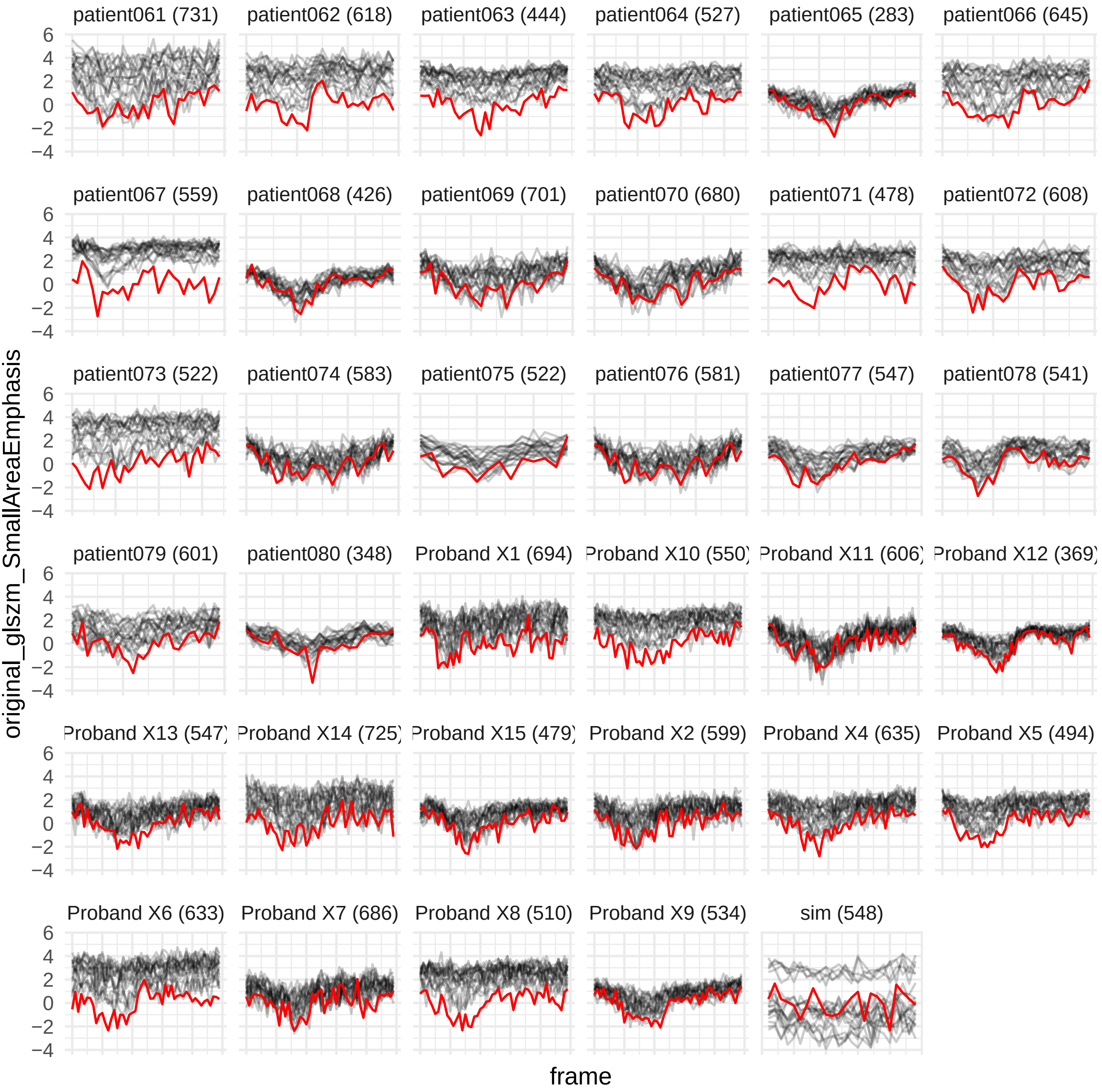
patient061 (731)
patient062 (618)
patient063 (444)
patient064 (527)
patient065 (283)
patient066 (645)
patient067 (559)
patient068 (426)
patient069 (701)
patient070 (680)
patient071 (478)
patient072 (608)
patient073 (522)
patient074 (583)
patient075 (522)
patient076 (581)
patient077 (547)
patient078 (541)
patient079 (601)
patient080 (348)
Proband X1 (694)
Proband X10 (550)
Proband X11 (606)
Proband X12 (369)
Proband X13 (547)
Proband X14 (725)
Proband X15 (479)
Proband X2 (599)
Proband X4 (635)
Proband X5 (494)
Proband X6 (633)
Proband X7 (686)
Proband X8 (510)
Proband X9 (534)
sim (548)
6
4
2
0
−2
−4
original_glszm_SmallAreaEmphasis
frame

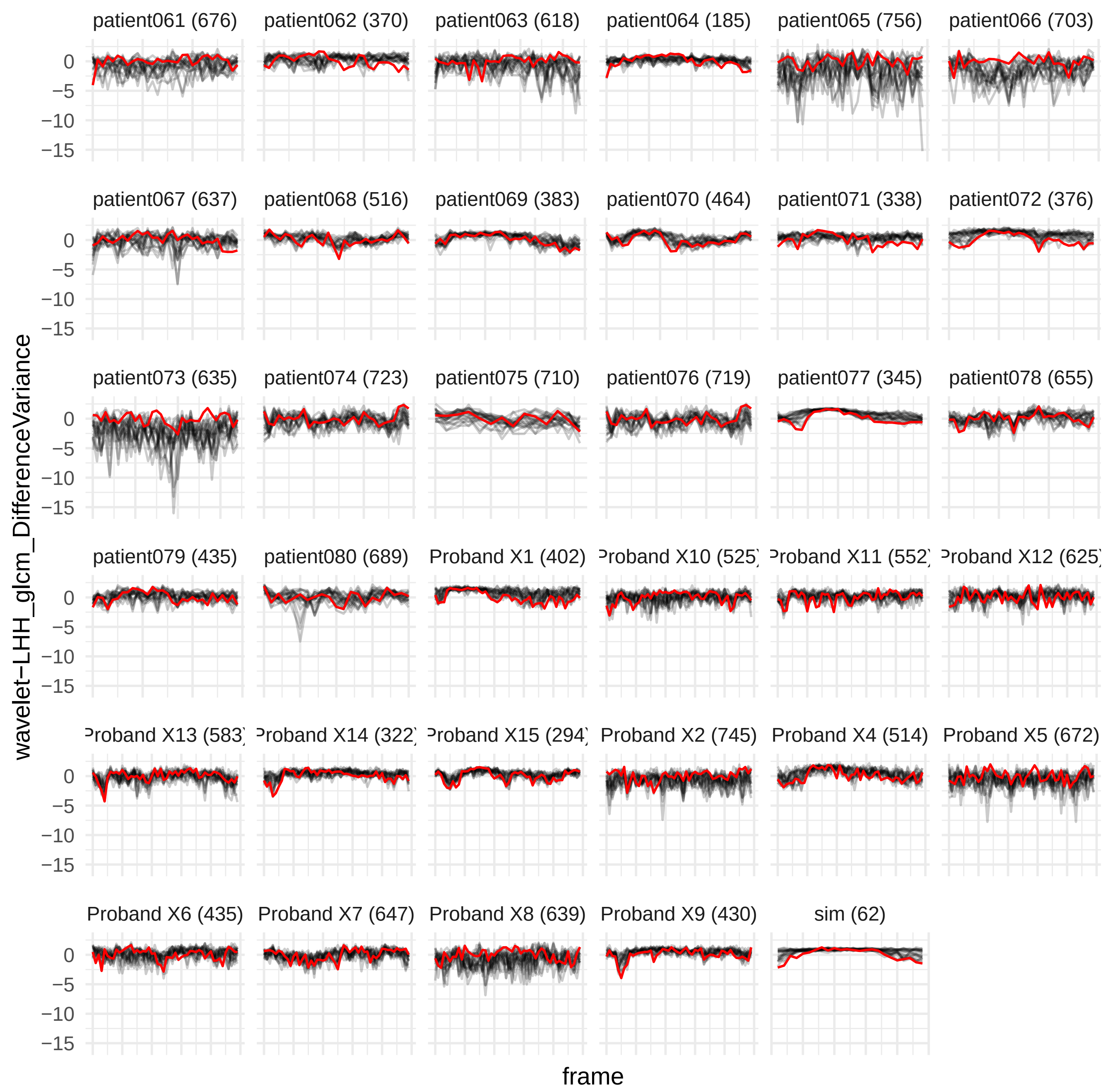

patient061 (676)
patient062 (370)
patient063 (618)
patient064 (185)
patient065 (756)
patient066 (703)
patient067 (637)
patient068 (516)
patient069 (383)
patient070 (464)
patient071 (338)
patient072 (376)
patient073 (635)
patient074 (723)
patient075 (710)
patient076 (719)
patient077 (345)
patient078 (655)
patient079 (435)
patient080 (689)
Proband X1 (402)
Proband X10 (525)
Proband X11 (552)
Proband X12 (625)
Proband X13 (583)
Proband X14 (322)
Proband X15 (294)
Proband X2 (745)
Proband X4 (514)
Proband X5 (672)
Proband X6 (435)
Proband X7 (647)
Proband X8 (639)
Proband X9 (430)
sim (62)
0
−5
−10
−15
wavelet−LHH_glcm_DifferenceVariance
frame

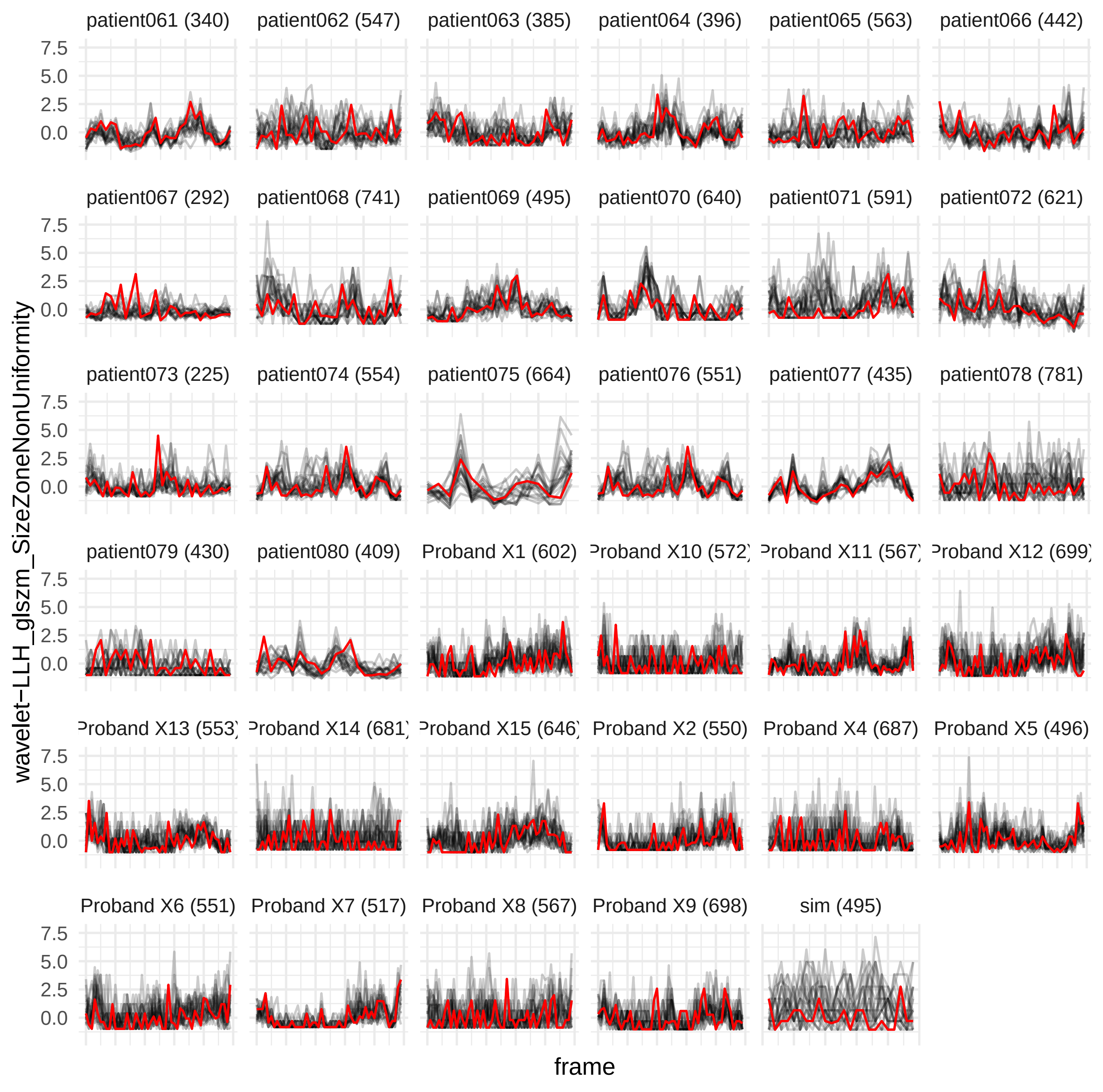
patient061 (340)
patient062 (547)
patient063 (385)
patient064 (396)
patient065 (563)
patient066 (442)
patient067 (292)
patient068 (741)
patient069 (495)
patient070 (640)
patient071 (591)
patient072 (621)
patient073 (225)
patient074 (554)
patient075 (664)
patient076 (551)
patient077 (435)
patient078 (781)
patient079 (430)
patient080 (409)
Proband X1 (602)
Proband X10 (572)
Proband X11 (567)
Proband X12 (699)
Proband X13 (553)
Proband X14 (681)
Proband X15 (646)
Proband X2 (550)
Proband X4 (687)
Proband X5 (496)
Proband X6 (551)
Proband X7 (517)
Proband X8 (567)
Proband X9 (698)
sim (495)
7.5
5.0
2.5
0.0
wavelet−LLH_glszm_SizeZoneNonUniformity
frame

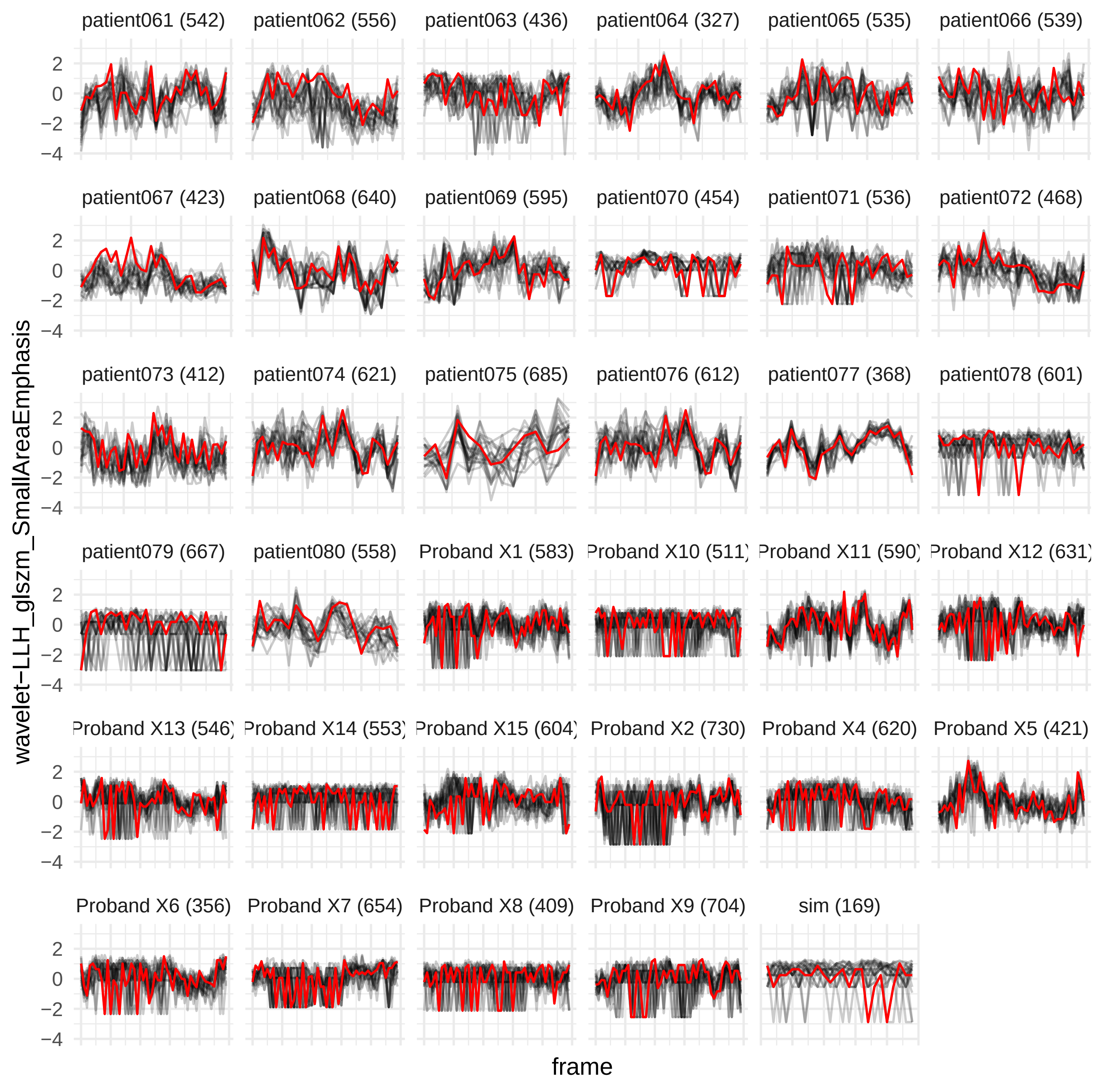

patient061 (542)
patient062 (556)
patient063 (436)
patient064 (327)
patient065 (535)
patient066 (539)
patient067 (423)
patient068 (640)
patient069 (595)
patient070 (454)
patient071 (536)
patient072 (468)
patient073 (412)
patient074 (621)
patient075 (685)
patient076 (612)
patient077 (368)
patient078 (601)
patient079 (667)
patient080 (558)
Proband X1 (583)
Proband X10 (511)
Proband X11 (590)
Proband X12 (631)
Proband X13 (546)
Proband X14 (553)
Proband X15 (604)
Proband X2 (730)
Proband X4 (620)
Proband X5 (421)
Proband X6 (356)
Proband X7 (654)
Proband X8 (409)
Proband X9 (704)
sim (169)
wavelet−LLH_glszm_SmallAreaEmphasis
frame
2
0
−2
−4

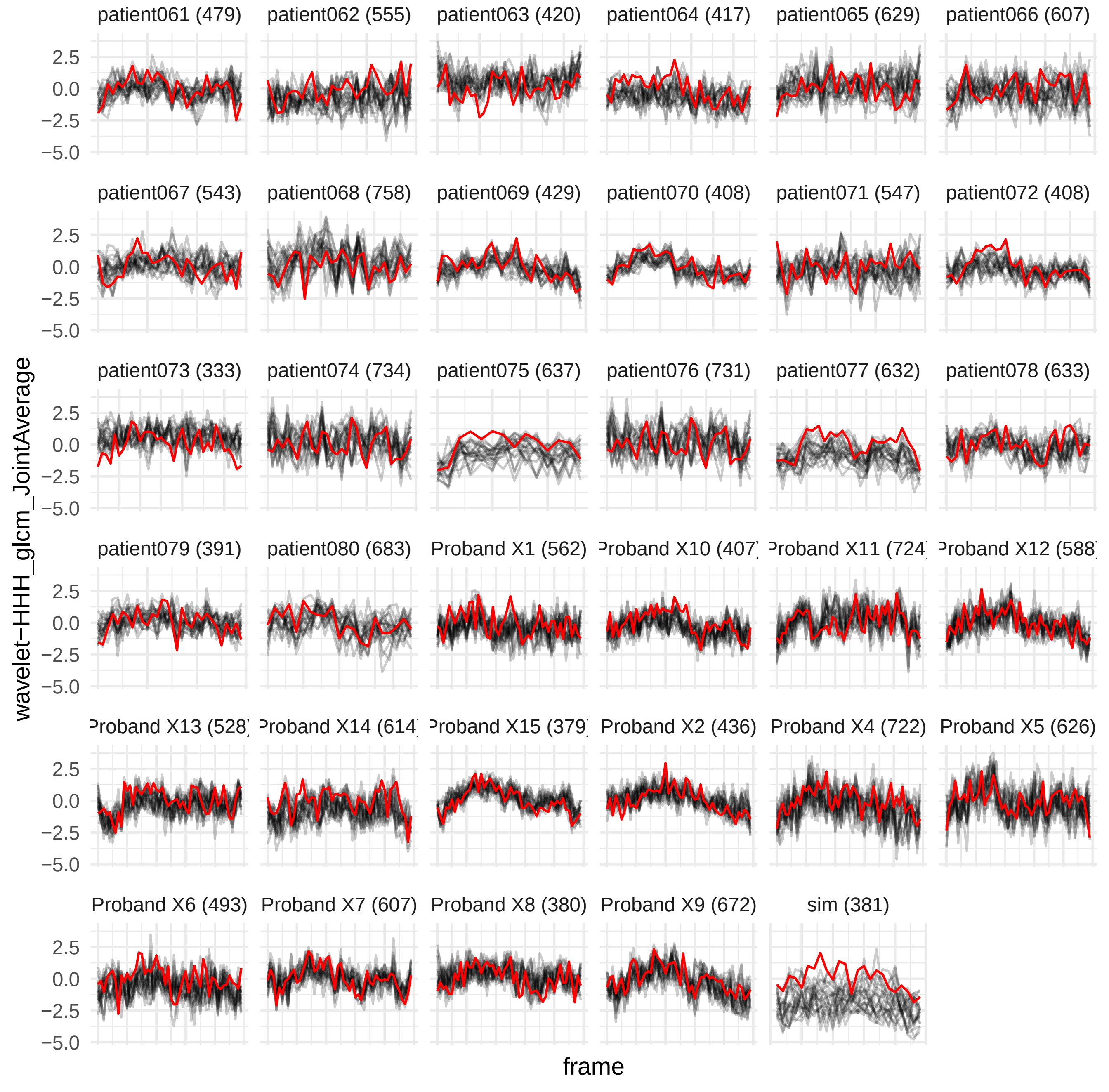

patient061 (479)
patient062 (555)
patient063 (420)
patient064 (417)
patient065 (629)
patient066 (607)
patient067 (543)
patient068 (758)
patient069 (429)
patient070 (408)
patient071 (547)
patient072 (408)
patient073 (333)
patient074 (734)
patient075 (637)
patient076 (731)
patient077 (632)
patient078 (633)
patient079 (391)
patient080 (683)
Proband X1 (562)
Proband X10 (407)
Proband X11 (724)
Proband X12 (588)
Proband X13 (528)
Proband X14 (614)
Proband X15 (379)
Proband X2 (436)
Proband X4 (722)
Proband X5 (626)
Proband X6 (493)
Proband X7 (607)
Proband X8 (380)
Proband X9 (672)
sim (381)
2.5
0.0
−2.5
−5.0
wavelet−HHH_glcm_JointAverage
frame

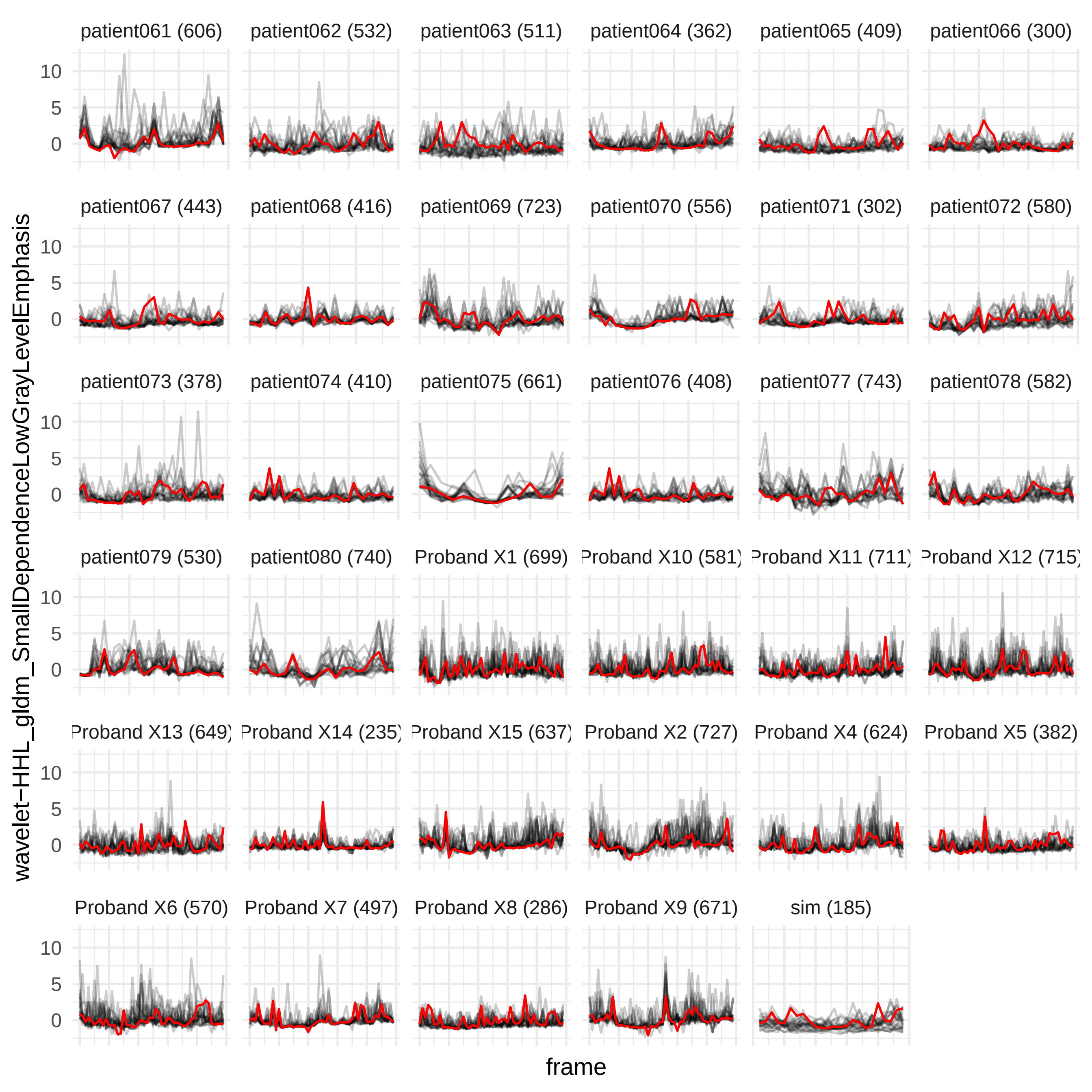

patient061 (606)
patient062 (532)
patient063 (511)
patient064 (362)
patient065 (409)
patient066 (300)
patient067 (443)
patient068 (416)
patient069 (723)
patient070 (556)
patient071 (302)
patient072 (580)
patient073 (378)
patient074 (410)
patient075 (661)
patient076 (408)
patient077 (743)
patient078 (582)
patient079 (530)
patient080 (740)
Proband X1 (699)
Proband X10 (581)
Proband X11 (711)
Proband X12 (715)
Proband X13 (649)
Proband X14 (235)
Proband X15 (637)
Proband X2 (727)
Proband X4 (624)
Proband X5 (382)
Proband X6 (570)
Proband X7 (497)
Proband X8 (286)
Proband X9 (671)
sim (185)
wavelet−HHL_gldm_SmallDependenceLowGrayLevelEmphasis
frame
10
5
0

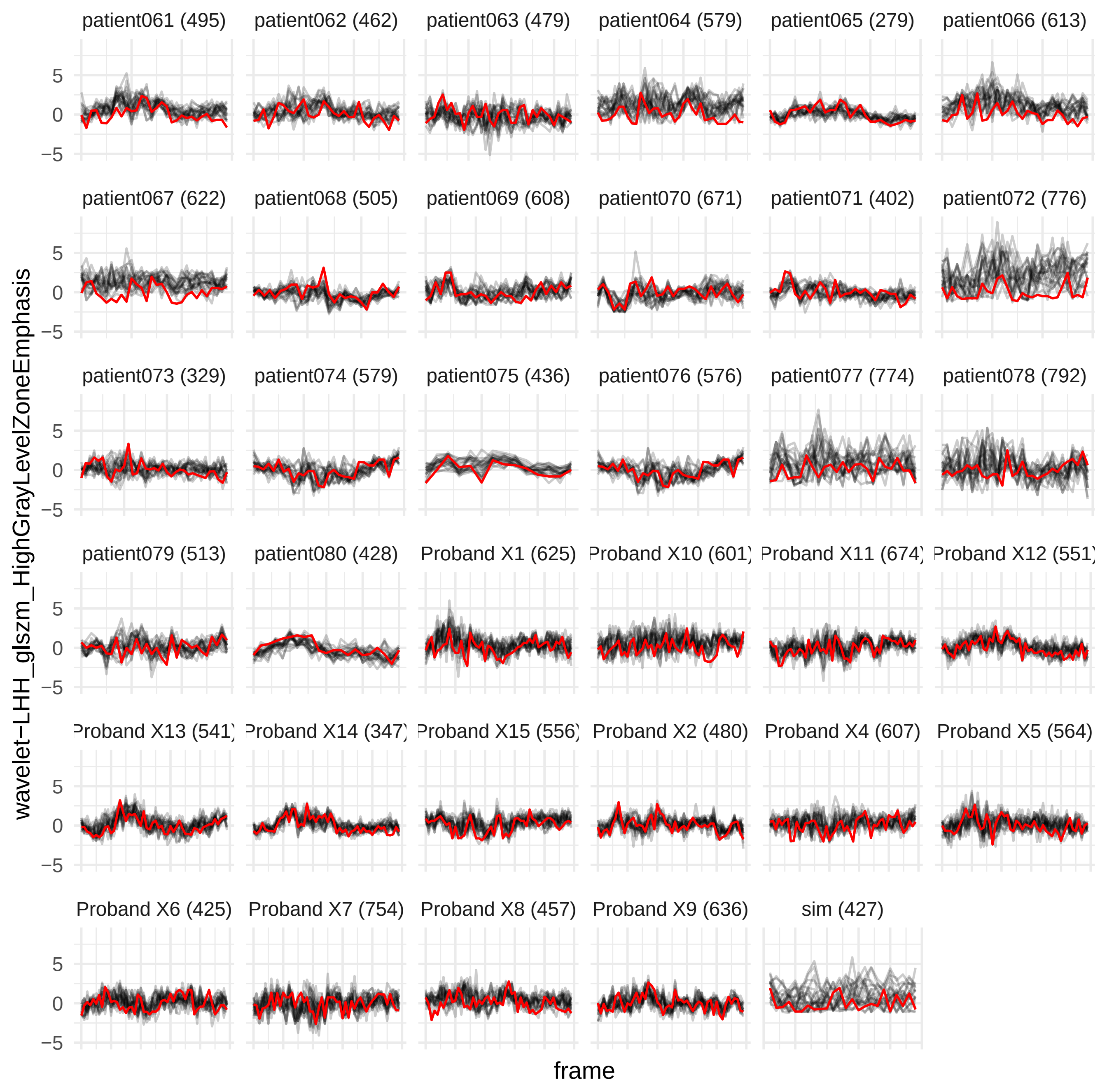

patient061 (495)
patient062 (462)
patient063 (479)
patient064 (579)
patient065 (279)
patient066 (613)
patient067 (622)
patient068 (505)
patient069 (608)
patient070 (671)
patient071 (402)
patient072 (776)
patient073 (329)
patient074 (579)
patient075 (436)
patient076 (576)
patient077 (774)
patient078 (792)
patient079 (513)
patient080 (428)
Proband X1 (625)
Proband X10 (601)
Proband X11 (674)
Proband X12 (551)
Proband X13 (541)
Proband X14 (347)
Proband X15 (556)
Proband X2 (480)
Proband X4 (607)
Proband X5 (564)
Proband X6 (425)
Proband X7 (754)
Proband X8 (457)
Proband X9 (636)
sim (427)
5
0
−5
wavelet−LHH_glszm_HighGrayLevelZoneEmphasis
frame

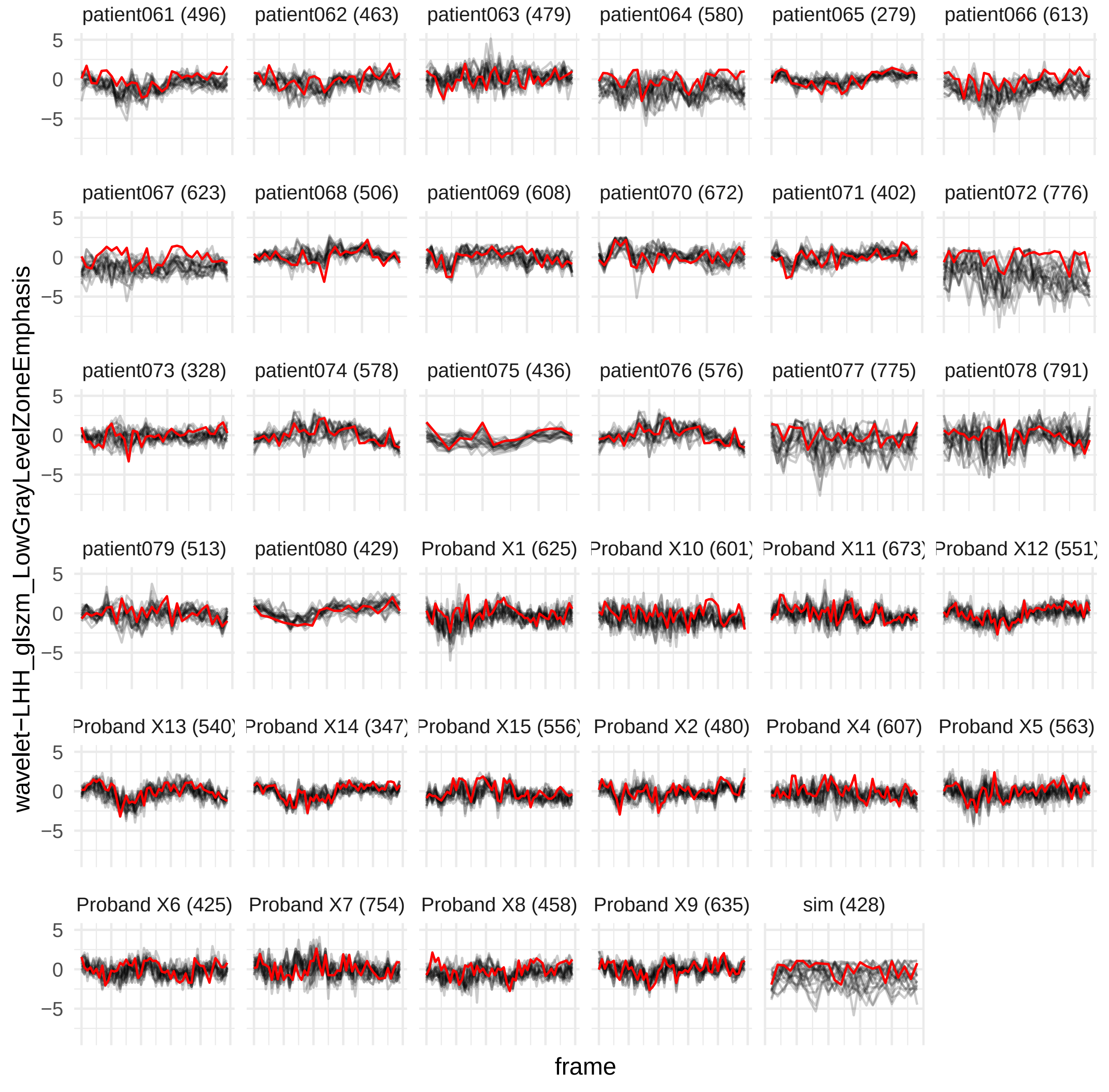
patient061 (496)
patient062 (463)
patient063 (479)
patient064 (580)
patient065 (279)
patient066 (613)
patient067 (623)
patient068 (506)
patient069 (608)
patient070 (672)
patient071 (402)
patient072 (776)
patient073 (328)
patient074 (578)
patient075 (436)
patient076 (576)
patient077 (775)
patient078 (791)
patient079 (513)
patient080 (429)
Proband X1 (625)
Proband X10 (601)
Proband X11 (673)
Proband X12 (551)
Proband X13 (540)
Proband X14 (347)
Proband X15 (556)
Proband X2 (480)
Proband X4 (607)
Proband X5 (563)
Proband X6 (425)
Proband X7 (754)
Proband X8 (458)
Proband X9 (635)
sim (428)
5
0
−5
wavelet−LHH_glszm_LowGrayLevelZoneEmphasis
frame

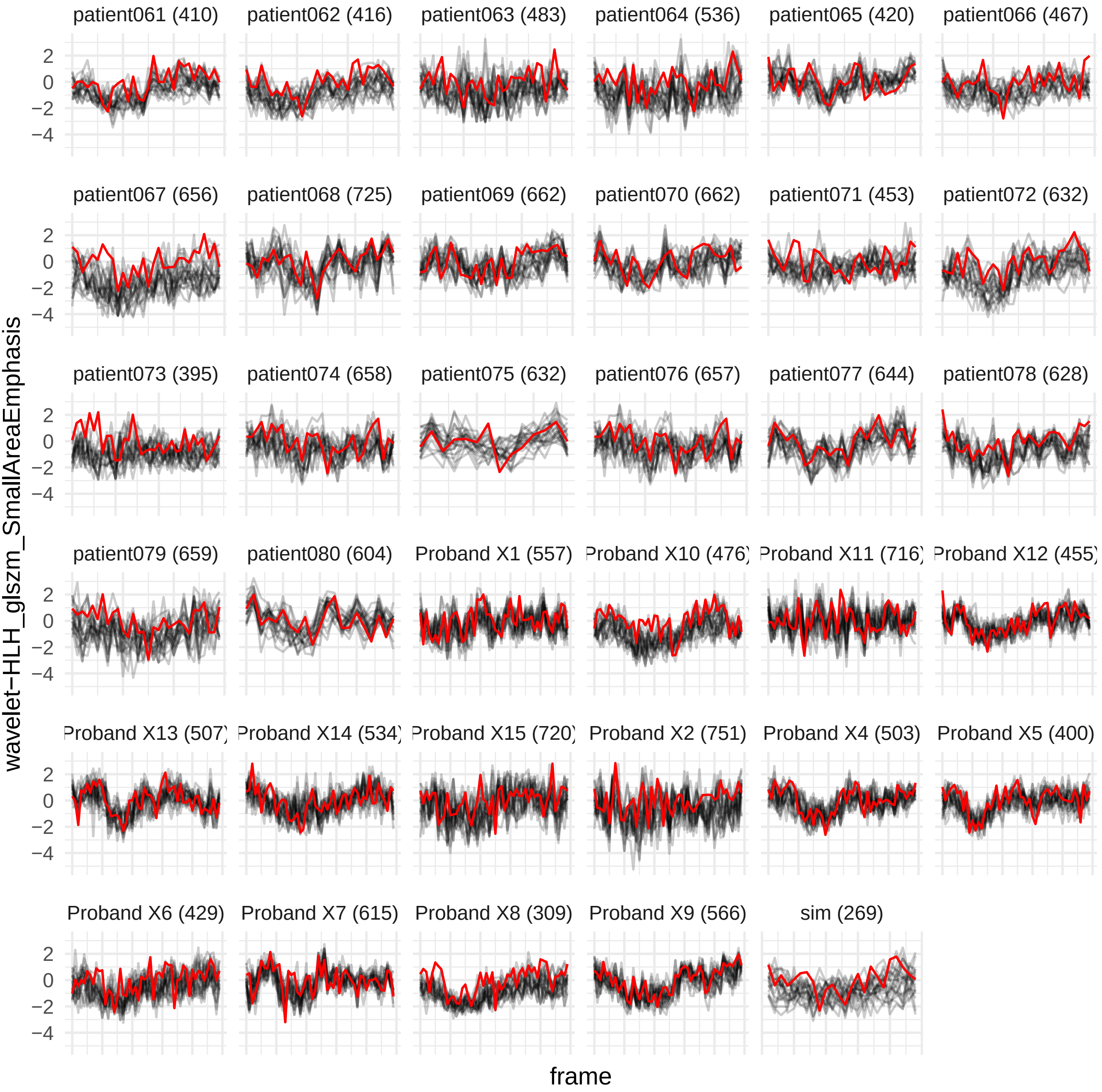

patient061 (410)
patient062 (416)
patient063 (483)
patient064 (536)
patient065 (420)
patient066 (467)
patient067 (656)
patient068 (725)
patient069 (662)
patient070 (662)
patient071 (453)
patient072 (632)
patient073 (395)
patient074 (658)
patient075 (632)
patient076 (657)
patient077 (644)
patient078 (628)
patient079 (659)
patient080 (604)
Proband X1 (557)
Proband X10 (476)
Proband X11 (716)
Proband X12 (455)
Proband X13 (507)
Proband X14 (534)
Proband X15 (720)
Proband X2 (751)
Proband X4 (503)
Proband X5 (400)
Proband X6 (429)
Proband X7 (615)
Proband X8 (309)
Proband X9 (566)
sim (269)
2
0
−2
−4
wavelet−HLH_glszm_SmallAreaEmphasis
frame

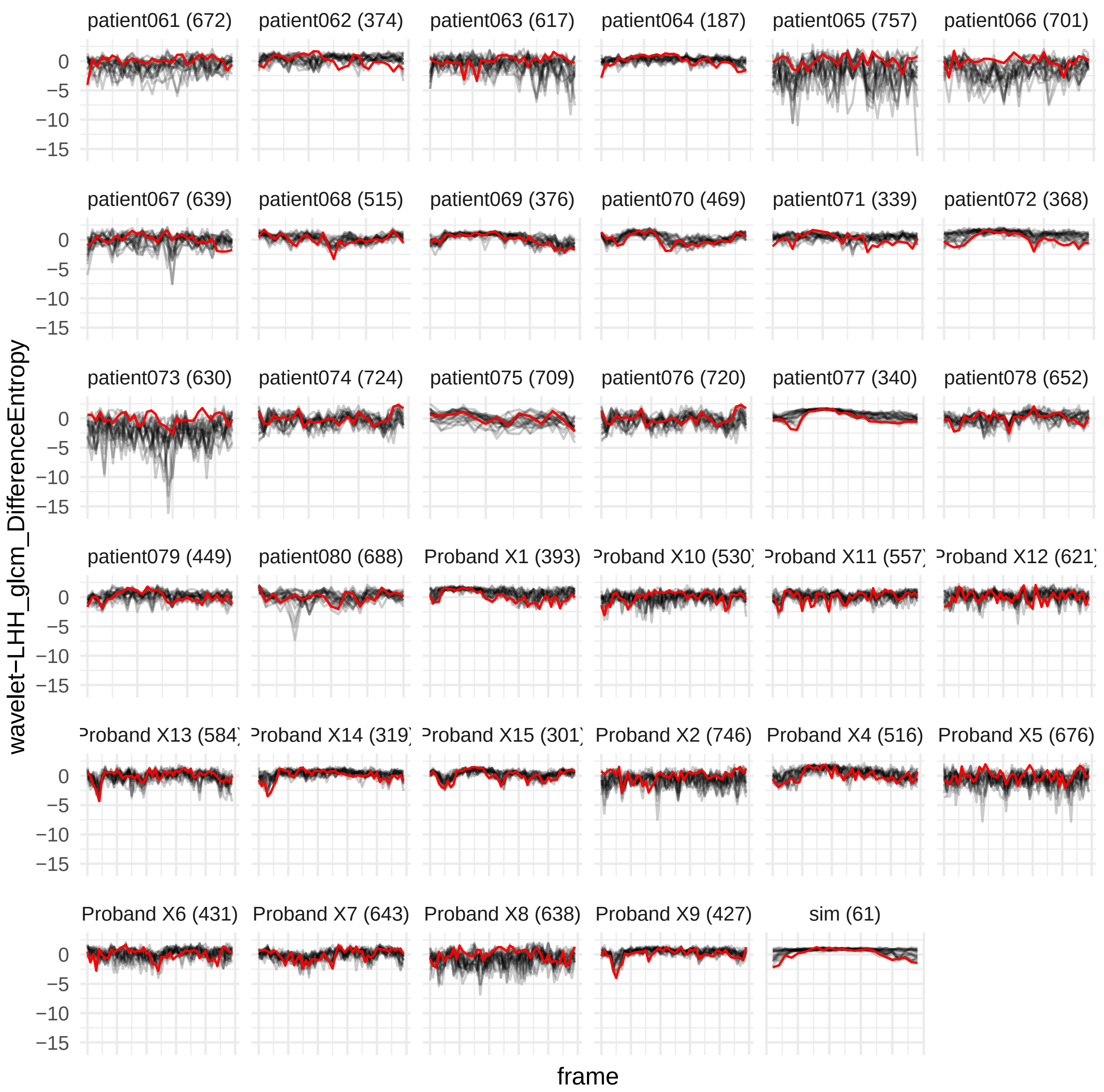

patient061 (672)
patient062 (374)
patient063 (617)
patient064 (187)
patient065 (757)
patient066 (701)
patient067 (639)
patient068 (515)
patient069 (376)
patient070 (469)
patient071 (339)
patient072 (368)
patient073 (630)
patient074 (724)
patient075 (709)
patient076 (720)
patient077 (340)
patient078 (652)
patient079 (449)
patient080 (688)
Proband X1 (393)
Proband X10 (530)
Proband X11 (557)
Proband X12 (621)
Proband X13 (584)
Proband X14 (319)
Proband X15 (301)
Proband X2 (746)
Proband X4 (516)
Proband X5 (676)
Proband X6 (431)
Proband X7 (643)
Proband X8 (638)
Proband X9 (427)
sim (61)
0
−5
−10
−15
wavelet−LHH_glcm_DifferenceEntropy
frame

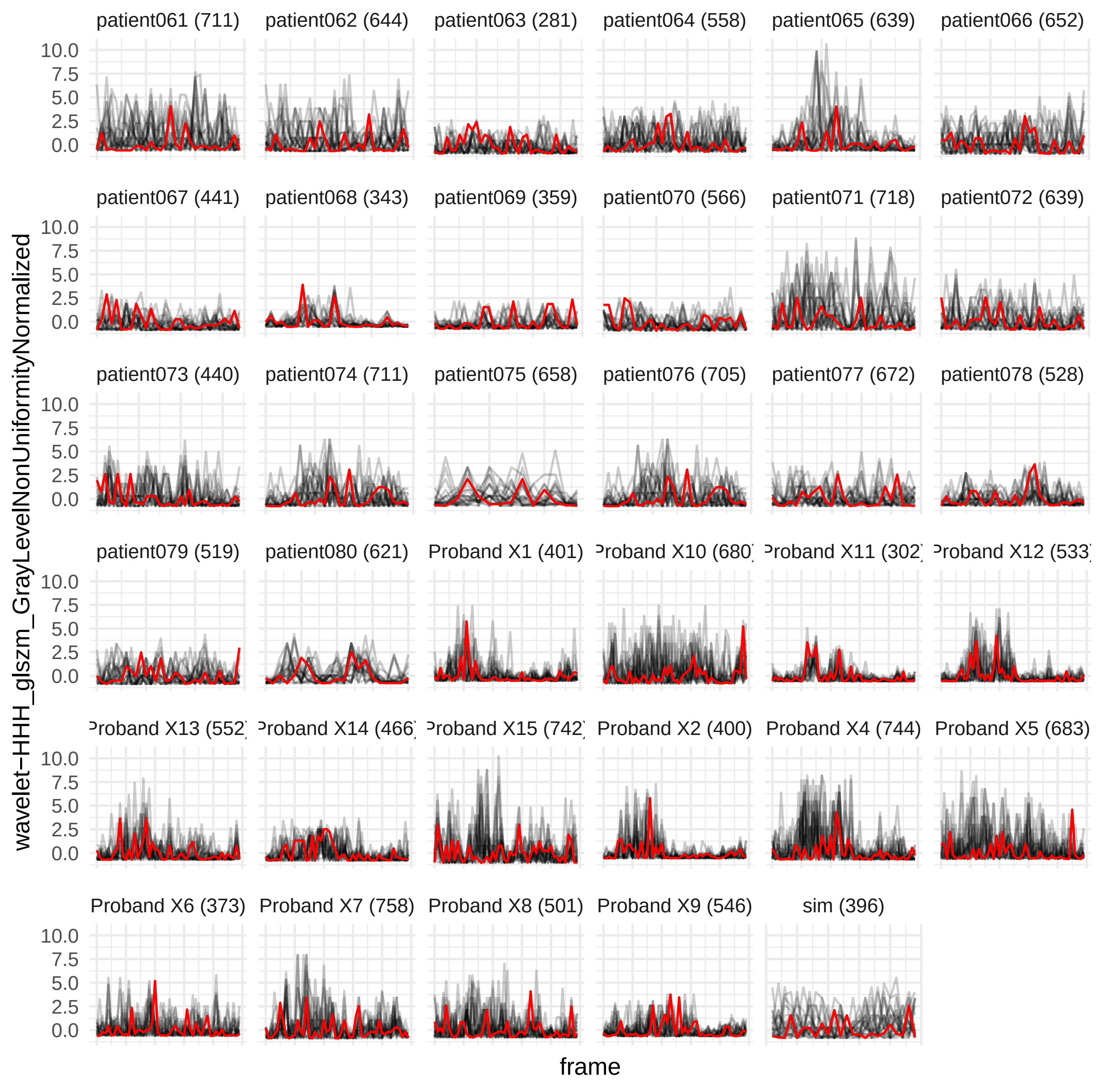

patient061 (711)
patient062 (644)
patient063 (281)
patient064 (558)
patient065 (639)
patient066 (652)
patient067 (441)
patient068 (343)
patient069 (359)
patient070 (566)
patient071 (718)
patient072 (639)
patient073 (440)
patient074 (711)
patient075 (658)
patient076 (705)
patient077 (672)
patient078 (528)
patient079 (519)
patient080 (621)
Proband X1 (401)
Proband X10 (680)
Proband X11 (302)
Proband X12 (533)
Proband X13 (552)
Proband X14 (466)
Proband X15 (742)
Proband X2 (400)
Proband X4 (744)
Proband X5 (683)
Proband X6 (373)
Proband X7 (758)
Proband X8 (501)
Proband X9 (546)
sim (396)
10.0
7.5
5.0
2.5
0.0
wavelet−HHH_glszm_GrayLevelNonUniformityNormalized
frame

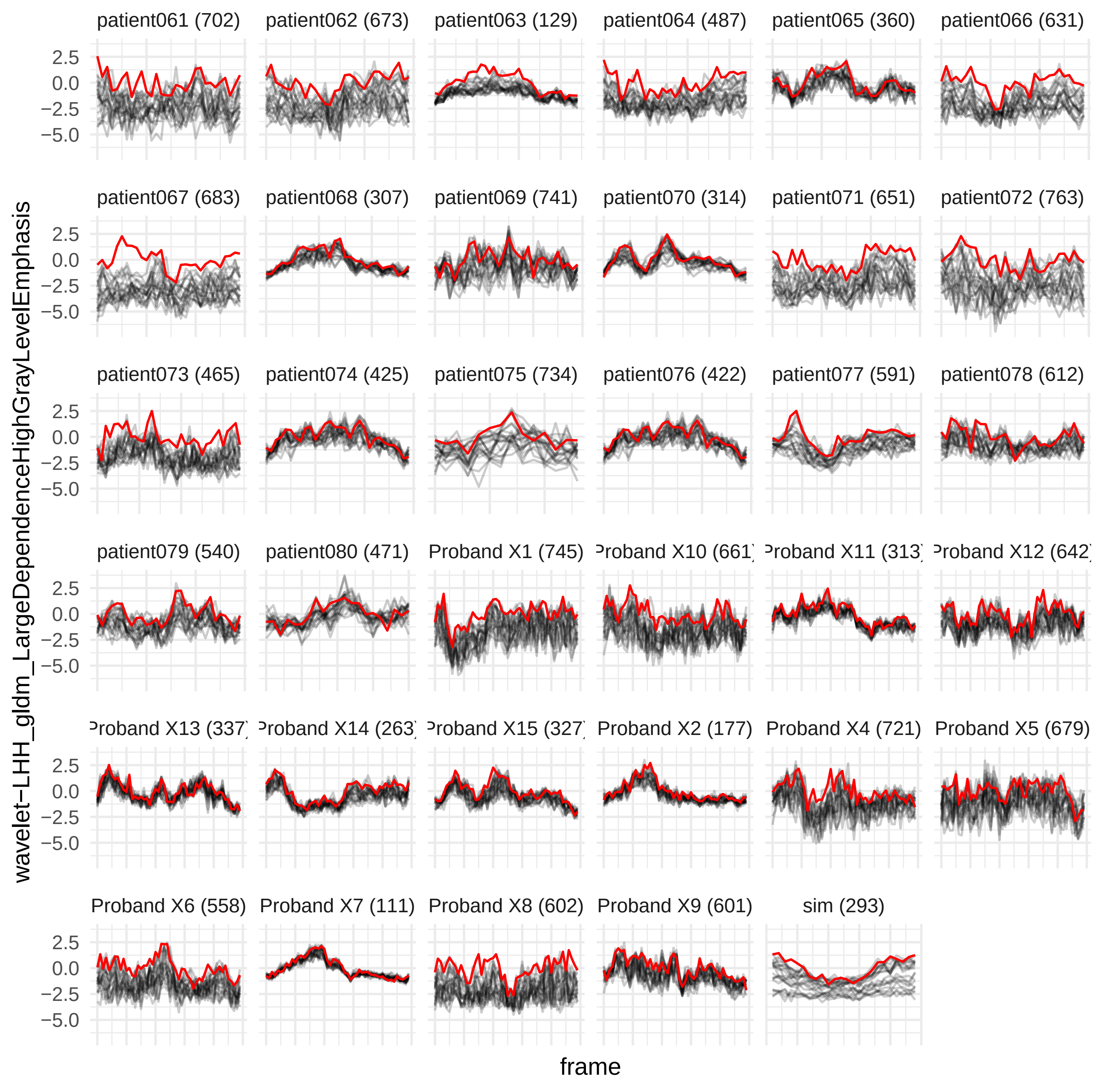

patient061 (702)
patient062 (673)
patient063 (129)
patient064 (487)
patient065 (360)
patient066 (631)
patient067 (683)
patient068 (307)
patient069 (741)
patient070 (314)
patient071 (651)
patient072 (763)
patient073 (465)
patient074 (425)
patient075 (734)
patient076 (422)
patient077 (591)
patient078 (612)
patient079 (540)
patient080 (471)
Proband X1 (745)
Proband X10 (661)
Proband X11 (313)
Proband X12 (642)
Proband X13 (337)
Proband X14 (263)
Proband X15 (327)
Proband X2 (177)
Proband X4 (721)
Proband X5 (679)
Proband X6 (558)
Proband X7 (111)
Proband X8 (602)
Proband X9 (601)
sim (293)
2.5
0.0
−2.5
−5.0
wavelet−LHH_gldm_LargeDependenceHighGrayLevelEmphasis
frame

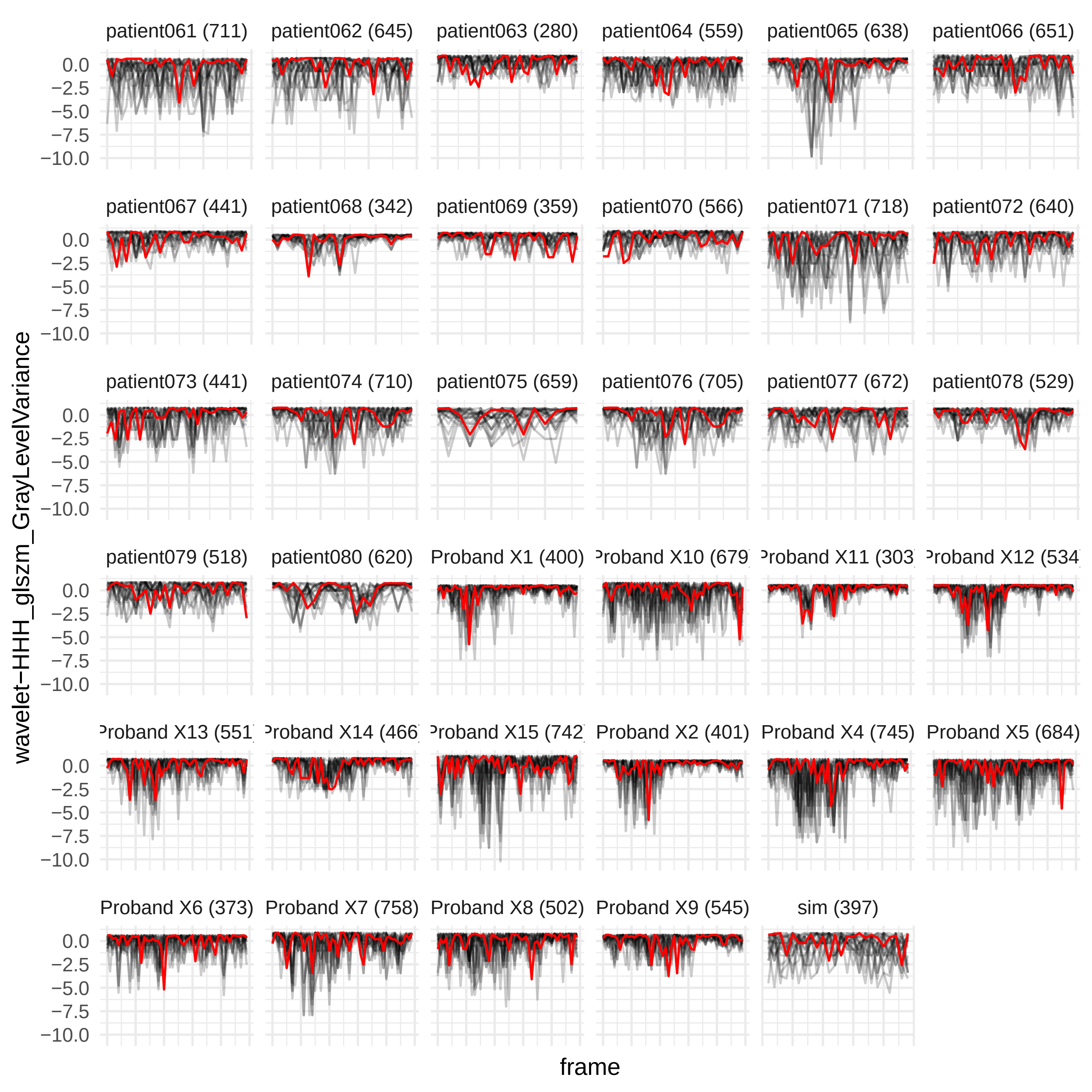

patient061 (711)
patient062 (645)
patient063 (280)
patient064 (559)
patient065 (638)
patient066 (651)
patient067 (441)
patient068 (342)
patient069 (359)
patient070 (566)
patient071 (718)
patient072 (640)
patient073 (441)
patient074 (710)
patient075 (659)
patient076 (705)
patient077 (672)
patient078 (529)
patient079 (518)
patient080 (620)
Proband X1 (400)
Proband X10 (679
Proband X11 (303
Proband X12 (534
Proband X13 (551
Proband X14 (466
Proband X15 (742
Proband X2 (401)
Proband X4 (745)
Proband X5 (684)
Proband X6 (373)
Proband X7 (758)
Proband X8 (502)
Proband X9 (545)
sim (397)
0.0
−2.5
−5.0
−7.5
−10.0
wavelet−HHH_glszm_GrayLevelVariance
frame

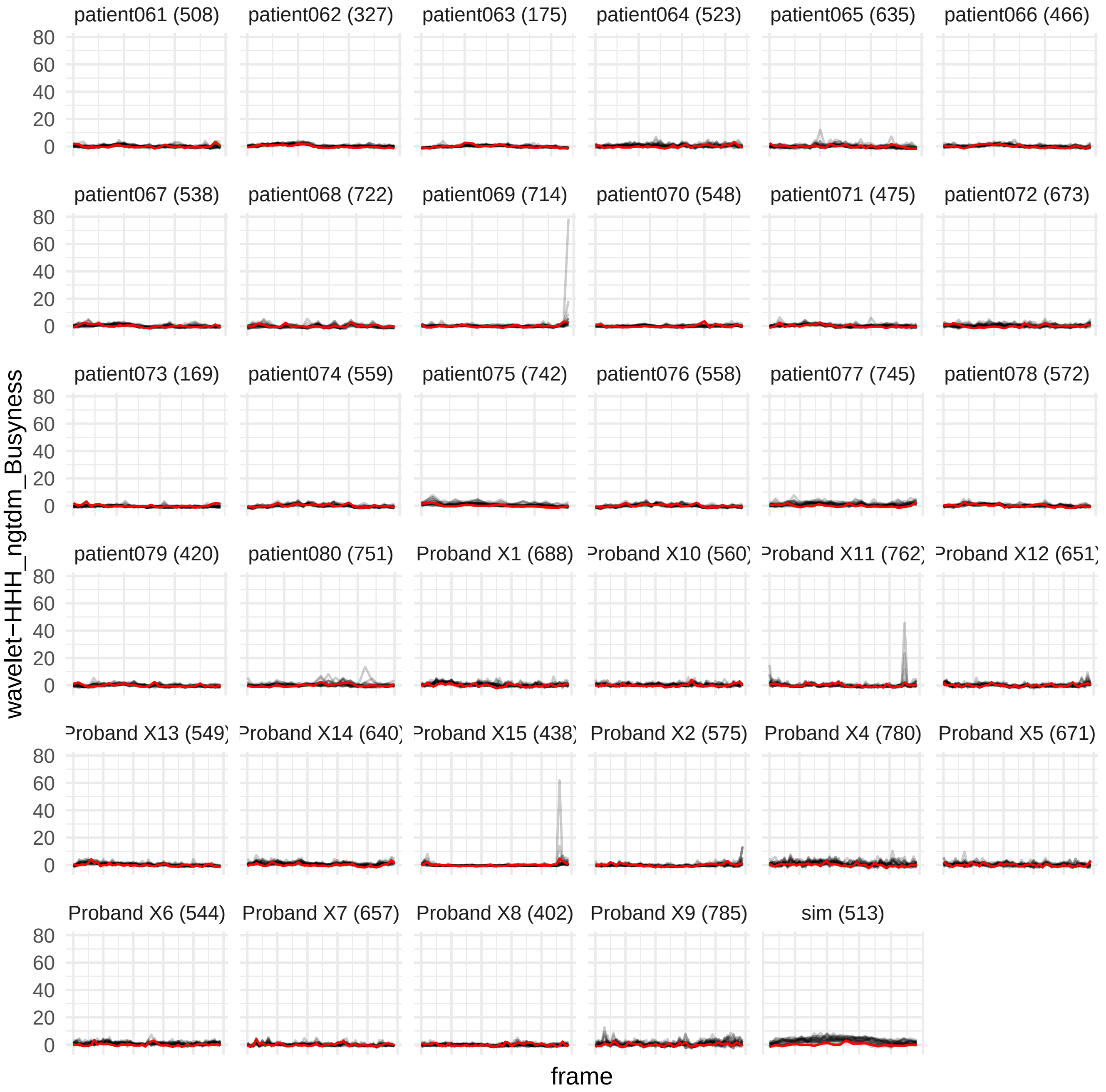

patient061 (508)
patient062 (327)
patient063 (175)
patient064 (523)
patient065 (635)
patient066 (466)
patient067 (538)
patient068 (722)
patient069 (714)
patient070 (548)
patient071 (475)
patient072 (673)
patient073 (169)
patient074 (559)
patient075 (742)
patient076 (558)
patient077 (745)
patient078 (572)
patient079 (420)
patient080 (751)
Proband X1 (688)
Proband X10 (560)
Proband X11 (762)
Proband X12 (651)
Proband X13 (549)
Proband X14 (640)
Proband X15 (438)
Proband X2 (575)
Proband X4 (780)
Proband X5 (671)
Proband X6 (544)
Proband X7 (657)
Proband X8 (402)
Proband X9 (785)
sim (513)
wavelet−HHH_ngtdm_Busyness
frame
0
20
40
60
80

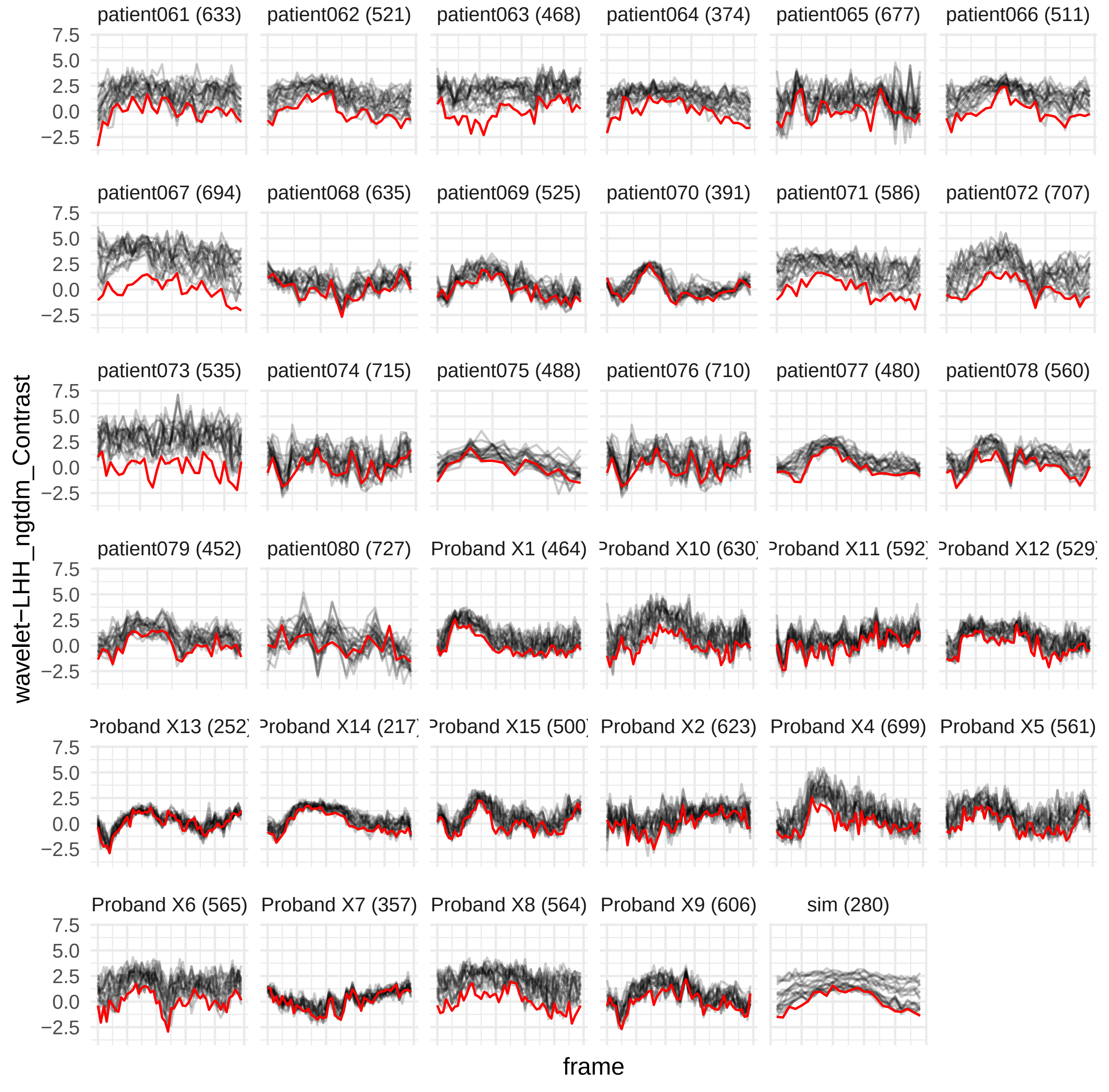

patient061 (633)
patient062 (521)
patient063 (468)
patient064 (374)
patient065 (677)
patient066 (511)
patient067 (694)
patient068 (635)
patient069 (525)
patient070 (391)
patient071 (586)
patient072 (707)
patient073 (535)
patient074 (715)
patient075 (488)
patient076 (710)
patient077 (480)
patient078 (560)
patient079 (452)
patient080 (727)
Proband X1 (464)
Proband X10 (630)
Proband X11 (592)
Proband X12 (529)
Proband X13 (252)
Proband X14 (217)
Proband X15 (500)
Proband X2 (623)
Proband X4 (699)
Proband X5 (561)
Proband X6 (565)
Proband X7 (357)
Proband X8 (564)
Proband X9 (606)
sim (280)
7.5
5.0
2.5
0.0
−2.5
wavelet−LHH_ngtdm_Contrast
frame

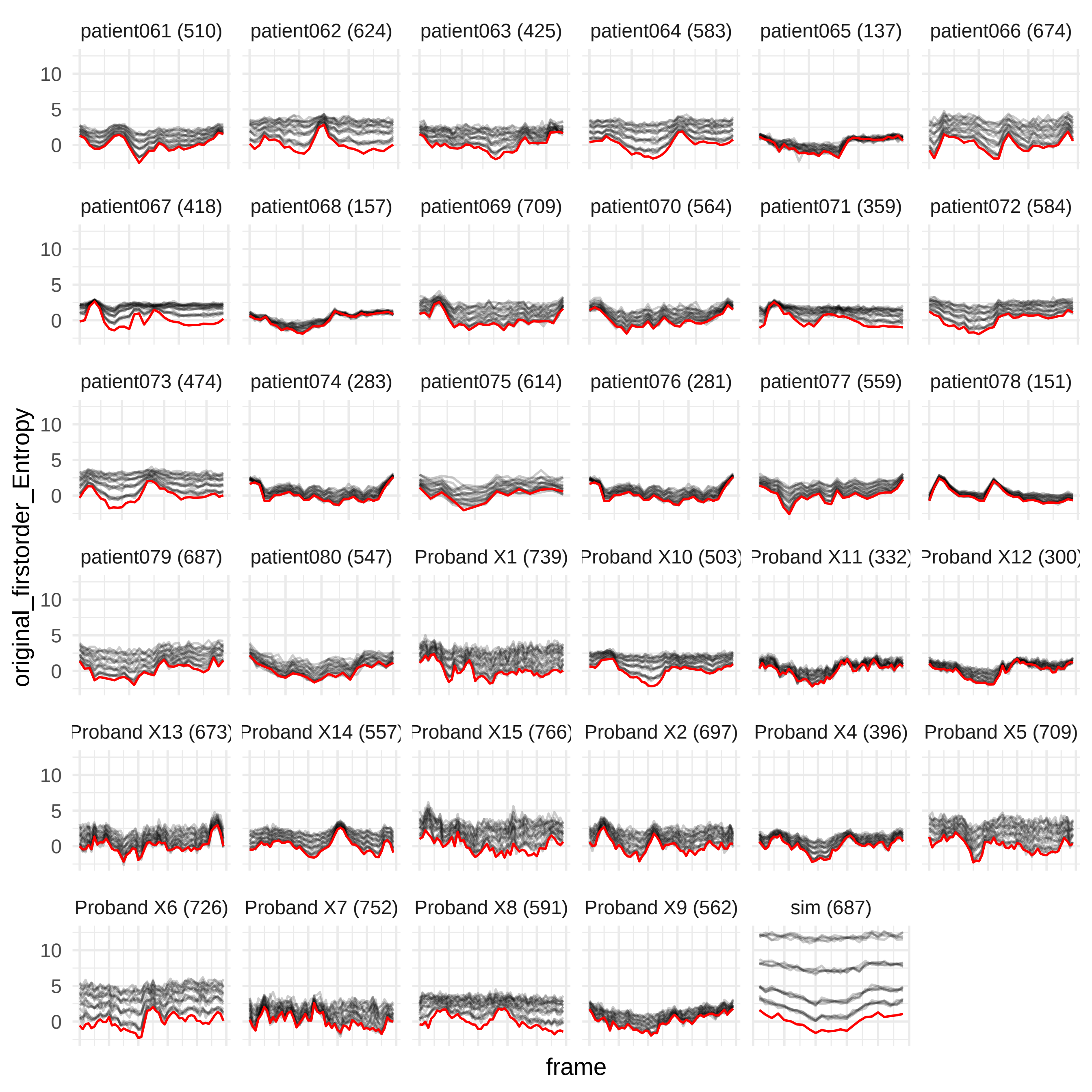
patient061 (510)
patient062 (624)
patient063 (425)
patient064 (583)
patient065 (137)
patient066 (674)
patient067 (418)
patient068 (157)
patient069 (709)
patient070 (564)
patient071 (359)
patient072 (584)
patient073 (474)
patient074 (283)
patient075 (614)
patient076 (281)
patient077 (559)
patient078 (151)
patient079 (687)
patient080 (547)
Proband X1 (739)
Proband X10 (503)
Proband X11 (332)
Proband X12 (300)
Proband X13 (673)
Proband X14 (557)
Proband X15 (766)
Proband X2 (697)
Proband X4 (396)
Proband X5 (709)
Proband X6 (726)
Proband X7 (752)
Proband X8 (591)
Proband X9 (562)
sim (687)
original_firstorder_Entropy
10
5
0
frame

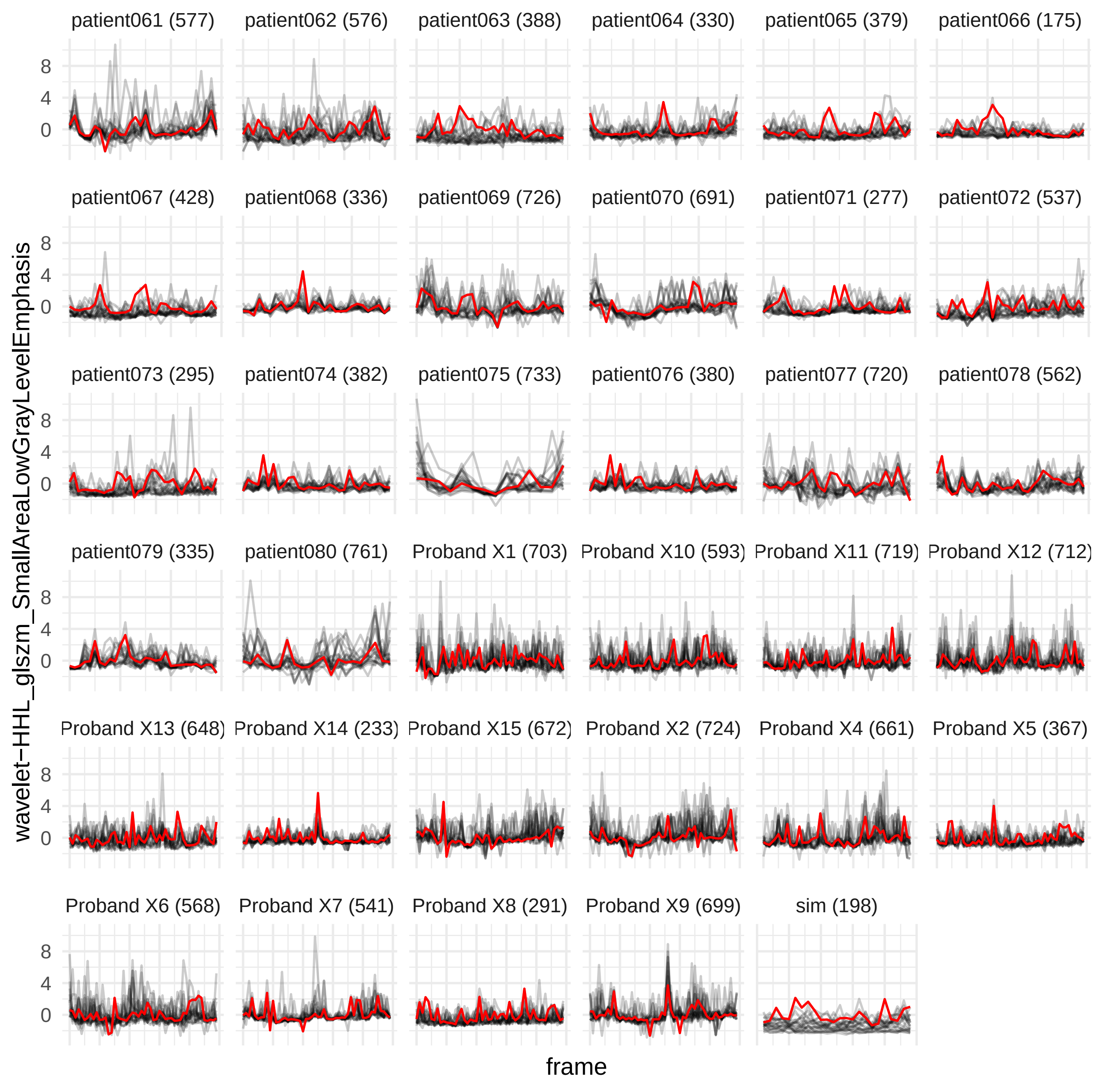

patient061 (577)
patient062 (576)
patient063 (388)
patient064 (330)
patient065 (379)
patient066 (175)
patient067 (428)
patient068 (336)
patient069 (726)
patient070 (691)
patient071 (277)
patient072 (537)
patient073 (295)
patient074 (382)
patient075 (733)
patient076 (380)
patient077 (720)
patient078 (562)
patient079 (335)
patient080 (761)
Proband X1 (703)
Proband X10 (593)
Proband X11 (719)
Proband X12 (712)
Proband X13 (648)
Proband X14 (233)
Proband X15 (672)
Proband X2 (724)
Proband X4 (661)
Proband X5 (367)
Proband X6 (568)
Proband X7 (541)
Proband X8 (291)
Proband X9 (699)
sim (198)
wavelet−HHL_glszm_SmallAreaLowGrayLevelEmphasis
frame
8
4
0

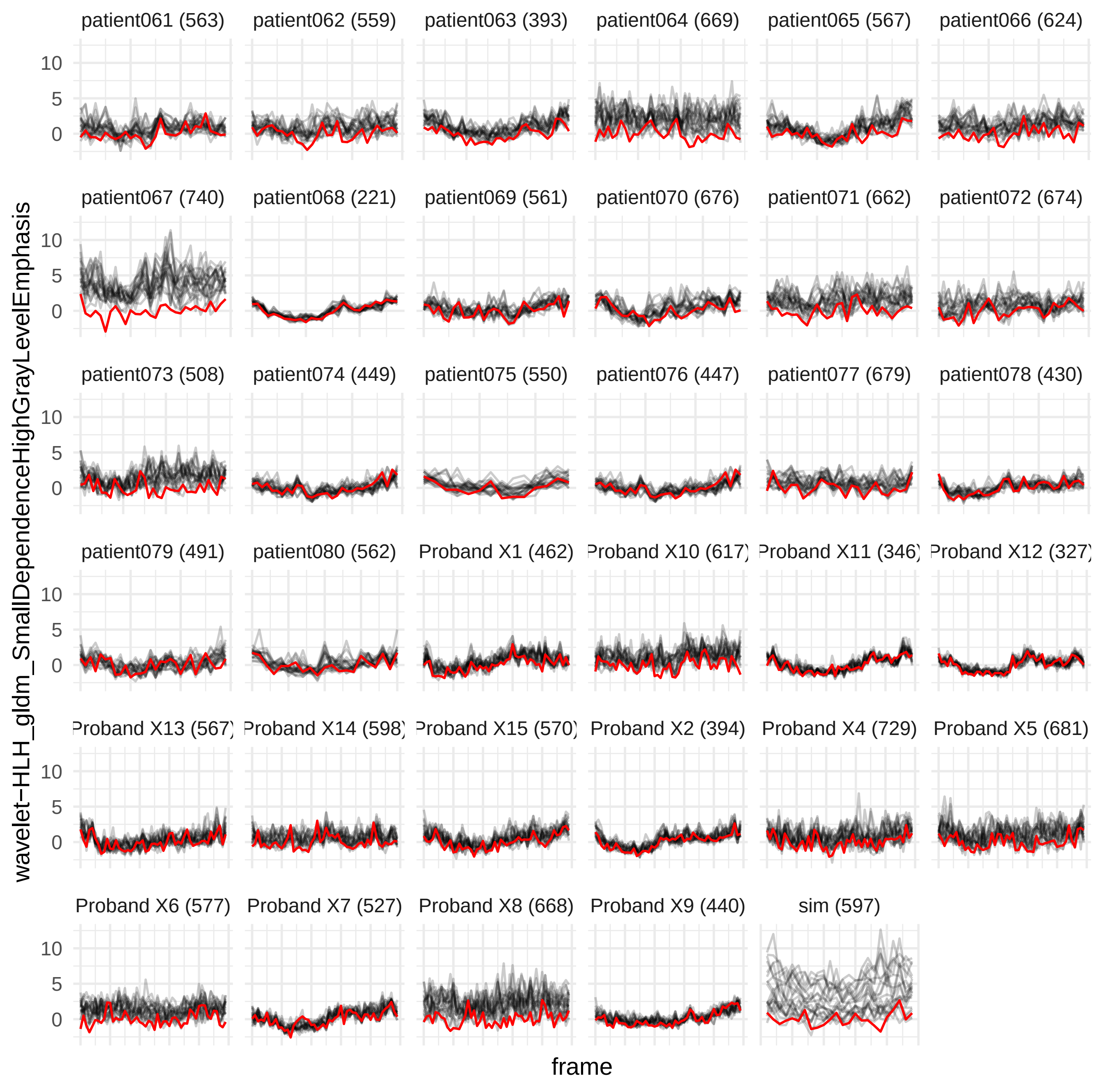
patient061 (563)
patient062 (559)
patient063 (393)
patient064 (669)
patient065 (567)
patient066 (624)
patient067 (740)
patient068 (221)
patient069 (561)
patient070 (676)
patient071 (662)
patient072 (674)
patient073 (508)
patient074 (449)
patient075 (550)
patient076 (447)
patient077 (679)
patient078 (430)
patient079 (491)
patient080 (562)
Proband X1 (462)
Proband X10 (617)
Proband X11 (346)
Proband X12 (327)
Proband X13 (567)
Proband X14 (598)
Proband X15 (570)
Proband X2 (394)
Proband X4 (729)
Proband X5 (681)
Proband X6 (577)
Proband X7 (527)
Proband X8 (668)
Proband X9 (440)
sim (597)
10
5
0
wavelet−HLH_gldm_SmallDependenceHighGrayLevelEmphasis
frame

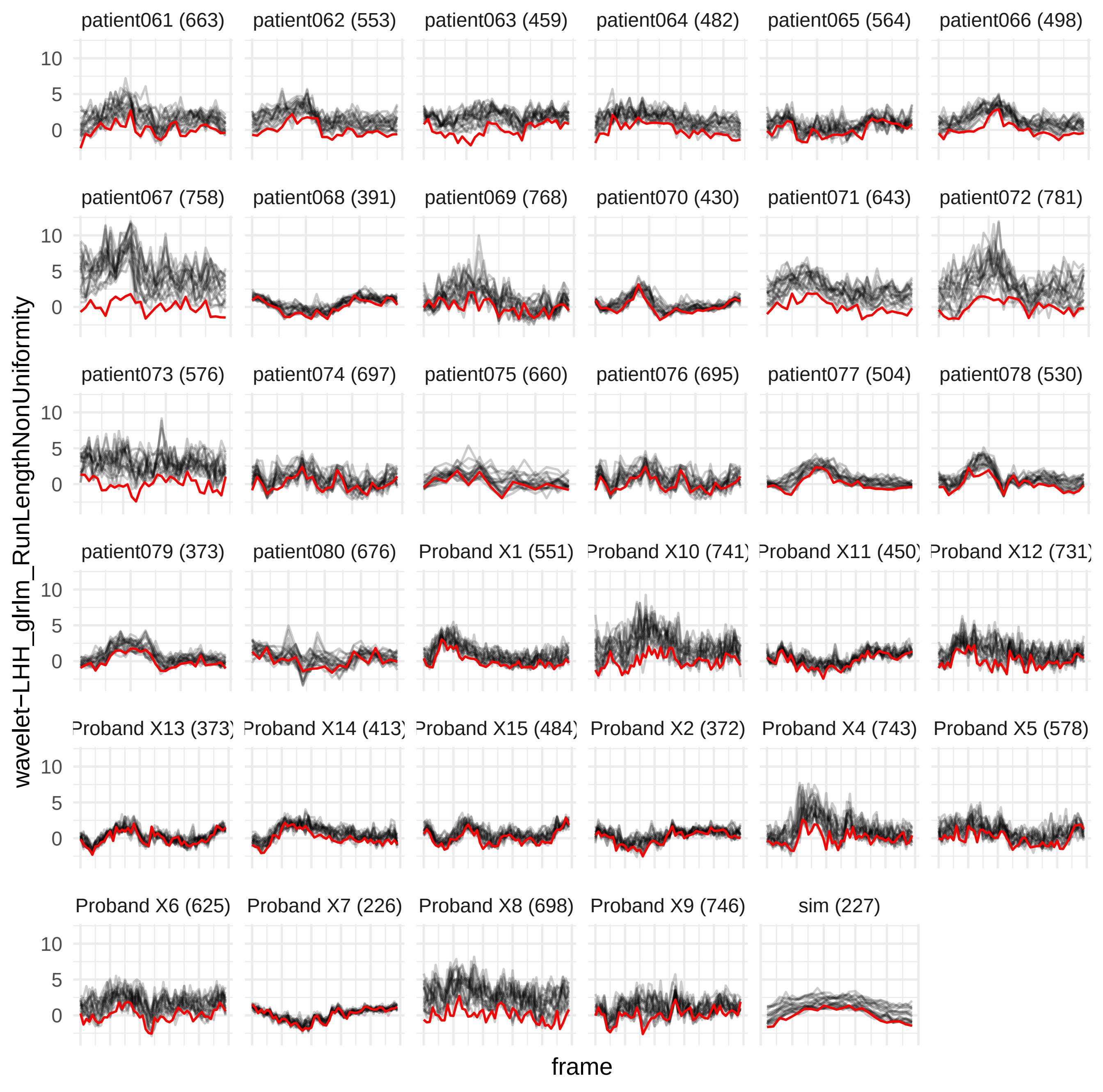
patient061 (663)
patient062 (553)
patient063 (459)
patient064 (482)
patient065 (564)
patient066 (498)
patient067 (758)
patient068 (391)
patient069 (768)
patient070 (430)
patient071 (643)
patient072 (781)
patient073 (576)
patient074 (697)
patient075 (660)
patient076 (695)
patient077 (504)
patient078 (530)
patient079 (373)
patient080 (676)
Proband X1 (551)
Proband X10 (741)
Proband X11 (450)
Proband X12 (731)
Proband X13 (373)
Proband X14 (413)
Proband X15 (484)
Proband X2 (372)
Proband X4 (743)
Proband X5 (578)
Proband X6 (625)
Proband X7 (226)
Proband X8 (698)
Proband X9 (746)
sim (227)
10
5
0
wavelet−LHH_glrlm_RunLengthNonUniformity
frame

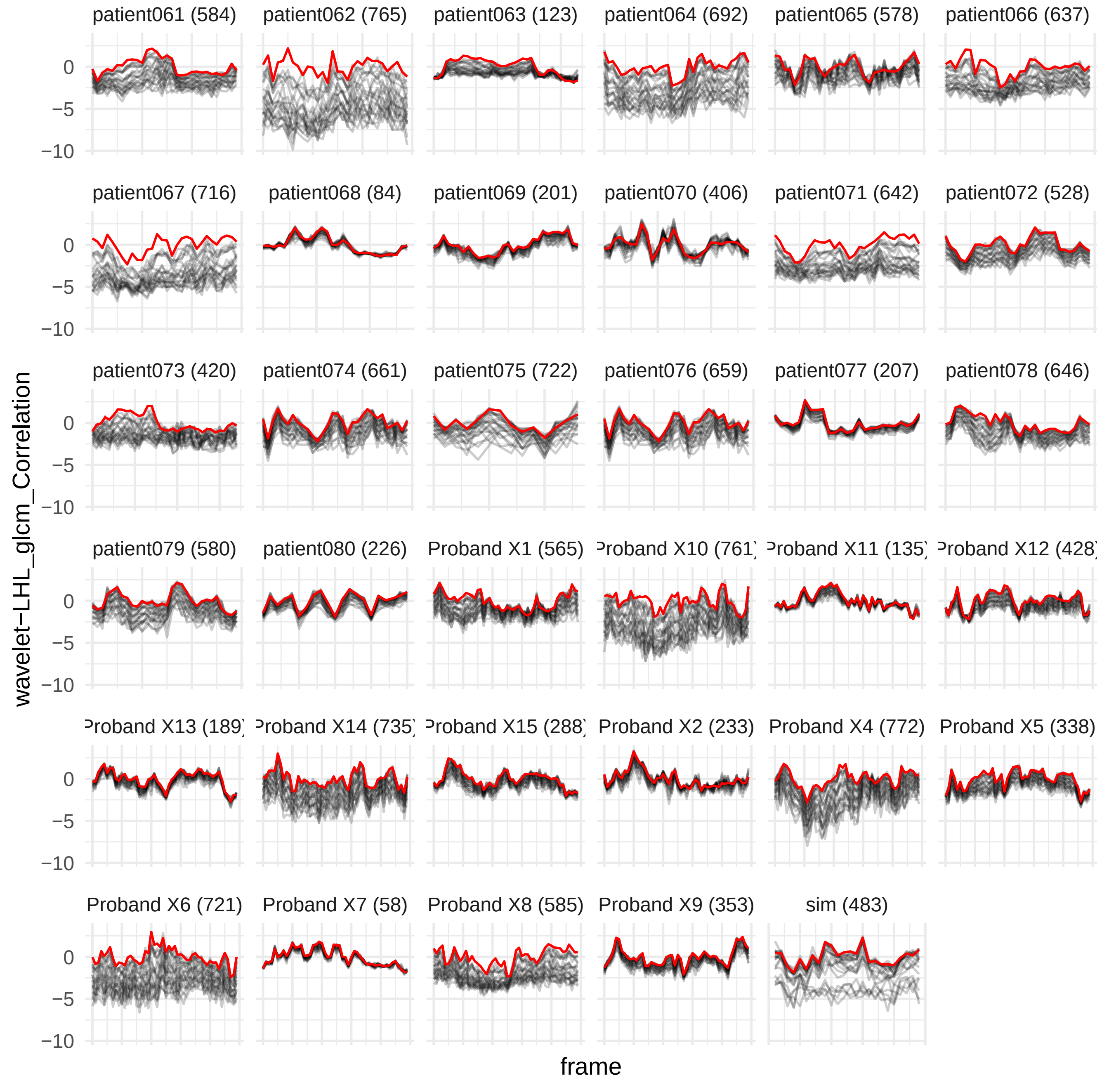

patient061 (584)
patient062 (765)
patient063 (123)
patient064 (692)
patient065 (578)
patient066 (637)
patient067 (716)
patient068 (84)
patient069 (201)
patient070 (406)
patient071 (642)
patient072 (528)
patient073 (420)
patient074 (661)
patient075 (722)
patient076 (659)
patient077 (207)
patient078 (646)
patient079 (580)
patient080 (226)
Proband X1 (565)
Proband X10 (761)
Proband X11 (135)
Proband X12 (428)
Proband X13 (189)
Proband X14 (735)
Proband X15 (288)
Proband X2 (233)
Proband X4 (772)
Proband X5 (338)
Proband X6 (721)
Proband X7 (58)
Proband X8 (585)
Proband X9 (353)
sim (483)
0
−5
−10
wavelet−LHL_glcm_Correlation
frame

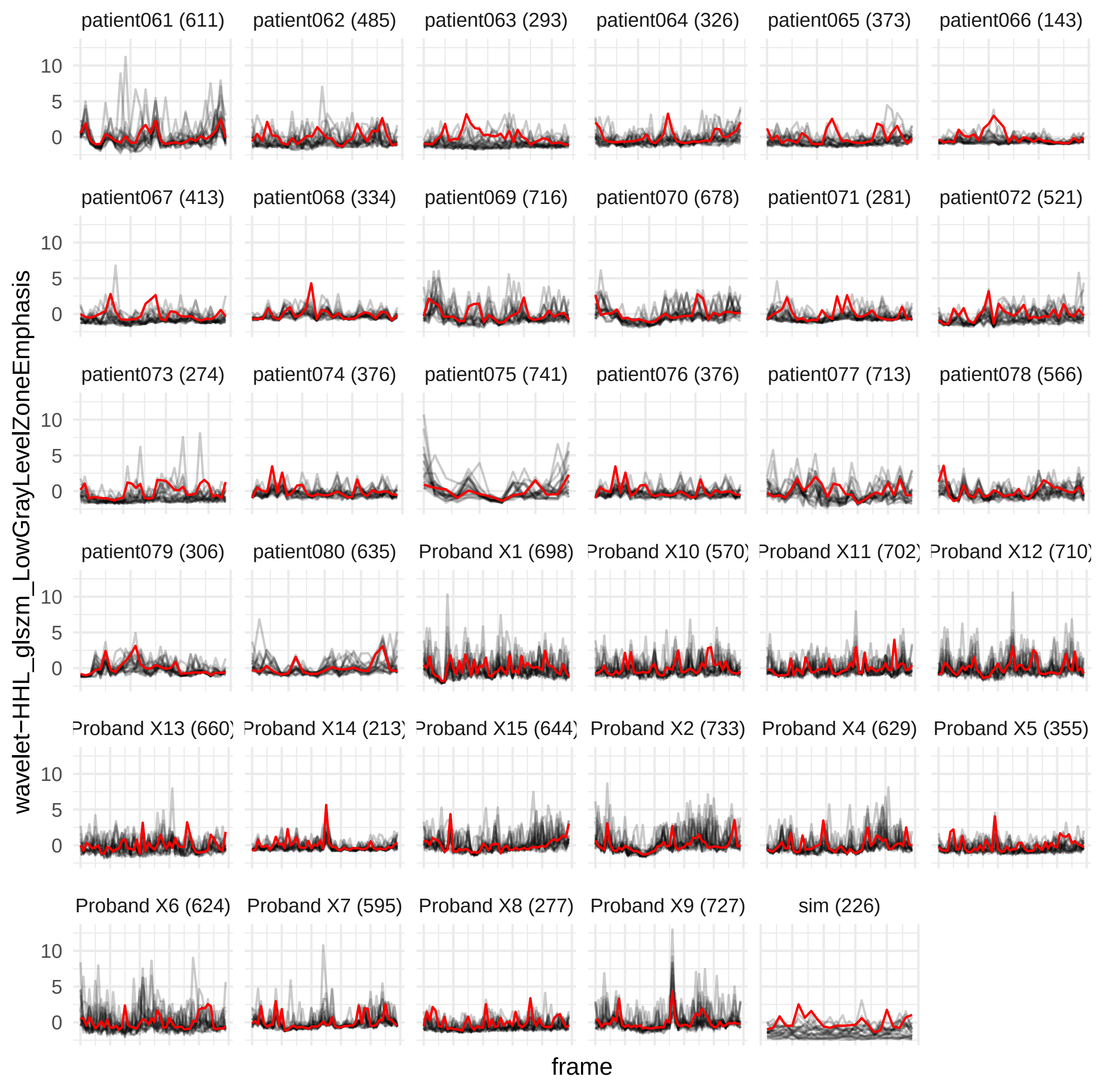

patient061 (611)
patient062 (485)
patient063 (293)
patient064 (326)
patient065 (373)
patient066 (143)
patient067 (413)
patient068 (334)
patient069 (716)
patient070 (678)
patient071 (281)
patient072 (521)
patient073 (274)
patient074 (376)
patient075 (741)
patient076 (376)
patient077 (713)
patient078 (566)
patient079 (306)
patient080 (635)
Proband X1 (698)
Proband X10 (570)
Proband X11 (702)
Proband X12 (710)
Proband X13 (660)
Proband X14 (213)
Proband X15 (644)
Proband X2 (733)
Proband X4 (629)
Proband X5 (355)
Proband X6 (624)
Proband X7 (595)
Proband X8 (277)
Proband X9 (727)
sim (226)
wavelet−HHL_glszm_LowGrayLevelZoneEmphasis
10
5
0
frame

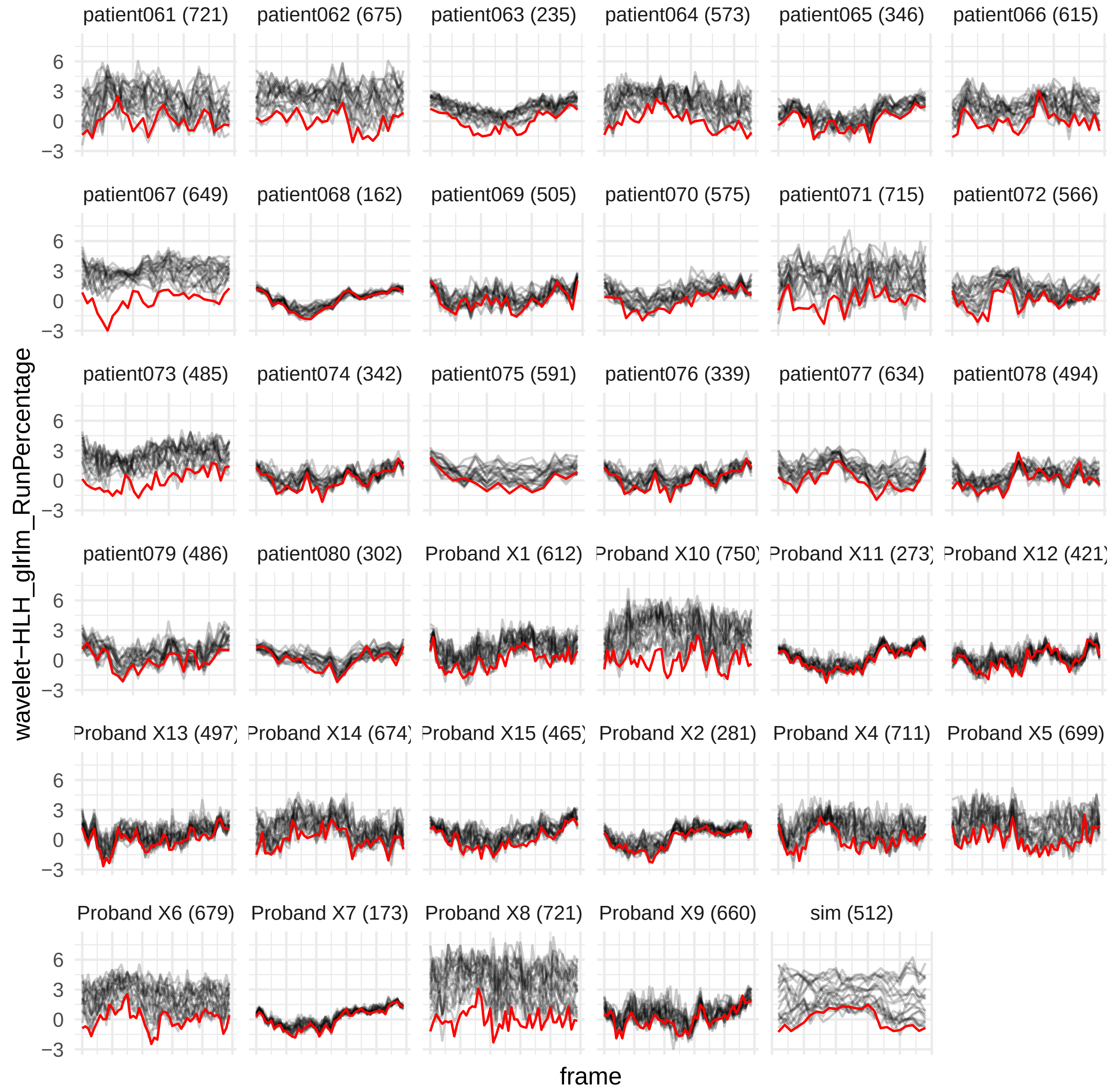

patient061 (721)
patient062 (675)
patient063 (235)
patient064 (573)
patient065 (346)
patient066 (615)
patient067 (649)
patient068 (162)
patient069 (505)
patient070 (575)
patient071 (715)
patient072 (566)
patient073 (485)
patient074 (342)
patient075 (591)
patient076 (339)
patient077 (634)
patient078 (494)
patient079 (486)
patient080 (302)
Proband X1 (612)
Proband X10 (750)
Proband X11 (273)
Proband X12 (421)
Proband X13 (497)
Proband X14 (674)
Proband X15 (465)
Proband X2 (281)
Proband X4 (711)
Proband X5 (699)
Proband X6 (679)
Proband X7 (173)
Proband X8 (721)
Proband X9 (660)
sim (512)
wavelet−HLH_glrlm_RunPercentage
frame
6
3
0
−3

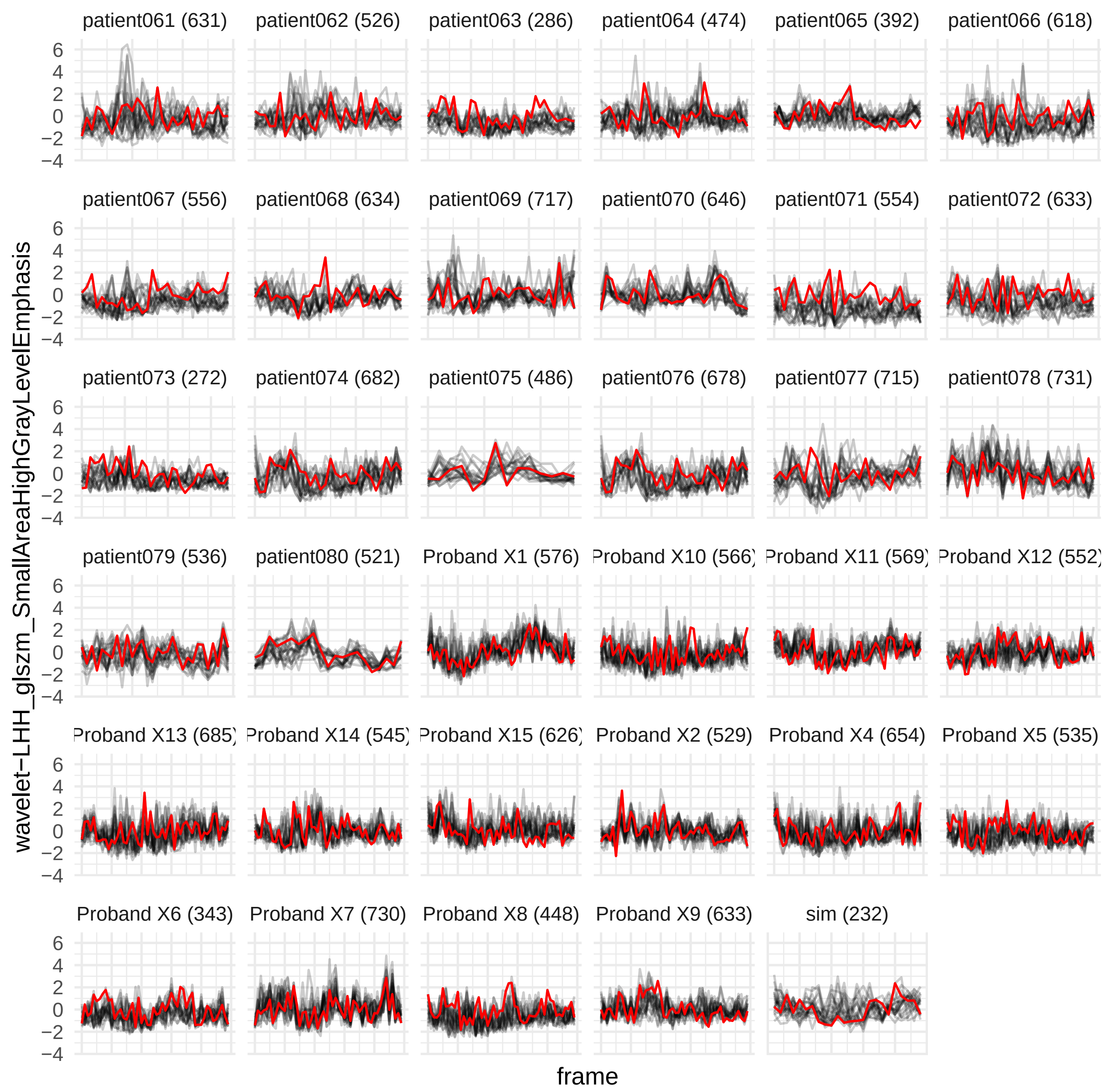

patient061 (631)
patient062 (526)
patient063 (286)
patient064 (474)
patient065 (392)
patient066 (618)
patient067 (556)
patient068 (634)
patient069 (717)
patient070 (646)
patient071 (554)
patient072 (633)
patient073 (272)
patient074 (682)
patient075 (486)
patient076 (678)
patient077 (715)
patient078 (731)
patient079 (536)
patient080 (521)
Proband X1 (576)
Proband X10 (566)
Proband X11 (569)
Proband X12 (552)
Proband X13 (685)
Proband X14 (545)
Proband X15 (626)
Proband X2 (529)
Proband X4 (654)
Proband X5 (535)
Proband X6 (343)
Proband X7 (730)
Proband X8 (448)
Proband X9 (633)
sim (232)
wavelet−LHH_glszm_SmallAreaHighGrayLevelEmphasis
6
4
2
0
−2
−4
frame

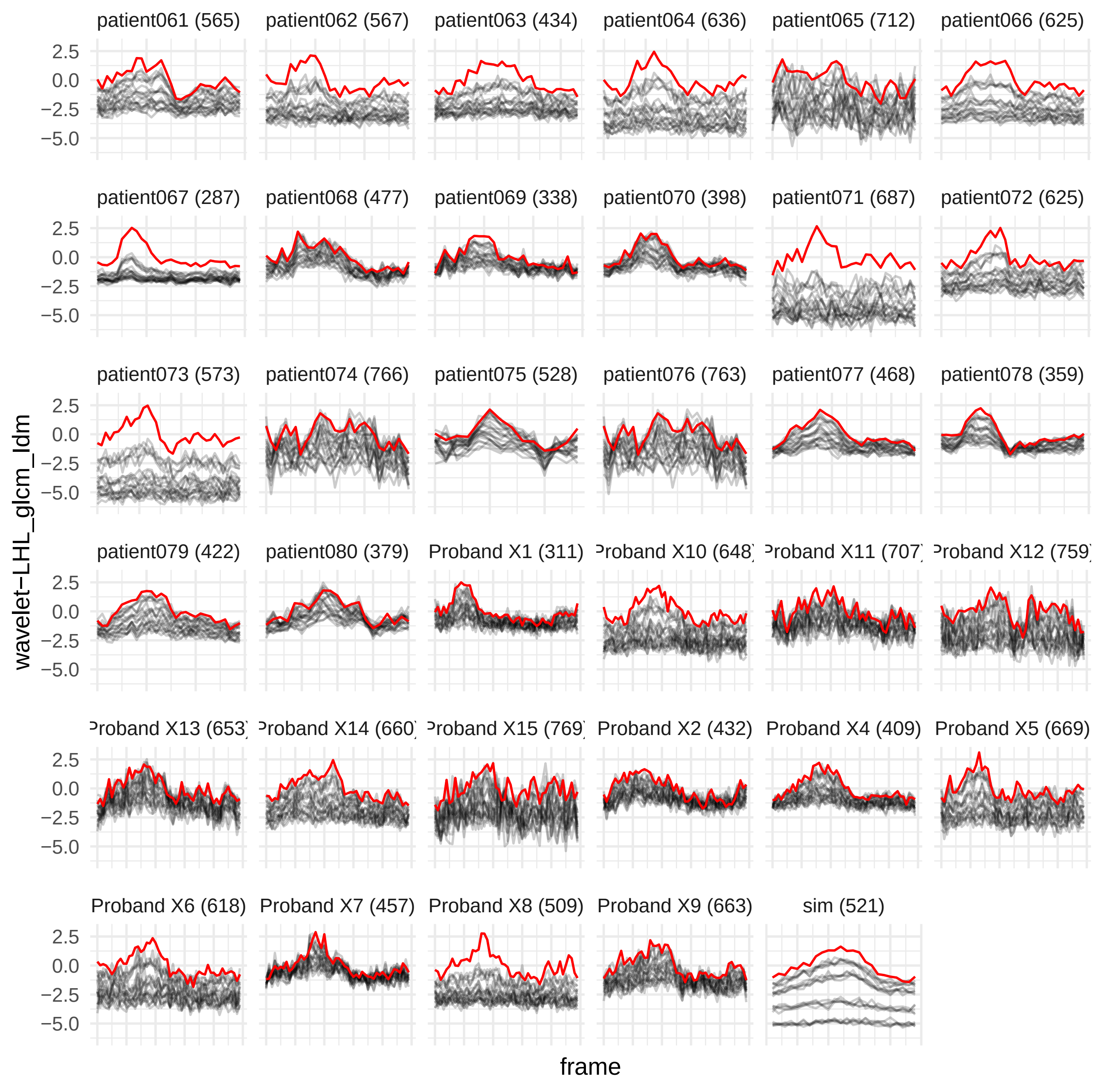

patient061 (565)
patient062 (567)
patient063 (434)
patient064 (636)
patient065 (712)
patient066 (625)
patient067 (287)
patient068 (477)
patient069 (338)
patient070 (398)
patient071 (687)
patient072 (625)
patient073 (573)
patient074 (766)
patient075 (528)
patient076 (763)
patient077 (468)
patient078 (359)
patient079 (422)
patient080 (379)
Proband X1 (311)
Proband X10 (648)
Proband X11 (707)
Proband X12 (759)
Proband X13 (653)
Proband X14 (660)
Proband X15 (769)
Proband X2 (432)
Proband X4 (409)
Proband X5 (669)
Proband X6 (618)
Proband X7 (457)
Proband X8 (509)
Proband X9 (663)
sim (521)
2.5
0.0
−2.5
−5.0
wavelet−LHL_glcm_ldm
frame

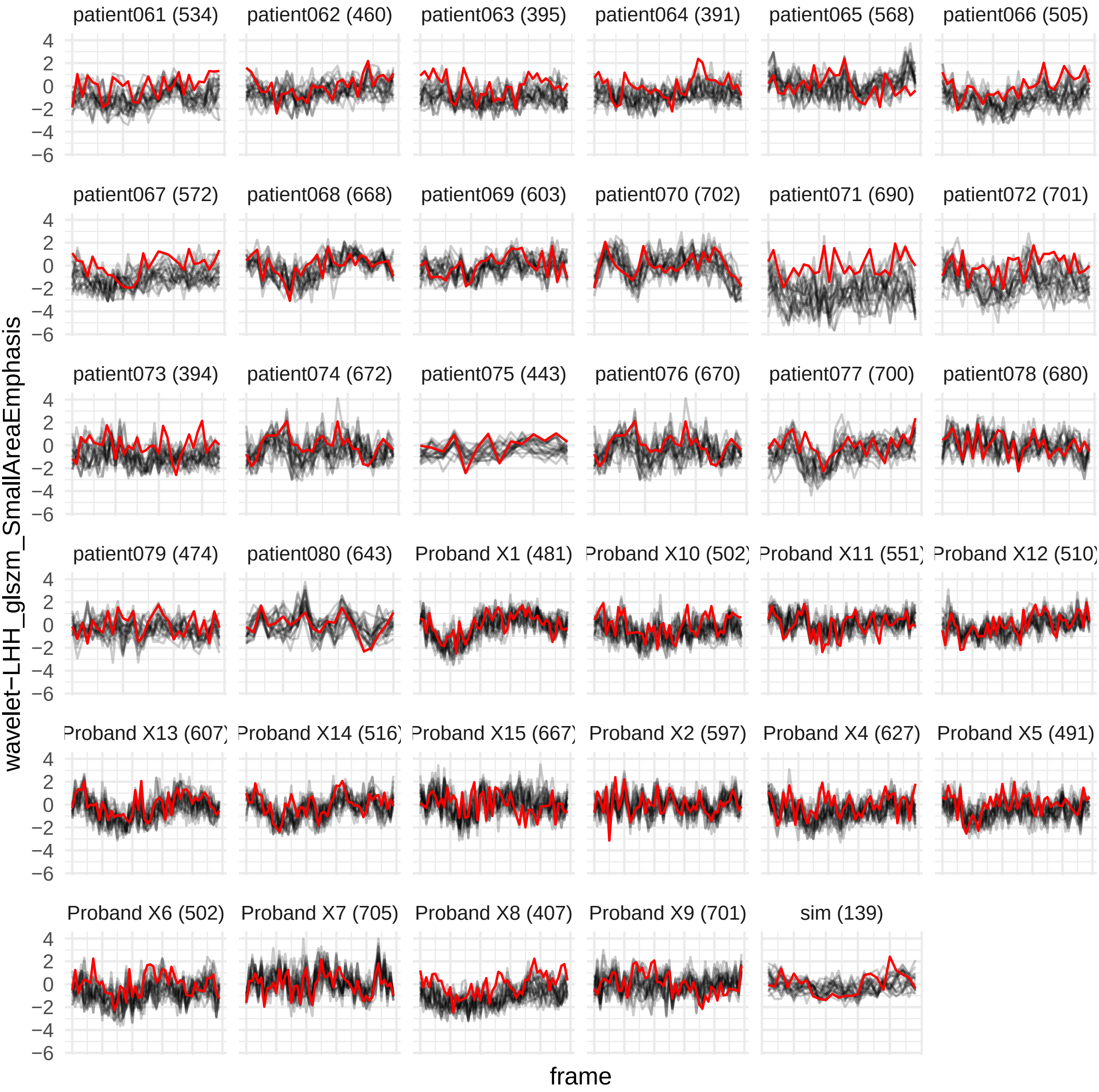

patient061 (534)
patient062 (460)
patient063 (395)
patient064 (391)
patient065 (568)
patient066 (505)
patient067 (572)
patient068 (668)
patient069 (603)
patient070 (702)
patient071 (690)
patient072 (701)
patient073 (394)
patient074 (672)
patient075 (443)
patient076 (670)
patient077 (700)
patient078 (680)
patient079 (474)
patient080 (643)
Proband X1 (481)
Proband X10 (502)
Proband X11 (551)
Proband X12 (510)
Proband X13 (607)
Proband X14 (516)
Proband X15 (667)
Proband X2 (597)
Proband X4 (627)
Proband X5 (491)
Proband X6 (502)
Proband X7 (705)
Proband X8 (407)
Proband X9 (701)
sim (139)
4
2
0
−2
−4
−6
wavelet−LHH_glszm_SmallAreaEmphasis
frame

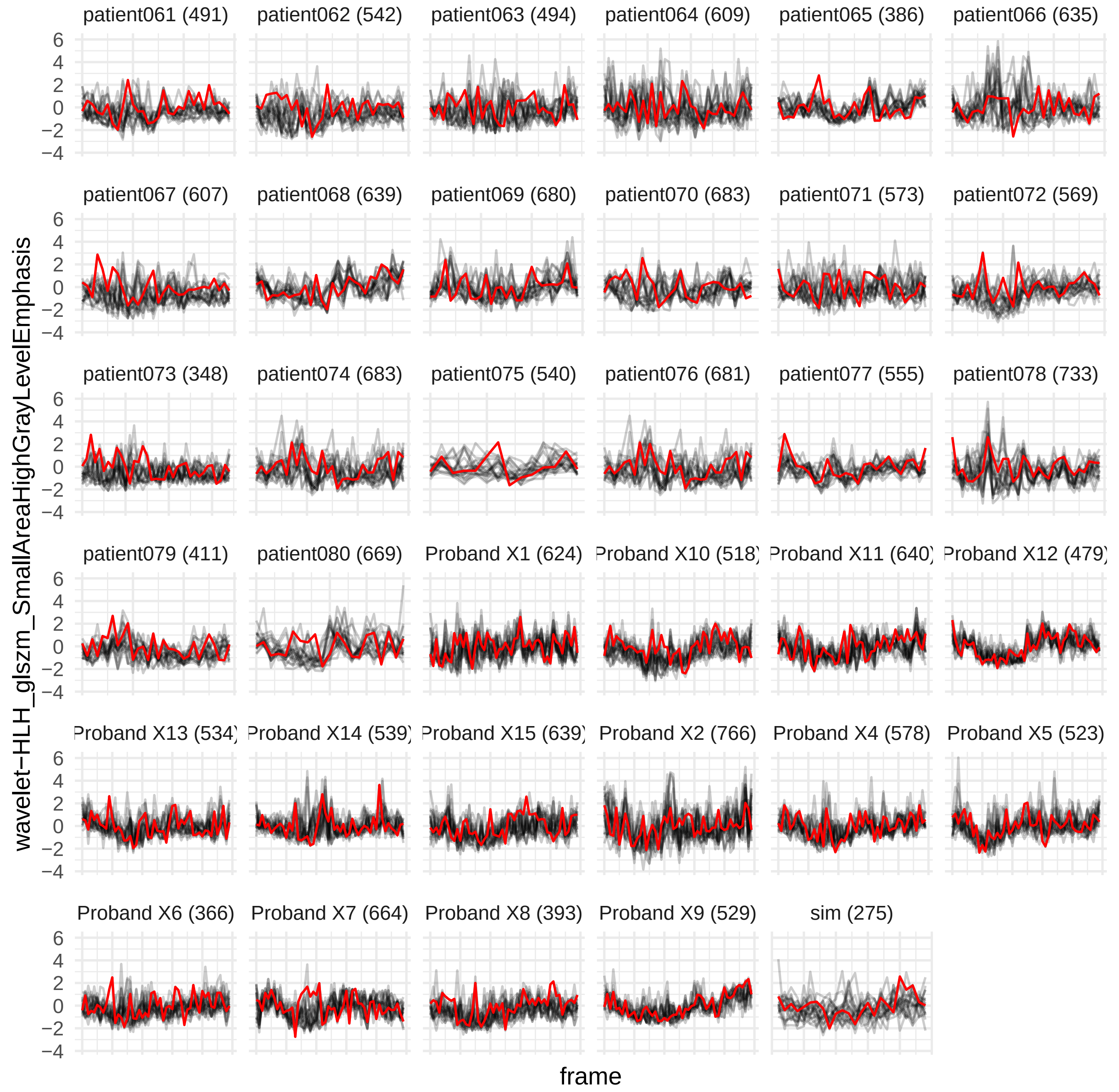

patient061 (491)
patient062 (542)
patient063 (494)
patient064 (609)
patient065 (386)
patient066 (635)
patient067 (607)
patient068 (639)
patient069 (680)
patient070 (683)
patient071 (573)
patient072 (569)
patient073 (348)
patient074 (683)
patient075 (540)
patient076 (681)
patient077 (555)
patient078 (733)
patient079 (411)
patient080 (669)
Proband X1 (624)
Proband X10 (518)
Proband X11 (640)
Proband X12 (479)
Proband X13 (534)
Proband X14 (539)
Proband X15 (639)
Proband X2 (766)
Proband X4 (578)
Proband X5 (523)
Proband X6 (366)
Proband X7 (664)
Proband X8 (393)
Proband X9 (529)
sim (275)
wavelet−HLH_glszm_SmallAreaHighGrayLevelEmphasis
frame

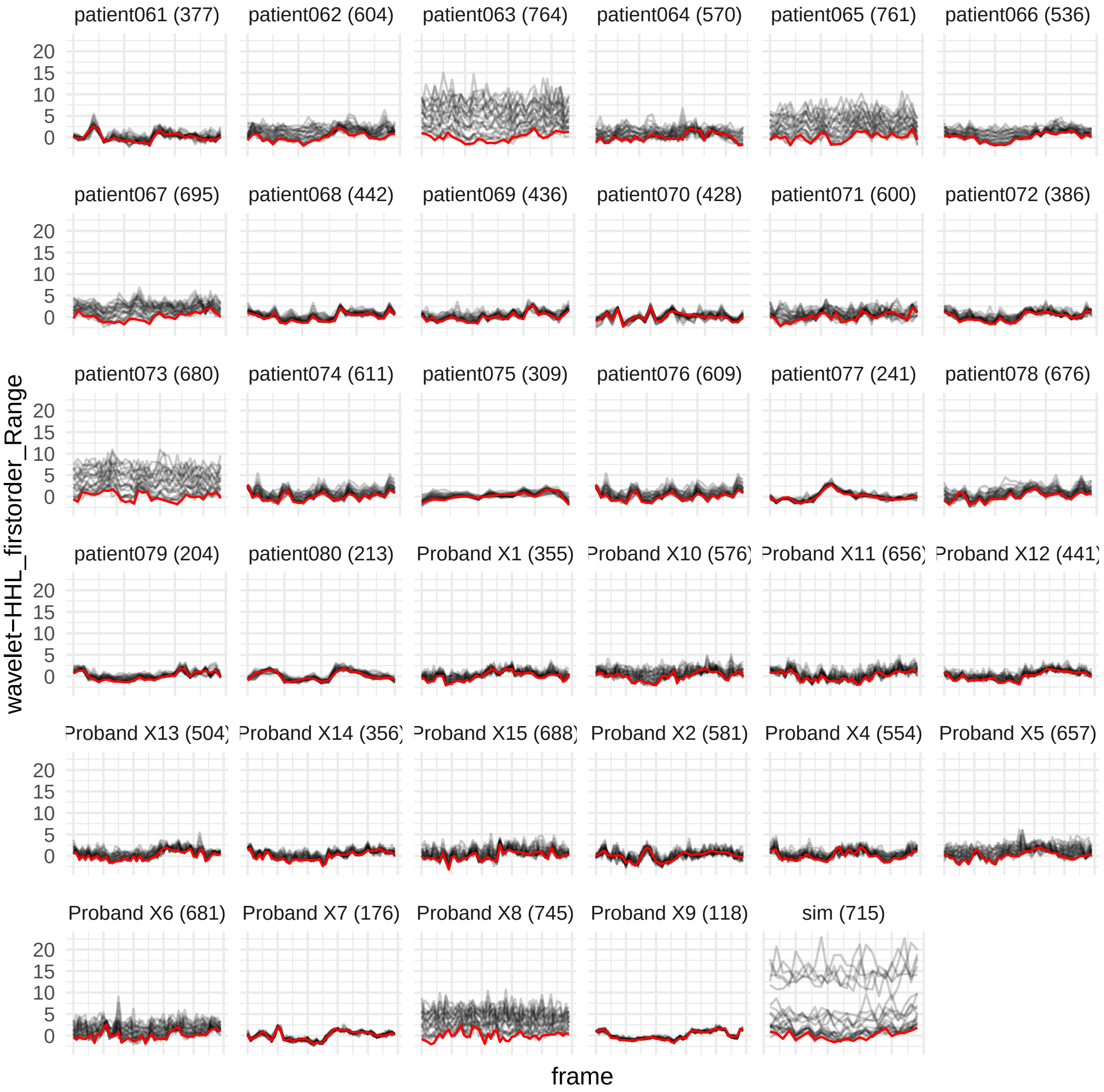

patient061 (377)
patient062 (604)
patient063 (764)
patient064 (570)
patient065 (761)
patient066 (536)
patient067 (695)
patient068 (442)
patient069 (436)
patient070 (428)
patient071 (600)
patient072 (386)
patient073 (680)
patient074 (611)
patient075 (309)
patient076 (609)
patient077 (241)
patient078 (676)
patient079 (204)
patient080 (213)
Proband X1 (355)
Proband X10 (576)
Proband X11 (656)
Proband X12 (441)
Proband X13 (504)
Proband X14 (356)
Proband X15 (688)
Proband X2 (581)
Proband X4 (554)
Proband X5 (657)
Proband X6 (681)
Proband X7 (176)
Proband X8 (745)
Proband X9 (118)
sim (715)
wavelet−HHL_firstorder_Range
20
15
10
5
0
frame

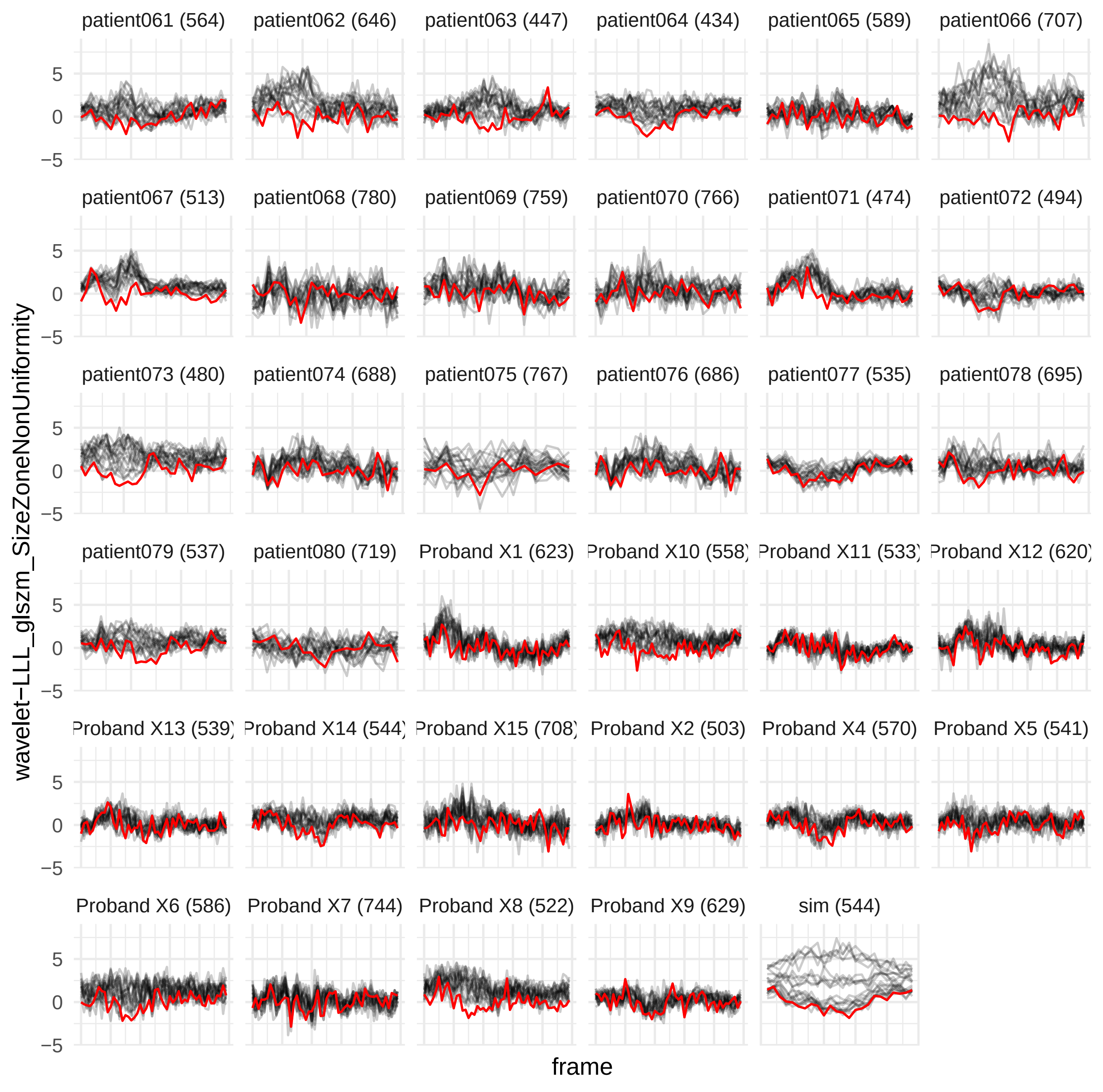
patient061 (564)
patient062 (646)
patient063 (447)
patient064 (434)
patient065 (589)
patient066 (707)
patient067 (513)
patient068 (780)
patient069 (759)
patient070 (766)
patient071 (474)
patient072 (494)
patient073 (480)
patient074 (688)
patient075 (767)
patient076 (686)
patient077 (535)
patient078 (695)
patient079 (537)
patient080 (719)
Proband X1 (623)
Proband X10 (558)
Proband X11 (533)
Proband X12 (620)
Proband X13 (539)
Proband X14 (544)
Proband X15 (708)
Proband X2 (503)
Proband X4 (570)
Proband X5 (541)
Proband X6 (586)
Proband X7 (744)
Proband X8 (522)
Proband X9 (629)
sim (544)
wavelet−LLL_glszm_SizeZoneNonUniformity
frame
5
0
−5

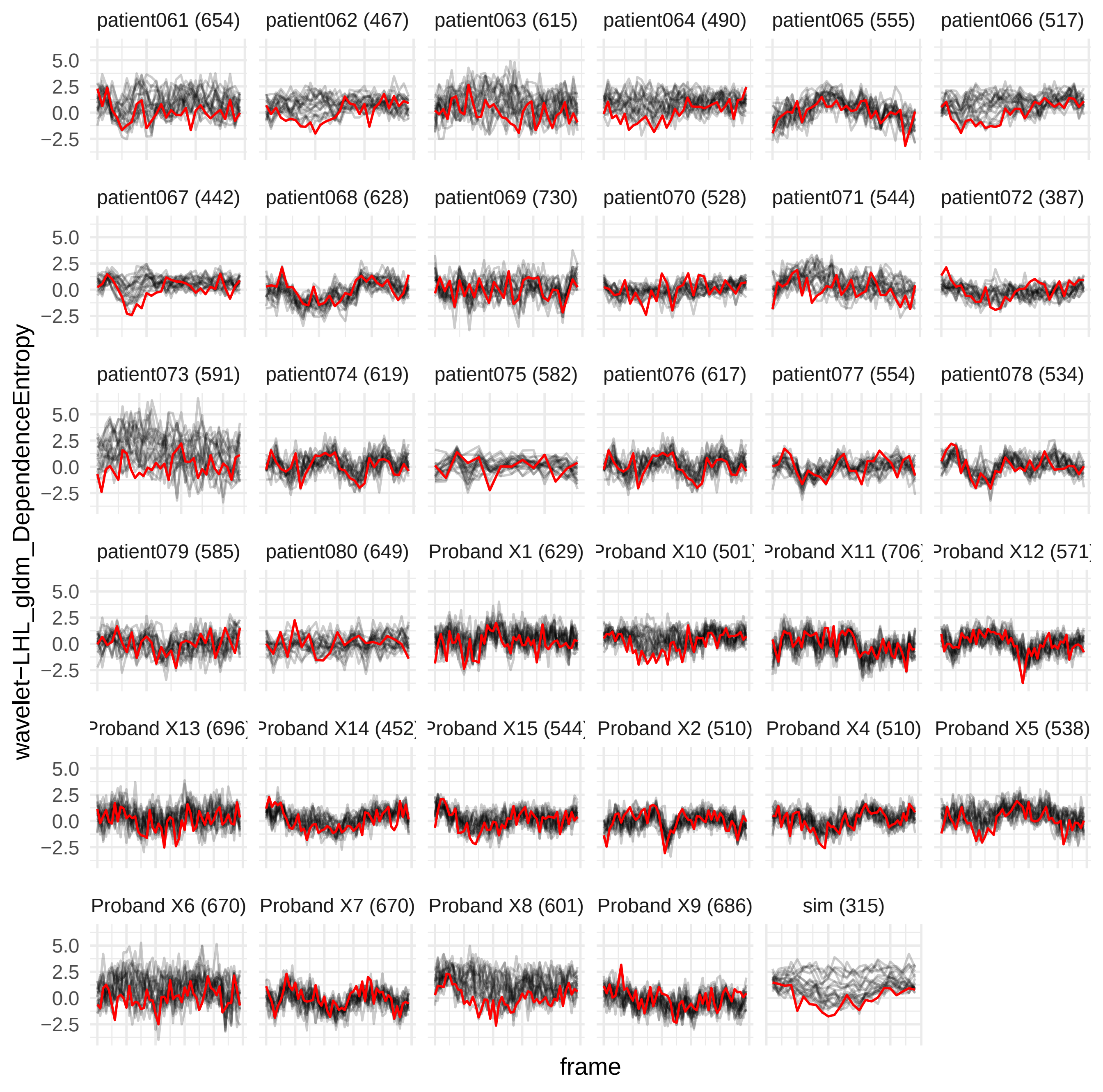

patient061 (654)
patient062 (467)
patient063 (615)
patient064 (490)
patient065 (555)
patient066 (517)
patient067 (442)
patient068 (628)
patient069 (730)
patient070 (528)
patient071 (544)
patient072 (387)
patient073 (591)
patient074 (619)
patient075 (582)
patient076 (617)
patient077 (554)
patient078 (534)
patient079 (585)
patient080 (649)
Proband X1 (629)
Proband X10 (501)
Proband X11 (706)
Proband X12 (571)
Proband X13 (696)
Proband X14 (452)
Proband X15 (544)
Proband X2 (510)
Proband X4 (510)
Proband X5 (538)
Proband X6 (670)
Proband X7 (670)
Proband X8 (601)
Proband X9 (686)
sim (315)
5.0
2.5
0.0
−2.5
wavelet−LHL_gldm_DependenceEntropy
frame

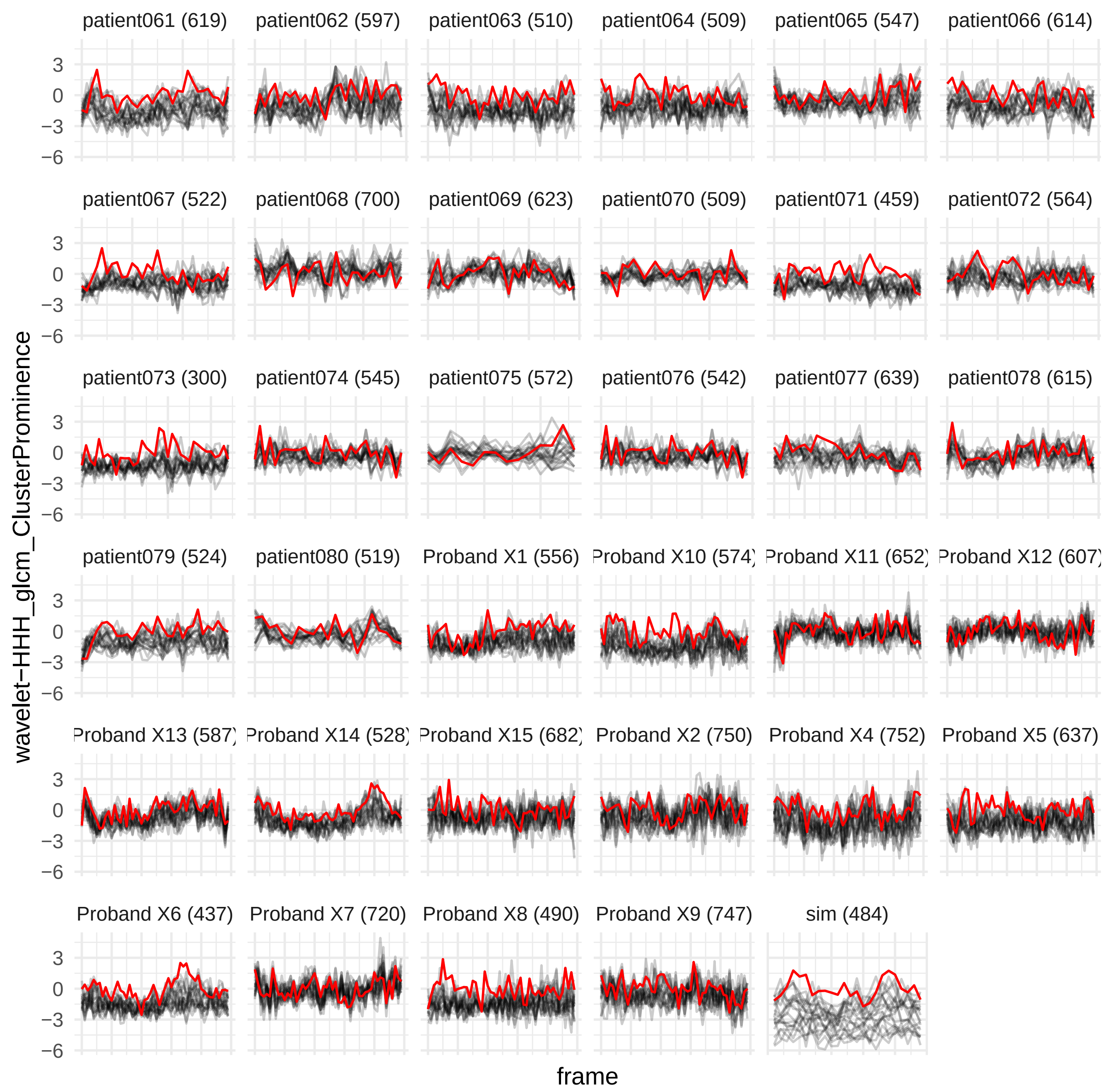

patient061 (619)
patient062 (597)
patient063 (510)
patient064 (509)
patient065 (547)
patient066 (614)
patient067 (522)
patient068 (700)
patient069 (623)
patient070 (509)
patient071 (459)
patient072 (564)
patient073 (300)
patient074 (545)
patient075 (572)
patient076 (542)
patient077 (639)
patient078 (615)
patient079 (524)
patient080 (519)
Proband X1 (556)
Proband X10 (574)
Proband X11 (652)
Proband X12 (607)
Proband X13 (587)
Proband X14 (528)
Proband X15 (682)
Proband X2 (750)
Proband X4 (752)
Proband X5 (637)
Proband X6 (437)
Proband X7 (720)
Proband X8 (490)
Proband X9 (747)
sim (484)
wavelet−HHH_glcm_ClusterProminence
frame
3
0
−3
−6

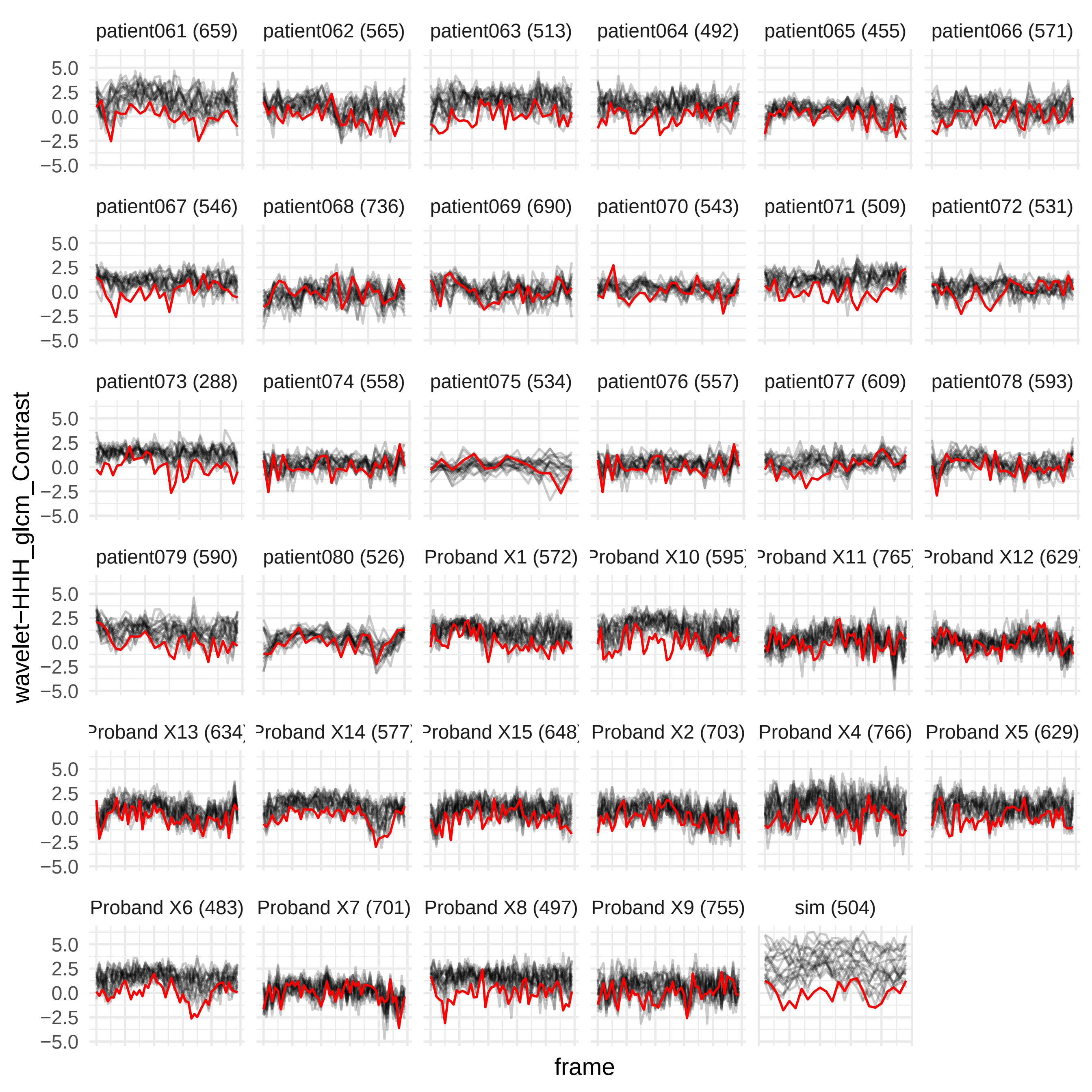

patient061 (659)
patient062 (565)
patient063 (513)
patient064 (492)
patient065 (455)
patient066 (571)
patient067 (546)
patient068 (736)
patient069 (690)
patient070 (543)
patient071 (509)
patient072 (531)
patient073 (288)
patient074 (558)
patient075 (534)
patient076 (557)
patient077 (609)
patient078 (593)
patient079 (590)
patient080 (526)
Proband X1 (572)
Proband X10 (595)
Proband X11 (765)
Proband X12 (629)
Proband X13 (634)
Proband X14 (577)
Proband X15 (648)
Proband X2 (703)
Proband X4 (766)
Proband X5 (629)
Proband X6 (483)
Proband X7 (701)
Proband X8 (497)
Proband X9 (755)
sim (504)
5.0
2.5
0.0
−2.5
−5.0
wavelet−HHH_glcm_Contrast
frame

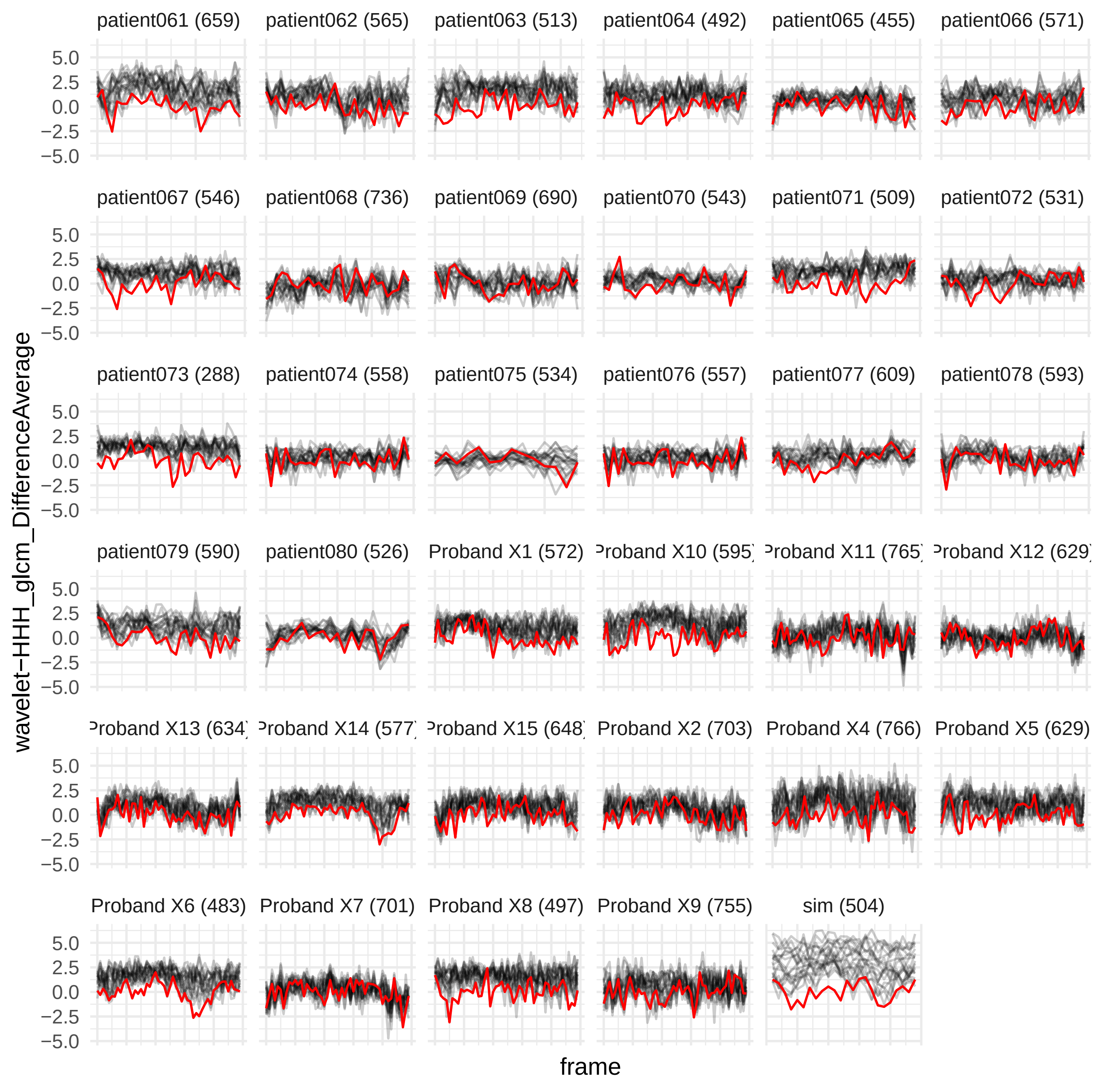
patient061 (659)
patient062 (565)
patient063 (513)
patient064 (492)
patient065 (455)
patient066 (571)
patient067 (546)
patient068 (736)
patient069 (690)
patient070 (543)
patient071 (509)
patient072 (531)
patient073 (288)
patient074 (558)
patient075 (534)
patient076 (557)
patient077 (609)
patient078 (593)
patient079 (590)
patient080 (526)
Proband X1 (572)
Proband X2 (703)
Proband X4 (766)
Proband X5 (629)
Proband X6 (483)
Proband X7 (701)
Proband X8 (497)
Proband X9 (755)
sim (504)
wavelet−HHH_glcm_DifferenceAverage
frame
5.0
2.5
0.0
−2.5
−5.0

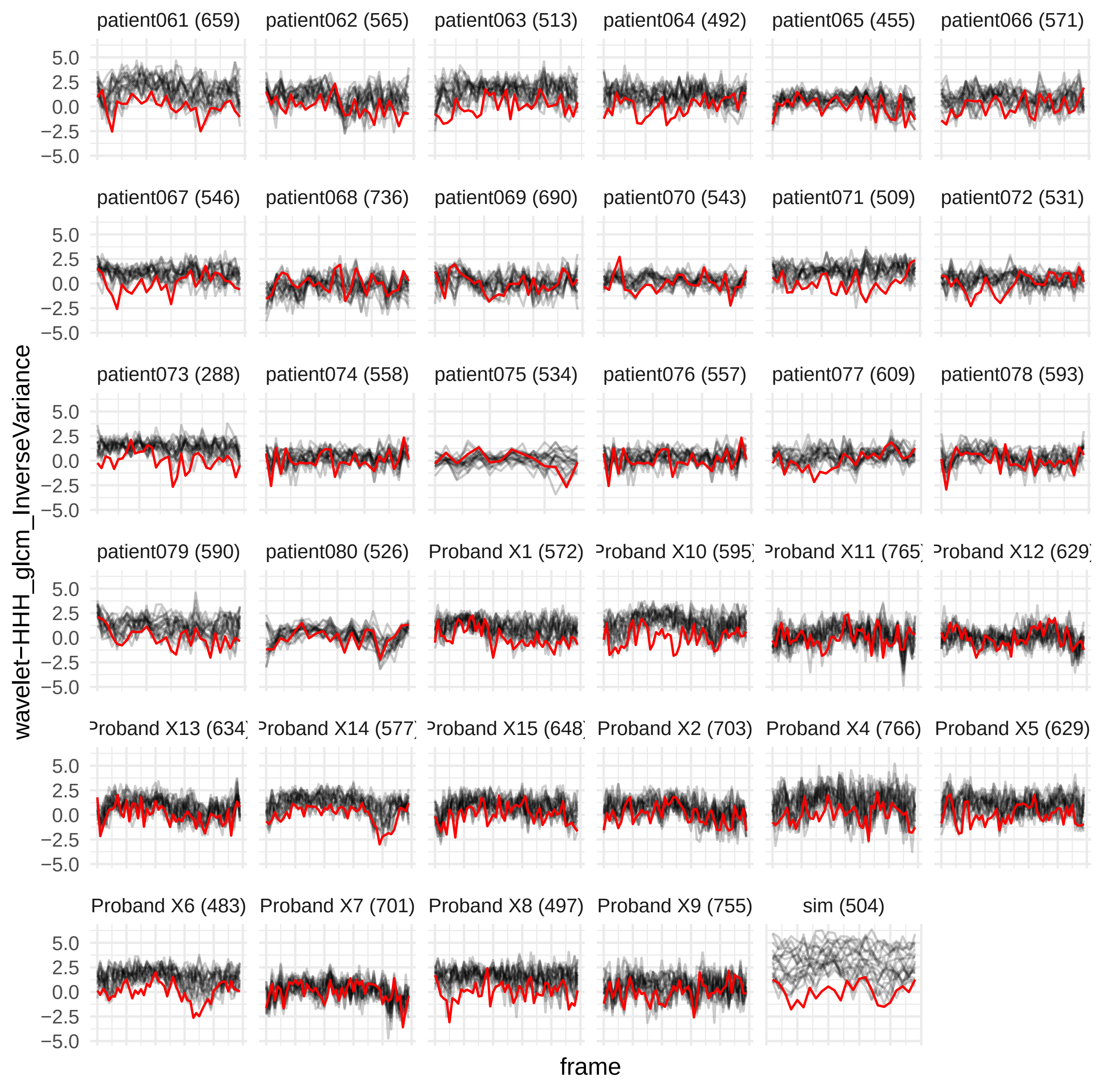

patient061 (659)
patient062 (565)
patient063 (513)
patient064 (492)
patient065 (455)
patient066 (571)
patient067 (546)
patient068 (736)
patient069 (690)
patient070 (543)
patient071 (509)
patient072 (531)
patient073 (288)
patient074 (558)
patient075 (534)
patient076 (557)
patient077 (609)
patient078 (593)
patient079 (590)
patient080 (526)
Proband X1 (572)
Proband X10 (595)
Proband X11 (765)
Proband X12 (629)
Proband X13 (634)
Proband X14 (577)
Proband X15 (648)
Proband X2 (703)
Proband X4 (766)
Proband X5 (629)
Proband X6 (483)
Proband X7 (701)
Proband X8 (497)
Proband X9 (755)
sim (504)
5.0
2.5
0.0
−2.5
−5.0
wavelet−HHH_glcm_InverseVariance
frame

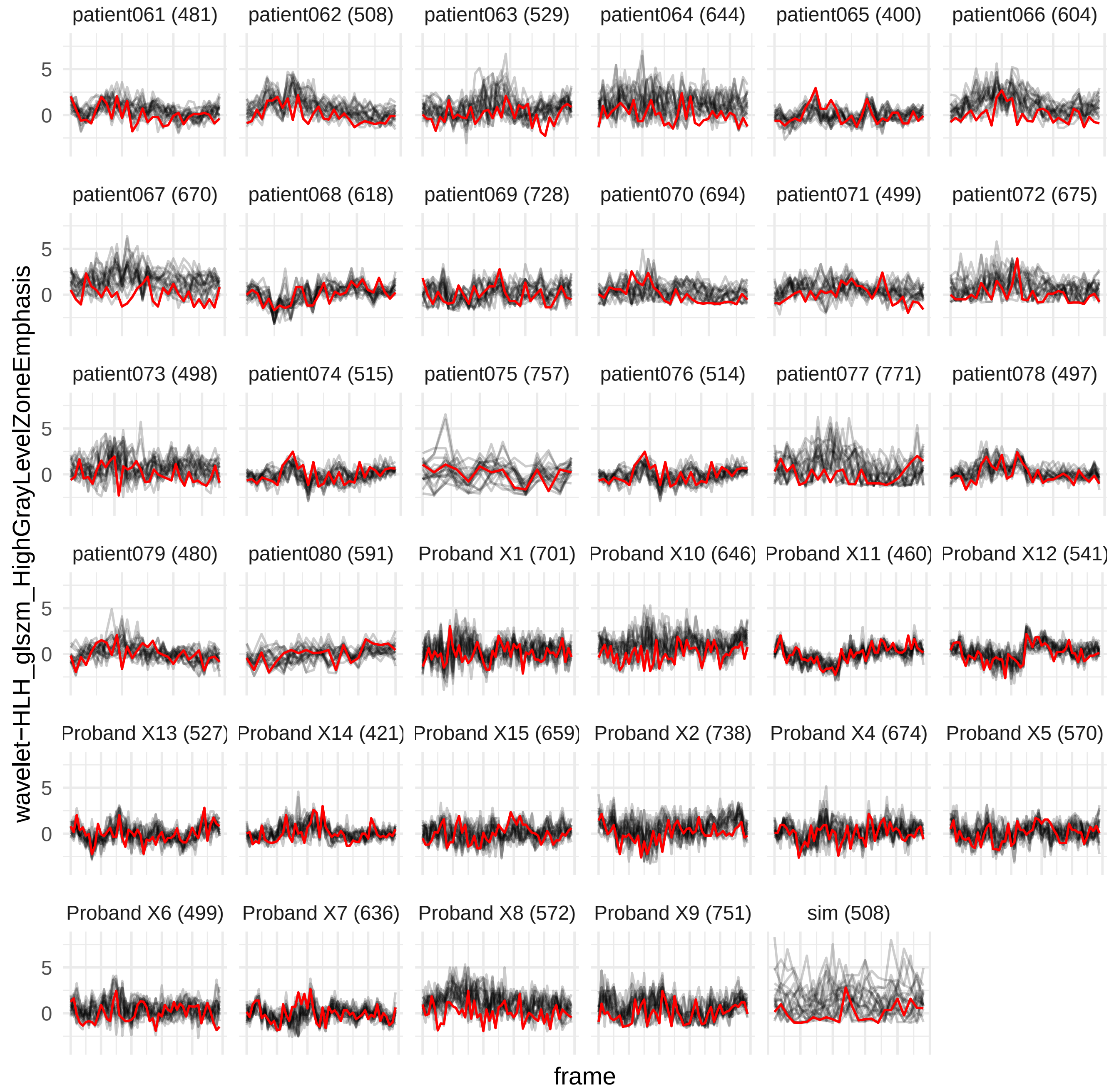

patient061 (481)
patient062 (508)
patient063 (529)
patient064 (644)
patient065 (400)
patient066 (604)
patient067 (670)
patient068 (618)
patient069 (728)
patient070 (694)
patient071 (499)
patient072 (675)
patient073 (498)
patient074 (515)
patient075 (757)
patient076 (514)
patient077 (771)
patient078 (497)
patient079 (480)
patient080 (591)
Proband X1 (701)
Proband X10 (646)
Proband X11 (460)
Proband X12 (541)
Proband X13 (527)
Proband X14 (421)
Proband X15 (659)
Proband X2 (738)
Proband X4 (674)
Proband X5 (570)
Proband X6 (499)
Proband X7 (636)
Proband X8 (572)
Proband X9 (751)
sim (508)
5
0
wavelet−HLH_glszm_HighGrayLevelZoneEmphasis
frame

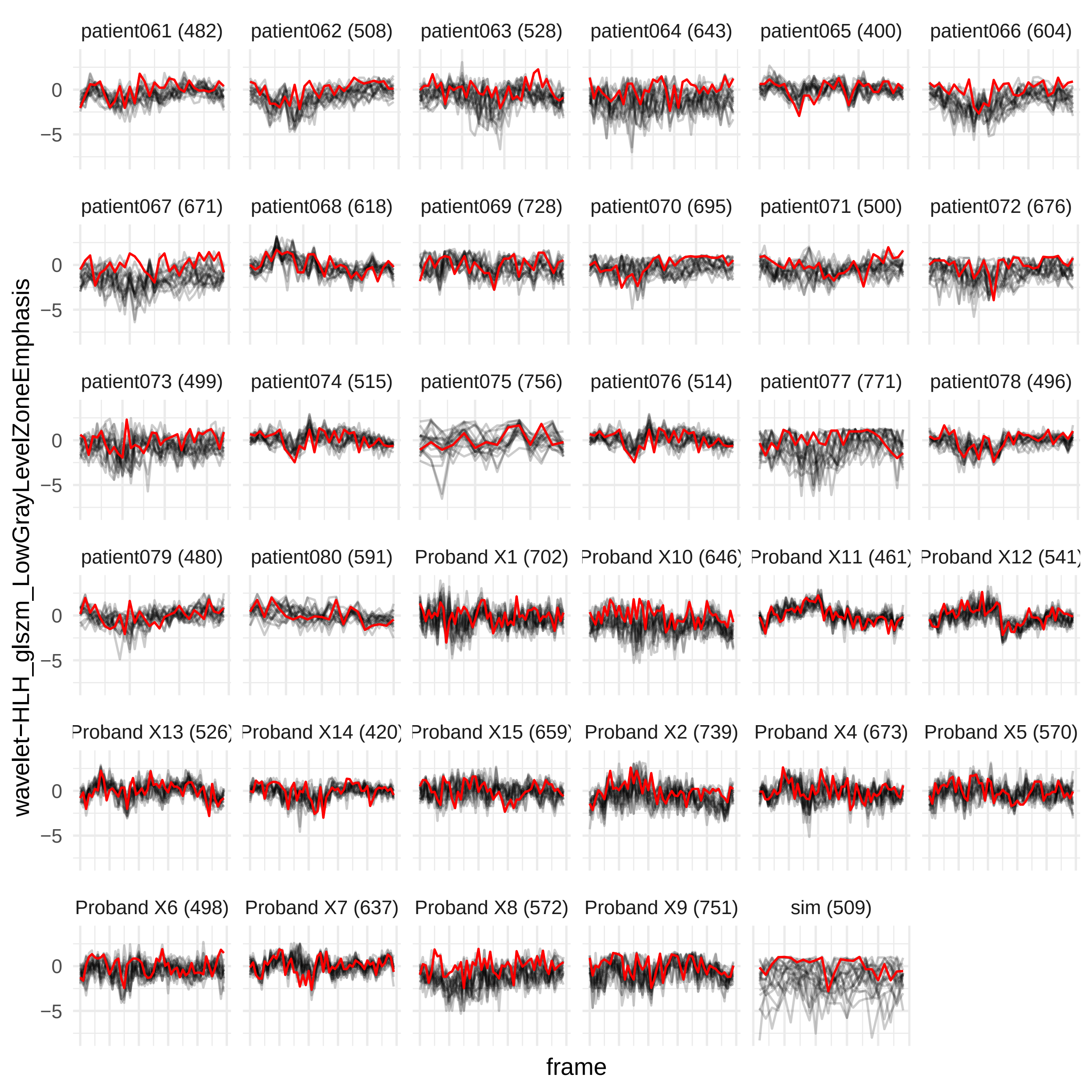
patient061 (482)
patient062 (508)
patient063 (528)
patient064 (643)
patient065 (400)
patient066 (604)
patient067 (671)
patient068 (618)
patient069 (728)
patient070 (695)
patient071 (500)
patient072 (676)
patient073 (499)
patient074 (515)
patient075 (756)
patient076 (514)
patient077 (771)
patient078 (496)
patient079 (480)
patient080 (591)
Proband X1 (702)
Proband X10 (646)
Proband X11 (461)
Proband X12 (541)
Proband X13 (526)
Proband X14 (420)
Proband X15 (659)
Proband X2 (739)
Proband X4 (673)
Proband X5 (570)
Proband X6 (498)
Proband X7 (637)
Proband X8 (572)
Proband X9 (751)
sim (509)
0
−5
wavelet−HLH_glszm_LowGrayLevelZoneEmphasis
frame

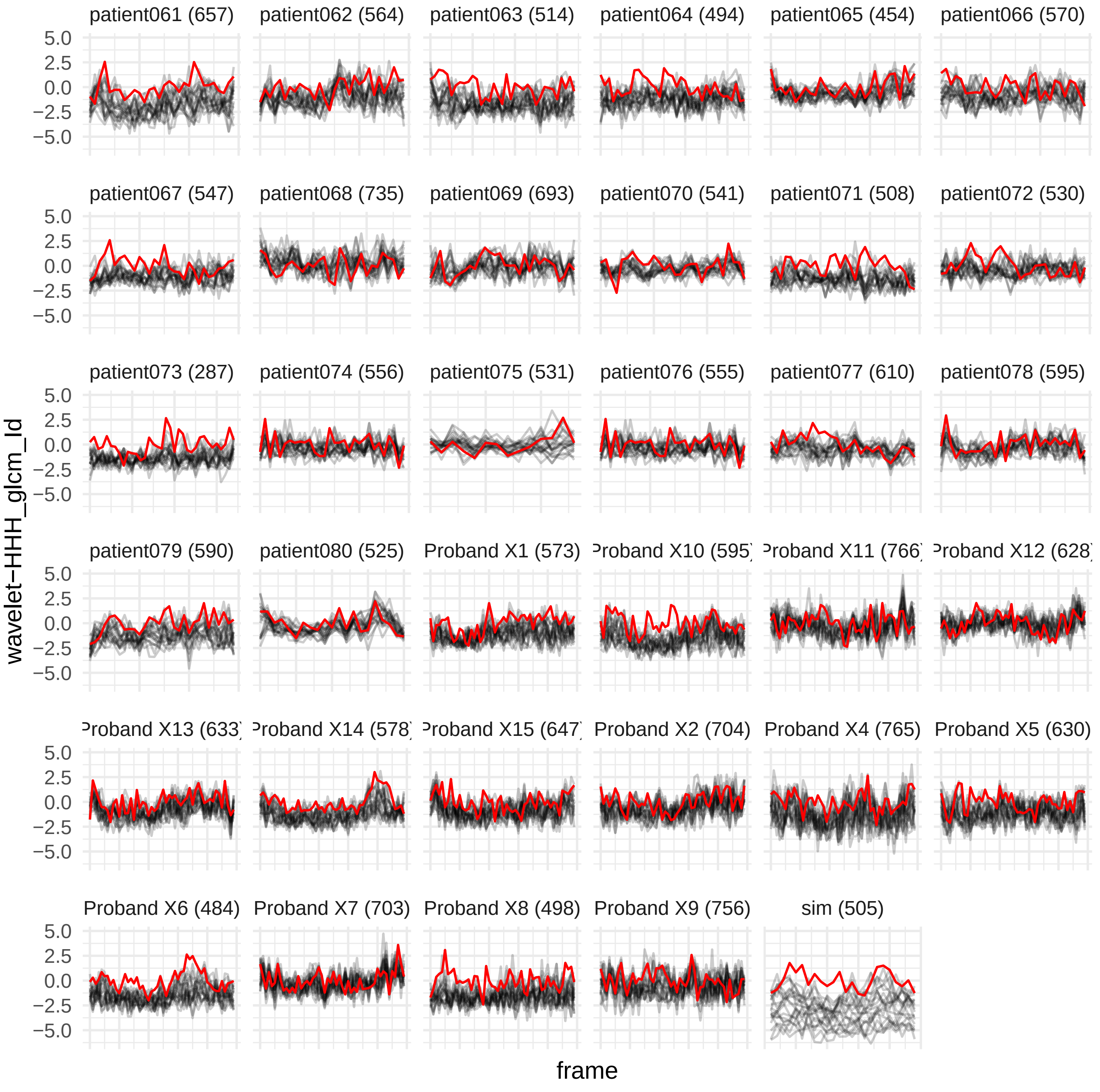
patient061 (657)
patient062 (564)
patient063 (514)
patient064 (494)
patient065 (454)
patient066 (570)
patient067 (547)
patient068 (735)
patient069 (693)
patient070 (541)
patient071 (508)
patient072 (530)
patient073 (287)
patient074 (556)
patient075 (531)
patient076 (555)
patient077 (610)
patient078 (595)
patient079 (590)
patient080 (525)
Proband X1 (573)
Proband X10 (595)
Proband X11 (766)
Proband X12 (628)
Proband X13 (633)
Proband X14 (578)
Proband X15 (647)
Proband X2 (704)
Proband X4 (765)
Proband X5 (630)
Proband X6 (484)
Proband X7 (703)
Proband X8 (498)
Proband X9 (756)
sim (505)
wavelet−HHH_glcm_Id
frame
5.0
2.5
0.0
−2.5
−5.0

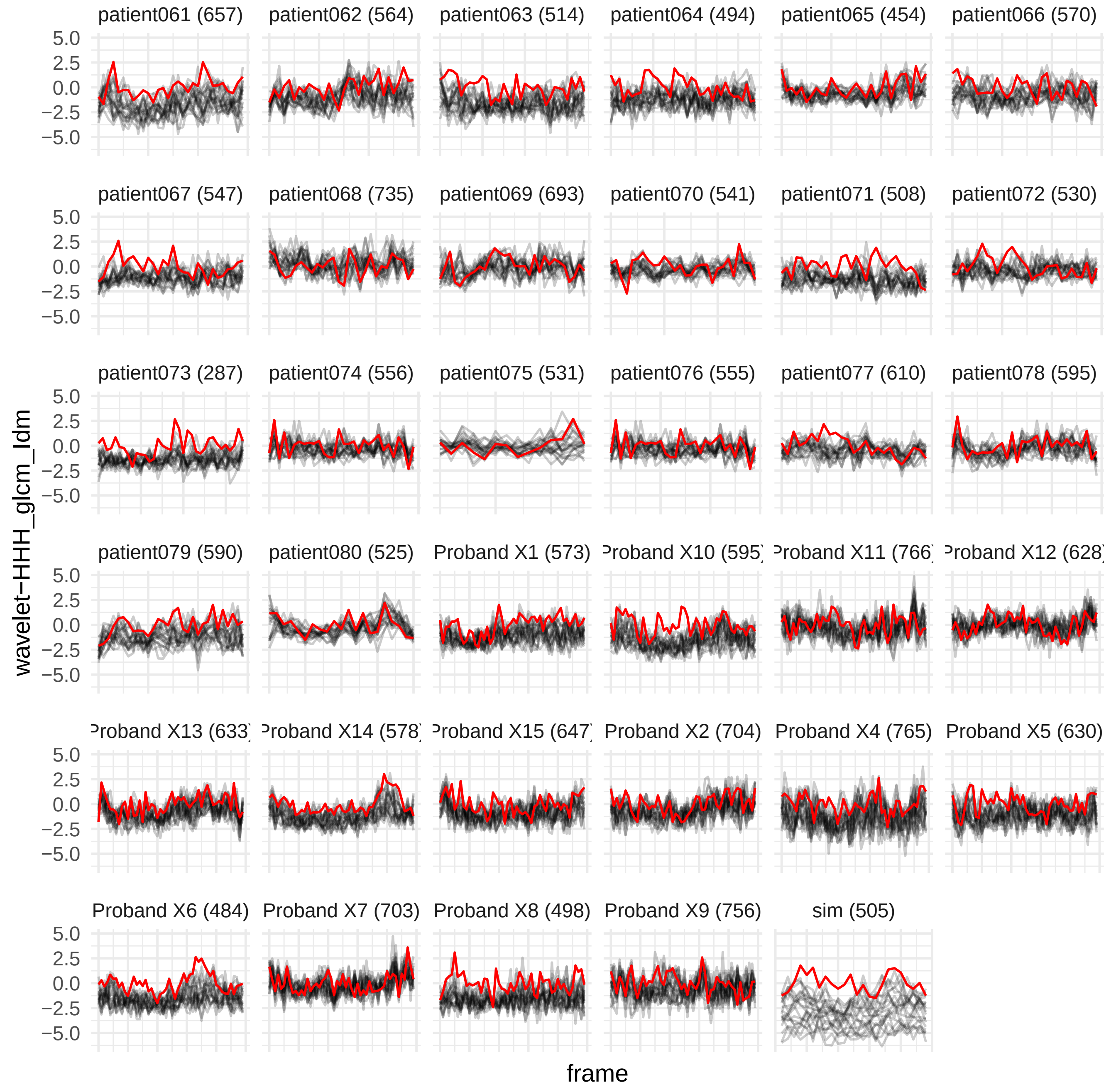

patient061 (657)
patient062 (564)
patient063 (514)
patient064 (494)
patient065 (454)
patient066 (570)
patient067 (547)
patient068 (735)
patient069 (693)
patient070 (541)
patient071 (508)
patient072 (530)
patient073 (287)
patient074 (556)
patient075 (531)
patient076 (555)
patient077 (610)
patient078 (595)
patient079 (590)
patient080 (525)
Proband X1 (573)
Proband X10 (595)
Proband X11 (766)
Proband X12 (628)
Proband X13 (633)
Proband X14 (578)
Proband X15 (647)
Proband X2 (704)
Proband X4 (765)
Proband X5 (630)
Proband X6 (484)
Proband X7 (703)
Proband X8 (498)
Proband X9 (756)
sim (505)
5.0
2.5
0.0
−2.5
−5.0
wavelet−HHH_glcm_ldm
frame

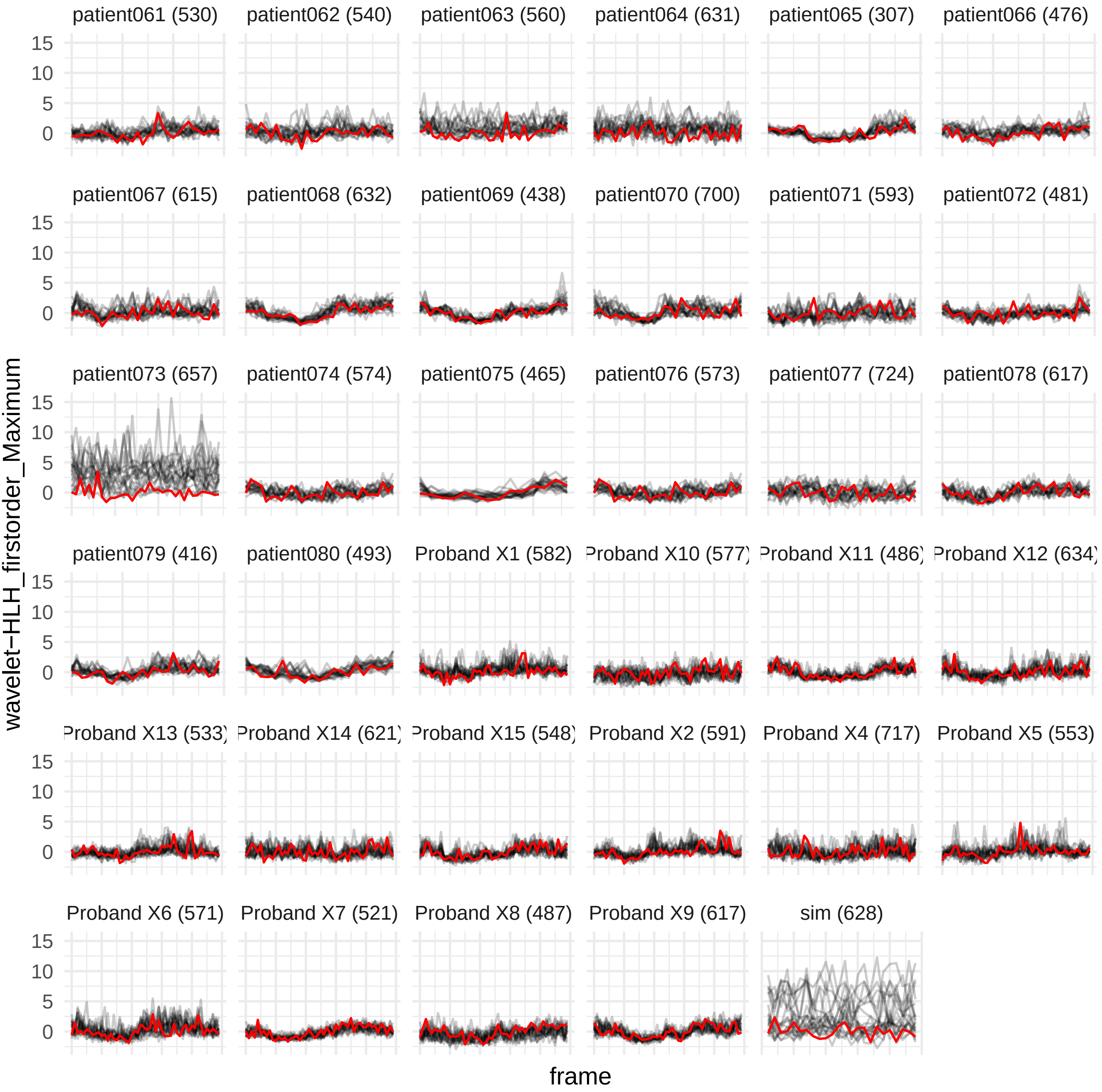

patient061 (530)
patient062 (540)
patient063 (560)
patient064 (631)
patient065 (307)
patient066 (476)
patient067 (615)
patient068 (632)
patient069 (438)
patient070 (700)
patient071 (593)
patient072 (481)
patient073 (657)
patient074 (574)
patient075 (465)
patient076 (573)
patient077 (724)
patient078 (617)
patient079 (416)
patient080 (493)
Proband X1 (582)
Proband X10 (577)
Proband X11 (486)
Proband X12 (634)
Proband X13 (533)
Proband X14 (621)
Proband X15 (548)
Proband X2 (591)
Proband X4 (717)
Proband X5 (553)
Proband X6 (571)
Proband X7 (521)
Proband X8 (487)
Proband X9 (617)
sim (628)
wavelet−HLH_firstorder_Maximum
frame
15
10
5
0

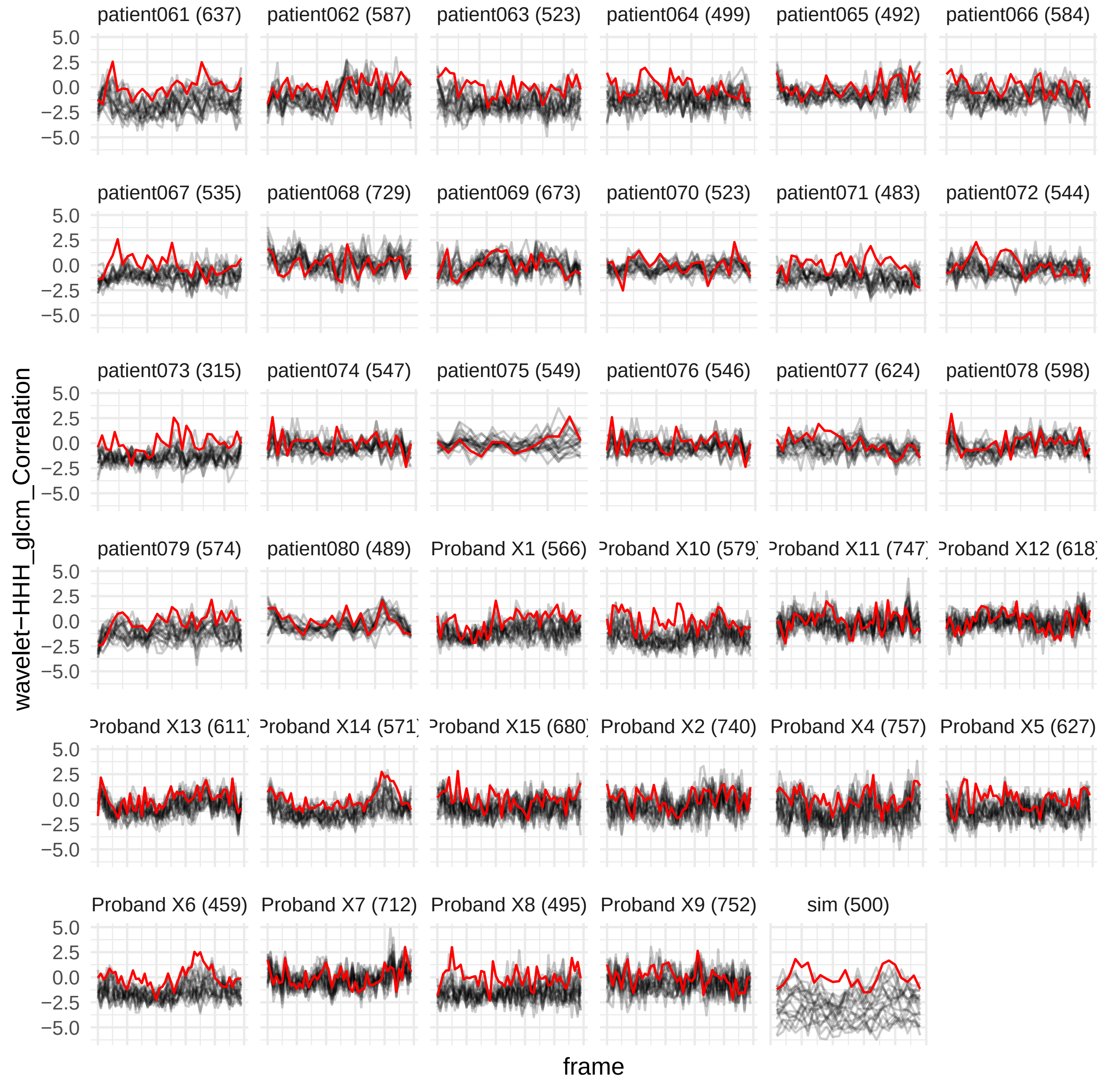
patient061 (637)
patient062 (587)
patient063 (523)
patient064 (499)
patient065 (492)
patient066 (584)
patient067 (535)
patient068 (729)
patient069 (673)
patient070 (523)
patient071 (483)
patient072 (544)
patient073 (315)
patient074 (547)
patient075 (549)
patient076 (546)
patient077 (624)
patient078 (598)
patient079 (574)
patient080 (489)
Proband X1 (566)
Proband X10 (579)
Proband X11 (747)
Proband X12 (618)
Proband X13 (611)
Proband X14 (571)
Proband X15 (680)
Proband X2 (740)
Proband X4 (757)
Proband X5 (627)
Proband X6 (459)
Proband X7 (712)
Proband X8 (495)
Proband X9 (752)
sim (500)
5.0
2.5
0.0
−2.5
−5.0
wavelet−HHH_glcm_Correlation
frame

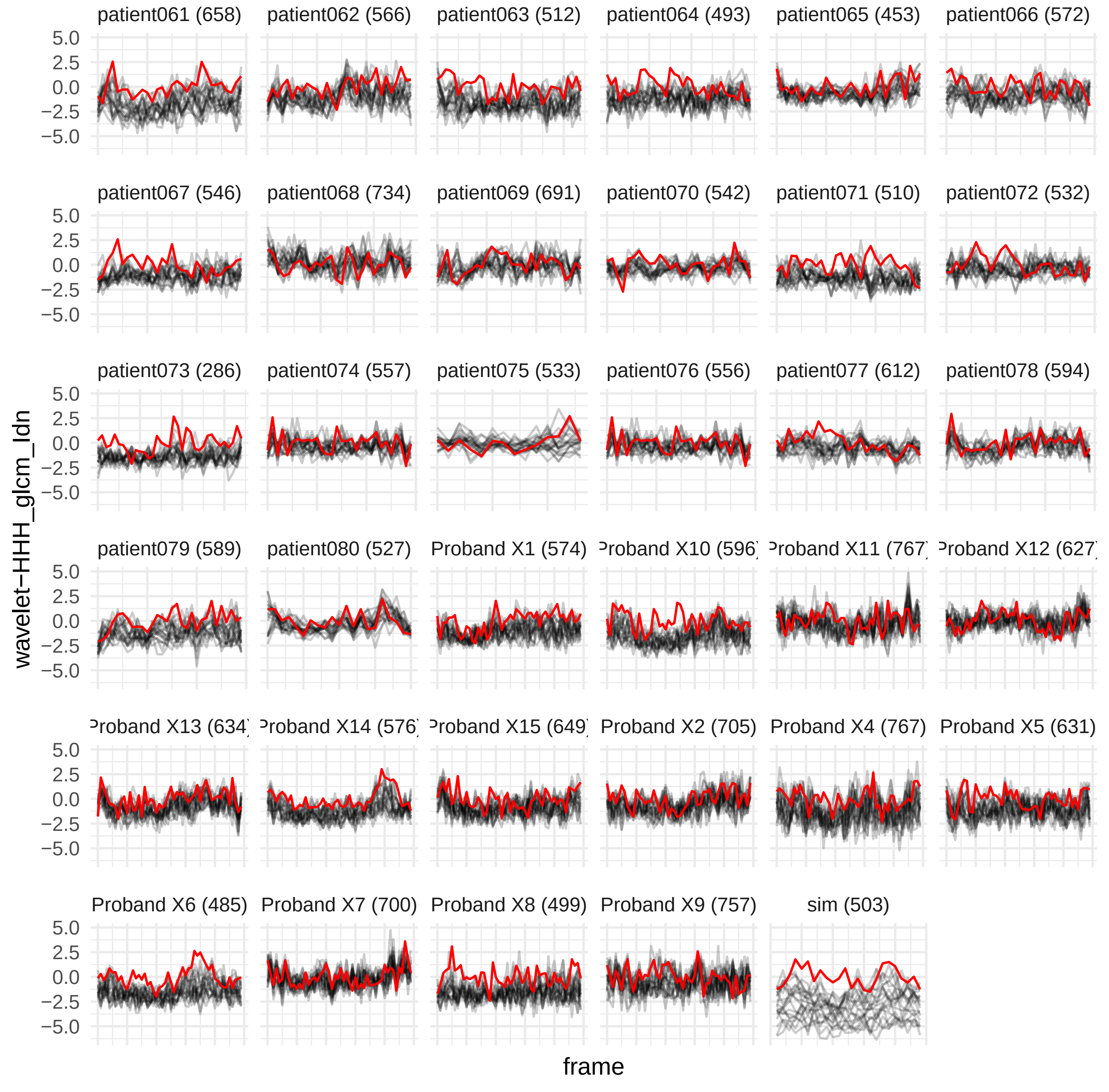

patient061 (658)
patient062 (566)
patient063 (512)
patient064 (493)
patient065 (453)
patient066 (572)
patient067 (546)
patient068 (734)
patient069 (691)
patient070 (542)
patient071 (510)
patient072 (532)
patient073 (286)
patient074 (557)
patient075 (533)
patient076 (556)
patient077 (612)
patient078 (594)
patient079 (589)
patient080 (527)
Proband X1 (574)
Proband X10 (596)
Proband X11 (767)
Proband X12 (627)
Proband X13 (634)
Proband X14 (576)
Proband X15 (649)
Proband X2 (705)
Proband X4 (767)
Proband X5 (631)
Proband X6 (485)
Proband X7 (700)
Proband X8 (499)
Proband X9 (757)
sim (503)
5.0
2.5
0.0
−2.5
−5.0
wavelet−HHH_glcm_Idn
frame

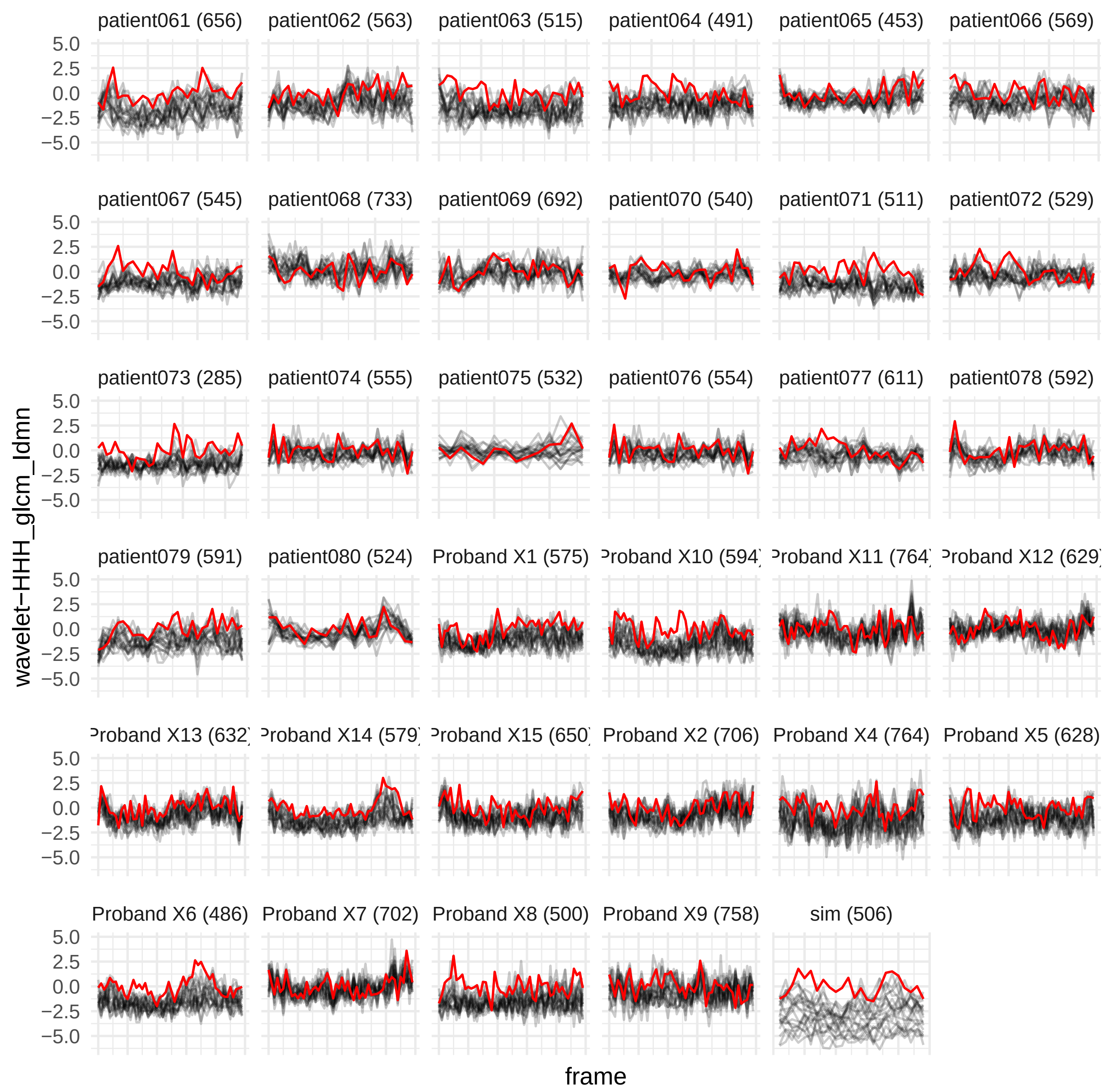

patient061 (656)
patient062 (563)
patient063 (515)
patient064 (491)
patient065 (453)
patient066 (569)
patient067 (545)
patient068 (733)
patient069 (692)
patient070 (540)
patient071 (511)
patient072 (529)
patient073 (285)
patient074 (555)
patient075 (532)
patient076 (554)
patient077 (611)
patient078 (592)
patient079 (591)
patient080 (524)
Proband X1 (575)
Proband X10 (594)
Proband X11 (764)
Proband X12 (629)
Proband X13 (632)
Proband X14 (579)
Proband X15 (650)
Proband X2 (706)
Proband X4 (764)
Proband X5 (628)
Proband X6 (486)
Proband X7 (702)
Proband X8 (500)
Proband X9 (758)
sim (506)
5.0
2.5
0.0
−2.5
−5.0
wavelet−HHH_glcm_Idmn
frame

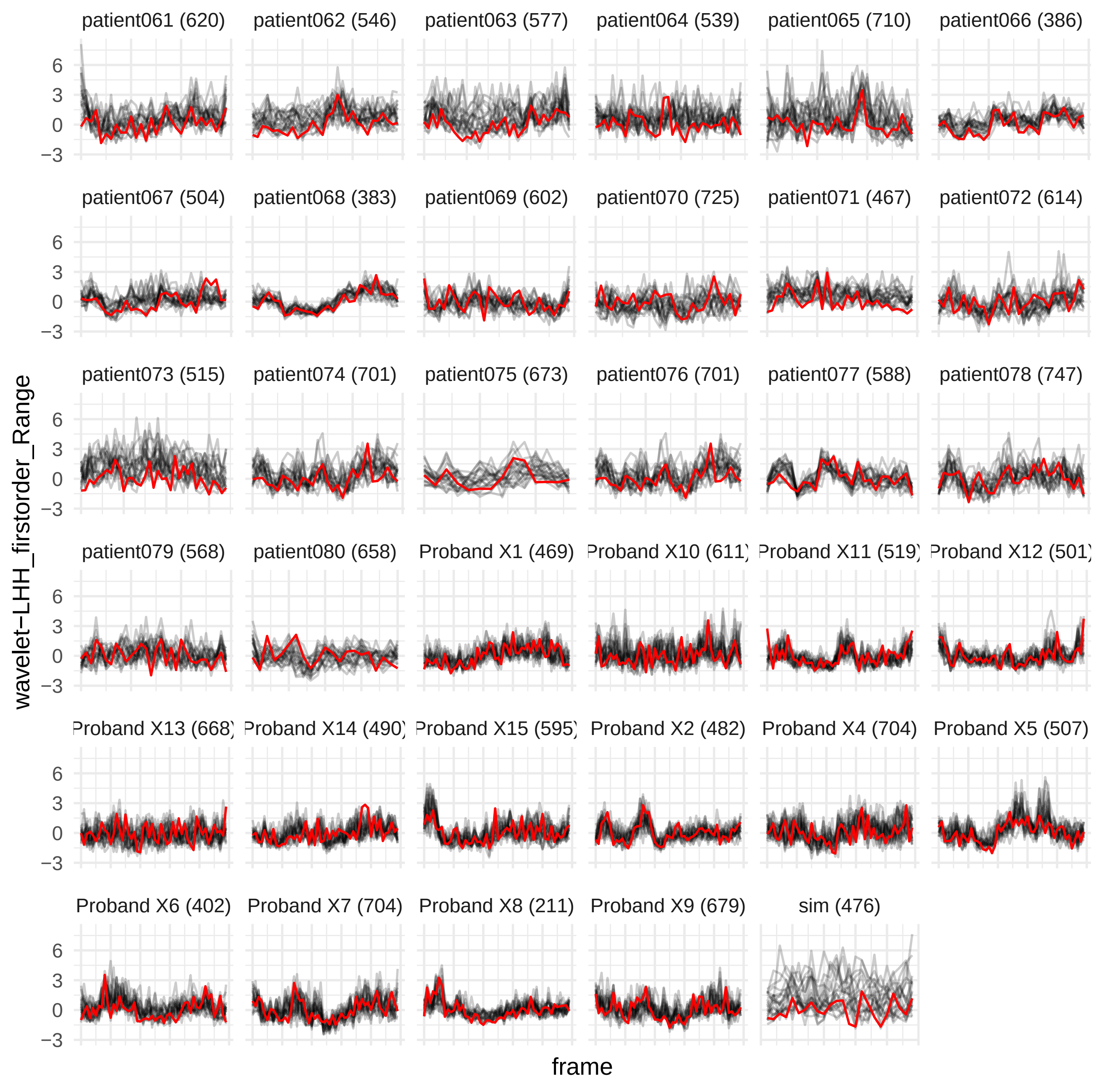

patient061 (620)
patient062 (546)
patient063 (577)
patient064 (539)
patient065 (710)
patient066 (386)
patient067 (504)
patient068 (383)
patient069 (602)
patient070 (725)
patient071 (467)
patient072 (614)
patient073 (515)
patient074 (701)
patient075 (673)
patient076 (701)
patient077 (588)
patient078 (747)
patient079 (568)
patient080 (658)
Proband X1 (469)
Proband X10 (611)
Proband X11 (519)
Proband X12 (501)
Proband X13 (668)
Proband X14 (490)
Proband X15 (595)
Proband X2 (482)
Proband X4 (704)
Proband X5 (507)
Proband X6 (402)
Proband X7 (704)
Proband X8 (211)
Proband X9 (679)
sim (476)
wavelet−LHH_firstorder_Range
frame
6
3
0
−3

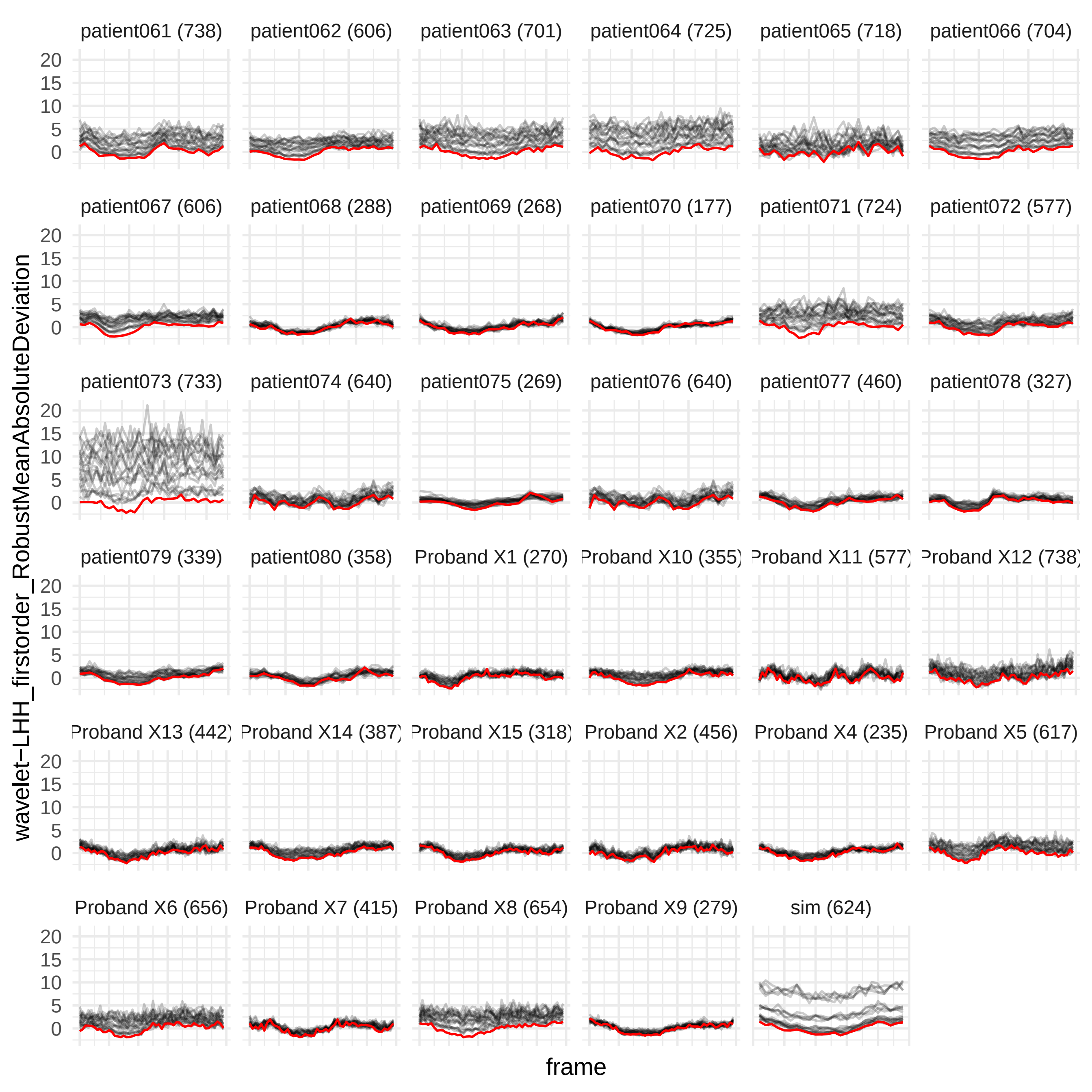

patient061 (738)
patient062 (606)
patient063 (701)
patient064 (725)
patient065 (718)
patient066 (704)
patient067 (606)
patient068 (288)
patient069 (268)
patient070 (177)
patient071 (724)
patient072 (577)
patient073 (733)
patient074 (640)
patient075 (269)
patient076 (640)
patient077 (460)
patient078 (327)
patient079 (339)
patient080 (358)
Proband X1 (270)
Proband X10 (355)
Proband X11 (577)
Proband X12 (738)
Proband X13 (442)
Proband X14 (387)
Proband X15 (318)
Proband X2 (456)
Proband X4 (235)
Proband X5 (617)
Proband X6 (656)
Proband X7 (415)
Proband X8 (654)
Proband X9 (279)
sim (624)
0
5
10
15
20
wavelet−LHH_firstorder_RobustMeanAbsoluteDeviation
frame

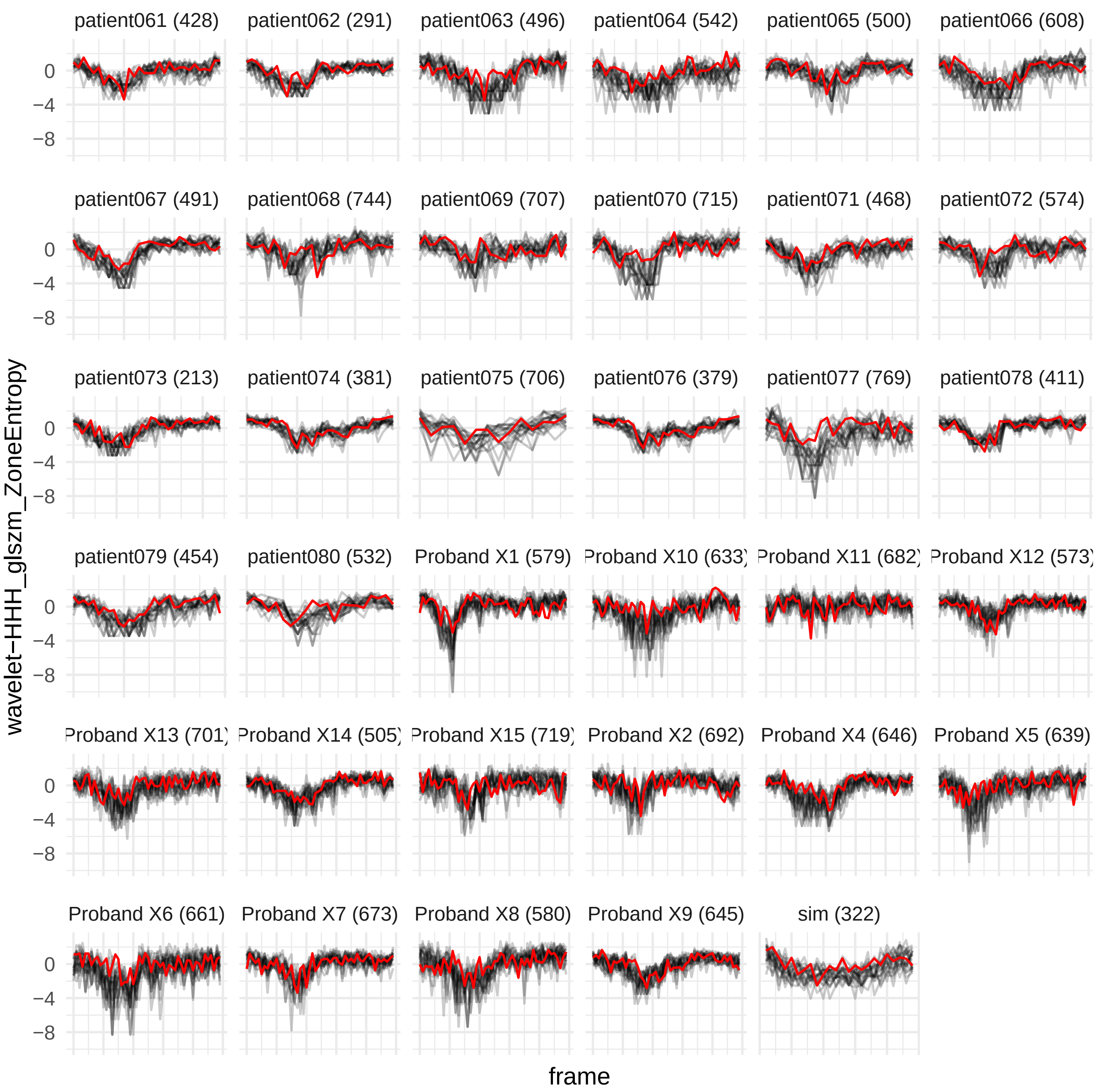
patient061 (428)
patient062 (291)
patient063 (496)
patient064 (542)
patient065 (500)
patient066 (608)
patient067 (491)
patient068 (744)
patient069 (707)
patient070 (715)
patient071 (468)
patient072 (574)
patient073 (213)
patient074 (381)
patient075 (706)
patient076 (379)
patient077 (769)
patient078 (411)
patient079 (454)
patient080 (532)
Proband X1 (579)
Proband X10 (633)
Proband X11 (682)
Proband X12 (573)
Proband X13 (701)
Proband X14 (505)
Proband X15 (719)
Proband X2 (692)
Proband X4 (646)
Proband X5 (639)
Proband X6 (661)
Proband X7 (673)
Proband X8 (580)
Proband X9 (645)
sim (322)
0
−4
−8
wavelet−HHH_glszm_ZoneEntropy
frame

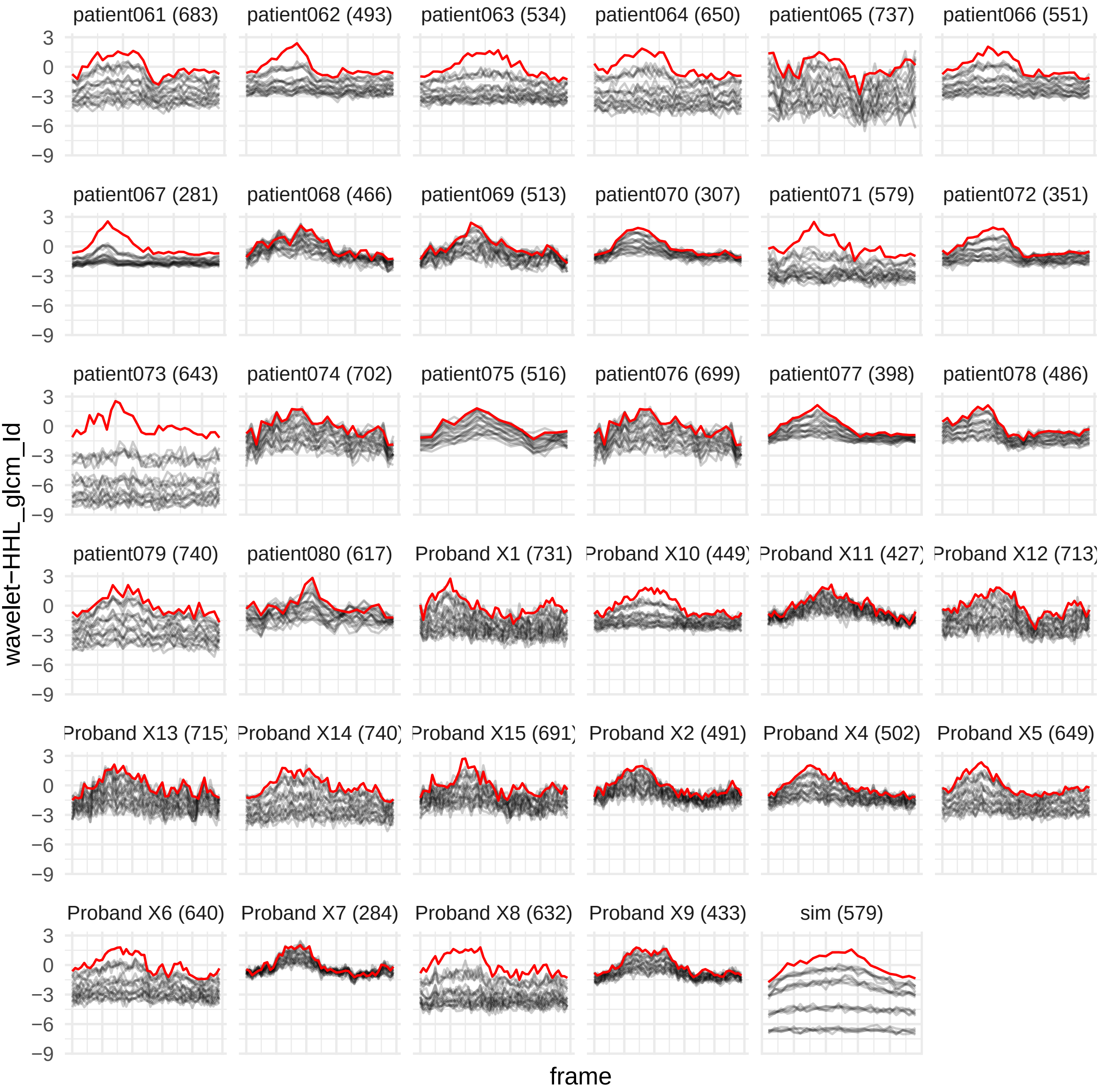
patient061 (683)
patient062 (493)
patient063 (534)
patient064 (650)
patient065 (737)
patient066 (551)
patient067 (281)
patient068 (466)
patient069 (513)
patient070 (307)
patient071 (579)
patient072 (351)
patient073 (643)
patient074 (702)
patient075 (516)
patient076 (699)
patient077 (398)
patient078 (486)
patient079 (740)
patient080 (617)
Proband X1 (731)
Proband X10 (449)
Proband X11 (427)
Proband X12 (713)
Proband X13 (715)
Proband X14 (740)
Proband X15 (691)
Proband X2 (491)
Proband X4 (502)
Proband X5 (649)
Proband X6 (640)
Proband X7 (284)
Proband X8 (632)
Proband X9 (433)
sim (579)
wavelet−HHL_glcm_Id
frame
3
0
−3
−6
−9

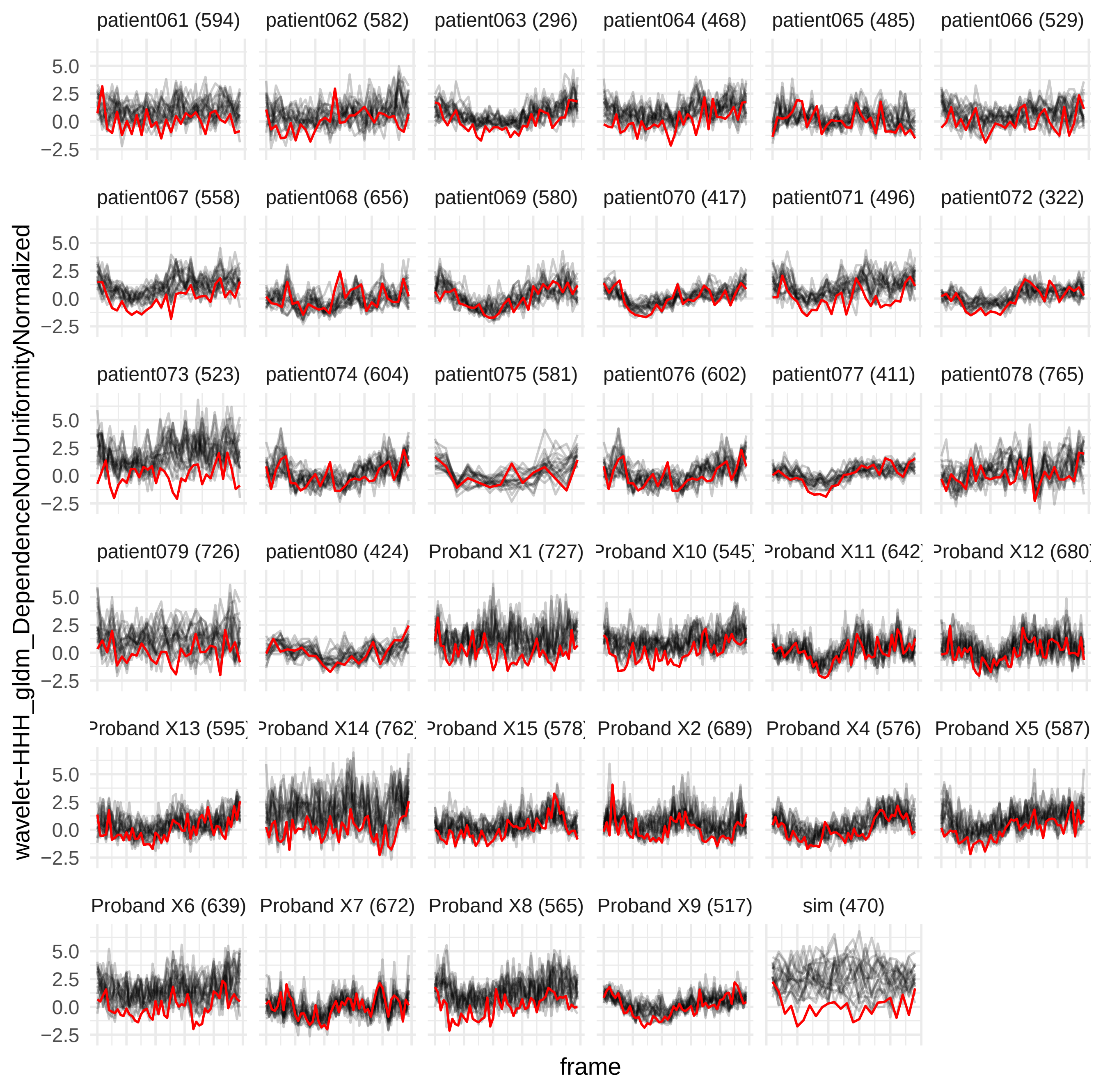
patient061 (594)
patient062 (582)
patient063 (296)
patient064 (468)
patient065 (485)
patient066 (529)
patient067 (558)
patient068 (656)
patient069 (580)
patient070 (417)
patient071 (496)
patient072 (322)
patient073 (523)
patient074 (604)
patient075 (581)
patient076 (602)
patient077 (411)
patient078 (765)
patient079 (726)
patient080 (424)
Proband X1 (727)
Proband X10 (545)
Proband X11 (642)
Proband X12 (680)
Proband X13 (595)
Proband X14 (762)
Proband X15 (578)
Proband X2 (689)
Proband X4 (576)
Proband X5 (587)
Proband X6 (639)
Proband X7 (672)
Proband X8 (565)
Proband X9 (517)
sim (470)
5.0
2.5
0.0
−2.5
wavelet−HHH_gldm_DependenceNonUniformityNormalized
frame

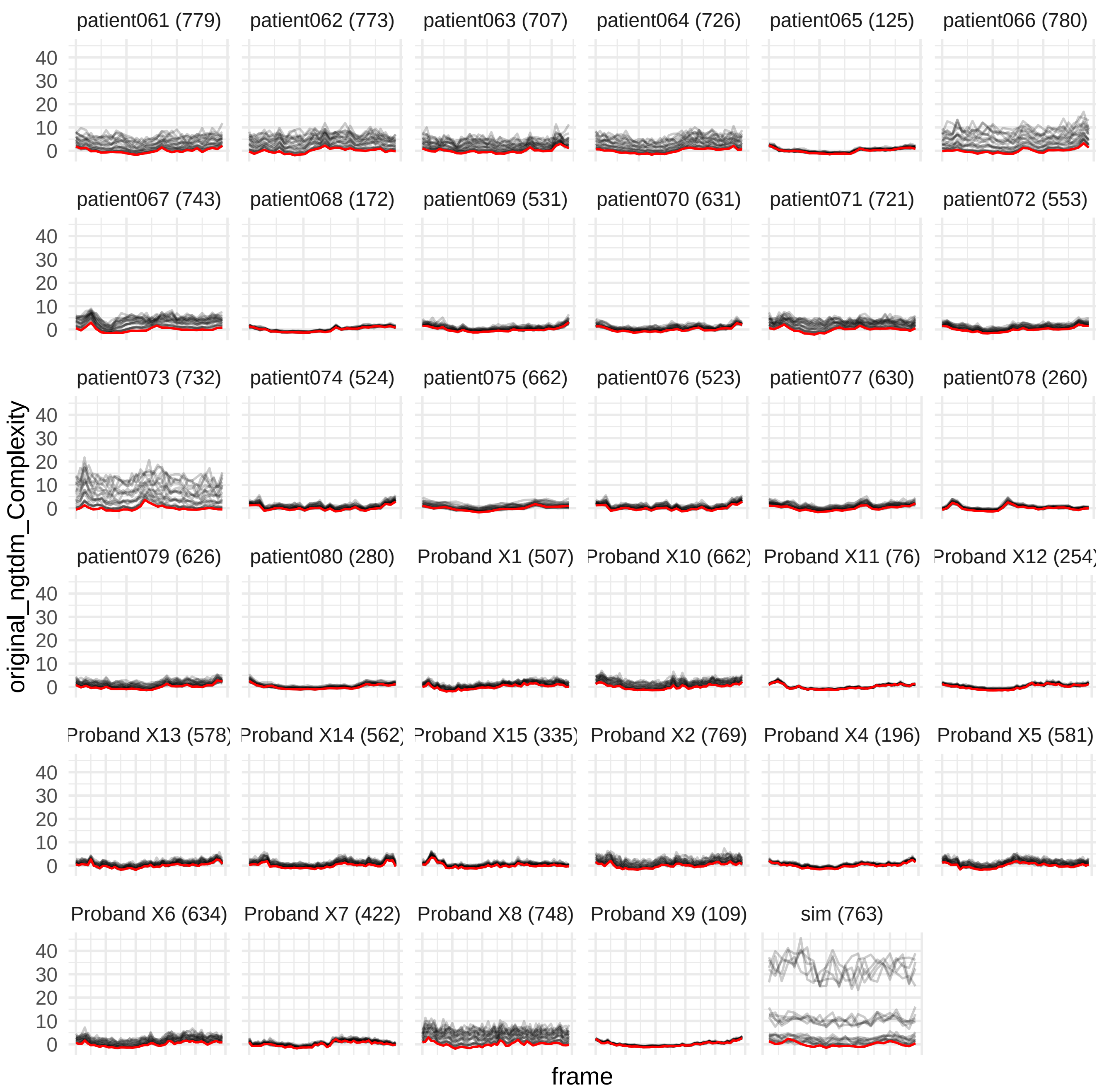
patient061 (779)
patient062 (773)
patient063 (707)
patient064 (726)
patient065 (125)
patient066 (780)
patient067 (743)
patient068 (172)
patient069 (531)
patient070 (631)
patient071 (721)
patient072 (553)
patient073 (732)
patient074 (524)
patient075 (662)
patient076 (523)
patient077 (630)
patient078 (260)
patient079 (626)
patient080 (280)
Proband X1 (507)
Proband X10 (662)
Proband X11 (76)
Proband X12 (254)
Proband X13 (578)
Proband X14 (562)
Proband X15 (335)
Proband X2 (769)
Proband X4 (196)
Proband X5 (581)
Proband X6 (634)
Proband X7 (422)
Proband X8 (748)
Proband X9 (109)
sim (763)
original_ngtdm_Complexity
frame
0
10
20
30
40

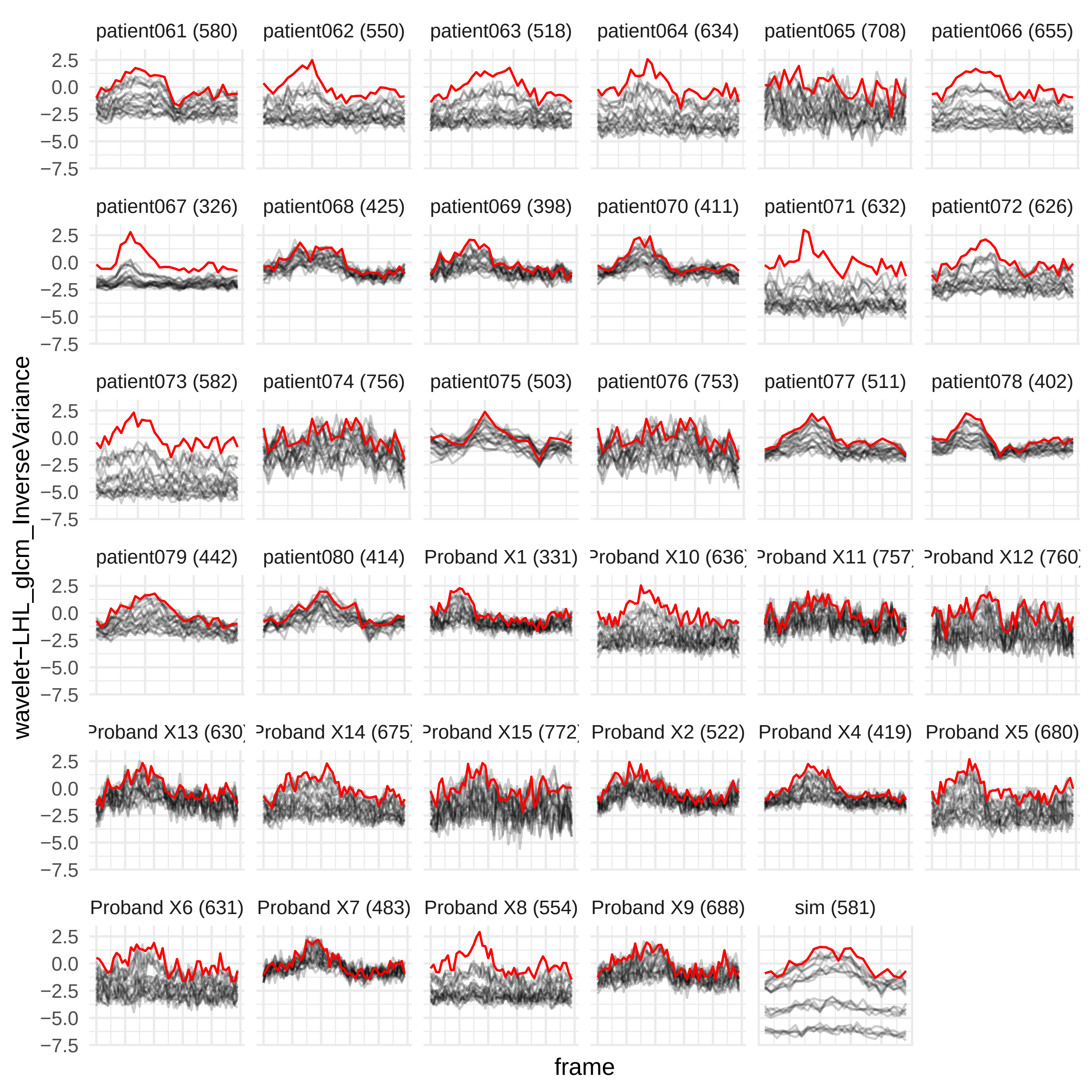

patient061 (580)
patient062 (550)
patient063 (518)
patient064 (634)
patient065 (708)
patient066 (655)
patient067 (326)
patient068 (425)
patient069 (398)
patient070 (411)
patient071 (632)
patient072 (626)
patient073 (582)
patient074 (756)
patient075 (503)
patient076 (753)
patient077 (511)
patient078 (402)
patient079 (442)
patient080 (414)
Proband X1 (331)
Proband X10 (636)
Proband X11 (757)
Proband X12 (760)
Proband X13 (630)
Proband X14 (675)
Proband X15 (772)
Proband X2 (522)
Proband X4 (419)
Proband X5 (680)
Proband X6 (631)
Proband X7 (483)
Proband X8 (554)
Proband X9 (688)
sim (581)
2.5
0.0
−2.5
−5.0
−7.5
wavelet−LHL_glcm_InverseVariance
frame

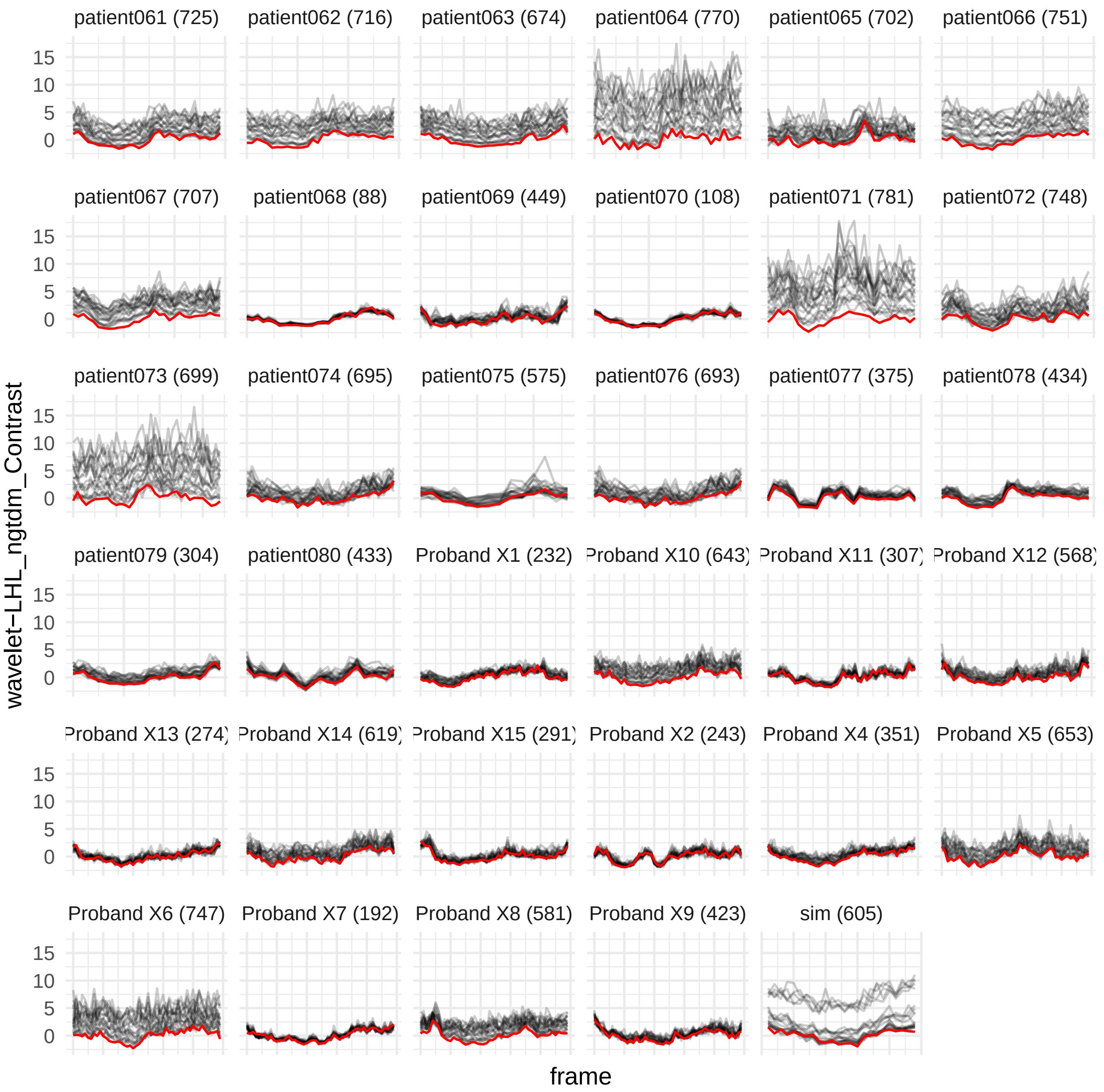
patient061 (725)
patient062 (716)
patient063 (674)
patient064 (770)
patient065 (702)
patient066 (751)
patient067 (707)
patient068 (88)
patient069 (449)
patient070 (108)
patient071 (781)
patient072 (748)
patient073 (699)
patient074 (695)
patient075 (575)
patient076 (693)
patient077 (375)
patient078 (434)
patient079 (304)
patient080 (433)
Proband X1 (232)
Proband X10 (643)
Proband X11 (307)
Proband X12 (568)
Proband X13 (274)
Proband X14 (619)
Proband X15 (291)
Proband X2 (243)
Proband X4 (351)
Proband X5 (653)
Proband X6 (747)
Proband X7 (192)
Proband X8 (581)
Proband X9 (423)
sim (605)
wavelet−LHL_ngtdm_Contrast
frame
0
5
10
15

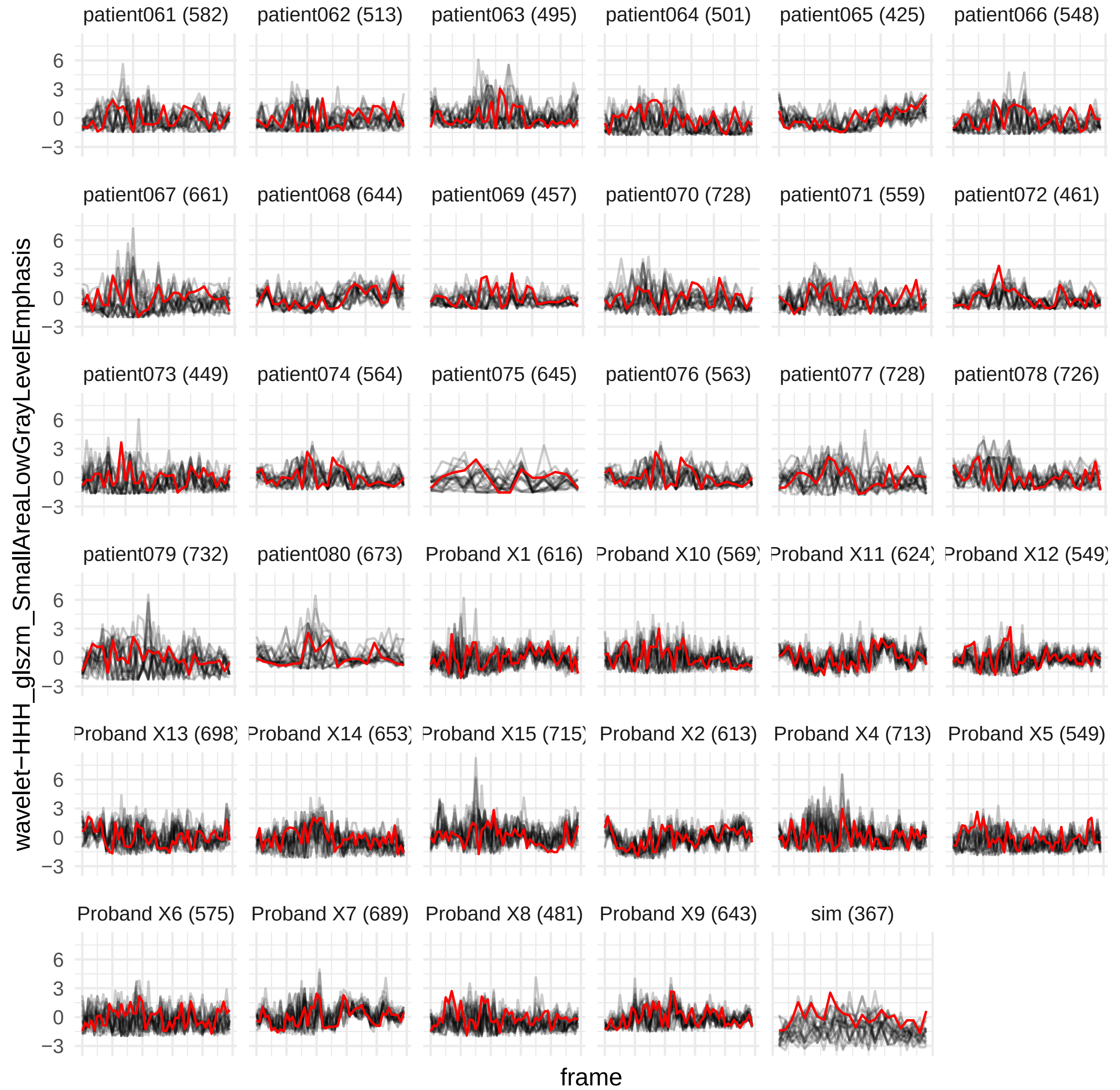

patient061 (582)
patient062 (513)
patient063 (495)
patient064 (501)
patient065 (425)
patient066 (548)
patient067 (661)
patient068 (644)
patient069 (457)
patient070 (728)
patient071 (559)
patient072 (461)
patient073 (449)
patient074 (564)
patient075 (645)
patient076 (563)
patient077 (728)
patient078 (726)
patient079 (732)
patient080 (673)
Proband X1 (616)
Proband X10 (569)
Proband X11 (624)
Proband X12 (549)
Proband X13 (698)
Proband X14 (653)
Proband X15 (715)
Proband X2 (613)
Proband X4 (713)
Proband X5 (549)
Proband X6 (575)
Proband X7 (689)
Proband X8 (481)
Proband X9 (643)
sim (367)
6
3
0
−3
wavelet−HHH_glszm_SmallAreaLowGrayLevelEmphasis
frame

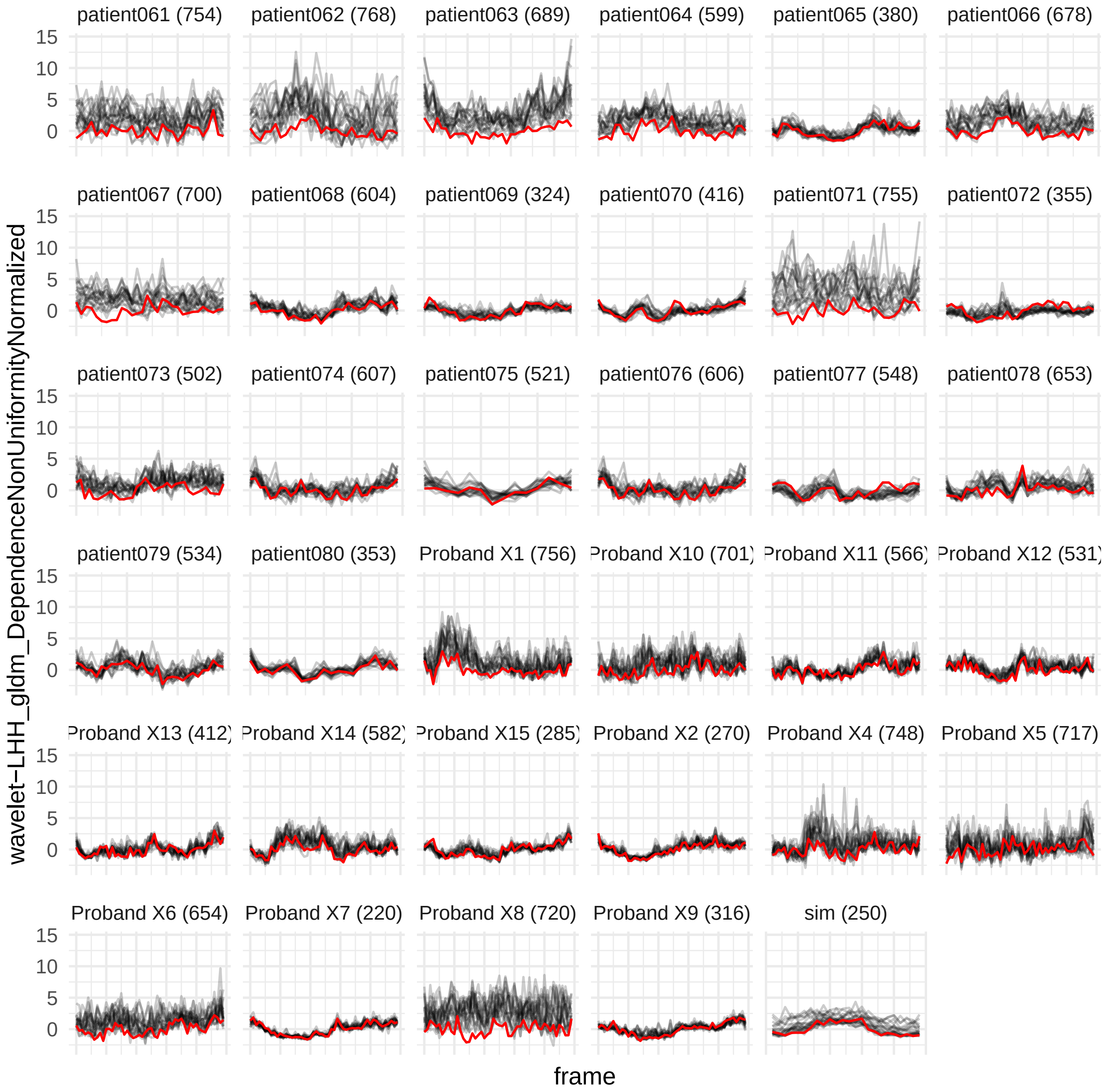
patient061 (754)
patient062 (768)
patient063 (689)
patient064 (599)
patient065 (380)
patient066 (678)
patient067 (700)
patient068 (604)
patient069 (324)
patient070 (416)
patient071 (755)
patient072 (355)
patient073 (502)
patient074 (607)
patient075 (521)
patient076 (606)
patient077 (548)
patient078 (653)
patient079 (534)
patient080 (353)
Proband X1 (756)
Proband X10 (701)
Proband X11 (566)
Proband X12 (531)
Proband X13 (412)
Proband X14 (582)
Proband X15 (285)
Proband X2 (270)
Proband X4 (748)
Proband X5 (717)
Proband X6 (654)
Proband X7 (220)
Proband X8 (720)
Proband X9 (316)
sim (250)
wavelet−LHH_gldm_DependenceNonUniformityNormalized
frame
15
10
5
0

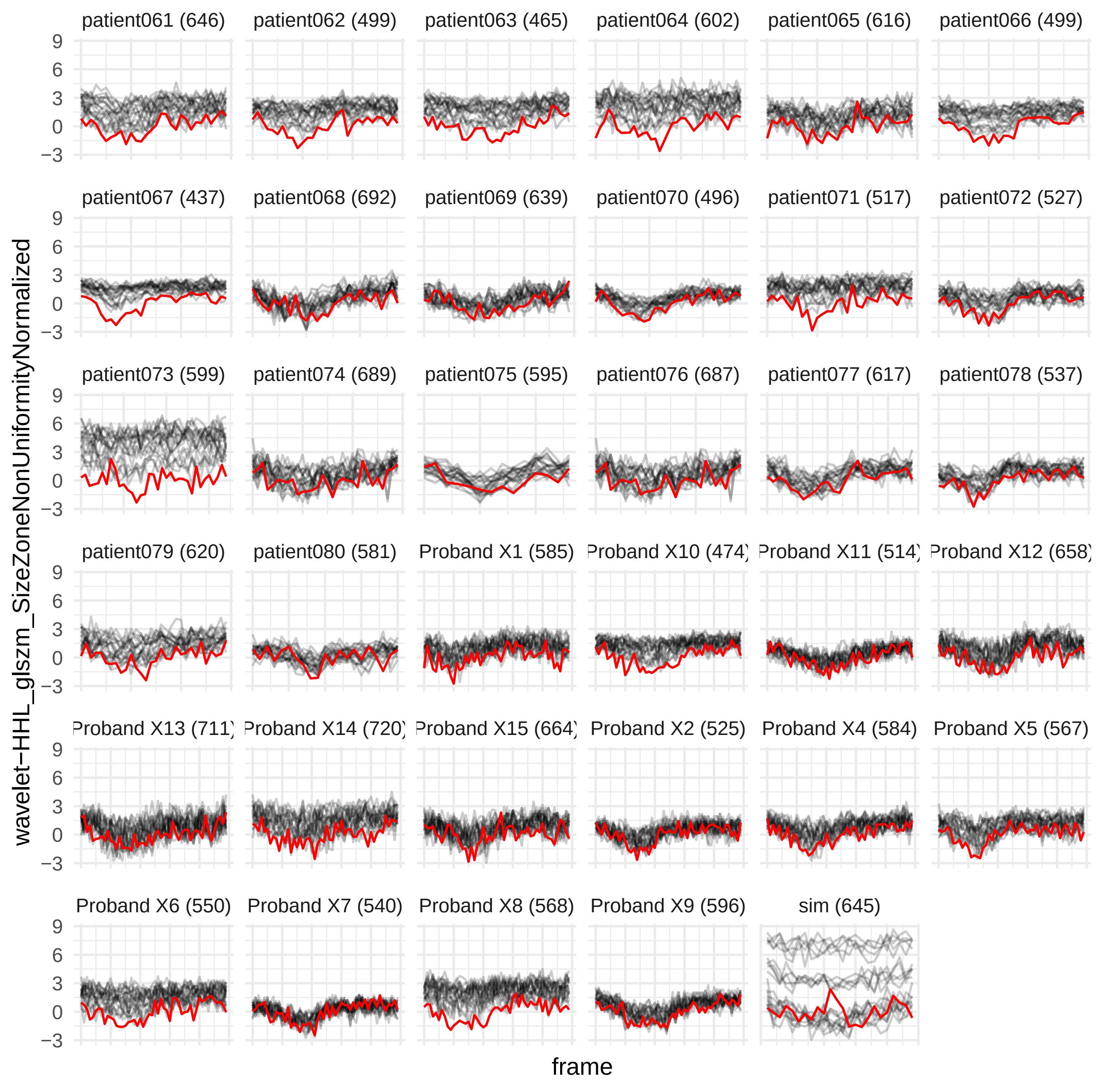
patient061 (646)
patient062 (499)
patient063 (465)
patient064 (602)
patient065 (616)
patient066 (499)
patient067 (437)
patient068 (692)
patient069 (639)
patient070 (496)
patient071 (517)
patient072 (527)
patient073 (599)
patient074 (689)
patient075 (595)
patient076 (687)
patient077 (617)
patient078 (537)
patient079 (620)
patient080 (581)
Proband X1 (585)
Proband X10 (474)
Proband X11 (514)
Proband X12 (658)
Proband X13 (711)
Proband X14 (720)
Proband X15 (664)
Proband X2 (525)
Proband X4 (584)
Proband X5 (567)
Proband X6 (550)
Proband X7 (540)
Proband X8 (568)
Proband X9 (596)
sim (645)
9
6
3
0
−3
wavelet−HHL_glszm_SizeZoneNonUniformityNormalized
frame

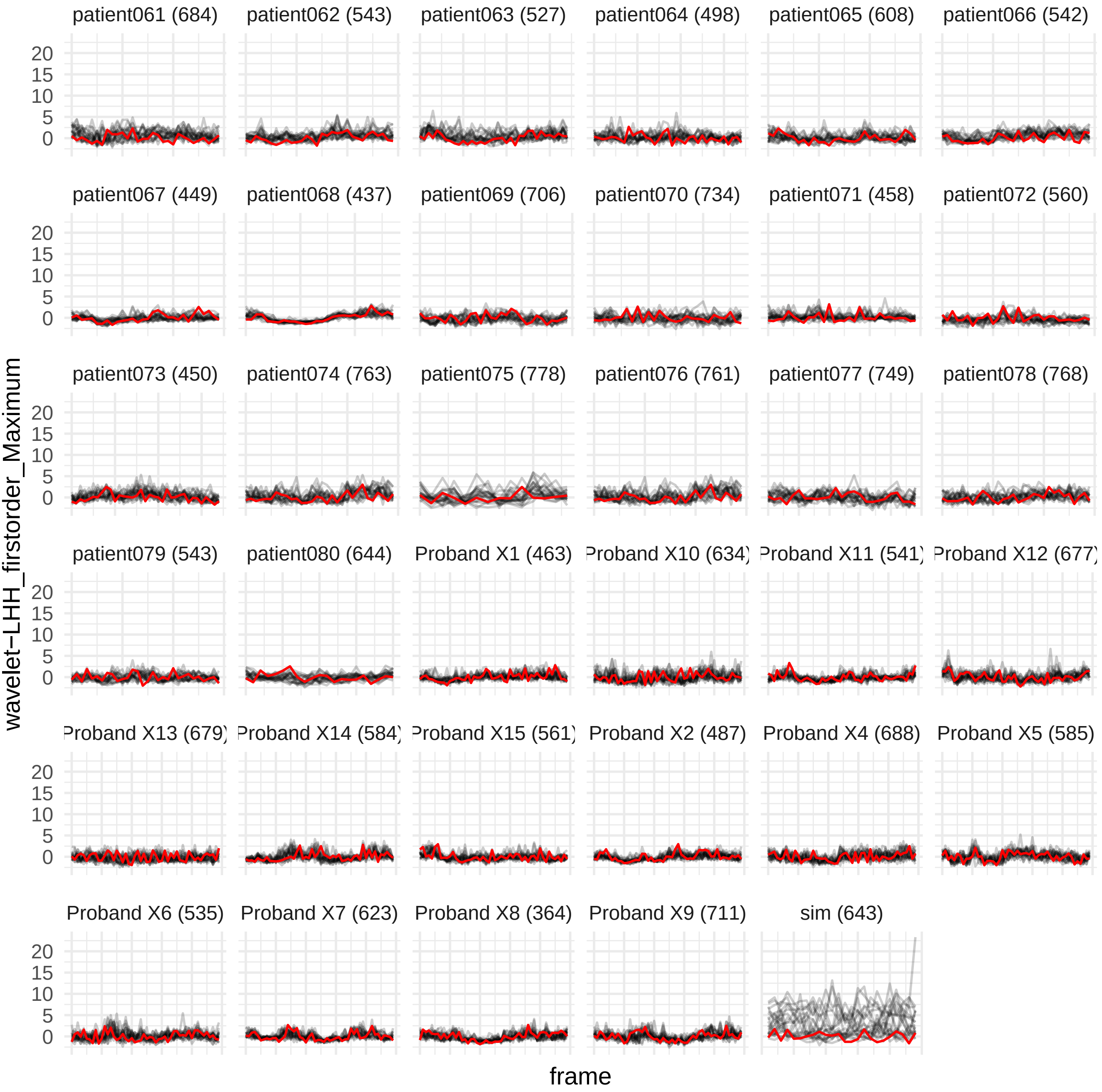

patient061 (684)
patient062 (543)
patient063 (527)
patient064 (498)
patient065 (608)
patient066 (542)
patient067 (449)
patient068 (437)
patient069 (706)
patient070 (734)
patient071 (458)
patient072 (560)
patient073 (450)
patient074 (763)
patient075 (778)
patient076 (761)
patient077 (749)
patient078 (768)
patient079 (543)
patient080 (644)
Proband X1 (463)
Proband X10 (634)
Proband X11 (541)
Proband X12 (677)
Proband X13 (679)
Proband X14 (584)
Proband X15 (561)
Proband X2 (487)
Proband X4 (688)
Proband X5 (585)
Proband X6 (535)
Proband X7 (623)
Proband X8 (364)
Proband X9 (711)
sim (643)
20
15
10
5
0
wavelet−LHH_firstorder_Maximum
frame

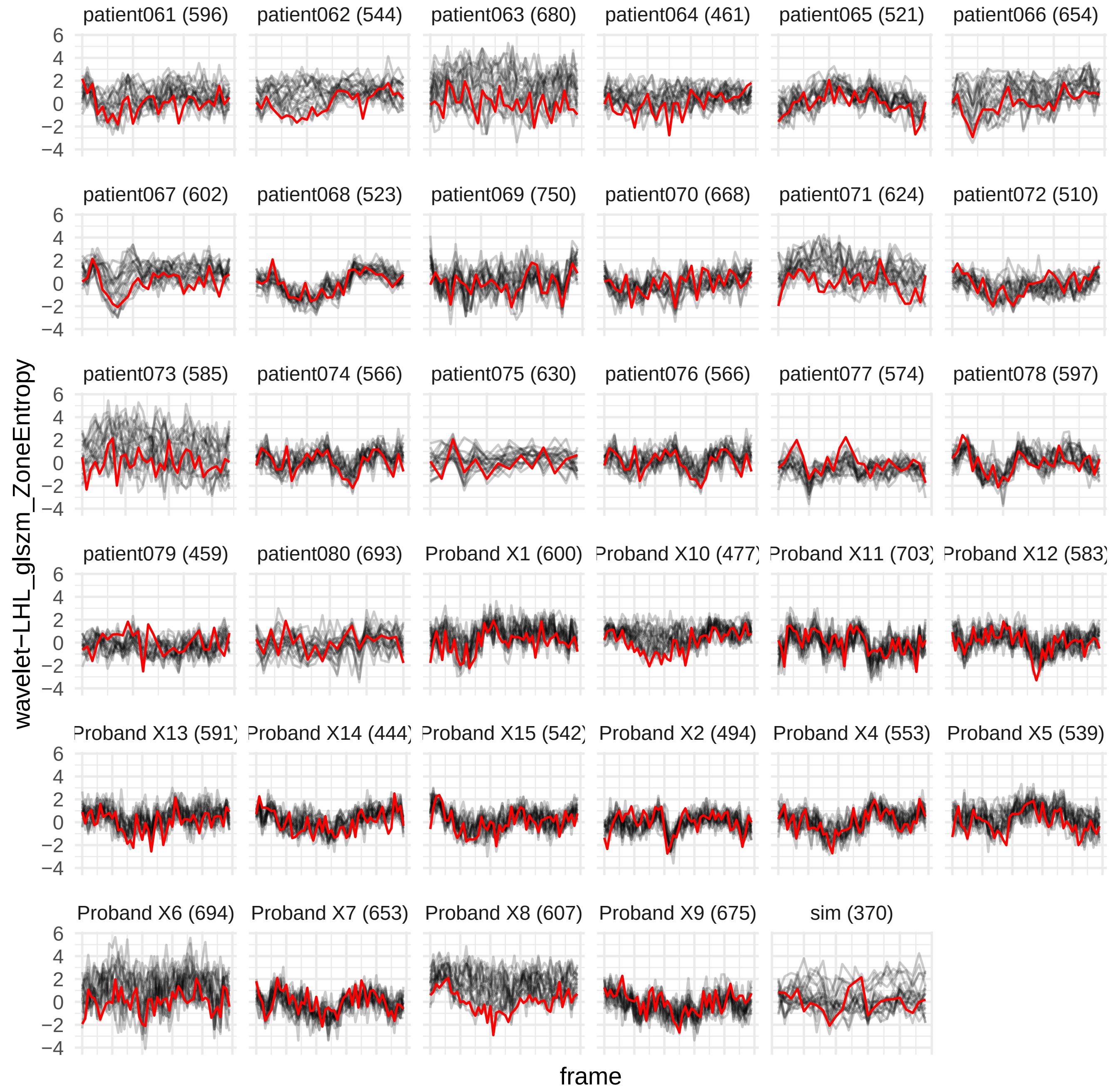
patient061 (596)
patient062 (544)
patient063 (680)
patient064 (461)
patient065 (521)
patient066 (654)
patient067 (602)
patient068 (523)
patient069 (750)
patient070 (668)
patient071 (624)
patient072 (510)
patient073 (585)
patient074 (566)
patient075 (630)
patient076 (566)
patient077 (574)
patient078 (597)
patient079 (459)
patient080 (693)
Proband X1 (600)
Proband X10 (477)
Proband X11 (703)
Proband X12 (583)
Proband X13 (591)
Proband X14 (444)
Proband X15 (542)
Proband X2 (494)
Proband X4 (553)
Proband X5 (539)
Proband X6 (694)
Proband X7 (653)
Proband X8 (607)
Proband X9 (675)
sim (370)
6
4
2
0
−2
−4
wavelet−LHL_glszm_ZoneEntropy
frame

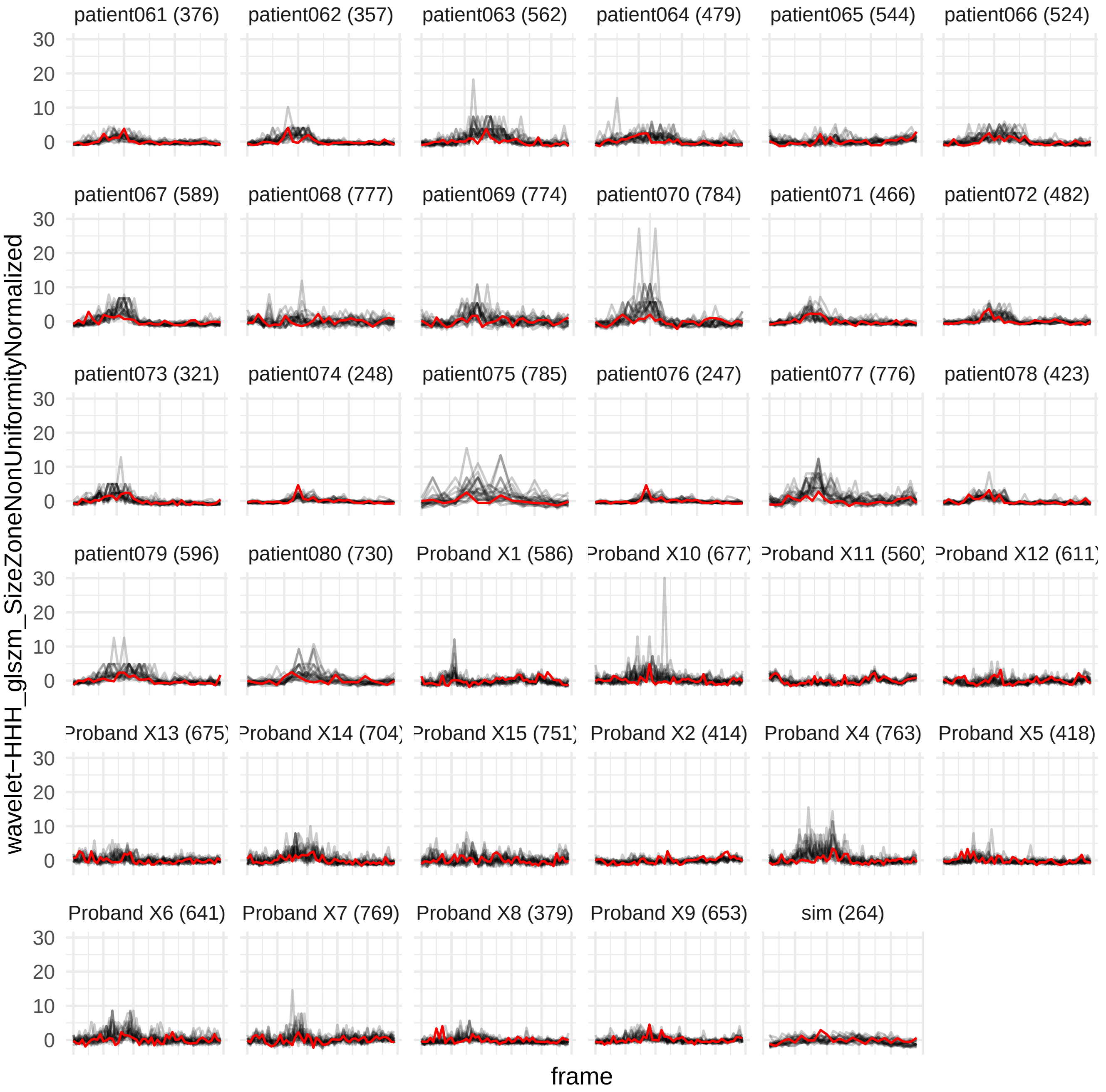
patient061 (376)
patient062 (357)
patient063 (562)
patient064 (479)
patient065 (544)
patient066 (524)
patient067 (589)
patient068 (777)
patient069 (774)
patient070 (784)
patient071 (466)
patient072 (482)
patient073 (321)
patient074 (248)
patient075 (785)
patient076 (247)
patient077 (776)
patient078 (423)
patient079 (596)
patient080 (730)
Proband X1 (586)
Proband X10 (677)
Proband X11 (560)
Proband X12 (611)
Proband X13 (675)
Proband X14 (704)
Proband X15 (751)
Proband X2 (414)
Proband X4 (763)
Proband X5 (418)
Proband X6 (641)
Proband X7 (769)
Proband X8 (379)
Proband X9 (653)
sim (264)
0
10
20
30
wavelet−HHH_glszm_SizeZoneNonUniformityNormalized
frame

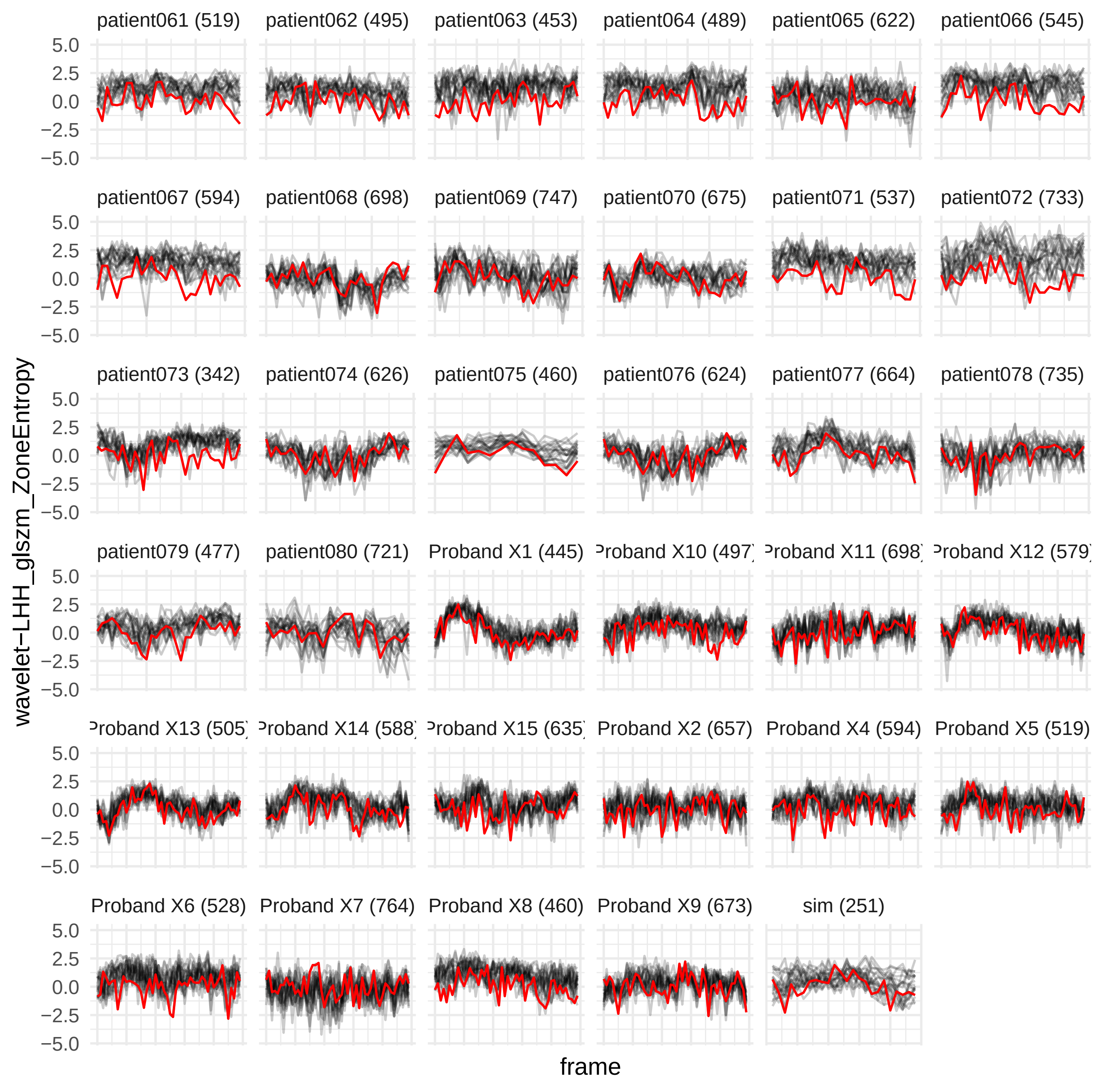
patient061 (519)
patient062 (495)
patient063 (453)
patient064 (489)
patient065 (622)
patient066 (545)
patient067 (594)
patient068 (698)
patient069 (747)
patient070 (675)
patient071 (537)
patient072 (733)
patient073 (342)
patient074 (626)
patient075 (460)
patient076 (624)
patient077 (664)
patient078 (735)
patient079 (477)
patient080 (721)
Proband X1 (445)
Proband X10 (497)
Proband X11 (698)
Proband X12 (579)
Proband X13 (505)
Proband X14 (588)
Proband X15 (635)
Proband X2 (657)
Proband X4 (594)
Proband X5 (519)
Proband X6 (528)
Proband X7 (764)
Proband X8 (460)
Proband X9 (673)
sim (251)
5.0
2.5
0.0
−2.5
−5.0
wavelet−LHH_glszm_ZoneEntropy
frame

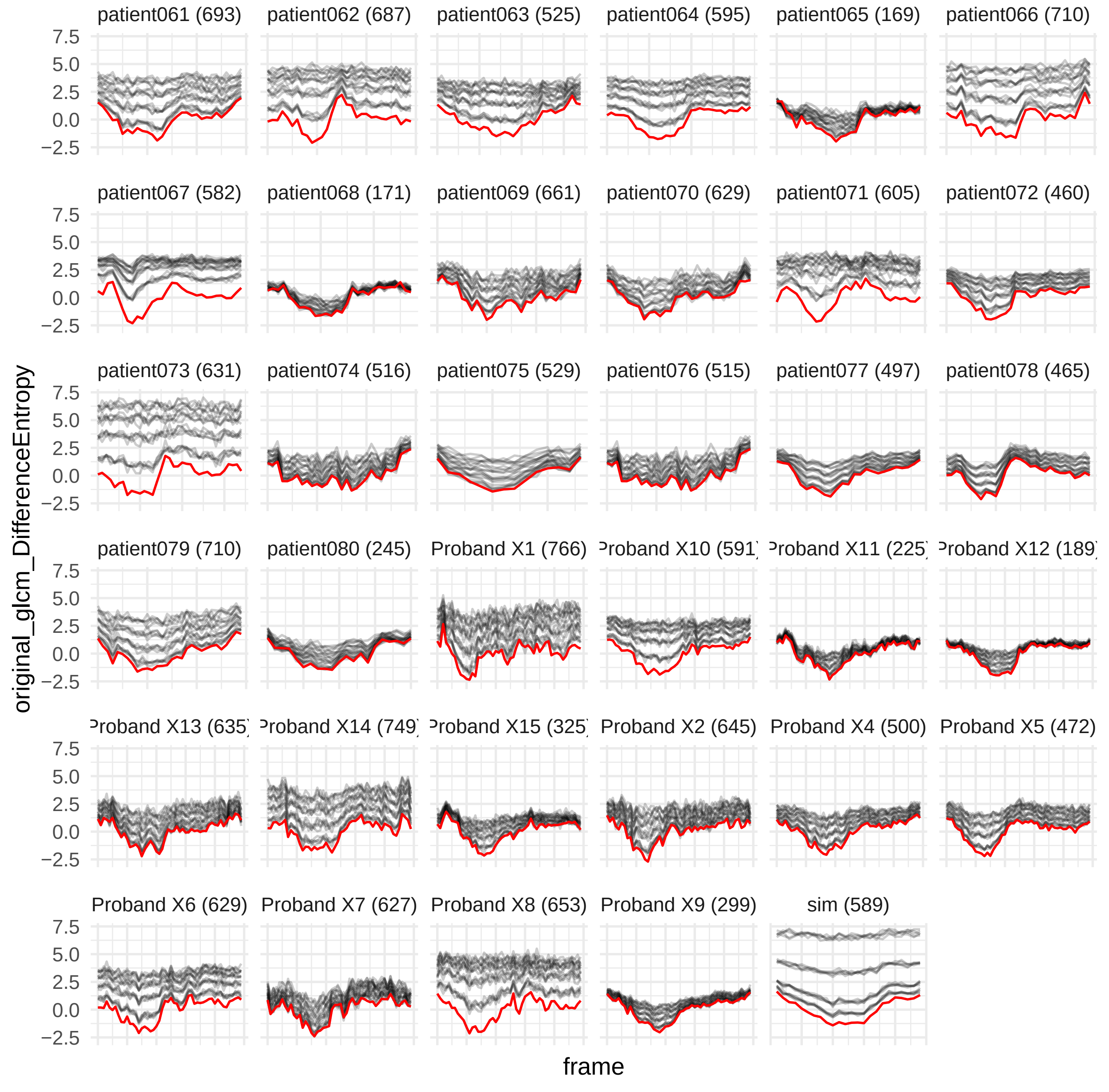

patient061 (693)
patient062 (687)
patient063 (525)
patient064 (595)
patient065 (169)
patient066 (710)
patient067 (582)
patient068 (171)
patient069 (661)
patient070 (629)
patient071 (605)
patient072 (460)
patient073 (631)
patient074 (516)
patient075 (529)
patient076 (515)
patient077 (497)
patient078 (465)
patient079 (710)
patient080 (245)
Proband X1 (766)
Proband X10 (591)
Proband X11 (225)
Proband X12 (189)
Proband X13 (635)
Proband X14 (749)
Proband X15 (325)
Proband X2 (645)
Proband X4 (500)
Proband X5 (472)
Proband X6 (629)
Proband X7 (627)
Proband X8 (653)
Proband X9 (299)
sim (589)
7.5
5.0
2.5
0.0
−2.5
original_glcm_DifferenceEntropy
frame

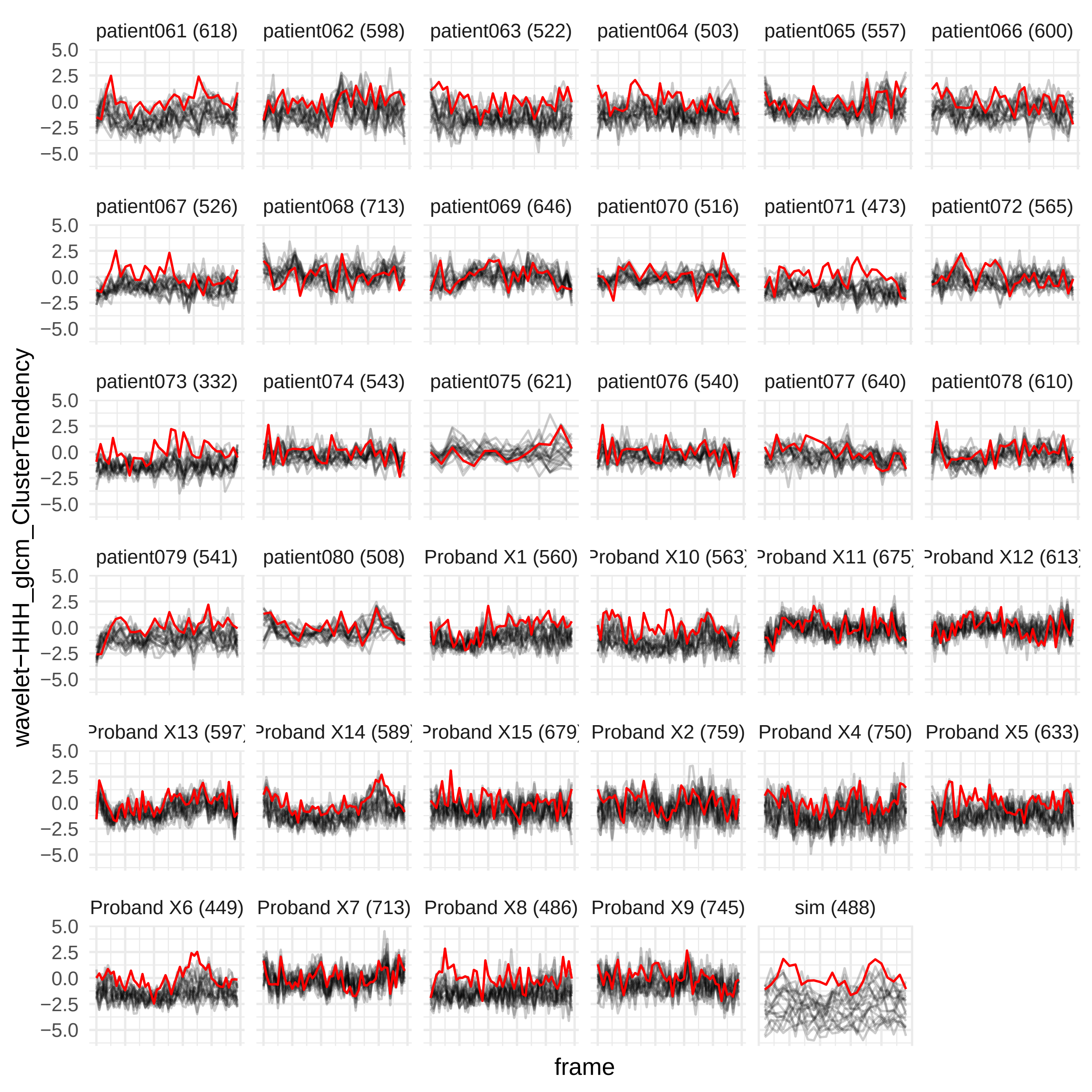

patient061 (618)
patient062 (598)
patient063 (522)
patient064 (503)
patient065 (557)
patient066 (600)
patient067 (526)
patient068 (713)
patient069 (646)
patient070 (516)
patient071 (473)
patient072 (565)
patient073 (332)
patient074 (543)
patient075 (621)
patient076 (540)
patient077 (640)
patient078 (610)
patient079 (541)
patient080 (508)
Proband X1 (560)
Proband X10 (563)
Proband X11 (675)
Proband X12 (613)
Proband X13 (597)
Proband X14 (589)
Proband X15 (679)
Proband X2 (759)
Proband X4 (750)
Proband X5 (633)
Proband X6 (449)
Proband X7 (713)
Proband X8 (486)
Proband X9 (745)
sim (488)
5.0
2.5
0.0
−2.5
−5.0
wavelet−HHH_glcm_ClusterTendency
frame

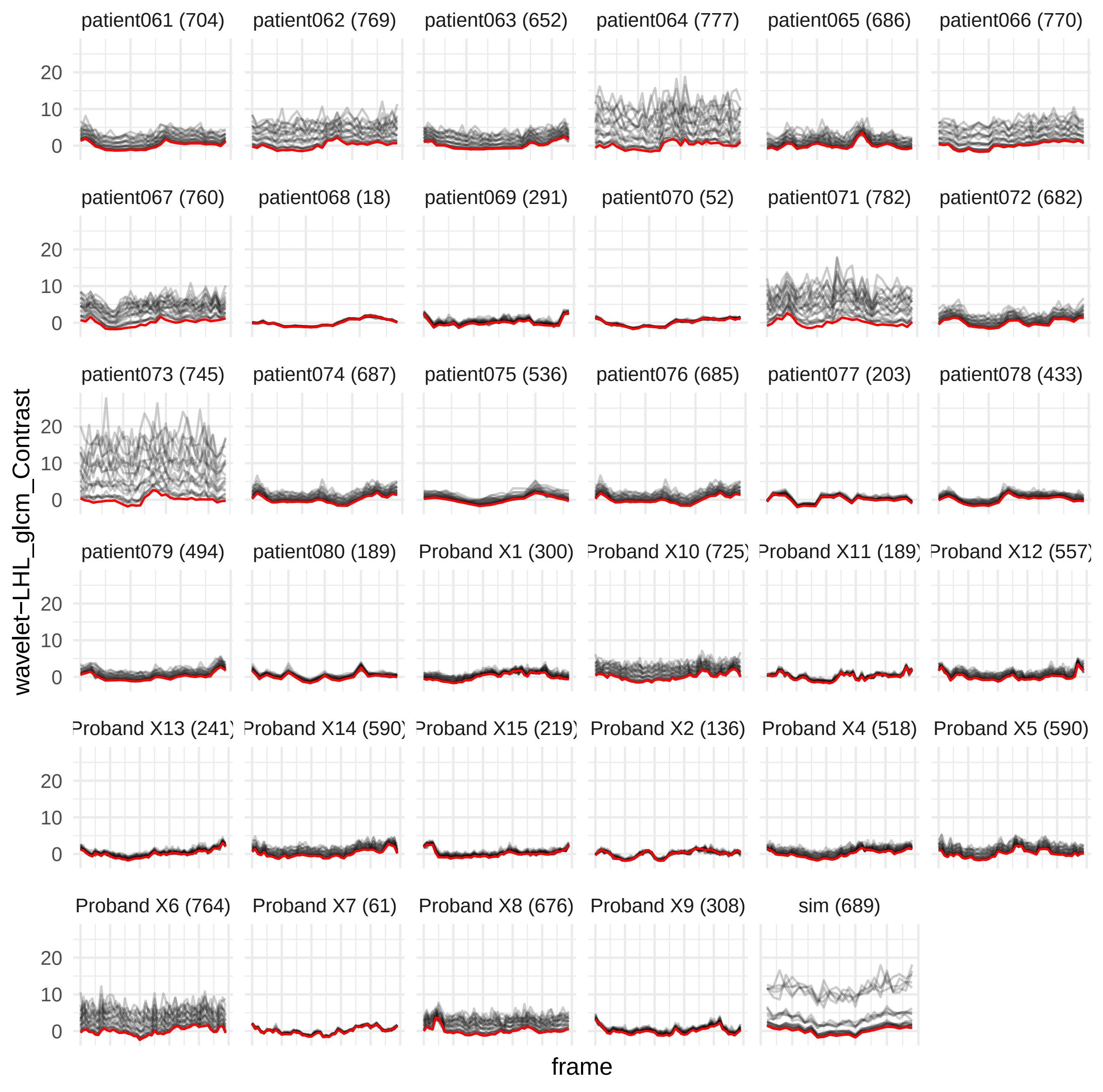
patient061 (704)
patient062 (769)
patient063 (652)
patient064 (777)
patient065 (686)
patient066 (770)
patient067 (760)
patient068 (18)
patient069 (291)
patient070 (52)
patient071 (782)
patient072 (682)
patient073 (745)
patient074 (687)
patient075 (536)
patient076 (685)
patient077 (203)
patient078 (433)
patient079 (494)
patient080 (189)
Proband X1 (300)
Proband X10 (725)
Proband X11 (189)
Proband X12 (557)
Proband X13 (241)
Proband X14 (590)
Proband X15 (219)
Proband X2 (136)
Proband X4 (518)
Proband X5 (590)
Proband X6 (764)
Proband X7 (61)
Proband X8 (676)
Proband X9 (308)
sim (689)
20
10
0
wavelet−LHL_glcm_Contrast
frame

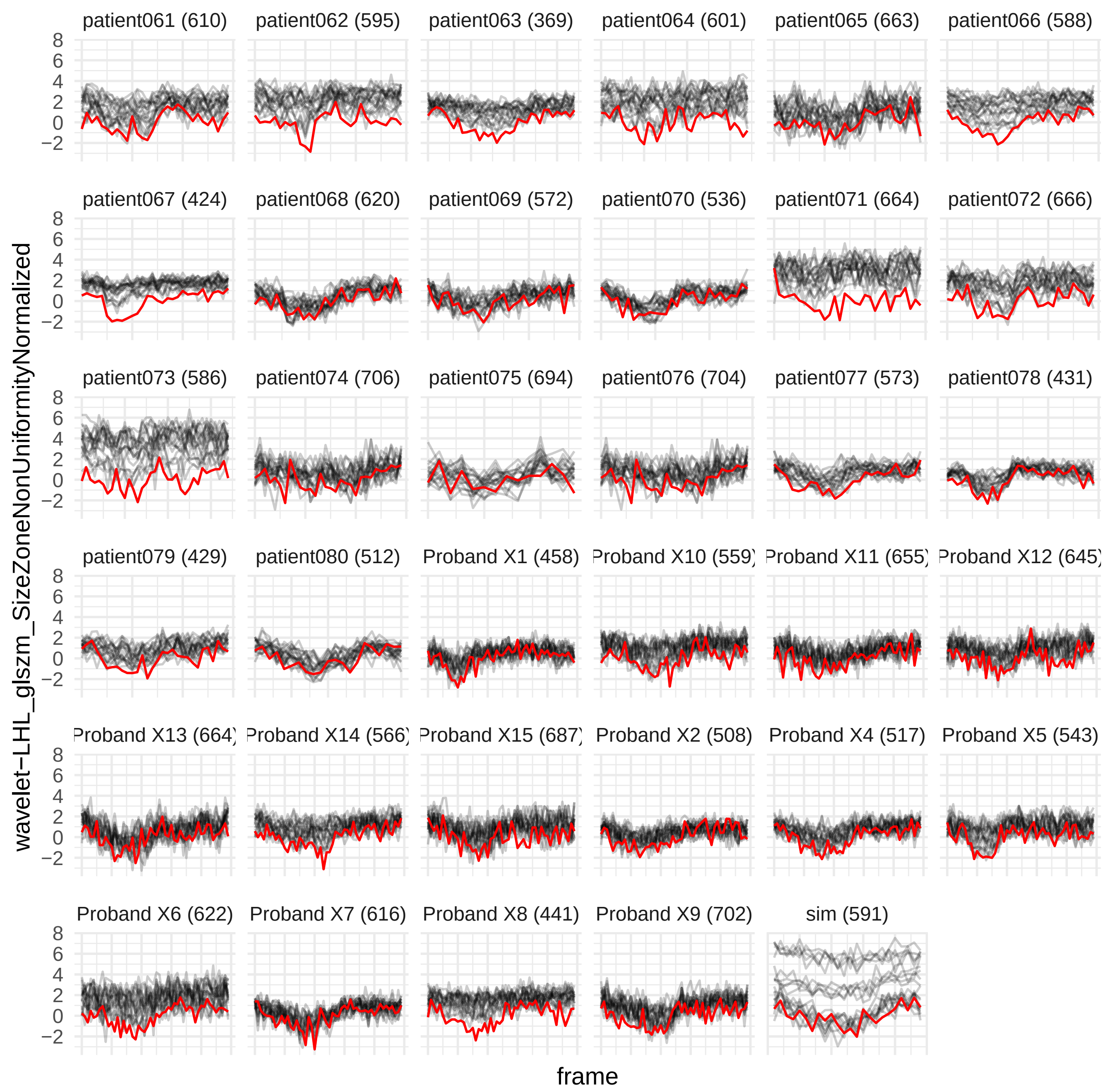

patient061 (610)
patient062 (595)
patient063 (369)
patient064 (601)
patient065 (663)
patient066 (588)
patient067 (424)
patient068 (620)
patient069 (572)
patient070 (536)
patient071 (664)
patient072 (666)
patient073 (586)
patient074 (706)
patient075 (694)
patient076 (704)
patient077 (573)
patient078 (431)
patient079 (429)
patient080 (512)
Proband X1 (458)
Proband X10 (559)
Proband X11 (655)
Proband X12 (645)
Proband X13 (664)
Proband X14 (566)
Proband X15 (687)
Proband X2 (508)
Proband X4 (517)
Proband X5 (543)
Proband X6 (622)
Proband X7 (616)
Proband X8 (441)
Proband X9 (702)
sim (591)
wavelet−LHL_glszm_SizeZoneNonUniformityNormalized
frame
8
6
4
2
0
−2

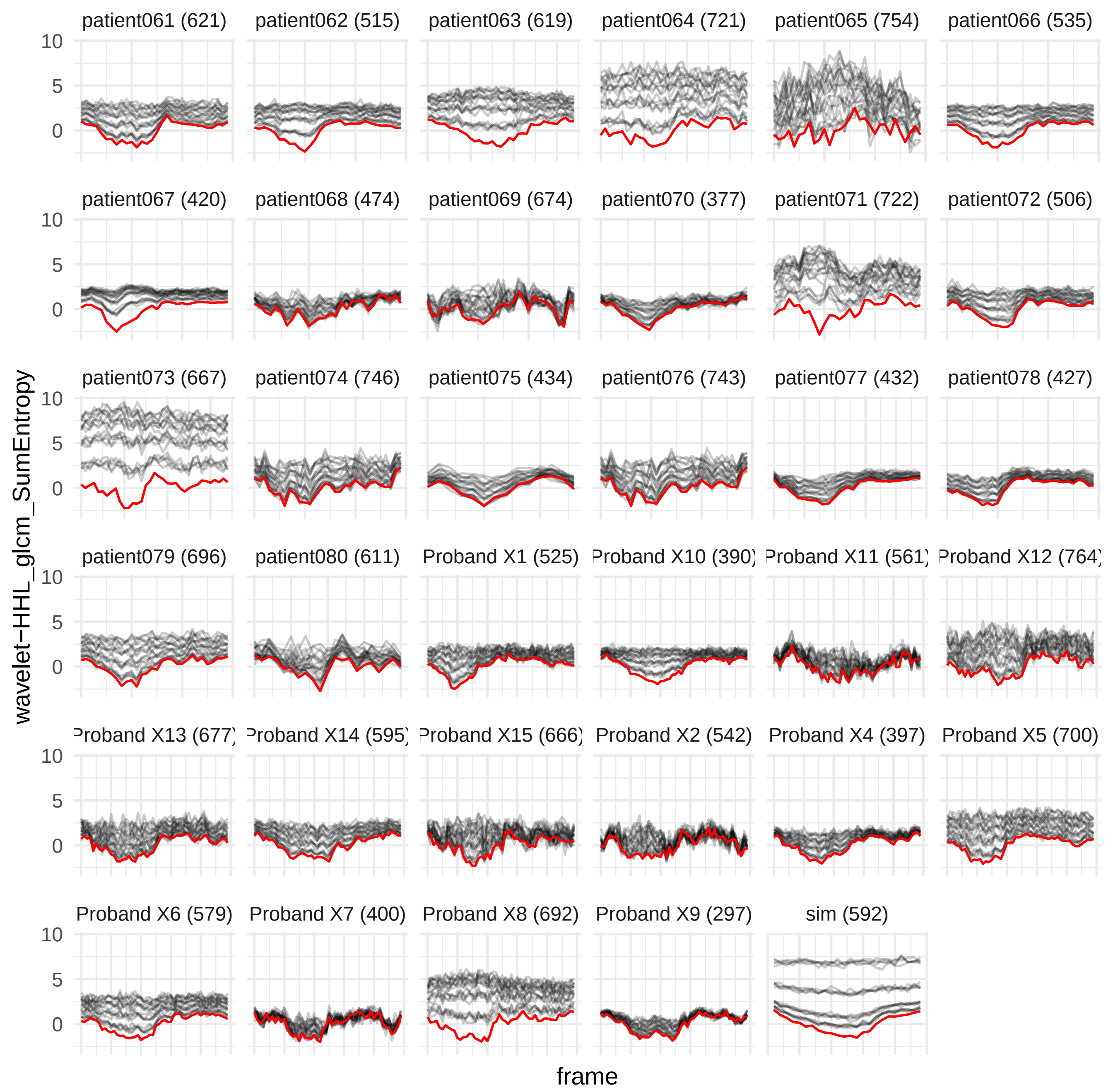

patient061 (621)
patient062 (515)
patient063 (619)
patient064 (721)
patient065 (754)
patient066 (535)
patient067 (420)
patient068 (474)
patient069 (674)
patient070 (377)
patient071 (722)
patient072 (506)
patient073 (667)
patient074 (746)
patient075 (434)
patient076 (743)
patient077 (432)
patient078 (427)
patient079 (696)
patient080 (611)
Proband X1 (525)
Proband X10 (390)
Proband X11 (561)
Proband X12 (764)
Proband X13 (677)
Proband X14 (595)
Proband X15 (666)
Proband X2 (542)
Proband X4 (397)
Proband X5 (700)
Proband X6 (579)
Proband X7 (400)
Proband X8 (692)
Proband X9 (297)
sim (592)
10
5
0
wavelet−HHL_glcm_SumEntropy
frame

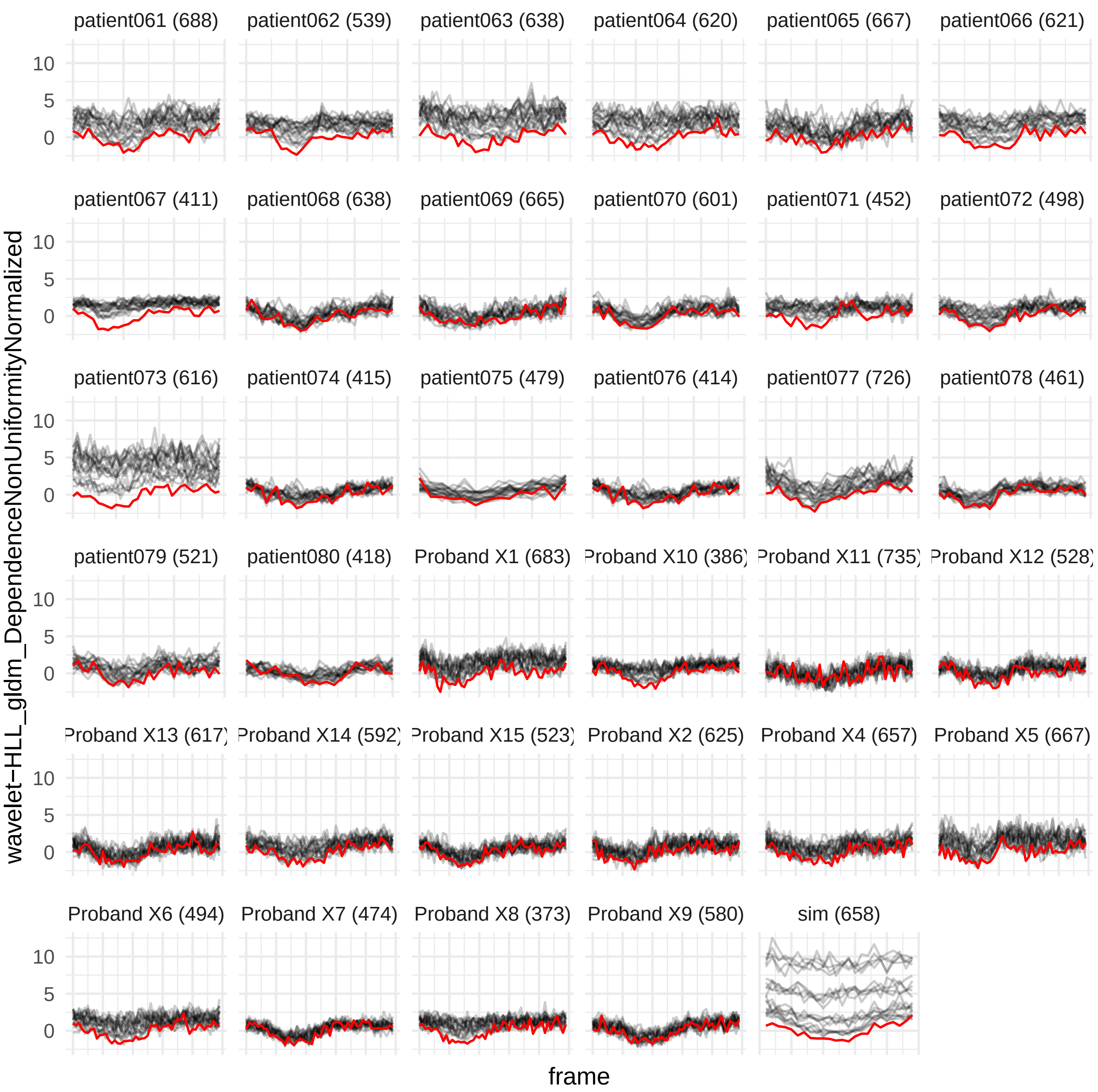
patient061 (688)
patient062 (539)
patient063 (638)
patient064 (620)
patient065 (667)
patient066 (621)
patient067 (411)
patient068 (638)
patient069 (665)
patient070 (601)
patient071 (452)
patient072 (498)
patient073 (616)
patient074 (415)
patient075 (479)
patient076 (414)
patient077 (726)
patient078 (461)
patient079 (521)
patient080 (418)
Proband X1 (683)
Proband X10 (386)
Proband X11 (735)
Proband X12 (528)
Proband X13 (617)
Proband X14 (592)
Proband X15 (523)
Proband X2 (625)
Proband X4 (657)
Proband X5 (667)
Proband X6 (494)
Proband X7 (474)
Proband X8 (373)
Proband X9 (580)
sim (658)
10
5
0
wavelet−HLL_gldm_DependenceNonUniformityNormalized
frame

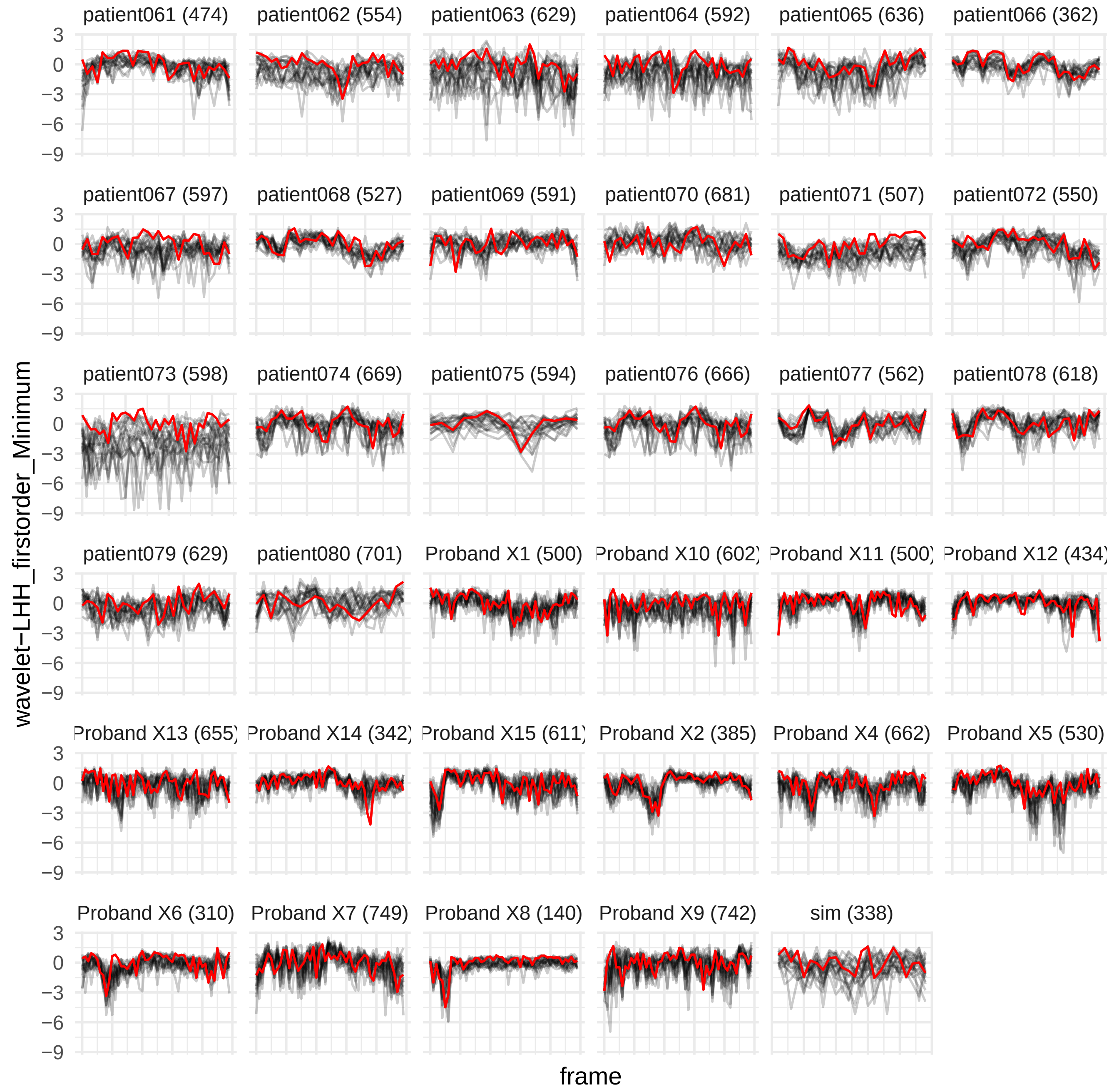

patient061 (474)
patient062 (554)
patient063 (629)
patient064 (592)
patient065 (636)
patient066 (362)
patient067 (597)
patient068 (527)
patient069 (591)
patient070 (681)
patient071 (507)
patient072 (550)
patient073 (598)
patient074 (669)
patient075 (594)
patient076 (666)
patient077 (562)
patient078 (618)
patient079 (629)
patient080 (701)
Proband X1 (500)
Proband X10 (602)
Proband X11 (500)
Proband X12 (434)
Proband X13 (655)
Proband X14 (342)
Proband X15 (611)
Proband X2 (385)
Proband X4 (662)
Proband X5 (530)
Proband X6 (310)
Proband X7 (749)
Proband X8 (140)
Proband X9 (742)
sim (338)
3
0
−3
−6
−9
wavelet−LHH_firstorder_Minimum
frame

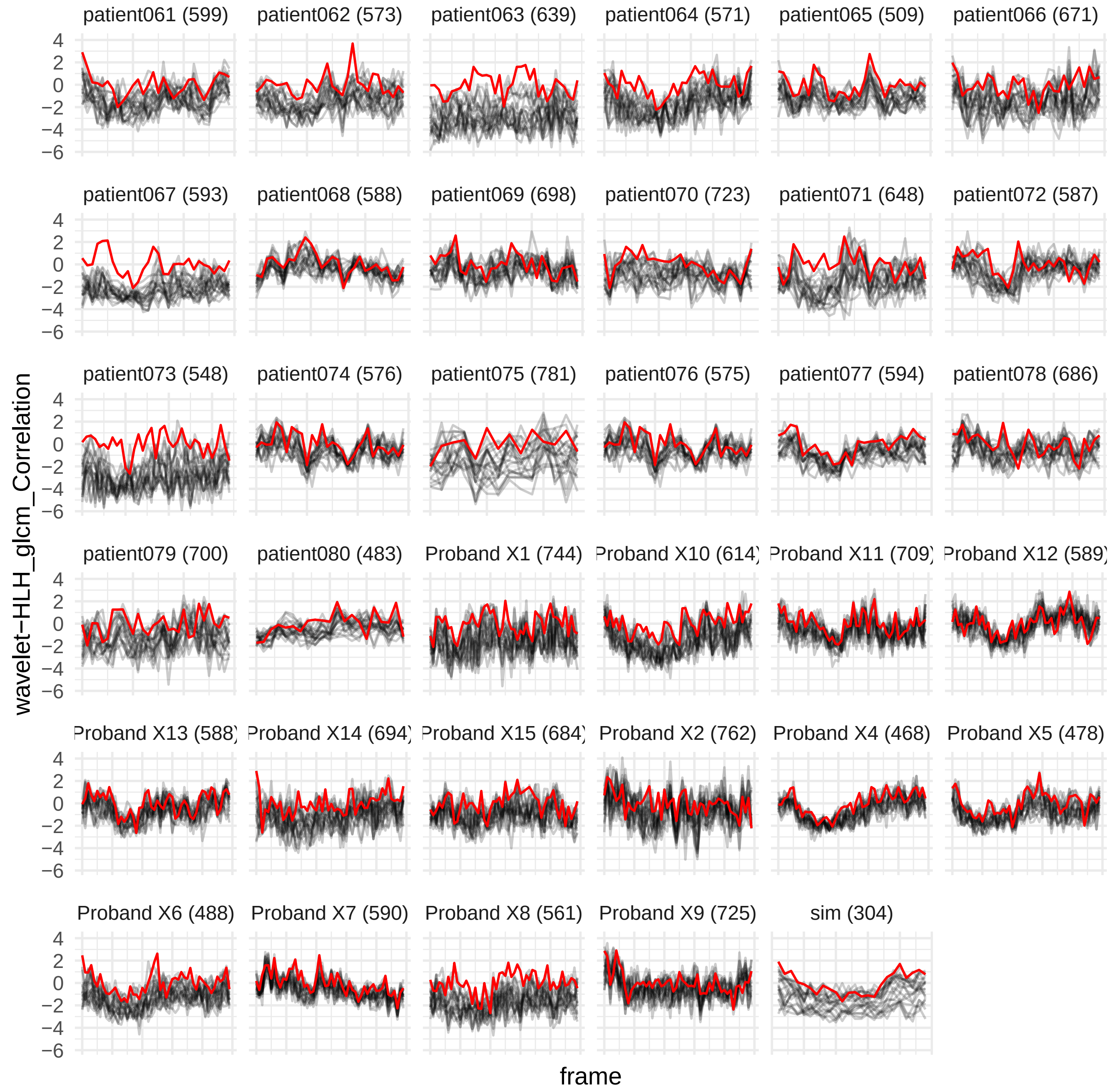
patient061 (599)
patient062 (573)
patient063 (639)
patient064 (571)
patient065 (509)
patient066 (671)
patient067 (593)
patient068 (588)
patient069 (698)
patient070 (723)
patient071 (648)
patient072 (587)
patient073 (548)
patient074 (576)
patient075 (781)
patient076 (575)
patient077 (594)
patient078 (686)
patient079 (700)
patient080 (483)
Proband X1 (744)
Proband X10 (614)
Proband X11 (709)
Proband X12 (589)
Proband X13 (588)
Proband X14 (694)
Proband X15 (684)
Proband X2 (762)
Proband X4 (468)
Proband X5 (478)
Proband X6 (488)
Proband X7 (590)
Proband X8 (561)
Proband X9 (725)
sim (304)
wavelet−HLH_glcm_Correlation
frame

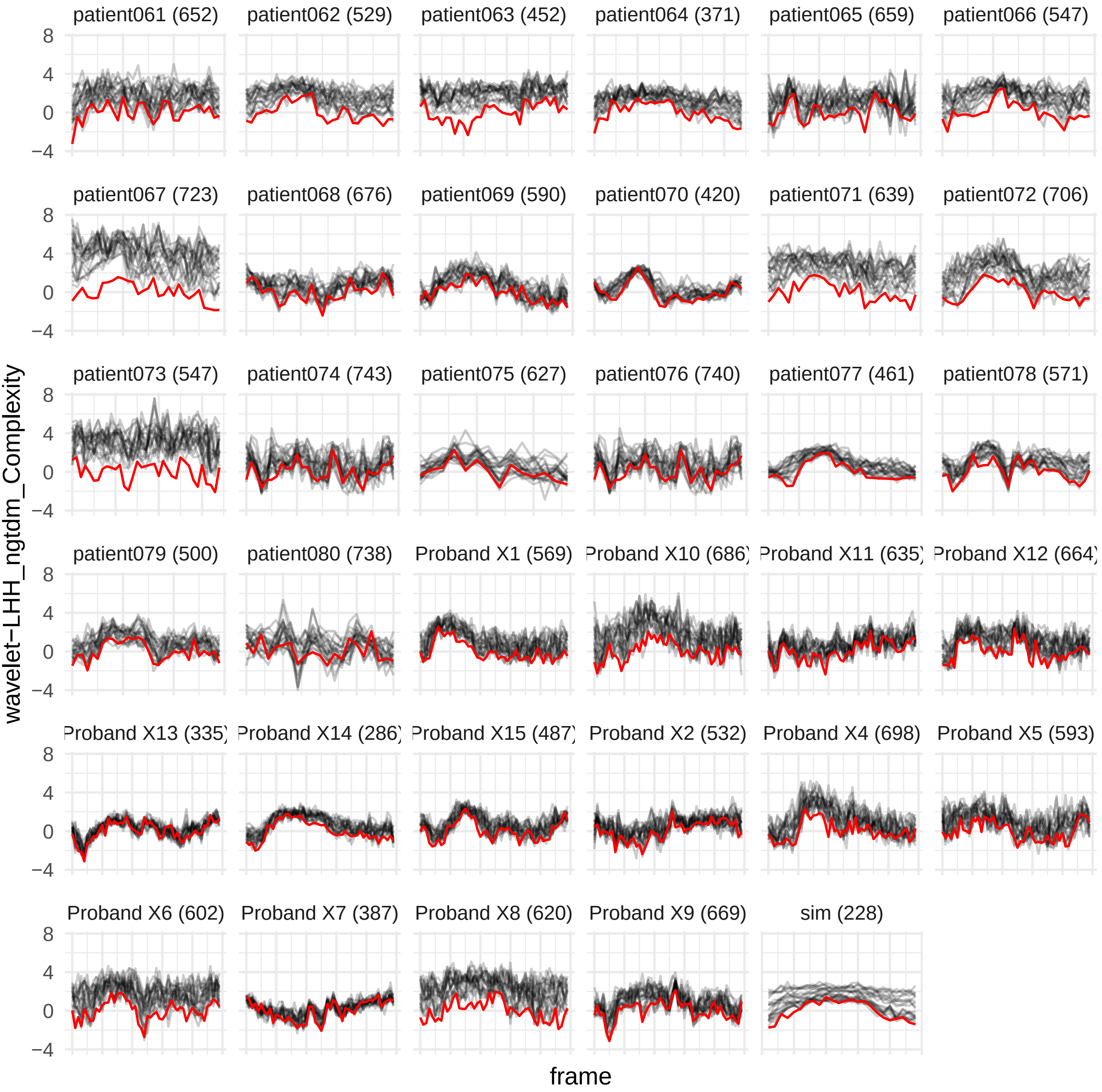
patient061 (652)
patient062 (529)
patient063 (452)
patient064 (371)
patient065 (659)
patient066 (547)
patient067 (723)
patient068 (676)
patient069 (590)
patient070 (420)
patient071 (639)
patient072 (706)
patient073 (547)
patient074 (743)
patient075 (627)
patient076 (740)
patient077 (461)
patient078 (571)
patient079 (500)
patient080 (738)
Proband X1 (569)
Proband X10 (686)
Proband X11 (635)
Proband X12 (664)
Proband X13 (335)
Proband X14 (286)
Proband X15 (487)
Proband X2 (532)
Proband X4 (698)
Proband X5 (593)
Proband X6 (602)
Proband X7 (387)
Proband X8 (620)
Proband X9 (669)
sim (228)
8
4
0
−4
wavelet−LHH_ngtdm_Complexity
frame

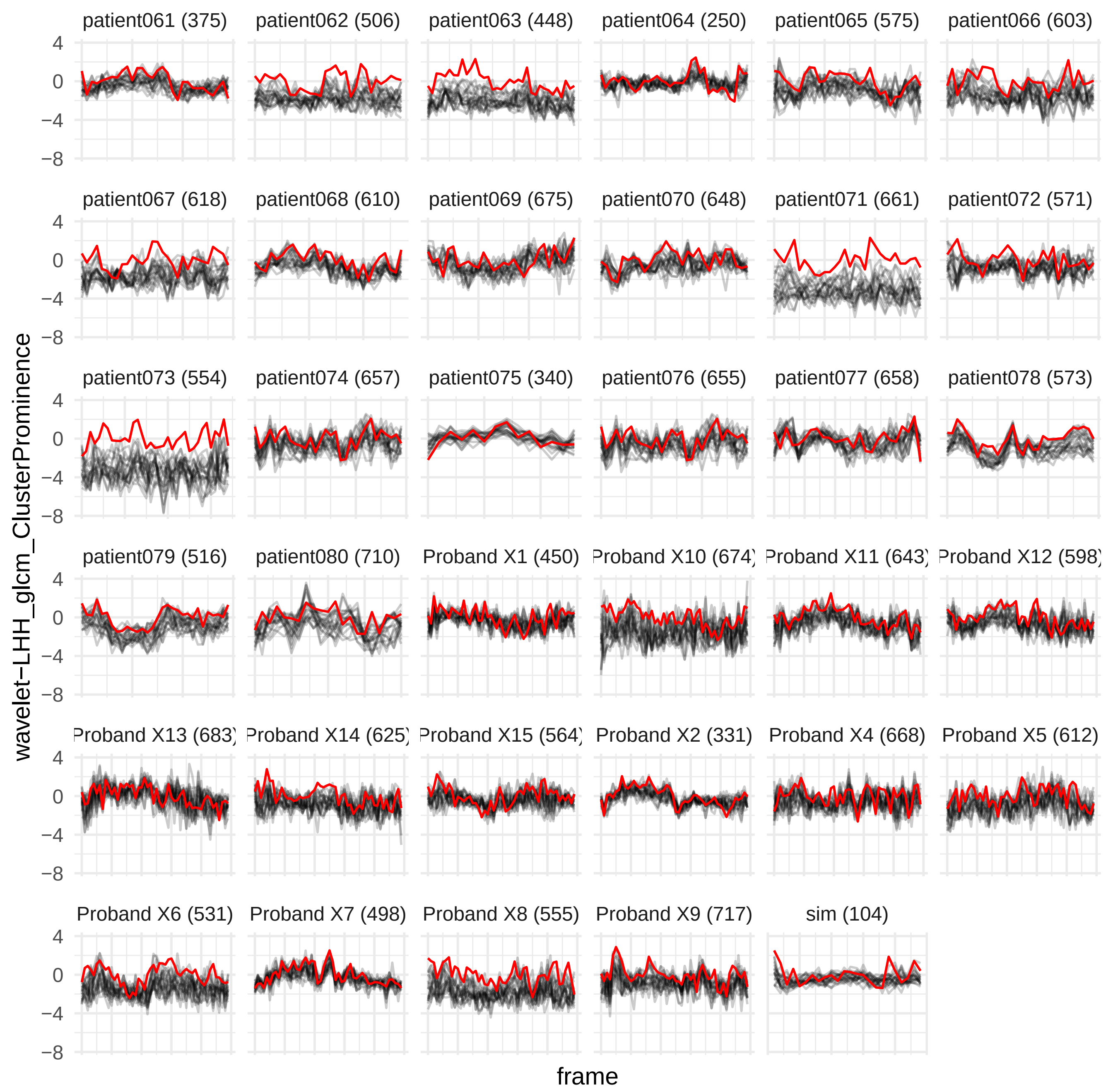

patient061 (375)
patient062 (506)
patient063 (448)
patient064 (250)
patient065 (575)
patient066 (603)
patient067 (618)
patient068 (610)
patient069 (675)
patient070 (648)
patient071 (661)
patient072 (571)
patient073 (554)
patient074 (657)
patient075 (340)
patient076 (655)
patient077 (658)
patient078 (573)
patient079 (516)
patient080 (710)
Proband X1 (450)
Proband X10 (674)
Proband X11 (643)
Proband X12 (598)
Proband X13 (683)
Proband X14 (625)
Proband X15 (564)
Proband X2 (331)
Proband X4 (668)
Proband X5 (612)
Proband X6 (531)
Proband X7 (498)
Proband X8 (555)
Proband X9 (717)
sim (104)
4
0
−4
−8
wavelet−LHH_glcm_ClusterProminence
frame

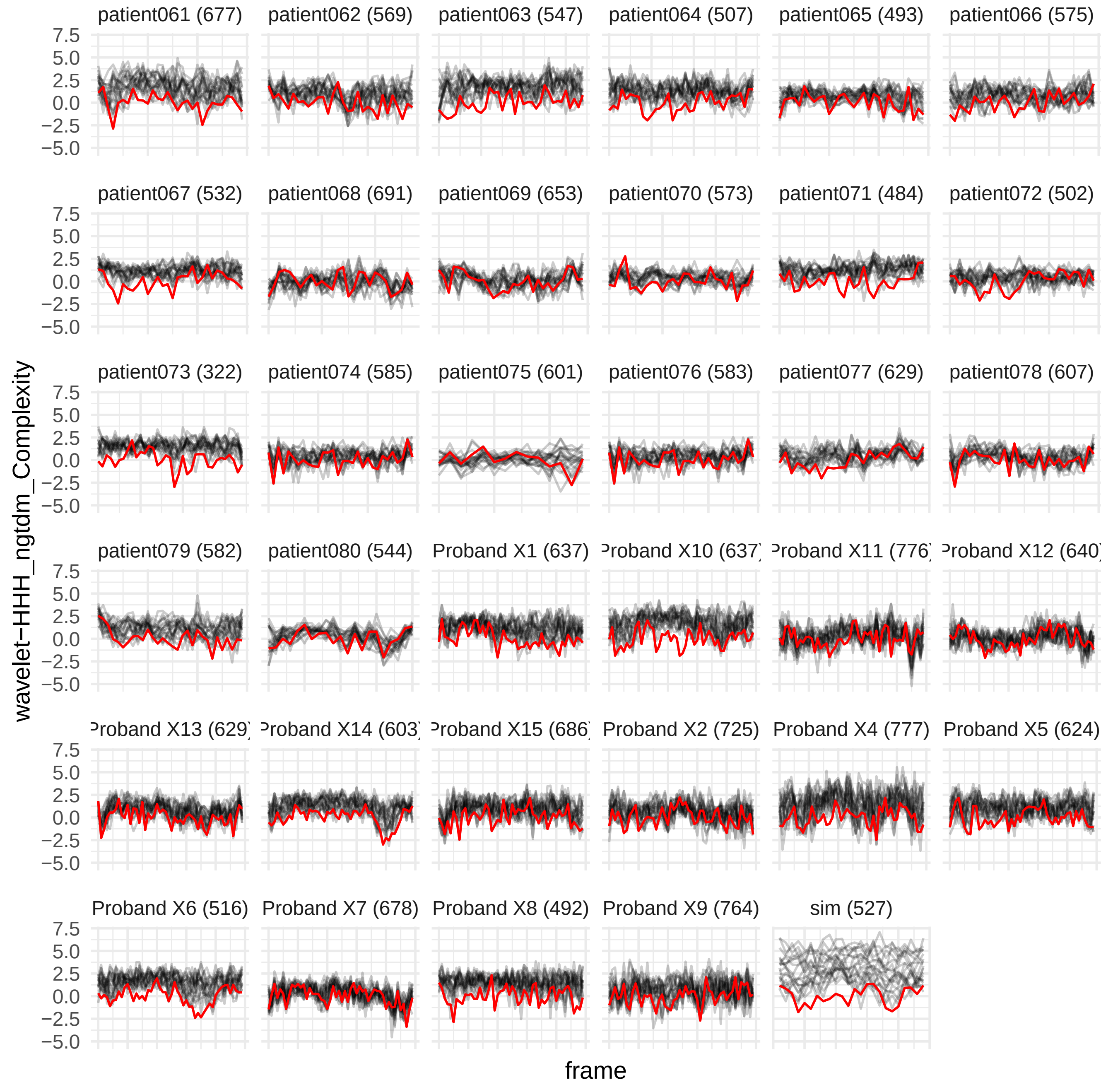

patient061 (677)
patient062 (569)
patient063 (547)
patient064 (507)
patient065 (493)
patient066 (575)
patient067 (532)
patient068 (691)
patient069 (653)
patient070 (573)
patient071 (484)
patient072 (502)
patient073 (322)
patient074 (585)
patient075 (601)
patient076 (583)
patient077 (629)
patient078 (607)
patient079 (582)
patient080 (544)
Proband X1 (637)
Proband X10 (637)
Proband X11 (776)
Proband X12 (640)
Proband X13 (629)
Proband X14 (603)
Proband X15 (686)
Proband X2 (725)
Proband X4 (777)
Proband X5 (624)
Proband X6 (516)
Proband X7 (678)
Proband X8 (492)
Proband X9 (764)
sim (527)
7.5
5.0
2.5
0.0
−2.5
−5.0
wavelet−HHH_ngtdm_Complexity
frame

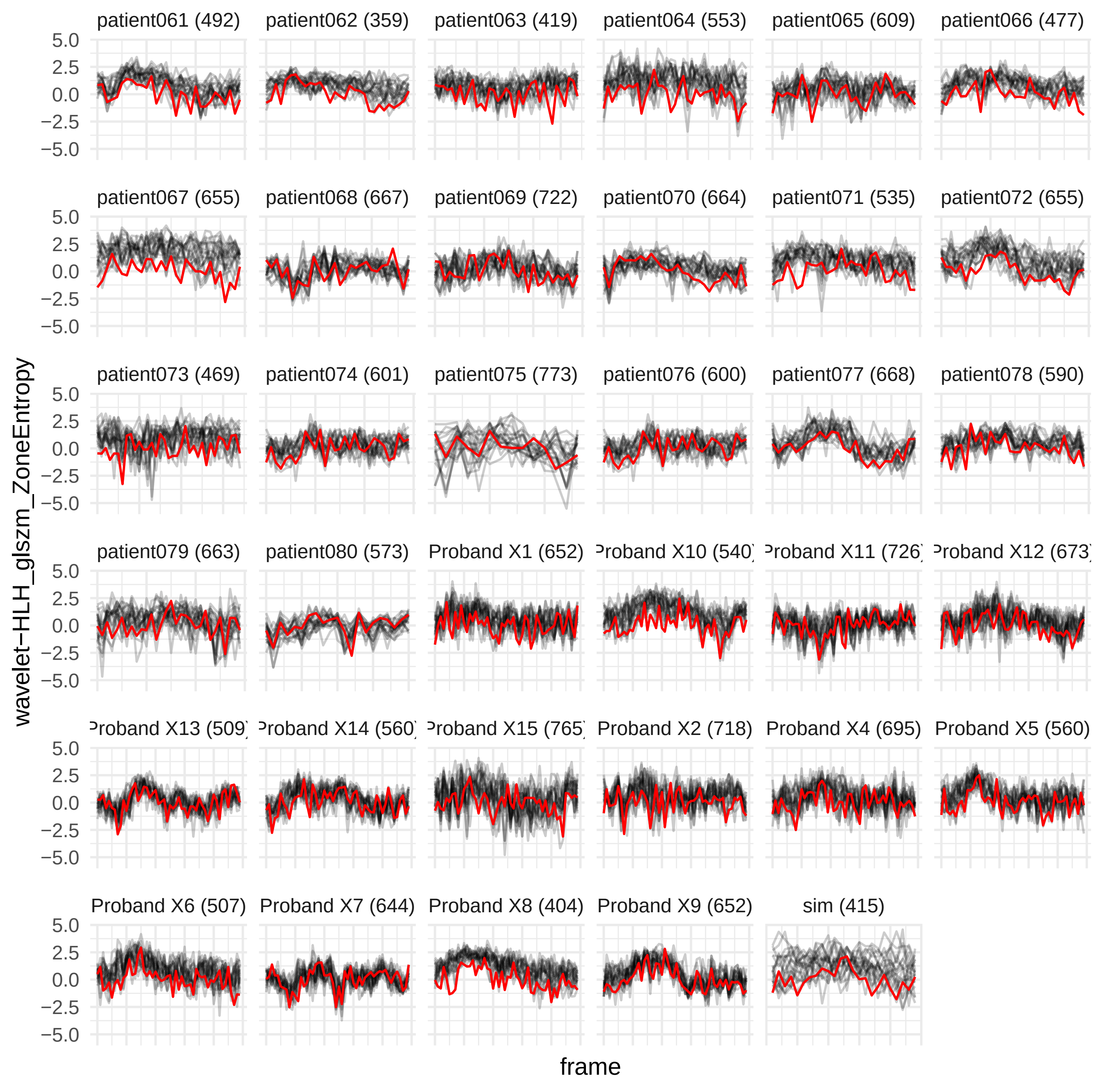
patient061 (492)
patient062 (359)
patient063 (419)
patient064 (553)
patient065 (609)
patient066 (477)
patient067 (655)
patient068 (667)
patient069 (722)
patient070 (664)
patient071 (535)
patient072 (655)
patient073 (469)
patient074 (601)
patient075 (773)
patient076 (600)
patient077 (668)
patient078 (590)
patient079 (663)
patient080 (573)
Proband X1 (652)
Proband X10 (540)
Proband X11 (726)
Proband X12 (673)
Proband X13 (509)
Proband X14 (560)
Proband X15 (765)
Proband X2 (718)
Proband X4 (695)
Proband X5 (560)
Proband X6 (507)
Proband X7 (644)
Proband X8 (404)
Proband X9 (652)
sim (415)
5.0
2.5
0.0
−2.5
−5.0
wavelet−HLH_glszm_ZoneEntropy
frame

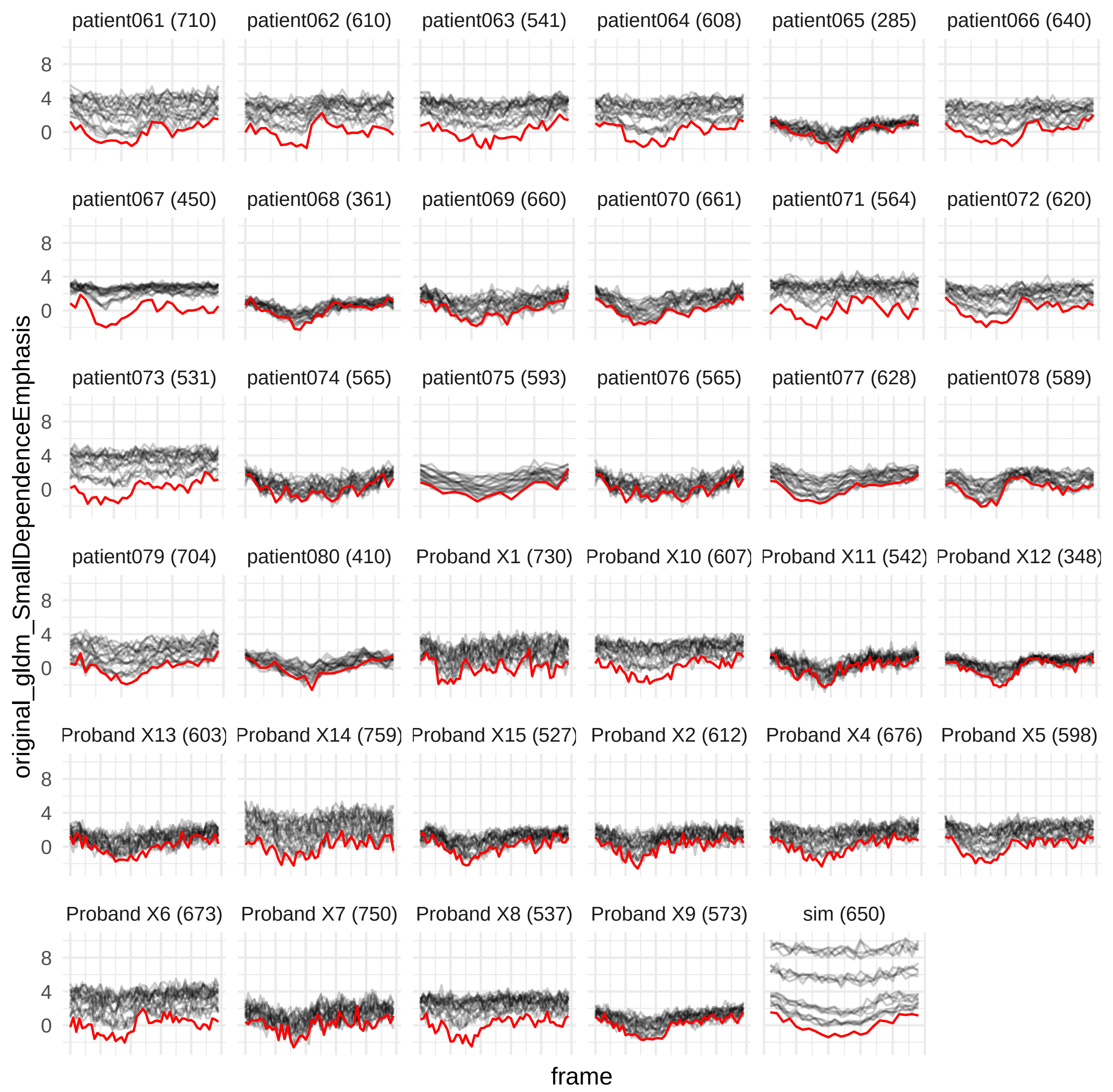

patient061 (710)
patient062 (610)
patient063 (541)
patient064 (608)
patient065 (285)
patient066 (640)
patient067 (450)
patient068 (361)
patient069 (660)
patient070 (661)
patient071 (564)
patient072 (620)
patient073 (531)
patient074 (565)
patient075 (593)
patient076 (565)
patient077 (628)
patient078 (589)
patient079 (704)
patient080 (410)
Proband X1 (730)
Proband X10 (607)
Proband X11 (542)
Proband X12 (348)
Proband X13 (603)
Proband X14 (759)
Proband X15 (527)
Proband X2 (612)
Proband X4 (676)
Proband X5 (598)
Proband X6 (673)
Proband X7 (750)
Proband X8 (537)
Proband X9 (573)
sim (650)
original_gldm_SmallDependenceEmphasis
frame
8
4
0

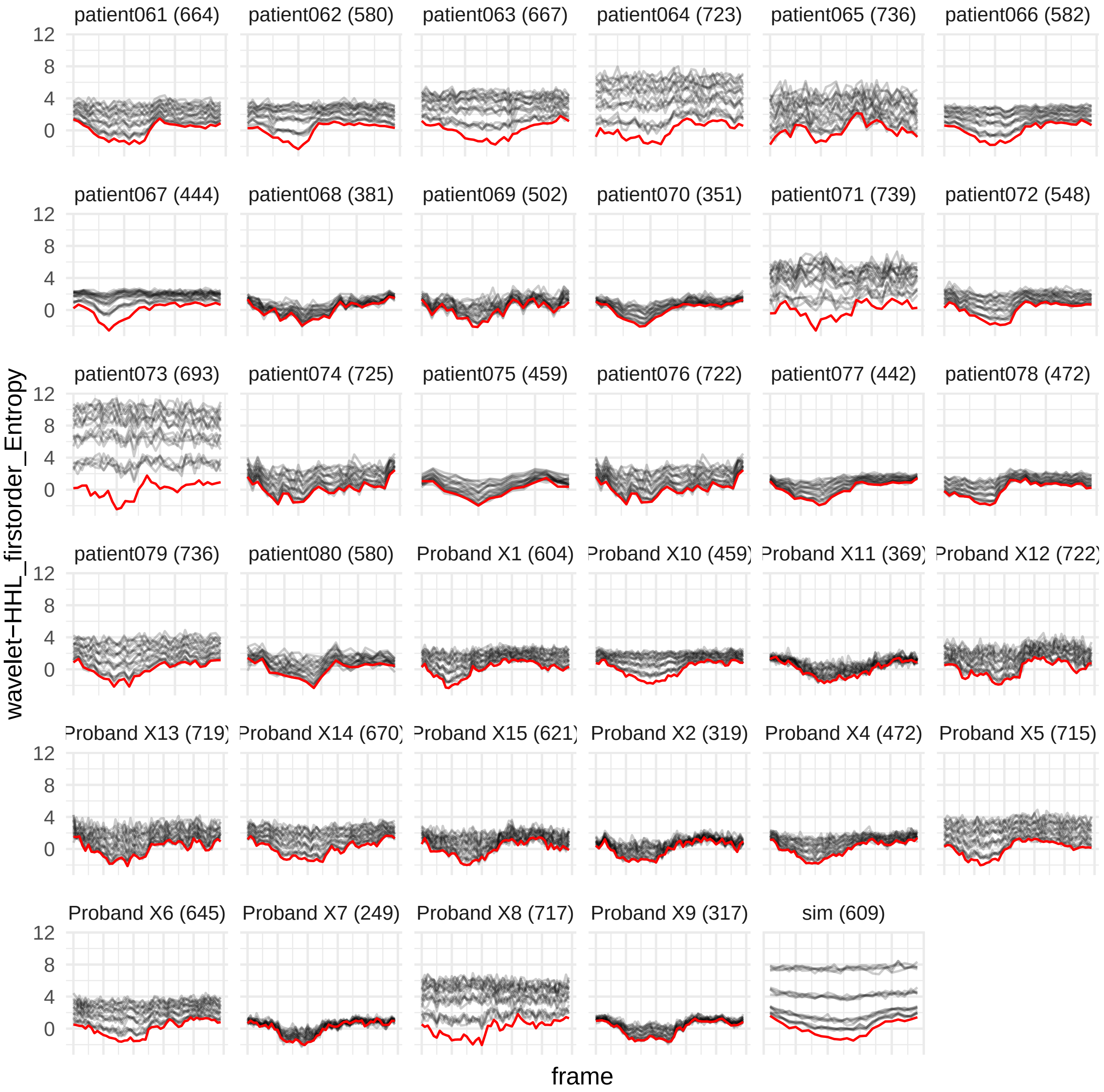
patient061 (664)
patient062 (580)
patient063 (667)
patient064 (723)
patient065 (736)
patient066 (582)
patient067 (444)
patient068 (381)
patient069 (502)
patient070 (351)
patient071 (739)
patient072 (548)
patient073 (693)
patient074 (725)
patient075 (459)
patient076 (722)
patient077 (442)
patient078 (472)
patient079 (736)
patient080 (580)
Proband X1 (604)
Proband X10 (459)
Proband X11 (369)
Proband X12 (722)
Proband X13 (719)
Proband X14 (670)
Proband X15 (621)
Proband X2 (319)
Proband X4 (472)
Proband X5 (715)
Proband X6 (645)
Proband X7 (249)
Proband X8 (717)
Proband X9 (317)
sim (609)
12
8
4
0
wavelet−HHL_firstorder_Entropy
frame

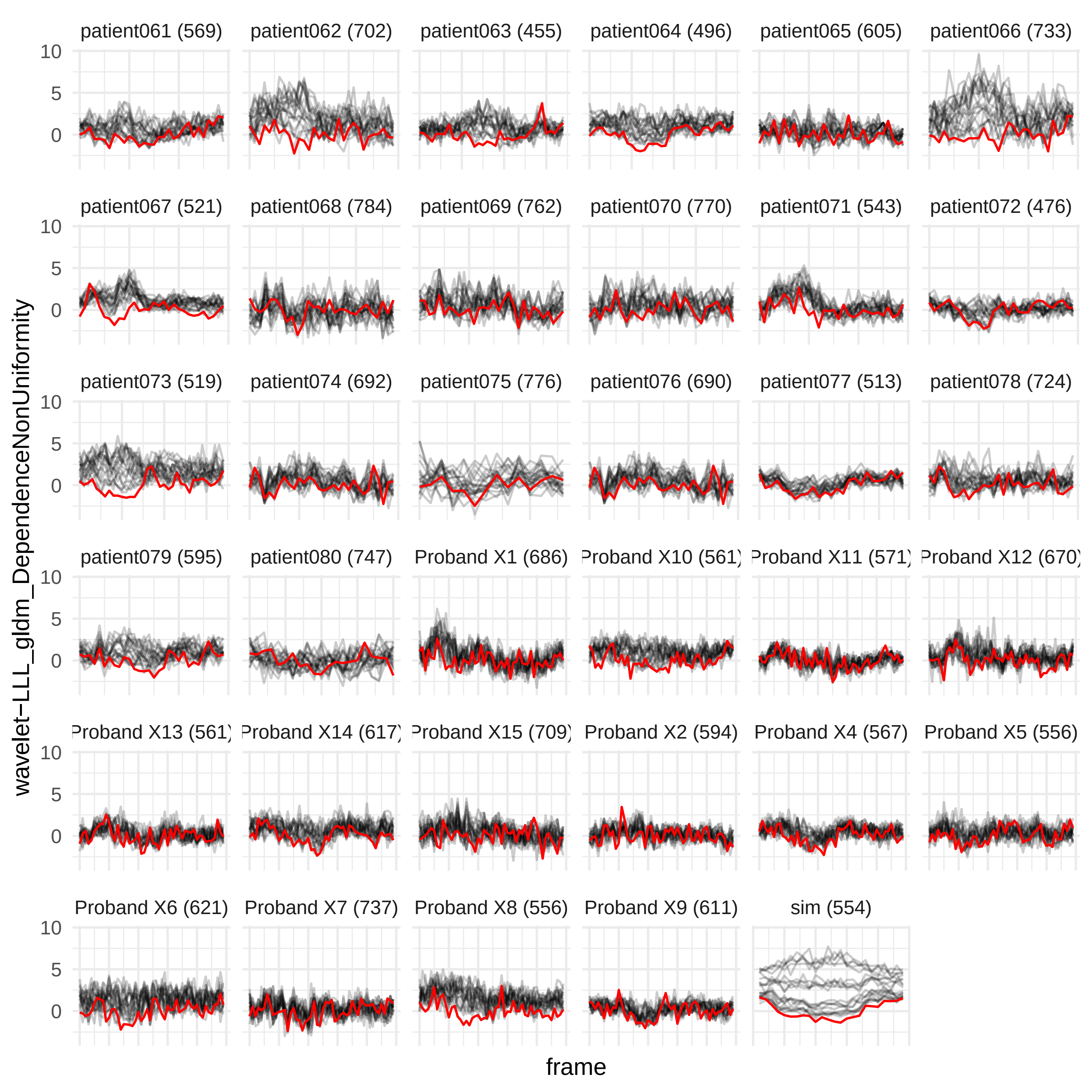

patient061 (569)
patient062 (702)
patient063 (455)
patient064 (496)
patient065 (605)
patient066 (733)
patient067 (521)
patient068 (784)
patient069 (762)
patient070 (770)
patient071 (543)
patient072 (476)
patient073 (519)
patient074 (692)
patient075 (776)
patient076 (690)
patient077 (513)
patient078 (724)
patient079 (595)
patient080 (747)
Proband X1 (686)
Proband X10 (561)
Proband X11 (571)
Proband X12 (670)
Proband X13 (561)
Proband X14 (617)
Proband X15 (709)
Proband X2 (594)
Proband X4 (567)
Proband X5 (556)
Proband X6 (621)
Proband X7 (737)
Proband X8 (556)
Proband X9 (611)
sim (554)
wavelet−LLL_gldm_DependenceNonUniformity
frame
10
5
0

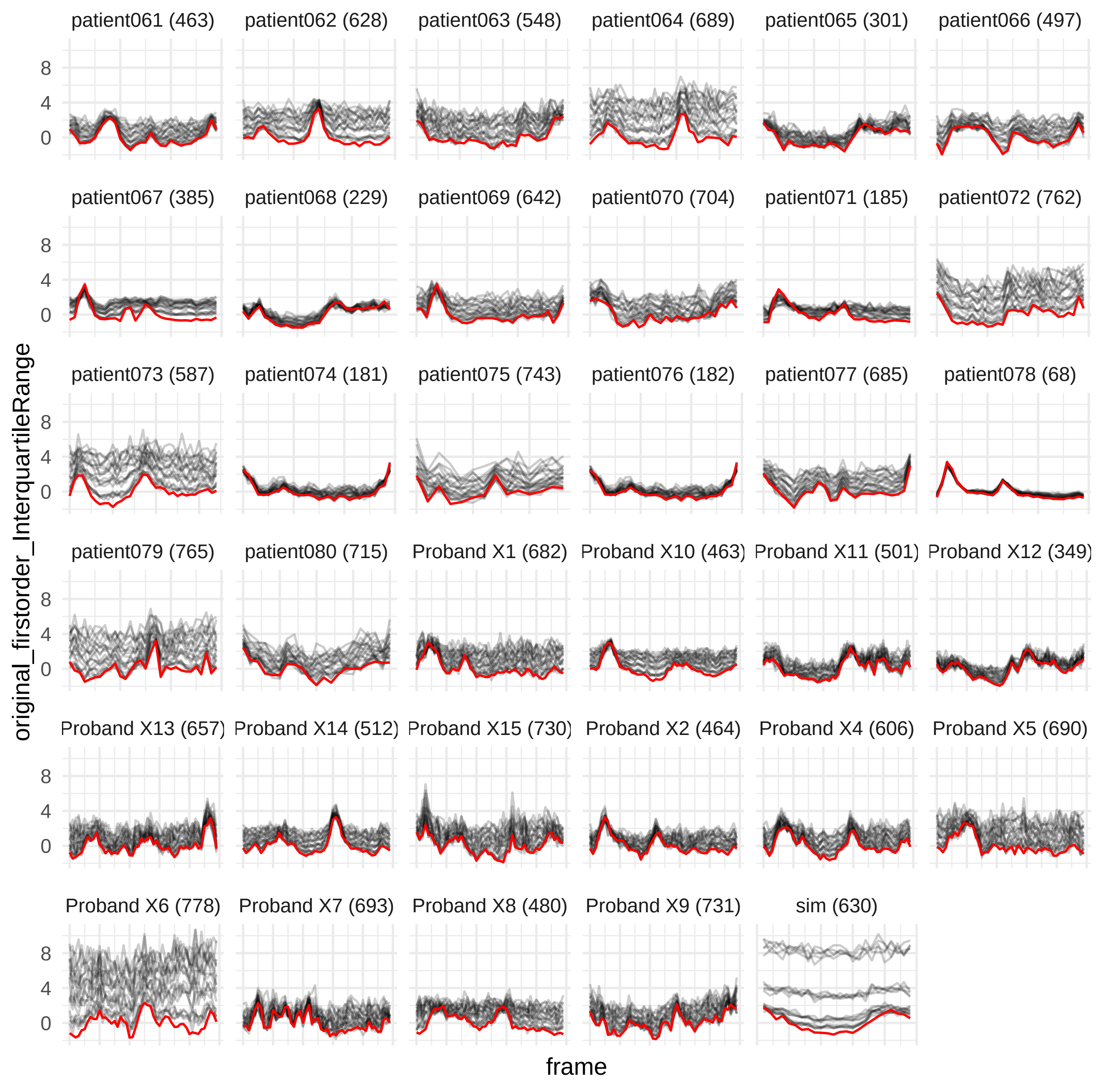
patient061 (463)
patient062 (628)
patient063 (548)
patient064 (689)
patient065 (301)
patient066 (497)
patient067 (385)
patient068 (229)
patient069 (642)
patient070 (704)
patient071 (185)
patient072 (762)
patient073 (587)
patient074 (181)
patient075 (743)
patient076 (182)
patient077 (685)
patient078 (68)
patient079 (765)
patient080 (715)
Proband X1 (682)
Proband X10 (463)
Proband X11 (501)
Proband X12 (349)
Proband X13 (657)
Proband X14 (512)
Proband X15 (730)
Proband X2 (464)
Proband X4 (606)
Proband X5 (690)
Proband X6 (778)
Proband X7 (693)
Proband X8 (480)
Proband X9 (731)
sim (630)
0
4
8
original_firstorder_InterquartileRange
frame

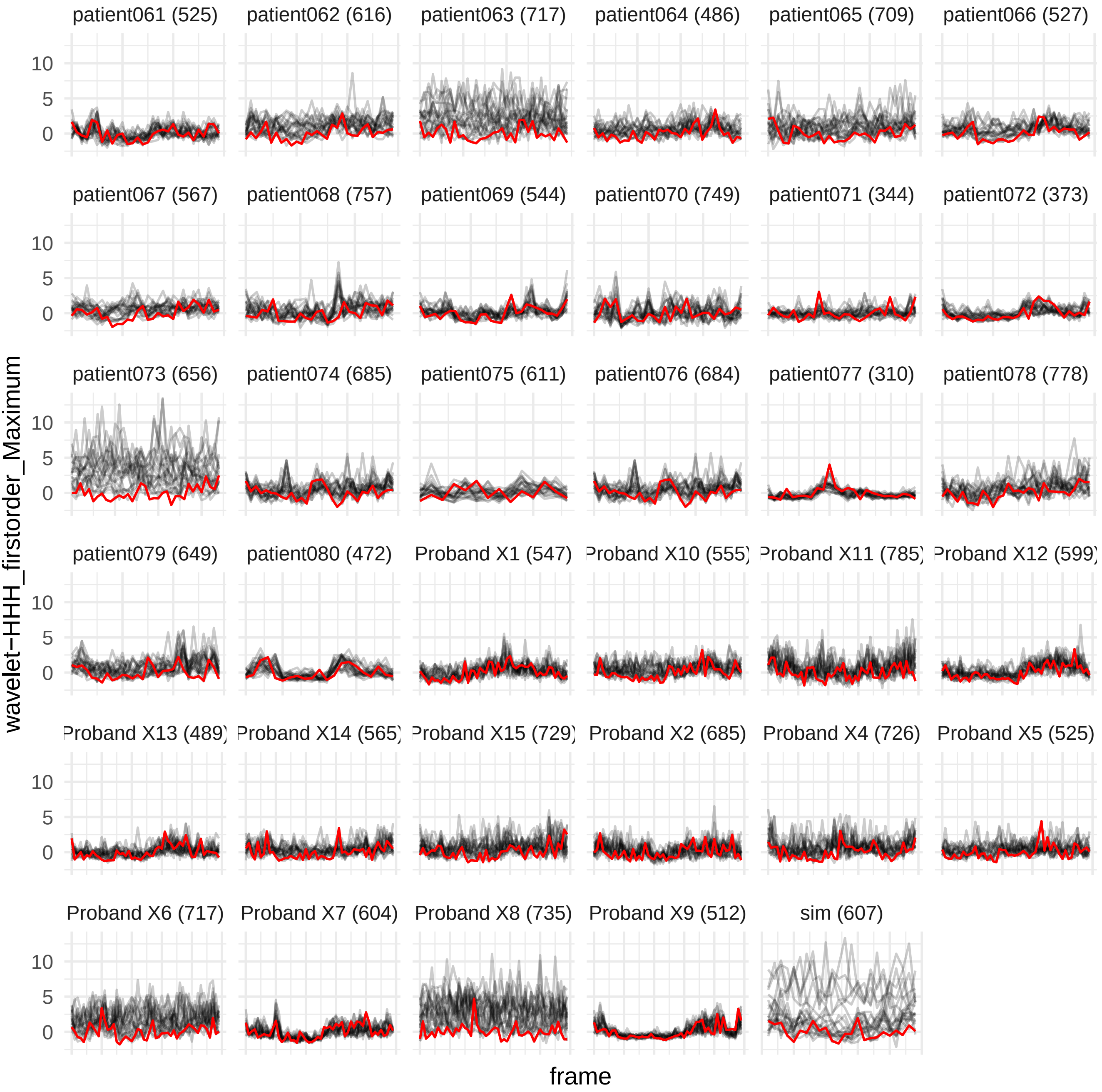
patient061 (525)
patient062 (616)
patient063 (717)
patient064 (486)
patient065 (709)
patient066 (527)
patient067 (567)
patient068 (757)
patient069 (544)
patient070 (749)
patient071 (344)
patient072 (373)
patient073 (656)
patient074 (685)
patient075 (611)
patient076 (684)
patient077 (310)
patient078 (778)
patient079 (649)
patient080 (472)
Proband X1 (547)
Proband X10 (555)
Proband X11 (785)
Proband X12 (599)
Proband X13 (489)
Proband X14 (565)
Proband X15 (729)
Proband X2 (685)
Proband X4 (726)
Proband X5 (525)
Proband X6 (717)
Proband X7 (604)
Proband X8 (735)
Proband X9 (512)
sim (607)
wavelet−HHH_firstorder_Maximum
frame
0
5
10

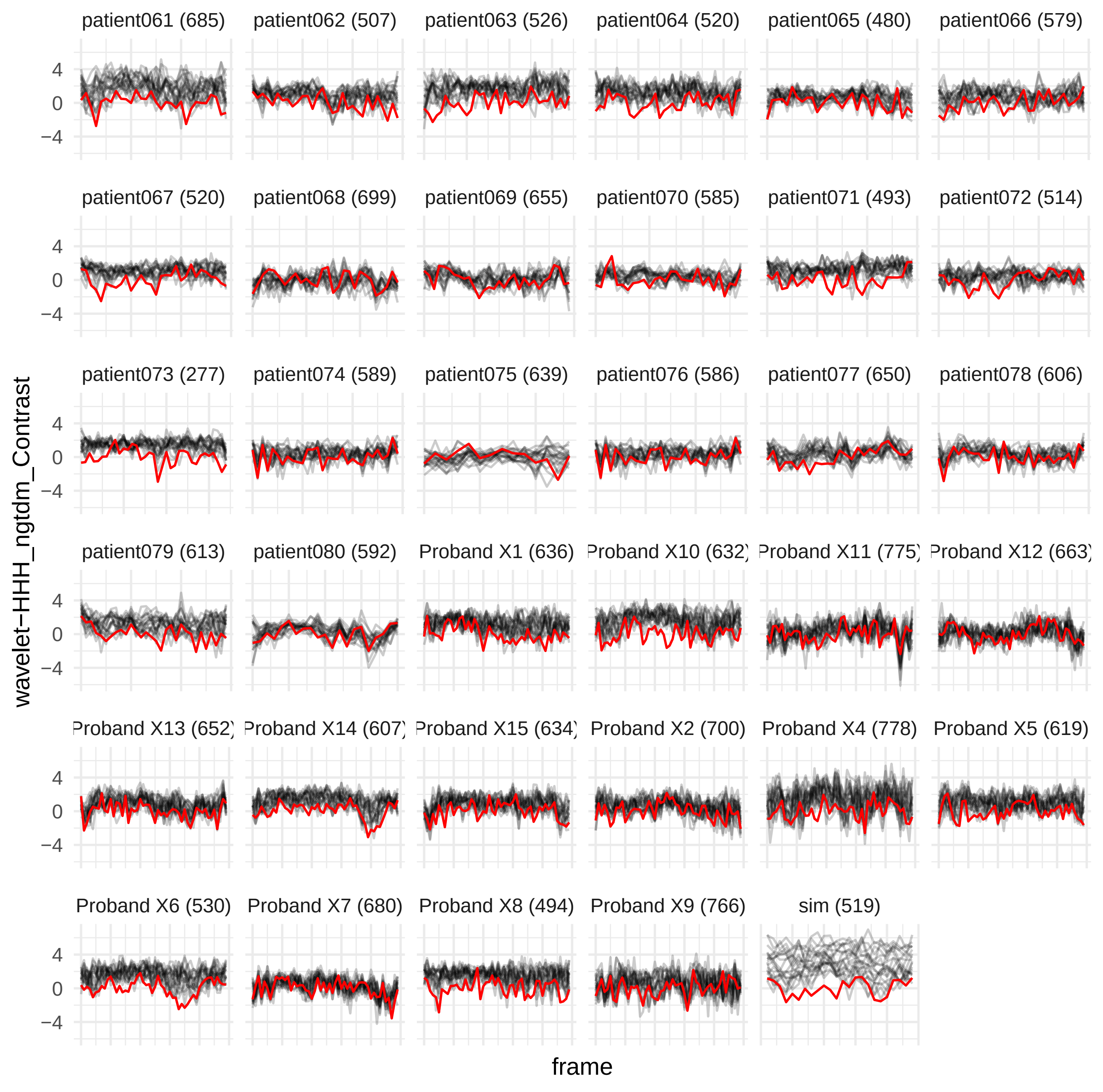
patient061 (685)
patient062 (507)
patient063 (526)
patient064 (520)
patient065 (480)
patient066 (579)
patient067 (520)
patient068 (699)
patient069 (655)
patient070 (585)
patient071 (493)
patient072 (514)
patient073 (277)
patient074 (589)
patient075 (639)
patient076 (586)
patient077 (650)
patient078 (606)
patient079 (613)
patient080 (592)
Proband X1 (636)
Proband X10 (632)
Proband X11 (775)
Proband X12 (663)
Proband X13 (652)
Proband X14 (607)
Proband X15 (634)
Proband X2 (700)
Proband X4 (778)
Proband X5 (619)
Proband X6 (530)
Proband X7 (680)
Proband X8 (494)
Proband X9 (766)
sim (519)
4
0
−4
wavelet−HHH_ngtdm_Contrast
frame

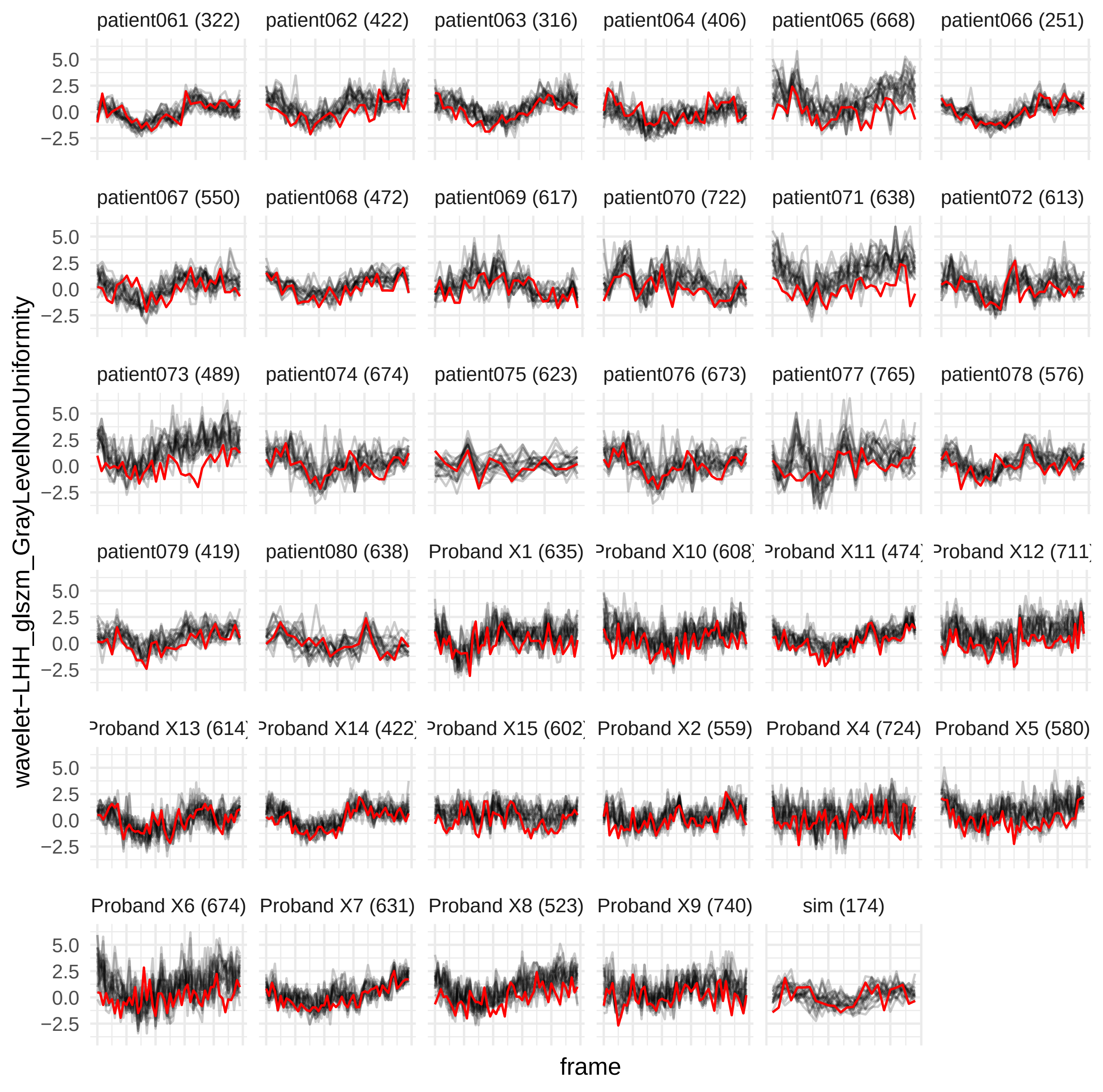

patient061 (322)
patient062 (422)
patient063 (316)
patient064 (406)
patient065 (668)
patient066 (251)
patient067 (550)
patient068 (472)
patient069 (617)
patient070 (722)
patient071 (638)
patient072 (613)
patient073 (489)
patient074 (674)
patient075 (623)
patient076 (673)
patient077 (765)
patient078 (576)
patient079 (419)
patient080 (638)
Proband X1 (635)
Proband X10 (608)
Proband X11 (474)
Proband X12 (711)
Proband X13 (614)
Proband X14 (422)
Proband X15 (602)
Proband X2 (559)
Proband X4 (724)
Proband X5 (580)
Proband X6 (674)
Proband X7 (631)
Proband X8 (523)
Proband X9 (740)
sim (174)
5.0
2.5
0.0
−2.5
wavelet−LHH_glszm_GrayLevelNonUniformity
frame

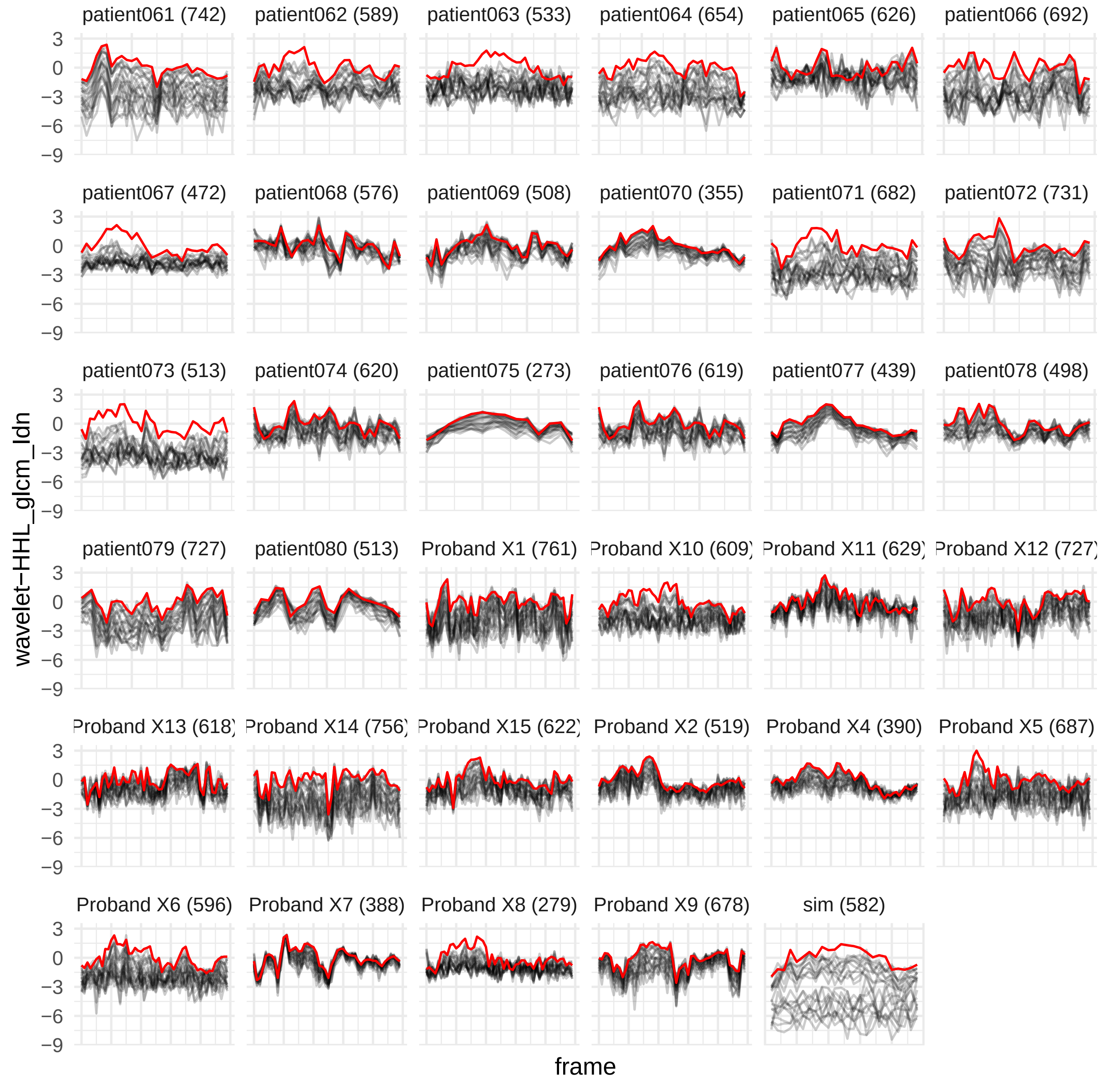
patient061 (742)
patient062 (589)
patient063 (533)
patient064 (654)
patient065 (626)
patient066 (692)
patient067 (472)
patient068 (576)
patient069 (508)
patient070 (355)
patient071 (682)
patient072 (731)
patient073 (513)
patient074 (620)
patient075 (273)
patient076 (619)
patient077 (439)
patient078 (498)
patient079 (727)
patient080 (513)
Proband X1 (761)
Proband X10 (609)
Proband X11 (629)
Proband X12 (727)
Proband X13 (618)
Proband X14 (756)
Proband X15 (622)
Proband X2 (519)
Proband X4 (390)
Proband X5 (687)
Proband X6 (596)
Proband X7 (388)
Proband X8 (279)
Proband X9 (678)
sim (582)
3
0
−3
−6
−9
wavelet−HHL_glcm_Idn
frame

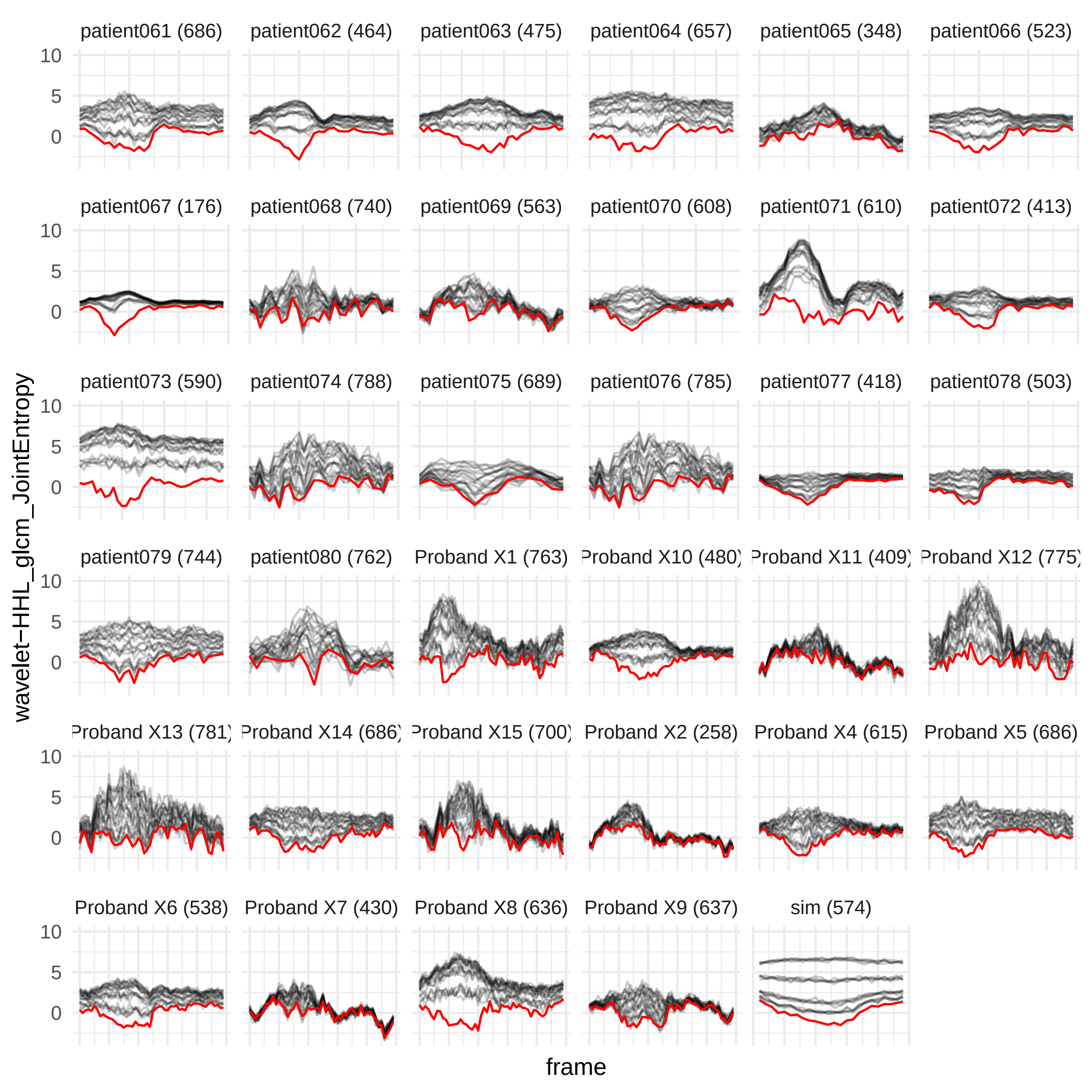

patient061 (686)
patient062 (464)
patient063 (475)
patient064 (657)
patient065 (348)
patient066 (523)
patient067 (176)
patient068 (740)
patient069 (563)
patient070 (608)
patient071 (610)
patient072 (413)
patient073 (590)
patient074 (788)
patient075 (689)
patient076 (785)
patient077 (418)
patient078 (503)
patient079 (744)
patient080 (762)
Proband X1 (763)
Proband X10 (480)
Proband X11 (409)
Proband X12 (775)
Proband X13 (781)
Proband X14 (686)
Proband X15 (700)
Proband X2 (258)
Proband X4 (615)
Proband X5 (686)
Proband X6 (538)
Proband X7 (430)
Proband X8 (636)
Proband X9 (637)
sim (574)
10
5
0
wavelet−HHL_glcm_JointEntropy
frame

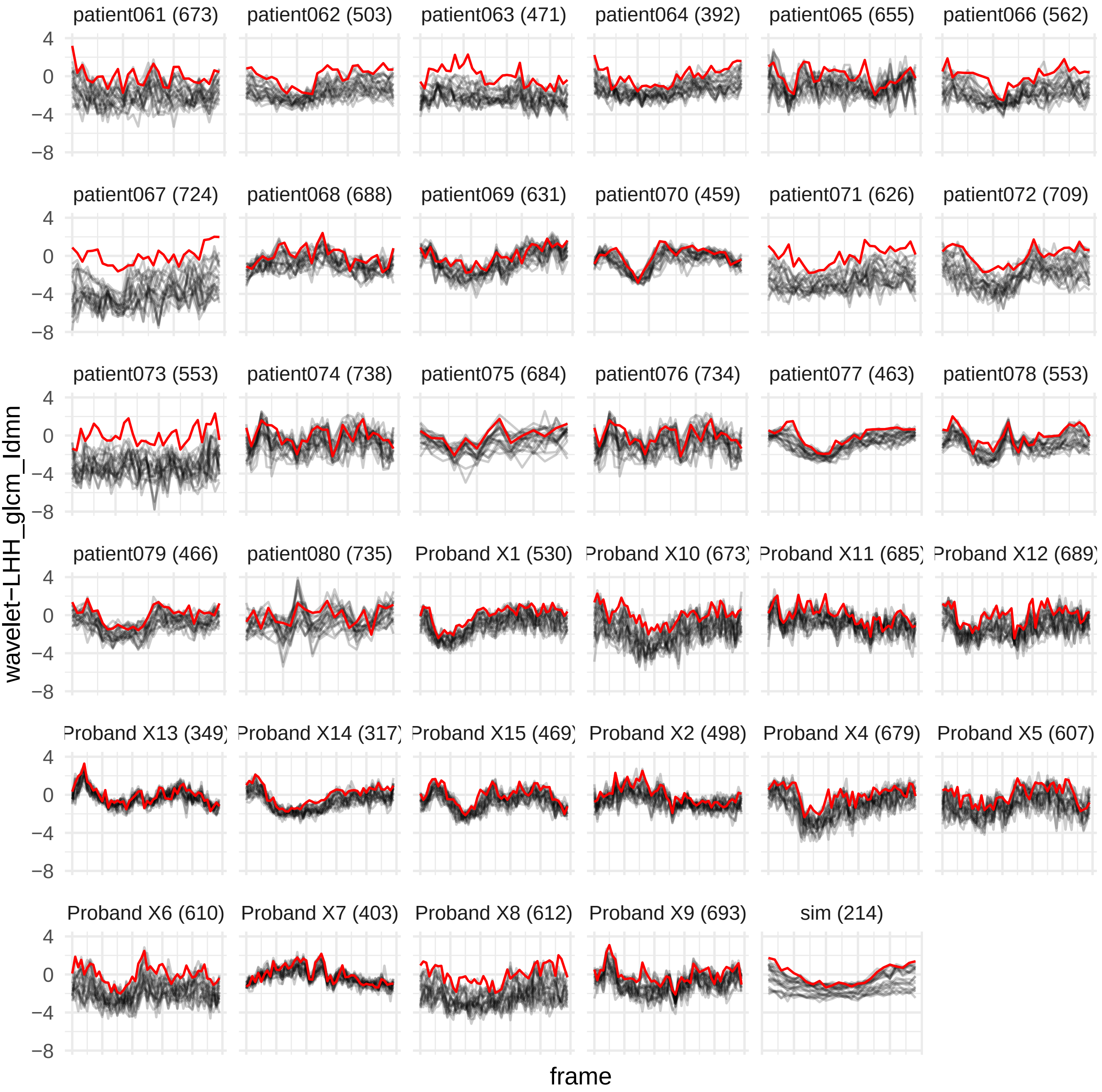

patient061 (673)
patient062 (503)
patient063 (471)
patient064 (392)
patient065 (655)
patient066 (562)
patient067 (724)
patient068 (688)
patient069 (631)
patient070 (459)
patient071 (626)
patient072 (709)
patient073 (553)
patient074 (738)
patient075 (684)
patient076 (734)
patient077 (463)
patient078 (553)
patient079 (466)
patient080 (735)
Proband X1 (530)
Proband X10 (673)
Proband X11 (685)
Proband X12 (689)
Proband X13 (349)
Proband X14 (317)
Proband X15 (469)
Proband X2 (498)
Proband X4 (679)
Proband X5 (607)
Proband X6 (610)
Proband X7 (403)
Proband X8 (612)
Proband X9 (693)
sim (214)
4
0
−4
−8
wavelet−LHH_glcm_Idmn
frame

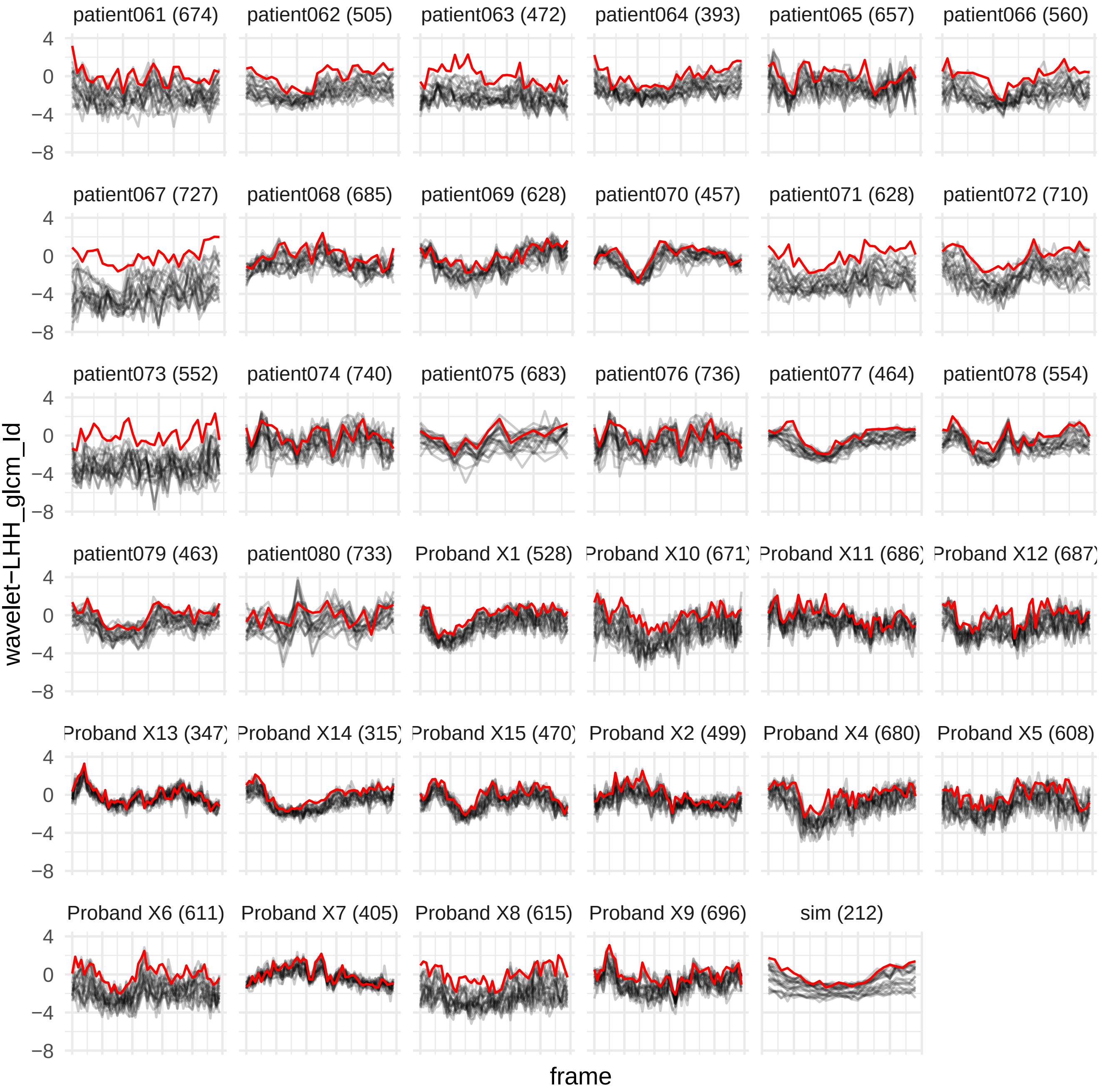
patient061 (674)
patient062 (505)
patient063 (472)
patient064 (393)
patient065 (657)
patient066 (560)
patient067 (727)
patient068 (685)
patient069 (628)
patient070 (457)
patient071 (628)
patient072 (710)
patient073 (552)
patient074 (740)
patient075 (683)
patient076 (736)
patient077 (464)
patient078 (554)
patient079 (463)
patient080 (733)
Proband X1 (528)
Proband X10 (671)
Proband X11 (686)
Proband X12 (687)
Proband X13 (347)
Proband X14 (315)
Proband X15 (470)
Proband X2 (499)
Proband X4 (680)
Proband X5 (608)
Proband X6 (611)
Proband X7 (405)
Proband X8 (615)
Proband X9 (696)
sim (212)
4
0
−4
−8
wavelet−LHH_glcm_Id
frame

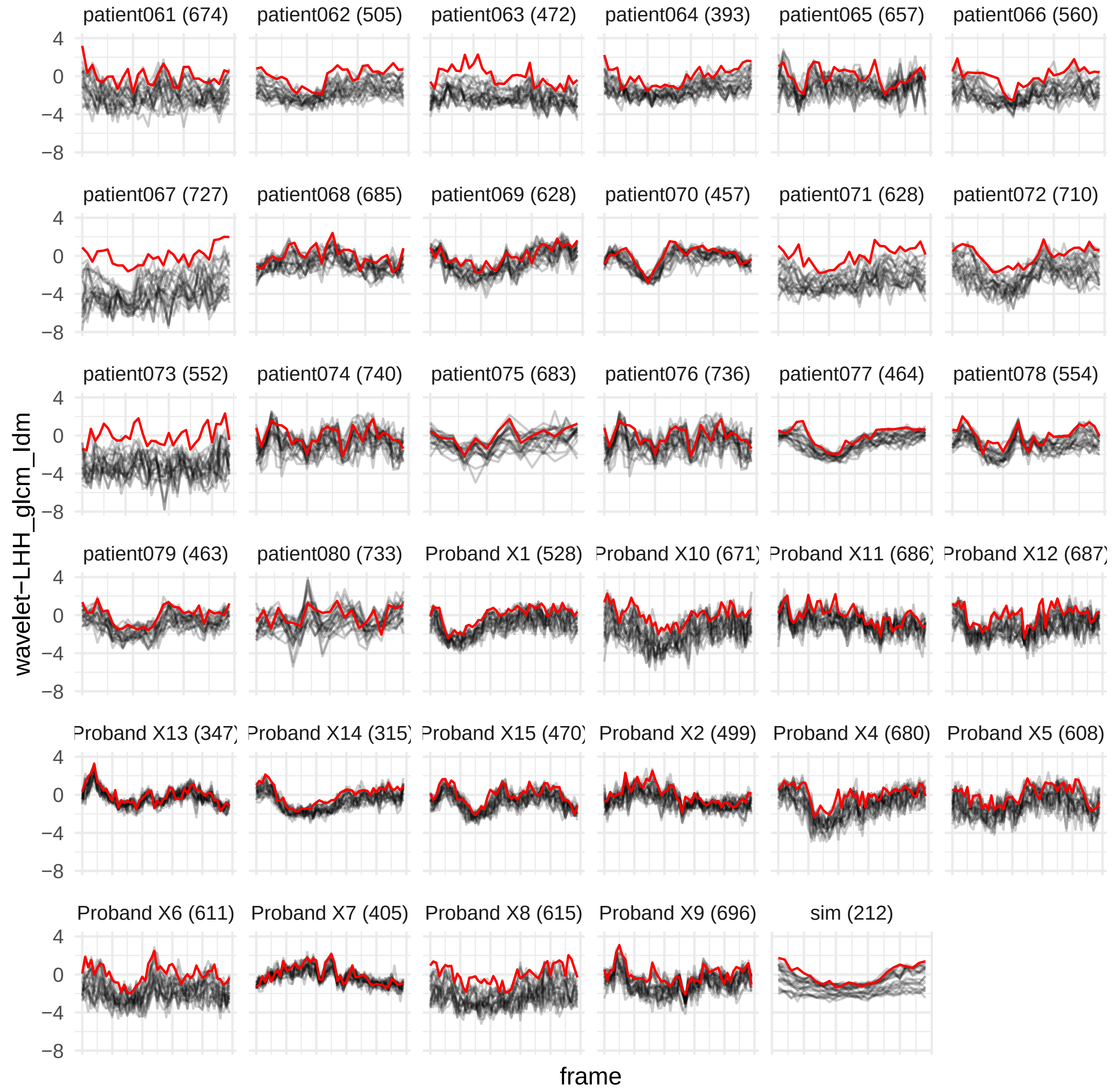

patient061 (674)
patient062 (505)
patient063 (472)
patient064 (393)
patient065 (657)
patient066 (560)
patient067 (727)
patient068 (685)
patient069 (628)
patient070 (457)
patient071 (628)
patient072 (710)
patient073 (552)
patient074 (740)
patient075 (683)
patient076 (736)
patient077 (464)
patient078 (554)
patient079 (463)
patient080 (733)
Proband X1 (528)
Proband X10 (671)
Proband X11 (686)
Proband X12 (687)
Proband X13 (347)
Proband X14 (315)
Proband X15 (470)
Proband X2 (499)
Proband X4 (680)
Proband X5 (608)
Proband X6 (611)
Proband X7 (405)
Proband X8 (615)
Proband X9 (696)
sim (212)
4
0
−4
−8
wavelet−LHH_glcm_Idm
frame

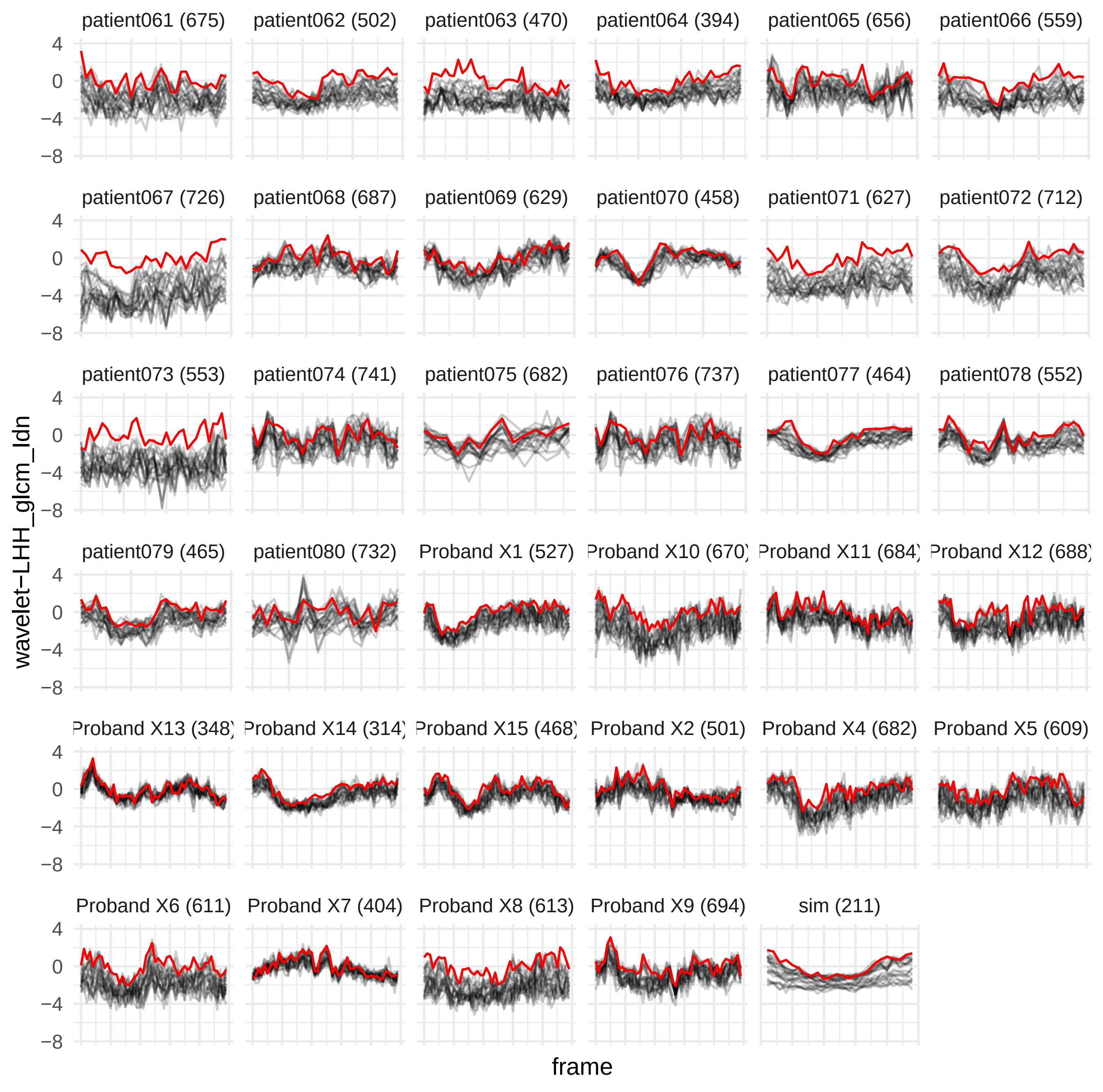
patient061 (675)
patient062 (502)
patient063 (470)
patient064 (394)
patient065 (656)
patient066 (559)
patient067 (726)
patient068 (687)
patient069 (629)
patient070 (458)
patient071 (627)
patient072 (712)
patient073 (553)
patient074 (741)
patient075 (682)
patient076 (737)
patient077 (464)
patient078 (552)
patient079 (465)
patient080 (732)
Proband X1 (527)
Proband X10 (670)
Proband X11 (684)
Proband X12 (688)
Proband X13 (348)
Proband X14 (314)
Proband X15 (468)
Proband X2 (501)
Proband X4 (682)
Proband X5 (609)
Proband X6 (611)
Proband X7 (404)
Proband X8 (613)
Proband X9 (694)
sim (211)
4
0
−4
−8
wavelet−LHH_glcm_Idn
frame

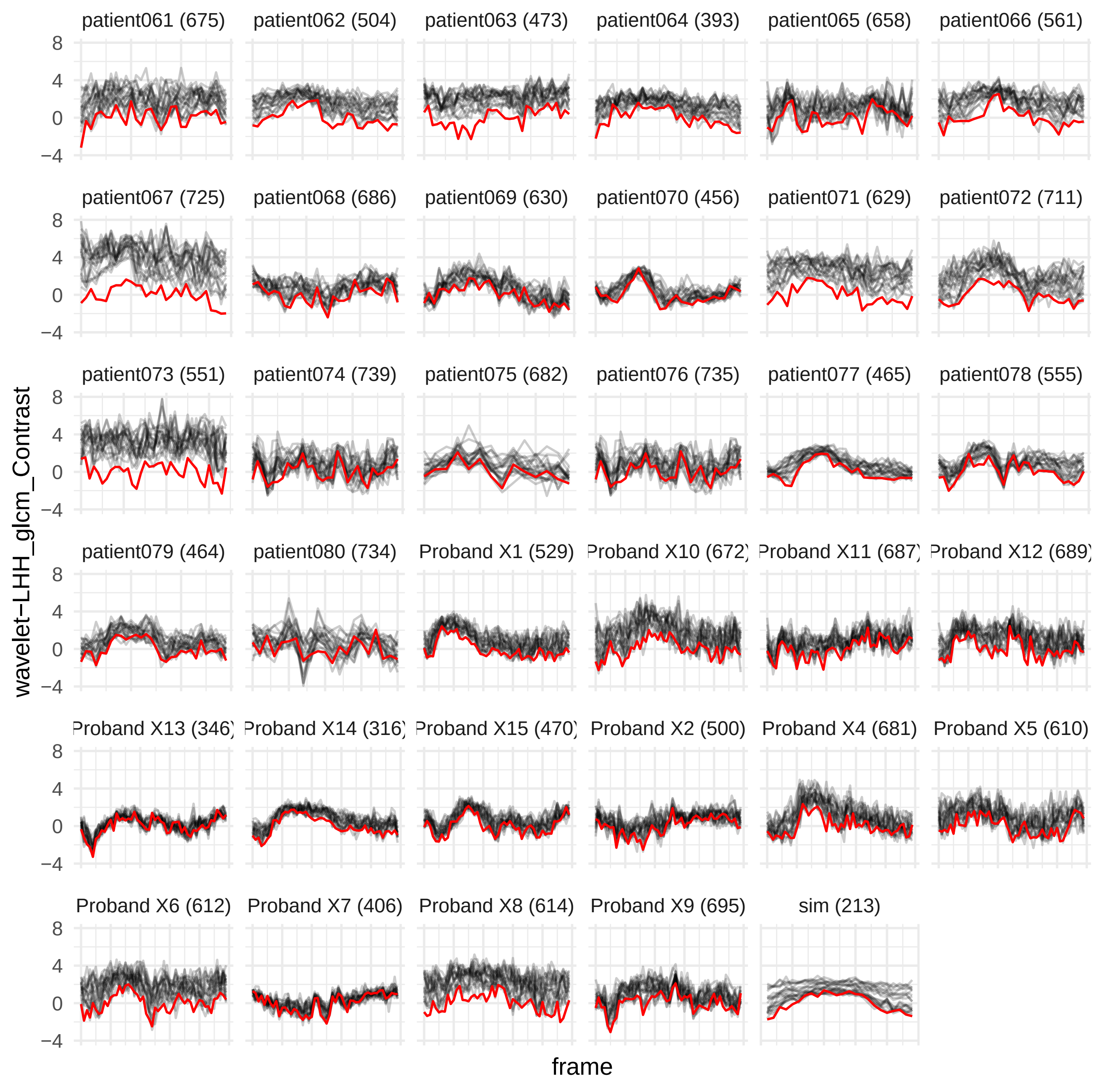
patient061 (675)
patient062 (504)
patient063 (473)
patient064 (393)
patient065 (658)
patient066 (561)
patient067 (725)
patient068 (686)
patient069 (630)
patient070 (456)
patient071 (629)
patient072 (711)
patient073 (551)
patient074 (739)
patient075 (682)
patient076 (735)
patient077 (465)
patient078 (555)
patient079 (464)
patient080 (734)
Proband X1 (529)
Proband X10 (672)
Proband X11 (687)
Proband X12 (689)
Proband X13 (346)
Proband X14 (316)
Proband X15 (470)
Proband X2 (500)
Proband X4 (681)
Proband X5 (610)
Proband X6 (612)
Proband X7 (406)
Proband X8 (614)
Proband X9 (695)
sim (213)
8
4
0
−4
wavelet−LHH_glcm_Contrast
frame

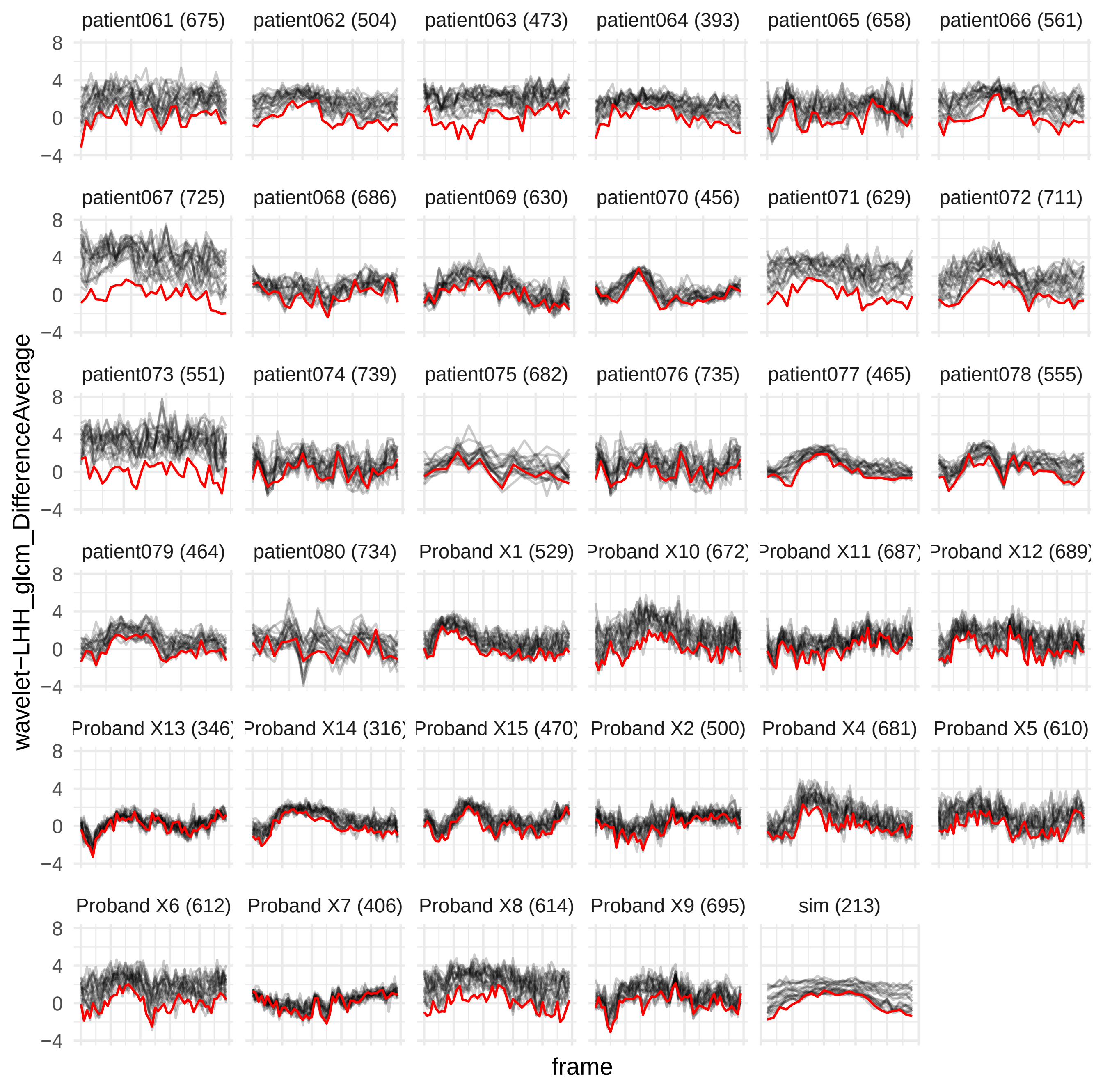

patient061 (675)
patient062 (504)
patient063 (473)
patient064 (393)
patient065 (658)
patient066 (561)
patient067 (725)
patient068 (686)
patient069 (630)
patient070 (456)
patient071 (629)
patient072 (711)
patient073 (551)
patient074 (739)
patient075 (682)
patient076 (735)
patient077 (465)
patient078 (555)
patient079 (464)
patient080 (734)
Proband X1 (529)
Proband X10 (672)
Proband X11 (687)
Proband X12 (689)
Proband X13 (346)
Proband X14 (316)
Proband X15 (470)
Proband X2 (500)
Proband X4 (681)
Proband X5 (610)
Proband X6 (612)
Proband X7 (406)
Proband X8 (614)
Proband X9 (695)
sim (213)
8
4
0
−4
wavelet−LHH_glcm_DifferenceAverage
frame

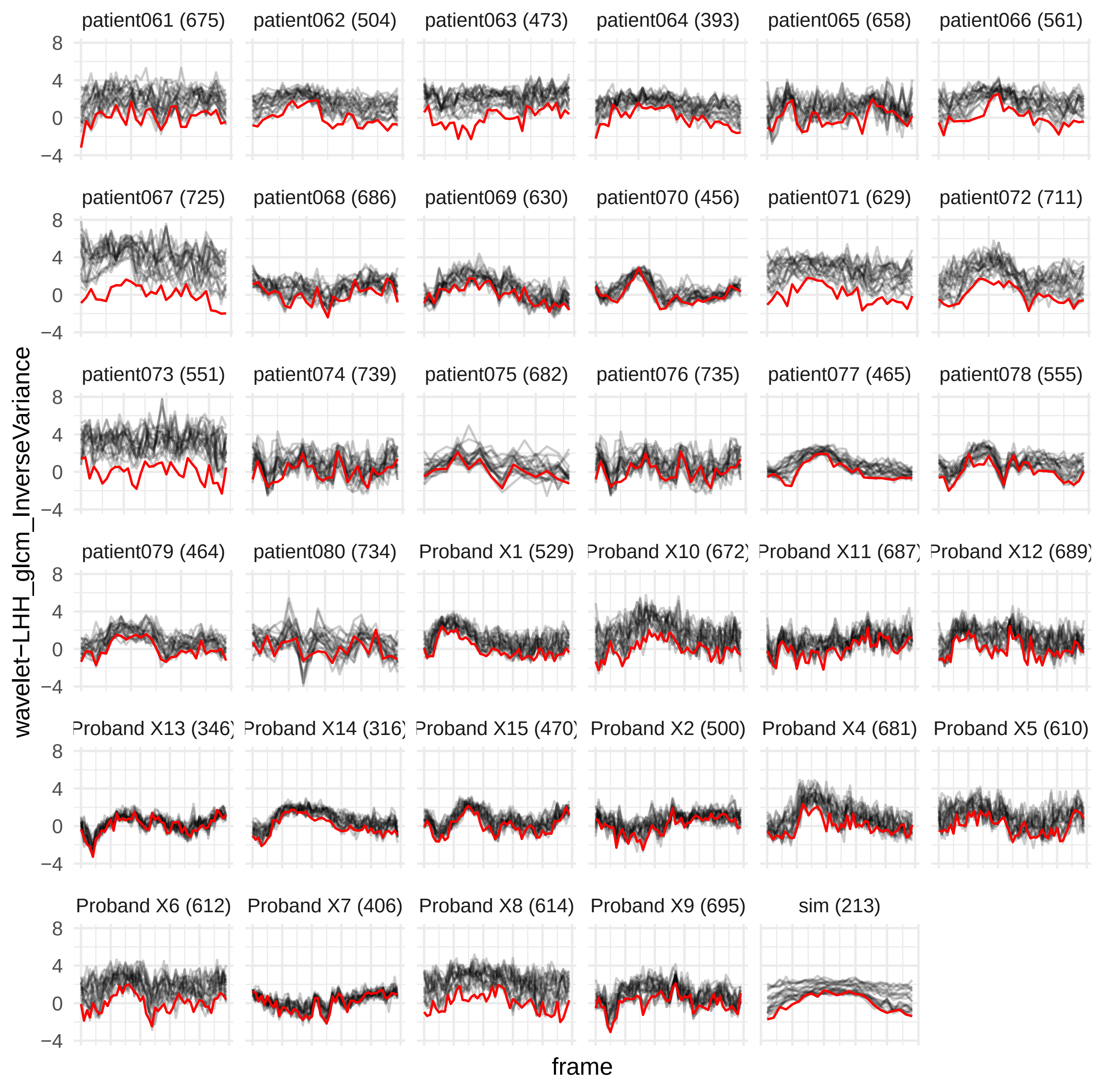
patient061 (675)
patient062 (504)
patient063 (473)
patient064 (393)
patient065 (658)
patient066 (561)
patient067 (725)
patient068 (686)
patient069 (630)
patient070 (456)
patient071 (629)
patient072 (711)
patient073 (551)
patient074 (739)
patient075 (682)
patient076 (735)
patient077 (465)
patient078 (555)
patient079 (464)
patient080 (734)
Proband X1 (529)
Proband X10 (672)
Proband X11 (687)
Proband X12 (689)
Proband X13 (346)
Proband X14 (316)
Proband X15 (470)
Proband X2 (500)
Proband X4 (681)
Proband X5 (610)
Proband X6 (612)
Proband X7 (406)
Proband X8 (614)
Proband X9 (695)
sim (213)
8
4
0
−4
wavelet−LHH_glcm_InverseVariance
frame

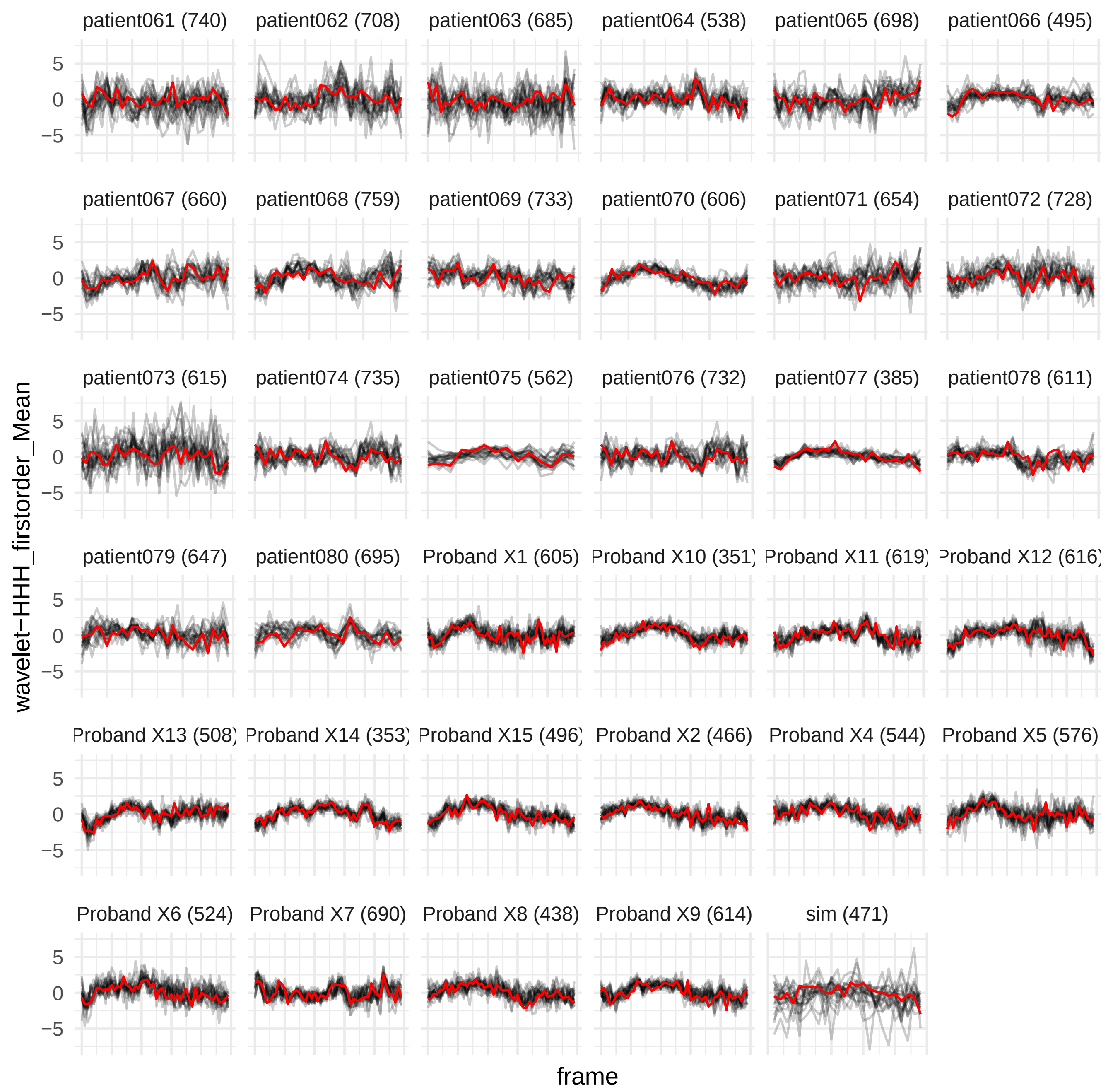
patient061 (740)
patient062 (708)
patient063 (685)
patient064 (538)
patient065 (698)
patient066 (495)
patient067 (660)
patient068 (759)
patient069 (733)
patient070 (606)
patient071 (654)
patient072 (728)
patient073 (615)
patient074 (735)
patient075 (562)
patient076 (732)
patient077 (385)
patient078 (611)
patient079 (647)
patient080 (695)
Proband X1 (605)
Proband X10 (351)
Proband X11 (619)
Proband X12 (616)
Proband X13 (508)
Proband X14 (353)
Proband X15 (496)
Proband X2 (466)
Proband X4 (544)
Proband X5 (576)
Proband X6 (524)
Proband X7 (690)
Proband X8 (438)
Proband X9 (614)
sim (471)
5
0
−5
wavelet−HHH_firstorder_Mean
frame

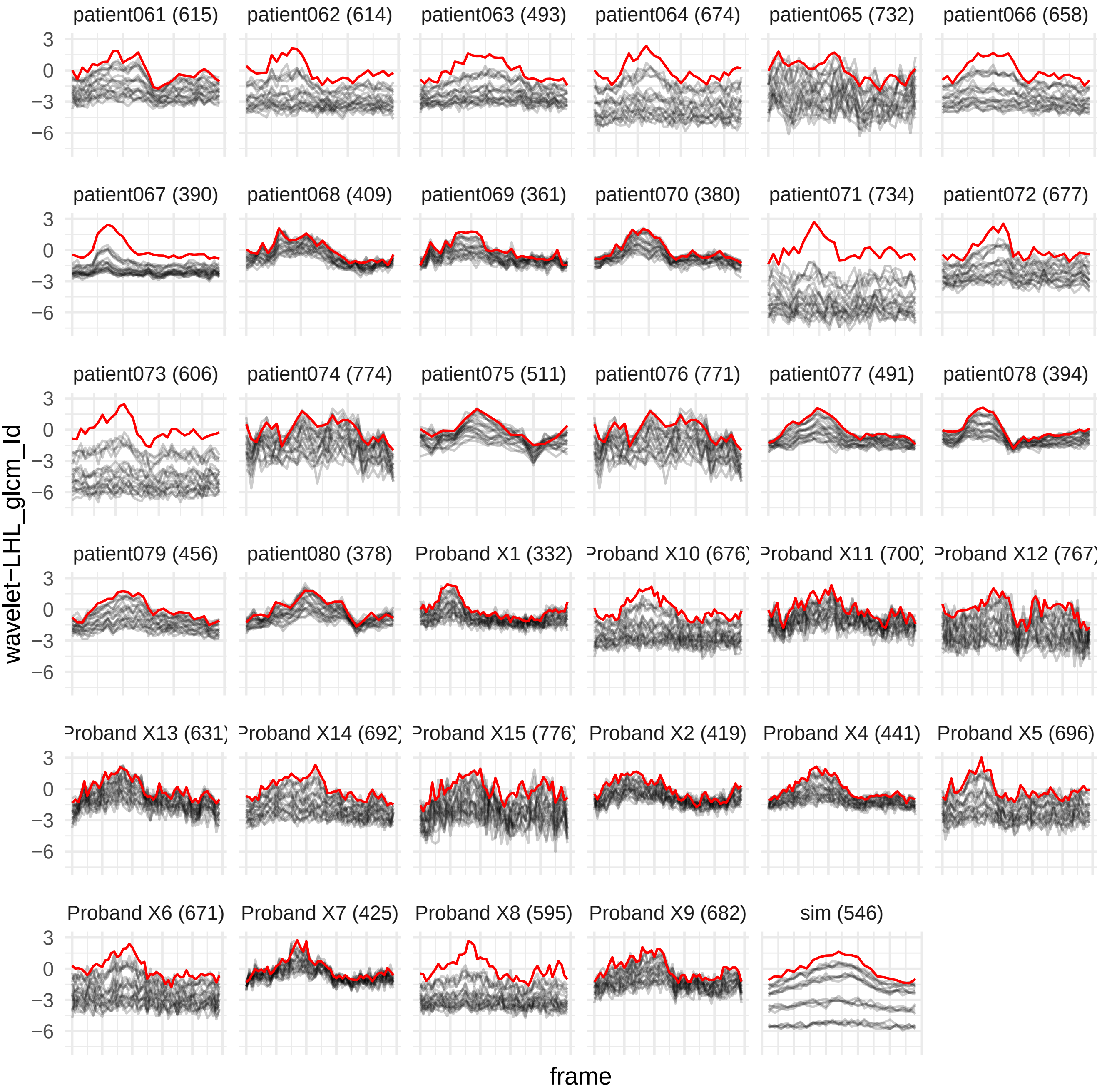

patient061 (615)
patient062 (614)
patient063 (493)
patient064 (674)
patient065 (732)
patient066 (658)
patient067 (390)
patient068 (409)
patient069 (361)
patient070 (380)
patient071 (734)
patient072 (677)
patient073 (606)
patient074 (774)
patient075 (511)
patient076 (771)
patient077 (491)
patient078 (394)
patient079 (456)
patient080 (378)
Proband X1 (332)
Proband X10 (676)
Proband X11 (700)
Proband X12 (767)
Proband X13 (631)
Proband X14 (692)
Proband X15 (776)
Proband X2 (419)
Proband X4 (441)
Proband X5 (696)
Proband X6 (671)
Proband X7 (425)
Proband X8 (595)
Proband X9 (682)
sim (546)
3
0
−3
−6
wavelet−LHL_glcm_Id
frame

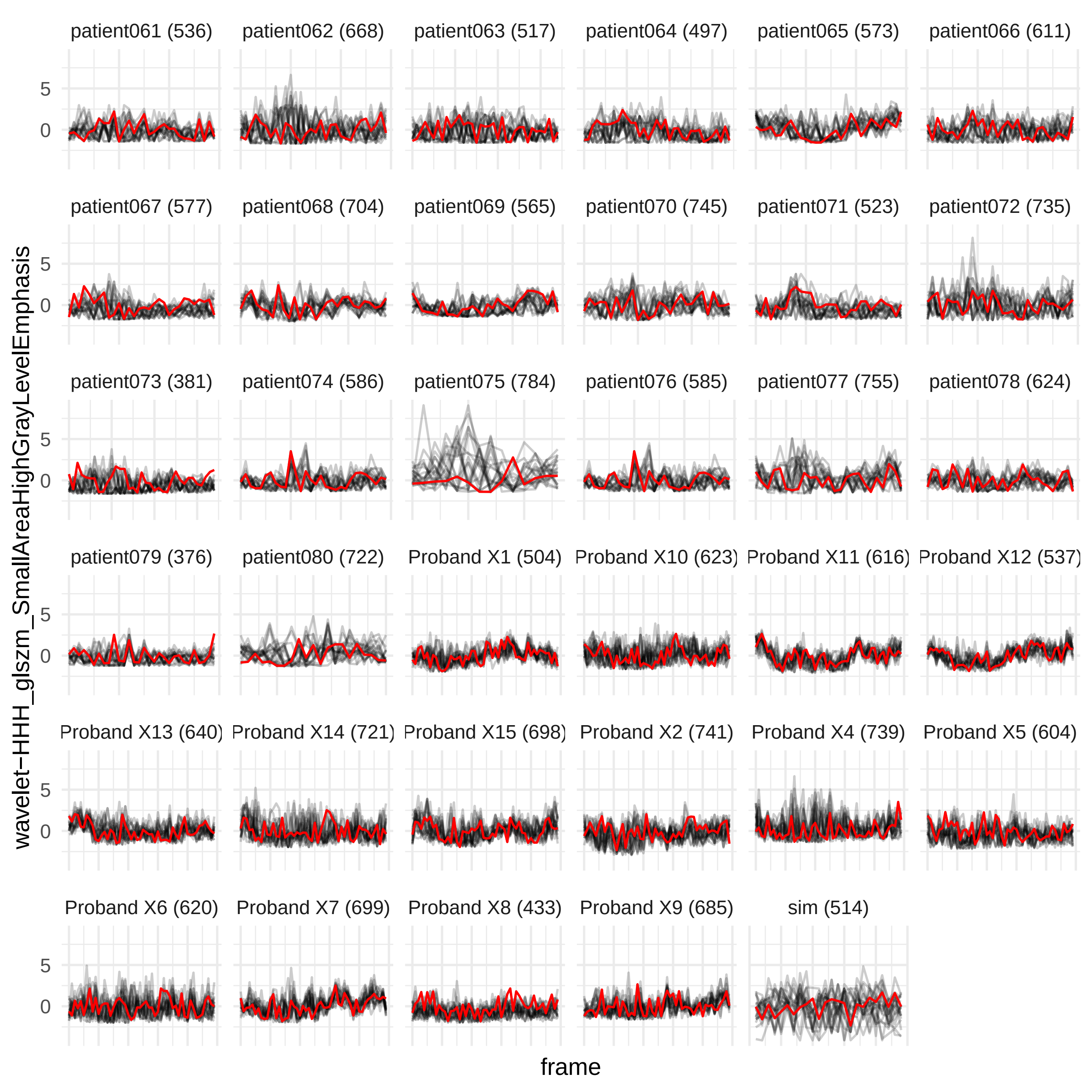
patient061 (536)
patient062 (668)
patient063 (517)
patient064 (497)
patient065 (573)
patient066 (611)
patient067 (577)
patient068 (704)
patient069 (565)
patient070 (745)
patient071 (523)
patient072 (735)
patient073 (381)
patient074 (586)
patient075 (784)
patient076 (585)
patient077 (755)
patient078 (624)
patient079 (376)
patient080 (722)
Proband X1 (504)
Proband X10 (623)
Proband X11 (616)
Proband X12 (537)
Proband X13 (640)
Proband X14 (721)
Proband X15 (698)
Proband X2 (741)
Proband X4 (739)
Proband X5 (604)
Proband X6 (620)
Proband X7 (699)
Proband X8 (433)
Proband X9 (685)
sim (514)
wavelet−HHH_glszm_SmallAreaHighGrayLevelEmphasis
frame

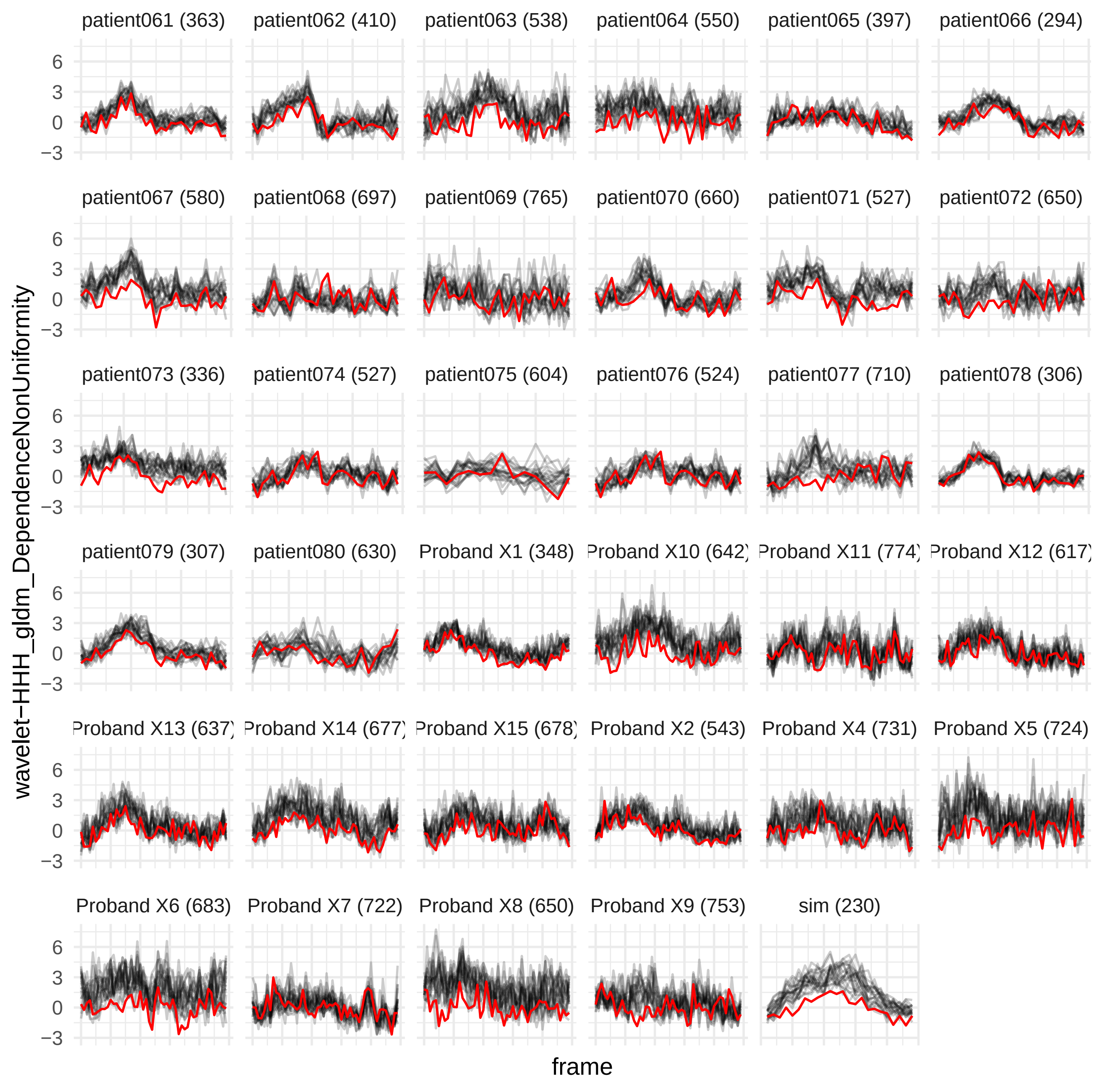
patient061 (363)
patient062 (410)
patient063 (538)
patient064 (550)
patient065 (397)
patient066 (294)
patient067 (580)
patient068 (697)
patient069 (765)
patient070 (660)
patient071 (527)
patient072 (650)
patient073 (336)
patient074 (527)
patient075 (604)
patient076 (524)
patient077 (710)
patient078 (306)
patient079 (307)
patient080 (630)
Proband X1 (348)
Proband X10 (642)
Proband X11 (774)
Proband X12 (617)
Proband X13 (637)
Proband X14 (677)
Proband X15 (678)
Proband X2 (543)
Proband X4 (731)
Proband X5 (724)
Proband X6 (683)
Proband X7 (722)
Proband X8 (650)
Proband X9 (753)
sim (230)
6
3
0
−3
wavelet−HHH_gldm_DependenceNonUniformity
frame

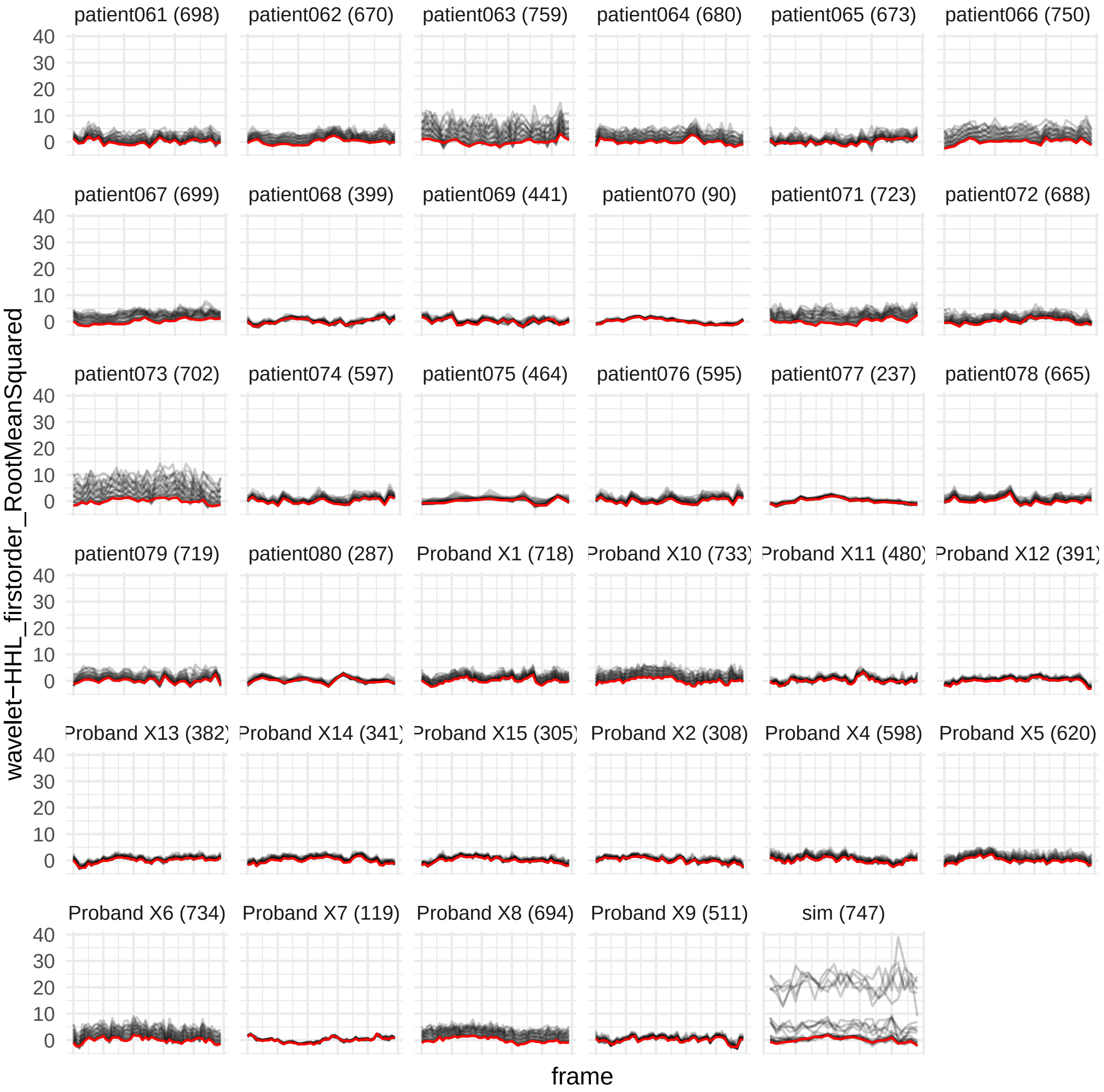

patient061 (698)
patient062 (670)
patient063 (759)
patient064 (680)
patient065 (673)
patient066 (750)
patient067 (699)
patient068 (399)
patient069 (441)
patient070 (90)
patient071 (723)
patient072 (688)
patient073 (702)
patient074 (597)
patient075 (464)
patient076 (595)
patient077 (237)
patient078 (665)
patient079 (719)
patient080 (287)
Proband X1 (718)
Proband X10 (733)
Proband X11 (480)
Proband X12 (391)
Proband X13 (382)
Proband X14 (341)
Proband X15 (305)
Proband X2 (308)
Proband X4 (598)
Proband X5 (620)
Proband X6 (734)
Proband X7 (119)
Proband X8 (694)
Proband X9 (511)
sim (747)
40
30
20
10
0
wavelet−HHL_firstorder_RootMeanSquared
frame

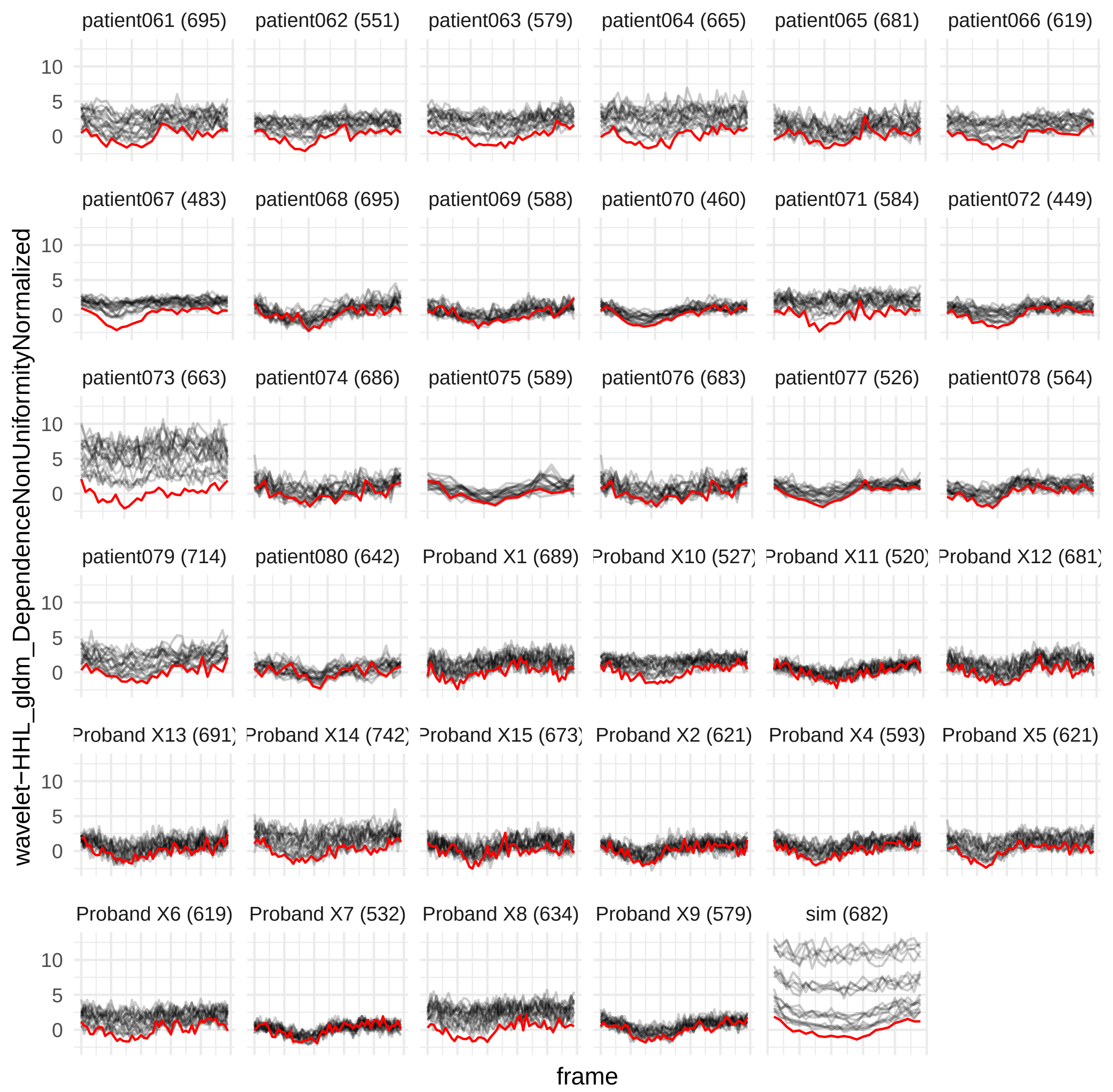

patient061 (695)
patient062 (551)
patient063 (579)
patient064 (665)
patient065 (681)
patient066 (619)
patient067 (483)
patient068 (695)
patient069 (588)
patient070 (460)
patient071 (584)
patient072 (449)
patient073 (663)
patient074 (686)
patient075 (589)
patient076 (683)
patient077 (526)
patient078 (564)
patient079 (714)
patient080 (642)
Proband X1 (689)
Proband X10 (527)
Proband X11 (520)
Proband X12 (681)
Proband X13 (691)
Proband X14 (742)
Proband X15 (673)
Proband X2 (621)
Proband X4 (593)
Proband X5 (621)
Proband X6 (619)
Proband X7 (532)
Proband X8 (634)
Proband X9 (579)
sim (682)
wavelet−HHL_gldm_DependenceNonUniformityNormalized
frame

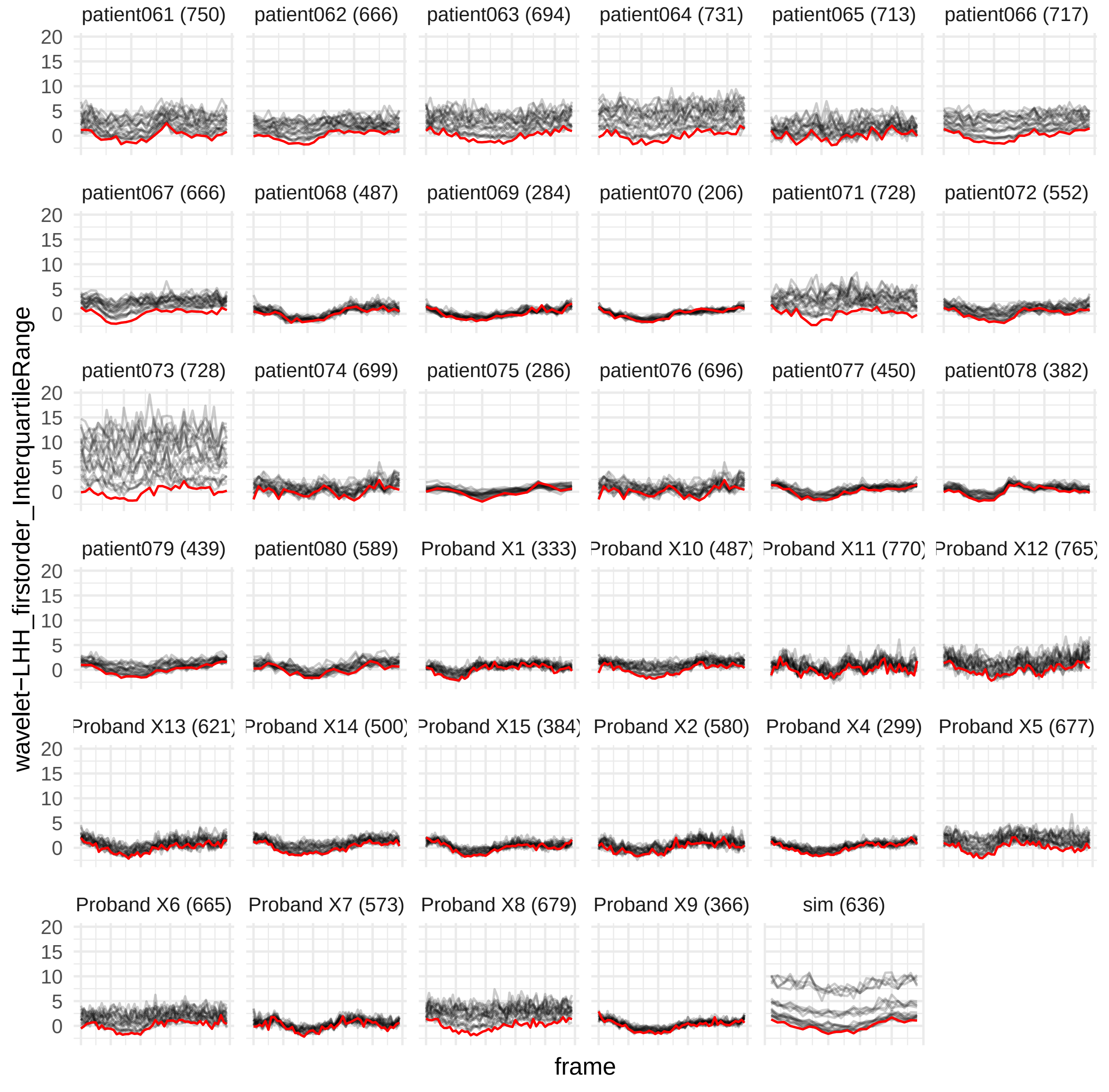

patient061 (750)
patient062 (666)
patient063 (694)
patient064 (731)
patient065 (713)
patient066 (717)
patient067 (666)
patient068 (487)
patient069 (284)
patient070 (206)
patient071 (728)
patient072 (552)
patient073 (728)
patient074 (699)
patient075 (286)
patient076 (696)
patient077 (450)
patient078 (382)
patient079 (439)
patient080 (589)
Proband X1 (333)
Proband X10 (487)
Proband X11 (770)
Proband X12 (765)
Proband X13 (621)
Proband X14 (500)
Proband X15 (384)
Proband X2 (580)
Proband X4 (299)
Proband X5 (677)
Proband X6 (665)
Proband X7 (573)
Proband X8 (679)
Proband X9 (366)
sim (636)
wavelet−LHH_firstorder_InterquartileRange
frame
0
5
10
15
20

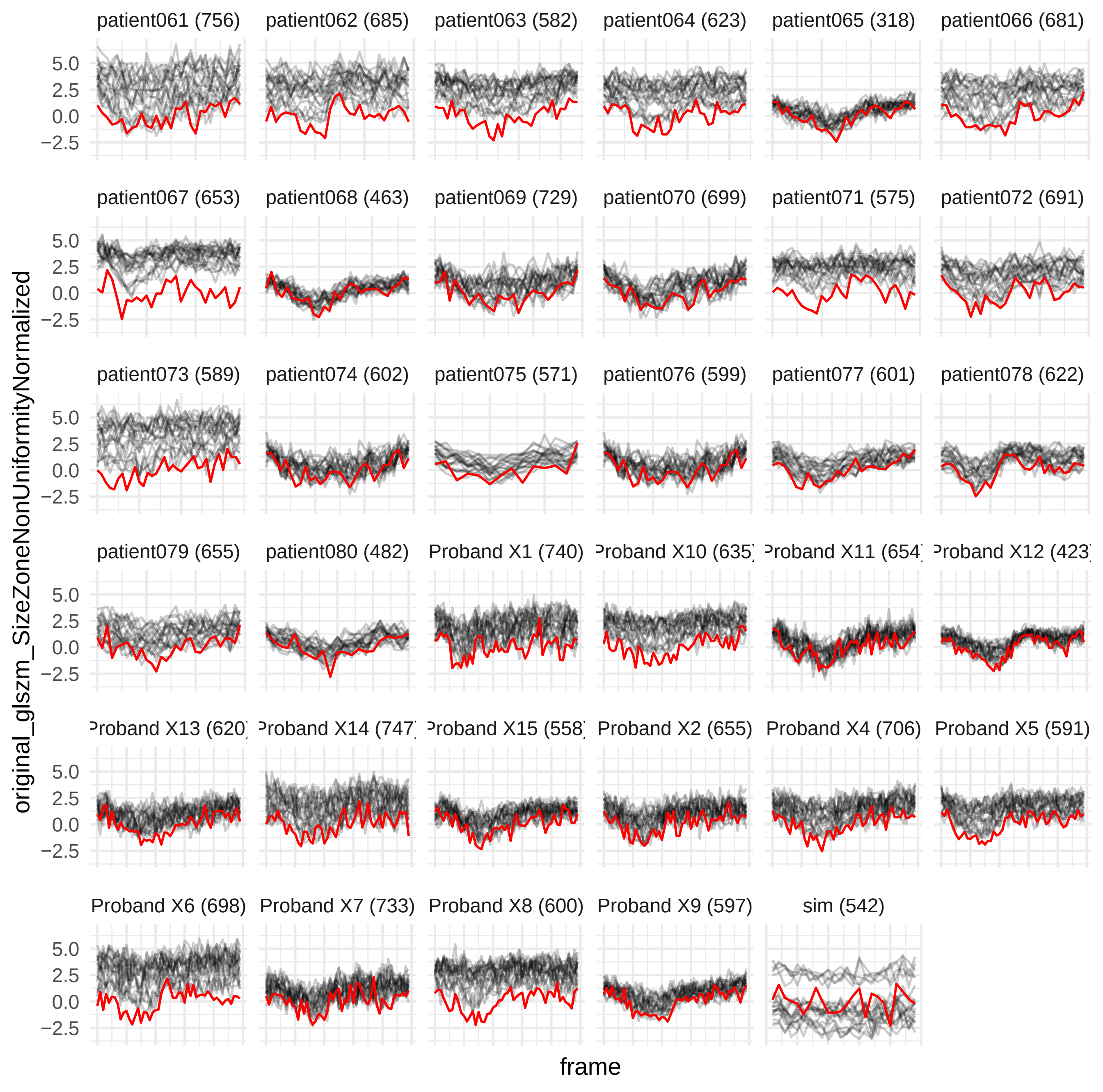
patient061 (756)
patient062 (685)
patient063 (582)
patient064 (623)
patient065 (318)
patient066 (681)
patient067 (653)
patient068 (463)
patient069 (729)
patient070 (699)
patient071 (575)
patient072 (691)
patient073 (589)
patient074 (602)
patient075 (571)
patient076 (599)
patient077 (601)
patient078 (622)
patient079 (655)
patient080 (482)
Proband X1 (740)
Proband X10 (635)
Proband X11 (654)
Proband X12 (423)
Proband X13 (620)
Proband X14 (747)
Proband X15 (558)
Proband X2 (655)
Proband X4 (706)
Proband X5 (591)
Proband X6 (698)
Proband X7 (733)
Proband X8 (600)
Proband X9 (597)
sim (542)
5.0
2.5
0.0
−2.5
original_glszm_SizeZoneNonUniformityNormalized
frame

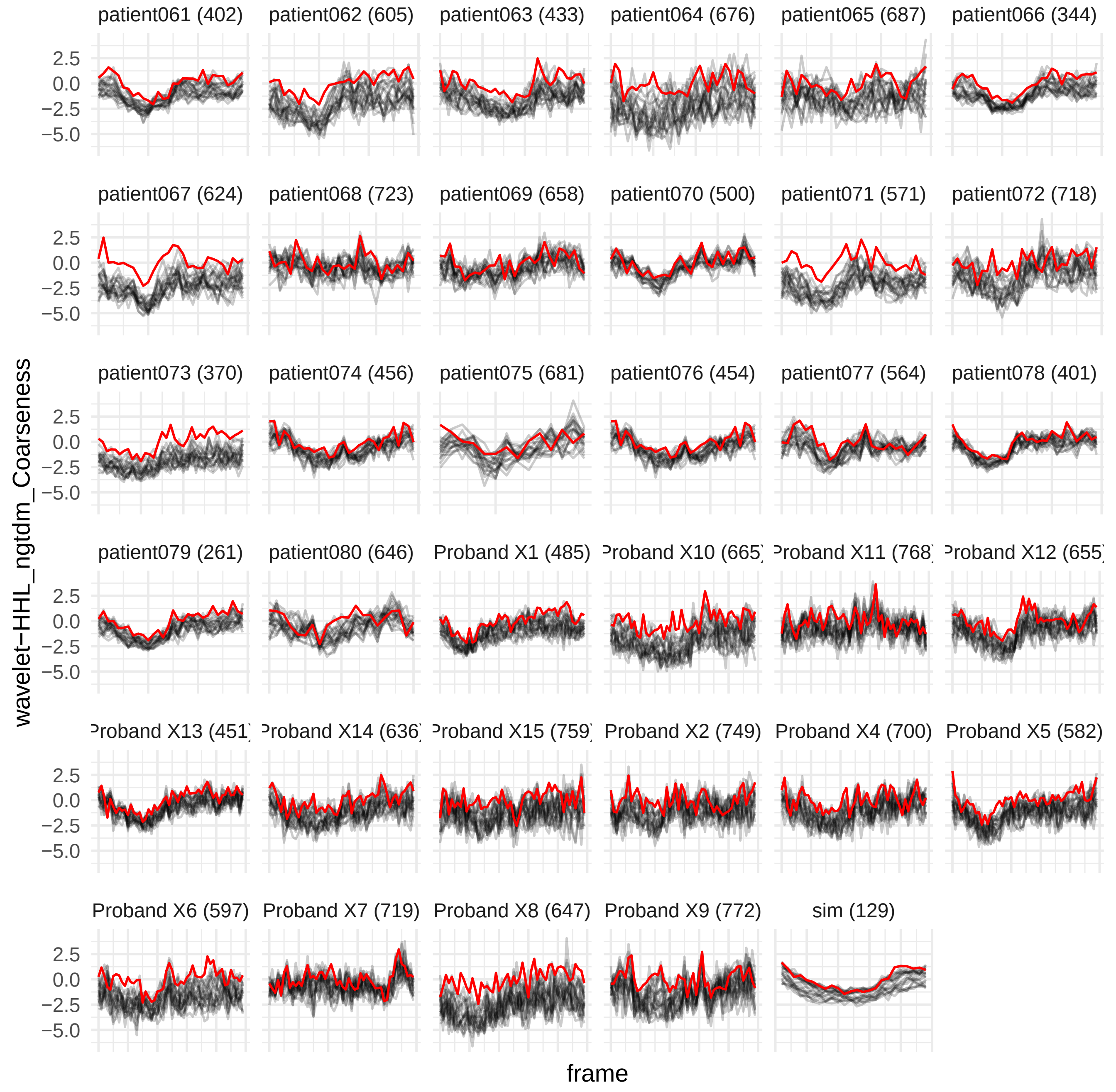
patient061 (402)
patient062 (605)
patient063 (433)
patient064 (676)
patient065 (687)
patient066 (344)
patient067 (624)
patient068 (723)
patient069 (658)
patient070 (500)
patient071 (571)
patient072 (718)
patient073 (370)
patient074 (456)
patient075 (681)
patient076 (454)
patient077 (564)
patient078 (401)
patient079 (261)
patient080 (646)
Proband X1 (485)
Proband X10 (665)
Proband X11 (768)
Proband X12 (655)
Proband X13 (451)
Proband X14 (636)
Proband X15 (759)
Proband X2 (749)
Proband X4 (700)
Proband X5 (582)
Proband X6 (597)
Proband X7 (719)
Proband X8 (647)
Proband X9 (772)
sim (129)
2.5
0.0
−2.5
−5.0
wavelet−HHL_ngtdm_Coarseness
frame

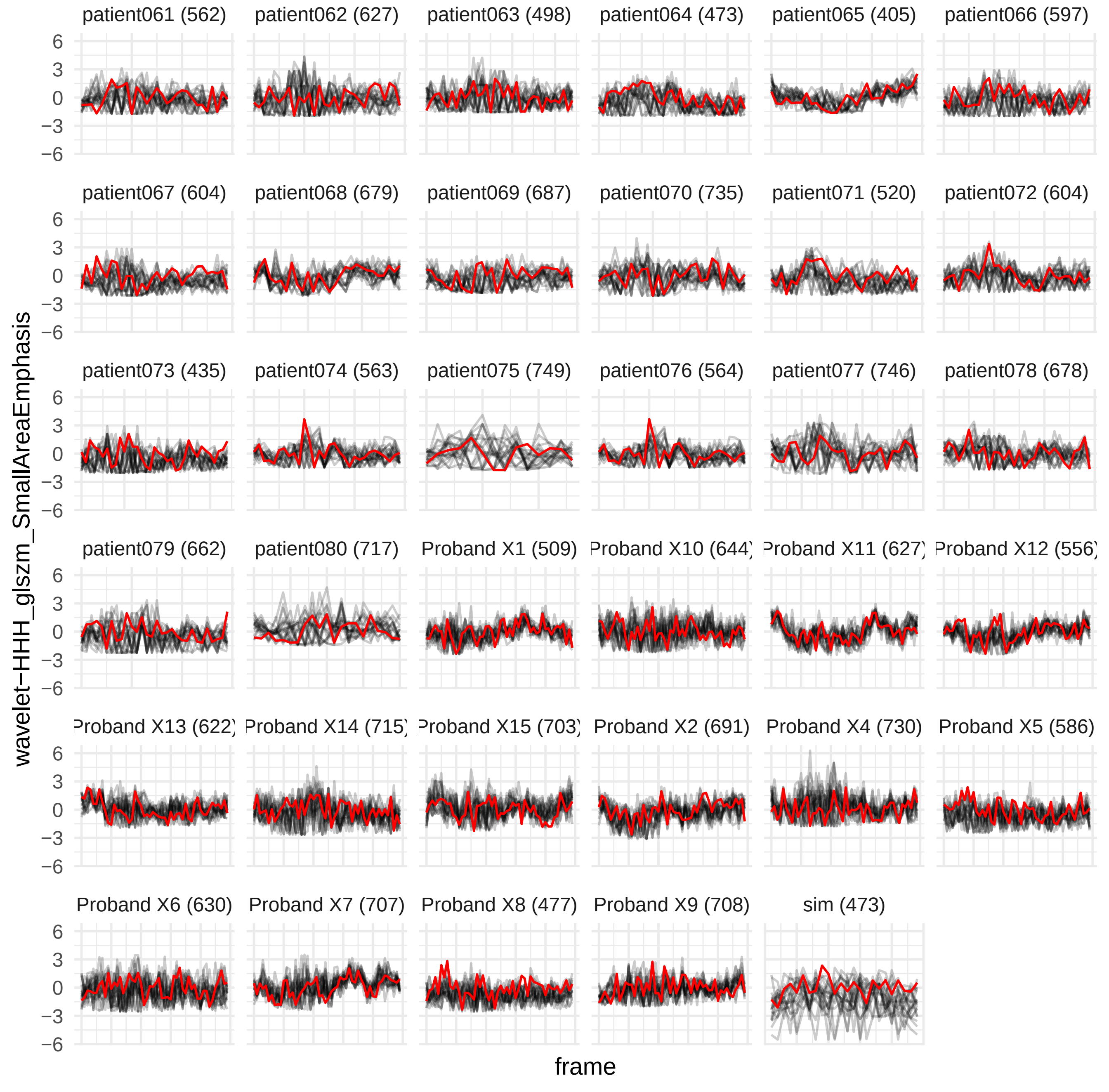
patient061 (562)
patient062 (627)
patient063 (498)
patient064 (473)
patient065 (405)
patient066 (597)
patient067 (604)
patient068 (679)
patient069 (687)
patient070 (735)
patient071 (520)
patient072 (604)
patient073 (435)
patient074 (563)
patient075 (749)
patient076 (564)
patient077 (746)
patient078 (678)
patient079 (662)
patient080 (717)
Proband X1 (509)
Proband X10 (644)
Proband X11 (627)
Proband X12 (556)
Proband X13 (622)
Proband X14 (715)
Proband X15 (703)
Proband X2 (691)
Proband X4 (730)
Proband X5 (586)
Proband X6 (630)
Proband X7 (707)
Proband X8 (477)
Proband X9 (708)
sim (473)
6
3
0
−3
−6
wavelet−HHH_glszm_SmallAreaEmphasis
frame

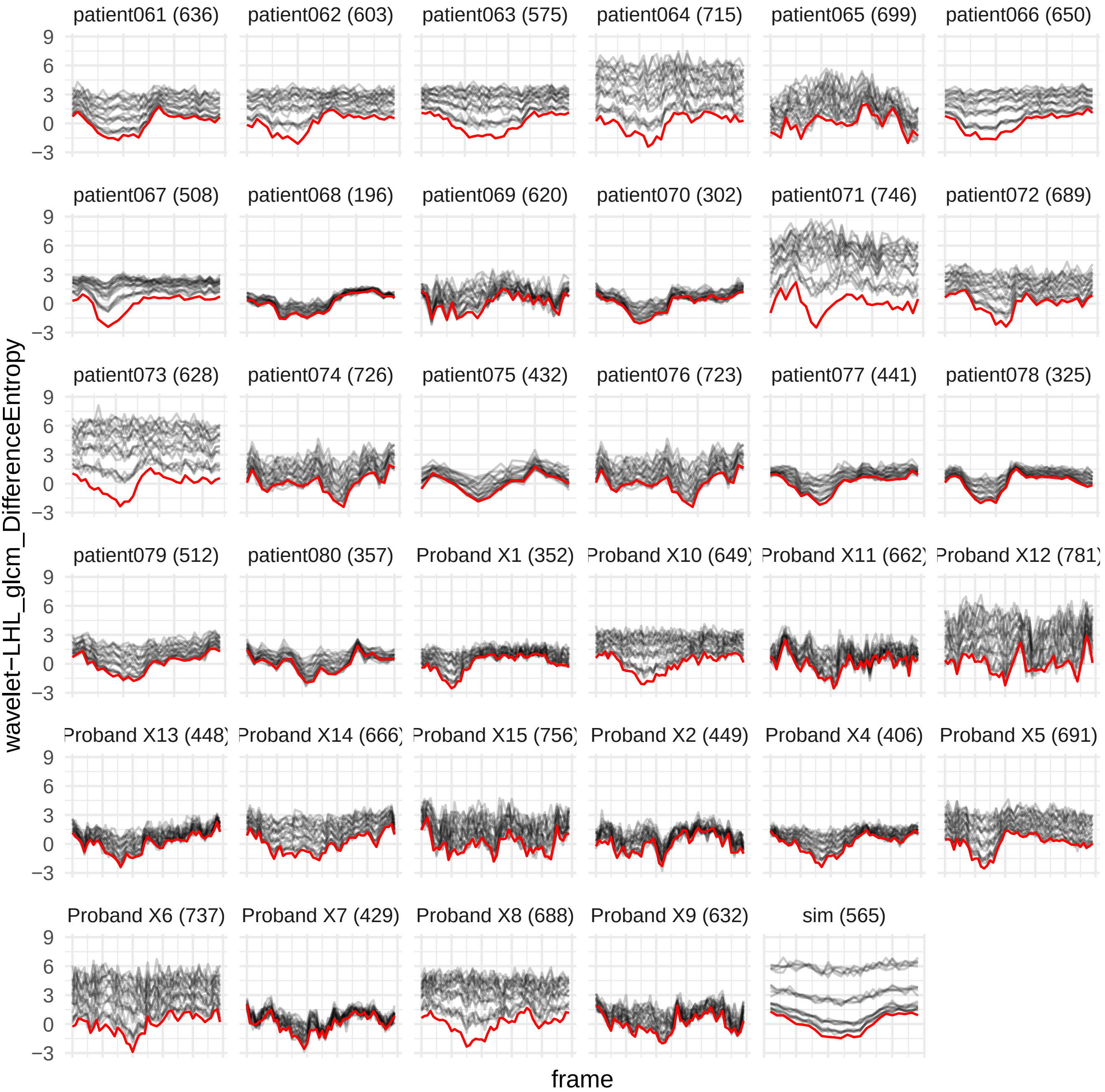

patient061 (636)
patient062 (603)
patient063 (575)
patient064 (715)
patient065 (699)
patient066 (650)
patient067 (508)
patient068 (196)
patient069 (620)
patient070 (302)
patient071 (746)
patient072 (689)
patient073 (628)
patient074 (726)
patient075 (432)
patient076 (723)
patient077 (441)
patient078 (325)
patient079 (512)
patient080 (357)
Proband X1 (352)
Proband X10 (649)
Proband X11 (662)
Proband X12 (781)
Proband X13 (448)
Proband X14 (666)
Proband X15 (756)
Proband X2 (449)
Proband X4 (406)
Proband X5 (691)
Proband X6 (737)
Proband X7 (429)
Proband X8 (688)
Proband X9 (632)
sim (565)
9
6
3
0
−3
wavelet−LHL_glcm_DifferenceEntropy
frame

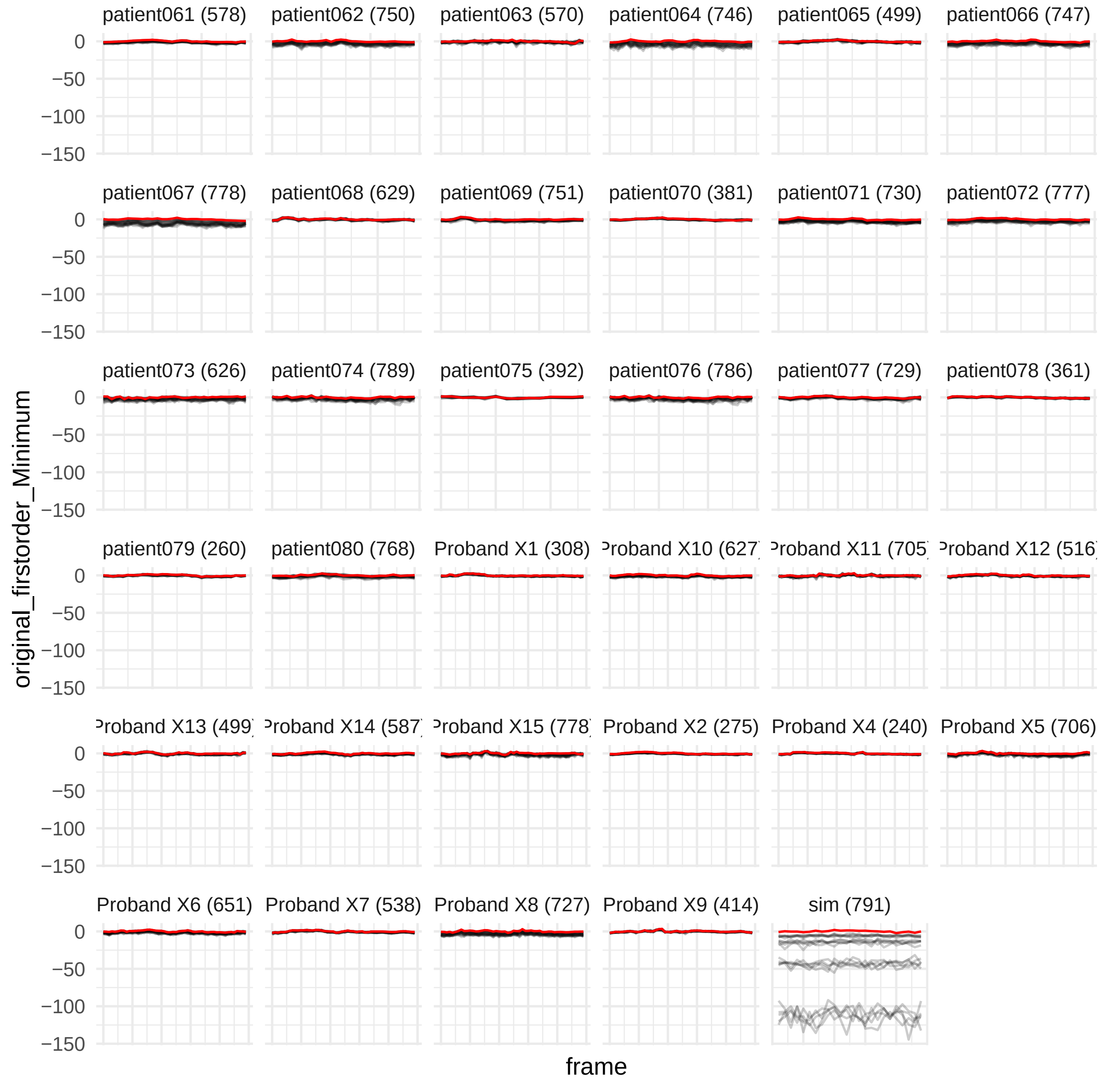

patient061 (578)
patient062 (750)
patient063 (570)
patient064 (746)
patient065 (499)
patient066 (747)
patient067 (778)
patient068 (629)
patient069 (751)
patient070 (381)
patient071 (730)
patient072 (777)
patient073 (626)
patient074 (789)
patient075 (392)
patient076 (786)
patient077 (729)
patient078 (361)
patient079 (260)
patient080 (768)
Proband X1 (308)
Proband X10 (627)
Proband X11 (705)
Proband X12 (516)
Proband X13 (499)
Proband X14 (587)
Proband X15 (778)
Proband X2 (275)
Proband X4 (240)
Proband X5 (706)
Proband X6 (651)
Proband X7 (538)
Proband X8 (727)
Proband X9 (414)
sim (791)
0
−50
−100
−150
original_firstorder_Minimum
frame

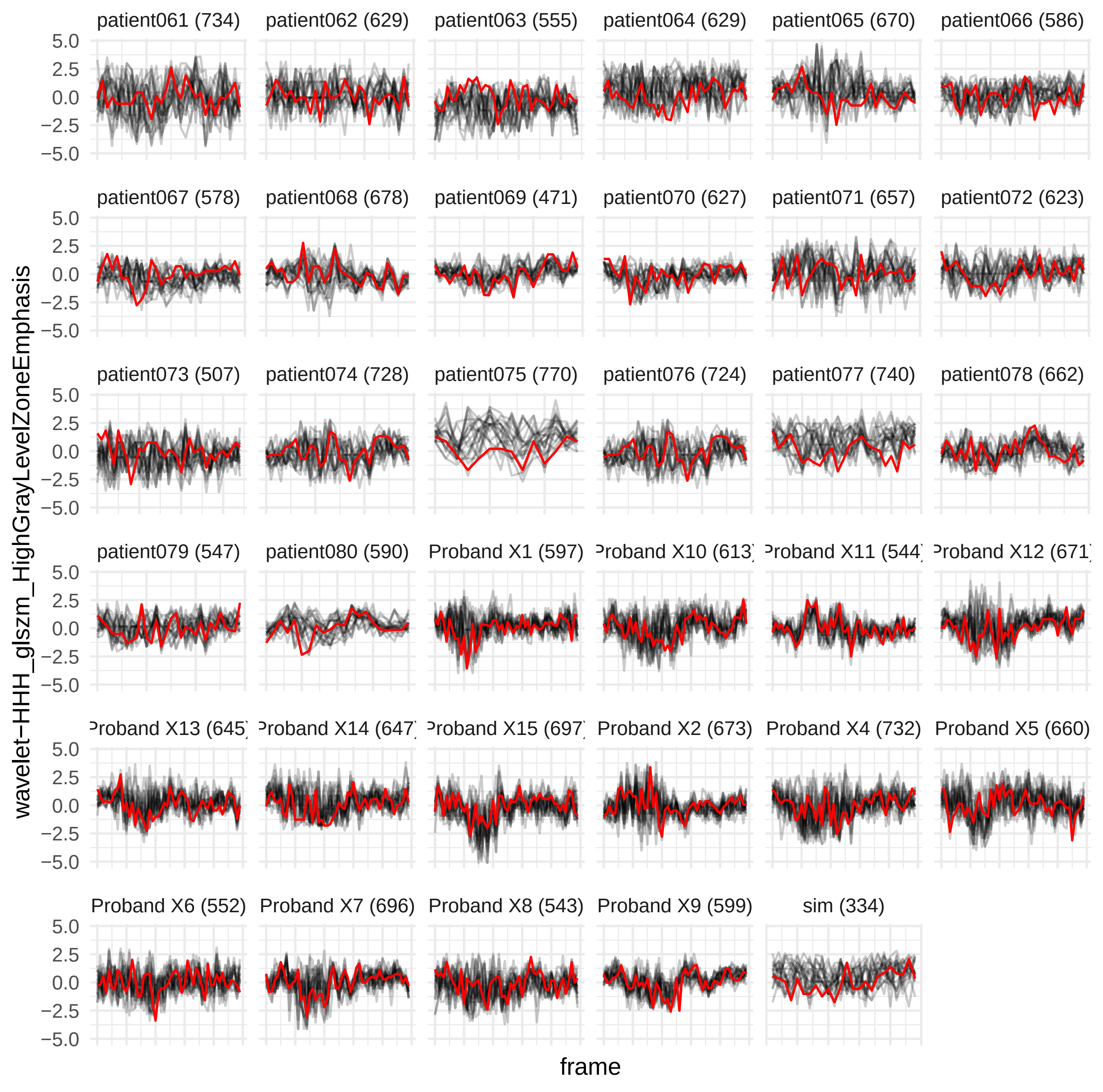

patient061 (734)
patient062 (629)
patient063 (555)
patient064 (629)
patient065 (670)
patient066 (586)
patient067 (578)
patient068 (678)
patient069 (471)
patient070 (627)
patient071 (657)
patient072 (623)
patient073 (507)
patient074 (728)
patient075 (770)
patient076 (724)
patient077 (740)
patient078 (662)
patient079 (547)
patient080 (590)
Proband X1 (597)
Proband X10 (613)
Proband X11 (544)
Proband X12 (671)
Proband X13 (645)
Proband X14 (647)
Proband X15 (697)
Proband X2 (673)
Proband X4 (732)
Proband X5 (660)
Proband X6 (552)
Proband X7 (696)
Proband X8 (543)
Proband X9 (599)
sim (334)
5.0
2.5
0.0
−2.5
−5.0
wavelet−HHH_glszm_HighGrayLevelZoneEmphasis
frame

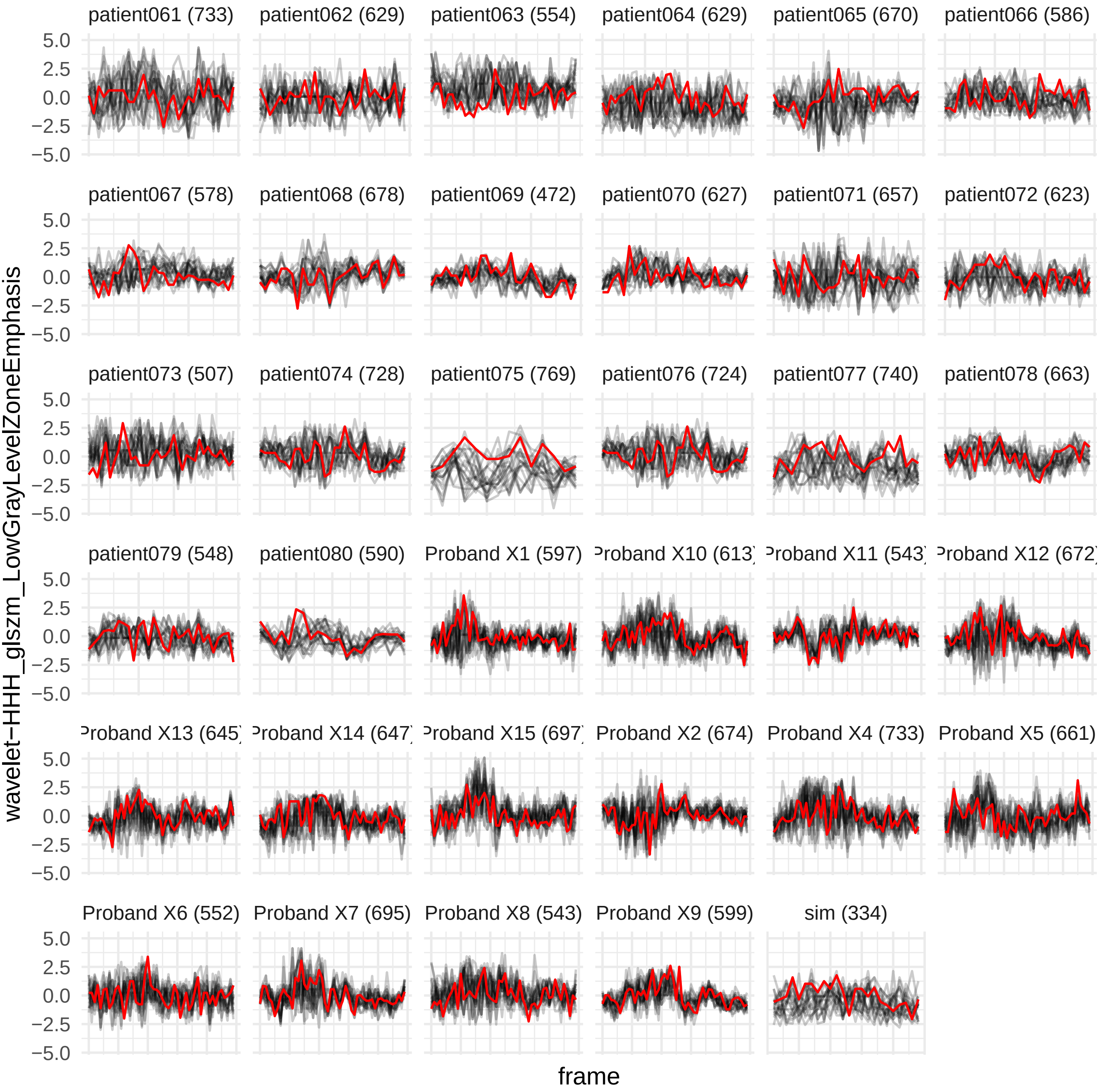

patient061 (733)
patient062 (629)
patient063 (554)
patient064 (629)
patient065 (670)
patient066 (586)
patient067 (578)
patient068 (678)
patient069 (472)
patient070 (627)
patient071 (657)
patient072 (623)
patient073 (507)
patient074 (728)
patient075 (769)
patient076 (724)
patient077 (740)
patient078 (663)
patient079 (548)
patient080 (590)
Proband X1 (597)
Proband X10 (613)
Proband X11 (543)
Proband X12 (672)
Proband X13 (645)
Proband X14 (647)
Proband X15 (697)
Proband X2 (674)
Proband X4 (733)
Proband X5 (661)
Proband X6 (552)
Proband X7 (695)
Proband X8 (543)
Proband X9 (599)
sim (334)
5.0
2.5
0.0
−2.5
−5.0
wavelet−HHH_glszm_LowGrayLevelZoneEmphasis
frame

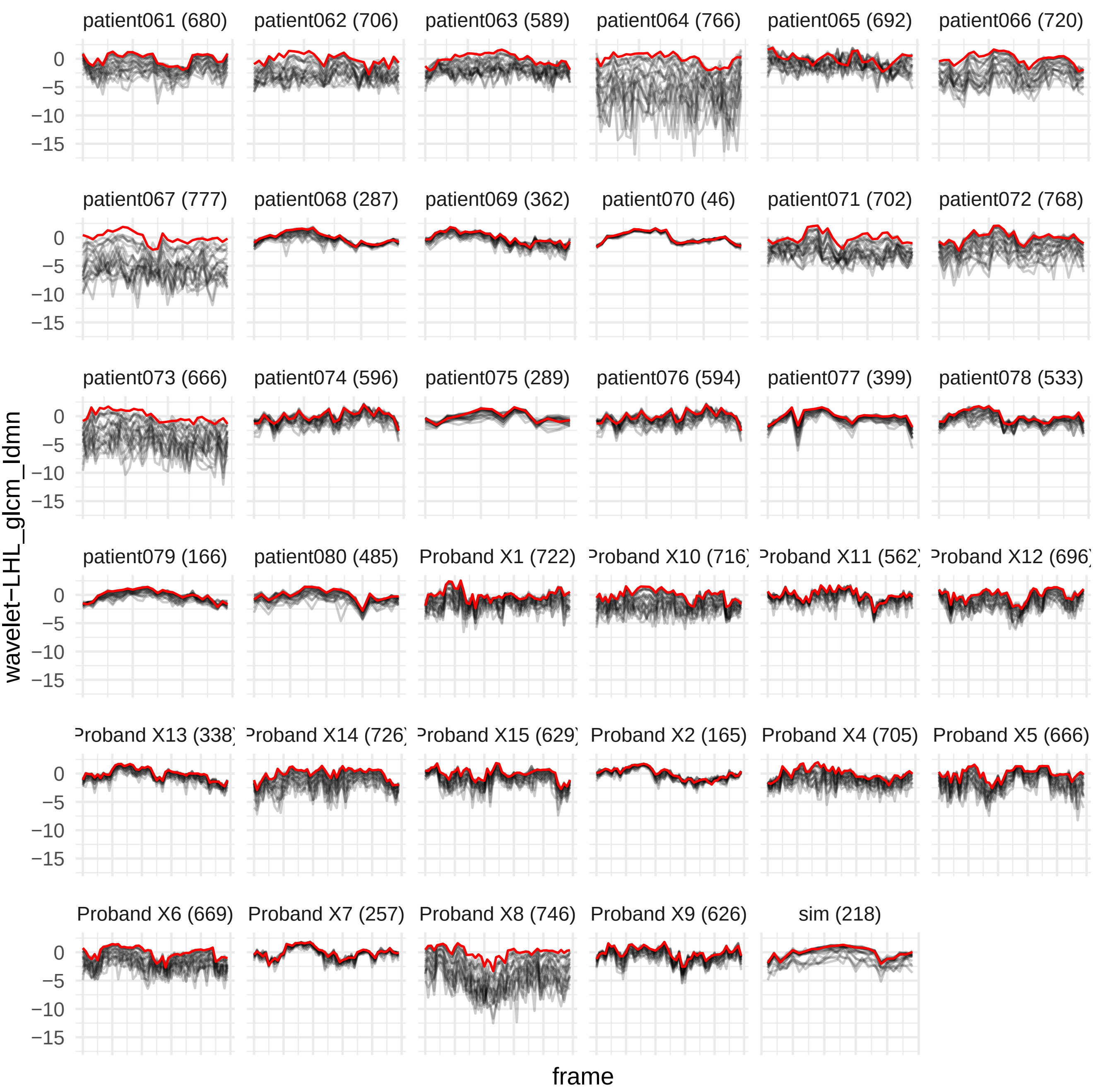

patient061 (680)
patient062 (706)
patient063 (589)
patient064 (766)
patient065 (692)
patient066 (720)
patient067 (777)
patient068 (287)
patient069 (362)
patient070 (46)
patient071 (702)
patient072 (768)
patient073 (666)
patient074 (596)
patient075 (289)
patient076 (594)
patient077 (399)
patient078 (533)
patient079 (166)
patient080 (485)
Proband X1 (722)
Proband X10 (716)
Proband X11 (562)
Proband X12 (696)
Proband X13 (338)
Proband X14 (726)
Proband X15 (629)
Proband X2 (165)
Proband X4 (705)
Proband X5 (666)
Proband X6 (669)
Proband X7 (257)
Proband X8 (746)
Proband X9 (626)
sim (218)
0
−5
−10
−15
wavelet−LHL_glcm_Idmn
frame

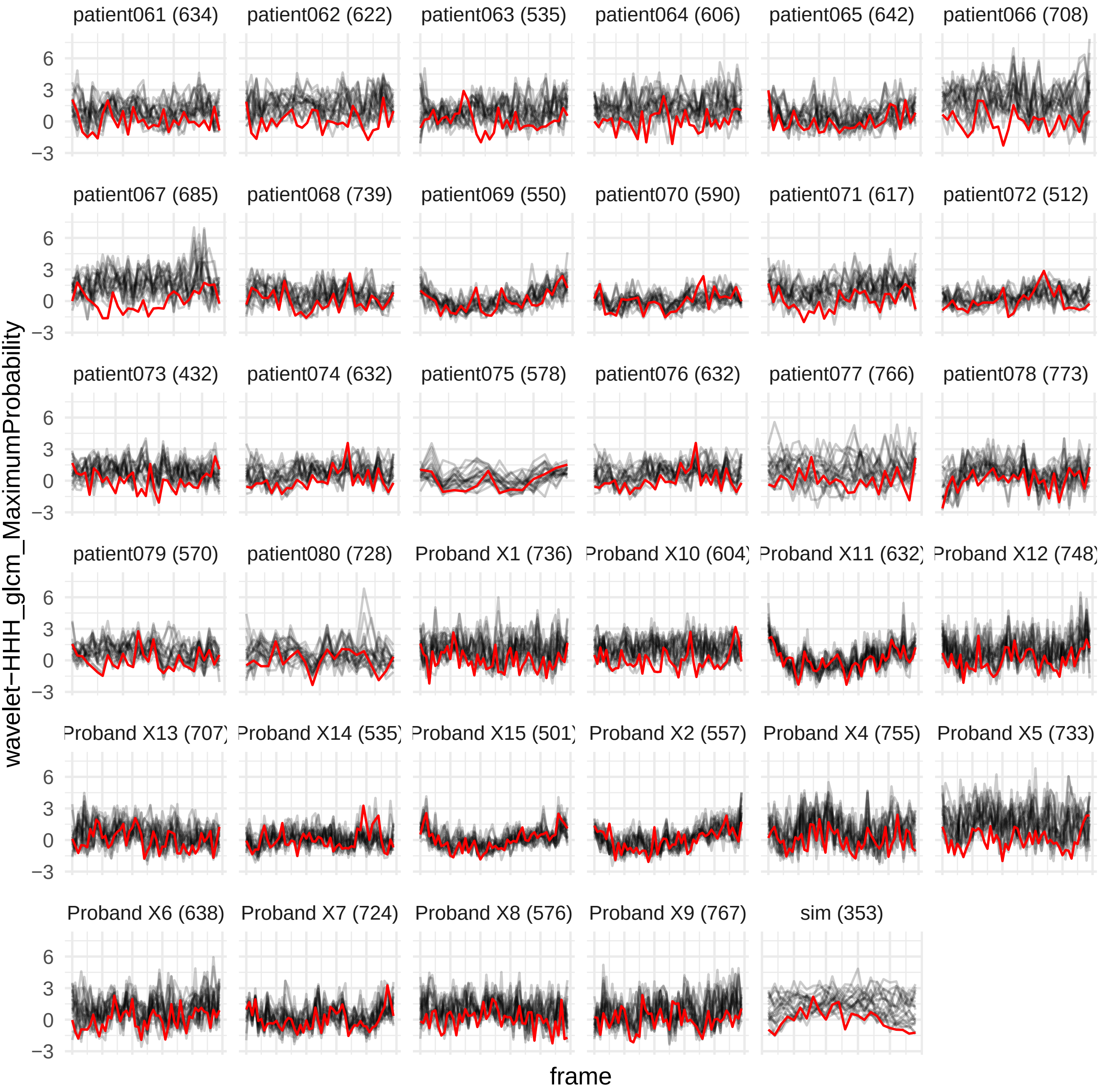

patient061 (634)
patient062 (622)
patient063 (535)
patient064 (606)
patient065 (642)
patient066 (708)
patient067 (685)
patient068 (739)
patient069 (550)
patient070 (590)
patient071 (617)
patient072 (512)
patient073 (432)
patient074 (632)
patient075 (578)
patient076 (632)
patient077 (766)
patient078 (773)
patient079 (570)
patient080 (728)
Proband X1 (736)
Proband X10 (604)
Proband X11 (632)
Proband X12 (748)
Proband X13 (707)
Proband X14 (535)
Proband X15 (501)
Proband X2 (557)
Proband X4 (755)
Proband X5 (733)
Proband X6 (638)
Proband X7 (724)
Proband X8 (576)
Proband X9 (767)
sim (353)
6
3
0
−3
wavelet−HHH_glcm_MaximumProbability
frame

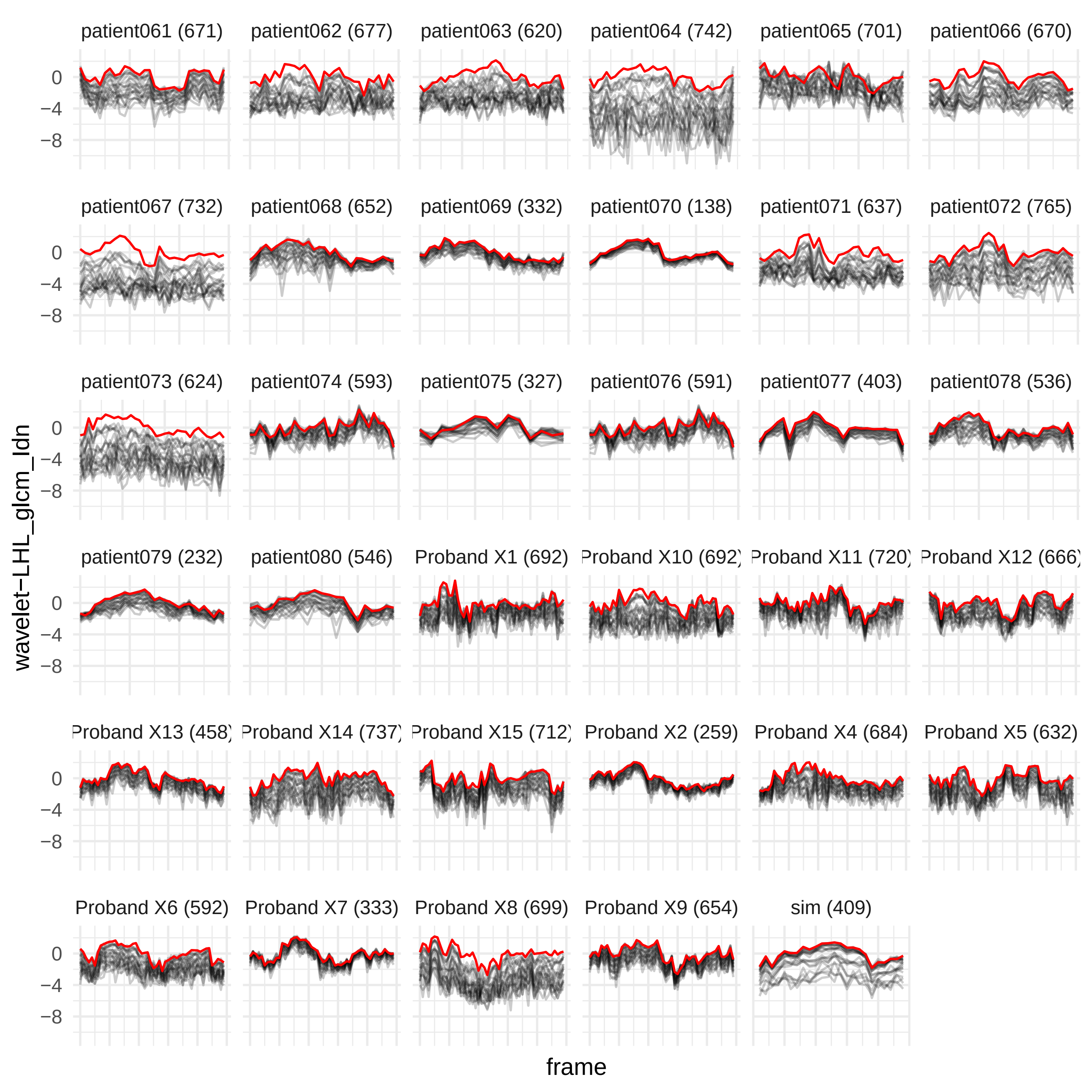

patient061 (671)
patient062 (677)
patient063 (620)
patient064 (742)
patient065 (701)
patient066 (670)
patient067 (732)
patient068 (652)
patient069 (332)
patient070 (138)
patient071 (637)
patient072 (765)
patient073 (624)
patient074 (593)
patient075 (327)
patient076 (591)
patient077 (403)
patient078 (536)
patient079 (232)
patient080 (546)
Proband X1 (692)
Proband X10 (692)
Proband X11 (720)
Proband X12 (666)
Proband X13 (458)
Proband X14 (737)
Proband X15 (712)
Proband X2 (259)
Proband X4 (684)
Proband X5 (632)
Proband X6 (592)
Proband X7 (333)
Proband X8 (699)
Proband X9 (654)
sim (409)
wavelet−LHL_glcm_Idn
0
−4
−8
frame

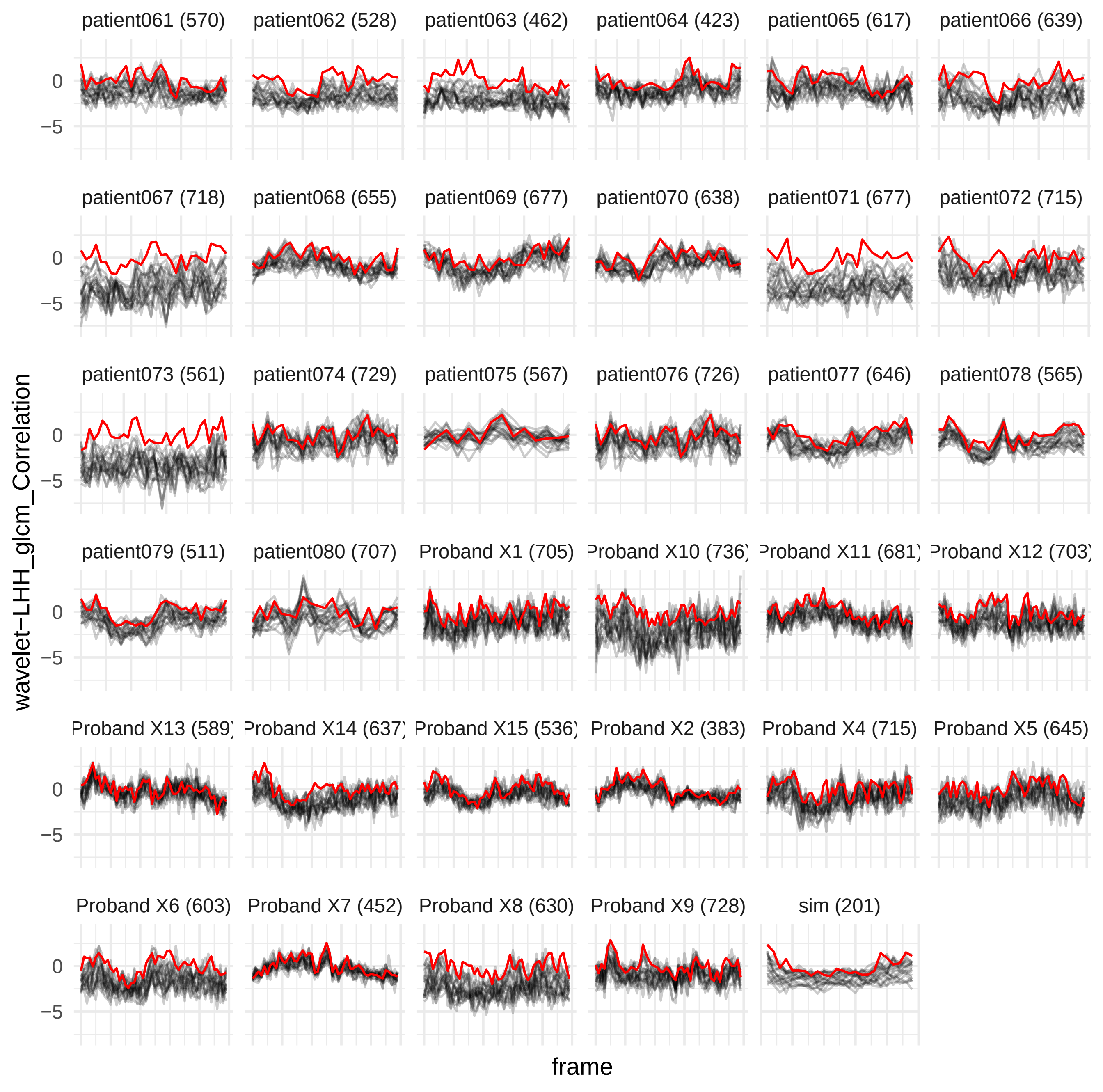
patient061 (570)
patient062 (528)
patient063 (462)
patient064 (423)
patient065 (617)
patient066 (639)
patient067 (718)
patient068 (655)
patient069 (677)
patient070 (638)
patient071 (677)
patient072 (715)
patient073 (561)
patient074 (729)
patient075 (567)
patient076 (726)
patient077 (646)
patient078 (565)
patient079 (511)
patient080 (707)
Proband X1 (705)
Proband X10 (736)
Proband X11 (681)
Proband X12 (703)
Proband X13 (589)
Proband X14 (637)
Proband X15 (536)
Proband X2 (383)
Proband X4 (715)
Proband X5 (645)
Proband X6 (603)
Proband X7 (452)
Proband X8 (630)
Proband X9 (728)
sim (201)
0
−5
wavelet−LHH_glcm_Correlation
frame

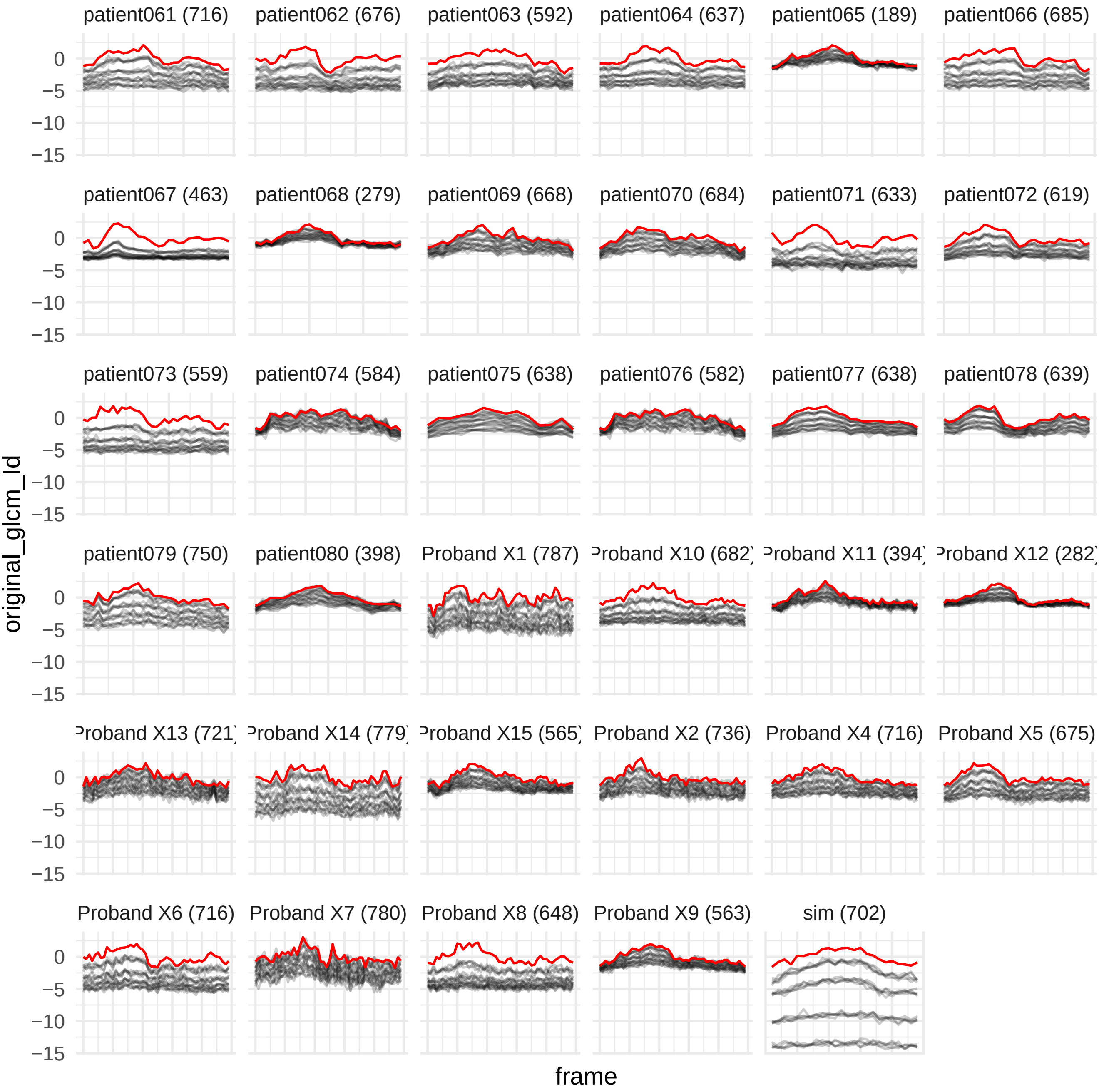

patient061 (716)
patient062 (676)
patient063 (592)
patient064 (637)
patient065 (189)
patient066 (685)
patient067 (463)
patient068 (279)
patient069 (668)
patient070 (684)
patient071 (633)
patient072 (619)
patient073 (559)
patient074 (584)
patient075 (638)
patient076 (582)
patient077 (638)
patient078 (639)
patient079 (750)
patient080 (398)
Proband X1 (787)
Proband X10 (682)
Proband X11 (394)
Proband X12 (282)
Proband X13 (721)
Proband X14 (779)
Proband X15 (565)
Proband X2 (736)
Proband X4 (716)
Proband X5 (675)
Proband X6 (716)
Proband X7 (780)
Proband X8 (648)
Proband X9 (563)
sim (702)
0
−5
−10
−15
original_glcm_Id
frame

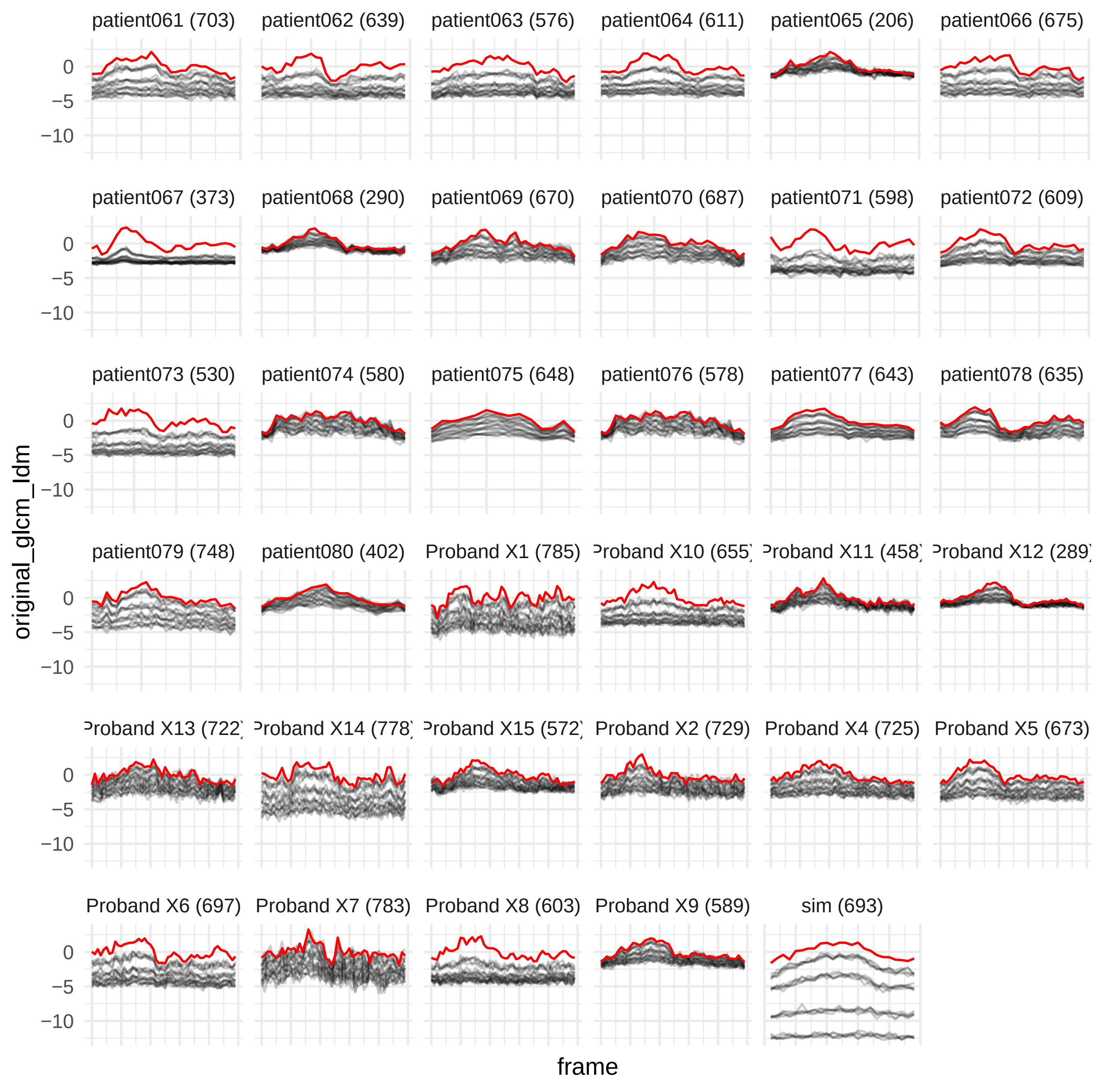
patient061 (703)
patient062 (639)
patient063 (576)
patient064 (611)
patient065 (206)
patient066 (675)
patient067 (373)
patient068 (290)
patient069 (670)
patient070 (687)
patient071 (598)
patient072 (609)
patient073 (530)
patient074 (580)
patient075 (648)
patient076 (578)
patient077 (643)
patient078 (635)
patient079 (748)
patient080 (402)
Proband X1 (785)
Proband X10 (655)
Proband X11 (458)
Proband X12 (289)
Proband X13 (722)
Proband X14 (778)
Proband X15 (572)
Proband X2 (729)
Proband X4 (725)
Proband X5 (673)
Proband X6 (697)
Proband X7 (783)
Proband X8 (603)
Proband X9 (589)
sim (693)
0
−5
−10
original_glcm_ldm
frame

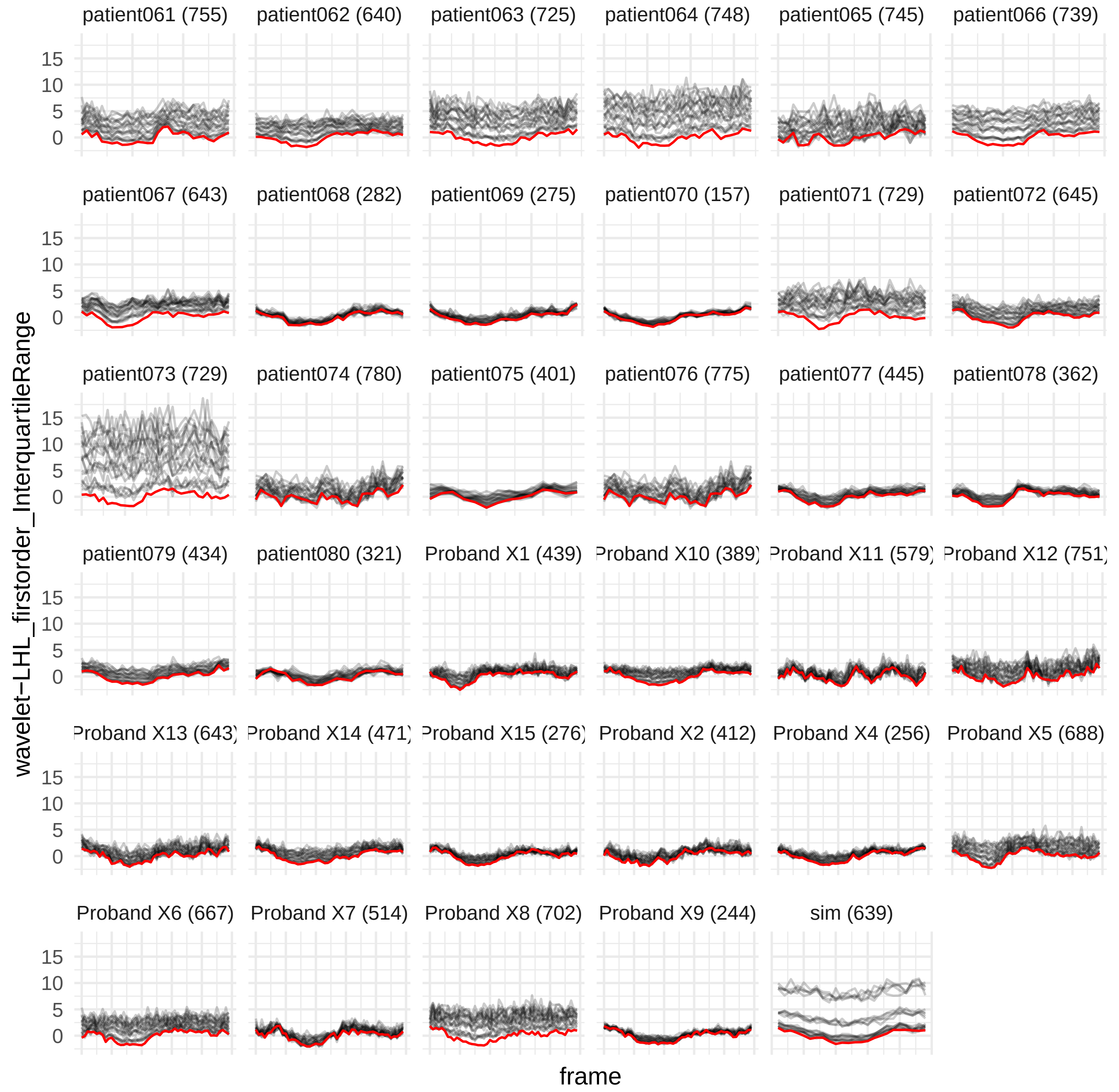
patient061 (755)
patient062 (640)
patient063 (725)
patient064 (748)
patient065 (745)
patient066 (739)
patient067 (643)
patient068 (282)
patient069 (275)
patient070 (157)
patient071 (729)
patient072 (645)
patient073 (729)
patient074 (780)
patient075 (401)
patient076 (775)
patient077 (445)
patient078 (362)
patient079 (434)
patient080 (321)
Proband X1 (439)
Proband X10 (389)
Proband X11 (579)
Proband X12 (751)
Proband X13 (643)
Proband X14 (471)
Proband X15 (276)
Proband X2 (412)
Proband X4 (256)
Proband X5 (688)
Proband X6 (667)
Proband X7 (514)
Proband X8 (702)
Proband X9 (244)
sim (639)
wavelet−LHL_firstorder_InterquartileRange
frame
15
10
5
0

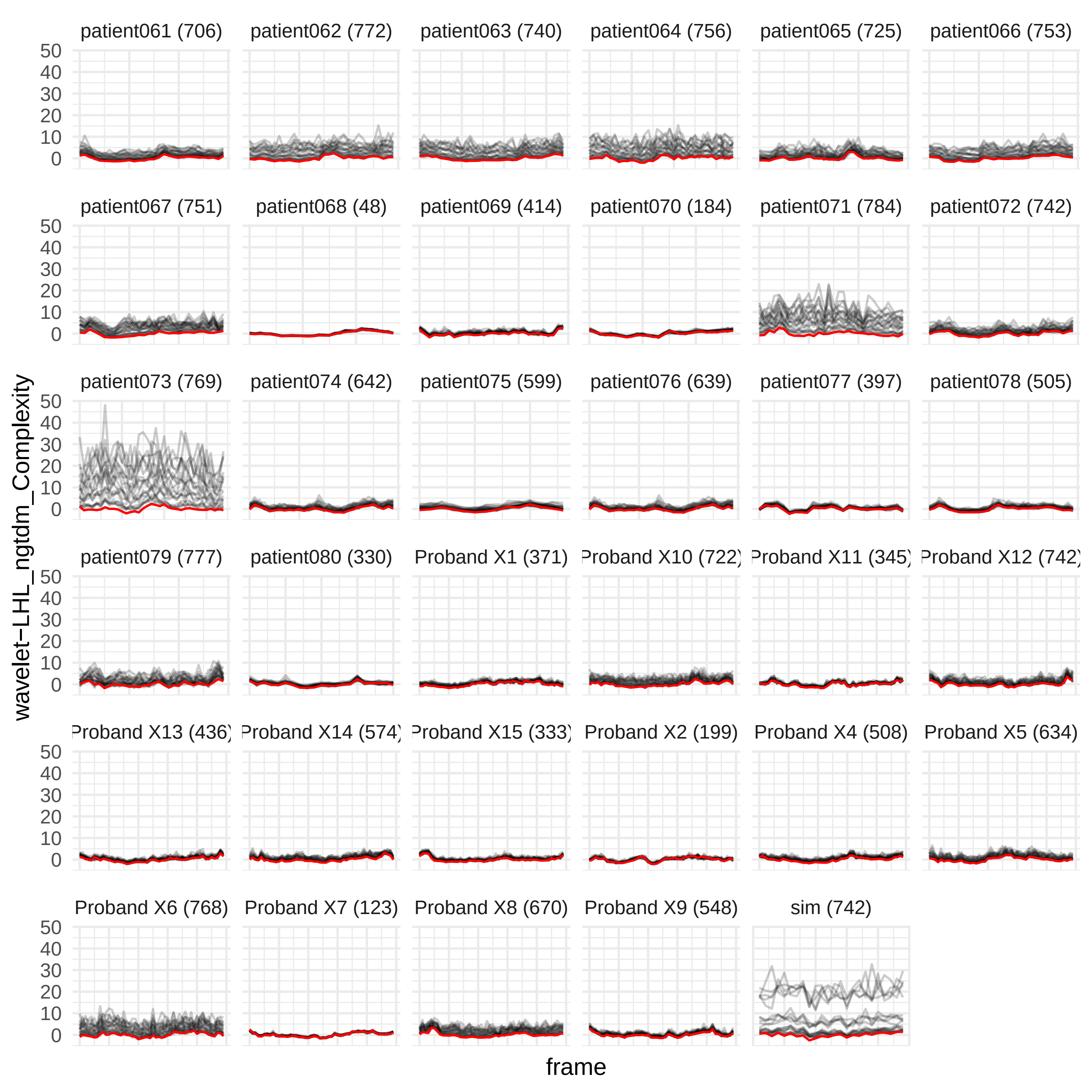

patient061 (706)
patient062 (772)
patient063 (740)
patient064 (756)
patient065 (725)
patient066 (753)
patient067 (751)
patient068 (48)
patient069 (414)
patient070 (184)
patient071 (784)
patient072 (742)
patient073 (769)
patient074 (642)
patient075 (599)
patient076 (639)
patient077 (397)
patient078 (505)
patient079 (777)
patient080 (330)
Proband X1 (371)
Proband X10 (722)
Proband X11 (345)
Proband X12 (742)
Proband X13 (436)
Proband X14 (574)
Proband X15 (333)
Proband X2 (199)
Proband X4 (508)
Proband X5 (634)
Proband X6 (768)
Proband X7 (123)
Proband X8 (670)
Proband X9 (548)
sim (742)
wavelet−LHL_ngtdm_Complexity
0
10
20
30
40
50
frame

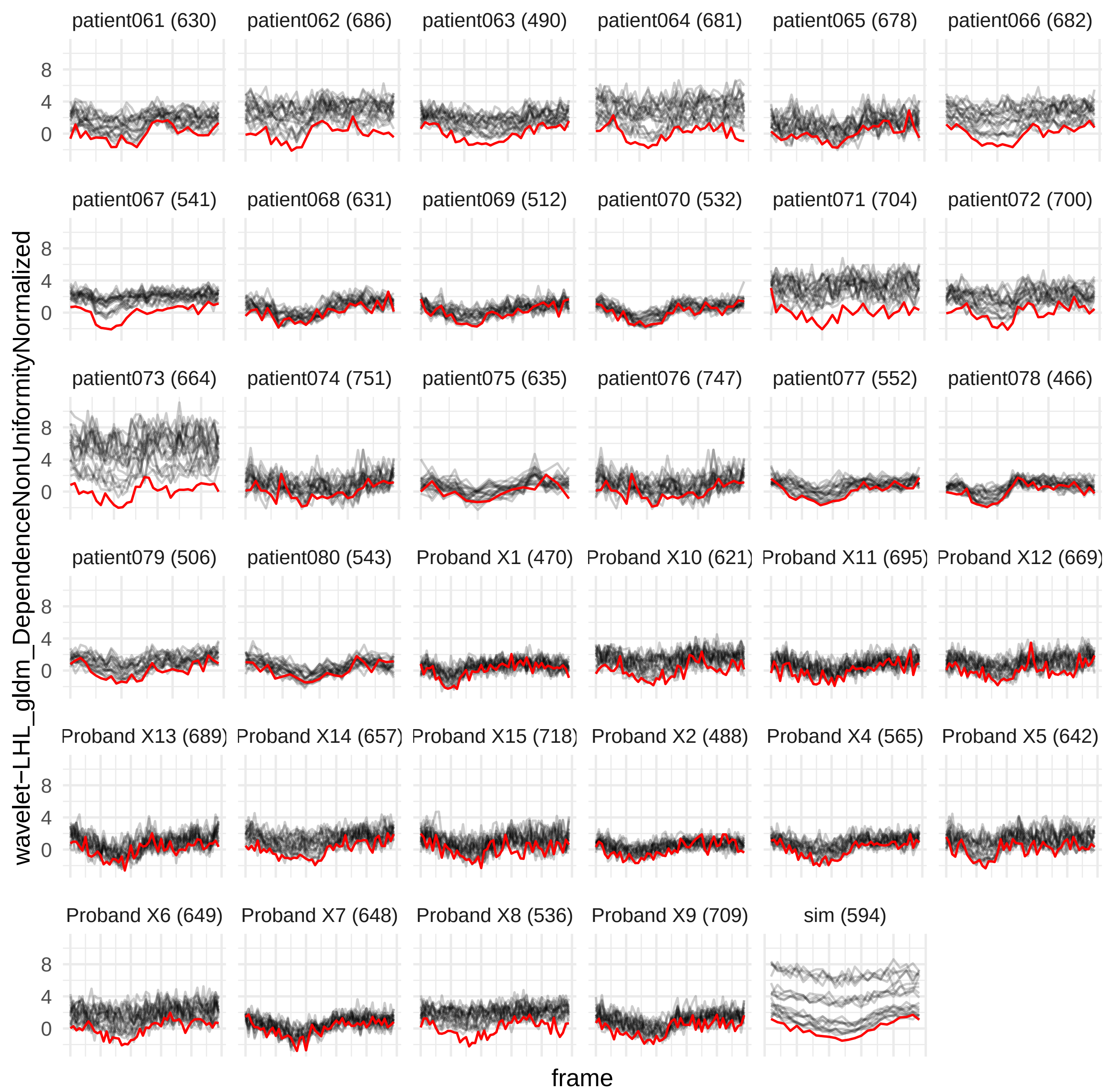

patient061 (630)
patient062 (686)
patient063 (490)
patient064 (681)
patient065 (678)
patient066 (682)
patient067 (541)
patient068 (631)
patient069 (512)
patient070 (532)
patient071 (704)
patient072 (700)
patient073 (664)
patient074 (751)
patient075 (635)
patient076 (747)
patient077 (552)
patient078 (466)
patient079 (506)
patient080 (543)
Proband X1 (470)
Proband X10 (621)
Proband X11 (695)
Proband X12 (669)
Proband X13 (689)
Proband X14 (657)
Proband X15 (718)
Proband X2 (488)
Proband X4 (565)
Proband X5 (642)
Proband X6 (649)
Proband X7 (648)
Proband X8 (536)
Proband X9 (709)
sim (594)
wavelet−LHL_gldm_DependenceNonUniformityNormalized
8
4
0
frame

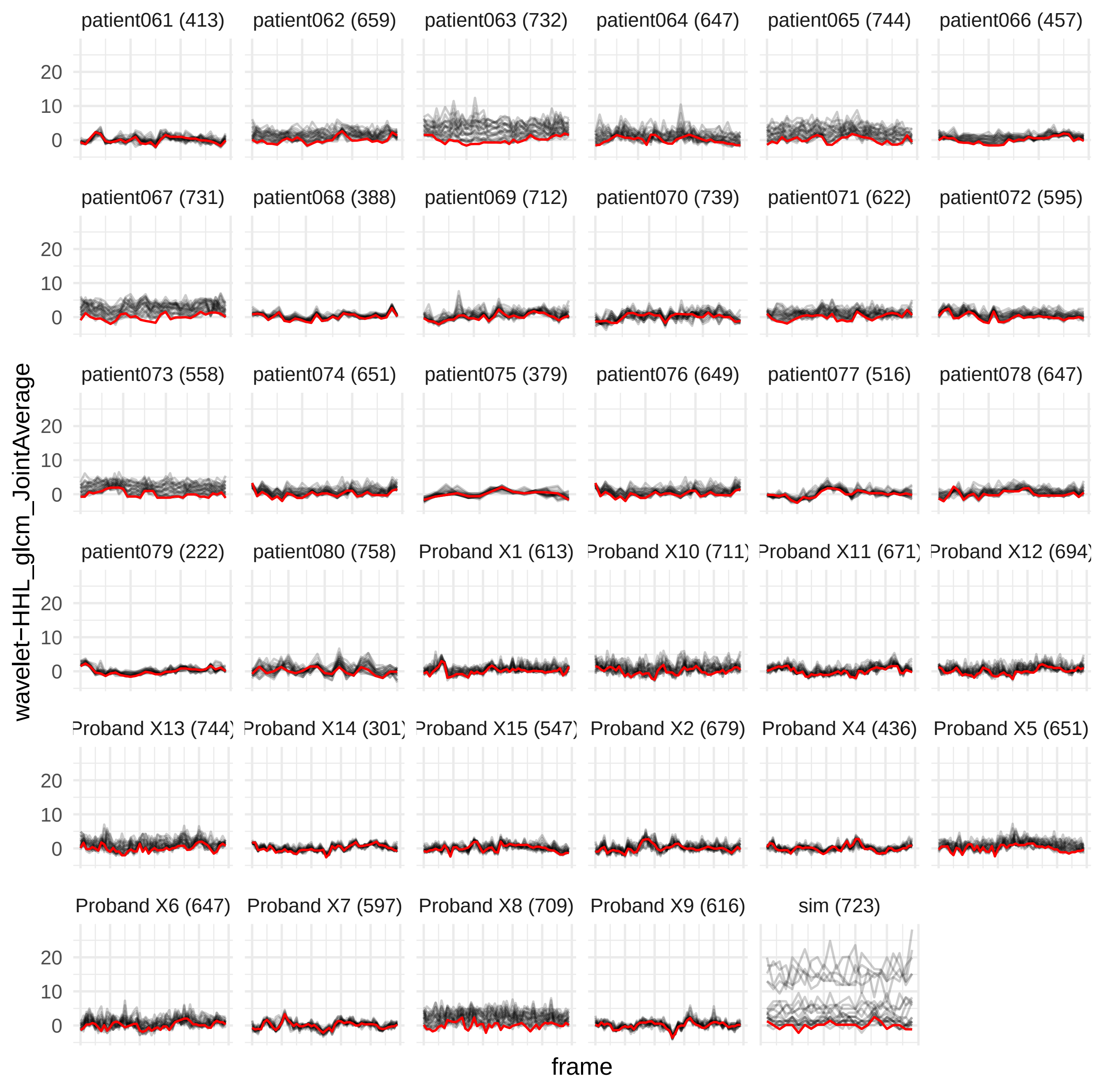

patient061 (413)
patient062 (659)
patient063 (732)
patient064 (647)
patient065 (744)
patient066 (457)
patient067 (731)
patient068 (388)
patient069 (712)
patient070 (739)
patient071 (622)
patient072 (595)
patient073 (558)
patient074 (651)
patient075 (379)
patient076 (649)
patient077 (516)
patient078 (647)
patient079 (222)
patient080 (758)
Proband X1 (613)
Proband X10 (711)
Proband X11 (671)
Proband X12 (694)
Proband X13 (744)
Proband X14 (301)
Proband X15 (547)
Proband X2 (679)
Proband X4 (436)
Proband X5 (651)
Proband X6 (647)
Proband X7 (597)
Proband X8 (709)
Proband X9 (616)
sim (723)
20
10
0
wavelet−HHL_glcm_JointAverage
frame

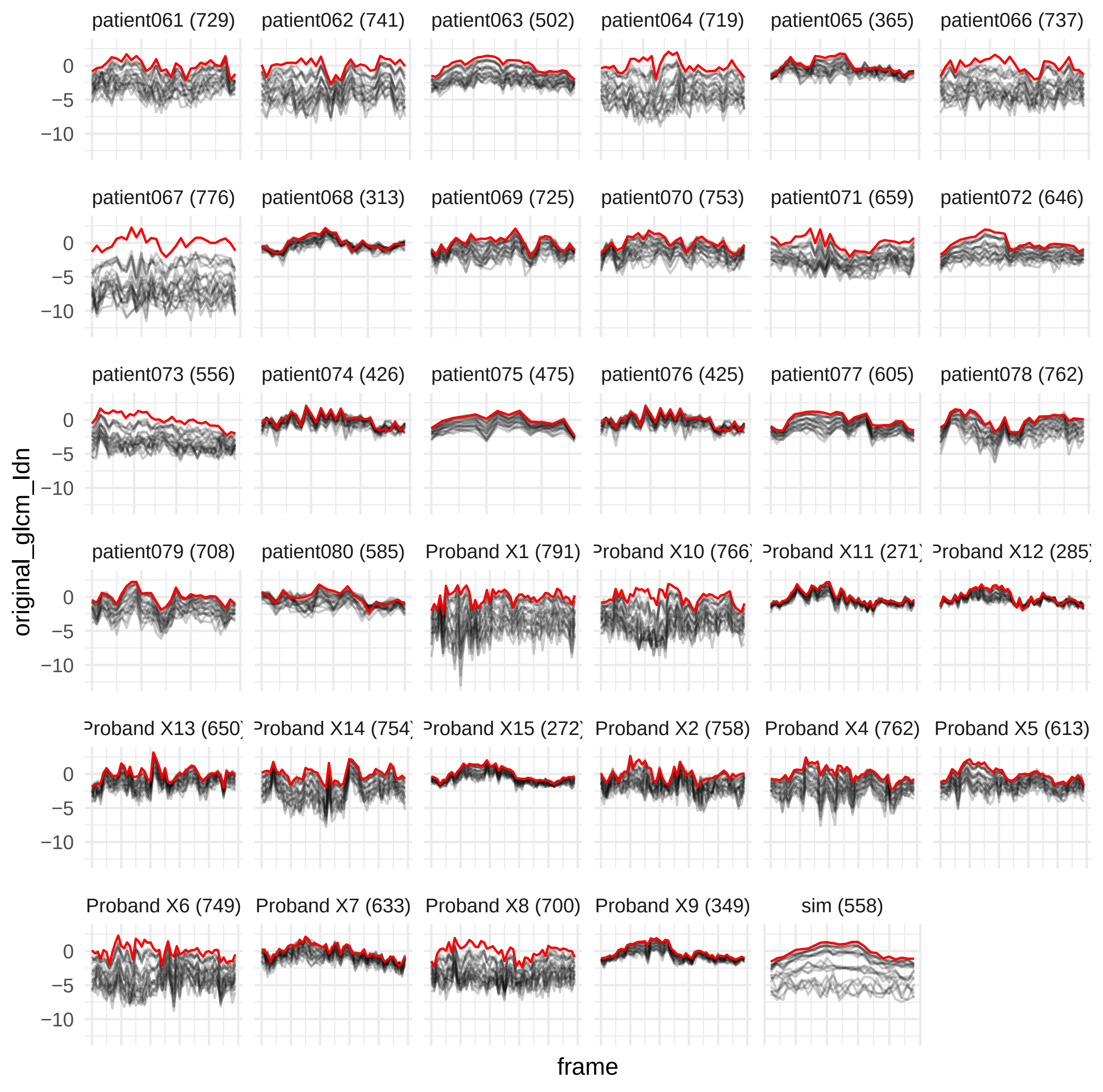
patient061 (729)
patient062 (741)
patient063 (502)
patient064 (719)
patient065 (365)
patient066 (737)
patient067 (776)
patient068 (313)
patient069 (725)
patient070 (753)
patient071 (659)
patient072 (646)
patient073 (556)
patient074 (426)
patient075 (475)
patient076 (425)
patient077 (605)
patient078 (762)
patient079 (708)
patient080 (585)
Proband X1 (791)
Proband X10 (766)
Proband X11 (271)
Proband X12 (285)
Proband X13 (650)
Proband X14 (754)
Proband X15 (272)
Proband X2 (758)
Proband X4 (762)
Proband X5 (613)
Proband X6 (749)
Proband X7 (633)
Proband X8 (700)
Proband X9 (349)
sim (558)
original_glcm_Idn
0
−5
−10
frame

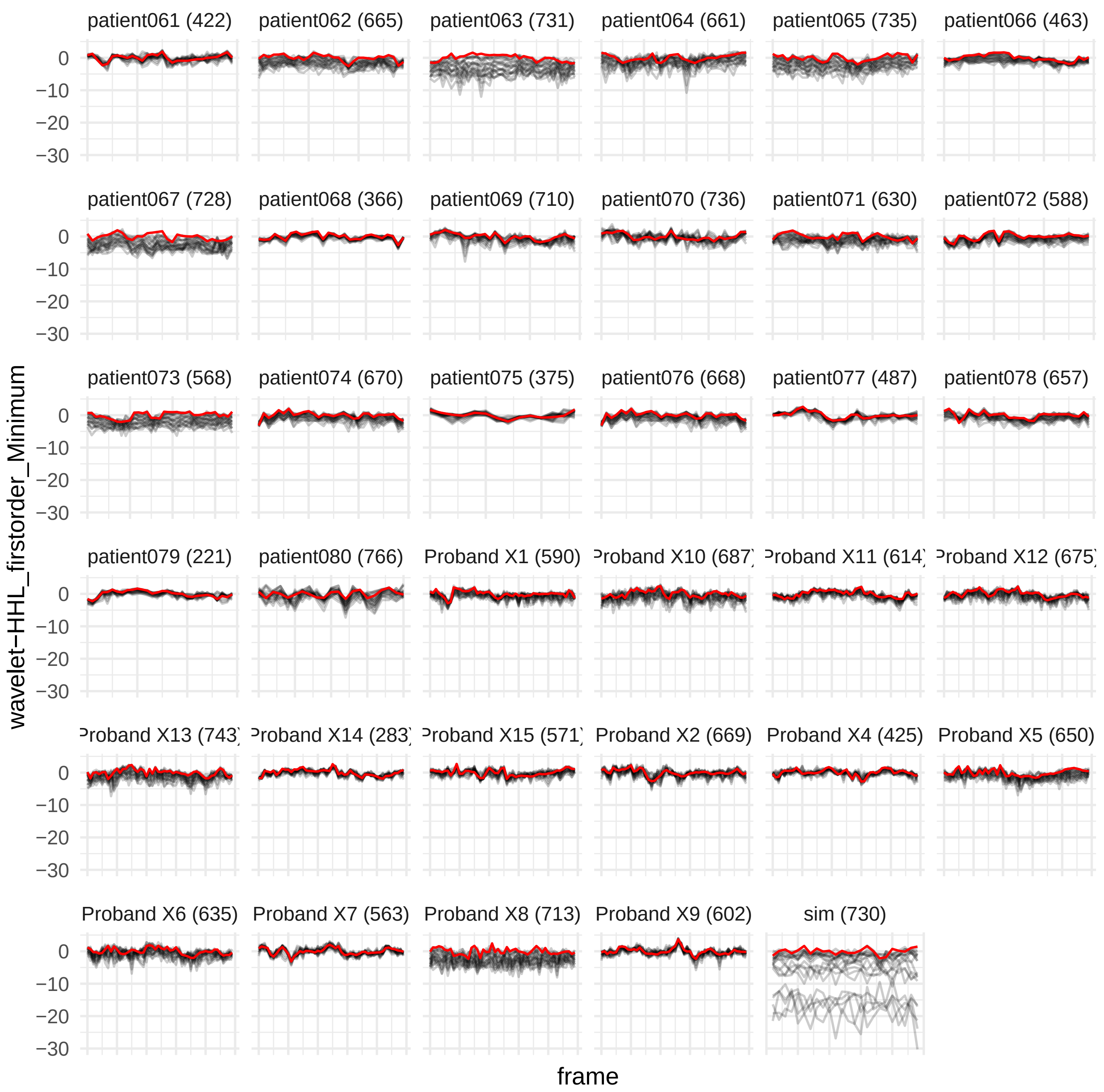

patient061 (422)
patient062 (665)
patient063 (731)
patient064 (661)
patient065 (735)
patient066 (463)
patient067 (728)
patient068 (366)
patient069 (710)
patient070 (736)
patient071 (630)
patient072 (588)
patient073 (568)
patient074 (670)
patient075 (375)
patient076 (668)
patient077 (487)
patient078 (657)
patient079 (221)
patient080 (766)
Proband X1 (590)
Proband X10 (687)
Proband X11 (614)
Proband X12 (675)
Proband X13 (743)
Proband X14 (283)
Proband X15 (571)
Proband X2 (669)
Proband X4 (425)
Proband X5 (650)
Proband X6 (635)
Proband X7 (563)
Proband X8 (713)
Proband X9 (602)
sim (730)
0
−10
−20
−30
wavelet−HHL_firstorder_Minimum
frame

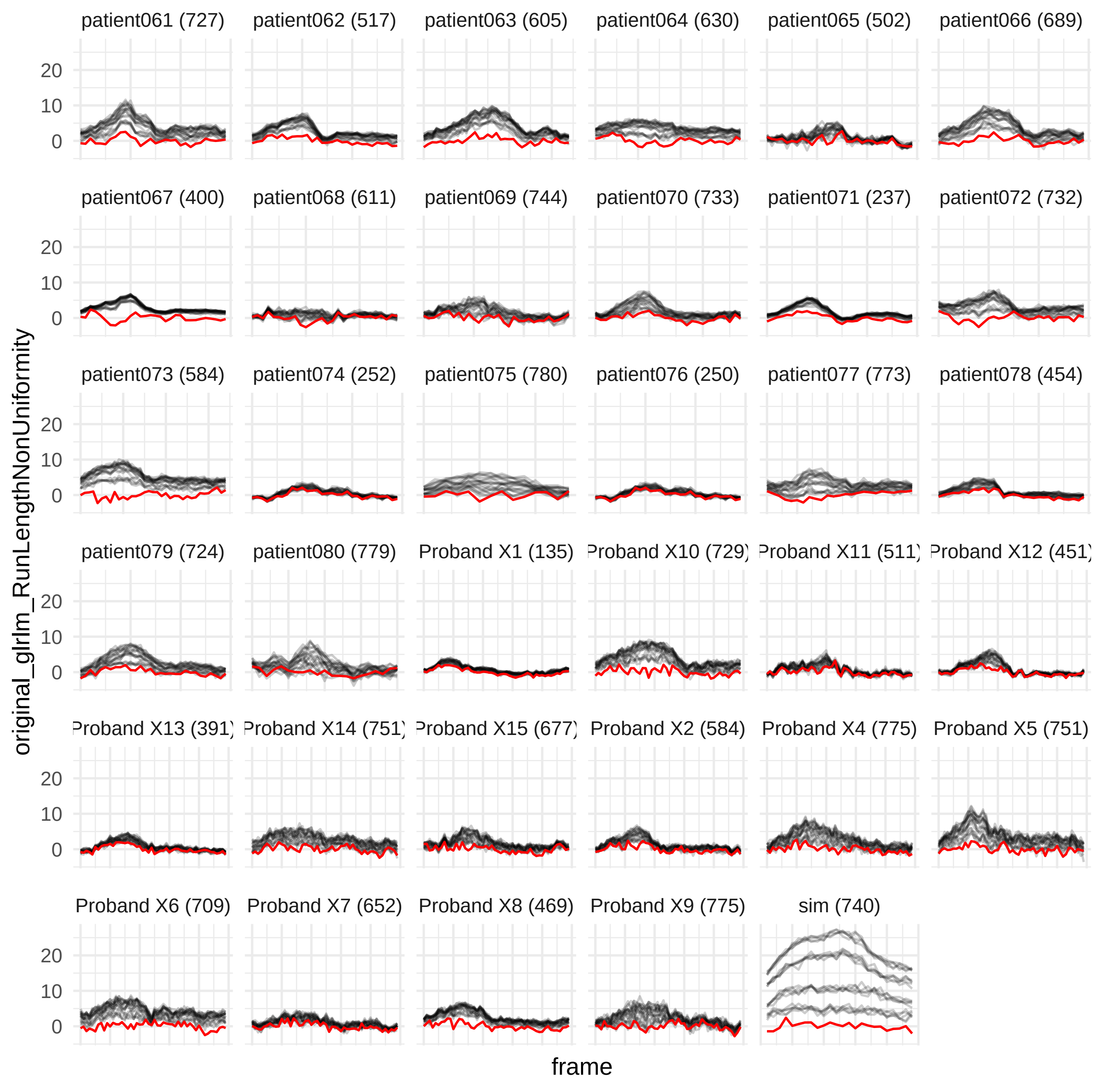
patient061 (727)
patient062 (517)
patient063 (605)
patient064 (630)
patient065 (502)
patient066 (689)
patient067 (400)
patient068 (611)
patient069 (744)
patient070 (733)
patient071 (237)
patient072 (732)
patient073 (584)
patient074 (252)
patient075 (780)
patient076 (250)
patient077 (773)
patient078 (454)
patient079 (724)
patient080 (779)
Proband X1 (135)
Proband X10 (729)
Proband X11 (511)
Proband X12 (451)
Proband X13 (391)
Proband X14 (751)
Proband X15 (677)
Proband X2 (584)
Proband X4 (775)
Proband X5 (751)
Proband X6 (709)
Proband X7 (652)
Proband X8 (469)
Proband X9 (775)
sim (740)
original_glrlm_RunLengthNonUniformity
20
10
0
frame

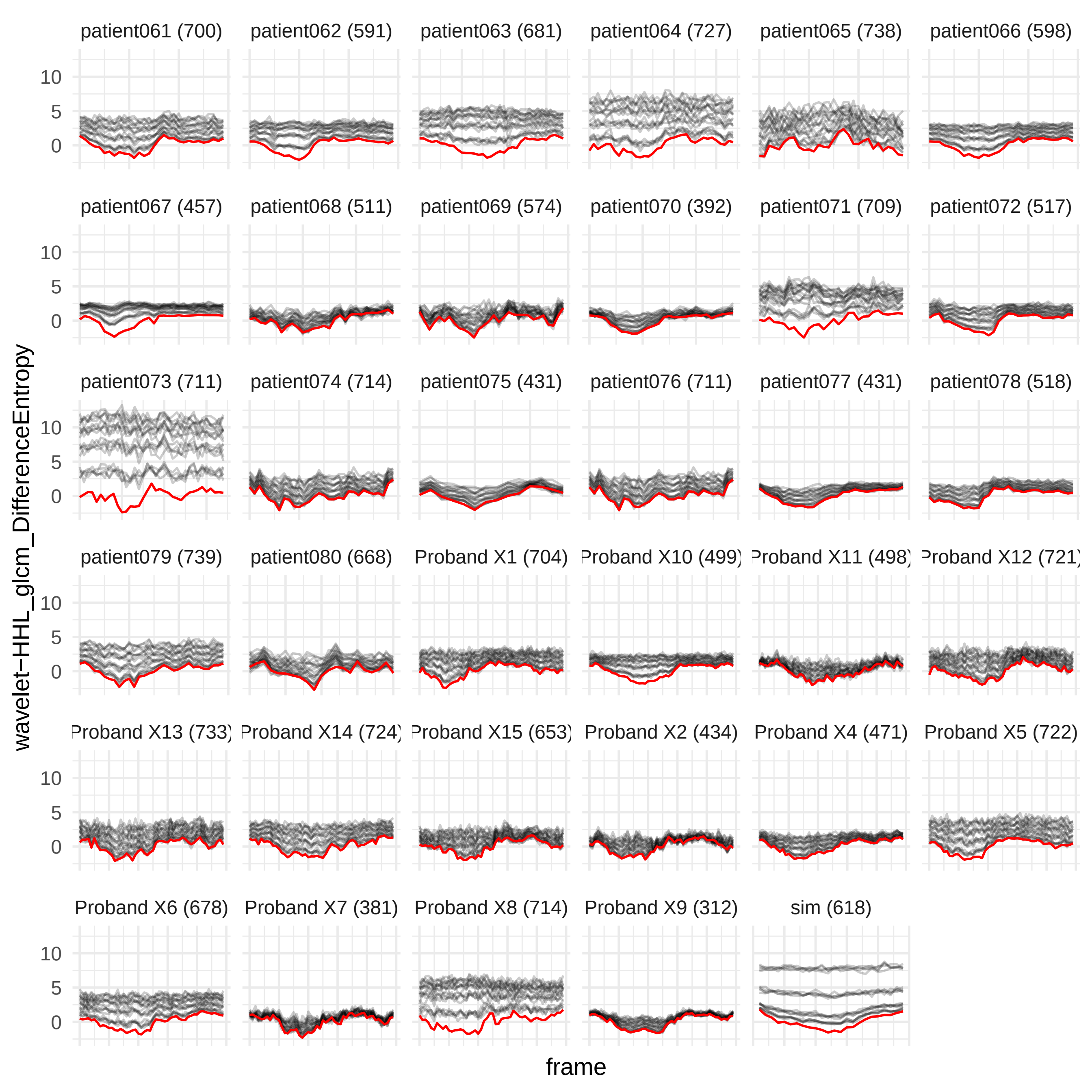

patient061 (700)
patient062 (591)
patient063 (681)
patient064 (727)
patient065 (738)
patient066 (598)
patient067 (457)
patient068 (511)
patient069 (574)
patient070 (392)
patient071 (709)
patient072 (517)
patient073 (711)
patient074 (714)
patient075 (431)
patient076 (711)
patient077 (431)
patient078 (518)
patient079 (739)
patient080 (668)
Proband X1 (704)
Proband X10 (499)
Proband X11 (498)
Proband X12 (721)
Proband X13 (733)
Proband X14 (724)
Proband X15 (653)
Proband X2 (434)
Proband X4 (471)
Proband X5 (722)
Proband X6 (678)
Proband X7 (381)
Proband X8 (714)
Proband X9 (312)
sim (618)
10
5
0
wavelet−HHL_glcm_DifferenceEntropy
frame

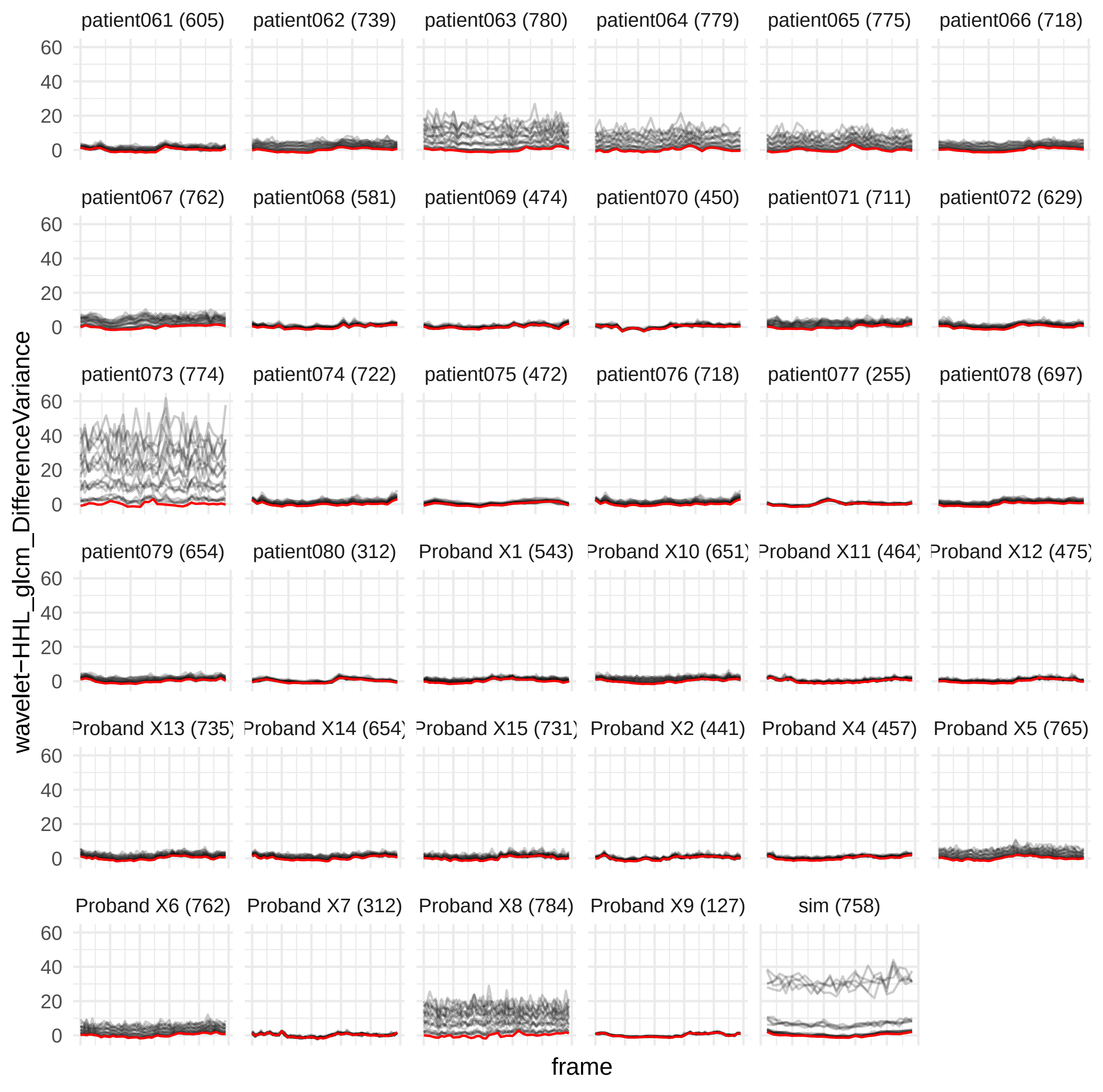

patient061 (605)
patient062 (739)
patient063 (780)
patient064 (779)
patient065 (775)
patient066 (718)
patient067 (762)
patient068 (581)
patient069 (474)
patient070 (450)
patient071 (711)
patient072 (629)
patient073 (774)
patient074 (722)
patient075 (472)
patient076 (718)
patient077 (255)
patient078 (697)
patient079 (654)
patient080 (312)
Proband X1 (543)
Proband X10 (651)
Proband X11 (464)
Proband X12 (475)
Proband X13 (735)
Proband X14 (654)
Proband X15 (731)
Proband X2 (441)
Proband X4 (457)
Proband X5 (765)
Proband X6 (762)
Proband X7 (312)
Proband X8 (784)
Proband X9 (127)
sim (758)
60
40
20
0
wavelet−HHL_glcm_DifferenceVariance
frame

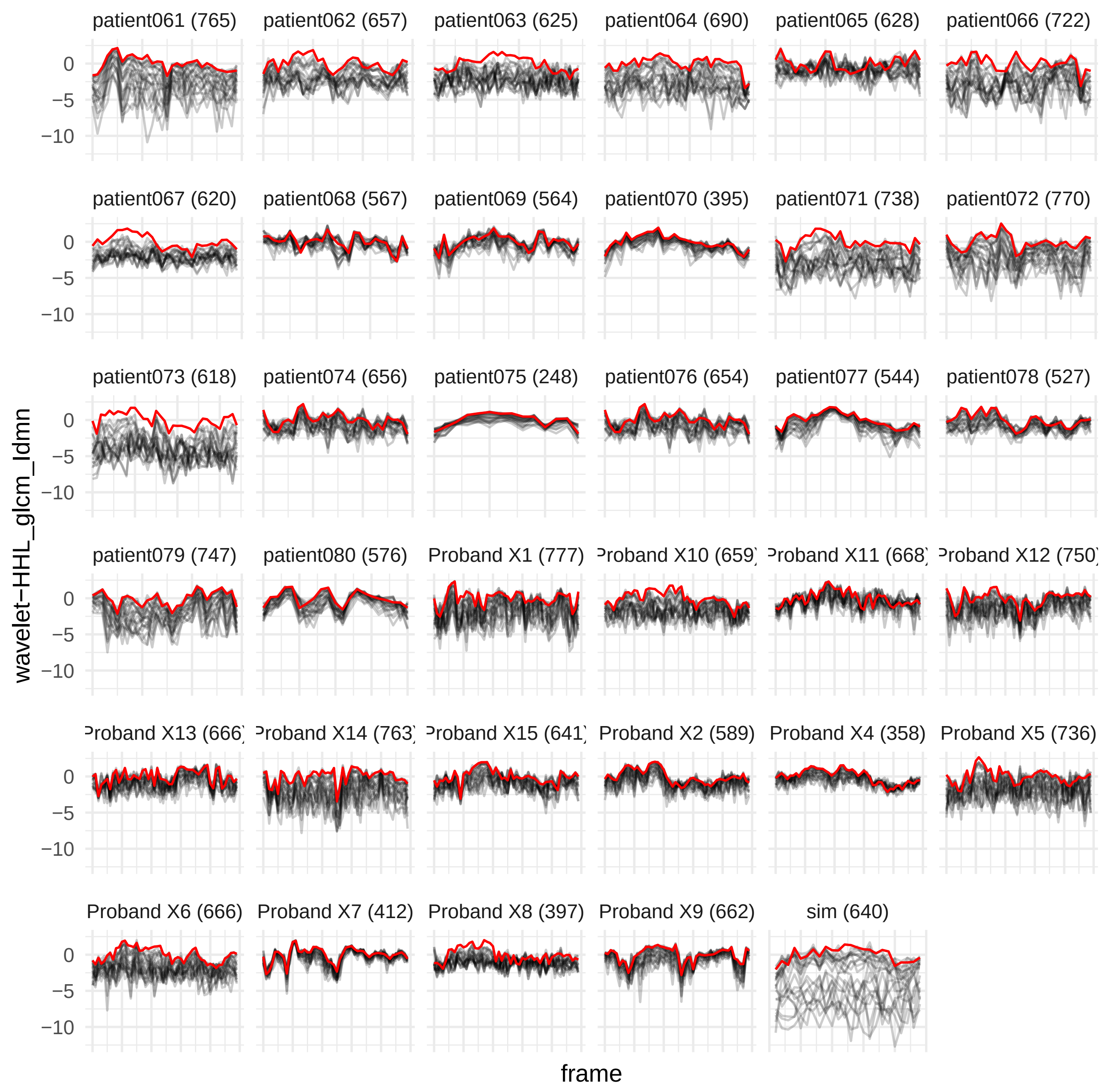

patient061 (765)
patient062 (657)
patient063 (625)
patient064 (690)
patient065 (628)
patient066 (722)
patient067 (620)
patient068 (567)
patient069 (564)
patient070 (395)
patient071 (738)
patient072 (770)
patient073 (618)
patient074 (656)
patient075 (248)
patient076 (654)
patient077 (544)
patient078 (527)
patient079 (747)
patient080 (576)
Proband X1 (777)
Proband X10 (659)
Proband X11 (668)
Proband X12 (750)
Proband X13 (666)
Proband X14 (763)
Proband X15 (641)
Proband X2 (589)
Proband X4 (358)
Proband X5 (736)
Proband X6 (666)
Proband X7 (412)
Proband X8 (397)
Proband X9 (662)
sim (640)
0
−5
−10
wavelet−HHL_glcm_ldmn
frame

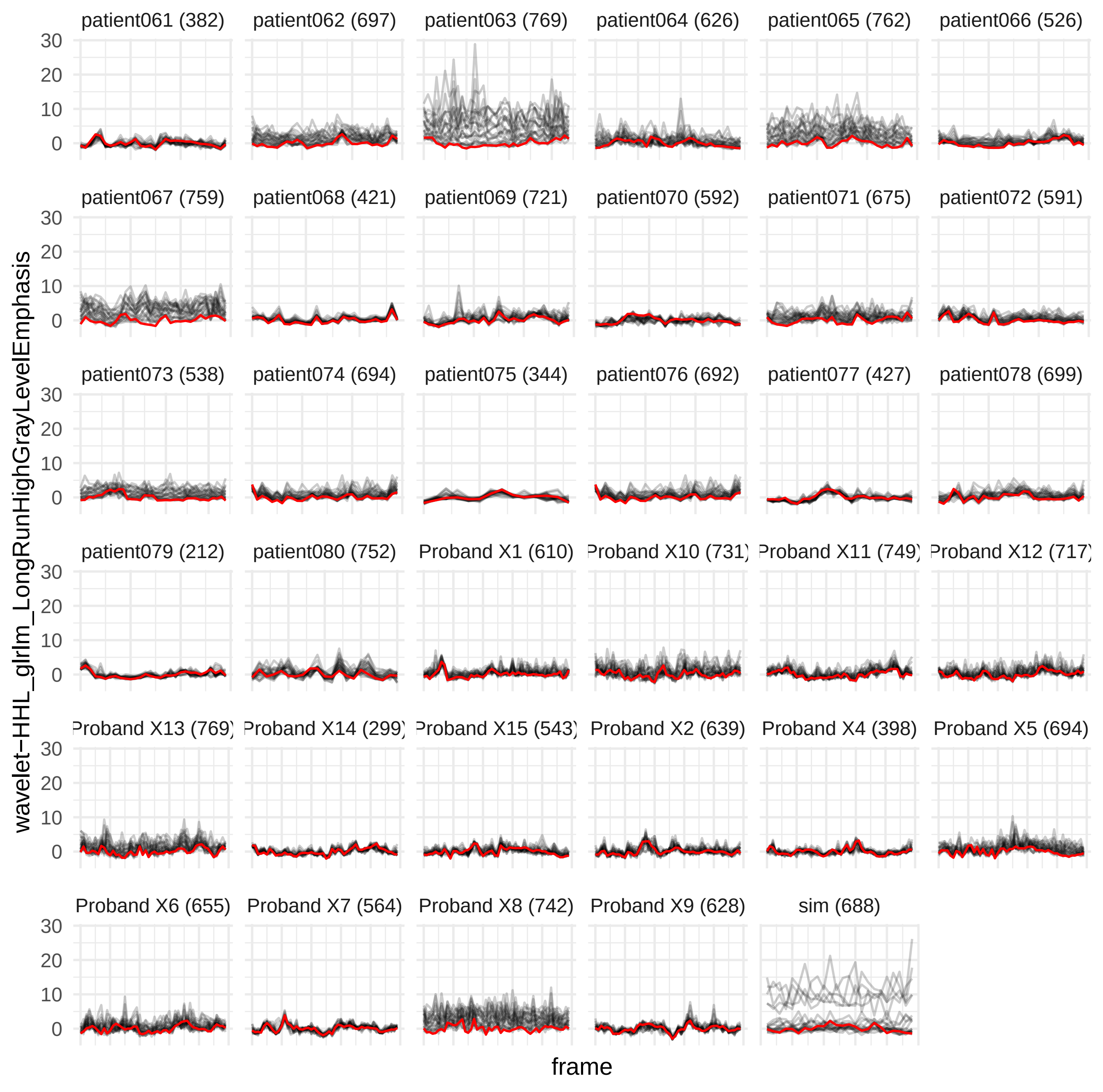

patient061 (382)
patient062 (697)
patient063 (769)
patient064 (626)
patient065 (762)
patient066 (526)
patient067 (759)
patient068 (421)
patient069 (721)
patient070 (592)
patient071 (675)
patient072 (591)
patient073 (538)
patient074 (694)
patient075 (344)
patient076 (692)
patient077 (427)
patient078 (699)
patient079 (212)
patient080 (752)
Proband X1 (610)
Proband X10 (731)
Proband X11 (749)
Proband X12 (717)
Proband X13 (769)
Proband X14 (299)
Proband X15 (543)
Proband X2 (639)
Proband X4 (398)
Proband X5 (694)
Proband X6 (655)
Proband X7 (564)
Proband X8 (742)
Proband X9 (628)
sim (688)
0
10
20
30
wavelet−HHL_glrlm_LongRunHighGrayLevelEmphasis
frame

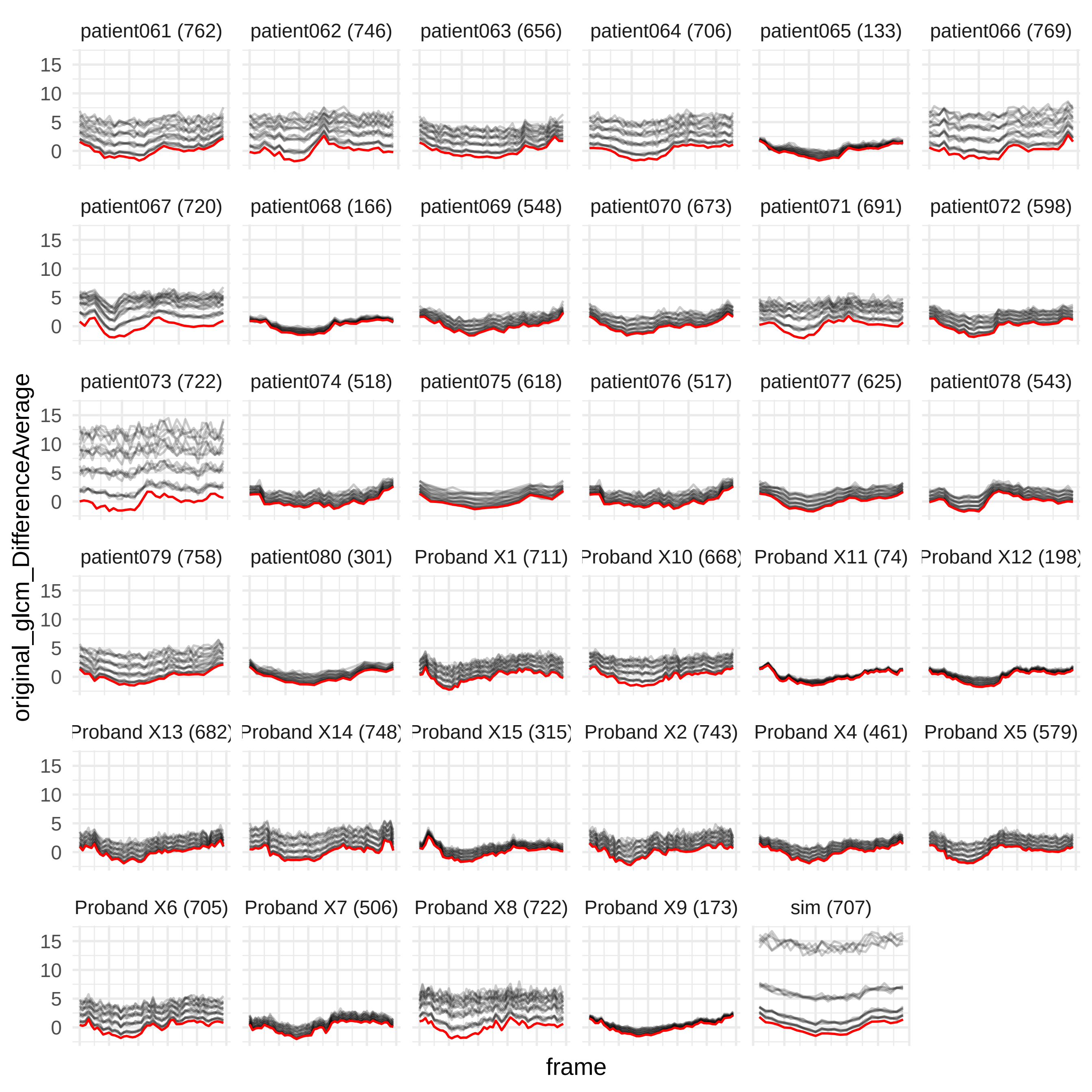

patient061 (762)
patient062 (746)
patient063 (656)
patient064 (706)
patient065 (133)
patient066 (769)
patient067 (720)
patient068 (166)
patient069 (548)
patient070 (673)
patient071 (691)
patient072 (598)
patient073 (722)
patient074 (518)
patient075 (618)
patient076 (517)
patient077 (625)
patient078 (543)
patient079 (758)
patient080 (301)
Proband X1 (711)
Proband X10 (668)
Proband X11 (74)
Proband X12 (198)
Proband X13 (682)
Proband X14 (748)
Proband X15 (315)
Proband X2 (743)
Proband X4 (461)
Proband X5 (579)
Proband X6 (705)
Proband X7 (506)
Proband X8 (722)
Proband X9 (173)
sim (707)
original_glcm_DifferenceAverage
frame

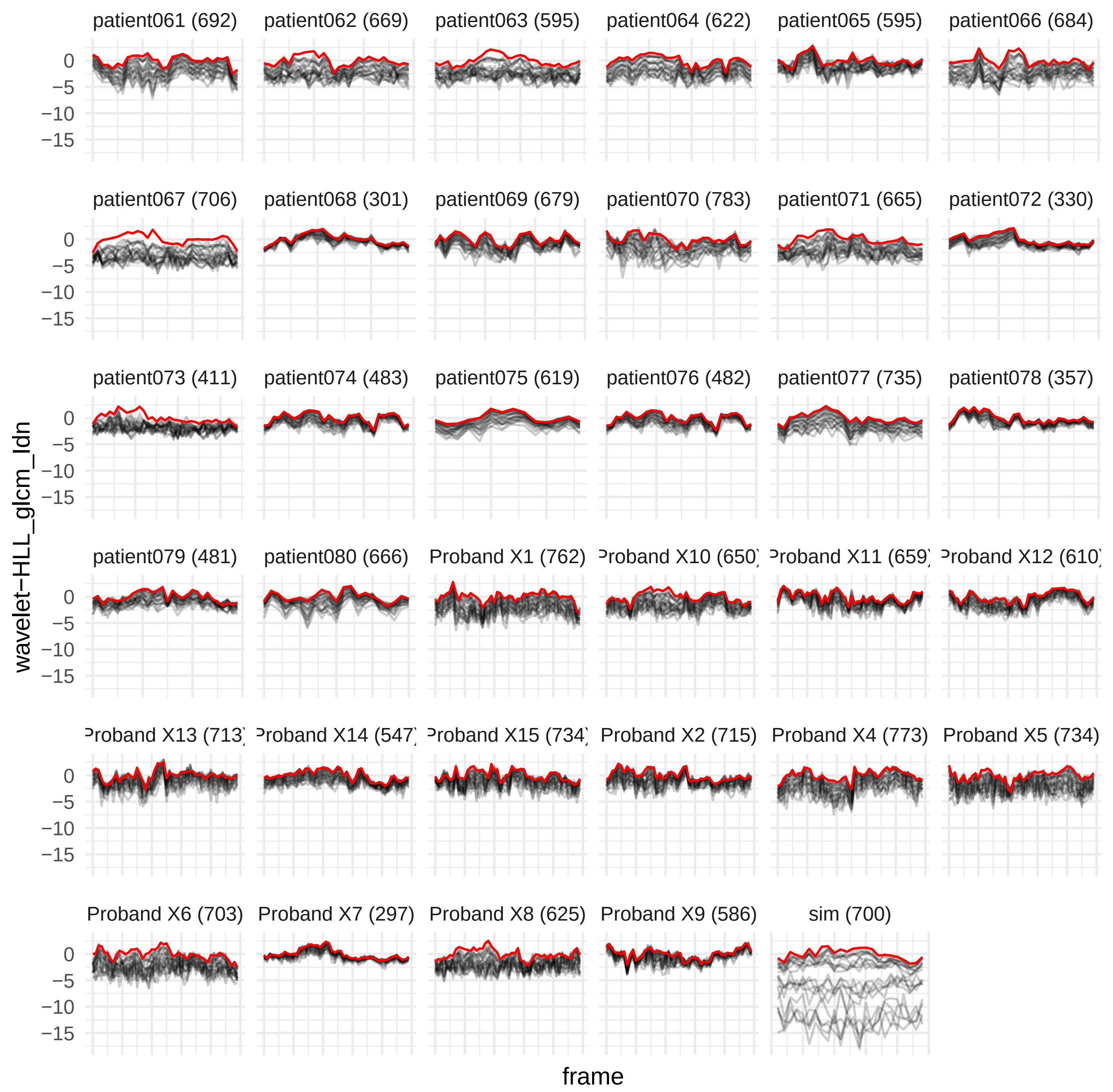

patient061 (692)
patient062 (669)
patient063 (595)
patient064 (622)
patient065 (595)
patient066 (684)
patient067 (706)
patient068 (301)
patient069 (679)
patient070 (783)
patient071 (665)
patient072 (330)
patient073 (411)
patient074 (483)
patient075 (619)
patient076 (482)
patient077 (735)
patient078 (357)
patient079 (481)
patient080 (666)
Proband X1 (762)
Proband X10 (650)
Proband X11 (659)
Proband X12 (610)
Proband X13 (713)
Proband X14 (547)
Proband X15 (734)
Proband X2 (715)
Proband X4 (773)
Proband X5 (734)
Proband X6 (703)
Proband X7 (297)
Proband X8 (625)
Proband X9 (586)
sim (700)
wavelet−HLL_glcm_Idn
0
−5
−10
−15
frame

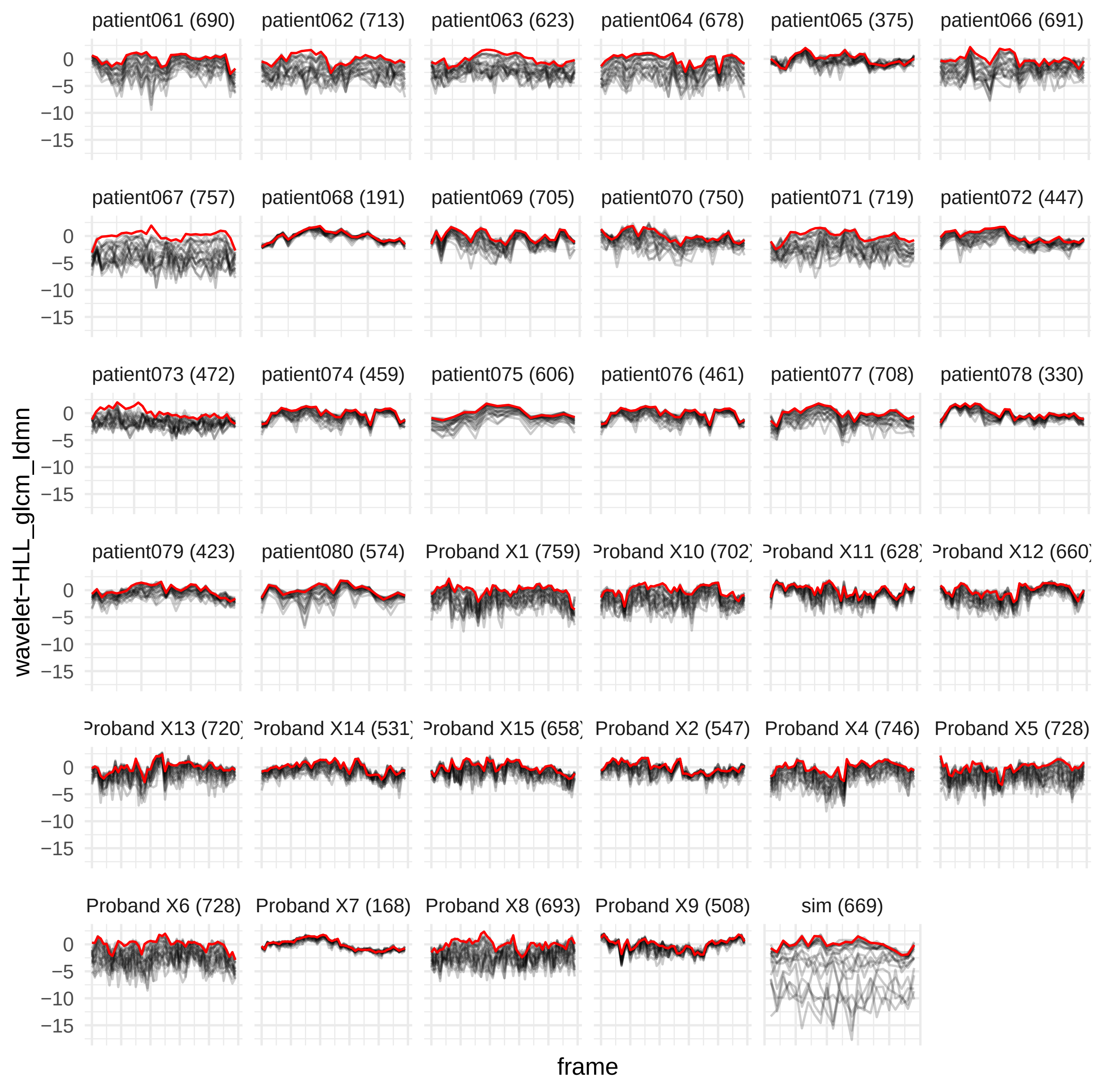

patient061 (690)
patient062 (713)
patient063 (623)
patient064 (678)
patient065 (375)
patient066 (691)
patient067 (757)
patient068 (191)
patient069 (705)
patient070 (750)
patient071 (719)
patient072 (447)
patient073 (472)
patient074 (459)
patient075 (606)
patient076 (461)
patient077 (708)
patient078 (330)
patient079 (423)
patient080 (574)
Proband X1 (759)
Proband X10 (702)
Proband X11 (628)
Proband X12 (660)
Proband X13 (720)
Proband X14 (531)
Proband X15 (658)
Proband X2 (547)
Proband X4 (746)
Proband X5 (728)
Proband X6 (728)
Proband X7 (168)
Proband X8 (693)
Proband X9 (508)
sim (669)
0
−5
−10
−15
wavelet−HLL_glcm_Idmn
frame

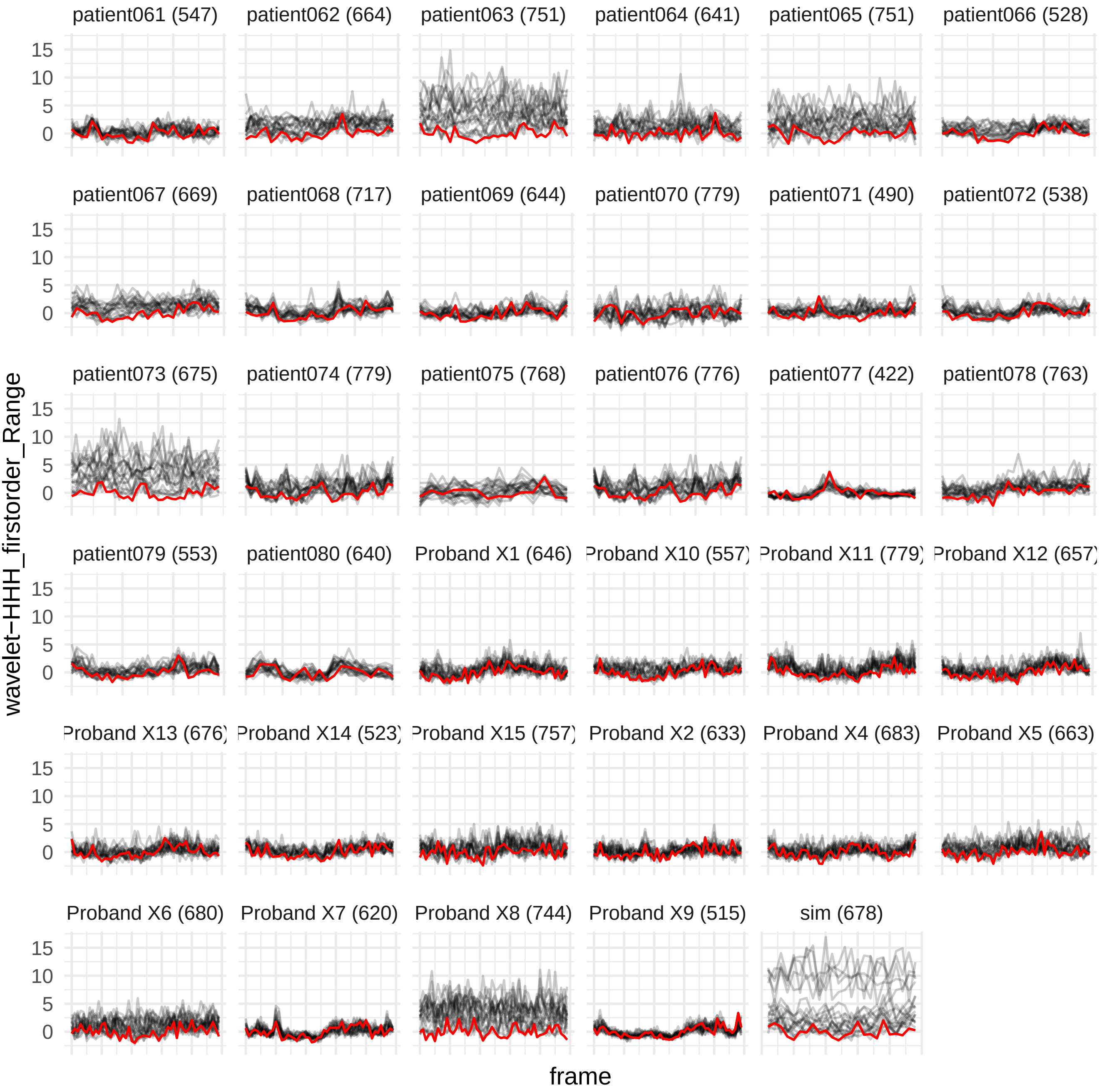
patient061 (547)
patient062 (664)
patient063 (751)
patient064 (641)
patient065 (751)
patient066 (528)
patient067 (669)
patient068 (717)
patient069 (644)
patient070 (779)
patient071 (490)
patient072 (538)
patient073 (675)
patient074 (779)
patient075 (768)
patient076 (776)
patient077 (422)
patient078 (763)
patient079 (553)
patient080 (640)
Proband X1 (646)
Proband X10 (557)
Proband X11 (779)
Proband X12 (657)
Proband X13 (676)
Proband X14 (523)
Proband X15 (757)
Proband X2 (633)
Proband X4 (683)
Proband X5 (663)
Proband X6 (680)
Proband X7 (620)
Proband X8 (744)
Proband X9 (515)
sim (678)
wavelet−HHH_firstorder_Range
15
10
5
0
frame

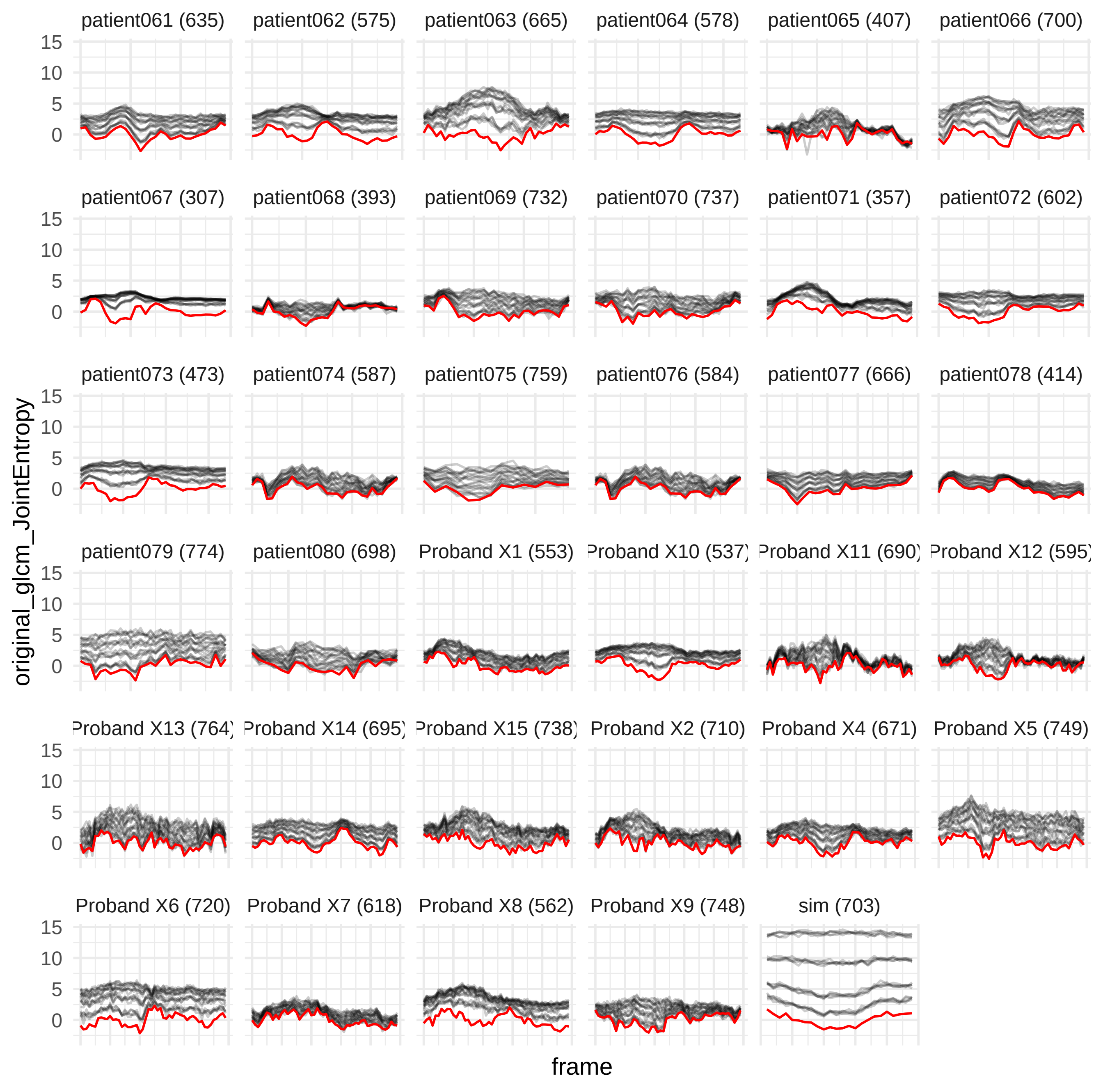

patient061 (635)
patient062 (575)
patient063 (665)
patient064 (578)
patient065 (407)
patient066 (700)
patient067 (307)
patient068 (393)
patient069 (732)
patient070 (737)
patient071 (357)
patient072 (602)
patient073 (473)
patient074 (587)
patient075 (759)
patient076 (584)
patient077 (666)
patient078 (414)
patient079 (774)
patient080 (698)
Proband X1 (553)
Proband X10 (537)
Proband X11 (690)
Proband X12 (595)
Proband X13 (764)
Proband X14 (695)
Proband X15 (738)
Proband X2 (710)
Proband X4 (671)
Proband X5 (749)
Proband X6 (720)
Proband X7 (618)
Proband X8 (562)
Proband X9 (748)
sim (703)
original_glcm_JointEntropy
frame
15
10
5
0

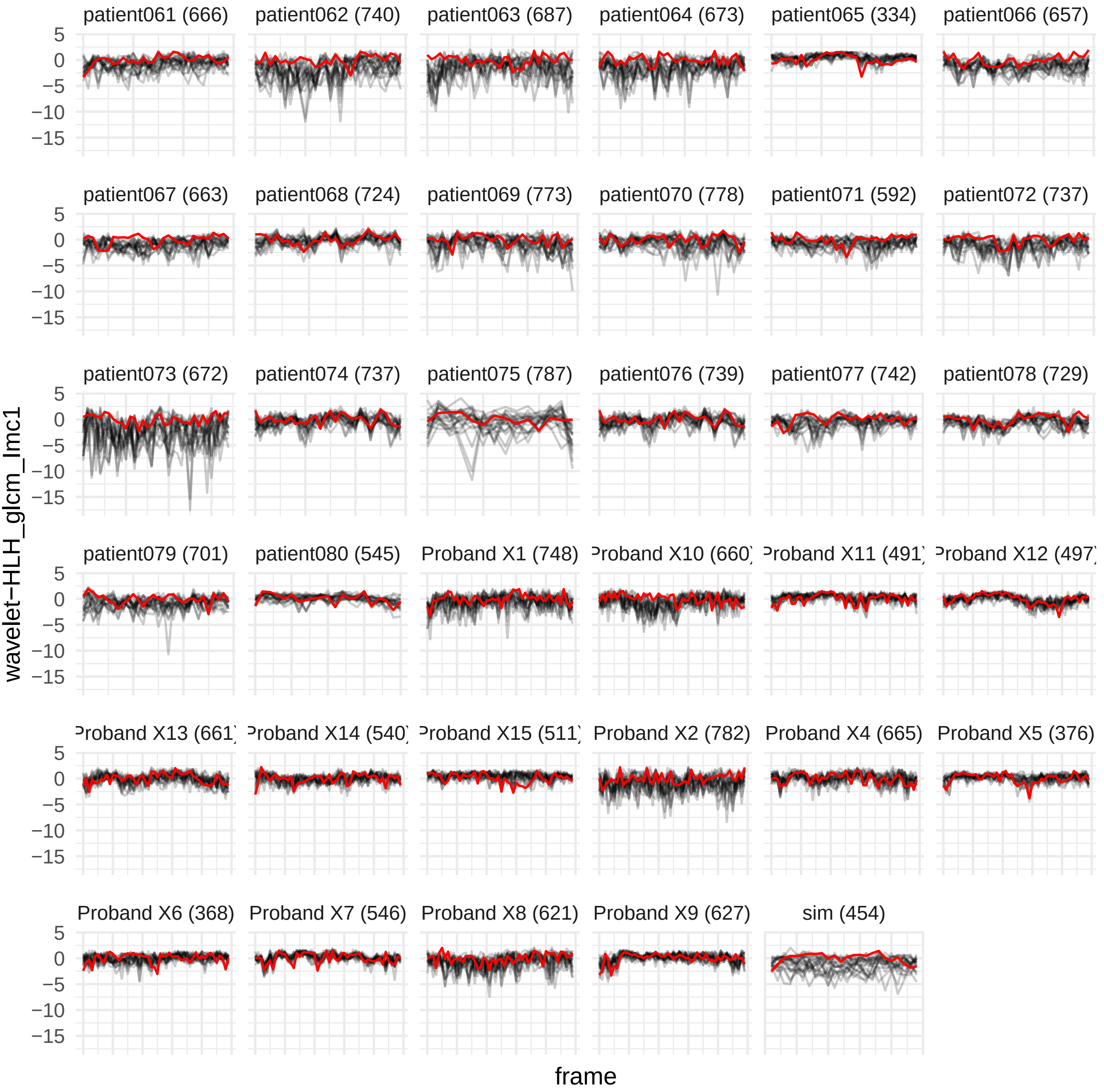

patient061 (666)
patient062 (740)
patient063 (687)
patient064 (673)
patient065 (334)
patient066 (657)
patient067 (663)
patient068 (724)
patient069 (773)
patient070 (778)
patient071 (592)
patient072 (737)
patient073 (672)
patient074 (737)
patient075 (787)
patient076 (739)
patient077 (742)
patient078 (729)
patient079 (701)
patient080 (545)
Proband X1 (748)
Proband X10 (660)
Proband X11 (491)
Proband X12 (497)
Proband X13 (661)
Proband X14 (540)
Proband X15 (511)
Proband X2 (782)
Proband X4 (665)
Proband X5 (376)
Proband X6 (368)
Proband X7 (546)
Proband X8 (621)
Proband X9 (627)
sim (454)
5
0
−5
−10
−15
wavelet−HLH_glcm_Imc1
frame

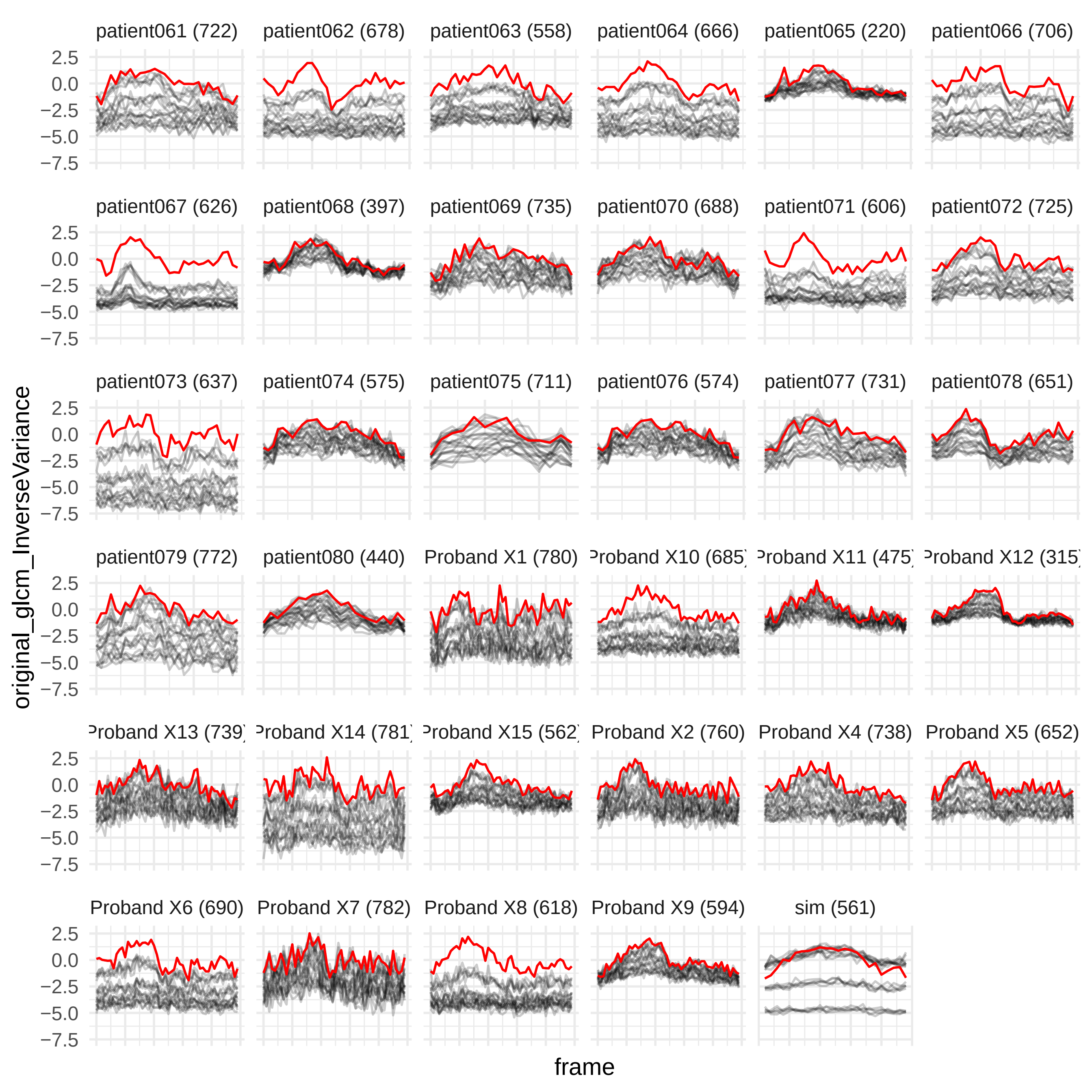

patient061 (722)
patient062 (678)
patient063 (558)
patient064 (666)
patient065 (220)
patient066 (706)
patient067 (626)
patient068 (397)
patient069 (735)
patient070 (688)
patient071 (606)
patient072 (725)
patient073 (637)
patient074 (575)
patient075 (711)
patient076 (574)
patient077 (731)
patient078 (651)
patient079 (772)
patient080 (440)
Proband X1 (780)
Proband X10 (685)
Proband X11 (475)
Proband X12 (315)
Proband X13 (739)
Proband X14 (781)
Proband X15 (562)
Proband X2 (760)
Proband X4 (738)
Proband X5 (652)
Proband X6 (690)
Proband X7 (782)
Proband X8 (618)
Proband X9 (594)
sim (561)
2.5
0.0
−2.5
−5.0
−7.5
original_glcm_InverseVariance
frame

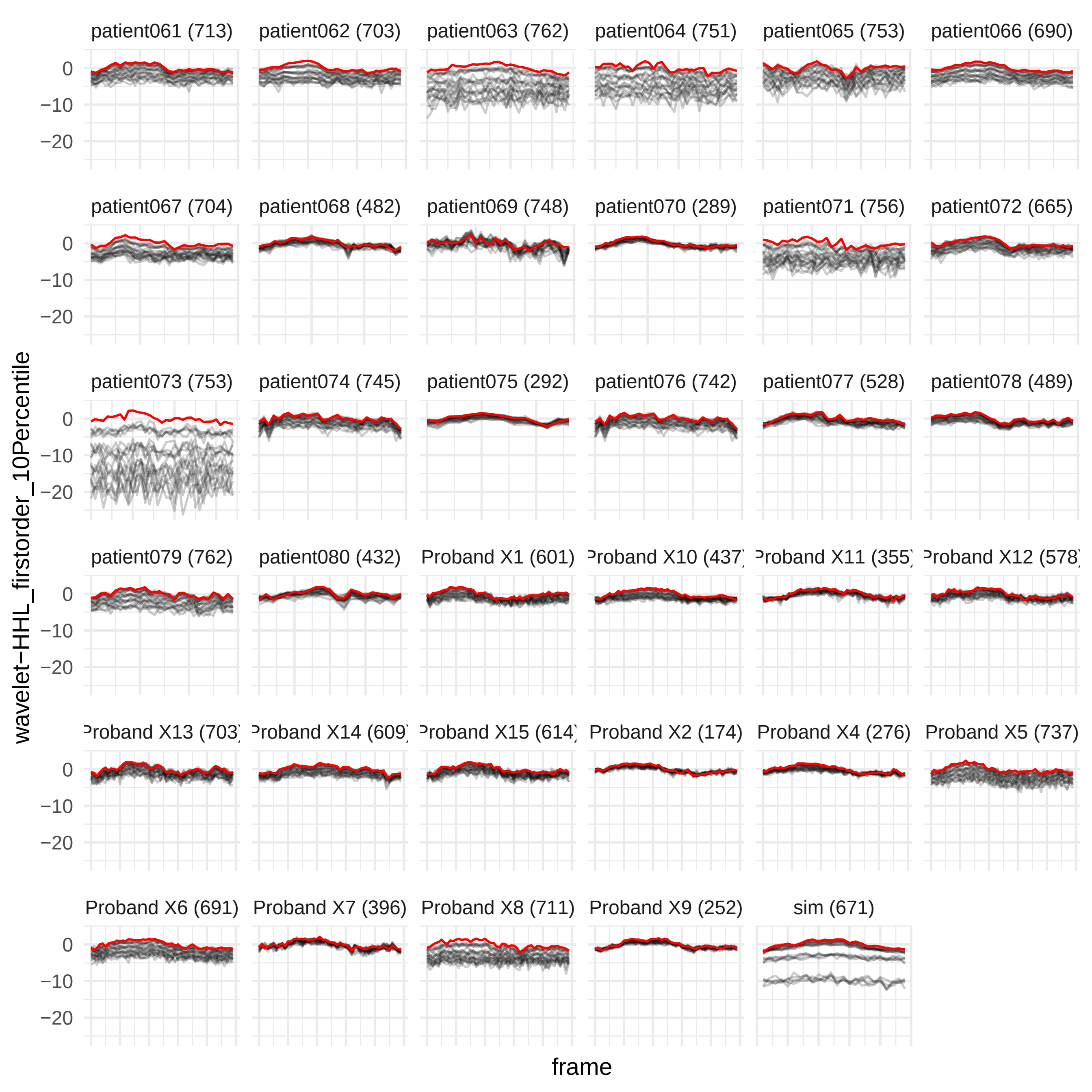
patient061 (713)
patient062 (703)
patient063 (762)
patient064 (751)
patient065 (753)
patient066 (690)
patient067 (704)
patient068 (482)
patient069 (748)
patient070 (289)
patient071 (756)
patient072 (665)
patient073 (753)
patient074 (745)
patient075 (292)
patient076 (742)
patient077 (528)
patient078 (489)
patient079 (762)
patient080 (432)
Proband X1 (601)
Proband X10 (437)
Proband X11 (355)
Proband X12 (578)
Proband X13 (703)
Proband X14 (609)
Proband X15 (614)
Proband X2 (174)
Proband X4 (276)
Proband X5 (737)
Proband X6 (691)
Proband X7 (396)
Proband X8 (711)
Proband X9 (252)
sim (671)
0
−10
−20
wavelet−HHL_firstorder_10Percentile
frame

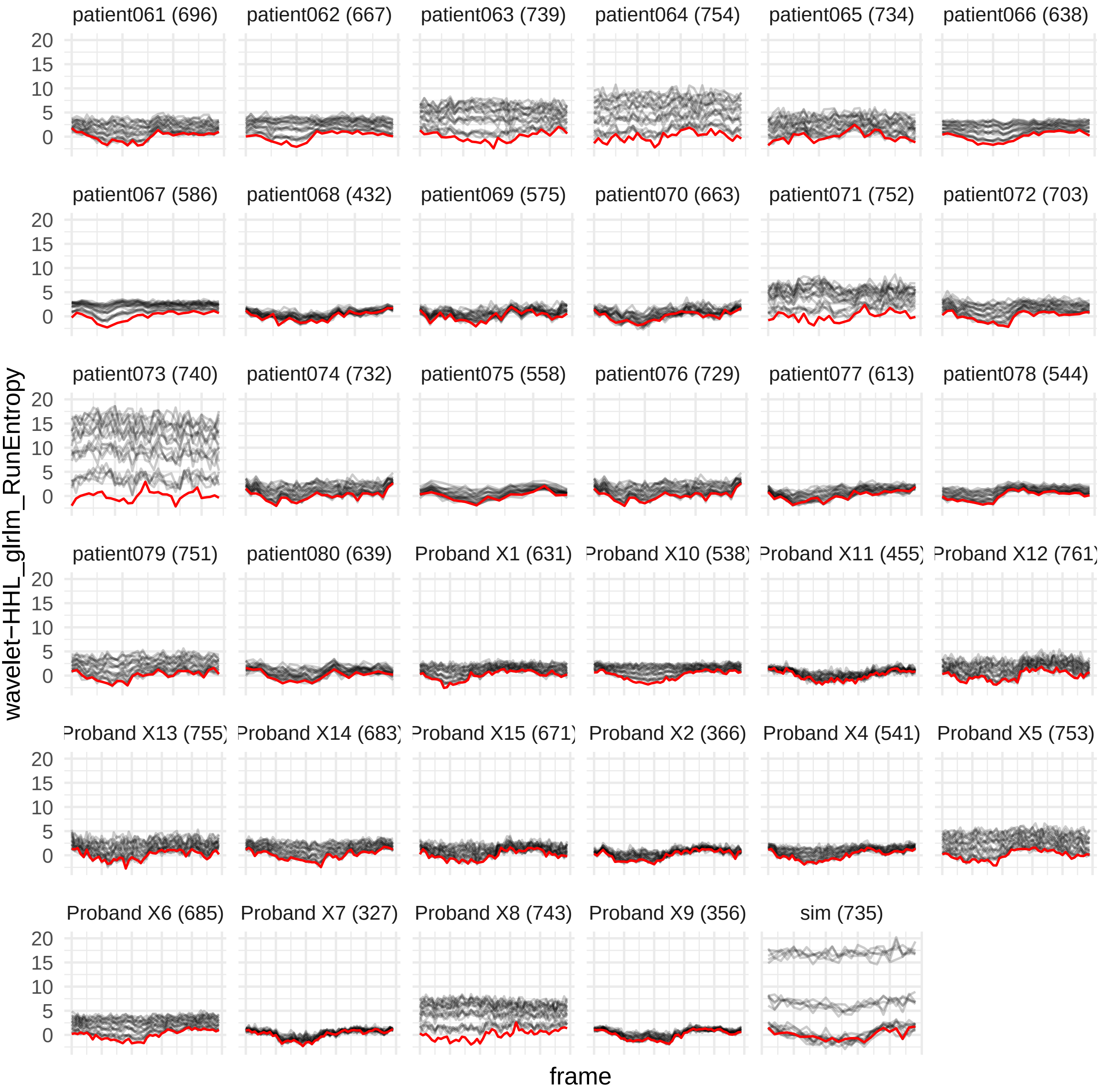

patient061 (696)
patient062 (667)
patient063 (739)
patient064 (754)
patient065 (734)
patient066 (638)
patient067 (586)
patient068 (432)
patient069 (575)
patient070 (663)
patient071 (752)
patient072 (703)
patient073 (740)
patient074 (732)
patient075 (558)
patient076 (729)
patient077 (613)
patient078 (544)
patient079 (751)
patient080 (639)
Proband X1 (631)
Proband X10 (538)
Proband X11 (455)
Proband X12 (761)
Proband X13 (755)
Proband X14 (683)
Proband X15 (671)
Proband X2 (366)
Proband X4 (541)
Proband X5 (753)
Proband X6 (685)
Proband X7 (327)
Proband X8 (743)
Proband X9 (356)
sim (735)
0
5
10
15
20
wavelet−HHL_glrlm_RunEntropy
frame

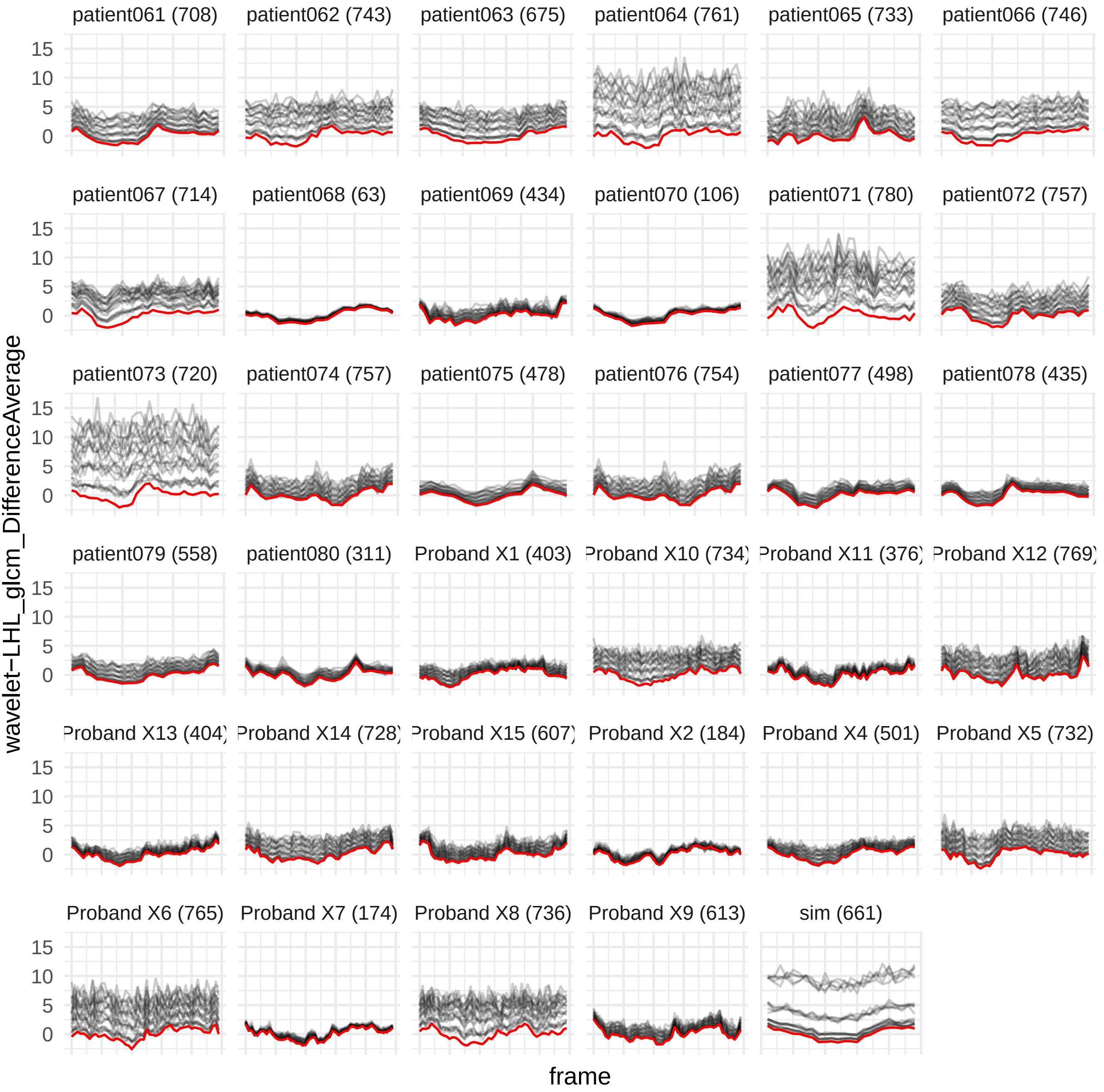

patient061 (708)
patient062 (743)
patient063 (675)
patient064 (761)
patient065 (733)
patient066 (746)
patient067 (714)
patient068 (63)
patient069 (434)
patient070 (106)
patient071 (780)
patient072 (757)
patient073 (720)
patient074 (757)
patient075 (478)
patient076 (754)
patient077 (498)
patient078 (435)
patient079 (558)
patient080 (311)
Proband X1 (403)
Proband X10 (734)
Proband X11 (376)
Proband X12 (769)
Proband X13 (404)
Proband X14 (728)
Proband X15 (607)
Proband X2 (184)
Proband X4 (501)
Proband X5 (732)
Proband X6 (765)
Proband X7 (174)
Proband X8 (736)
Proband X9 (613)
sim (661)
15
10
5
0
wavelet−LHL_glcm_DifferenceAverage
frame

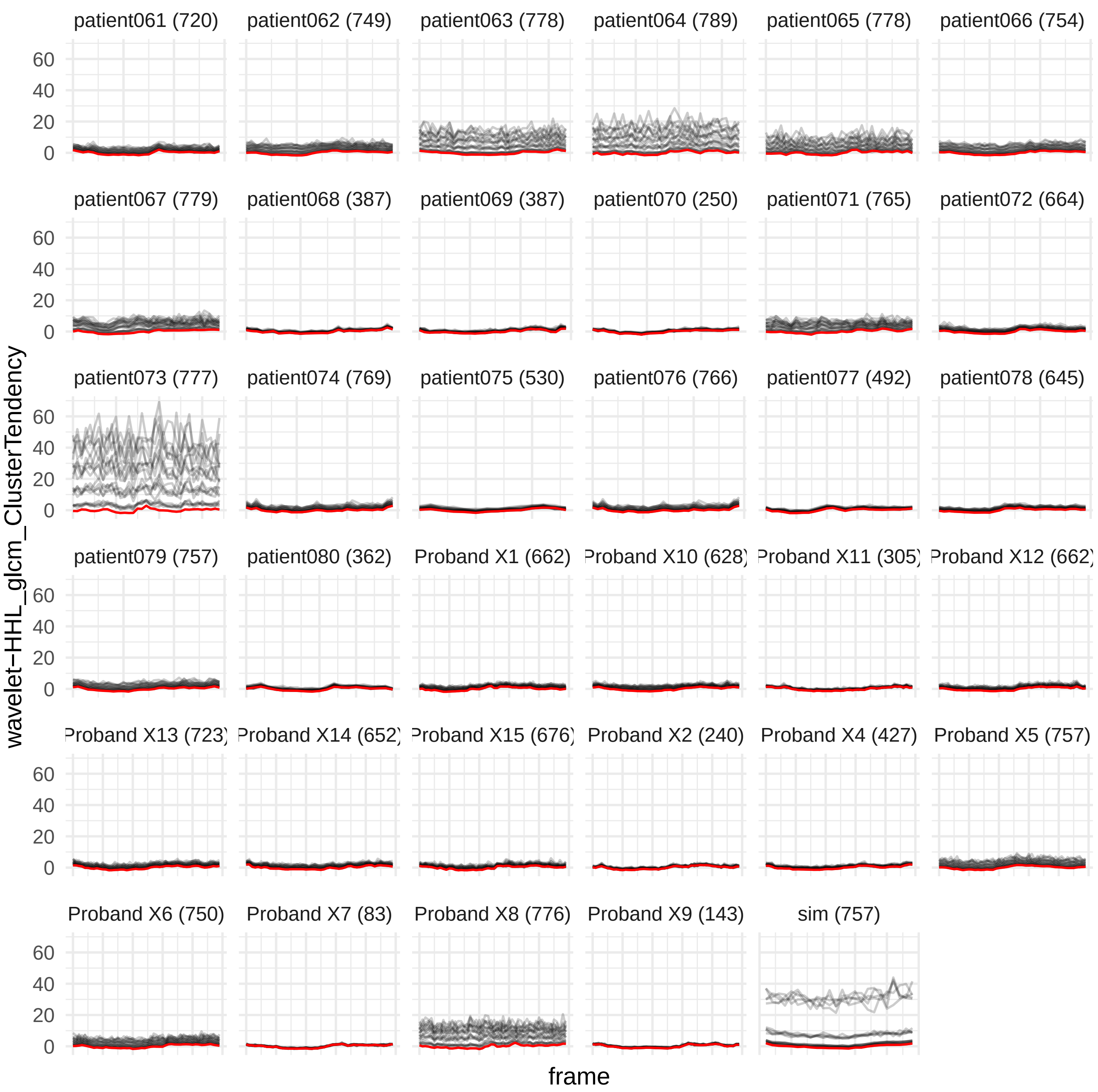

patient061 (720)
patient062 (749)
patient063 (778)
patient064 (789)
patient065 (778)
patient066 (754)
patient067 (779)
patient068 (387)
patient069 (387)
patient070 (250)
patient071 (765)
patient072 (664)
patient073 (777)
patient074 (769)
patient075 (530)
patient076 (766)
patient077 (492)
patient078 (645)
patient079 (757)
patient080 (362)
Proband X1 (662)
Proband X10 (628)
Proband X11 (305)
Proband X12 (662)
Proband X13 (723)
Proband X14 (652)
Proband X15 (676)
Proband X2 (240)
Proband X4 (427)
Proband X5 (757)
Proband X6 (750)
Proband X7 (83)
Proband X8 (776)
Proband X9 (143)
sim (757)
0
20
40
60
wavelet−HHL_glcm_ClusterTendency
frame

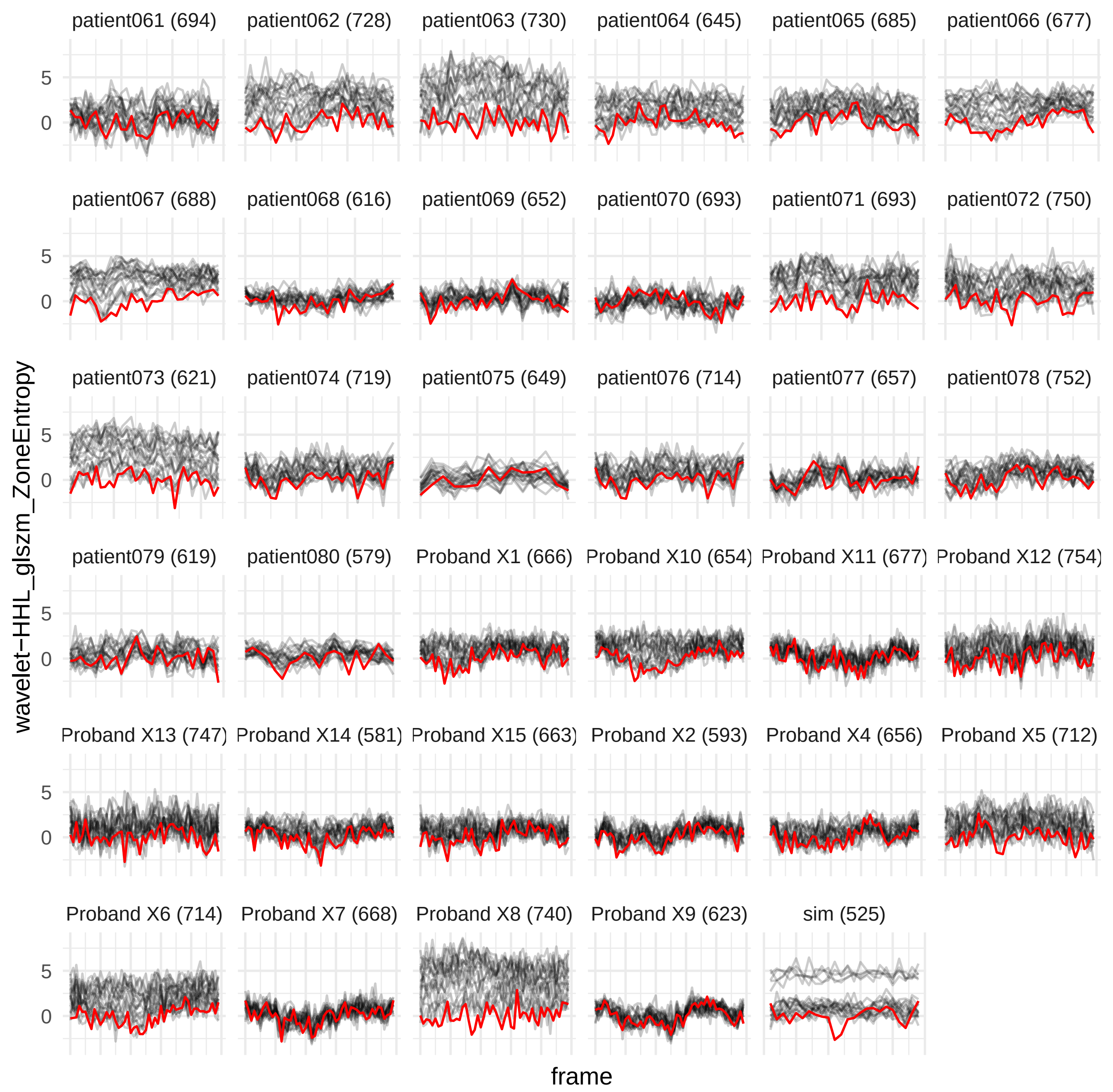
patient061 (694)
patient062 (728)
patient063 (730)
patient064 (645)
patient065 (685)
patient066 (677)
patient067 (688)
patient068 (616)
patient069 (652)
patient070 (693)
patient071 (693)
patient072 (750)
patient073 (621)
patient074 (719)
patient075 (649)
patient076 (714)
patient077 (657)
patient078 (752)
patient079 (619)
patient080 (579)
Proband X1 (666)
Proband X10 (654)
Proband X11 (677)
Proband X12 (754)
Proband X13 (747)
Proband X14 (581)
Proband X15 (663)
Proband X2 (593)
Proband X4 (656)
Proband X5 (712)
Proband X6 (714)
Proband X7 (668)
Proband X8 (740)
Proband X9 (623)
sim (525)
5
0
wavelet−HHL_glszm_ZoneEntropy
frame

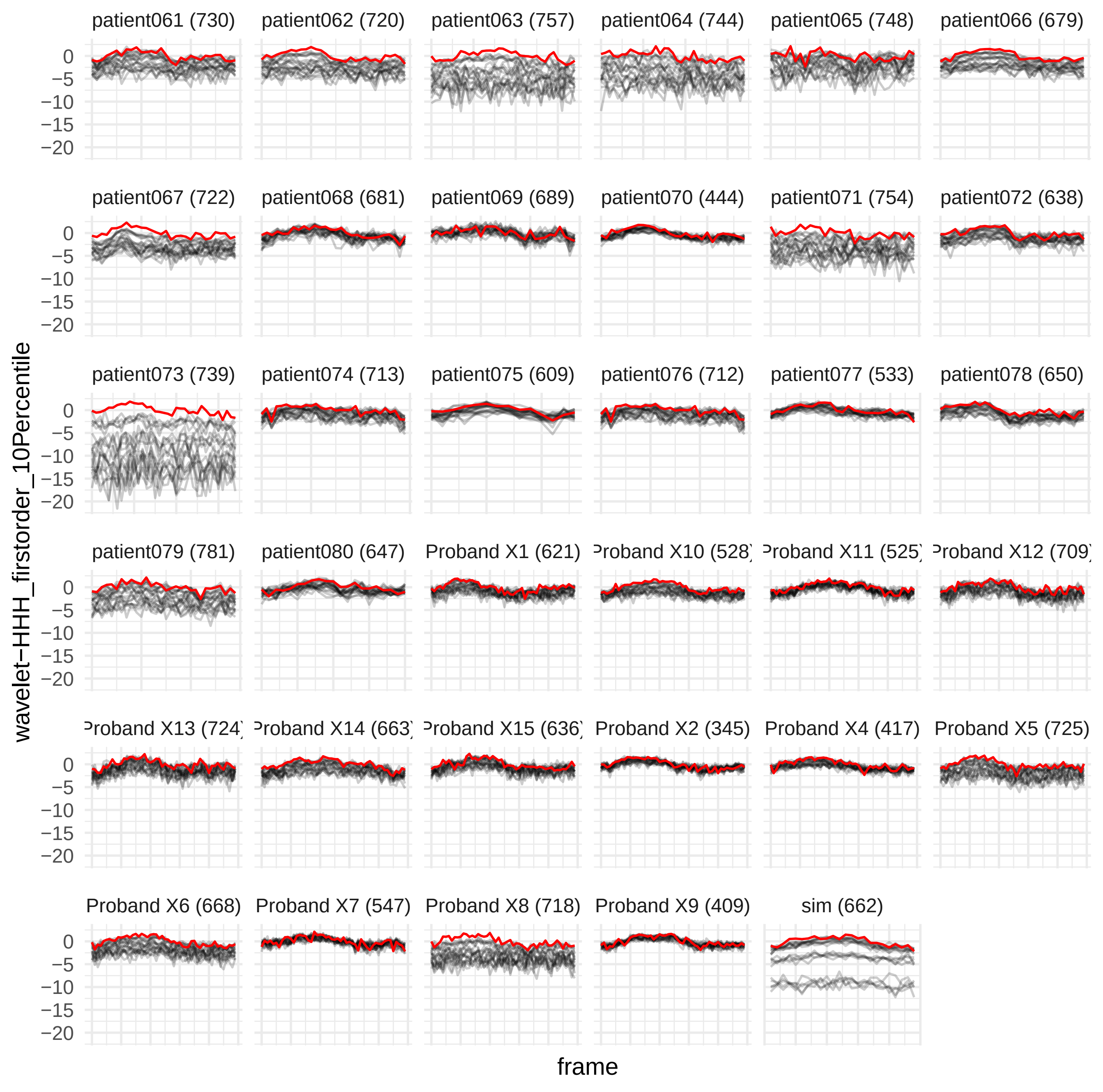

patient061 (730)
patient062 (720)
patient063 (757)
patient064 (744)
patient065 (748)
patient066 (679)
patient067 (722)
patient068 (681)
patient069 (689)
patient070 (444)
patient071 (754)
patient072 (638)
patient073 (739)
patient074 (713)
patient075 (609)
patient076 (712)
patient077 (533)
patient078 (650)
patient079 (781)
patient080 (647)
Proband X1 (621)
Proband X10 (528)
Proband X11 (525)
Proband X12 (709)
Proband X13 (724)
Proband X14 (663)
Proband X15 (636)
Proband X2 (345)
Proband X4 (417)
Proband X5 (725)
Proband X6 (668)
Proband X7 (547)
Proband X8 (718)
Proband X9 (409)
sim (662)
0
−5
−10
−15
−20
wavelet−HHH_firstorder_10Percentile
frame

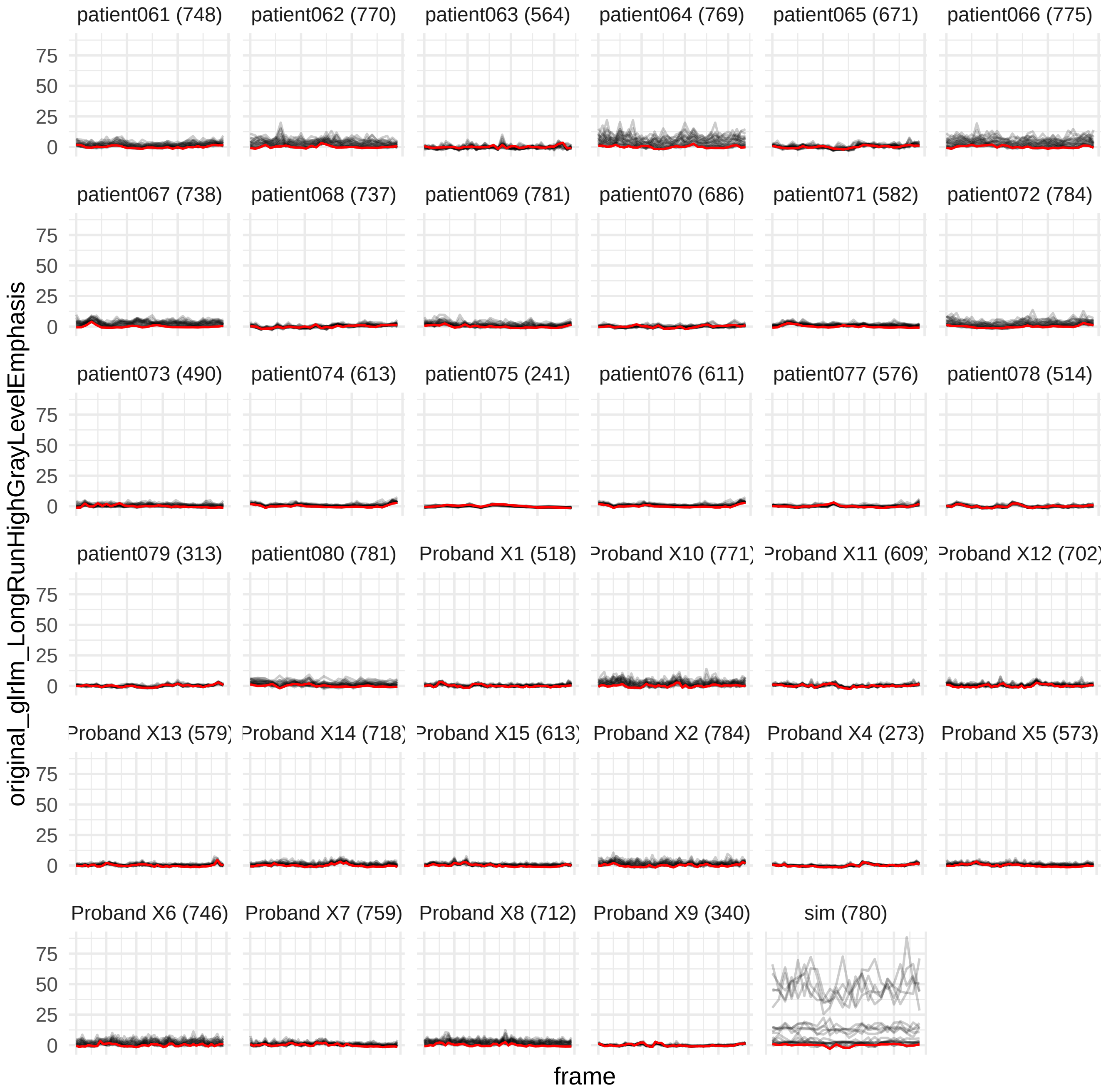

patient061 (748)
patient062 (770)
patient063 (564)
patient064 (769)
patient065 (671)
patient066 (775)
patient067 (738)
patient068 (737)
patient069 (781)
patient070 (686)
patient071 (582)
patient072 (784)
patient073 (490)
patient074 (613)
patient075 (241)
patient076 (611)
patient077 (576)
patient078 (514)
patient079 (313)
patient080 (781)
Proband X1 (518)
Proband X10 (771)
Proband X11 (609)
Proband X12 (702)
Proband X13 (579)
Proband X14 (718)
Proband X15 (613)
Proband X2 (784)
Proband X4 (273)
Proband X5 (573)
Proband X6 (746)
Proband X7 (759)
Proband X8 (712)
Proband X9 (340)
sim (780)
0
25
50
75
original_glrlm_LongRunHighGrayLevelEmphasis
frame

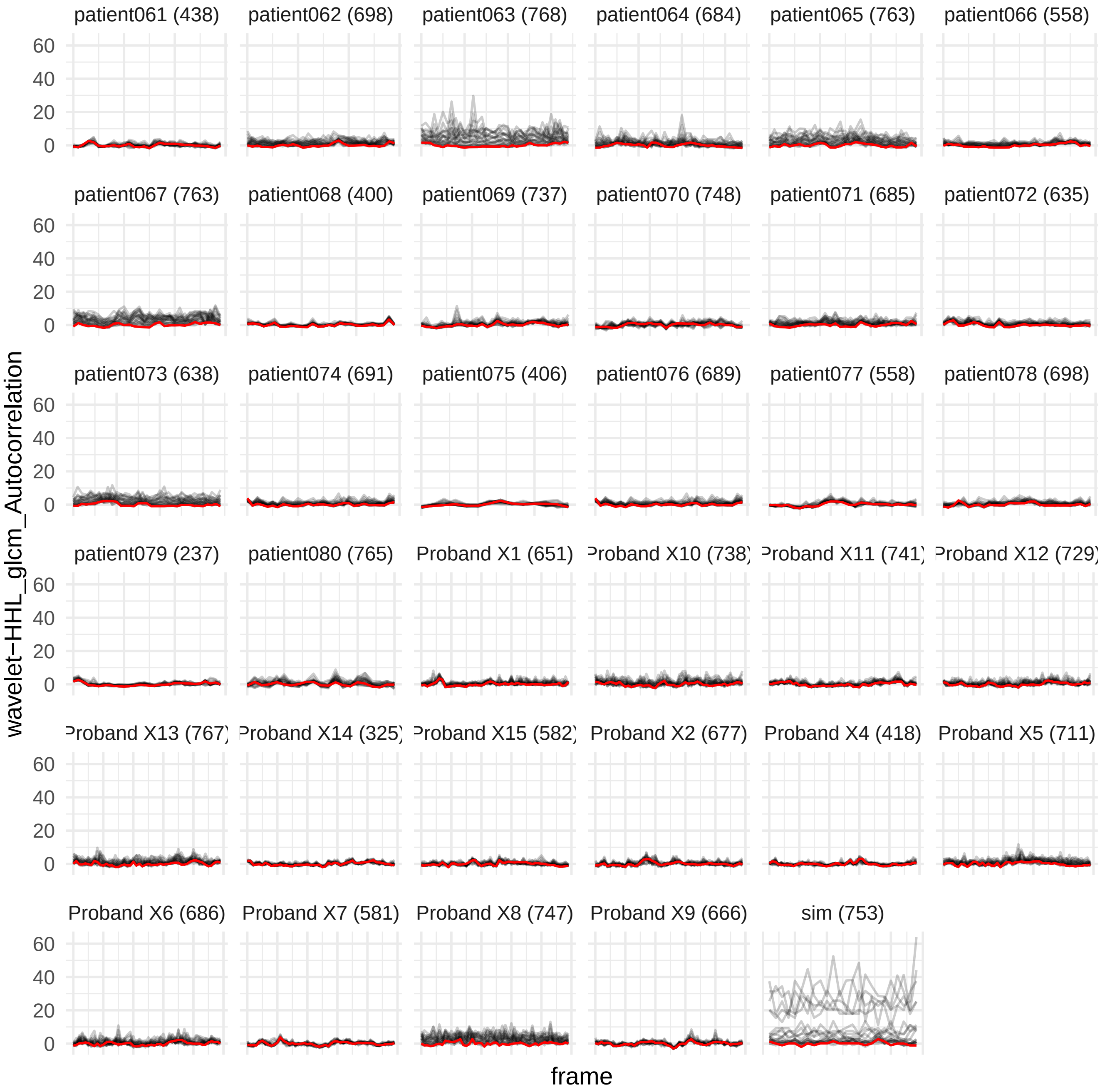
patient061 (438)
patient062 (698)
patient063 (768)
patient064 (684)
patient065 (763)
patient066 (558)
patient067 (763)
patient068 (400)
patient069 (737)
patient070 (748)
patient071 (685)
patient072 (635)
patient073 (638)
patient074 (691)
patient075 (406)
patient076 (689)
patient077 (558)
patient078 (698)
patient079 (237)
patient080 (765)
Proband X1 (651)
Proband X10 (738)
Proband X11 (741)
Proband X12 (729)
Proband X13 (767)
Proband X14 (325)
Proband X15 (582)
Proband X2 (677)
Proband X4 (418)
Proband X5 (711)
Proband X6 (686)
Proband X7 (581)
Proband X8 (747)
Proband X9 (666)
sim (753)
60
40
20
0
wavelet−HHL_glcm_Autocorrelation
frame

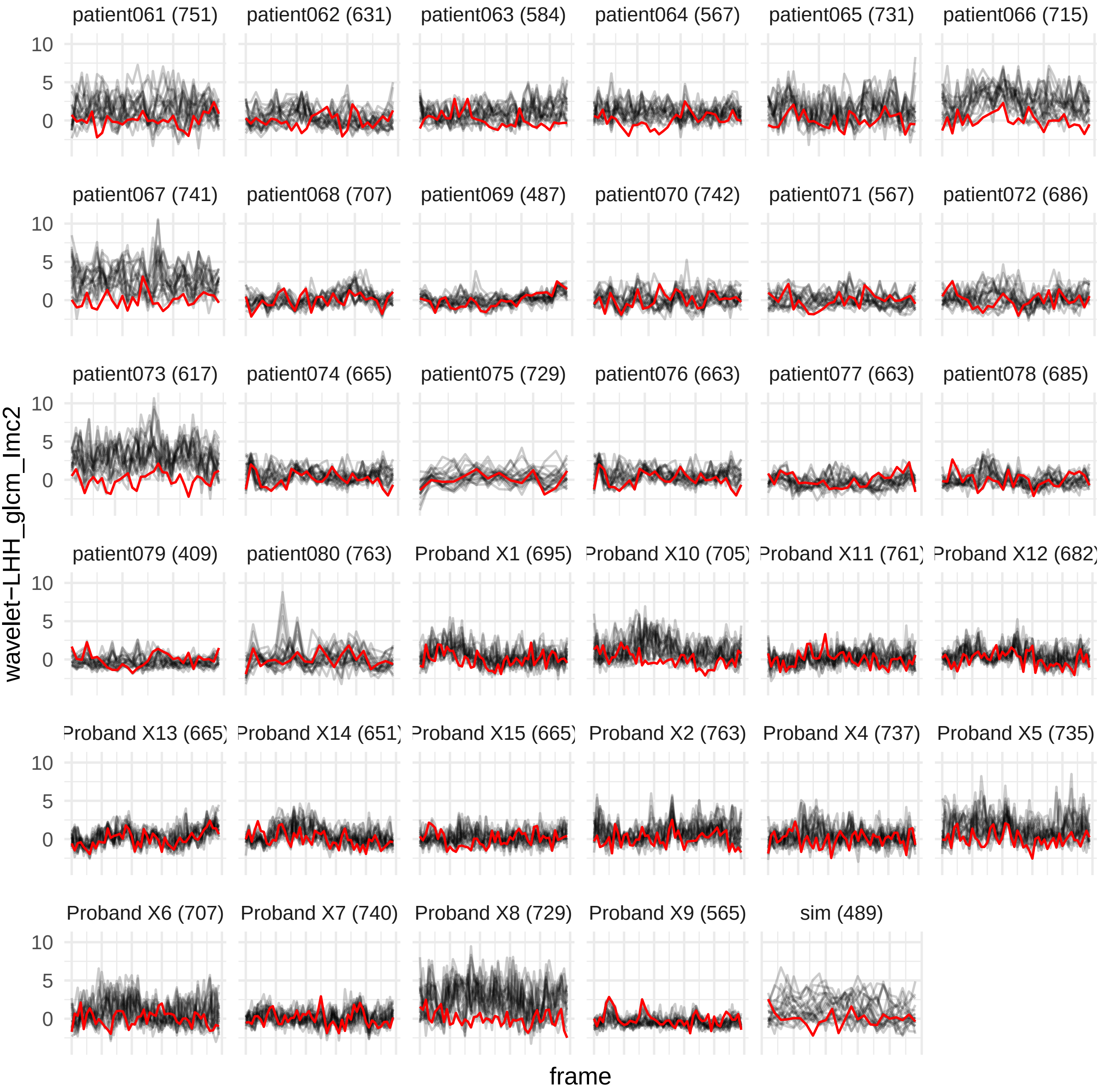

patient061 (751)
patient062 (631)
patient063 (584)
patient064 (567)
patient065 (731)
patient066 (715)
patient067 (741)
patient068 (707)
patient069 (487)
patient070 (742)
patient071 (567)
patient072 (686)
patient073 (617)
patient074 (665)
patient075 (729)
patient076 (663)
patient077 (663)
patient078 (685)
patient079 (409)
patient080 (763)
Proband X1 (695)
Proband X10 (705)
Proband X11 (761)
Proband X12 (682)
Proband X13 (665)
Proband X14 (651)
Proband X15 (665)
Proband X2 (763)
Proband X4 (737)
Proband X5 (735)
Proband X6 (707)
Proband X7 (740)
Proband X8 (729)
Proband X9 (565)
sim (489)
10
5
0
wavelet−LHH_glcm_Imc2
frame

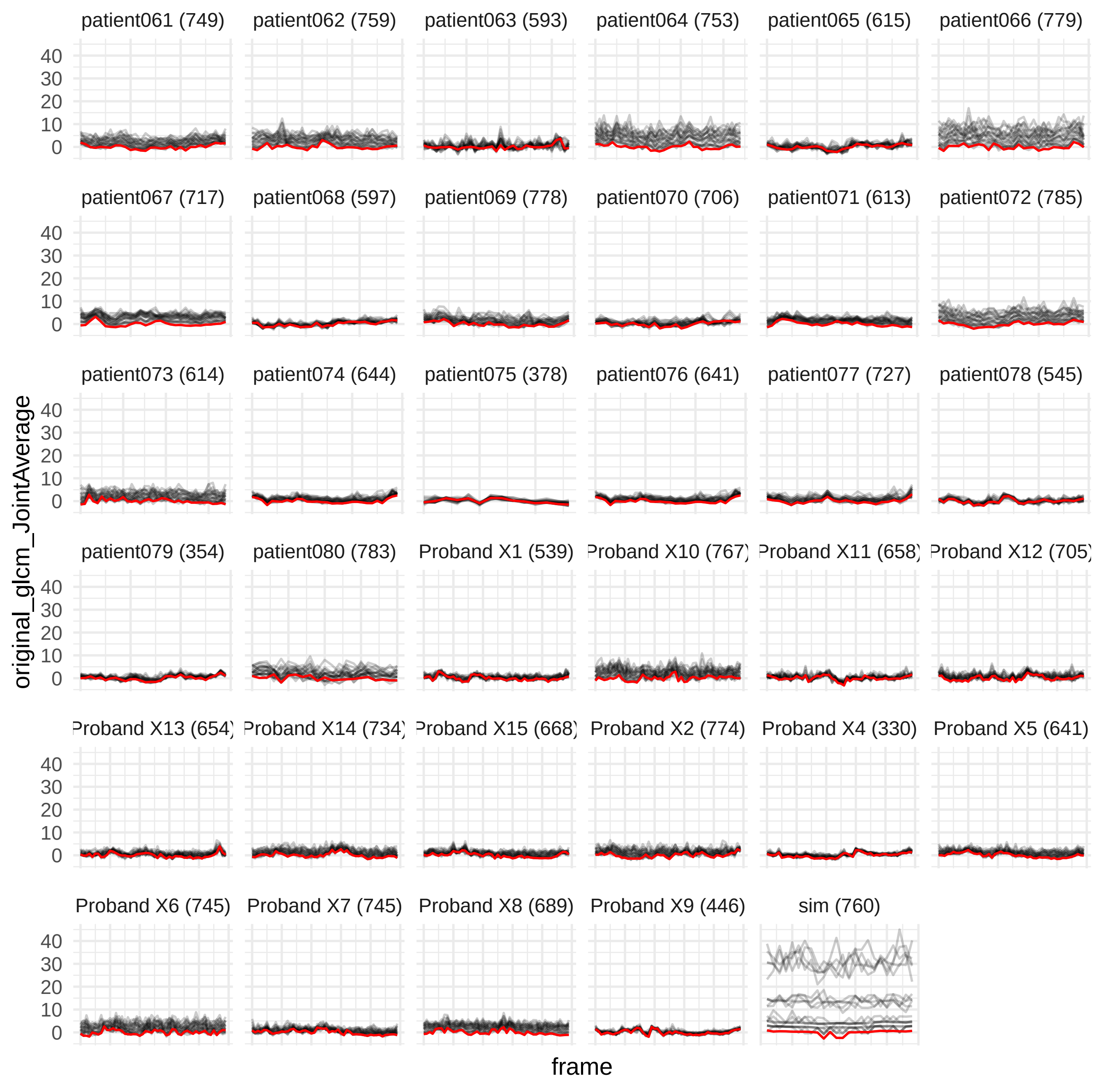
patient061 (749)
patient062 (759)
patient063 (593)
patient064 (753)
patient065 (615)
patient066 (779)
patient067 (717)
patient068 (597)
patient069 (778)
patient070 (706)
patient071 (613)
patient072 (785)
patient073 (614)
patient074 (644)
patient075 (378)
patient076 (641)
patient077 (727)
patient078 (545)
patient079 (354)
patient080 (783)
Proband X1 (539)
Proband X10 (767)
Proband X11 (658)
Proband X12 (705)
Proband X13 (654)
Proband X14 (734)
Proband X15 (668)
Proband X2 (774)
Proband X4 (330)
Proband X5 (641)
Proband X6 (745)
Proband X7 (745)
Proband X8 (689)
Proband X9 (446)
sim (760)
original_glcm_JointAverage
frame
40
30
20
10
0

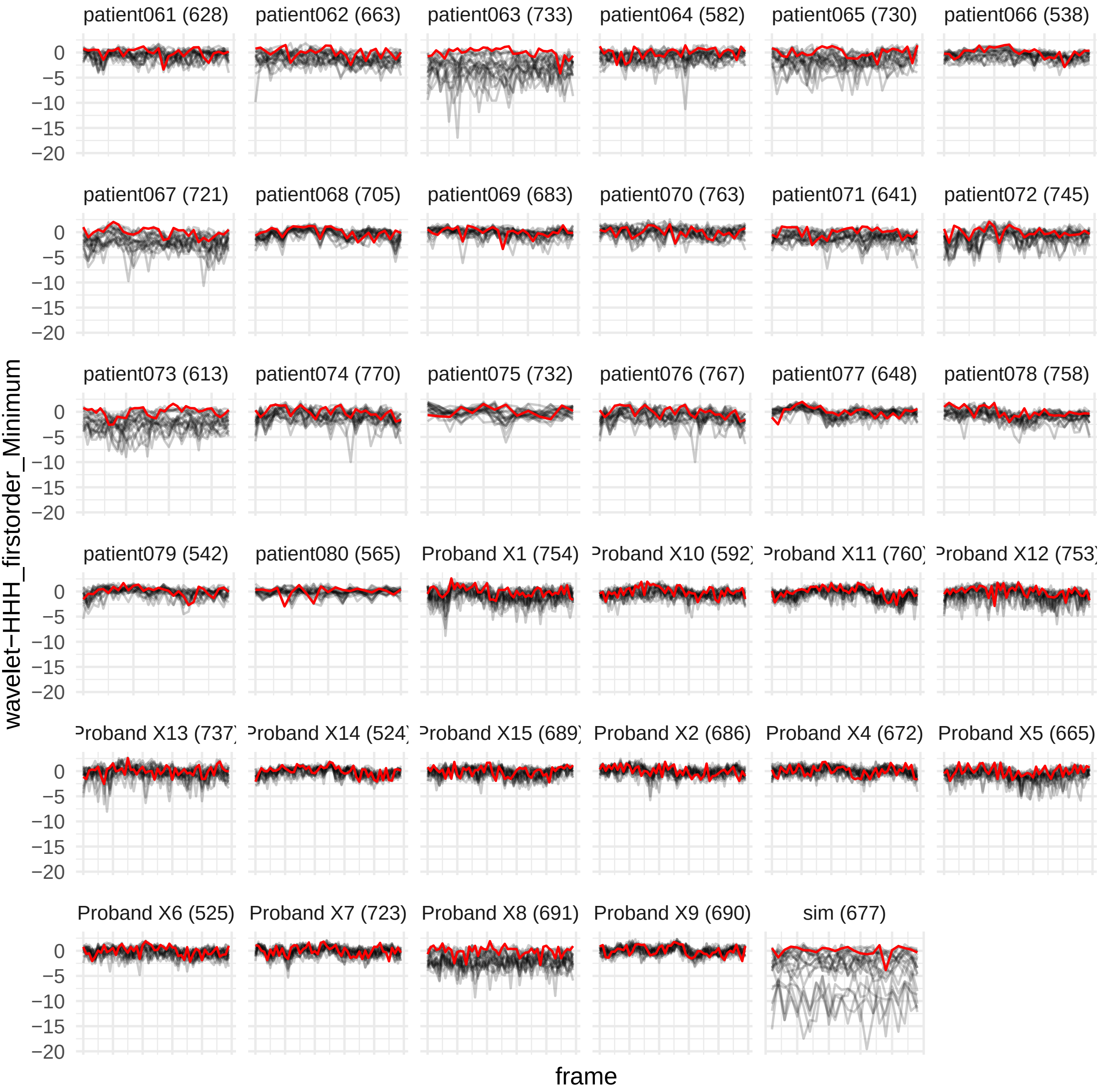

patient061 (628)
patient062 (663)
patient063 (733)
patient064 (582)
patient065 (730)
patient066 (538)
patient067 (721)
patient068 (705)
patient069 (683)
patient070 (763)
patient071 (641)
patient072 (745)
patient073 (613)
patient074 (770)
patient075 (732)
patient076 (767)
patient077 (648)
patient078 (758)
patient079 (542)
patient080 (565)
Proband X1 (754)
Proband X10 (592)
Proband X11 (760)
Proband X12 (753)
Proband X13 (737)
Proband X14 (524)
Proband X15 (689)
Proband X2 (686)
Proband X4 (672)
Proband X5 (665)
Proband X6 (525)
Proband X7 (723)
Proband X8 (691)
Proband X9 (690)
sim (677)
0
−5
−10
−15
−20
wavelet−HHH_firstorder_Minimum
frame

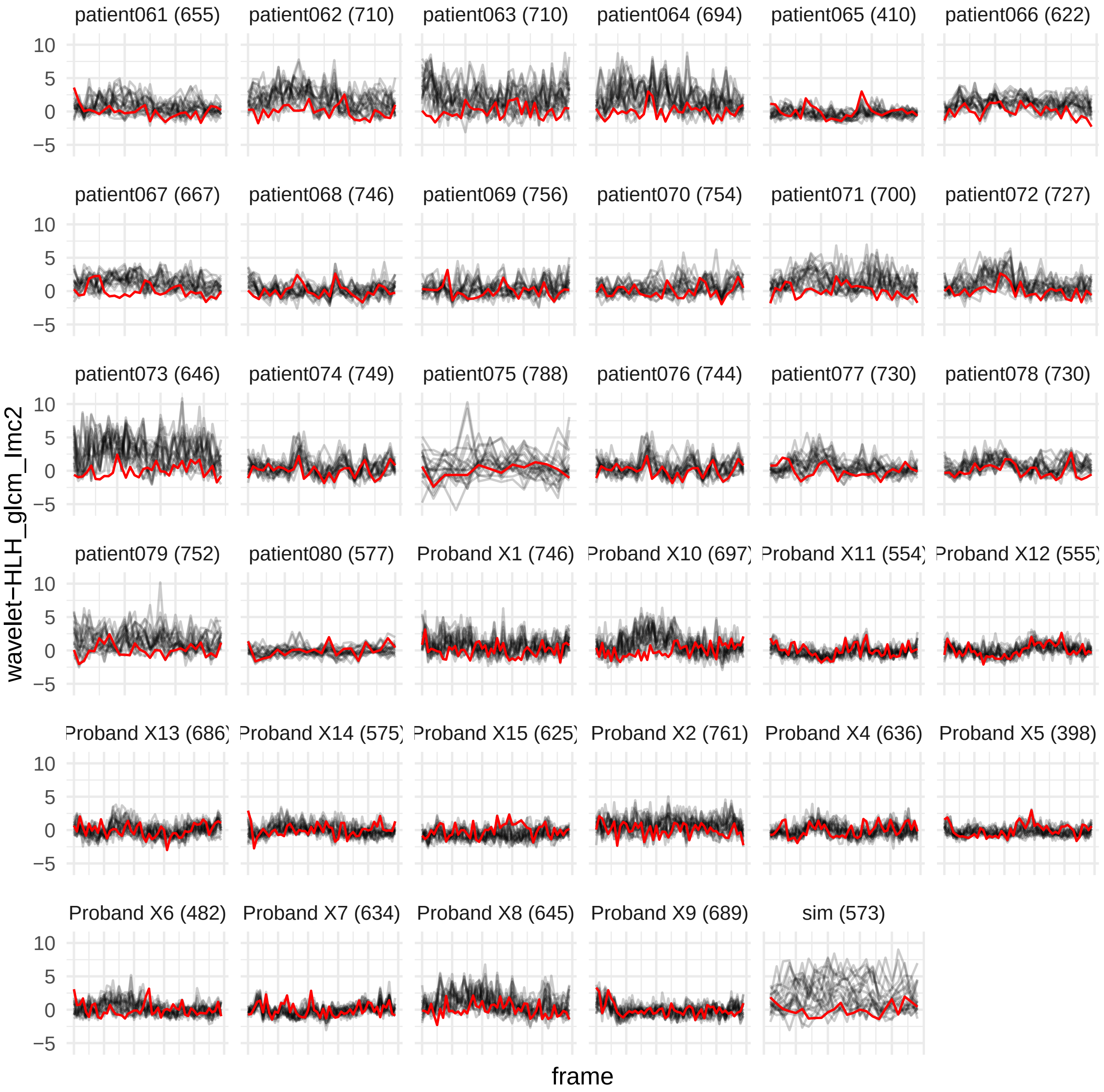

patient061 (655)
patient062 (710)
patient063 (710)
patient064 (694)
patient065 (410)
patient066 (622)
patient067 (667)
patient068 (746)
patient069 (756)
patient070 (754)
patient071 (700)
patient072 (727)
patient073 (646)
patient074 (749)
patient075 (788)
patient076 (744)
patient077 (730)
patient078 (730)
patient079 (752)
patient080 (577)
Proband X1 (746)
Proband X10 (697)
Proband X11 (554)
Proband X12 (555)
Proband X13 (686)
Proband X14 (575)
Proband X15 (625)
Proband X2 (761)
Proband X4 (636)
Proband X5 (398)
Proband X6 (482)
Proband X7 (634)
Proband X8 (645)
Proband X9 (689)
sim (573)
wavelet−HLH_glcm_Imc2
10
5
0
−5
frame

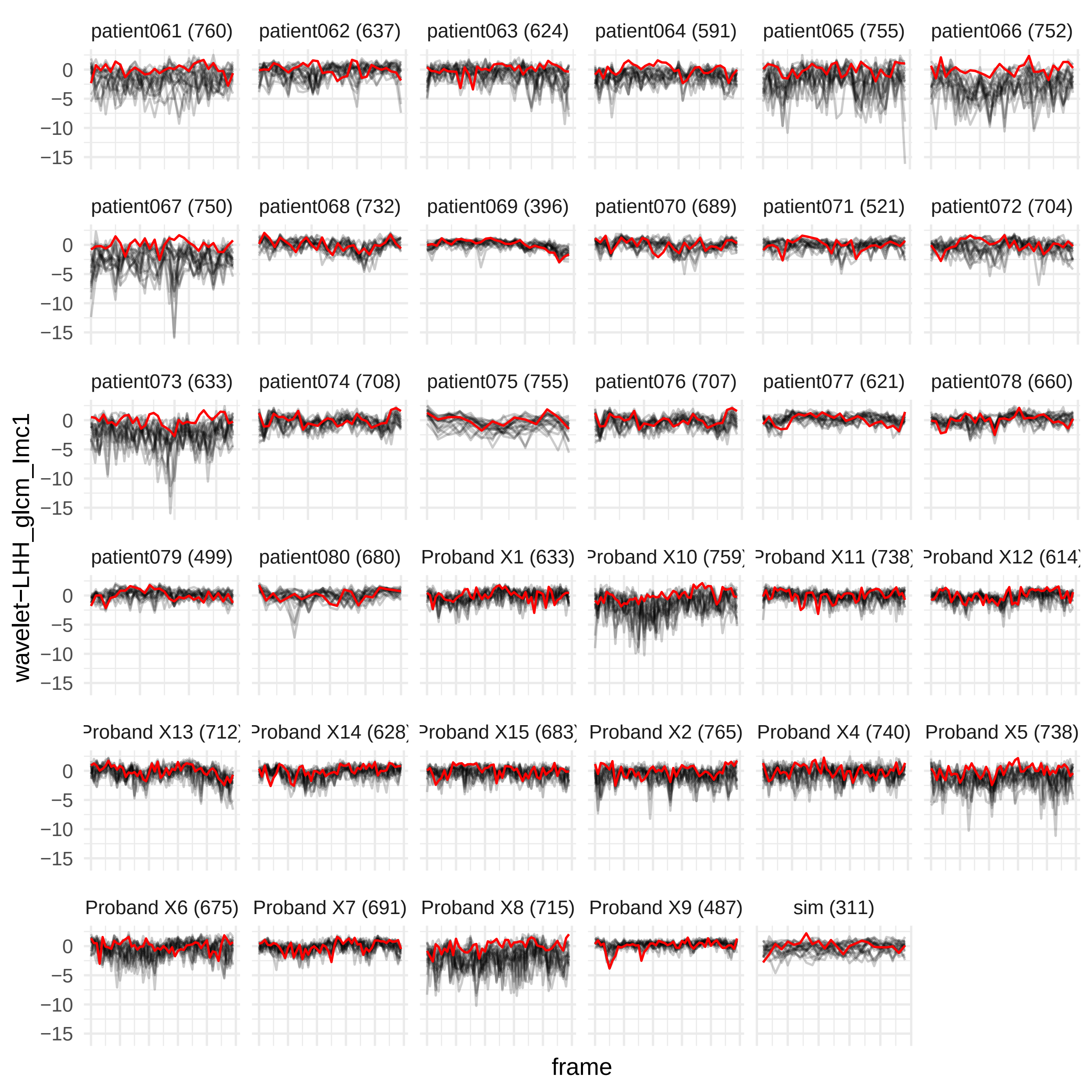

patient061 (760)
patient062 (637)
patient063 (624)
patient064 (591)
patient065 (755)
patient066 (752)
patient067 (750)
patient068 (732)
patient069 (396)
patient070 (689)
patient071 (521)
patient072 (704)
patient073 (633)
patient074 (708)
patient075 (755)
patient076 (707)
patient077 (621)
patient078 (660)
patient079 (499)
patient080 (680)
Proband X1 (633)
Proband X10 (759)
Proband X11 (738)
Proband X12 (614)
Proband X13 (712)
Proband X14 (628)
Proband X15 (683)
Proband X2 (765)
Proband X4 (740)
Proband X5 (738)
Proband X6 (675)
Proband X7 (691)
Proband X8 (715)
Proband X9 (487)
sim (311)
0
−5
−10
−15
wavelet−LHH_glcm_Imc1
frame

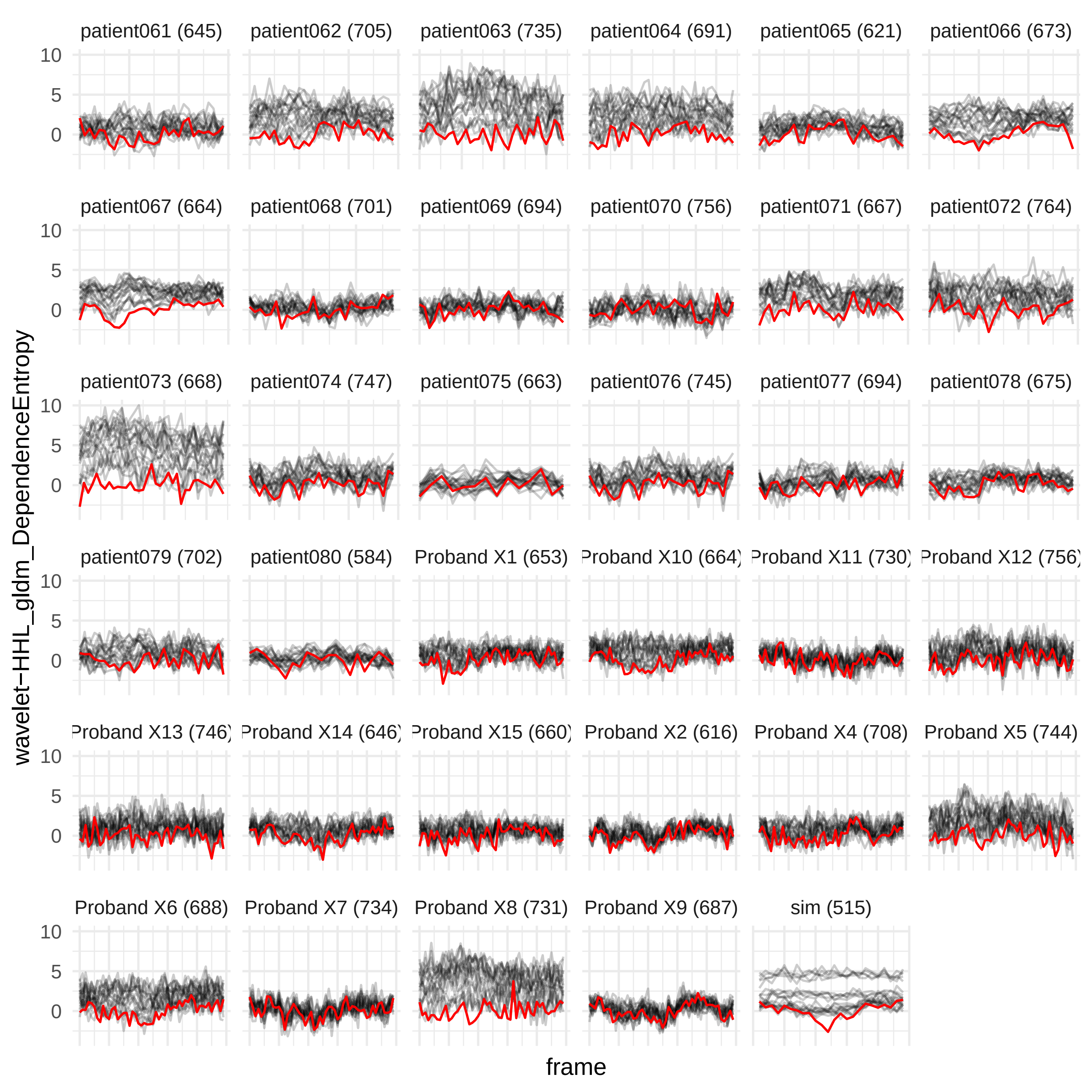

patient061 (645)
patient062 (705)
patient063 (735)
patient064 (691)
patient065 (621)
patient066 (673)
patient067 (664)
patient068 (701)
patient069 (694)
patient070 (756)
patient071 (667)
patient072 (764)
patient073 (668)
patient074 (747)
patient075 (663)
patient076 (745)
patient077 (694)
patient078 (675)
patient079 (702)
patient080 (584)
Proband X1 (653)
Proband X10 (664)
Proband X11 (730)
Proband X12 (756)
Proband X13 (746)
Proband X14 (646)
Proband X15 (660)
Proband X2 (616)
Proband X4 (708)
Proband X5 (744)
Proband X6 (688)
Proband X7 (734)
Proband X8 (731)
Proband X9 (687)
sim (515)
10
5
0
wavelet−HHL_gldm_DependenceEntropy
frame

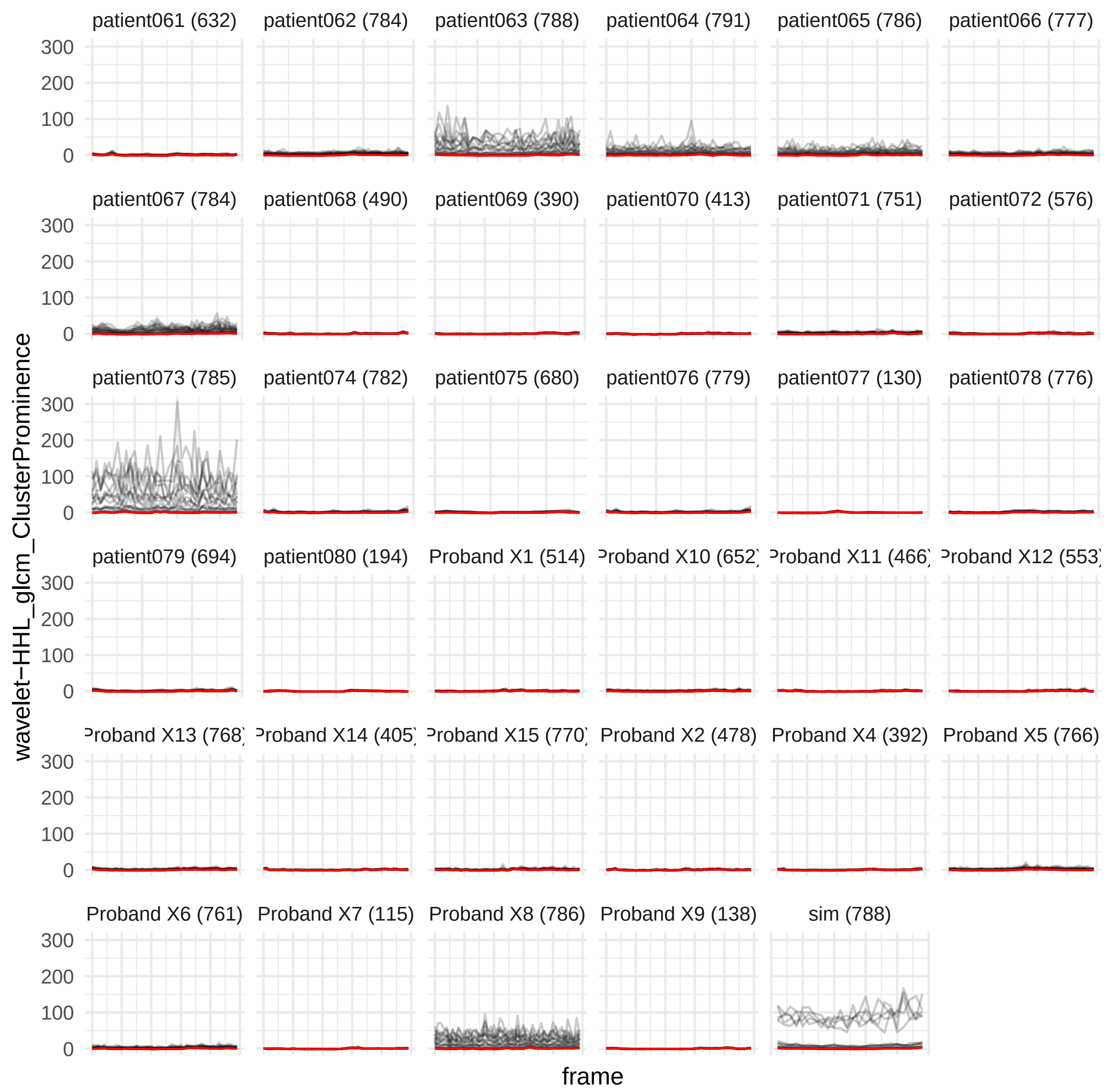
patient061 (632)
patient062 (784)
patient063 (788)
patient064 (791)
patient065 (786)
patient066 (777)
patient067 (784)
patient068 (490)
patient069 (390)
patient070 (413)
patient071 (751)
patient072 (576)
patient073 (785)
patient074 (782)
patient075 (680)
patient076 (779)
patient077 (130)
patient078 (776)
patient079 (694)
patient080 (194)
Proband X1 (514)
Proband X10 (652)
Proband X11 (466)
Proband X12 (553)
Proband X13 (768)
Proband X14 (405)
Proband X15 (770)
Proband X2 (478)
Proband X4 (392)
Proband X5 (766)
Proband X6 (761)
Proband X7 (115)
Proband X8 (786)
Proband X9 (138)
sim (788)
300
200
100
0
wavelet−HHL_glcm_ClusterProminence
frame

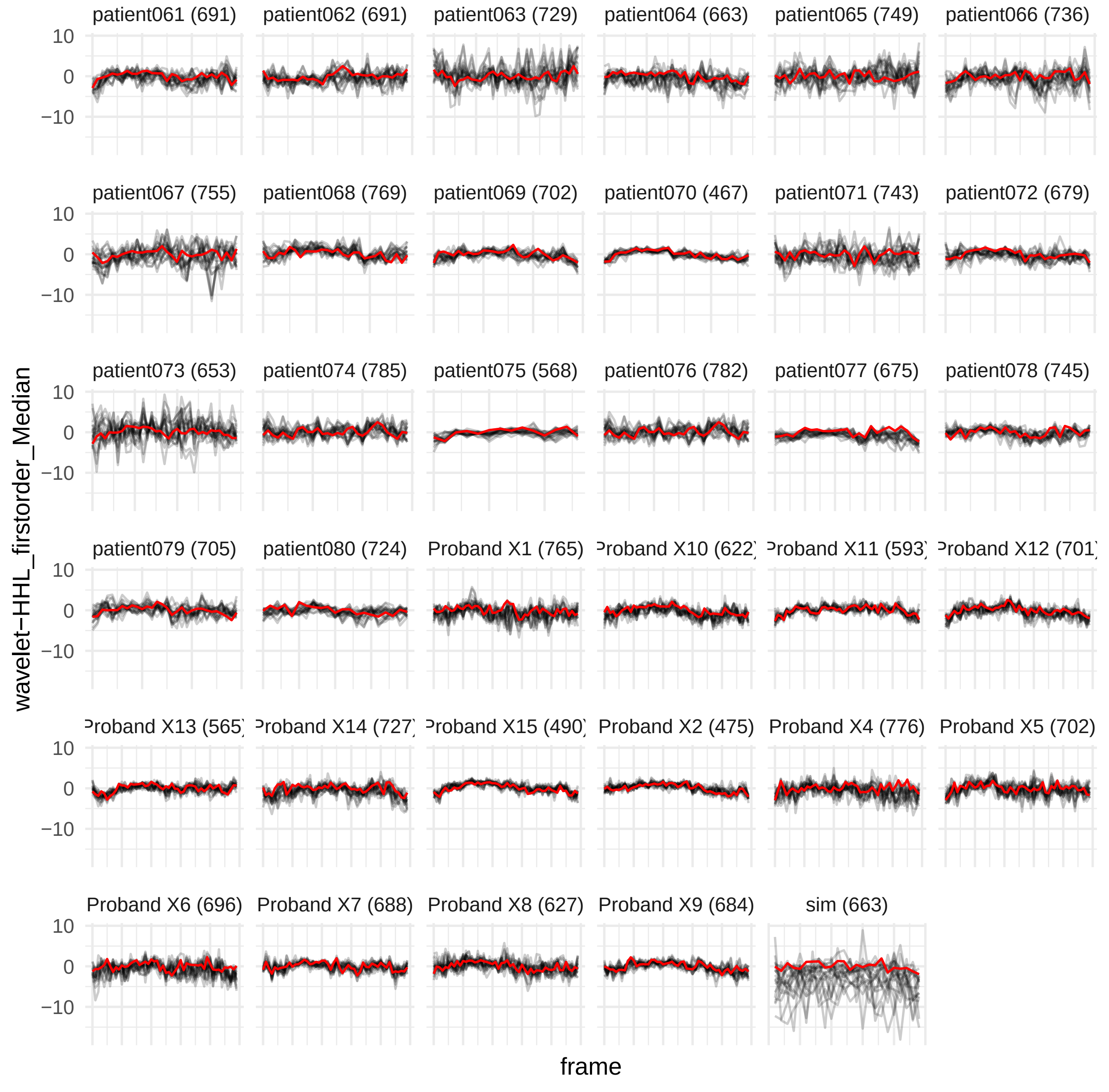

patient061 (691)
patient062 (691)
patient063 (729)
patient064 (663)
patient065 (749)
patient066 (736)
patient067 (755)
patient068 (769)
patient069 (702)
patient070 (467)
patient071 (743)
patient072 (679)
patient073 (653)
patient074 (785)
patient075 (568)
patient076 (782)
patient077 (675)
patient078 (745)
patient079 (705)
patient080 (724)
Proband X1 (765)
Proband X10 (622)
Proband X11 (593)
Proband X12 (701)
Proband X13 (565)
Proband X14 (727)
Proband X15 (490)
Proband X2 (475)
Proband X4 (776)
Proband X5 (702)
Proband X6 (696)
Proband X7 (688)
Proband X8 (627)
Proband X9 (684)
sim (663)
10
0
−10
wavelet−HHL_firstorder_Median
frame

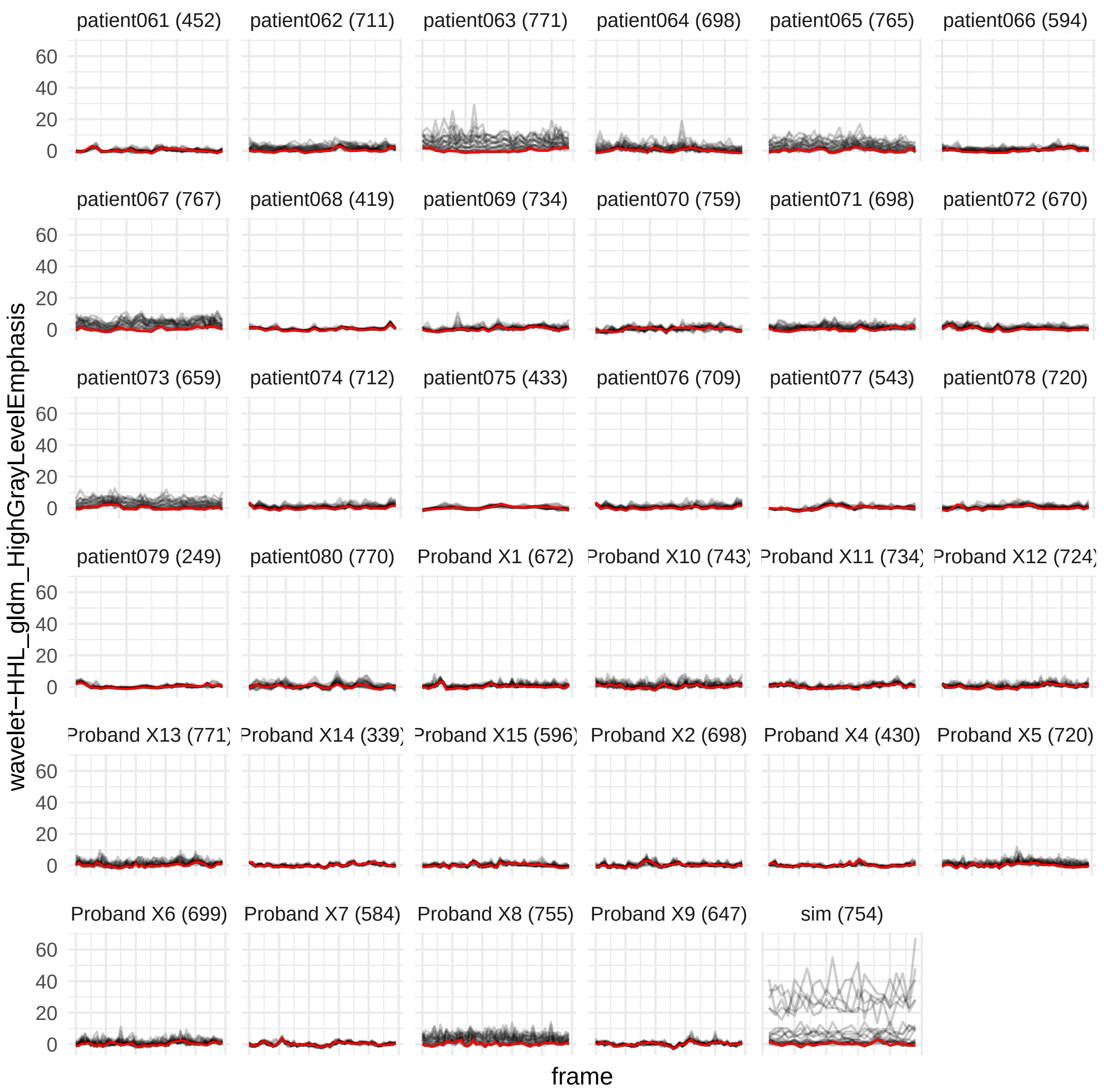

patient061 (452)
patient062 (711)
patient063 (771)
patient064 (698)
patient065 (765)
patient066 (594)
patient067 (767)
patient068 (419)
patient069 (734)
patient070 (759)
patient071 (698)
patient072 (670)
patient073 (659)
patient074 (712)
patient075 (433)
patient076 (709)
patient077 (543)
patient078 (720)
patient079 (249)
patient080 (770)
Proband X1 (672)
Proband X10 (743)
Proband X11 (734)
Proband X12 (724)
Proband X13 (771)
Proband X14 (339)
Proband X15 (596)
Proband X2 (698)
Proband X4 (430)
Proband X5 (720)
Proband X6 (699)
Proband X7 (584)
Proband X8 (755)
Proband X9 (647)
sim (754)
wavelet−HHL_gldm_HighGrayLevelEmphasis
frame
0
20
40
60

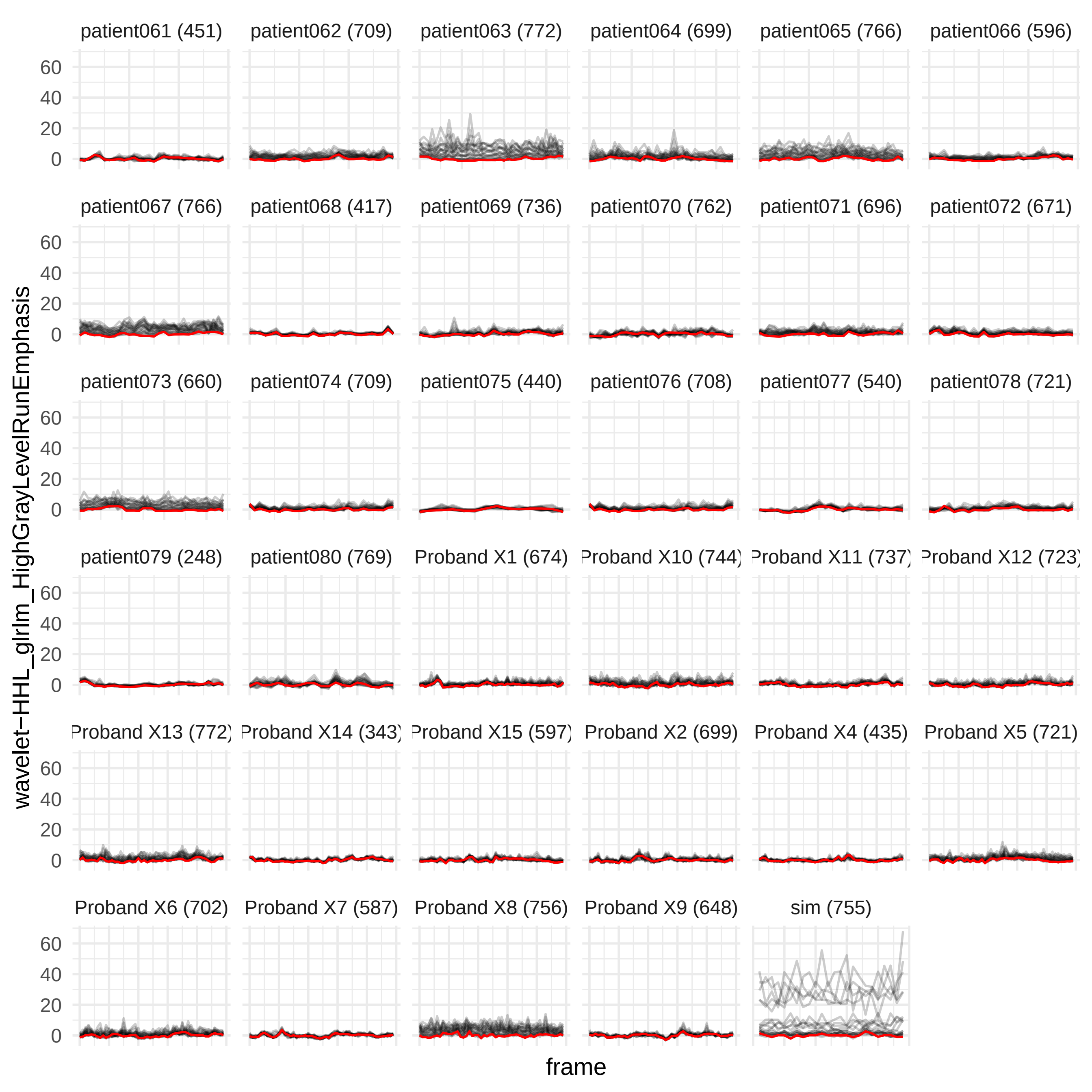
patient061 (451)
patient062 (709)
patient063 (772)
patient064 (699)
patient065 (766)
patient066 (596)
patient067 (766)
patient068 (417)
patient069 (736)
patient070 (762)
patient071 (696)
patient072 (671)
patient073 (660)
patient074 (709)
patient075 (440)
patient076 (708)
patient077 (540)
patient078 (721)
patient079 (248)
patient080 (769)
Proband X1 (674)
Proband X10 (744)
Proband X11 (737)
Proband X12 (723)
Proband X13 (772)
Proband X14 (343)
Proband X15 (597)
Proband X2 (699)
Proband X4 (435)
Proband X5 (721)
Proband X6 (702)
Proband X7 (587)
Proband X8 (756)
Proband X9 (648)
sim (755)
wavelet−HHL_glrlm_HighGrayLevelRunEmphasis
frame
0
20
40
60

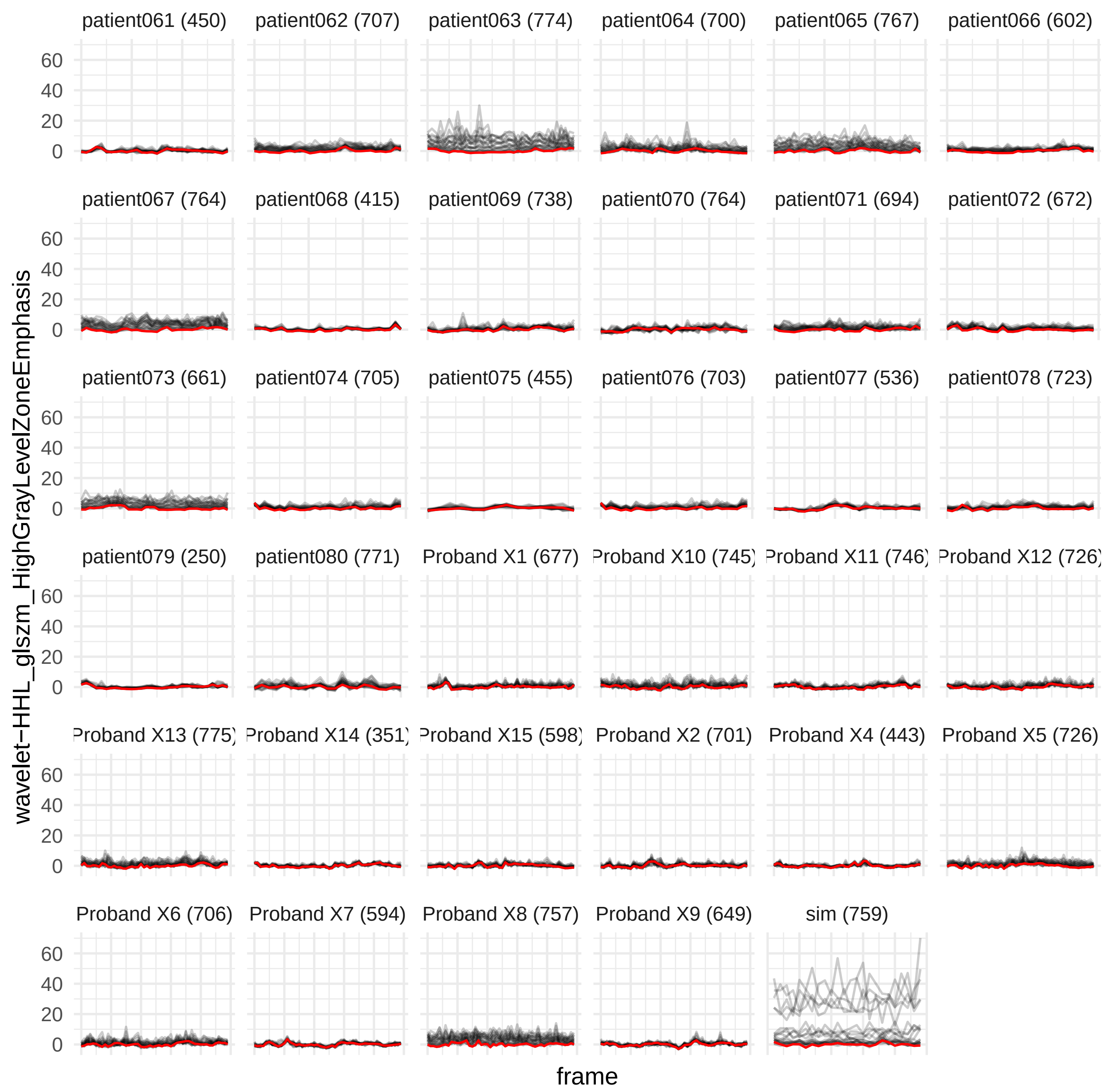

patient061 (450)
patient062 (707)
patient063 (774)
patient064 (700)
patient065 (767)
patient066 (602)
patient067 (764)
patient068 (415)
patient069 (738)
patient070 (764)
patient071 (694)
patient072 (672)
patient073 (661)
patient074 (705)
patient075 (455)
patient076 (703)
patient077 (536)
patient078 (723)
patient079 (250)
patient080 (771)
Proband X1 (677)
Proband X10 (745)
Proband X11 (746)
Proband X12 (726)
Proband X13 (775)
Proband X14 (351)
Proband X15 (598)
Proband X2 (701)
Proband X4 (443)
Proband X5 (726)
Proband X6 (706)
Proband X7 (594)
Proband X8 (757)
Proband X9 (649)
sim (759)
wavelet−HHL_glszm_HighGrayLevelZoneEmphasis
frame
0
20
40
60

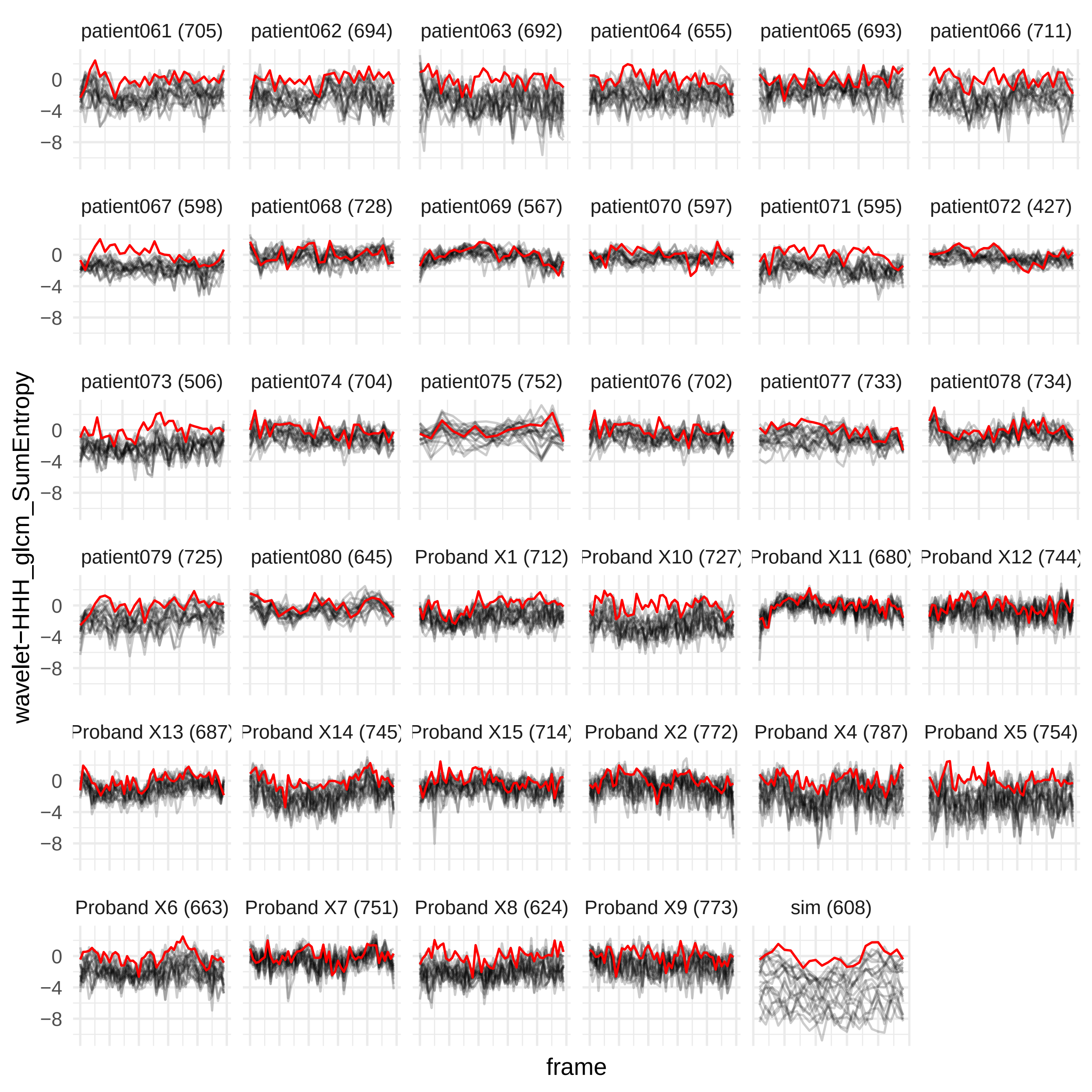

patient061 (705)
patient062 (694)
patient063 (692)
patient064 (655)
patient065 (693)
patient066 (711)
patient067 (598)
patient068 (728)
patient069 (567)
patient070 (597)
patient071 (595)
patient072 (427)
patient073 (506)
patient074 (704)
patient075 (752)
patient076 (702)
patient077 (733)
patient078 (734)
patient079 (725)
patient080 (645)
Proband X1 (712)
Proband X10 (727)
Proband X11 (680)
Proband X12 (744)
Proband X13 (687)
Proband X14 (745)
Proband X15 (714)
Proband X2 (772)
Proband X4 (787)
Proband X5 (754)
Proband X6 (663)
Proband X7 (751)
Proband X8 (624)
Proband X9 (773)
sim (608)
0
−4
−8
wavelet−HHH_glcm_SumEntropy
frame

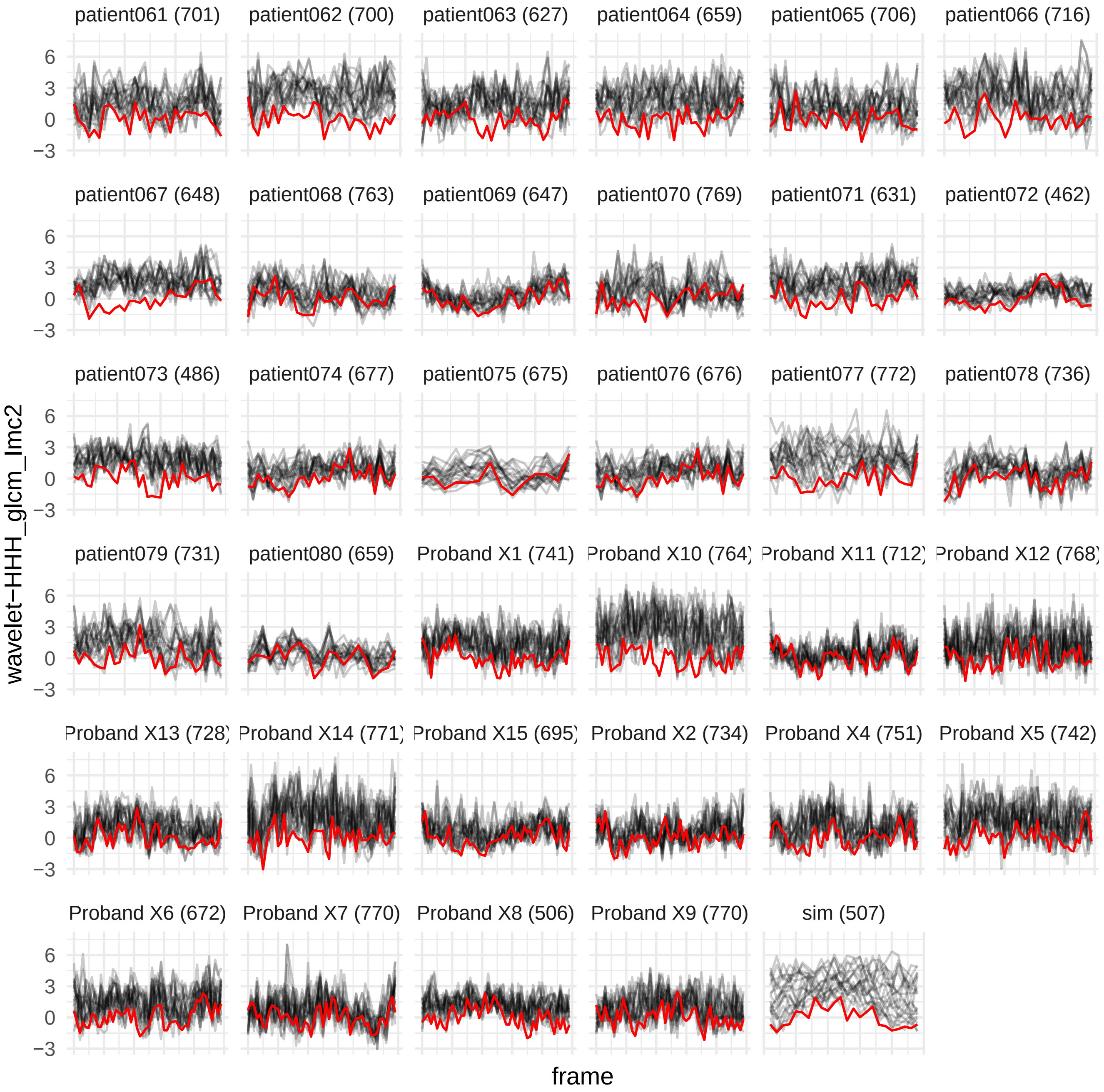

patient061 (701)
patient062 (700)
patient063 (627)
patient064 (659)
patient065 (706)
patient066 (716)
patient067 (648)
patient068 (763)
patient069 (647)
patient070 (769)
patient071 (631)
patient072 (462)
patient073 (486)
patient074 (677)
patient075 (675)
patient076 (676)
patient077 (772)
patient078 (736)
patient079 (731)
patient080 (659)
Proband X1 (741)
Proband X10 (764)
Proband X11 (712)
Proband X12 (768)
Proband X13 (728)
Proband X14 (771)
Proband X15 (695)
Proband X2 (734)
Proband X4 (751)
Proband X5 (742)
Proband X6 (672)
Proband X7 (770)
Proband X8 (506)
Proband X9 (770)
sim (507)
6
3
0
−3
wavelet−HHH_glcm_Imc2
frame

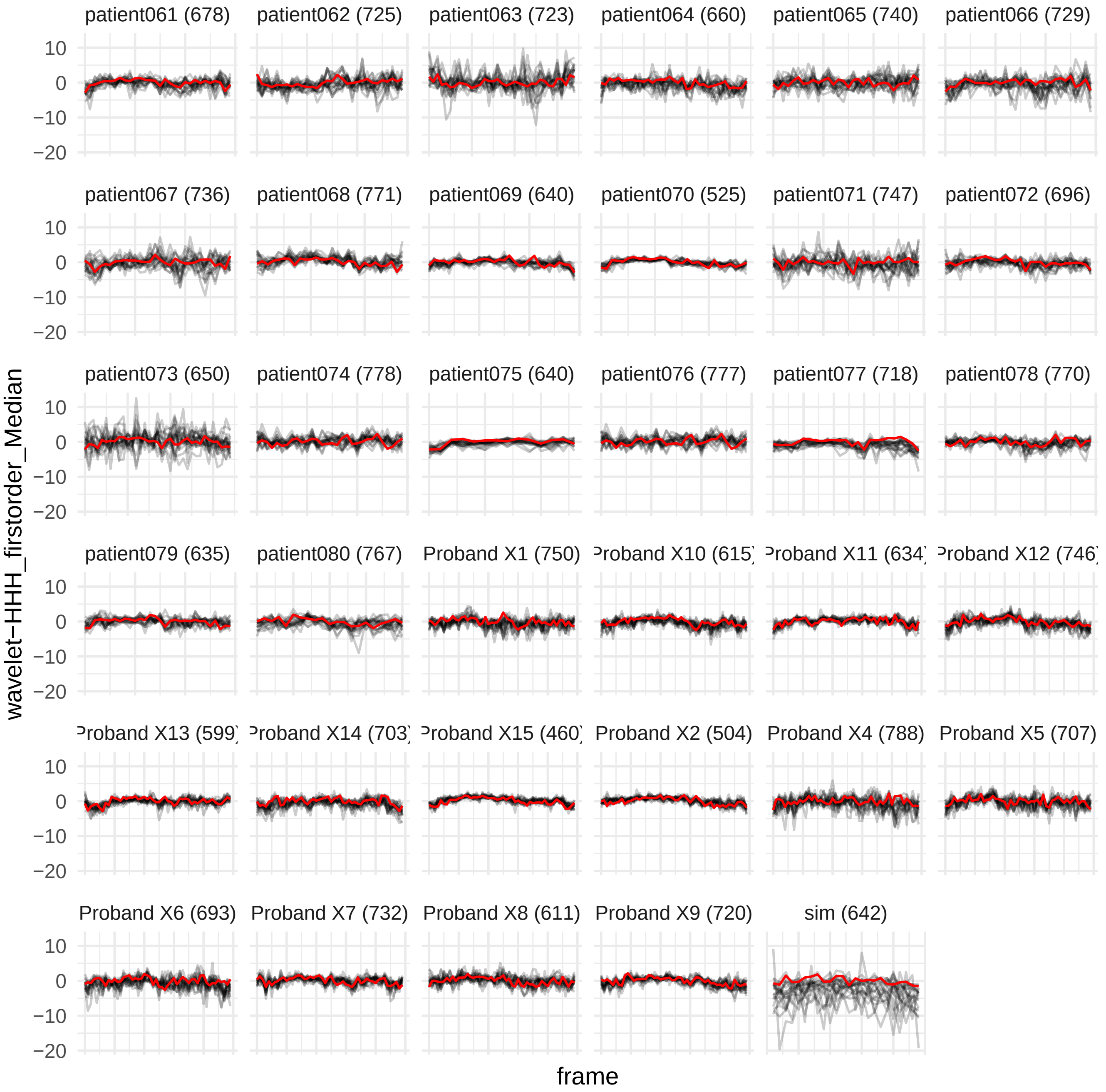

patient061 (678)
patient062 (725)
patient063 (723)
patient064 (660)
patient065 (740)
patient066 (729)
patient067 (736)
patient068 (771)
patient069 (640)
patient070 (525)
patient071 (747)
patient072 (696)
patient073 (650)
patient074 (778)
patient075 (640)
patient076 (777)
patient077 (718)
patient078 (770)
patient079 (635)
patient080 (767)
Proband X1 (750)
Proband X10 (615)
Proband X11 (634)
Proband X12 (746)
Proband X13 (599)
Proband X14 (703)
Proband X15 (460)
Proband X2 (504)
Proband X4 (788)
Proband X5 (707)
Proband X6 (693)
Proband X7 (732)
Proband X8 (611)
Proband X9 (720)
sim (642)
10
0
−10
−20
wavelet−HHH_firstorder_Median
frame

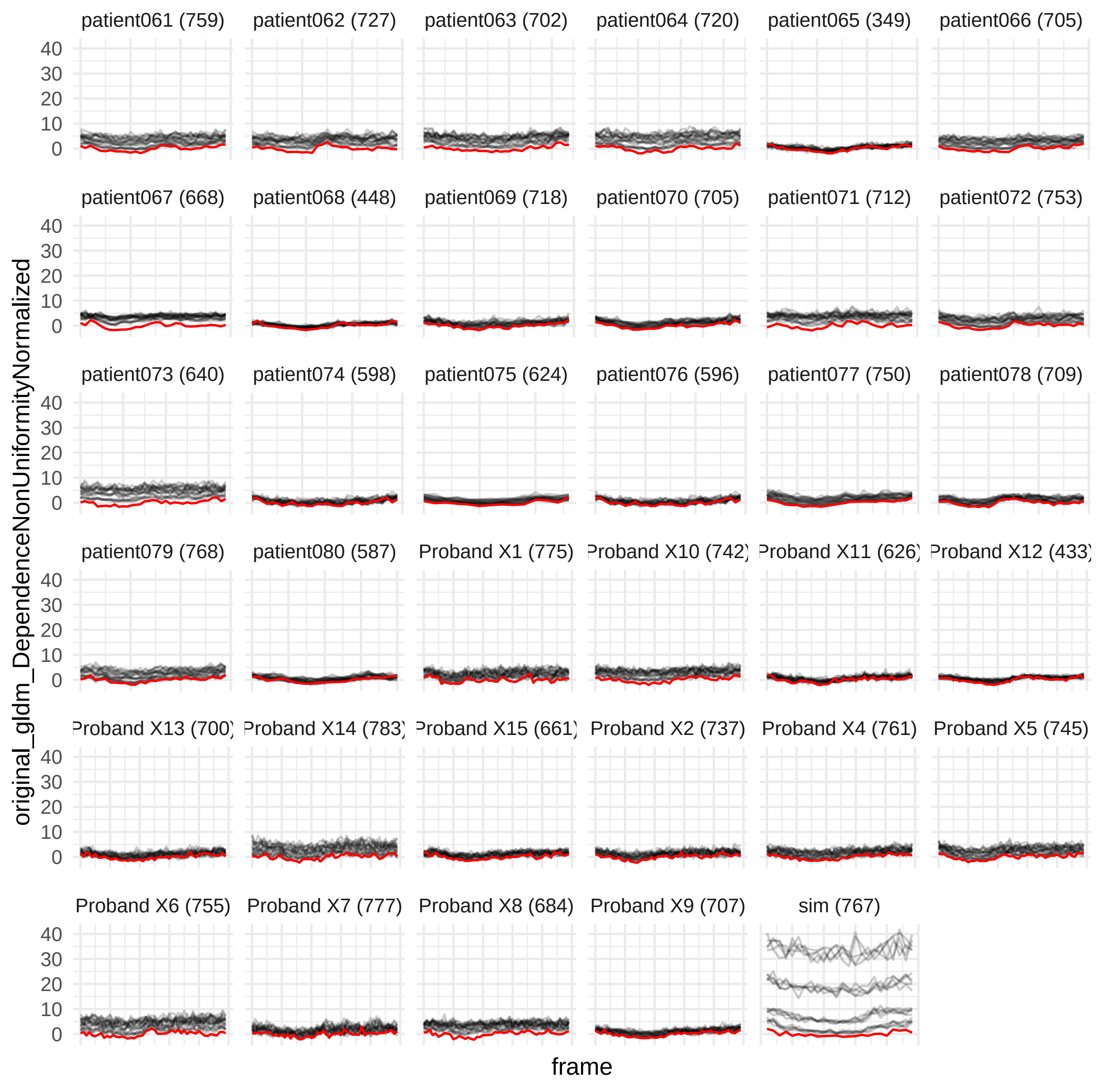

patient061 (759)
patient062 (727)
patient063 (702)
patient064 (720)
patient065 (349)
patient066 (705)
patient067 (668)
patient068 (448)
patient069 (718)
patient070 (705)
patient071 (712)
patient072 (753)
patient073 (640)
patient074 (598)
patient075 (624)
patient076 (596)
patient077 (750)
patient078 (709)
patient079 (768)
patient080 (587)
Proband X1 (775)
Proband X10 (742)
Proband X11 (626)
Proband X12 (433)
Proband X13 (700)
Proband X14 (783)
Proband X15 (661)
Proband X2 (737)
Proband X4 (761)
Proband X5 (745)
Proband X6 (755)
Proband X7 (777)
Proband X8 (684)
Proband X9 (707)
sim (767)
original_gldm_DependenceNonUniformityNormalized
frame
0
10
20
30
40

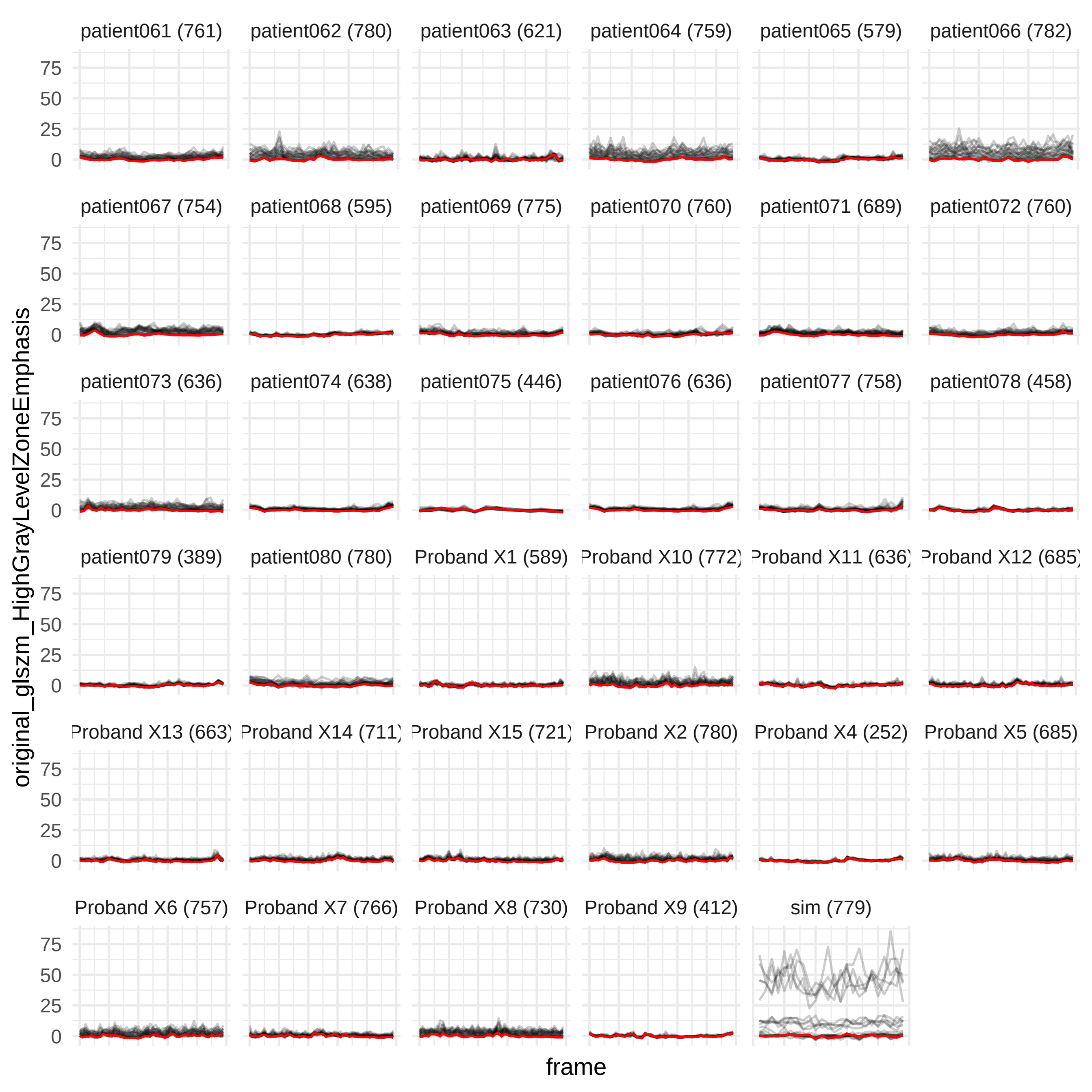

patient061 (761)
patient062 (780)
patient063 (621)
patient064 (759)
patient065 (579)
patient066 (782)
patient067 (754)
patient068 (595)
patient069 (775)
patient070 (760)
patient071 (689)
patient072 (760)
patient073 (636)
patient074 (638)
patient075 (446)
patient076 (636)
patient077 (758)
patient078 (458)
patient079 (389)
patient080 (780)
Proband X1 (589)
Proband X10 (772)
Proband X11 (636)
Proband X12 (685)
Proband X13 (663)
Proband X14 (711)
Proband X15 (721)
Proband X2 (780)
Proband X4 (252)
Proband X5 (685)
Proband X6 (757)
Proband X7 (766)
Proband X8 (730)
Proband X9 (412)
sim (779)
original_glszm_HighGrayLevelZoneEmphasis
0
25
50
75
frame

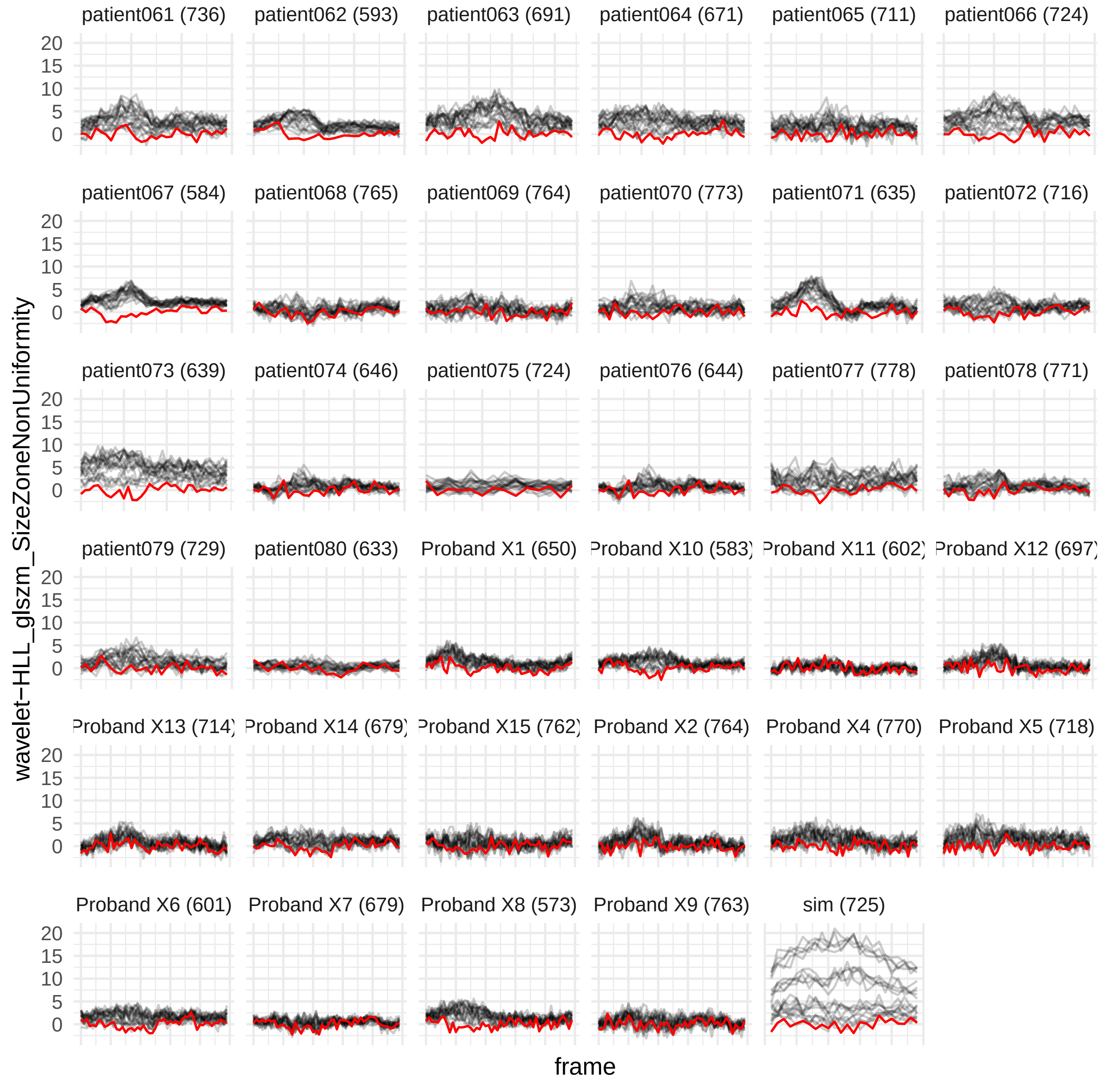

patient061 (736)
patient062 (593)
patient063 (691)
patient064 (671)
patient065 (711)
patient066 (724)
patient067 (584)
patient068 (765)
patient069 (764)
patient070 (773)
patient071 (635)
patient072 (716)
patient073 (639)
patient074 (646)
patient075 (724)
patient076 (644)
patient077 (778)
patient078 (771)
patient079 (729)
patient080 (633)
Proband X1 (650)
Proband X10 (583)
Proband X11 (602)
Proband X12 (697)
Proband X13 (714)
Proband X14 (679)
Proband X15 (762)
Proband X2 (764)
Proband X4 (770)
Proband X5 (718)
Proband X6 (601)
Proband X7 (679)
Proband X8 (573)
Proband X9 (763)
sim (725)
wavelet−HLL_glszm_SizeZoneNonUniformity
frame
0
5
10
15
20

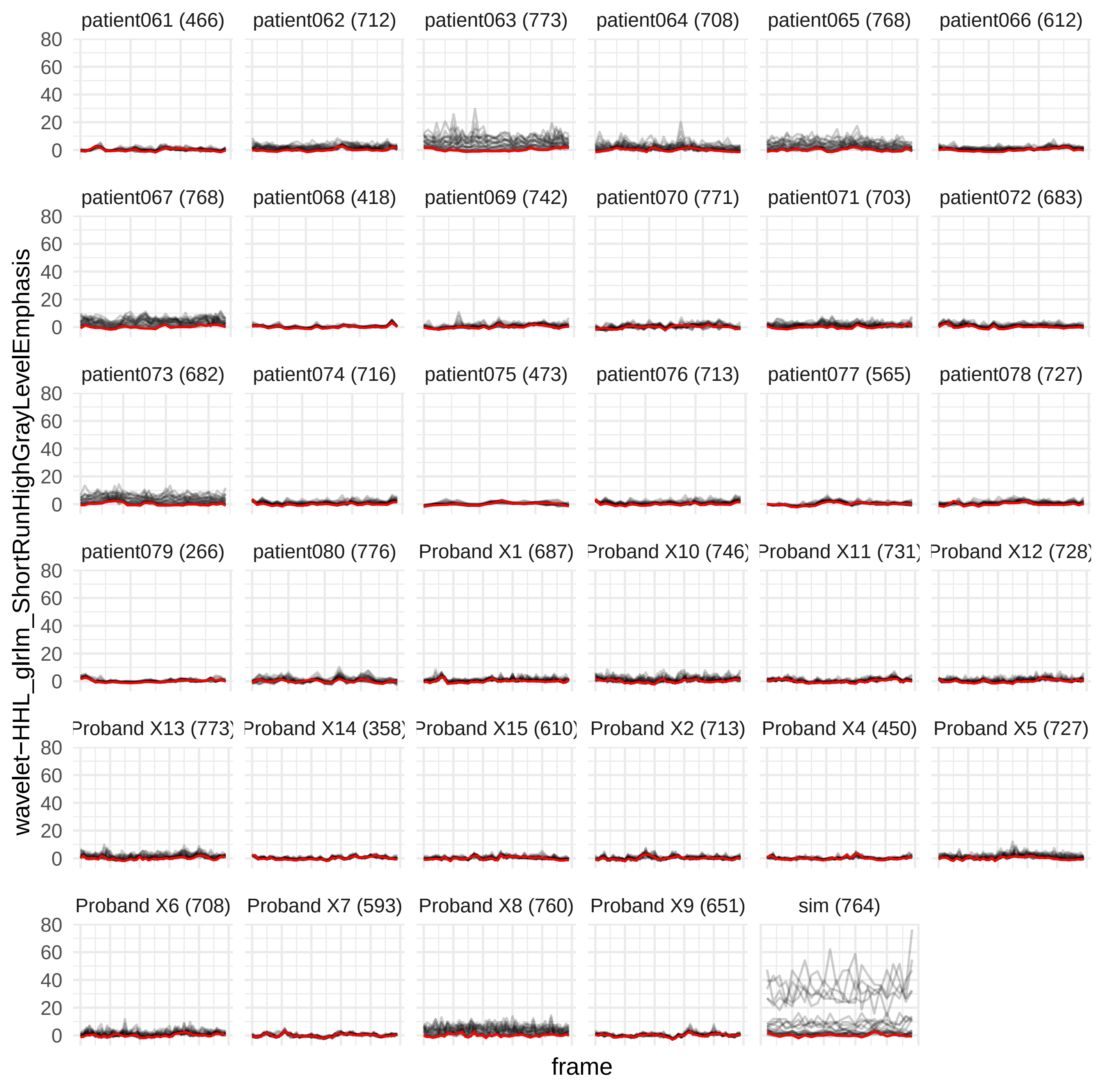
patient061 (466)
patient062 (712)
patient063 (773)
patient064 (708)
patient065 (768)
patient066 (612)
patient067 (768)
patient068 (418)
patient069 (742)
patient070 (771)
patient071 (703)
patient072 (683)
patient073 (682)
patient074 (716)
patient075 (473)
patient076 (713)
patient077 (565)
patient078 (727)
patient079 (266)
patient080 (776)
Proband X1 (687)
Proband X10 (746)
Proband X11 (731)
Proband X12 (728)
Proband X13 (773)
Proband X14 (358)
Proband X15 (610)
Proband X2 (713)
Proband X4 (450)
Proband X5 (727)
Proband X6 (708)
Proband X7 (593)
Proband X8 (760)
Proband X9 (651)
sim (764)
wavelet−HHL_glrlm_ShortRunHighGrayLevelEmphasis
frame
0
20
40
60
80

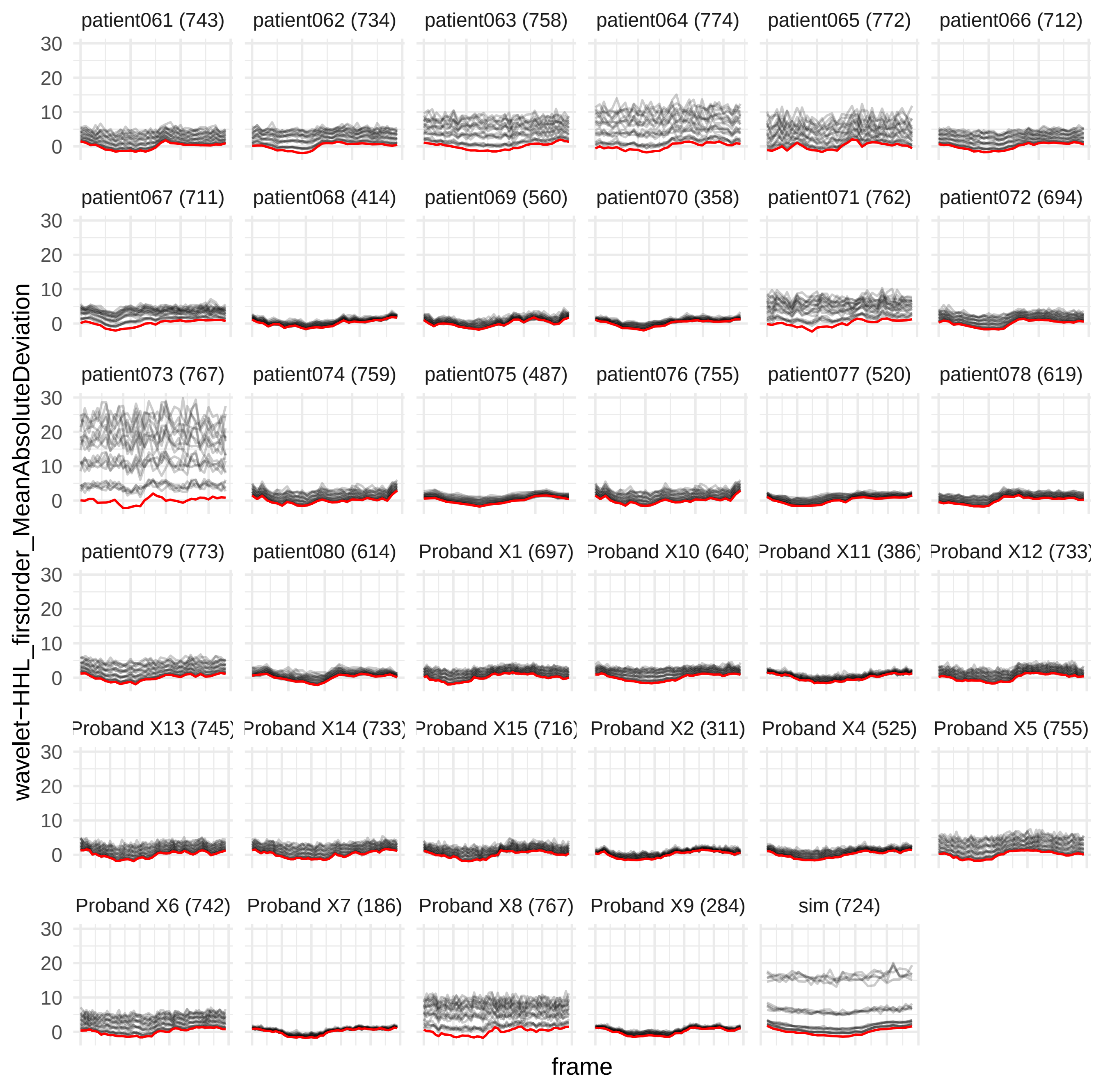

patient061 (743)
patient062 (734)
patient063 (758)
patient064 (774)
patient065 (772)
patient066 (712)
patient067 (711)
patient068 (414)
patient069 (560)
patient070 (358)
patient071 (762)
patient072 (694)
patient073 (767)
patient074 (759)
patient075 (487)
patient076 (755)
patient077 (520)
patient078 (619)
patient079 (773)
patient080 (614)
Proband X1 (697)
Proband X10 (640)
Proband X11 (386)
Proband X12 (733)
Proband X13 (745)
Proband X14 (733)
Proband X15 (716)
Proband X2 (311)
Proband X4 (525)
Proband X5 (755)
Proband X6 (742)
Proband X7 (186)
Proband X8 (767)
Proband X9 (284)
sim (724)
wavelet−HHL_firstorder_MeanAbsoluteDeviation
frame

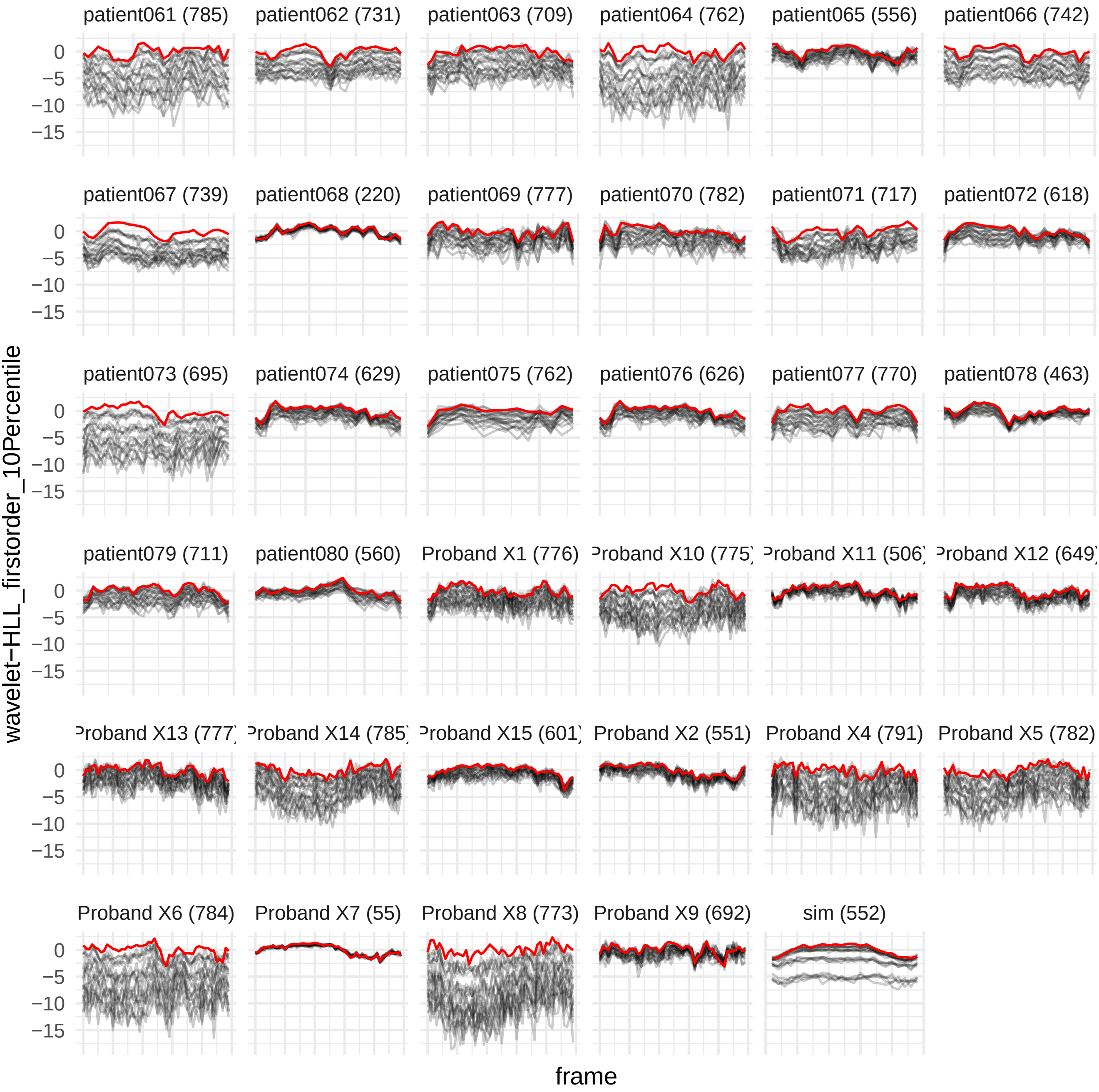

patient061 (785)
patient062 (731)
patient063 (709)
patient064 (762)
patient065 (556)
patient066 (742)
patient067 (739)
patient068 (220)
patient069 (777)
patient070 (782)
patient071 (717)
patient072 (618)
patient073 (695)
patient074 (629)
patient075 (762)
patient076 (626)
patient077 (770)
patient078 (463)
patient079 (711)
patient080 (560)
Proband X1 (776)
Proband X10 (775)
Proband X11 (506)
Proband X12 (649)
Proband X13 (777)
Proband X14 (785)
Proband X15 (601)
Proband X2 (551)
Proband X4 (791)
Proband X5 (782)
Proband X6 (784)
Proband X7 (55)
Proband X8 (773)
Proband X9 (692)
sim (552)
0
−5
−10
−15
wavelet−HLL_firstorder_10Percentile
frame

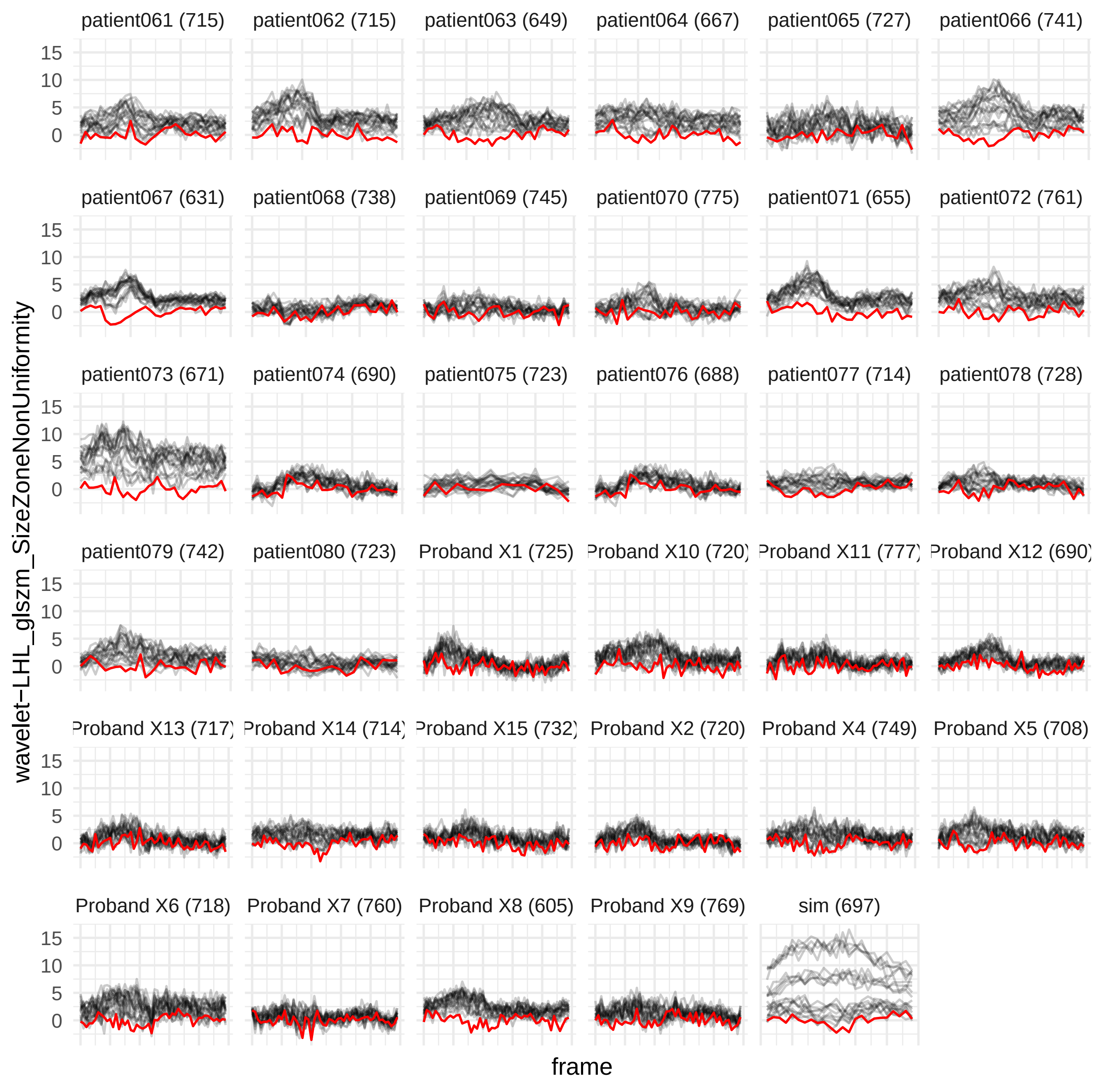
patient061 (715)
patient062 (715)
patient063 (649)
patient064 (667)
patient065 (727)
patient066 (741)
patient067 (631)
patient068 (738)
patient069 (745)
patient070 (775)
patient071 (655)
patient072 (761)
patient073 (671)
patient074 (690)
patient075 (723)
patient076 (688)
patient077 (714)
patient078 (728)
patient079 (742)
patient080 (723)
Proband X1 (725)
Proband X10 (720)
Proband X11 (777)
Proband X12 (690)
Proband X13 (717)
Proband X14 (714)
Proband X15 (732)
Proband X2 (720)
Proband X4 (749)
Proband X5 (708)
Proband X6 (718)
Proband X7 (760)
Proband X8 (605)
Proband X9 (769)
sim (697)
wavelet−LHL_glszm_SizeZoneNonUniformity
frame
15
10
5
0

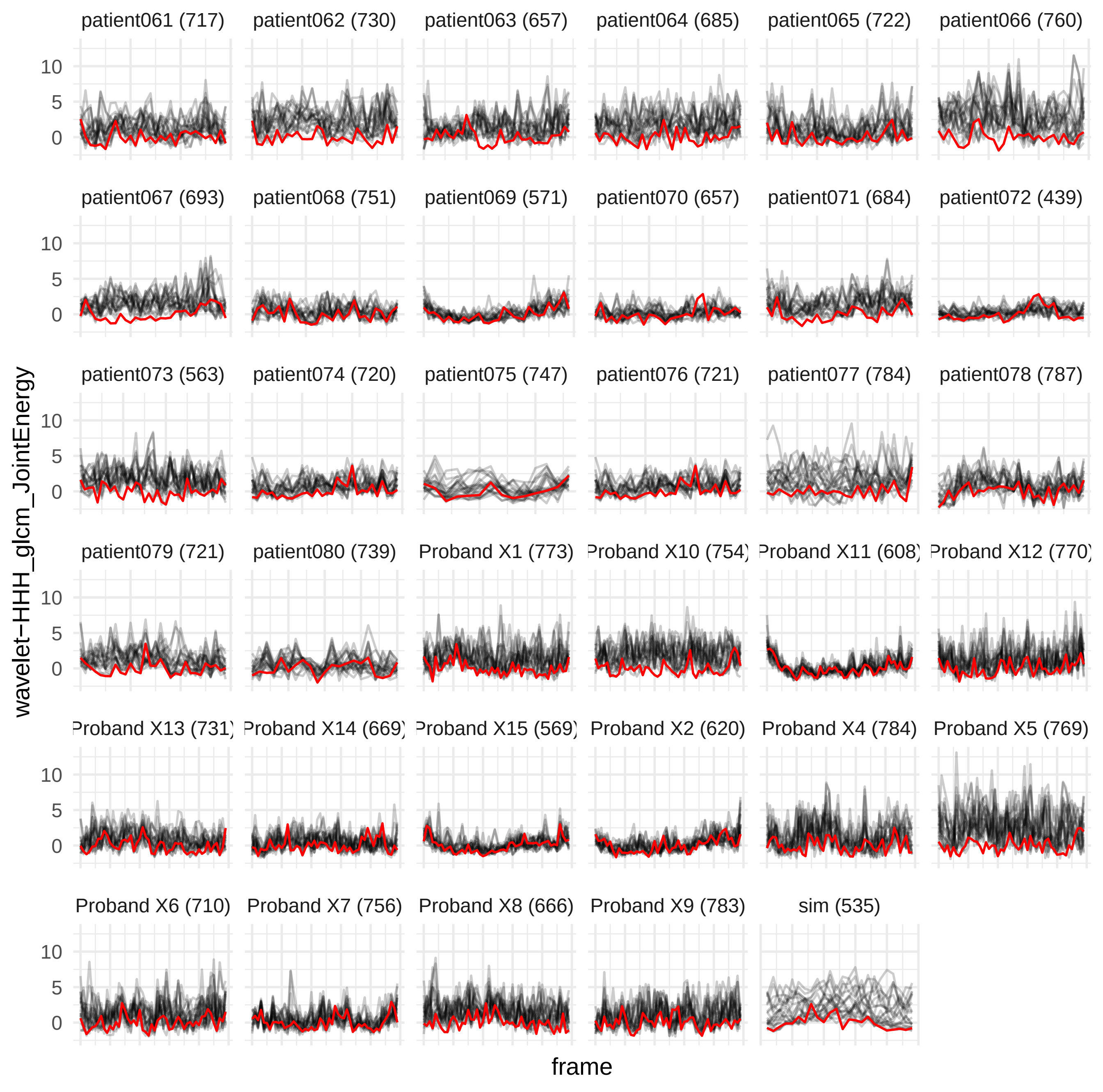

patient061 (717)
patient062 (730)
patient063 (657)
patient064 (685)
patient065 (722)
patient066 (760)
patient067 (693)
patient068 (751)
patient069 (571)
patient070 (657)
patient071 (684)
patient072 (439)
patient073 (563)
patient074 (720)
patient075 (747)
patient076 (721)
patient077 (784)
patient078 (787)
patient079 (721)
patient080 (739)
Proband X1 (773)
Proband X10 (754)
Proband X11 (608)
Proband X12 (770)
Proband X13 (731)
Proband X14 (669)
Proband X15 (569)
Proband X2 (620)
Proband X4 (784)
Proband X5 (769)
Proband X6 (710)
Proband X7 (756)
Proband X8 (666)
Proband X9 (783)
sim (535)
10
5
0
wavelet−HHH_glcm_JointEnergy
frame

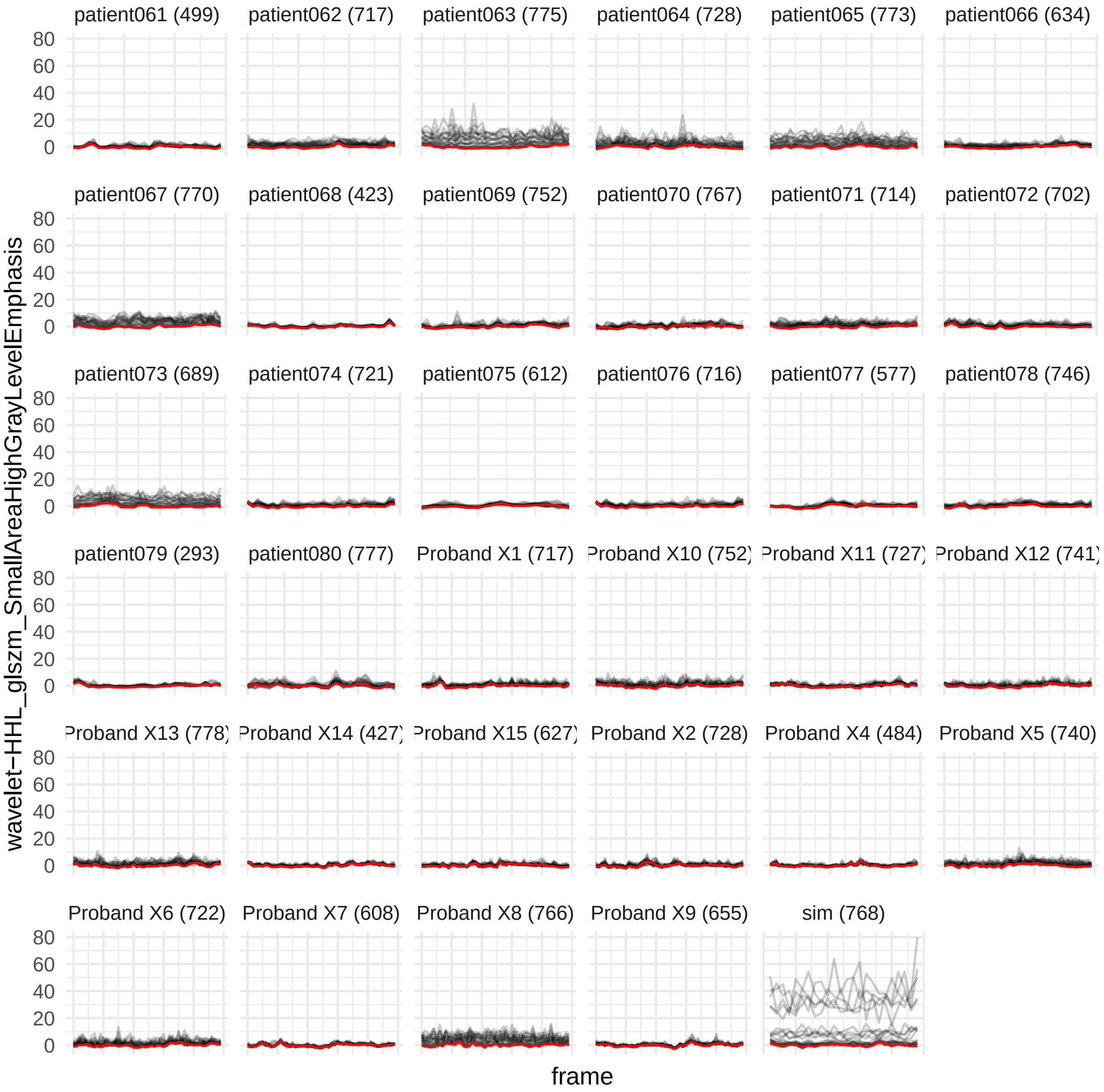

patient061 (499)
patient062 (717)
patient063 (775)
patient064 (728)
patient065 (773)
patient066 (634)
patient067 (770)
patient068 (423)
patient069 (752)
patient070 (767)
patient071 (714)
patient072 (702)
patient073 (689)
patient074 (721)
patient075 (612)
patient076 (716)
patient077 (577)
patient078 (746)
patient079 (293)
patient080 (777)
Proband X1 (717)
Proband X10 (752)
Proband X11 (727)
Proband X12 (741)
Proband X13 (778)
Proband X14 (427)
Proband X15 (627)
Proband X2 (728)
Proband X4 (484)
Proband X5 (740)
Proband X6 (722)
Proband X7 (608)
Proband X8 (766)
Proband X9 (655)
sim (768)
wavelet−HHL_glszm_SmallAreaHighGrayLevelEmphasis
frame
0
20
40
60
80

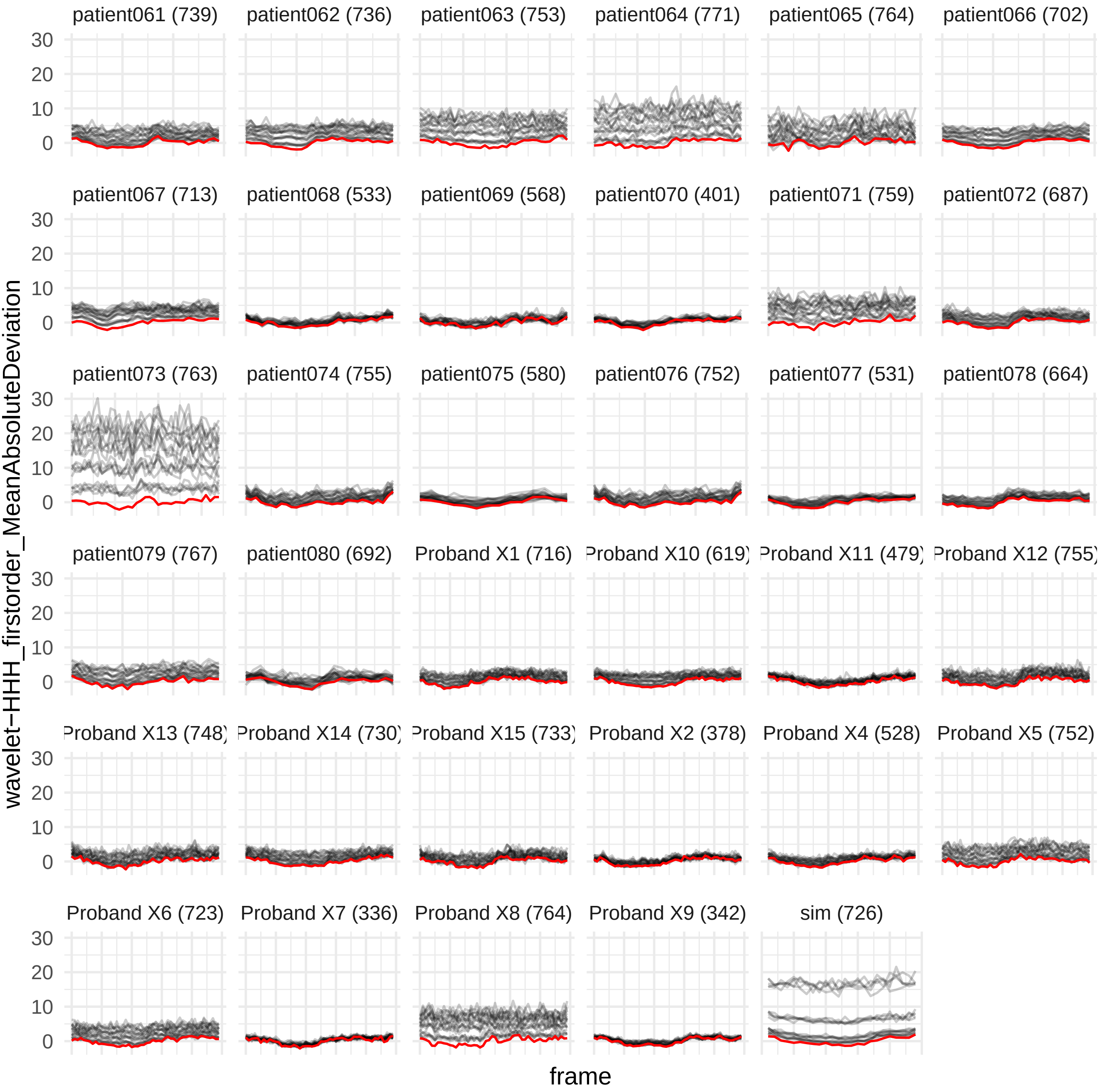

patient061 (739)
patient062 (736)
patient063 (753)
patient064 (771)
patient065 (764)
patient066 (702)
patient067 (713)
patient068 (533)
patient069 (568)
patient070 (401)
patient071 (759)
patient072 (687)
patient073 (763)
patient074 (755)
patient075 (580)
patient076 (752)
patient077 (531)
patient078 (664)
patient079 (767)
patient080 (692)
Proband X1 (716)
Proband X10 (619)
Proband X11 (479)
Proband X12 (755)
Proband X13 (748)
Proband X14 (730)
Proband X15 (733)
Proband X2 (378)
Proband X4 (528)
Proband X5 (752)
Proband X6 (723)
Proband X7 (336)
Proband X8 (764)
Proband X9 (342)
sim (726)
0
10
20
30
wavelet−HHH_firstorder_MeanAbsoluteDeviation
frame

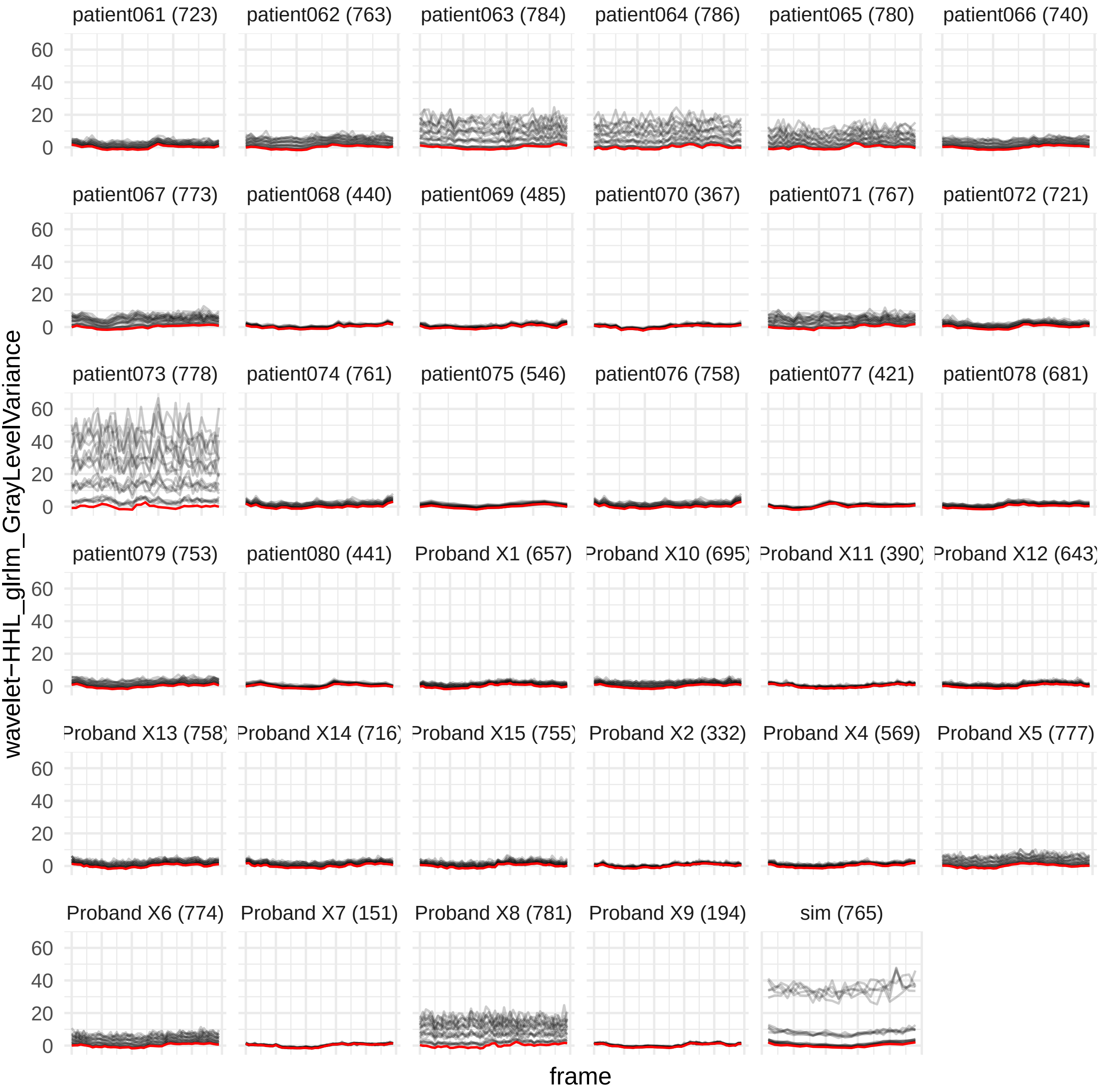
patient061 (723)
patient062 (763)
patient063 (784)
patient064 (786)
patient065 (780)
patient066 (740)
patient067 (773)
patient068 (440)
patient069 (485)
patient070 (367)
patient071 (767)
patient072 (721)
patient073 (778)
patient074 (761)
patient075 (546)
patient076 (758)
patient077 (421)
patient078 (681)
patient079 (753)
patient080 (441)
Proband X1 (657)
Proband X10 (695)
Proband X11 (390)
Proband X12 (643)
Proband X13 (758)
Proband X14 (716)
Proband X15 (755)
Proband X2 (332)
Proband X4 (569)
Proband X5 (777)
Proband X6 (774)
Proband X7 (151)
Proband X8 (781)
Proband X9 (194)
sim (765)
0
20
40
60
wavelet−HHL_glrlm_GrayLevelVariance
frame

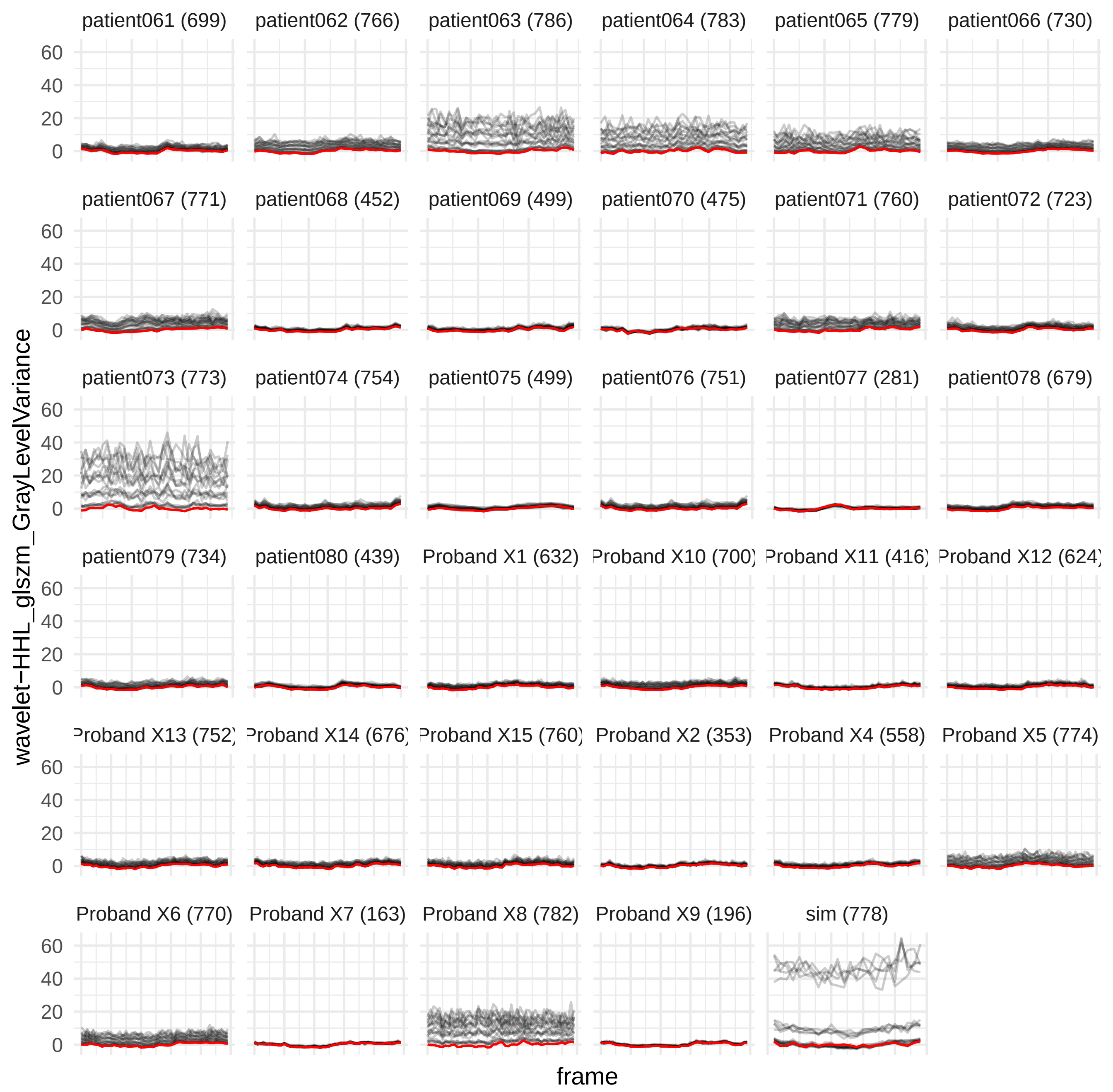

patient061 (699)
patient062 (766)
patient063 (786)
patient064 (783)
patient065 (779)
patient066 (730)
patient067 (771)
patient068 (452)
patient069 (499)
patient070 (475)
patient071 (760)
patient072 (723)
patient073 (773)
patient074 (754)
patient075 (499)
patient076 (751)
patient077 (281)
patient078 (679)
patient079 (734)
patient080 (439)
Proband X1 (632)
Proband X10 (700)
Proband X11 (416)
Proband X12 (624)
Proband X13 (752)
Proband X14 (676)
Proband X15 (760)
Proband X2 (353)
Proband X4 (558)
Proband X5 (774)
Proband X6 (770)
Proband X7 (163)
Proband X8 (782)
Proband X9 (196)
sim (778)
0
20
40
60
wavelet−HHL_glszm_GrayLevelVariance
frame

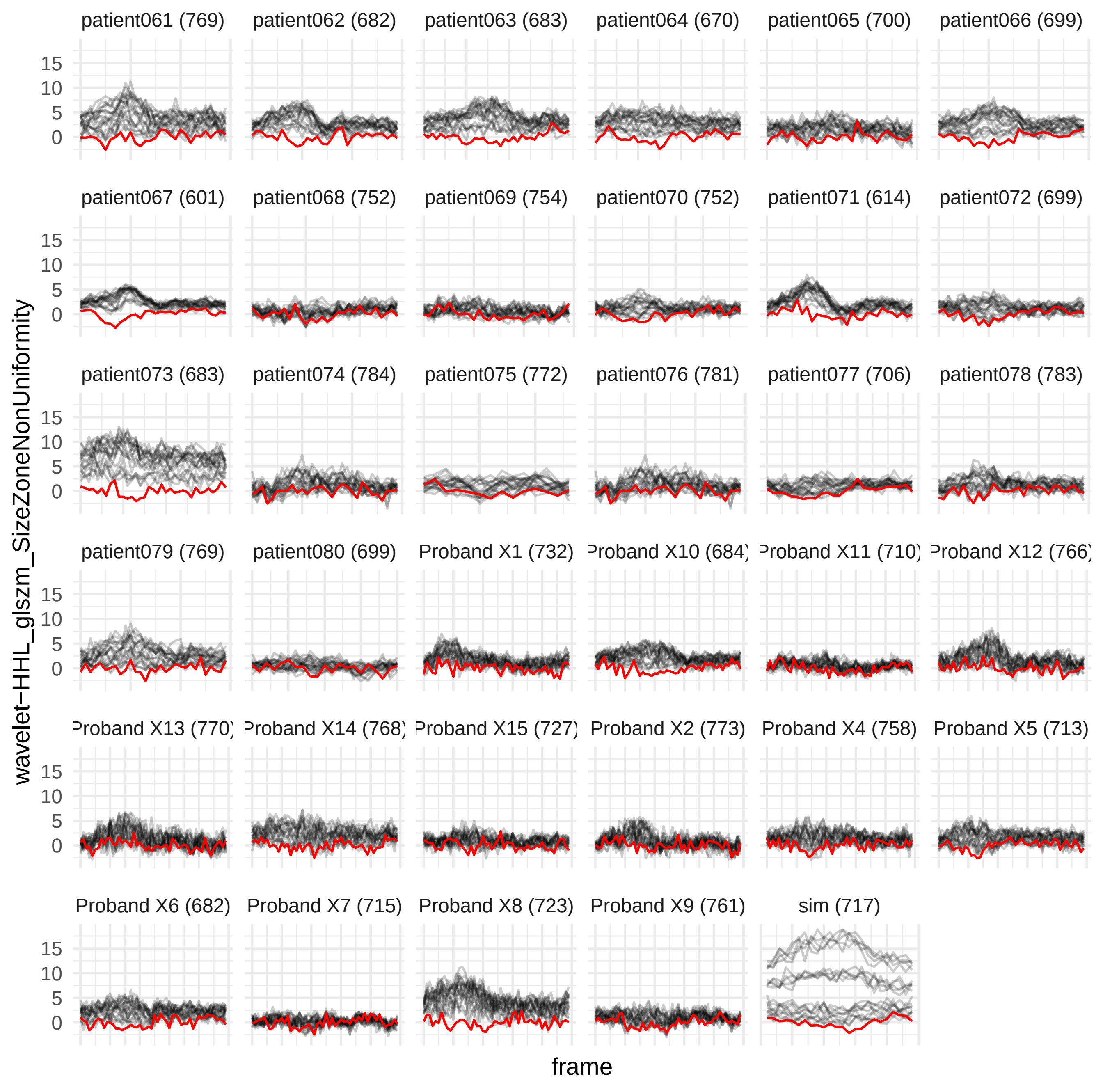
patient061 (769)
patient062 (682)
patient063 (683)
patient064 (670)
patient065 (700)
patient066 (699)
patient067 (601)
patient068 (752)
patient069 (754)
patient070 (752)
patient071 (614)
patient072 (699)
patient073 (683)
patient074 (784)
patient075 (772)
patient076 (781)
patient077 (706)
patient078 (783)
patient079 (769)
patient080 (699)
Proband X1 (732)
Proband X10 (684)
Proband X11 (710)
Proband X12 (766)
Proband X13 (770)
Proband X14 (768)
Proband X15 (727)
Proband X2 (773)
Proband X4 (758)
Proband X5 (713)
Proband X6 (682)
Proband X7 (715)
Proband X8 (723)
Proband X9 (761)
sim (717)
wavelet−HHL_glszm_SizeZoneNonUniformity
frame
0
5
10
15

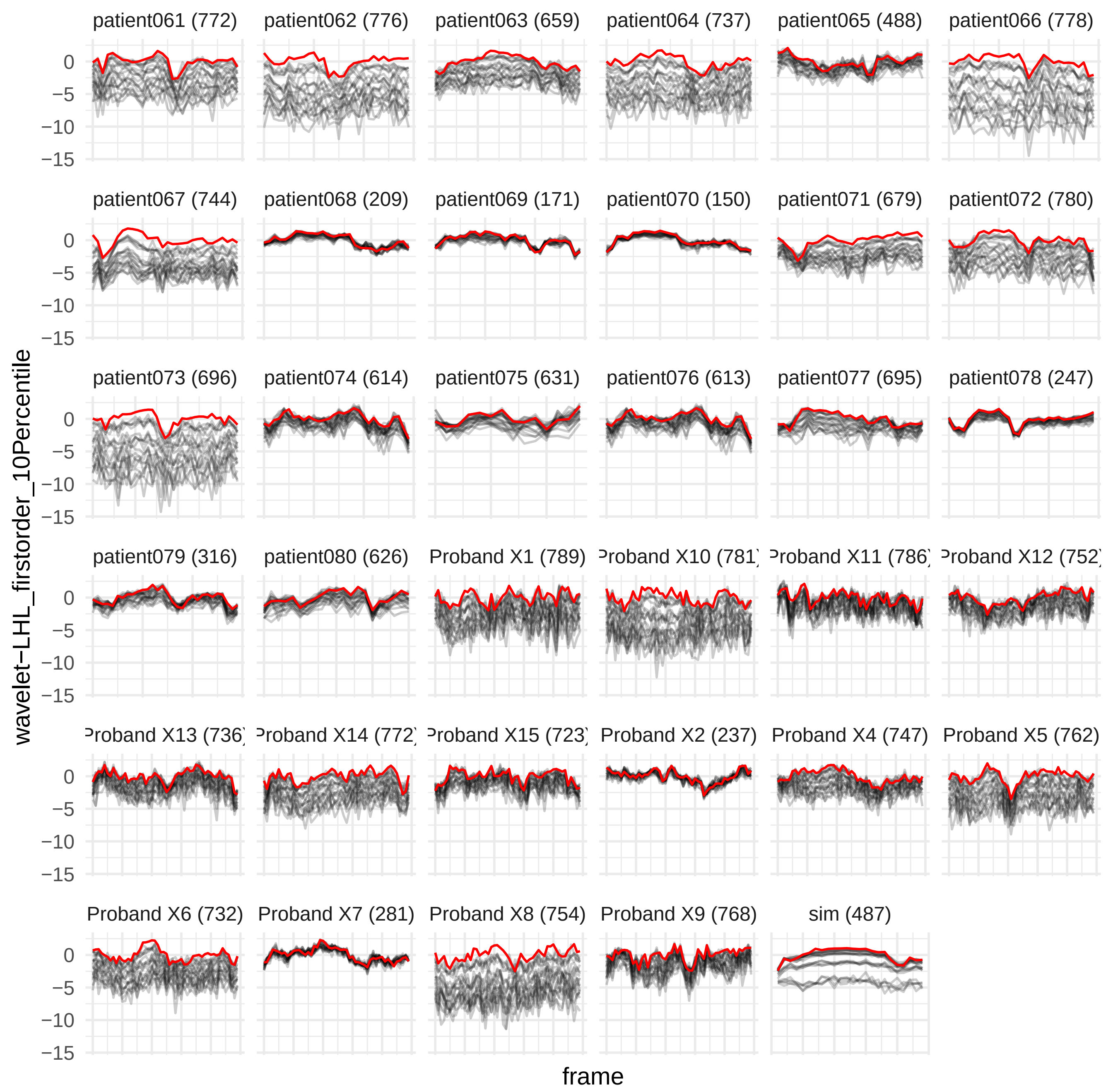

patient061 (772)
patient062 (776)
patient063 (659)
patient064 (737)
patient065 (488)
patient066 (778)
patient067 (744)
patient068 (209)
patient069 (171)
patient070 (150)
patient071 (679)
patient072 (780)
patient073 (696)
patient074 (614)
patient075 (631)
patient076 (613)
patient077 (695)
patient078 (247)
patient079 (316)
patient080 (626)
Proband X1 (789)
Proband X10 (781)
Proband X11 (786)
Proband X12 (752)
Proband X13 (736)
Proband X14 (772)
Proband X15 (723)
Proband X2 (237)
Proband X4 (747)
Proband X5 (762)
Proband X6 (732)
Proband X7 (281)
Proband X8 (754)
Proband X9 (768)
sim (487)
0
−5
−10
−15
wavelet−LHL_firstorder_10Percentile
frame

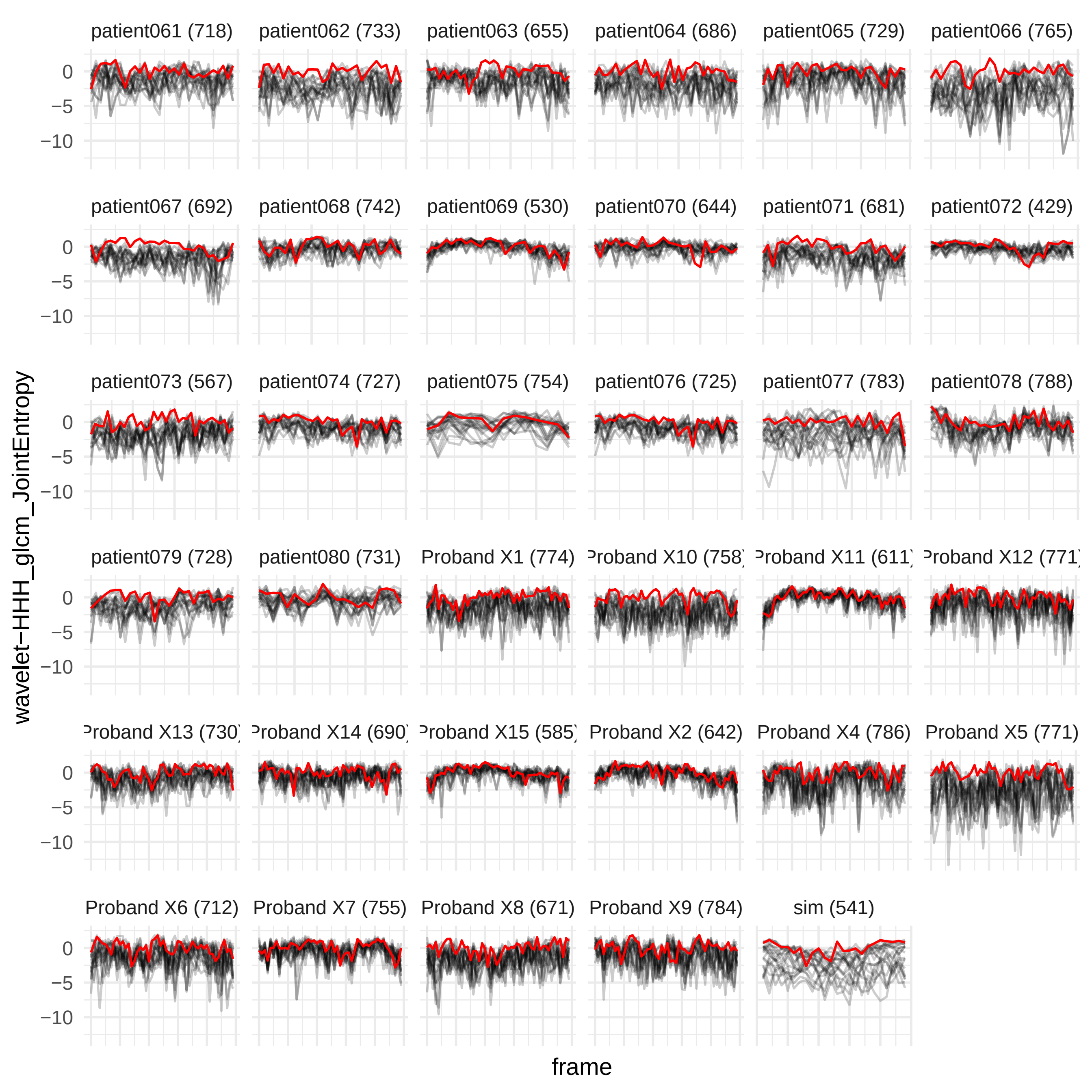
patient061 (718)
patient062 (733)
patient063 (655)
patient064 (686)
patient065 (729)
patient066 (765)
patient067 (692)
patient068 (742)
patient069 (530)
patient070 (644)
patient071 (681)
patient072 (429)
patient073 (567)
patient074 (727)
patient075 (754)
patient076 (725)
patient077 (783)
patient078 (788)
patient079 (728)
patient080 (731)
Proband X1 (774)
Proband X10 (758)
Proband X11 (611)
Proband X12 (771)
Proband X13 (730)
Proband X14 (690)
Proband X15 (585)
Proband X2 (642)
Proband X4 (786)
Proband X5 (771)
Proband X6 (712)
Proband X7 (755)
Proband X8 (671)
Proband X9 (784)
sim (541)
0
−5
−10
wavelet−HHH_glcm_JointEntropy
frame

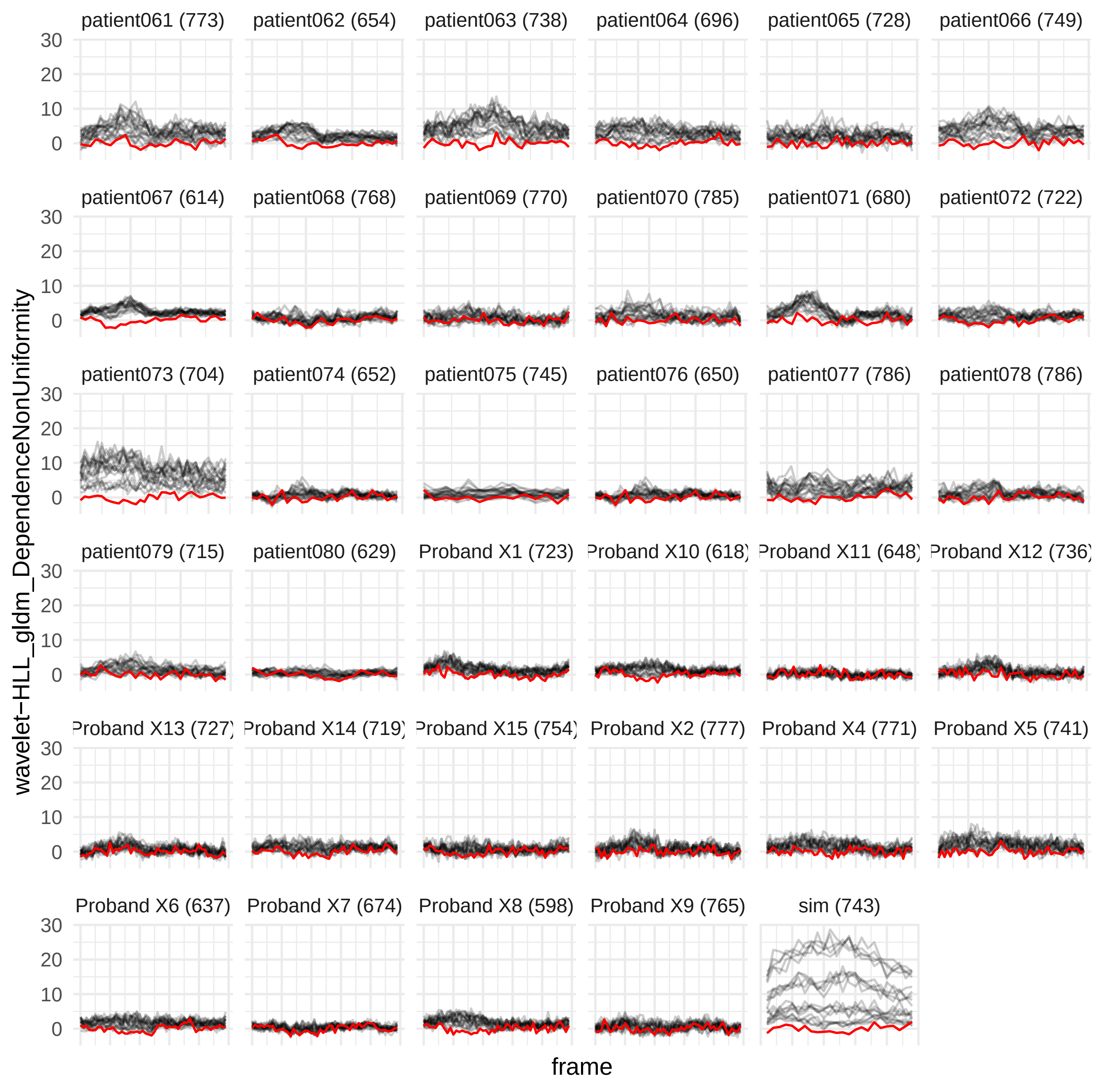

patient061 (773)
patient062 (654)
patient063 (738)
patient064 (696)
patient065 (728)
patient066 (749)
patient067 (614)
patient068 (768)
patient069 (770)
patient070 (785)
patient071 (680)
patient072 (722)
patient073 (704)
patient074 (652)
patient075 (745)
patient076 (650)
patient077 (786)
patient078 (786)
patient079 (715)
patient080 (629)
Proband X1 (723)
Proband X10 (618)
Proband X11 (648)
Proband X12 (736)
Proband X13 (727)
Proband X14 (719)
Proband X15 (754)
Proband X2 (777)
Proband X4 (771)
Proband X5 (741)
Proband X6 (637)
Proband X7 (674)
Proband X8 (598)
Proband X9 (765)
sim (743)
30
20
10
0
wavelet−HLL_gldm_DependenceNonUniformity
frame

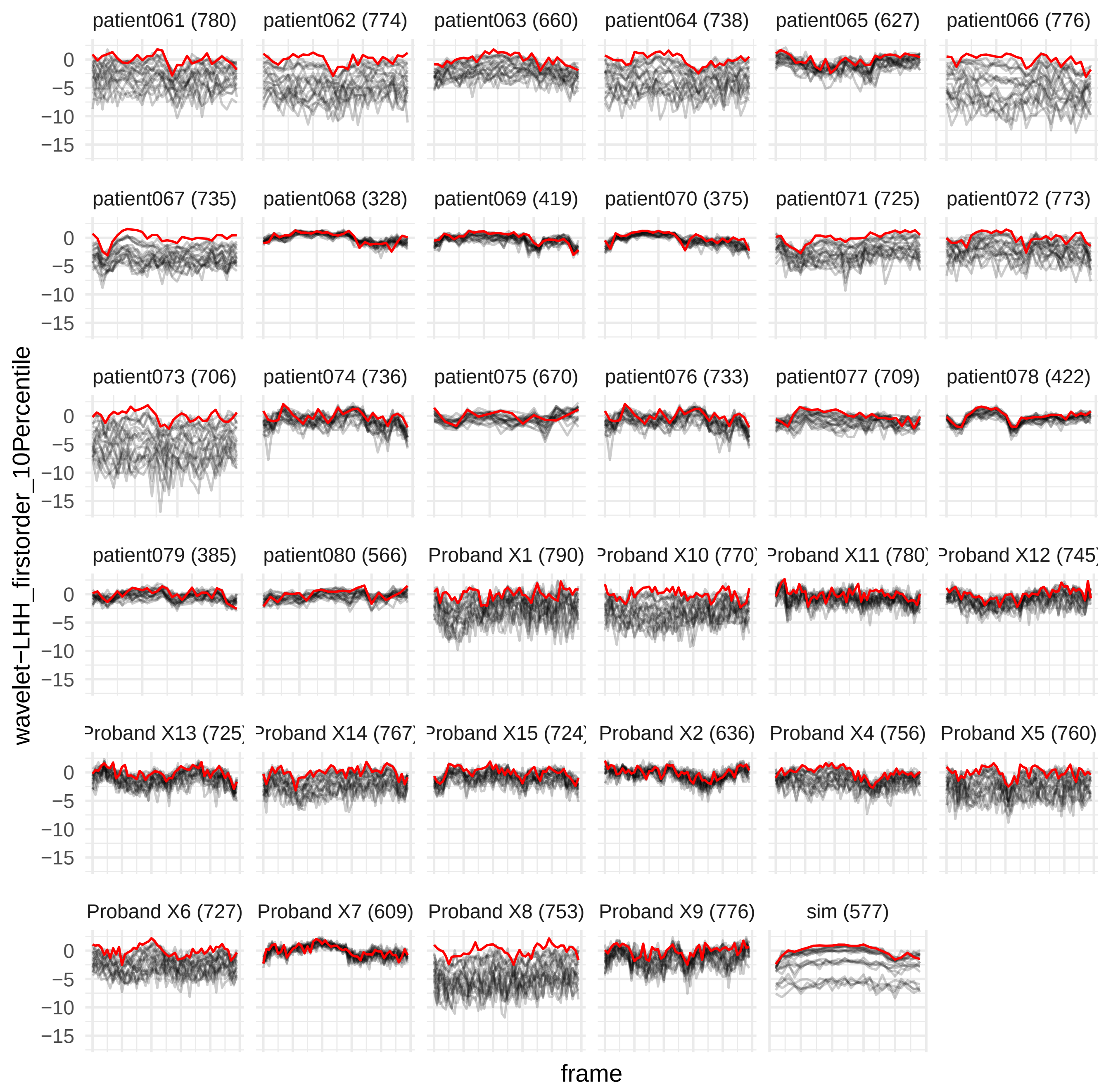

patient061 (780)
patient062 (774)
patient063 (660)
patient064 (738)
patient065 (627)
patient066 (776)
patient067 (735)
patient068 (328)
patient069 (419)
patient070 (375)
patient071 (725)
patient072 (773)
patient073 (706)
patient074 (736)
patient075 (670)
patient076 (733)
patient077 (709)
patient078 (422)
patient079 (385)
patient080 (566)
Proband X1 (790)
Proband X10 (770)
Proband X11 (780)
Proband X12 (745)
Proband X13 (725)
Proband X14 (767)
Proband X15 (724)
Proband X2 (636)
Proband X4 (756)
Proband X5 (760)
Proband X6 (727)
Proband X7 (609)
Proband X8 (753)
Proband X9 (776)
sim (577)
0
−5
−10
−15
wavelet−LHH_firstorder_10Percentile
frame

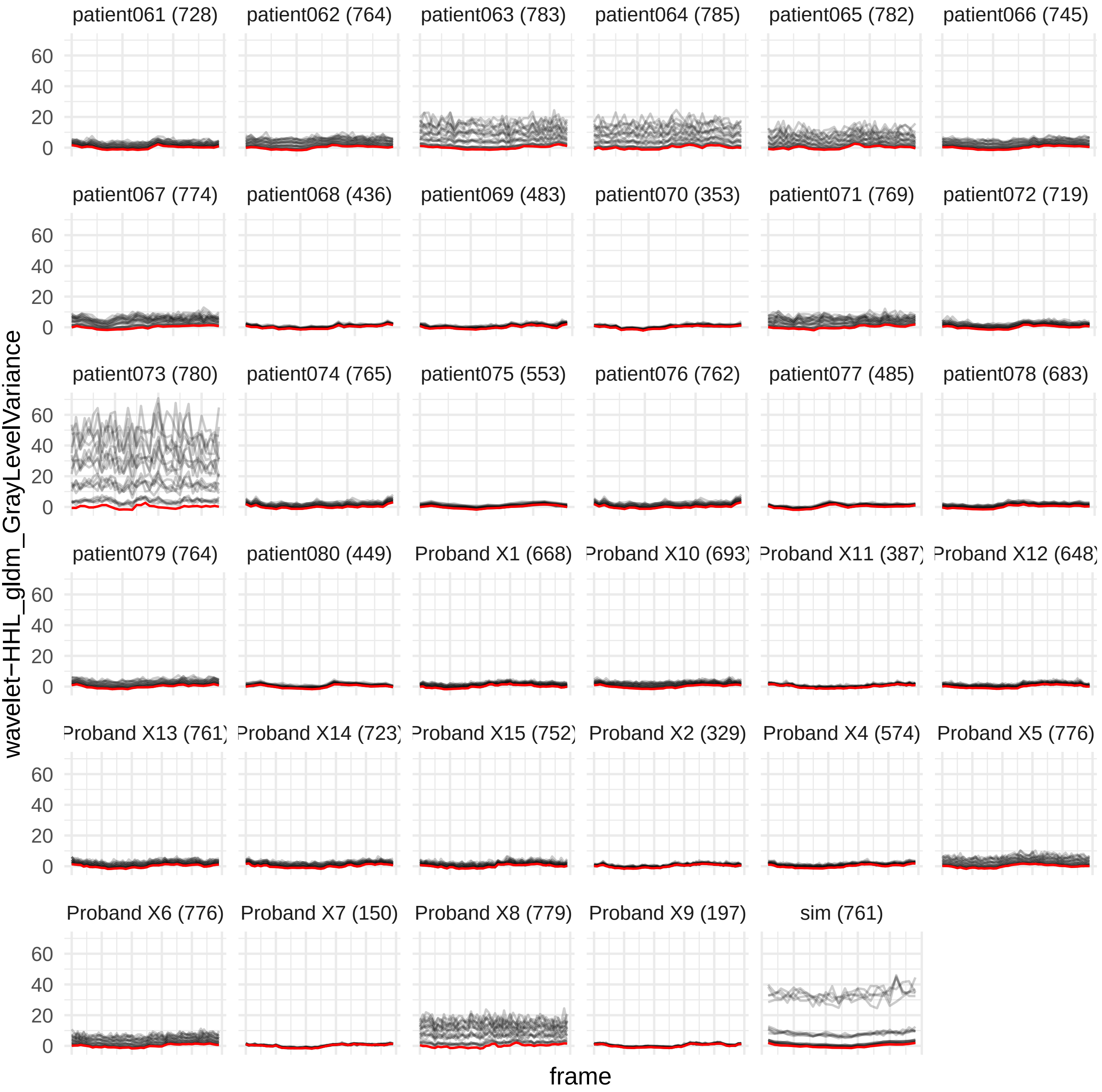

patient061 (728)
patient062 (764)
patient063 (783)
patient064 (785)
patient065 (782)
patient066 (745)
patient067 (774)
patient068 (436)
patient069 (483)
patient070 (353)
patient071 (769)
patient072 (719)
patient073 (780)
patient074 (765)
patient075 (553)
patient076 (762)
patient077 (485)
patient078 (683)
patient079 (764)
patient080 (449)
Proband X1 (668)
Proband X10 (693)
Proband X11 (387)
Proband X12 (648)
Proband X13 (761)
Proband X14 (723)
Proband X15 (752)
Proband X2 (329)
Proband X4 (574)
Proband X5 (776)
Proband X6 (776)
Proband X7 (150)
Proband X8 (779)
Proband X9 (197)
sim (761)
0
20
40
60
wavelet−HHL_gldm_GrayLevelVariance
frame

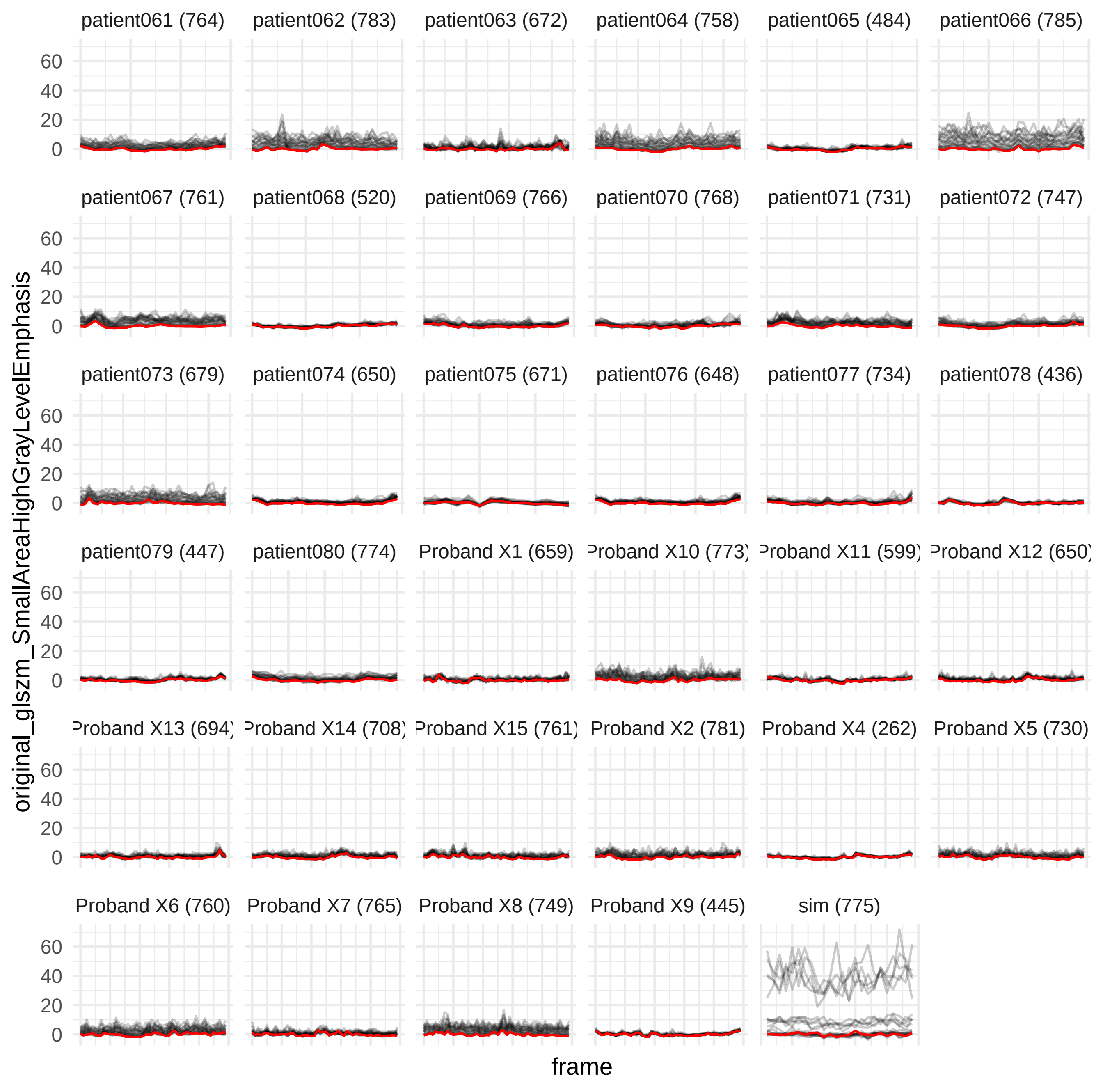
patient061 (764)
patient062 (783)
patient063 (672)
patient064 (758)
patient065 (484)
patient066 (785)
patient067 (761)
patient068 (520)
patient069 (766)
patient070 (768)
patient071 (731)
patient072 (747)
patient073 (679)
patient074 (650)
patient075 (671)
patient076 (648)
patient077 (734)
patient078 (436)
patient079 (447)
patient080 (774)
Proband X1 (659)
Proband X10 (773)
Proband X11 (599)
Proband X12 (650)
Proband X13 (694)
Proband X14 (708)
Proband X15 (761)
Proband X2 (781)
Proband X4 (262)
Proband X5 (730)
Proband X6 (760)
Proband X7 (765)
Proband X8 (749)
Proband X9 (445)
sim (775)
60
40
20
0
original_glszm_SmallAreaHighGrayLevelEmphasis
frame

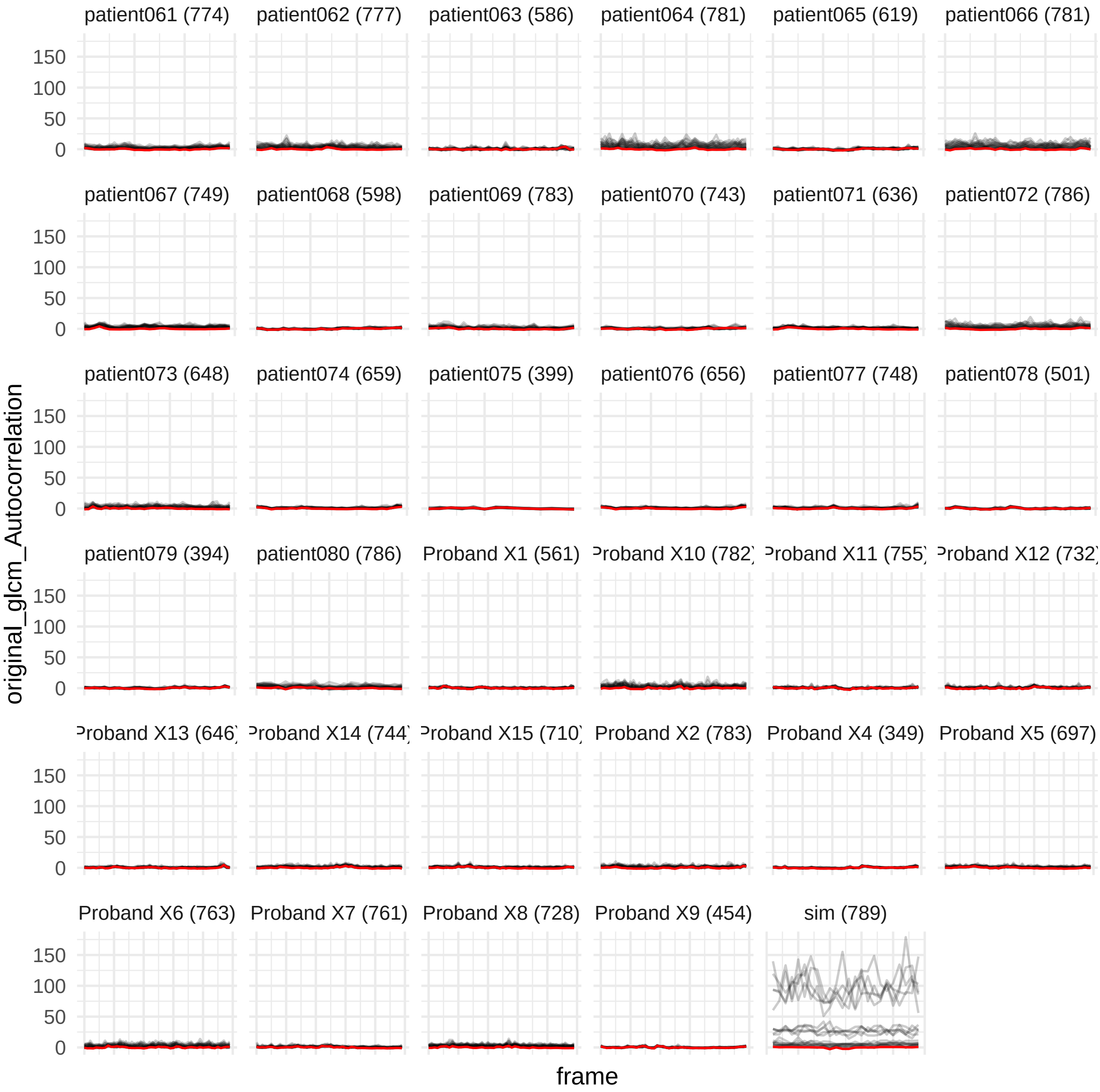
patient061 (774)
patient062 (777)
patient063 (586)
patient064 (781)
patient065 (619)
patient066 (781)
patient067 (749)
patient068 (598)
patient069 (783)
patient070 (743)
patient071 (636)
patient072 (786)
patient073 (648)
patient074 (659)
patient075 (399)
patient076 (656)
patient077 (748)
patient078 (501)
patient079 (394)
patient080 (786)
Proband X1 (561)
Proband X10 (782)
Proband X11 (755)
Proband X12 (732)
Proband X13 (646)
Proband X14 (744)
Proband X15 (710)
Proband X2 (783)
Proband X4 (349)
Proband X5 (697)
Proband X6 (763)
Proband X7 (761)
Proband X8 (728)
Proband X9 (454)
sim (789)
0
50
100
150
original_glcm_Autocorrelation
frame

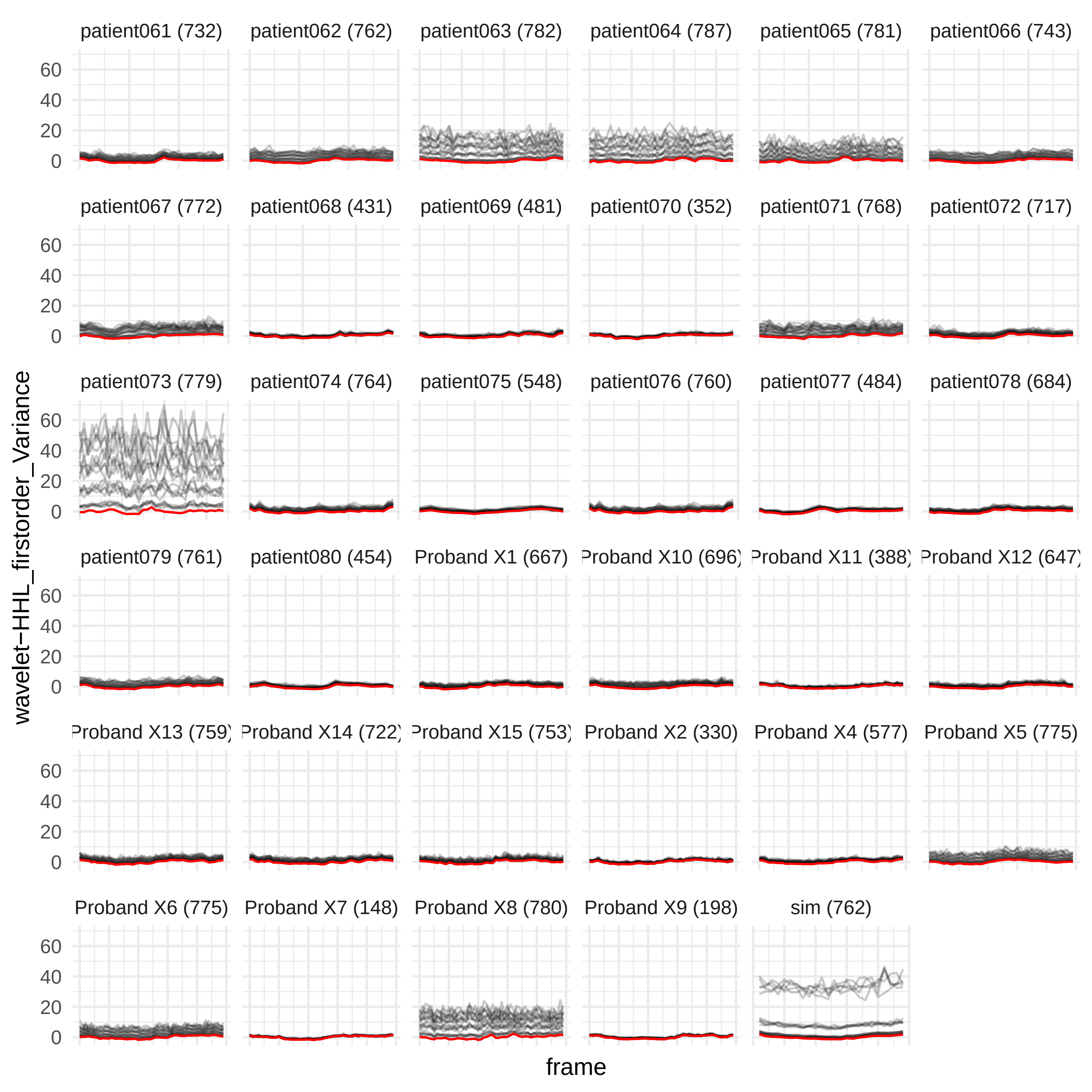

patient061 (732)
patient062 (762)
patient063 (782)
patient064 (787)
patient065 (781)
patient066 (743)
patient067 (772)
patient068 (431)
patient069 (481)
patient070 (352)
patient071 (768)
patient072 (717)
patient073 (779)
patient074 (764)
patient075 (548)
patient076 (760)
patient077 (484)
patient078 (684)
patient079 (761)
patient080 (454)
Proband X1 (667)
Proband X10 (696)
Proband X11 (388)
Proband X12 (647)
Proband X13 (759)
Proband X14 (722)
Proband X15 (753)
Proband X2 (330)
Proband X4 (577)
Proband X5 (775)
Proband X6 (775)
Proband X7 (148)
Proband X8 (780)
Proband X9 (198)
sim (762)
0
20
40
60
wavelet−HHL_firstorder_Variance
frame

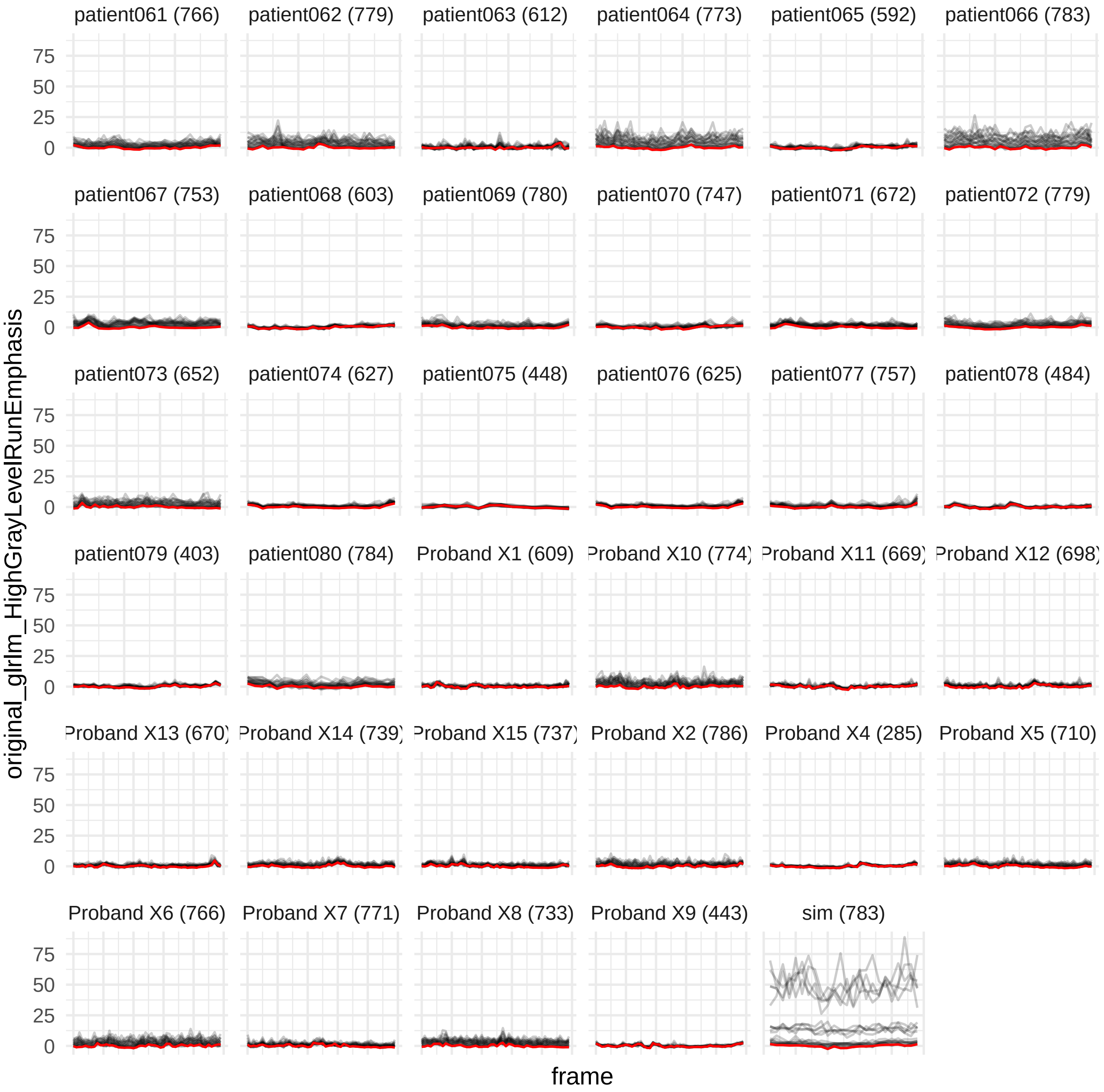

patient061 (766)
patient062 (779)
patient063 (612)
patient064 (773)
patient065 (592)
patient066 (783)
patient067 (753)
patient068 (603)
patient069 (780)
patient070 (747)
patient071 (672)
patient072 (779)
patient073 (652)
patient074 (627)
patient075 (448)
patient076 (625)
patient077 (757)
patient078 (484)
patient079 (403)
patient080 (784)
Proband X1 (609)
Proband X10 (774)
Proband X11 (669)
Proband X12 (698)
Proband X13 (670)
Proband X14 (739)
Proband X15 (737)
Proband X2 (786)
Proband X4 (285)
Proband X5 (710)
Proband X6 (766)
Proband X7 (771)
Proband X8 (733)
Proband X9 (443)
sim (783)
original_glrlm_HighGrayLevelRunEmphasis
0
25
50
75
frame

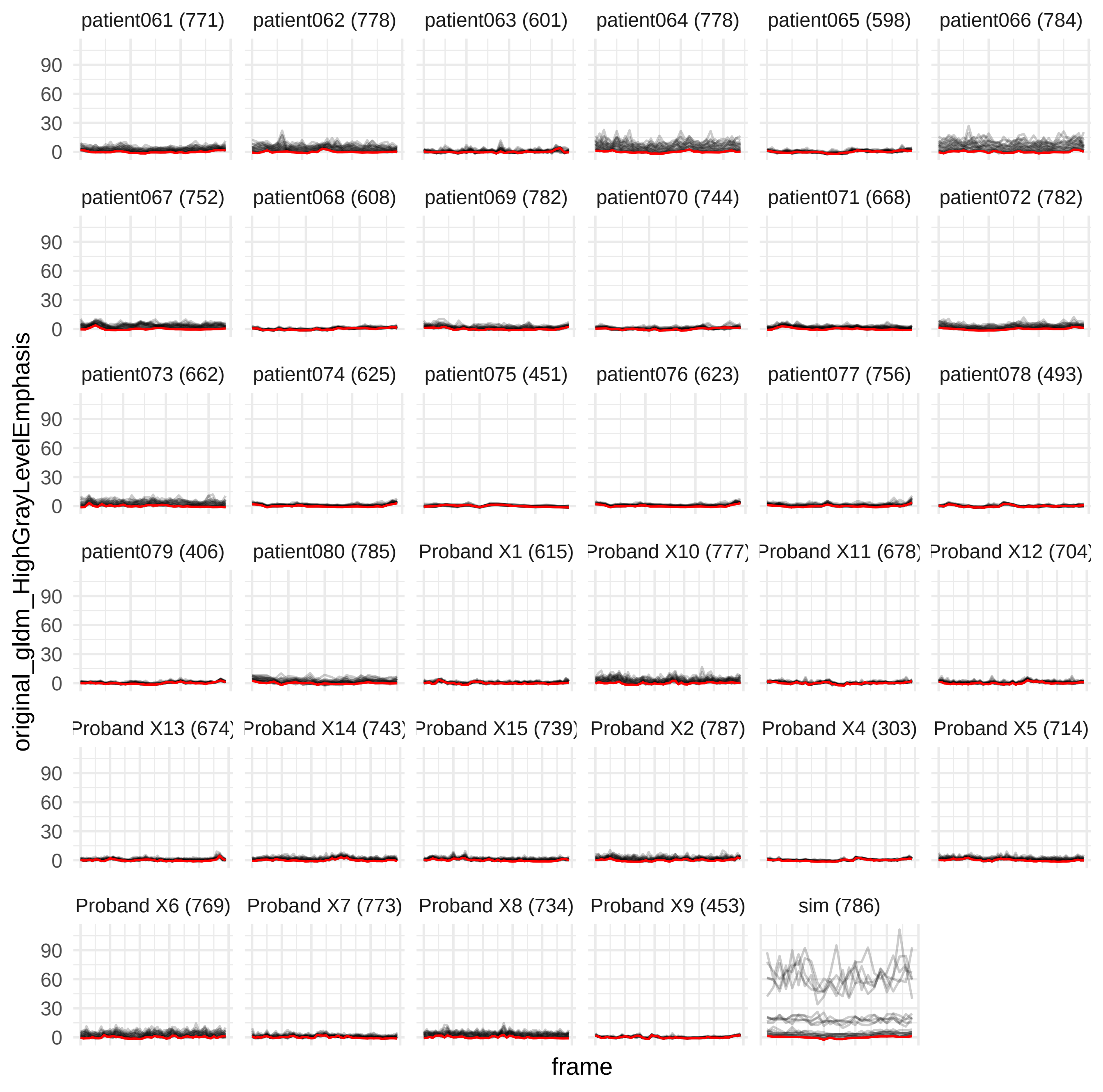
patient061 (771)
patient062 (778)
patient063 (601)
patient064 (778)
patient065 (598)
patient066 (784)
patient067 (752)
patient068 (608)
patient069 (782)
patient070 (744)
patient071 (668)
patient072 (782)
patient073 (662)
patient074 (625)
patient075 (451)
patient076 (623)
patient077 (756)
patient078 (493)
patient079 (406)
patient080 (785)
Proband X1 (615)
Proband X10 (777)
Proband X11 (678)
Proband X12 (704)
Proband X13 (674)
Proband X14 (743)
Proband X15 (739)
Proband X2 (787)
Proband X4 (303)
Proband X5 (714)
Proband X6 (769)
Proband X7 (773)
Proband X8 (734)
Proband X9 (453)
sim (786)
90
60
30
0
original_gldm_HighGrayLevelEmphasis
frame

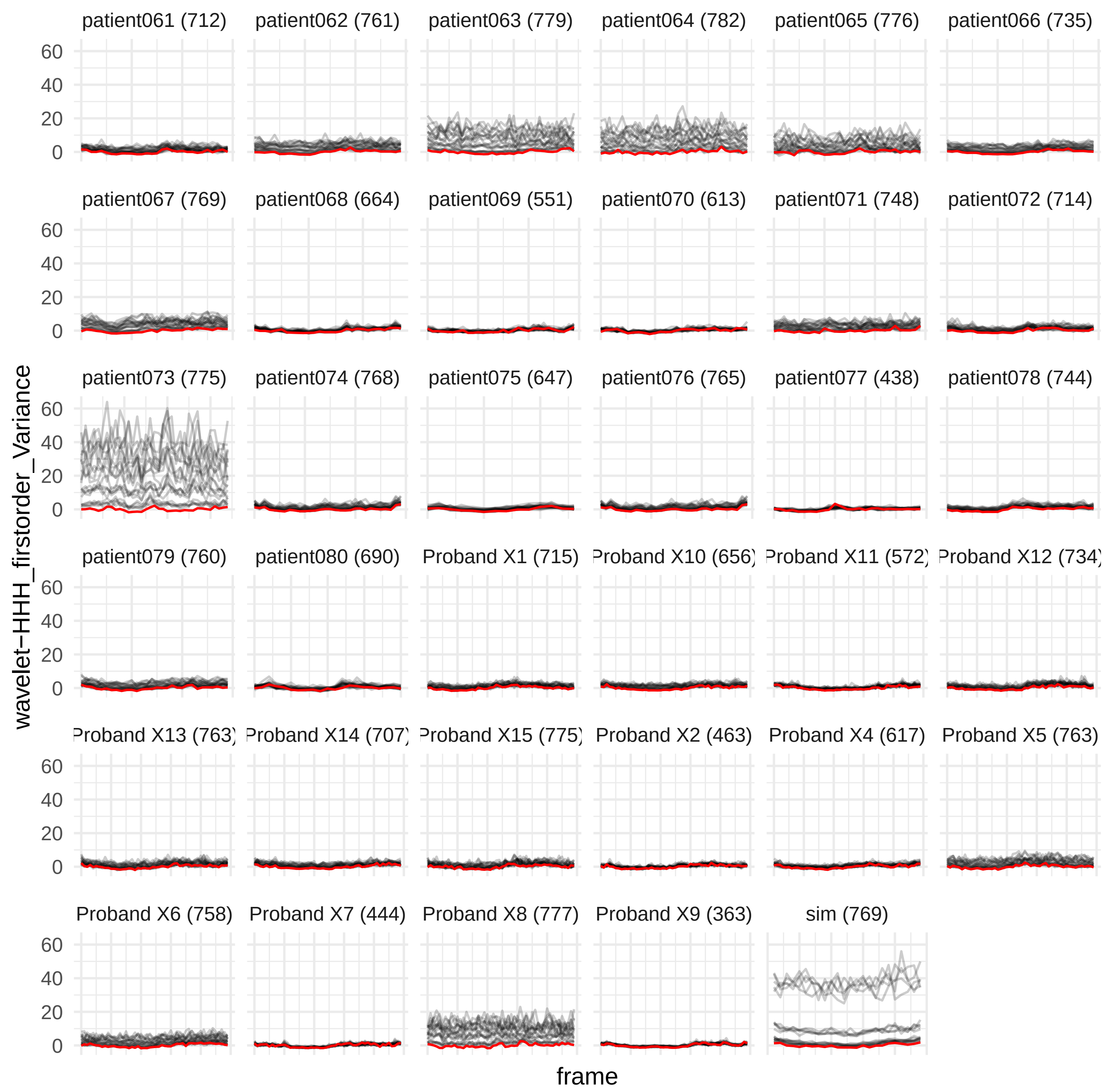
patient061 (712)
patient062 (761)
patient063 (779)
patient064 (782)
patient065 (776)
patient066 (735)
patient067 (769)
patient068 (664)
patient069 (551)
patient070 (613)
patient071 (748)
patient072 (714)
patient073 (775)
patient074 (768)
patient075 (647)
patient076 (765)
patient077 (438)
patient078 (744)
patient079 (760)
patient080 (690)
Proband X1 (715)
Proband X10 (656)
Proband X11 (572)
Proband X12 (734)
Proband X13 (763)
Proband X14 (707)
Proband X15 (775)
Proband X2 (463)
Proband X4 (617)
Proband X5 (763)
Proband X6 (758)
Proband X7 (444)
Proband X8 (777)
Proband X9 (363)
sim (769)
0
20
40
60
wavelet−HHH_firstorder_Variance
frame

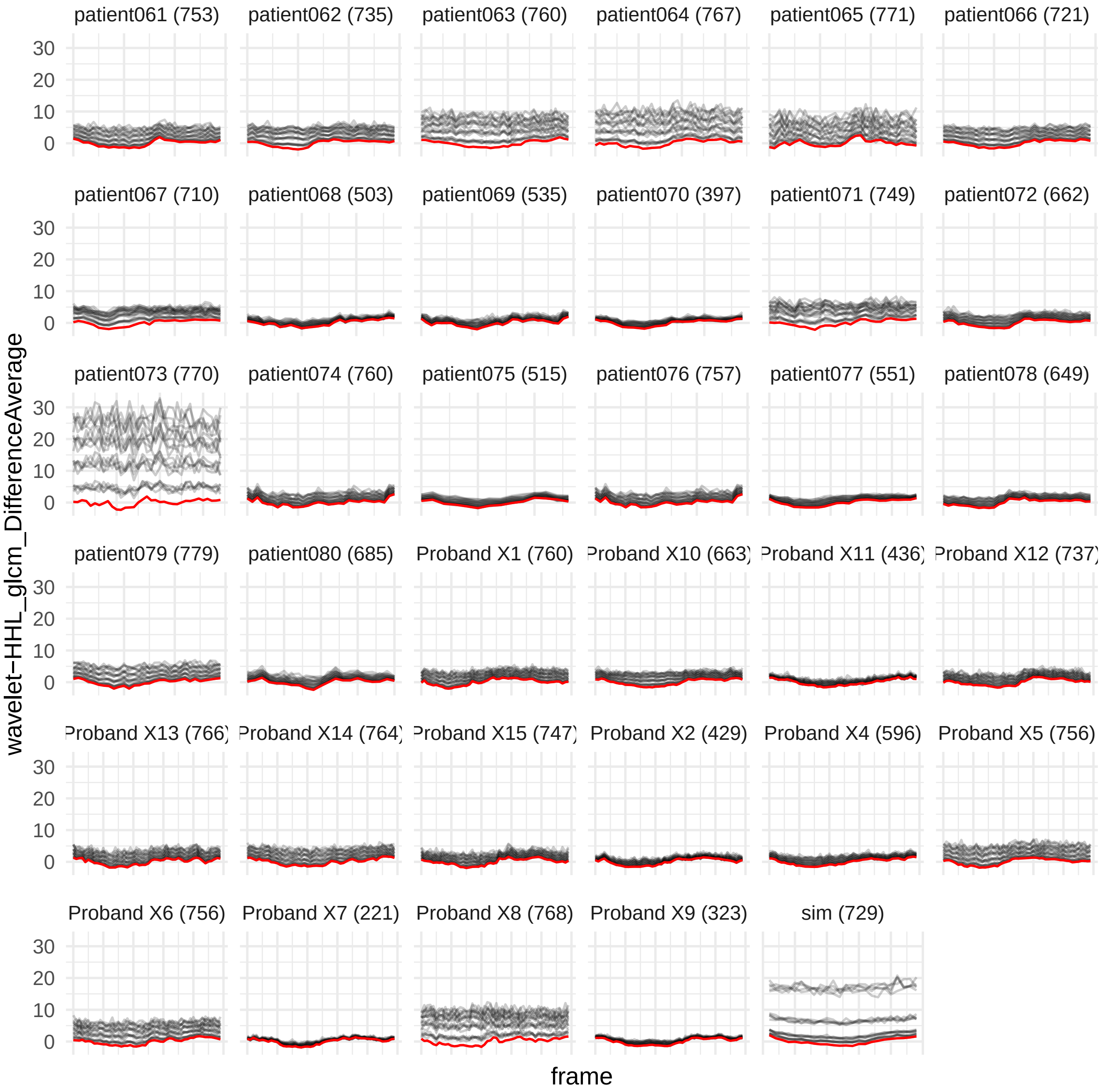
patient061 (753)
patient062 (735)
patient063 (760)
patient064 (767)
patient065 (771)
patient066 (721)
patient067 (710)
patient068 (503)
patient069 (535)
patient070 (397)
patient071 (749)
patient072 (662)
patient073 (770)
patient074 (760)
patient075 (515)
patient076 (757)
patient077 (551)
patient078 (649)
patient079 (779)
patient080 (685)
Proband X1 (760)
Proband X10 (663)
Proband X11 (436)
Proband X12 (737)
Proband X13 (766)
Proband X14 (764)
Proband X15 (747)
Proband X2 (429)
Proband X4 (596)
Proband X5 (756)
Proband X6 (756)
Proband X7 (221)
Proband X8 (768)
Proband X9 (323)
sim (729)
wavelet−HHL_glcm_DifferenceAverage
0
10
20
30
frame

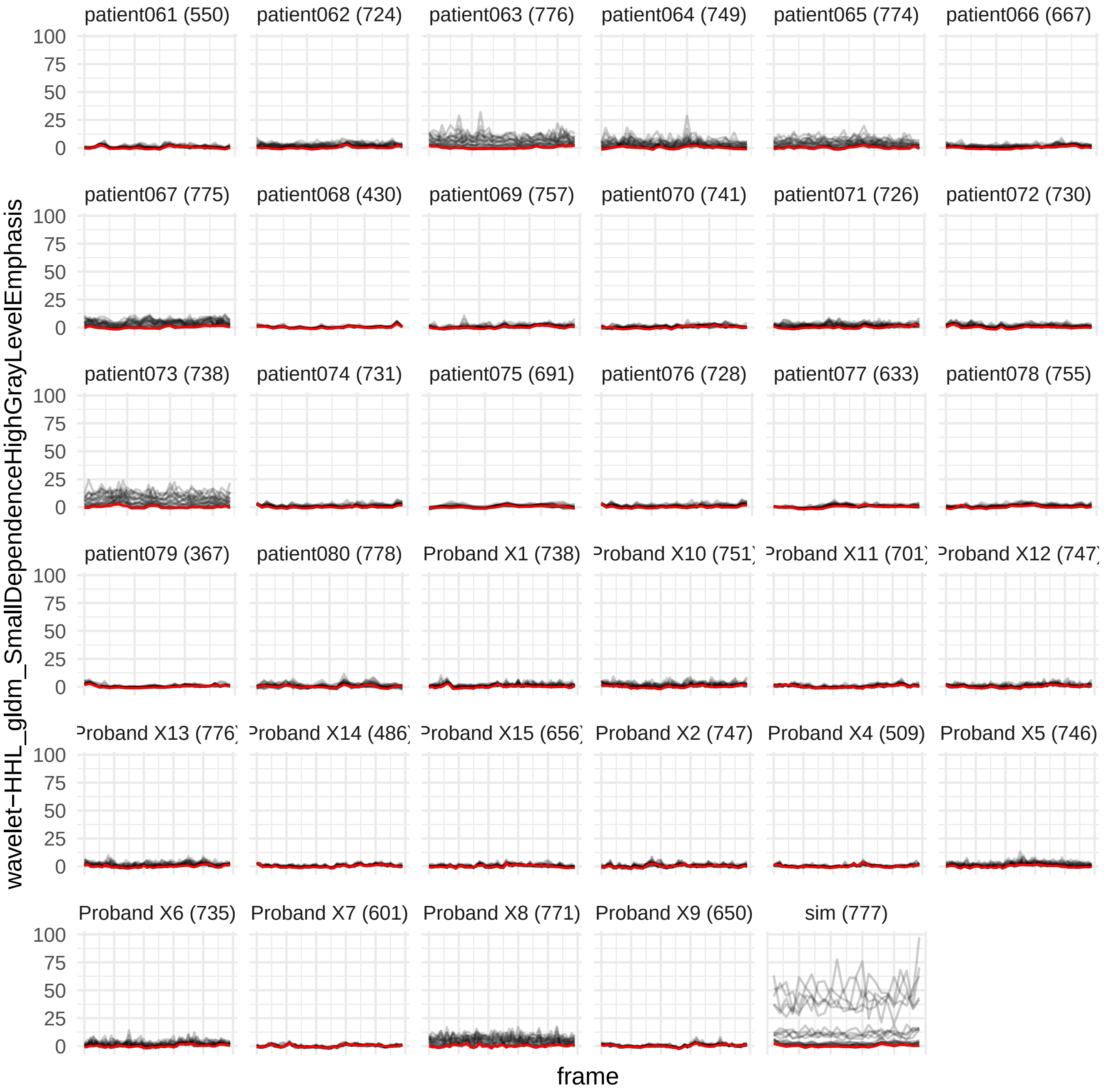

patient061 (550)
patient062 (724)
patient063 (776)
patient064 (749)
patient065 (774)
patient066 (667)
patient067 (775)
patient068 (430)
patient069 (757)
patient070 (741)
patient071 (726)
patient072 (730)
patient073 (738)
patient074 (731)
patient075 (691)
patient076 (728)
patient077 (633)
patient078 (755)
patient079 (367)
patient080 (778)
Proband X1 (738)
Proband X10 (751)
Proband X11 (701)
Proband X12 (747)
Proband X13 (776)
Proband X14 (486)
Proband X15 (656)
Proband X2 (747)
Proband X4 (509)
Proband X5 (746)
Proband X6 (735)
Proband X7 (601)
Proband X8 (771)
Proband X9 (650)
sim (777)
100
75
50
25
0
wavelet−HHL_gldm_SmallDependenceHighGrayLevelEmphasis
frame

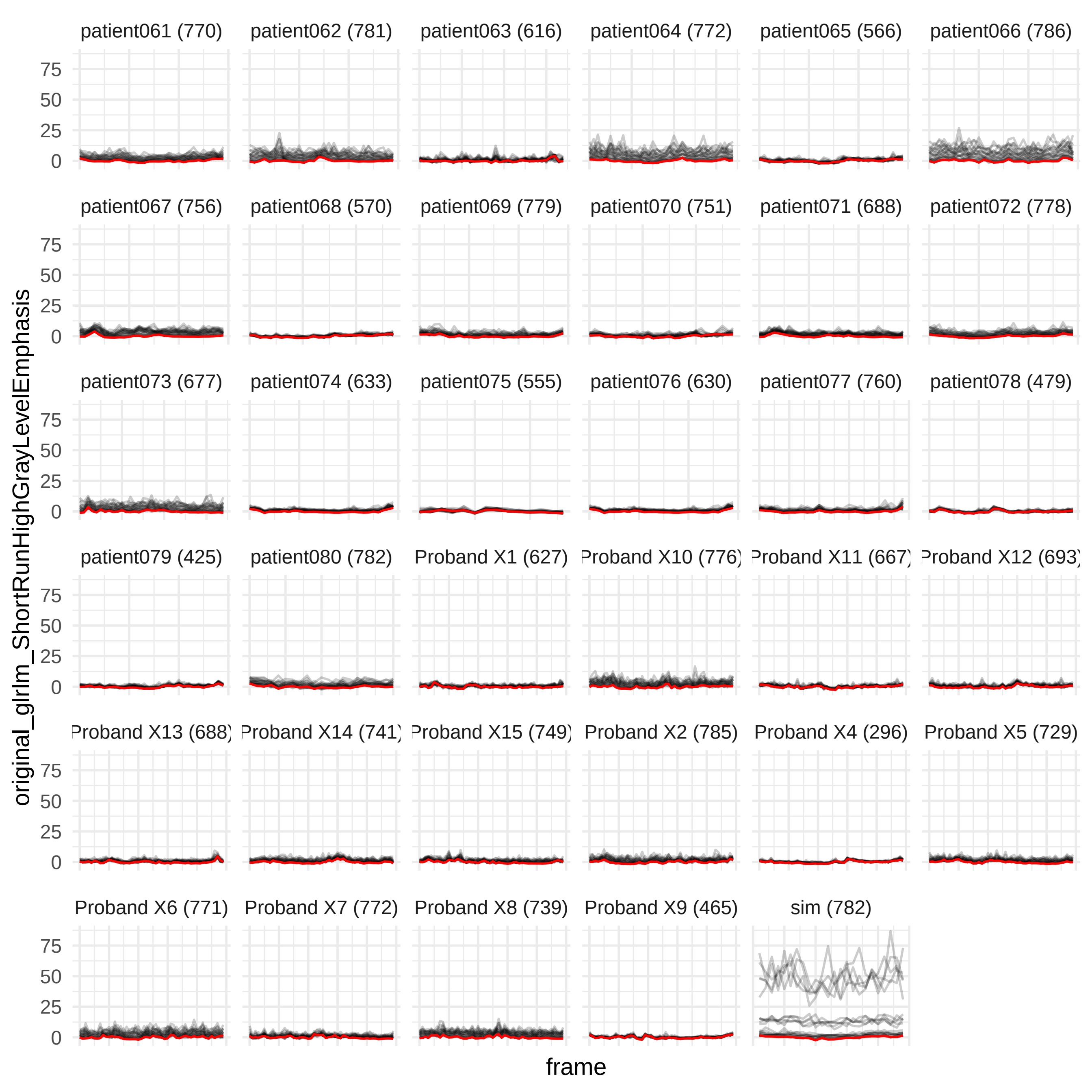
patient061 (770)
patient062 (781)
patient063 (616)
patient064 (772)
patient065 (566)
patient066 (786)
patient067 (756)
patient068 (570)
patient069 (779)
patient070 (751)
patient071 (688)
patient072 (778)
patient073 (677)
patient074 (633)
patient075 (555)
patient076 (630)
patient077 (760)
patient078 (479)
patient079 (425)
patient080 (782)
Proband X1 (627)
Proband X10 (776)
Proband X11 (667)
Proband X12 (693)
Proband X13 (688)
Proband X14 (741)
Proband X15 (749)
Proband X2 (785)
Proband X4 (296)
Proband X5 (729)
Proband X6 (771)
Proband X7 (772)
Proband X8 (739)
Proband X9 (465)
sim (782)
original_glrlm_ShortRunHighGrayLevelEmphasis
frame
0
25
50
75

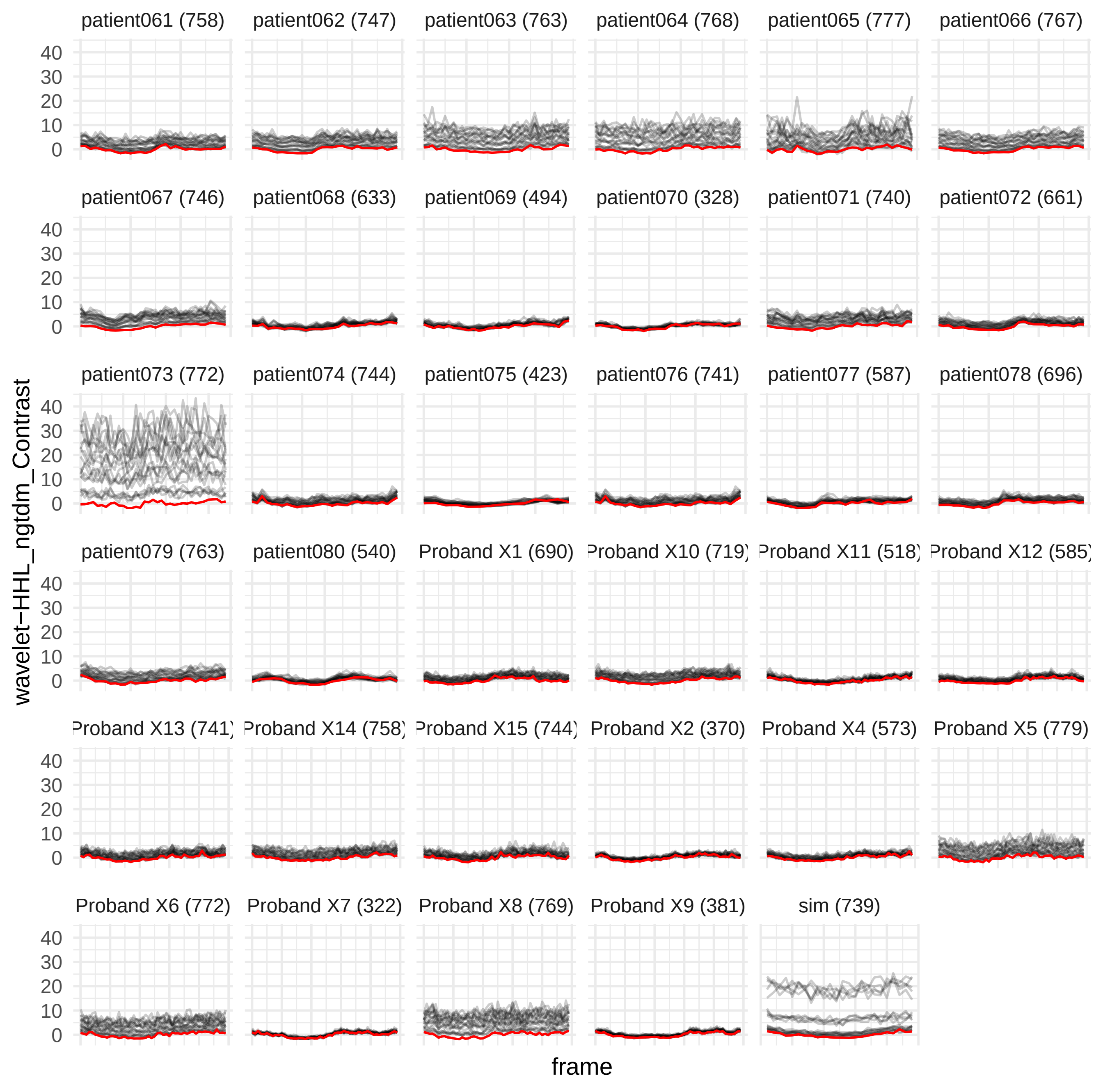

patient061 (758)
patient062 (747)
patient063 (763)
patient064 (768)
patient065 (777)
patient066 (767)
patient067 (746)
patient068 (633)
patient069 (494)
patient070 (328)
patient071 (740)
patient072 (661)
patient073 (772)
patient074 (744)
patient075 (423)
patient076 (741)
patient077 (587)
patient078 (696)
patient079 (763)
patient080 (540)
Proband X1 (690)
Proband X10 (719)
Proband X11 (518)
Proband X12 (585)
Proband X13 (741)
Proband X14 (758)
Proband X15 (744)
Proband X2 (370)
Proband X4 (573)
Proband X5 (779)
Proband X6 (772)
Proband X7 (322)
Proband X8 (769)
Proband X9 (381)
sim (739)
0
10
20
30
40
wavelet−HHL_ngtdm_Contrast
frame

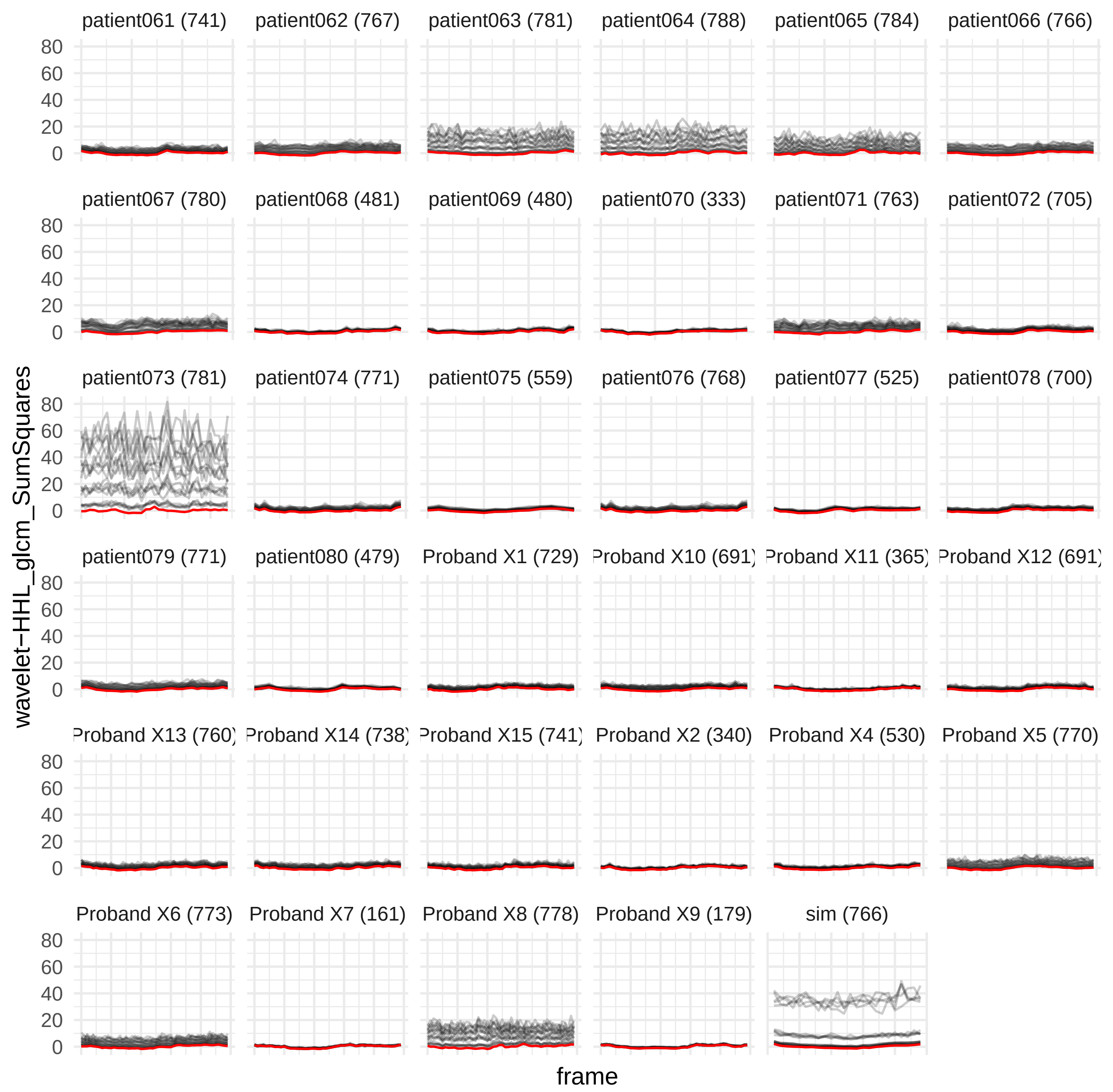
patient061 (741)
patient062 (767)
patient063 (781)
patient064 (788)
patient065 (784)
patient066 (766)
patient067 (780)
patient068 (481)
patient069 (480)
patient070 (333)
patient071 (763)
patient072 (705)
patient073 (781)
patient074 (771)
patient075 (559)
patient076 (768)
patient077 (525)
patient078 (700)
patient079 (771)
patient080 (479)
Proband X1 (729)
Proband X10 (691)
Proband X11 (365)
Proband X12 (691)
Proband X13 (760)
Proband X14 (738)
Proband X15 (741)
Proband X2 (340)
Proband X4 (530)
Proband X5 (770)
Proband X6 (773)
Proband X7 (161)
Proband X8 (778)
Proband X9 (179)
sim (766)
0
20
40
60
80
wavelet−HHL_glcm_SumSquares
frame

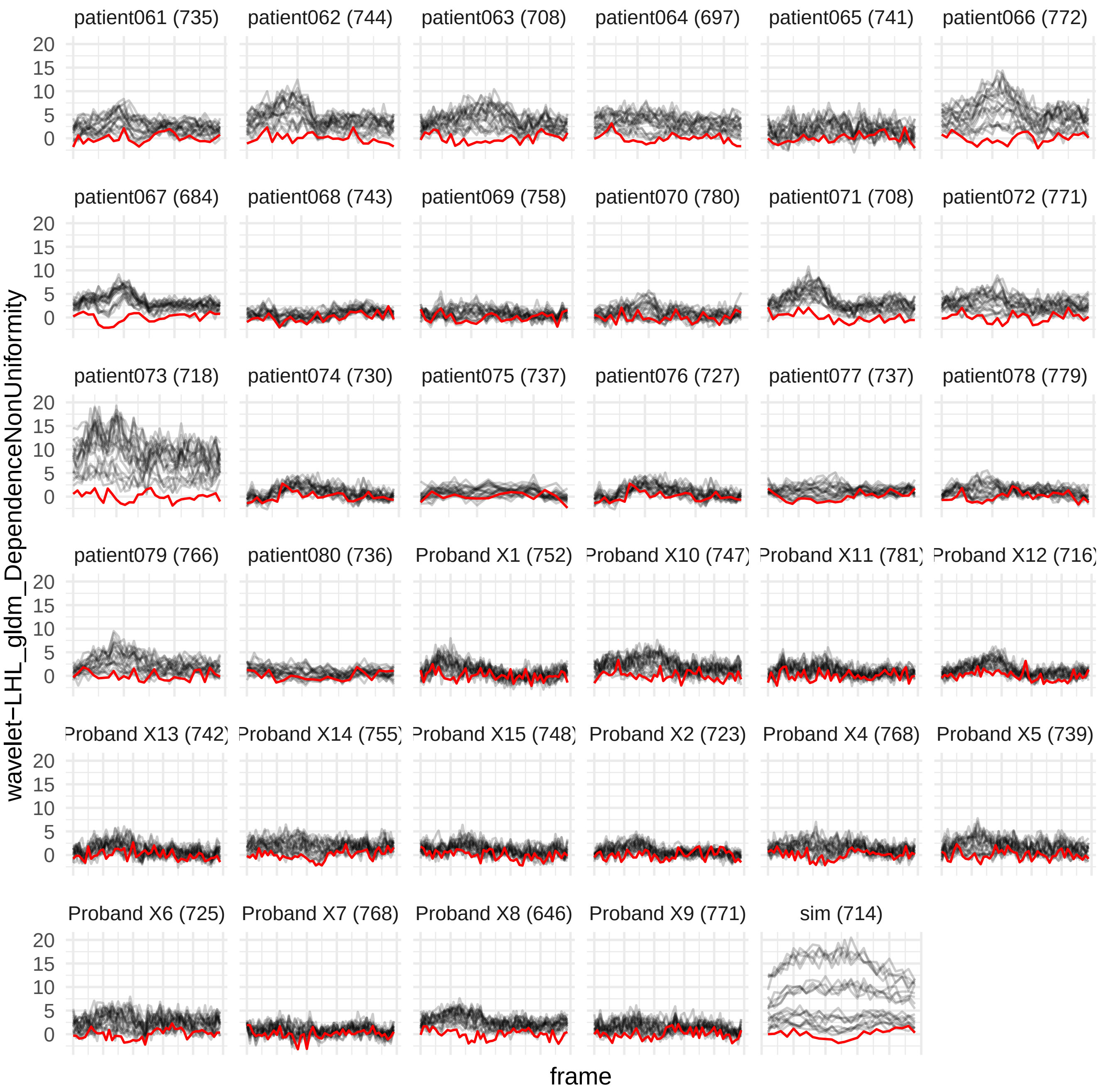
patient061 (735)
patient062 (744)
patient063 (708)
patient064 (697)
patient065 (741)
patient066 (772)
patient067 (684)
patient068 (743)
patient069 (758)
patient070 (780)
patient071 (708)
patient072 (771)
patient073 (718)
patient074 (730)
patient075 (737)
patient076 (727)
patient077 (737)
patient078 (779)
patient079 (766)
patient080 (736)
Proband X1 (752)
Proband X10 (747)
Proband X11 (781)
Proband X12 (716)
Proband X13 (742)
Proband X14 (755)
Proband X15 (748)
Proband X2 (723)
Proband X4 (768)
Proband X5 (739)
Proband X6 (725)
Proband X7 (768)
Proband X8 (646)
Proband X9 (771)
sim (714)
0
5
10
15
20
wavelet−LHL_gldm_DependenceNonUniformity
frame

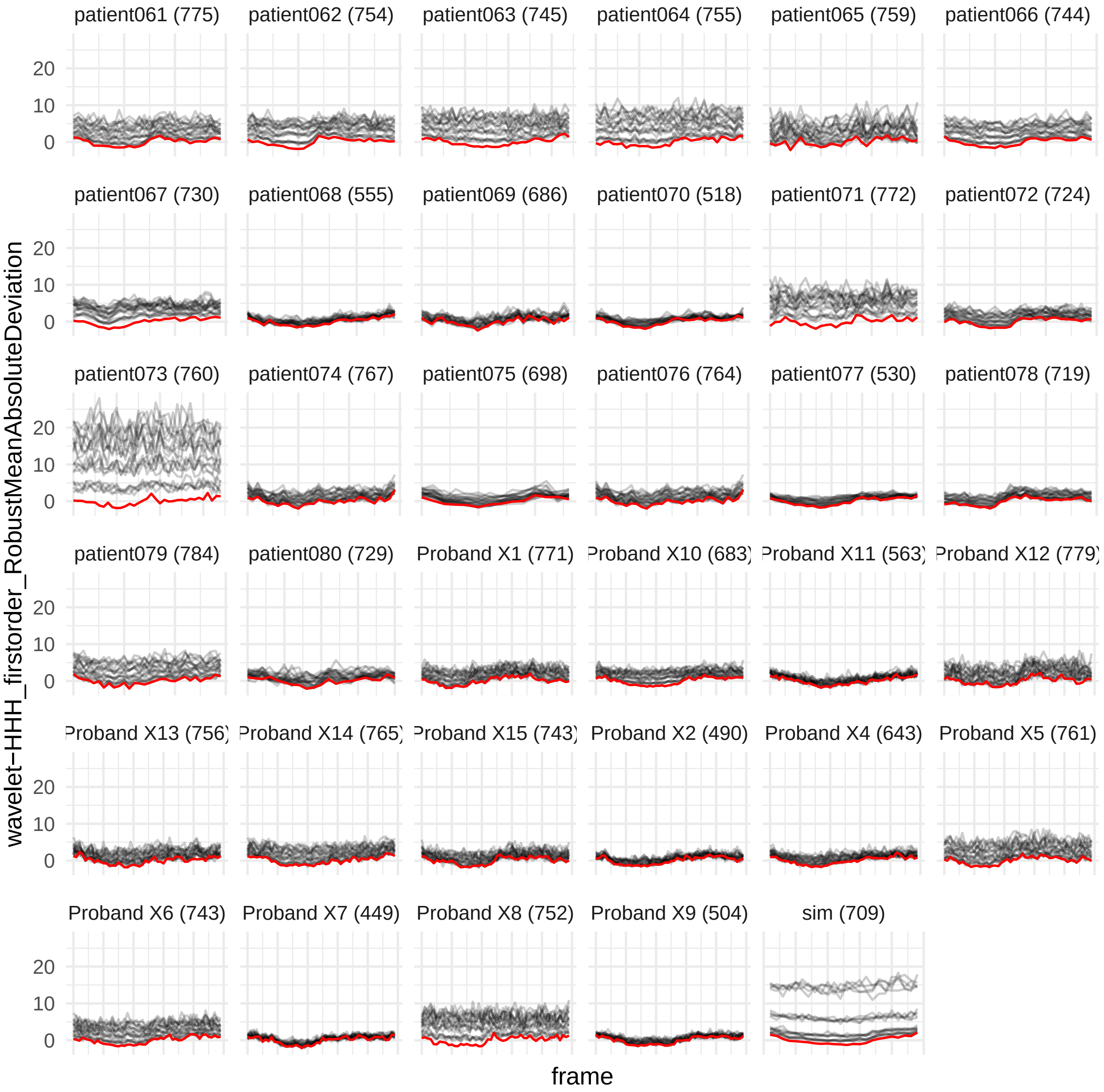
patient061 (775)
patient062 (754)
patient063 (745)
patient064 (755)
patient065 (759)
patient066 (744)
patient067 (730)
patient068 (555)
patient069 (686)
patient070 (518)
patient071 (772)
patient072 (724)
patient073 (760)
patient074 (767)
patient075 (698)
patient076 (764)
patient077 (530)
patient078 (719)
patient079 (784)
patient080 (729)
Proband X1 (771)
Proband X10 (683)
Proband X11 (563)
Proband X12 (779)
Proband X13 (756)
Proband X14 (765)
Proband X15 (743)
Proband X2 (490)
Proband X4 (643)
Proband X5 (761)
Proband X6 (743)
Proband X7 (449)
Proband X8 (752)
Proband X9 (504)
sim (709)
wavelet−HHH_firstorder_RobustMeanAbsoluteDeviation
frame
0
10
20

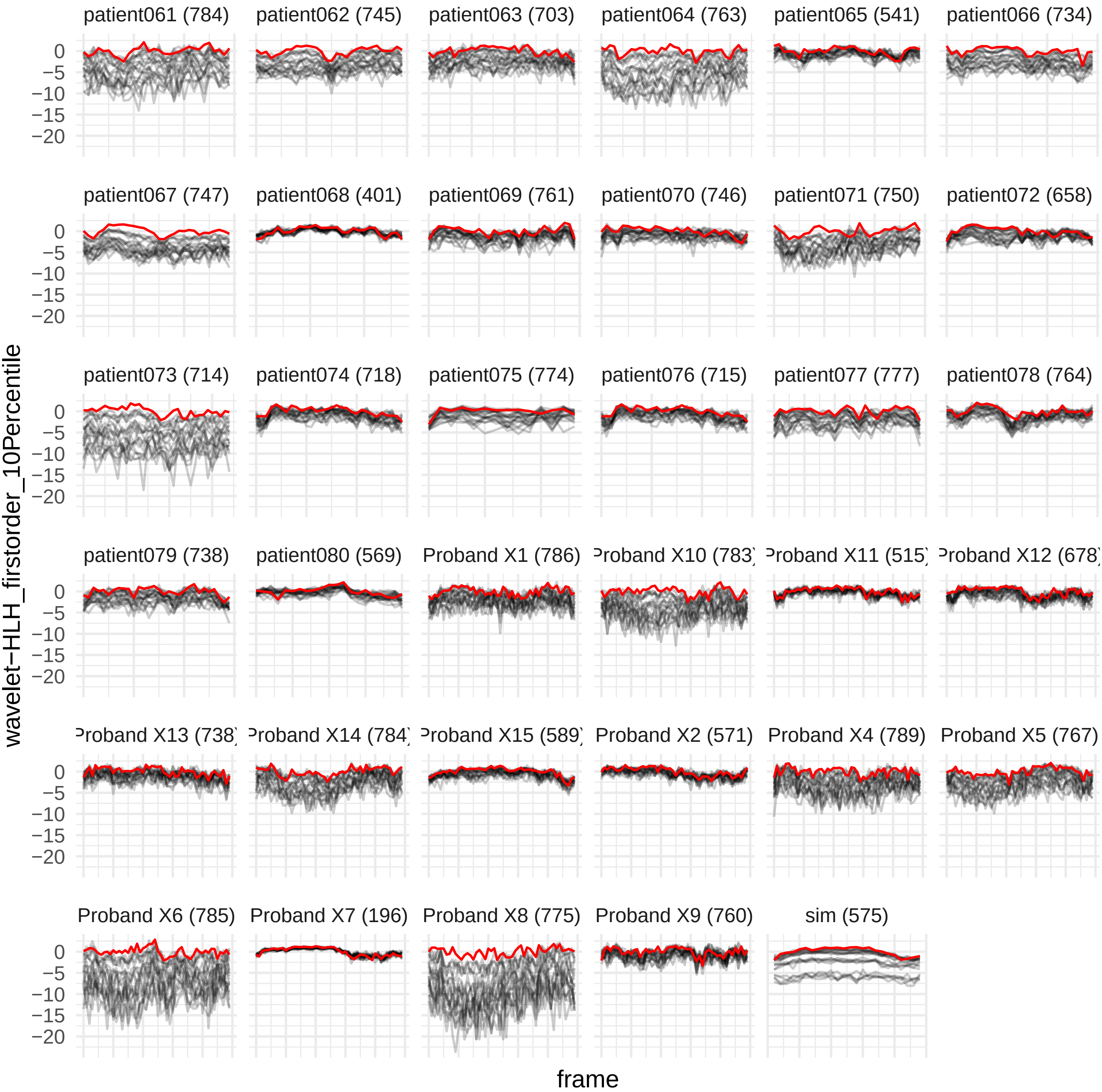

patient061 (784)
patient062 (745)
patient063 (703)
patient064 (763)
patient065 (541)
patient066 (734)
patient067 (747)
patient068 (401)
patient069 (761)
patient070 (746)
patient071 (750)
patient072 (658)
patient073 (714)
patient074 (718)
patient075 (774)
patient076 (715)
patient077 (777)
patient078 (764)
patient079 (738)
patient080 (569)
Proband X1 (786)
Proband X10 (783)
Proband X11 (515)
Proband X12 (678)
Proband X13 (738)
Proband X14 (784)
Proband X15 (589)
Proband X2 (571)
Proband X4 (789)
Proband X5 (767)
Proband X6 (785)
Proband X7 (196)
Proband X8 (775)
Proband X9 (760)
sim (575)
0
−5
−10
−15
−20
wavelet−HLH_firstorder_10Percentile
frame

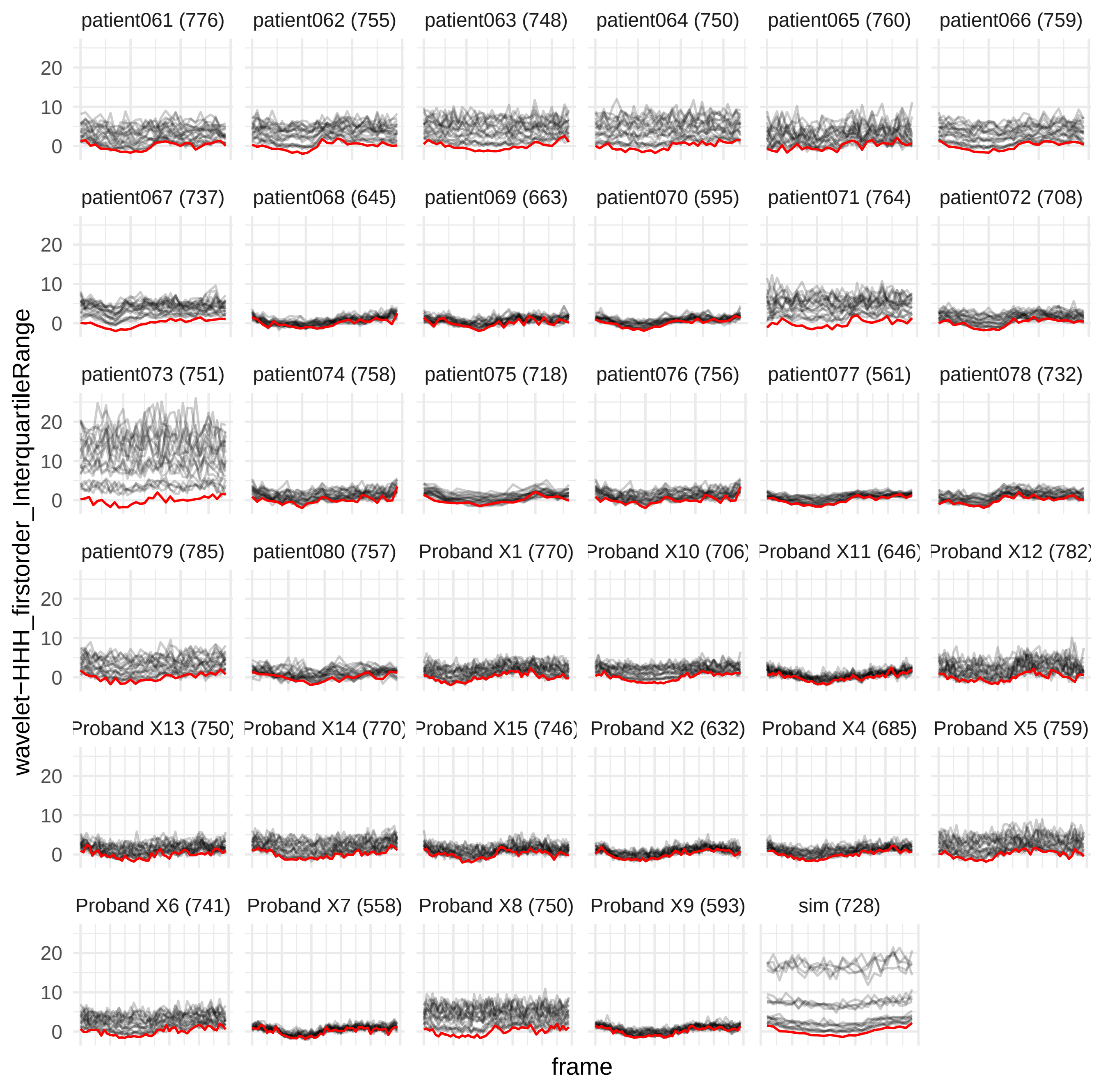
patient061 (776)
patient062 (755)
patient063 (748)
patient064 (750)
patient065 (760)
patient066 (759)
patient067 (737)
patient068 (645)
patient069 (663)
patient070 (595)
patient071 (764)
patient072 (708)
patient073 (751)
patient074 (758)
patient075 (718)
patient076 (756)
patient077 (561)
patient078 (732)
patient079 (785)
patient080 (757)
Proband X1 (770)
Proband X10 (706)
Proband X11 (646)
Proband X12 (782)
Proband X13 (750)
Proband X14 (770)
Proband X15 (746)
Proband X2 (632)
Proband X4 (685)
Proband X5 (759)
Proband X6 (741)
Proband X7 (558)
Proband X8 (750)
Proband X9 (593)
sim (728)
wavelet−HHH_firstorder_InterquartileRange
20
10
0
frame

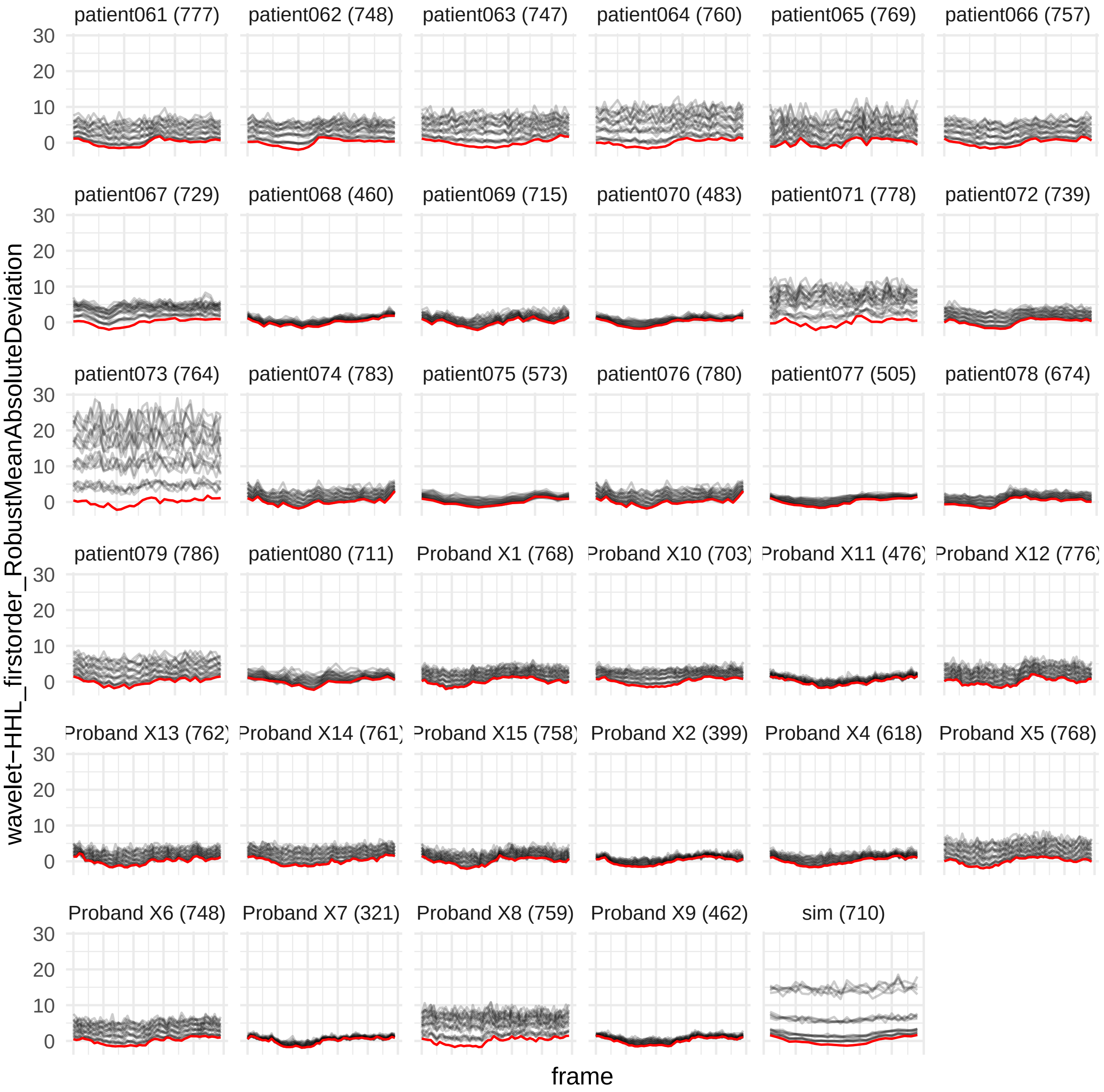

patient061 (777)
patient062 (748)
patient063 (747)
patient064 (760)
patient065 (769)
patient066 (757)
patient067 (729)
patient068 (460)
patient069 (715)
patient070 (483)
patient071 (778)
patient072 (739)
patient073 (764)
patient074 (783)
patient075 (573)
patient076 (780)
patient077 (505)
patient078 (674)
patient079 (786)
patient080 (711)
Proband X1 (768)
Proband X10 (703)
Proband X11 (476)
Proband X12 (776)
Proband X13 (762)
Proband X14 (761)
Proband X15 (758)
Proband X2 (399)
Proband X4 (618)
Proband X5 (768)
Proband X6 (748)
Proband X7 (321)
Proband X8 (759)
Proband X9 (462)
sim (710)
wavelet−HHL_firstorder_RobustMeanAbsoluteDeviation
frame

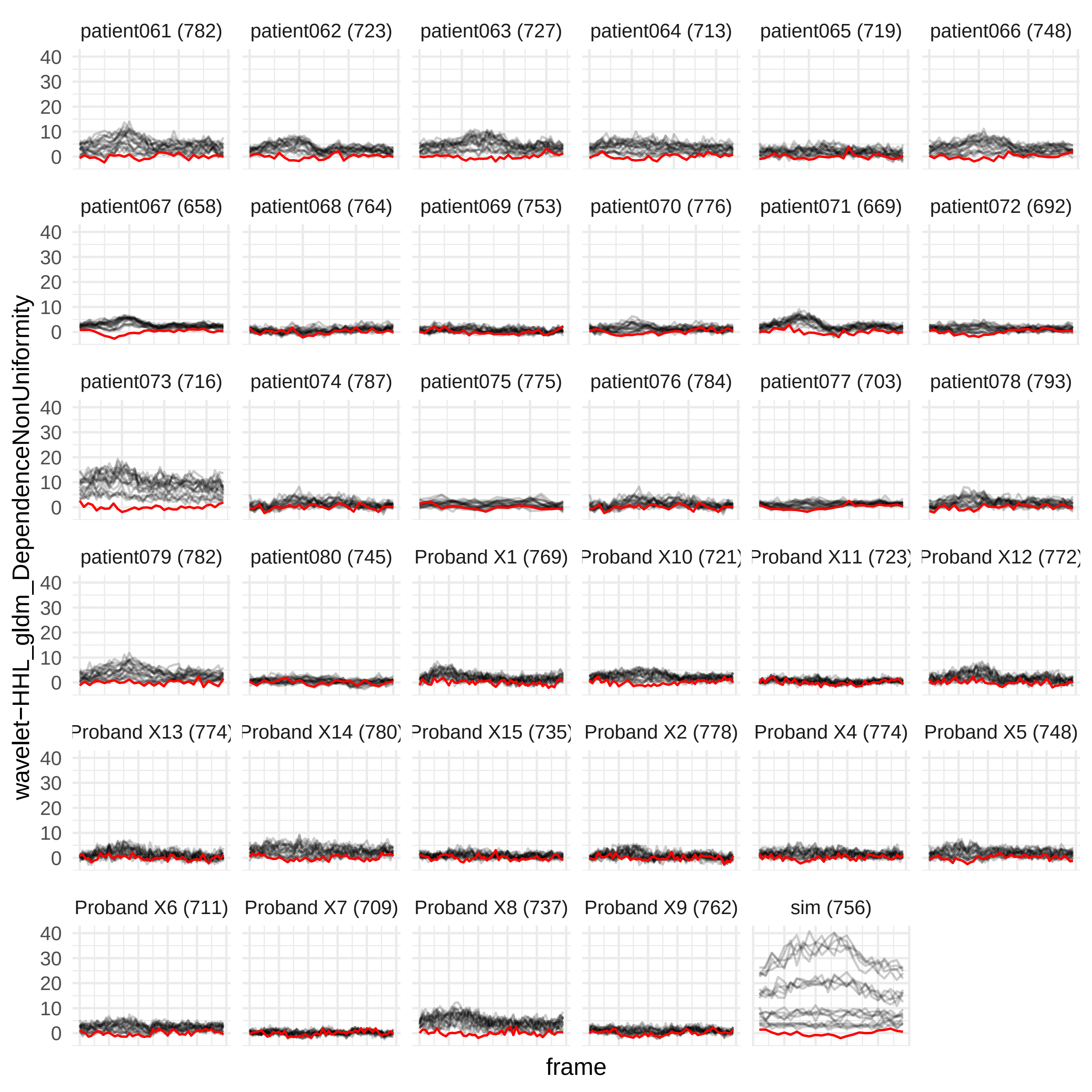

patient061 (782)
patient062 (723)
patient063 (727)
patient064 (713)
patient065 (719)
patient066 (748)
patient067 (658)
patient068 (764)
patient069 (753)
patient070 (776)
patient071 (669)
patient072 (692)
patient073 (716)
patient074 (787)
patient075 (775)
patient076 (784)
patient077 (703)
patient078 (793)
patient079 (782)
patient080 (745)
Proband X1 (769)
Proband X10 (721)
Proband X11 (723)
Proband X12 (772)
Proband X13 (774)
Proband X14 (780)
Proband X15 (735)
Proband X2 (778)
Proband X4 (774)
Proband X5 (748)
Proband X6 (711)
Proband X7 (709)
Proband X8 (737)
Proband X9 (762)
sim (756)
40
30
20
10
0
wavelet−HHL_gldm_DependenceNonUniformity
frame

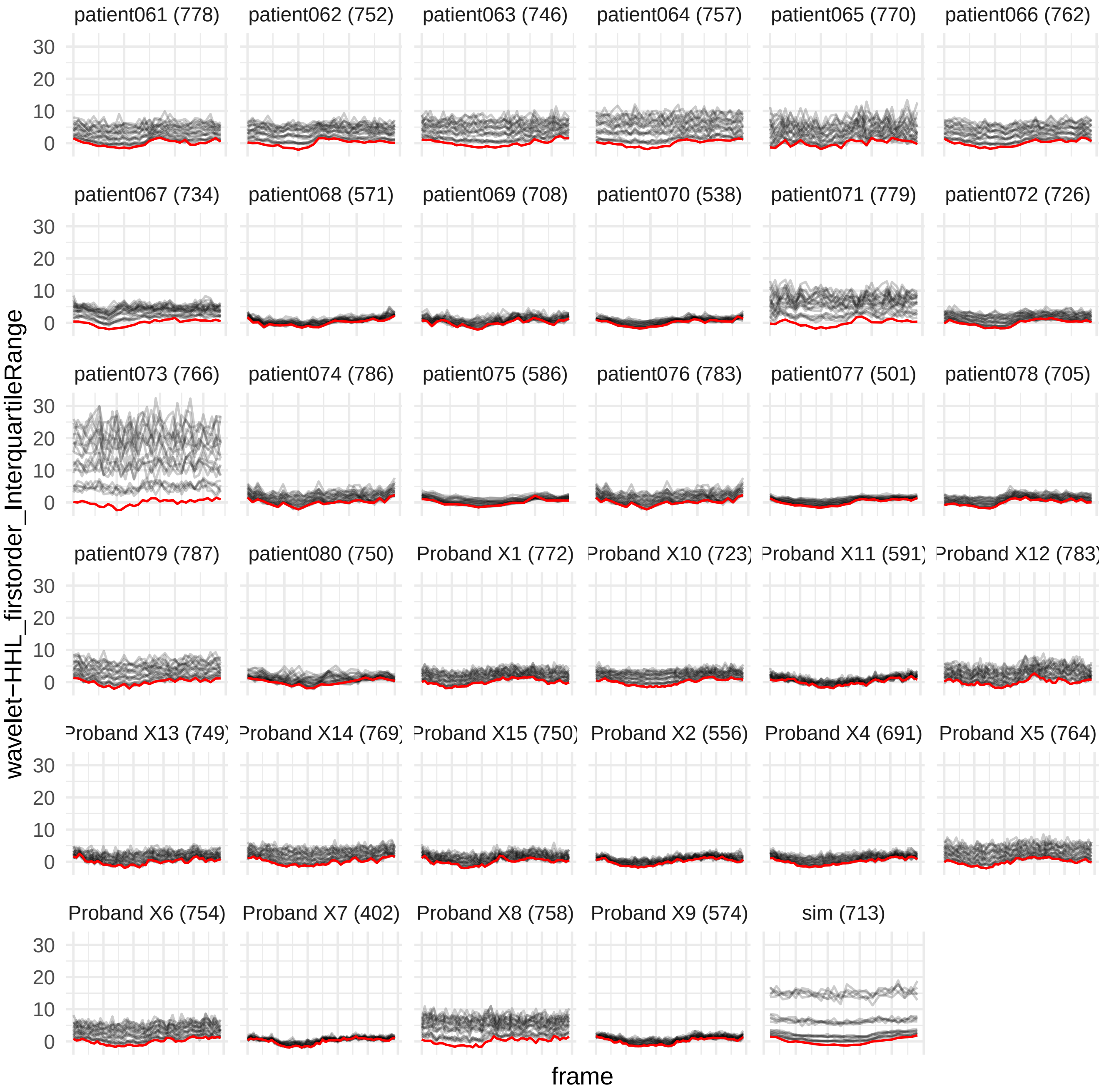
patient061 (778)
patient062 (752)
patient063 (746)
patient064 (757)
patient065 (770)
patient066 (762)
patient067 (734)
patient068 (571)
patient069 (708)
patient070 (538)
patient071 (779)
patient072 (726)
patient073 (766)
patient074 (786)
patient075 (586)
patient076 (783)
patient077 (501)
patient078 (705)
patient079 (787)
patient080 (750)
Proband X1 (772)
Proband X10 (723)
Proband X11 (591)
Proband X12 (783)
Proband X13 (749)
Proband X14 (769)
Proband X15 (750)
Proband X2 (556)
Proband X4 (691)
Proband X5 (764)
Proband X6 (754)
Proband X7 (402)
Proband X8 (758)
Proband X9 (574)
sim (713)
0
10
20
30
wavelet−HHL_firstorder_InterquartileRange
frame

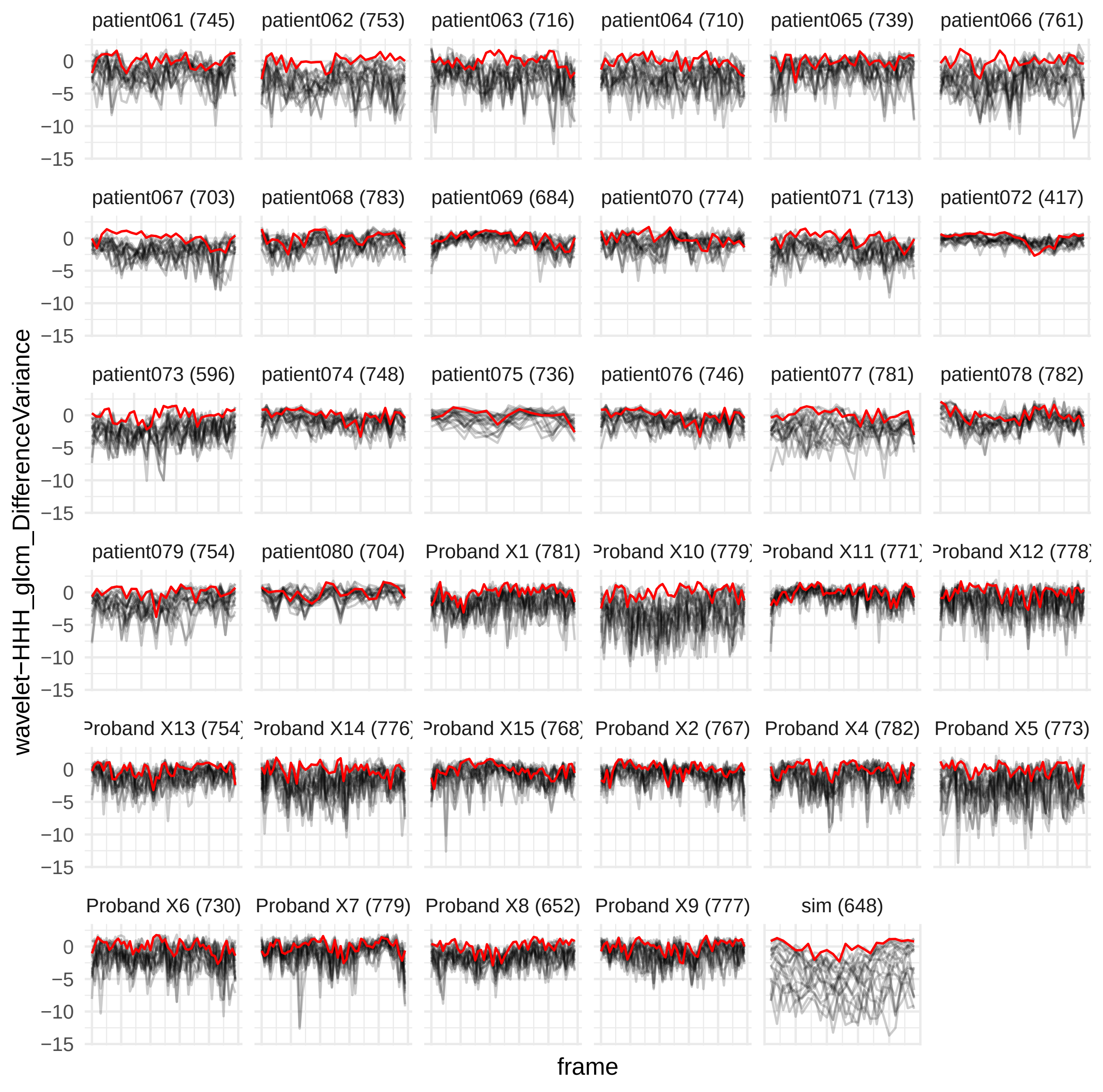
patient061 (745)
patient062 (753)
patient063 (716)
patient064 (710)
patient065 (739)
patient066 (761)
patient067 (703)
patient068 (783)
patient069 (684)
patient070 (774)
patient071 (713)
patient072 (417)
patient073 (596)
patient074 (748)
patient075 (736)
patient076 (746)
patient077 (781)
patient078 (782)
patient079 (754)
patient080 (704)
Proband X1 (781)
Proband X10 (779)
Proband X11 (771)
Proband X12 (778)
Proband X13 (754)
Proband X14 (776)
Proband X15 (768)
Proband X2 (767)
Proband X4 (782)
Proband X5 (773)
Proband X6 (730)
Proband X7 (779)
Proband X8 (652)
Proband X9 (777)
sim (648)
0
−5
−10
−15
wavelet−HHH_glcm_DifferenceVariance
frame

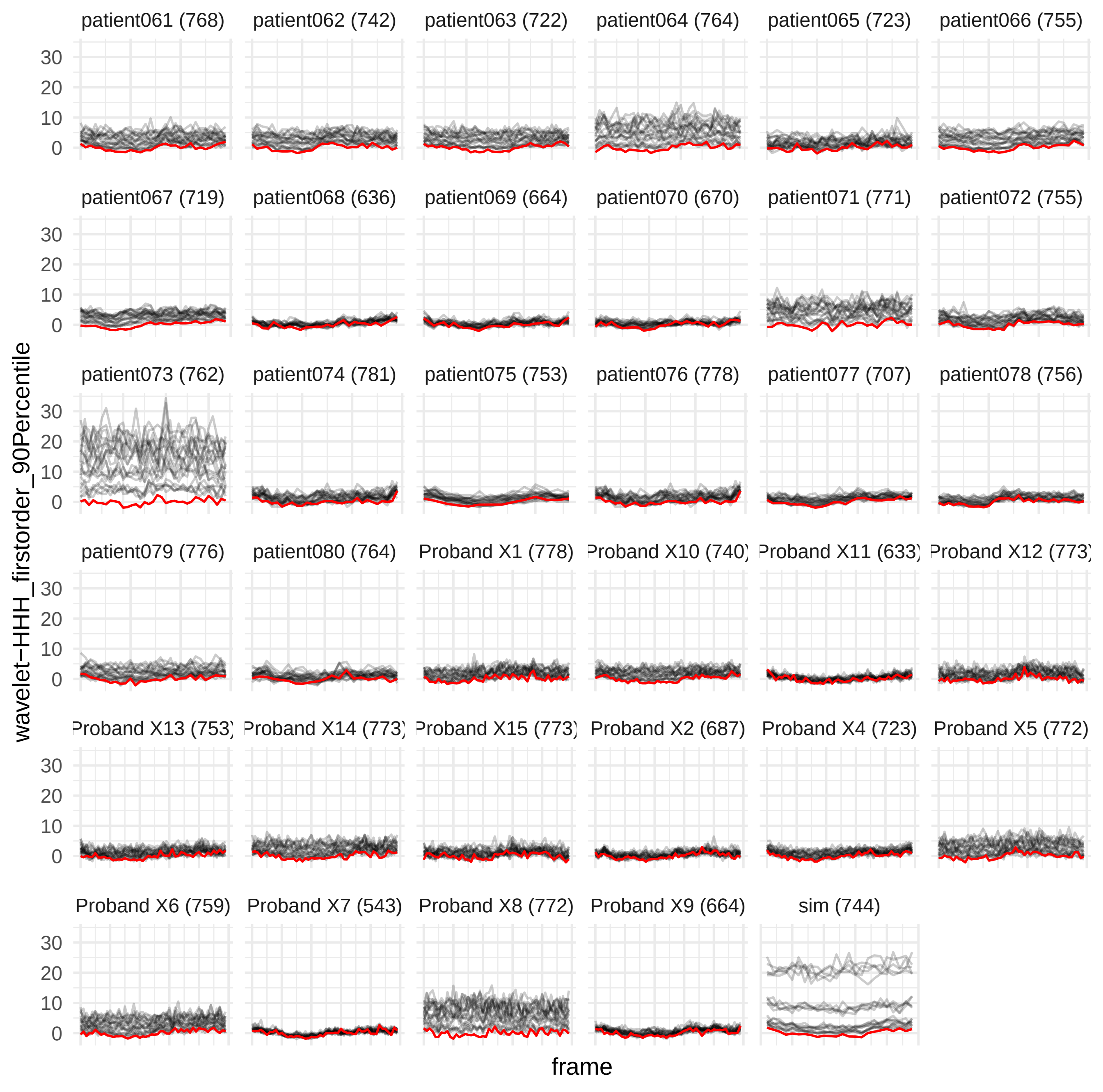
patient061 (768)
patient062 (742)
patient063 (722)
patient064 (764)
patient065 (723)
patient066 (755)
patient067 (719)
patient068 (636)
patient069 (664)
patient070 (670)
patient071 (771)
patient072 (755)
patient073 (762)
patient074 (781)
patient075 (753)
patient076 (778)
patient077 (707)
patient078 (756)
patient079 (776)
patient080 (764)
Proband X1 (778)
Proband X10 (740)
Proband X11 (633)
Proband X12 (773)
Proband X13 (753)
Proband X14 (773)
Proband X15 (773)
Proband X2 (687)
Proband X4 (723)
Proband X5 (772)
Proband X6 (759)
Proband X7 (543)
Proband X8 (772)
Proband X9 (664)
sim (744)
0
10
20
30
wavelet−HHH_firstorder_90Percentile
frame

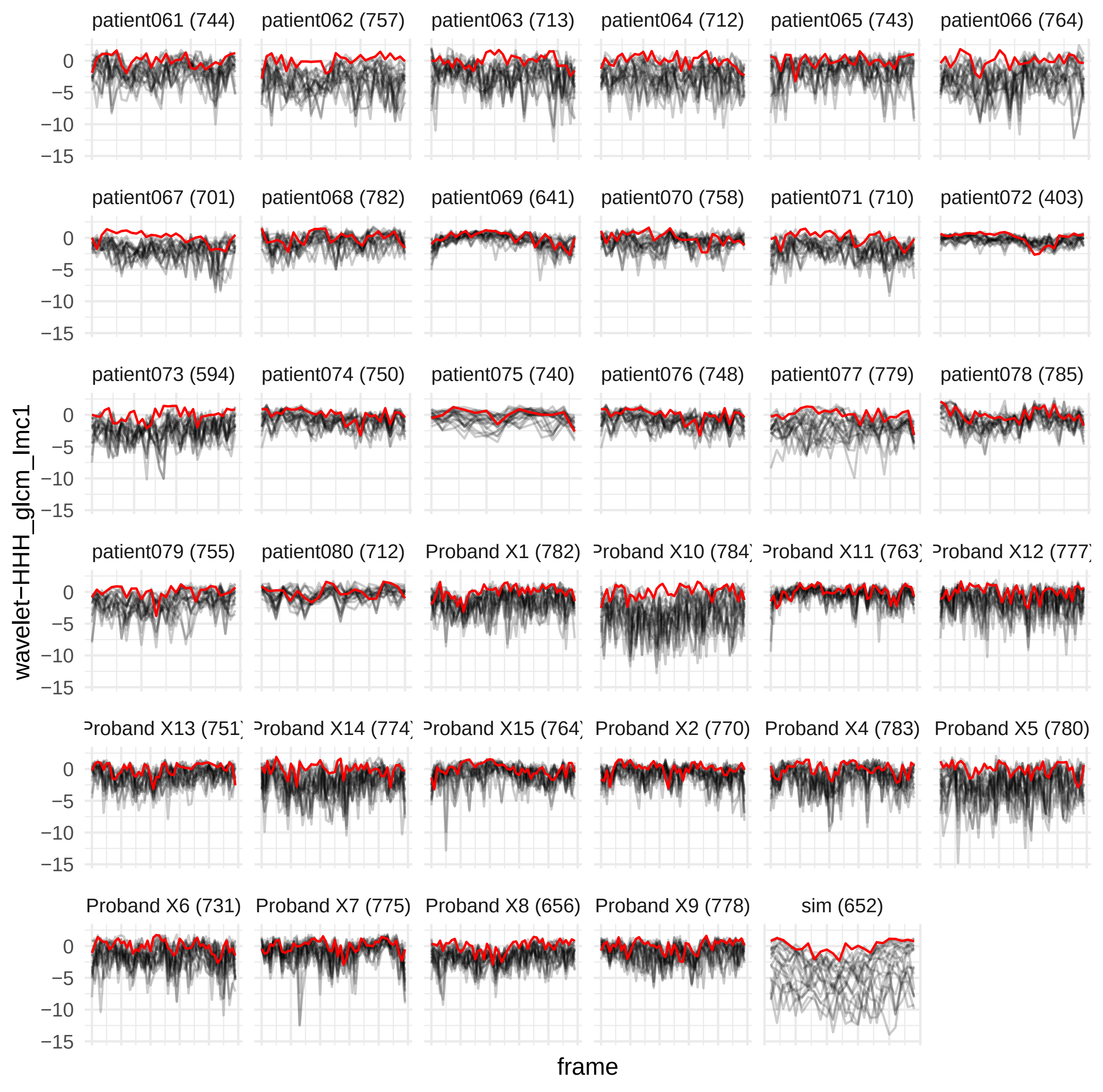

patient061 (744)
patient062 (757)
patient063 (713)
patient064 (712)
patient065 (743)
patient066 (764)
patient067 (701)
patient068 (782)
patient069 (641)
patient070 (758)
patient071 (710)
patient072 (403)
patient073 (594)
patient074 (750)
patient075 (740)
patient076 (748)
patient077 (779)
patient078 (785)
patient079 (755)
patient080 (712)
Proband X1 (782)
Proband X10 (784)
Proband X11 (763)
Proband X12 (777)
Proband X13 (751)
Proband X14 (774)
Proband X15 (764)
Proband X2 (770)
Proband X4 (783)
Proband X5 (780)
Proband X6 (731)
Proband X7 (775)
Proband X8 (656)
Proband X9 (778)
sim (652)
0
−5
−10
−15
wavelet−HHH_glcm_Imc1
frame

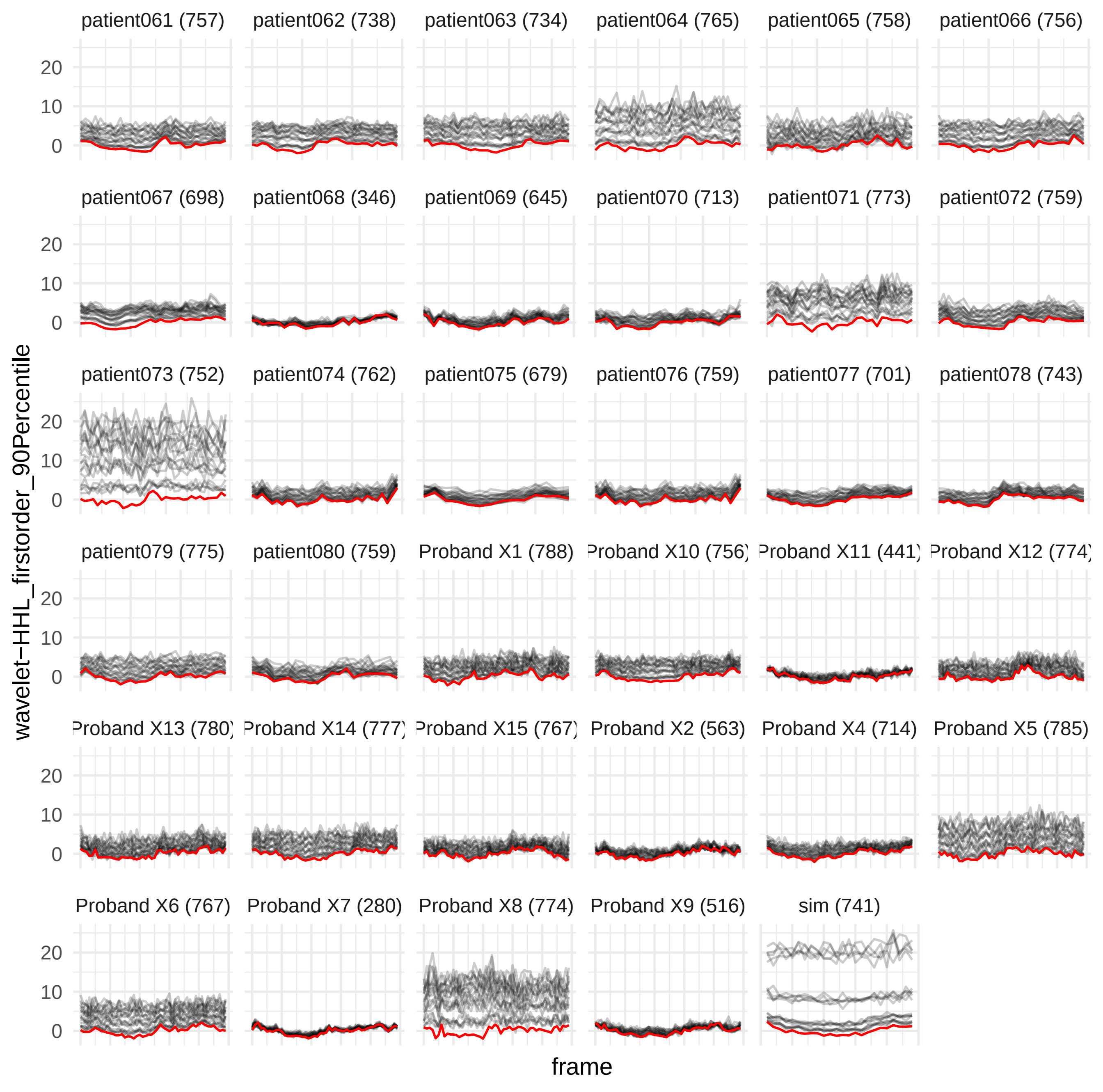

patient061 (757)
patient062 (738)
patient063 (734)
patient064 (765)
patient065 (758)
patient066 (756)
patient067 (698)
patient068 (346)
patient069 (645)
patient070 (713)
patient071 (773)
patient072 (759)
patient073 (752)
patient074 (762)
patient075 (679)
patient076 (759)
patient077 (701)
patient078 (743)
patient079 (775)
patient080 (759)
Proband X1 (788)
Proband X10 (756)
Proband X11 (441)
Proband X12 (774)
Proband X13 (780)
Proband X14 (777)
Proband X15 (767)
Proband X2 (563)
Proband X4 (714)
Proband X5 (785)
Proband X6 (767)
Proband X7 (280)
Proband X8 (774)
Proband X9 (516)
sim (741)
wavelet−HHL_firstorder_90Percentile
frame
0
10
20

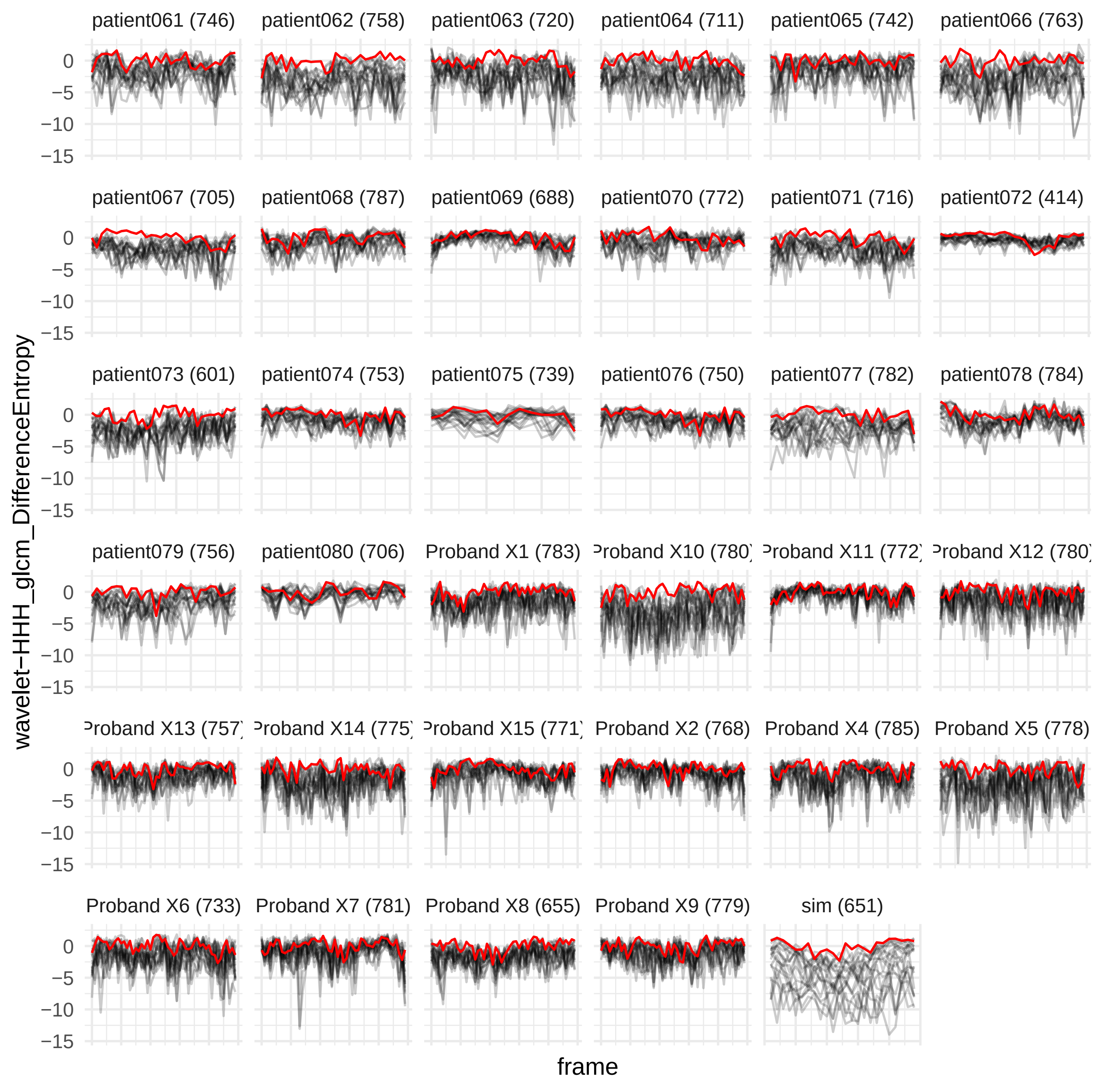
patient061 (746)
patient062 (758)
patient063 (720)
patient064 (711)
patient065 (742)
patient066 (763)
patient067 (705)
patient068 (787)
patient069 (688)
patient070 (772)
patient071 (716)
patient072 (414)
patient073 (601)
patient074 (753)
patient075 (739)
patient076 (750)
patient077 (782)
patient078 (784)
patient079 (756)
patient080 (706)
Proband X1 (783)
Proband X10 (780)
Proband X11 (772)
Proband X12 (780)
Proband X13 (757)
Proband X14 (775)
Proband X15 (771)
Proband X2 (768)
Proband X4 (785)
Proband X5 (778)
Proband X6 (733)
Proband X7 (781)
Proband X8 (655)
Proband X9 (779)
sim (651)
0
−5
−10
−15
wavelet−HHH_glcm_DifferenceEntropy
frame

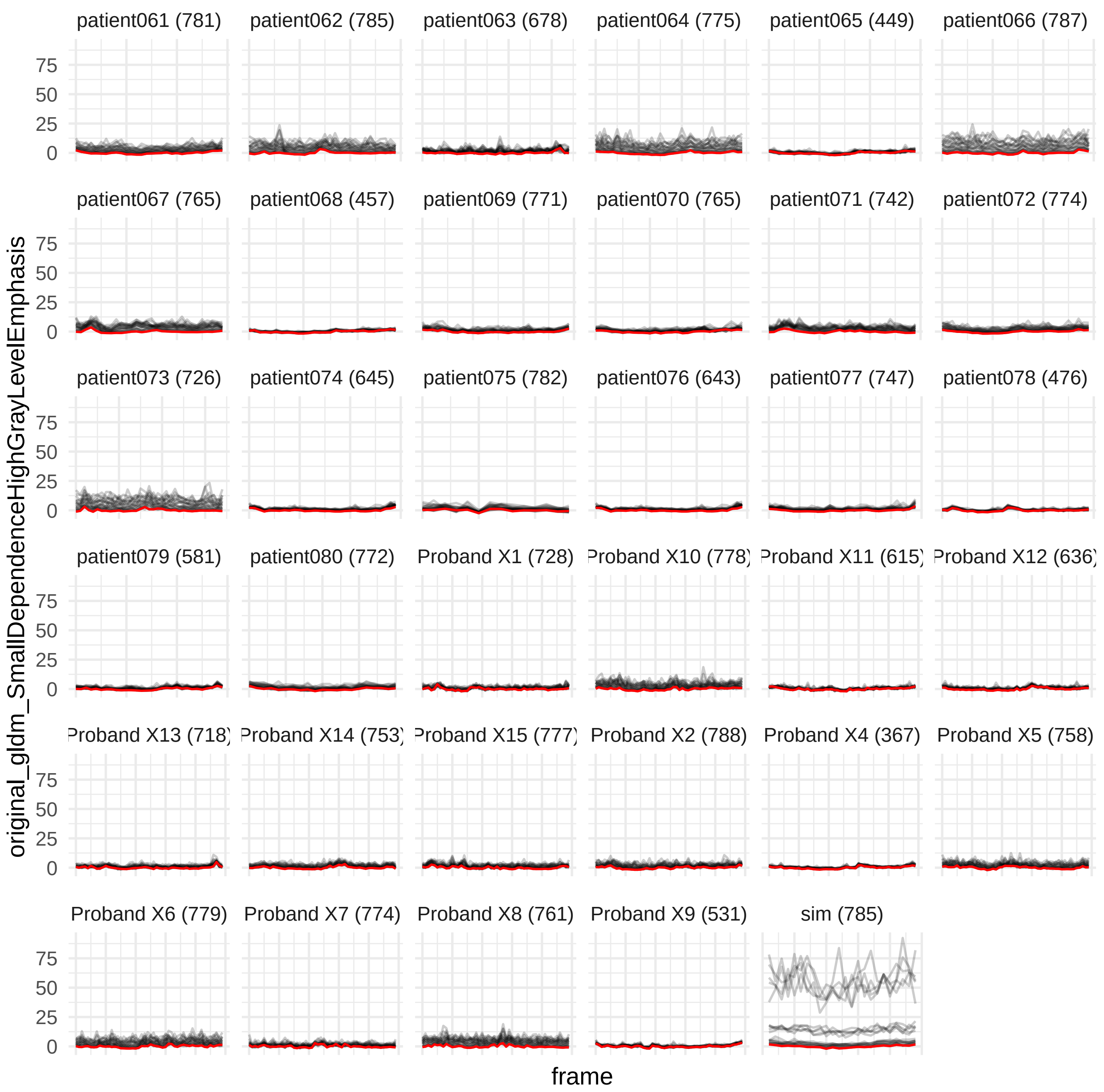

patient061 (781)
patient062 (785)
patient063 (678)
patient064 (775)
patient065 (449)
patient066 (787)
patient067 (765)
patient068 (457)
patient069 (771)
patient070 (765)
patient071 (742)
patient072 (774)
patient073 (726)
patient074 (645)
patient075 (782)
patient076 (643)
patient077 (747)
patient078 (476)
patient079 (581)
patient080 (772)
Proband X1 (728)
Proband X10 (778)
Proband X11 (615)
Proband X12 (636)
Proband X13 (718)
Proband X14 (753)
Proband X15 (777)
Proband X2 (788)
Proband X4 (367)
Proband X5 (758)
Proband X6 (779)
Proband X7 (774)
Proband X8 (761)
Proband X9 (531)
sim (785)
75
50
25
0
original_gldm_SmallDependenceHighGrayLevelEmphasis
frame

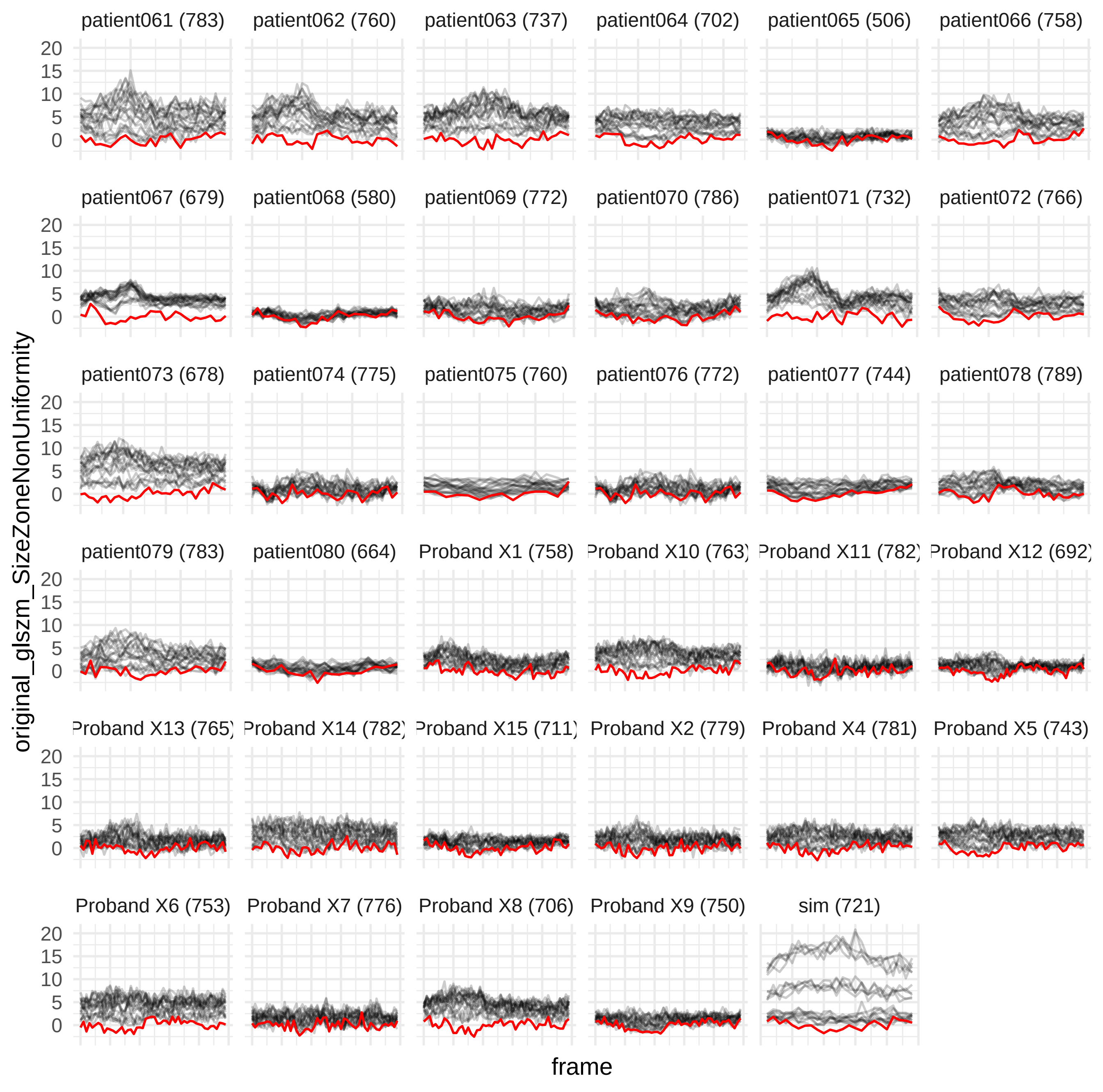
patient061 (783)
patient062 (760)
patient063 (737)
patient064 (702)
patient065 (506)
patient066 (758)
patient067 (679)
patient068 (580)
patient069 (772)
patient070 (786)
patient071 (732)
patient072 (766)
patient073 (678)
patient074 (775)
patient075 (760)
patient076 (772)
patient077 (744)
patient078 (789)
patient079 (783)
patient080 (664)
Proband X1 (758)
Proband X10 (763)
Proband X11 (782)
Proband X12 (692)
Proband X13 (765)
Proband X14 (782)
Proband X15 (711)
Proband X2 (779)
Proband X4 (781)
Proband X5 (743)
Proband X6 (753)
Proband X7 (776)
Proband X8 (706)
Proband X9 (750)
sim (721)
20
15
10
5
0
original_glszm_SizeZoneNonUniformity
frame

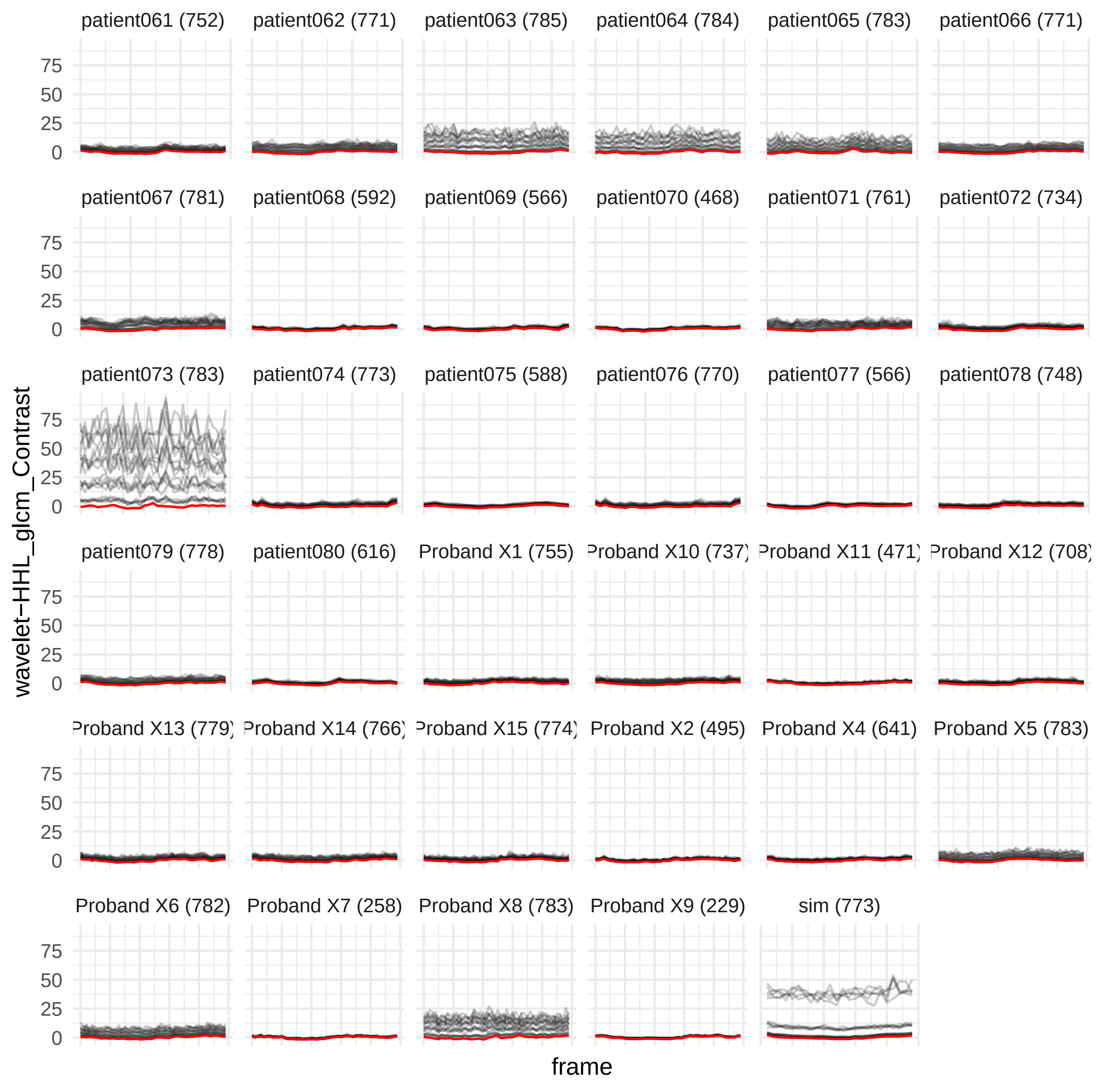

patient061 (752)
patient062 (771)
patient063 (785)
patient064 (784)
patient065 (783)
patient066 (771)
patient067 (781)
patient068 (592)
patient069 (566)
patient070 (468)
patient071 (761)
patient072 (734)
patient073 (783)
patient074 (773)
patient075 (588)
patient076 (770)
patient077 (566)
patient078 (748)
patient079 (778)
patient080 (616)
Proband X1 (755)
Proband X10 (737)
Proband X11 (471)
Proband X12 (708)
Proband X13 (779)
Proband X14 (766)
Proband X15 (774)
Proband X2 (495)
Proband X4 (641)
Proband X5 (783)
Proband X6 (782)
Proband X7 (258)
Proband X8 (783)
Proband X9 (229)
sim (773)
0
25
50
75
wavelet−HHL_glcm_Contrast
frame

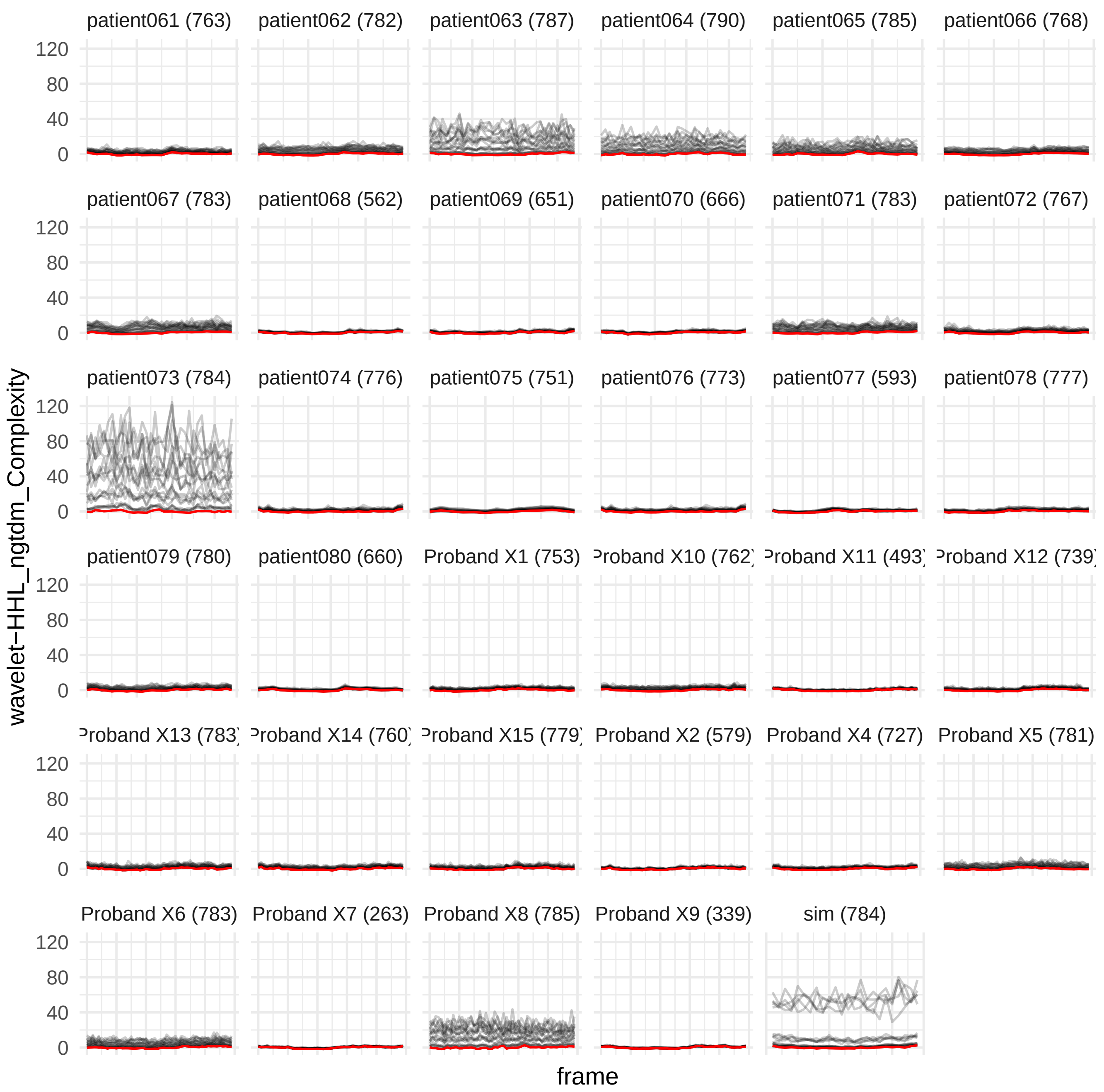

patient061 (763)
patient062 (782)
patient063 (787)
patient064 (790)
patient065 (785)
patient066 (768)
patient067 (783)
patient068 (562)
patient069 (651)
patient070 (666)
patient071 (783)
patient072 (767)
patient073 (784)
patient074 (776)
patient075 (751)
patient076 (773)
patient077 (593)
patient078 (777)
patient079 (780)
patient080 (660)
Proband X1 (753)
Proband X10 (762)
Proband X11 (493)
Proband X12 (739)
Proband X13 (783)
Proband X14 (760)
Proband X15 (779)
Proband X2 (579)
Proband X4 (727)
Proband X5 (781)
Proband X6 (783)
Proband X7 (263)
Proband X8 (785)
Proband X9 (339)
sim (784)
wavelet−HHL_ngtdm_Complexity
frame
0
40
80
120

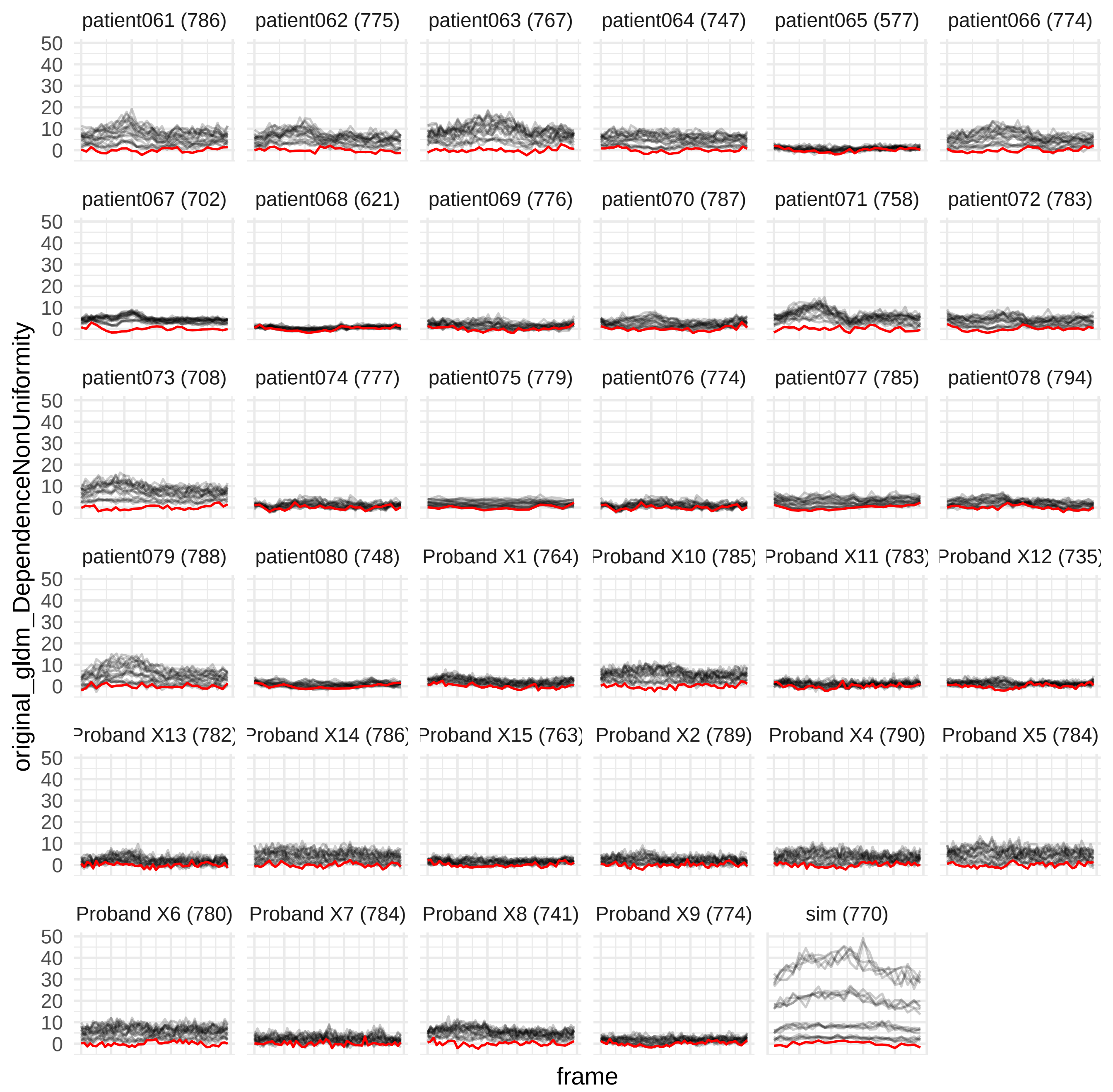
patient061 (786)
patient062 (775)
patient063 (767)
patient064 (747)
patient065 (577)
patient066 (774)
patient067 (702)
patient068 (621)
patient069 (776)
patient070 (787)
patient071 (758)
patient072 (783)
patient073 (708)
patient074 (777)
patient075 (779)
patient076 (774)
patient077 (785)
patient078 (794)
patient079 (788)
patient080 (748)
Proband X1 (764)
Proband X10 (785)
Proband X11 (783)
Proband X12 (735)
Proband X13 (782)
Proband X14 (786)
Proband X15 (763)
Proband X2 (789)
Proband X4 (790)
Proband X5 (784)
Proband X6 (780)
Proband X7 (784)
Proband X8 (741)
Proband X9 (774)
sim (770)
original_gldm_DependenceNonUniformity
frame
0
10
20
30
40
50
